# Differential contagiousness of respiratory disease across the United States

**DOI:** 10.1101/2022.09.15.22279948

**Authors:** Abhishek Mallela, Yen Ting Lin, William S. Hlavacek

## Abstract

The initial contagiousness of a communicable disease within a given population is quantified by the basic reproduction number, denoted *R*_0_. The value of *R*_0_ gives the expected number of new cases generated by an infectious person in a wholly susceptible population and depends on both pathogen and population properties. On the basis of compartmental models that reproduce Coronavirus Disease 2019 (COVID-19) surveillance data, we estimated region-specific *R*_0_ values for 280 of 384 metropolitan statistical areas (MSAs) in the United States (US), which account for 95% of the US population living in urban areas and 82% of the total population. Our estimates range from 1.9 to 7.7 and quantify the relative susceptibilities of regional populations to spread of respiratory diseases.

**One-Sentence Summary:** Initial contagiousness of Coronavirus Disease 2019 varied over a 4-fold range across urban areas of the United States.

LA-UR-22-29514

## Main Text

Public health surveillance efforts in the US during the COVID-19 pandemic were sweeping. In 2020 alone, approximately 254 million diagnostic tests for Severe Acute Respiratory Syndrome Coronavirus 2 (SARS-CoV-2) infection were administered *(1)*. The data generated from these surveillance efforts provide an unprecedented opportunity to gain insights into disease transmission dynamics in the US, especially for respiratory diseases similar to COVID-19, which is believed to be aerosol-transmitted *(2)*. To gain insights into the heterogeneity of disease transmission across the US, we attempted to use COVID-19 surveillance data, namely daily county-level reports of new cases, which have been collected in various repositories *(3, 4)*, to estimate the basic reproduction number of COVID-19 for as many distinct geographical regions in the US as possible.

The basic reproduction number, *R*_0_, is a dimensionless quantity corresponding to the expected number of secondary cases generated by an index case in a naïve population *(5)*. Although the contagiousness of a communicable disease is commonly characterized by an *R*_0_ estimate *(6)*, the value of *R*_0_ is in fact both pathogen- and population-specific *(7)*, meaning that it is a function of not only pathogen properties, including virulence factors, but also of population properties, including biological, sociobehavioral, and environmental factors *(8)*. The population properties that influence *R*_0_ are generally unknown.

Because many regions in the US were impacted by a common pathogen (i.e., SARS-CoV-2) around roughly the same time at the beginning of the COVID-19 pandemic *(9)*, we reasoned that a comparison of *R*_0_ estimates for COVID-19 in different regions would elucidate how the population properties of distinct regions combine to determine differential susceptibility to disease spread for COVID-19 and similar diseases. Thus, we undertook an effort to generate regional COVID-19 *R*_0_ estimates.

Estimation of *R*_0_ can be pursued in multiple ways *(10)*. Here, we adopted the approach of deriving *R*_0_ for a region of interest on the basis of a compartmental model parameterized for consistency with region-specific surveillance data *(11)*. This approach requires a tractable explanatory model *(12)*. An assumption of any compartmental model for an epidemic is that the population being considered is homogeneous *(13)*. With this limitation in mind, the population considered in a model should be carefully chosen: a population that is more uniformly impacted by an epidemic is a better choice for modeling.

We considered developing models for the populations of either states or metropolitan statistical areas (MSAs), the latter of which are delimited by the federal government on the basis of socioeconomic ties after each census *(14)*. In the US, there are currently 384 MSAs *(14)*, each encompassing a city with 50,000 or more inhabitants and surrounding communities. In the US, surveillance data are collected and reported by county-level public health authorities (or authorities of comparable jurisdictions) *(15)*, but county-level data can be aggregated to characterize disease transmission in MSAs and states. We did not develop models for county populations for two reasons. First, county-level surveillance data tend to be noisy, with noise arising from the stochastic nature of case detection combined with a small number of cases and/or because of irregularities in reporting. It is for these reasons that county-level data are commonly aggregated in epidemiological analyses *(16)*. Second, inspection of infection-risk curves suggested that counties within MSAs are similar, as illustrated in Fig. 1, which shows risk curves for four multi-county MSAs. Risk curves for the other multi-county MSAs are shown in Fig. S1.

**Figure 1.**
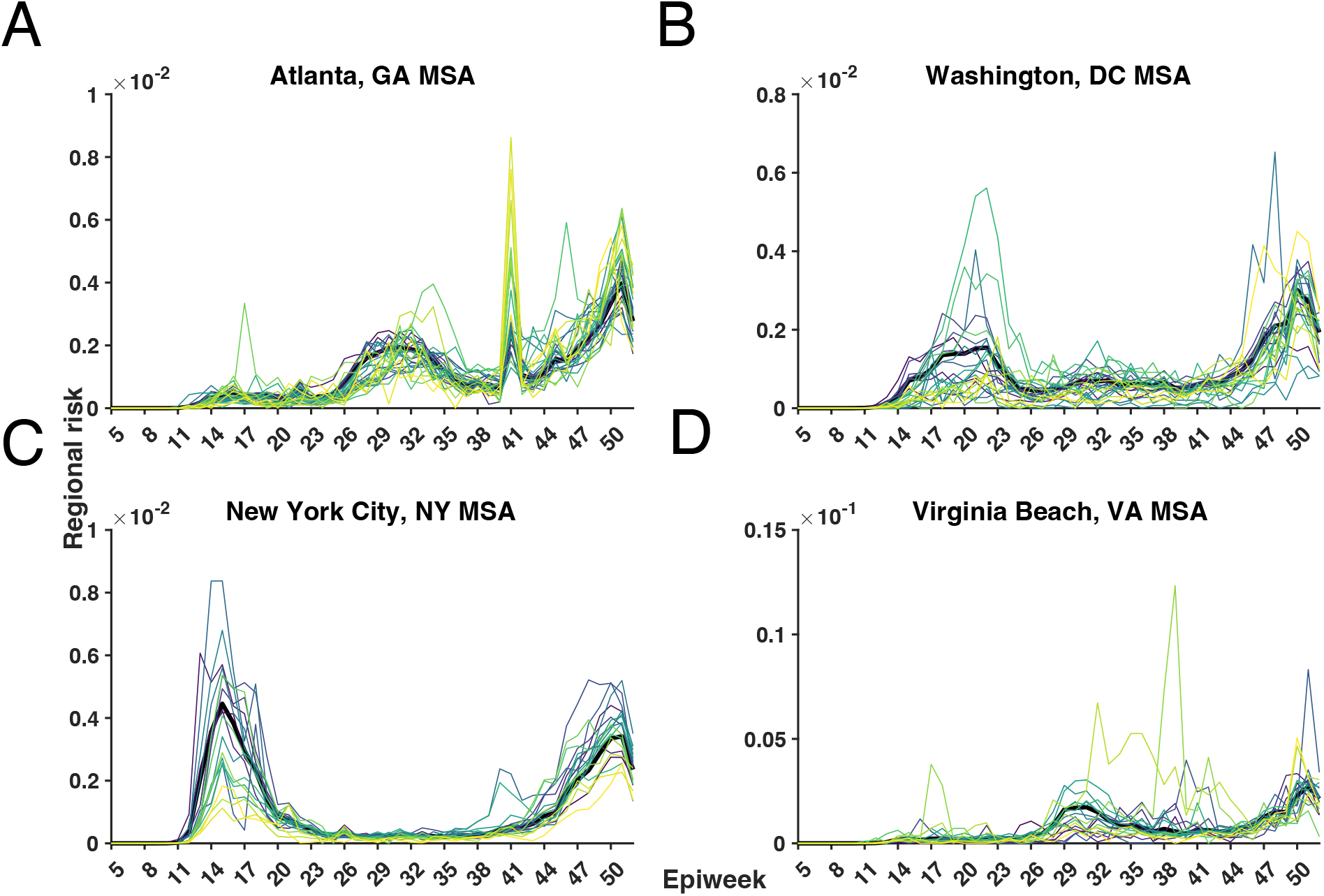
County- and MSA-level infection-risk curves for weekly-aggregated data in the MSAs encompassing (A) Atlanta, GA; (B) Washington, DC; (C) New York City, NY; and (D) Virginia Beach, VA from 26-January-2020 to 26-December-2020 (inclusive dates). Risk for a time period of interest in a given region is defined as the population fraction of new cases reported. MSA-level risk curves are shown in black and county-level risk curves are shown with a viridis color scheme. The four MSAs were chosen as the MSAs with the most counties. Atlanta, GA has 29 counties; Washington, DC has 25 counties or county equivalents; New York City, NY has 23 counties or county equivalents; and Virginia Beach, VA has 19 counties or county equivalents. States are indicated using two-letter US postal service abbreviations (https://about.usps.com/who-we-are/postal-history/state-abbreviations.pdf).

To ascertain whether disease transmission is more homogeneous across the counties of MSAs or states, we defined three different measures for variability in weekly disease incidence (i.e., infection risk over a 1-week period) across the counties of a given region (see Supplemental Methods). These measures are based on the Fano factor *(17)*, the Gini coefficient *(18)*, and the Wasserstein-1 distance *(19)*. We calculated variability measures for epidemiological weeks 5 through 52, a period starting on 26-January-2020 and ending on 26-December-2020, for all multi-county MSAs and the 50 states by aggregating confirmed COVID-19 daily county-level case-count data available in the GitHub repository maintained by *The New York Times* newspaper *(3)*.

Representative results of our risk variability analysis are shown in Fig. 2. In Fig. 2A, the Fano factor, Gini coefficient, and Wasserstein-1 distance variability measures are plotted as a function of epidemiological week for each of three selected MSAs and also for overlapping states. As can be seen, each variability measure tends to be less for an MSA than for an overlapping state. Results for other multi-county MSAs and overlapping states are shown in Figs. S2–S4. Histograms of time-averaged Fano factor, Gini coefficient, and Wasserstein-1 distance variability measures obtained for all multi-county MSAs and states are shown in Figs. 2B–2D. For each variability measure, the histogram for MSAs is shifted to the left of the histogram for states. These results indicate that county-level infection risks are more homogeneous for counties within MSAs than for counties within states. For this reason, we focused on developing models for MSA populations instead of state populations.

**Figure 2.**
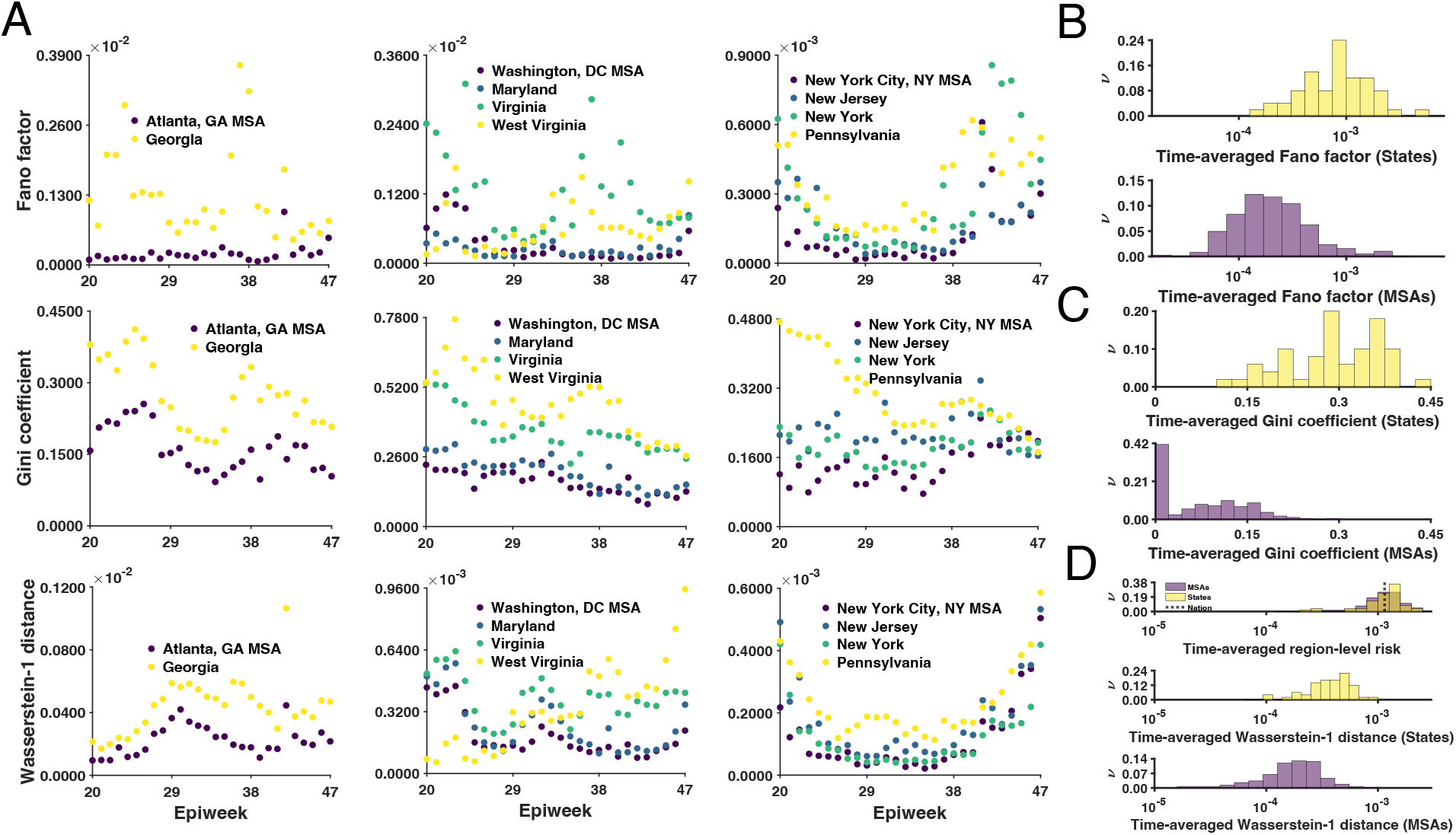
Counties in a given MSA are more homogeneous than counties in an overlapping state. (A) Plots of three infection-risk variability measures as a function of epidemiological week for three selected MSAs. The first, second, and third rows correspond to variability measures based on the Fano factor, Gini coefficient, and Wasserstein-1 distance, respectively. The first, second and third columns correspond to the MSAs encompassing Atlanta, GA; Washington, DC; and New York City, NY, respectively. (B) Time-averaged probability distributions of the Fano factor variability measure for states (top panel) and MSAs (bottom panel). (C) Time-averaged probability distributions of the Gini coefficient variability measure for states (top panel) and MSAs (bottom panel). (D) Time-averaged probability distributions of nation-, state-, and MSA-level infection risks (top panel), and time-averaged probability distributions of Wasserstein-1 distance variability measure for states (middle panel) and MSAs (bottom panel). In the panels of B, C, and D, the y-axis indicates relative frequency (*ν*).

In earlier work, we developed a compartmental model that is able to reproduce daily COVID-19 case count data for the 15 most populous MSAs in the US and all 50 states *(11, 20)*. Here, we found region-specific parameterizations of this model consistent with surveillance data for MSAs in the US having more than 200 cumulative cases reported before 21-May-2020 and at least 5 new cases on any given day between 21-January-2020 and 21-June-2020. 280 MSAs satisfy these criteria. For each of these MSAs, we applied a Bayesian inference approach described earlier *(11, 20)*, which is enabled by an adaptive Markov chain Monte Carlo (MCMC) sampling procedure. Inference job setup files for PyBioNetFit *(21)*, including files with MSA-specific surveillance data, are provided for each of the 280 MSAs (Data S1). To ensure that MCMC sampling converged, we visually inspected log-likelihood trace plots, parameter trace plots, and pairs plots. In addition, to ensure that regional parameterizations are explanatory, we compared posterior predictive distributions for case detection against daily counts of new cases (https://github.com/lanl/COVID-19-basic-reproduction-numbers). As illustrated in Fig. 3 for four selected MSAs, we were able to find parameterizations that are consistent with regional surveillance data.

**Figure 3.**
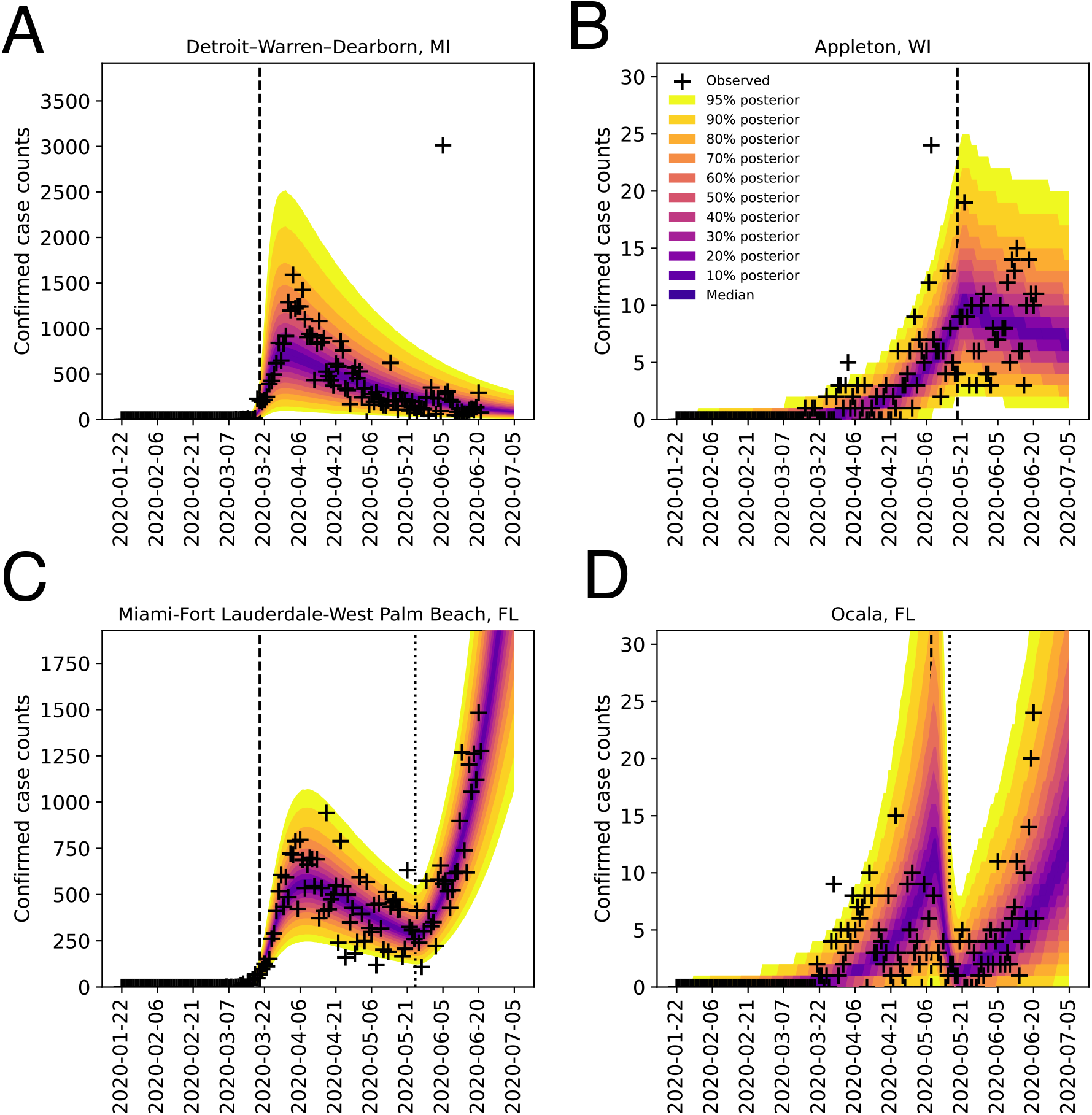
Bayesian posterior predictive distributions for daily confirmed COVID-19 case counts in the MSAs encompassing (A) Detroit, MI; (B) Appleton, WI; (C) Miami, FL; and (D) Ocala, FL from 21-January-2020 to 5-July-2020 (inclusive dates). The compartmental model used in analysis accounts for an initial period of social-distancing/nonpharmaceutical interventions (NPIs) (during which a fraction of the population adopts disease-avoiding behaviors and the remaining fraction mixes freely without taking special precautions to prevent infection) followed by *n* additional periods *(11)*. We considered *n* = 0, 1 and 2 and selected the best *n* using a model-selection procedure described previously *(11)*. Plus signs indicate daily case reports up to 21-June-2020, which were used in inference. The entire shaded region indicates a 95% credible interval accounting for prediction uncertainty arising from uncertainty in parameter estimates and inferred noise in detection of new cases. The color-coded bands within the shaded region indicate the median and credible intervals as indicated in the legend. In each panel, the vertical broken line indicates the onset time of the first social-distancing/NPI period. For MSAs with *n* = 1 (Miami and Ocala), there is an additional vertical dotted line, which indicates the onset time of the second social-distancing/NPI period.

For the compartmental model used in our analysis, we previously used the next-generation matrix method *(12)* to derive a formula that gives *R*_0_ in terms of model parameters *(20)*. According to this formula, the value of *R*_0_ depends on one inferred region-specific parameter, the contact rate parameter *β*, and seven fixed parameters describing within host-dynamics, which were estimated earlier and are expected to be universally applicable across regions of interest *(11)*. Using our earlier estimates for fixed parameters *(11)*, region-specific samples of the parametric posterior distribution for *β* obtained in Bayesian inference, and the formula for *R*_0_ *(20)*, for each of 280 MSAs, we found a maximum *a posteriori* (MAP) estimate for *R*_0_, which is equivalent to a maximum likelihood estimate because of the use of a uniform proper prior in Bayesian inference, and a 95% credible interval (Table S1). Similarly, through numerical solution of an equation obtained earlier *(20)*, for the same MSAs, we obtained a MAP estimate for the initial epidemic growth rate *λ* and a 95% credible interval (Table S1). Importantly, growth rate estimates are consistent with early surveillance data (https://github.com/lanl/COVID-19-basic-reproduction-numbers).

In Fig. 4, the *R*_0_ MAP estimates and 95% credible intervals that we obtained for 280 MSAs are presented in a rank order plot. The largest *R*_0_ estimate was 7.7 (for the MSA encompassing Detroit, Michigan) and the smallest was 1.9 (for the MSA encompassing Appleton, Wisconsin). These disparate estimates indicate that the population features contributing to initial disease spread are geographically heterogeneous and combine (in an unknown way) to yield a distribution of initial COVID-19 contagiousness that is spread over a 4-fold range. The absolute *R*_0_ values reported here are COVID-19-specific; however, the relative strengths or ratios quantify relative susceptibilities to respiratory disease spread arising from MSA-specific population features.

**Figure 4.**
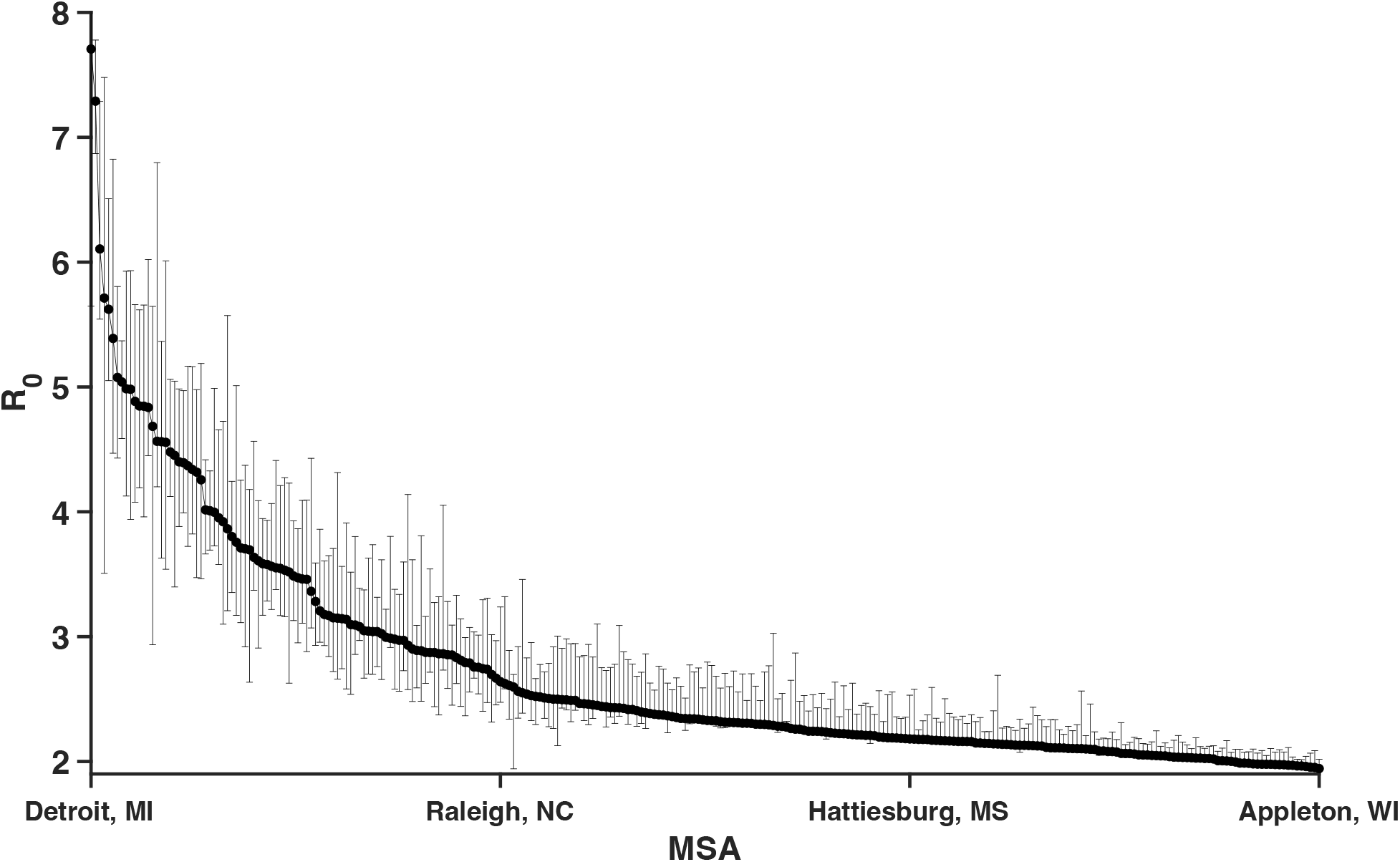
MAP estimates of the COVID-19 basic reproduction number *R*_0_ for the strains of SARS-CoV-2 emerging in 280 MSAs in 2020. MSA-specific estimates of *R*_0_ are sorted by MSA from largest to smallest values according to the *R*_0_ estimates inferred from surveillance data collected between 21-January-2020 and 21-June-2020. The whiskers associated with each filled circle indicate the 95% credible interval.

Using a mechanistic compartmental model in concert with COVID-19 surveillance data and Bayesian inference, we have obtained MSA-specific COVID-19 *R*_0_ estimates that quantify differences in population properties across urban areas in the US that affect disease contagiousness. This information may help mitigate future respiratory disease outbreaks, as some urban areas are evidently far more susceptible to rapid disease spread than others. Our findings may be helpful in identifying which population properties contribute to contagiousness, which is important because the population properties underlying our *R*_0_ estimates may eventually change with time.

## Data Availability

All data produced in the present work are contained in the manuscript.

## Acknowledgments

Computational resources used in this study included the FARM cluster at the University of California, Davis.

## Funding

2020 National Science Foundation Mathematical Sciences Graduate Internship program (AM)

Center for Nonlinear Studies, Los Alamos National Laboratory (AM)

Laboratory Directed Research and Development program at Los Alamos National Laboratory (project 20220268ER) (WSH, YTL)

National Institute of General Medical Sciences of the National Institutes of Health (grant R01GM111510) (WSH)

## Author contributions

Conceptualization: AM, WSH

Methodology: AM, YTL,

WSH Investigation: AM, YTL, WSH

Visualization: AM, YTL, WSH

Funding acquisition: YTL, WSH

Project administration: YTL, WSH

Supervision: YTL, WSH

Writing – original draft: AM

Writing – review & editing: AM, YTL, WSH

## Data and materials availability

All data are available in the main text or the supplementary materials.

## Supplementary Materials

Materials and Methods

Figs. S1 to S4

Table S1

Data S1

References (*3, 11, 20, 21*)

## Supplementary Materials for

### Materials and Methods

Here, we derive the measures discussed in the Main Text for infection risk variability across counties of a specified region. These variability measures are based on the Gini coefficient, Wasserstein-1 distance, and Fano factor. Infection risk for a time period of interest in a given region is defined as the population fraction of new cases reported.

#### Gini coefficient risk variability measure

Let us consider a population of *n* persons. The *i*^*th*^ person has infection risk *r*_*i*_. We want to know two quantities. The first quantity is MD, the mean absolute difference in risk. We calculate MD as the absolute difference in risk averaged over all distinct pairs of persons. The sum of absolute differences is 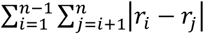, which equals 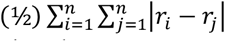. The number of distinct pairs of persons is 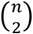, which is equal to *n*(*n* − 1)/2. Thus, MD is given by

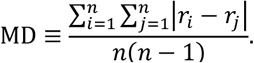

The second quantity of interest is AM, the arithmetic mean risk. AM is given by

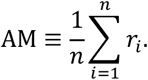

The ratio MD/AM is RMD, the relative mean absolute difference. RMD is given by

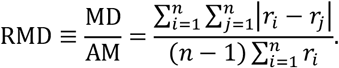

RMD is equal to twice the Gini coefficient, *G*. Thus *G* ≡ RMD/2 is given by

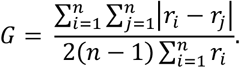

Let us consider the case where the *n* persons of interest reside in *k* ≤ *n* counties. All persons with the same county have the same risk. Let us denote the county-associated risks as *c*_1_, *c*_2_, …, *c*_*k*_. Let us use *n*_*i*_ to denote the population of the *i*^*th*^ county. It follows that 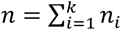. We want to calculate *G* in terms of county-level risks {*c*_1_, …, *c*_*k*_} instead of the individual-level risks {*r*_1_, …, *r*_*n*_}. The number of pairs of persons consisting of a person from county *I* and a person from a different county *j* ≠ *i* is given by *n*_*i*_*n*_j_. Thus,

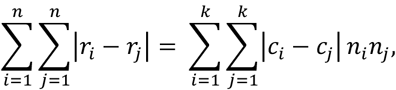

because persons in the same county share the same risk. We also have the following relation:

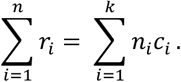

It follows that

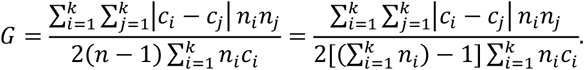

We can take *c*_*i*_ to be the COVID-19 incidence proportion for the *i*^*th*^ county over a given epidemiological week. If *n* ≫ 1,

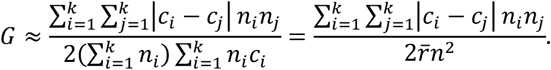

#### Wasserstein-1 distance risk variability measure

Let *X* denote a finite, non-empty, and ordered set. The Wasserstein-1 distance between two discrete distributions is given as *W*_1_(*p, q*) = ∑_*x*∈*X*_ |*F*_*p*_ (*x*) − *F*_*q*_ (*x*)| where *F*_*p*_(·) ∈ [0,1] and *F*_*q*_(·) ∈ [0,1] are the cumulative distribution functions of *p*(·) and *q*(·), respectively.

Let *k* denote the number of counties in a given region (i.e., an MSA or state) in a given epidemiological week. Let us consider the case where all persons within the same county have the same risk. Let us denote the county-associated risks as *c*_1_, *c*_2_, …, *c*_*k*_. Let us denote the population of the *i*^*th*^ county as *n*_*i*_. It follows that 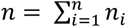, where *p* denotes the population of the aggregated region (i.e., an MSA or state.)

We take *c*_*i*_ to be the COVID-19 incidence proportion for the *i*^*th*^ county in a given epidemiological week. Let us denote our random variable by *C*, taking on the values *c*_1_, *c*_2_, …, *c*_*k*_. After separately sorting the values *c*_*i*_ and the populations (or weights) *p*_*i*_ in ascending order, it follows that 0 ≤ *c*_1_ < *c*_2_ < … < *c*_*k*_ ≤ 1 and 0 ≤ *p*_1_ < *p*_2_ < … < *p*_*k*_ ≤ *p*.

The weighted empirical cumulative distribution function is then constructed for a given MSA (or state) in a given epidemiological week, and is comprised of the values *c*_*i*_ and the normalized weights *p*_*i*_/*p*, ordered by the population (from small to large).

The reference empirical cumulative distribution function for a given MSA (or state), with averaged risk in a given epidemiological week, is comprised of the value 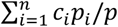 and the weight 1, with the value on the *x*-axis and the weight on the *y*-axis.

The Wasserstein-1 distance is computed between a given MSA (or state) and the corresponding reference distribution.

#### Fano factor risk variability measure

For a given region (i.e., MSA or state) and a given epidemiological week, the Fano factor is defined as the variance-to-mean ratio. The random variable of interest is given by the set of county-level risks, where the risk is defined as the confirmed number of new cases in a given week normalized by the county population.

#### Time-averaging

As presented in the Main Text (see Figs. 1B to 1D), we obtained average quantities for the infection risk variability measures above by averaging over the time domain (i.e., over the epidemiological weeks).

#### Bayesian inference

To infer region-specific values of adjustable model parameters (and *R*_1_ estimates), we followed the Bayesian inference approach of Lin et al. *(11)*. In inferences, we used all region-relevant confirmed COVID-19 case-count data available in the GitHub repository maintained by *The New York Times* newspaper *(3)* for the period starting on 21-January-2020 and ending on 21-June-2020 (inclusive dates). The first case in the US was reported on 21-January-2020. We focused on early surveillance data (vs. all available surveillance data up to the present time) so as to characterize COVID-19 transmission within populations that are nearly wholly susceptible. Markov Chain Monte Carlo (MCMC) sampling was performed using the Python code of Lin et al. *(11)* and a new release of PyBioNetFit *(21)*, version 1.1.9, which includes an implementation of the adaptive MCMC method used in the study of Lin et al. *(11)*. Inference job setup files for PyBioNetFit, including data files, are provided for each of 280 MSAs (Data S1). There are 384 MSAs in the US. We excluded 104 of 384 MSAs from analysis. In each of these cases, there was insufficient data to support parameterization; namely, each of these MSAs had fewer than 200 cumulative cases reported before 21-May-2020 or no more than 5 new cases reported on any given day between 21-January-2020 and 21-June-2020.

**Fig. S1.** County- and MSA-level infection-risk curves for weekly-aggregated data from 26-January-2020 to 26-December-2020 (inclusive dates) in each of 232 multi-county MSAs, except those encompassing Atlanta, GA; Washington, DC; New York City, NY; and Virginia Beach, VA, which are considered in Fig. 1 of the Main Text. MSA-level risk curves are shown in black and county-level risk curves are shown with a viridis color scheme. States are indicated using two-letter US postal service abbreviations.

**Fig. S2.** Plots of the infection-risk variability measures based on the Fano factor as a function of epidemiological week for each of 232 multi-county MSAs, except those encompassing Atlanta, GA; Washington, DC; and New York City, NY, which are considered in Fig. 2 of the Main Text.

**Fig. S3.** Plots of the infection-risk variability measures based on the Gini coefficient as a function of epidemiological week for each of 232 multi-county MSAs, except those encompassing Atlanta, GA; Washington, DC; and New York City, NY, which are considered in Fig. 2 of the Main Text.

**Fig. S4.** Plots of the infection-risk variability measures based on the Wasserstein-1 distance as a function of epidemiological week for each of 232 multi-county MSAs, except those encompassing Atlanta, GA; Washington, DC; and New York City, NY, which are considered in Fig. 2 of the Main Text.

## References and Notes

1) The COVID Tracking Project, “The Data: Totals for the US” (2022); https://covidtracking.com/data/national

2) N. Van Doremalen, T. Bushmaker, D. H. Morris, M. G. Holbrook, A. Gamble, B. N. Williamson, A. Tamin, J. L. Harcourt, N. J. Thornburg, S. I. Gerber, J. O. Lloyd-Smith, Aerosol and surface stability of SARS-CoV-2 as compared with SARS-CoV-1. N Engl J Med. 382, 1564–1567 (2020).

3) The New York Times COVID-19 Data Team, “Data from The New York Times.” (2020); https://github.com/nytimes/covid-19-data

4) Johns Hopkins University of Medicine, “Coronavirus Resource Center.” (2022); https://coronavirus.jhu.edu

5) P. van den Driessche, Reproduction numbers of infectious disease models. Infect Dis Model. 2, 288–303 (2017).

6) B. Ridenhour, J. M. Kowalik, D. K. Shay, Unraveling R_0_: Considerations for public health applications. Am J Public Health. 108, S445–S454 (2018).

7) H. E. Randolph, L. B. Barreiro, Herd Immunity: Understanding COVID-19. Immunity. 5, 737–741 (2020).

8) P. L. Delamater, E. J. Street, T. F. Leslie, Y. T. Yang, K. H. Jacobsen, Complexity of the basic reproduction number (R0). Emerg Infect Dis. 25, 1–4 (2019).

9) T. A. Perkins, S. M. Cavany, S. M. Moore, R. J. Oidtman, A. Lerch, M. Poterek, Estimating unobserved SARS-CoV-2 infections in the United States. Proc Natl Acad Sci. 117, 22597–22602 (2020).

10) C. T. Bauch, J. O. Lloyd-Smith, M. P. Coffee, A. P. Galvani, Dynamically modeling SARS and other newly emerging respiratory illnesses: past, present, and future. Epidemiology. 16, 791–801 (2005).

11) Y. T. Lin, J. Neumann, E. F. Miller, R. G. Posner, A. Mallela, C. Safta, J. Ray, G. Thakur, S. Chinthavali, W. S. Hlavacek, Daily forecasting of regional epidemics of Coronavirus Disease with Bayesian uncertainty quantification, United States. Emerg Infect Dis. 27, 767 (2021).

12) O. Diekmann, J. A. Heesterbeek, M. G. Roberts, The construction of next-generation matrices for compartmental epidemic models. J R Soc Interface. 7, 873–875 (2010).

13) M. J. Keeling, P. Rohani, Modeling Infectious Diseases in Humans and Animals (Princeton university Press, Princeton, NJ, 2008).

14) United States Census Bureau, “Metropolitan and Micropolitan.” (2022); https://www.census.gov/programs-surveys/metro-micro.html

15) Centers for Disease Control and Prevention, “National Notifiable Diseases Surveillance System (NNDSS): What is Case Surveillance?” (2022); https://www.cdc.gov/nndss/about/index.html

16) A. H. Auchincloss, S. Y. Gebreab, C. Mair, A. V. Diez Roux, A review of spatial methods in epidemiology, 2000–2010. Annu Rev Public Health. 33, 107–122 (2012).

17) A. A. Faisal, L. P. Selen, D. M. Wolpert, Noise in the nervous system. Nat Rev Neurosci. 9, 292–303 (2008).

18) L. Ceriani, P. Verme, The origins of the Gini index: extracts from Variabilità e Mutabilità (1912) by Corrado Gini. J Econ Inequal. 10, 421–443 (2012).

19) Y. Rubner, C. Tomasi, L. J. Guibas, The earth mover’s distance as a metric for image retrieval. Int J Comput Vis. 40, 99–121 (2000).

20) A. Mallela, J. Neumann, E. F. Miller, R. G. Posner, Y. T. Lin, W. S. Hlavacek, Bayesian inference of state-level COVID-19 basic reproduction numbers across the United States. Viruses. 14, 157 (2022).

21) J. Neumann, Y. T. Lin, A. Mallela, E. F. Miller, J. Colvin, A. T. Duprat, Y. Chen, W. S. Hlavacek, R. G. Posner, Implementation of a practical Markov chain Monte Carlo sampling algorithm in PyBioNetFit. Bioinformatics. 38, 1770–1772 (2022).

