## Supplementary material for "Differential contagiousness of respiratory disease across the United States": Figure S1

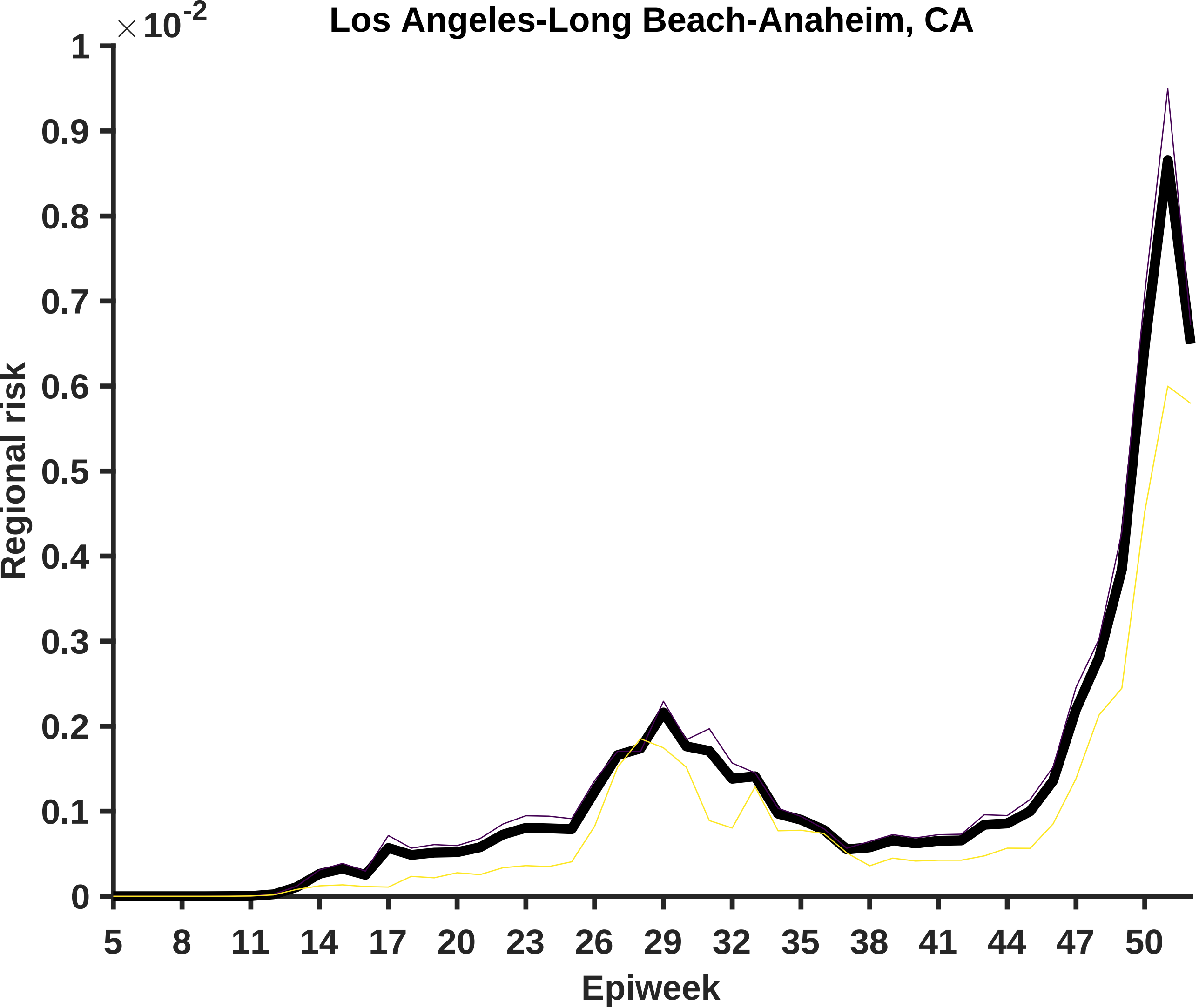

Los Angeles-Long Beach-Anaheim, CA
×10^-2
1
0.9
0.8
0.7
0.6
0.5
0.4
0.3
0.2
0.1
0
Regional risk
5
8
11
14
17
20
23
26
29
32
35
38
41
44
47
50
Epiweek

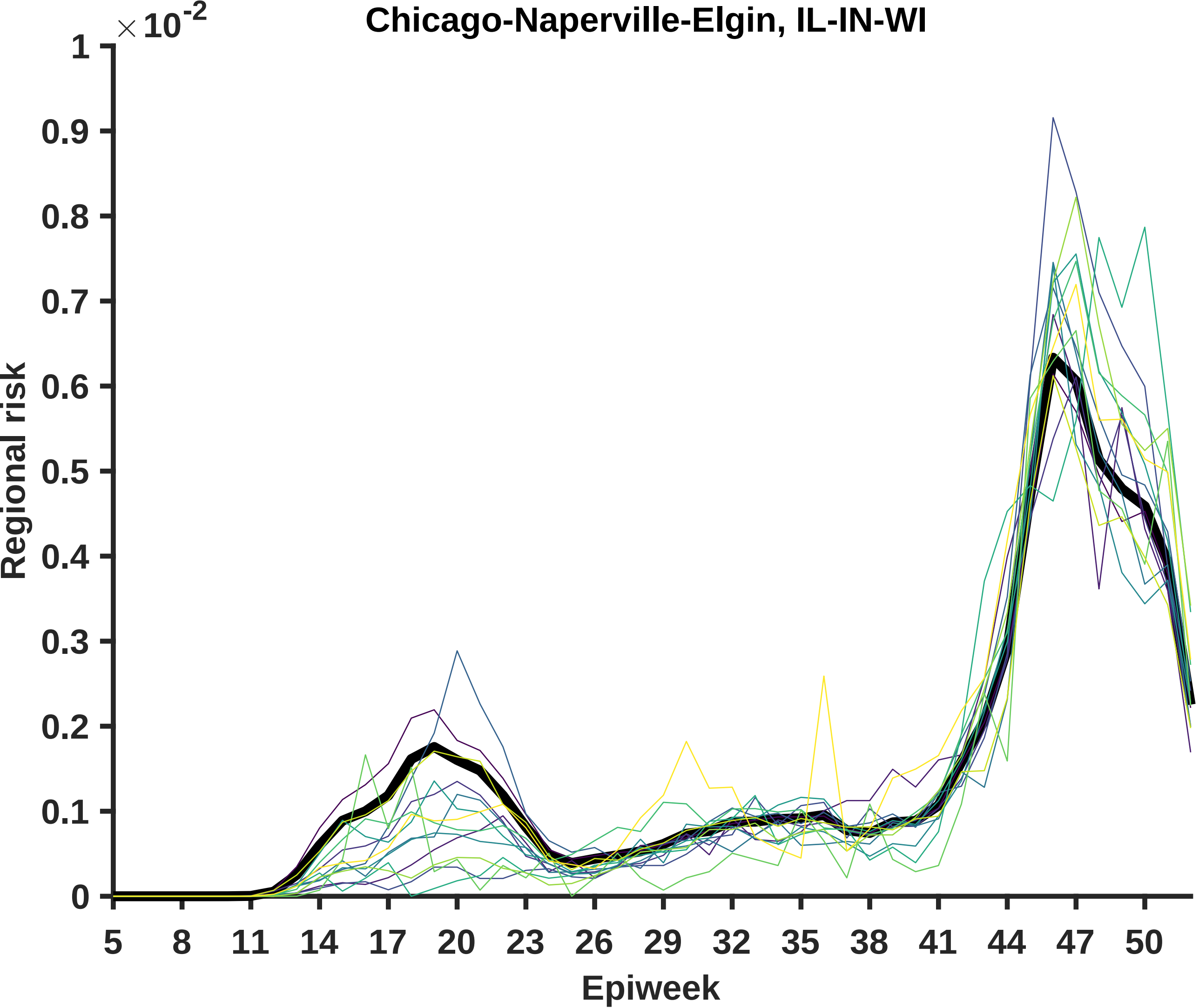

Chicago-Naperville-Elgin, IL-IN-WI
× 10^-2
1
0.9
0.8
0.7
0.6
0.5
0.4
0.3
0.2
0.1
0
Regional risk
5
8
11
14
17
20
23
26
29
32
35
38
41
44
47
50
Epiweek

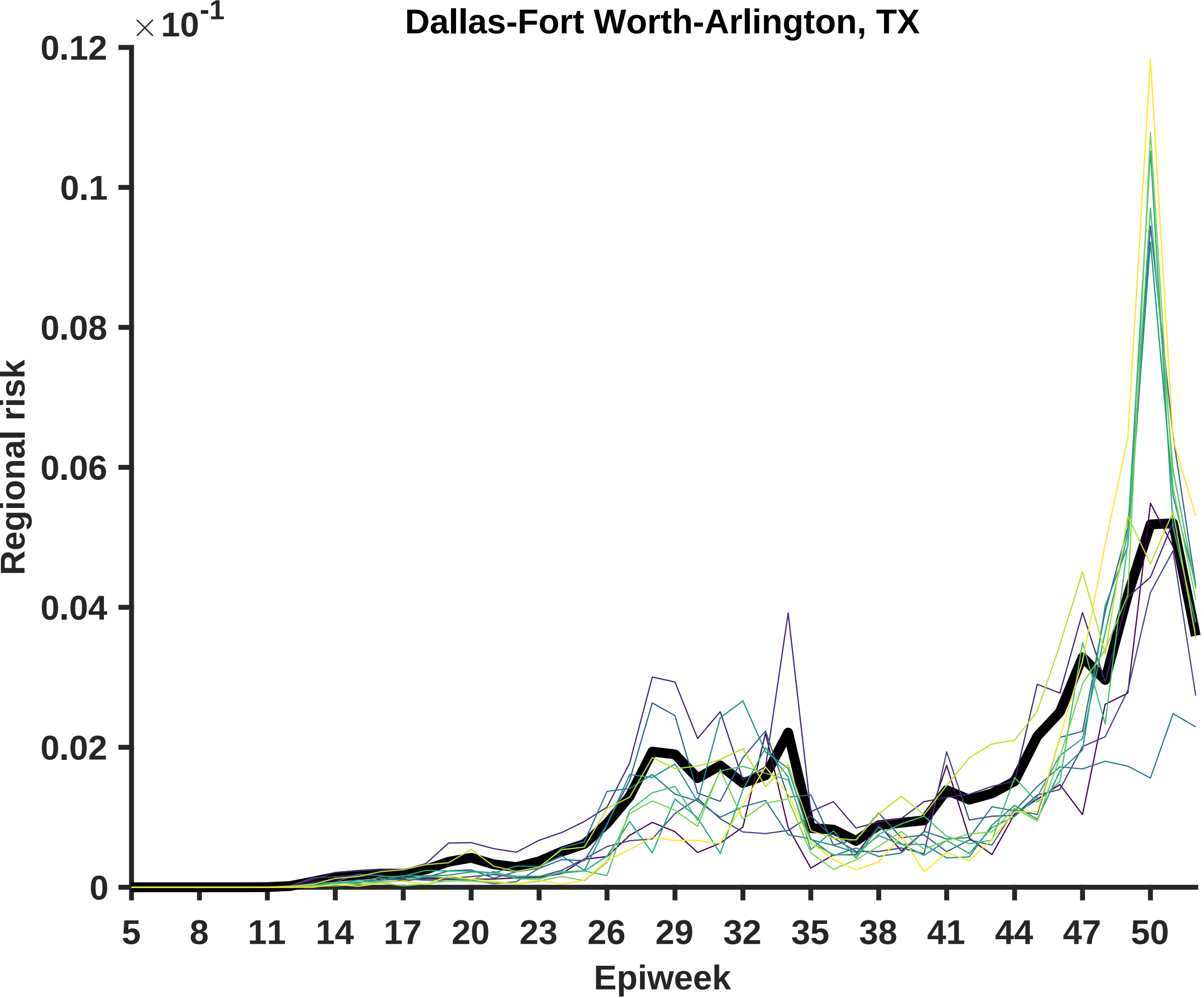
Dallas-Fort Worth-Arlington, TX
×10^-1
0.12
0.1
0.08
0.06
0.04
0.02
0
Regional risk
5
8
11
14
17
20
23
26
29
32
35
38
41
44
47
50
Epiweek

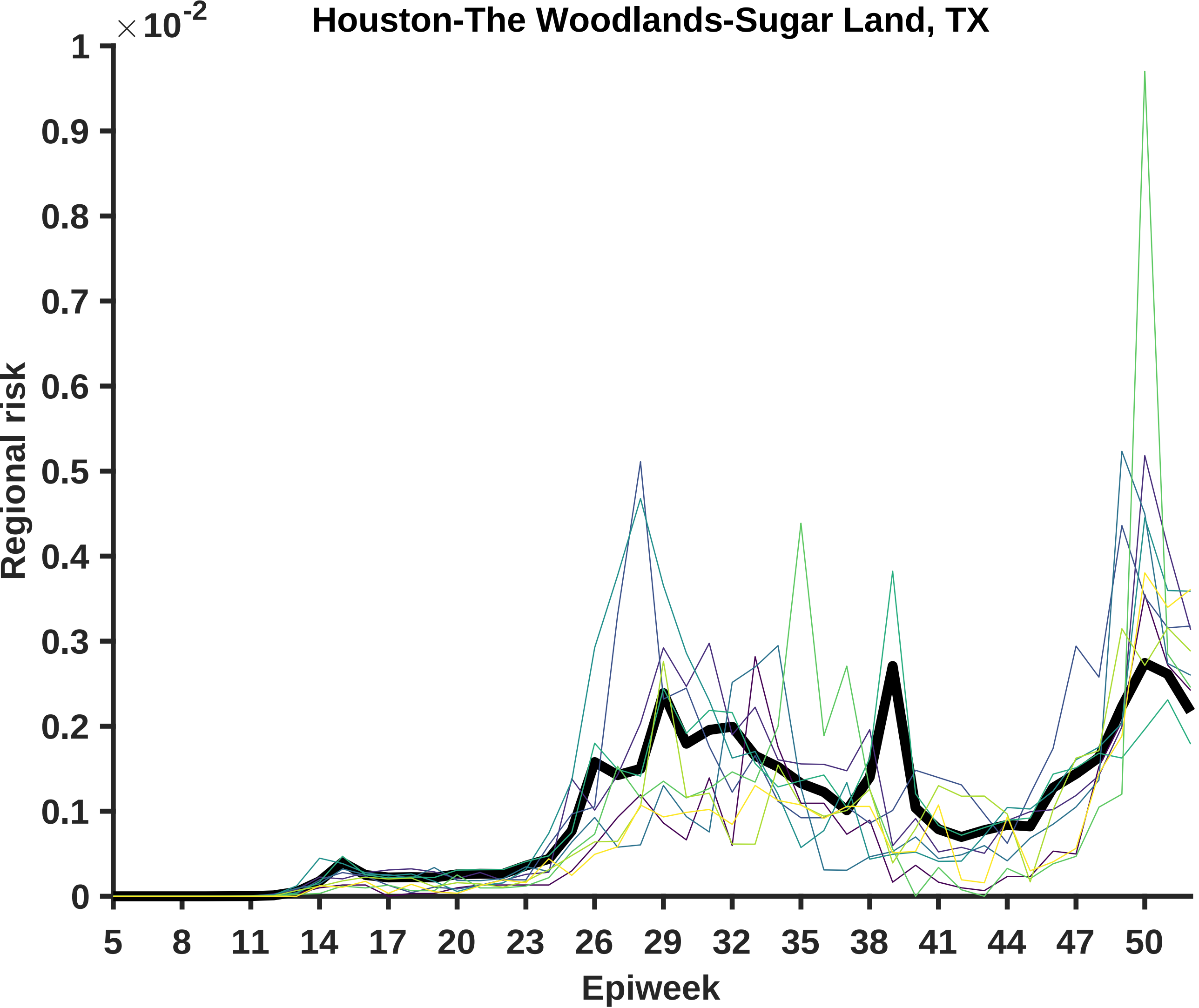
Houston-The Woodlands-Sugar Land, TX
$\times 10^{-2}$
Regional risk
0
0.1
0.2
0.3
0.4
0.5
0.6
0.7
0.8
0.9
1
5
8
11
14
17
20
23
26
29
32
35
38
41
44
47
50
Epiweek

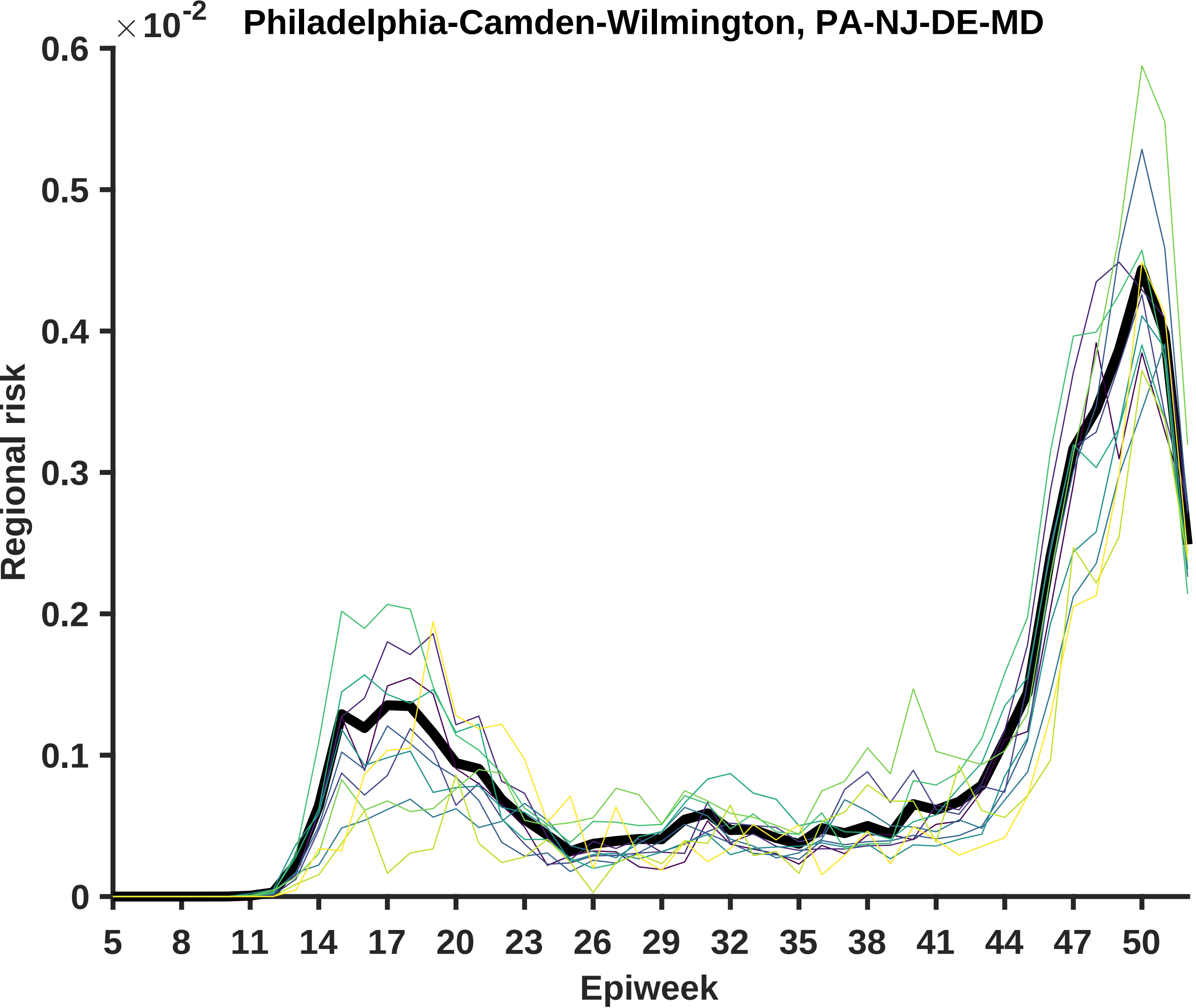

Philadelphia-Camden-Wilmington, PA-NJ-DE-MD
×10-2
0.6
0.5
0.4
0.3
0.2
0.1
0
Regional risk
5
8
11
14
17
20
23
26
29
32
35
38
41
44
47
50
Epiweek

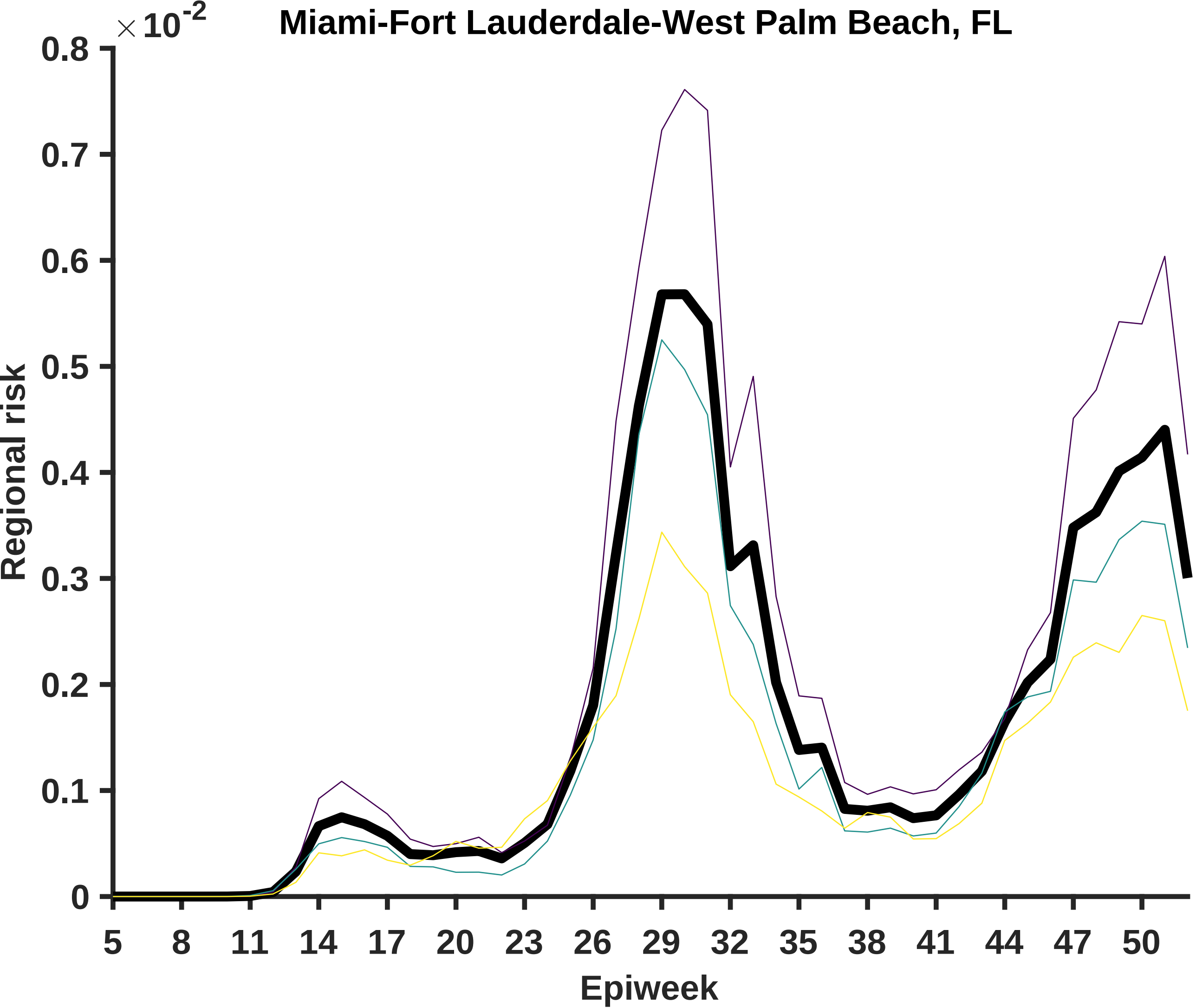

Miami-Fort Lauderdale-West Palm Beach, FL
$\times 10^{-2}$
0.8
0.7
0.6
0.5
0.4
0.3
0.2
0.1
0
Regional risk
5
8
11
14
17
20
23
26
29
32
35
38
41
44
47
50
Epiweek

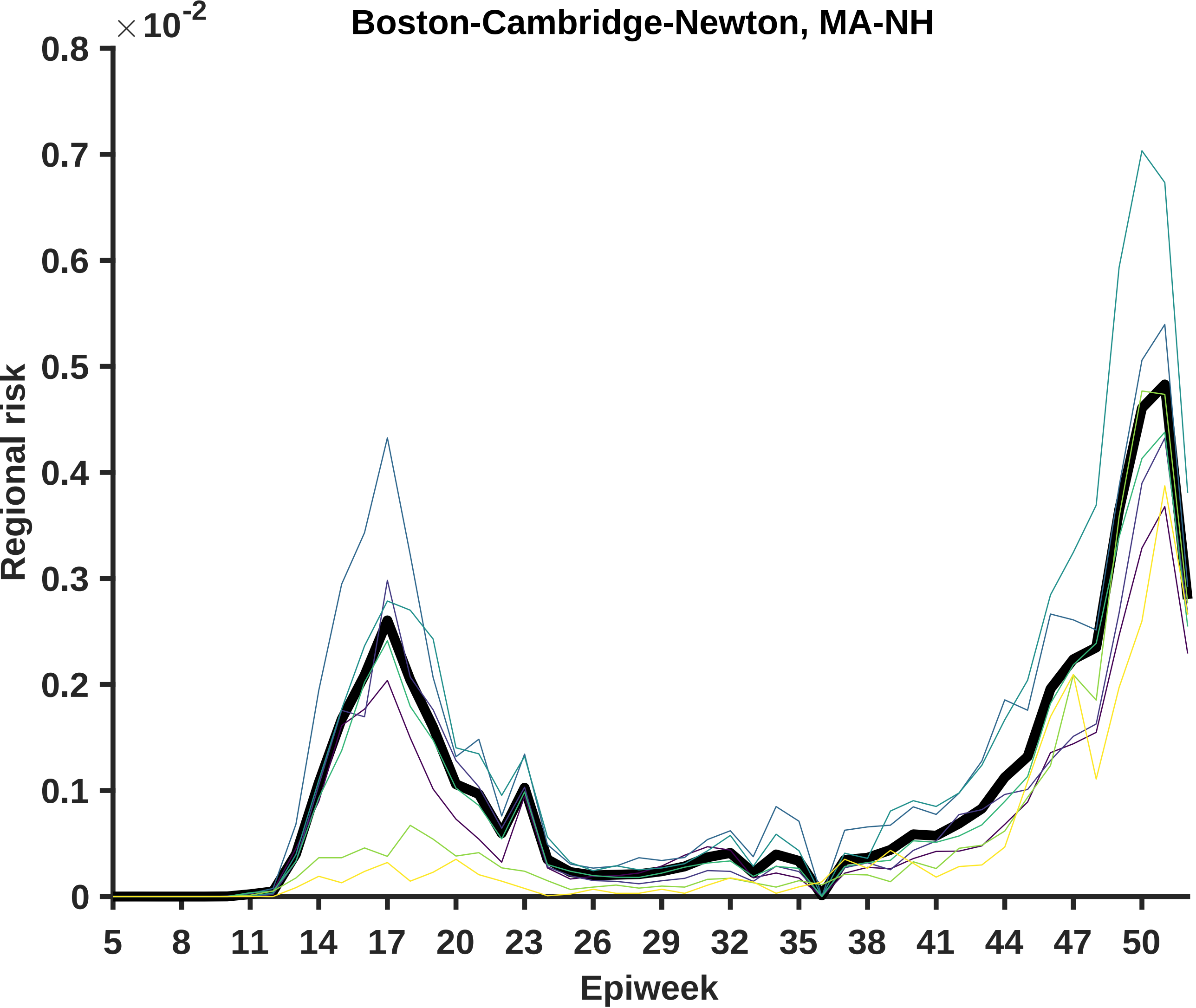
Boston-Cambridge-Newton, MA-NH
×10^-2
0.8
0.7
0.6
0.5
0.4
0.3
0.2
0.1
0
Regional risk
5
8
11
14
17
20
23
26
29
32
35
38
41
44
47
50
Epiweek

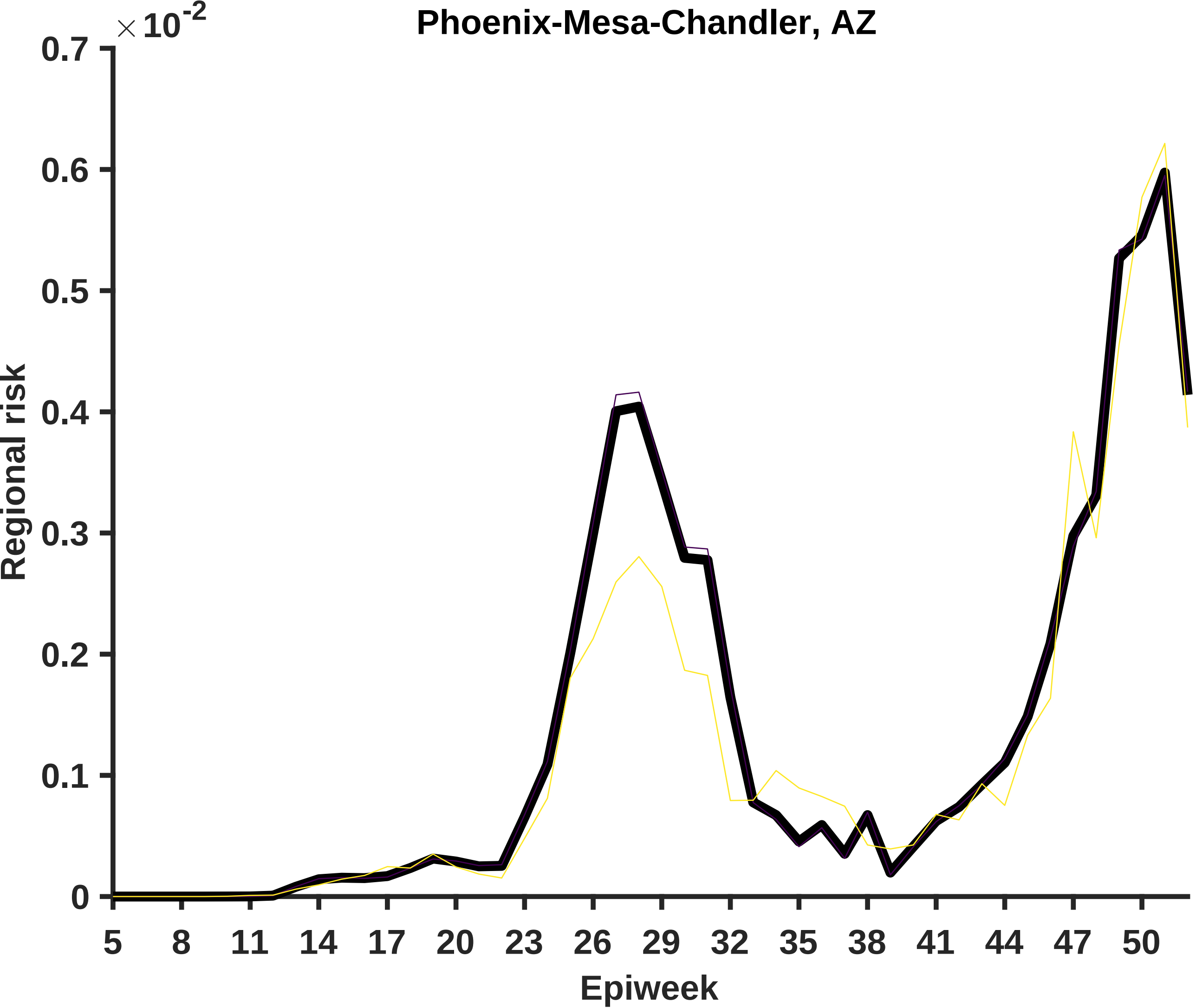

Phoenix-Mesa-Chandler, AZ
×10^-2
0.7
0.6
0.5
0.4
0.3
0.2
0.1
0
Regional risk
5
8
11
14
17
20
23
26
29
32
35
38
41
44
47
50
Epiweek

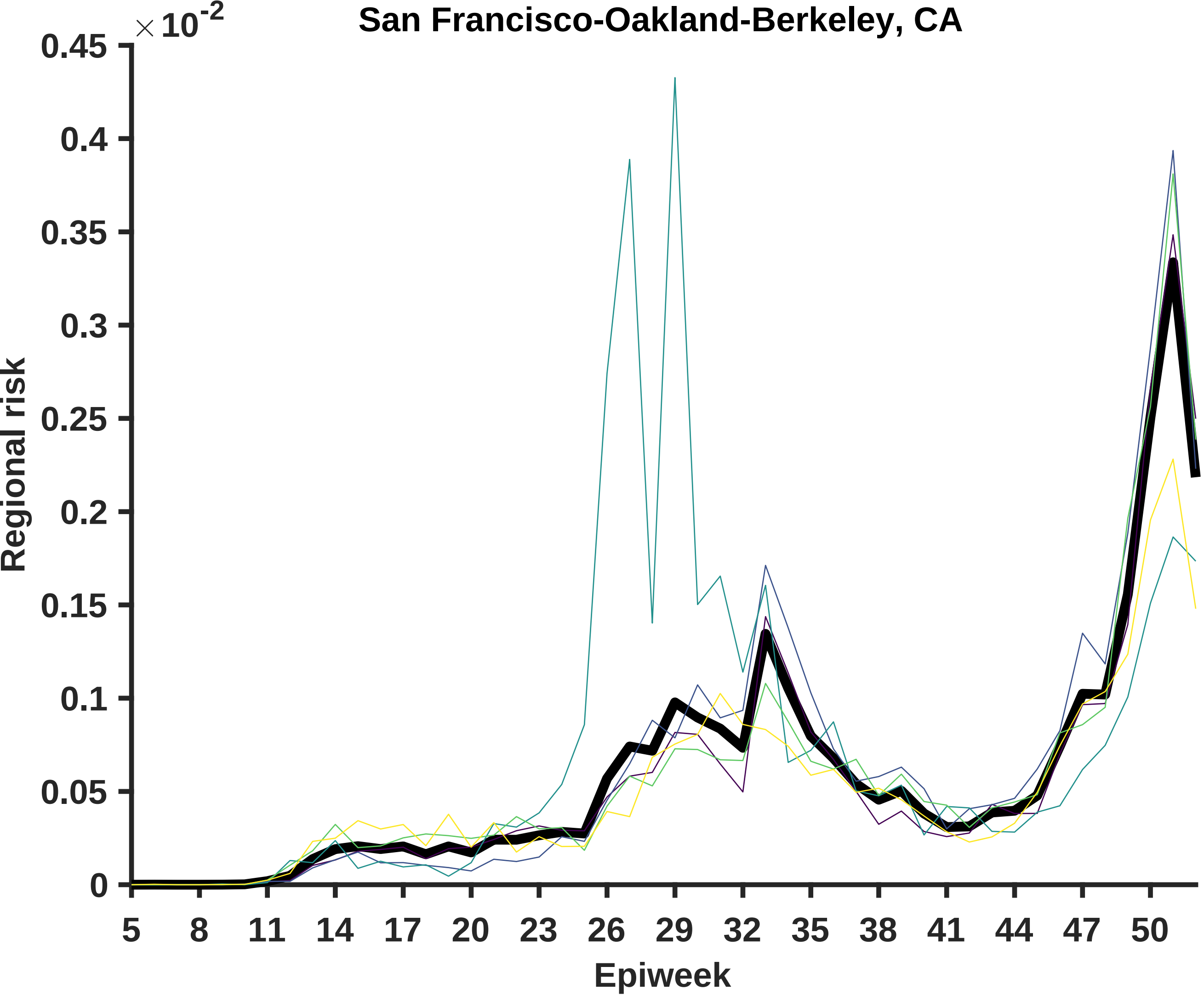

San Francisco-Oakland-Berkeley, CA
×10^-2
0.45
0.4
0.35
0.3
0.25
0.2
0.15
0.1
0.05
0
Regional risk
5
8
11
14
17
20
23
26
29
32
35
38
41
44
47
50
Epiweek

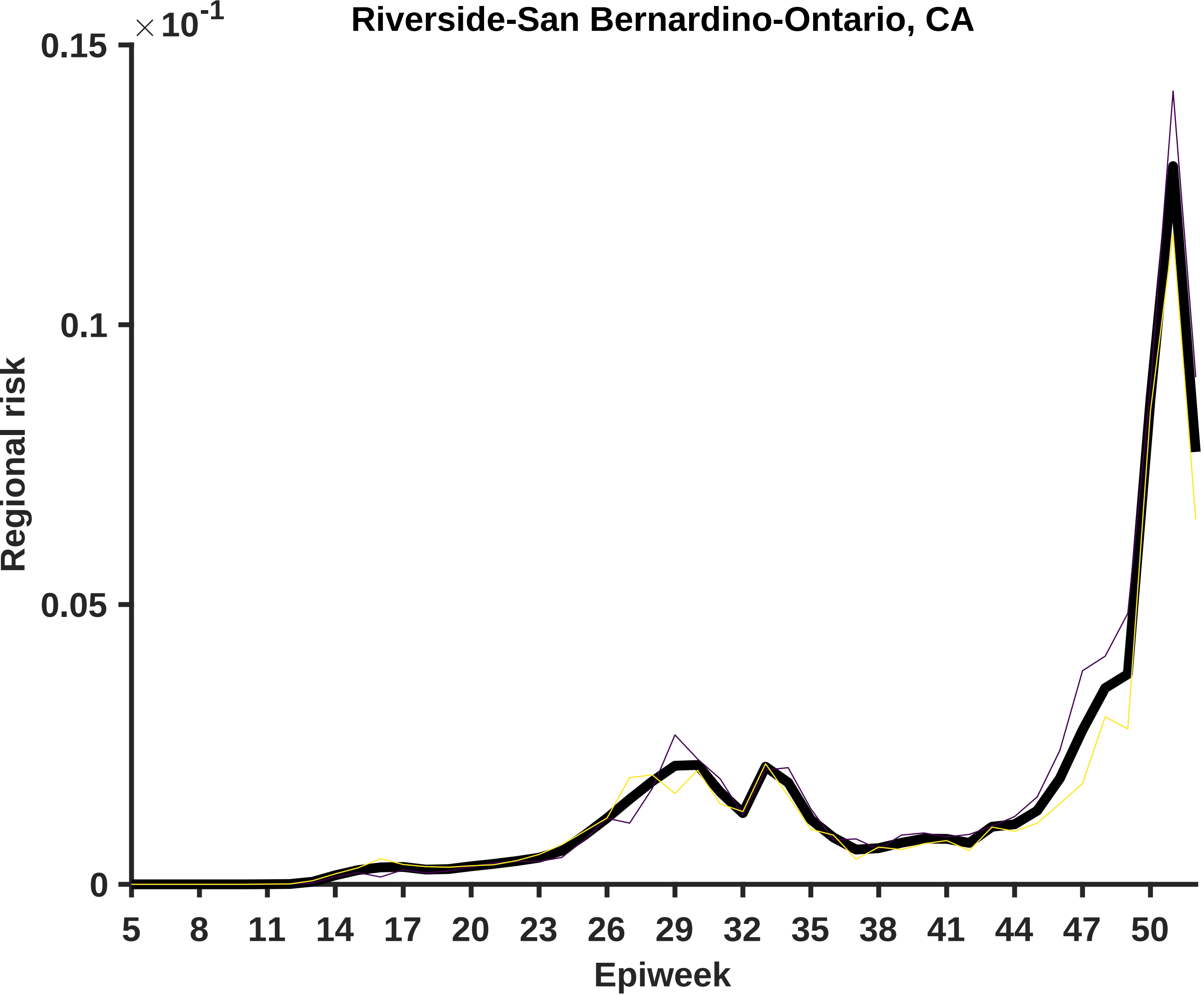
Riverside-San Bernardino-Ontario, CA
× 10^-1
0.15
0.1
0.05
0
Regional risk
5
8
11
14
17
20
23
26
29
32
35
38
41
44
47
50
Epiweek

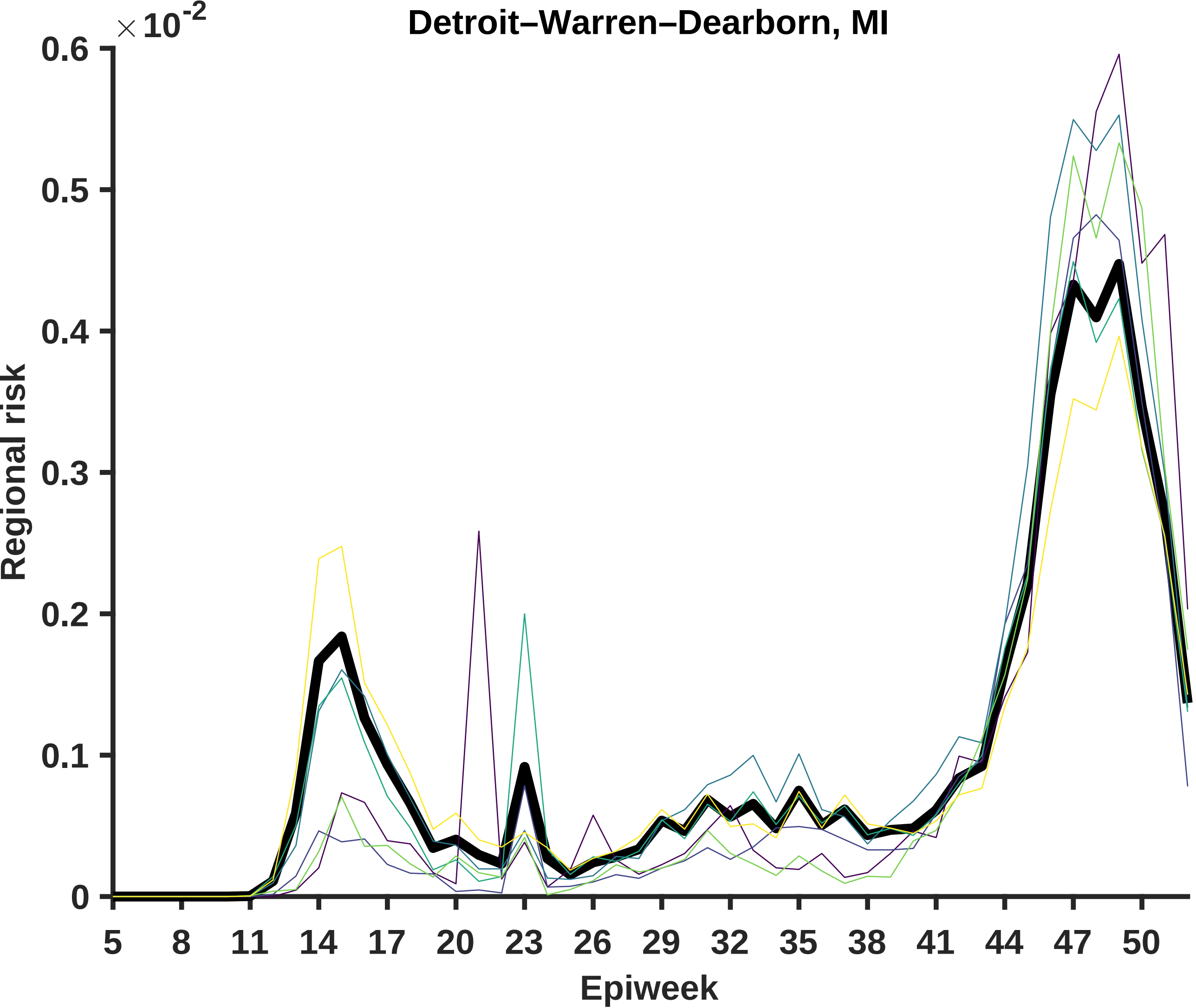

Detroit–Warren–Dearborn, MI
×10^-2
0.6
0.5
0.4
0.3
0.2
0.1
0
Regional risk
5
8
11
14
17
20
23
26
29
32
35
38
41
44
47
50
Epiweek

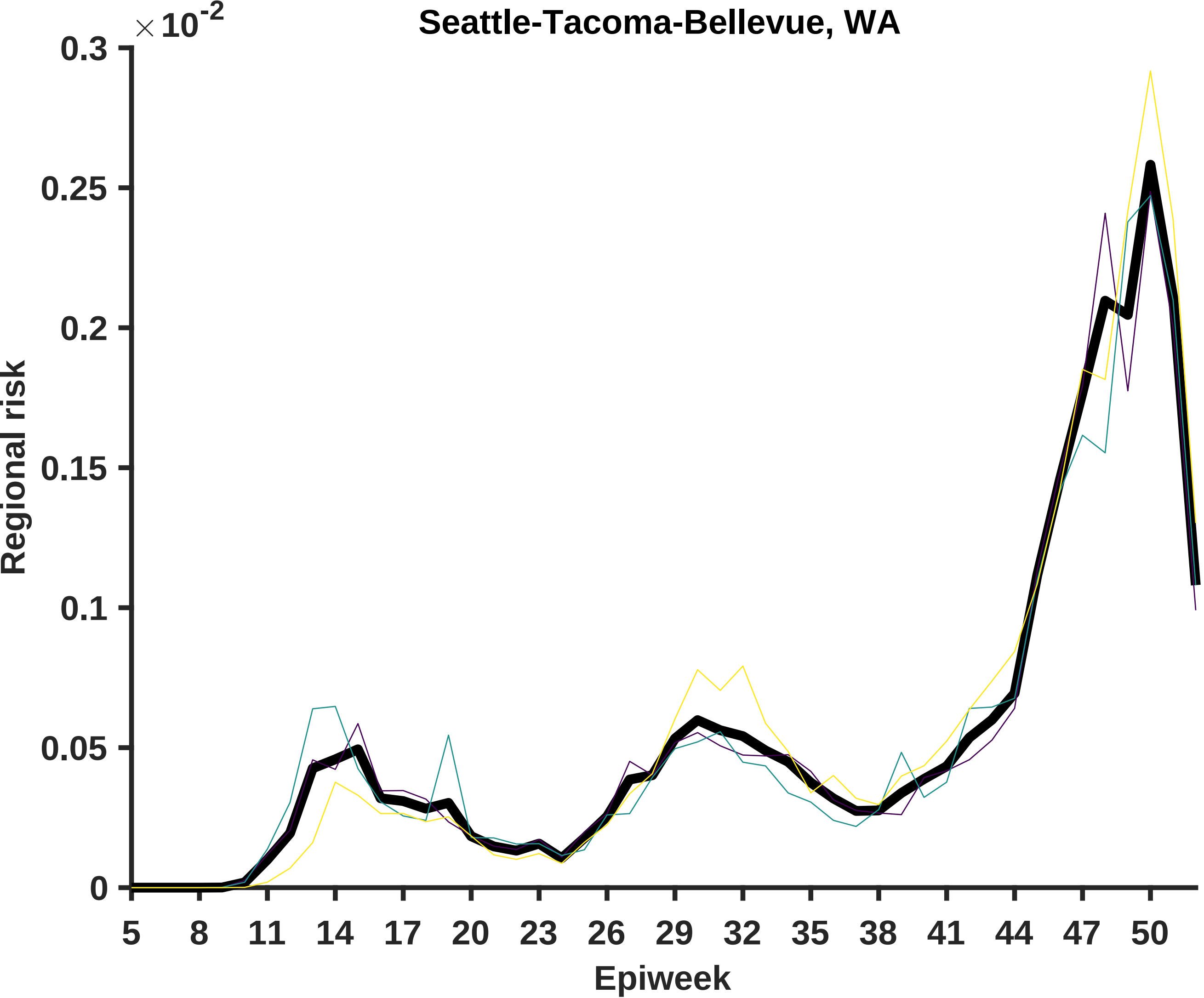

Seattle-Tacoma-Bellevue, WA
×10^-2
0.3
0.25
0.2
0.15
0.1
0.05
0
Regional risk
5
8
11
14
17
20
23
26
29
32
35
38
41
44
47
50
Epiweek

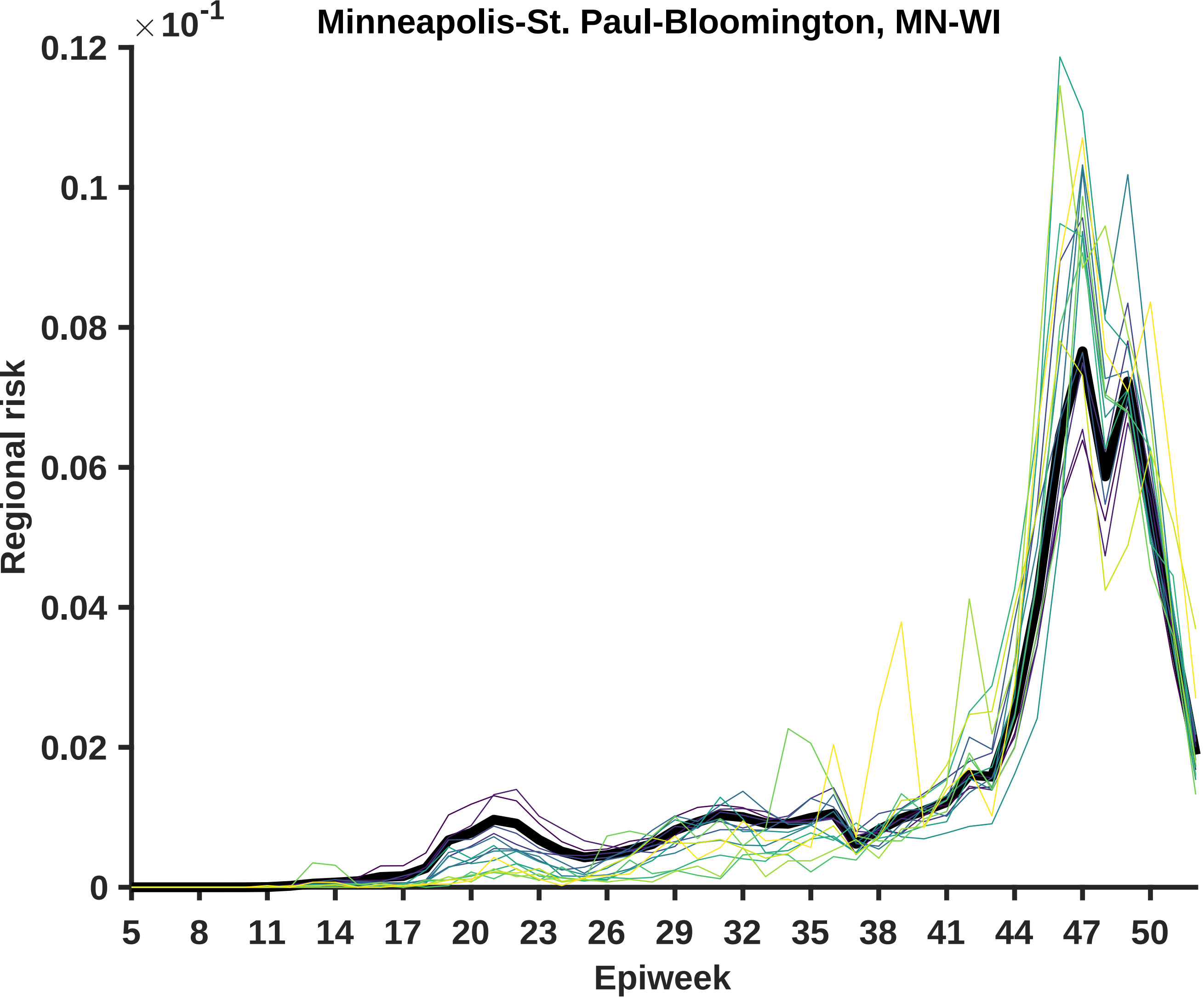

Minneapolis-St. Paul-Bloomington, MN-WI
$\times 10^{-1}$
0.12
0.1
0.08
0.06
0.04
0.02
0
Regional risk
5
8
11
14
17
20
23
26
29
32
35
38
41
44
47
50
Epiweek

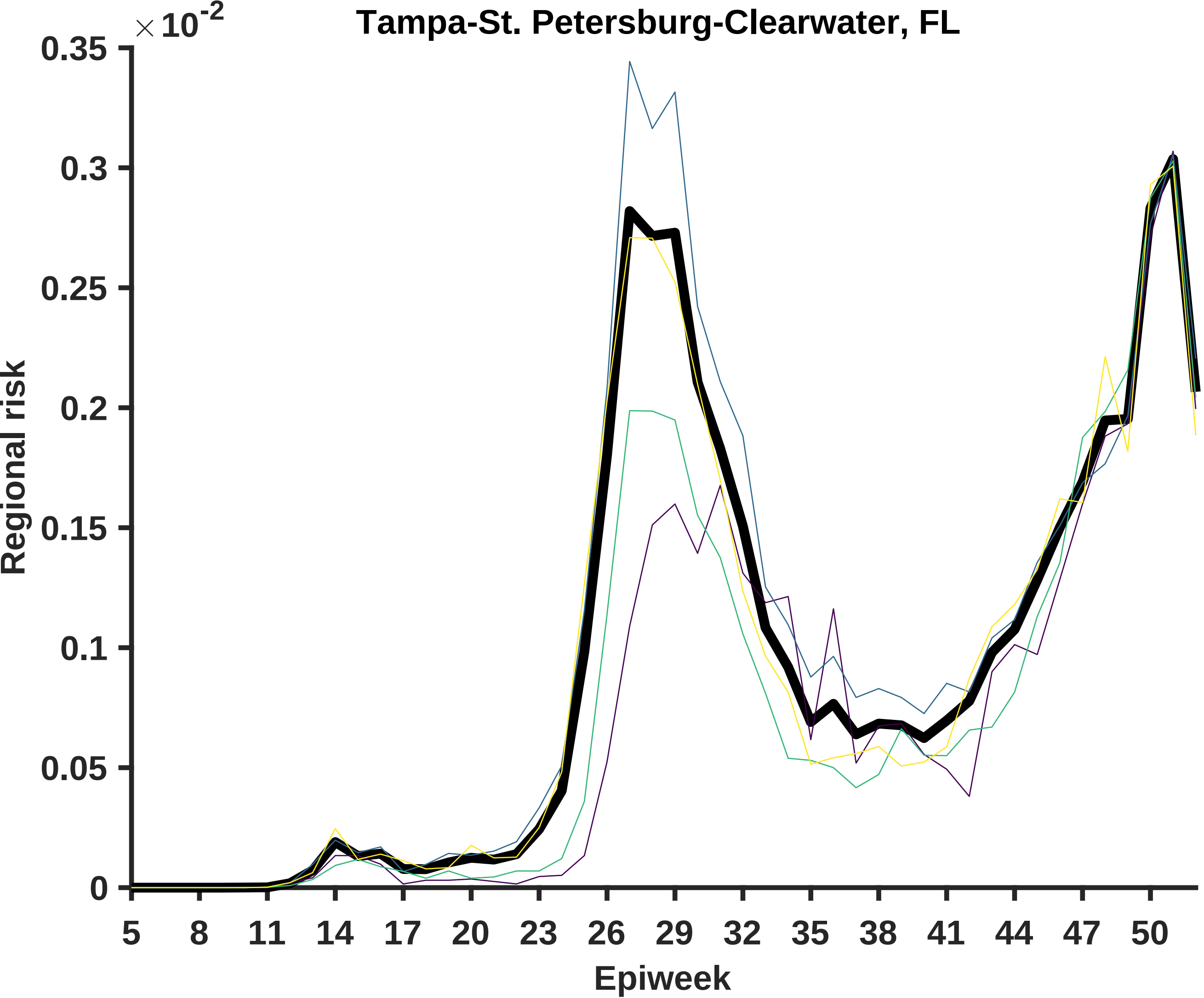

Tampa-St. Petersburg-Clearwater, FL
×10^-2
0.35
0.3
0.25
0.2
0.15
0.1
0.05
0
Regional risk
5
8
11
14
17
20
23
26
29
32
35
38
41
44
47
50
Epiweek

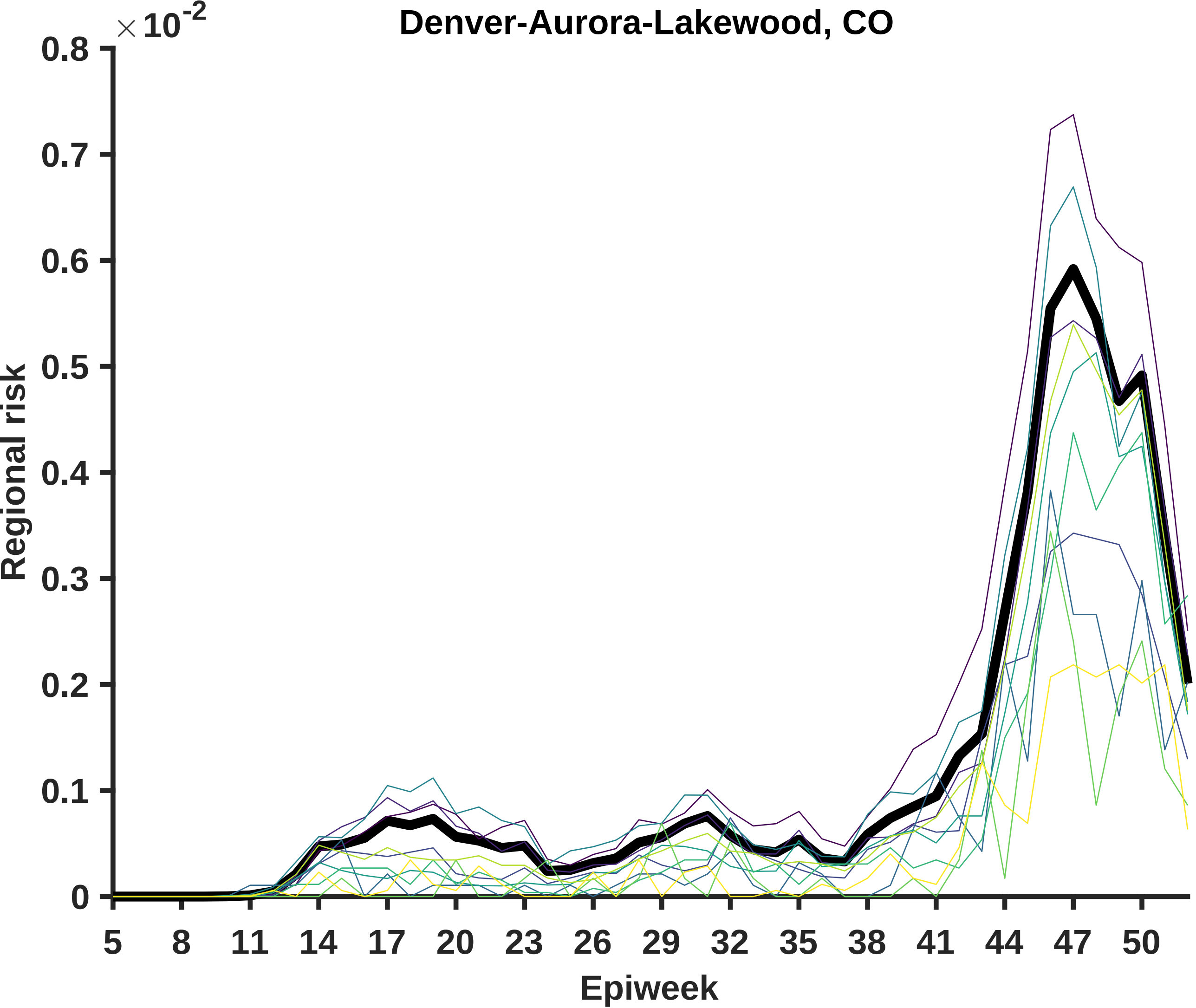
Denver-Aurora-Lakewood, CO
×10^-2
0.8
0.7
0.6
0.5
0.4
0.3
0.2
0.1
0
Regional risk
5
8
11
14
17
20
23
26
29
32
35
38
41
44
47
50
Epiweek

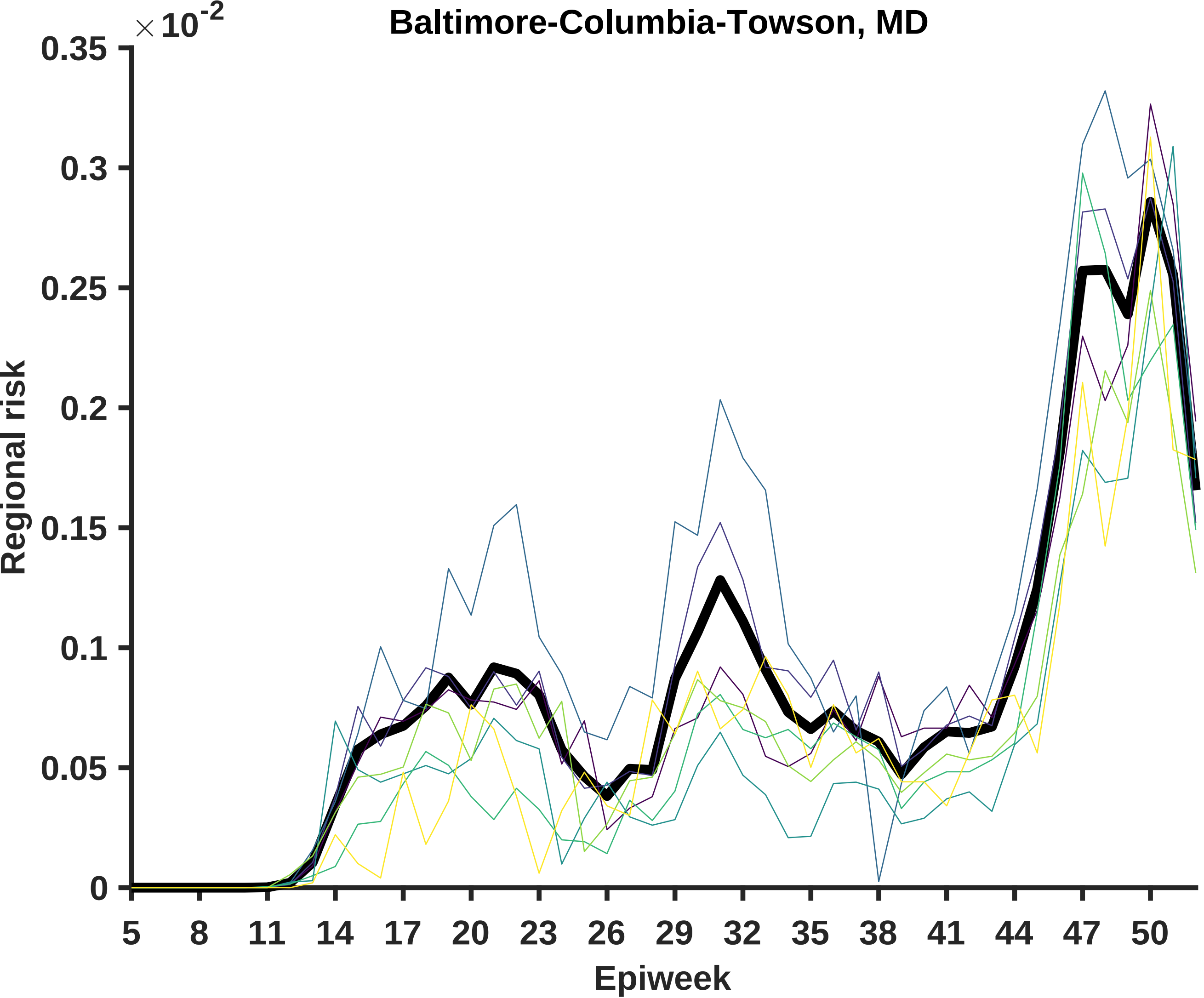
Baltimore-Columbia-Towson, MD
× 10^-2
0.35
0.3
0.25
0.2
0.15
0.1
0.05
0
Regional risk
5
8
11
14
17
20
23
26
29
32
35
38
41
44
47
50
Epiweek

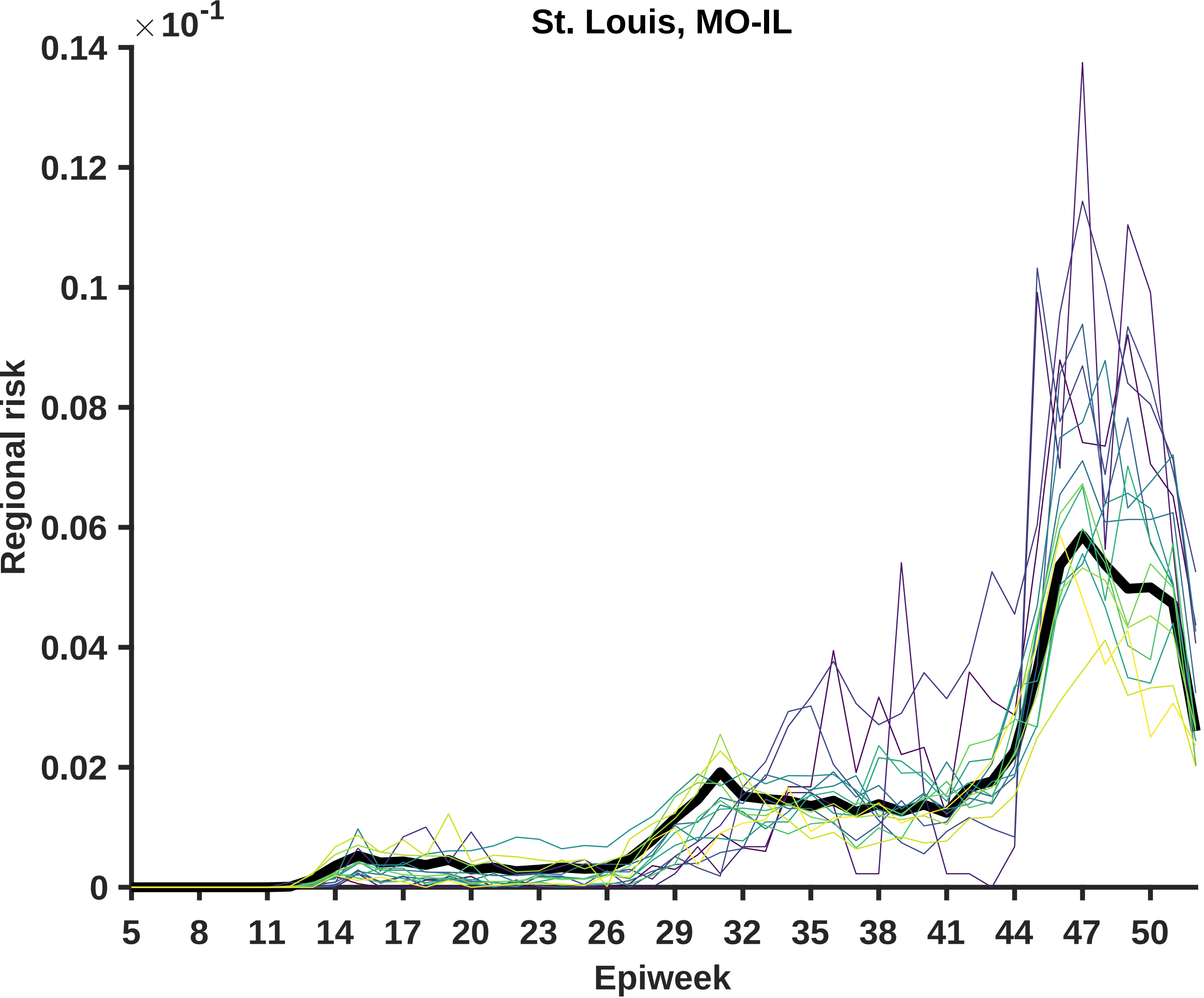

St. Louis, MO-IL
×10^-1
0.14
0.12
0.1
0.08
0.06
0.04
0.02
0
Regional risk
5
8
11
14
17
20
23
26
29
32
35
38
41
44
47
50
Epiweek

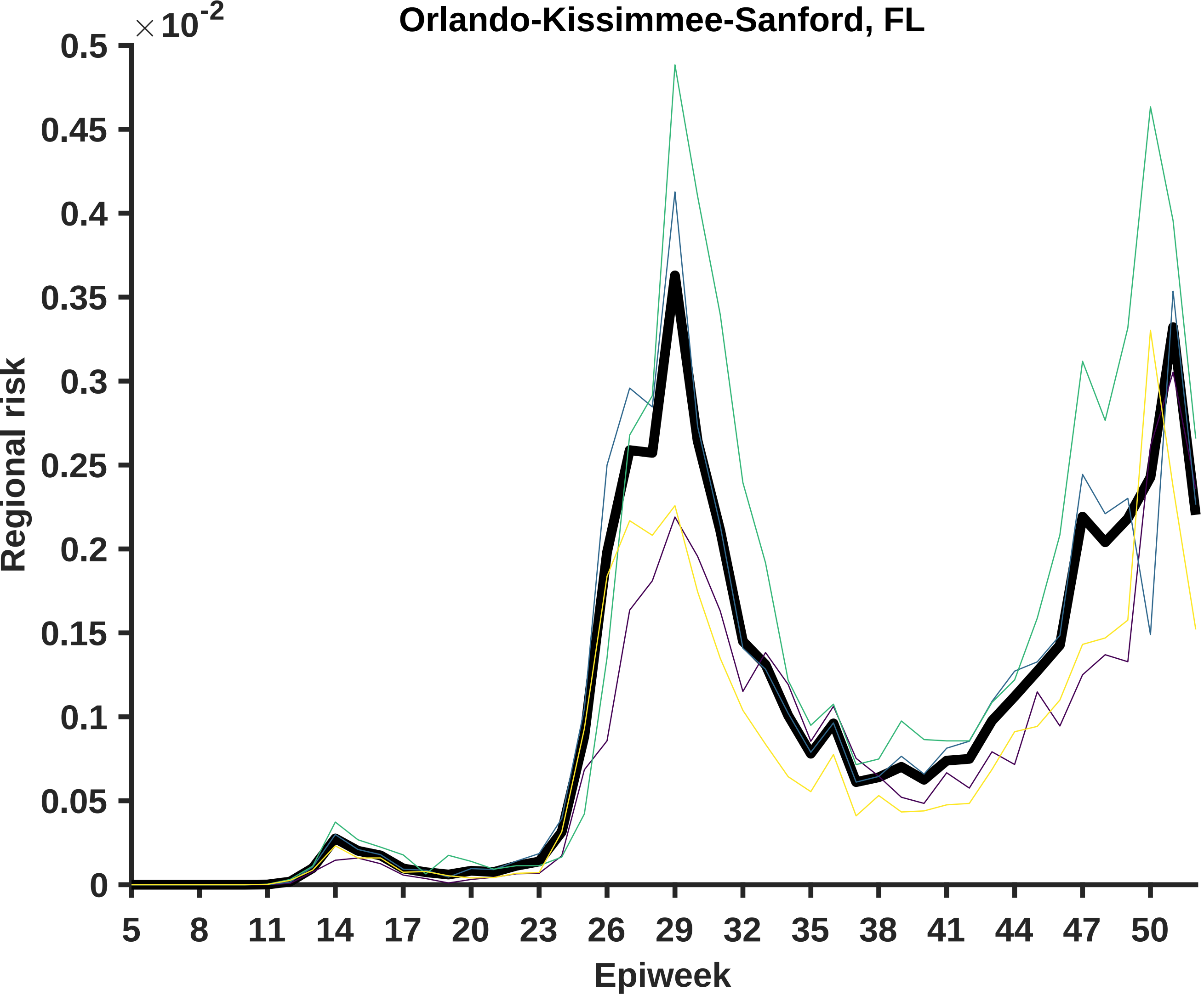

Orlando-Kissimmee-Sanford, FL
×10^-2
Regional risk
Epiweek
0
0.05
0.1
0.15
0.2
0.25
0.3
0.35
0.4
0.45
0.5
5
8
11
14
17
20
23
26
29
32
35
38
41
44
47
50

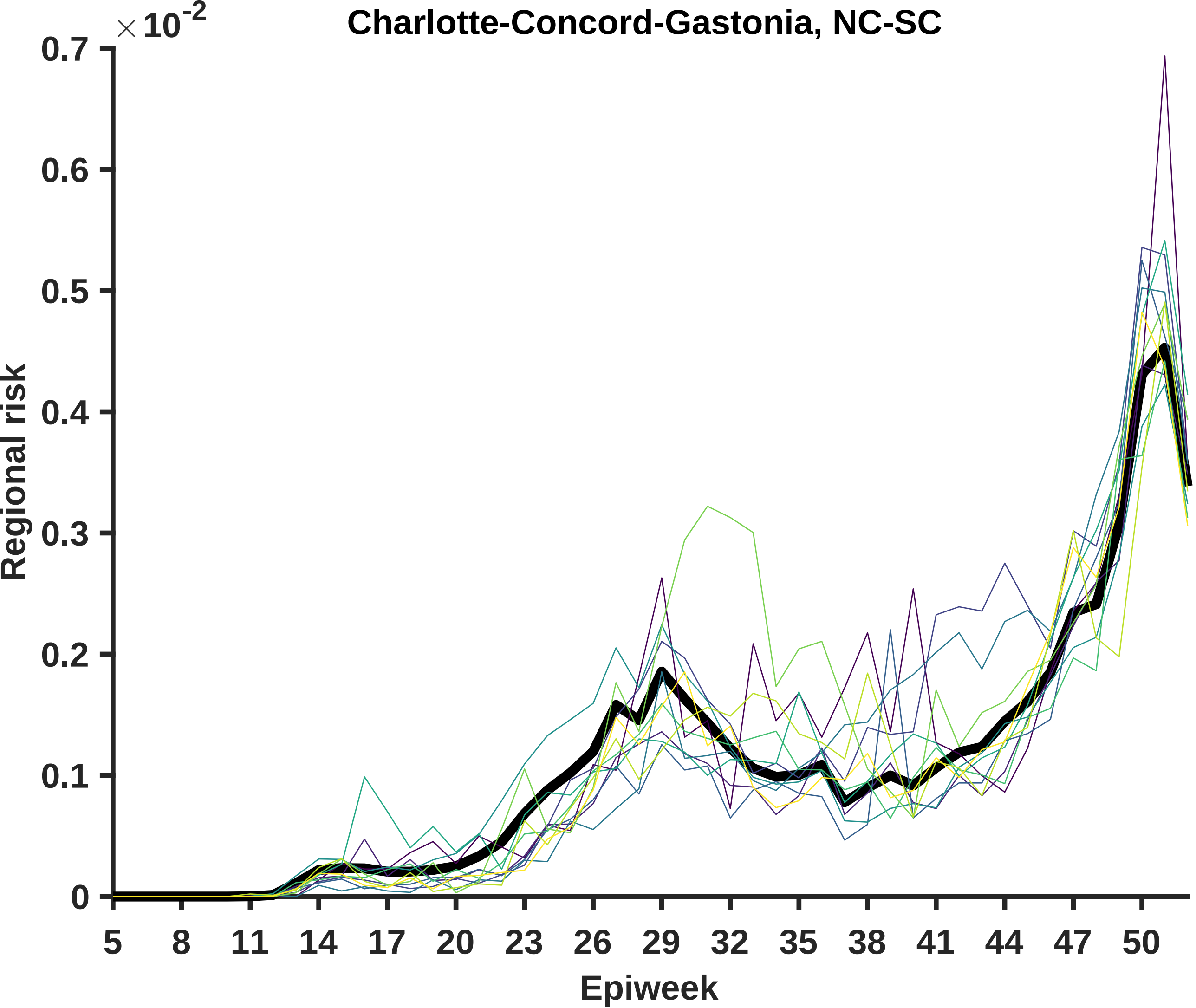

Charlotte-Concord-Gastonia, NC-SC
×10^-2
0.7
0.6
0.5
0.4
0.3
0.2
0.1
0
Regional risk
5
8
11
14
17
20
23
26
29
32
35
38
41
44
47
50
Epiweek

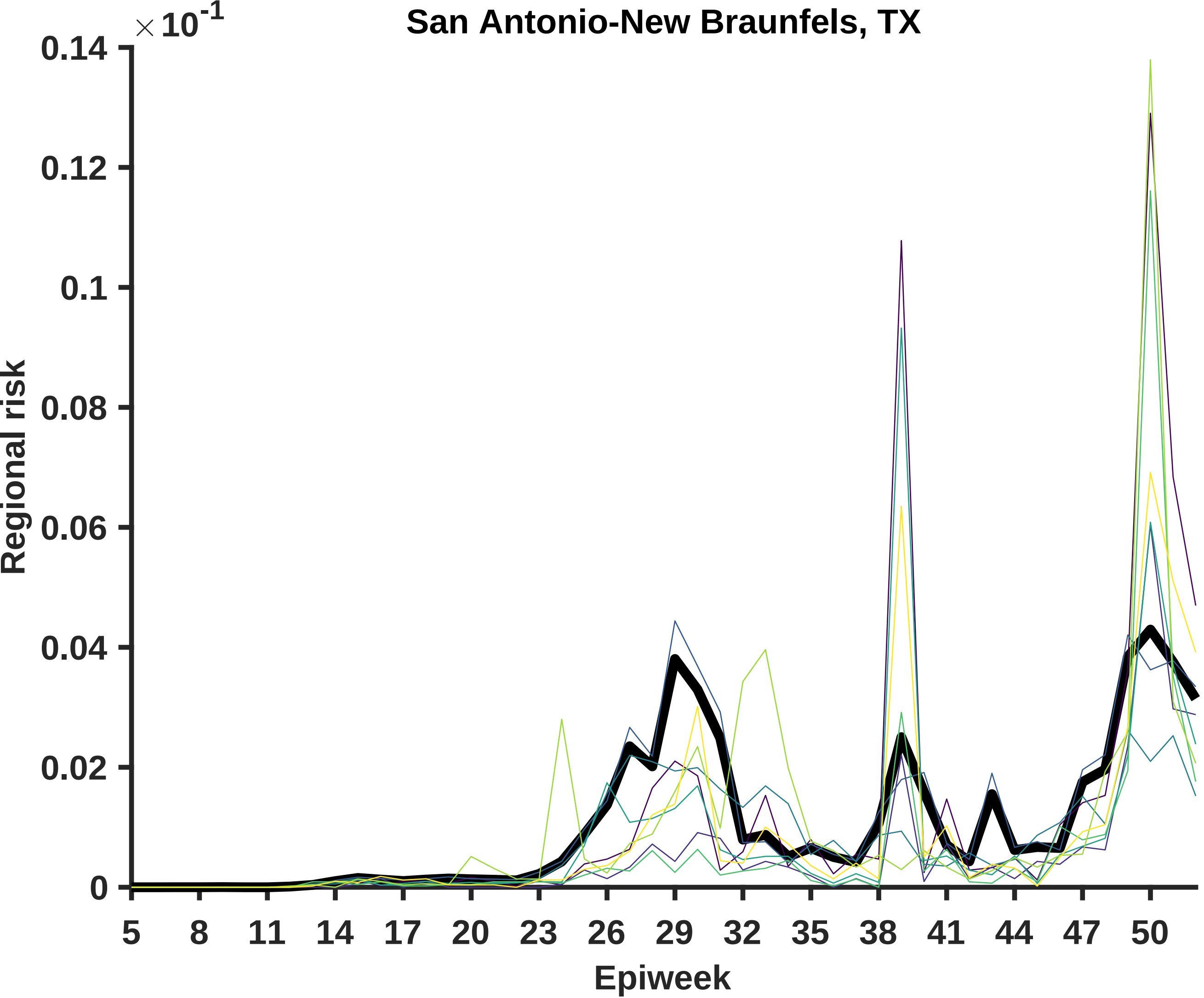

San Antonio-New Braunfels, TX
×10^-1
0.14
0.12
0.1
0.08
0.06
0.04
0.02
0
Regional risk
5
8
11
14
17
20
23
26
29
32
35
38
41
44
47
50
Epiweek

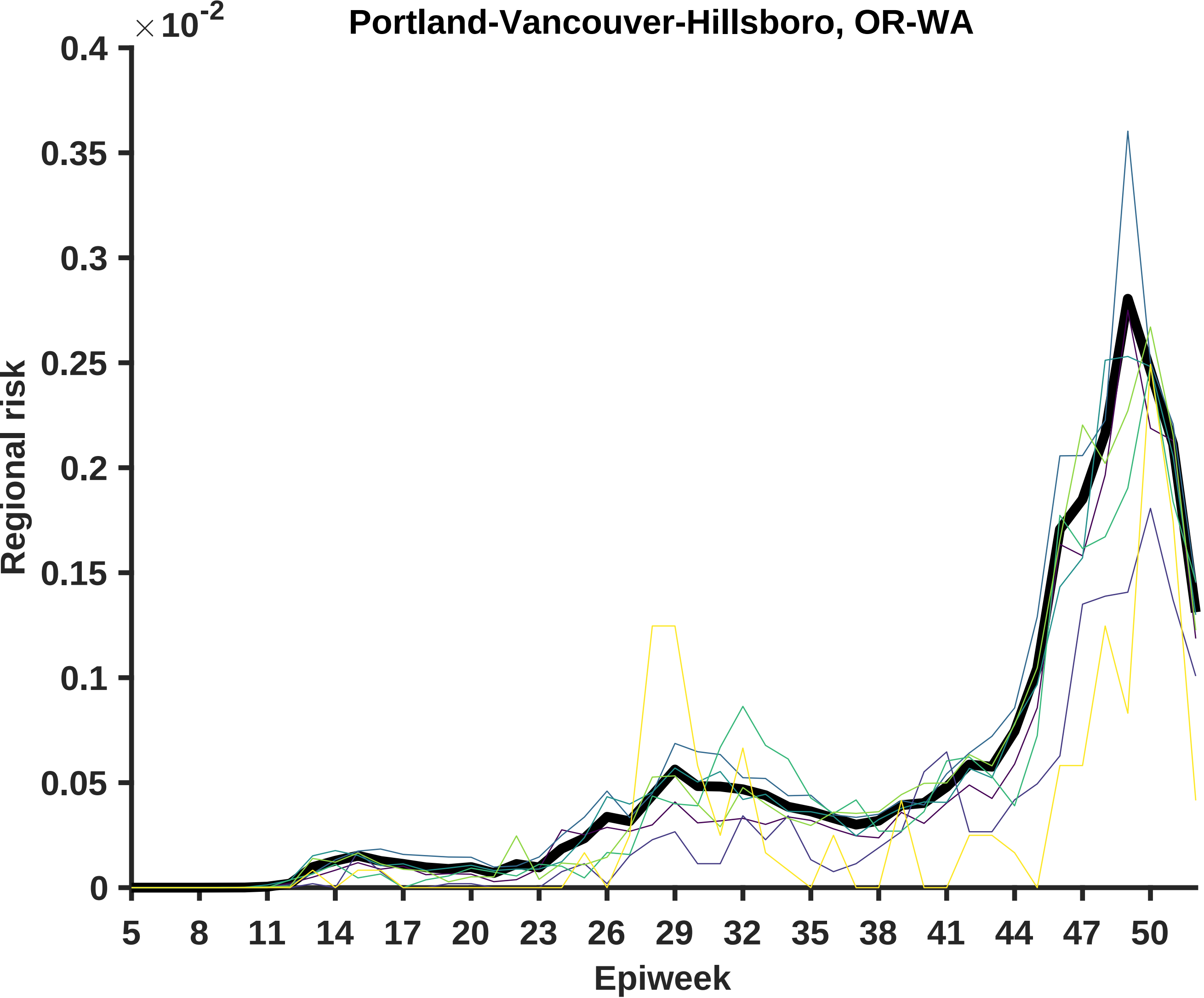
Portland-Vancouver-Hillsboro, OR-WA
×10-2
0.4
0.35
0.3
0.25
0.2
0.15
0.1
0.05
0
Regional risk
5
8
11
14
17
20
23
26
29
32
35
38
41
44
47
50
Epiweek

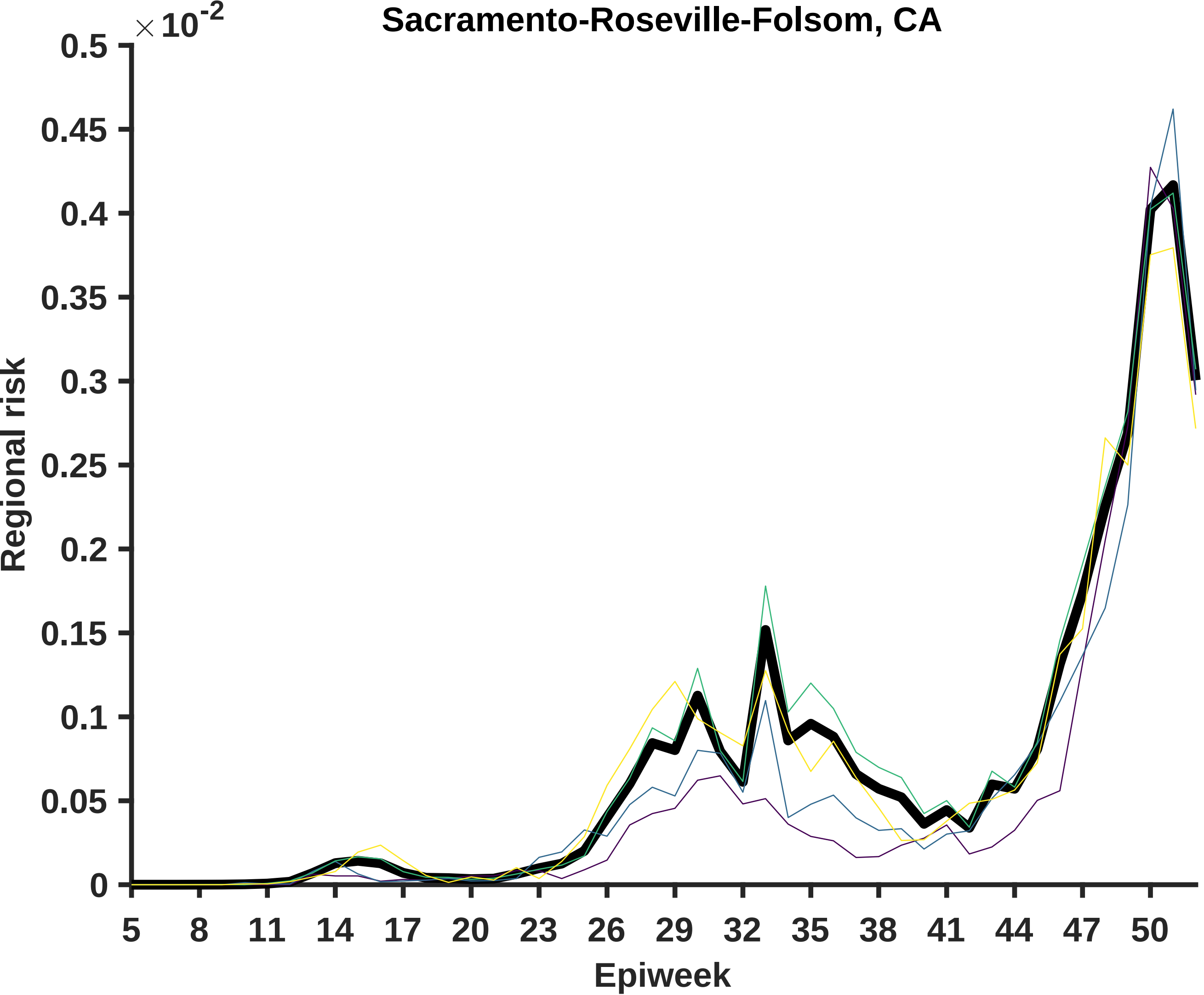

Sacramento-Roseville-Folsom, CA
$\times 10^{-2}$
0.5
0.45
0.4
0.35
0.3
0.25
0.2
0.15
0.1
0.05
0
Regional risk
5
8
11
14
17
20
23
26
29
32
35
38
41
44
47
50
Epiweek

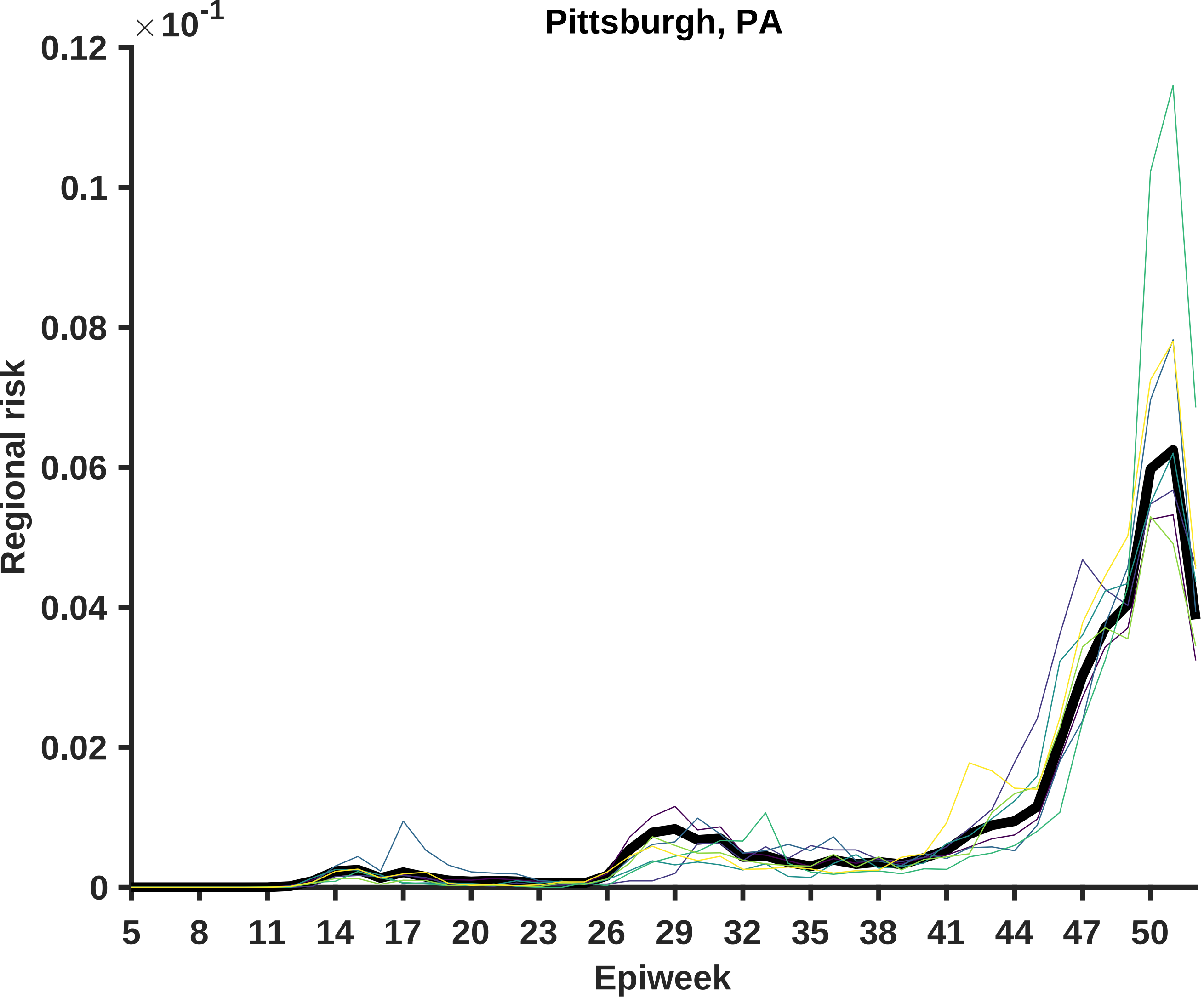
Pittsburgh, PA
×10^-1
0.12
0.1
0.08
0.06
0.04
0.02
0
Regional risk
5
8
11
14
17
20
23
26
29
32
35
38
41
44
47
50
Epiweek

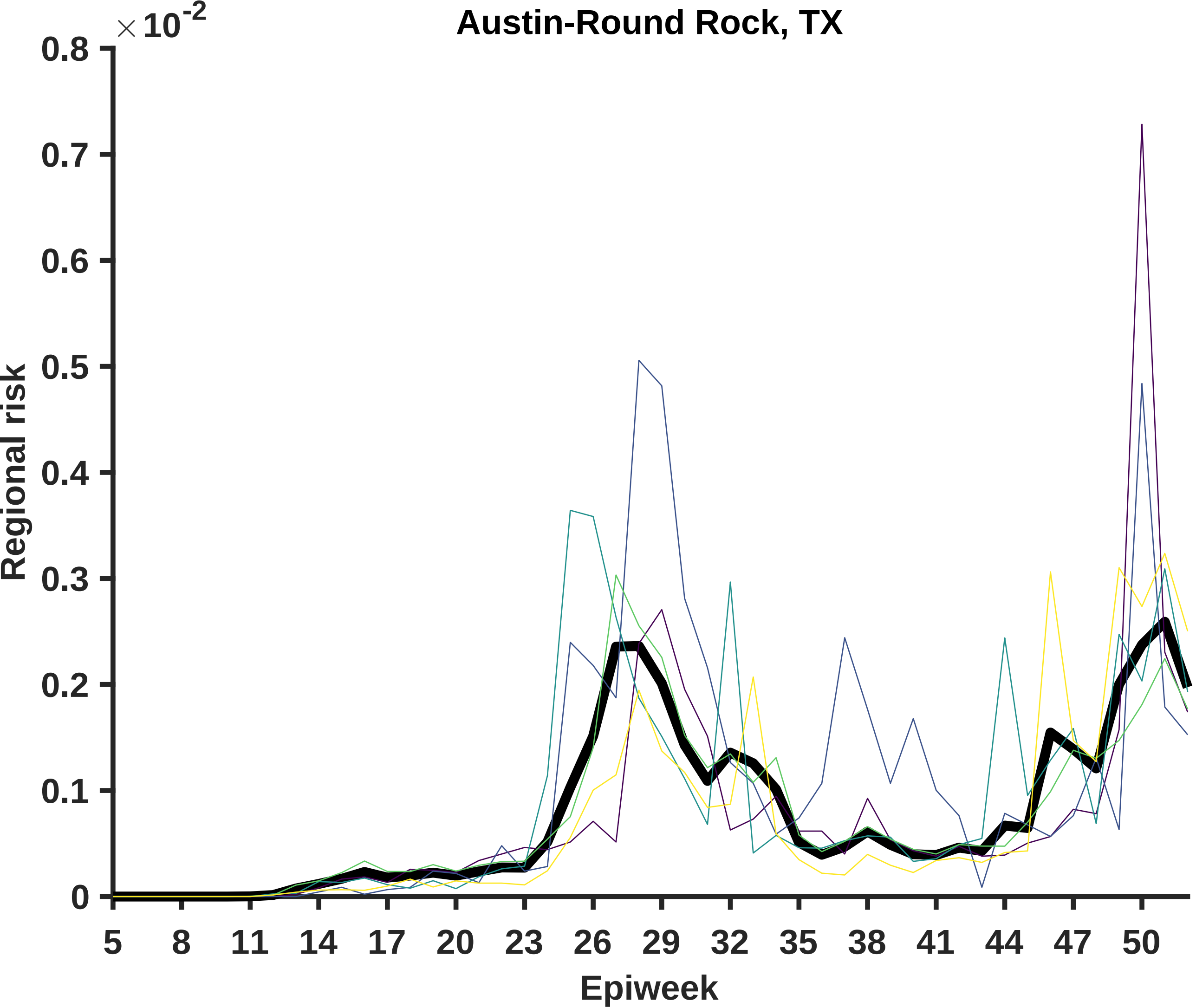

Austin-Round Rock, TX
×10^-2
0.8
0.7
0.6
0.5
0.4
0.3
0.2
0.1
0
Regional risk
5
8
11
14
17
20
23
26
29
32
35
38
41
44
47
50
Epiweek

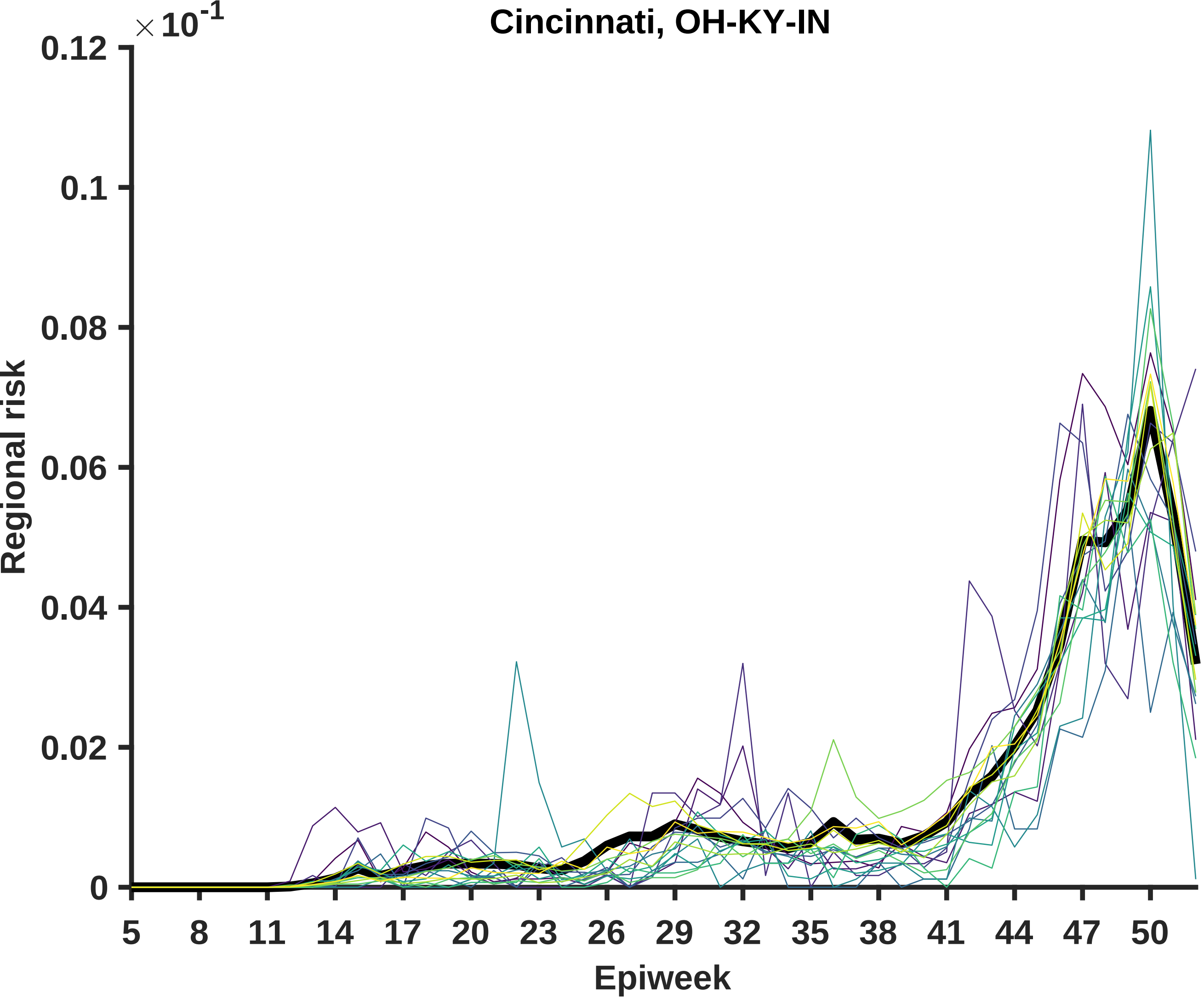

Cincinnati, OH-KY-IN
× 10^-1
Regional risk
0.12
0.1
0.08
0.06
0.04
0.02
0
5
8
11
14
17
20
23
26
29
32
35
38
41
44
47
50
Epiweek

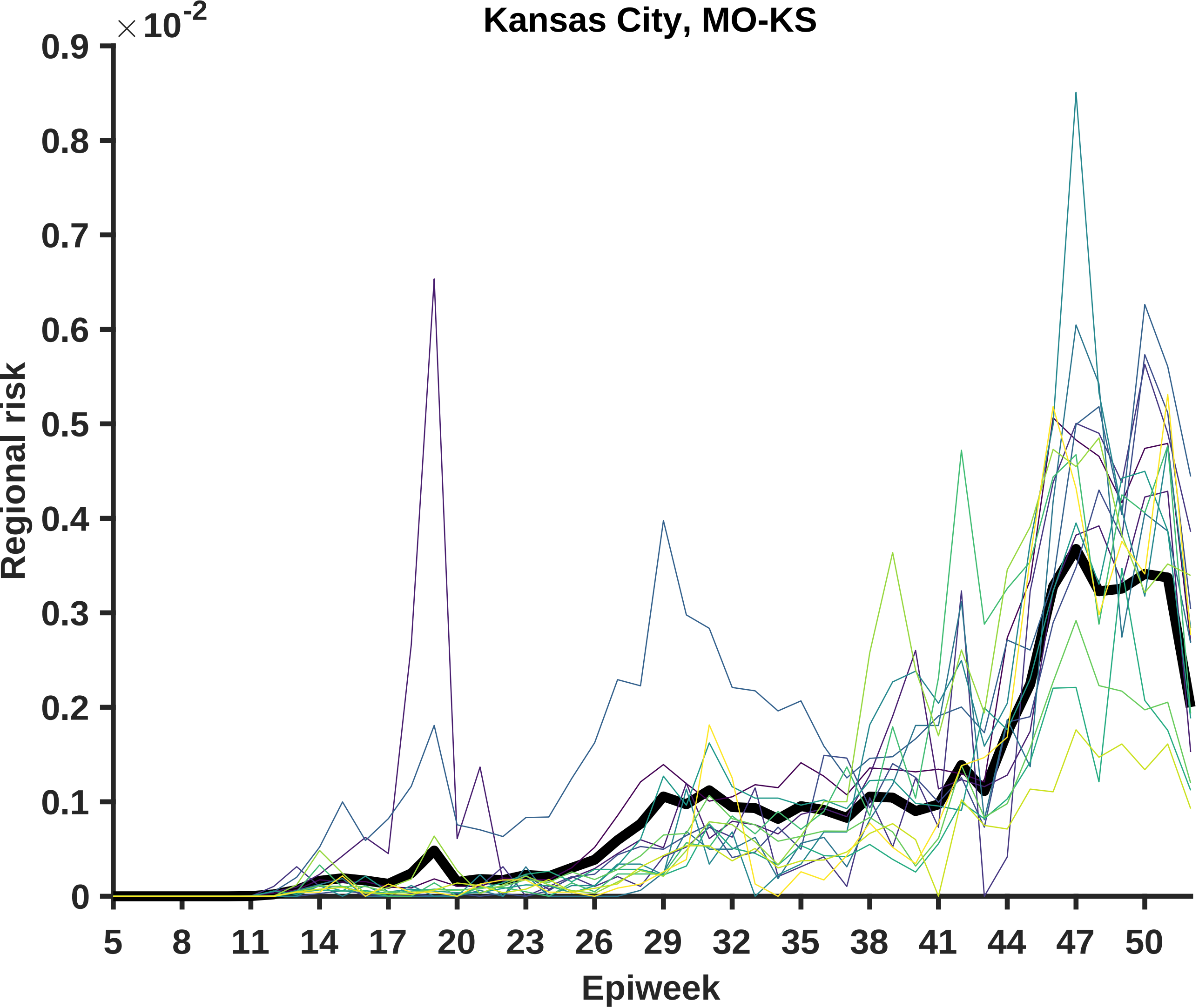

Kansas City, MO-KS
×10^-2
0.9
0.8
0.7
0.6
0.5
0.4
0.3
0.2
0.1
0
Regional risk
5
8
11
14
17
20
23
26
29
32
35
38
41
44
47
50
Epiweek

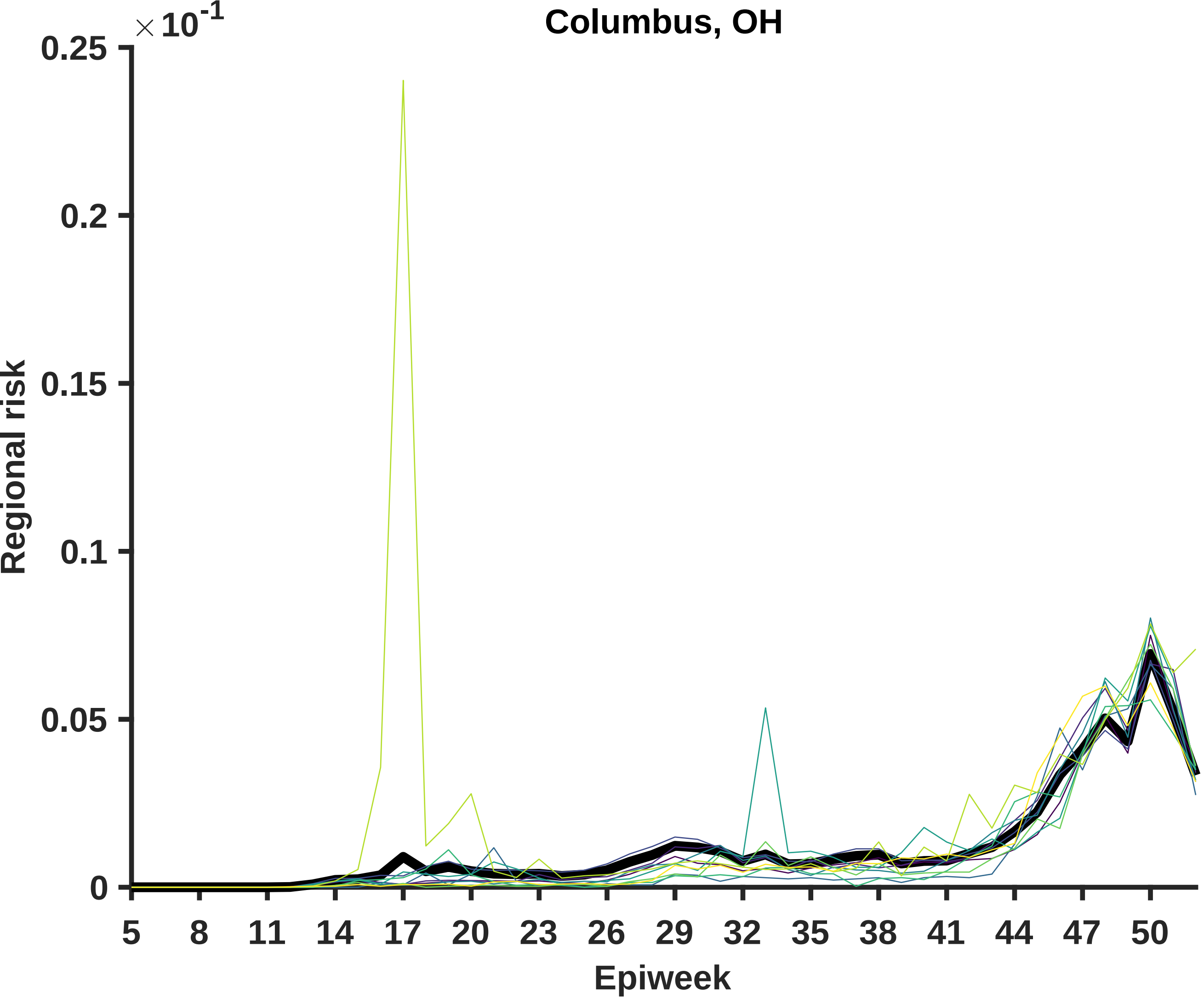

Columbus, OH
×10^-1
0.25
0.2
0.15
0.1
0.05
0
Regional risk
5
8
11
14
17
20
23
26
29
32
35
38
41
44
47
50
Epiweek

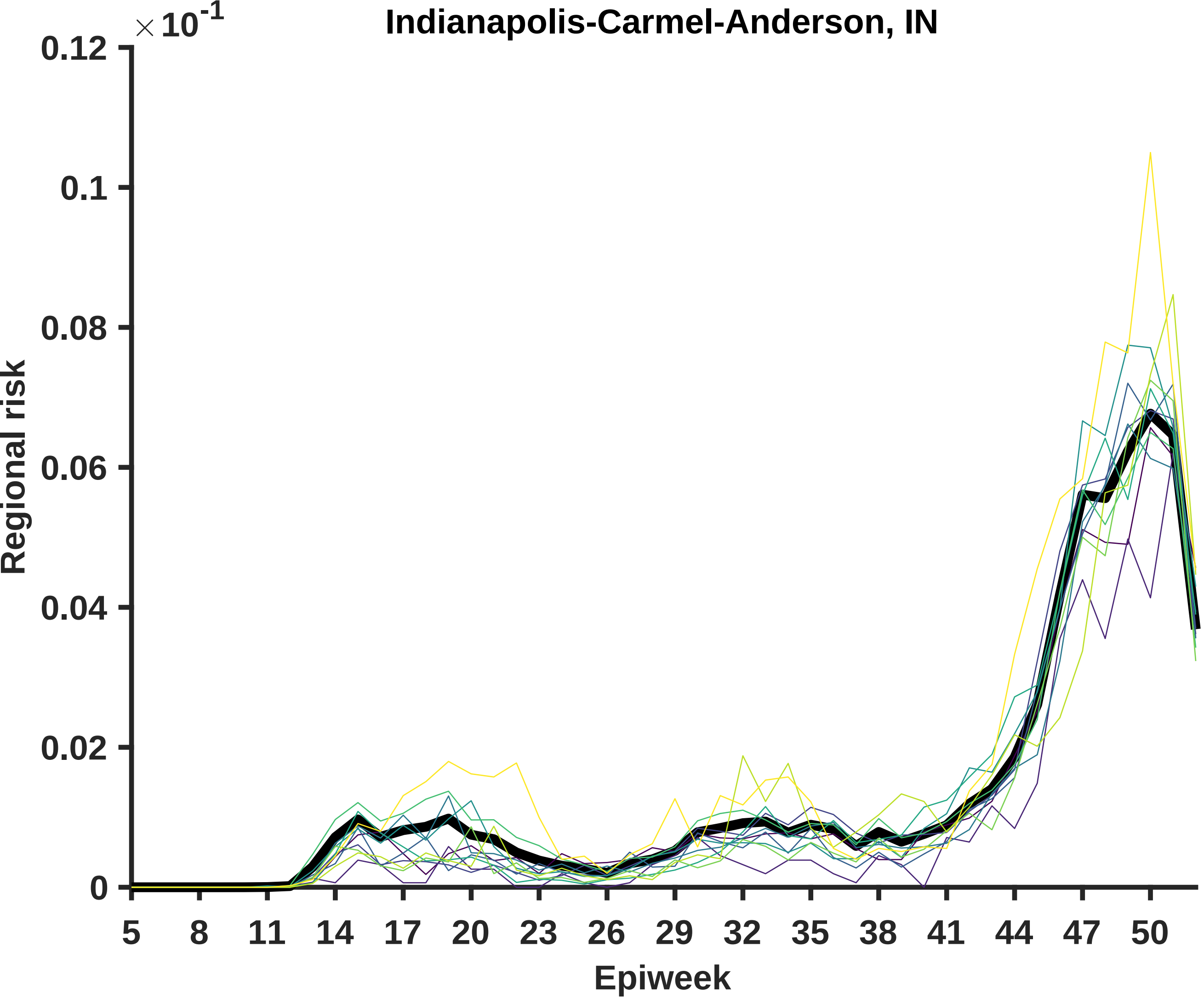

Indianapolis-Carmel-Anderson, IN
× 10^-1
0.12
0.1
0.08
0.06
0.04
0.02
0
Regional risk
5
8
11
14
17
20
23
26
29
32
35
38
41
44
47
50
Epiweek

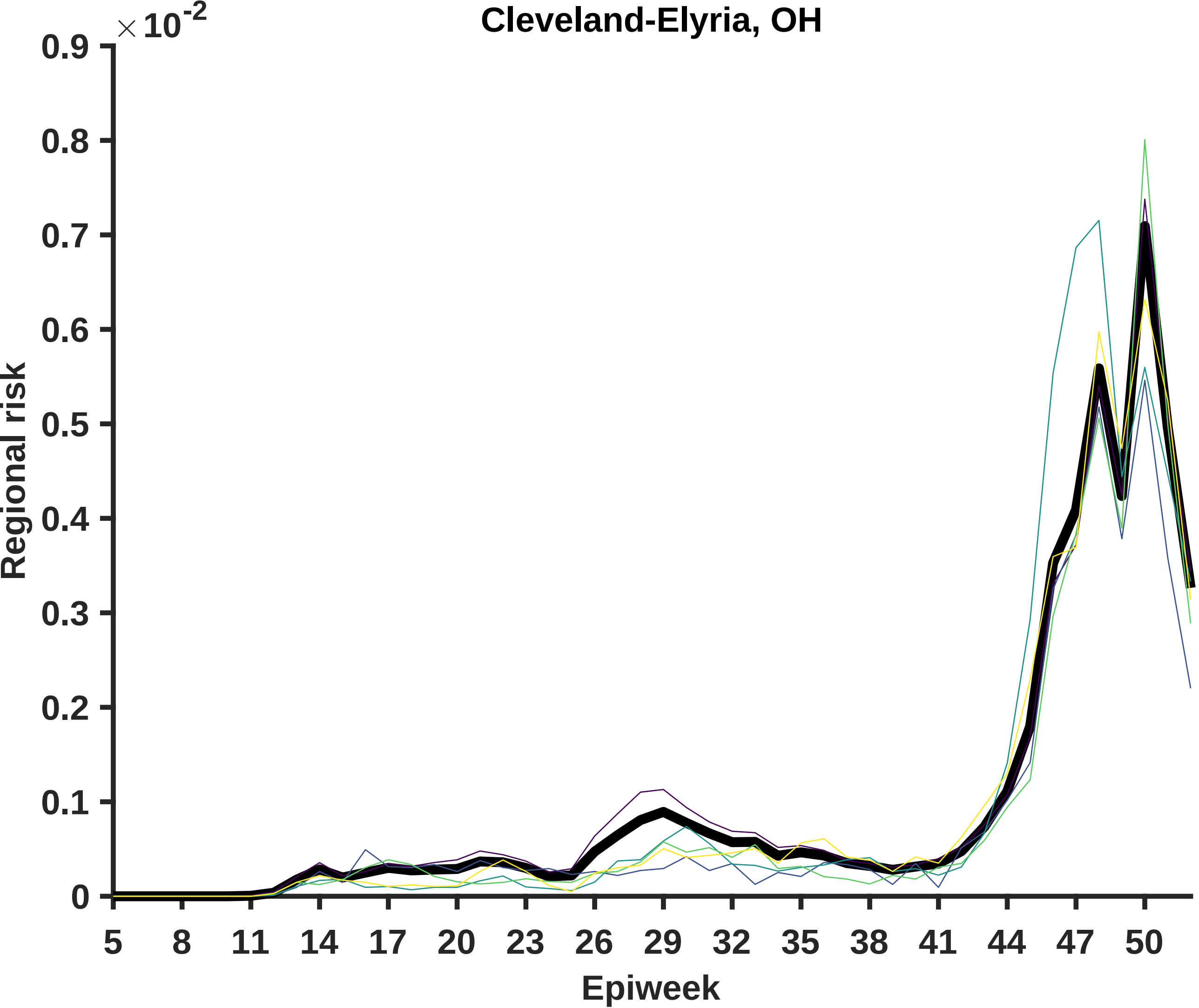

Cleveland-Elyria, OH
×10^-2
0.9
0.8
0.7
0.6
0.5
0.4
0.3
0.2
0.1
0
Regional risk
5
8
11
14
17
20
23
26
29
32
35
38
41
44
47
50
Epiweek

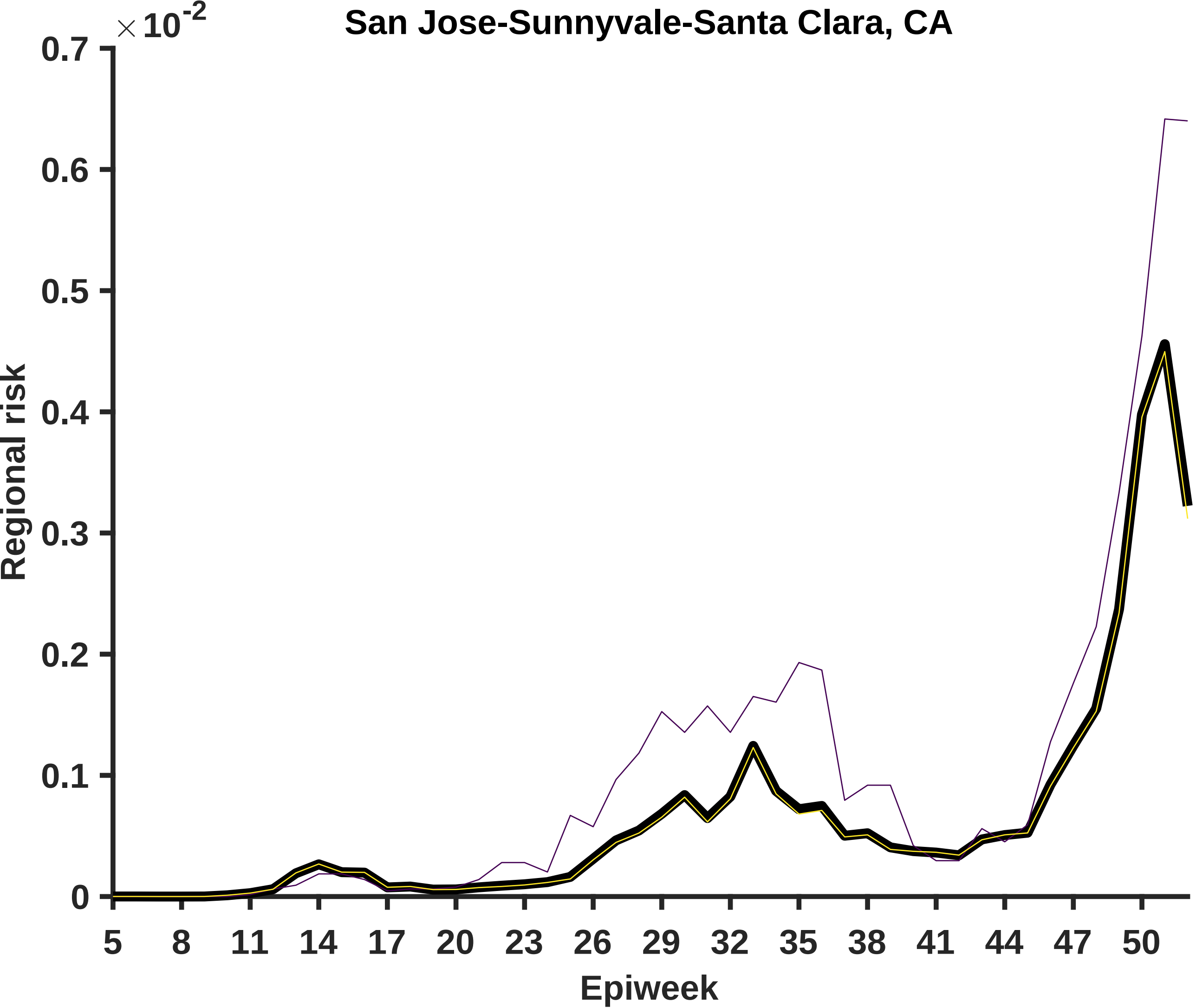
San Jose-Sunnyvale-Santa Clara, CA
×10-2
0.7
0.6
0.5
0.4
0.3
0.2
0.1
0
Regional risk
5
8
11
14
17
20
23
26
29
32
35
38
41
44
47
50
Epiweek

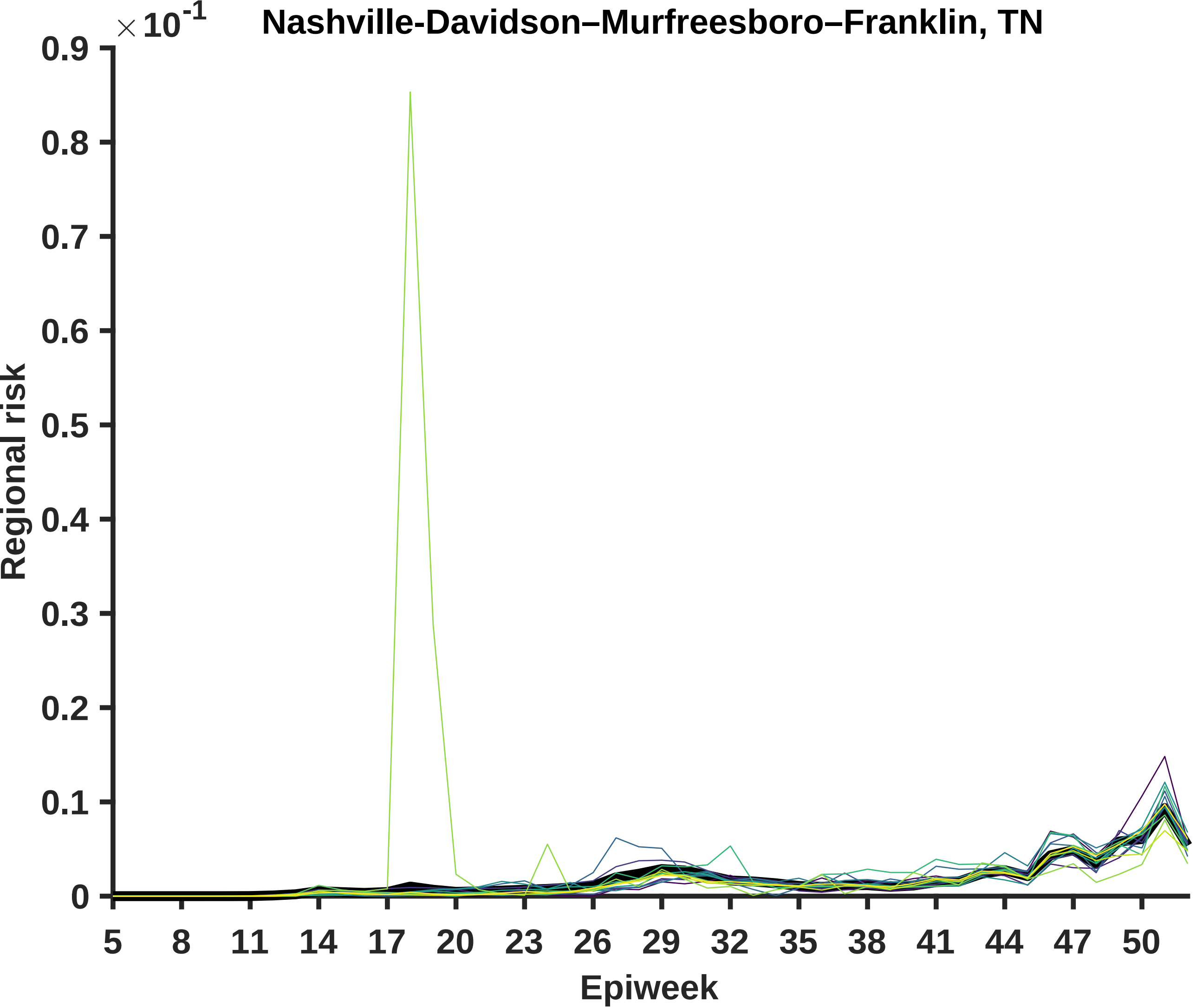
Nashville-Davidson–Murfreesboro–Franklin, TN
× 10^-1
0.9
0.8
0.7
0.6
0.5
0.4
0.3
0.2
0.1
0
Regional risk
5
8
11
14
17
20
23
26
29
32
35
38
41
44
47
50
Epiweek

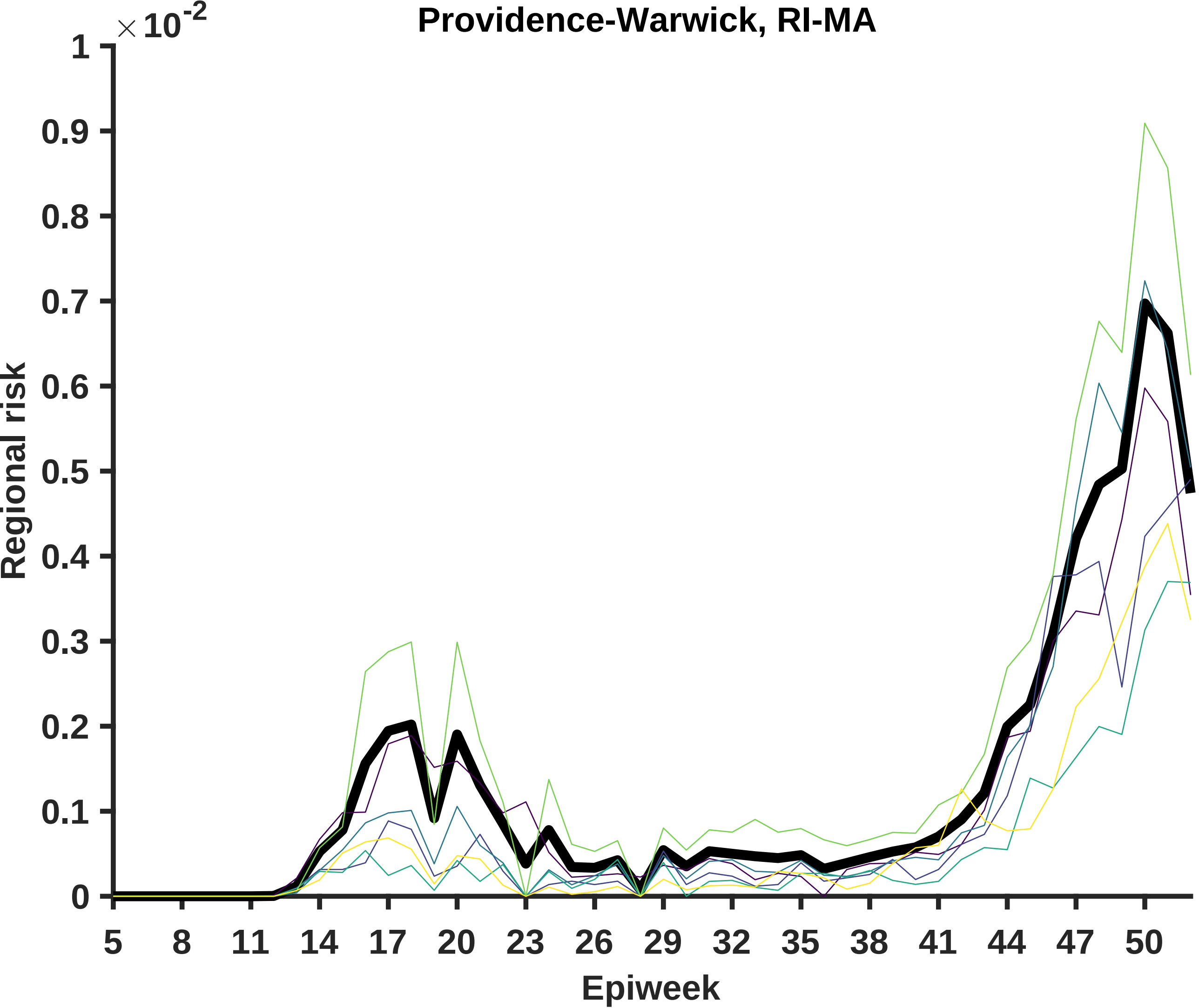

Providence-Warwick, RI-MA
×10^-2
1
0.9
0.8
0.7
0.6
0.5
0.4
0.3
0.2
0.1
0
Regional risk
5
8
11
14
17
20
23
26
29
32
35
38
41
44
47
50
Epiweek

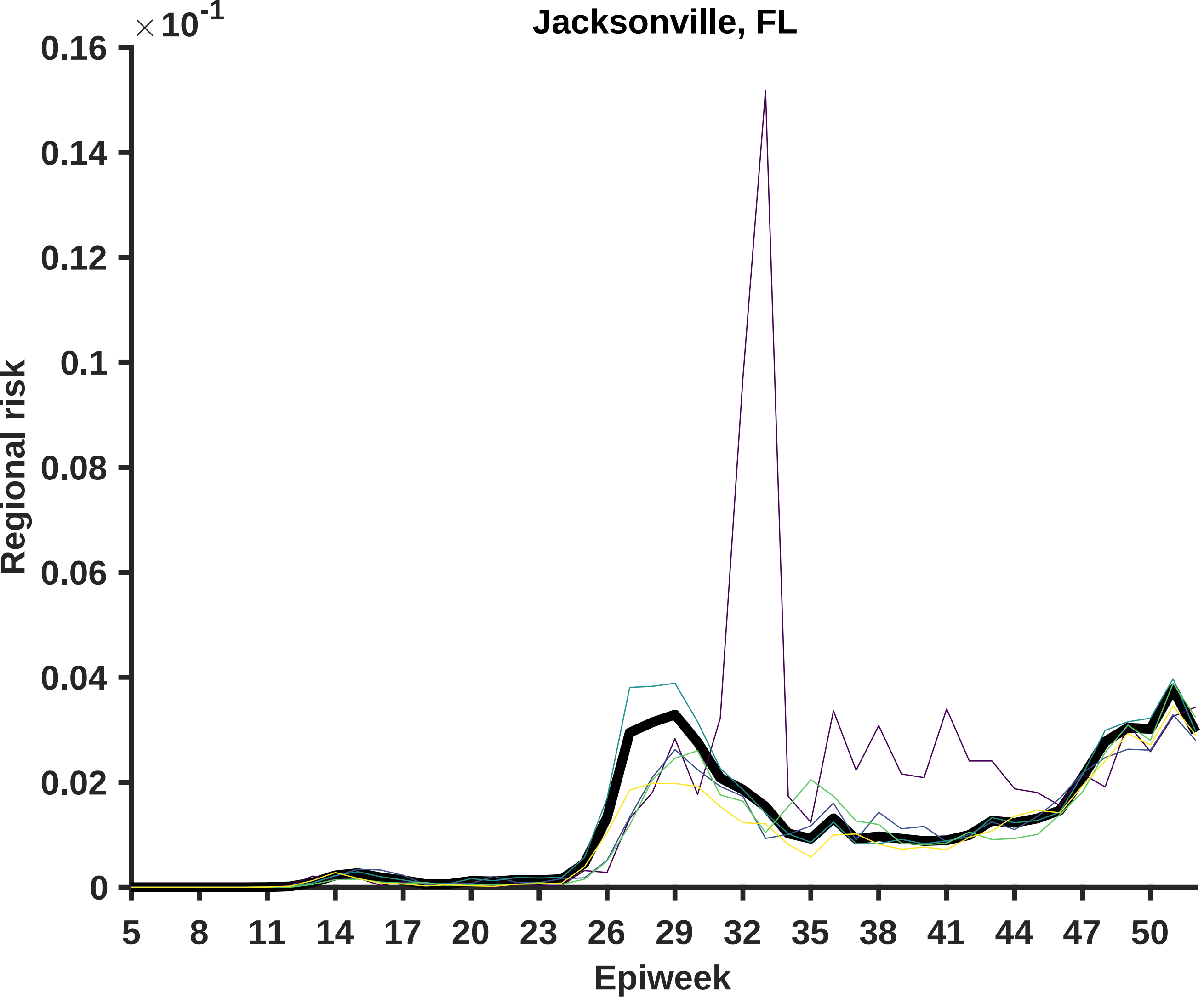

Jacksonville, FL
×10^-1
0.16
0.14
0.12
0.1
0.08
0.06
0.04
0.02
0
Regional risk
5
8
11
14
17
20
23
26
29
32
35
38
41
44
47
50
Epiweek

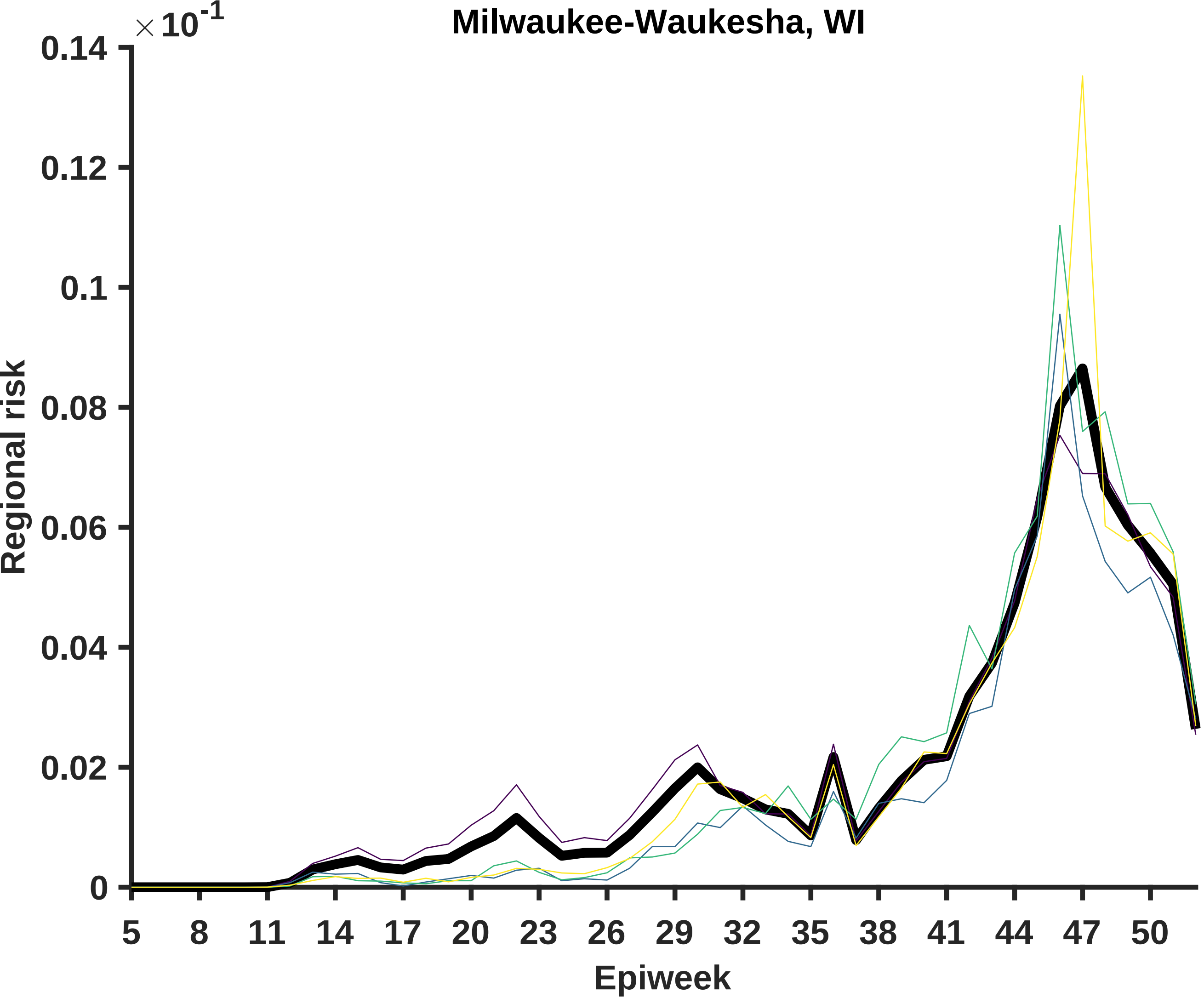

Milwaukee-Waukesha, WI
×10^-1
0.14
0.12
0.1
0.08
0.06
0.04
0.02
0
Regional risk
5
8
11
14
17
20
23
26
29
32
35
38
41
44
47
50
Epiweek

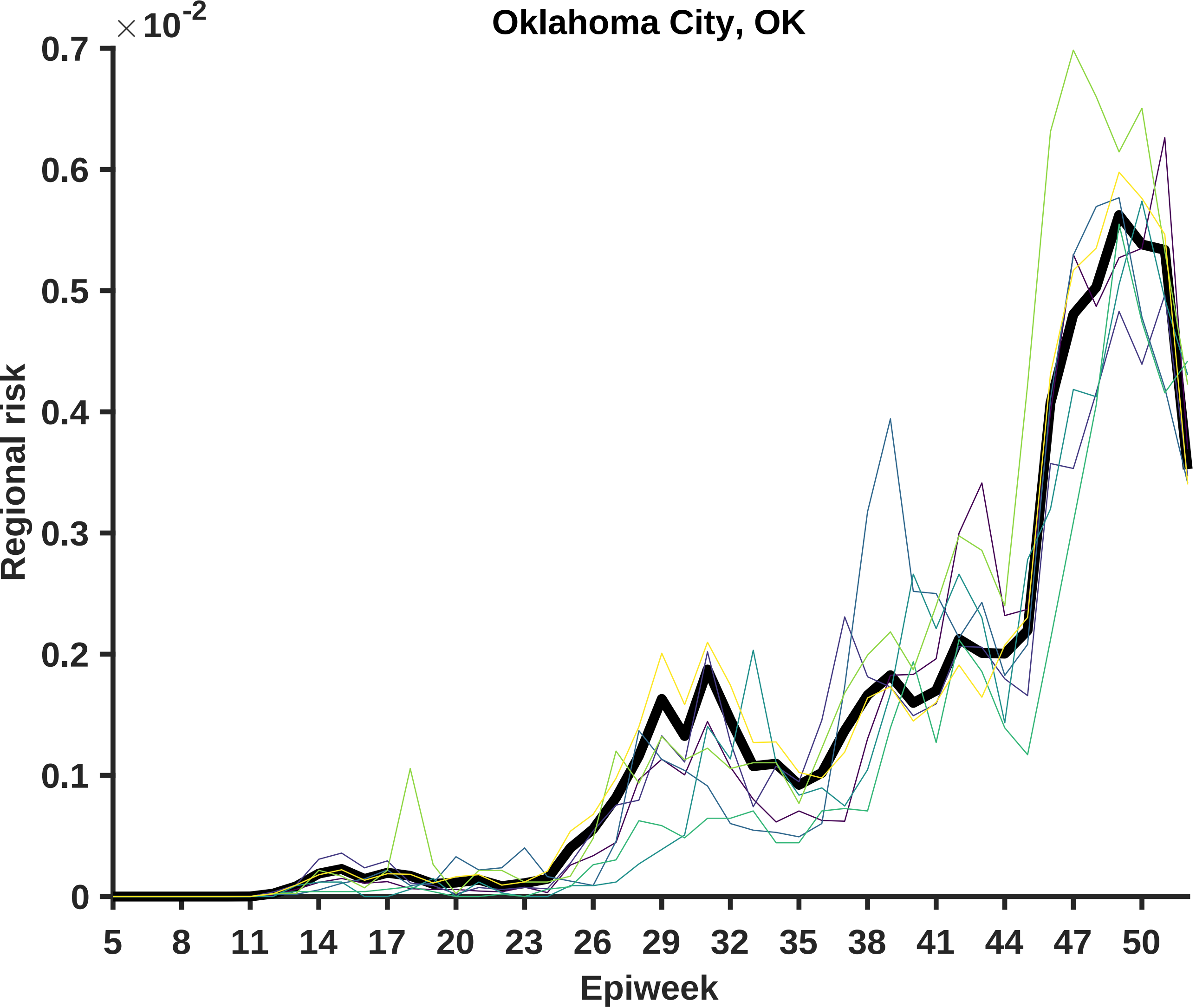

Oklahoma City, OK
×10^-2
0.7
0.6
0.5
0.4
0.3
0.2
0.1
0
Regional risk
5
8
11
14
17
20
23
26
29
32
35
38
41
44
47
50
Epiweek

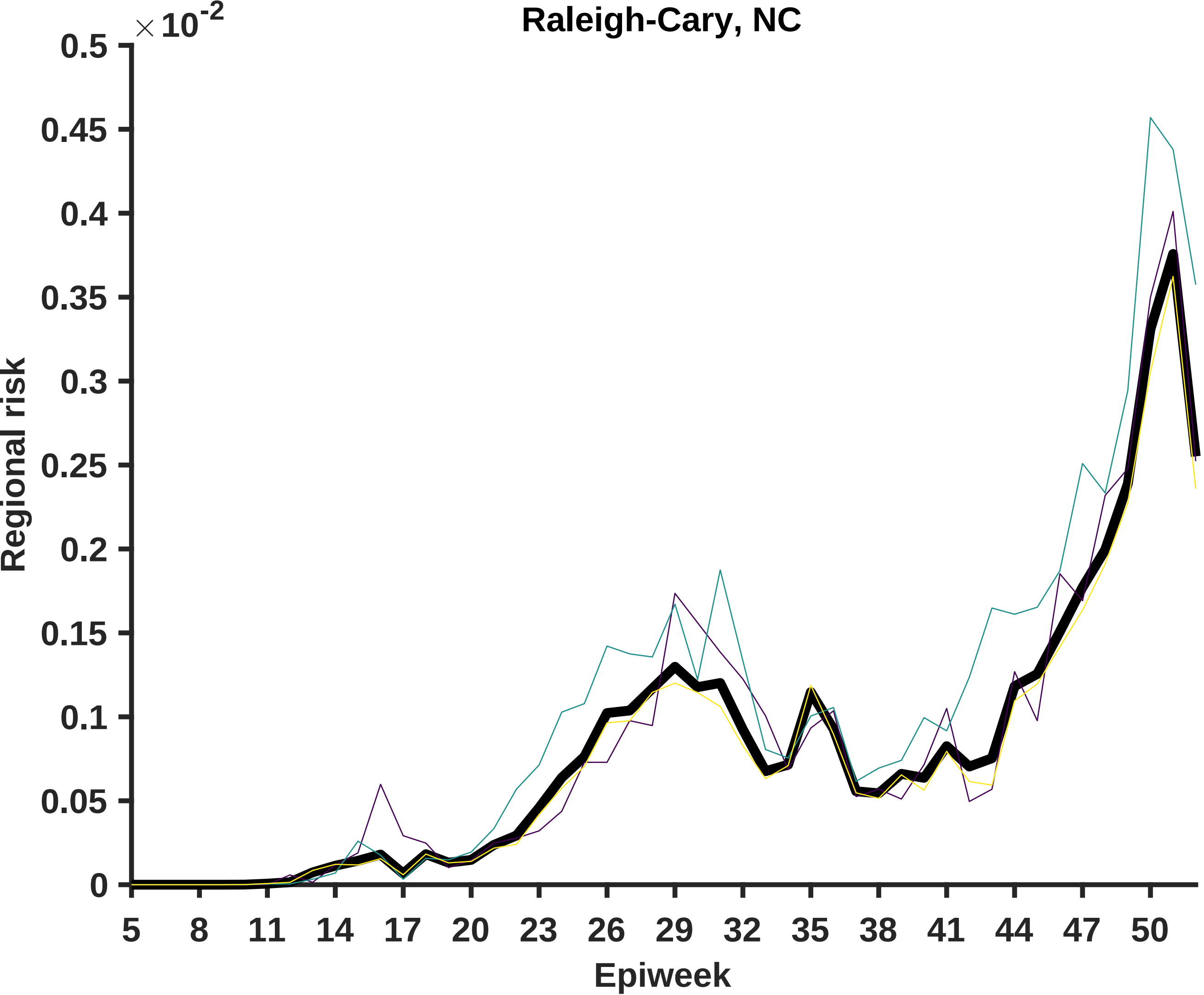

Raleigh-Cary, NC
×10^-2
0.5
0.45
0.4
0.35
0.3
0.25
0.2
0.15
0.1
0.05
0
Regional risk
5
8
11
14
17
20
23
26
29
32
35
38
41
44
47
50
Epiweek

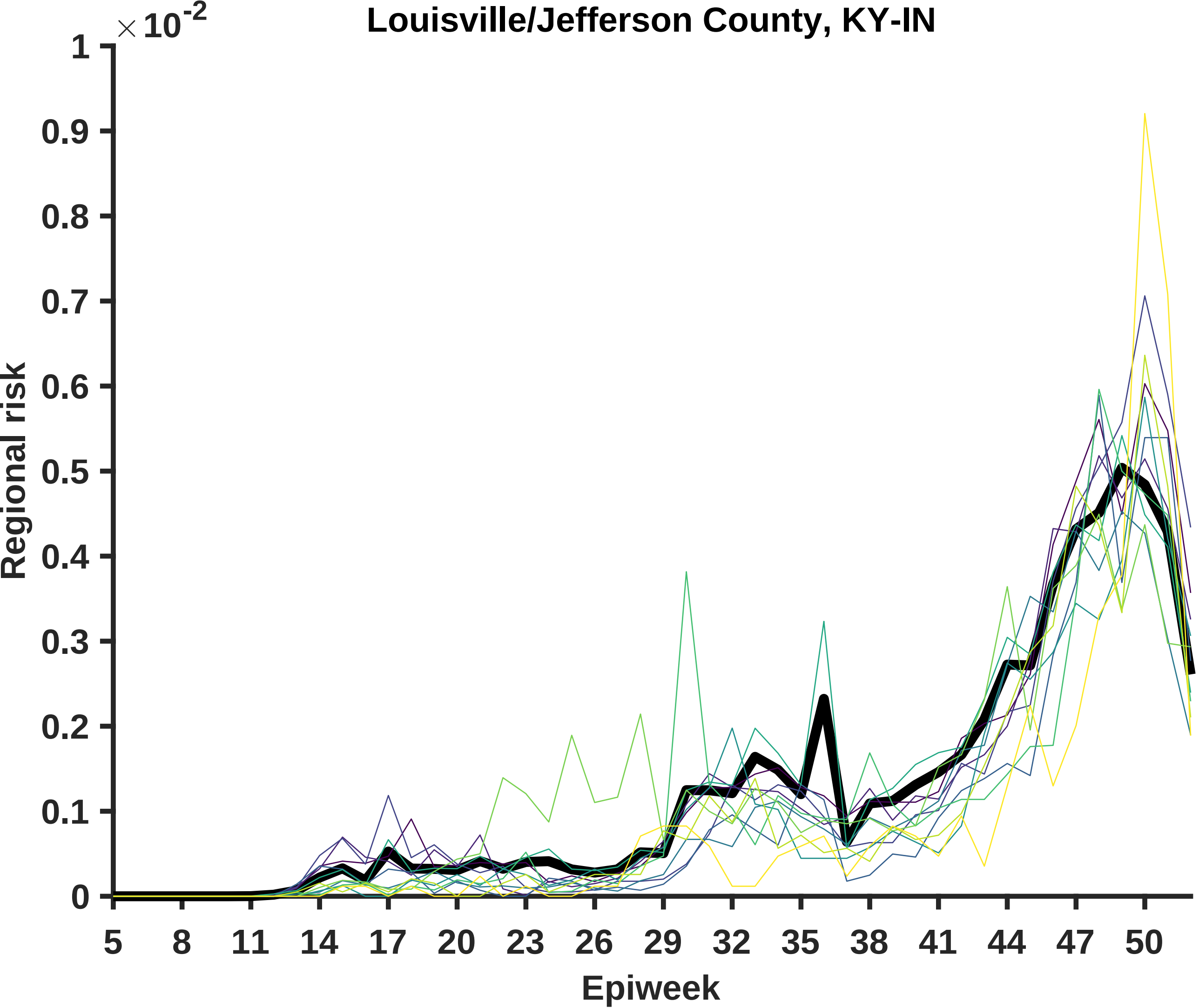

Louisville/Jefferson County, KY-IN
×10^-2
1
0.9
0.8
0.7
0.6
0.5
0.4
0.3
0.2
0.1
0
Regional risk
5
8
11
14
17
20
23
26
29
32
35
38
41
44
47
50
Epiweek

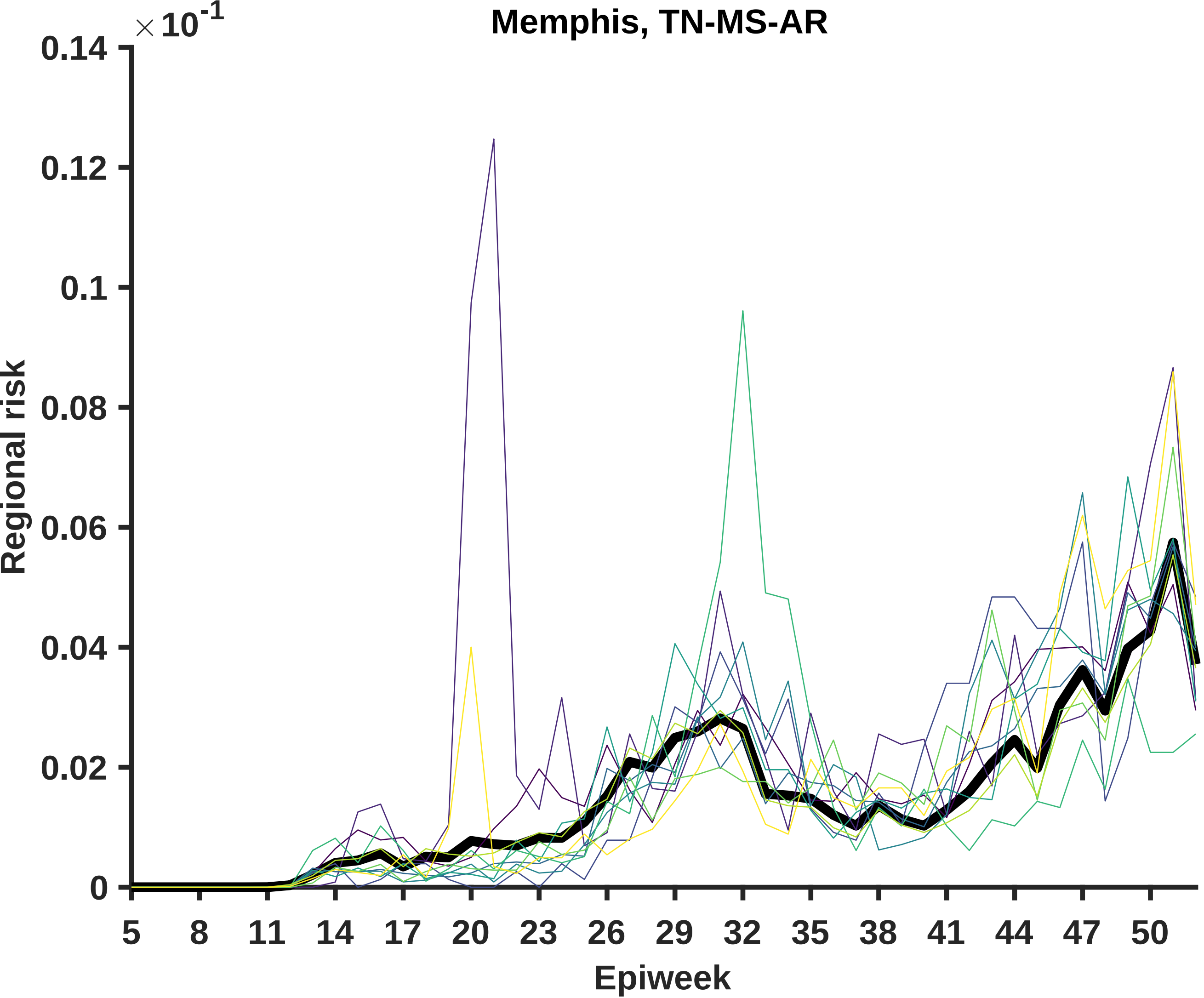
Memphis, TN-MS-AR
×10-1
0.14
0.12
0.1
0.08
0.06
0.04
0.02
0
Regional risk
5
8
11
14
17
20
23
26
29
32
35
38
41
44
47
50
Epiweek

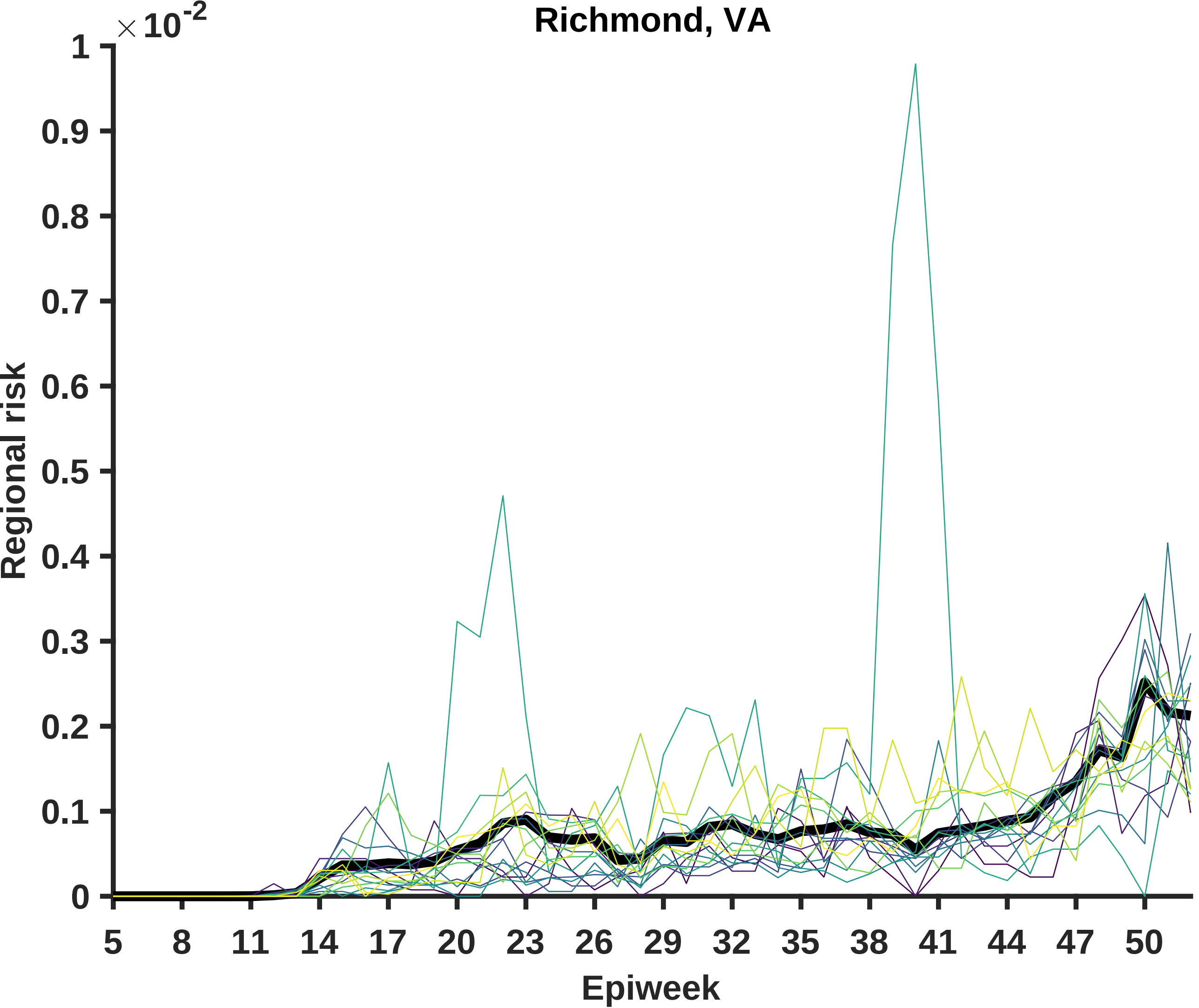

Richmond, VA
×10^-2
Regional risk
Epiweek
0
0.1
0.2
0.3
0.4
0.5
0.6
0.7
0.8
0.9
1
5
8
11
14
17
20
23
26
29
32
35
38
41
44
47
50

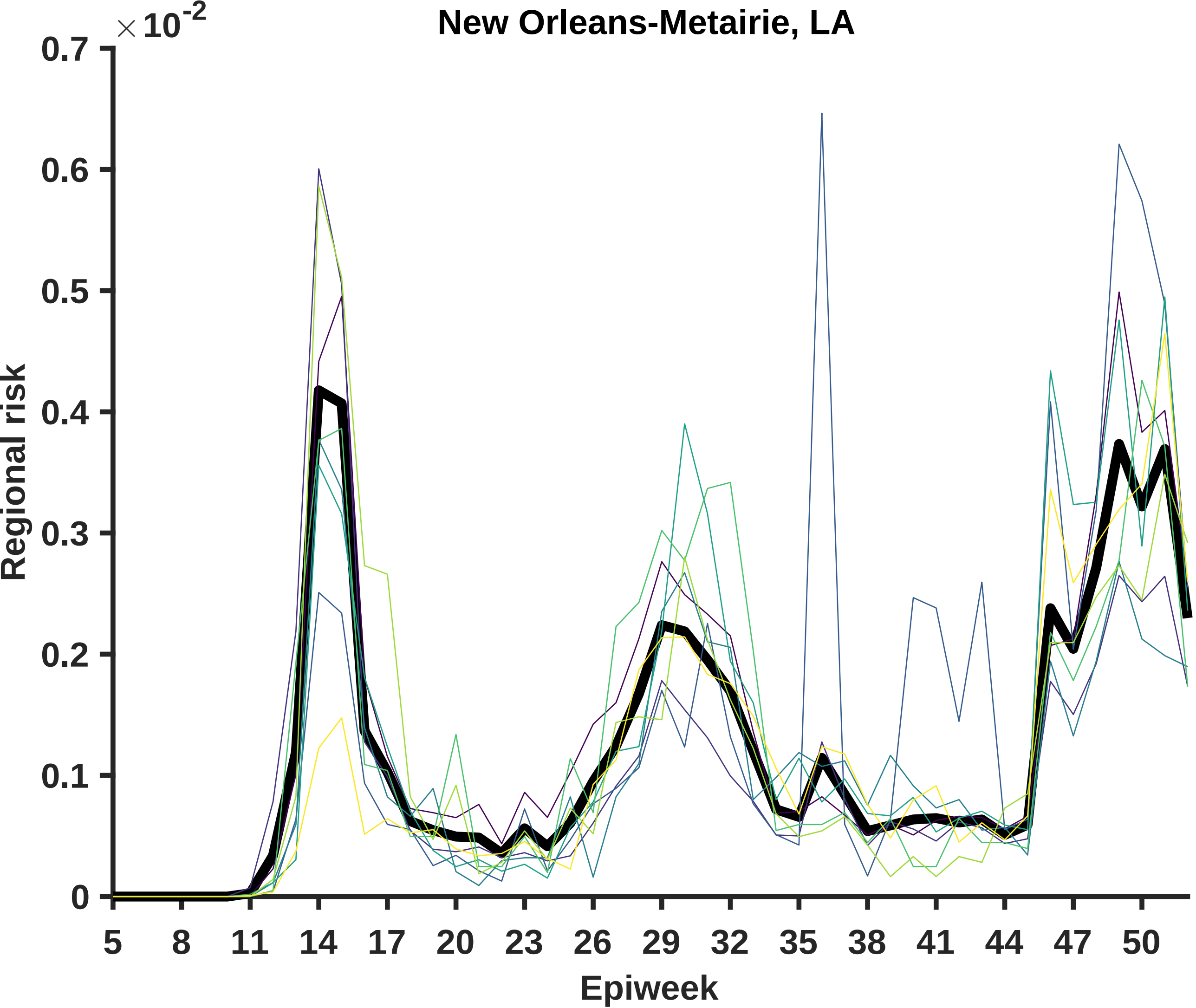
New Orleans-Metairie, LA
×10-2
0.7
0.6
0.5
0.4
0.3
0.2
0.1
0
Regional risk
5
8
11
14
17
20
23
26
29
32
35
38
41
44
47
50
Epiweek

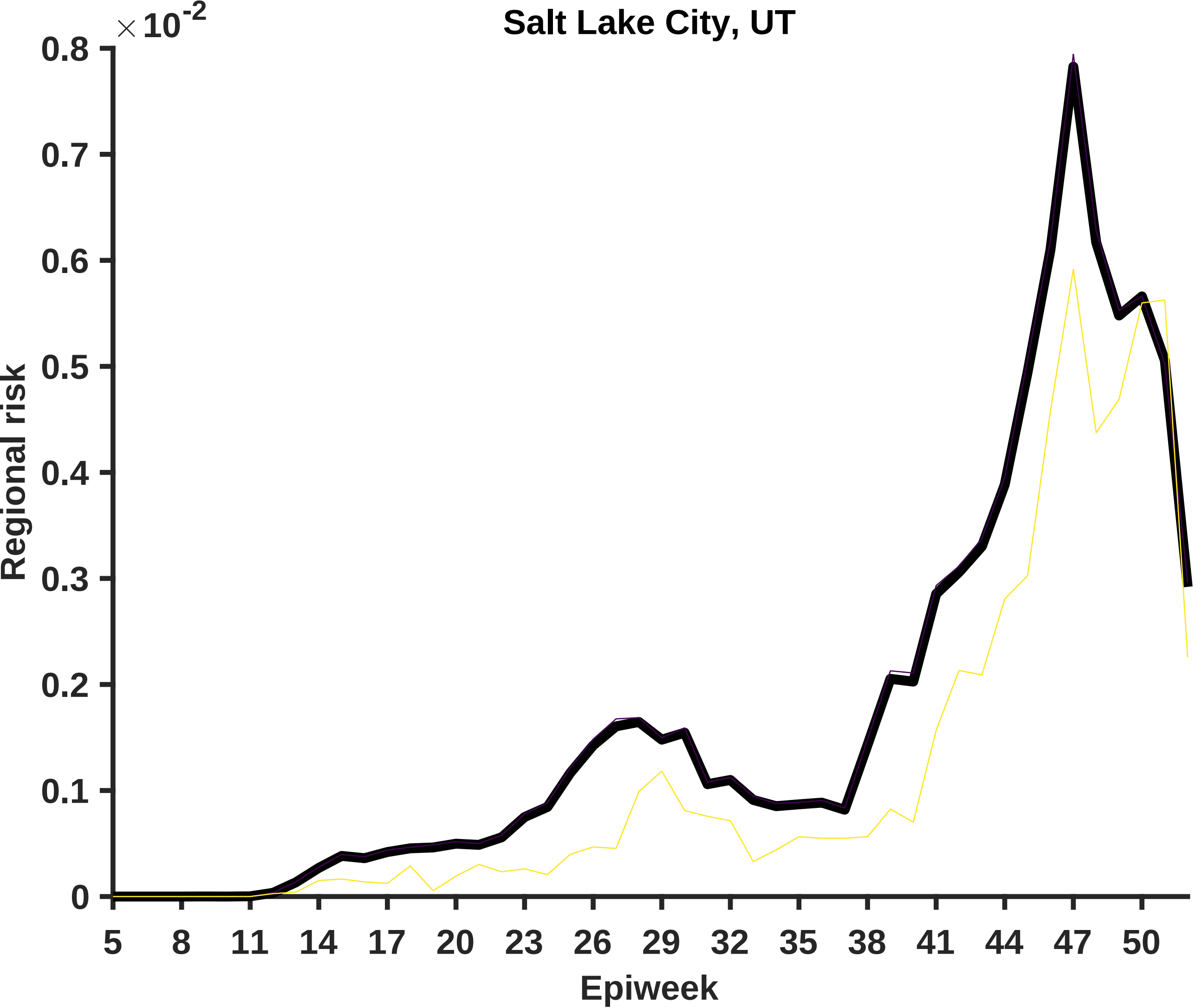

Salt Lake City, UT
×10^-2
0.8
0.7
0.6
0.5
0.4
0.3
0.2
0.1
0
Regional risk
5
8
11
14
17
20
23
26
29
32
35
38
41
44
47
50
Epiweek

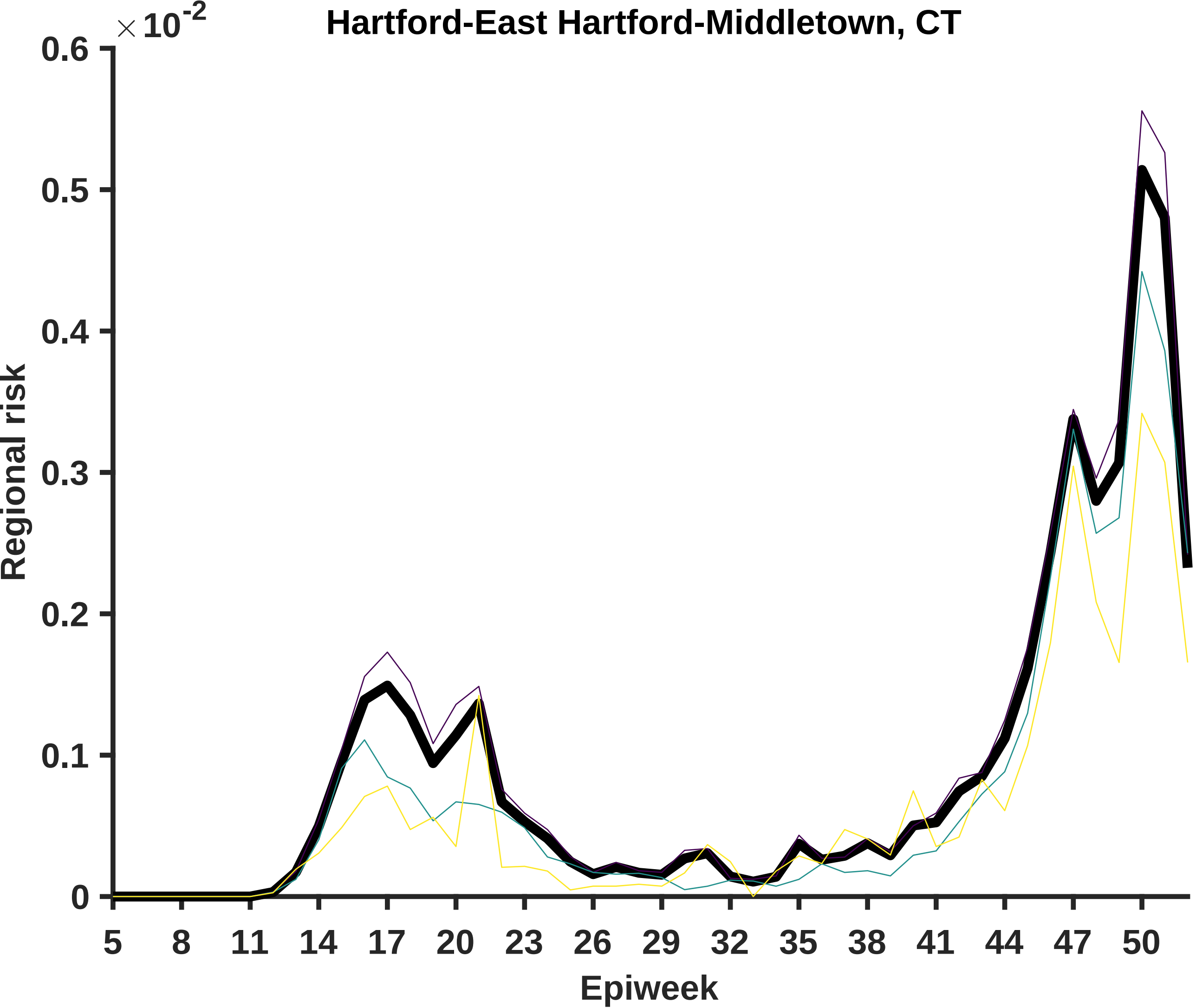

Hartford-East Hartford-Middletown, CT
×10^-2
0.6
0.5
0.4
0.3
0.2
0.1
0
Regional risk
5
8
11
14
17
20
23
26
29
32
35
38
41
44
47
50
Epiweek

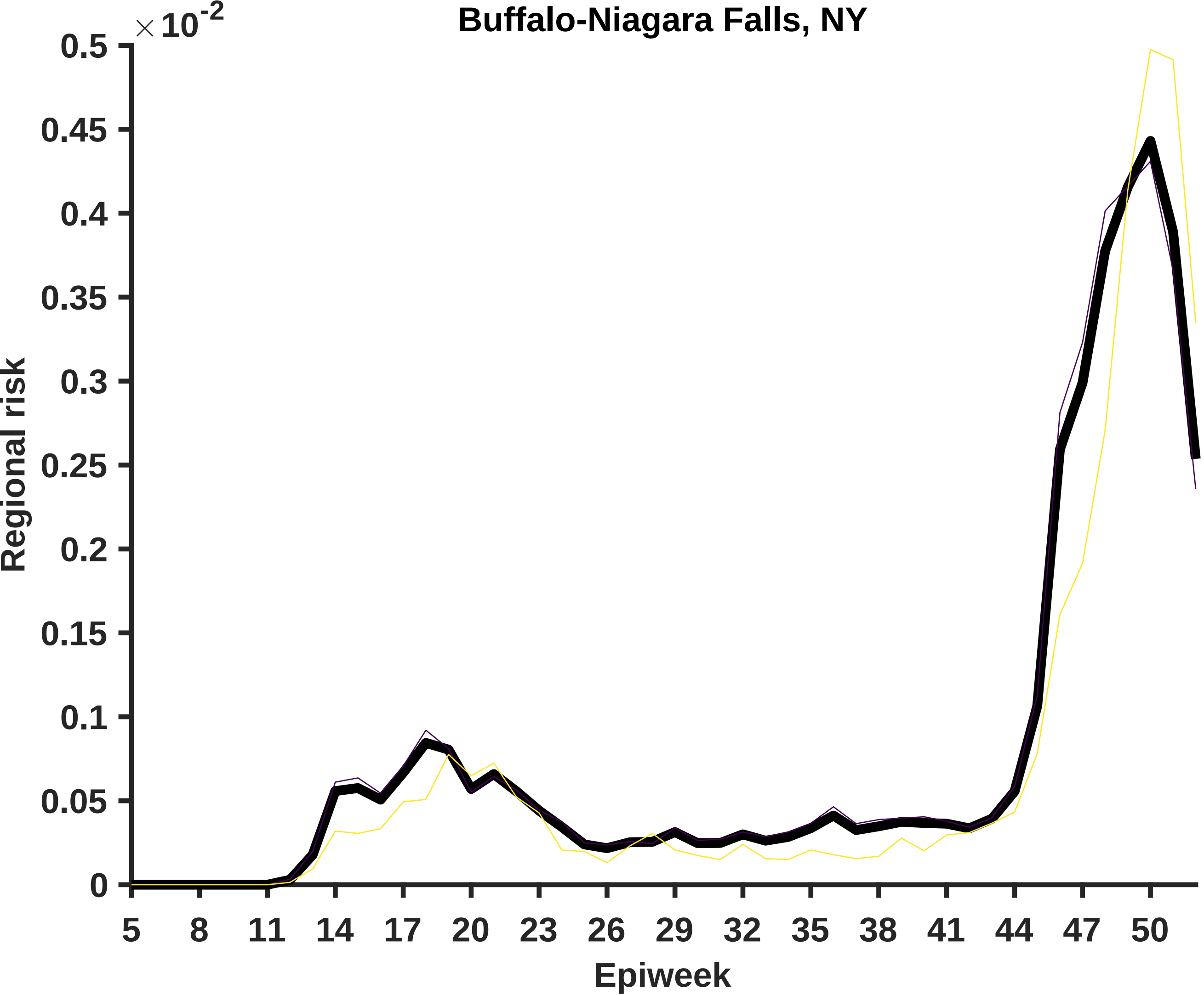

Buffalo-Niagara Falls, NY
×10-2
0.5
0.45
0.4
0.35
0.3
0.25
0.2
0.15
0.1
0.05
0
Regional risk
5
8
11
14
17
20
23
26
29
32
35
38
41
44
47
50
Epiweek

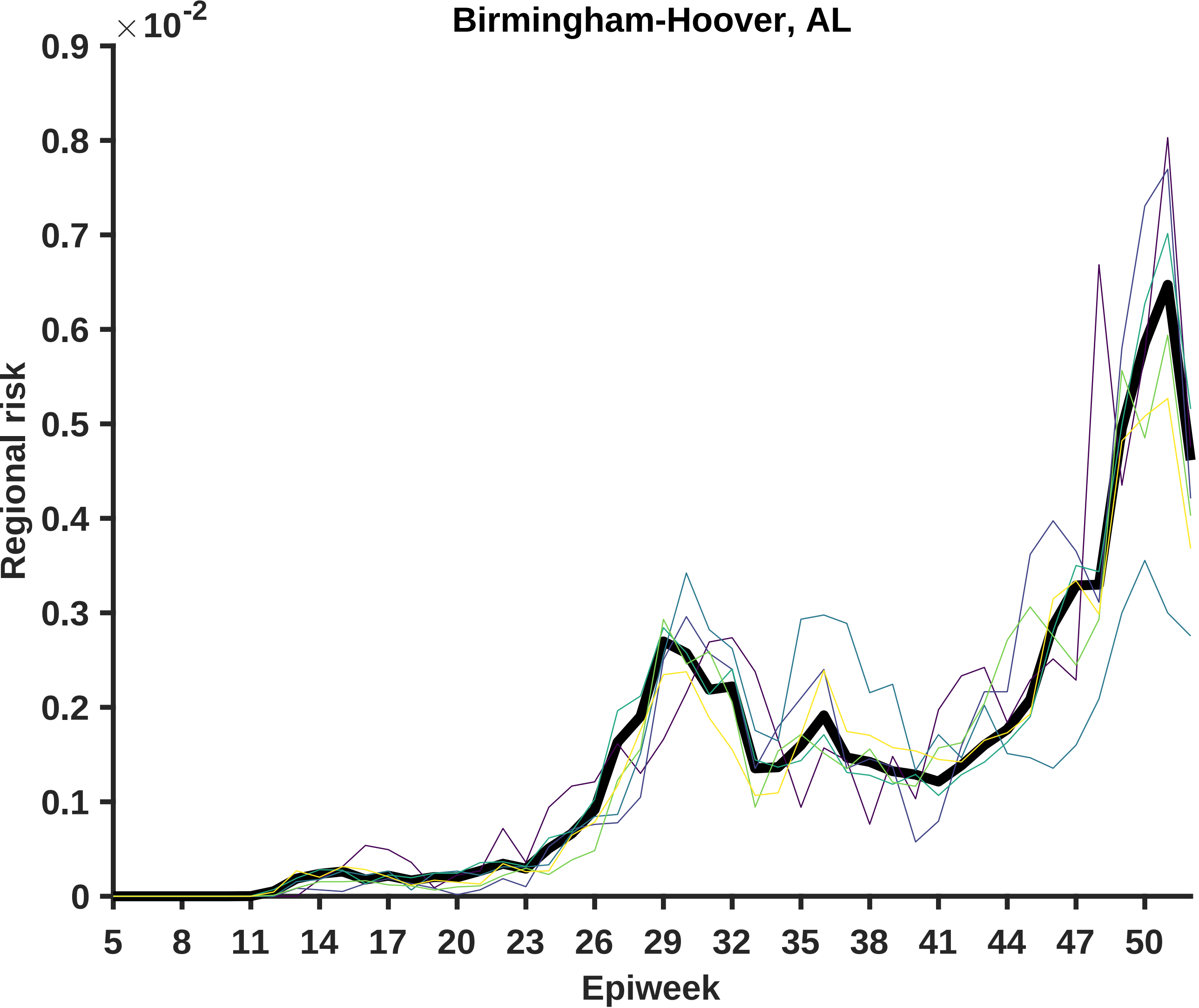
Birmingham-Hoover, AL
×10^-2
Regional risk
Epiweek
0.9
0.8
0.7
0.6
0.5
0.4
0.3
0.2
0.1
0
5
8
11
14
17
20
23
26
29
32
35
38
41
44
47
50

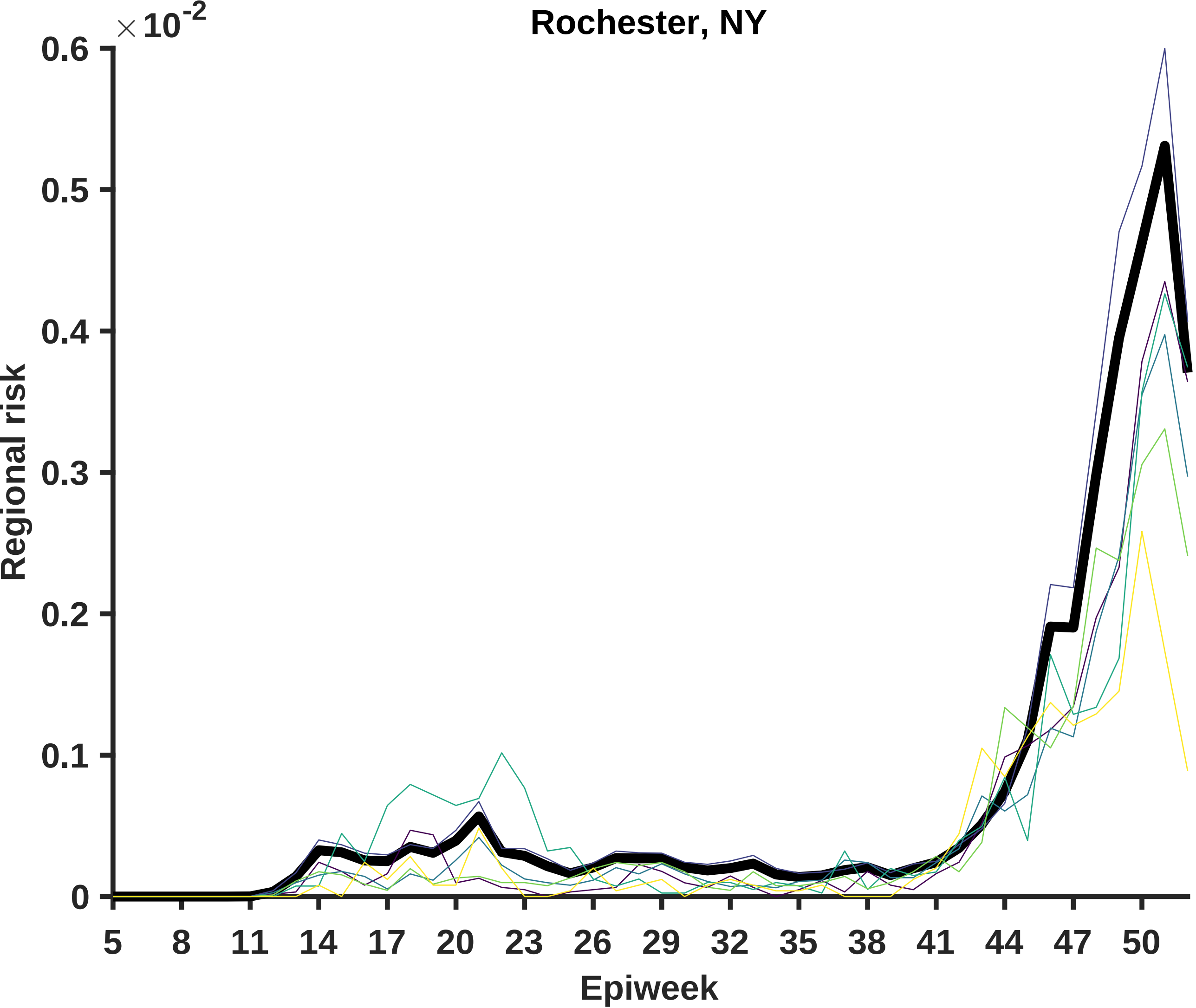

Rochester, NY
×10-2
0.6
0.5
0.4
0.3
0.2
0.1
0
Regional risk
5
8
11
14
17
20
23
26
29
32
35
38
41
44
47
50
Epiweek

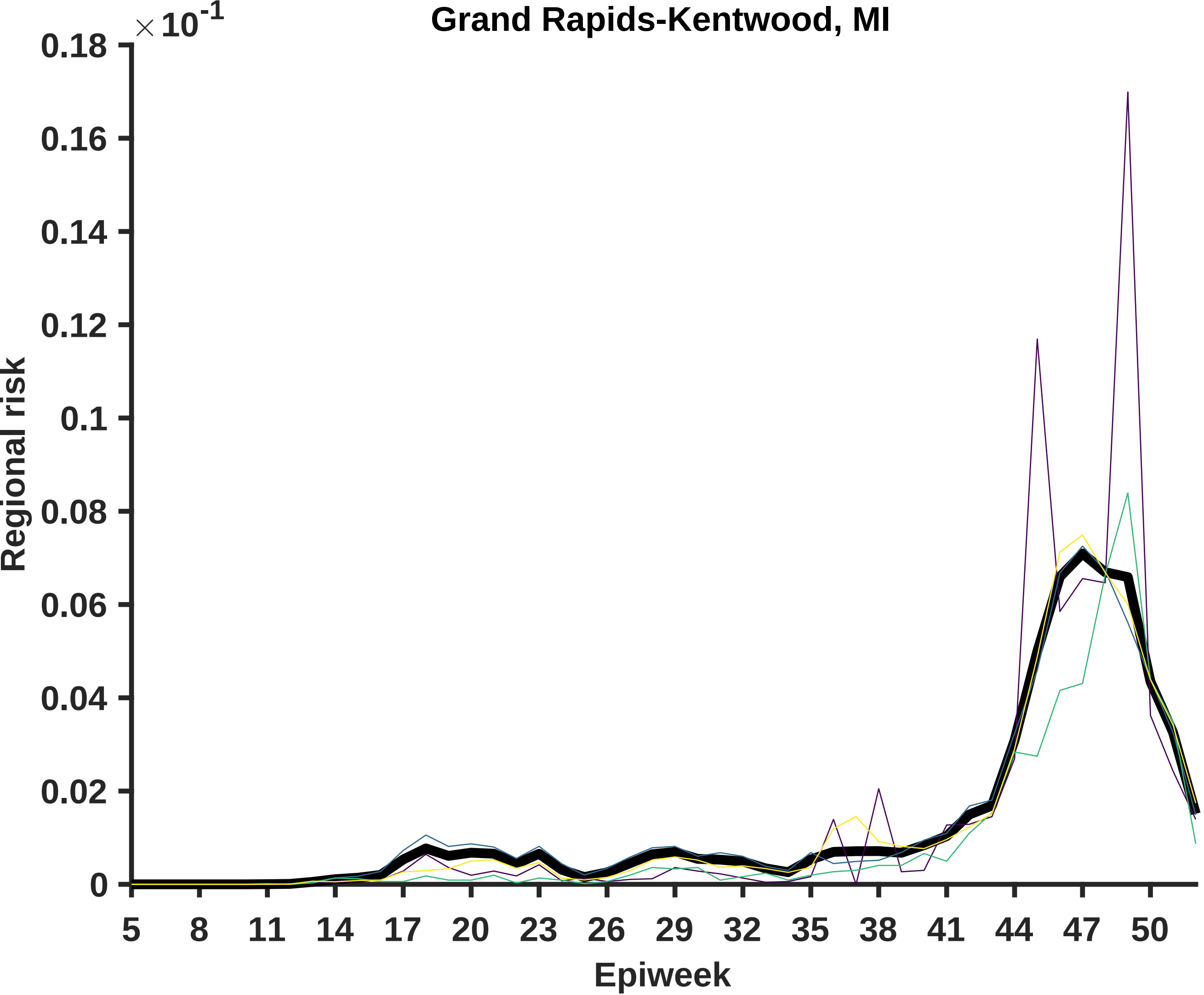

Grand Rapids-Kentwood, MI
× 10^-1
Regional risk
0.18
0.16
0.14
0.12
0.1
0.08
0.06
0.04
0.02
0
5
8
11
14
17
20
23
26
29
32
35
38
41
44
47
50
Epiweek

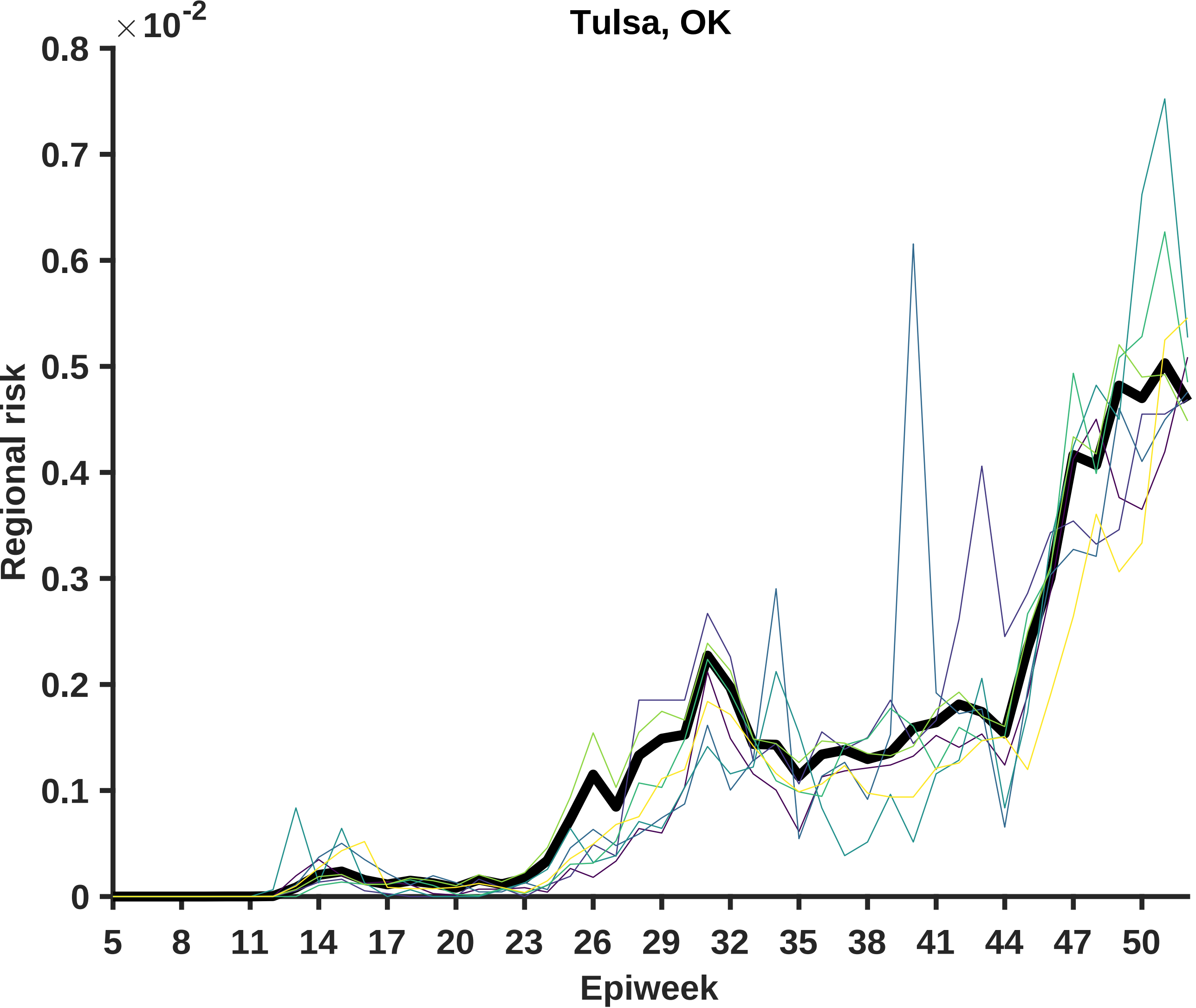

Tulsa, OK
×10^-2
0.8
0.7
0.6
0.5
0.4
0.3
0.2
0.1
0
Regional risk
5
8
11
14
17
20
23
26
29
32
35
38
41
44
47
50
Epiweek

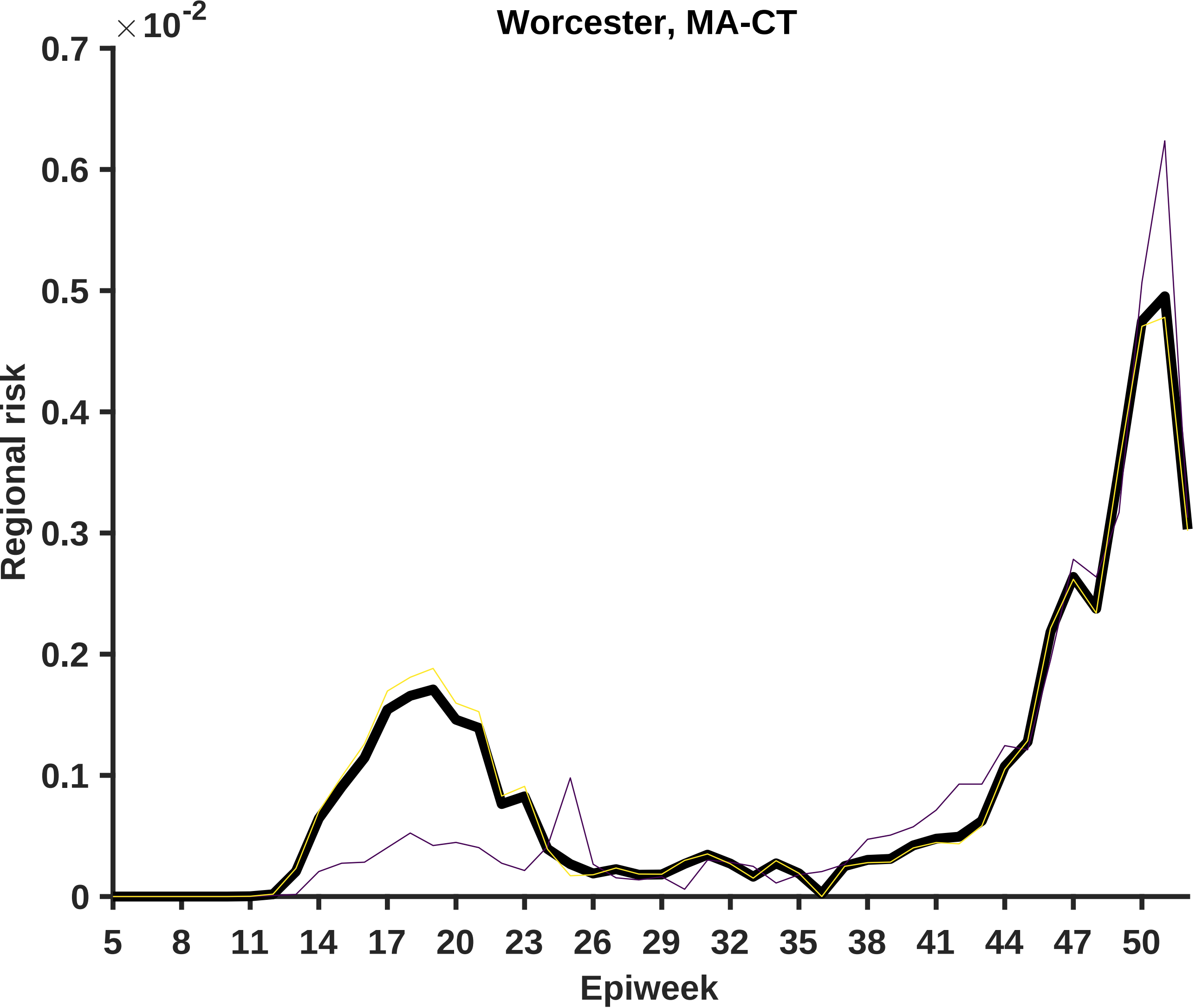
Worcester, MA-CT
×10^-2
0.7
0.6
0.5
0.4
0.3
0.2
0.1
0
Regional risk
5
8
11
14
17
20
23
26
29
32
35
38
41
44
47
50
Epiweek

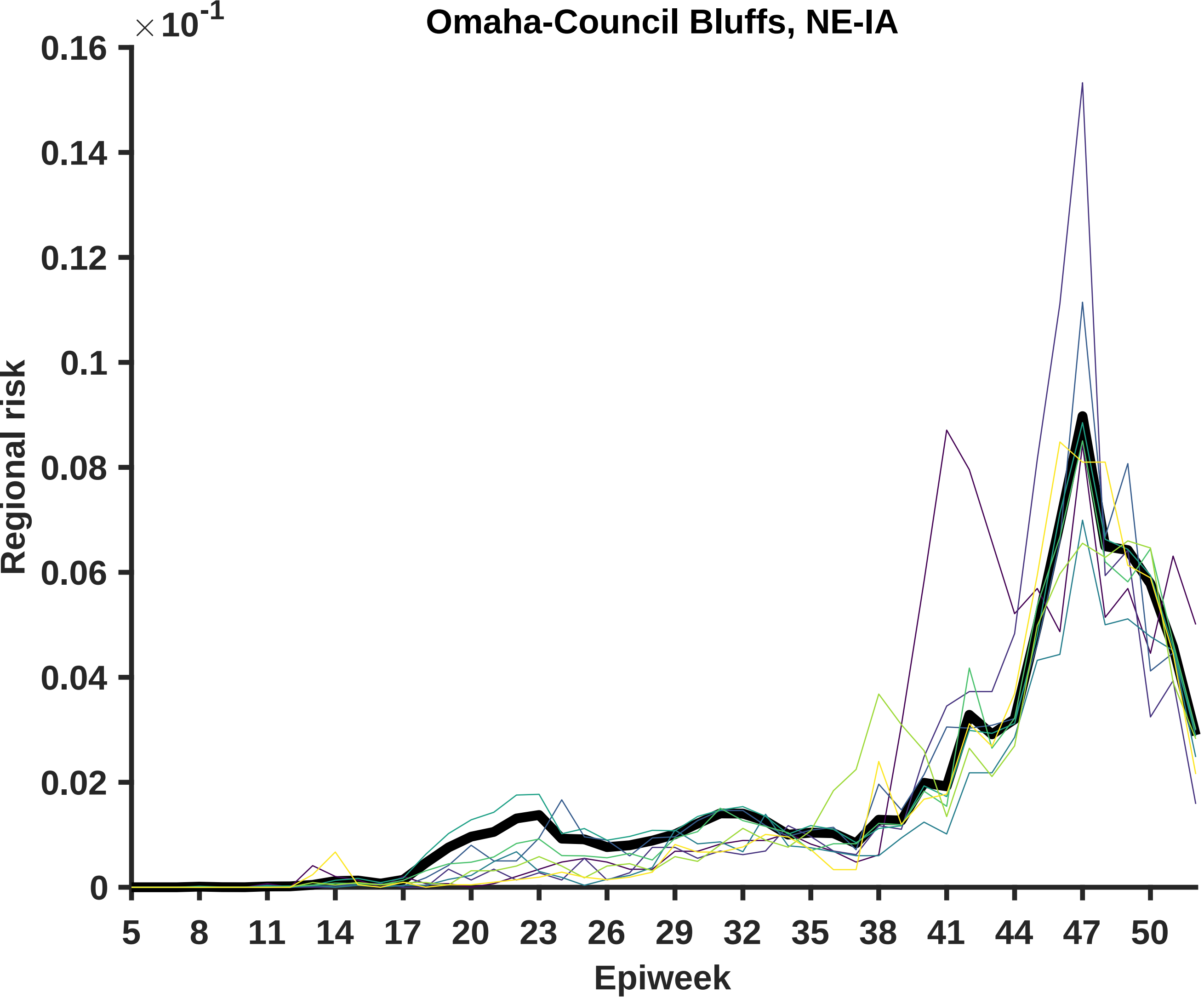

Omaha-Council Bluffs, NE-IA
×10-1
0.16
0.14
0.12
0.1
0.08
0.06
0.04
0.02
0
Regional risk
5
8
11
14
17
20
23
26
29
32
35
38
41
44
47
50
Epiweek

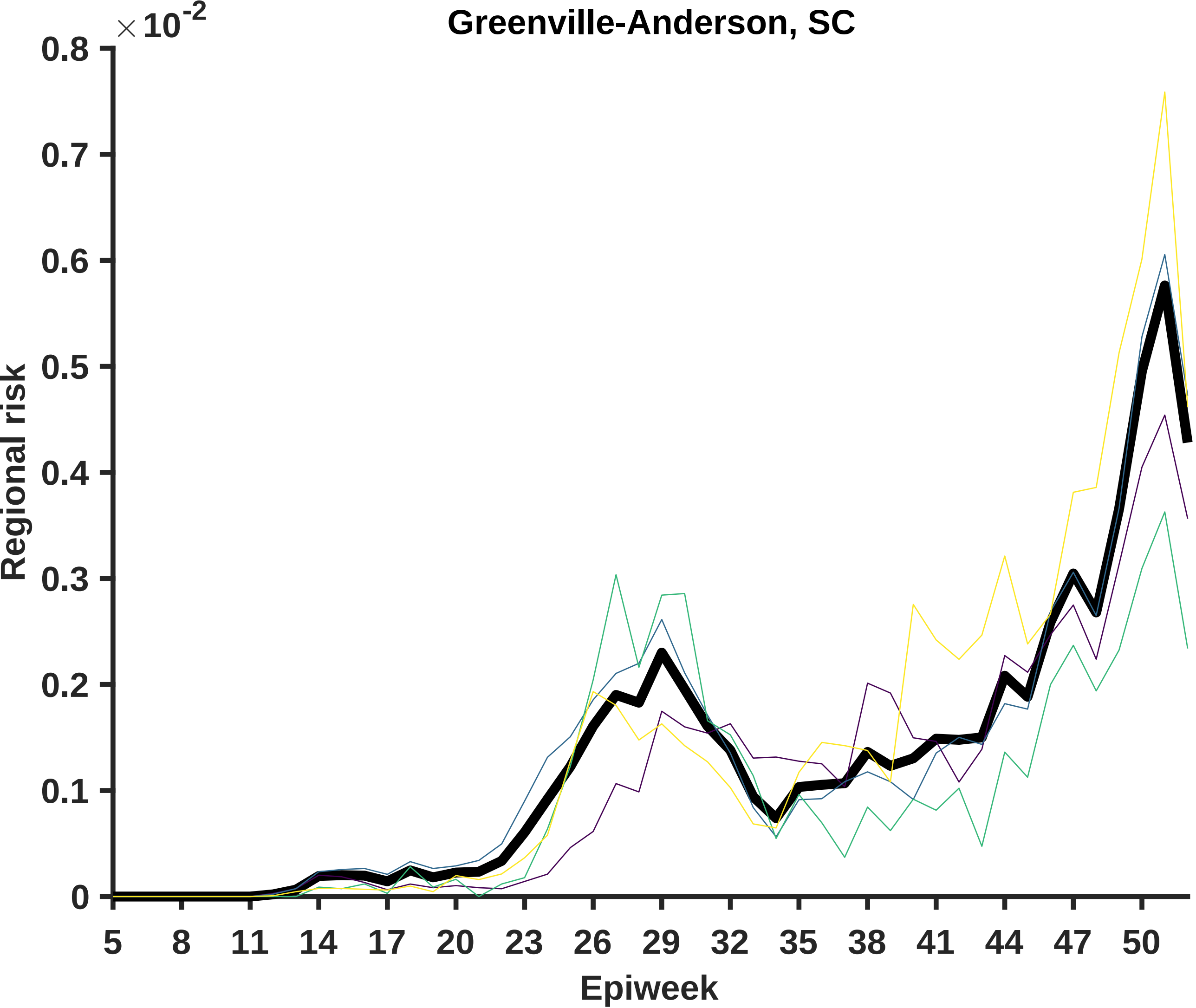
Greenville-Anderson, SC
×10^-2
0.8
0.7
0.6
0.5
0.4
0.3
0.2
0.1
0
Regional risk
5
8
11
14
17
20
23
26
29
32
35
38
41
44
47
50
Epiweek

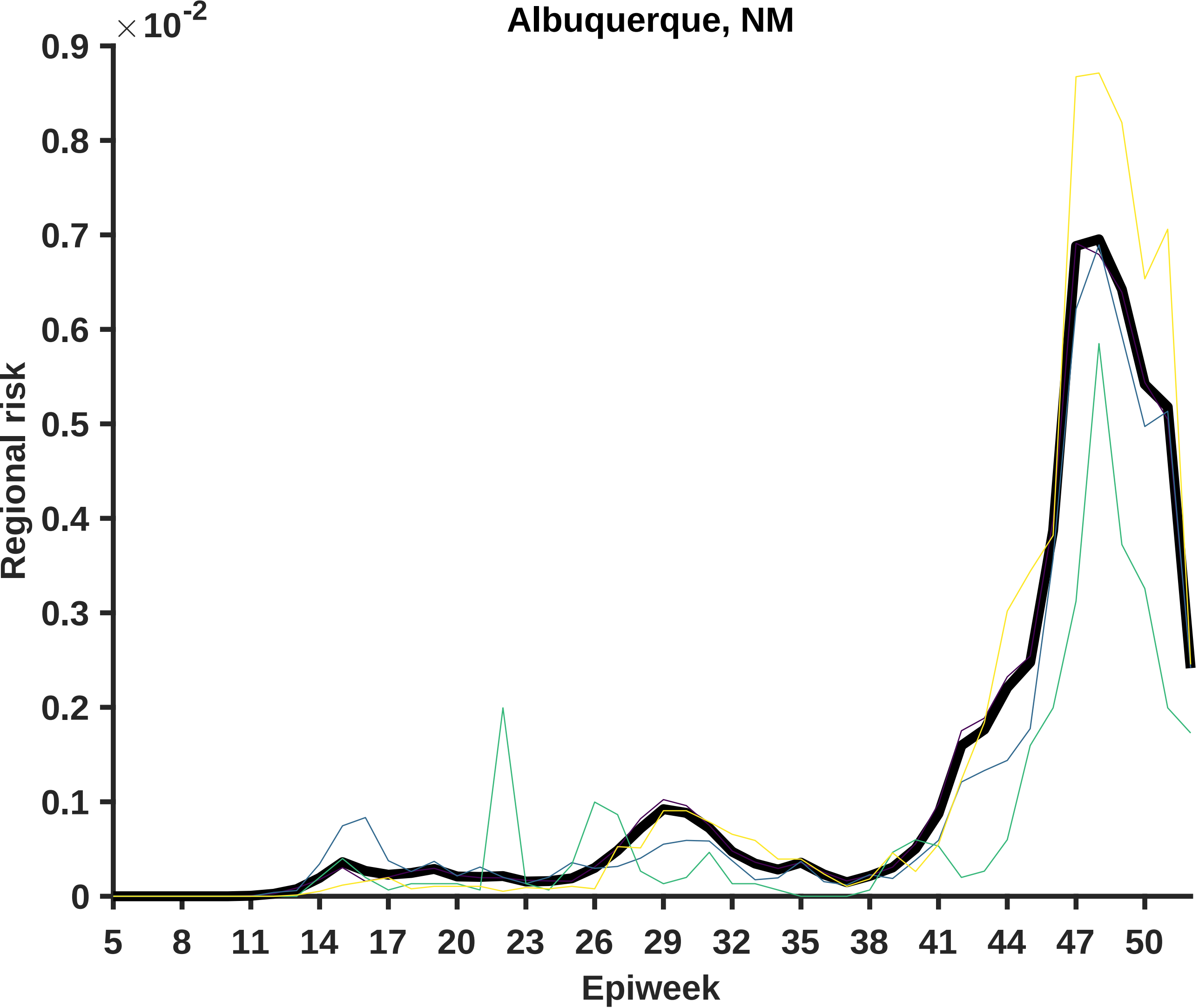

Albuquerque, NM
×10^-2
0.9
0.8
0.7
0.6
0.5
0.4
0.3
0.2
0.1
0
Regional risk
5
8
11
14
17
20
23
26
29
32
35
38
41
44
47
50
Epiweek

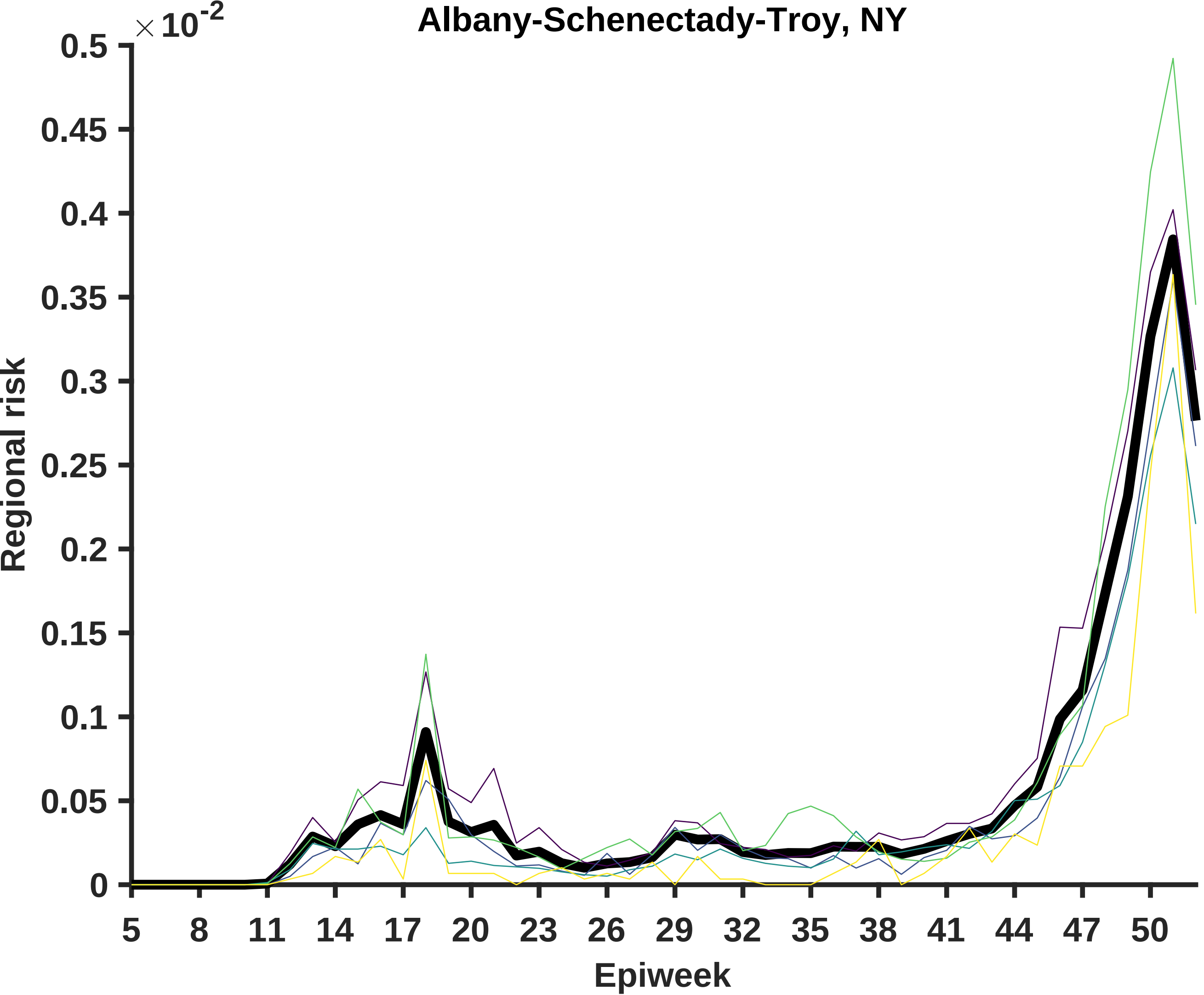

Albany-Schenectady-Troy, NY
× 10^-2
0.5
0.45
0.4
0.35
0.3
0.25
0.2
0.15
0.1
0.05
0
Regional risk
5
8
11
14
17
20
23
26
29
32
35
38
41
44
47
50
Epiweek

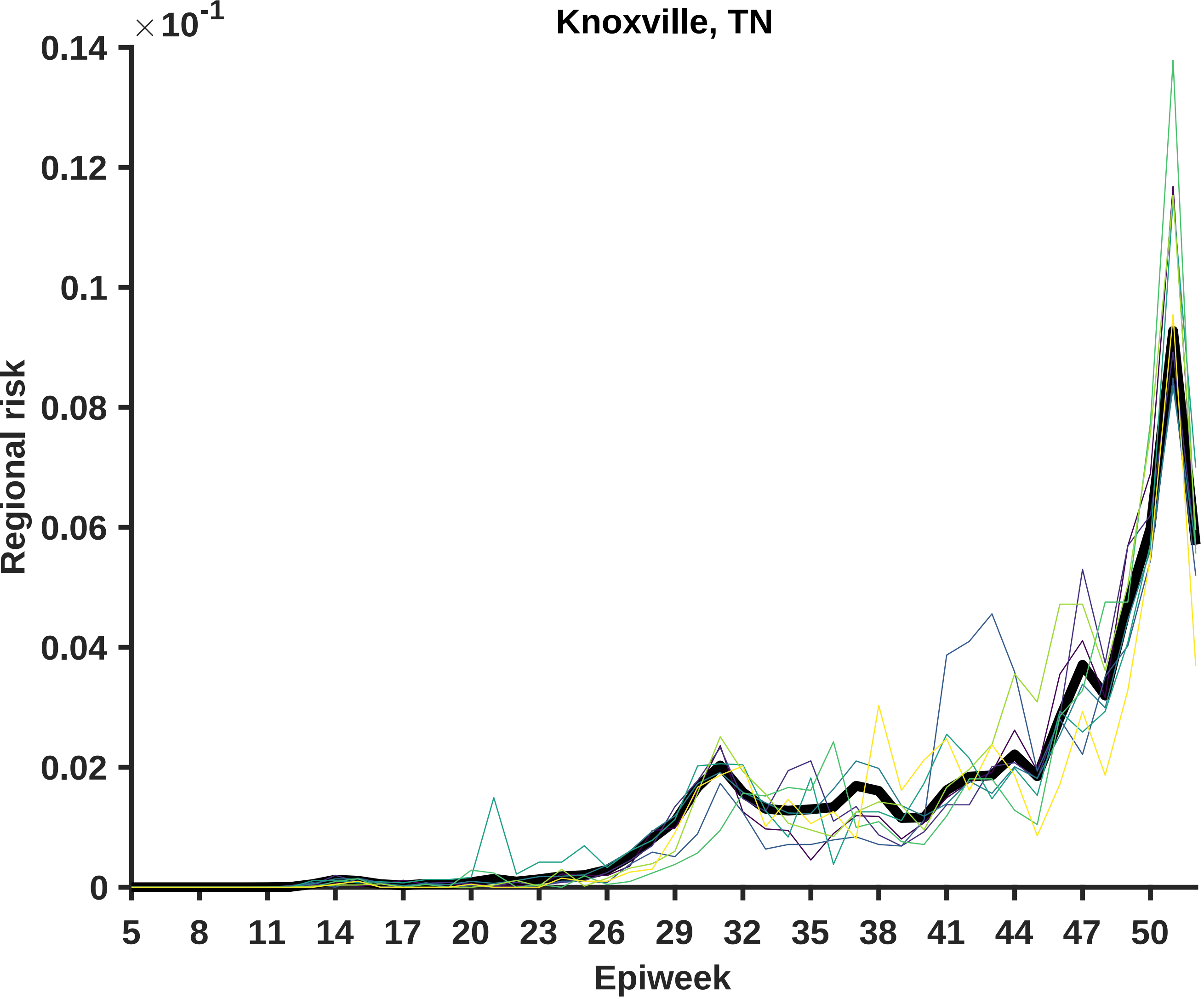
Knoxville, TN
×10^-1
0.14
0.12
0.1
0.08
0.06
0.04
0.02
0
Regional risk
5
8
11
14
17
20
23
26
29
32
35
38
41
44
47
50
Epiweek

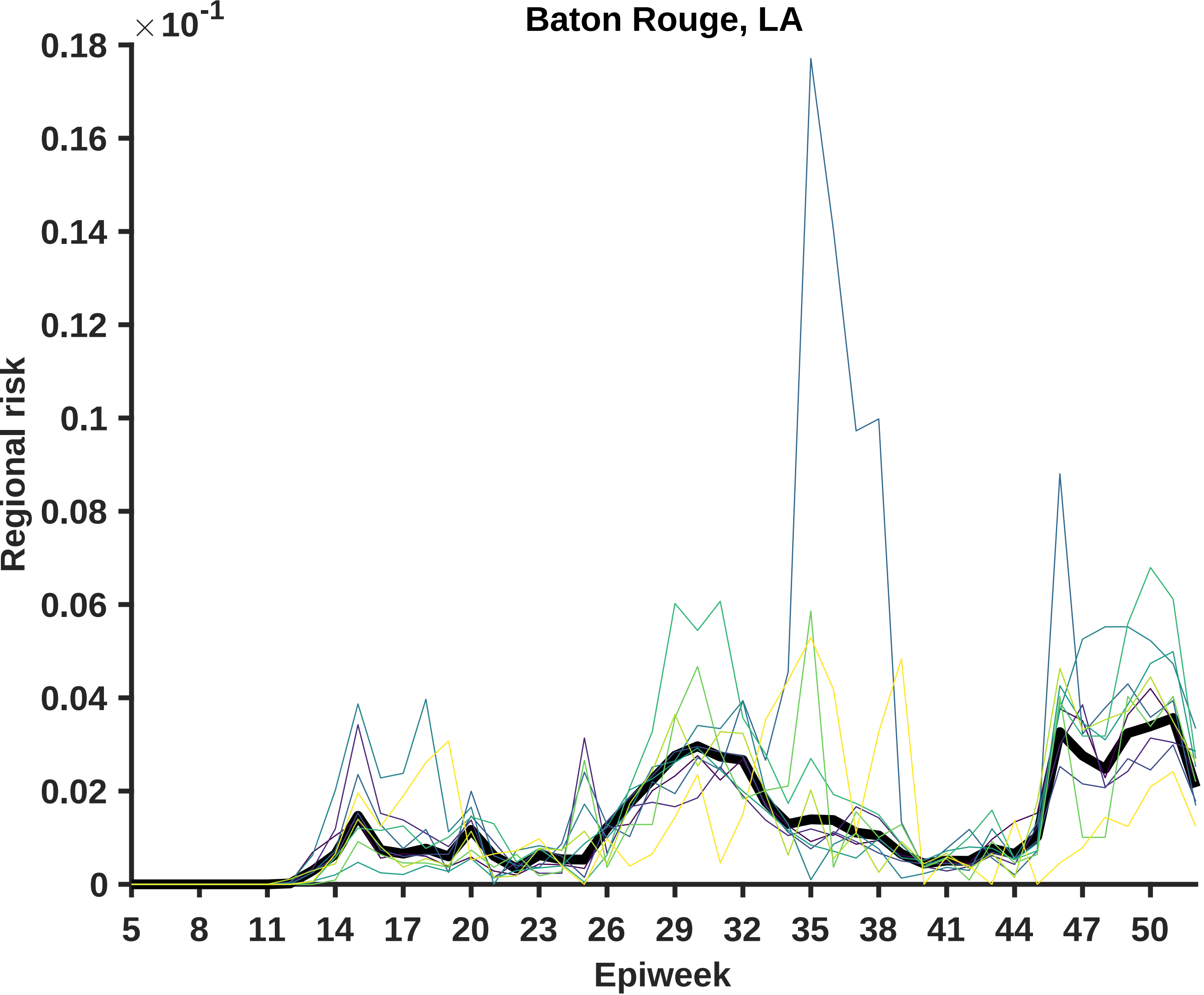

Baton Rouge, LA
× 10^-1
0.18
0.16
0.14
0.12
0.1
0.08
0.06
0.04
0.02
0
Regional risk
5
8
11
14
17
20
23
26
29
32
35
38
41
44
47
50
Epiweek

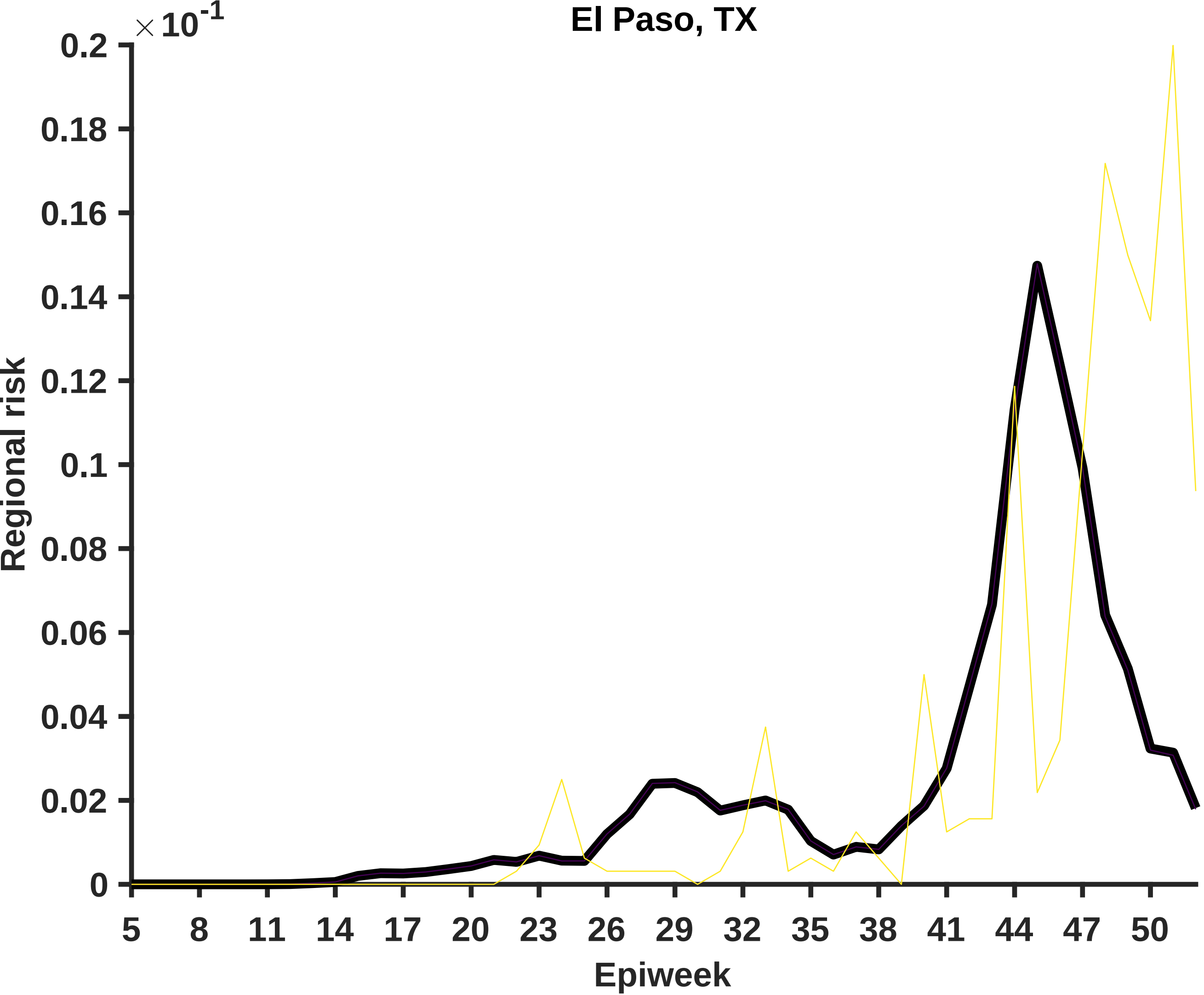

El Paso, TX
× 10^-1
0.2
0.18
0.16
0.14
0.12
0.1
0.08
0.06
0.04
0.02
0
Regional risk
5
8
11
14
17
20
23
26
29
32
35
38
41
44
47
50
Epiweek

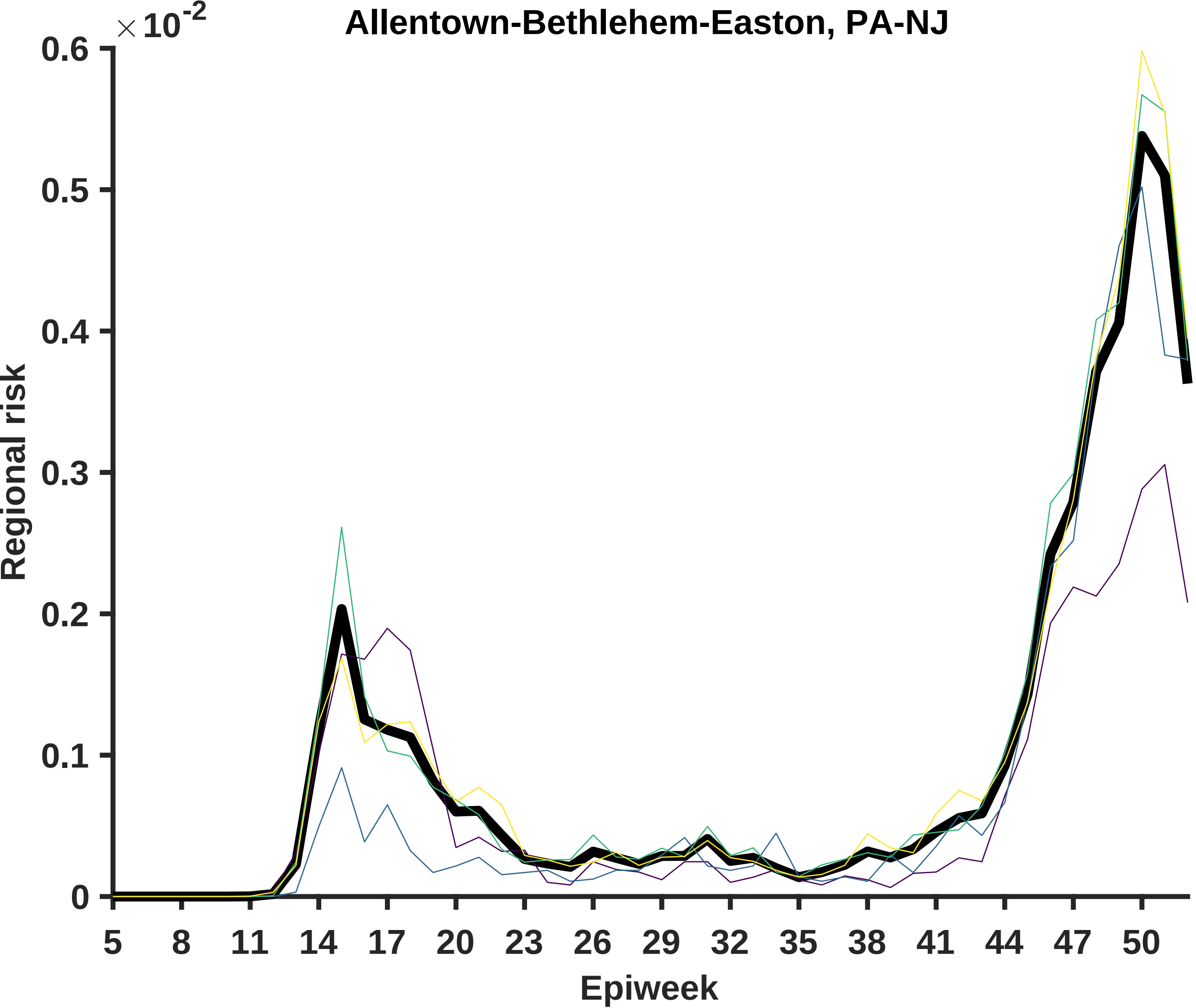

Allentown-Bethlehem-Easton, PA-NJ
×10^-2
0.6
0.5
0.4
0.3
0.2
0.1
0
Regional risk
5
8
11
14
17
20
23
26
29
32
35
38
41
44
47
50
Epiweek

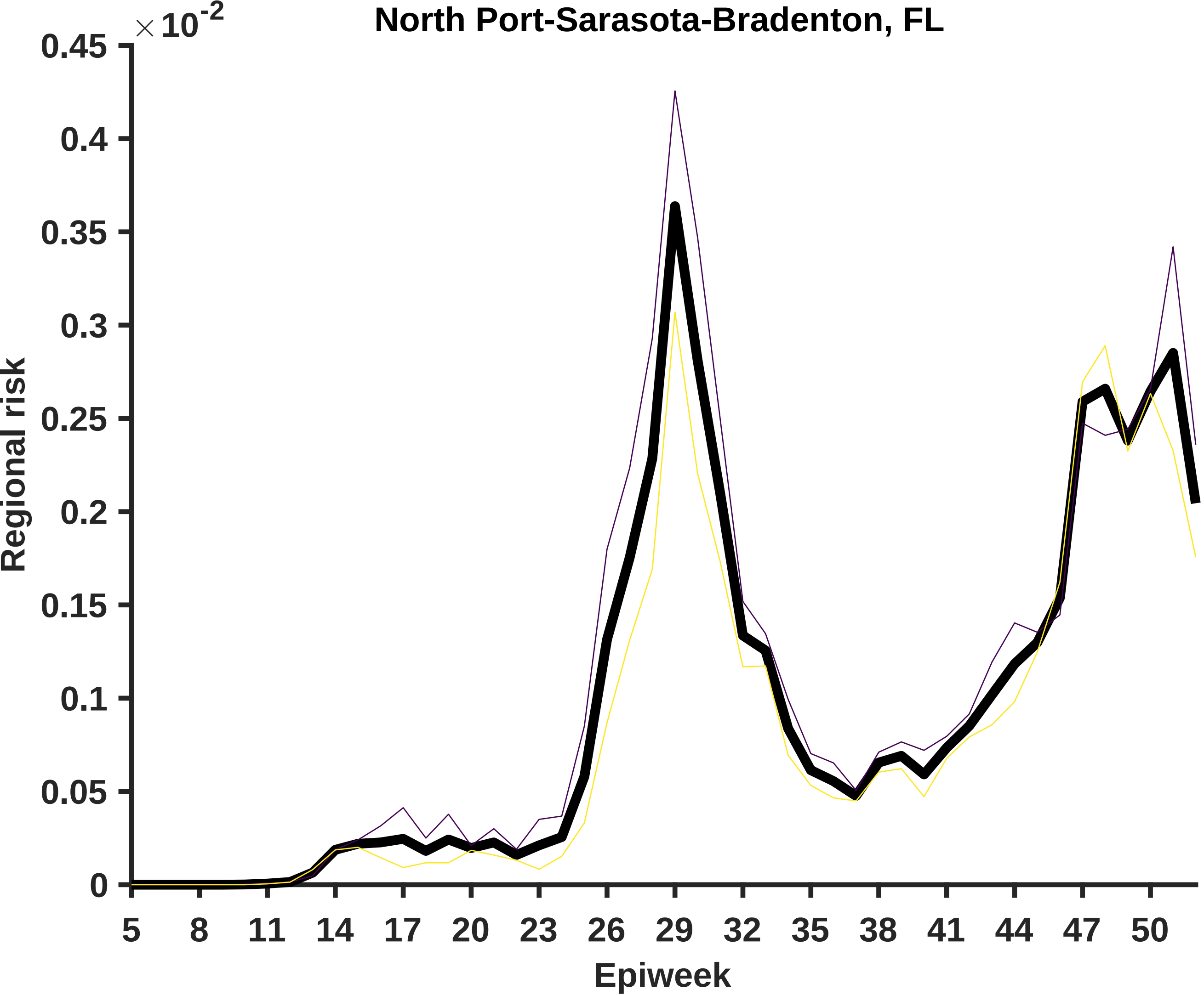

North Port-Sarasota-Bradenton, FL
× 10^-2
0.45
0.4
0.35
0.3
0.25
0.2
0.15
0.1
0.05
0
Regional risk
5
8
11
14
17
20
23
26
29
32
35
38
41
44
47
50
Epiweek

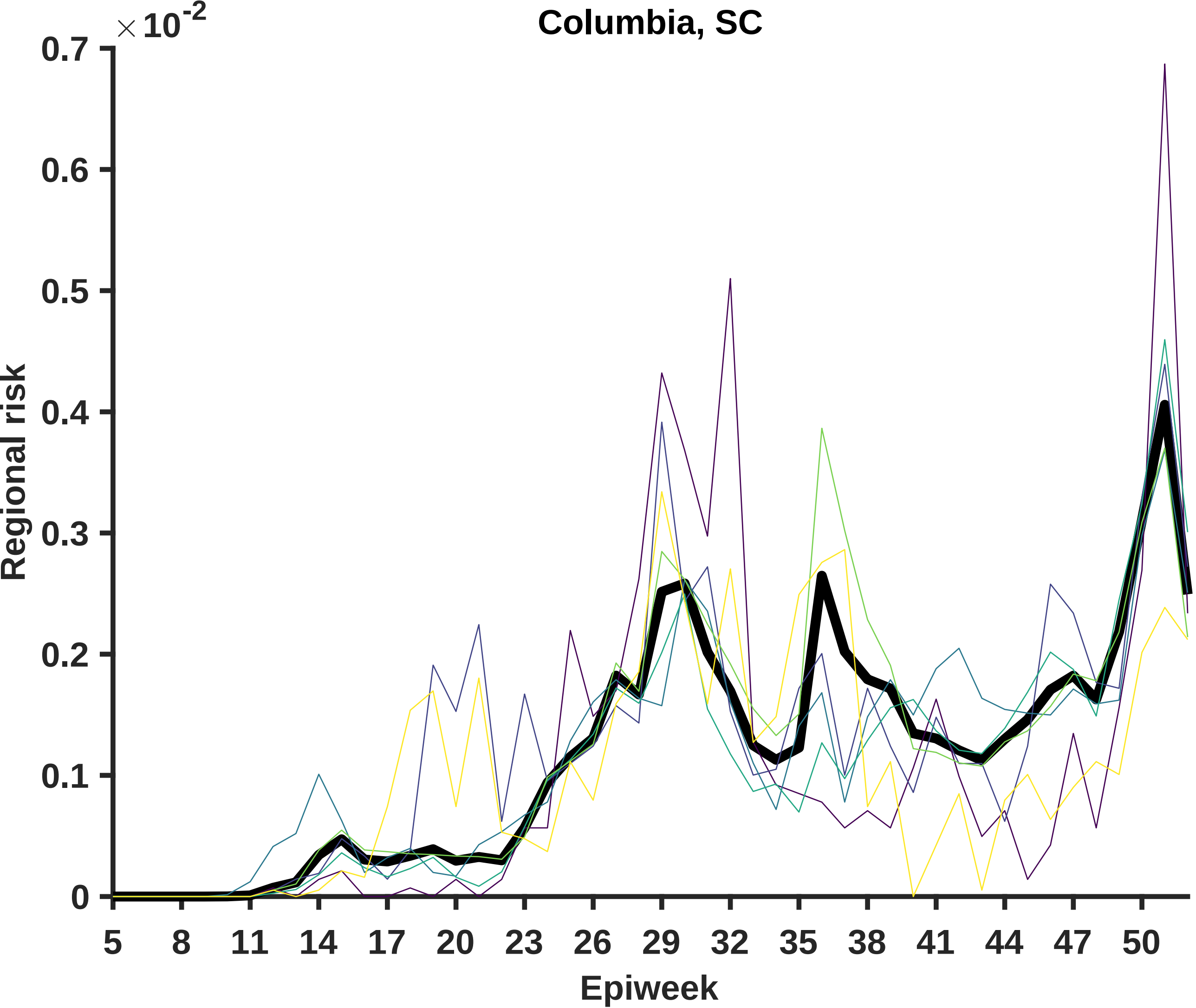

Columbia, SC
×10^-2
0.7
0.6
0.5
0.4
0.3
0.2
0.1
0
Regional risk
5
8
11
14
17
20
23
26
29
32
35
38
41
44
47
50
Epiweek

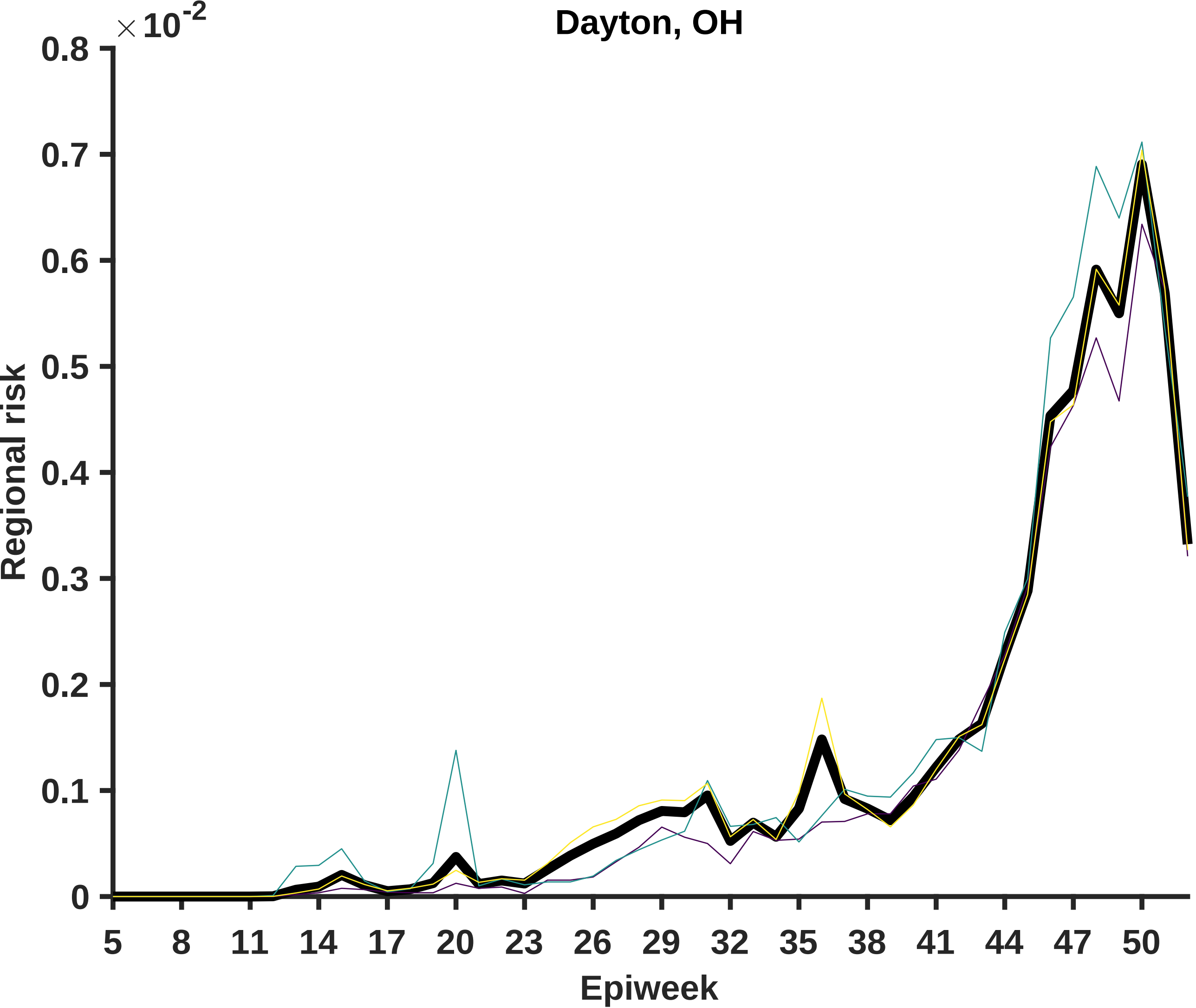

Dayton, OH
×10^-2
0.8
0.7
0.6
0.5
0.4
0.3
0.2
0.1
0
Regional risk
5
8
11
14
17
20
23
26
29
32
35
38
41
44
47
50
Epiweek

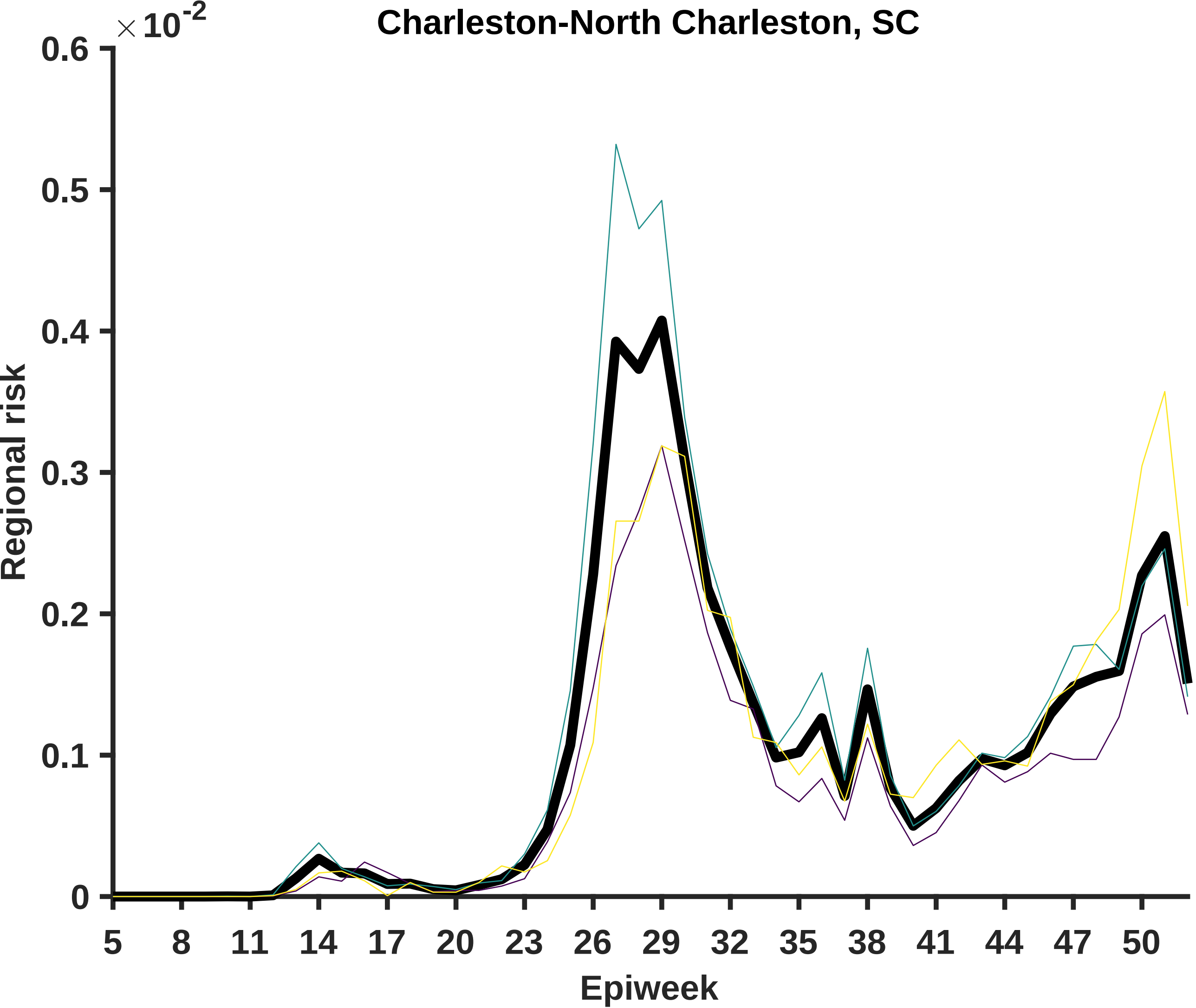

Charleston-North Charleston, SC
×10^-2
0.6
0.5
0.4
0.3
0.2
0.1
0
Regional risk
5
8
11
14
17
20
23
26
29
32
35
38
41
44
47
50
Epiweek

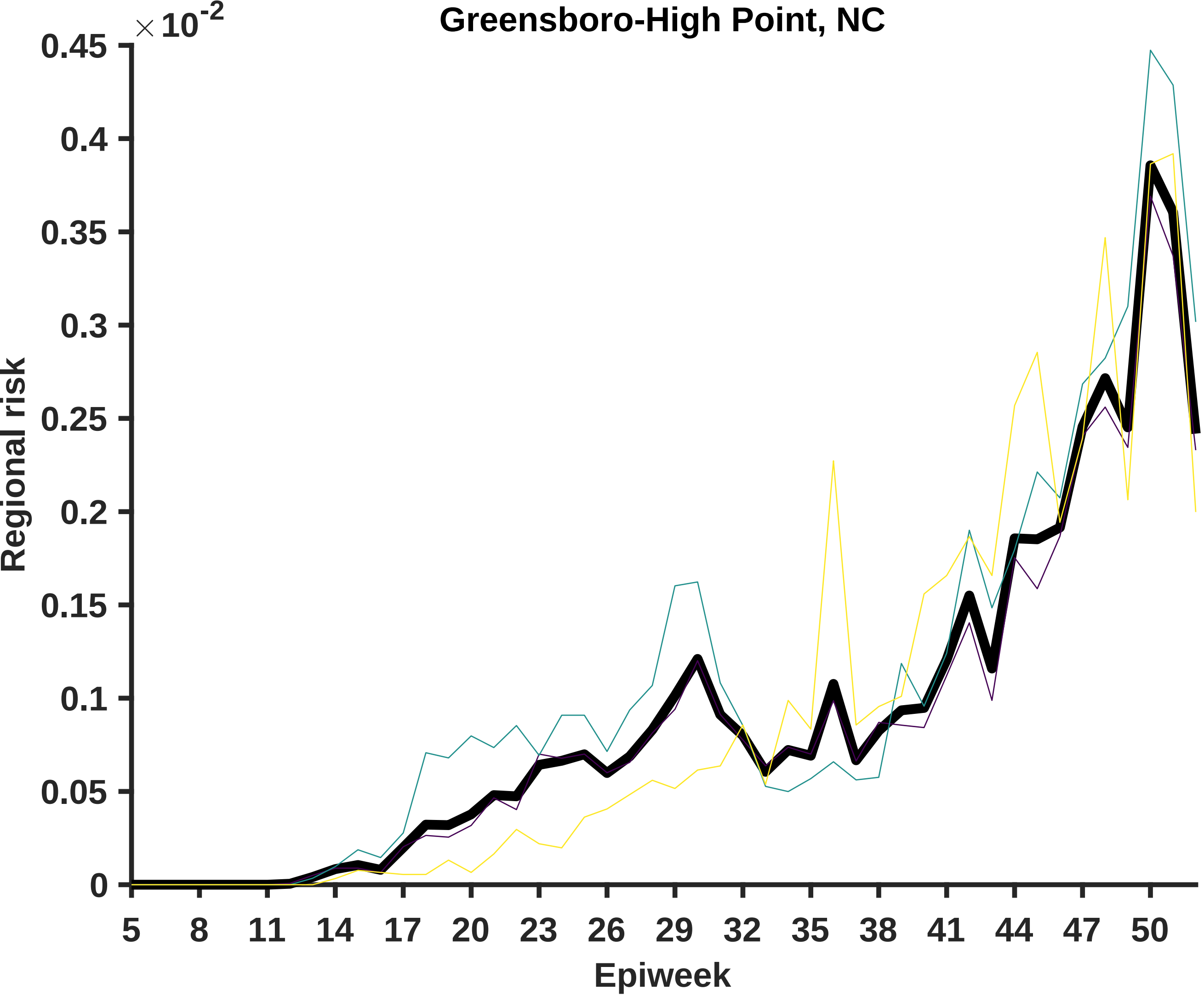

Greensboro-High Point, NC
× 10^-2
0.45
0.4
0.35
0.3
0.25
0.2
0.15
0.1
0.05
0
Regional risk
5
8
11
14
17
20
23
26
29
32
35
38
41
44
47
50
Epiweek

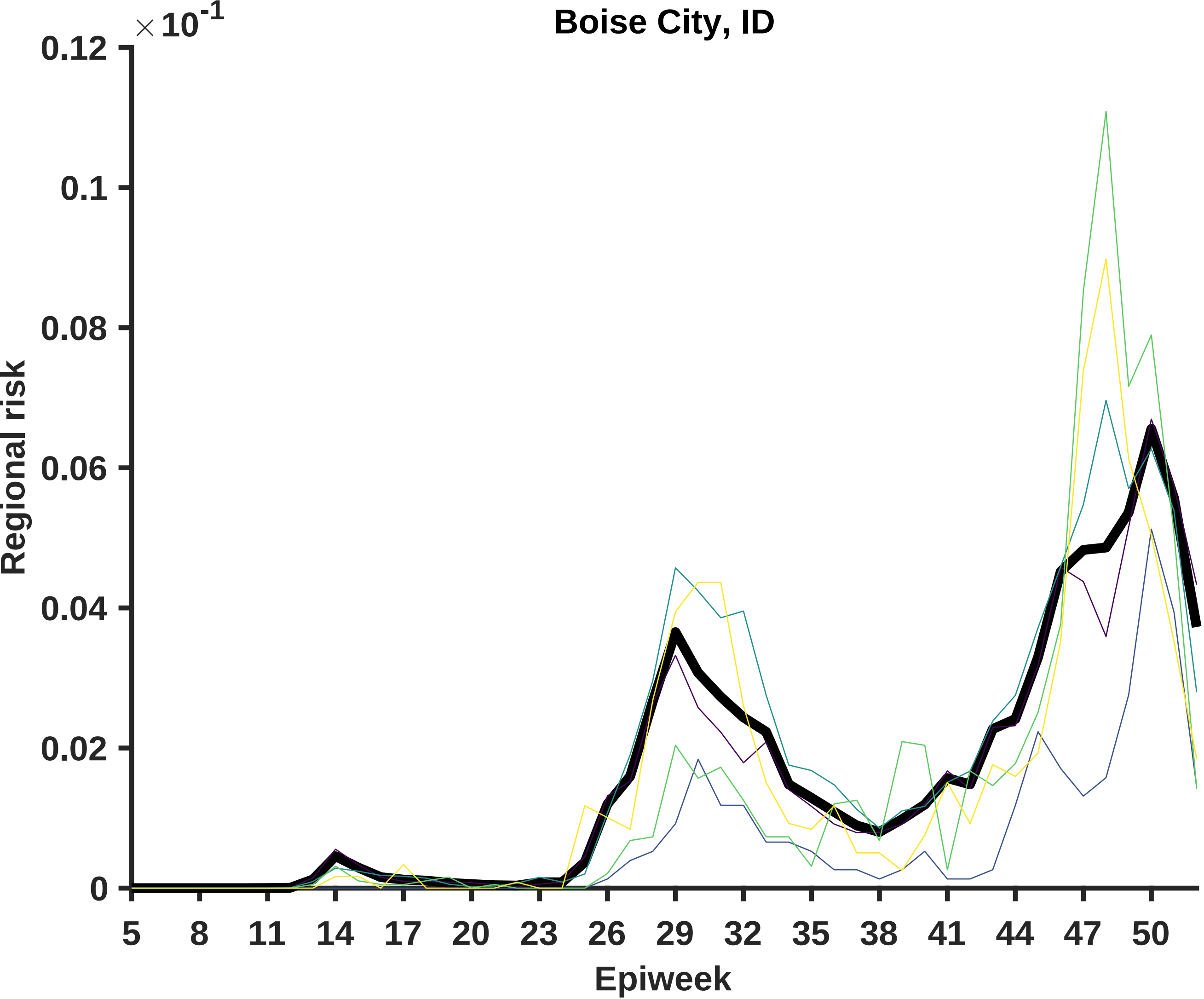

Boise City, ID
×10^-1
0.12
0.1
0.08
0.06
0.04
0.02
0
Regional risk
5
8
11
14
17
20
23
26
29
32
35
38
41
44
47
50
Epiweek

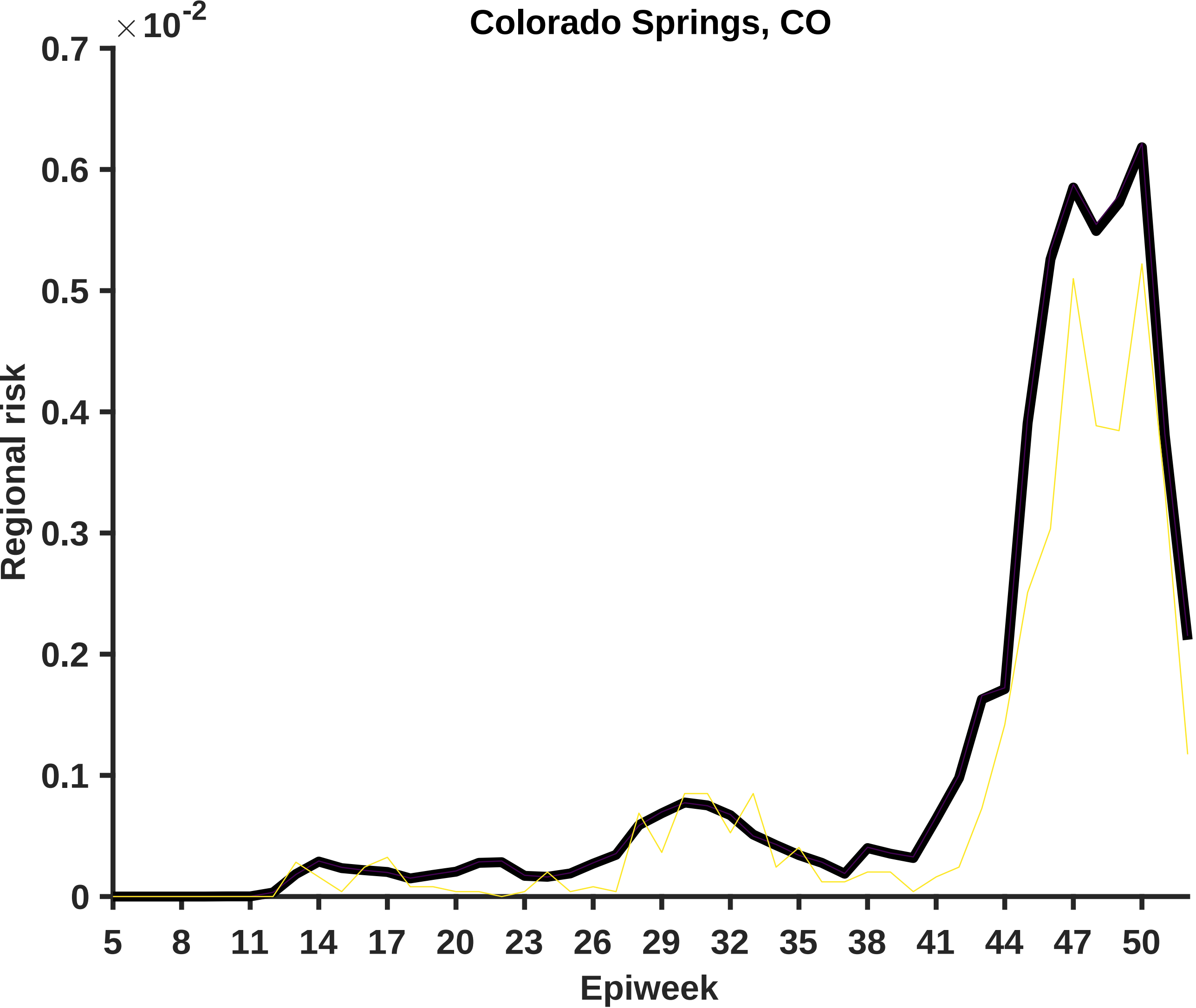

Colorado Springs, CO
×10^-2
0.7
0.6
0.5
0.4
0.3
0.2
0.1
0
Regional risk
5
8
11
14
17
20
23
26
29
32
35
38
41
44
47
50
Epiweek

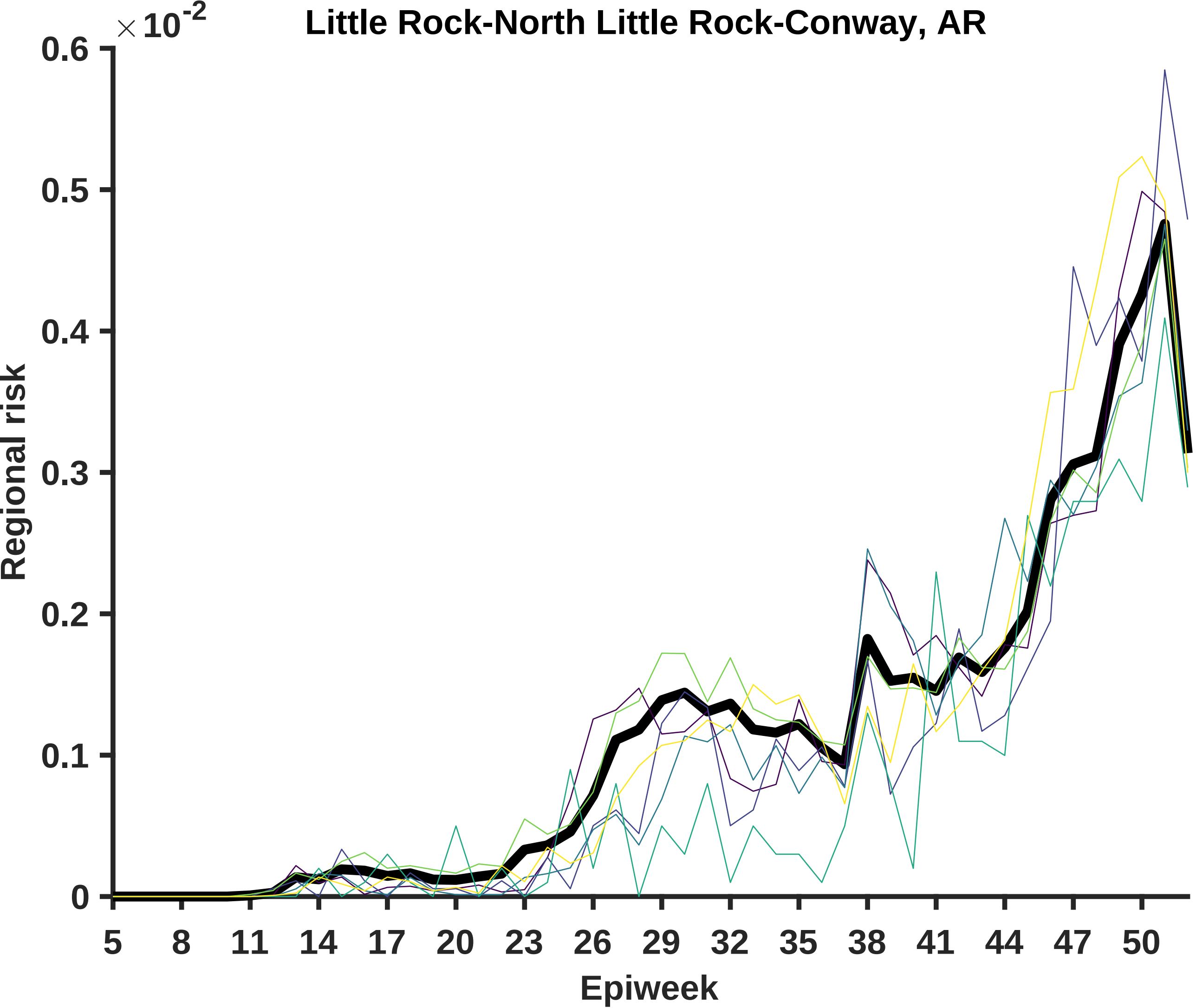

Little Rock-North Little Rock-Conway, AR
× 10^-2
0.6
0.5
0.4
0.3
0.2
0.1
0
Regional risk
5
8
11
14
17
20
23
26
29
32
35
38
41
44
47
50
Epiweek

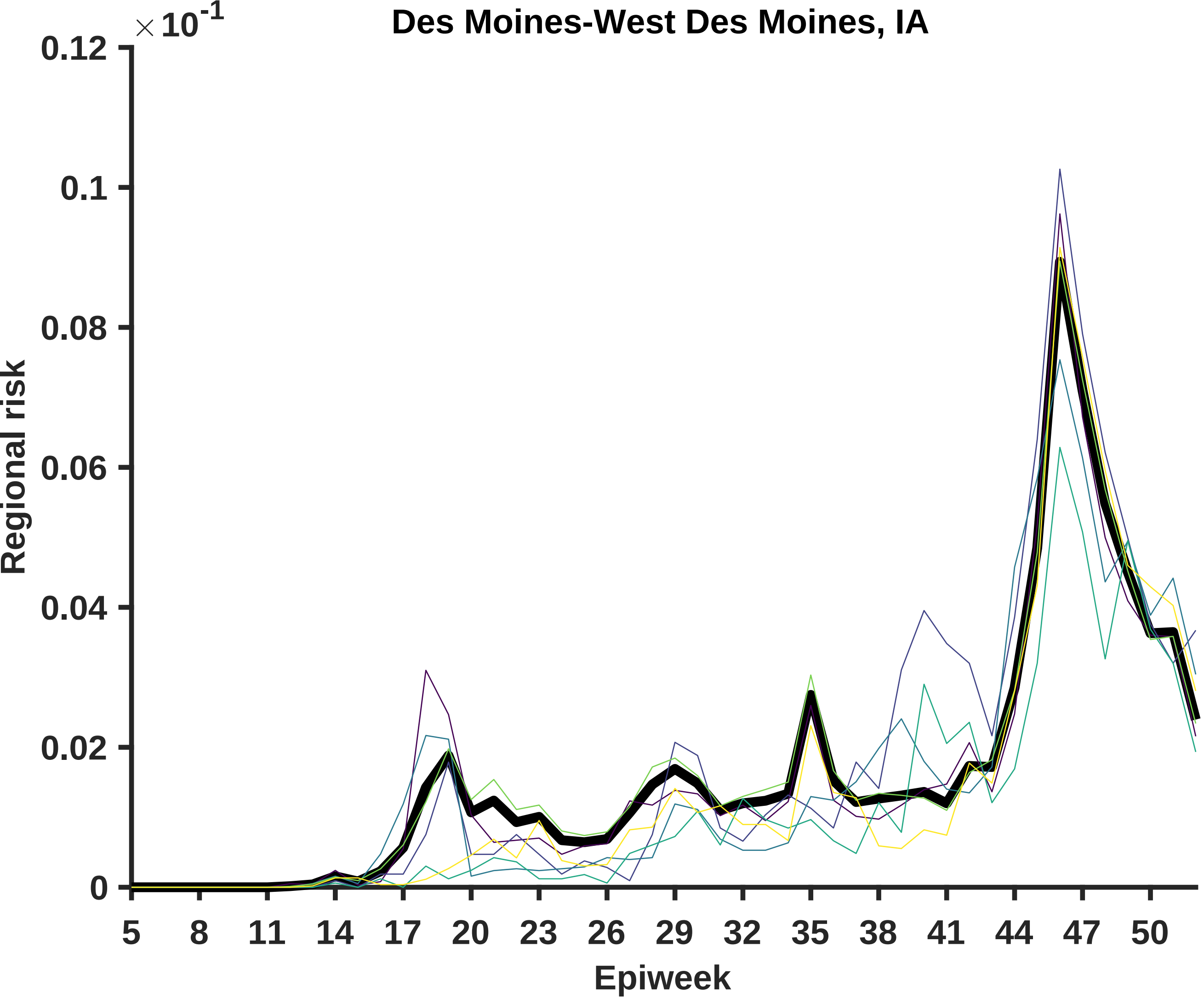

Des Moines-West Des Moines, IA
×10-1
0.12
0.1
0.08
0.06
0.04
0.02
0
Regional risk
5
8
11
14
17
20
23
26
29
32
35
38
41
44
47
50
Epiweek

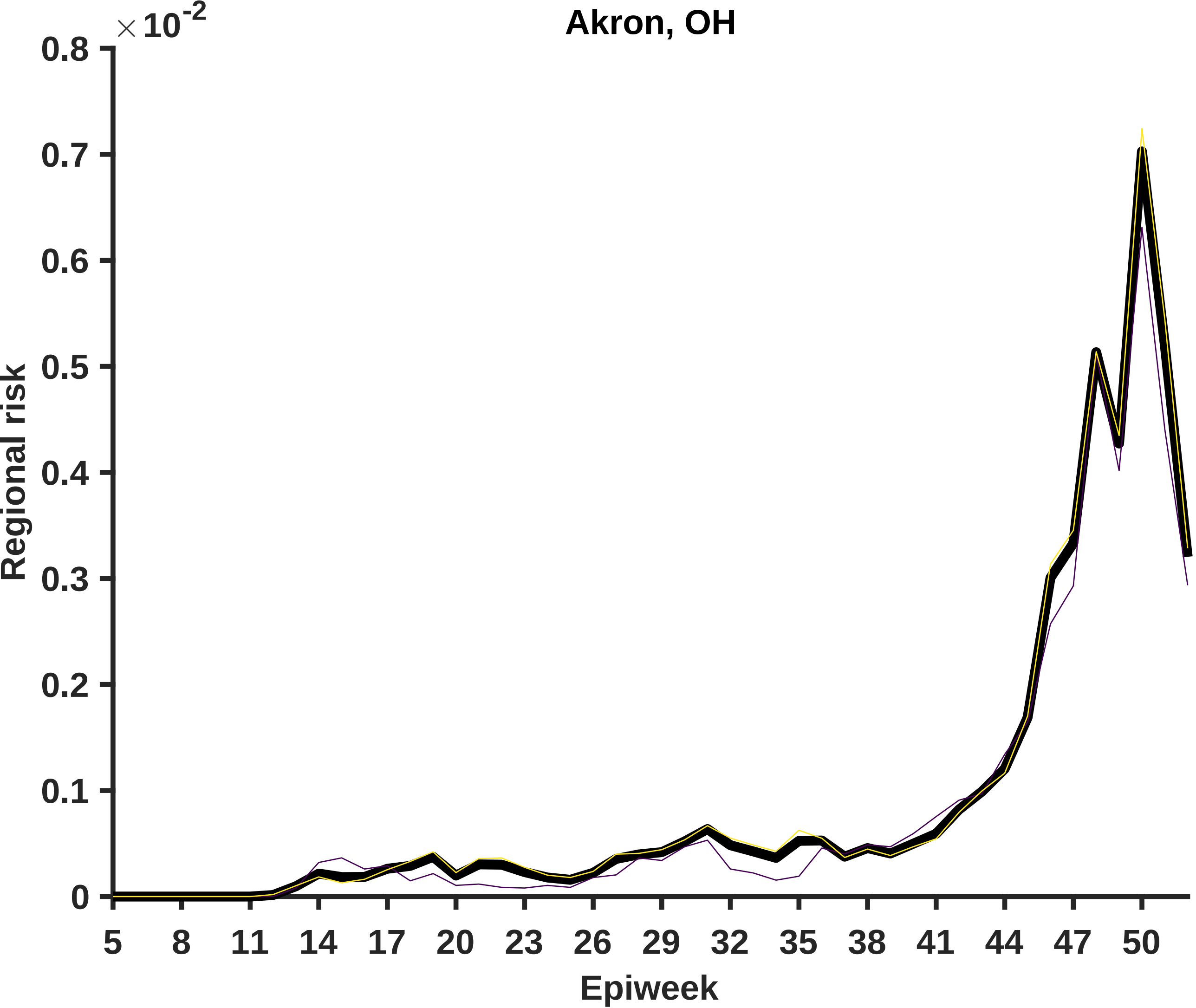

Akron, OH
×10^-2
0.8
0.7
0.6
0.5
0.4
0.3
0.2
0.1
0
Regional risk
5
8
11
14
17
20
23
26
29
32
35
38
41
44
47
50
Epiweek

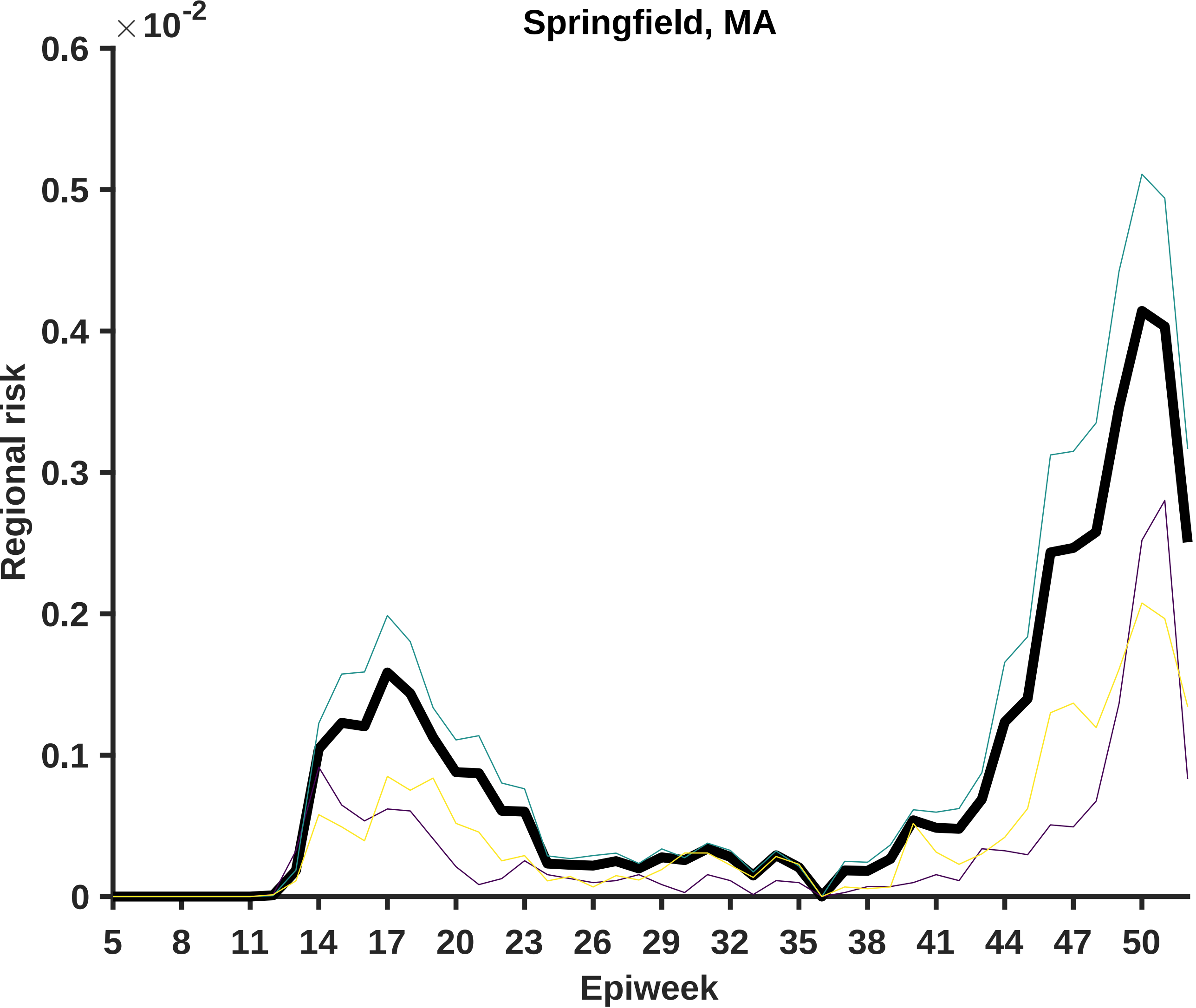

Springfield, MA
×10^-2
0.6
0.5
0.4
0.3
0.2
0.1
0
Regional risk
5
8
11
14
17
20
23
26
29
32
35
38
41
44
47
50
Epiweek

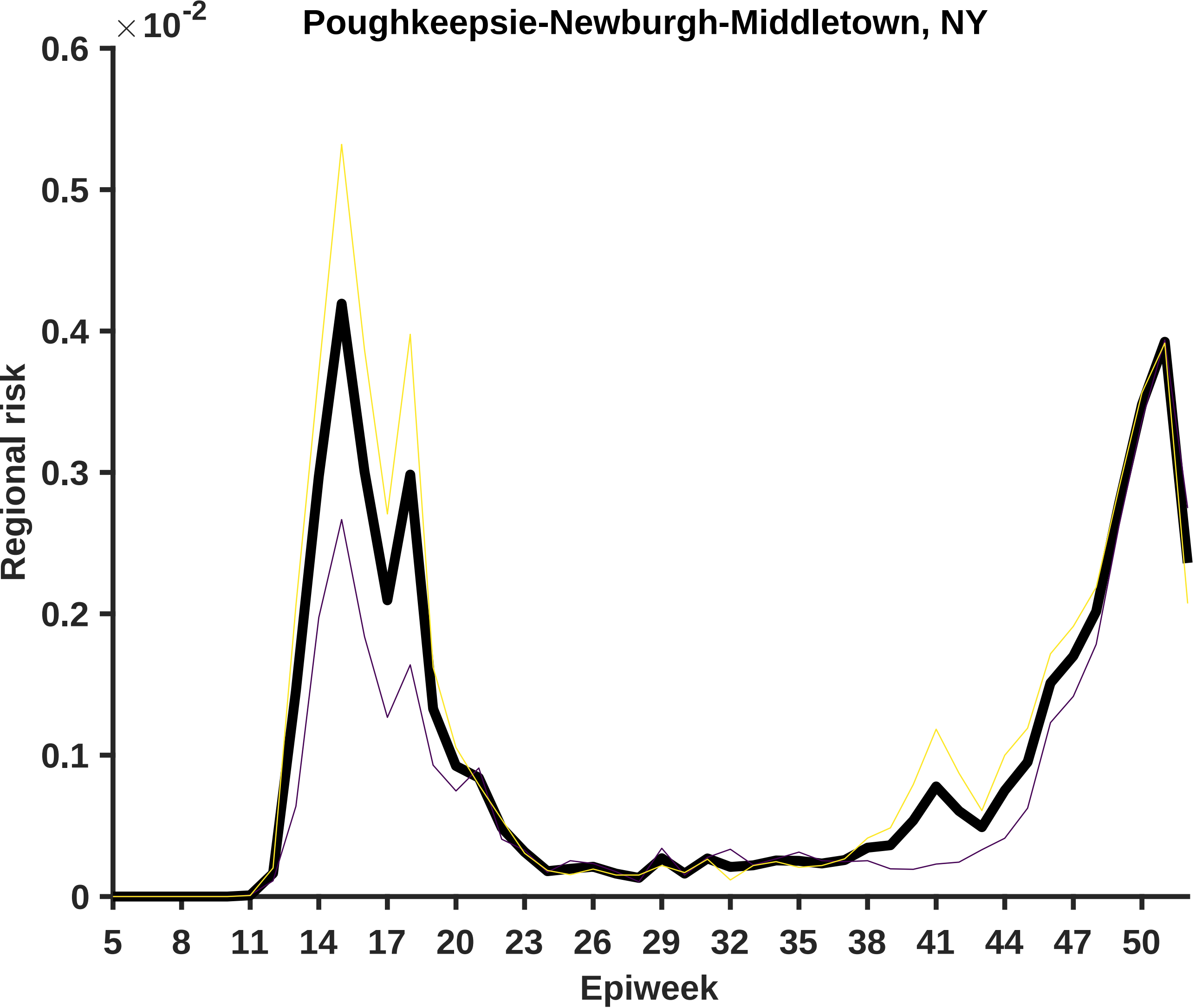

Poughkeepsie-Newburgh-Middletown, NY
×10-2
0.6
0.5
0.4
0.3
0.2
0.1
0
Regional risk
5
8
11
14
17
20
23
26
29
32
35
38
41
44
47
50
Epiweek

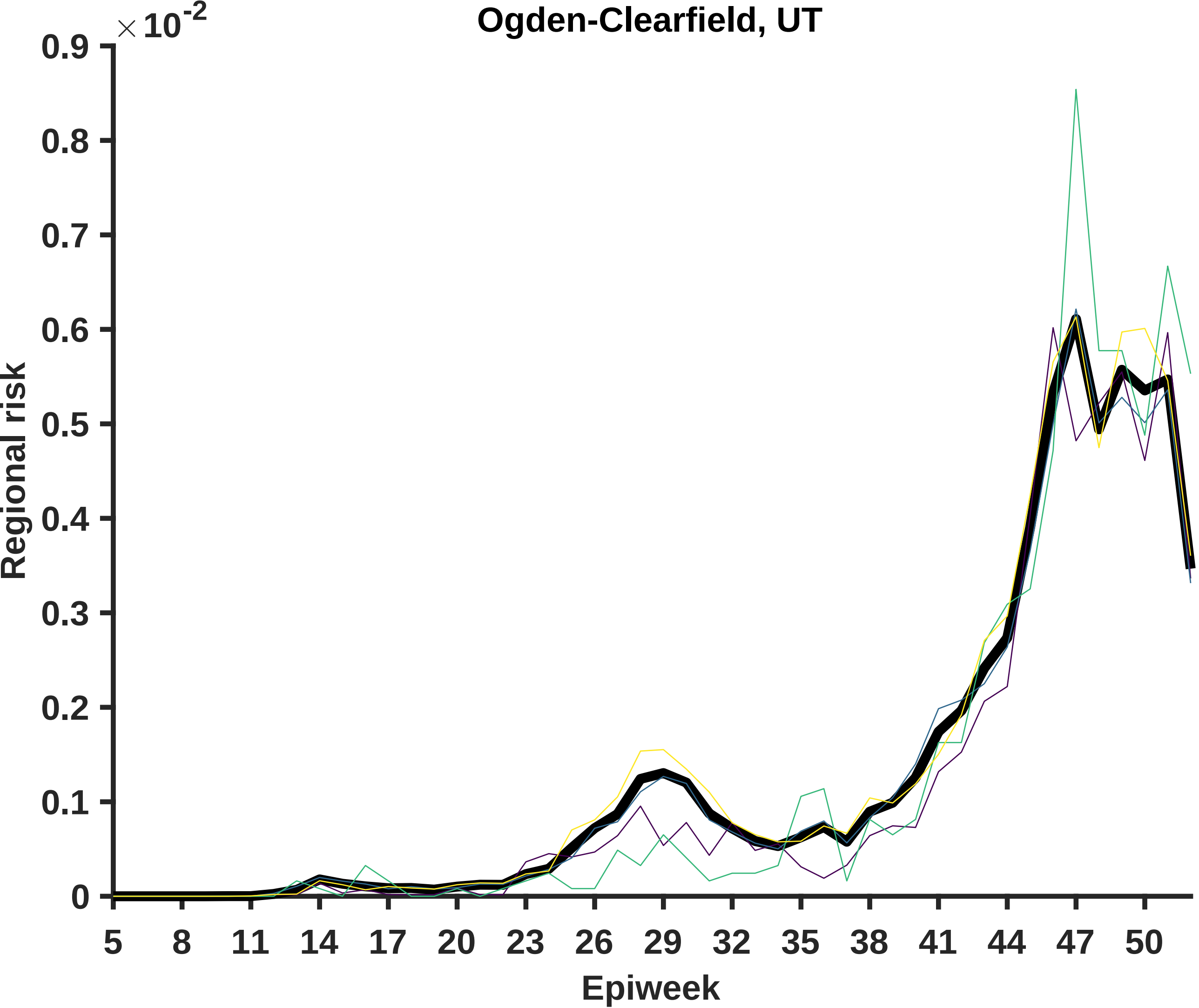

Ogden-Clearfield, UT
×10^-2
0.9
0.8
0.7
0.6
0.5
0.4
0.3
0.2
0.1
0
Regional risk
5
8
11
14
17
20
23
26
29
32
35
38
41
44
47
50
Epiweek

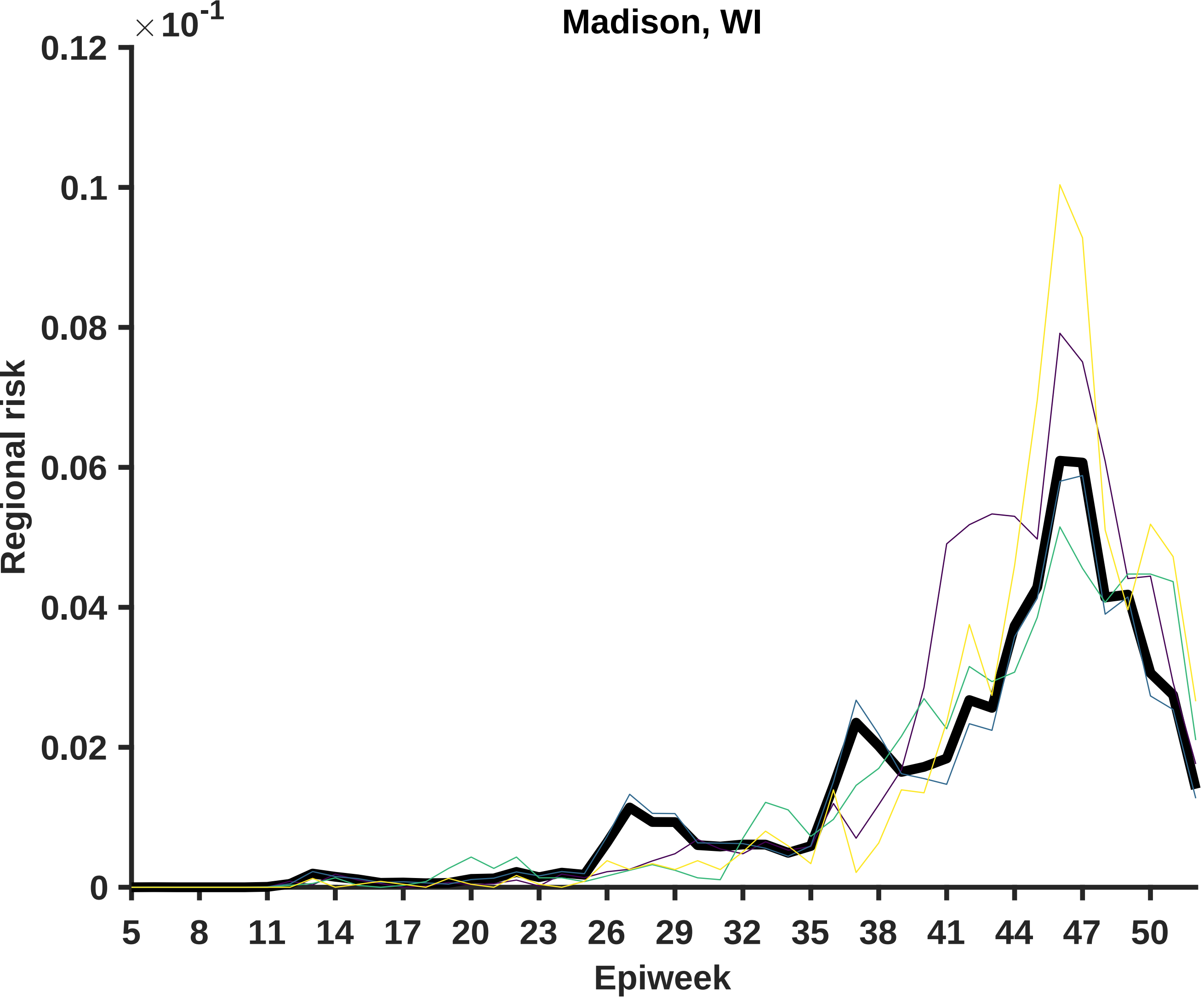

Madison, WI
× 10^-1
0.12
0.1
0.08
0.06
0.04
0.02
0
Regional risk
5
8
11
14
17
20
23
26
29
32
35
38
41
44
47
50
Epiweek

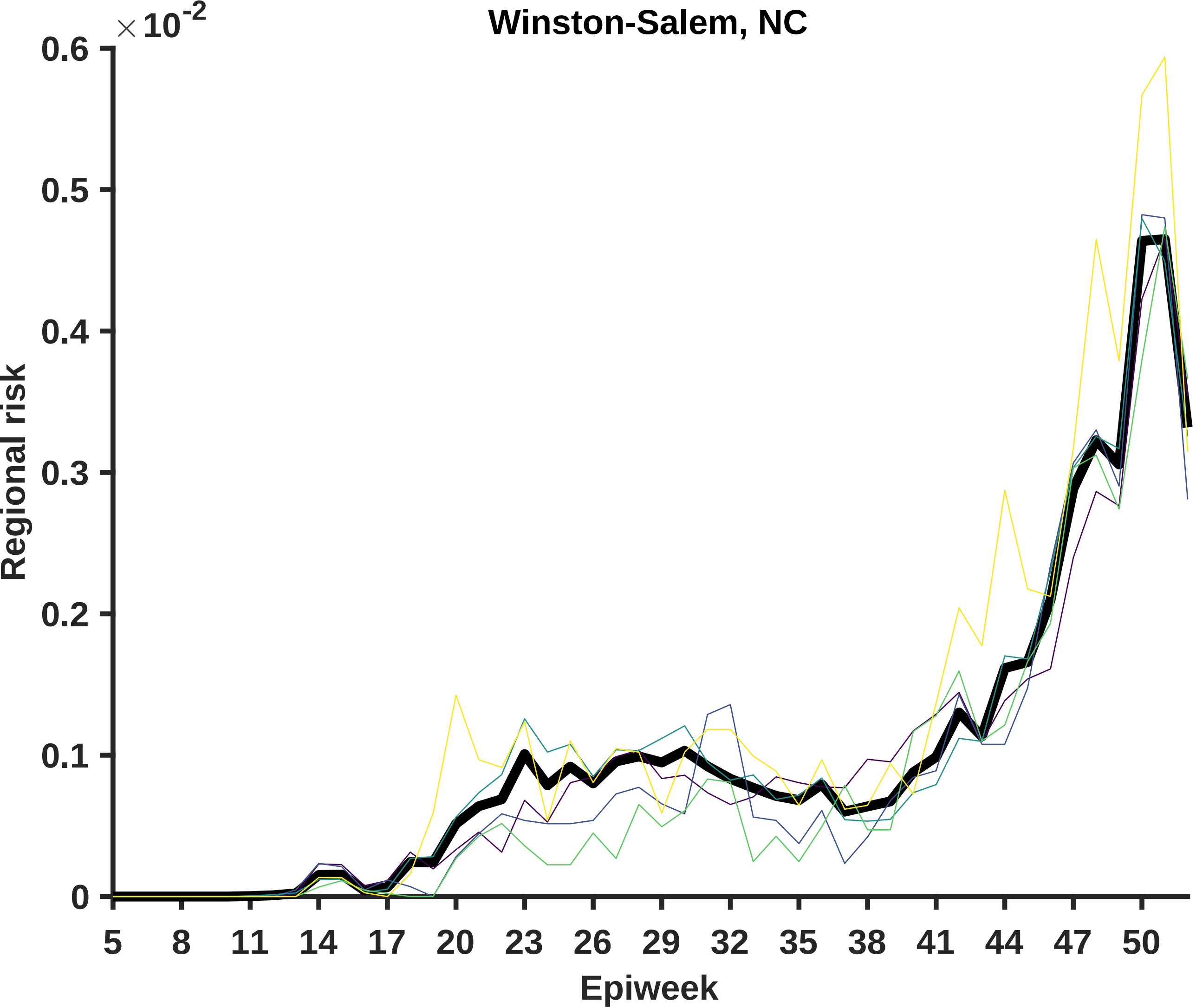
Winston-Salem, NC
$\times 10^{-2}$
0.6
0.5
0.4
0.3
0.2
0.1
0
Regional risk
5
8
11
14
17
20
23
26
29
32
35
38
41
44
47
50
Epiweek

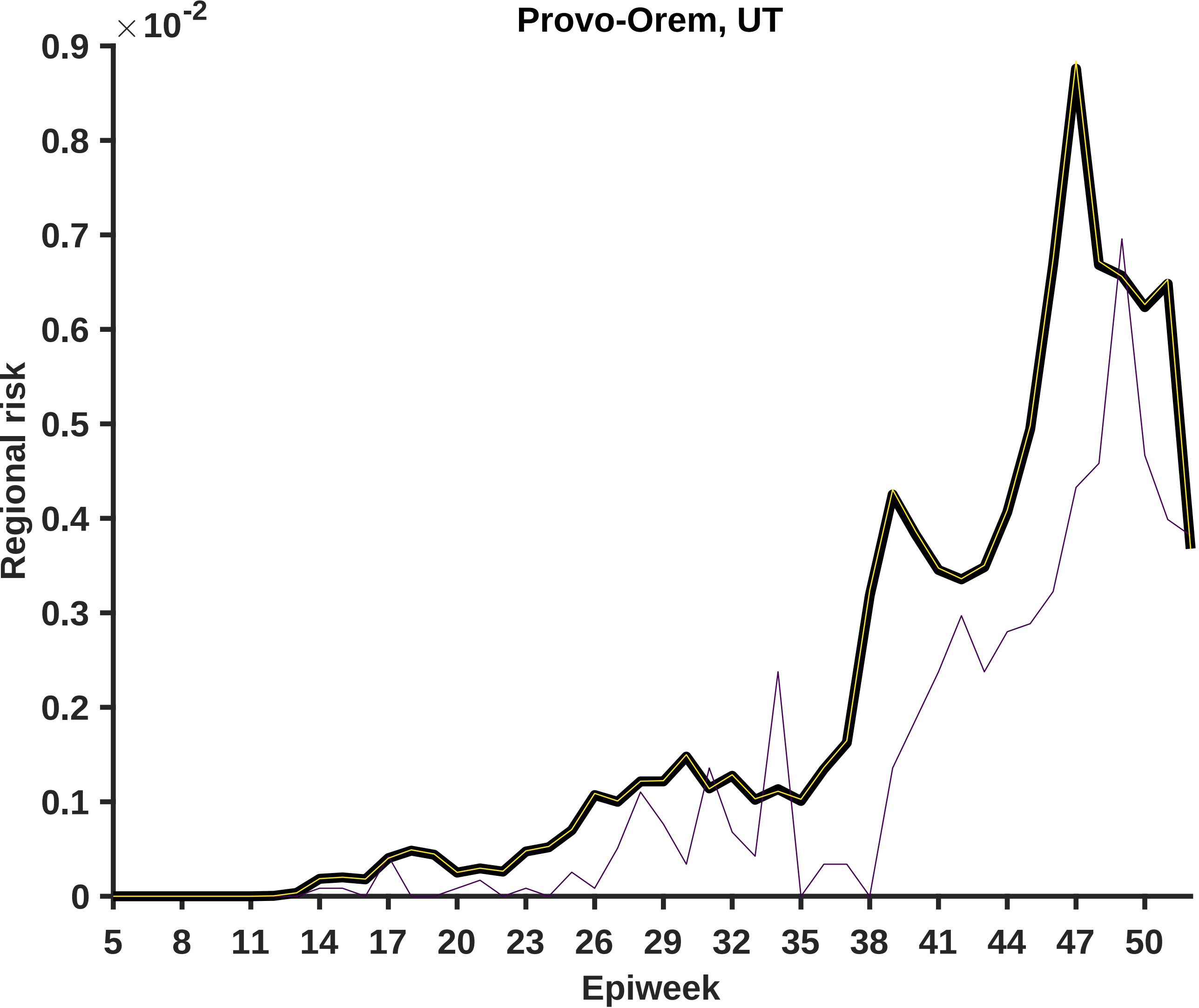

Provo-Orem, UT
×10^-2
0.9
0.8
0.7
0.6
0.5
0.4
0.3
0.2
0.1
0
Regional risk
5
8
11
14
17
20
23
26
29
32
35
38
41
44
47
50
Epiweek

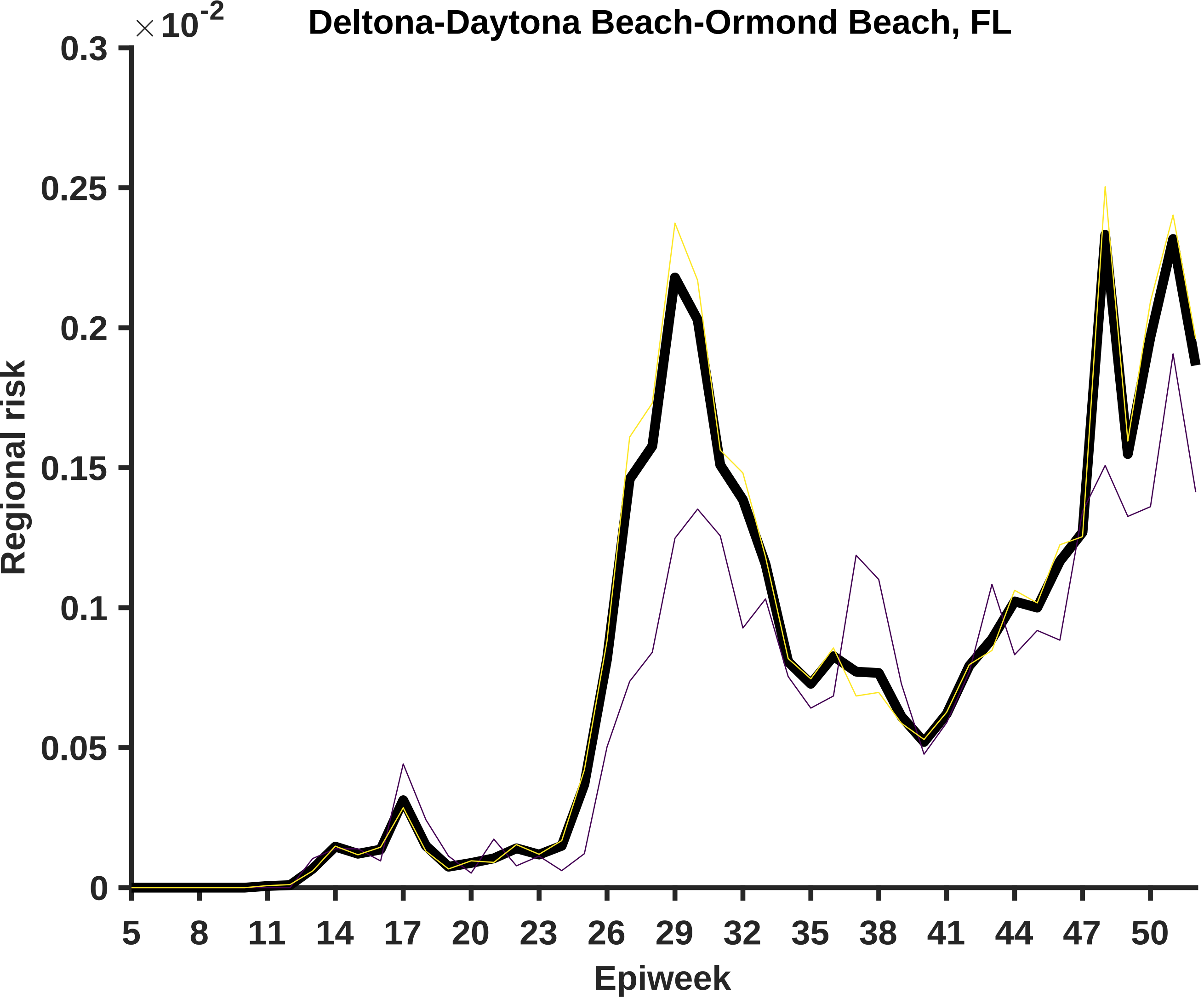

Deltona-Daytona Beach-Ormond Beach, FL
× 10^-2
0.3
0.25
0.2
0.15
0.1
0.05
0
Regional risk
5
8
11
14
17
20
23
26
29
32
35
38
41
44
47
50
Epiweek

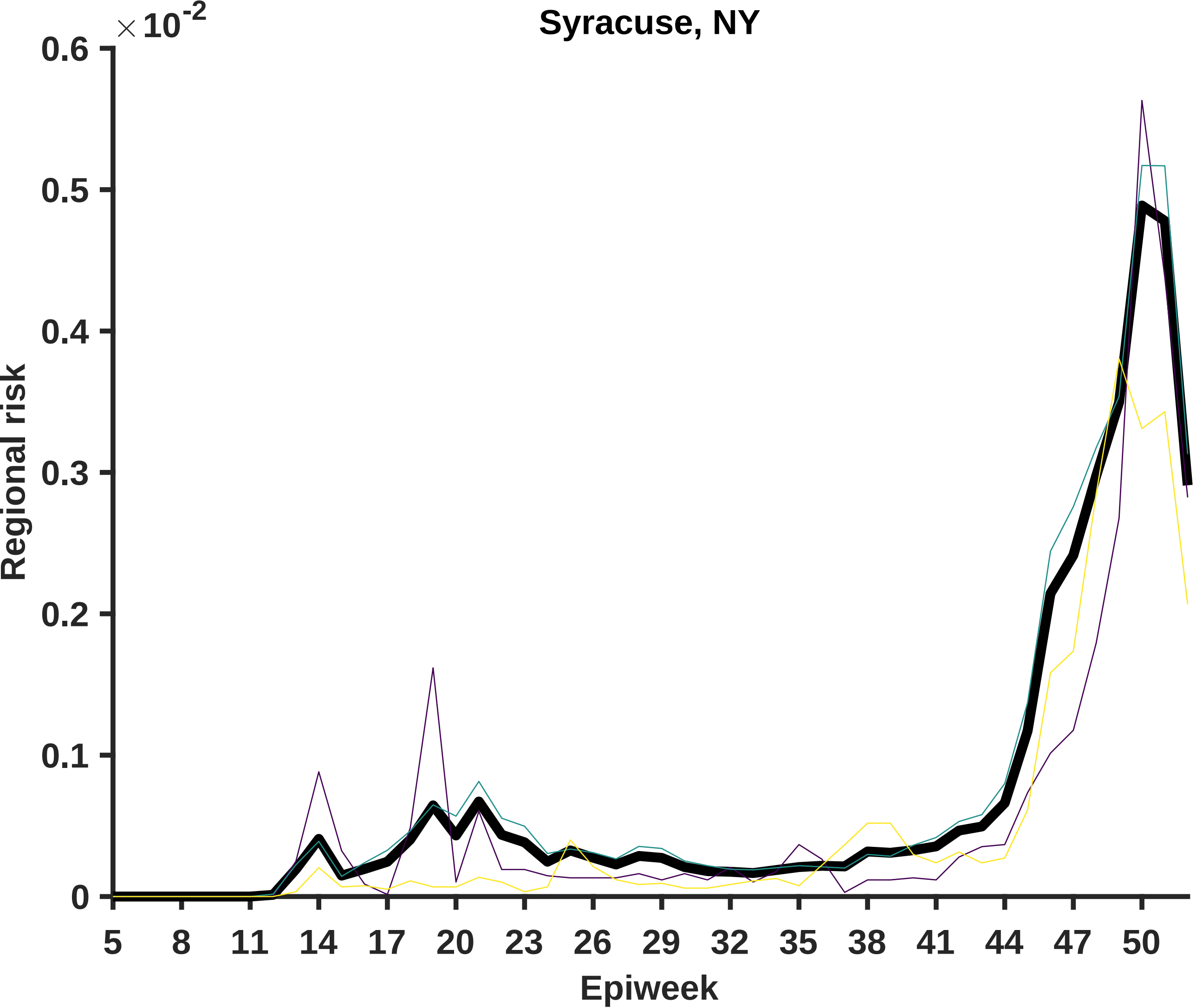

Syracuse, NY
×10^-2
0.6
0.5
0.4
0.3
0.2
0.1
0
Regional risk
5
8
11
14
17
20
23
26
29
32
35
38
41
44
47
50
Epiweek

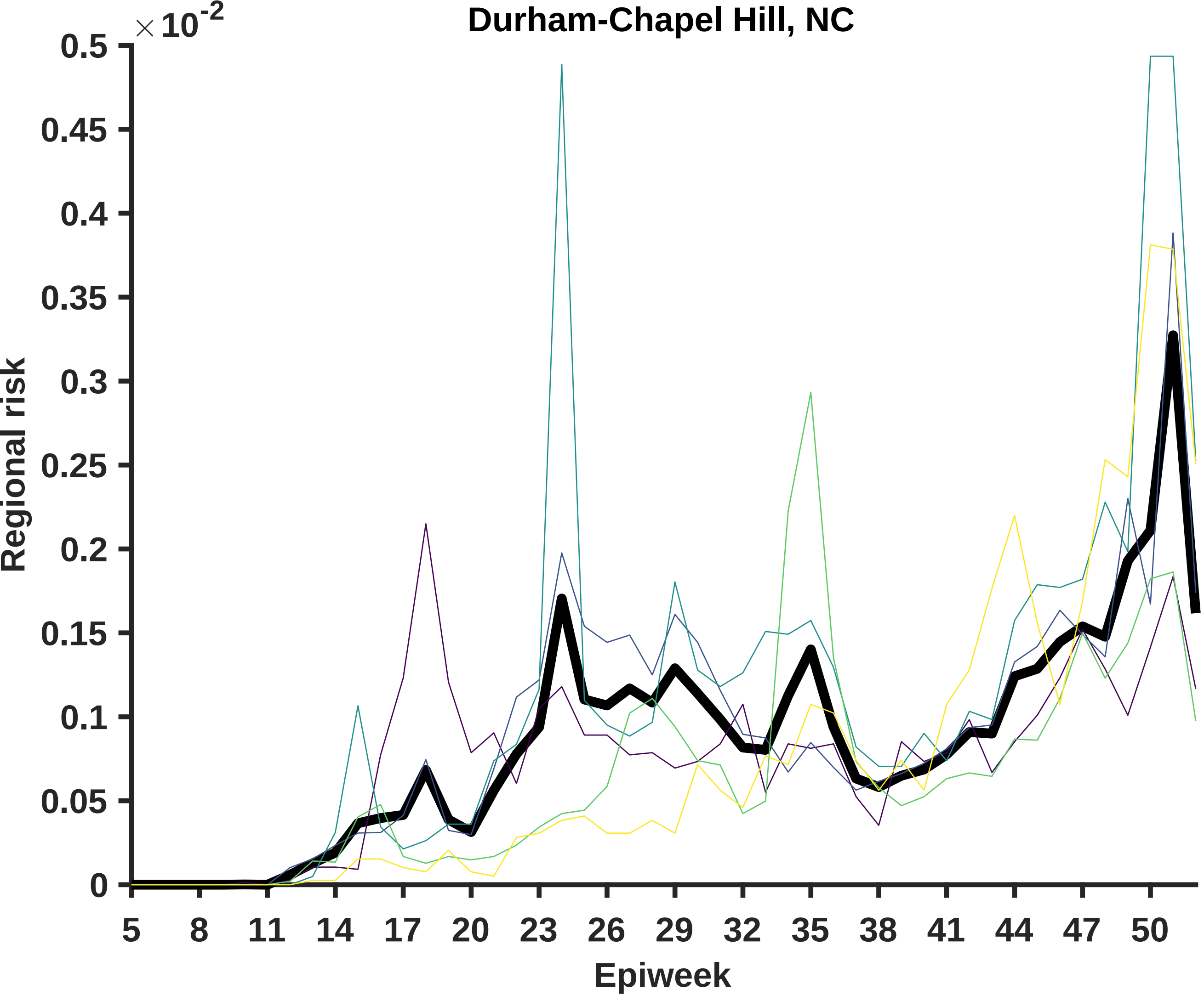
Durham-Chapel Hill, NC
× 10^-2
0.5
0.45
0.4
0.35
0.3
0.25
0.2
0.15
0.1
0.05
0
Regional risk
5
8
11
14
17
20
23
26
29
32
35
38
41
44
47
50
Epiweek

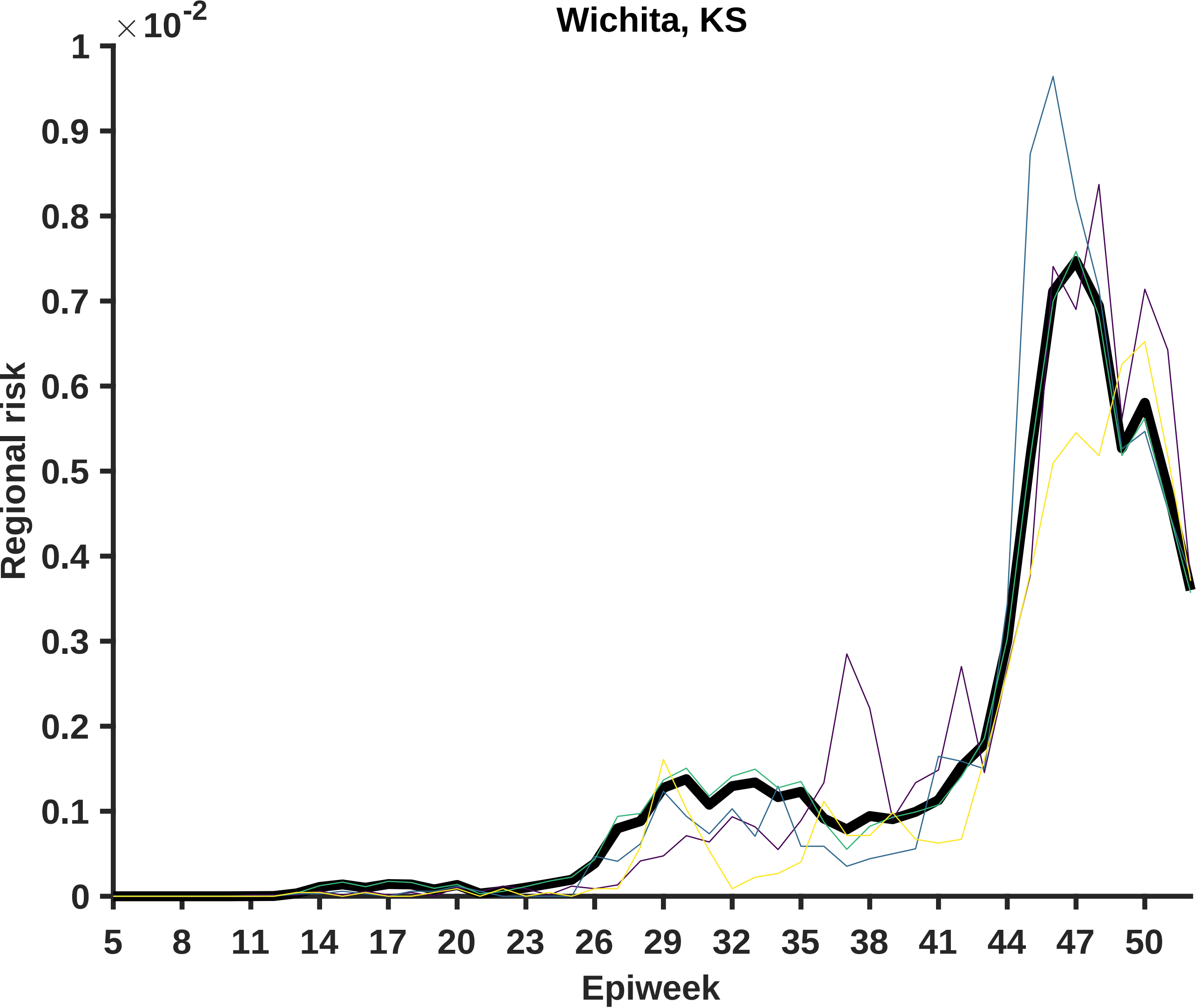
Wichita, KS
×10^-2
1
0.9
0.8
0.7
0.6
0.5
0.4
0.3
0.2
0.1
0
Regional risk
5
8
11
14
17
20
23
26
29
32
35
38
41
44
47
50
Epiweek

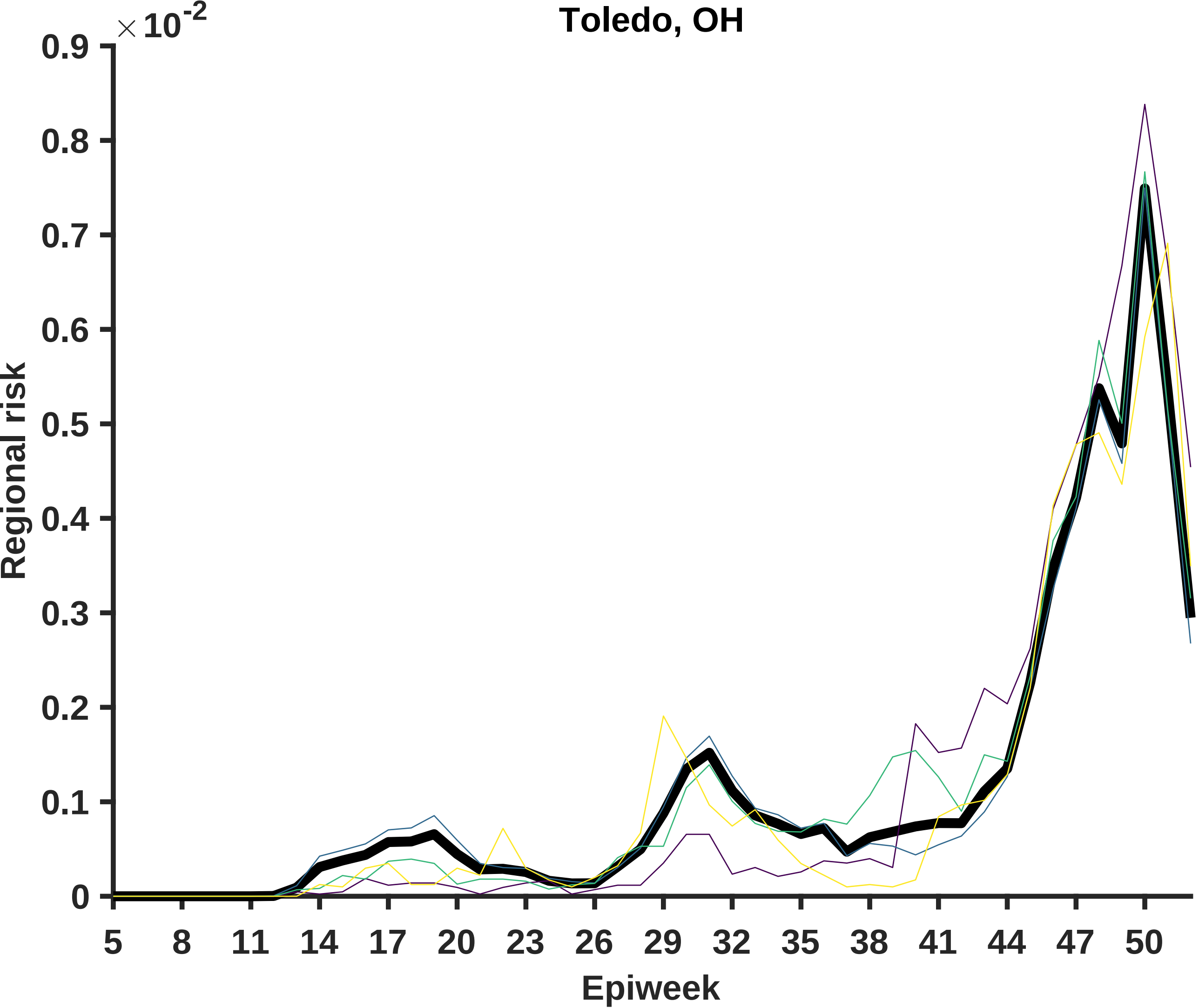
Toledo, OH
×10^-2
Regional risk
0.9
0.8
0.7
0.6
0.5
0.4
0.3
0.2
0.1
0
Epiweek
5
8
11
14
17
20
23
26
29
32
35
38
41
44
47
50

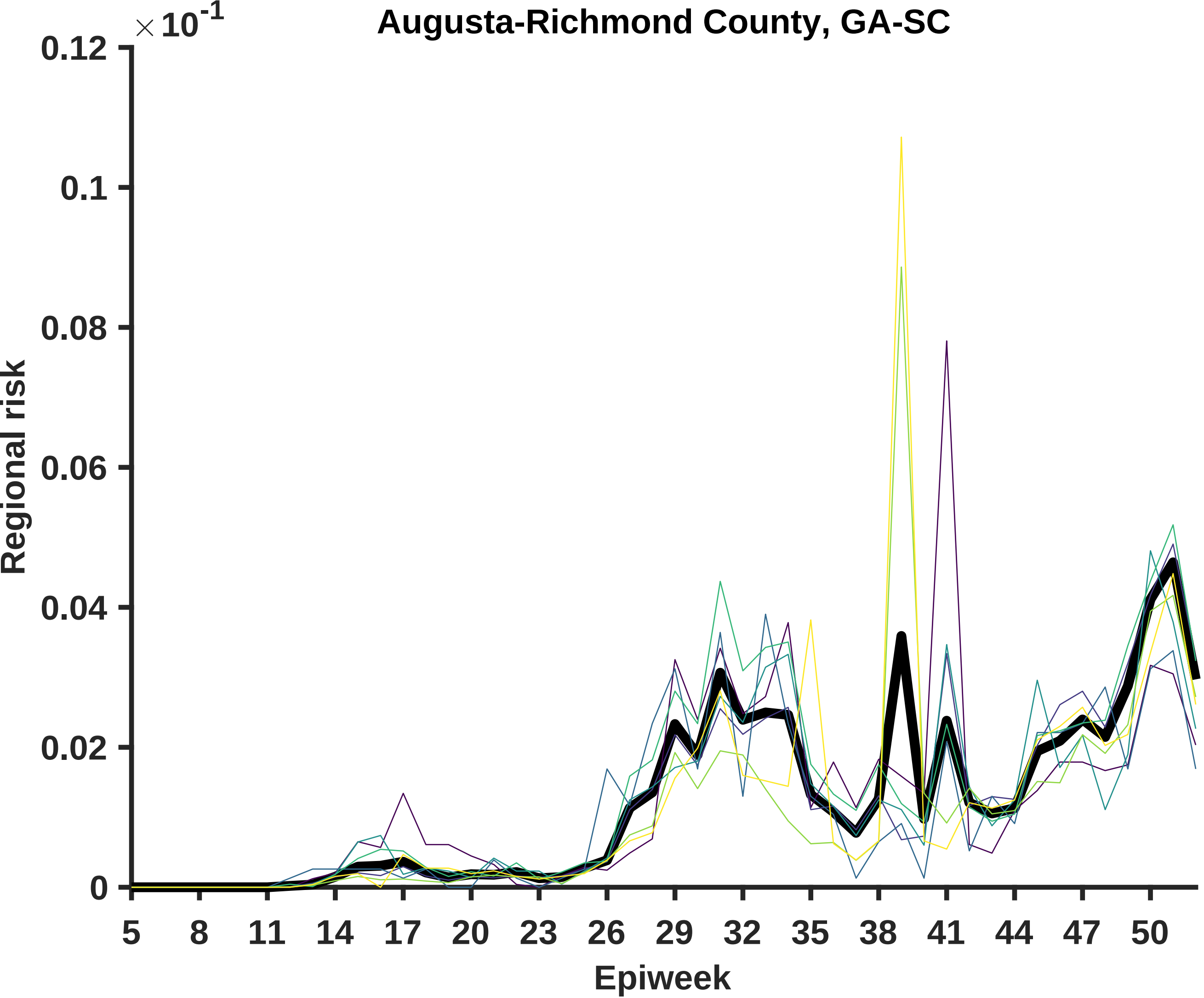

Augusta-Richmond County, GA-SC
×10^-1
Regional risk
0.12
0.1
0.08
0.06
0.04
0.02
0
5
8
11
14
17
20
23
26
29
32
35
38
41
44
47
50
Epiweek

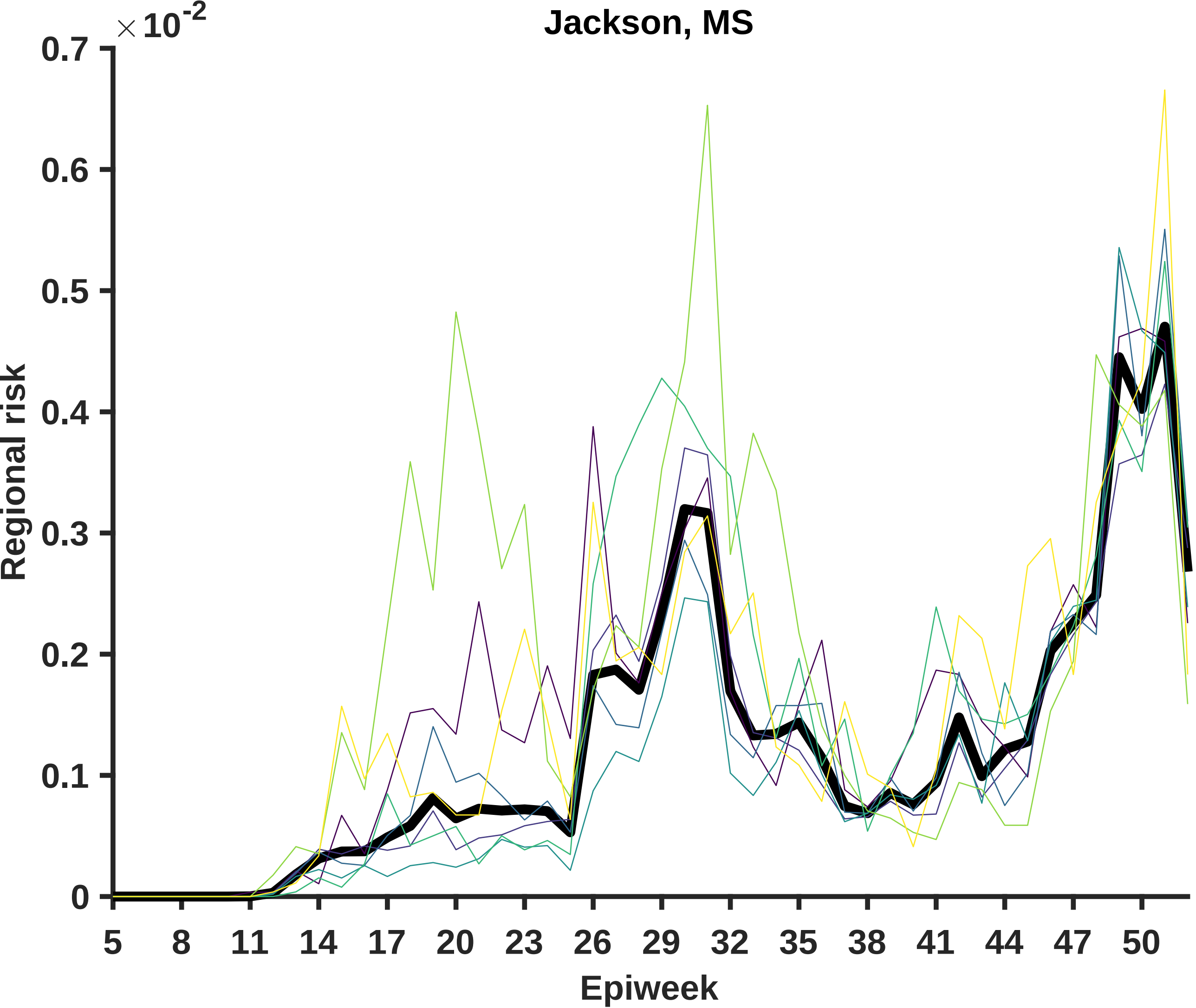

Jackson, MS
×10^-2
0.7
0.6
0.5
0.4
0.3
0.2
0.1
0
Regional risk
5
8
11
14
17
20
23
26
29
32
35
38
41
44
47
50
Epiweek

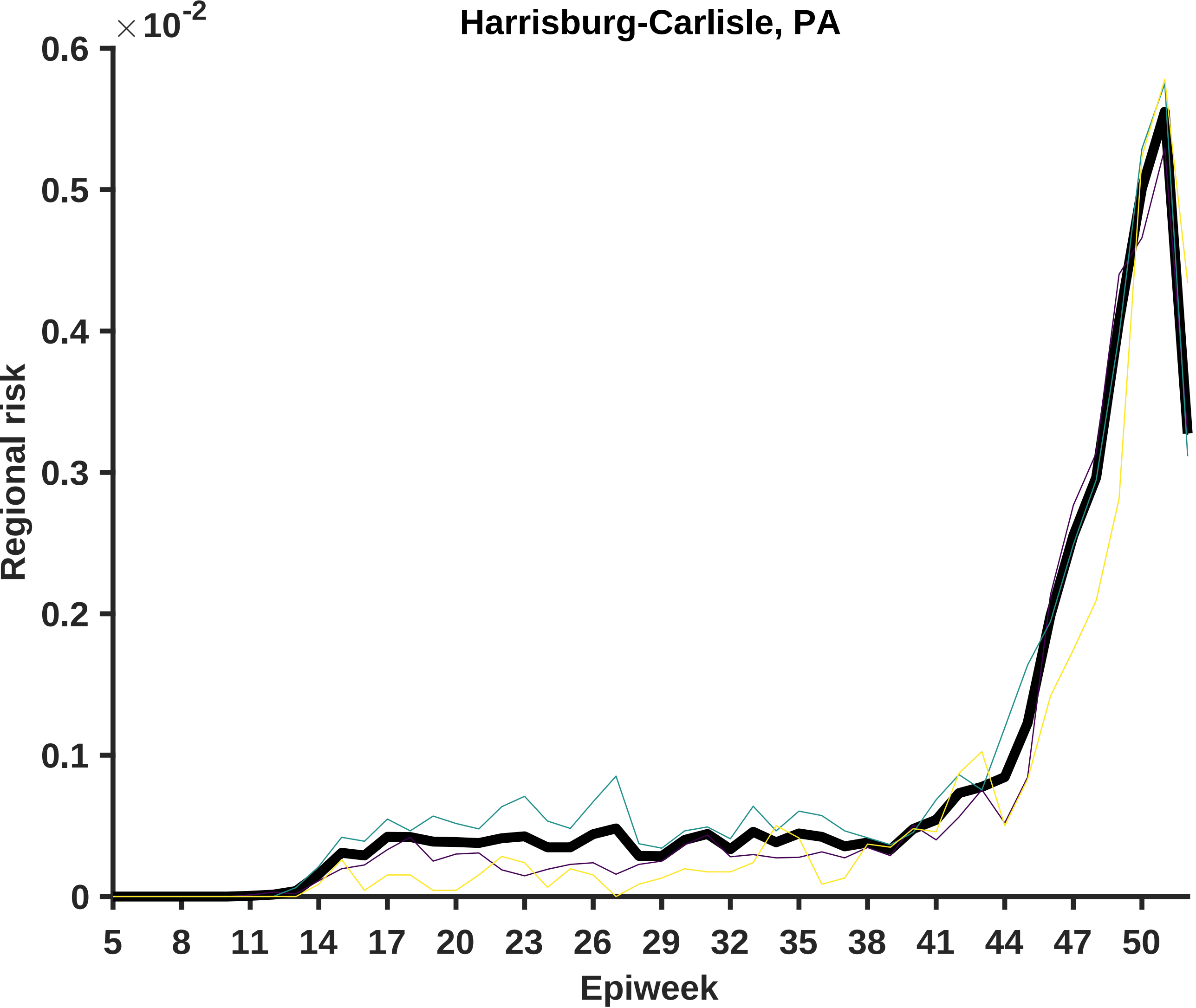
Harrisburg-Carlisle, PA
×10^-2
0.6
0.5
0.4
0.3
0.2
0.1
0
Regional risk
5
8
11
14
17
20
23
26
29
32
35
38
41
44
47
50
Epiweek

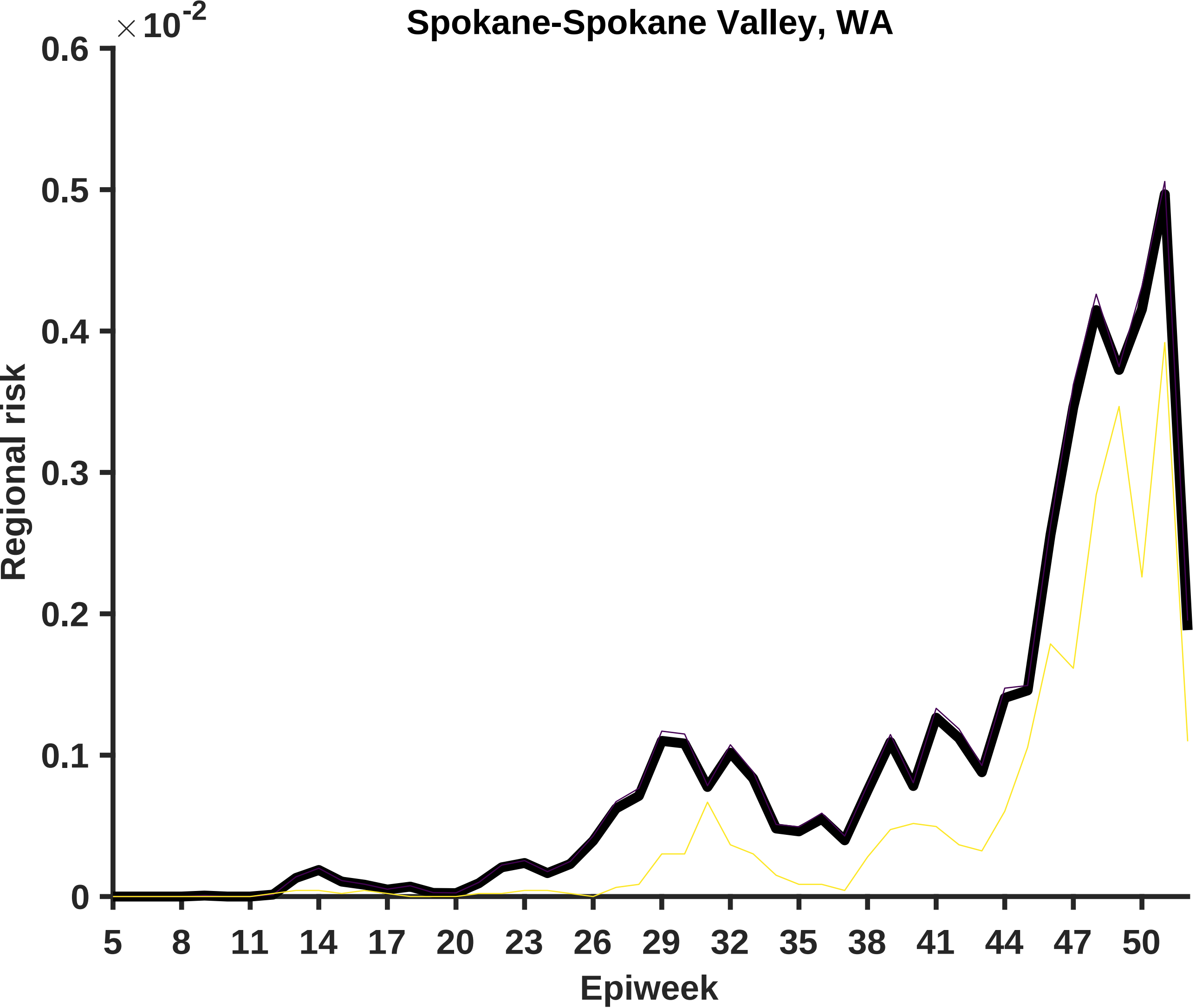
Spokane-Spokane Valley, WA
× 10^-2
0.6
0.5
0.4
0.3
0.2
0.1
0
Regional risk
5
8
11
14
17
20
23
26
29
32
35
38
41
44
47
50
Epiweek

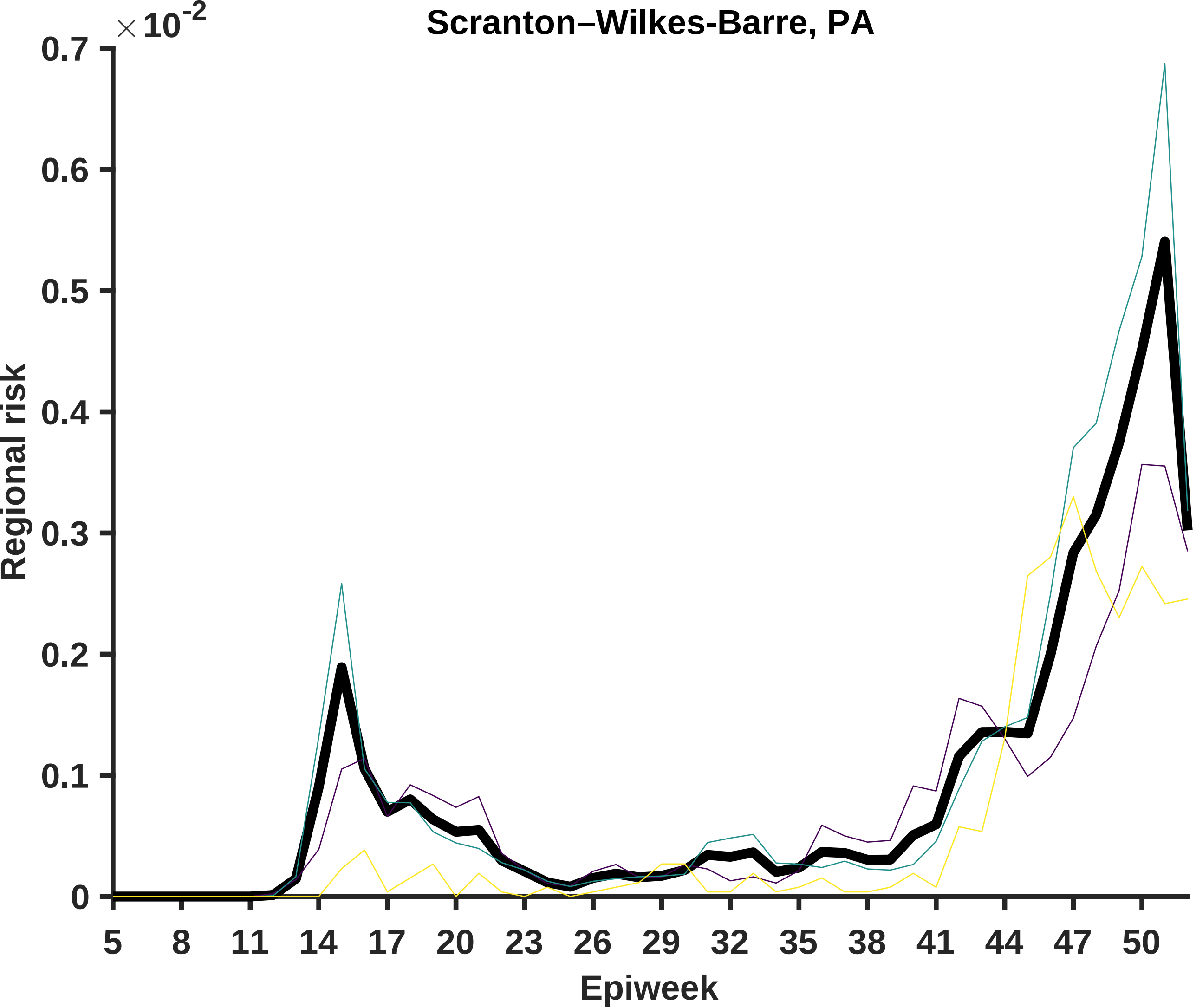

Scranton–Wilkes-Barre, PA
×10^-2
0.7
0.6
0.5
0.4
0.3
0.2
0.1
0
Regional risk
5
8
11
14
17
20
23
26
29
32
35
38
41
44
47
50
Epiweek

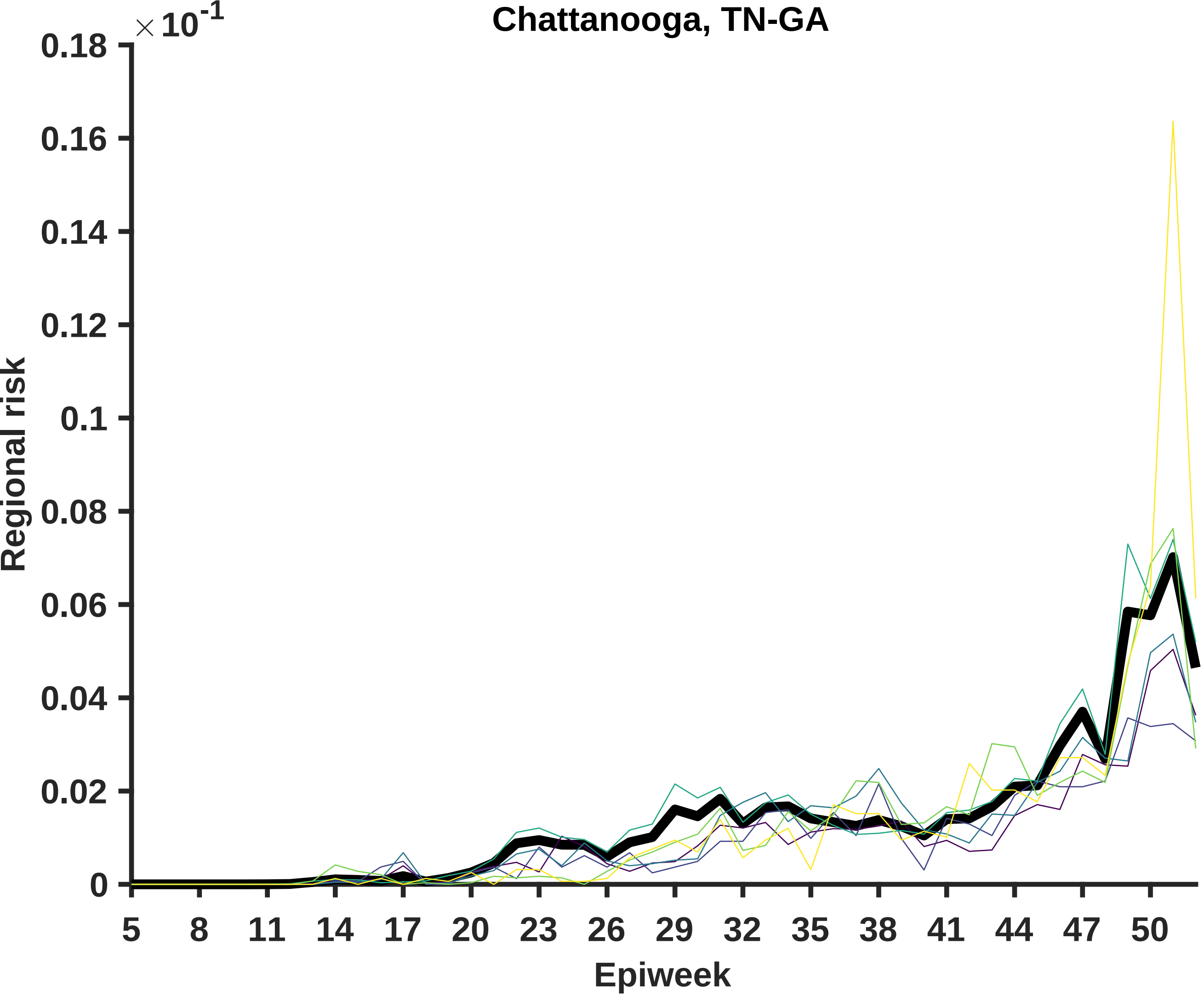

Chattanooga, TN-GA
× 10^-1
0.18
0.16
0.14
0.12
0.1
0.08
0.06
0.04
0.02
0
Regional risk
5
8
11
14
17
20
23
26
29
32
35
38
41
44
47
50
Epiweek

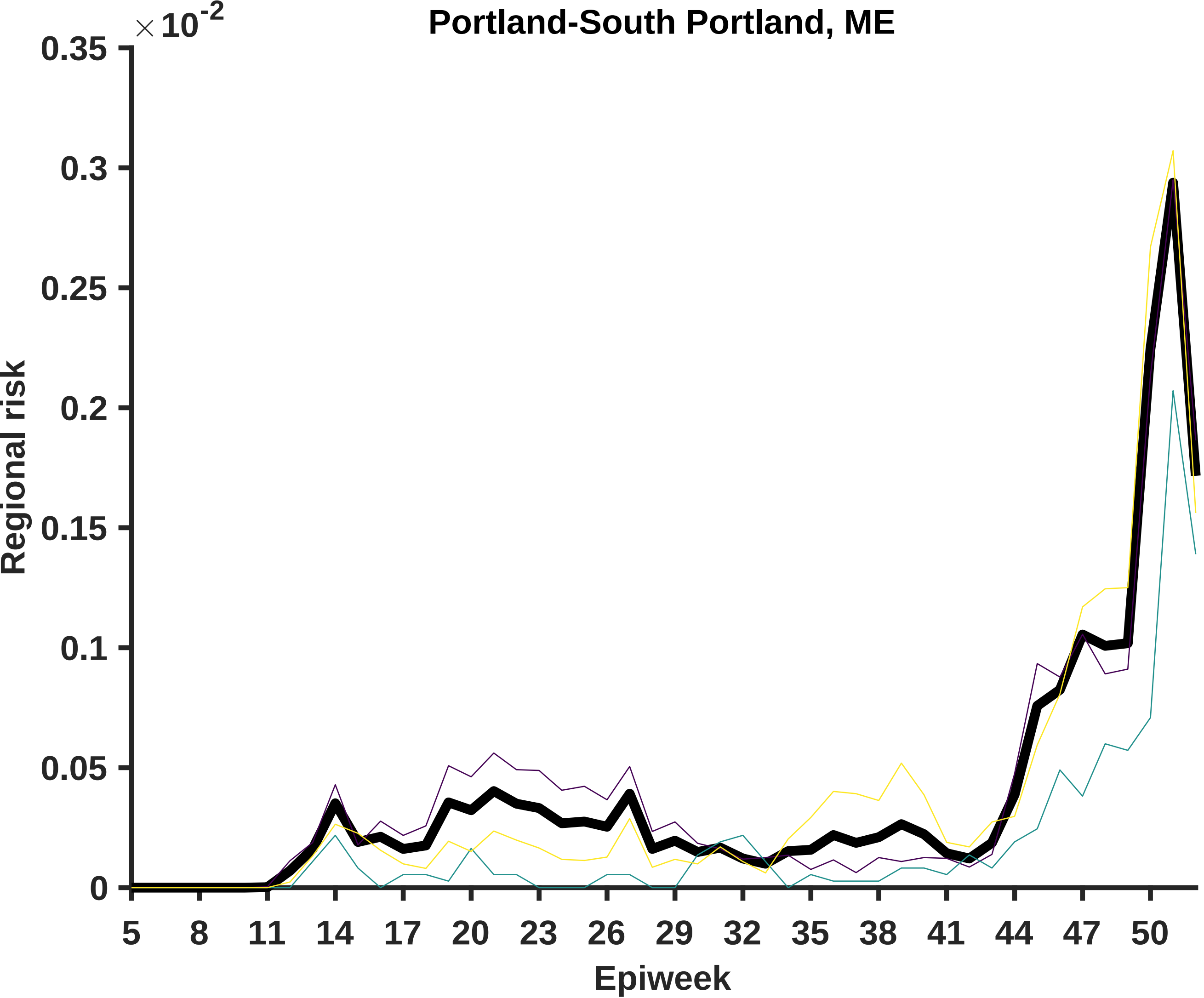
Portland-South Portland, ME
×10^-2
0.35
0.3
0.25
0.2
0.15
0.1
0.05
0
Regional risk
5
8
11
14
17
20
23
26
29
32
35
38
41
44
47
50
Epiweek

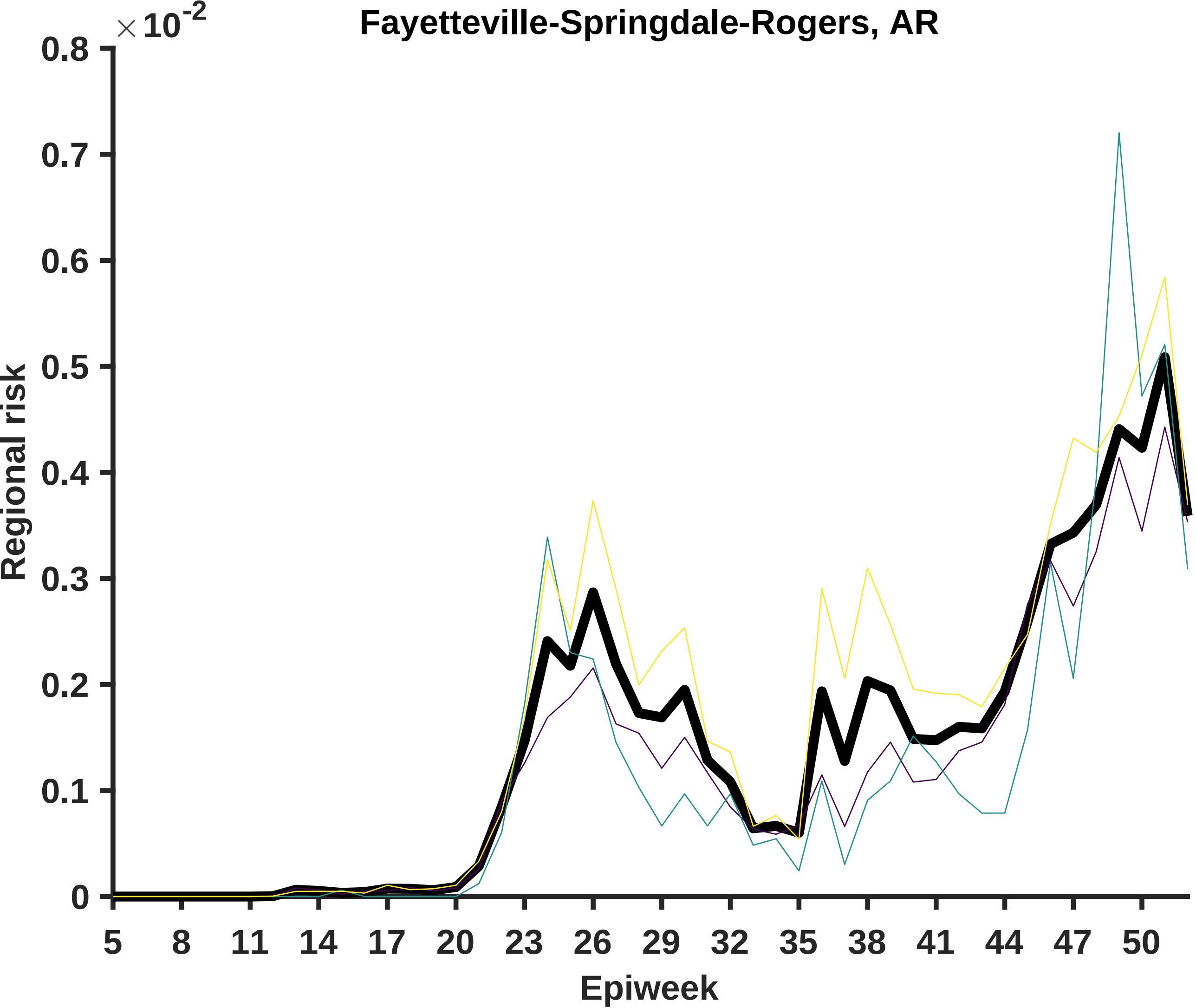

Fayetteville-Springdale-Rogers, AR
× 10^-2
0.8
0.7
0.6
0.5
0.4
0.3
0.2
0.1
0
Regional risk
5
8
11
14
17
20
23
26
29
32
35
38
41
44
47
50
Epiweek

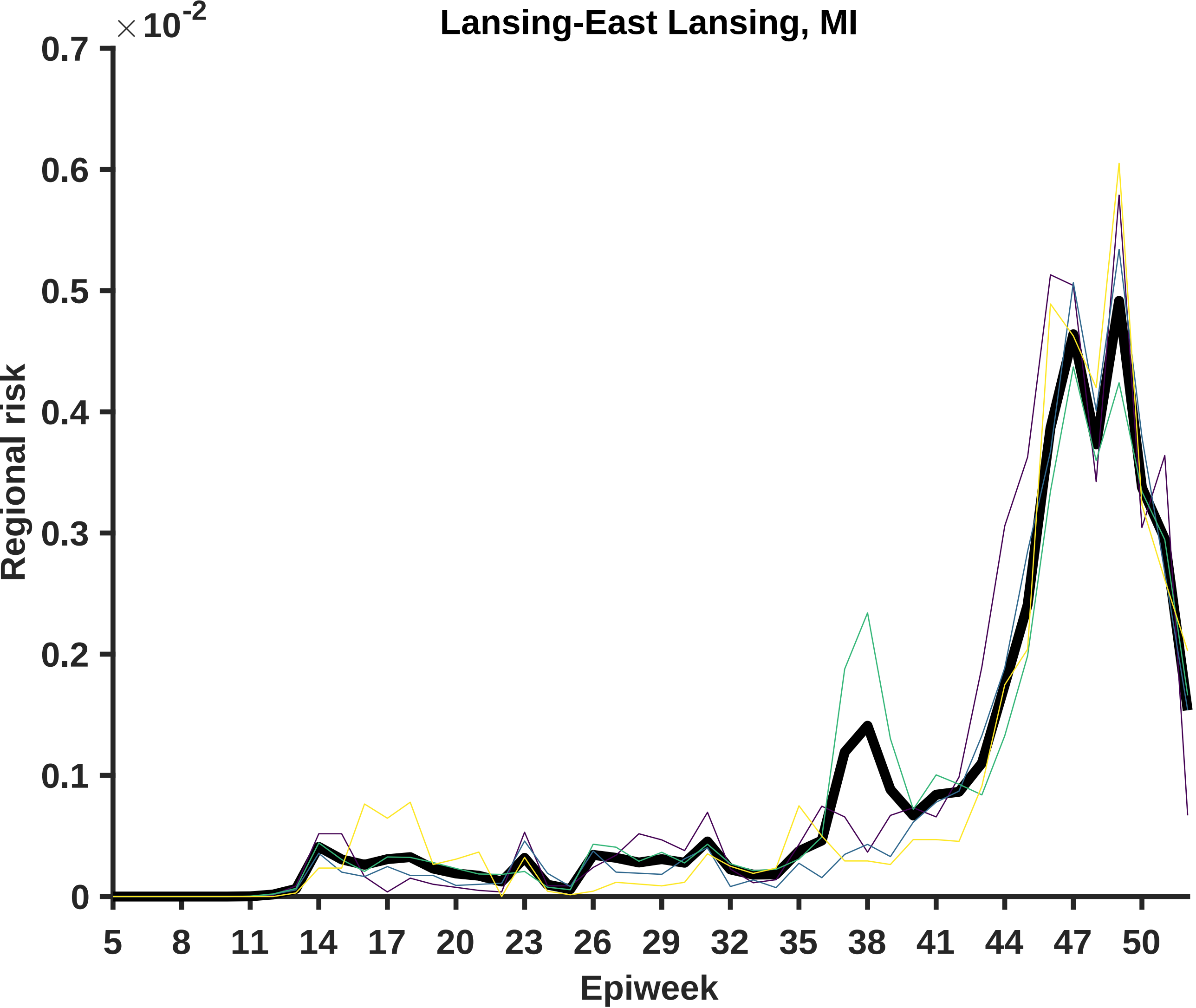

Lansing-East Lansing, MI
×10^-2
0.7
0.6
0.5
0.4
0.3
0.2
0.1
0
Regional risk
5
8
11
14
17
20
23
26
29
32
35
38
41
44
47
50
Epiweek

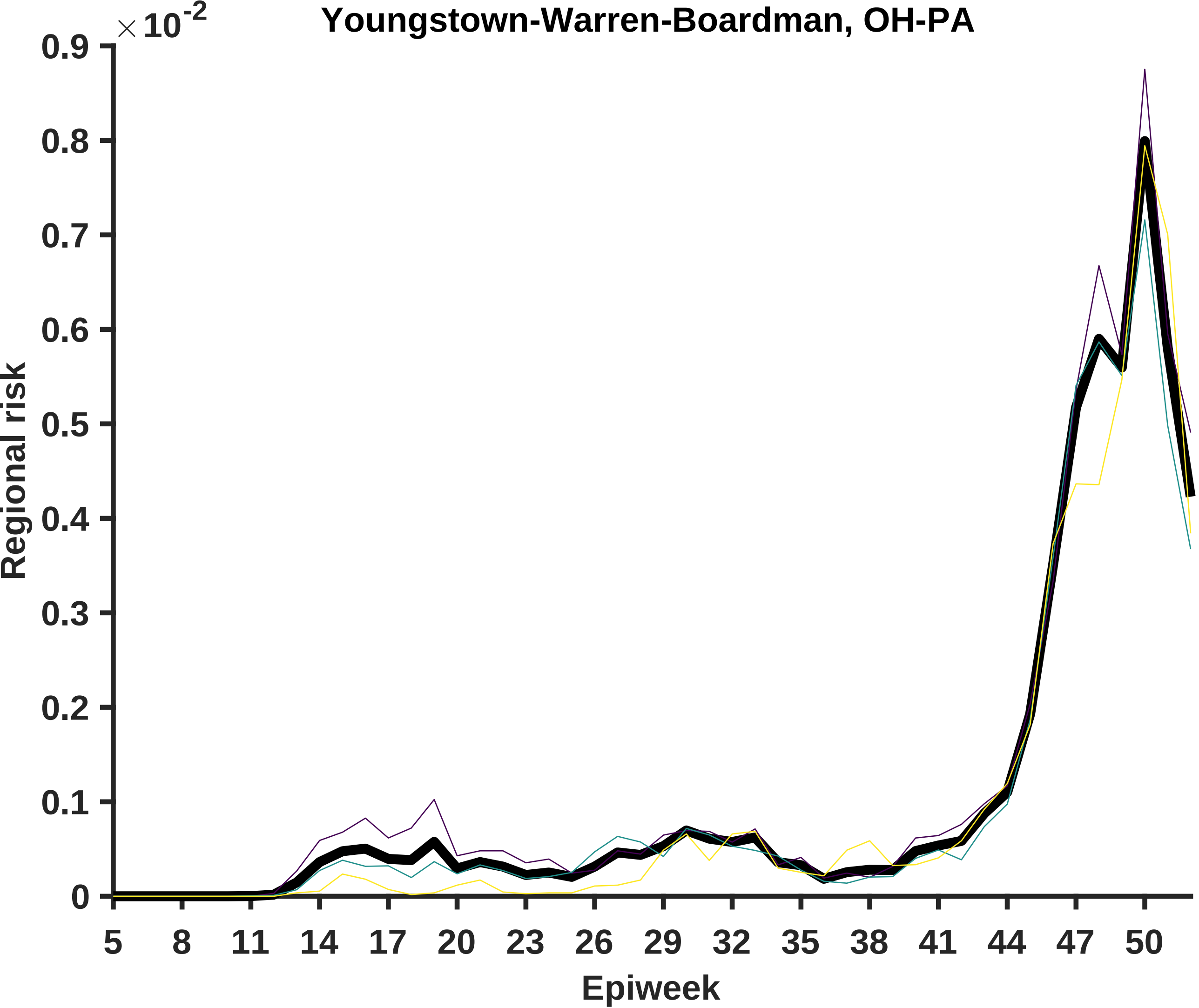

Youngstown-Warren-Boardman, OH-PA
×10-2
0.9
0.8
0.7
0.6
0.5
0.4
0.3
0.2
0.1
0
Regional risk
5
8
11
14
17
20
23
26
29
32
35
38
41
44
47
50
Epiweek

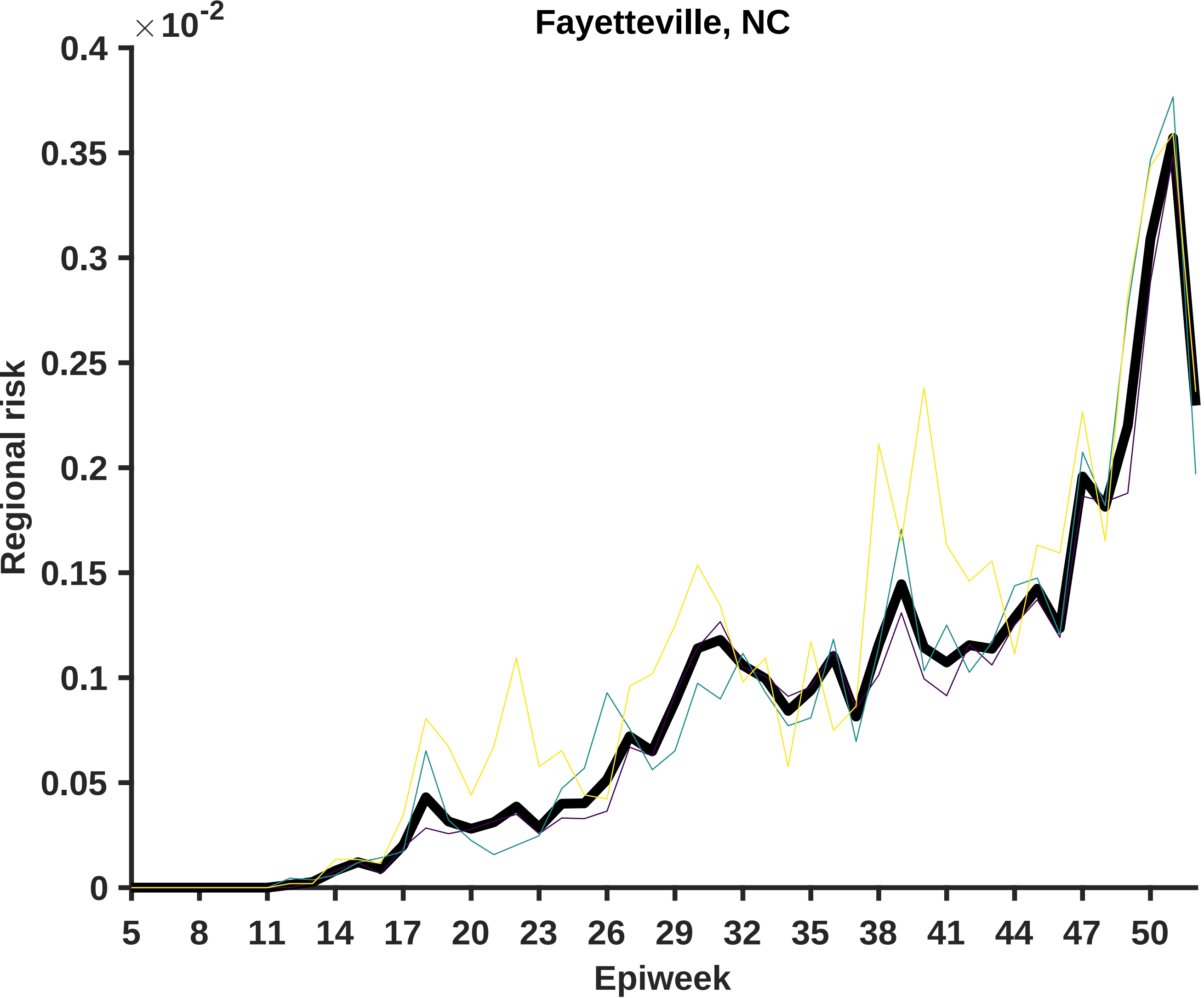
Fayetteville, NC
×10^-2
0.4
0.35
0.3
0.25
0.2
0.15
0.1
0.05
0
Regional risk
5
8
11
14
17
20
23
26
29
32
35
38
41
44
47
50
Epiweek

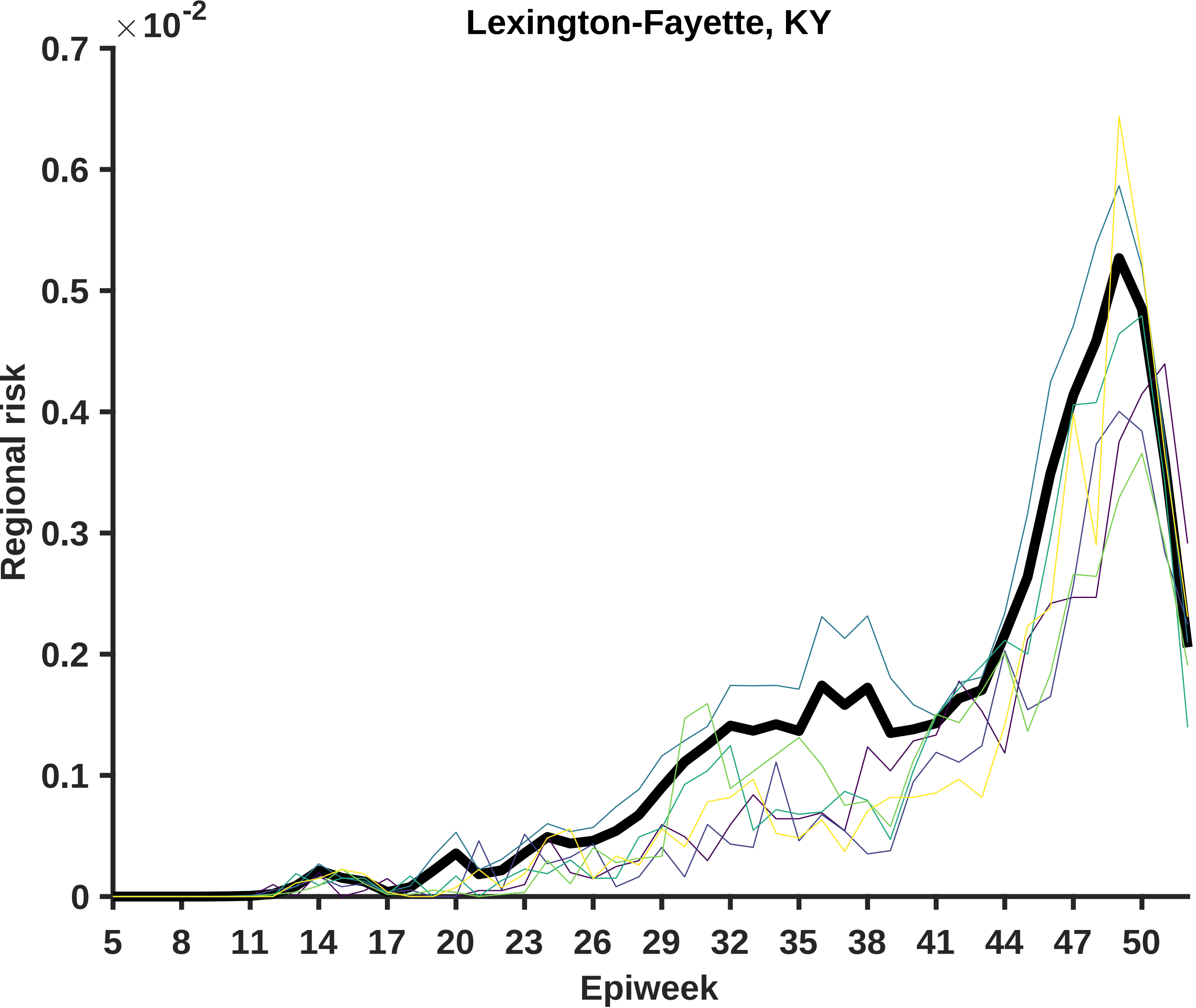
Lexington-Fayette, KY
×10^-2
0.7
0.6
0.5
0.4
0.3
0.2
0.1
0
Regional risk
5
8
11
14
17
20
23
26
29
32
35
38
41
44
47
50
Epiweek

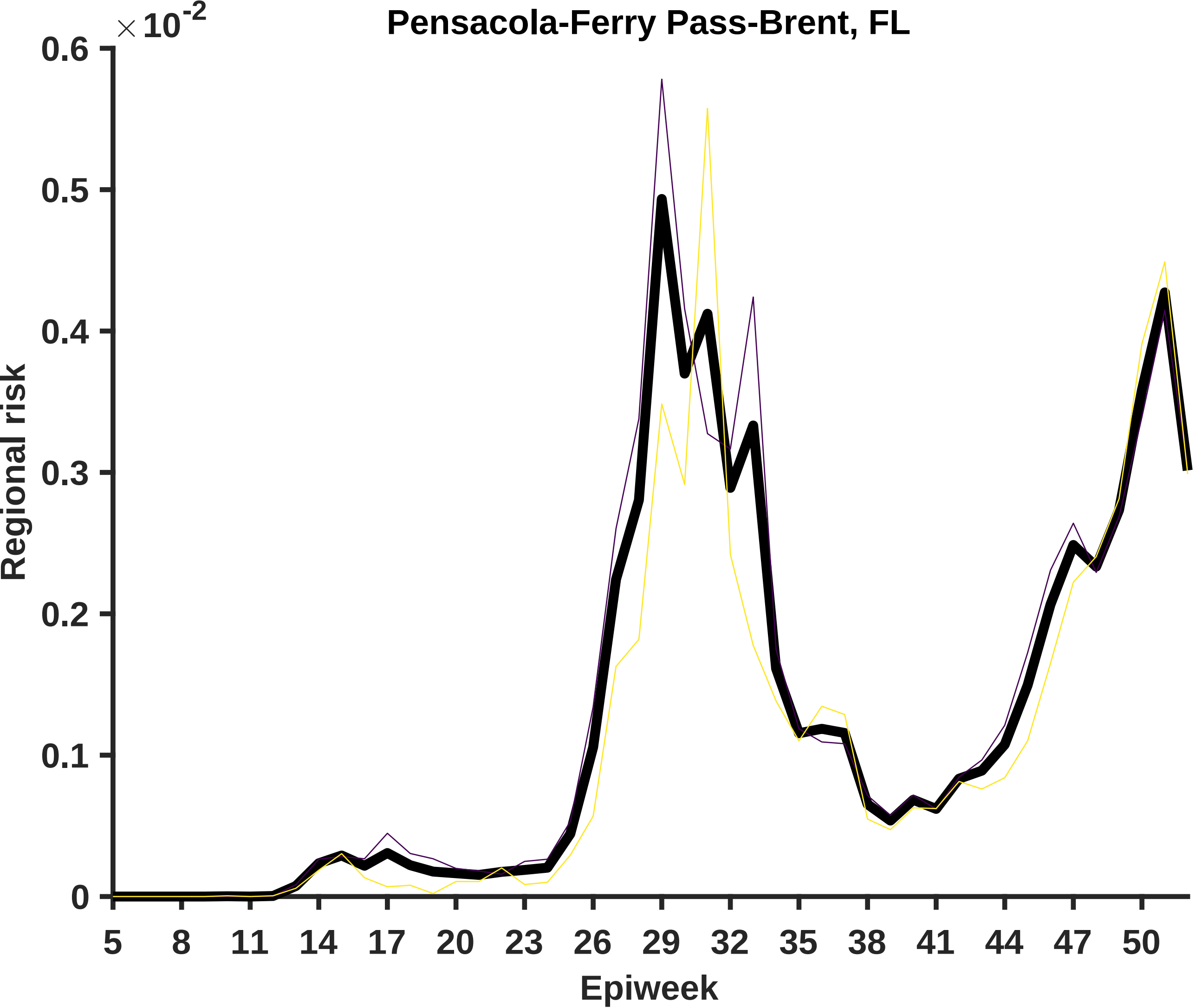
Pensacola-Ferry Pass-Brent, FL
×10^-2
0.6
0.5
0.4
0.3
0.2
0.1
0
Regional risk
5
8
11
14
17
20
23
26
29
32
35
38
41
44
47
50
Epiweek

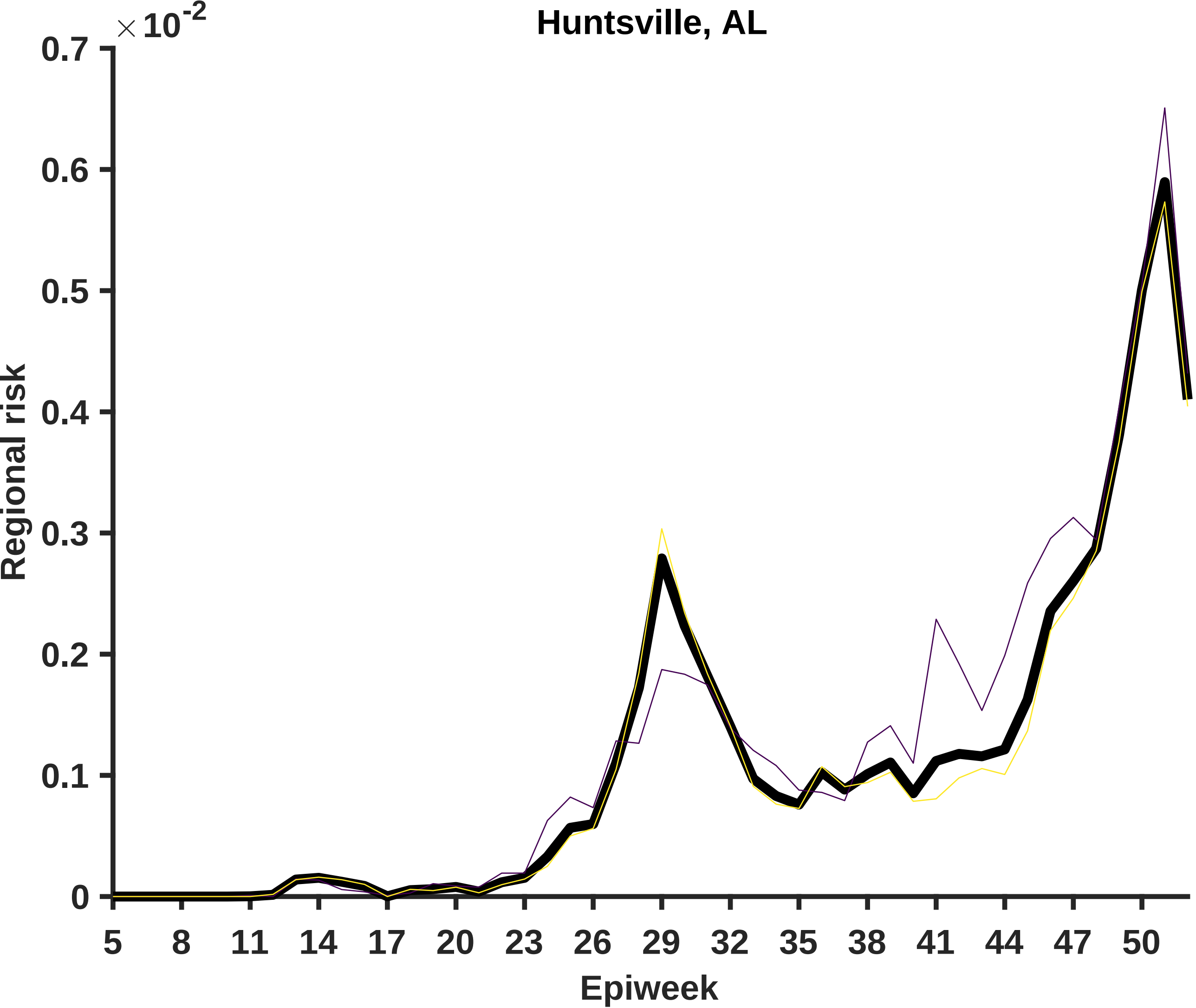

Huntsville, AL
×10⁻²
0.7
0.6
0.5
0.4
0.3
0.2
0.1
0
Regional risk
5
8
11
14
17
20
23
26
29
32
35
38
41
44
47
50
Epiweek

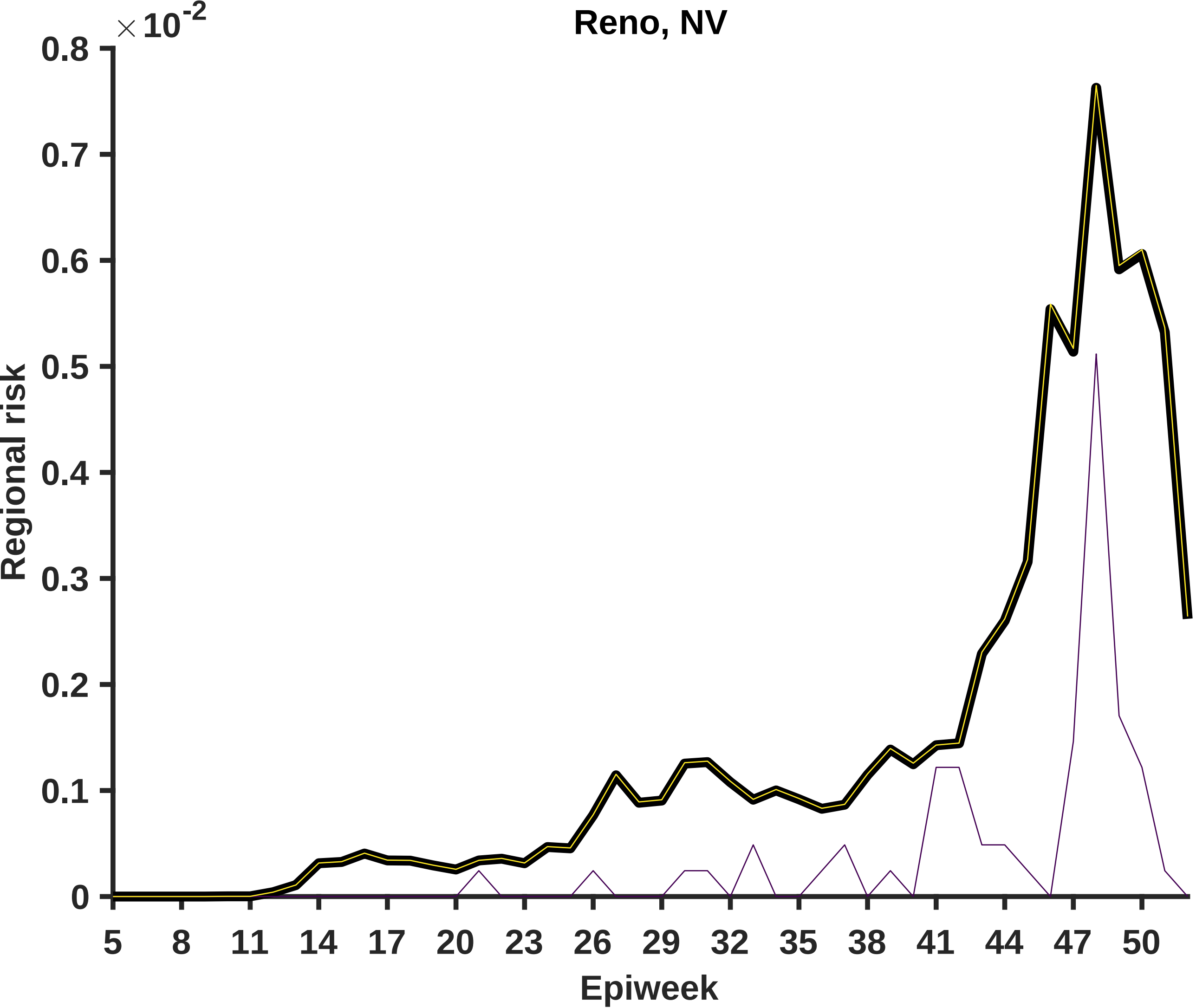

Reno, NV
×10^-2
Regional risk
Epiweek
0.8
0.7
0.6
0.5
0.4
0.3
0.2
0.1
0
5
8
11
14
17
20
23
26
29
32
35
38
41
44
47
50

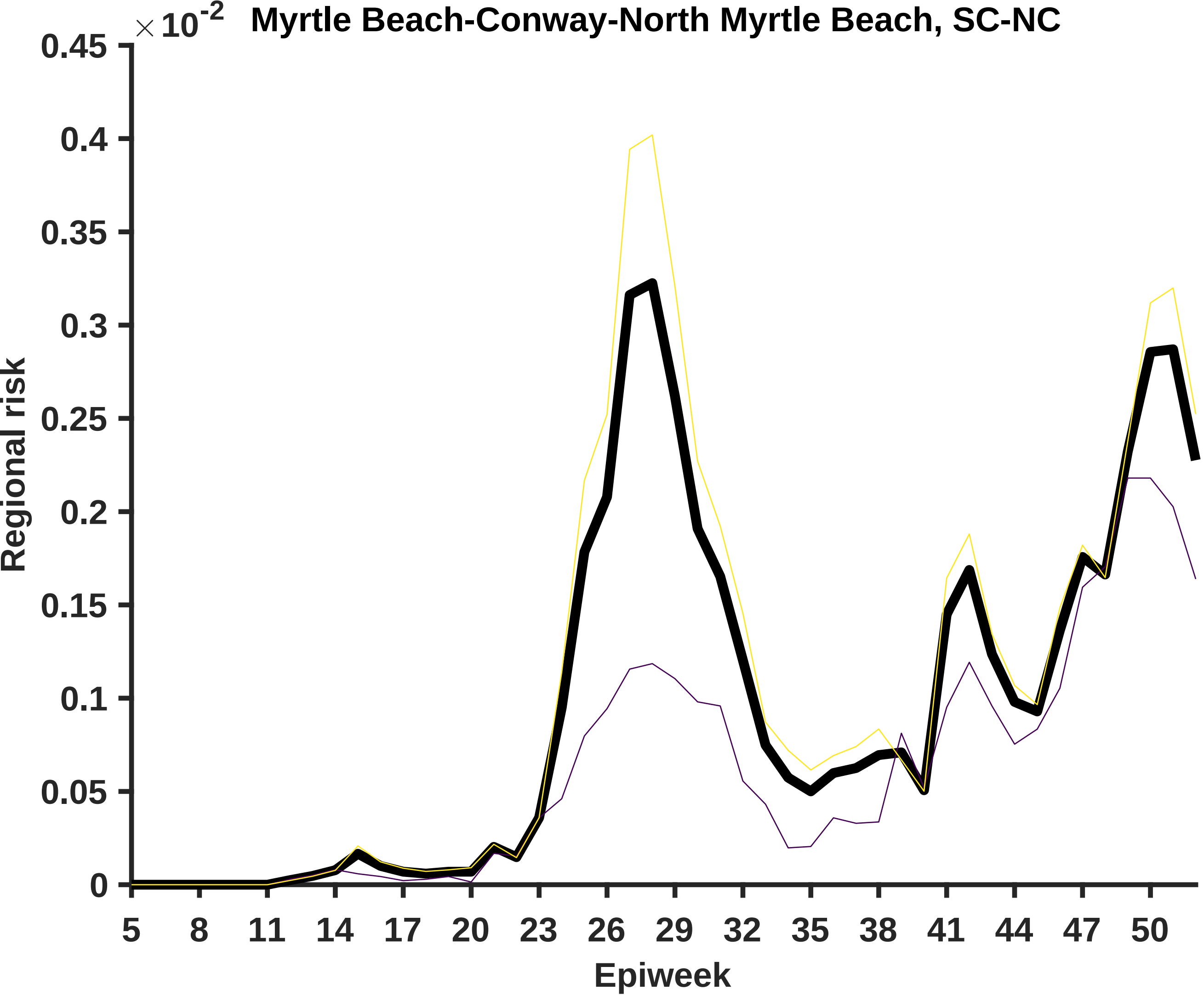

Myrtle Beach-Conway-North Myrtle Beach, SC-NC
× 10-2
0.45
0.4
0.35
0.3
0.25
0.2
0.15
0.1
0.05
0
Regional risk
5
8
11
14
17
20
23
26
29
32
35
38
41
44
47
50
Epiweek

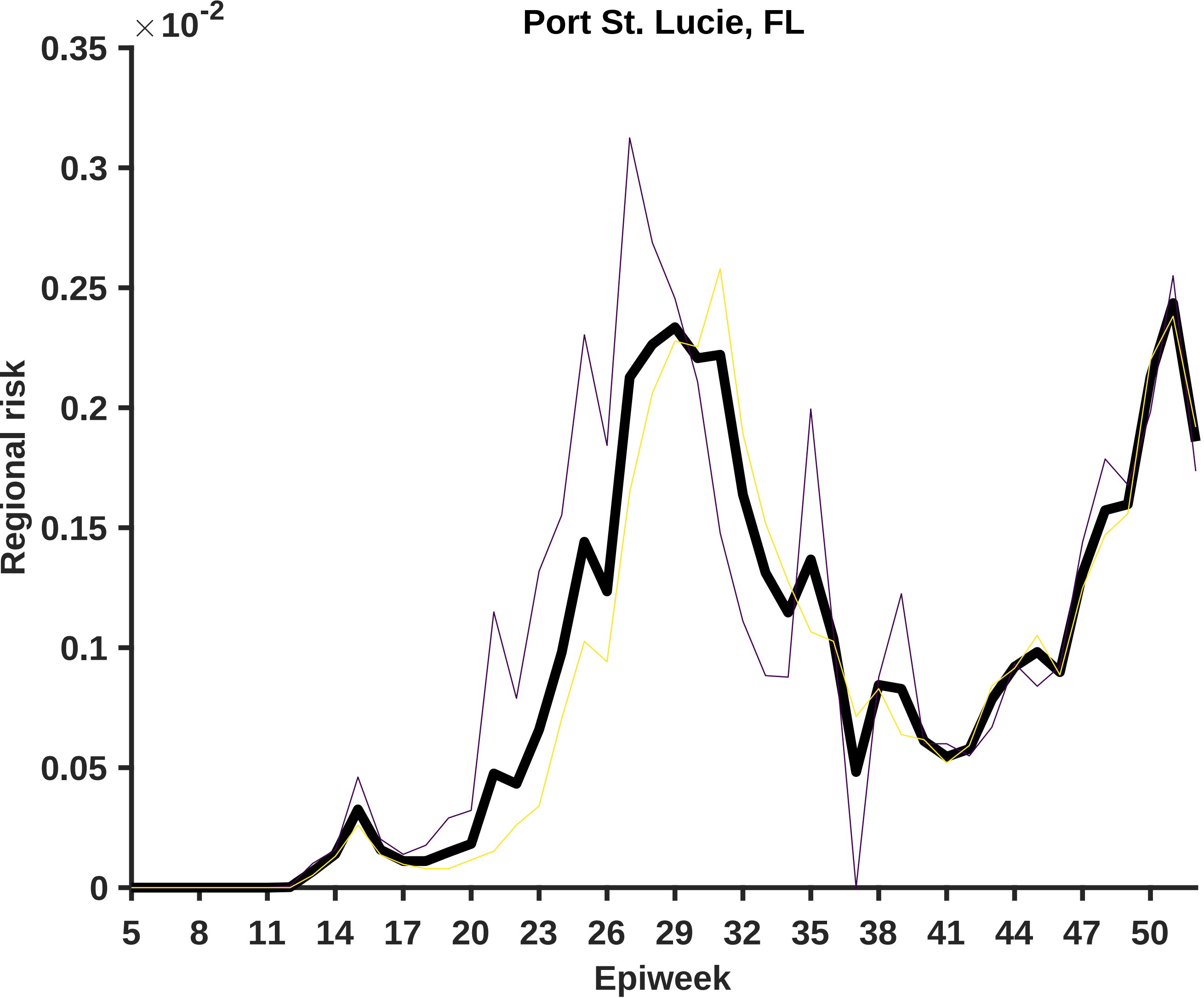

Port St. Lucie, FL
×10^-2
0.35
0.3
0.25
0.2
0.15
0.1
0.05
0
Regional risk
5
8
11
14
17
20
23
26
29
32
35
38
41
44
47
50
Epiweek

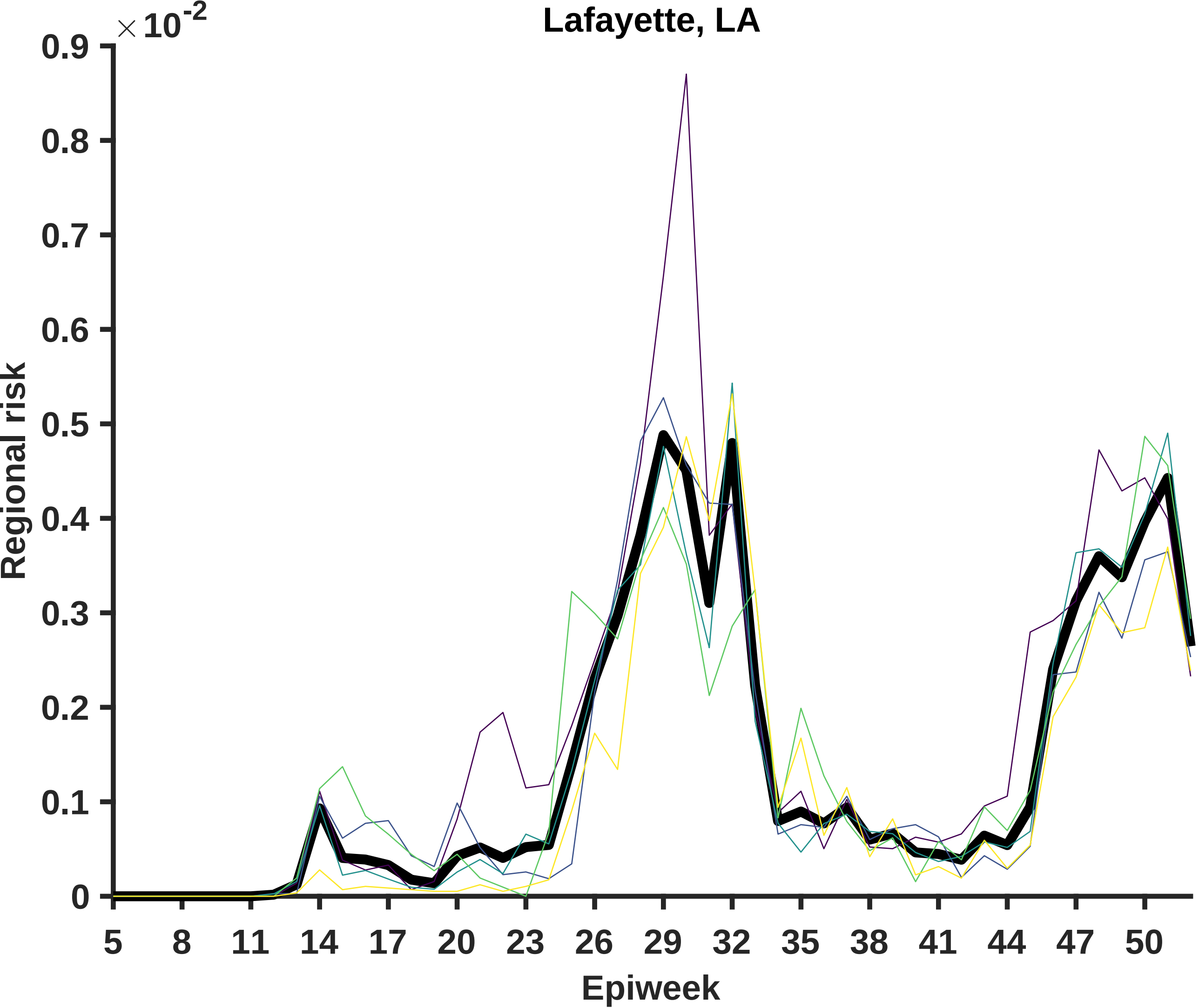
Lafayette, LA
×10^-2
0.9
0.8
0.7
0.6
0.5
0.4
0.3
0.2
0.1
0
Regional risk
5
8
11
14
17
20
23
26
29
32
35
38
41
44
47
50
Epiweek

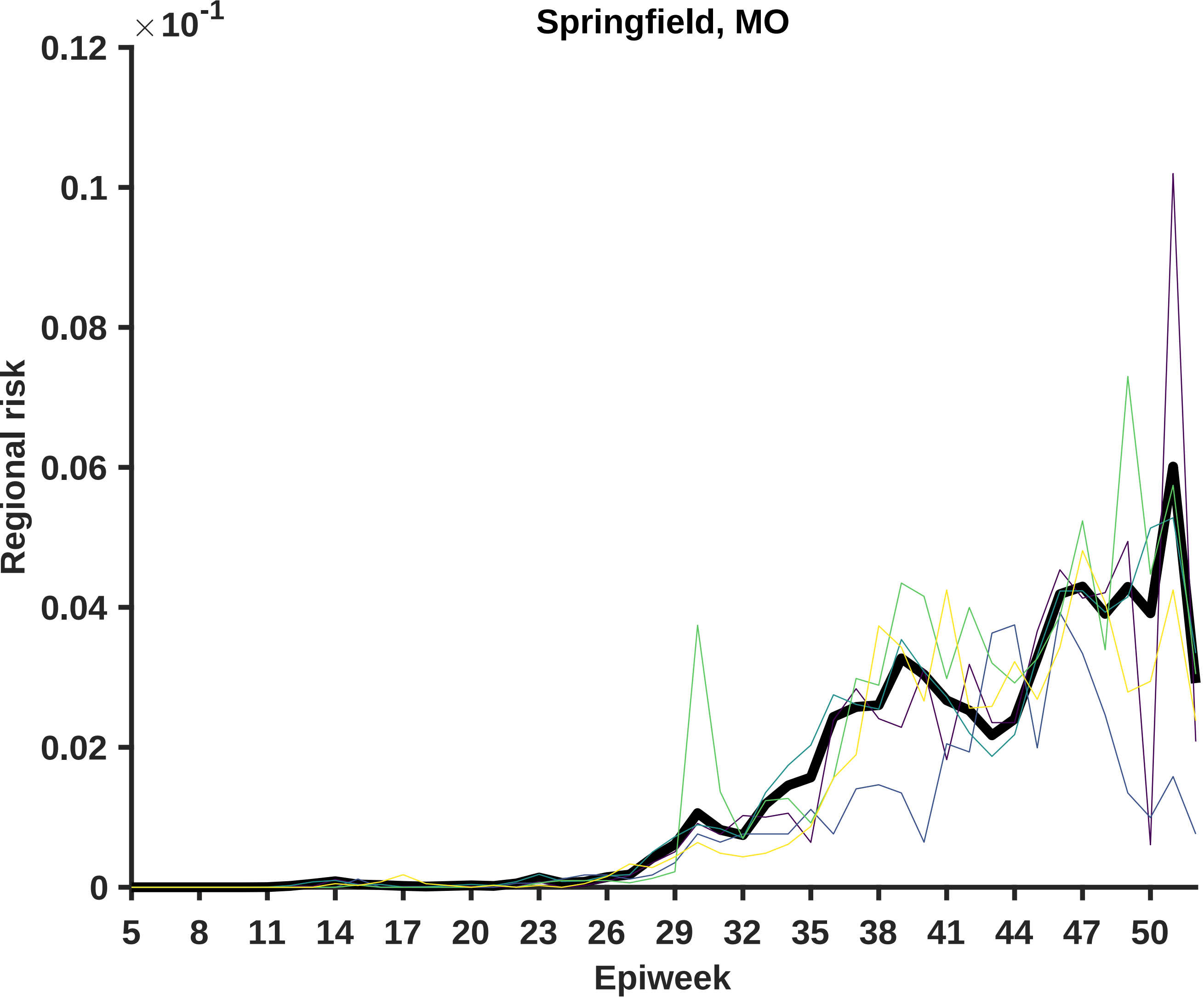
Springfield, MO
× 10^-1
0.12
0.1
0.08
0.06
0.04
0.02
0
Regional risk
5
8
11
14
17
20
23
26
29
32
35
38
41
44
47
50
Epiweek

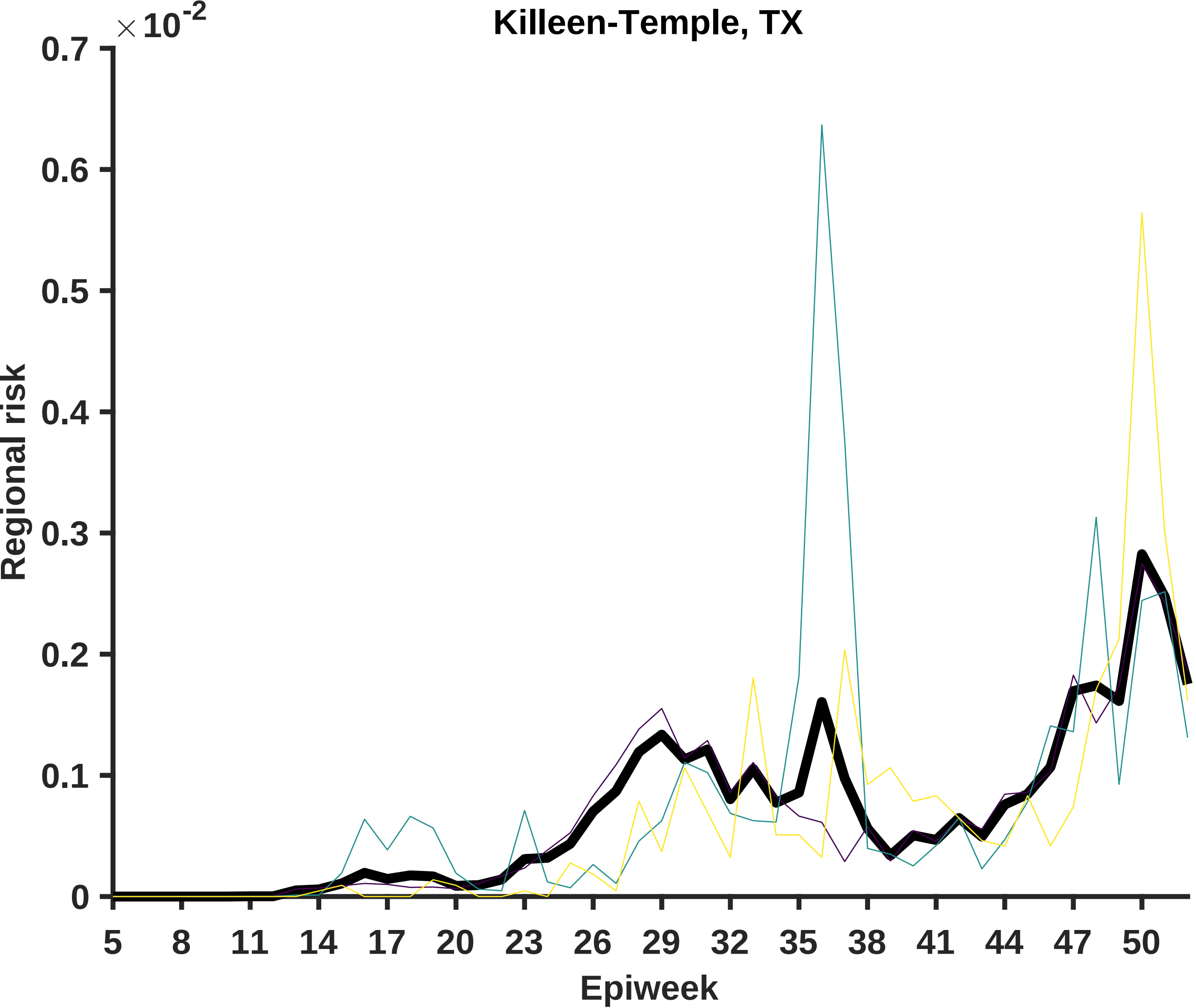

Killeen-Temple, TX
×10^-2
0.7
0.6
0.5
0.4
0.3
0.2
0.1
0
Regional risk
5
8
11
14
17
20
23
26
29
32
35
38
41
44
47
50
Epiweek

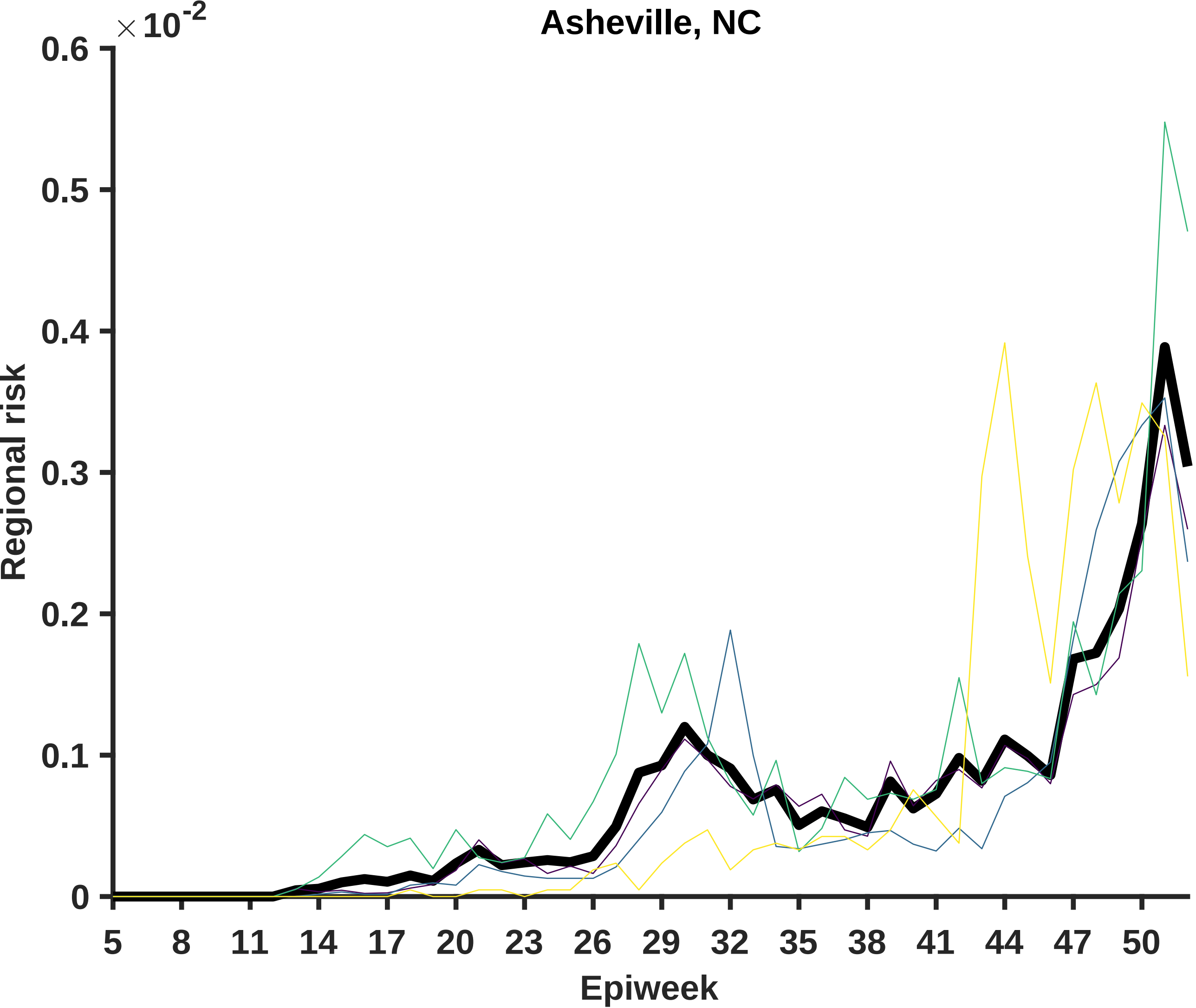

Asheville, NC
× 10^-2
0.6
0.5
0.4
0.3
0.2
0.1
0
Regional risk
5
8
11
14
17
20
23
26
29
32
35
38
41
44
47
50
Epiweek

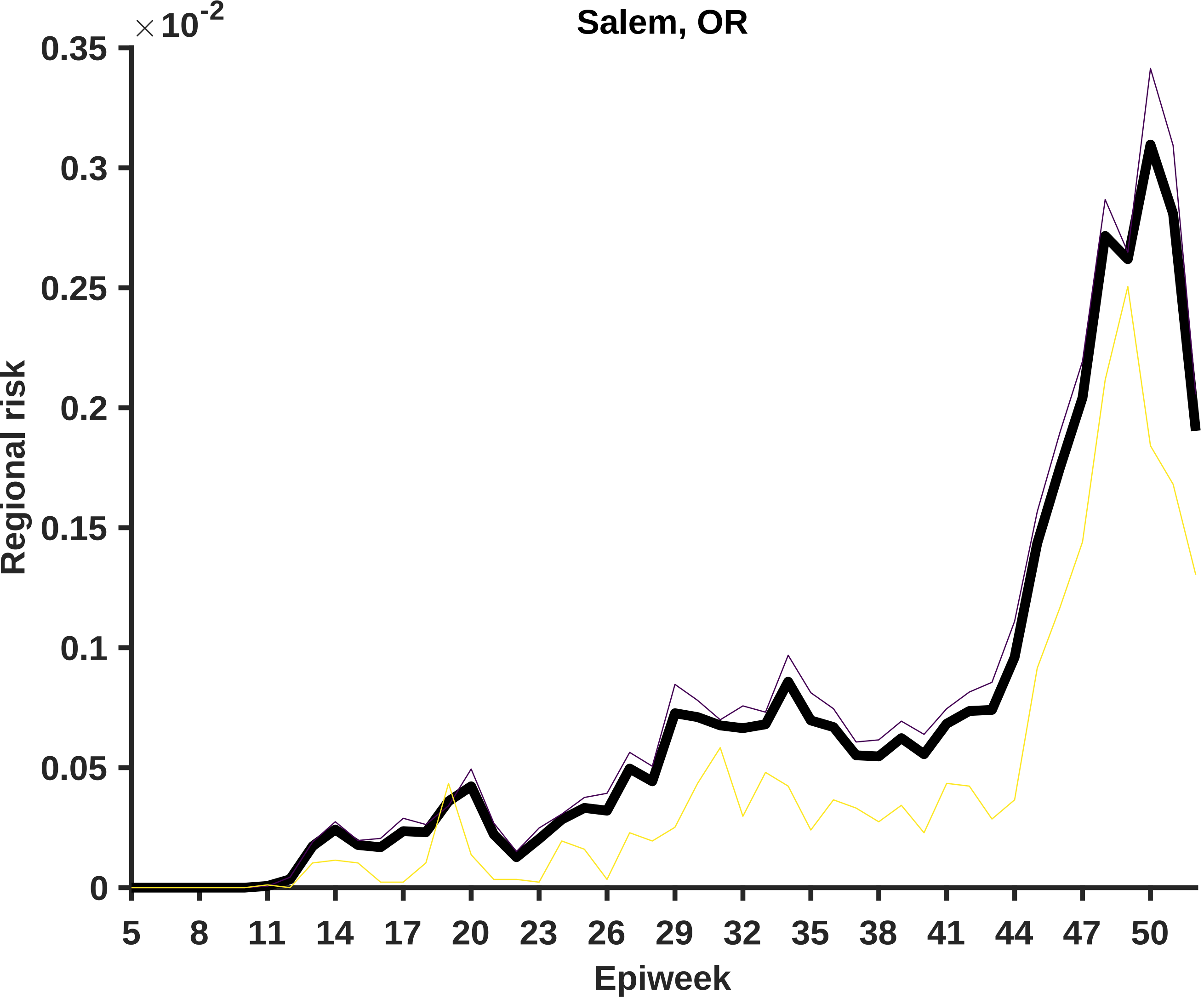
Salem, OR
×10^-2
Regional risk
0.35
0.3
0.25
0.2
0.15
0.1
0.05
0
5
8
11
14
17
20
23
26
29
32
35
38
41
44
47
50
Epiweek

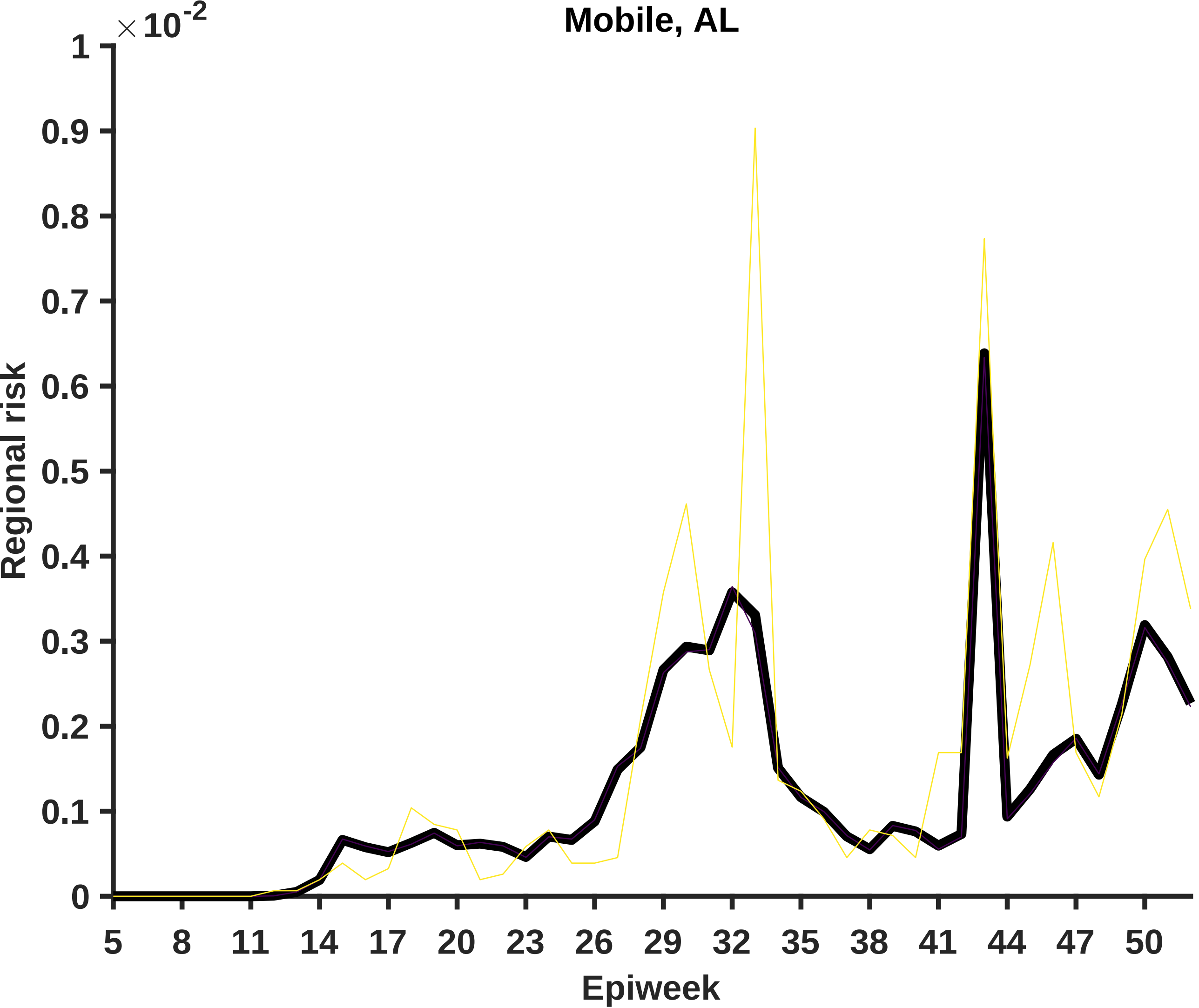

Mobile, AL
×10^-2
Regional risk
Epiweek
0
0.1
0.2
0.3
0.4
0.5
0.6
0.7
0.8
0.9
1
5
8
11
14
17
20
23
26
29
32
35
38
41
44
47
50

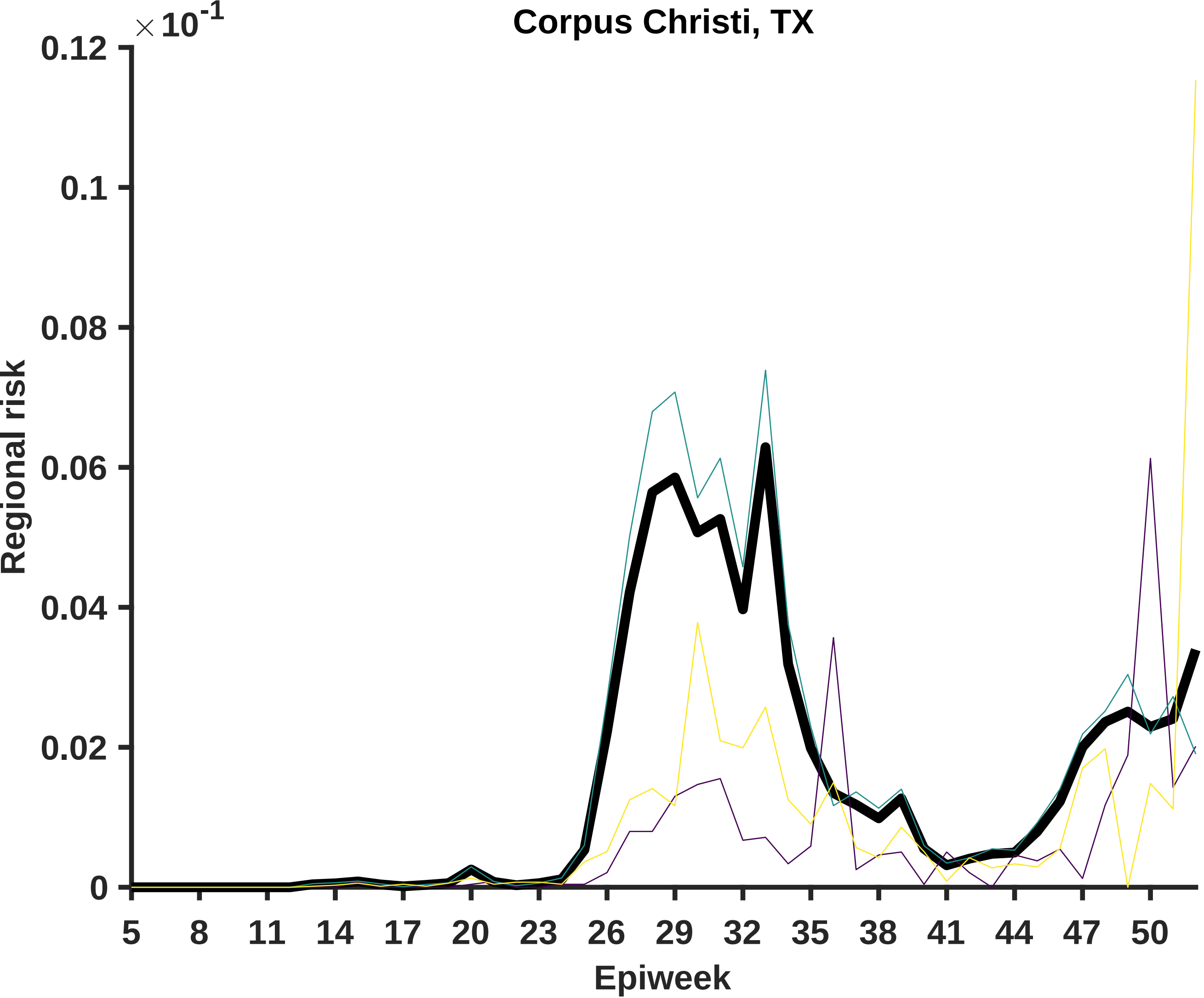

Corpus Christi, TX
× 10^-1
0.12
0.1
0.08
0.06
0.04
0.02
0
Regional risk
5
8
11
14
17
20
23
26
29
32
35
38
41
44
47
50
Epiweek

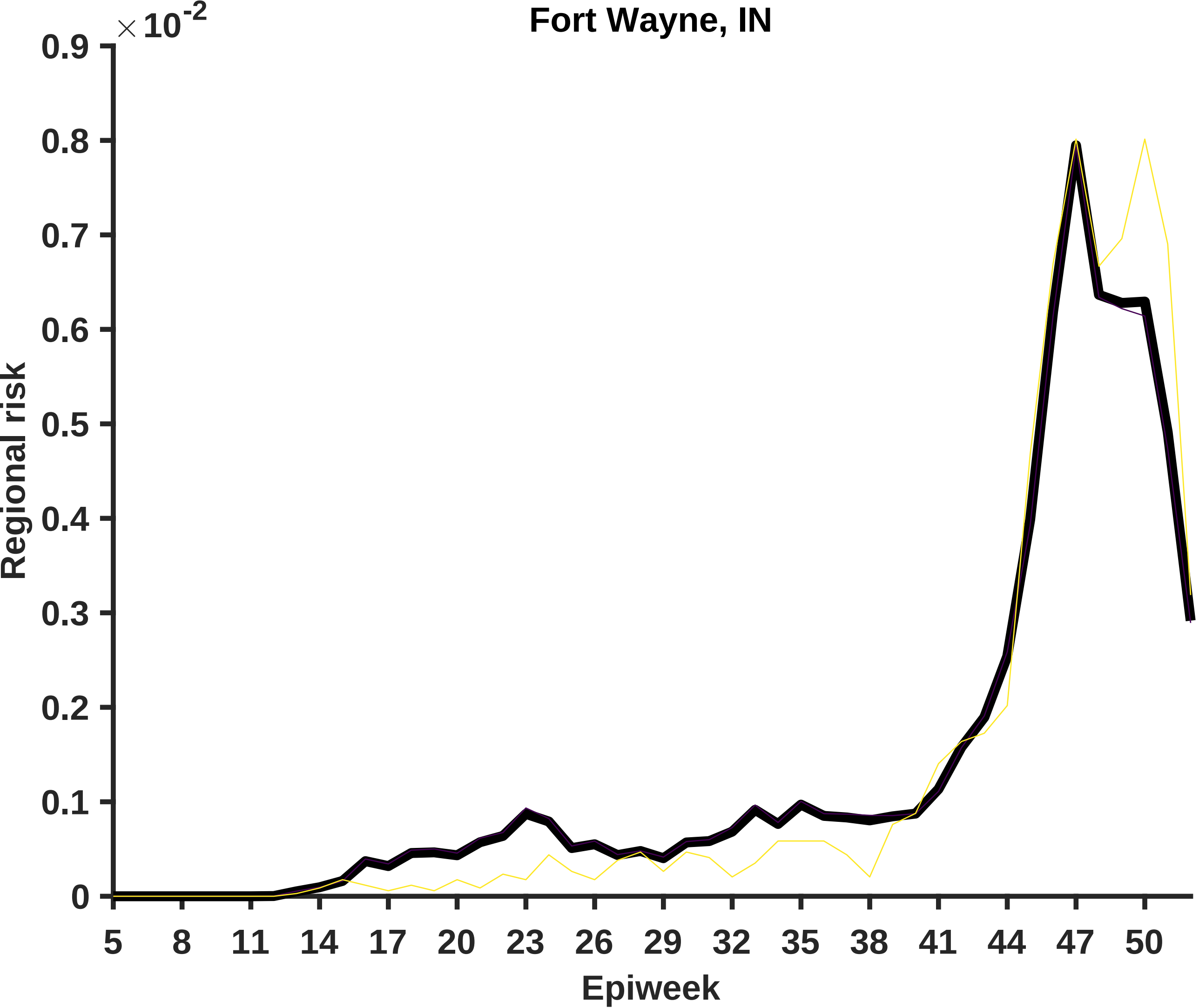
Fort Wayne, IN
×10^-2
0.9
0.8
0.7
0.6
0.5
0.4
0.3
0.2
0.1
0
Regional risk
5
8
11
14
17
20
23
26
29
32
35
38
41
44
47
50
Epiweek

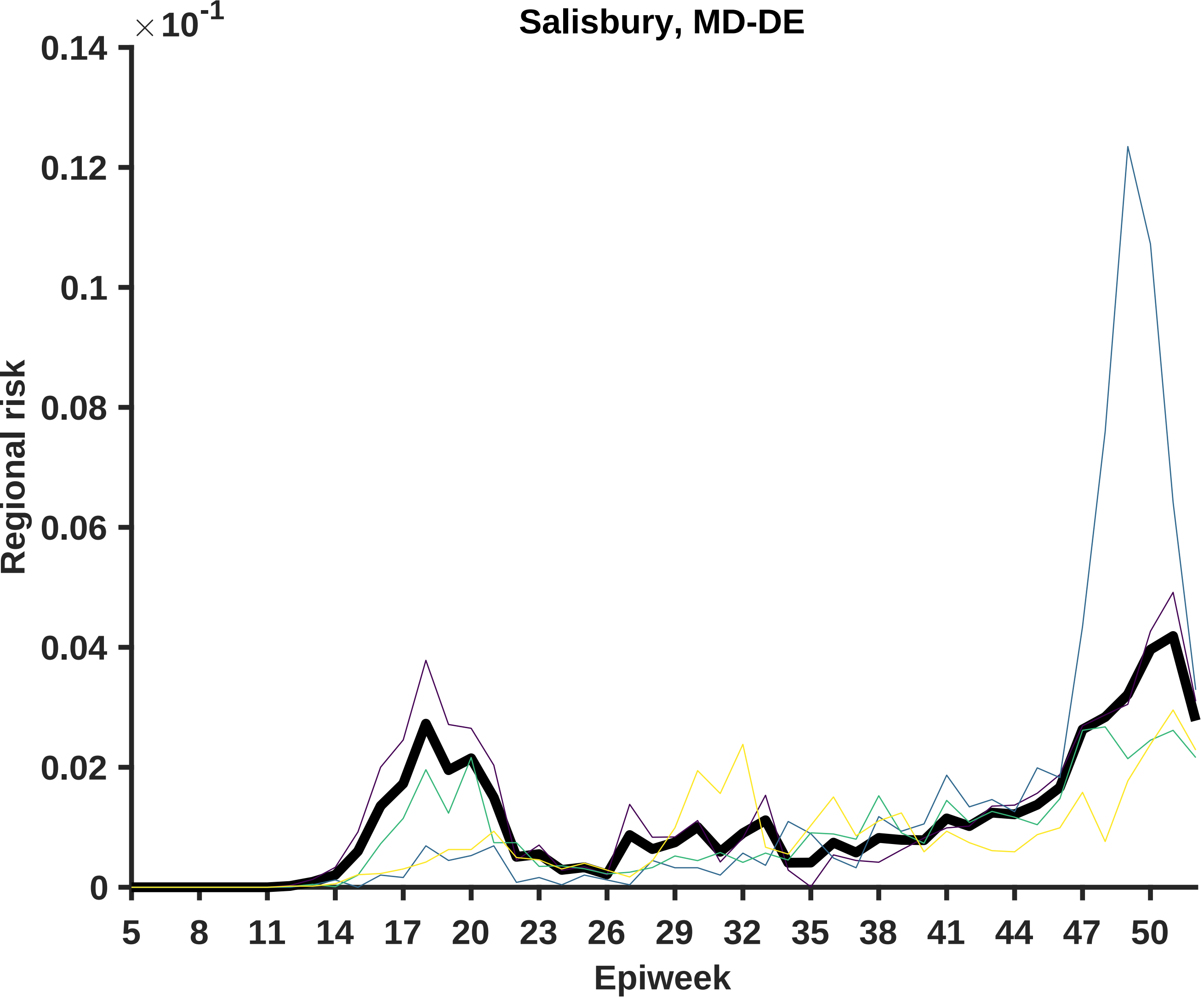

Salisbury, MD-DE
×10-1
0.14
0.12
0.1
0.08
0.06
0.04
0.02
0
Regional risk
5
8
11
14
17
20
23
26
29
32
35
38
41
44
47
50
Epiweek

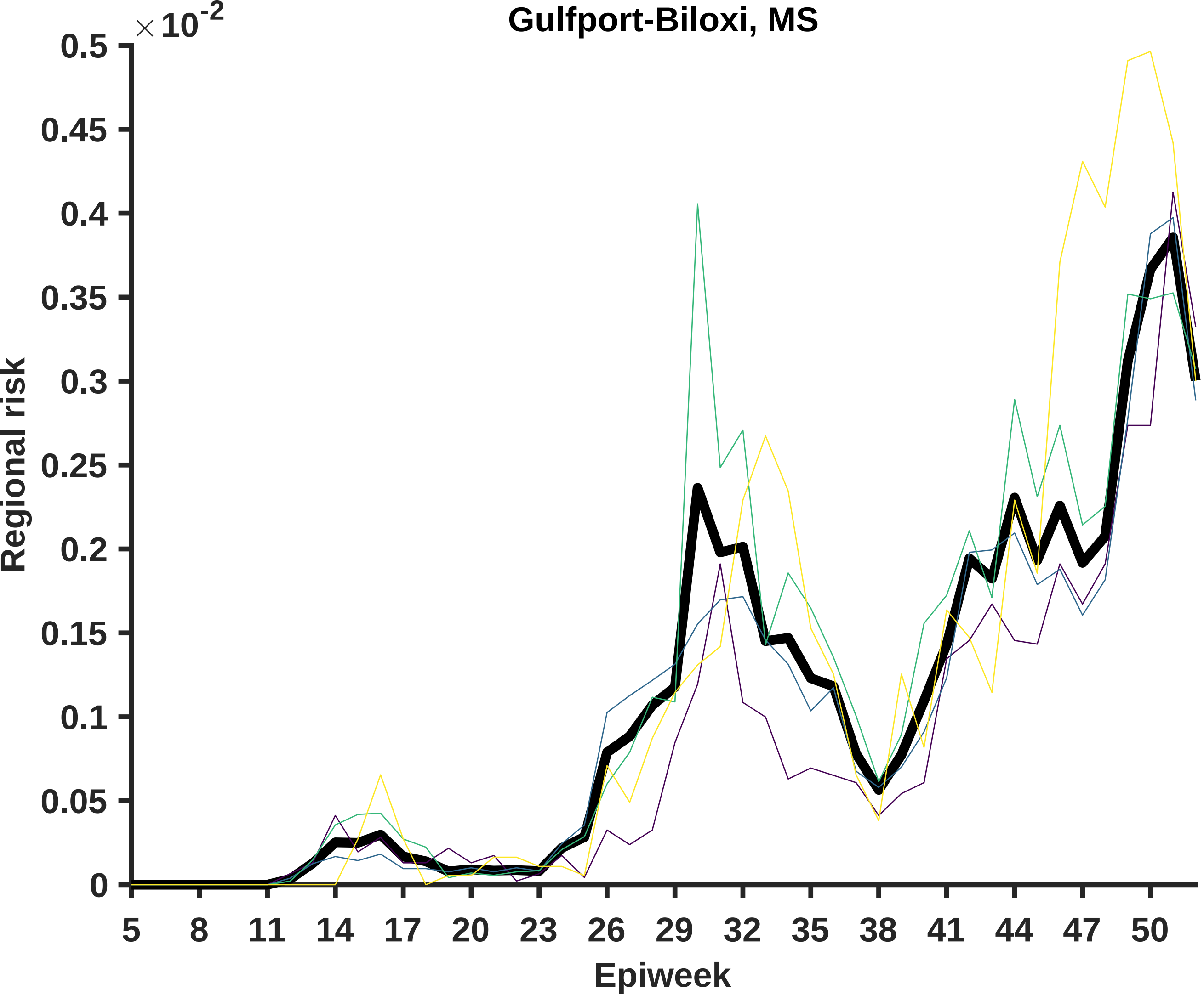
Gulfport-Biloxi, MS
× 10^-2
0.5
0.45
0.4
0.35
0.3
0.25
0.2
0.15
0.1
0.05
0
Regional risk
5
8
11
14
17
20
23
26
29
32
35
38
41
44
47
50
Epiweek

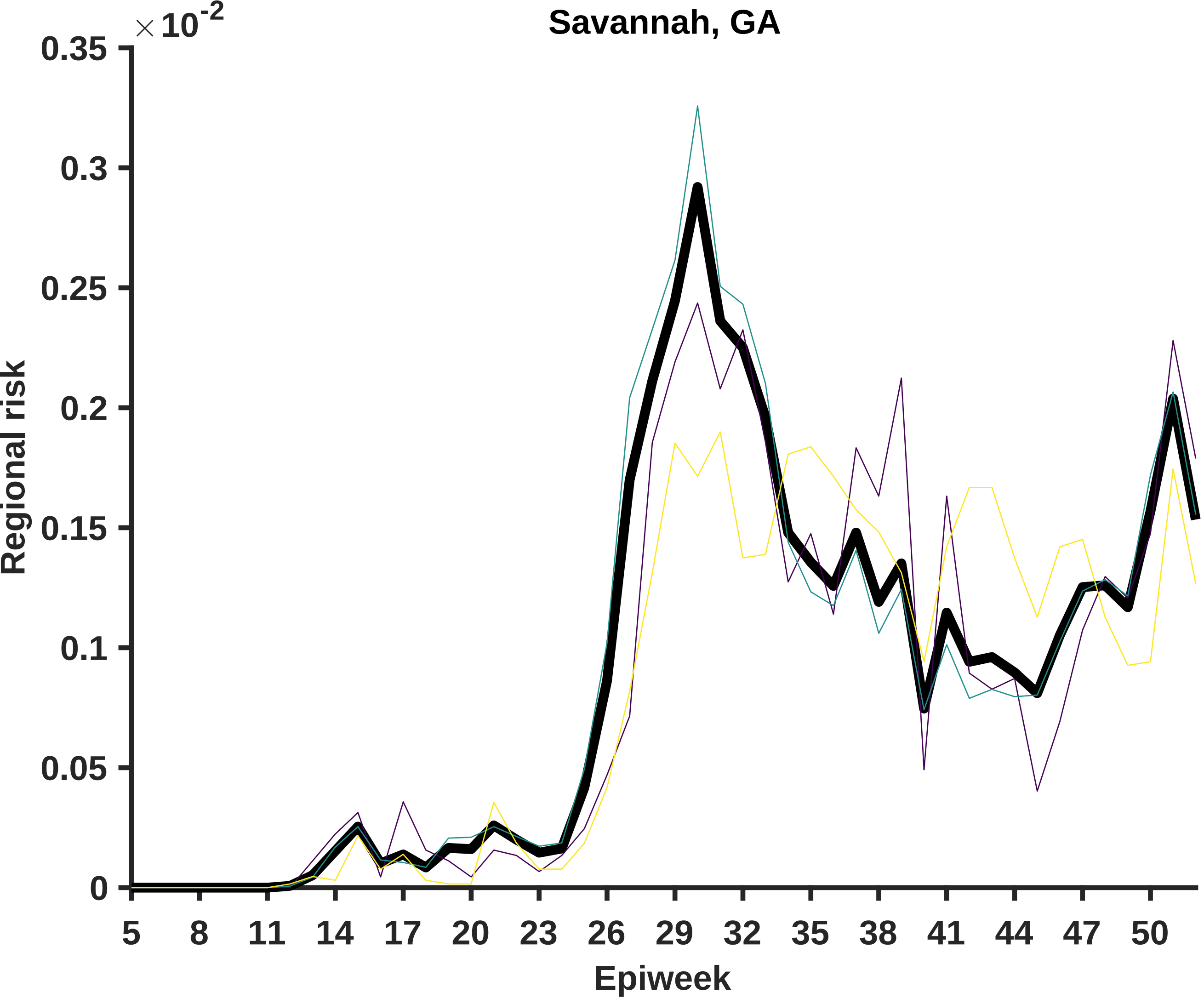
Savannah, GA
×10^-2
0.35
0.3
0.25
0.2
0.15
0.1
0.05
0
Regional risk
5
8
11
14
17
20
23
26
29
32
35
38
41
44
47
50
Epiweek

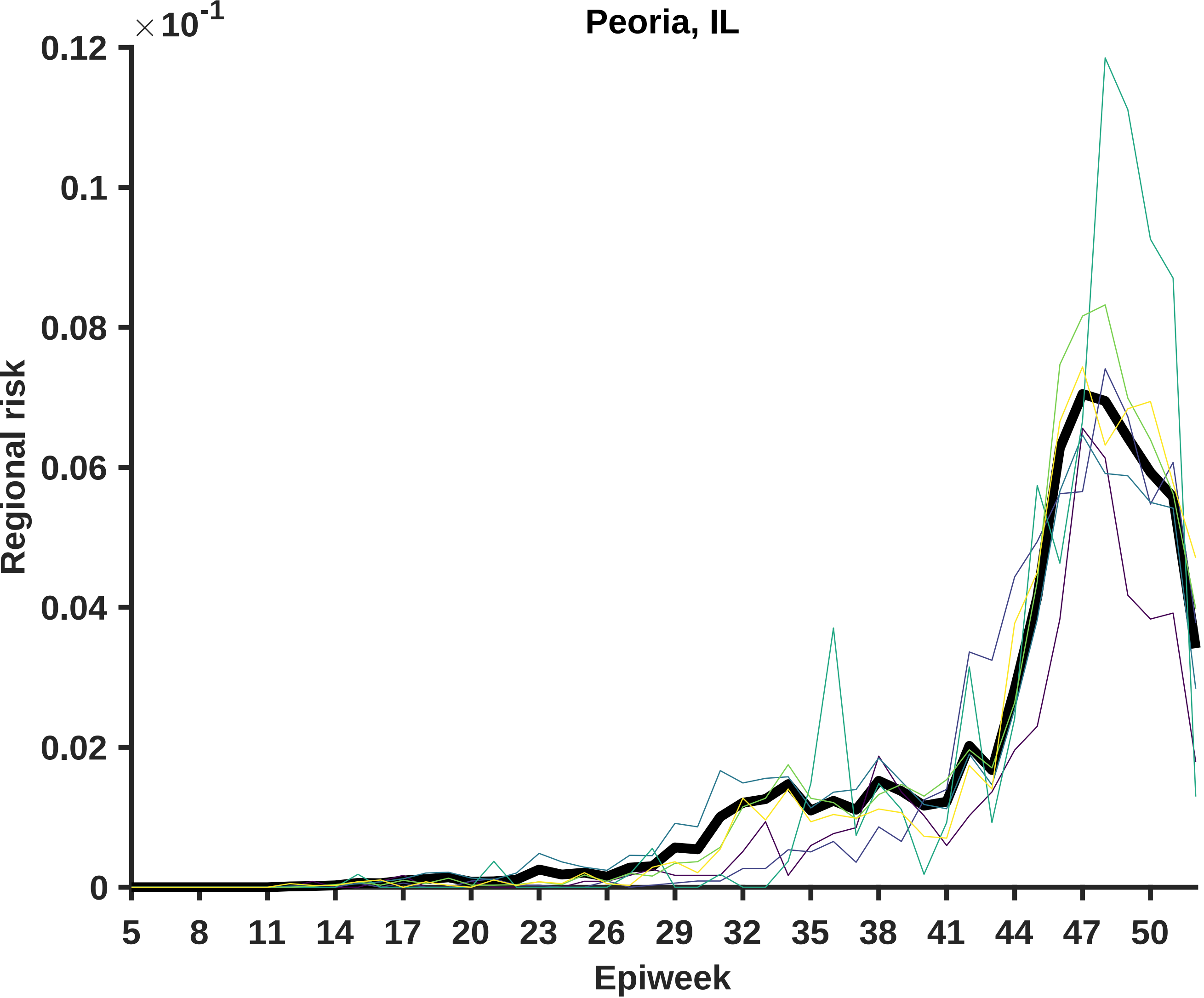
Peoria, IL
×10^-1
Regional risk
0.12
0.1
0.08
0.06
0.04
0.02
0
5
8
11
14
17
20
23
26
29
32
35
38
41
44
47
50
Epiweek

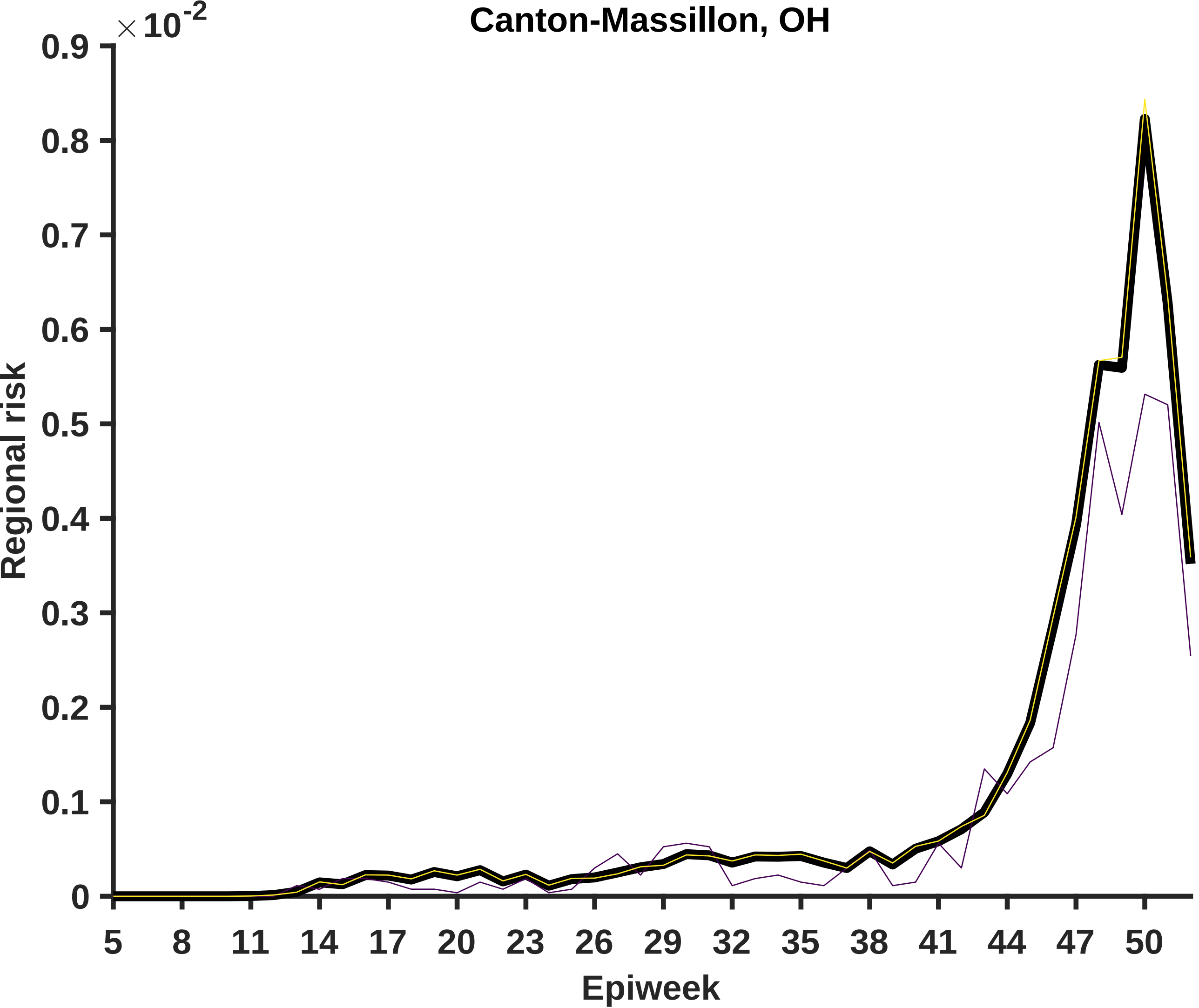
Canton-Massillon, OH
×10^-2
0.9
0.8
0.7
0.6
0.5
0.4
0.3
0.2
0.1
0
Regional risk
5
8
11
14
17
20
23
26
29
32
35
38
41
44
47
50
Epiweek

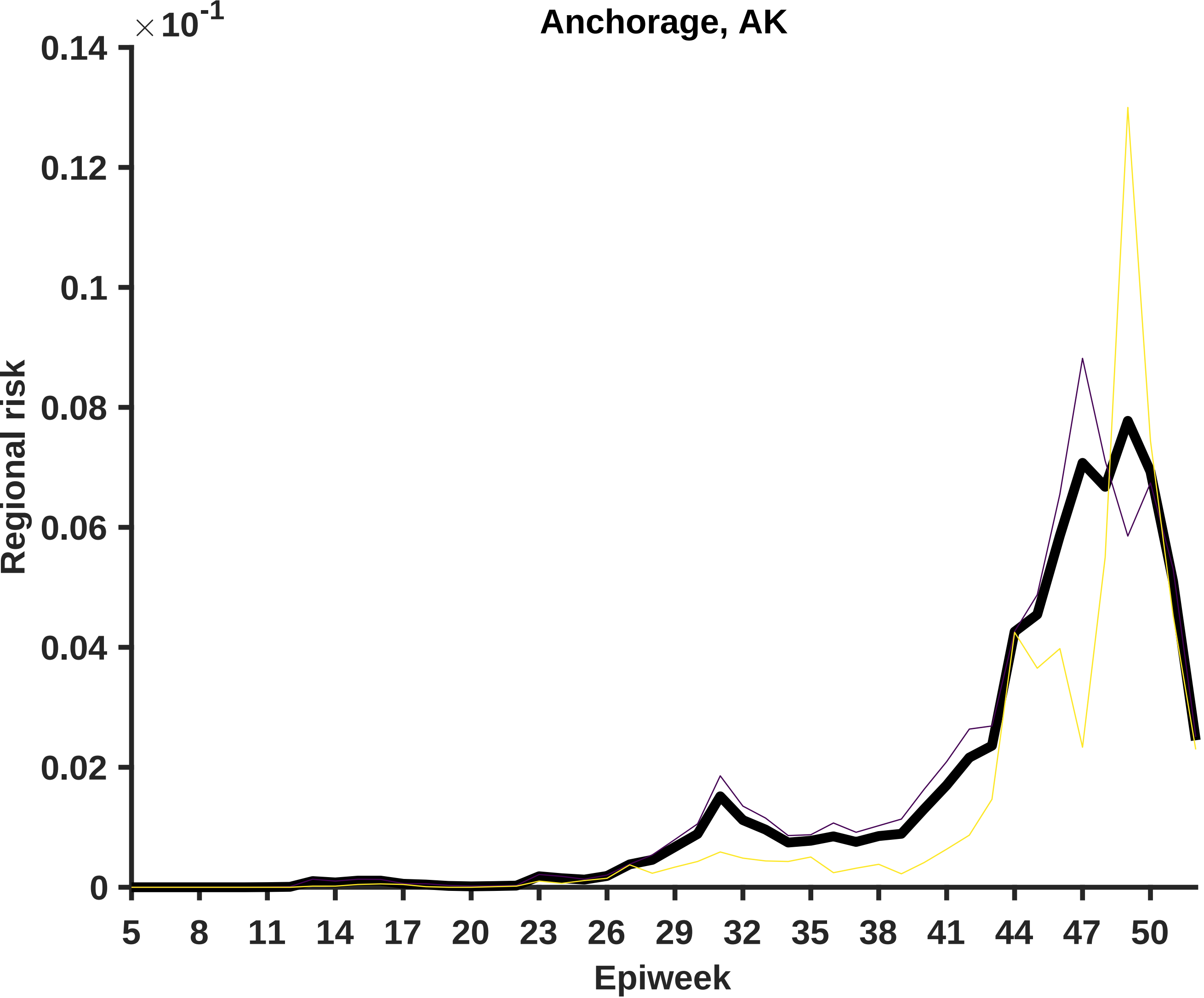

Anchorage, AK
×10^-1
Regional risk
0.14
0.12
0.1
0.08
0.06
0.04
0.02
0
5
8
11
14
17
20
23
26
29
32
35
38
41
44
47
50
Epiweek

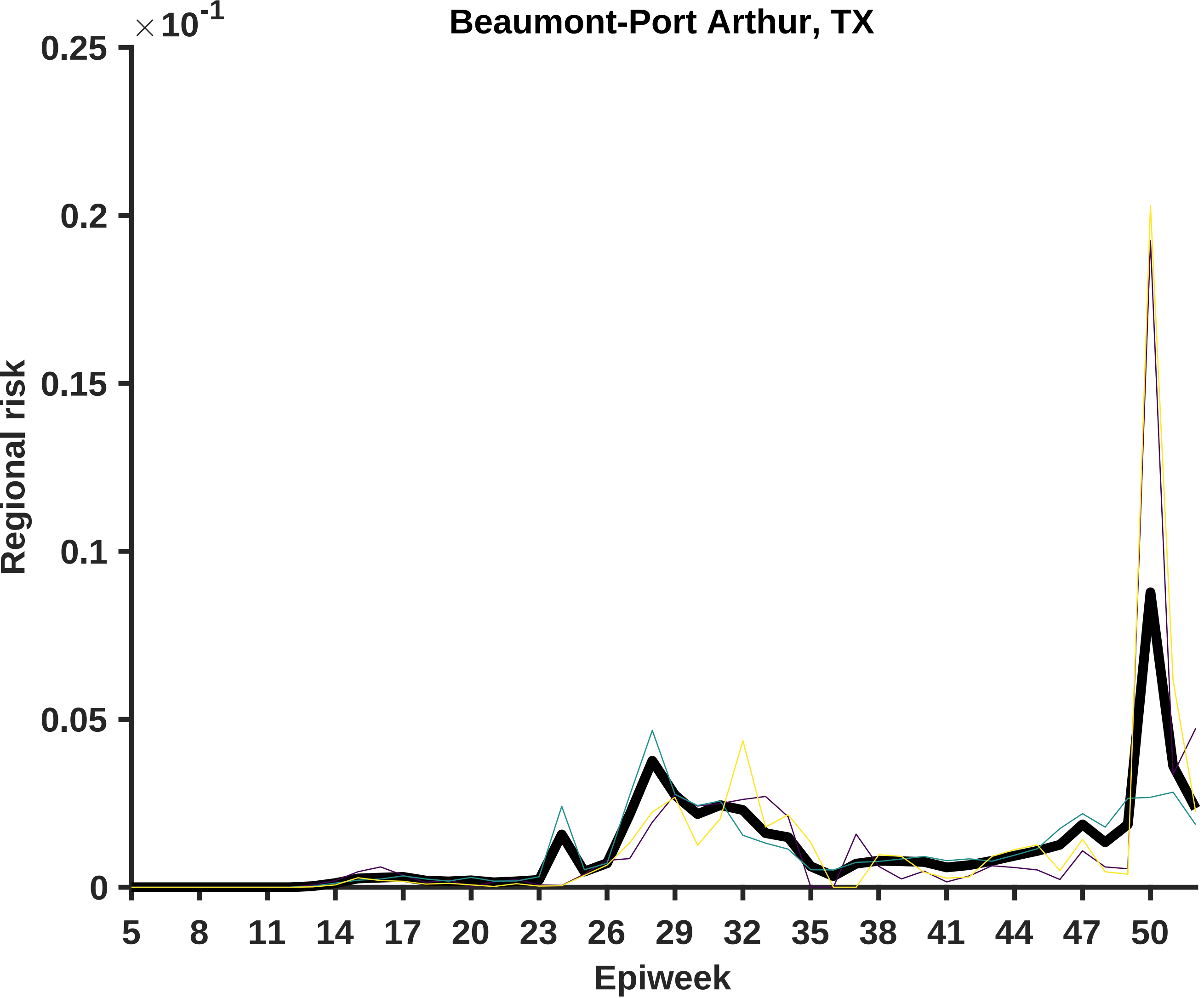

Beaumont-Port Arthur, TX
×10^-1
0.25
0.2
0.15
0.1
0.05
0
Regional risk
5
8
11
14
17
20
23
26
29
32
35
38
41
44
47
50
Epiweek

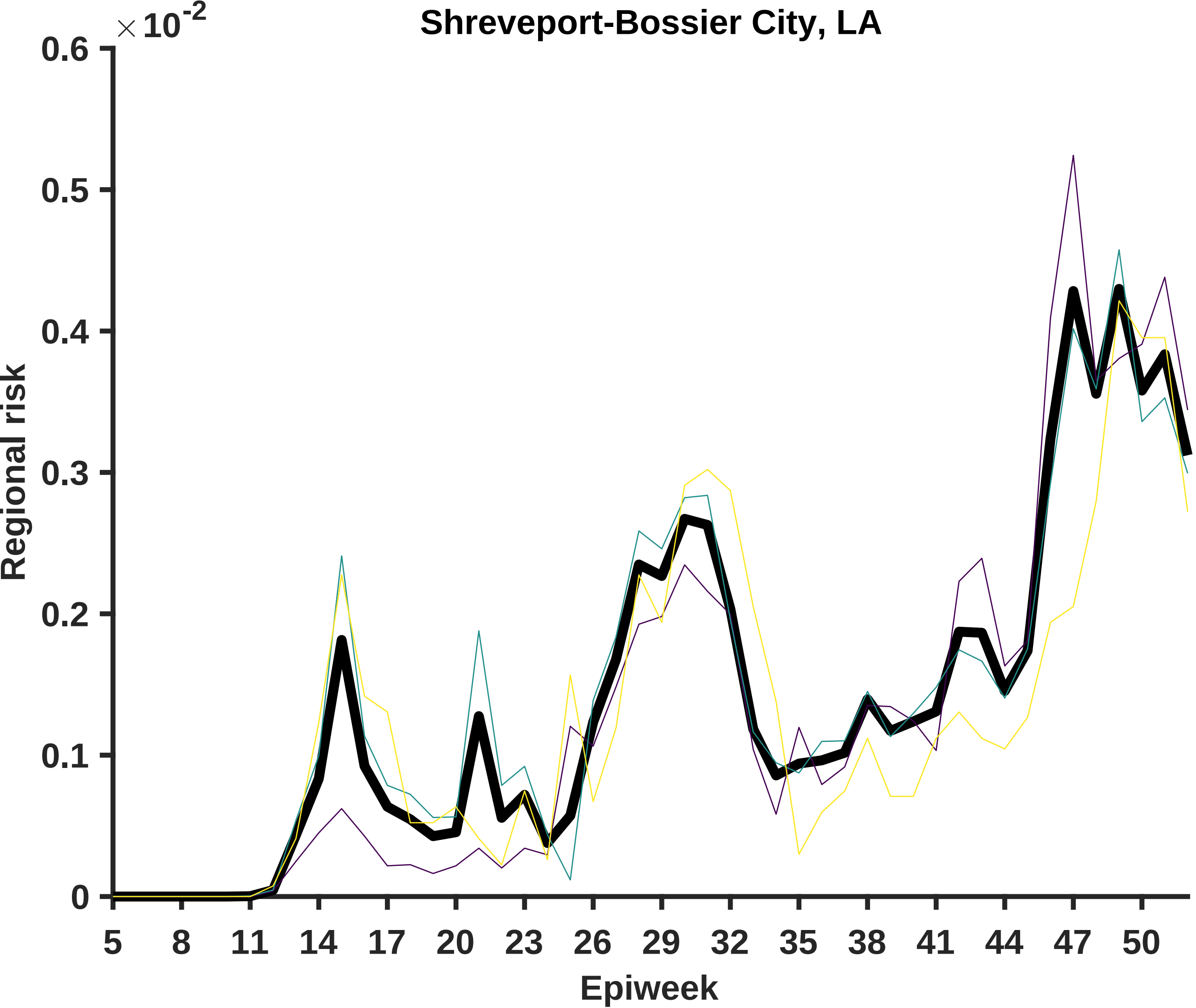
Shreveport-Bossier City, LA
$\times 10^{-2}$
0.6
0.5
0.4
0.3
0.2
0.1
0
Regional risk
5
8
11
14
17
20
23
26
29
32
35
38
41
44
47
50
Epiweek

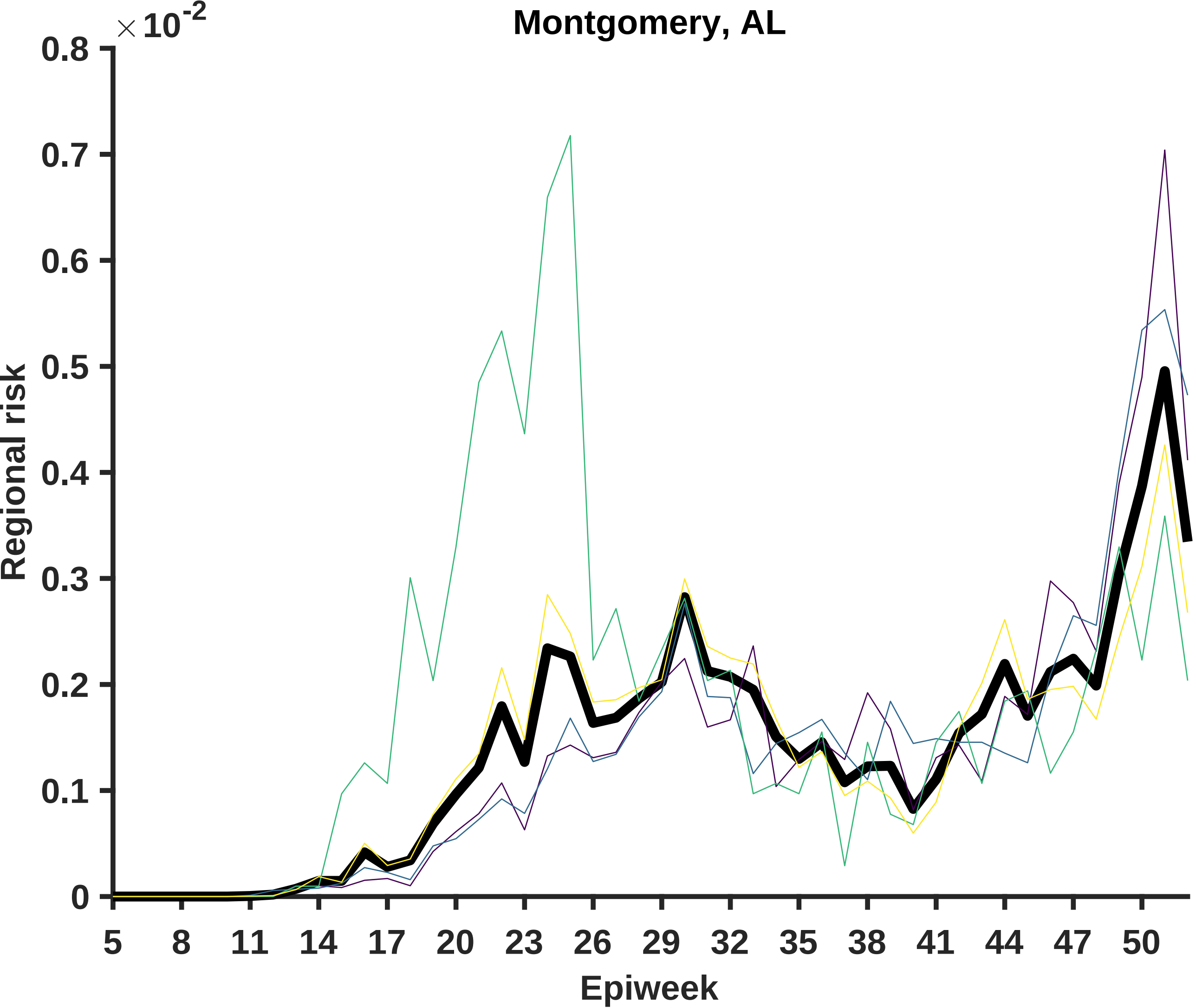

×10^-2
Montgomery, AL
Regional risk
0
0.1
0.2
0.3
0.4
0.5
0.6
0.7
0.8
5
8
11
14
17
20
23
26
29
32
35
38
41
44
47
50
Epiweek

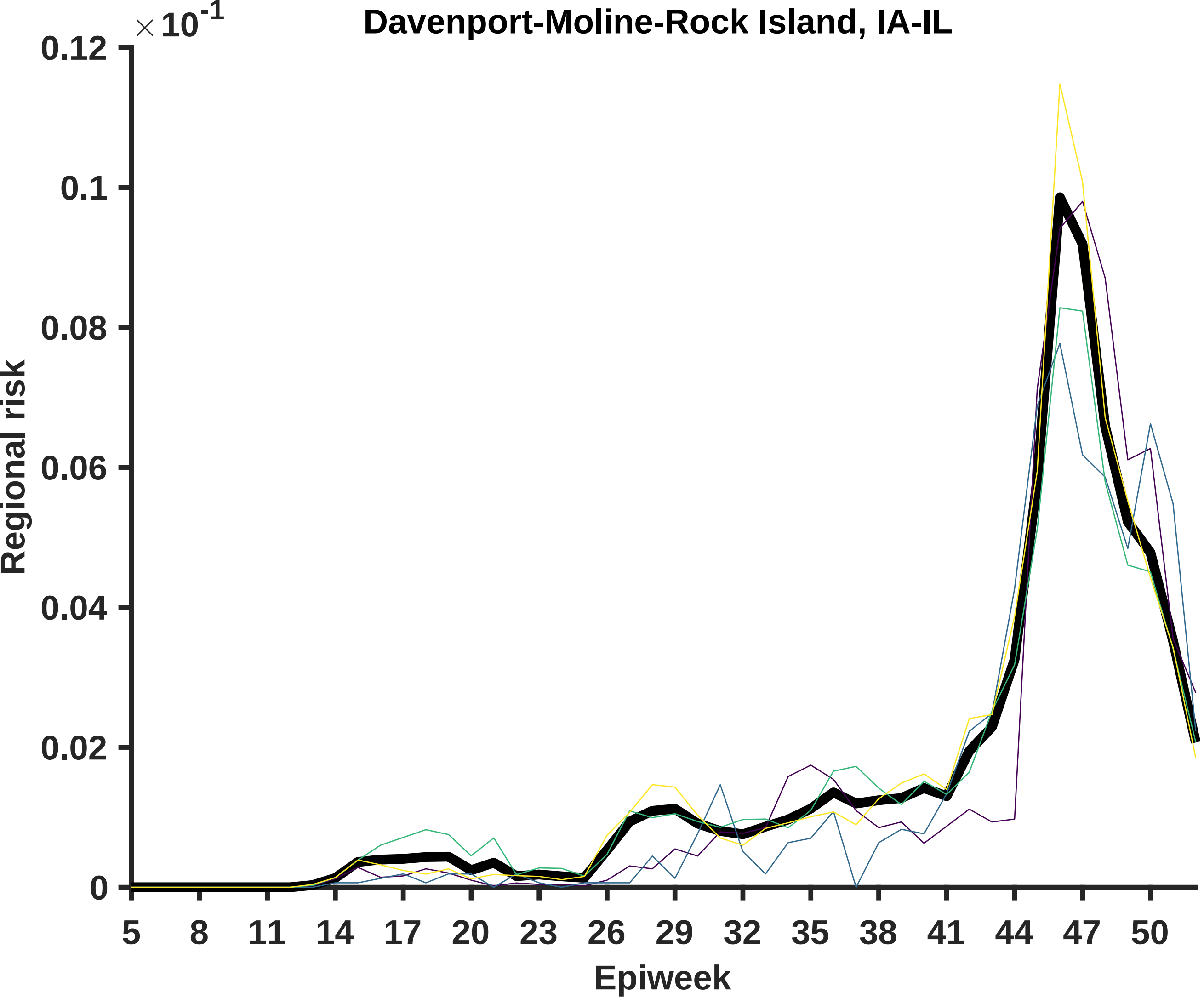

Davenport-Moline-Rock Island, IA-IL
×10^-1
0.12
0.1
0.08
0.06
0.04
0.02
0
Regional risk
5
8
11
14
17
20
23
26
29
32
35
38
41
44
47
50
Epiweek

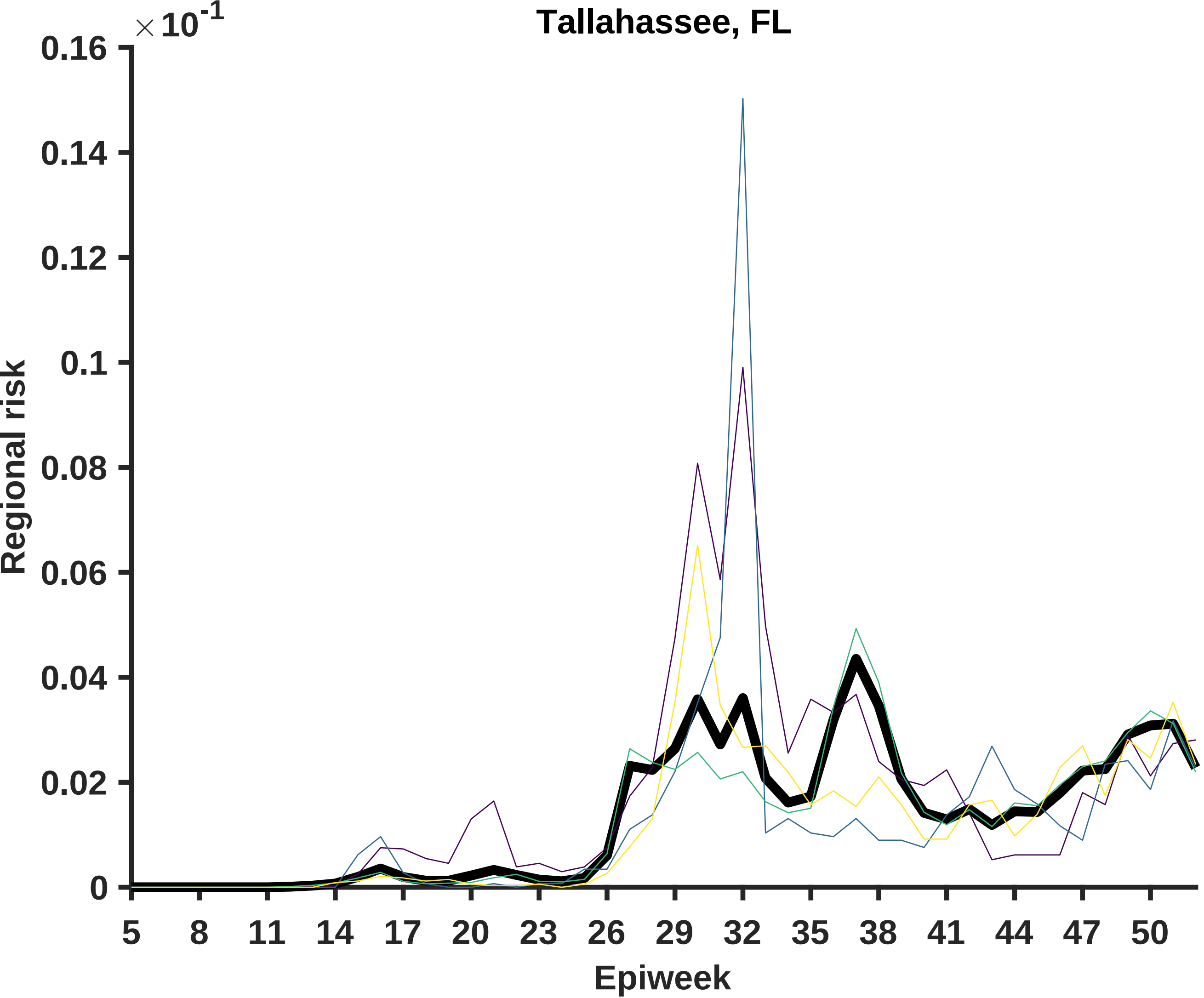

Tallahassee, FL
×10^-1
0.16
0.14
0.12
0.1
0.08
0.06
0.04
0.02
0
Regional risk
5
8
11
14
17
20
23
26
29
32
35
38
41
44
47
50
Epiweek

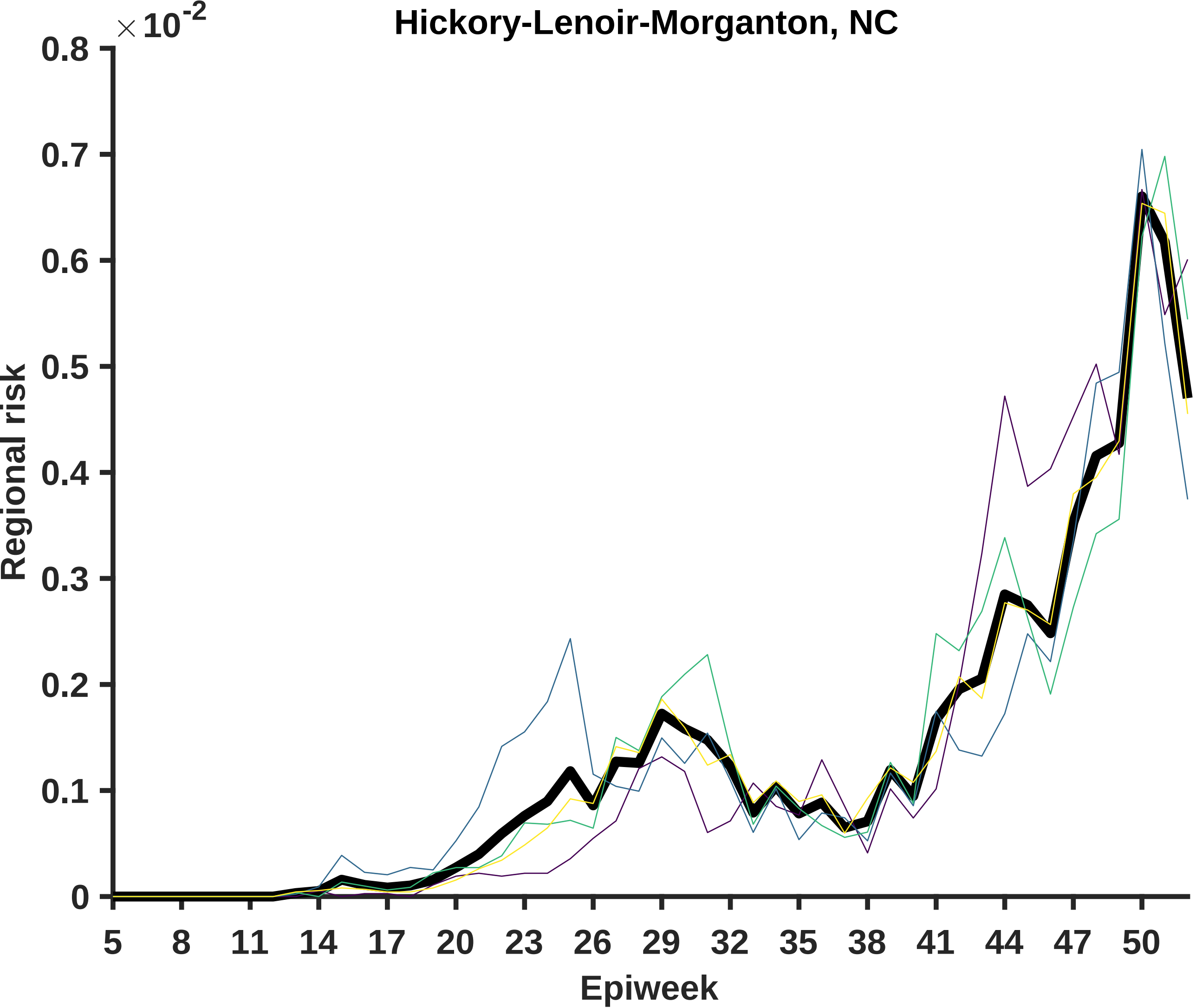

Hickory-Lenoir-Morganton, NC
× 10-2
0.8
0.7
0.6
0.5
0.4
0.3
0.2
0.1
0
Regional risk
5
8
11
14
17
20
23
26
29
32
35
38
41
44
47
50
Epiweek

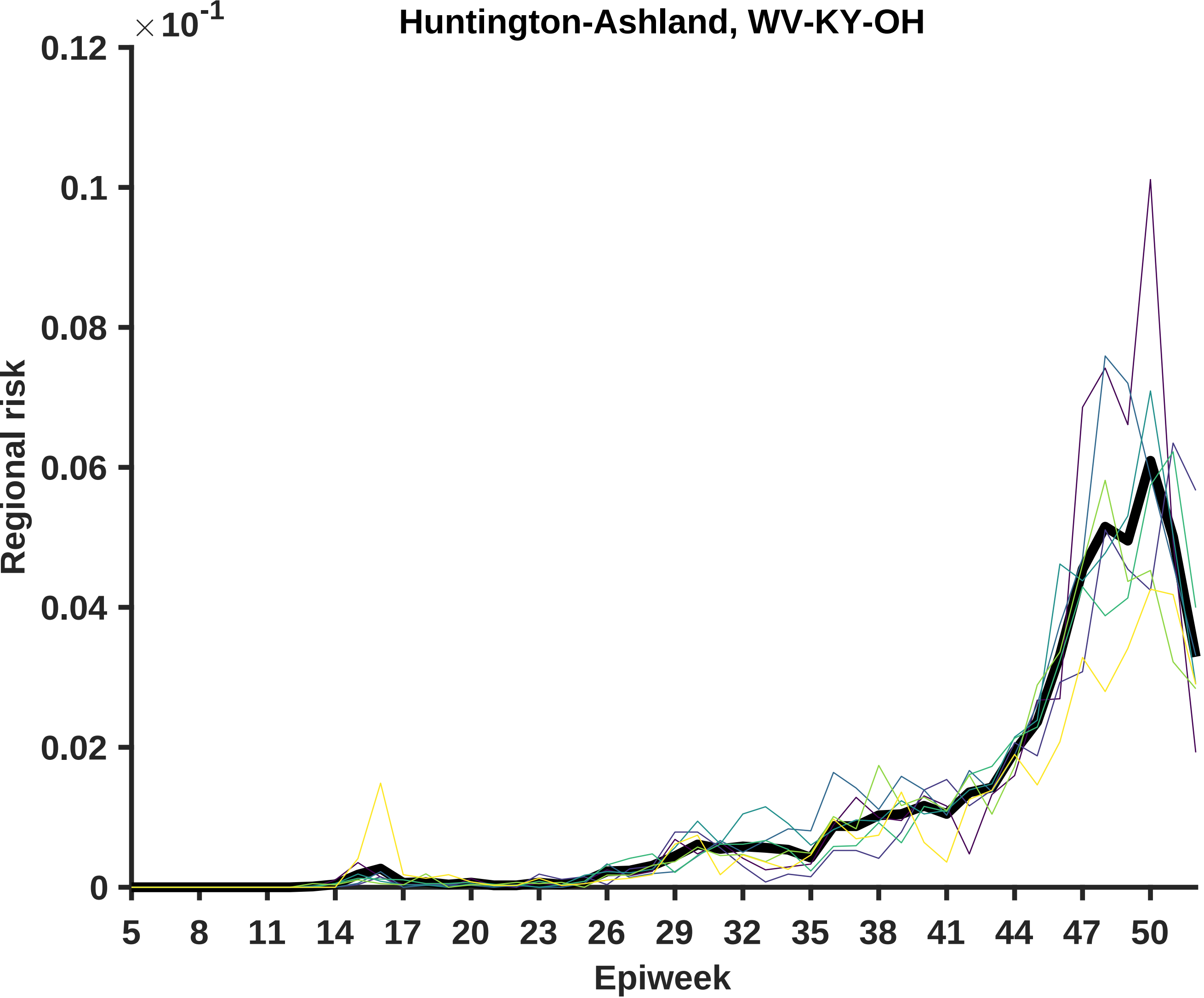

Huntington-Ashland, WV-KY-OH
× 10-1
0.12
0.1
0.08
0.06
0.04
0.02
0
Regional risk
5
8
11
14
17
20
23
26
29
32
35
38
41
44
47
50
Epiweek

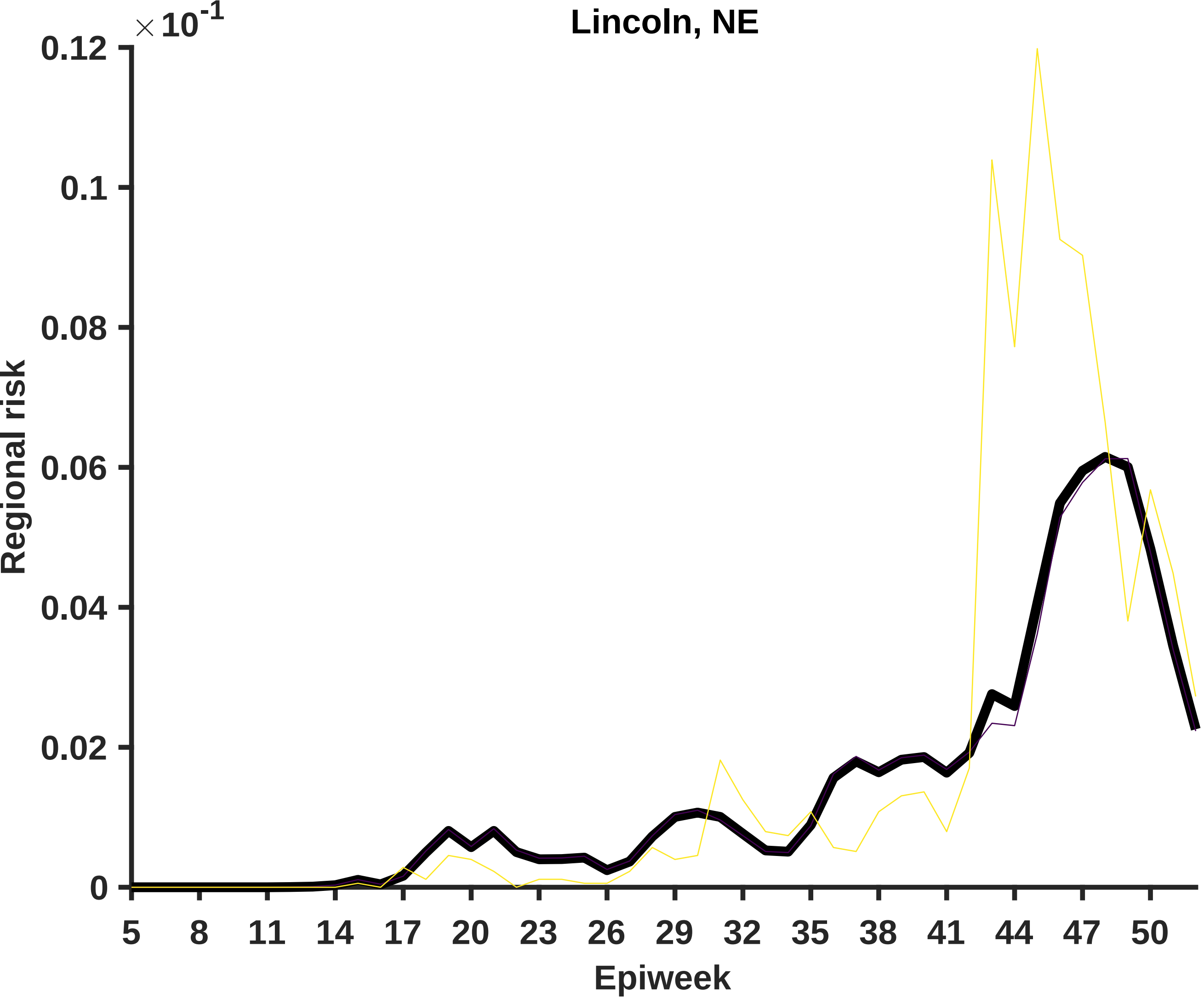
Lincoln, NE
×10^-1
0.12
0.1
0.08
0.06
0.04
0.02
0
Regional risk
5
8
11
14
17
20
23
26
29
32
35
38
41
44
47
50
Epiweek

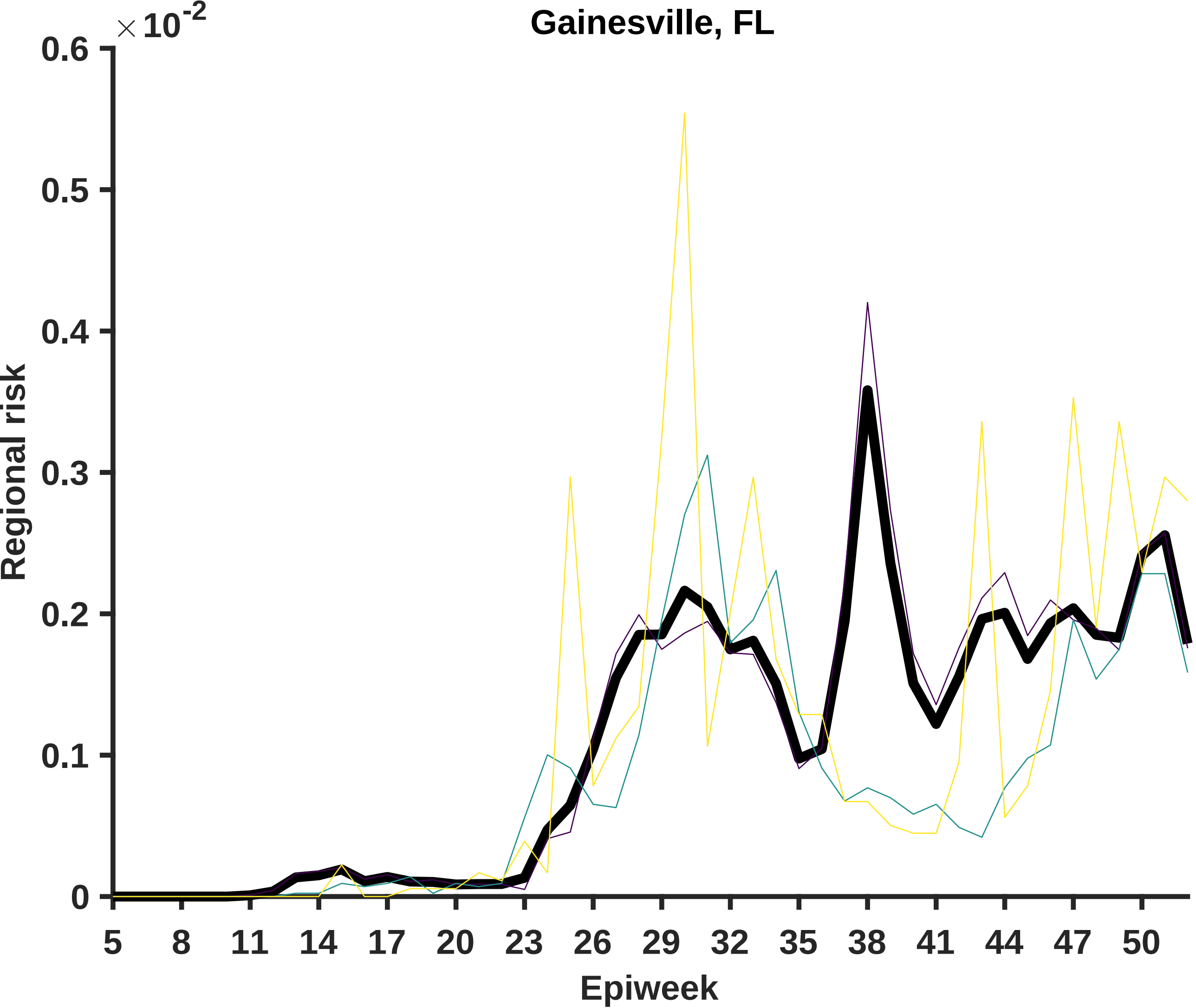

Gainesville, FL
× 10^-2
0.6
0.5
0.4
0.3
0.2
0.1
0
Regional risk
5
8
11
14
17
20
23
26
29
32
35
38
41
44
47
50
Epiweek

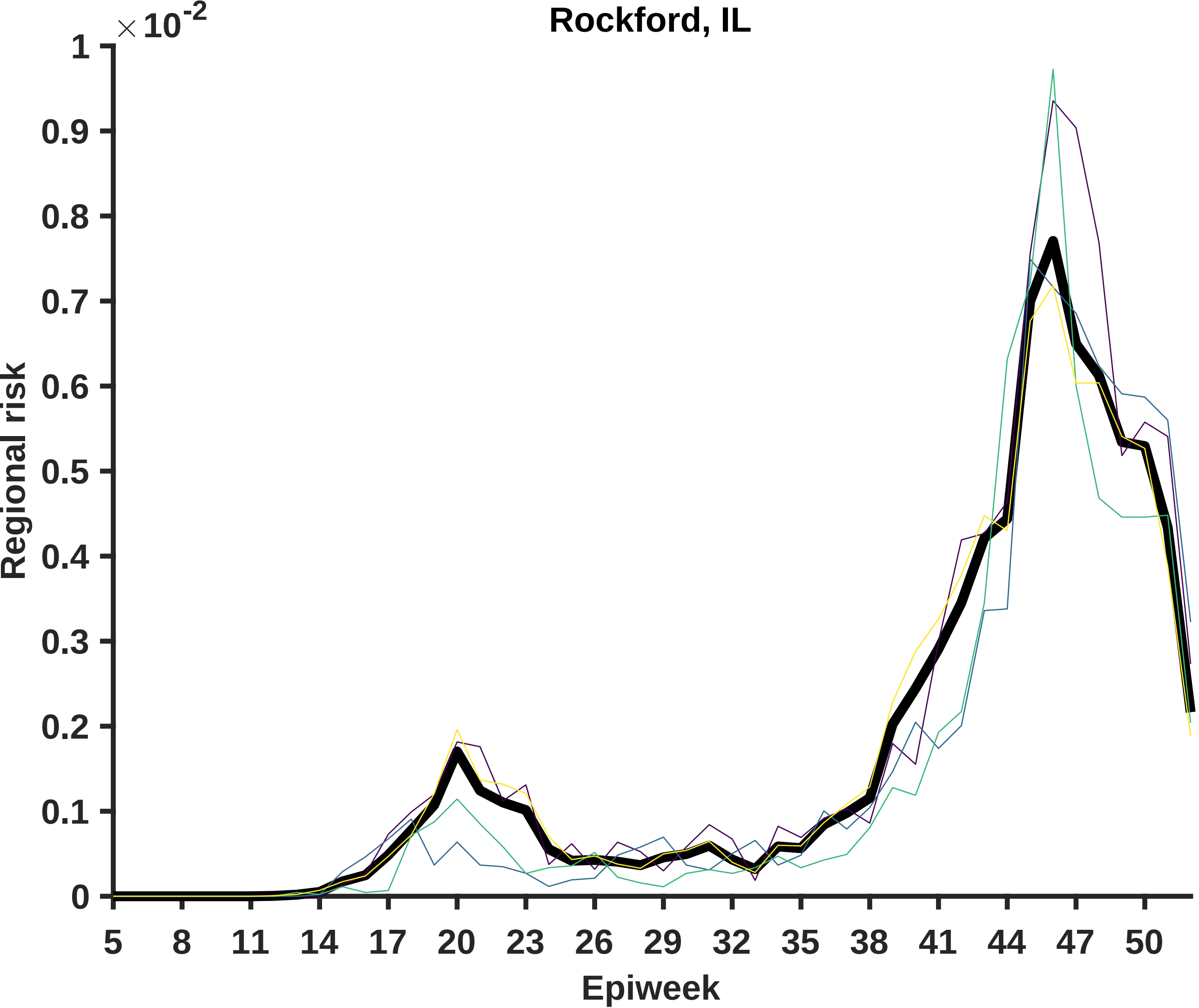

Rockford, IL
×10^-2
Regional risk
Epiweek
1
0.9
0.8
0.7
0.6
0.5
0.4
0.3
0.2
0.1
0
5
8
11
14
17
20
23
26
29
32
35
38
41
44
47
50

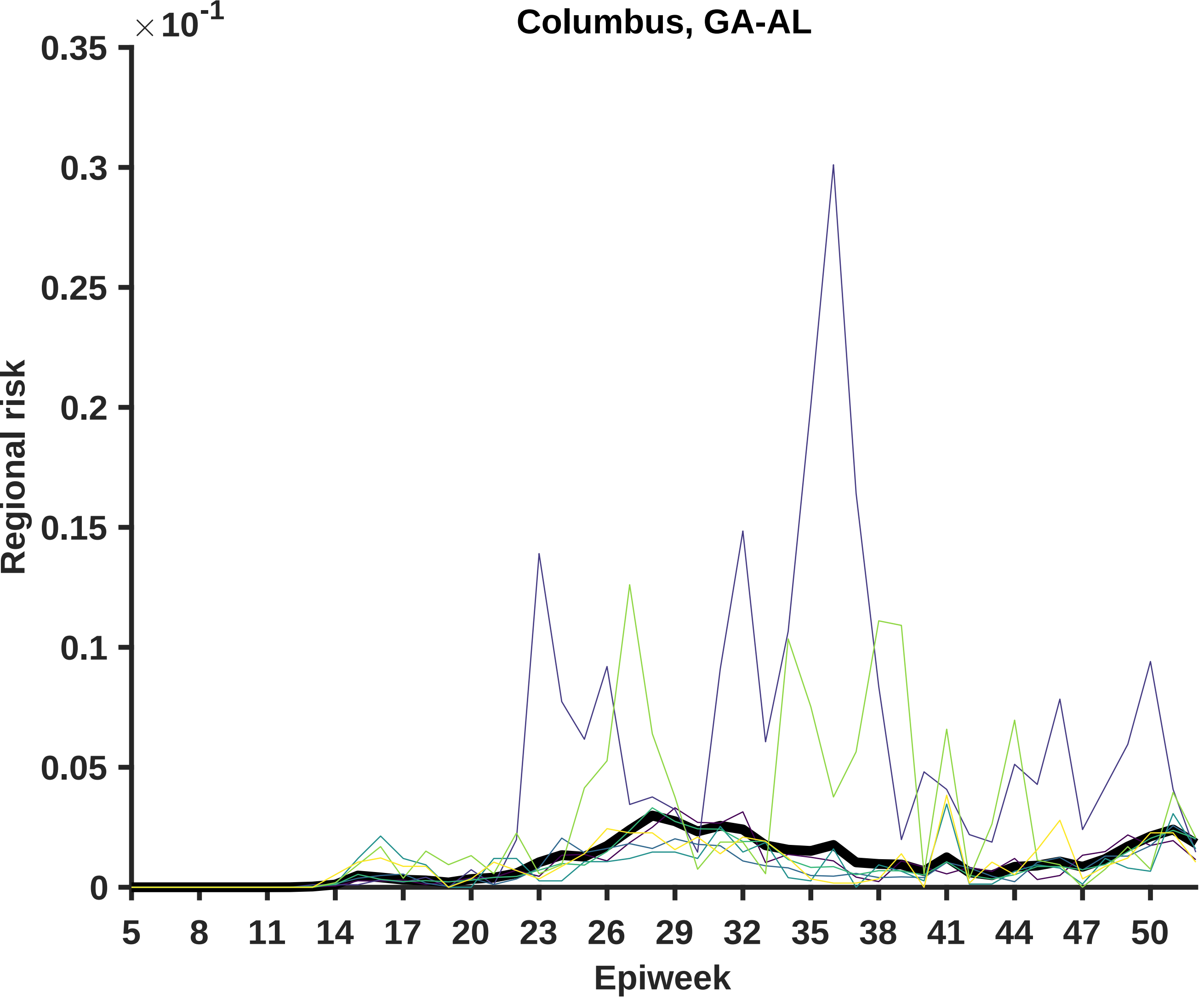

Columbus, GA-AL
×10^-1
0.35
0.3
0.25
0.2
0.15
0.1
0.05
0
Regional risk
5
8
11
14
17
20
23
26
29
32
35
38
41
44
47
50
Epiweek

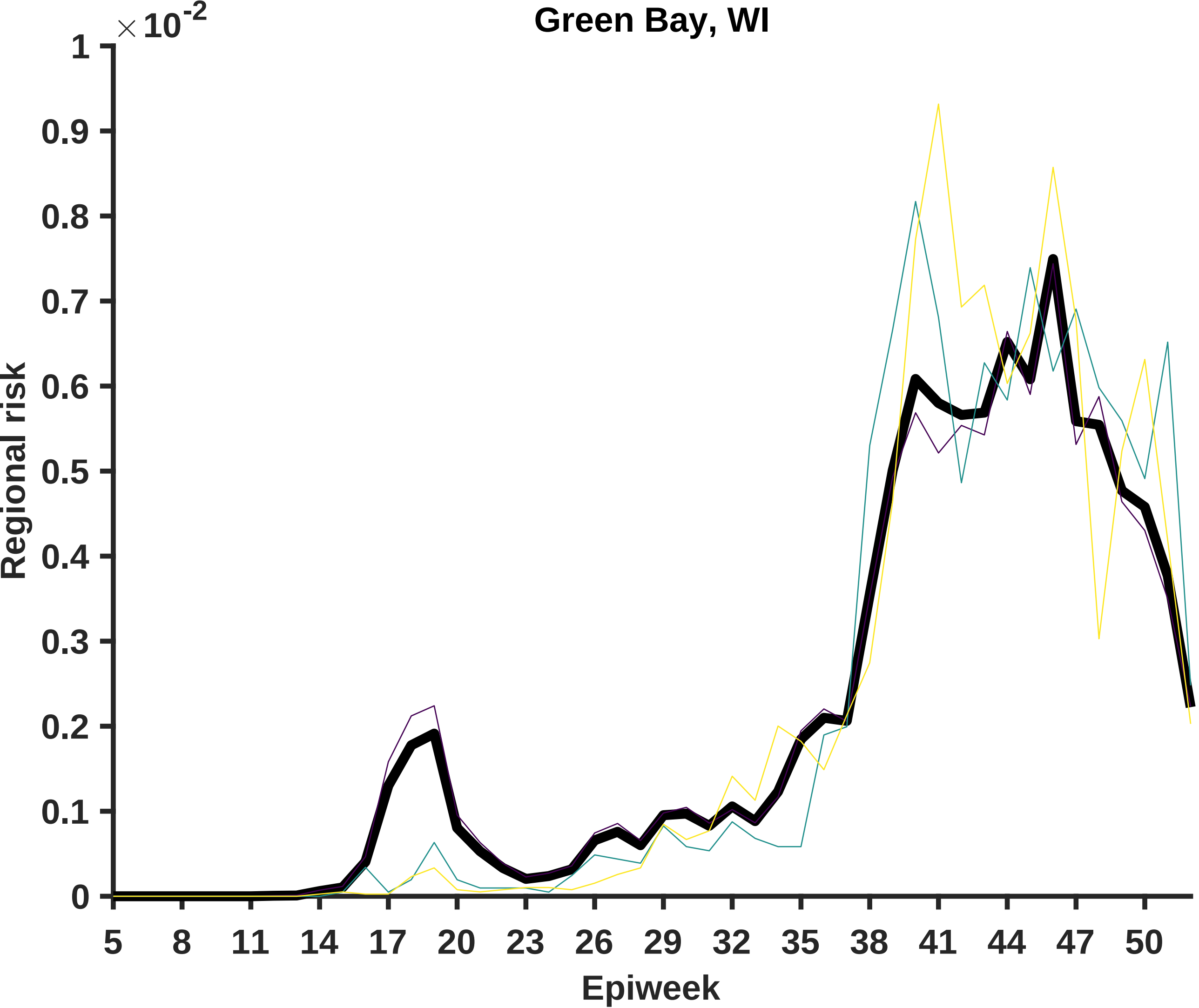

Green Bay, WI
×10^-2
Regional risk
Epiweek
0
0.1
0.2
0.3
0.4
0.5
0.6
0.7
0.8
0.9
1
5
8
11
14
17
20
23
26
29
32
35
38
41
44
47
50

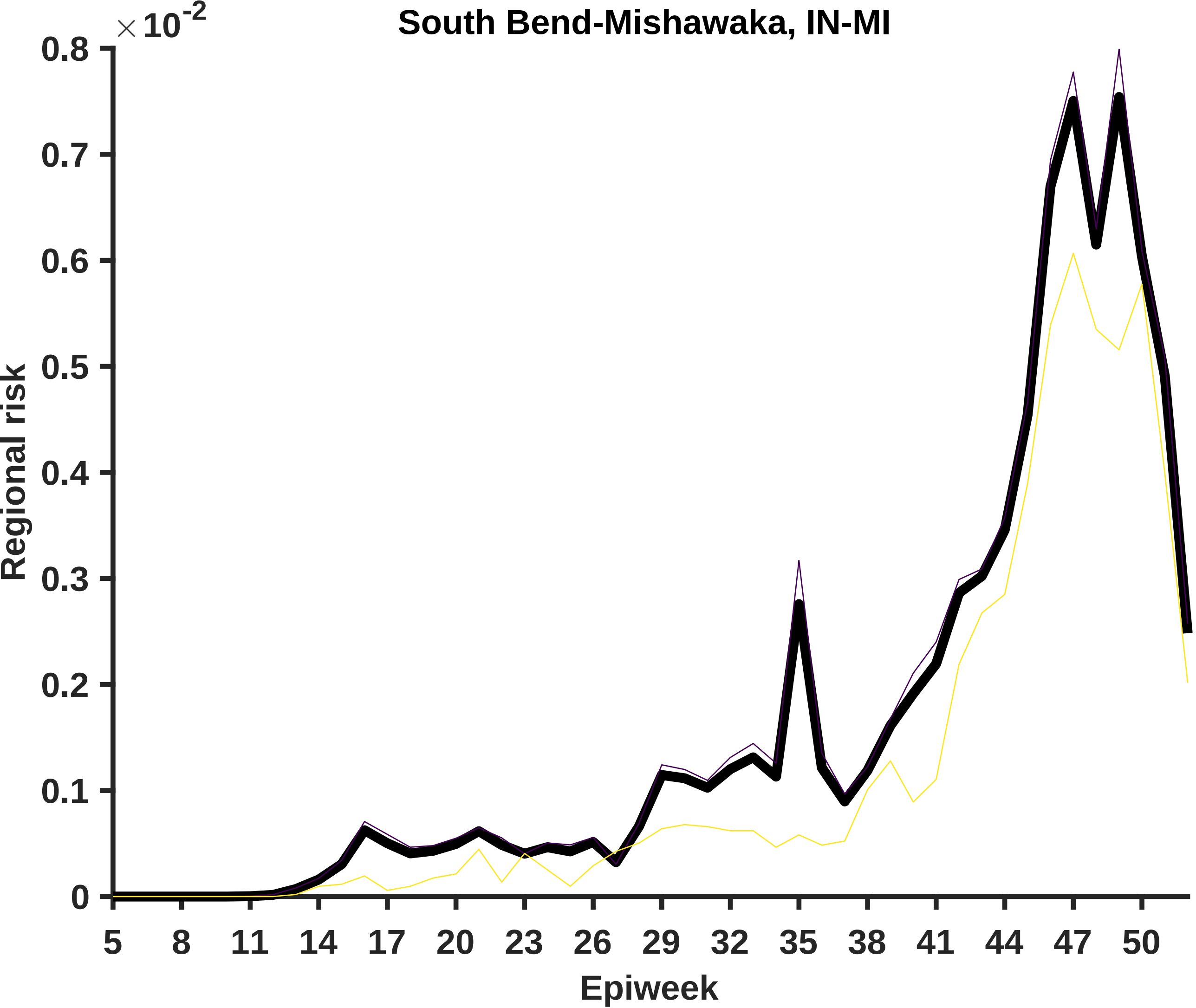
South Bend-Mishawaka, IN-MI
×10^-2
0.8
0.7
0.6
0.5
0.4
0.3
0.2
0.1
0
Regional risk
5
8
11
14
17
20
23
26
29
32
35
38
41
44
47
50
Epiweek

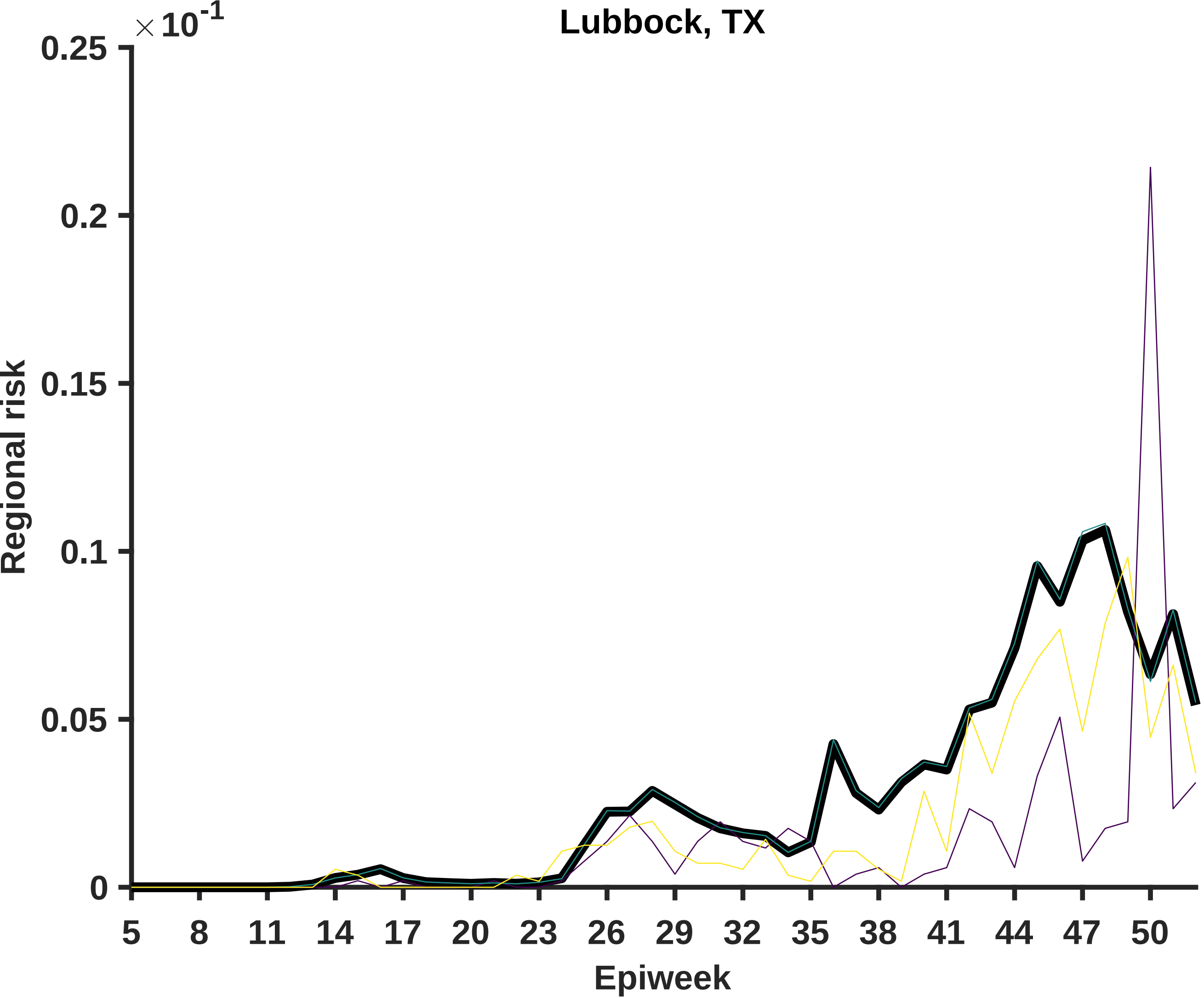

Lubbock, TX
×10^-1
0.25
0.2
0.15
0.1
0.05
0
Regional risk
5
8
11
14
17
20
23
26
29
32
35
38
41
44
47
50
Epiweek

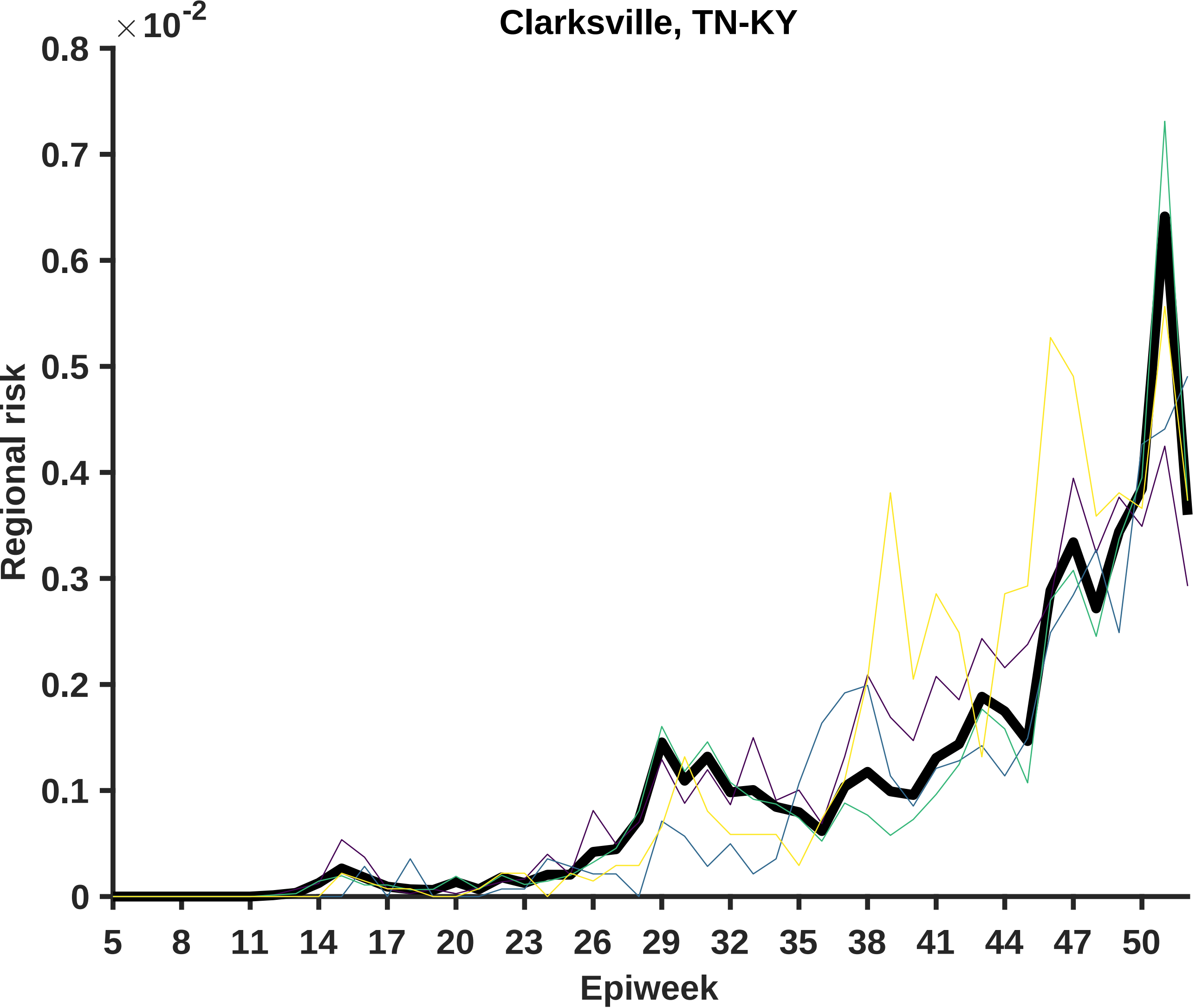

Clarksville, TN-KY
× 10^-2
0.8
0.7
0.6
0.5
0.4
0.3
0.2
0.1
0
Regional risk
5
8
11
14
17
20
23
26
29
32
35
38
41
44
47
50
Epiweek

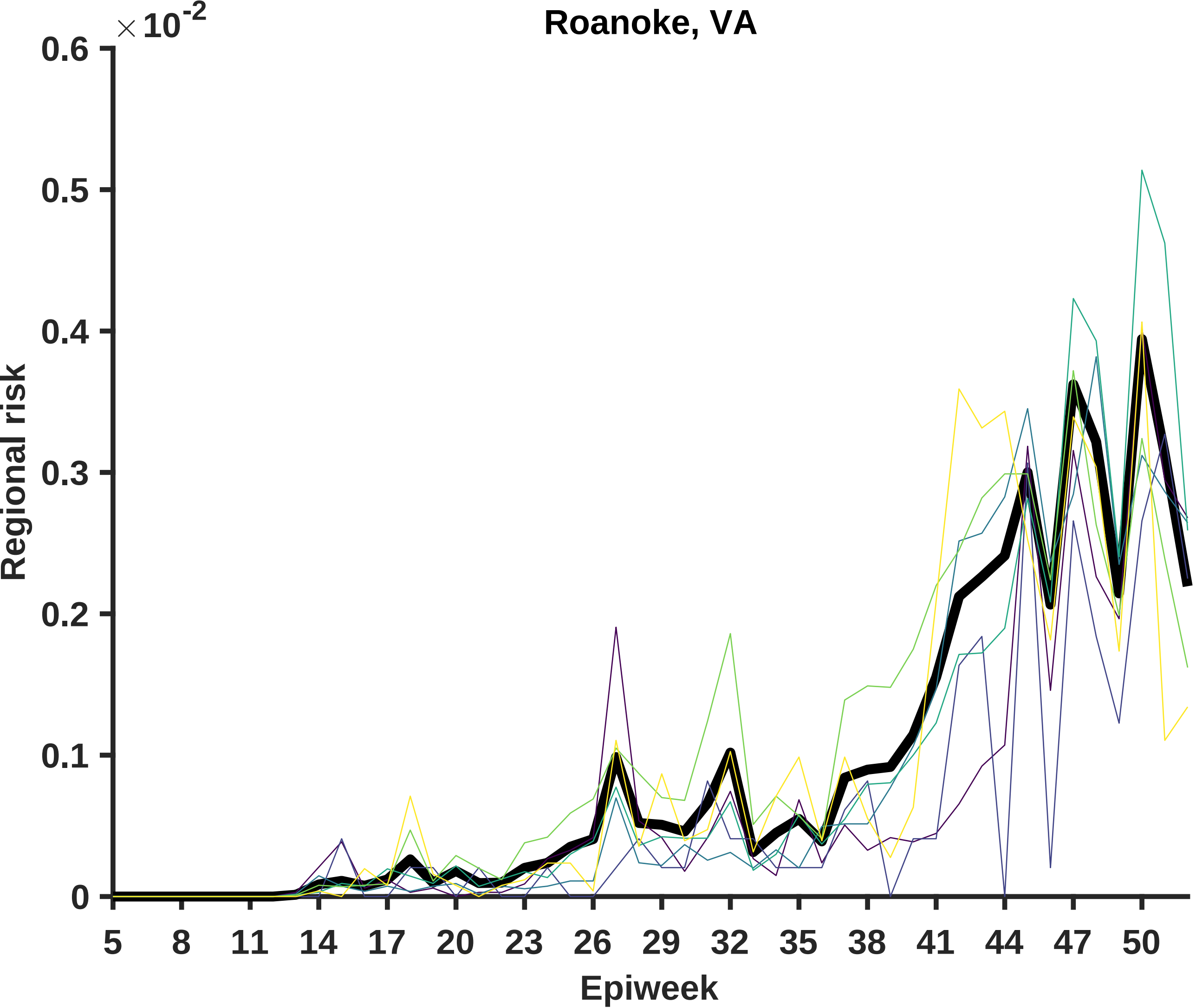
Roanoke, VA
×10^-2
0.6
0.5
0.4
0.3
0.2
0.1
0
Regional risk
5
8
11
14
17
20
23
26
29
32
35
38
41
44
47
50
Epiweek

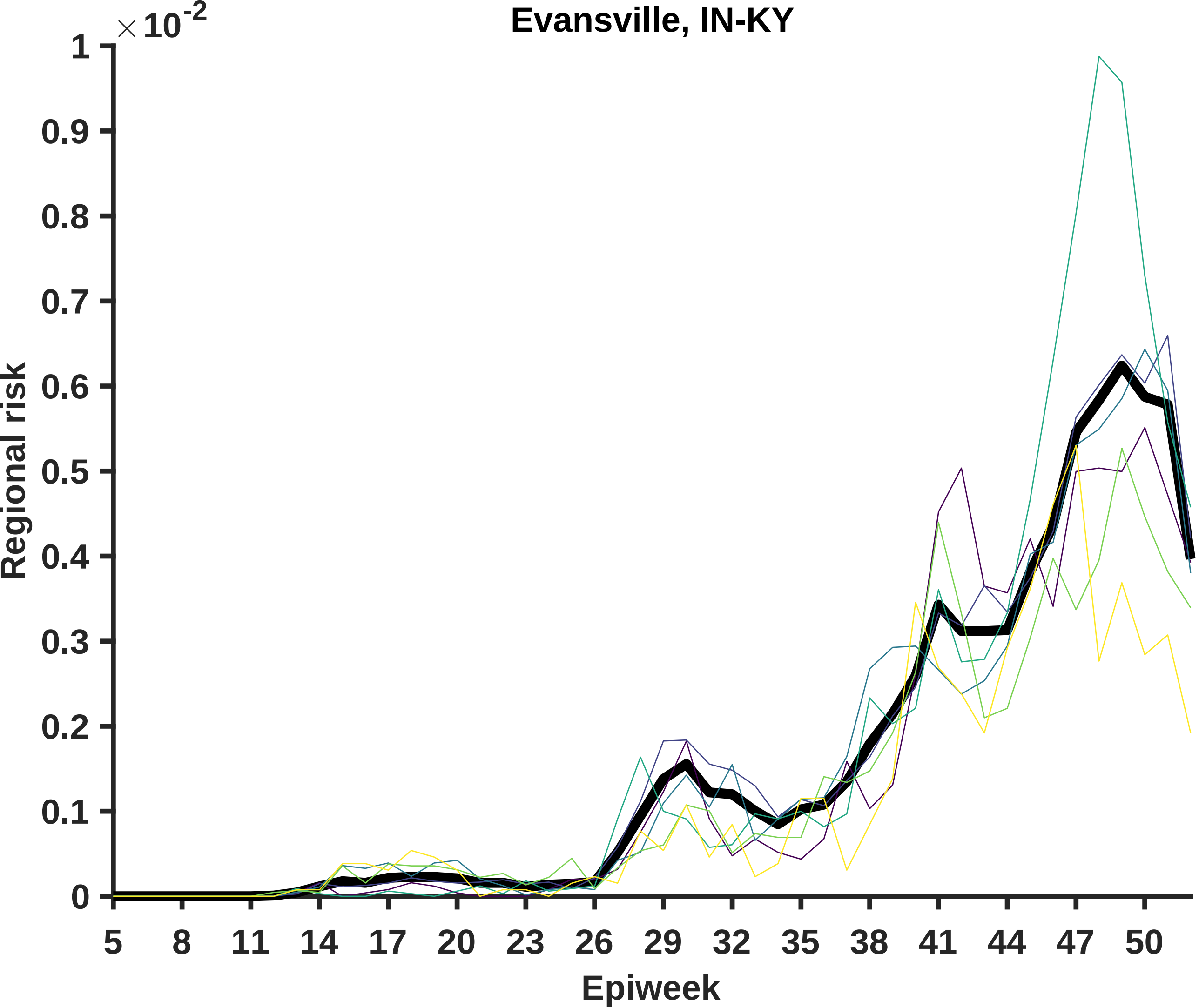

Evansville, IN-KY
×10^-2
Regional risk
0
0.1
0.2
0.3
0.4
0.5
0.6
0.7
0.8
0.9
1
Epiweek
5
8
11
14
17
20
23
26
29
32
35
38
41
44
47
50

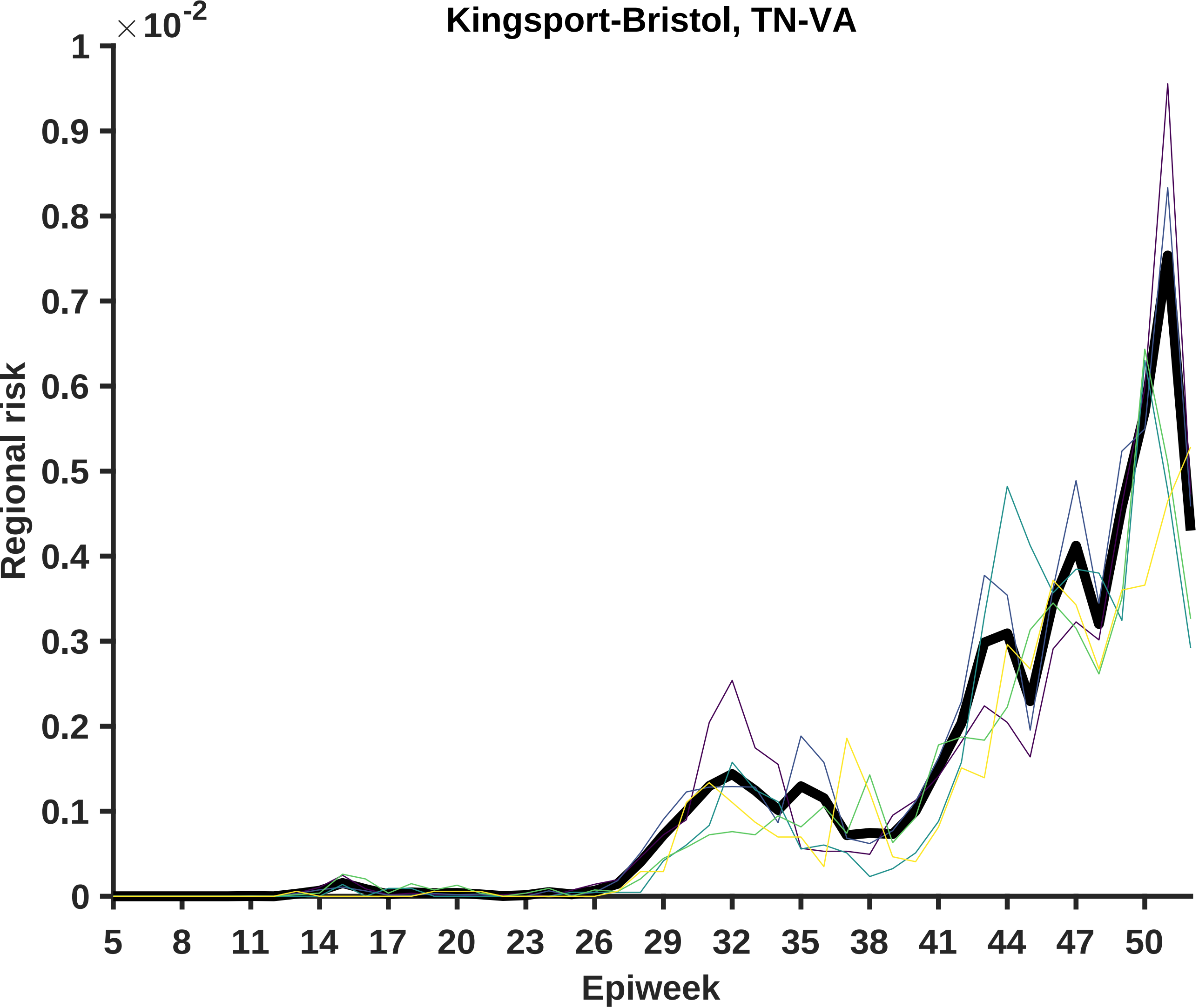

Kingsport-Bristol, TN-VA
×10^-2
1
0.9
0.8
0.7
0.6
0.5
0.4
0.3
0.2
0.1
0
Regional risk
5
8
11
14
17
20
23
26
29
32
35
38
41
44
47
50
Epiweek

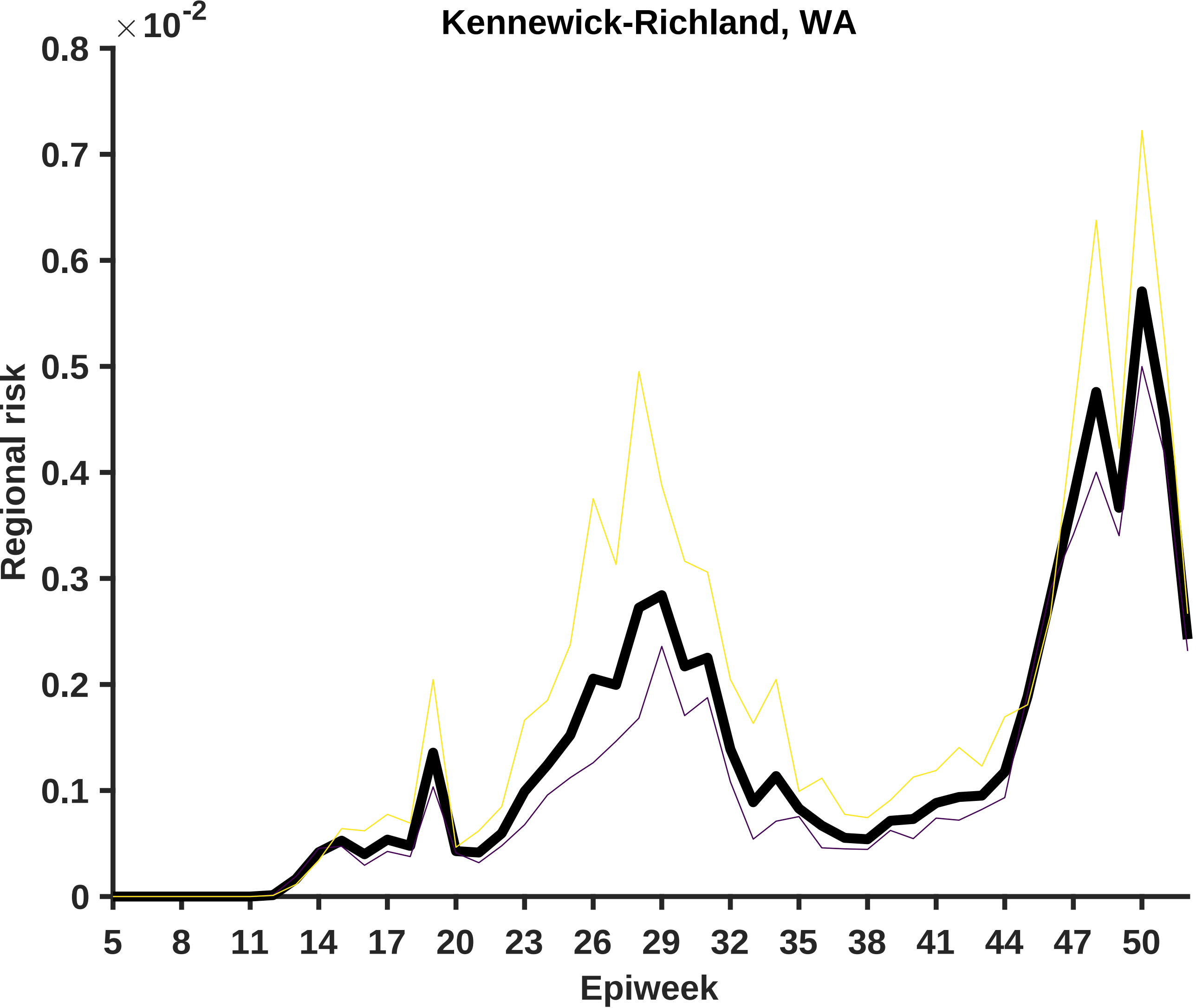

Kennewick-Richland, WA
×10^-2
0.8
0.7
0.6
0.5
0.4
0.3
0.2
0.1
0
Regional risk
5
8
11
14
17
20
23
26
29
32
35
38
41
44
47
50
Epiweek

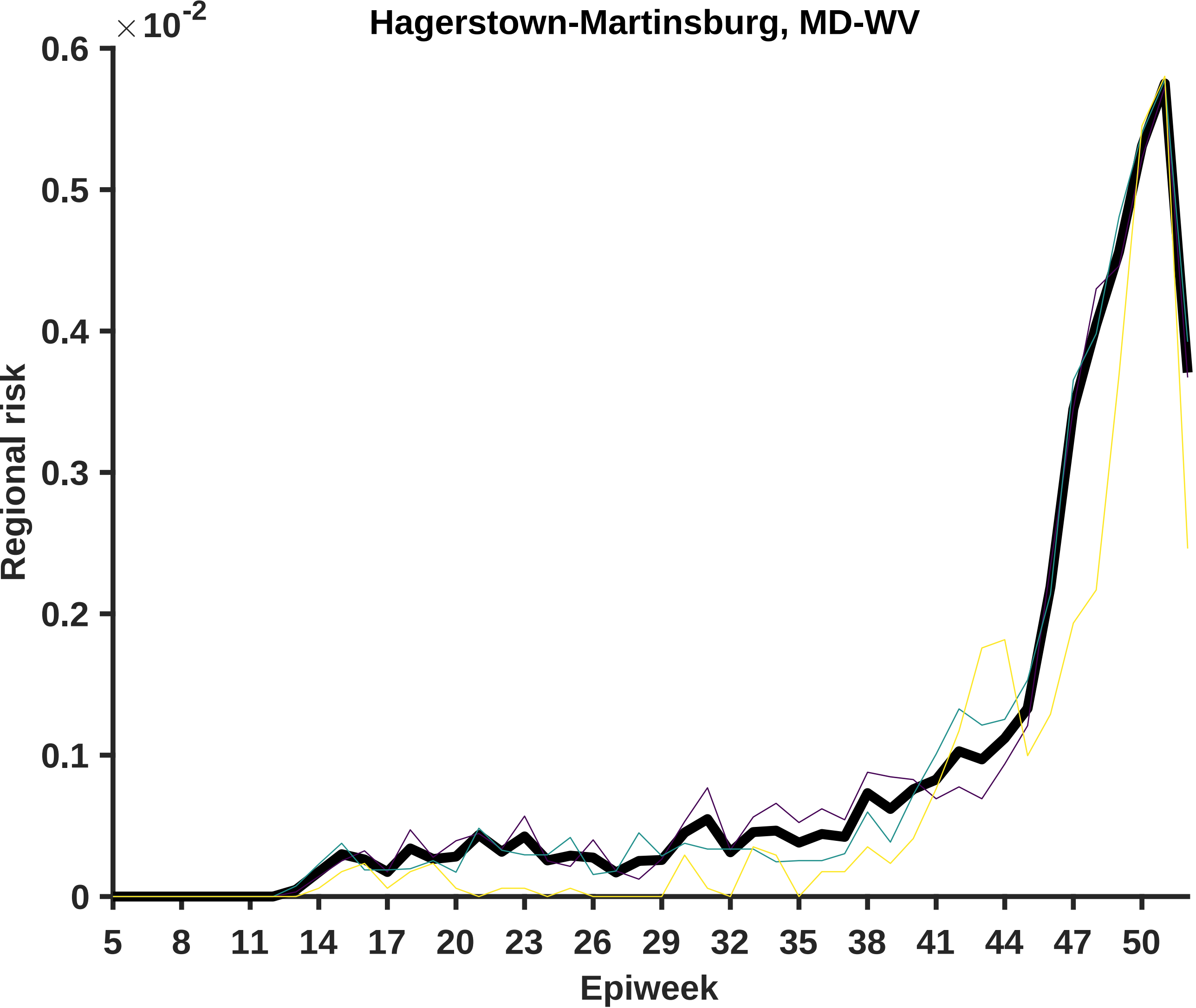

Hagerstown-Martinsburg, MD-WV
× 10^-2
0.6
0.5
0.4
0.3
0.2
0.1
0
Regional risk
5
8
11
14
17
20
23
26
29
32
35
38
41
44
47
50
Epiweek

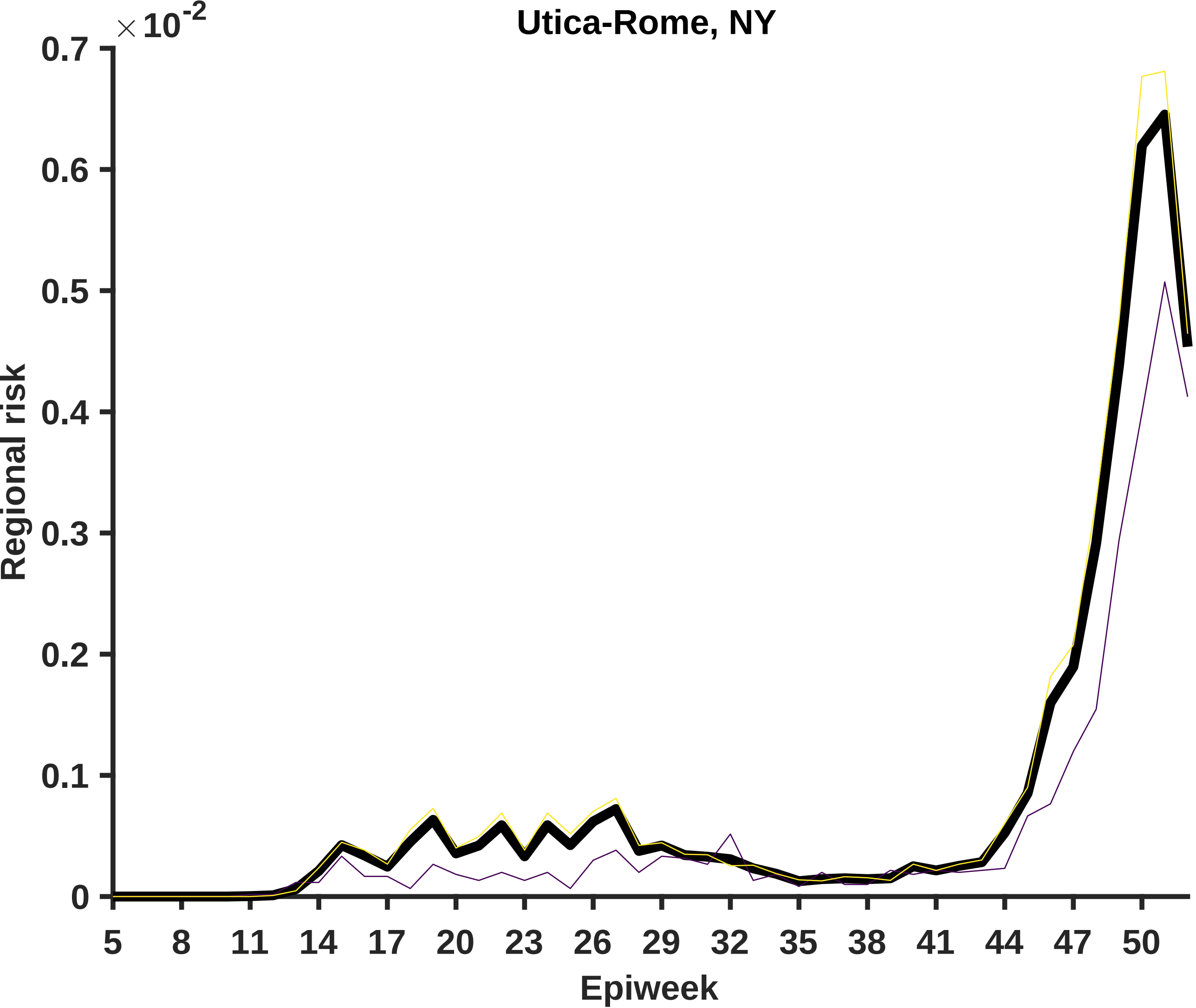

Utica-Rome, NY
×10^-2
0.7
0.6
0.5
0.4
0.3
0.2
0.1
0
Regional risk
5
8
11
14
17
20
23
26
29
32
35
38
41
44
47
50
Epiweek

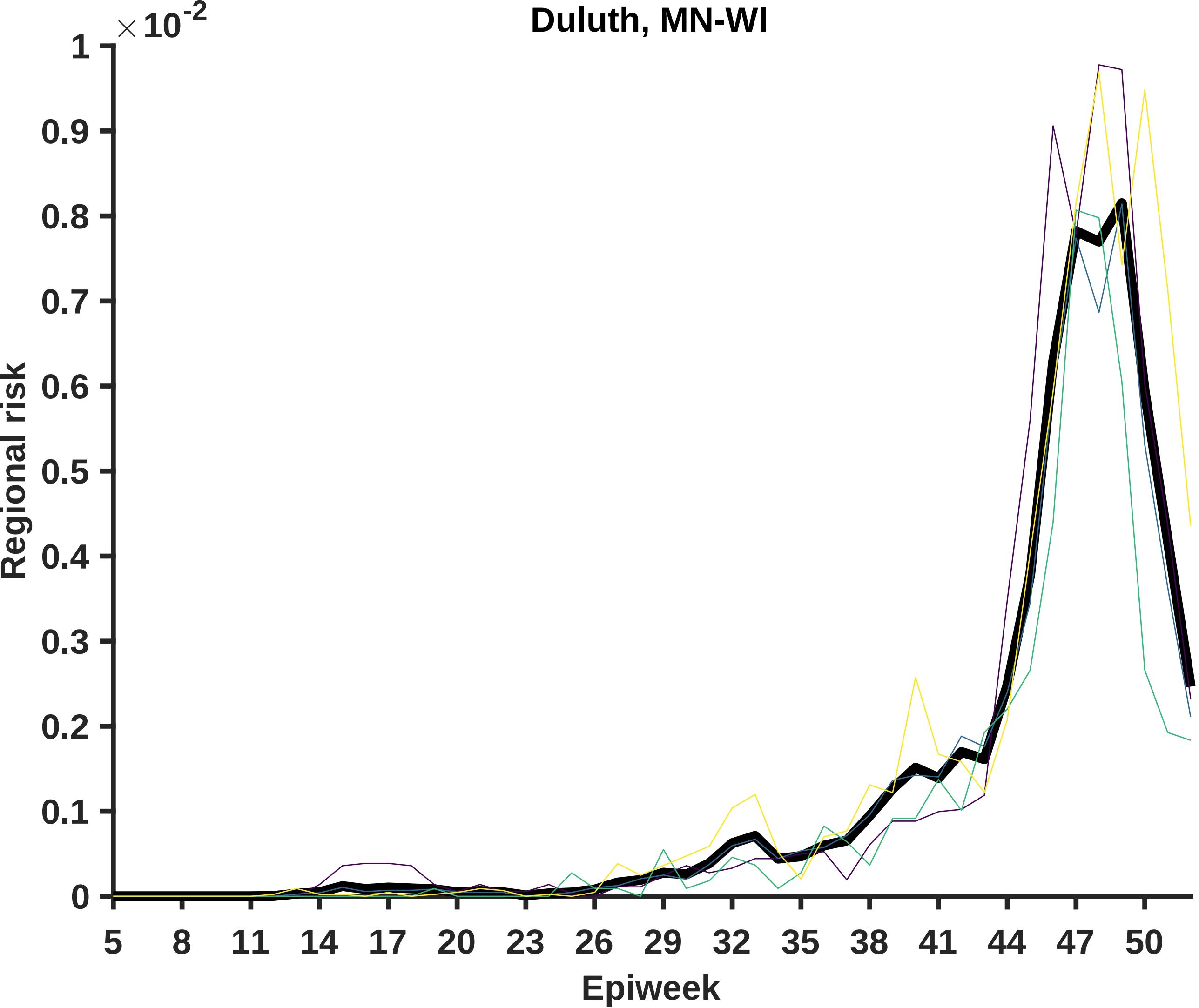
Duluth, MN-WI
×10^-2
1
0.9
0.8
0.7
0.6
0.5
0.4
0.3
0.2
0.1
0
Regional risk
5
8
11
14
17
20
23
26
29
32
35
38
41
44
47
50
Epiweek

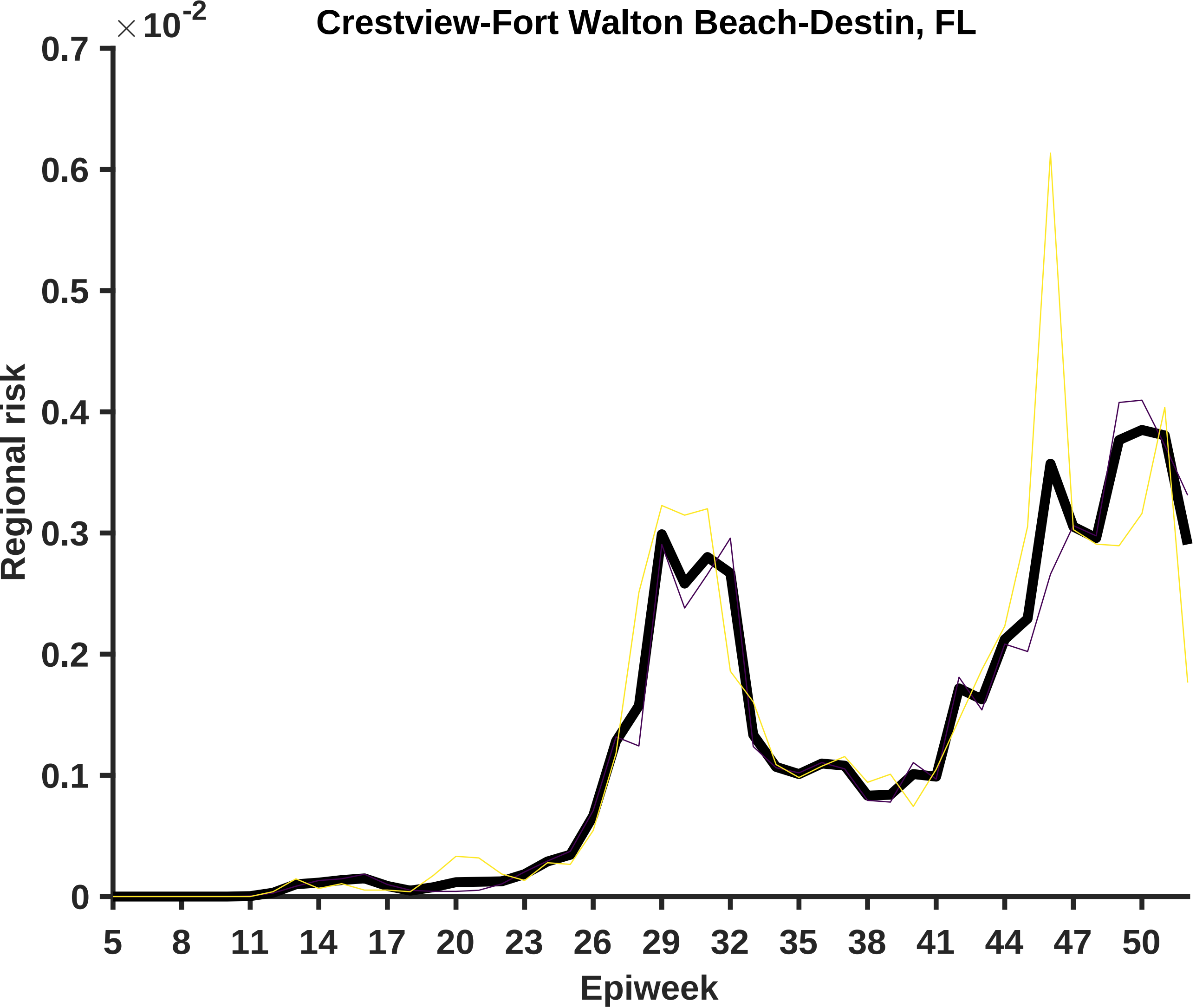

Crestview-Fort Walton Beach-Destin, FL
×10^-2
0.7
0.6
0.5
0.4
0.3
0.2
0.1
0
Regional risk
5
8
11
14
17
20
23
26
29
32
35
38
41
44
47
50
Epiweek

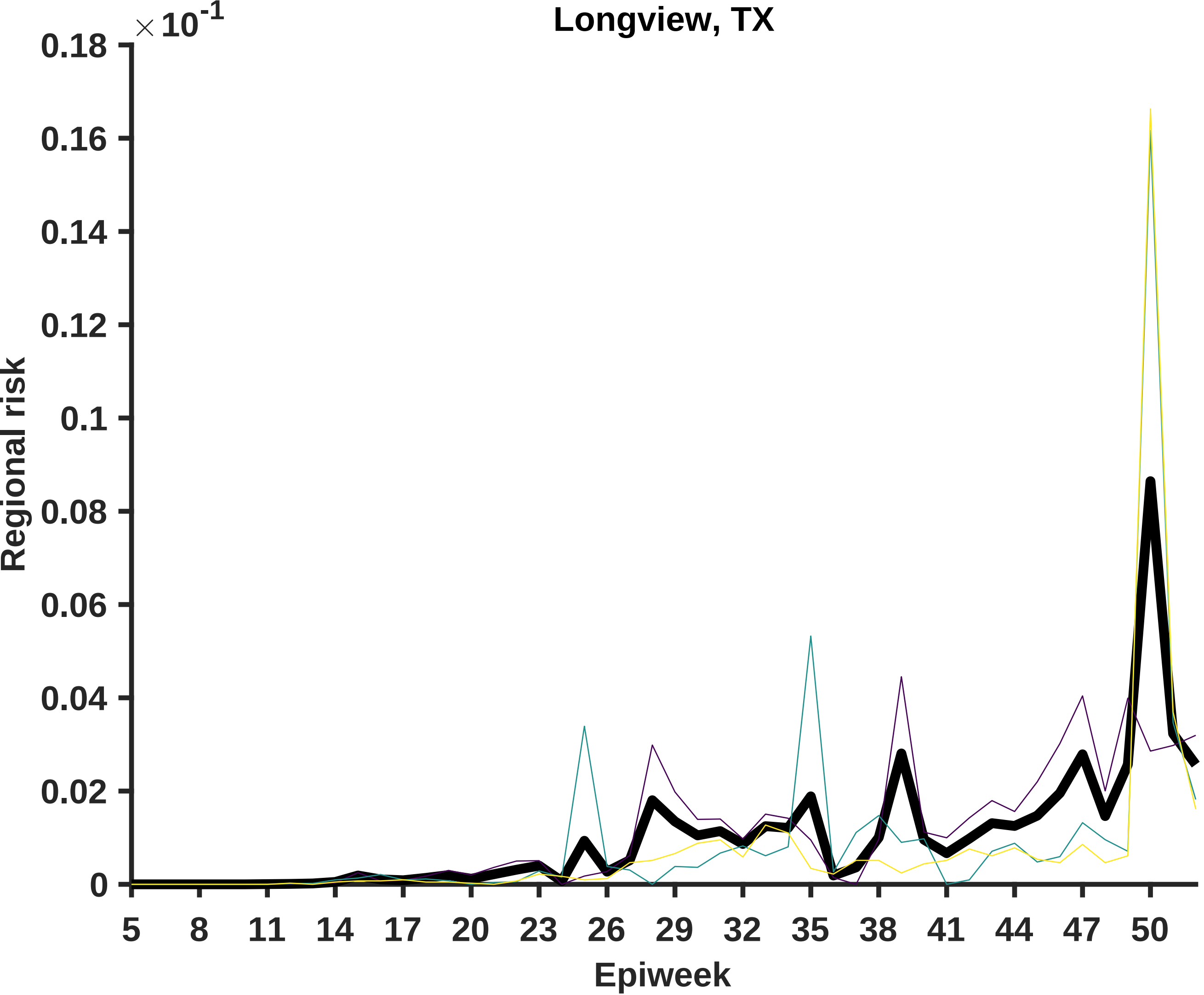

Longview, TX
× 10^-1
0.18
0.16
0.14
0.12
0.1
0.08
0.06
0.04
0.02
0
Regional risk
5
8
11
14
17
20
23
26
29
32
35
38
41
44
47
50
Epiweek

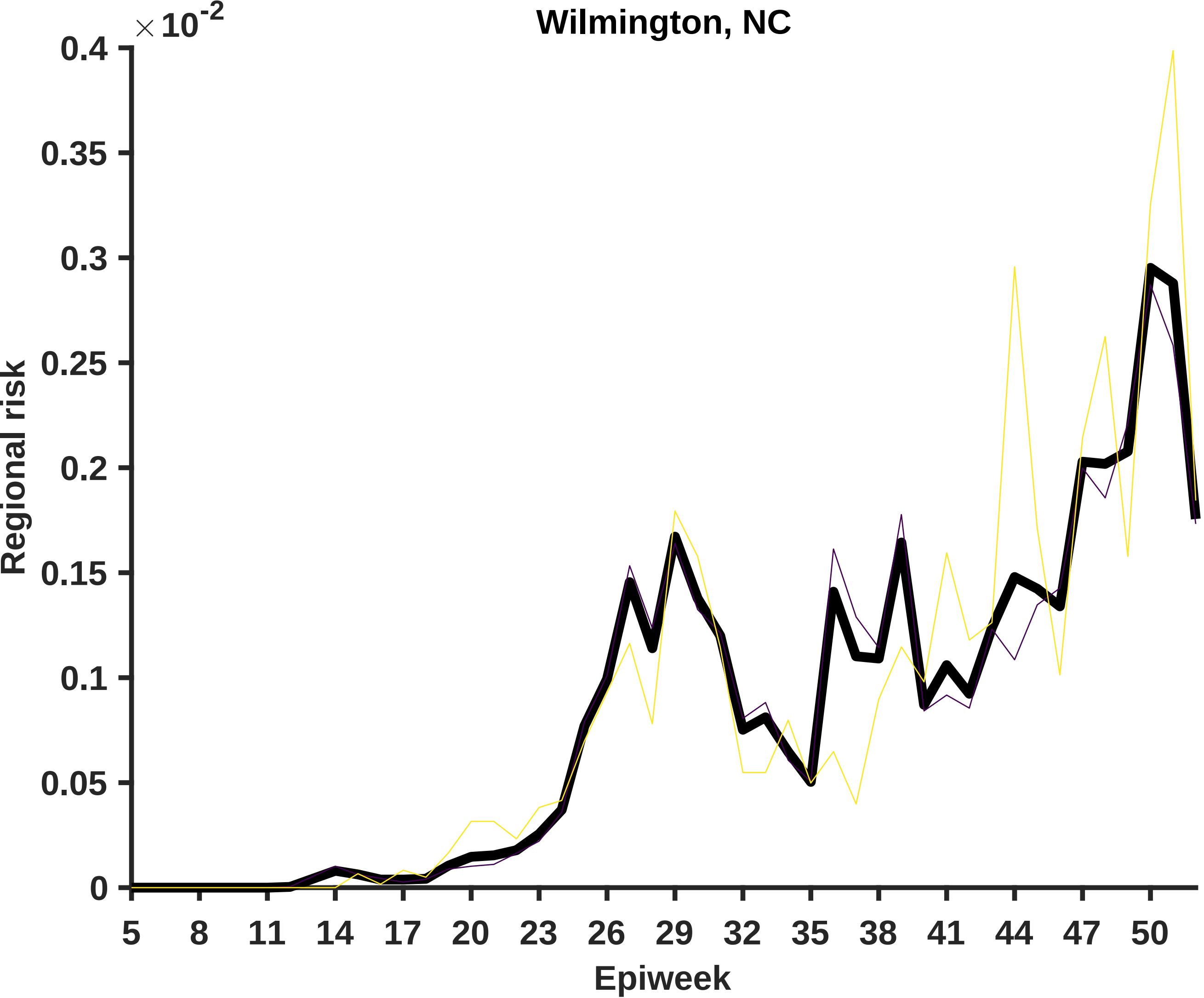

Wilmington, NC
× 10^-2
0.4
0.35
0.3
0.25
0.2
0.15
0.1
0.05
0
Regional risk
5
8
11
14
17
20
23
26
29
32
35
38
41
44
47
50
Epiweek

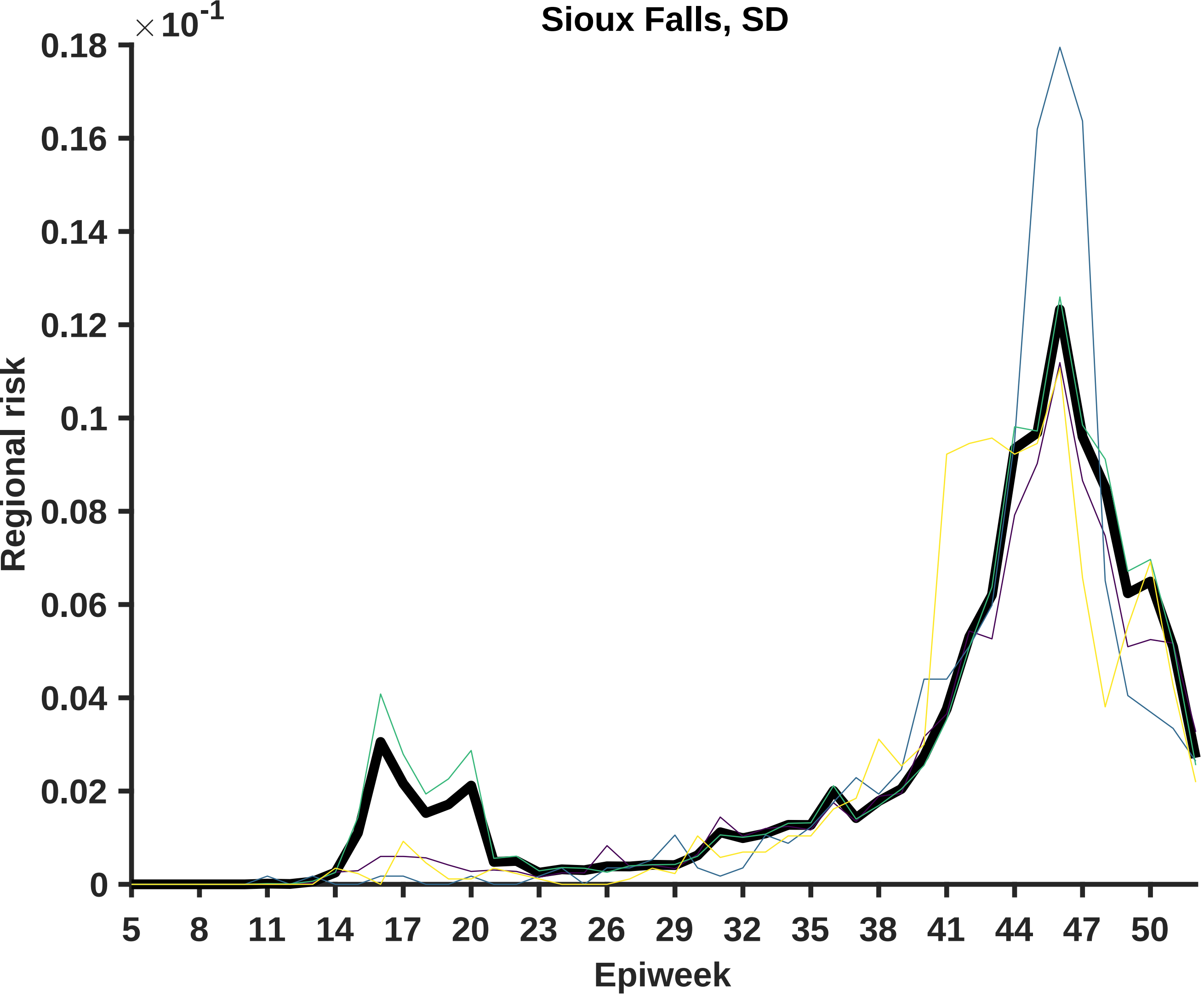

Sioux Falls, SD
× 10^-1
0.18
0.16
0.14
0.12
0.1
0.08
0.06
0.04
0.02
0
Regional risk
5
8
11
14
17
20
23
26
29
32
35
38
41
44
47
50
Epiweek

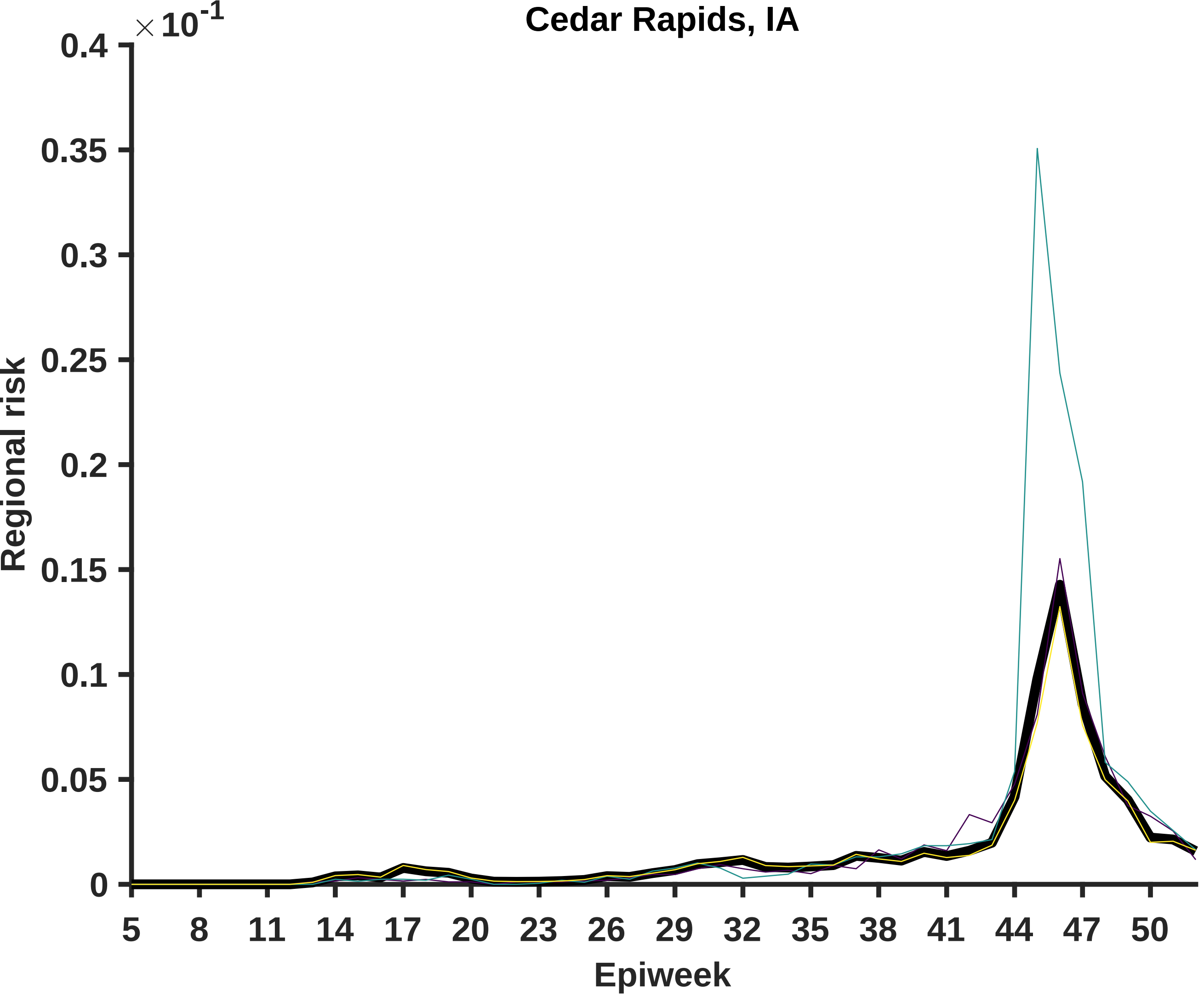

Cedar Rapids, IA
×10^-1
0.4
0.35
0.3
0.25
0.2
0.15
0.1
0.05
0
Regional risk
5
8
11
14
17
20
23
26
29
32
35
38
41
44
47
50
Epiweek

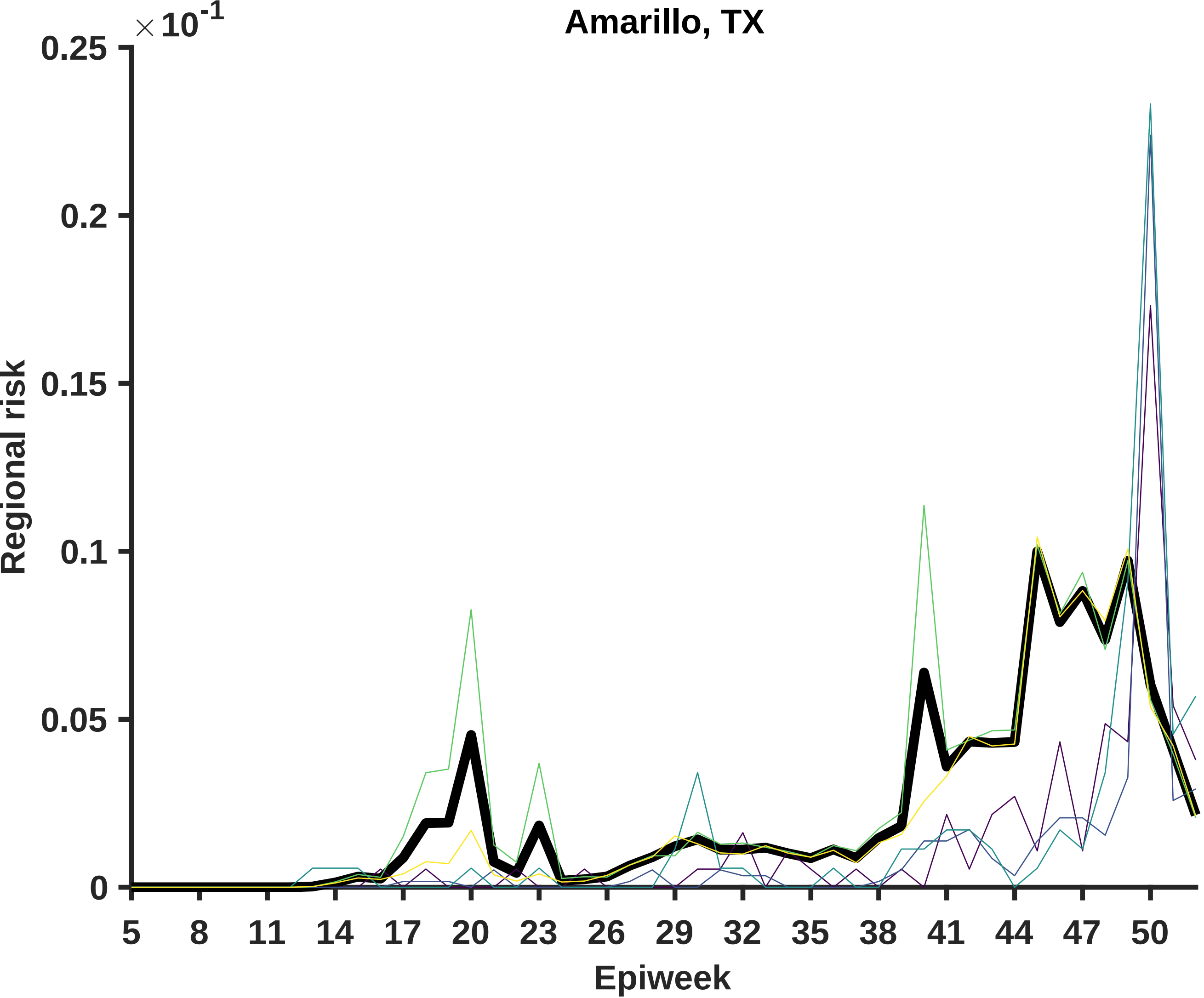

Amarillo, TX
×10^-1
0.25
0.2
0.15
0.1
0.05
0
Regional risk
5
8
11
14
17
20
23
26
29
32
35
38
41
44
47
50
Epiweek

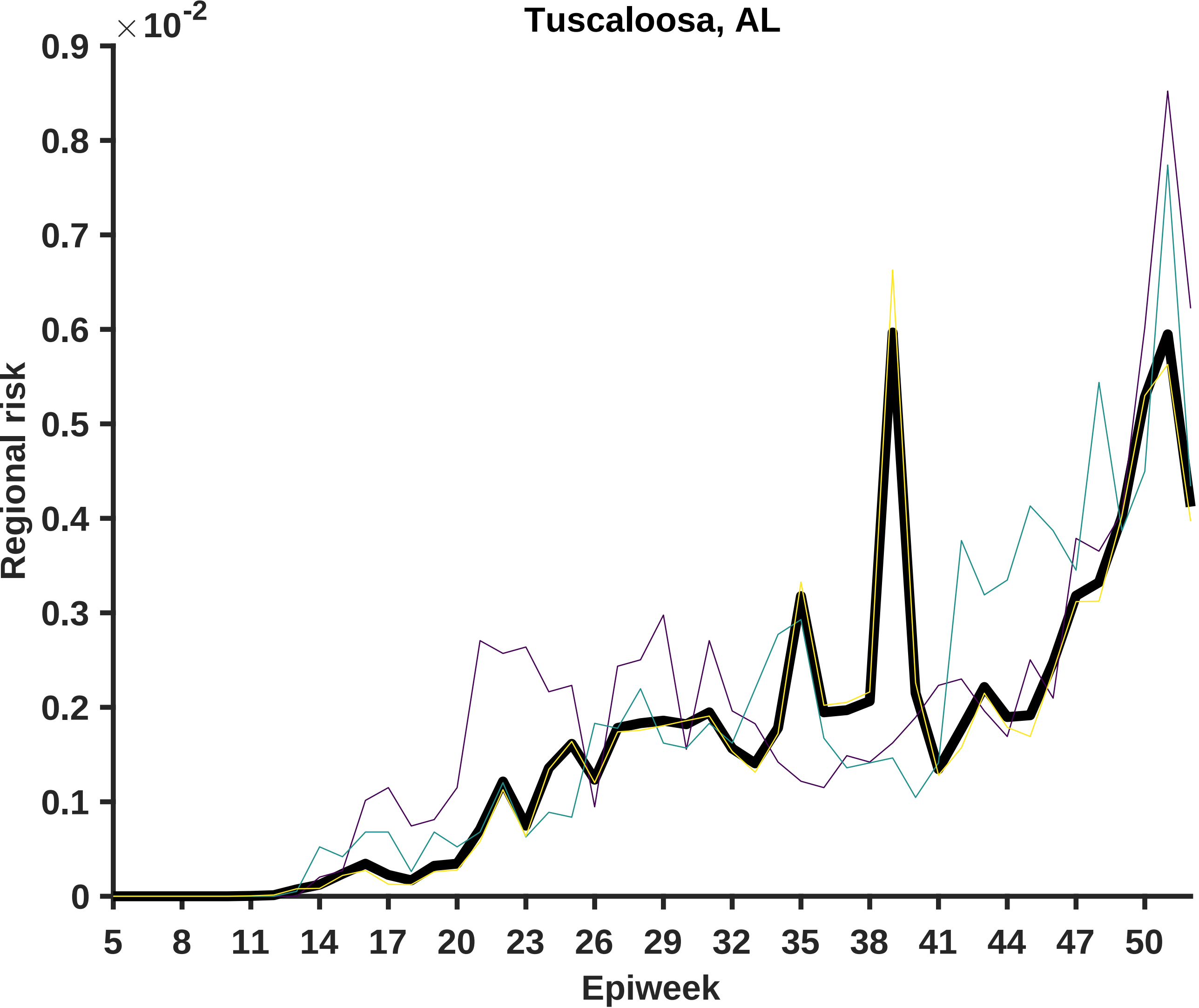

Tuscaloosa, AL
× 10^-2
0.9
0.8
0.7
0.6
0.5
0.4
0.3
0.2
0.1
0
Regional risk
5
8
11
14
17
20
23
26
29
32
35
38
41
44
47
50
Epiweek

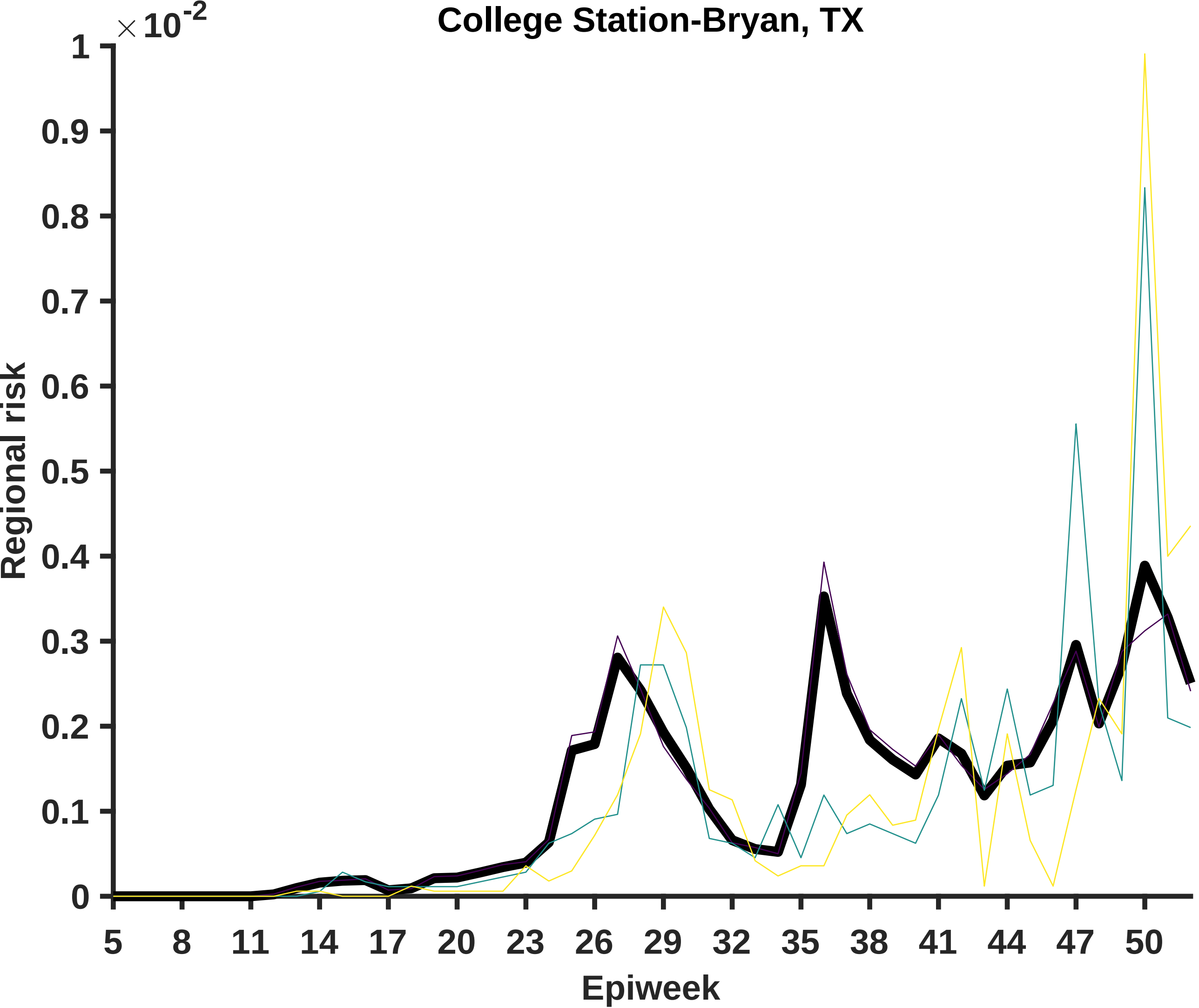

College Station-Bryan, TX
× 10^-2
1
0.9
0.8
0.7
0.6
0.5
0.4
0.3
0.2
0.1
0
Regional risk
5
8
11
14
17
20
23
26
29
32
35
38
41
44
47
50
Epiweek

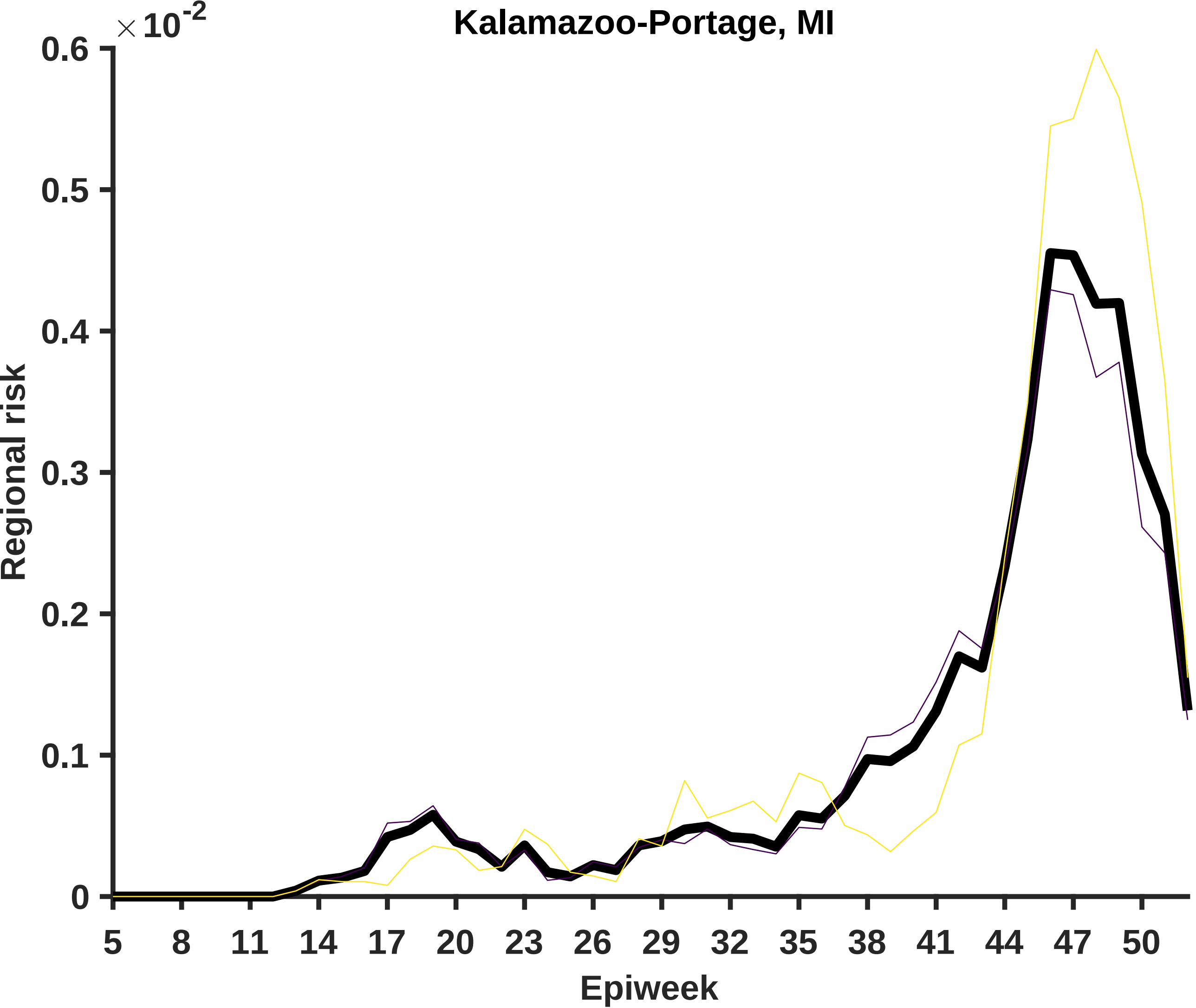

Kalamazoo-Portage, MI
×10^-2
0.6
0.5
0.4
0.3
0.2
0.1
0
Regional risk
5
8
11
14
17
20
23
26
29
32
35
38
41
44
47
50
Epiweek

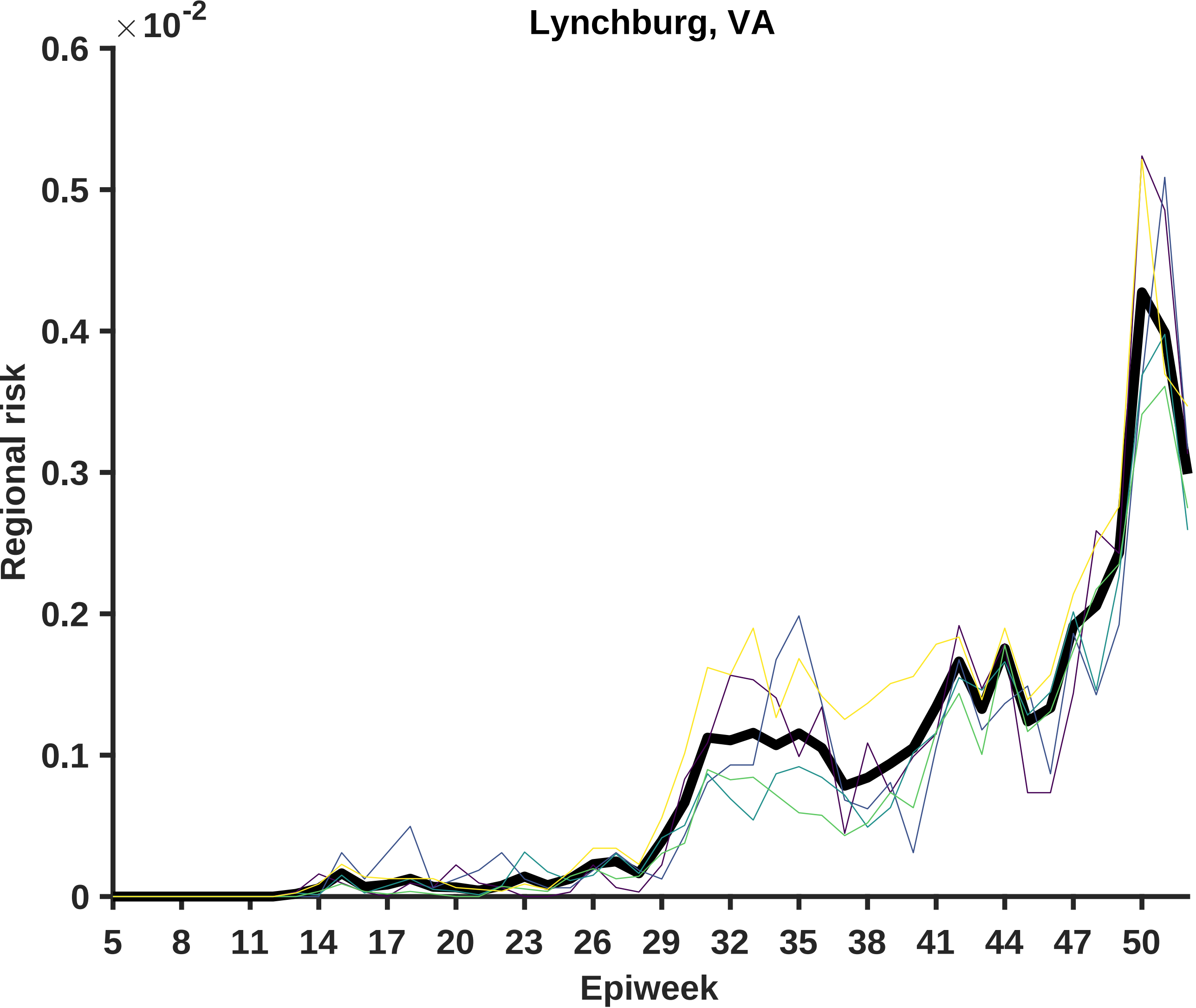

Lynchburg, VA
×10^-2
0.6
0.5
0.4
0.3
0.2
0.1
0
Regional risk
5
8
11
14
17
20
23
26
29
32
35
38
41
44
47
50
Epiweek

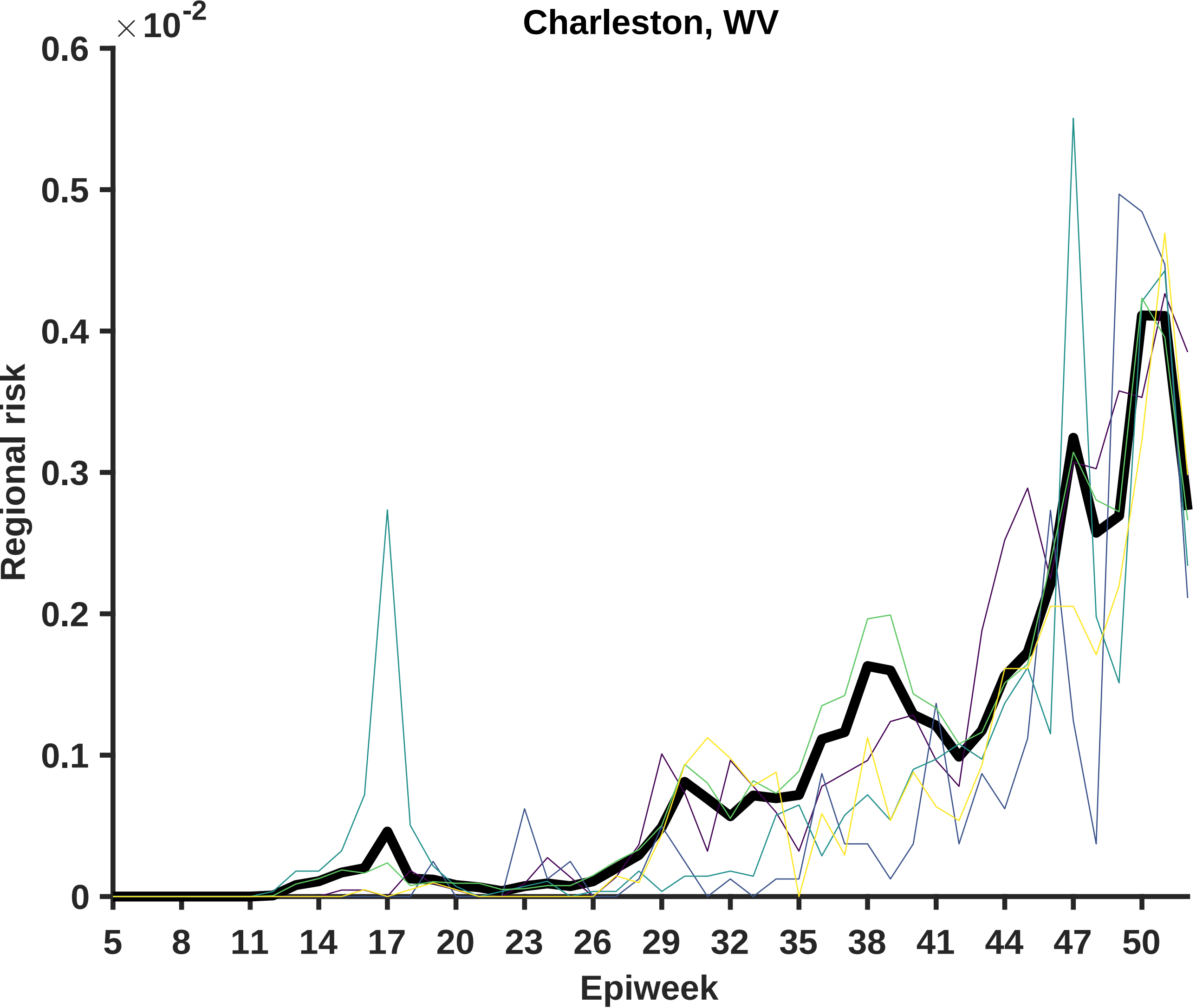

Charleston, WV
×10^-2
0.6
0.5
0.4
0.3
0.2
0.1
0
Regional risk
5
8
11
14
17
20
23
26
29
32
35
38
41
44
47
50
Epiweek

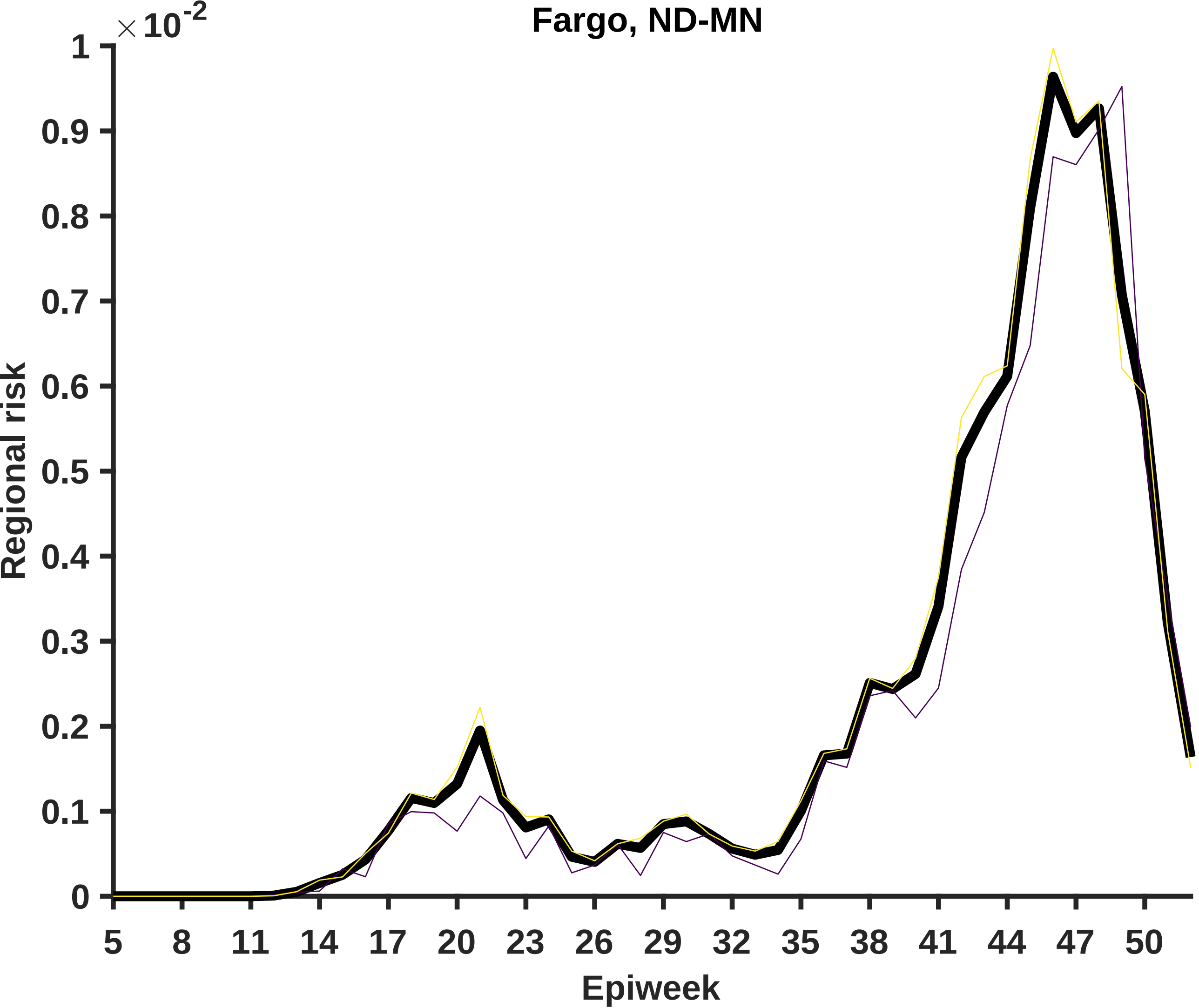

Fargo, ND-MN
×10^-2
1
0.9
0.8
0.7
0.6
0.5
0.4
0.3
0.2
0.1
0
Regional risk
5
8
11
14
17
20
23
26
29
32
35
38
41
44
47
50
Epiweek

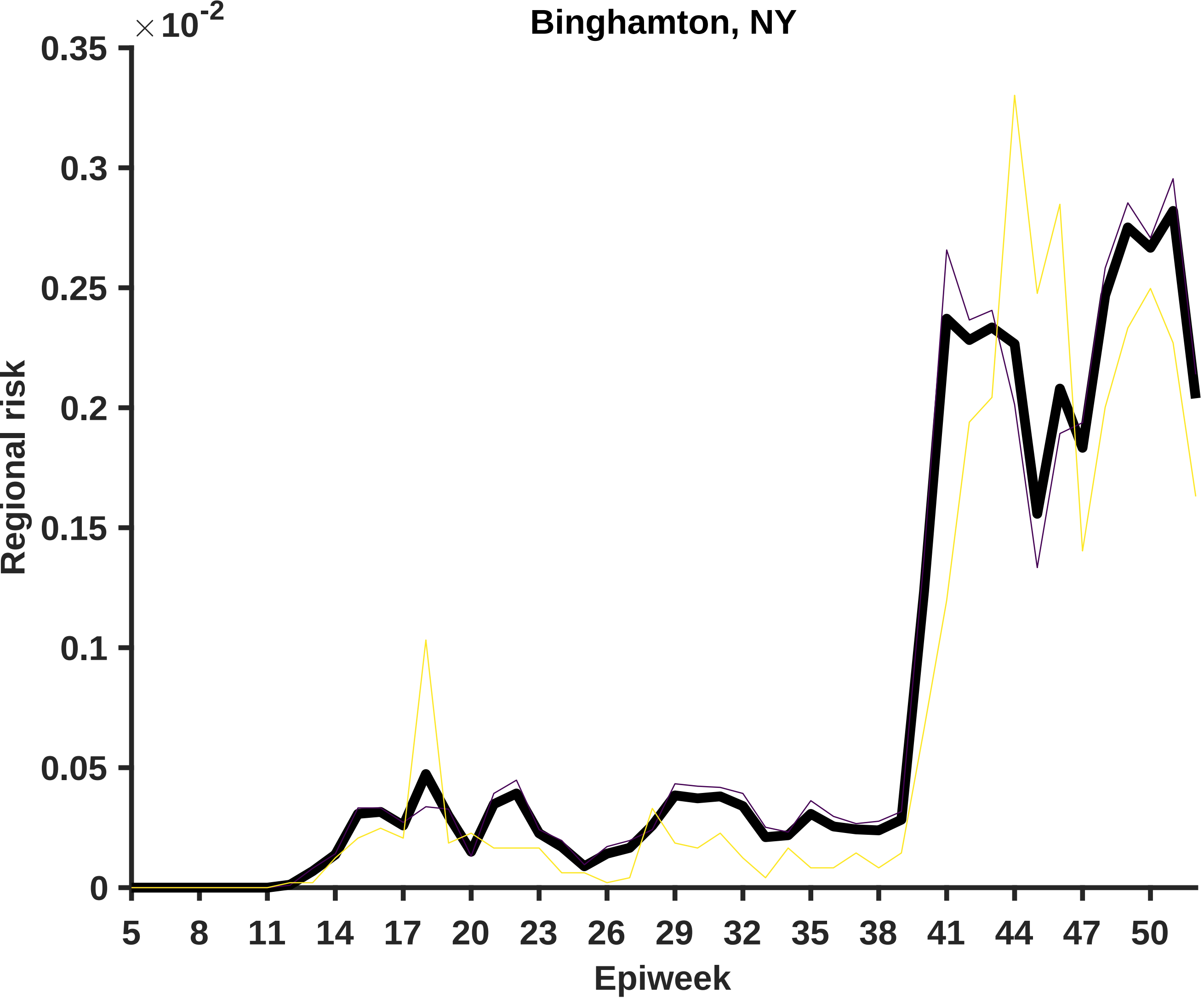

Binghamton, NY
× 10^-2
0.35
0.3
0.25
0.2
0.15
0.1
0.05
0
Regional risk
5
8
11
14
17
20
23
26
29
32
35
38
41
44
47
50
Epiweek

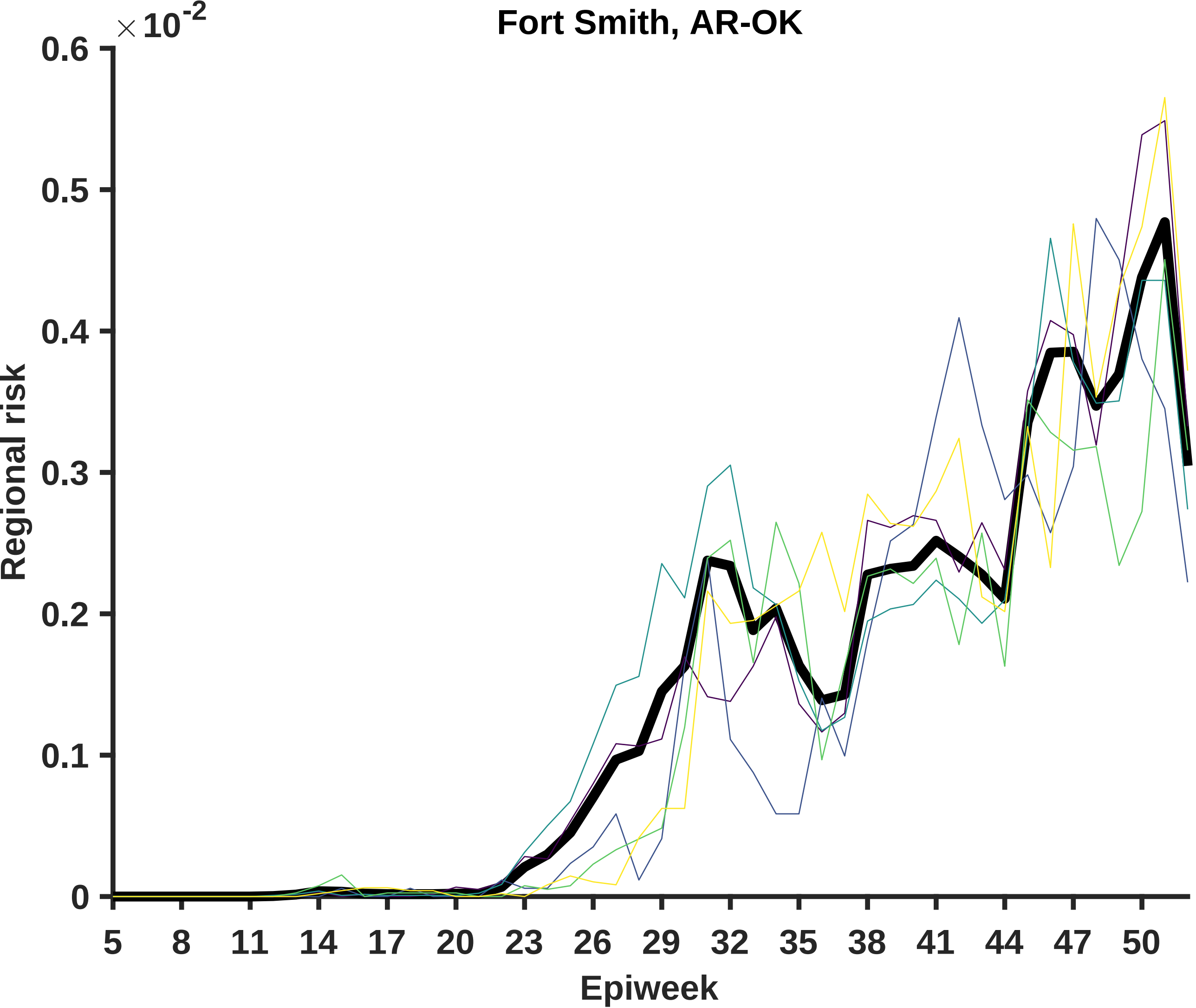

Fort Smith, AR-OK
×10^-2
0.6
0.5
0.4
0.3
0.2
0.1
0
Regional risk
5
8
11
14
17
20
23
26
29
32
35
38
41
44
47
50
Epiweek

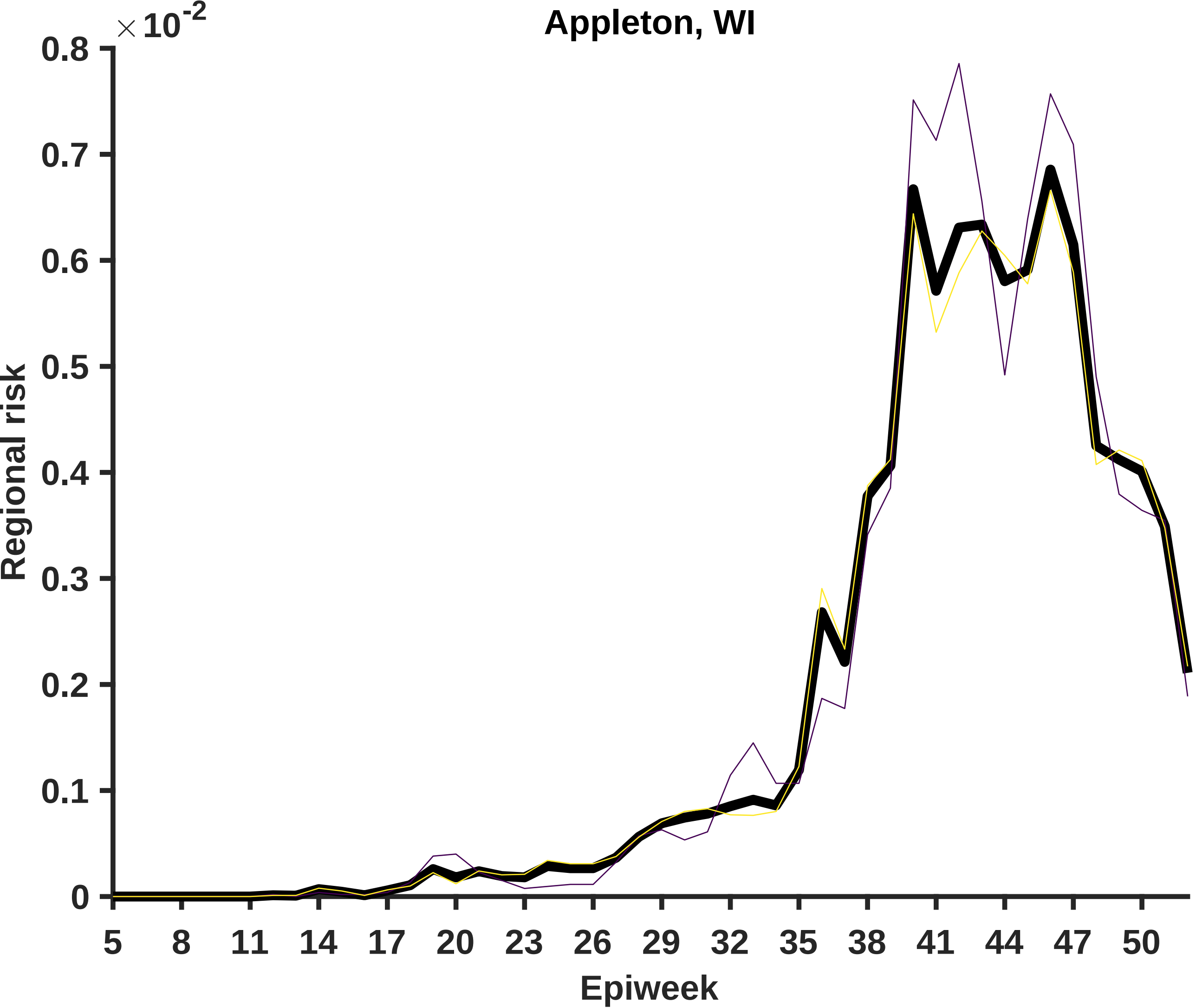

Appleton, WI
×10^-2
0.8
0.7
0.6
0.5
0.4
0.3
0.2
0.1
0
Regional risk
5
8
11
14
17
20
23
26
29
32
35
38
41
44
47
50
Epiweek

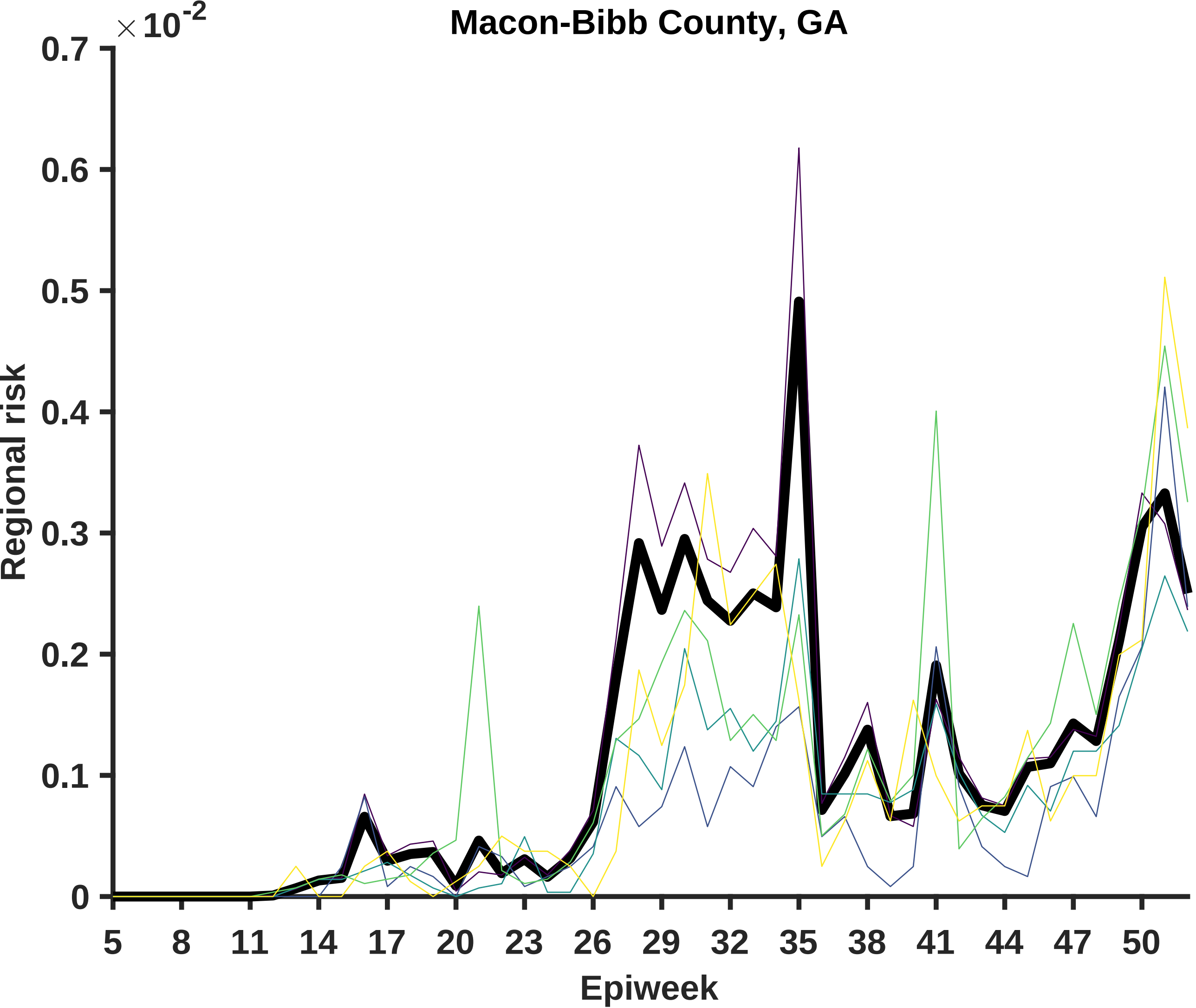

Macon-Bibb County, GA
×10^-2
0.7
0.6
0.5
0.4
0.3
0.2
0.1
0
Regional risk
5
8
11
14
17
20
23
26
29
32
35
38
41
44
47
50
Epiweek

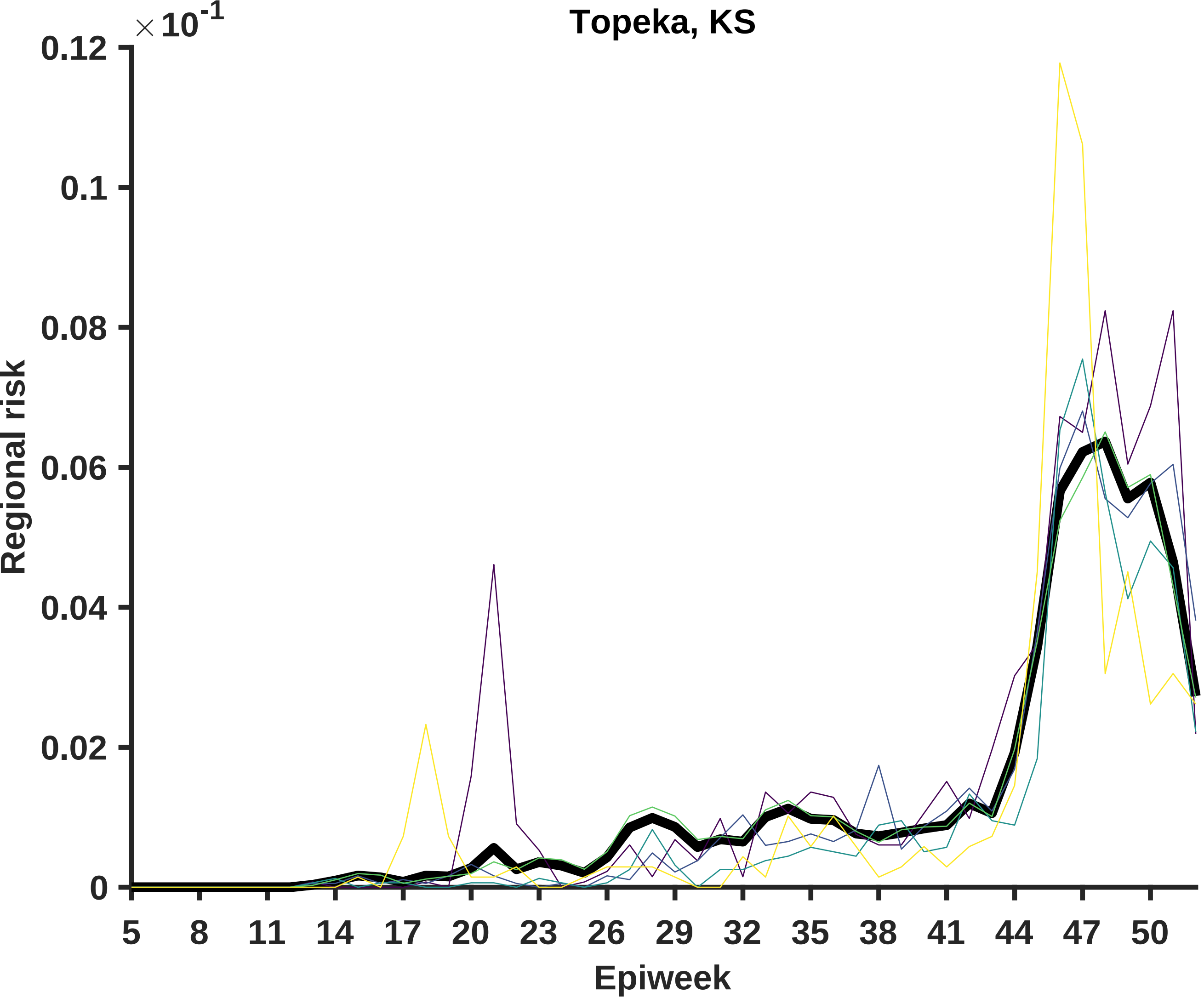

Topeka, KS
× 10^-1
0.12
0.1
0.08
0.06
0.04
0.02
0
Regional risk
5
8
11
14
17
20
23
26
29
32
35
38
41
44
47
50
Epiweek

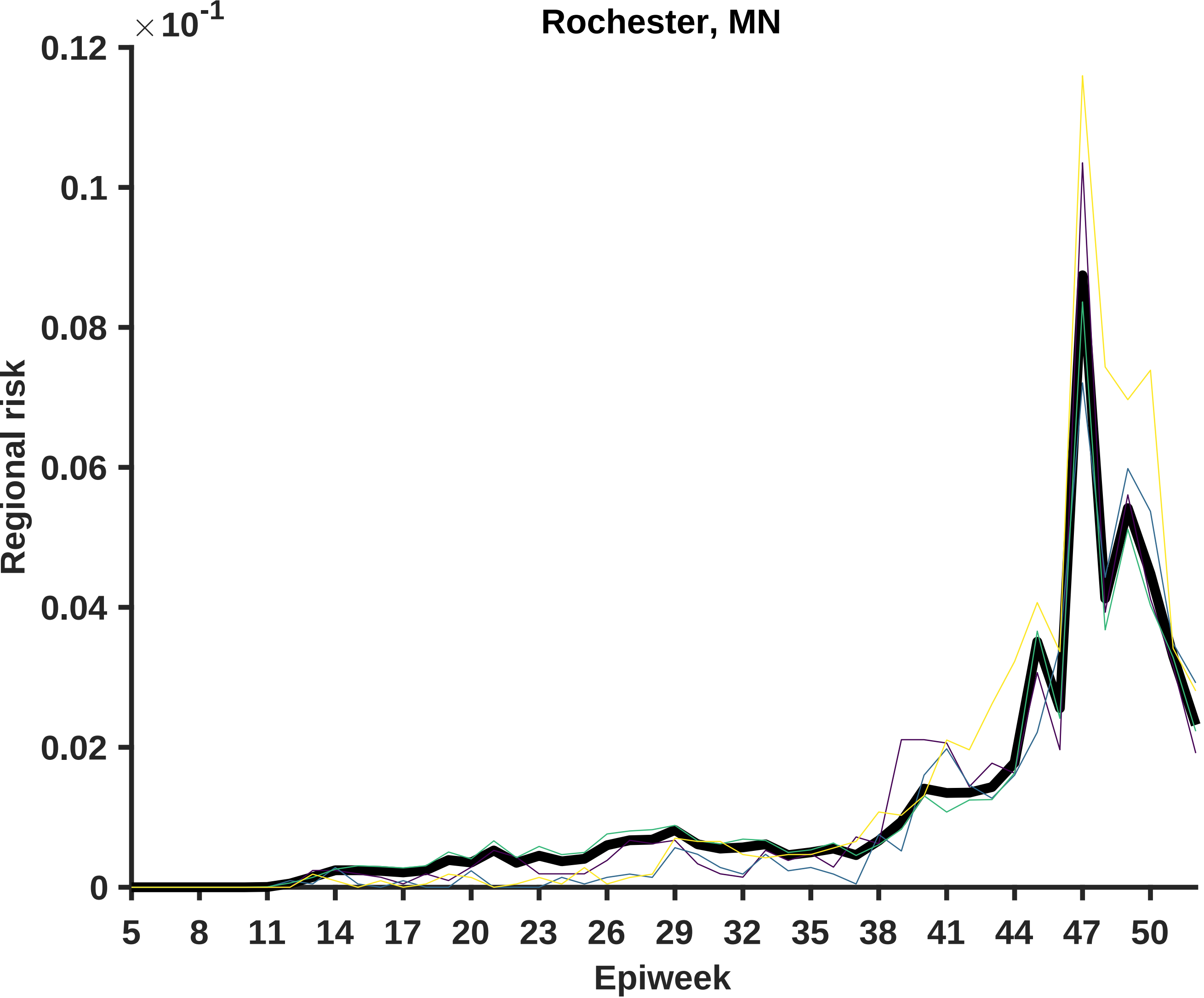
Rochester, MN
× 10^-1
0.12
0.1
0.08
0.06
0.04
0.02
0
Regional risk
5
8
11
14
17
20
23
26
29
32
35
38
41
44
47
50
Epiweek

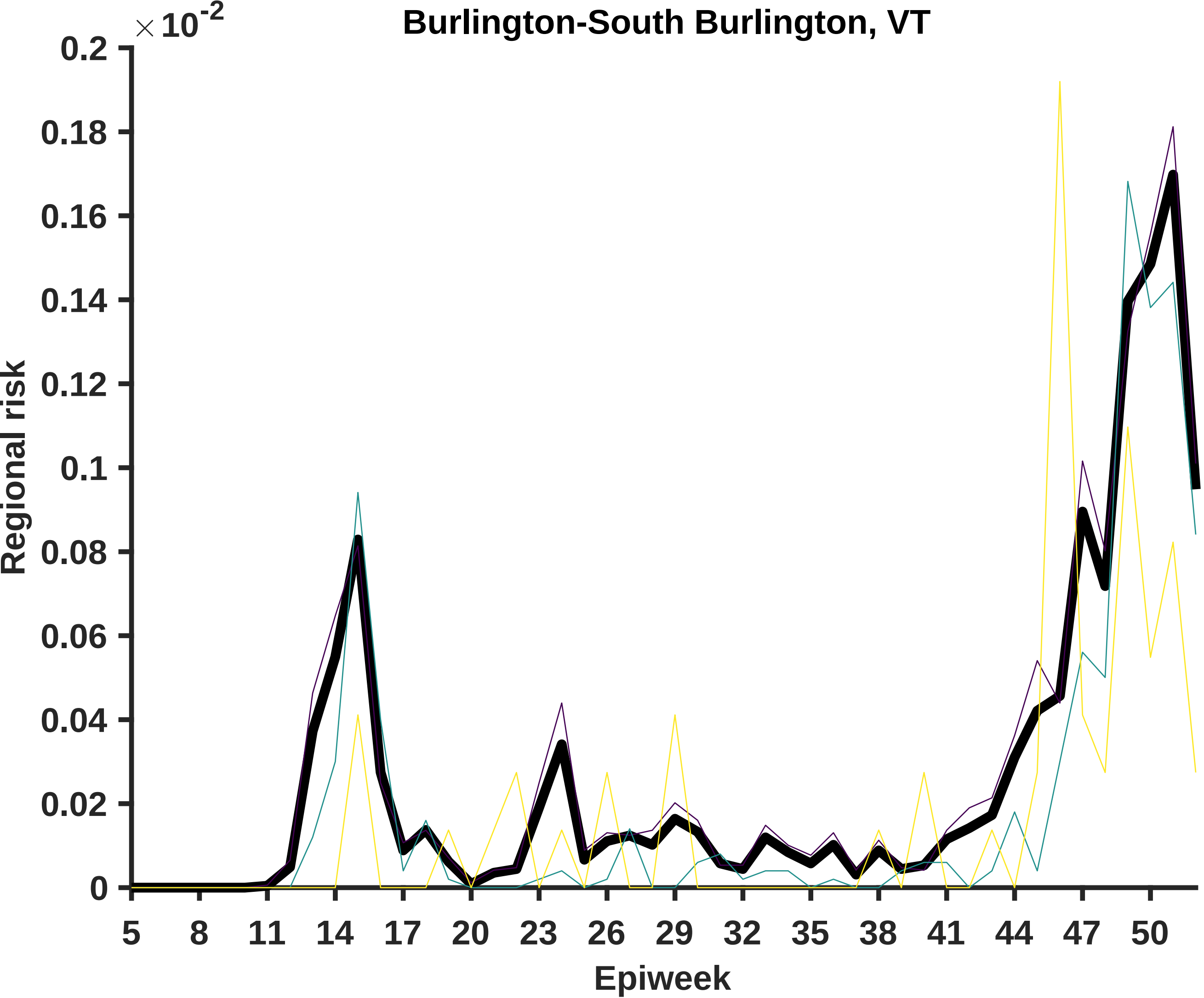

Burlington-South Burlington, VT
× 10^-2
0.2
0.18
0.16
0.14
0.12
0.1
0.08
0.06
0.04
0.02
0
Regional risk
5
8
11
14
17
20
23
26
29
32
35
38
41
44
47
50
Epiweek

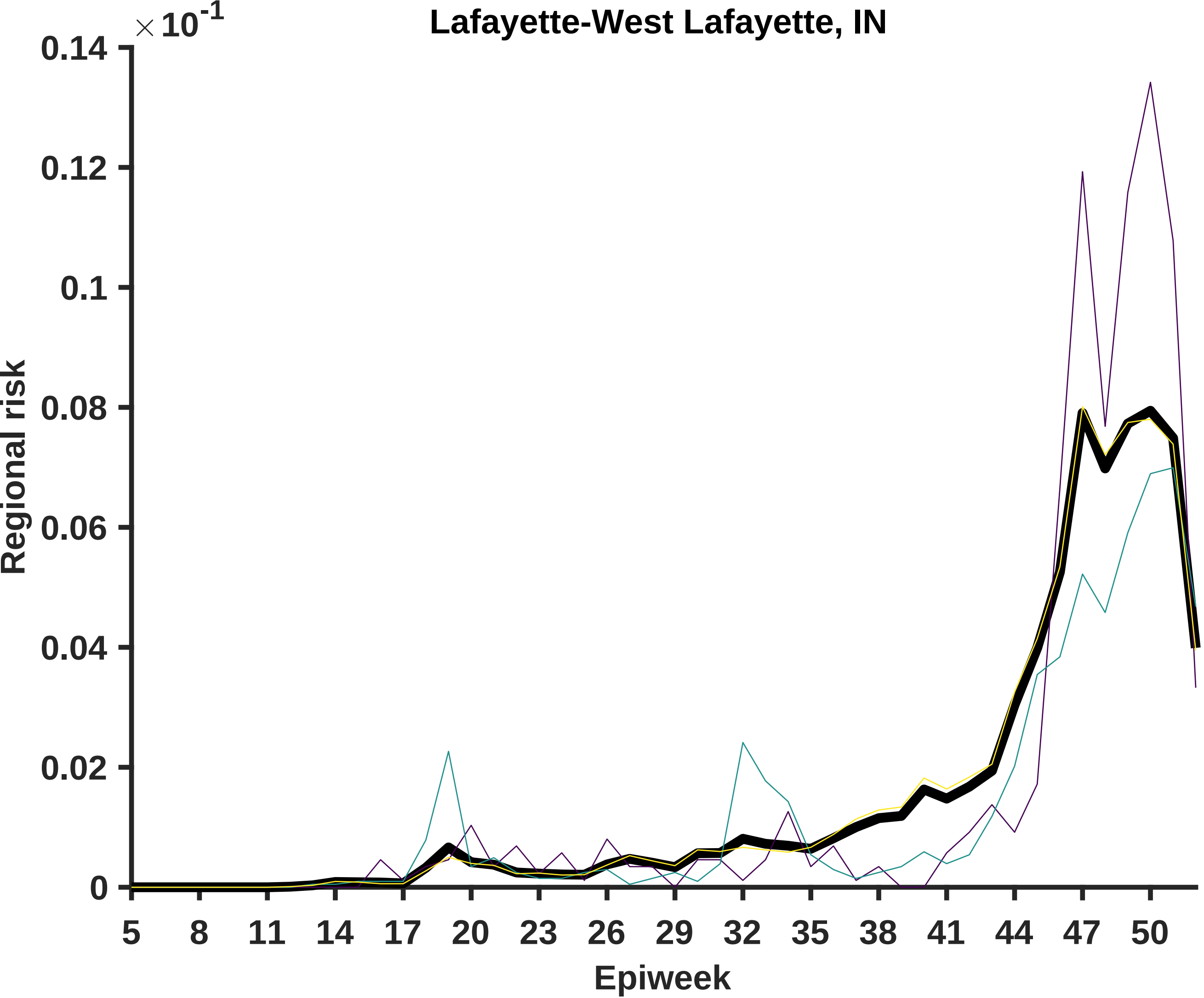

Lafayette-West Lafayette, IN
×10^-1
0.14
0.12
0.1
0.08
0.06
0.04
0.02
0
Regional risk
5
8
11
14
17
20
23
26
29
32
35
38
41
44
47
50
Epiweek

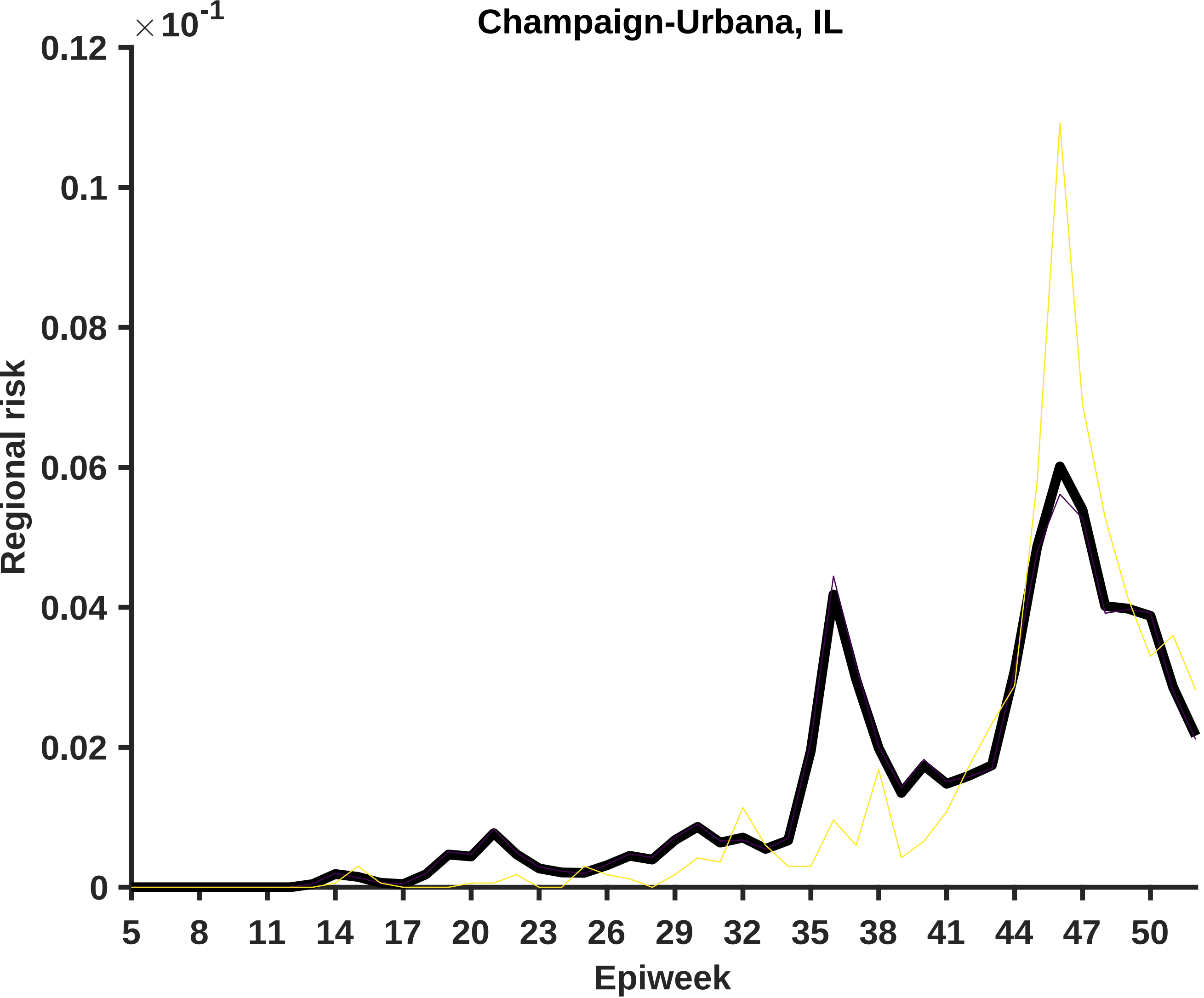

Champaign-Urbana, IL
× 10^-1
Regional risk
0.12
0.1
0.08
0.06
0.04
0.02
0
5
8
11
14
17
20
23
26
29
32
35
38
41
44
47
50
Epiweek

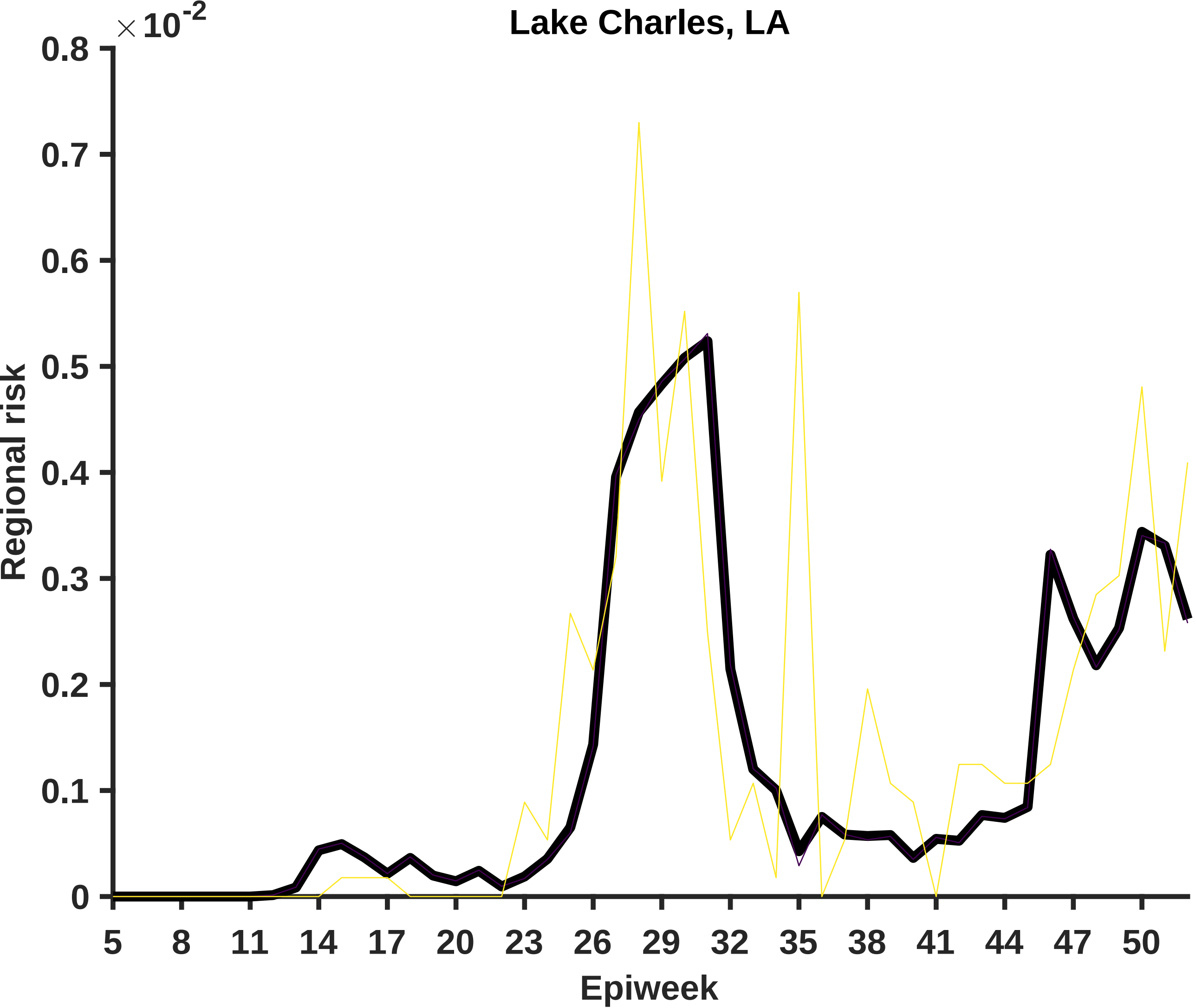

Lake Charles, LA
×10^-2
0.8
0.7
0.6
0.5
0.4
0.3
0.2
0.1
0
Regional risk
5
8
11
14
17
20
23
26
29
32
35
38
41
44
47
50
Epiweek

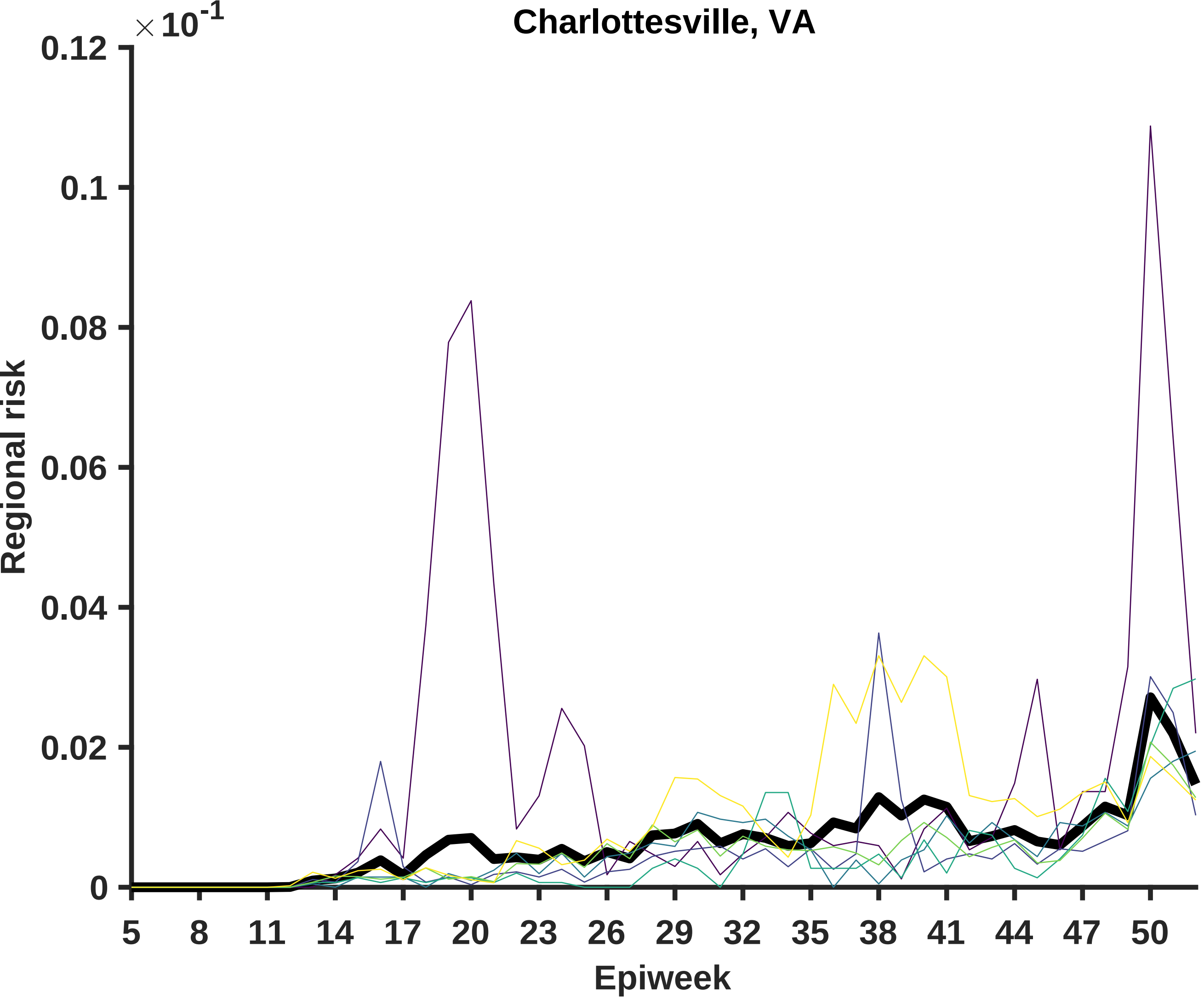

Charlottesville, VA
×10^-1
0.12
0.1
0.08
0.06
0.04
0.02
0
Regional risk
5
8
11
14
17
20
23
26
29
32
35
38
41
44
47
50
Epiweek

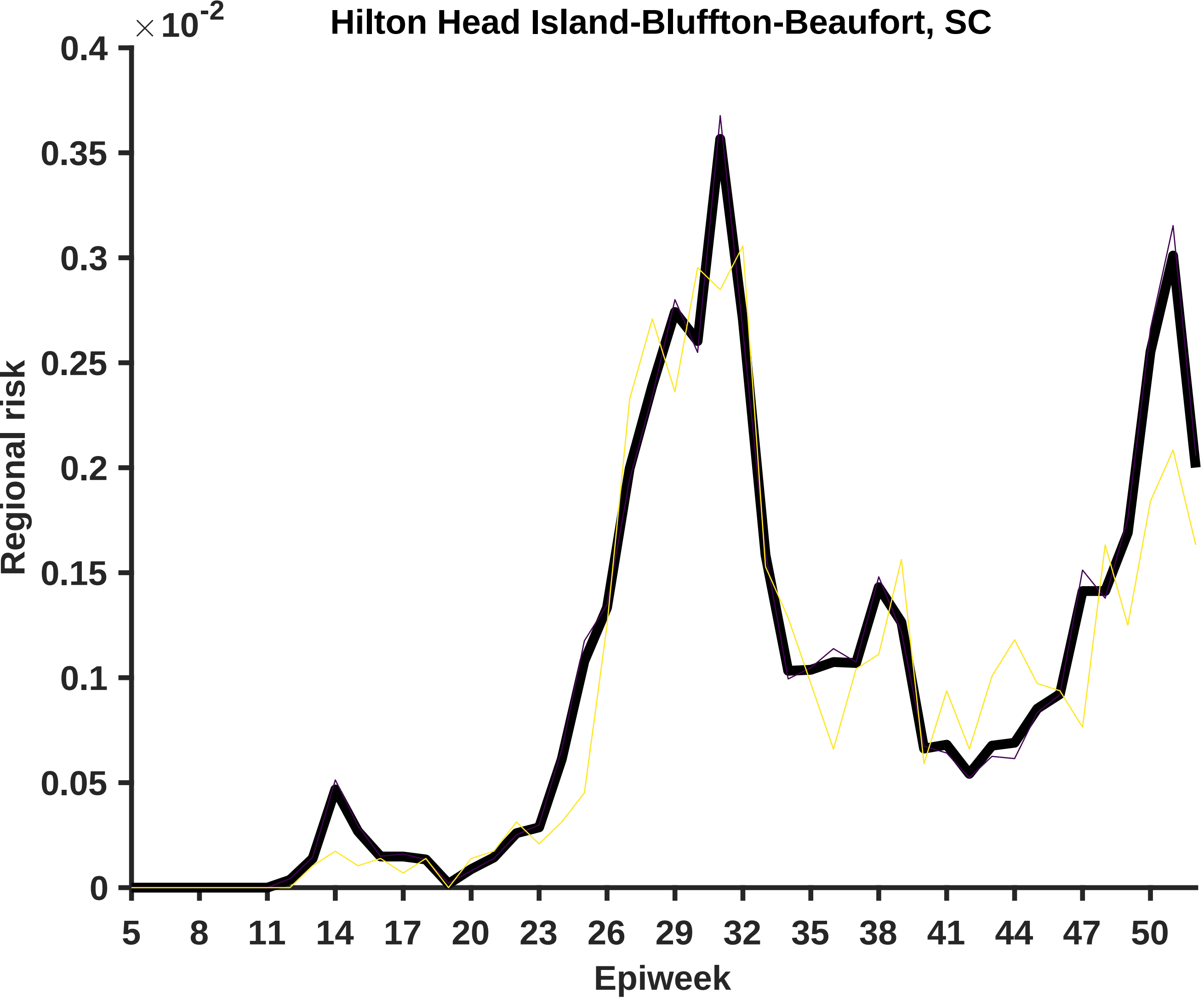

$\times 10^{-2}$
Hilton Head Island-Bluffton-Beaufort, SC
0.4
0.35
0.3
0.25
0.2
0.15
0.1
0.05
0
Regional risk
5
8
11
14
17
20
23
26
29
32
35
38
41
44
47
50
Epiweek

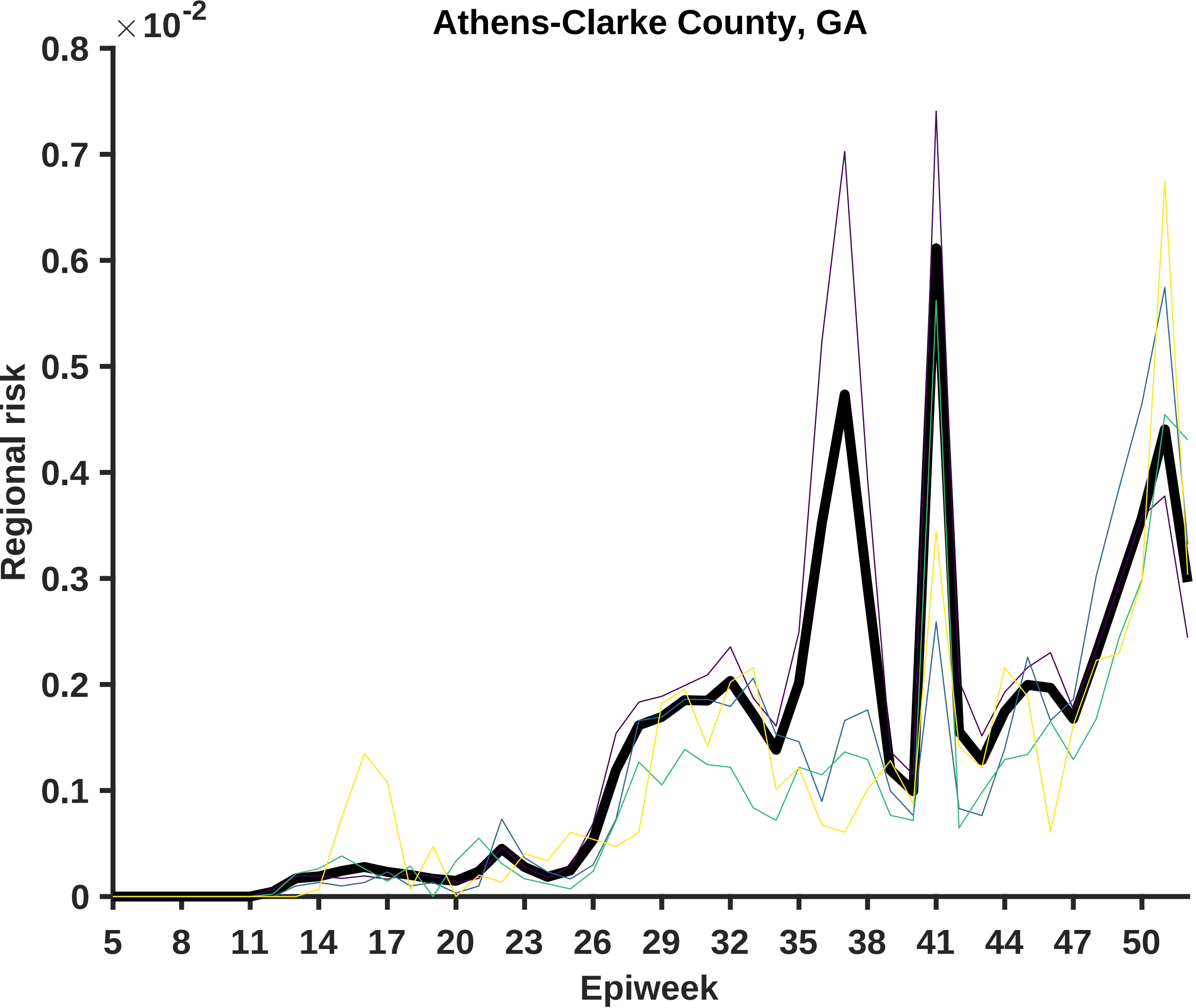

Athens-Clarke County, GA
×10^-2
0.8
0.7
0.6
0.5
0.4
0.3
0.2
0.1
0
Regional risk
5
8
11
14
17
20
23
26
29
32
35
38
41
44
47
50
Epiweek

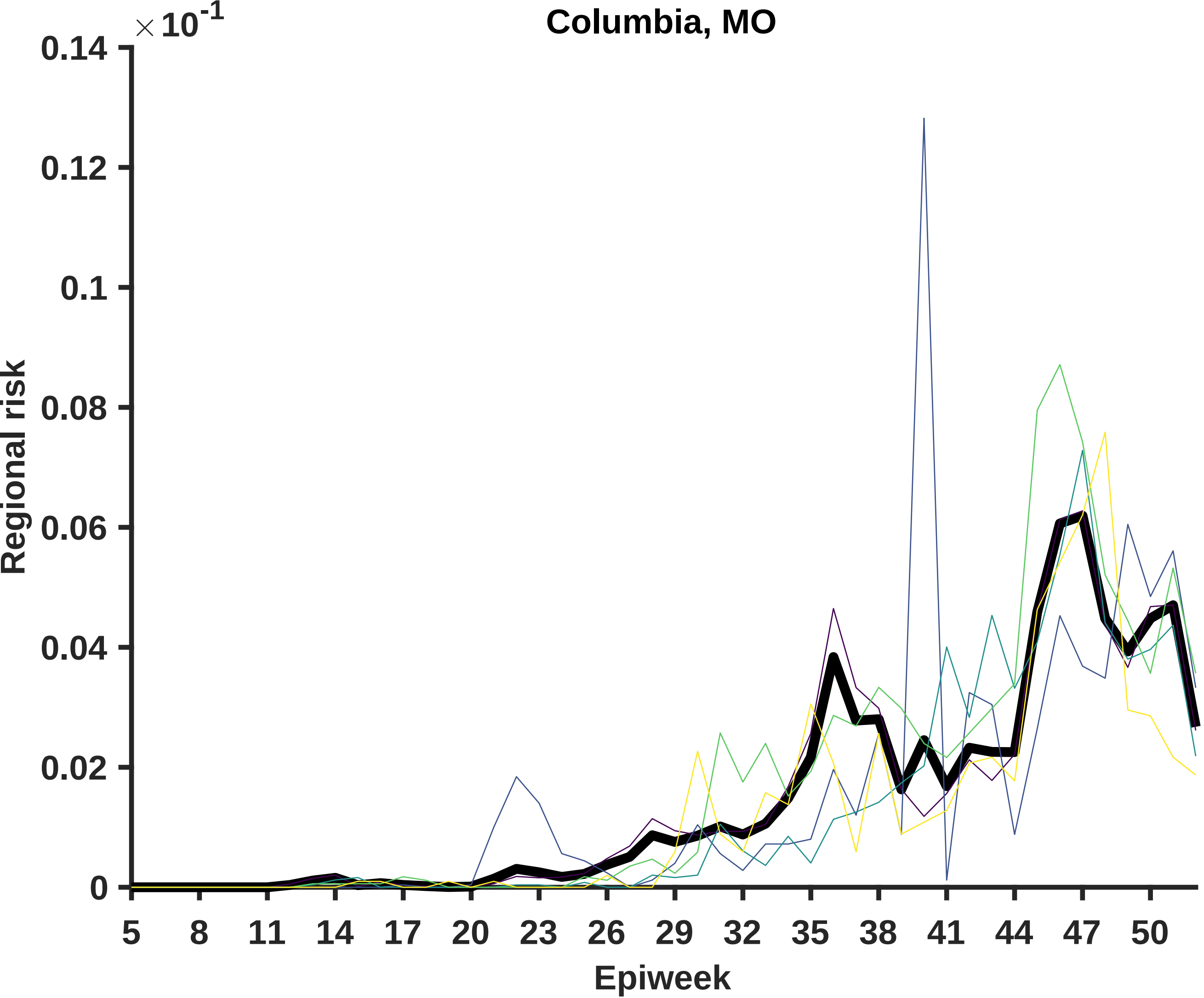
Columbia, MO
×10^-1
0.14
0.12
0.1
0.08
0.06
0.04
0.02
0
Regional risk
5
8
11
14
17
20
23
26
29
32
35
38
41
44
47
50
Epiweek

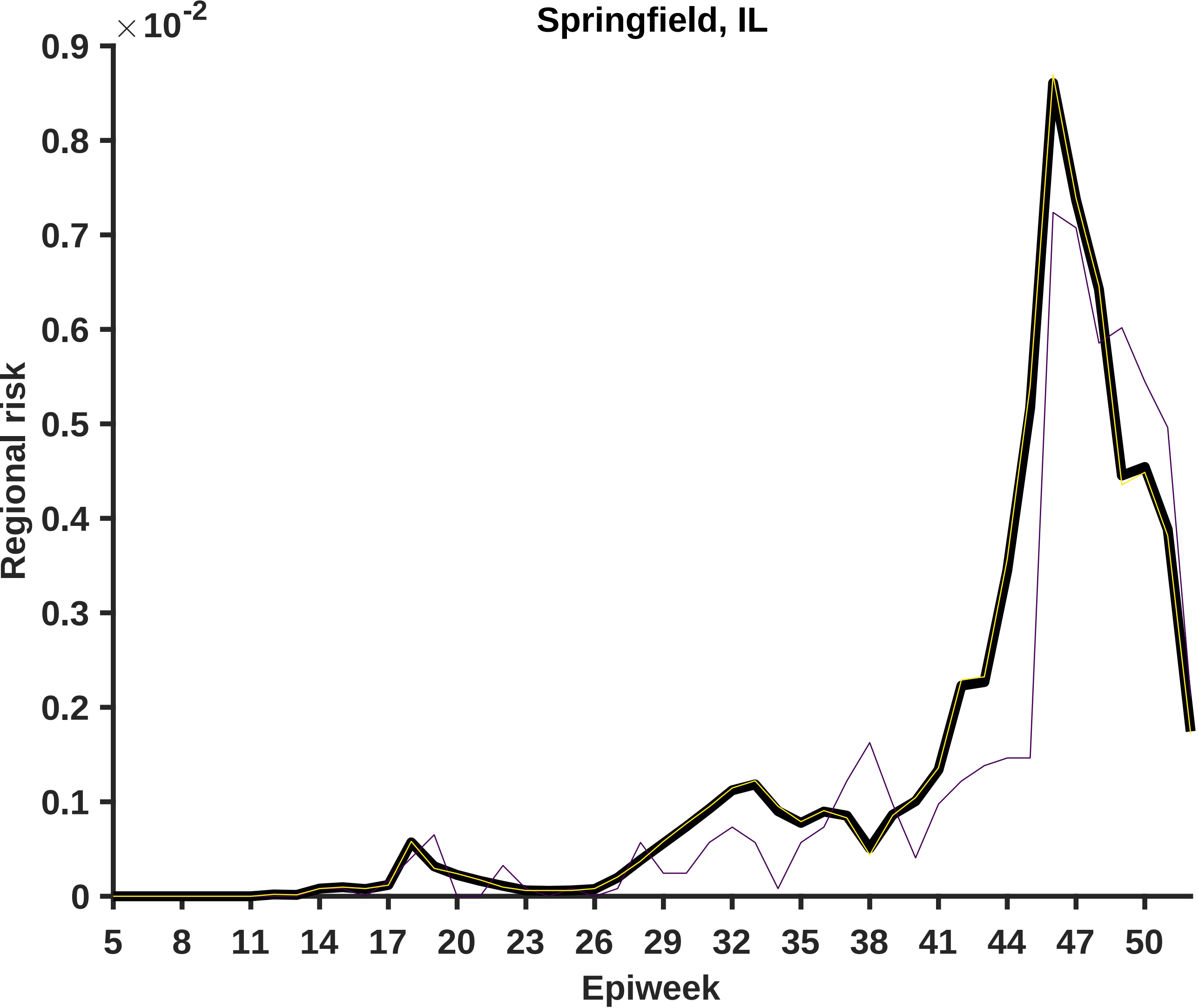

Springfield, IL
×10^-2
0.9
0.8
0.7
0.6
0.5
0.4
0.3
0.2
0.1
0
Regional risk
5
8
11
14
17
20
23
26
29
32
35
38
41
44
47
50
Epiweek

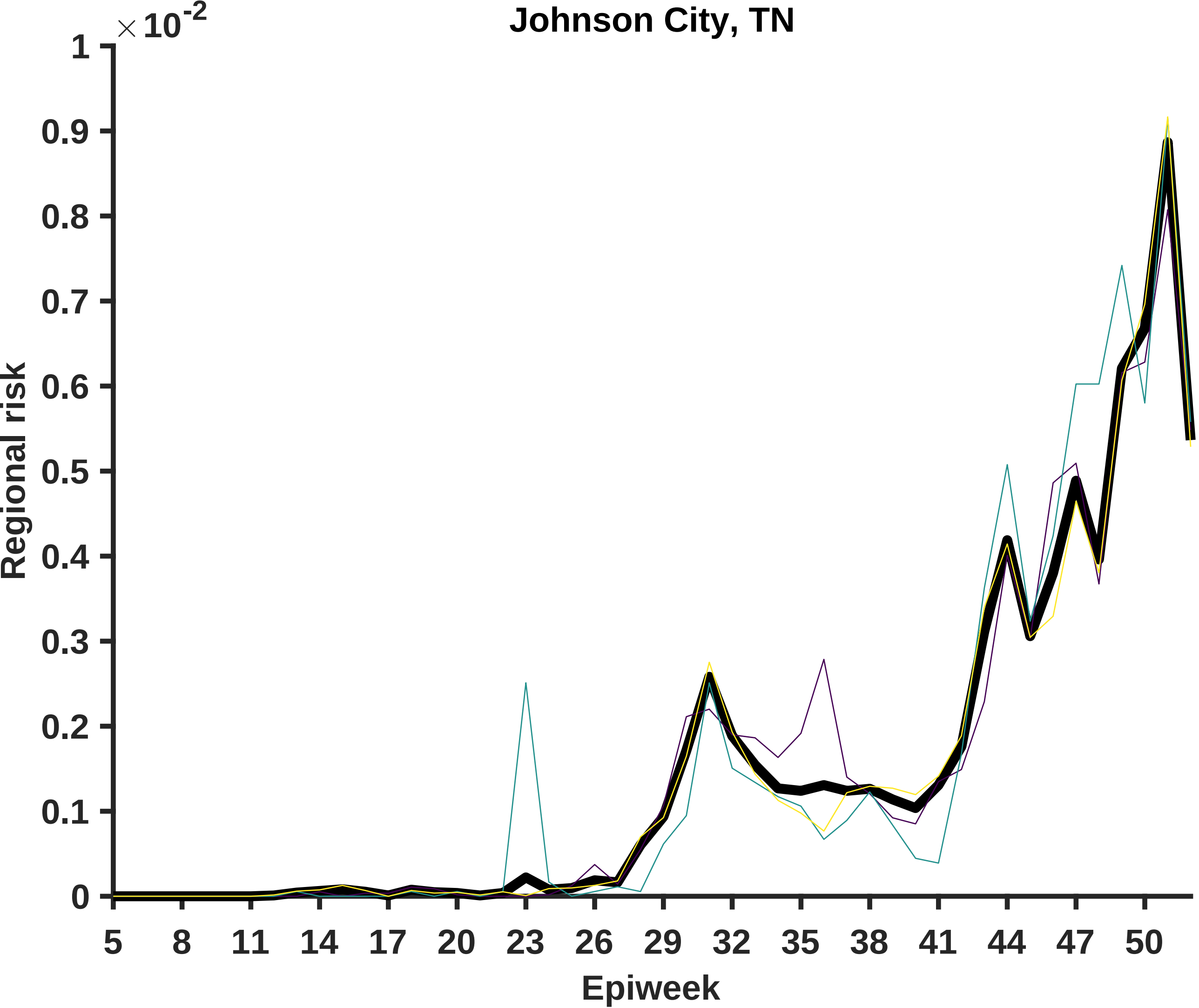
Johnson City, TN
×10^-2
1
0.9
0.8
0.7
0.6
0.5
0.4
0.3
0.2
0.1
0
Regional risk
5
8
11
14
17
20
23
26
29
32
35
38
41
44
47
50
Epiweek

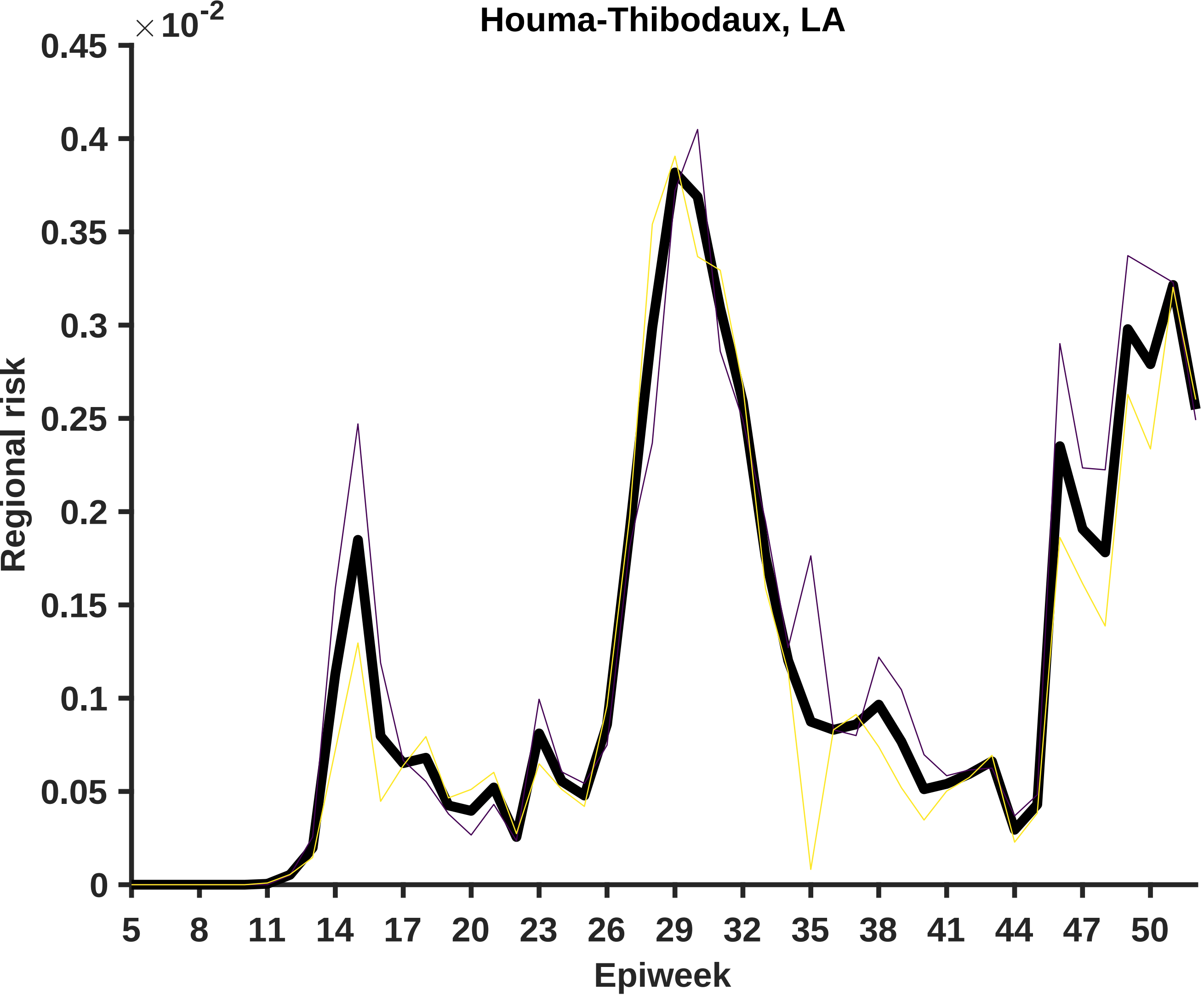
Houma-Thibodaux, LA
×10^-2
Regional risk
0.45
0.4
0.35
0.3
0.25
0.2
0.15
0.1
0.05
0
5
8
11
14
17
20
23
26
29
32
35
38
41
44
47
50
Epiweek

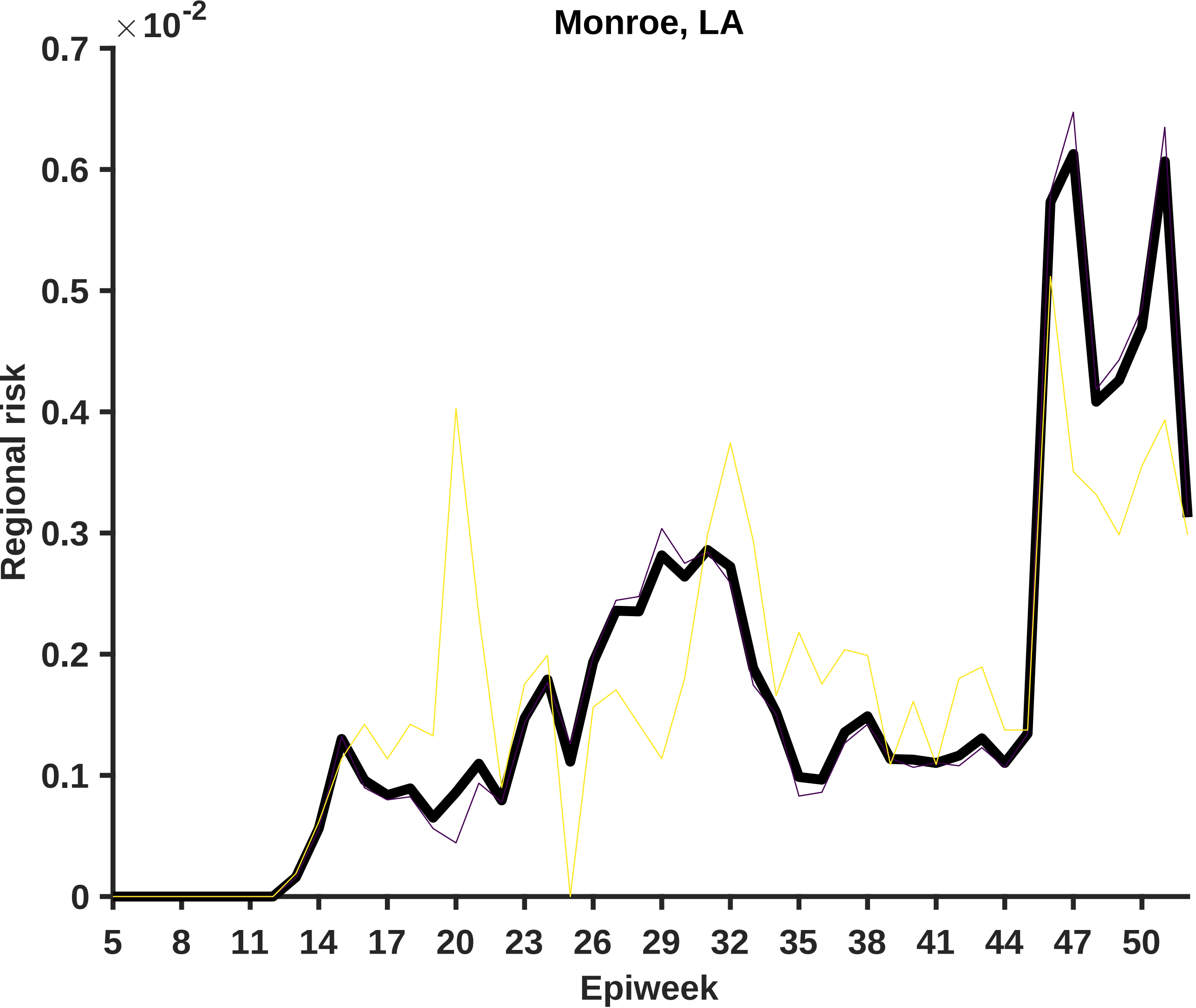
Monroe, LA
×10^-2
0.7
0.6
0.5
0.4
0.3
0.2
0.1
0
Regional risk
5
8
11
14
17
20
23
26
29
32
35
38
41
44
47
50
Epiweek

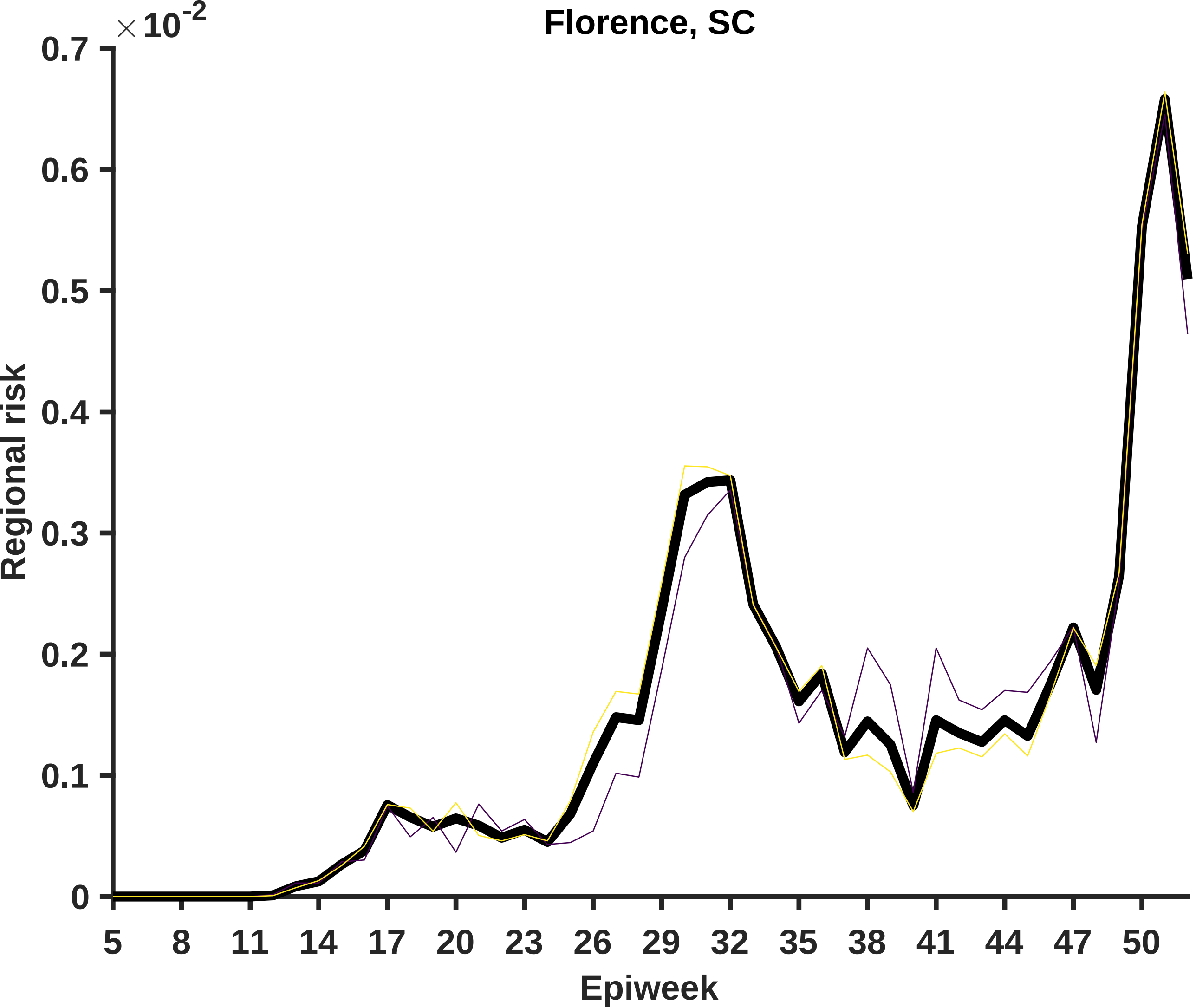

Florence, SC
×10^-2
0.7
0.6
0.5
0.4
0.3
0.2
0.1
0
Regional risk
5
8
11
14
17
20
23
26
29
32
35
38
41
44
47
50
Epiweek

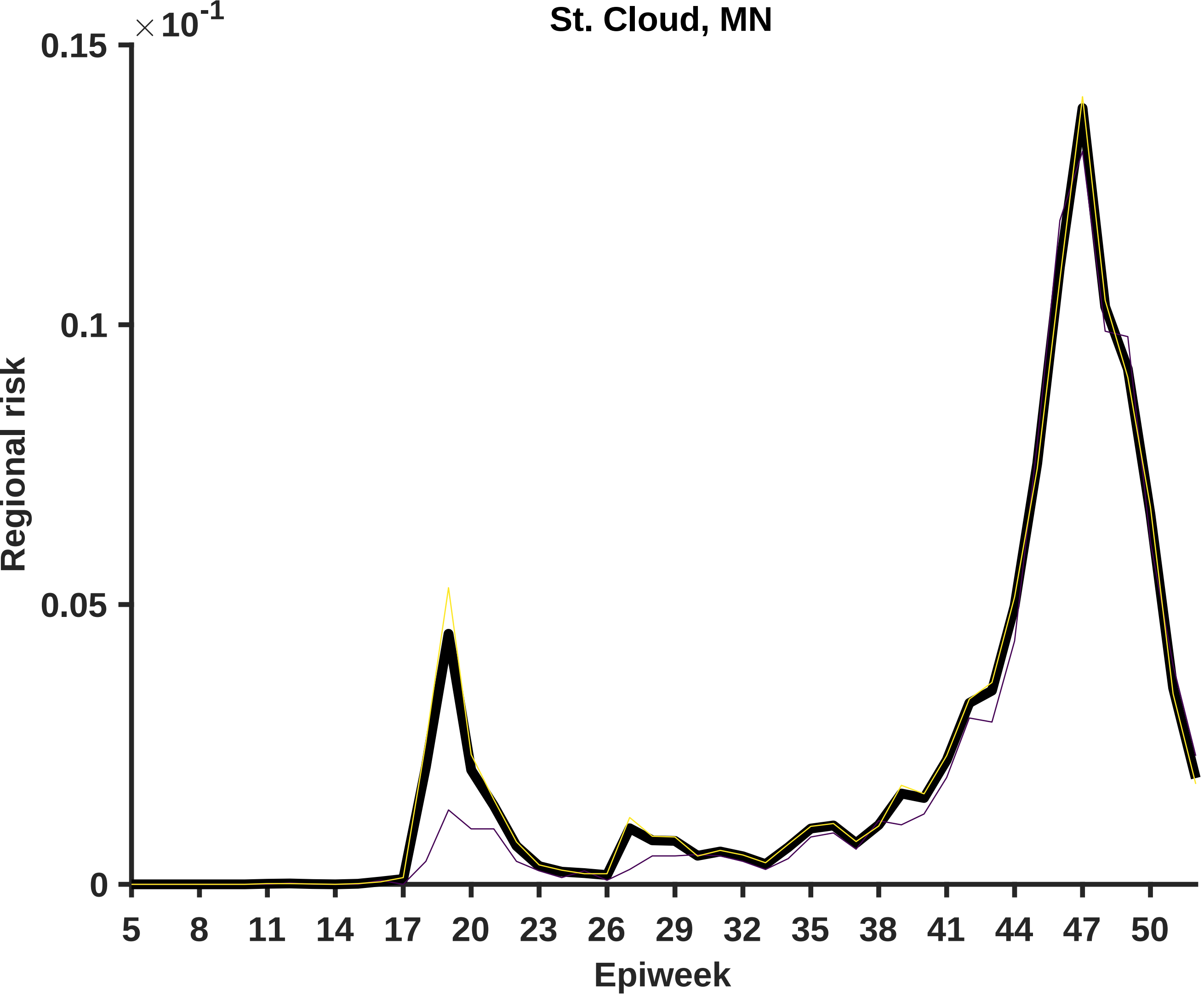

St. Cloud, MN
×10^-1
0.15
0.1
0.05
0
Regional risk
5
8
11
14
17
20
23
26
29
32
35
38
41
44
47
50
Epiweek

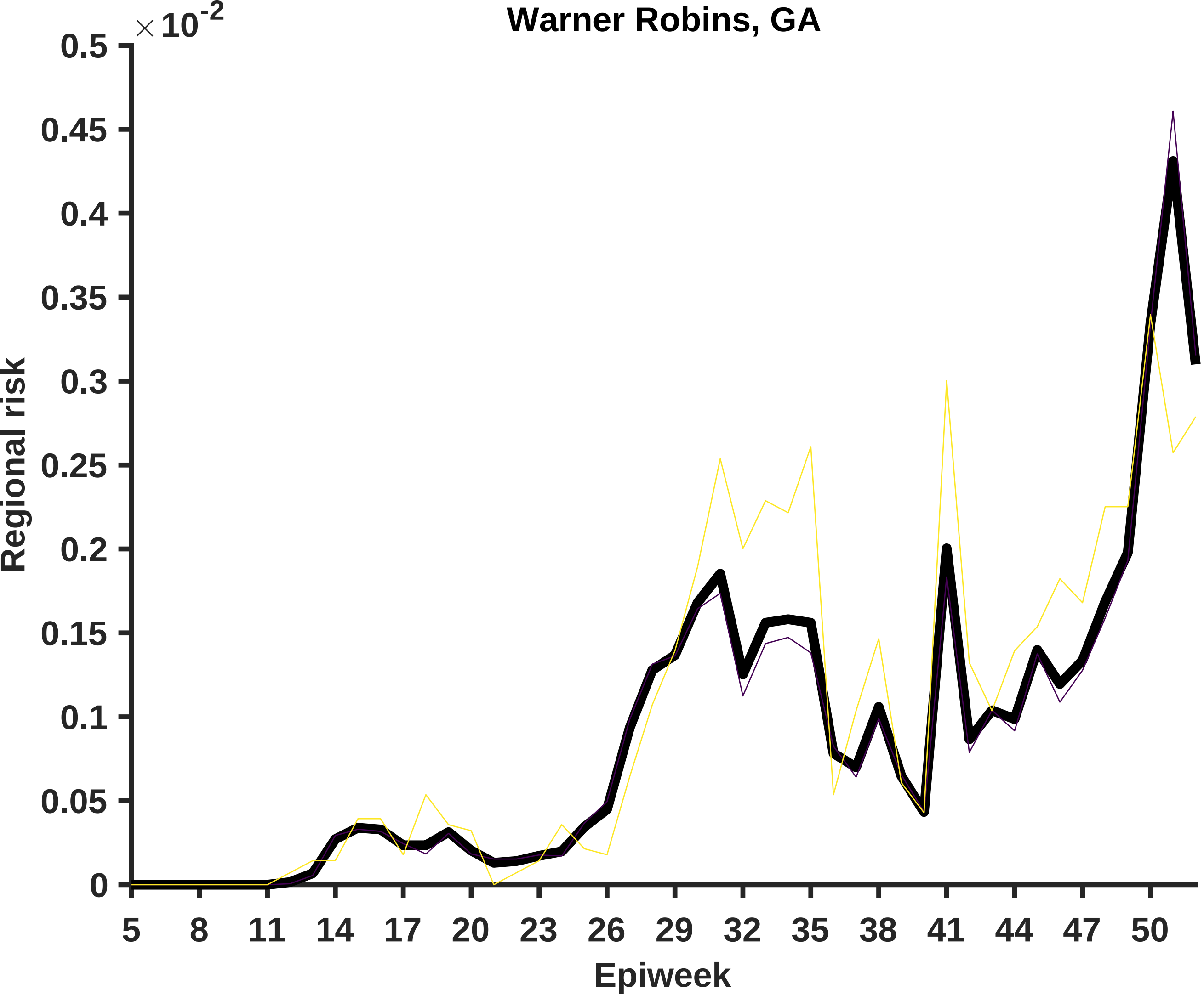

Warner Robins, GA
× 10^-2
0.5
0.45
0.4
0.35
0.3
0.25
0.2
0.15
0.1
0.05
0
Regional risk
5
8
11
14
17
20
23
26
29
32
35
38
41
44
47
50
Epiweek

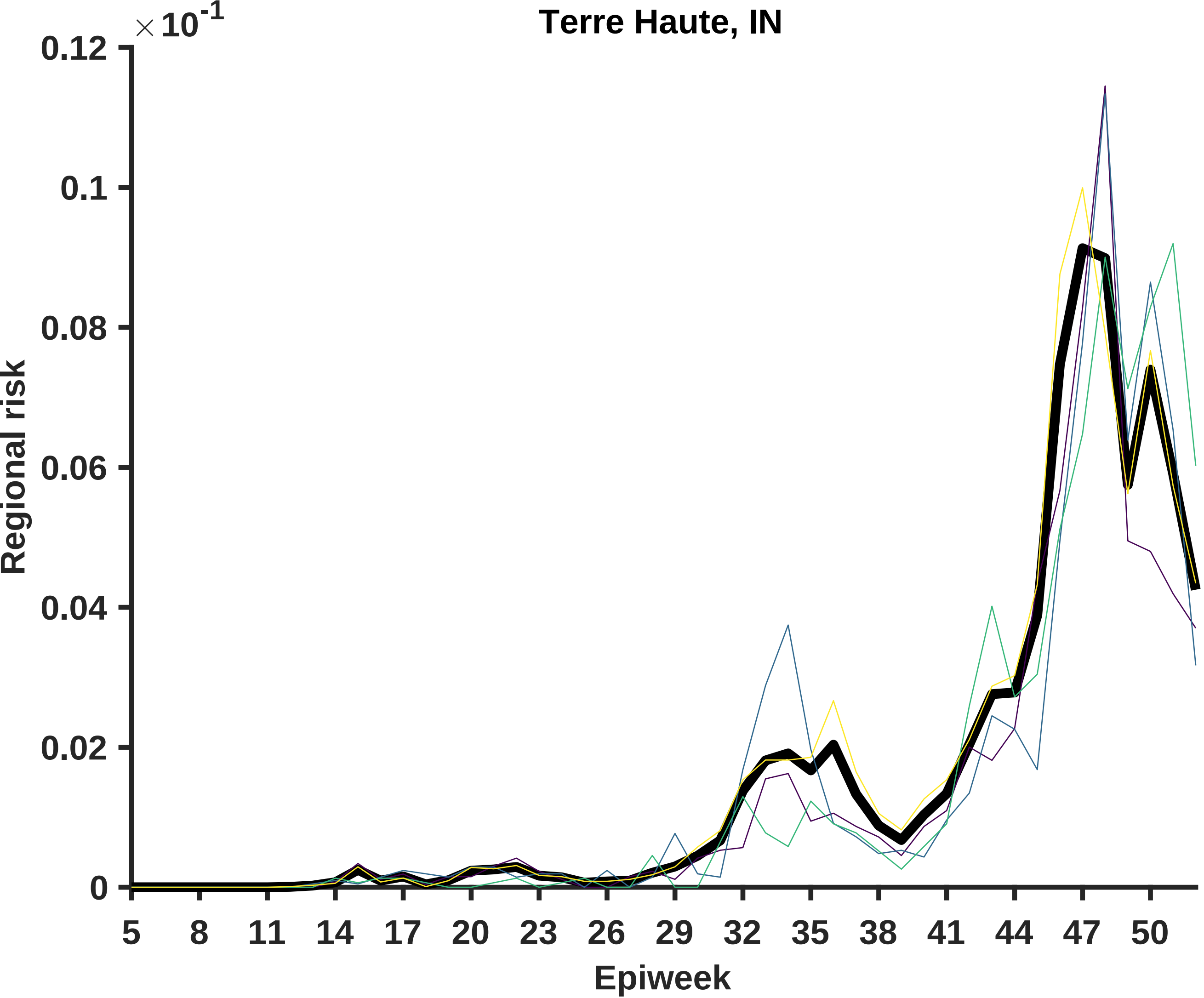
Terre Haute, IN
× 10^-1
0.12
0.1
0.08
0.06
0.04
0.02
0
Regional risk
5
8
11
14
17
20
23
26
29
32
35
38
41
44
47
50
Epiweek

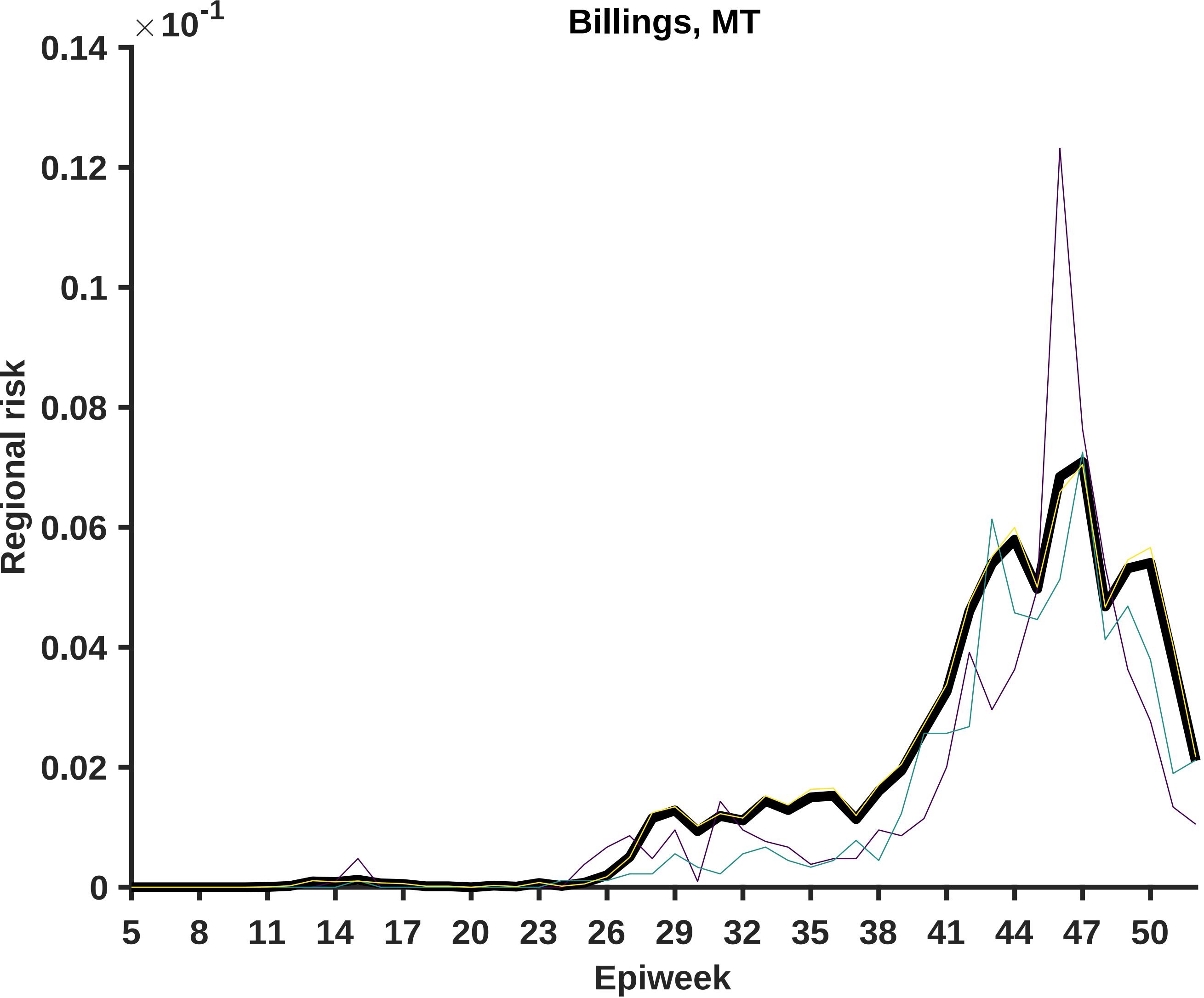

Billings, MT
×10-1
Regional risk
0.14
0.12
0.1
0.08
0.06
0.04
0.02
0
5
8
11
14
17
20
23
26
29
32
35
38
41
44
47
50
Epiweek

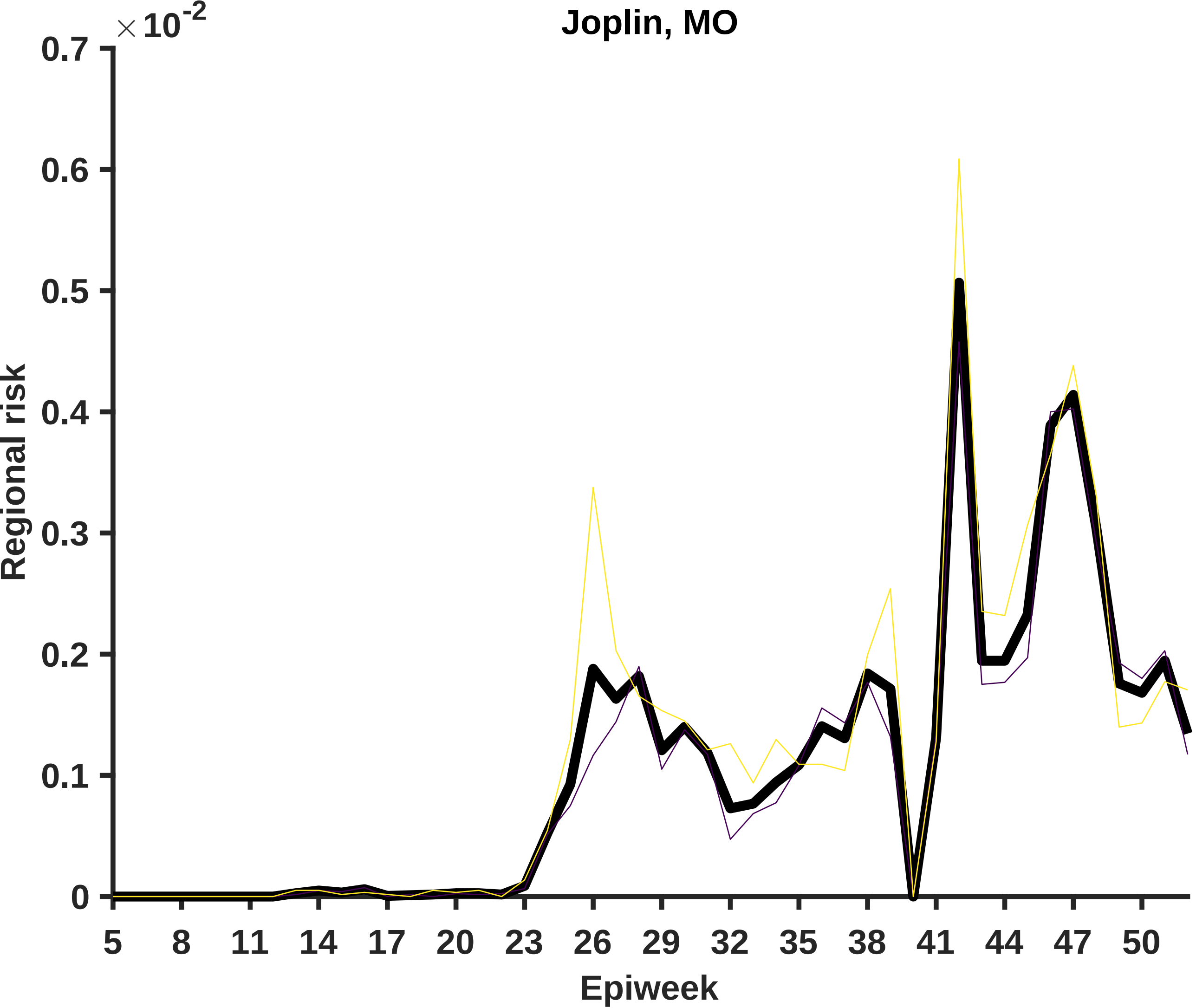

Joplin, MO
×10^-2
0.7
0.6
0.5
0.4
0.3
0.2
0.1
0
Regional risk
5
8
11
14
17
20
23
26
29
32
35
38
41
44
47
50
Epiweek

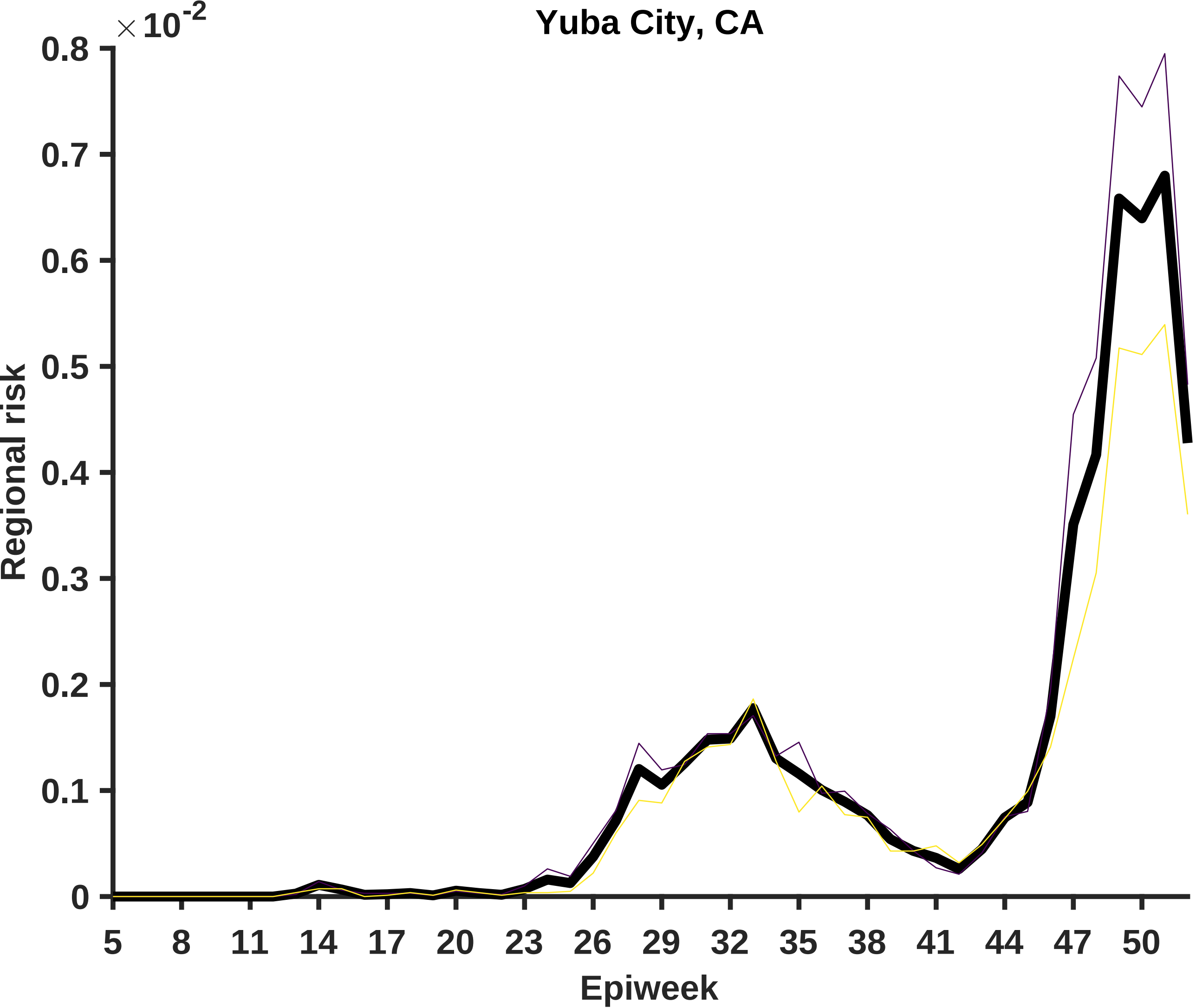

Yuba City, CA
×10-2
0.8
0.7
0.6
0.5
0.4
0.3
0.2
0.1
0
Regional risk
5
8
11
14
17
20
23
26
29
32
35
38
41
44
47
50
Epiweek

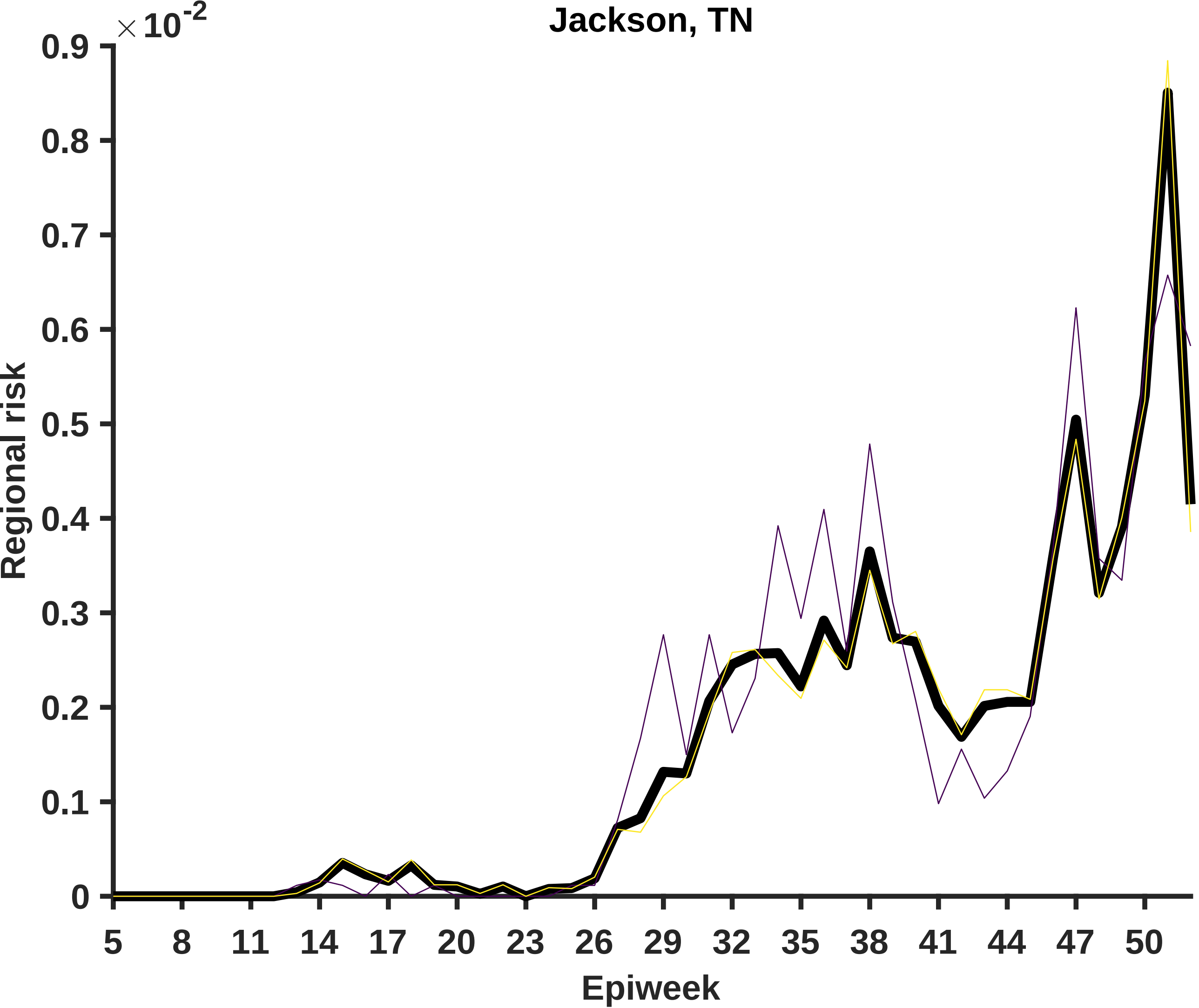

Jackson, TN
×10^-2
0.9
0.8
0.7
0.6
0.5
0.4
0.3
0.2
0.1
0
Regional risk
5
8
11
14
17
20
23
26
29
32
35
38
41
44
47
50
Epiweek

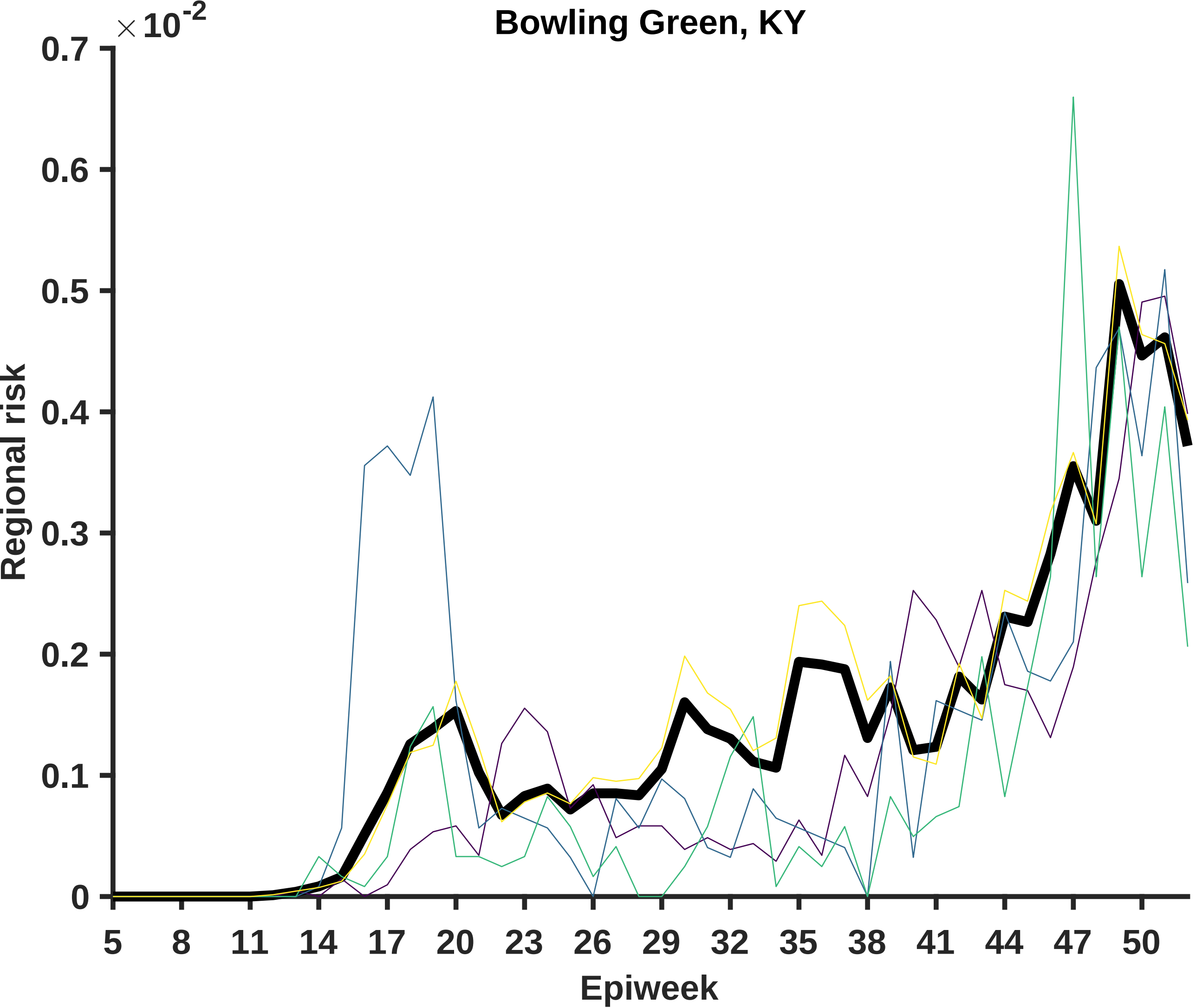
Bowling Green, KY
×10^-2
Regional risk
0.7
0.6
0.5
0.4
0.3
0.2
0.1
0
5
8
11
14
17
20
23
26
29
32
35
38
41
44
47
50
Epiweek

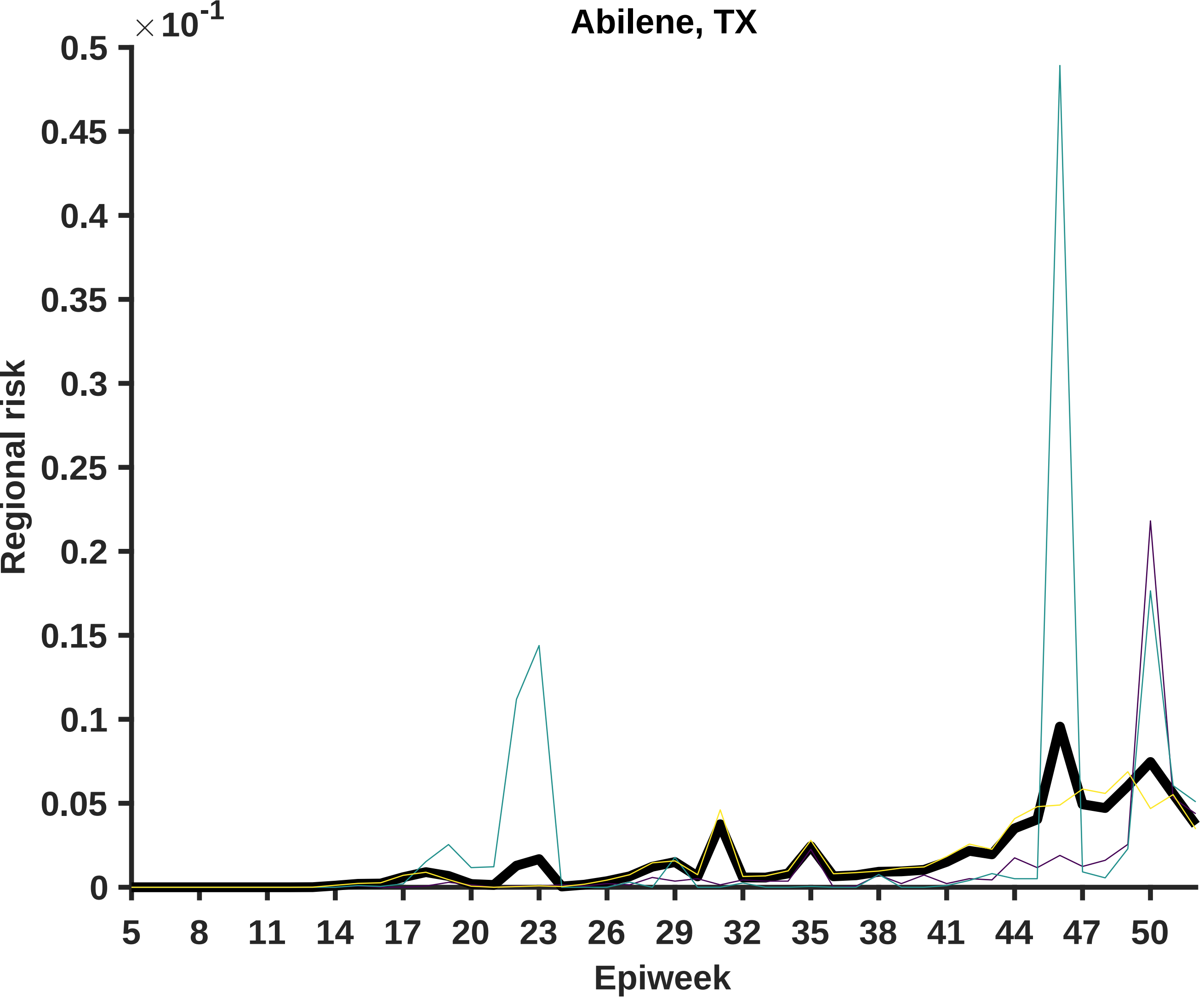

Abilene, TX
×10^-1
Regional risk
0.5
0.45
0.4
0.35
0.3
0.25
0.2
0.15
0.1
0.05
0
Epiweek
5
8
11
14
17
20
23
26
29
32
35
38
41
44
47
50

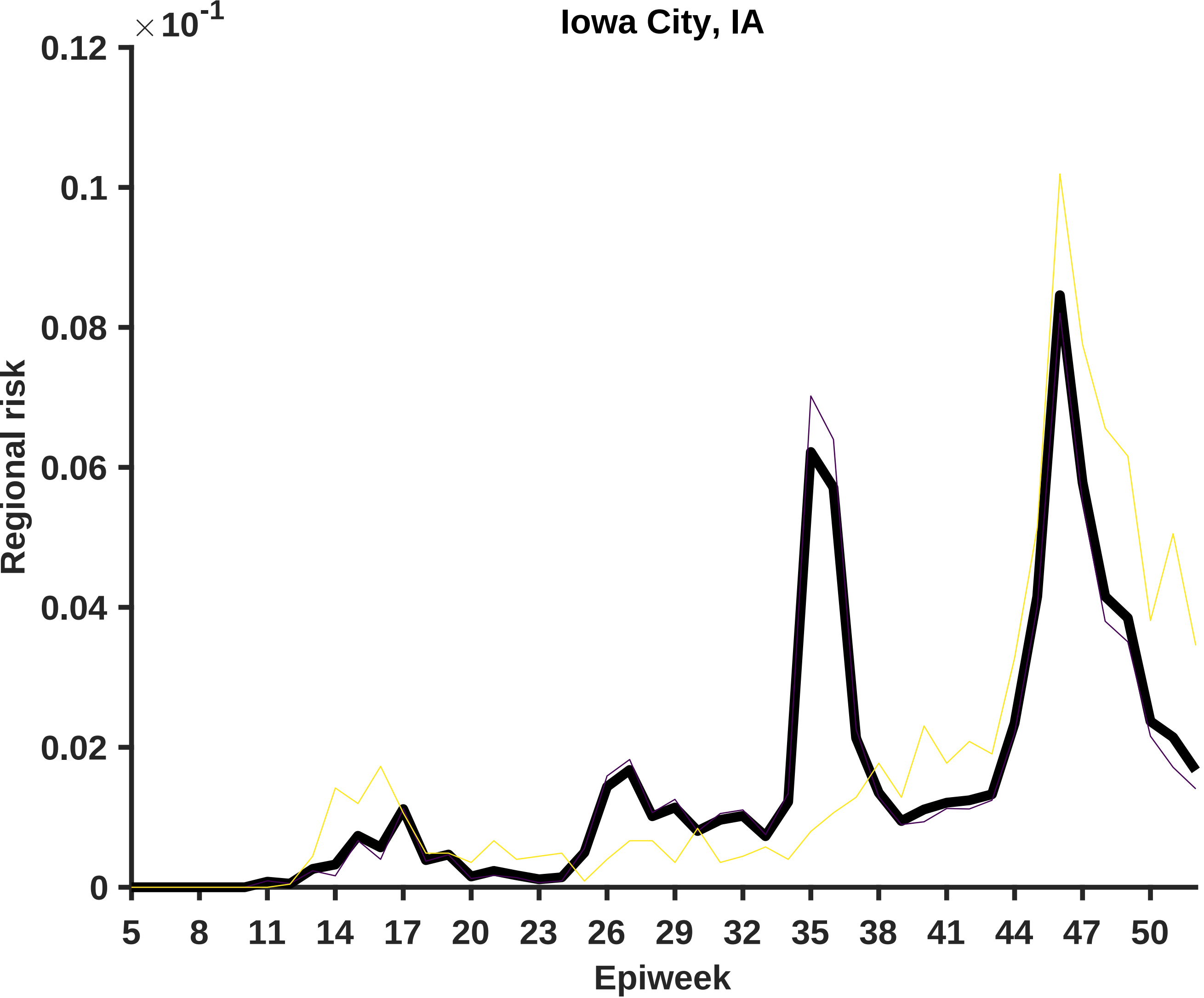

Iowa City, IA
×10^-1
0.12
0.1
0.08
0.06
0.04
0.02
0
Regional risk
5
8
11
14
17
20
23
26
29
32
35
38
41
44
47
50
Epiweek

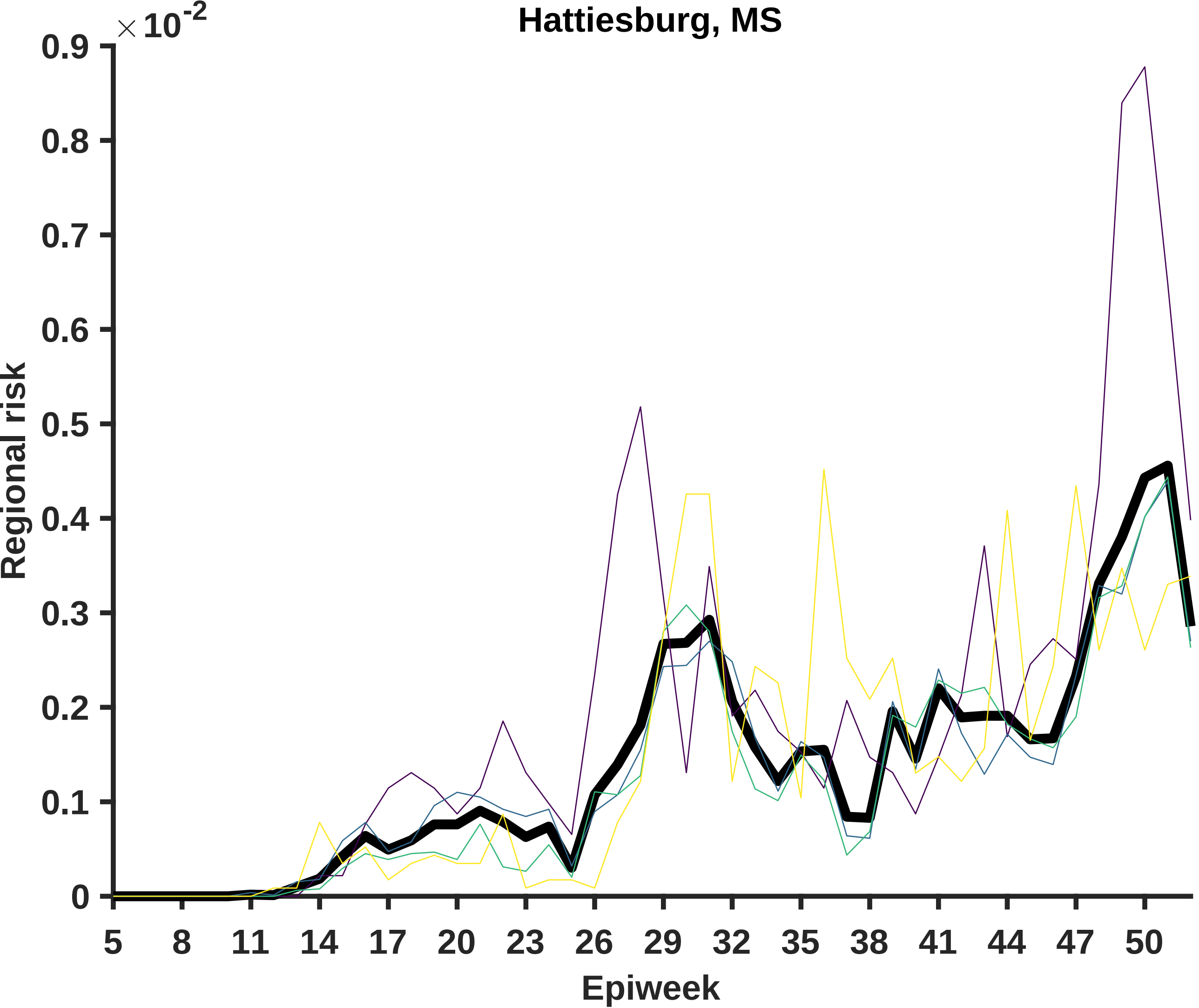
Hattiesburg, MS
$\times 10^{-2}$
0.9
0.8
0.7
0.6
0.5
0.4
0.3
0.2
0.1
0
Regional risk
5
8
11
14
17
20
23
26
29
32
35
38
41
44
47
50
Epiweek

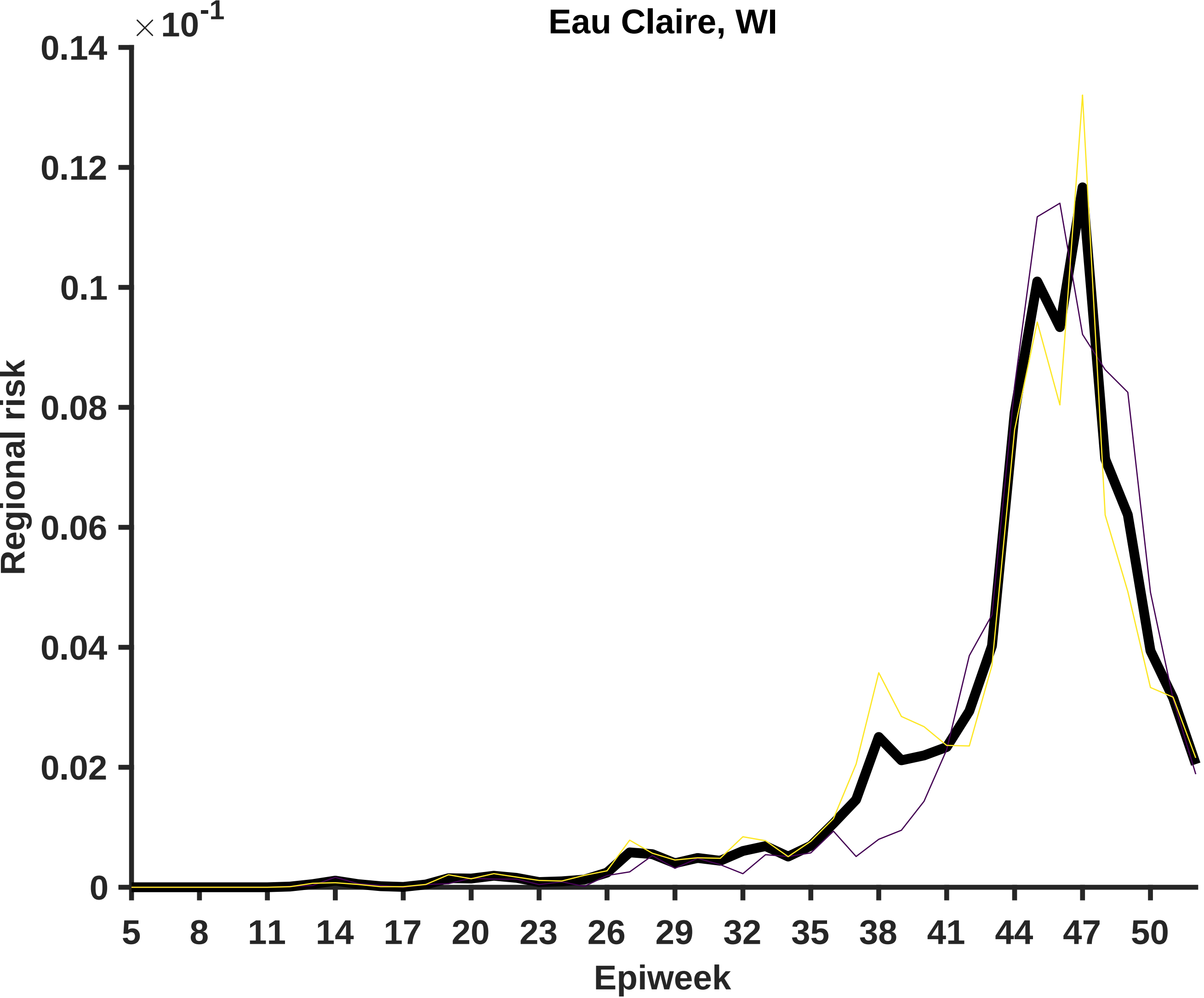
Eau Claire, WI
×10^-1
0.14
0.12
0.1
0.08
0.06
0.04
0.02
0
Regional risk
5
8
11
14
17
20
23
26
29
32
35
38
41
44
47
50
Epiweek

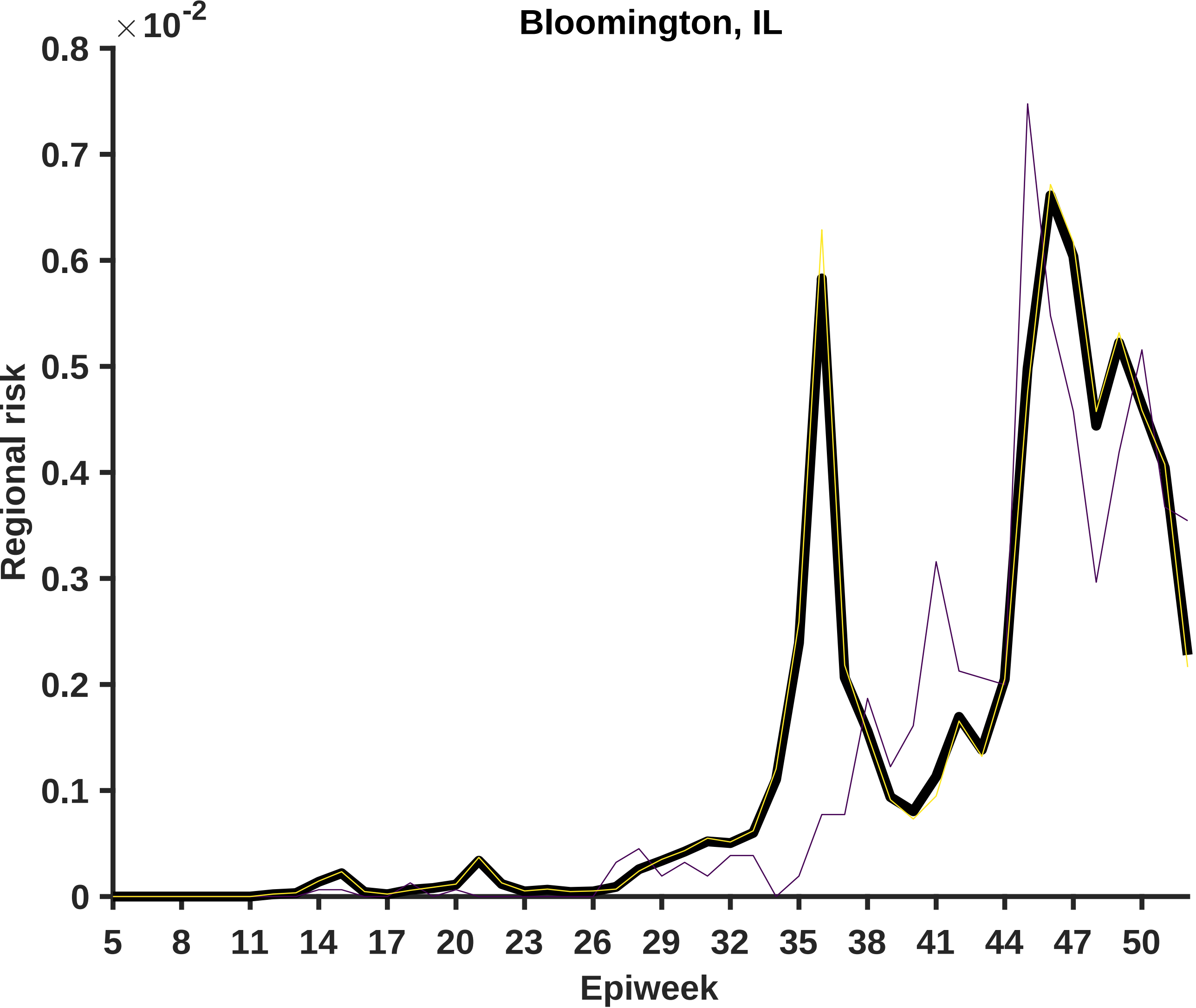

Bloomington, IL
×10^-2
Regional risk
0
0.1
0.2
0.3
0.4
0.5
0.6
0.7
0.8
5
8
11
14
17
20
23
26
29
32
35
38
41
44
47
50
Epiweek

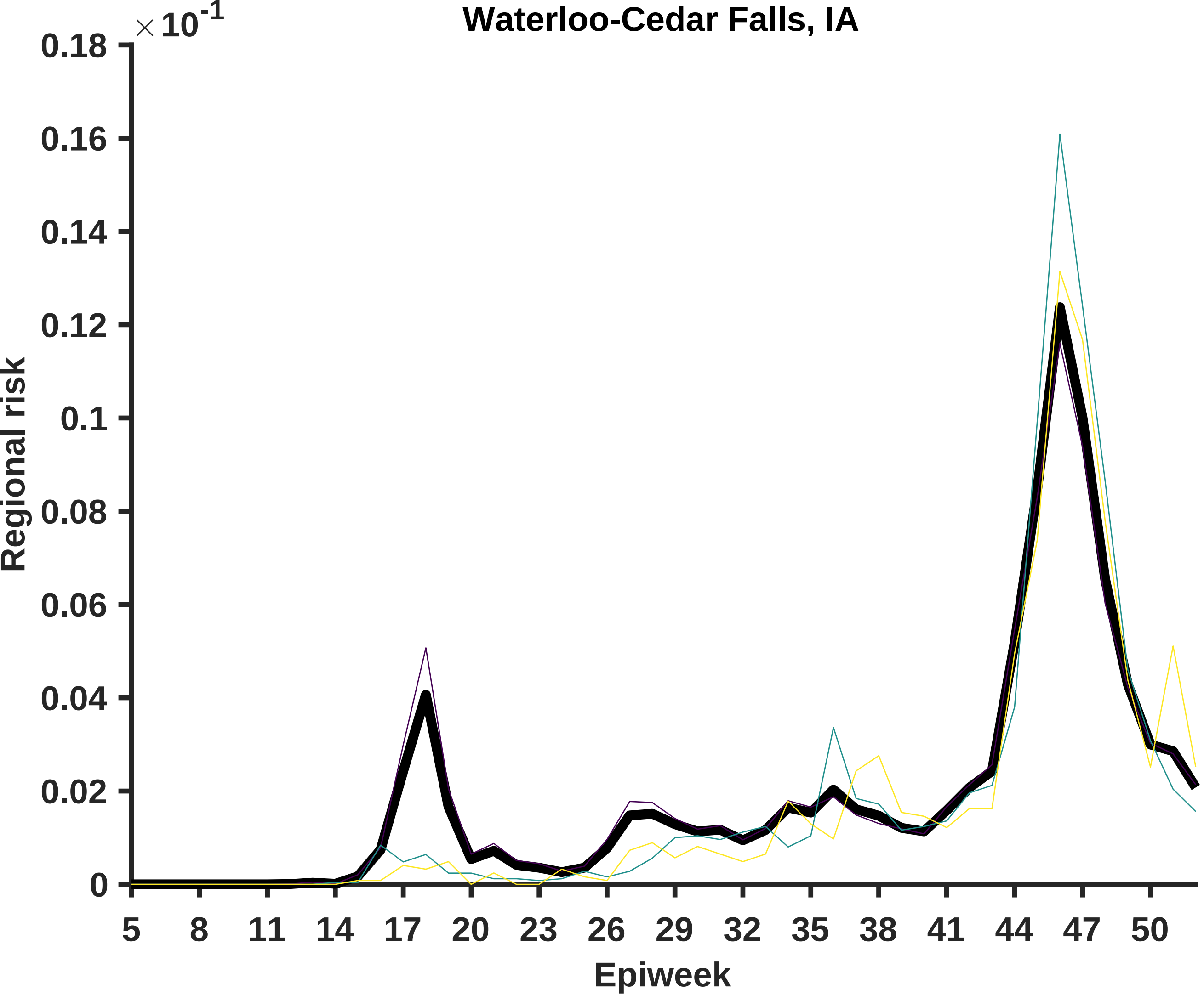

Waterloo-Cedar Falls, IA
$\times 10^{-1}$
0.18
0.16
0.14
0.12
0.1
0.08
0.06
0.04
0.02
0
Regional risk
5
8
11
14
17
20
23
26
29
32
35
38
41
44
47
50
Epiweek

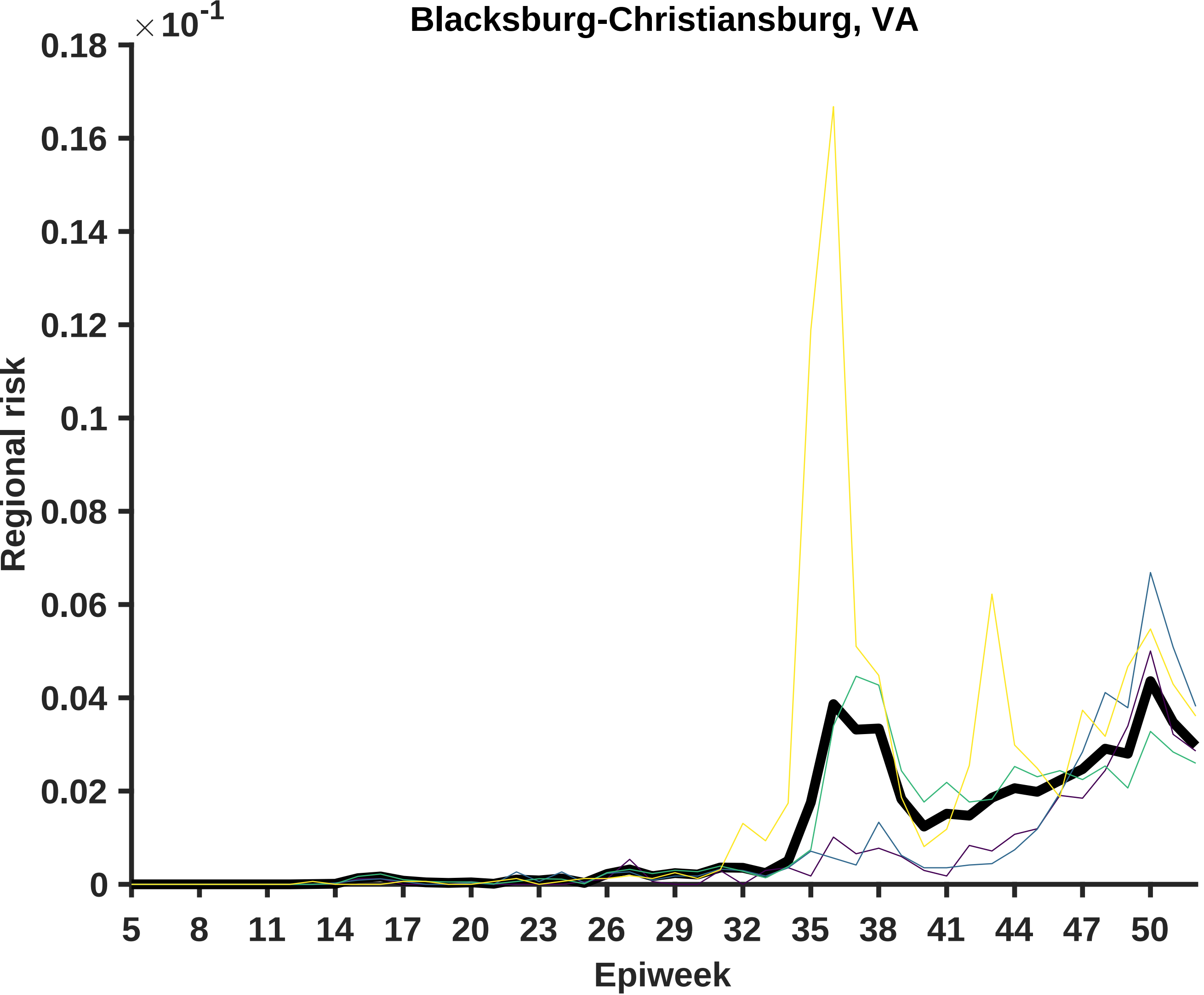

Blacksburg-Christiansburg, VA
× 10^-1
0.18
0.16
0.14
0.12
0.1
0.08
0.06
0.04
0.02
0
Regional risk
5
8
11
14
17
20
23
26
29
32
35
38
41
44
47
50
Epiweek

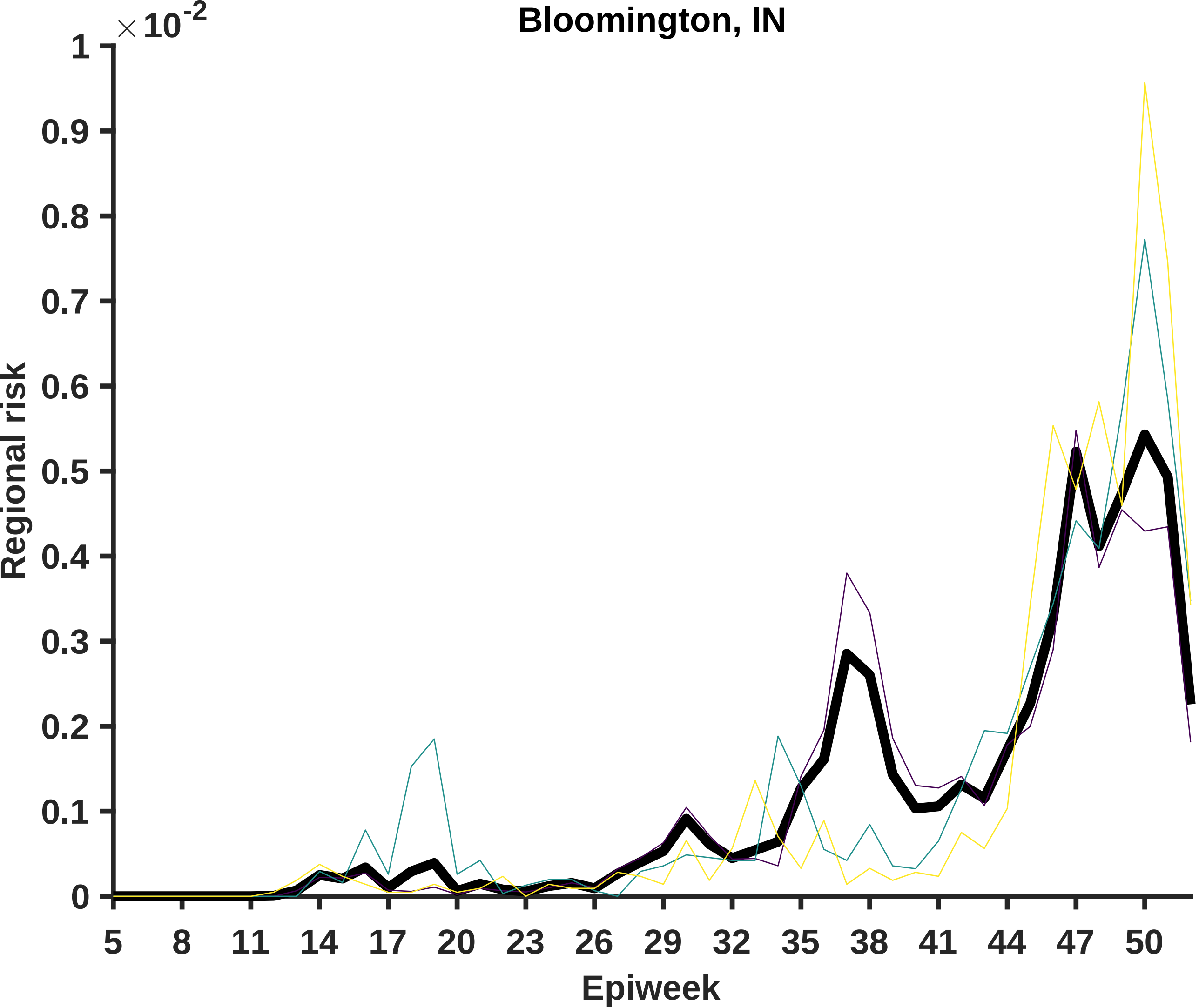

Bloomington, IN
×10^-2
Regional risk
Epiweek
1
0.9
0.8
0.7
0.6
0.5
0.4
0.3
0.2
0.1
0
5
8
11
14
17
20
23
26
29
32
35
38
41
44
47
50

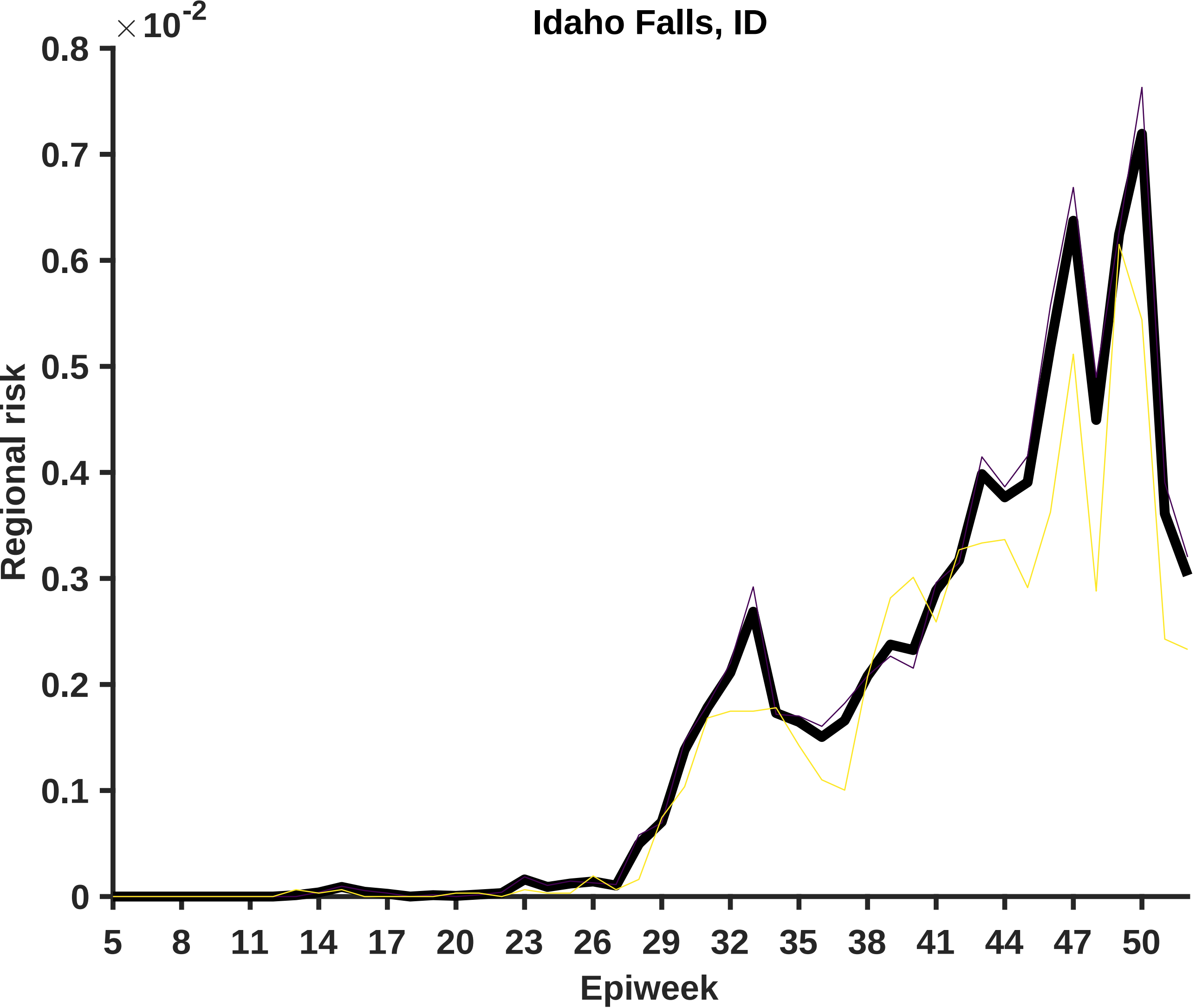

Idaho Falls, ID
×10^-2
Regional risk
Epiweek
0
0.1
0.2
0.3
0.4
0.5
0.6
0.7
0.8
5
8
11
14
17
20
23
26
29
32
35
38
41
44
47
50

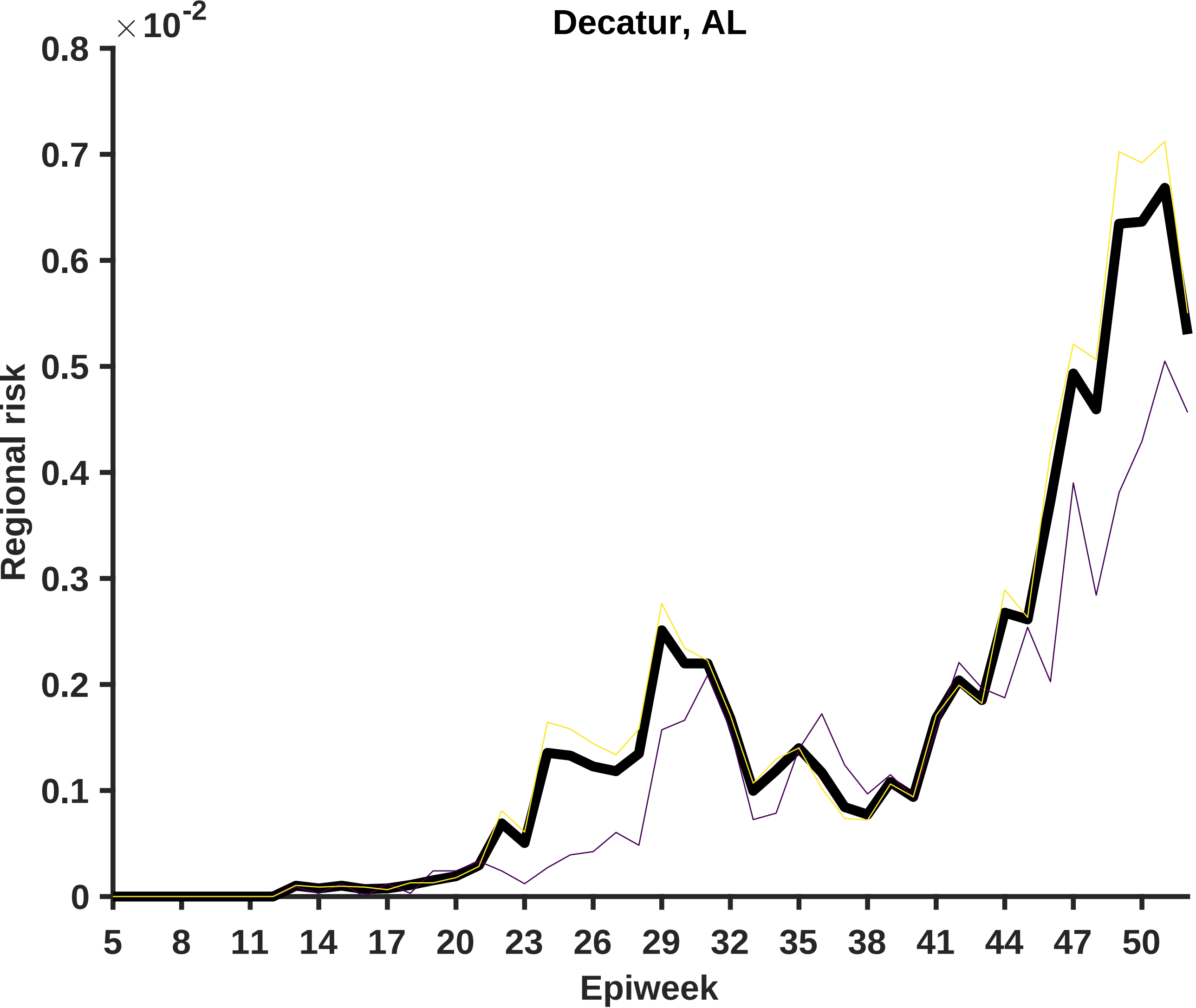

Decatur, AL
×10^-2
0.8
0.7
0.6
0.5
0.4
0.3
0.2
0.1
0
Regional risk
5
8
11
14
17
20
23
26
29
32
35
38
41
44
47
50
Epiweek

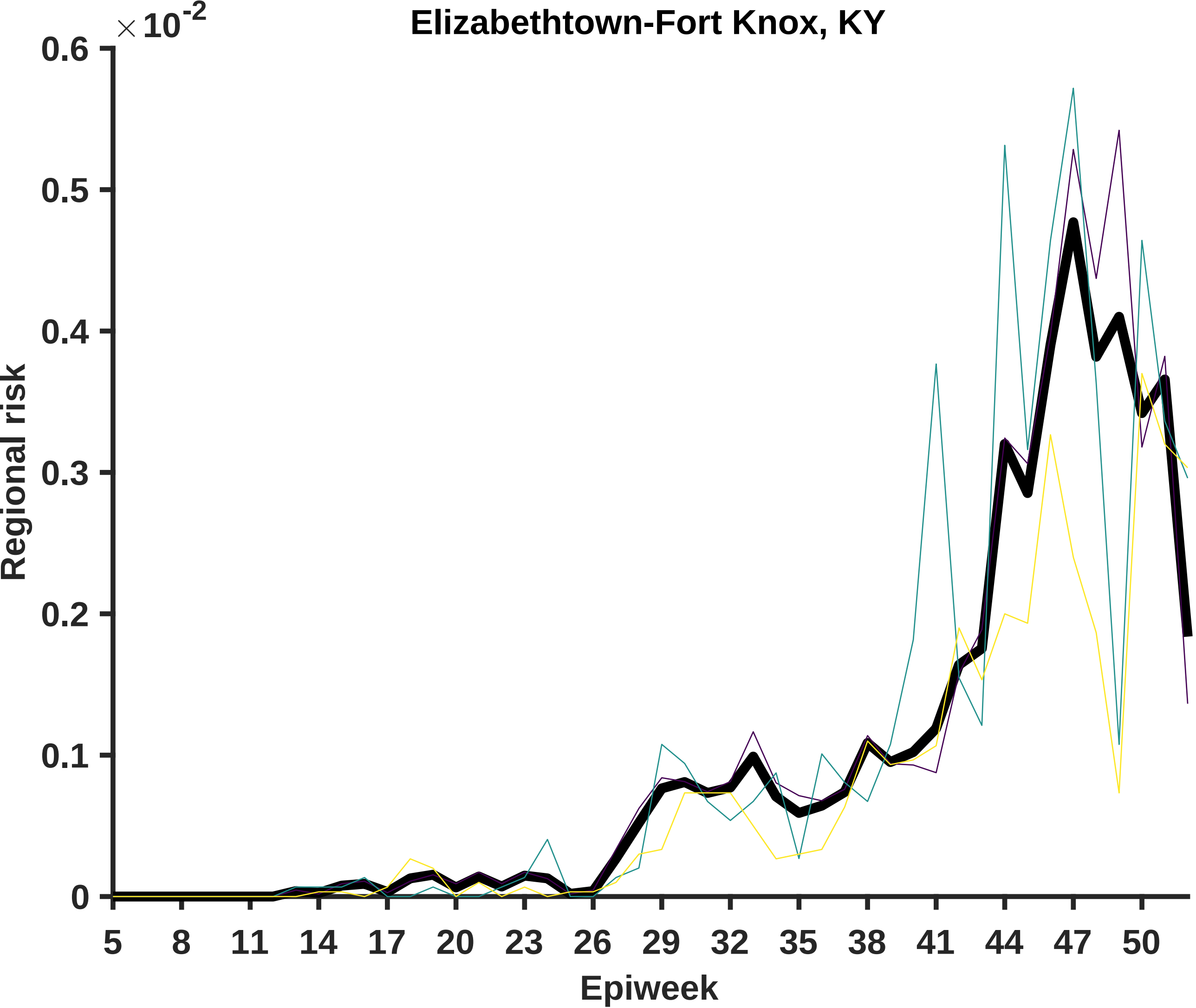

Elizabethtown-Fort Knox, KY
×10^-2
0.6
0.5
0.4
0.3
0.2
0.1
0
Regional risk
5
8
11
14
17
20
23
26
29
32
35
38
41
44
47
50
Epiweek

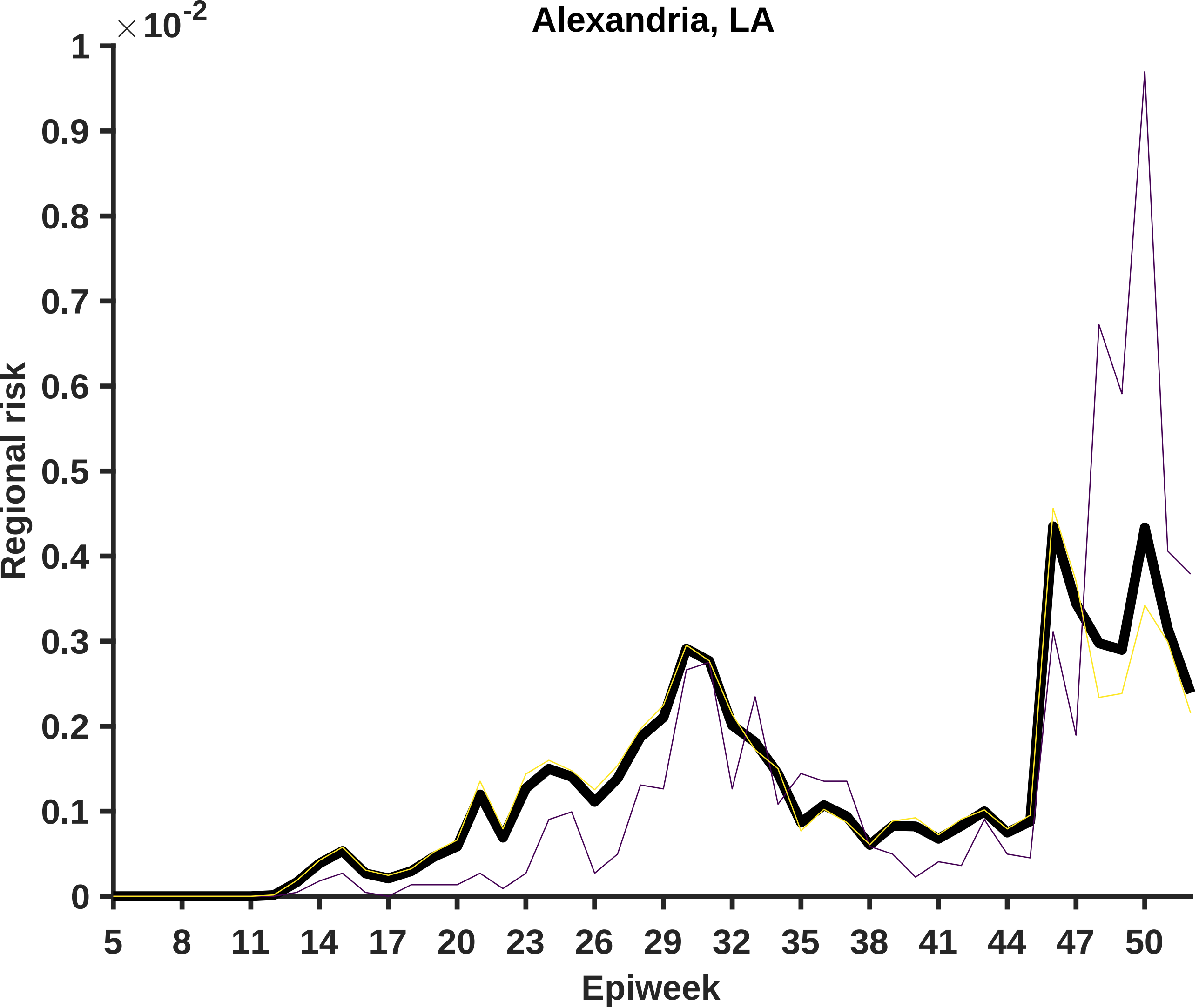

Alexandria, LA
×10^-2
1
0.9
0.8
0.7
0.6
0.5
0.4
0.3
0.2
0.1
0
Regional risk
5
8
11
14
17
20
23
26
29
32
35
38
41
44
47
50
Epiweek

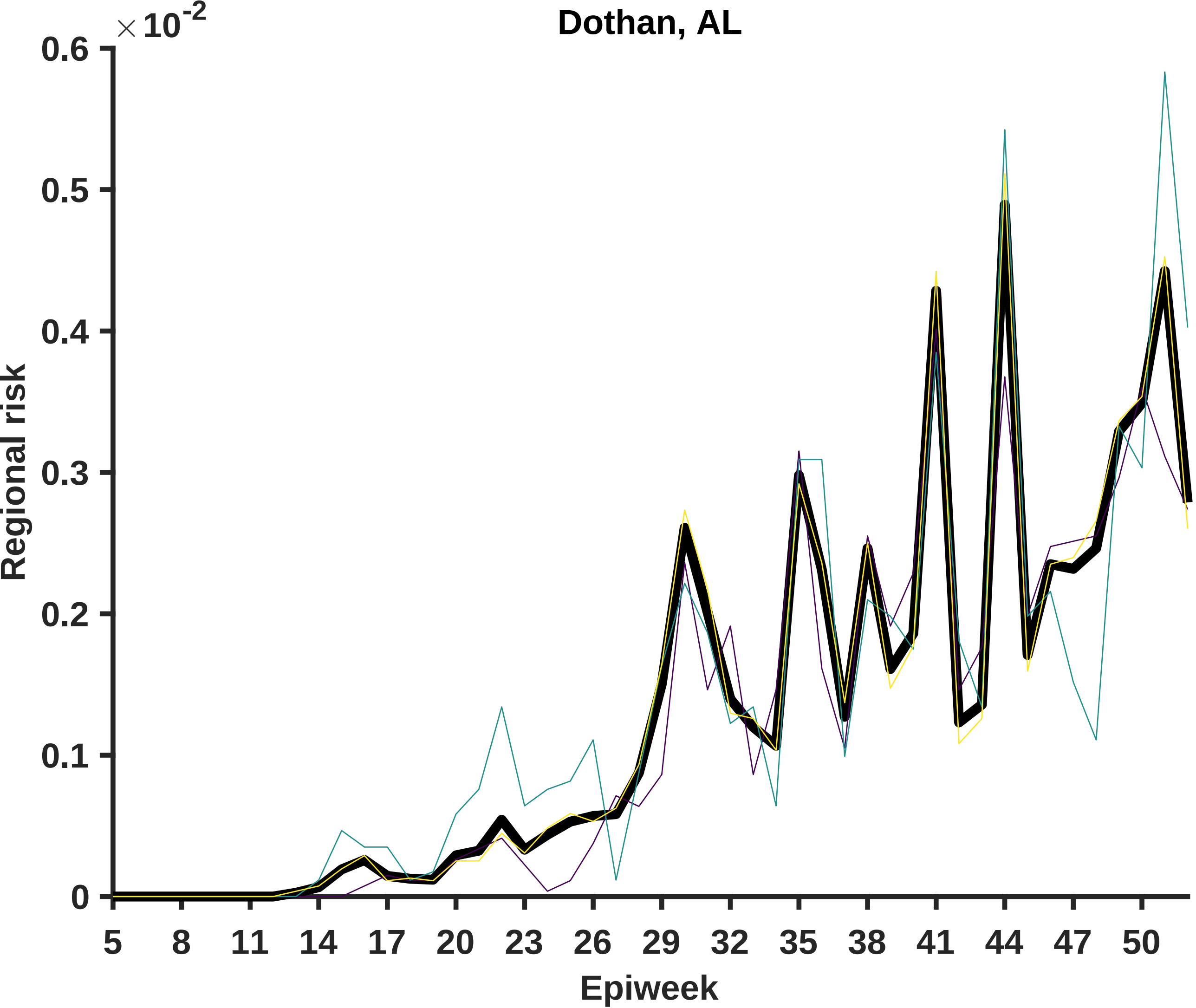

Dothan, AL
×10^-2
0.6
0.5
0.4
0.3
0.2
0.1
0
Regional risk
5
8
11
14
17
20
23
26
29
32
35
38
41
44
47
50
Epiweek

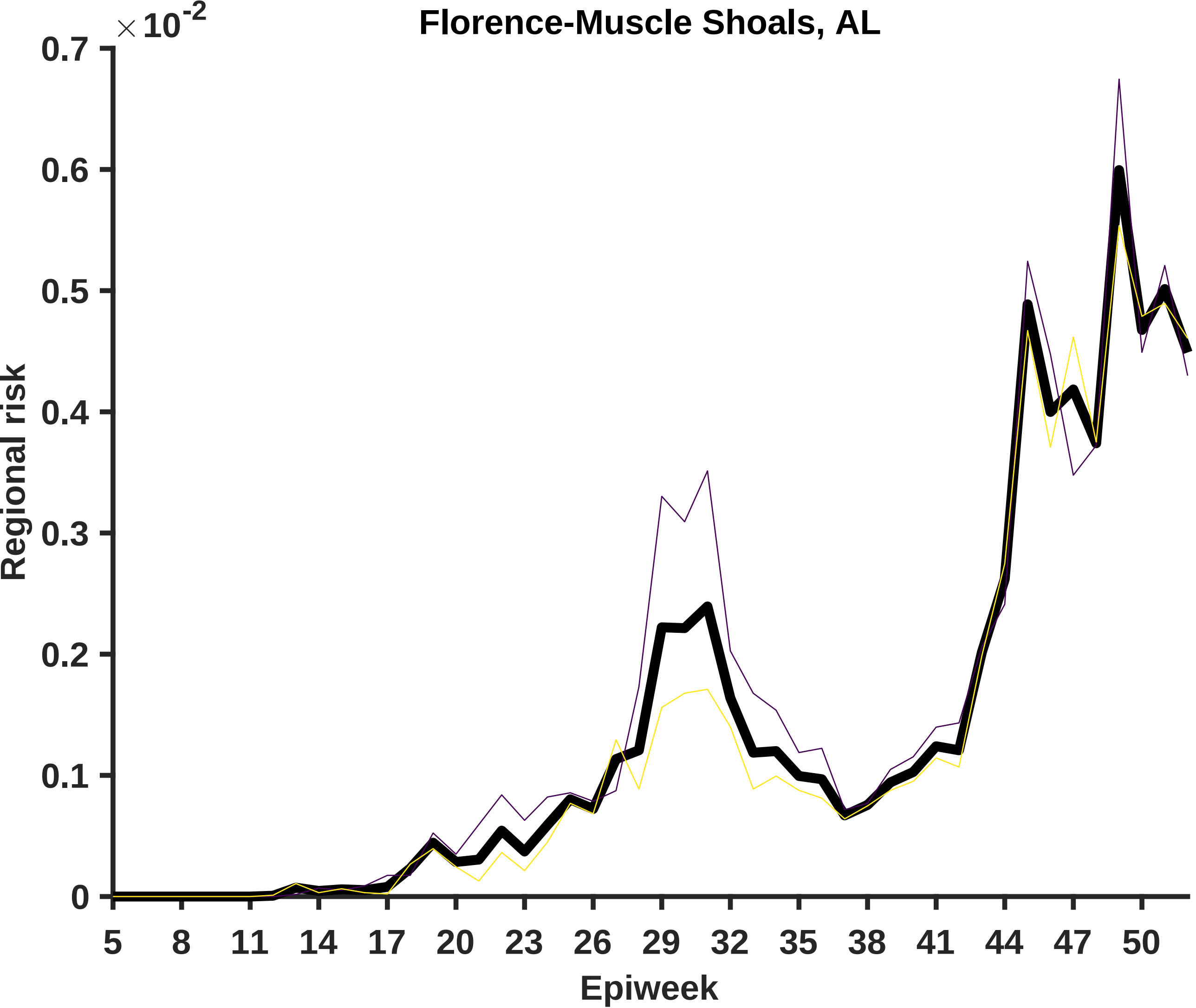

Florence-Muscle Shoals, AL
×10^-2
0.7
0.6
0.5
0.4
0.3
0.2
0.1
0
Regional risk
5
8
11
14
17
20
23
26
29
32
35
38
41
44
47
50
Epiweek

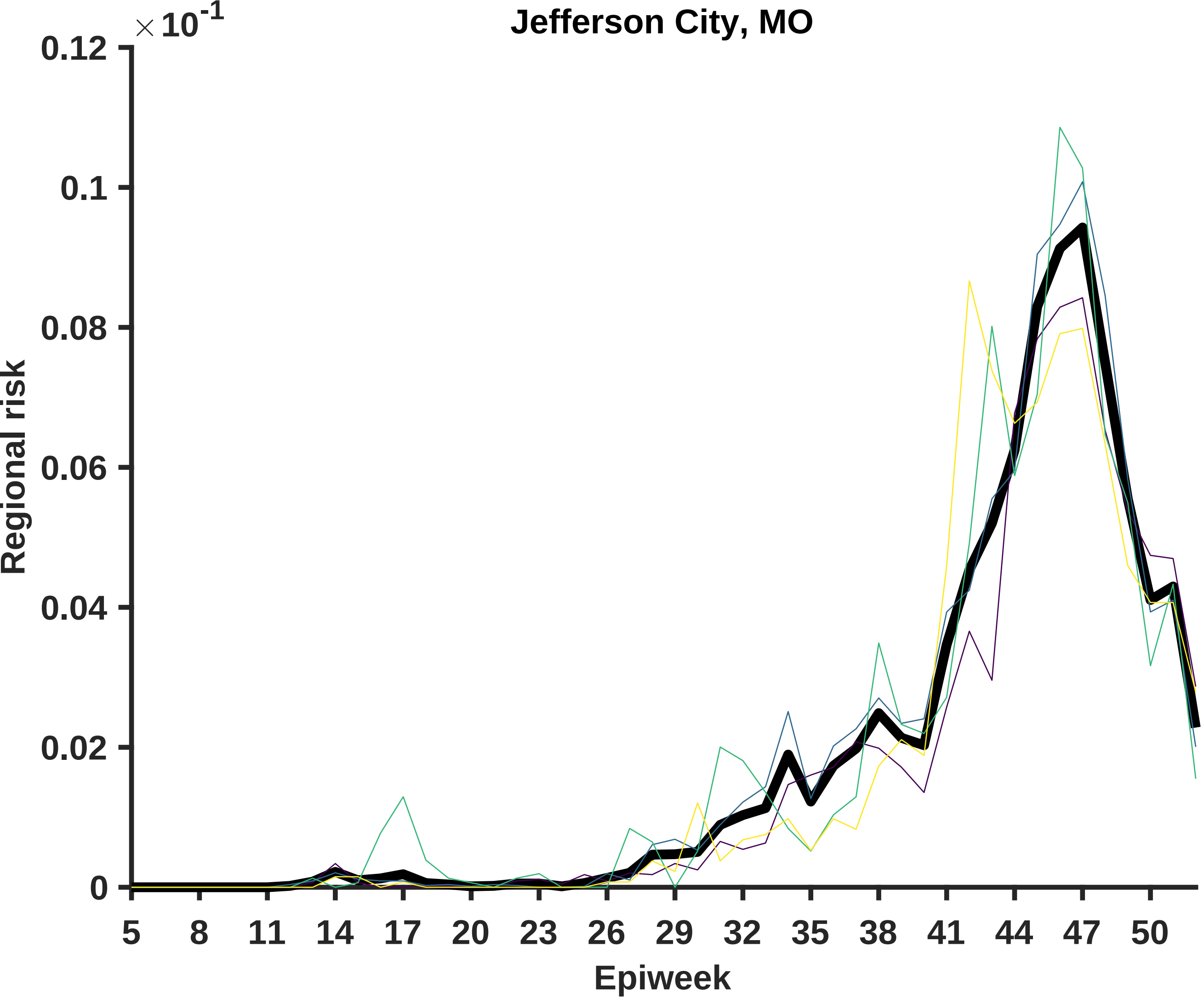

Jefferson City, MO
×10^-1
0.12
0.1
0.08
0.06
0.04
0.02
0
Regional risk
5
8
11
14
17
20
23
26
29
32
35
38
41
44
47
50
Epiweek

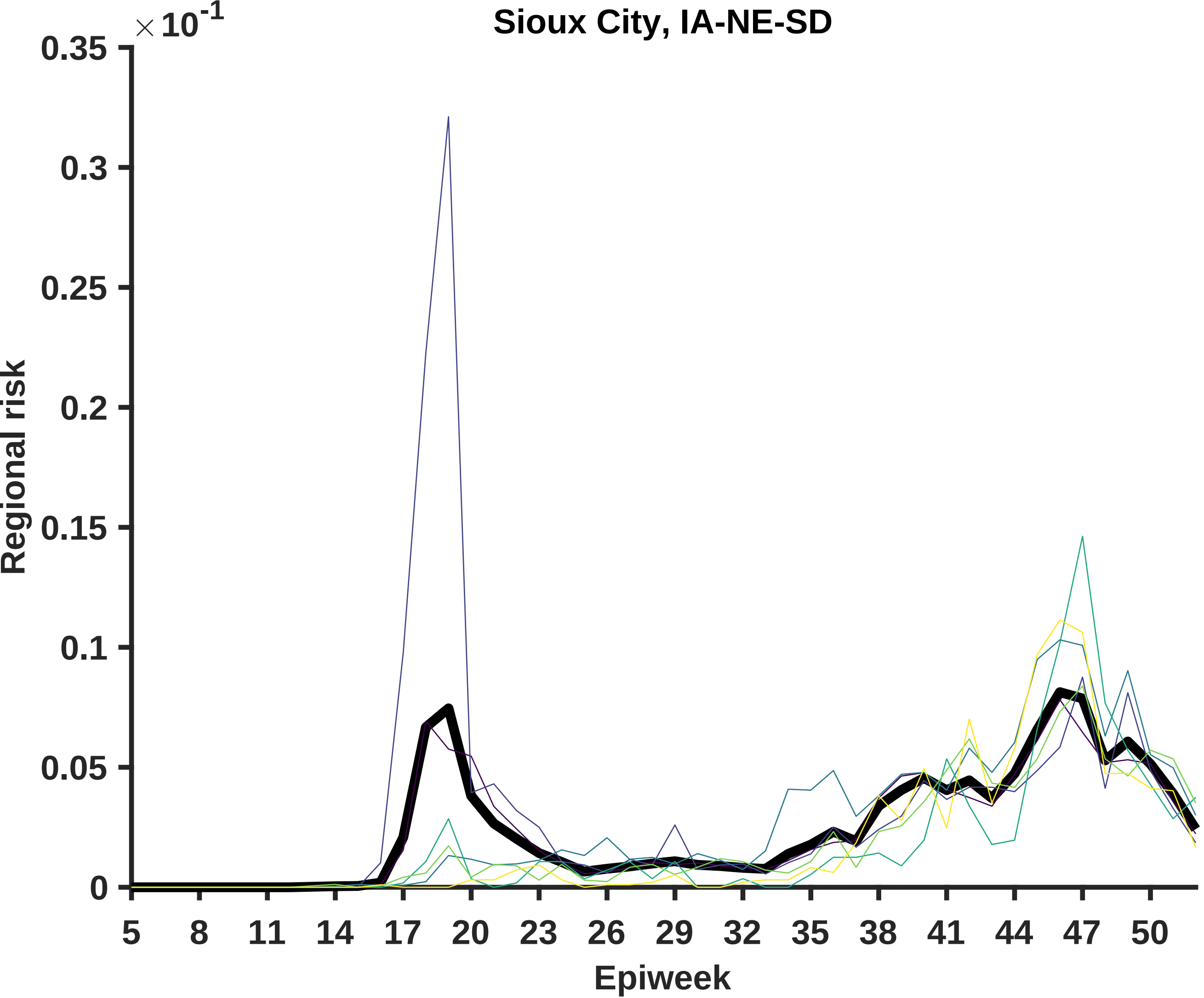

Sioux City, IA-NE-SD
×10^-1
0.35
0.3
0.25
0.2
0.15
0.1
0.05
0
Regional risk
5
8
11
14
17
20
23
26
29
32
35
38
41
44
47
50
Epiweek

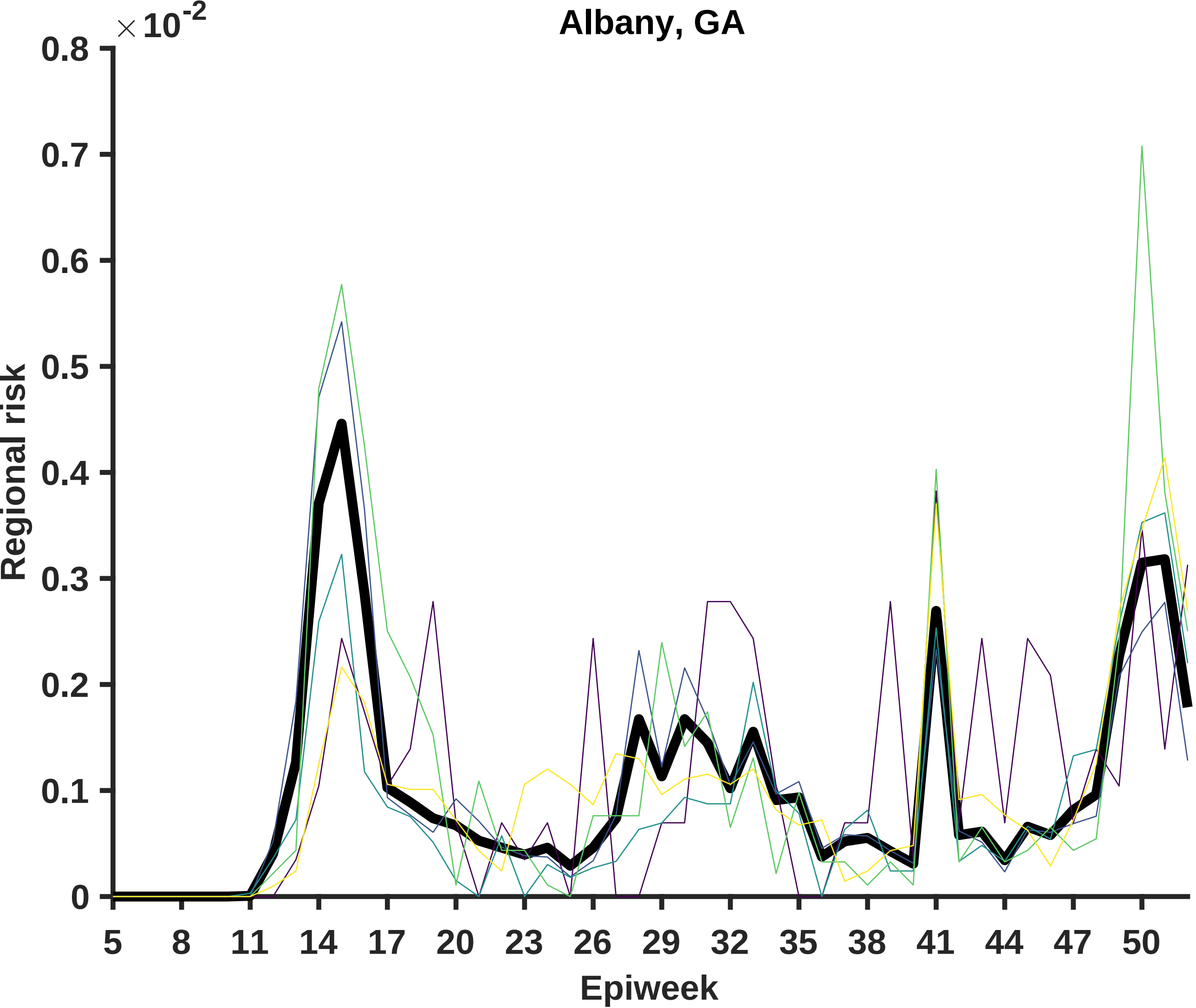

Albany, GA
×10^-2
0.8
0.7
0.6
0.5
0.4
0.3
0.2
0.1
0
Regional risk
5
8
11
14
17
20
23
26
29
32
35
38
41
44
47
50
Epiweek

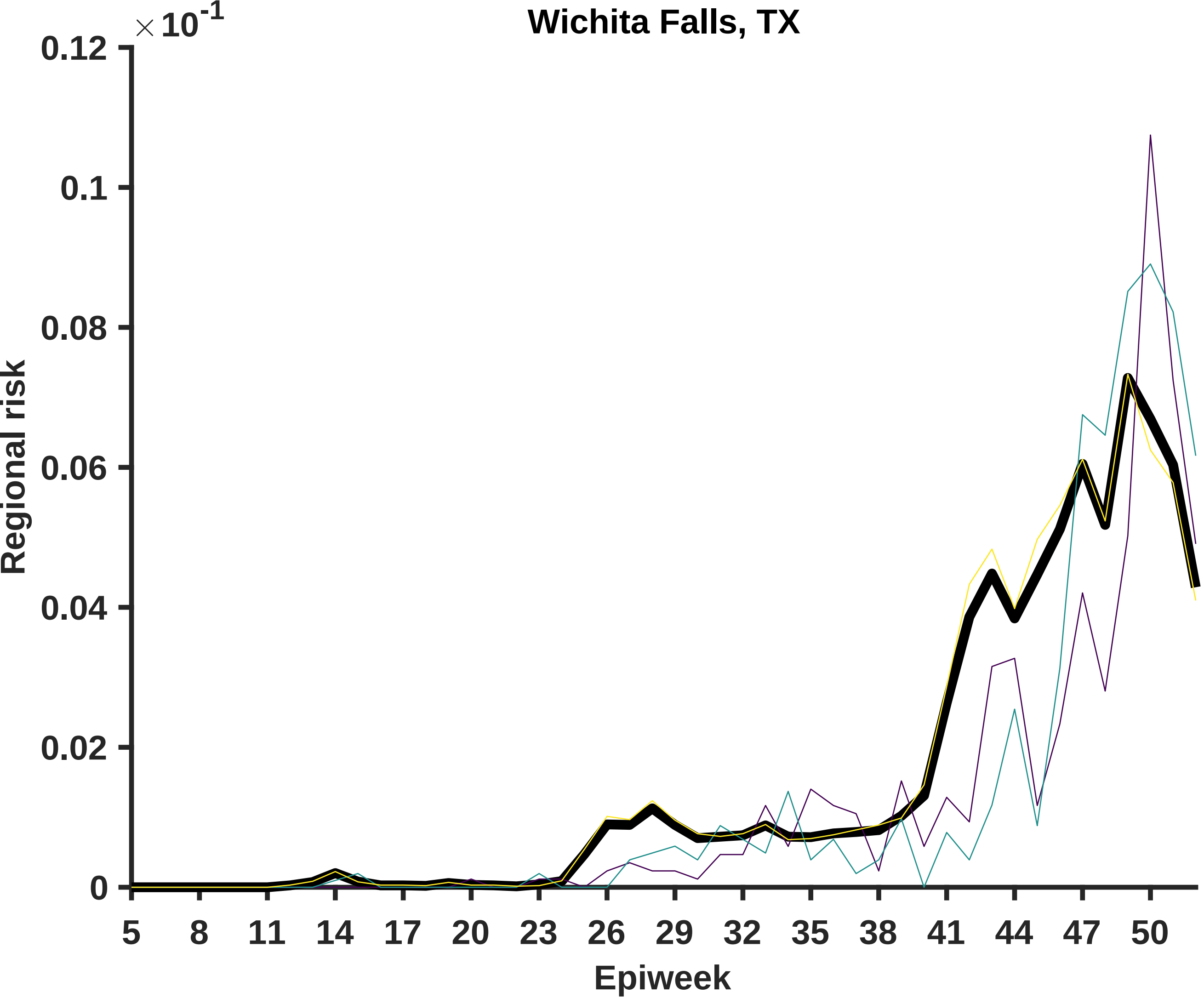

Wichita Falls, TX
×10^-1
0.12
0.1
0.08
0.06
0.04
0.02
0
Regional risk
5
8
11
14
17
20
23
26
29
32
35
38
41
44
47
50
Epiweek

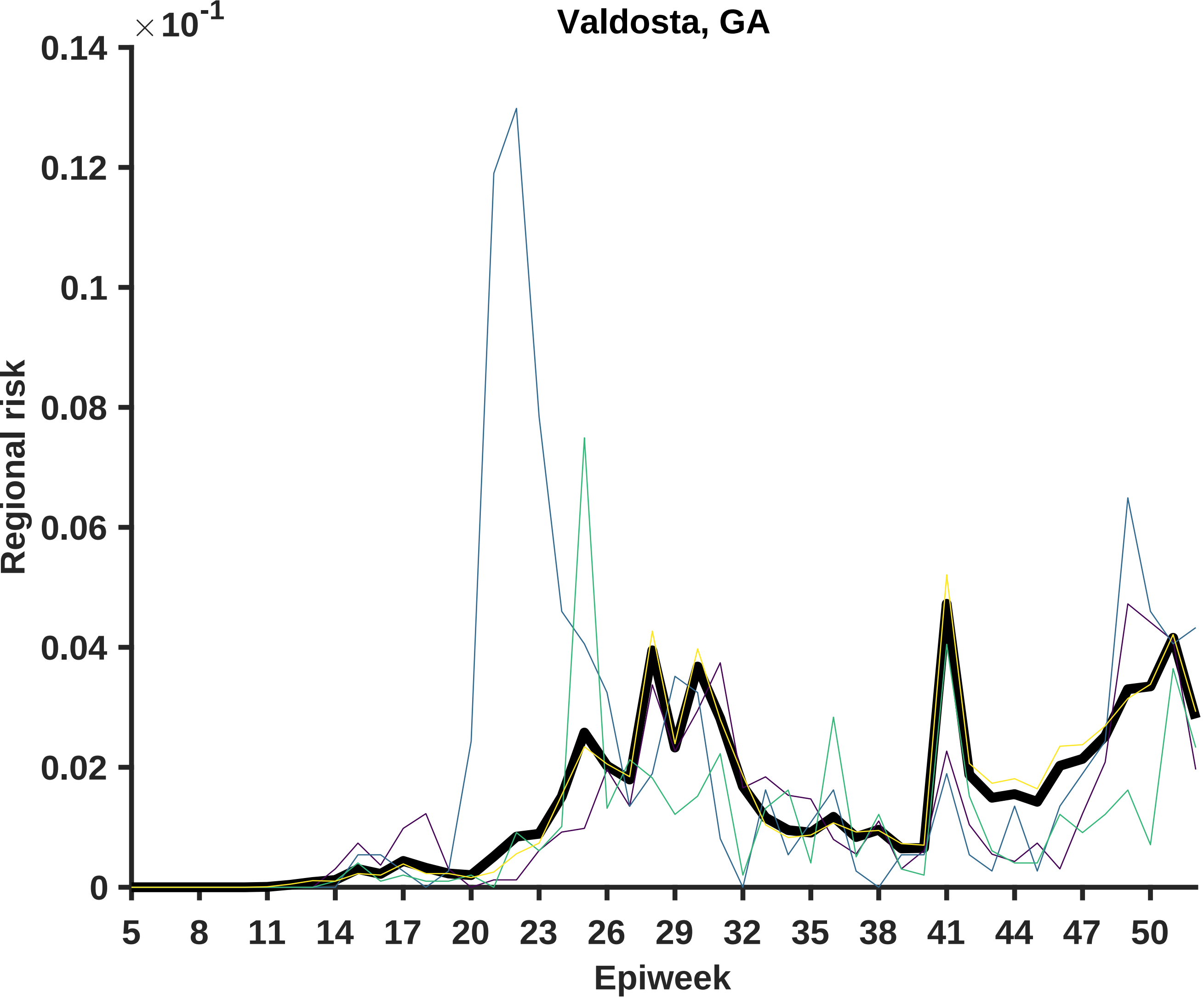

Valdosta, GA
×10^-1
0.14
0.12
0.1
0.08
0.06
0.04
0.02
0
Regional risk
5
8
11
14
17
20
23
26
29
32
35
38
41
44
47
50
Epiweek

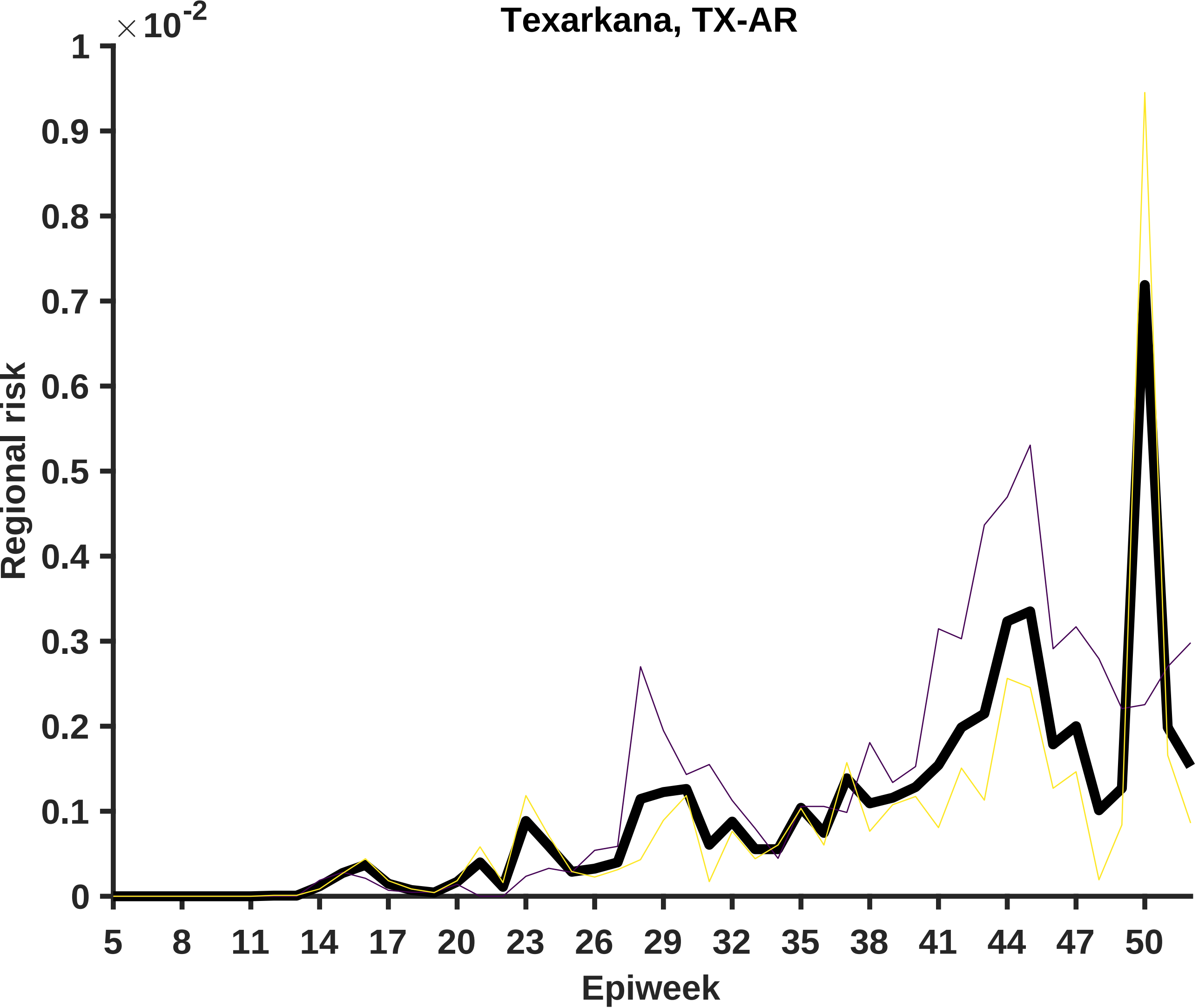

Texarkana, TX-AR
×10^-2
Regional risk
0
0.1
0.2
0.3
0.4
0.5
0.6
0.7
0.8
0.9
1
5
8
11
14
17
20
23
26
29
32
35
38
41
44
47
50
Epiweek

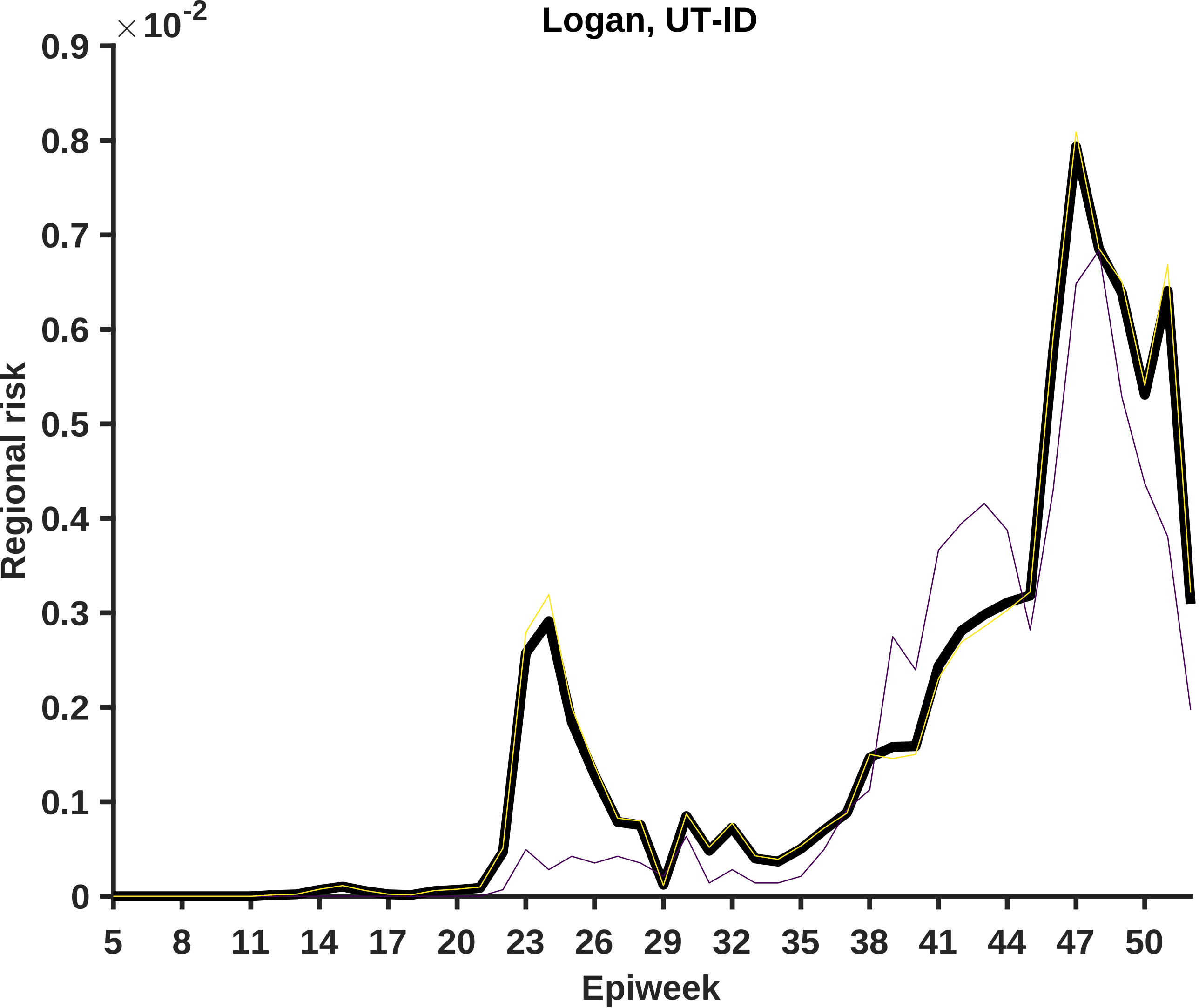

Logan, UT-ID
×10^-2
0.9
0.8
0.7
0.6
0.5
0.4
0.3
0.2
0.1
0
Regional risk
5
8
11
14
17
20
23
26
29
32
35
38
41
44
47
50
Epiweek

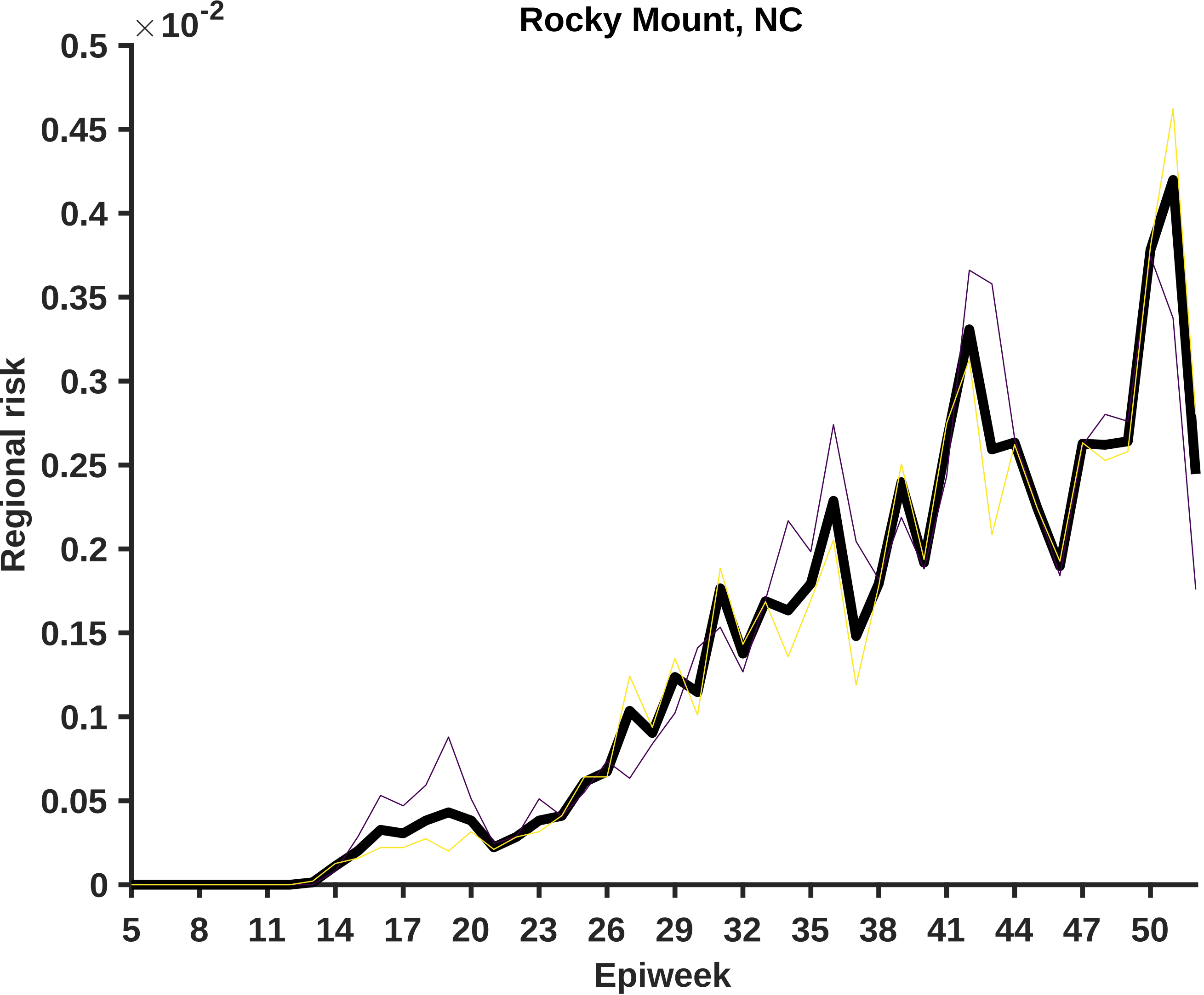
Rocky Mount, NC
×10^-2
0.5
0.45
0.4
0.35
0.3
0.25
0.2
0.15
0.1
0.05
0
Regional risk
5
8
11
14
17
20
23
26
29
32
35
38
41
44
47
50
Epiweek

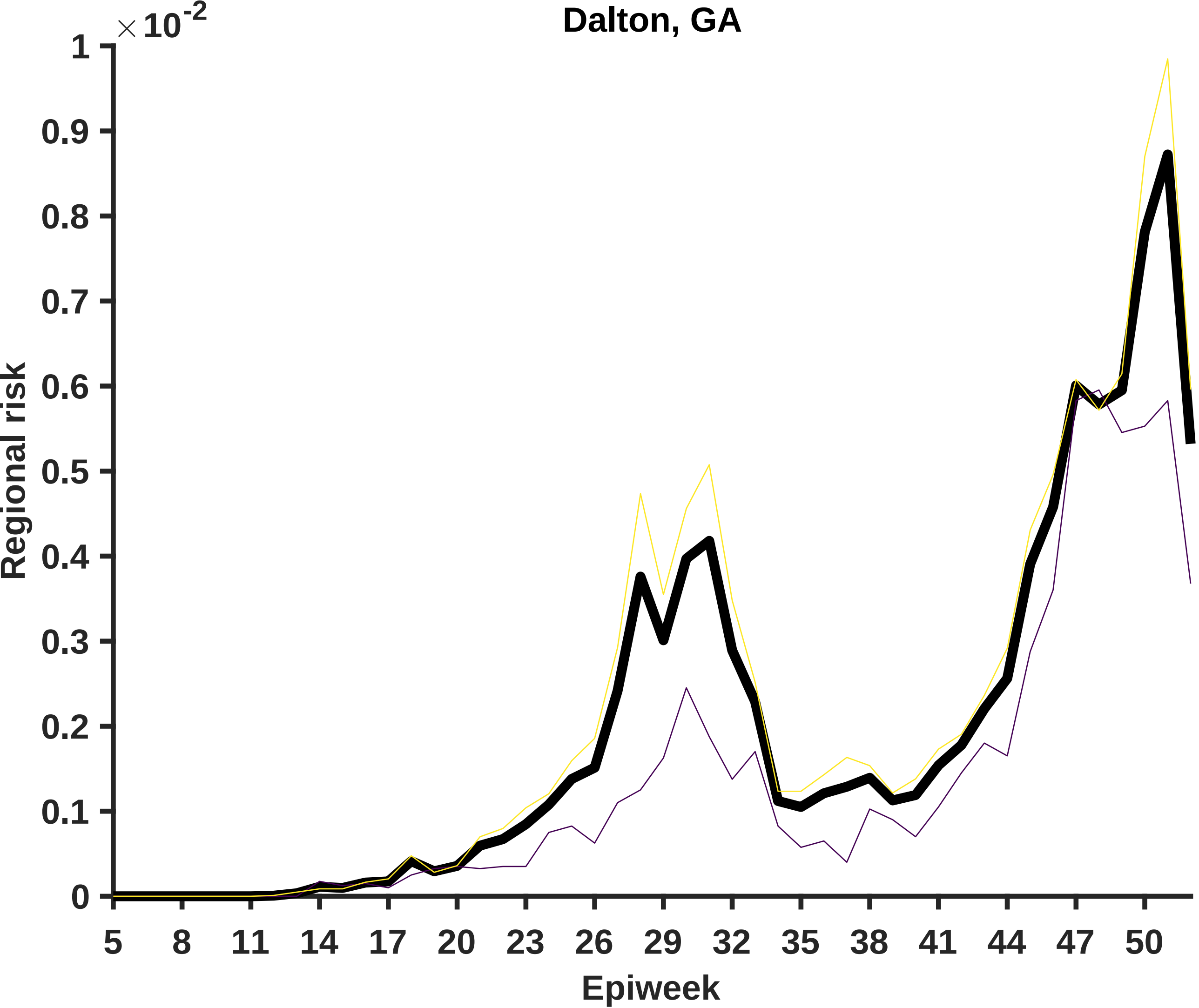

Dalton, GA
×10^-2
Regional risk
Epiweek
1
0.9
0.8
0.7
0.6
0.5
0.4
0.3
0.2
0.1
0
5
8
11
14
17
20
23
26
29
32
35
38
41
44
47
50

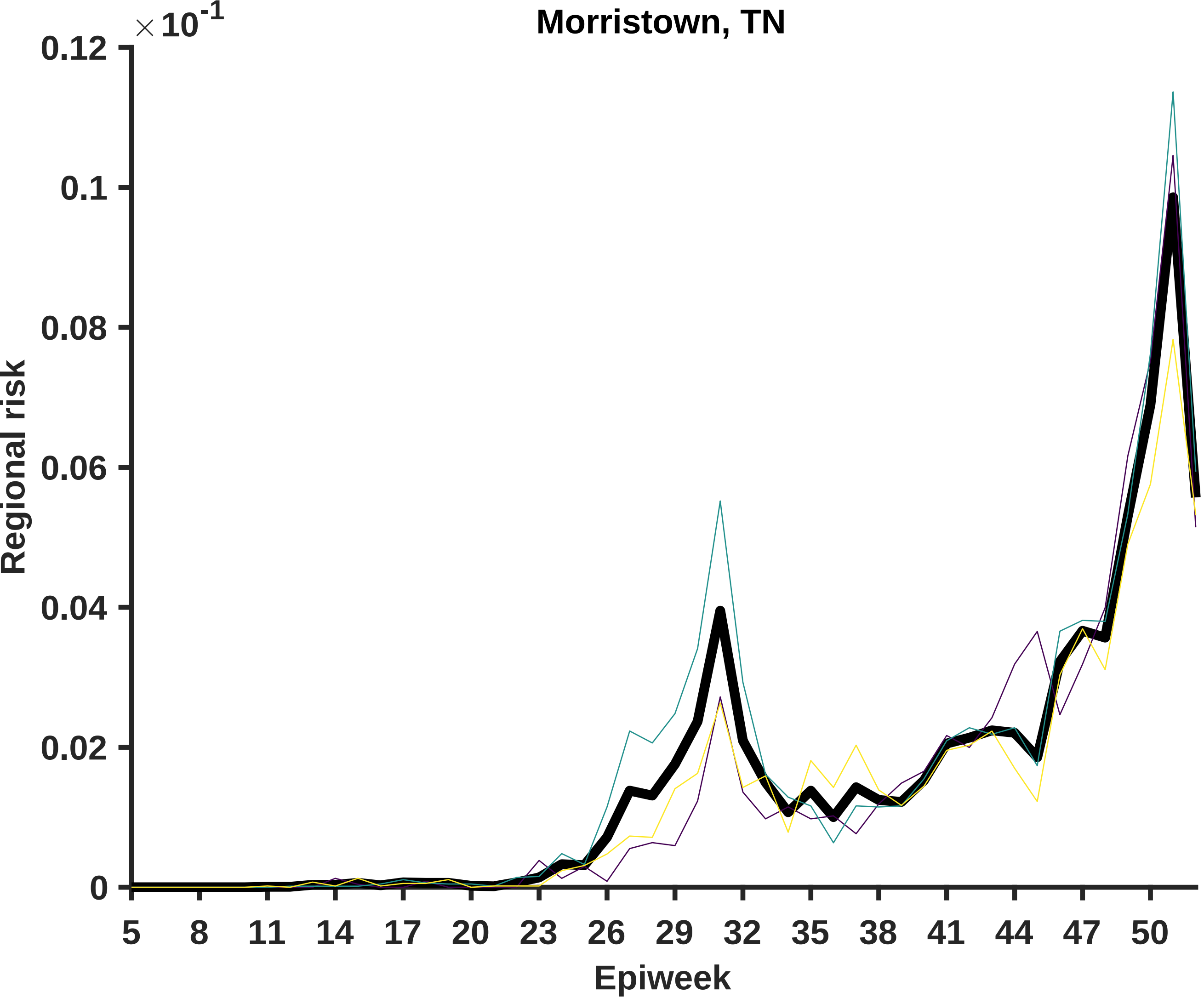

Morristown, TN
× 10^-1
0.12
0.1
0.08
0.06
0.04
0.02
0
Regional risk
5
8
11
14
17
20
23
26
29
32
35
38
41
44
47
50
Epiweek

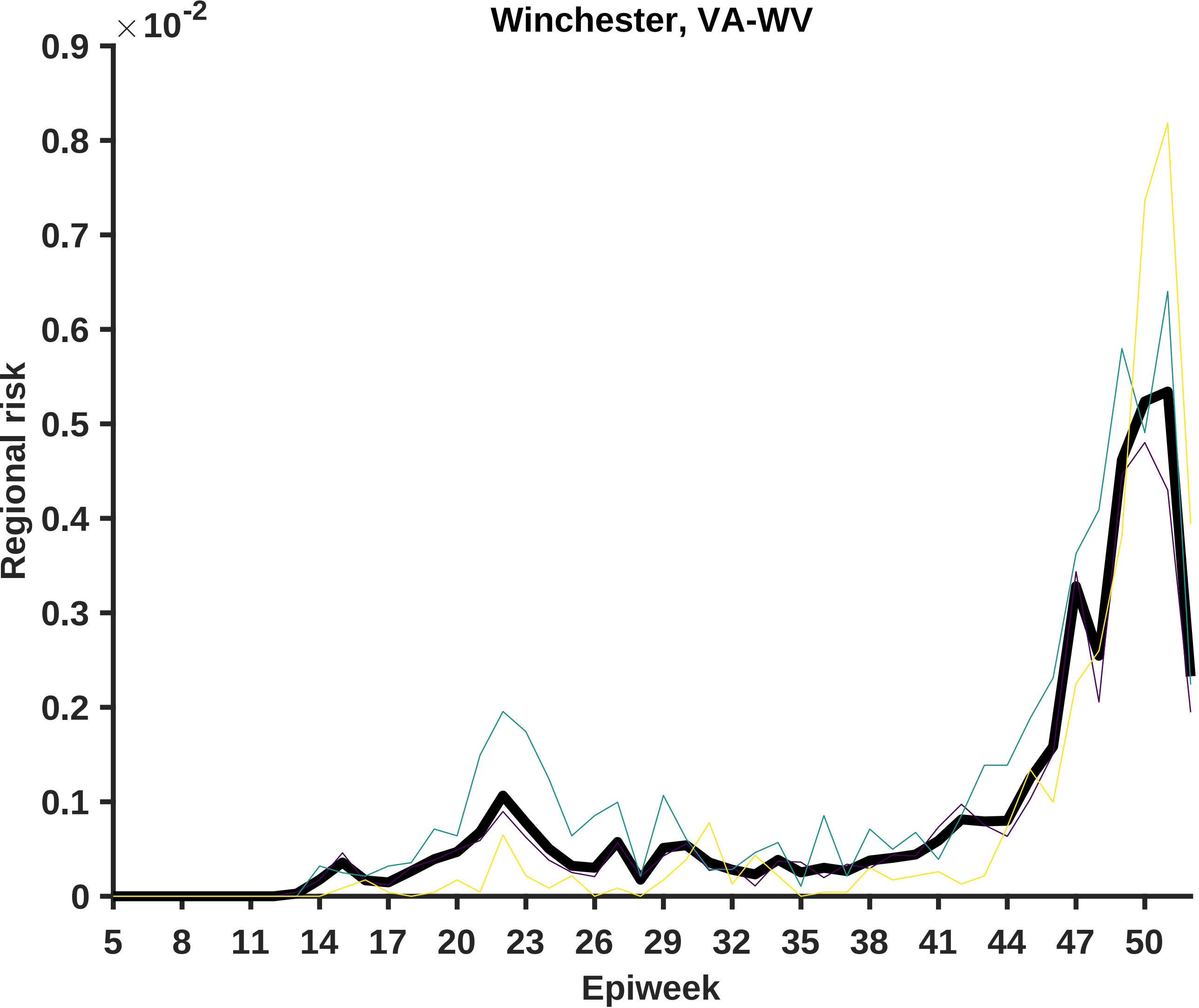
Winchester, VA-WV
×10^-2
0.9
0.8
0.7
0.6
0.5
0.4
0.3
0.2
0.1
0
Regional risk
5
8
11
14
17
20
23
26
29
32
35
38
41
44
47
50
Epiweek

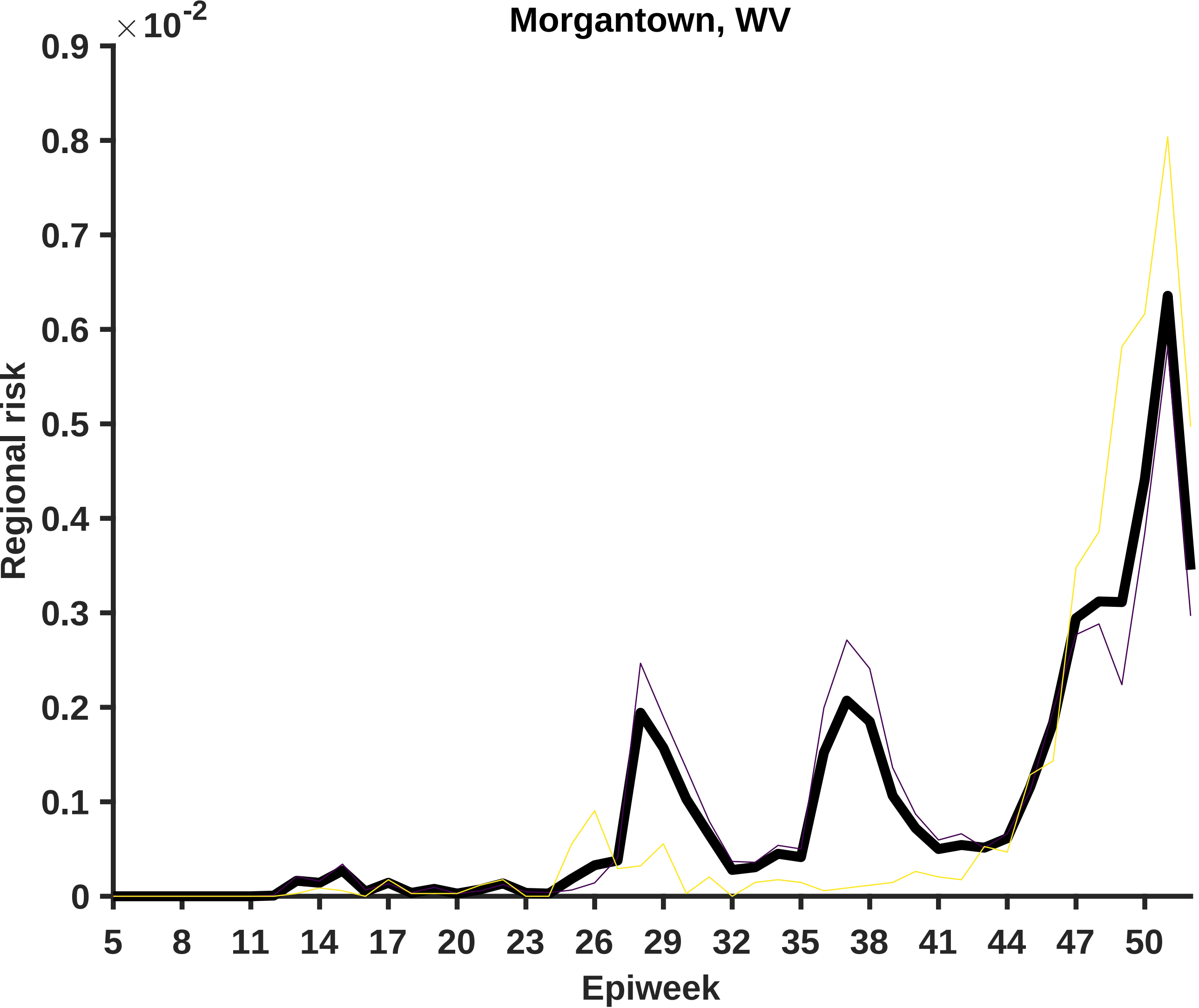

Morgantown, WV
×10^-2
0.9
0.8
0.7
0.6
0.5
0.4
0.3
0.2
0.1
0
Regional risk
5
8
11
14
17
20
23
26
29
32
35
38
41
44
47
50
Epiweek

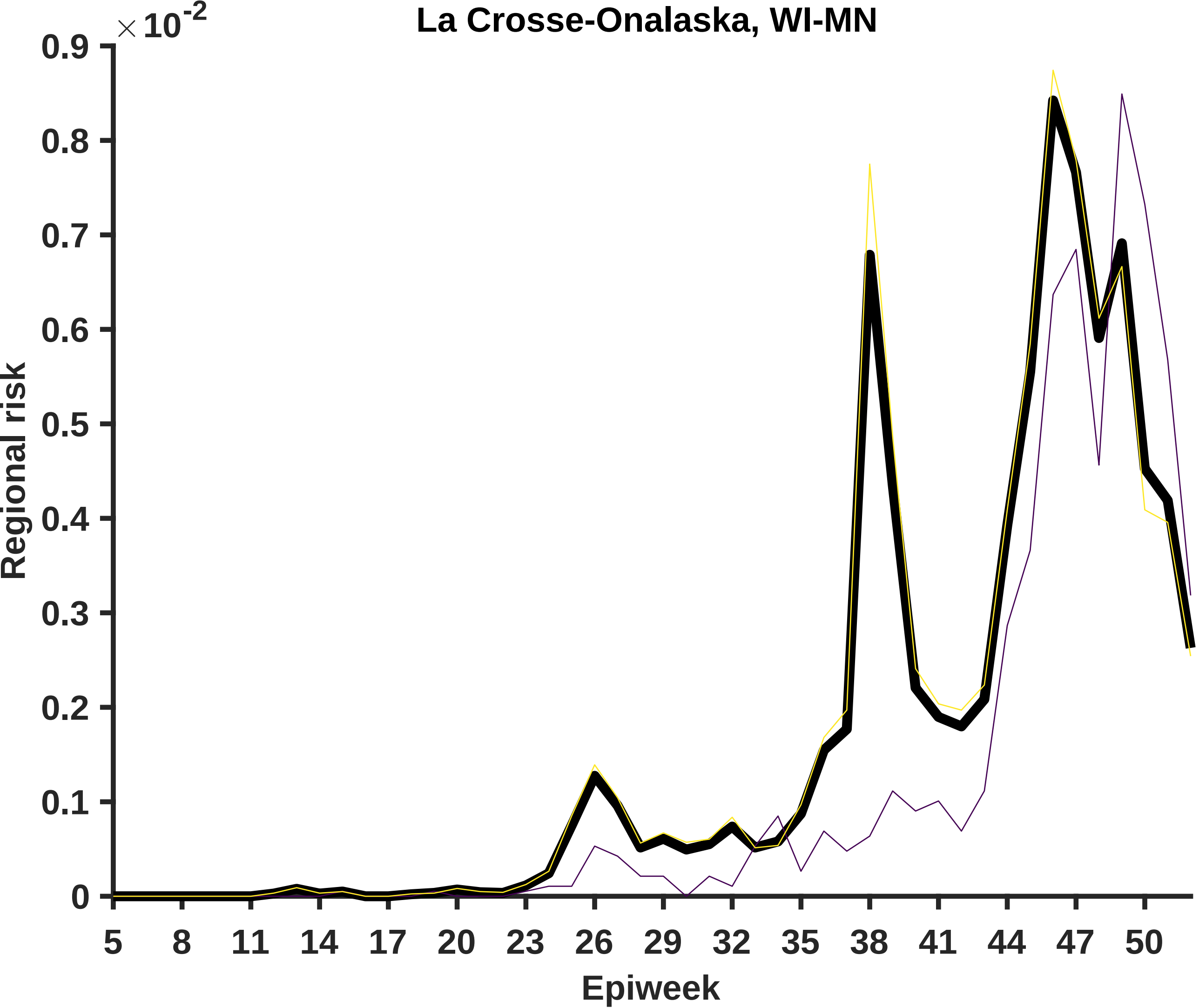
La Crosse-Onalaska, WI-MN
×10^-2
0.9
0.8
0.7
0.6
0.5
0.4
0.3
0.2
0.1
0
Regional risk
5
8
11
14
17
20
23
26
29
32
35
38
41
44
47
50
Epiweek

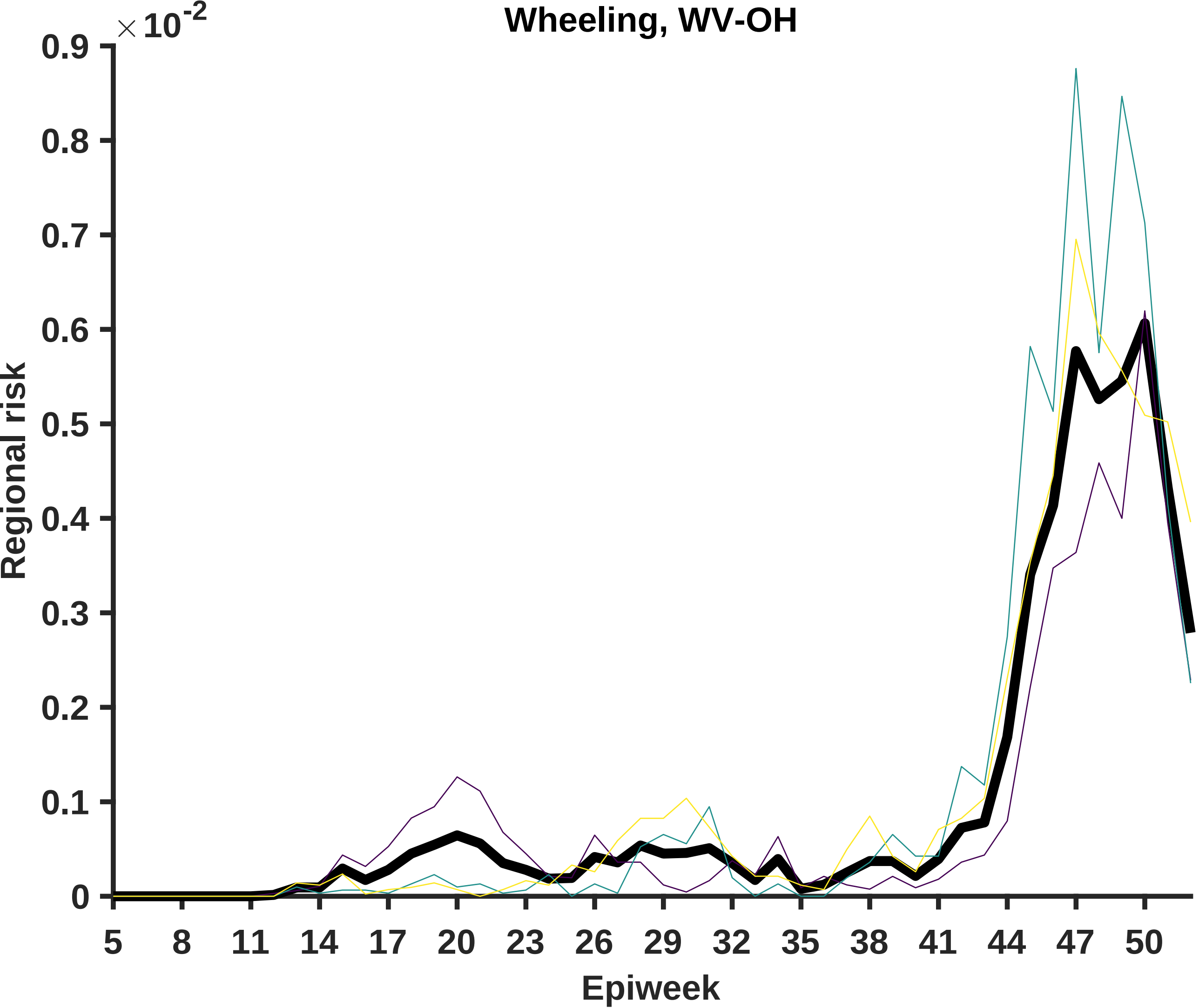

Wheeling, WV-OH
×10^-2
0.9
0.8
0.7
0.6
0.5
0.4
0.3
0.2
0.1
0
Regional risk
5
8
11
14
17
20
23
26
29
32
35
38
41
44
47
50
Epiweek

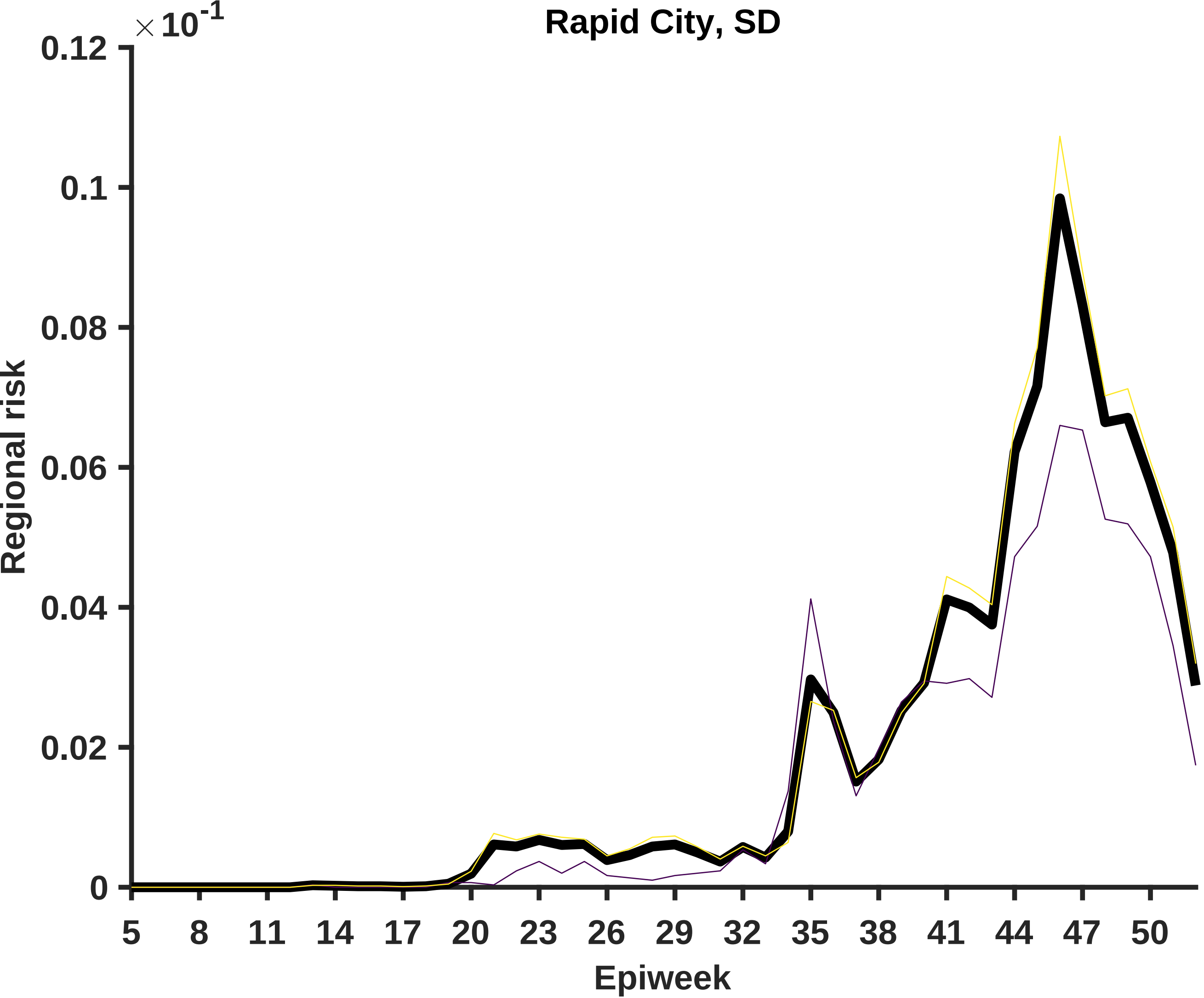
Rapid City, SD
×10^-1
0.12
0.1
0.08
0.06
0.04
0.02
0
Regional risk
5
8
11
14
17
20
23
26
29
32
35
38
41
44
47
50
Epiweek

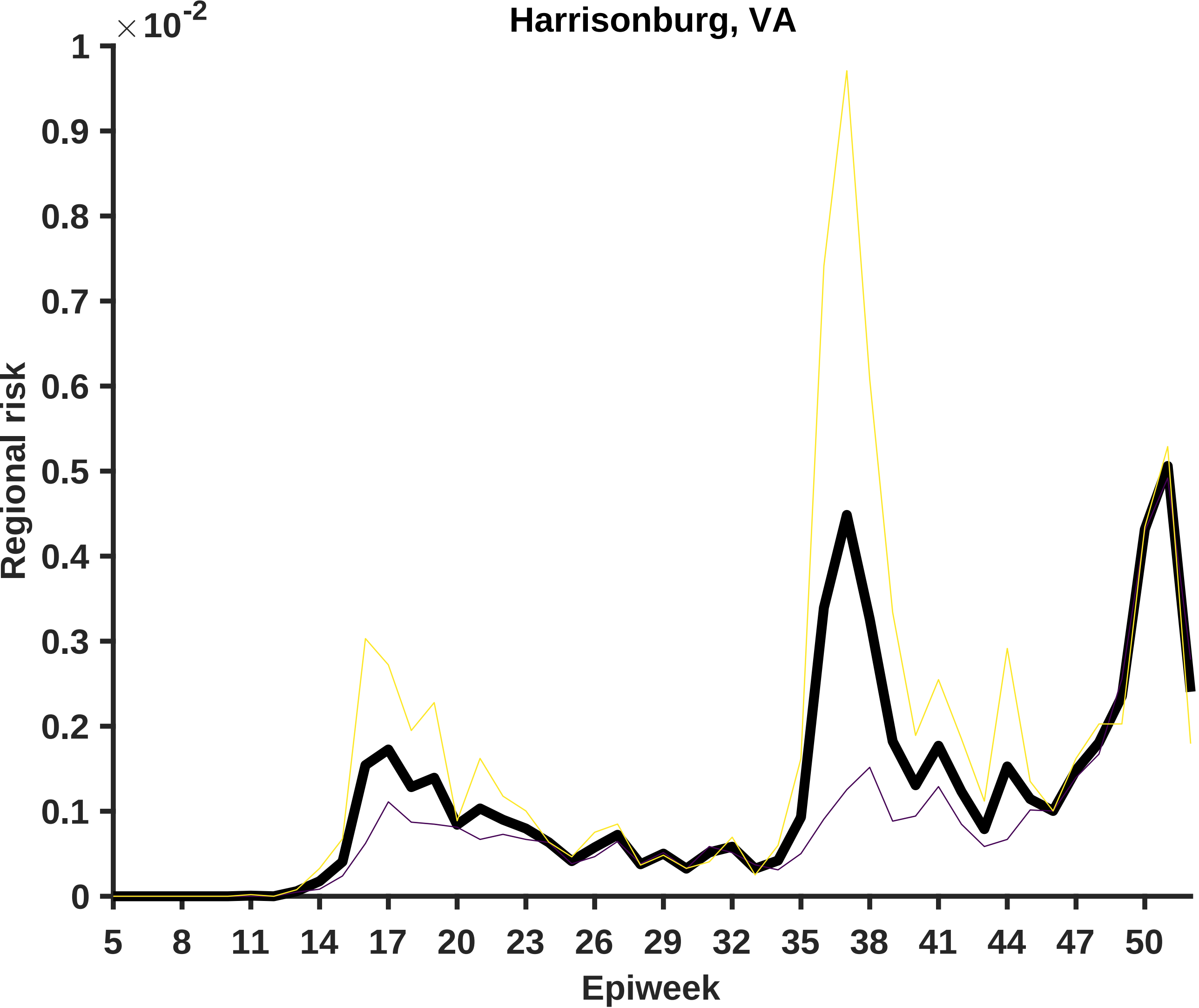

Harrisonburg, VA
×10^-2
1
0.9
0.8
0.7
0.6
0.5
0.4
0.3
0.2
0.1
0
Regional risk
5
8
11
14
17
20
23
26
29
32
35
38
41
44
47
50
Epiweek

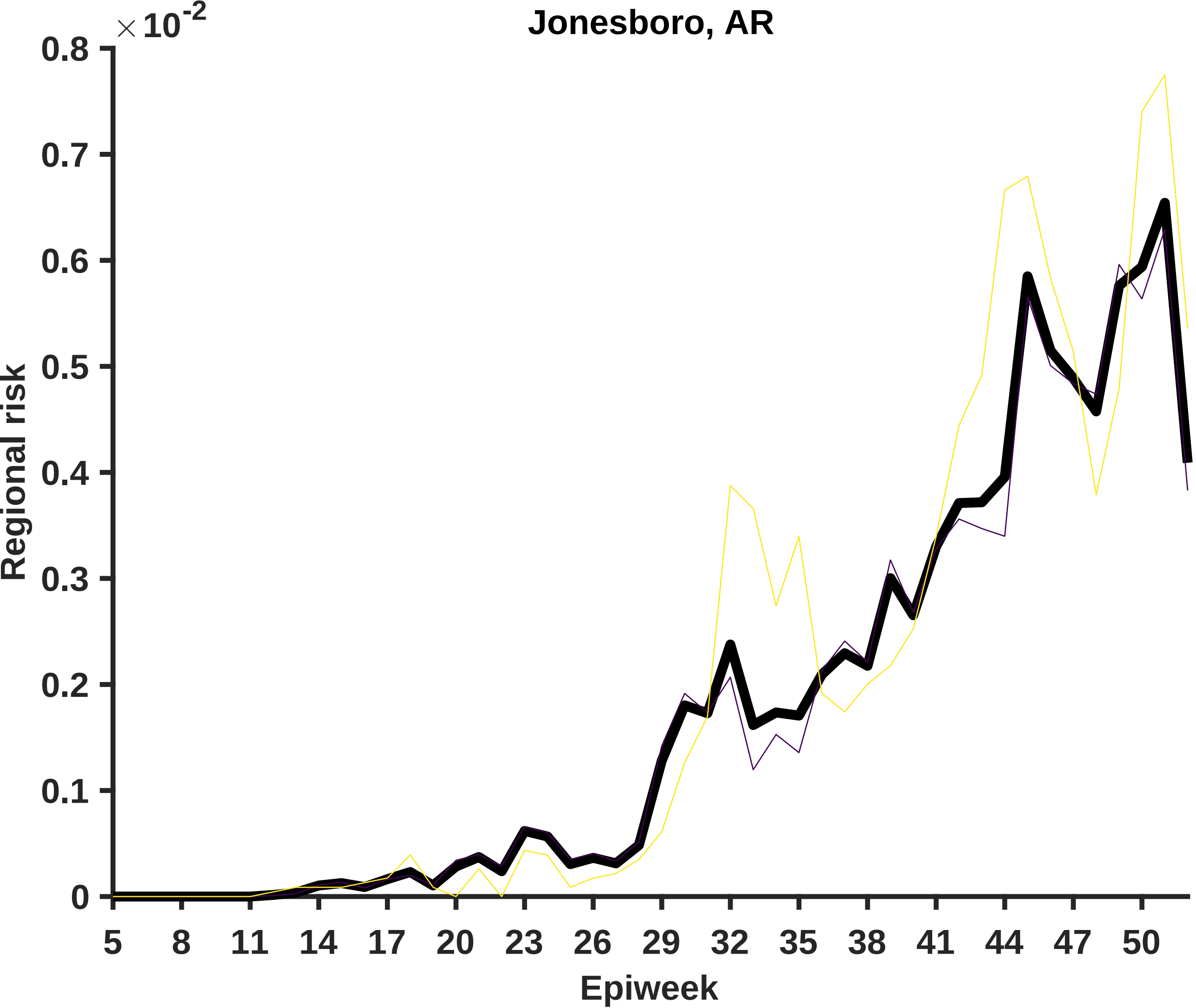
Jonesboro, AR
×10^-2
0.8
0.7
0.6
0.5
0.4
0.3
0.2
0.1
0
Regional risk
5
8
11
14
17
20
23
26
29
32
35
38
41
44
47
50
Epiweek

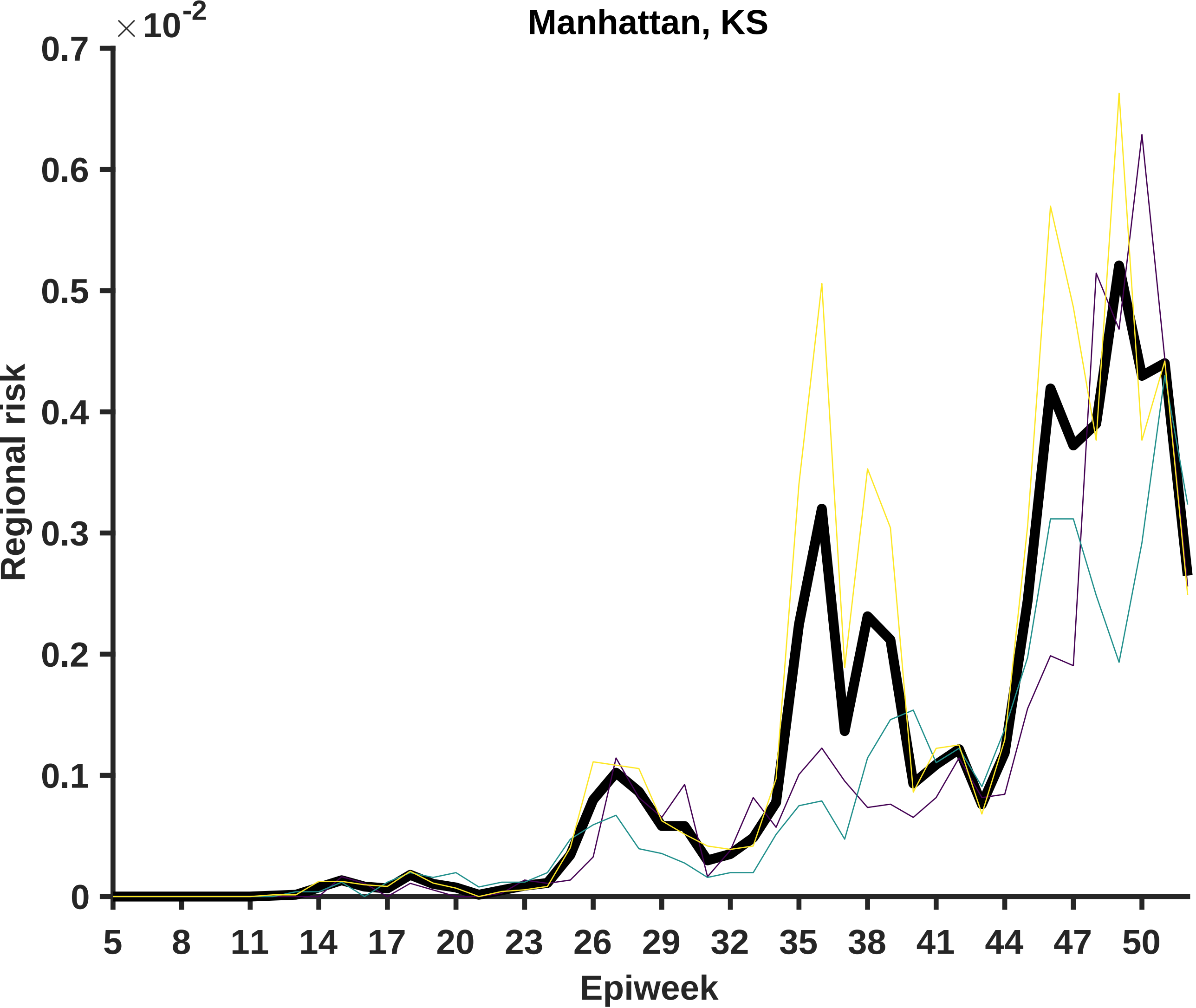

Manhattan, KS
×10^-2
0.7
0.6
0.5
0.4
0.3
0.2
0.1
0
Regional risk
5
8
11
14
17
20
23
26
29
32
35
38
41
44
47
50
Epiweek

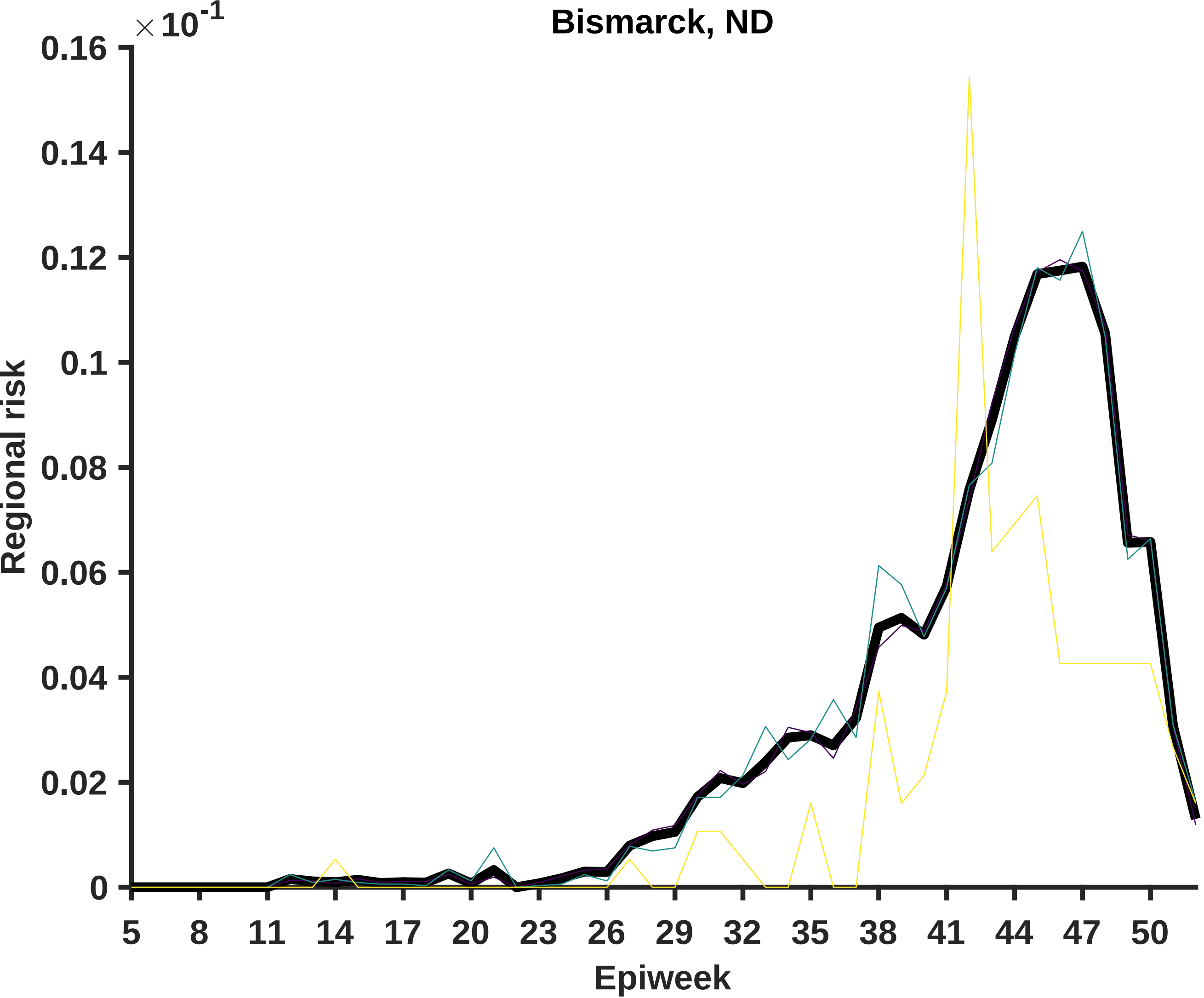

Bismarck, ND
× 10^-1
0.16
0.14
0.12
0.1
0.08
0.06
0.04
0.02
0
Regional risk
5
8
11
14
17
20
23
26
29
32
35
38
41
44
47
50
Epiweek

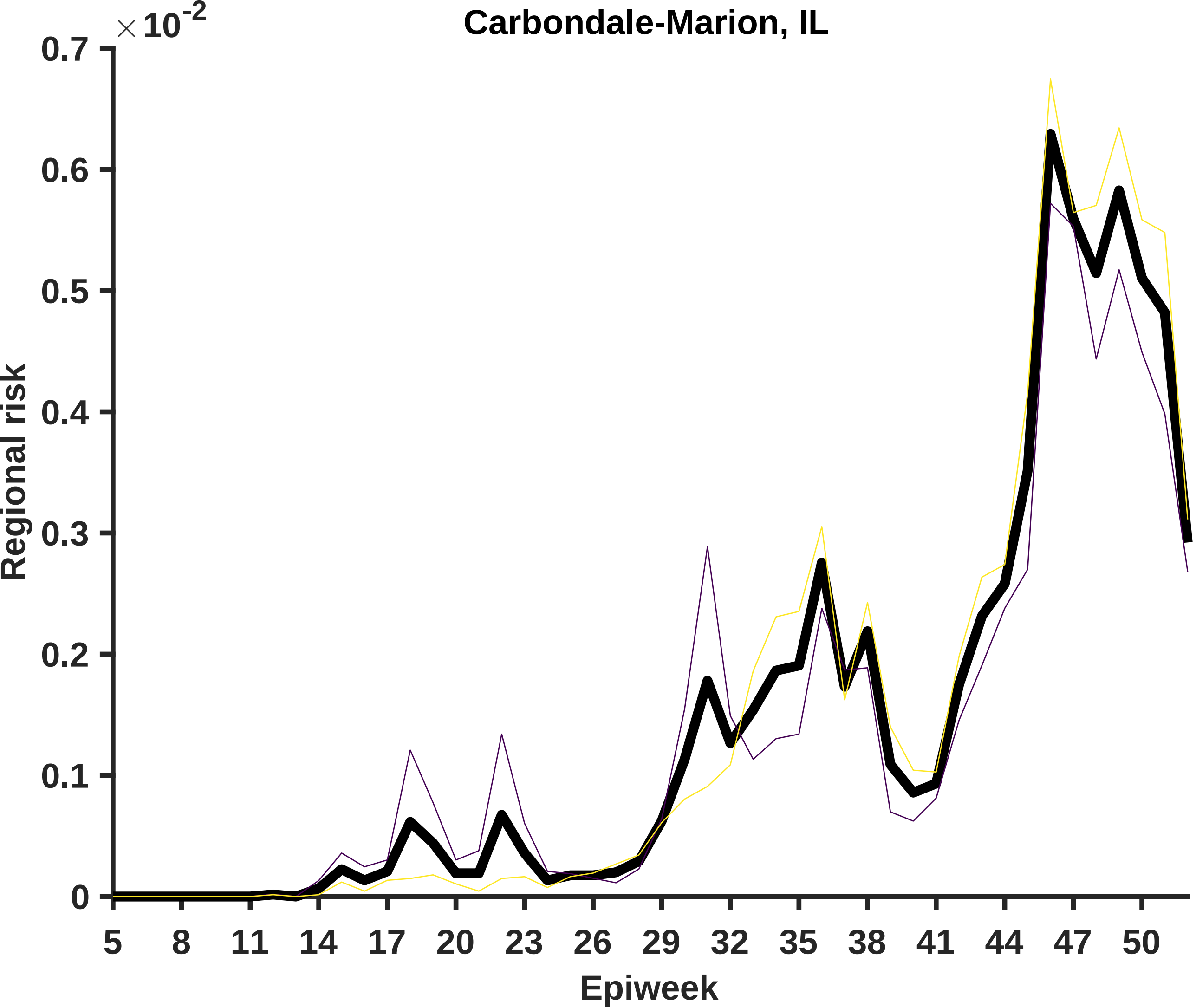

Carbondale-Marion, IL
×10^-2
0.7
0.6
0.5
0.4
0.3
0.2
0.1
0
Regional risk
5
8
11
14
17
20
23
26
29
32
35
38
41
44
47
50
Epiweek

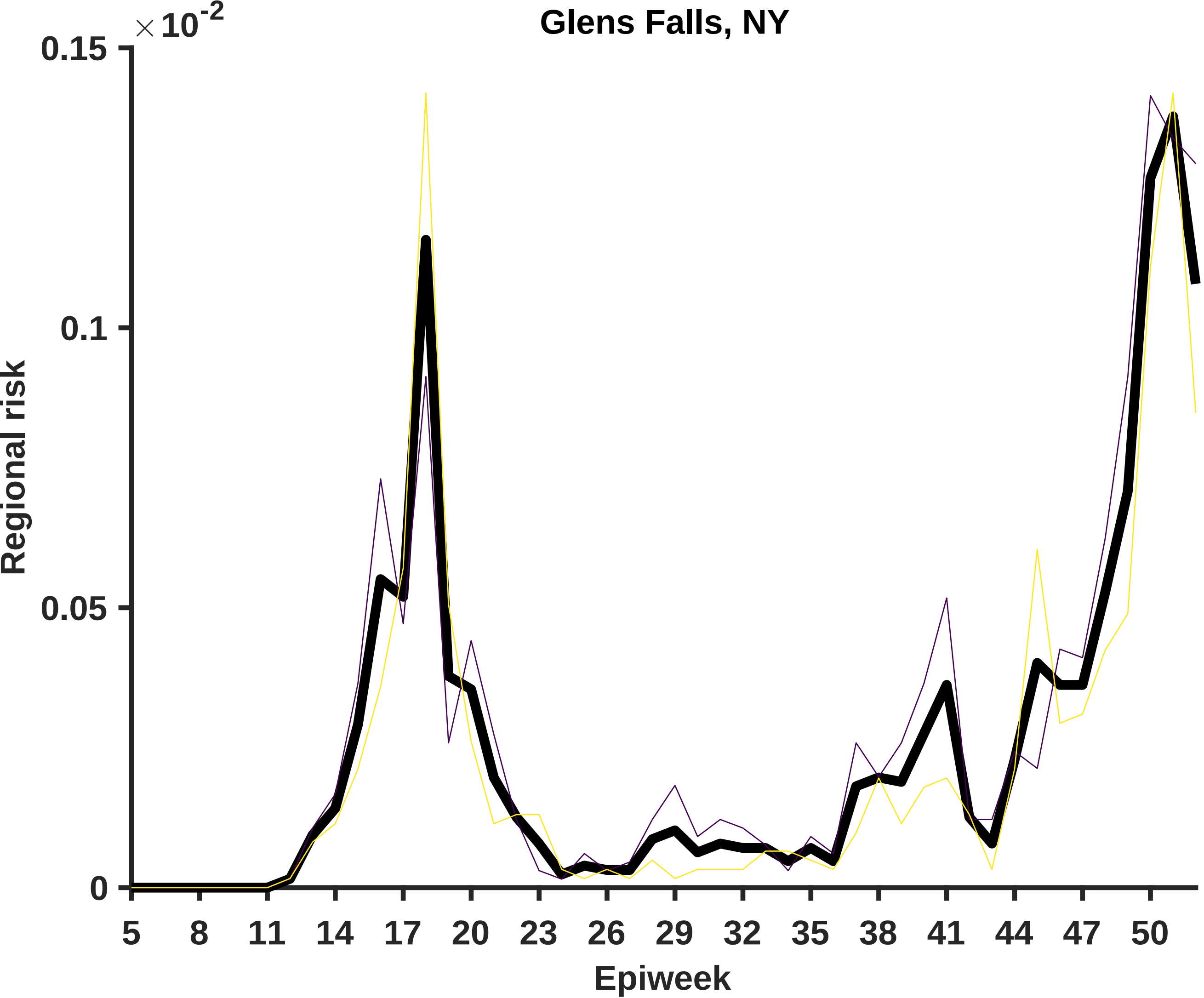
Glens Falls, NY
×10^-2
0.15
0.1
0.05
0
Regional risk
5
8
11
14
17
20
23
26
29
32
35
38
41
44
47
50
Epiweek

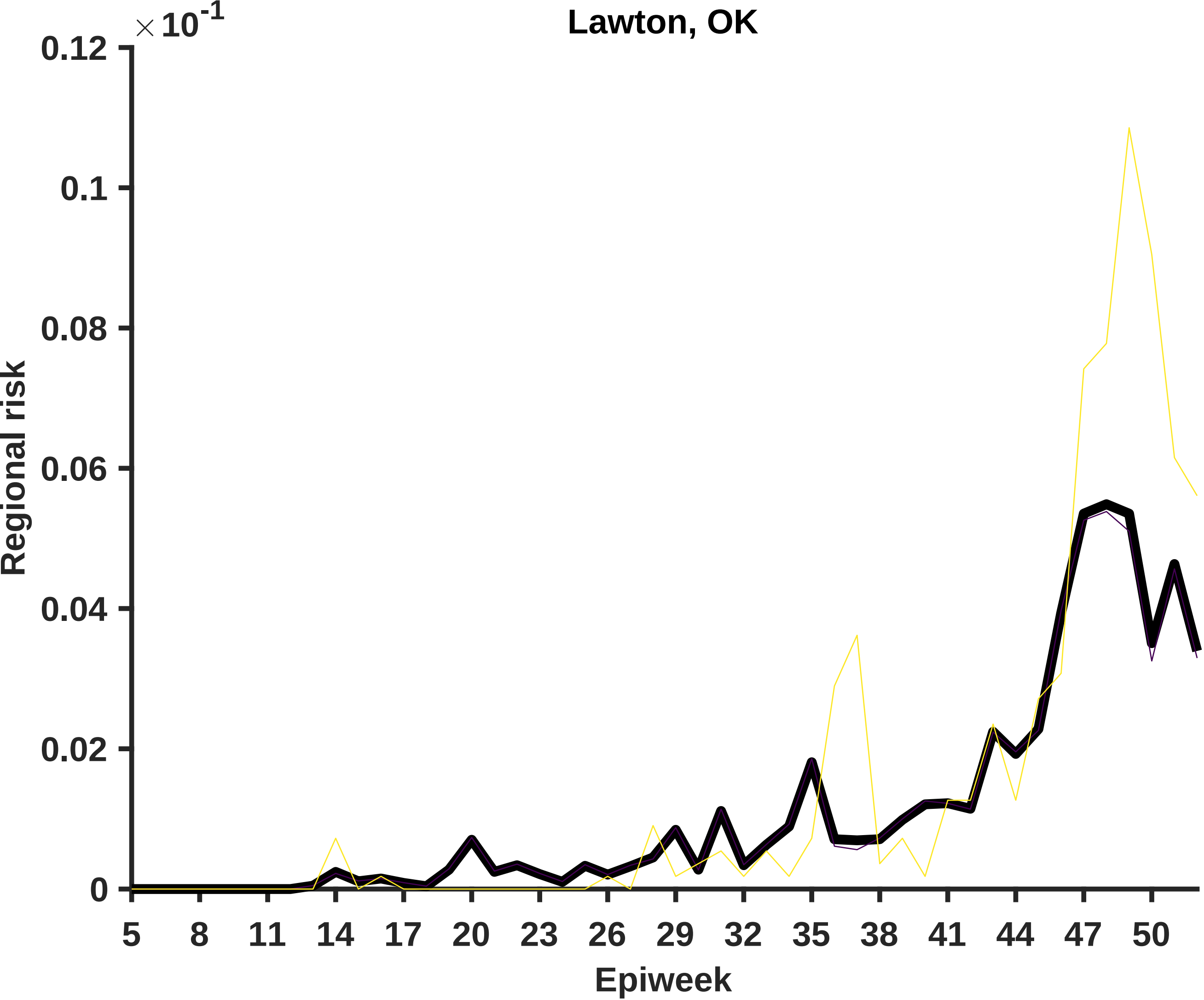

Lawton, OK
× 10^-1
0.12
0.1
0.08
0.06
0.04
0.02
0
Regional risk
5
8
11
14
17
20
23
26
29
32
35
38
41
44
47
50
Epiweek

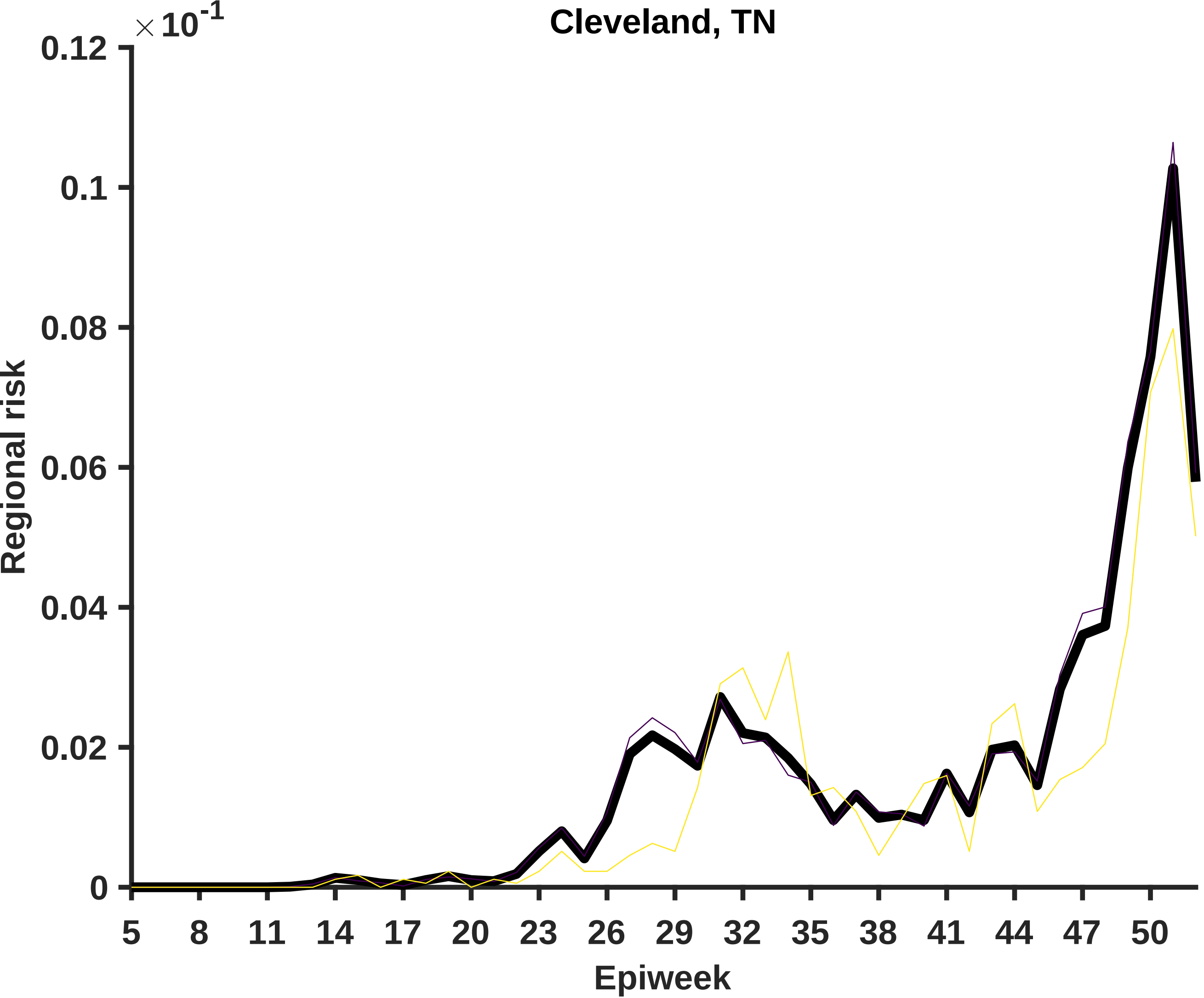

Cleveland, TN
× 10^-1
0.12
0.1
0.08
0.06
0.04
0.02
0
Regional risk
5
8
11
14
17
20
23
26
29
32
35
38
41
44
47
50
Epiweek

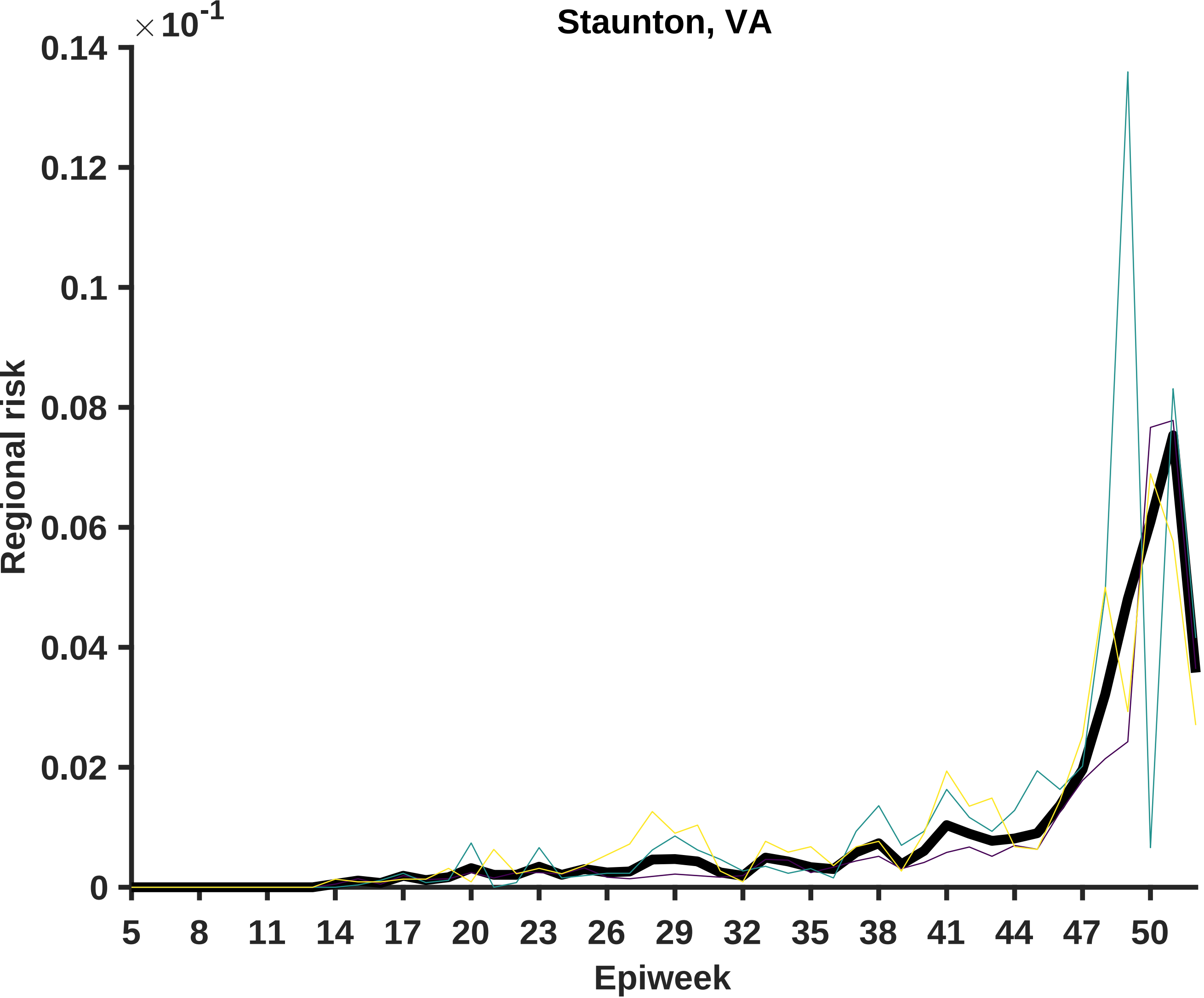

Staunton, VA
×10^-1
0.14
0.12
0.1
0.08
0.06
0.04
0.02
0
Regional risk
5
8
11
14
17
20
23
26
29
32
35
38
41
44
47
50
Epiweek

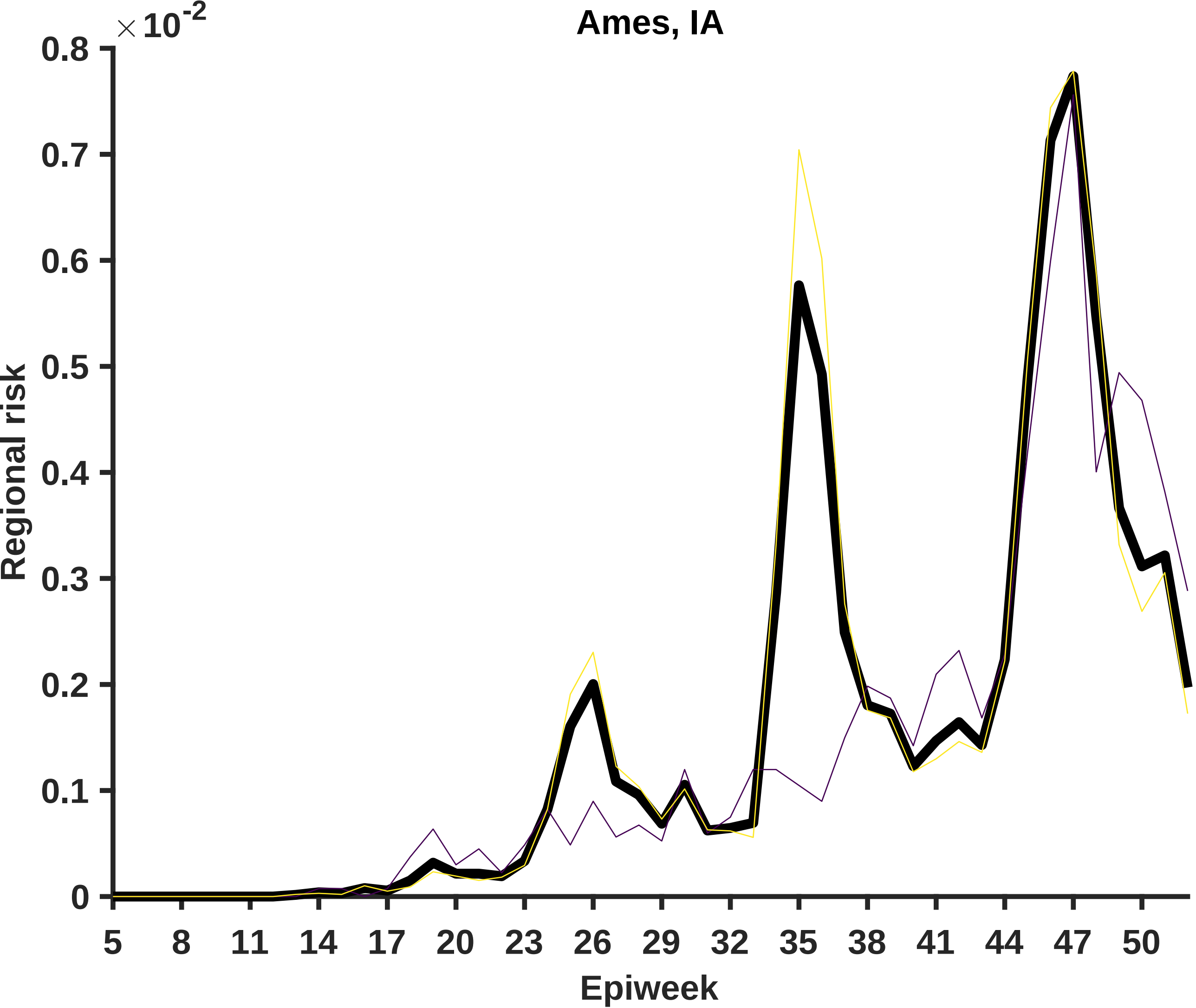

Ames, IA
×10^-2
0.8
0.7
0.6
0.5
0.4
0.3
0.2
0.1
0
Regional risk
5
8
11
14
17
20
23
26
29
32
35
38
41
44
47
50
Epiweek

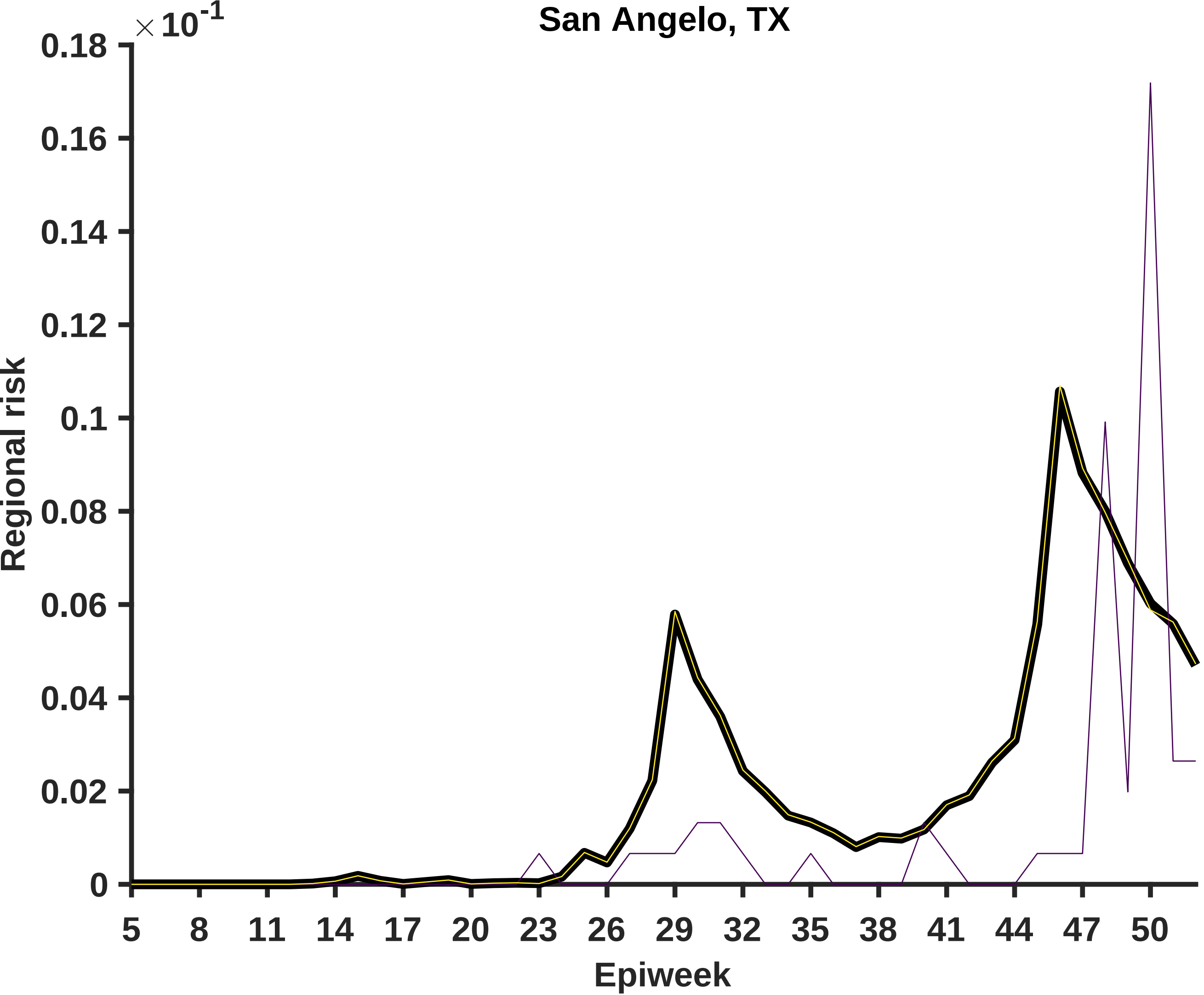

San Angelo, TX
×10^-1
0.18
0.16
0.14
0.12
0.1
0.08
0.06
0.04
0.02
0
Regional risk
5
8
11
14
17
20
23
26
29
32
35
38
41
44
47
50
Epiweek

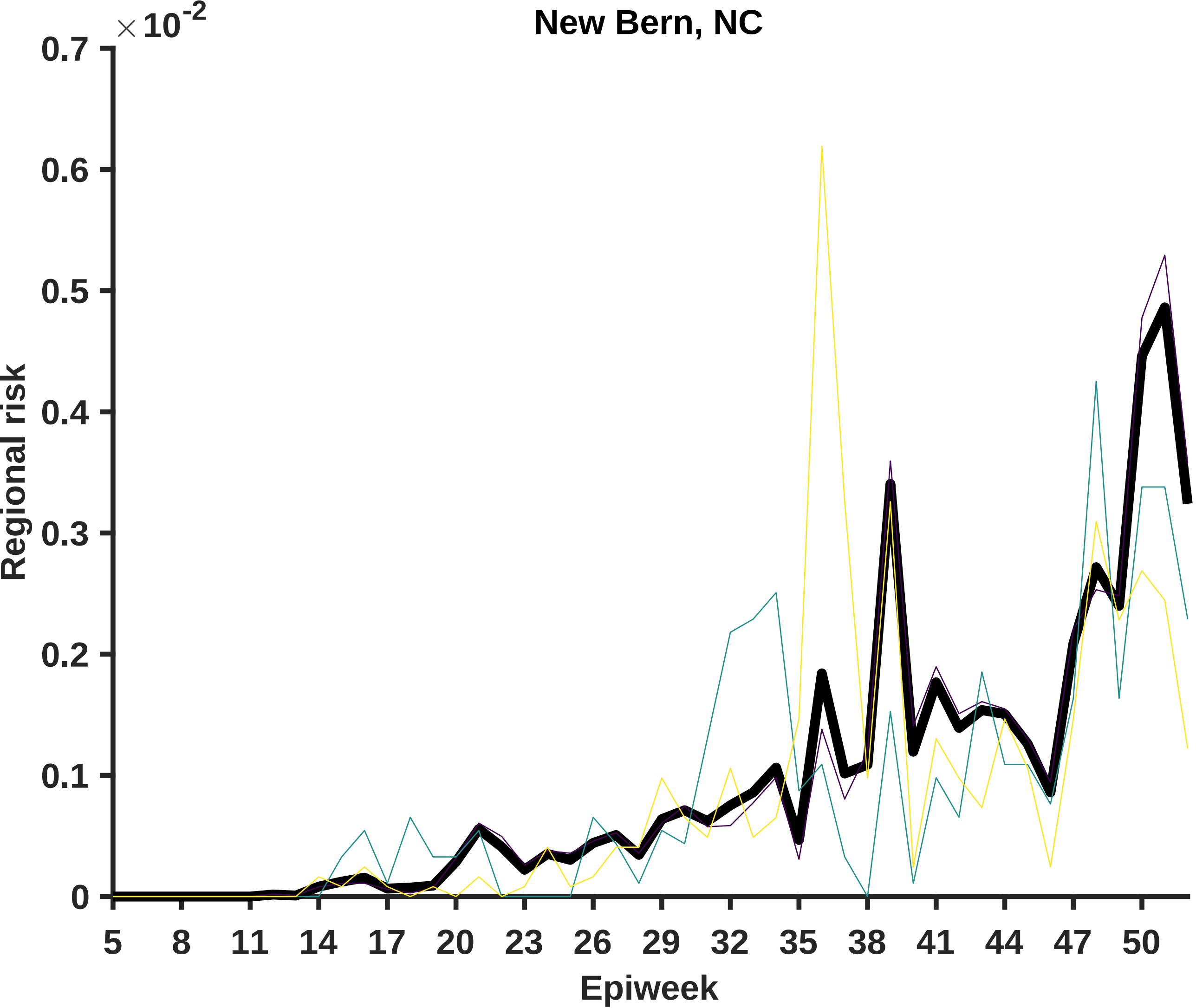
New Bern, NC
×10^-2
0.7
0.6
0.5
0.4
0.3
0.2
0.1
0
Regional risk
5
8
11
14
17
20
23
26
29
32
35
38
41
44
47
50
Epiweek

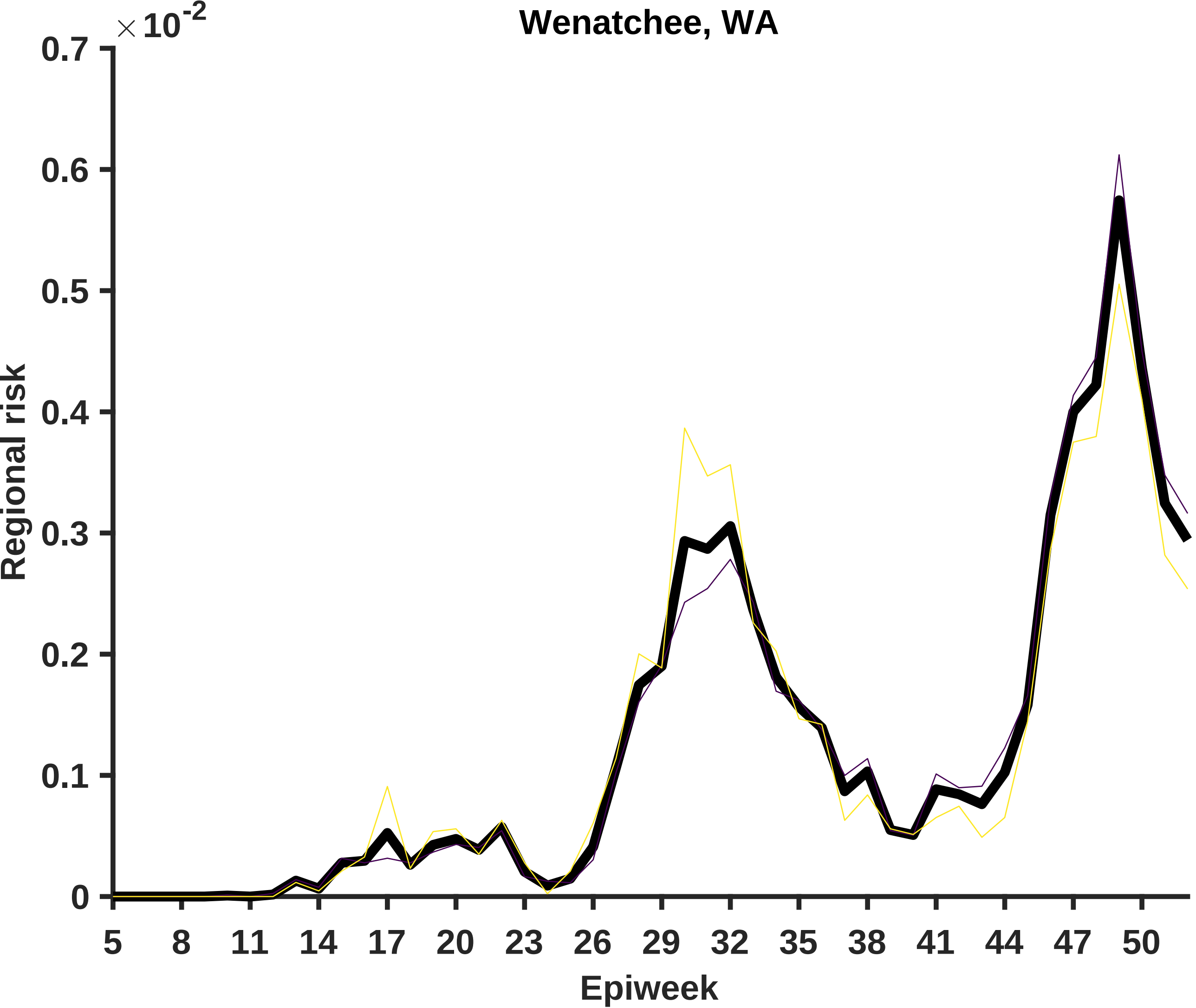
Wenatchee, WA
×10^-2
0.7
0.6
0.5
0.4
0.3
0.2
0.1
0
Regional risk
5
8
11
14
17
20
23
26
29
32
35
38
41
44
47
50
Epiweek

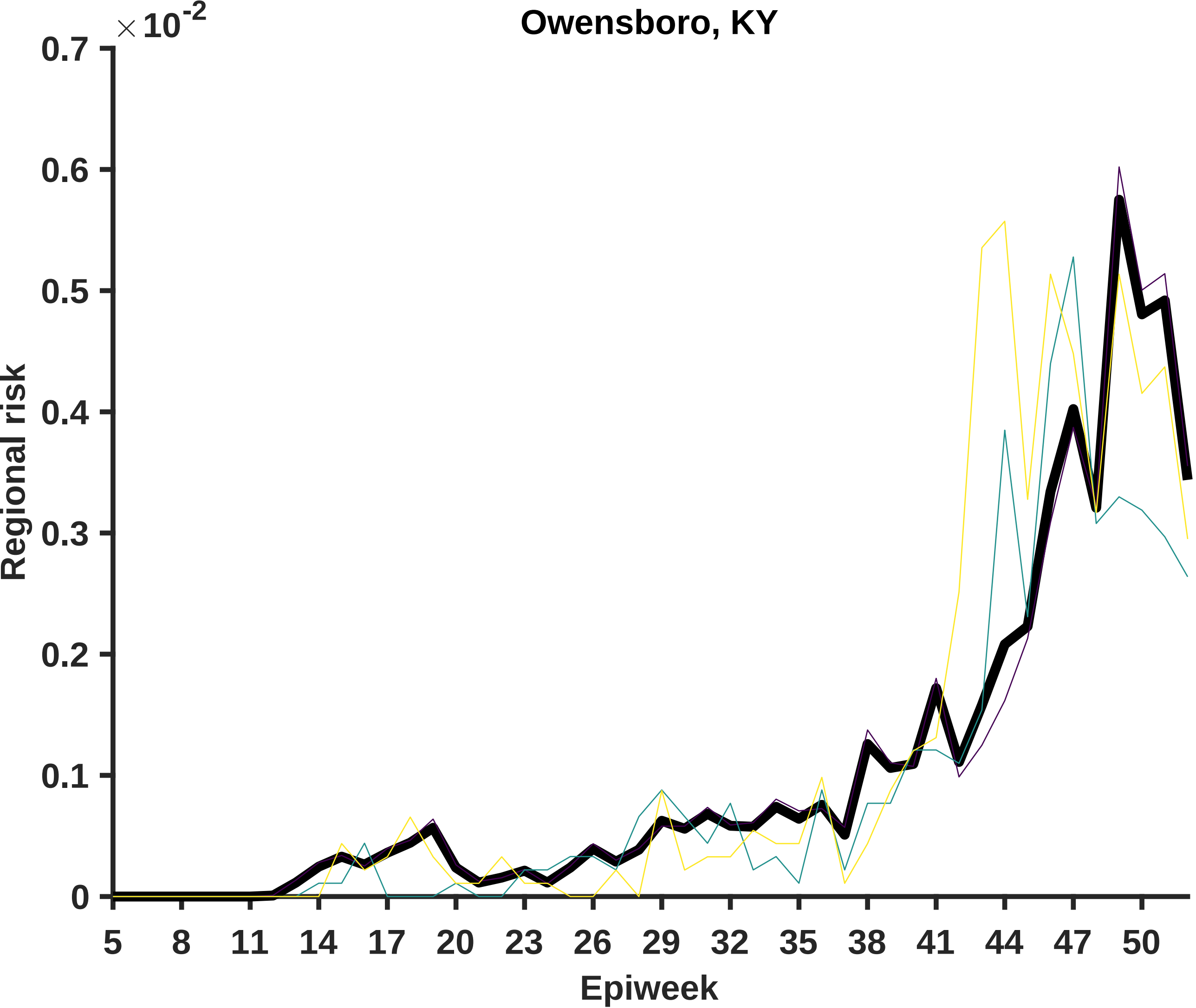

Owensboro, KY
×10^-2
0.7
0.6
0.5
0.4
0.3
0.2
0.1
0
Regional risk
5
8
11
14
17
20
23
26
29
32
35
38
41
44
47
50
Epiweek

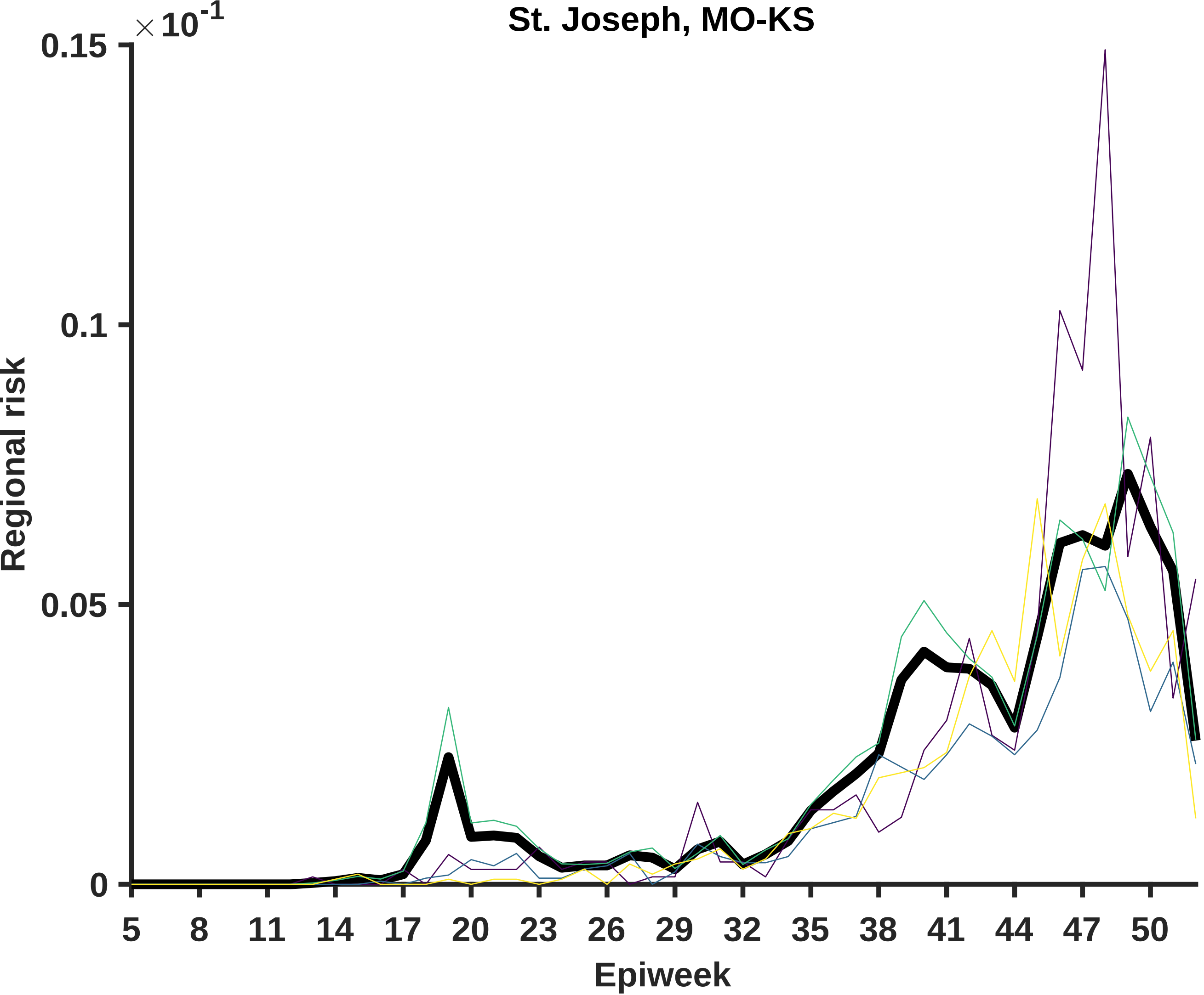
St. Joseph, MO-KS
×10-1
0.15
0.1
0.05
0
Regional risk
5
8
11
14
17
20
23
26
29
32
35
38
41
44
47
50
Epiweek

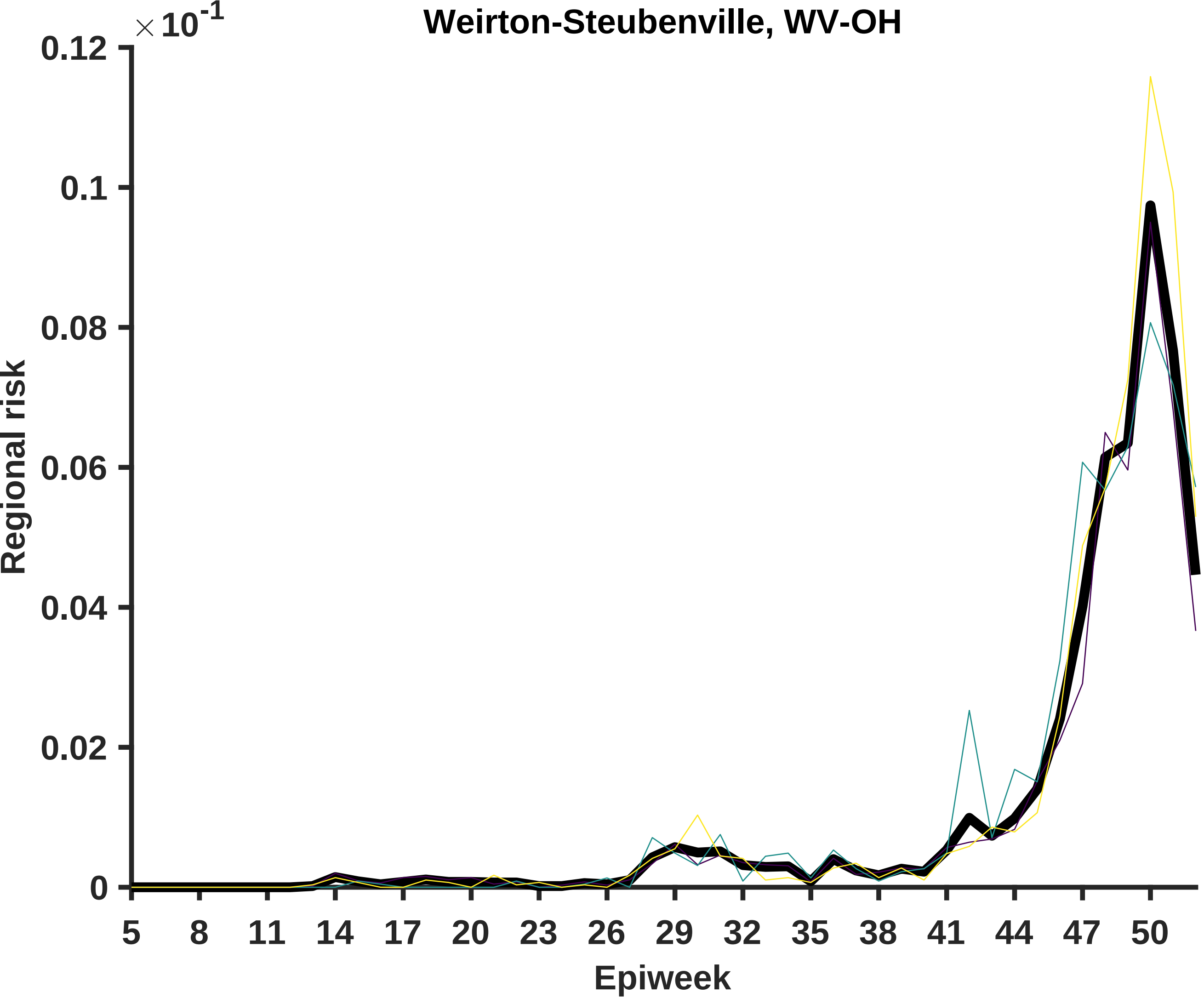

Weirton-Steubenville, WV-OH
×10^-1
0.12
0.1
0.08
0.06
0.04
0.02
0
Regional risk
5
8
11
14
17
20
23
26
29
32
35
38
41
44
47
50
Epiweek

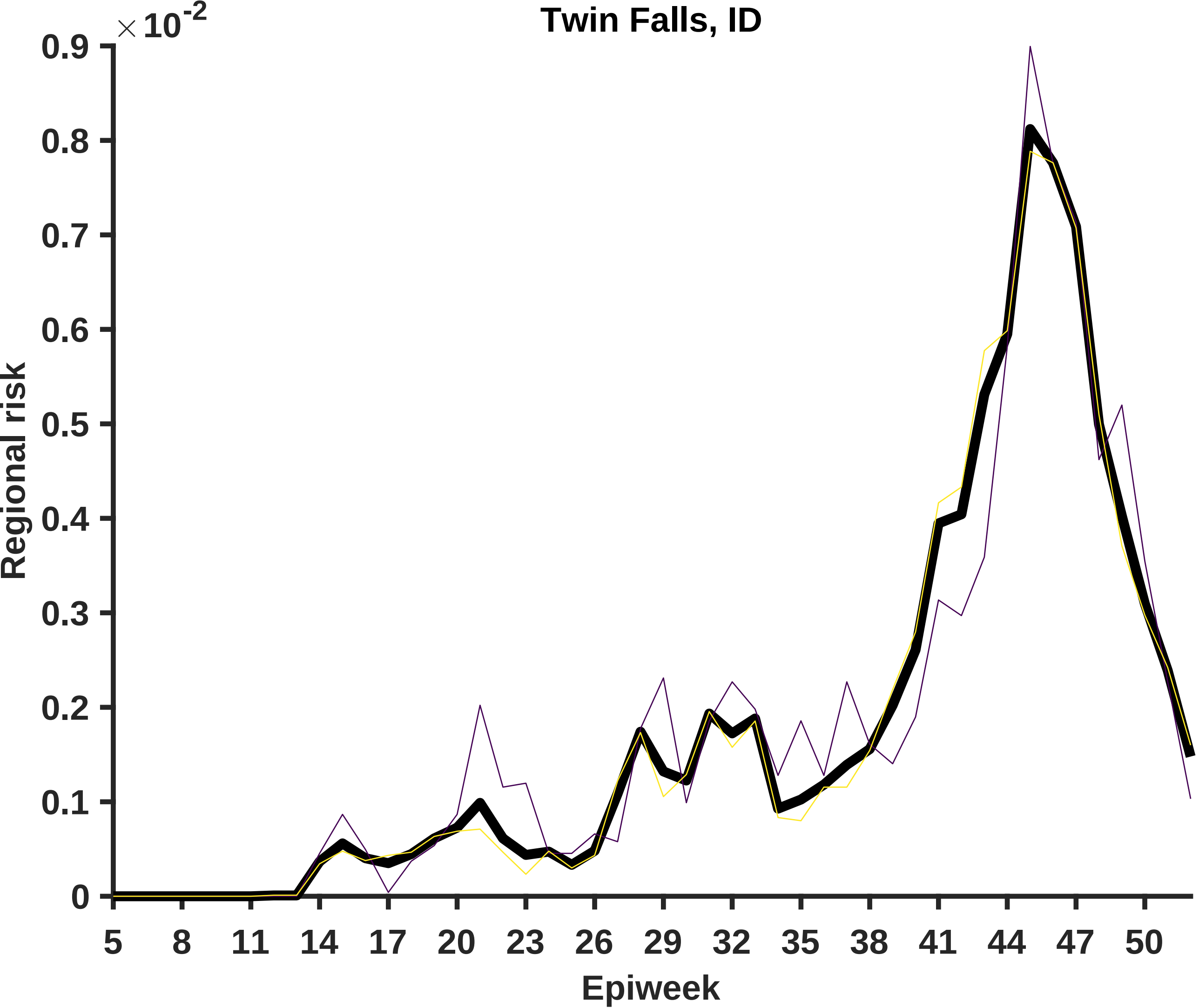

Twin Falls, ID
×10^-2
0.9
0.8
0.7
0.6
0.5
0.4
0.3
0.2
0.1
0
Regional risk
5
8
11
14
17
20
23
26
29
32
35
38
41
44
47
50
Epiweek

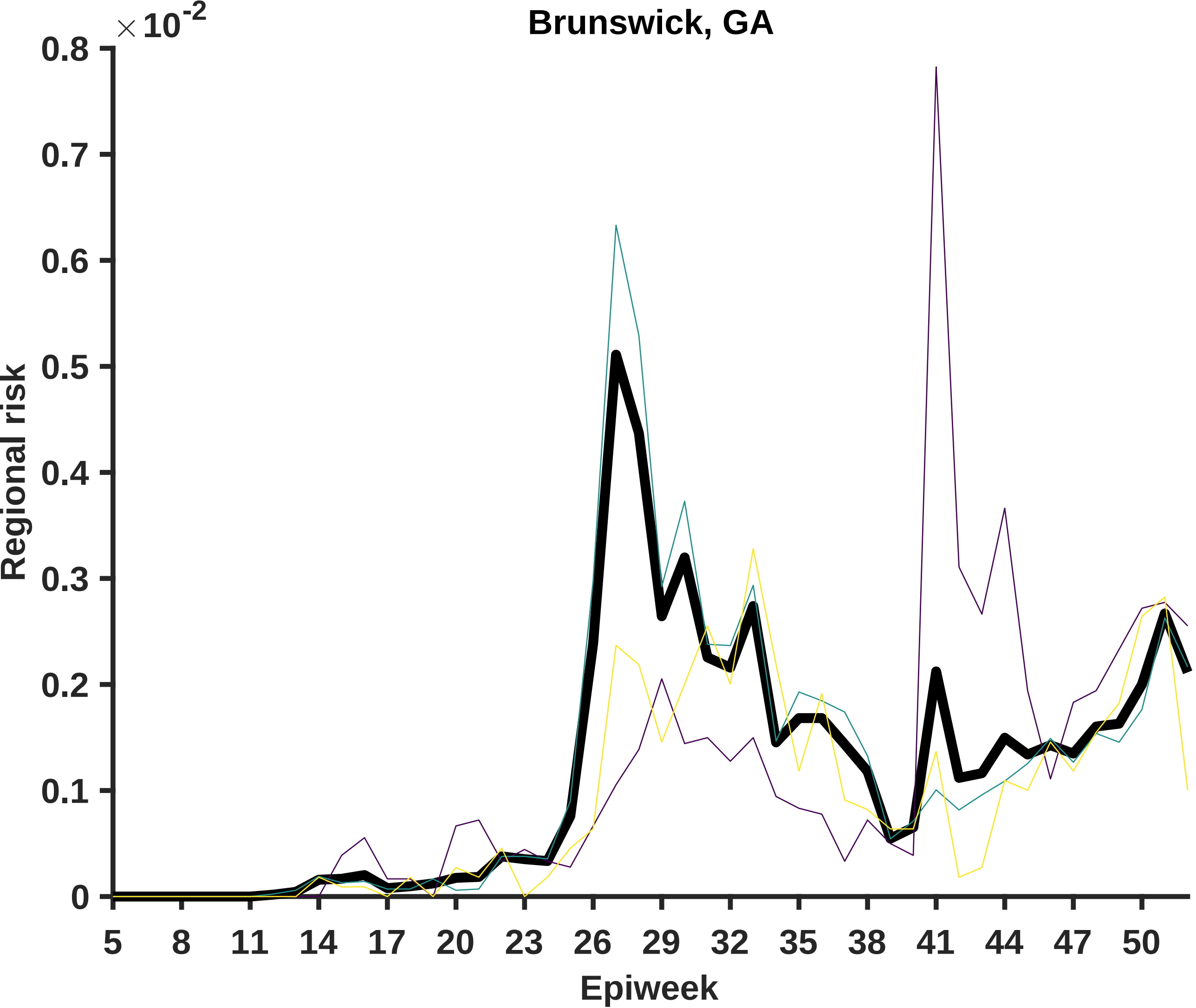

Brunswick, GA
×10^-2
0.8
0.7
0.6
0.5
0.4
0.3
0.2
0.1
0
Regional risk
5
8
11
14
17
20
23
26
29
32
35
38
41
44
47
50
Epiweek

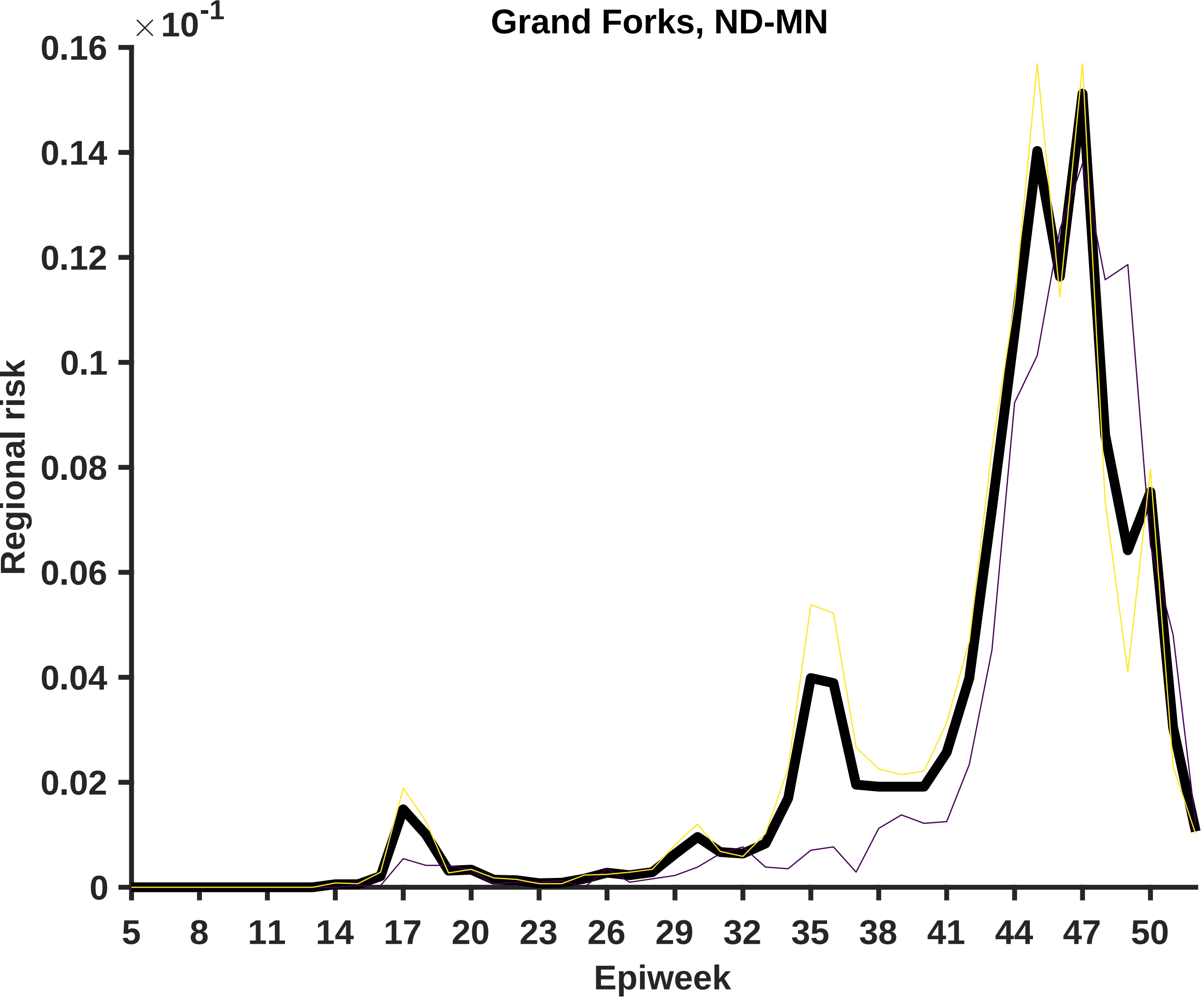

Grand Forks, ND-MN
×10^-1
0.16
0.14
0.12
0.1
0.08
0.06
0.04
0.02
0
Regional risk
5
8
11
14
17
20
23
26
29
32
35
38
41
44
47
50
Epiweek

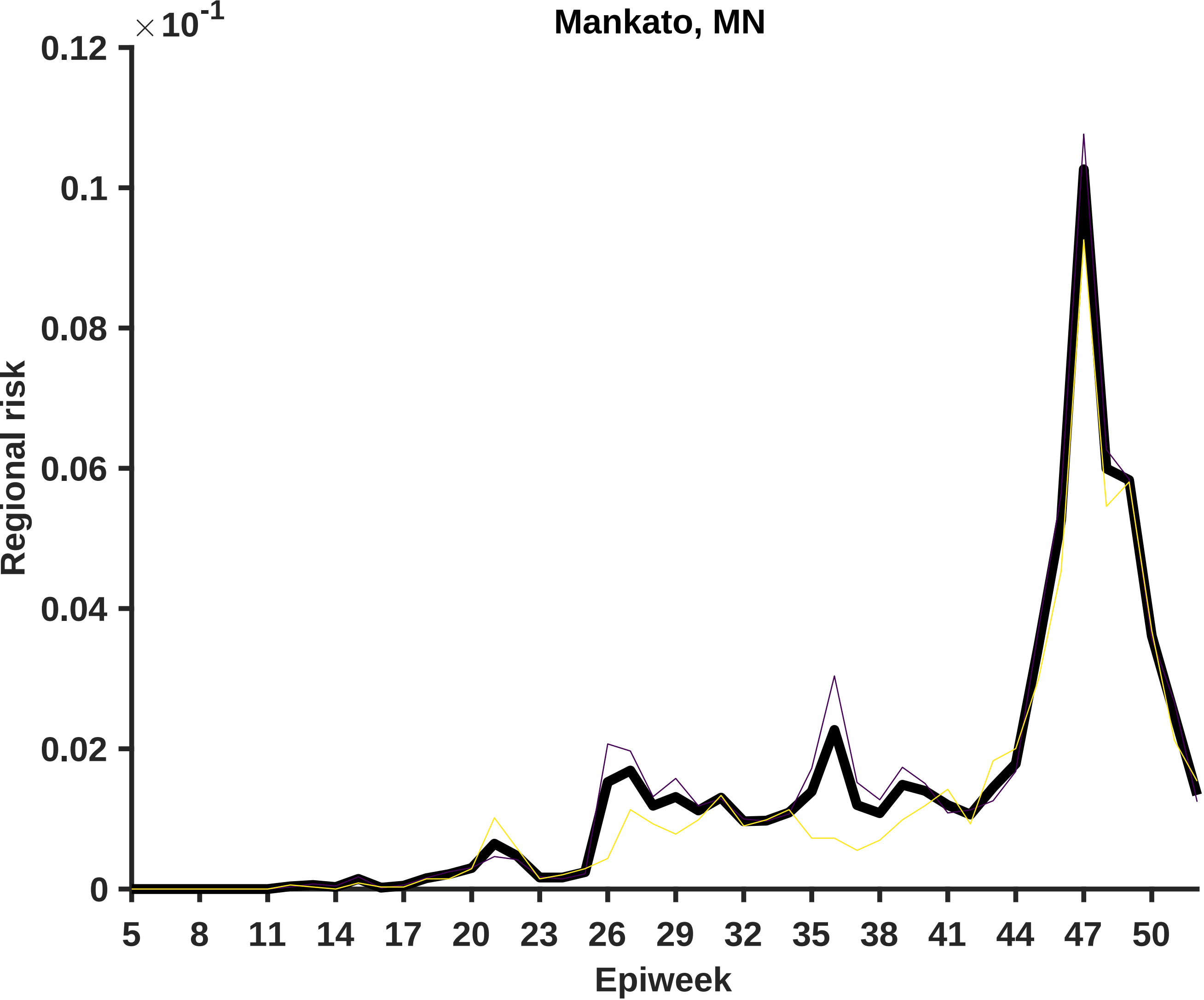

Mankato, MN
× 10^-1
0.12
0.1
0.08
0.06
0.04
0.02
0
Regional risk
5
8
11
14
17
20
23
26
29
32
35
38
41
44
47
50
Epiweek

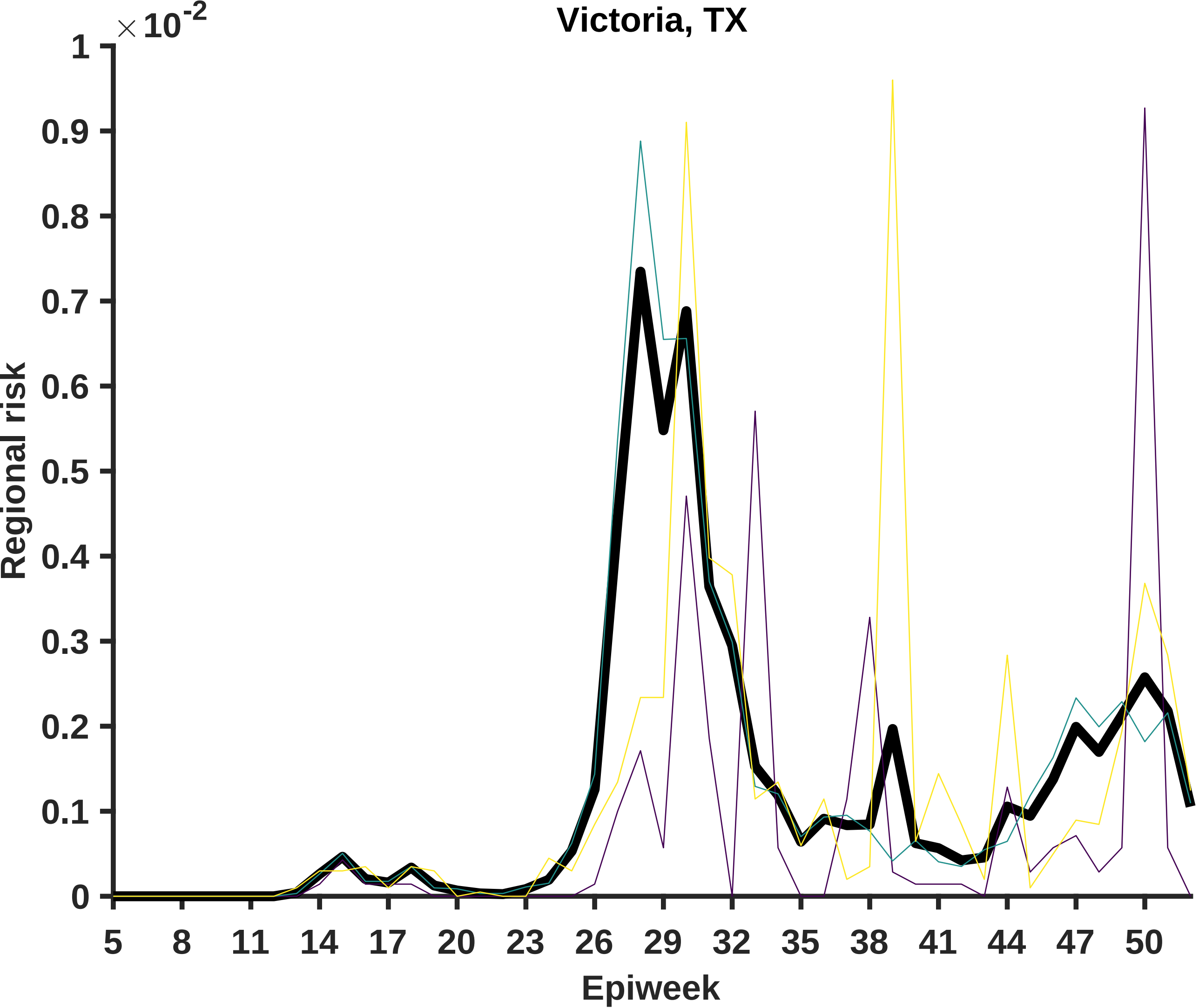

Victoria, TX
×10-2
1
0.9
0.8
0.7
0.6
0.5
0.4
0.3
0.2
0.1
0
Regional risk
5
8
11
14
17
20
23
26
29
32
35
38
41
44
47
50
Epiweek

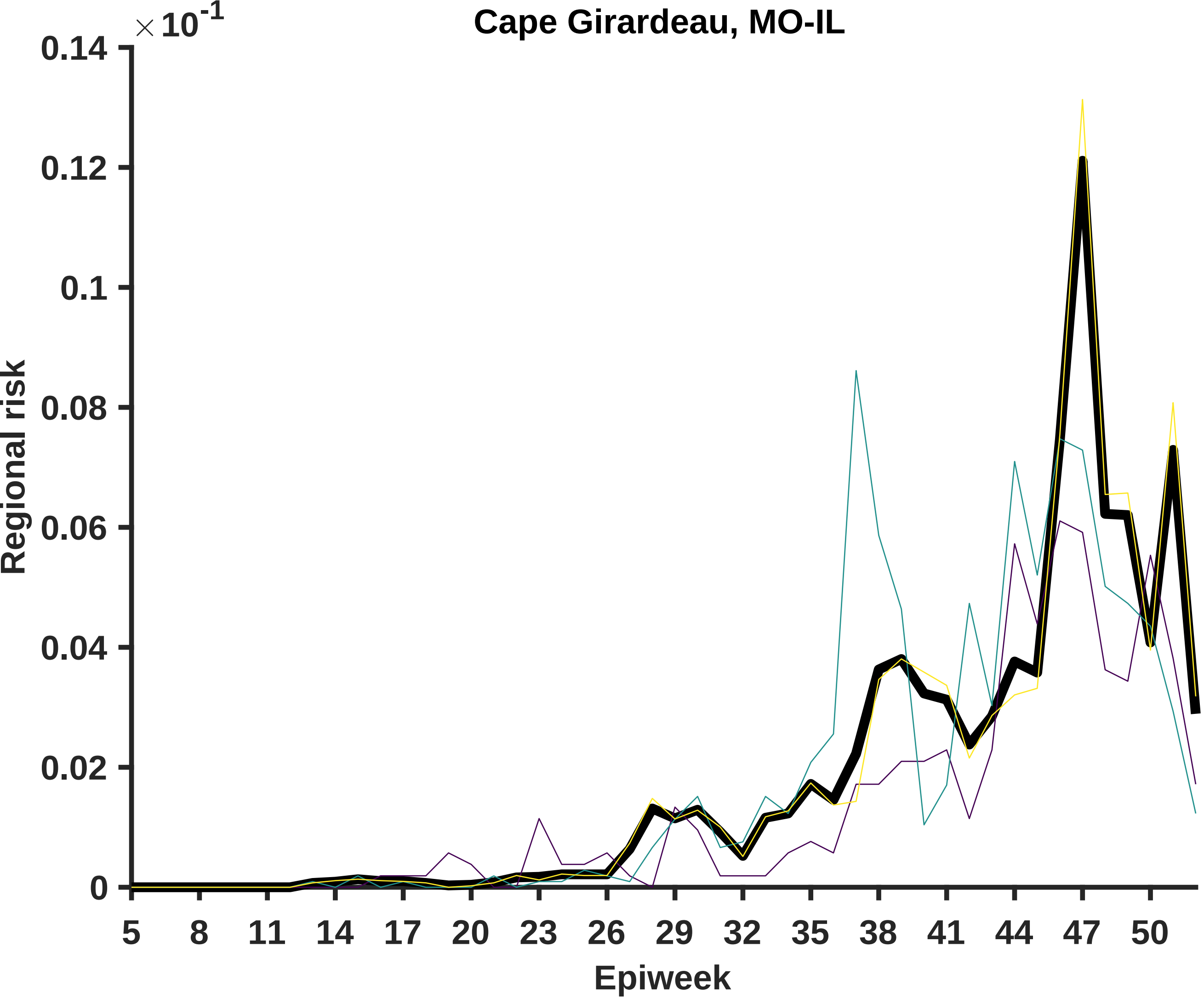
Cape Girardeau, MO-IL
× 10^-1
0.14
0.12
0.1
0.08
0.06
0.04
0.02
0
Regional risk
5
8
11
14
17
20
23
26
29
32
35
38
41
44
47
50
Epiweek

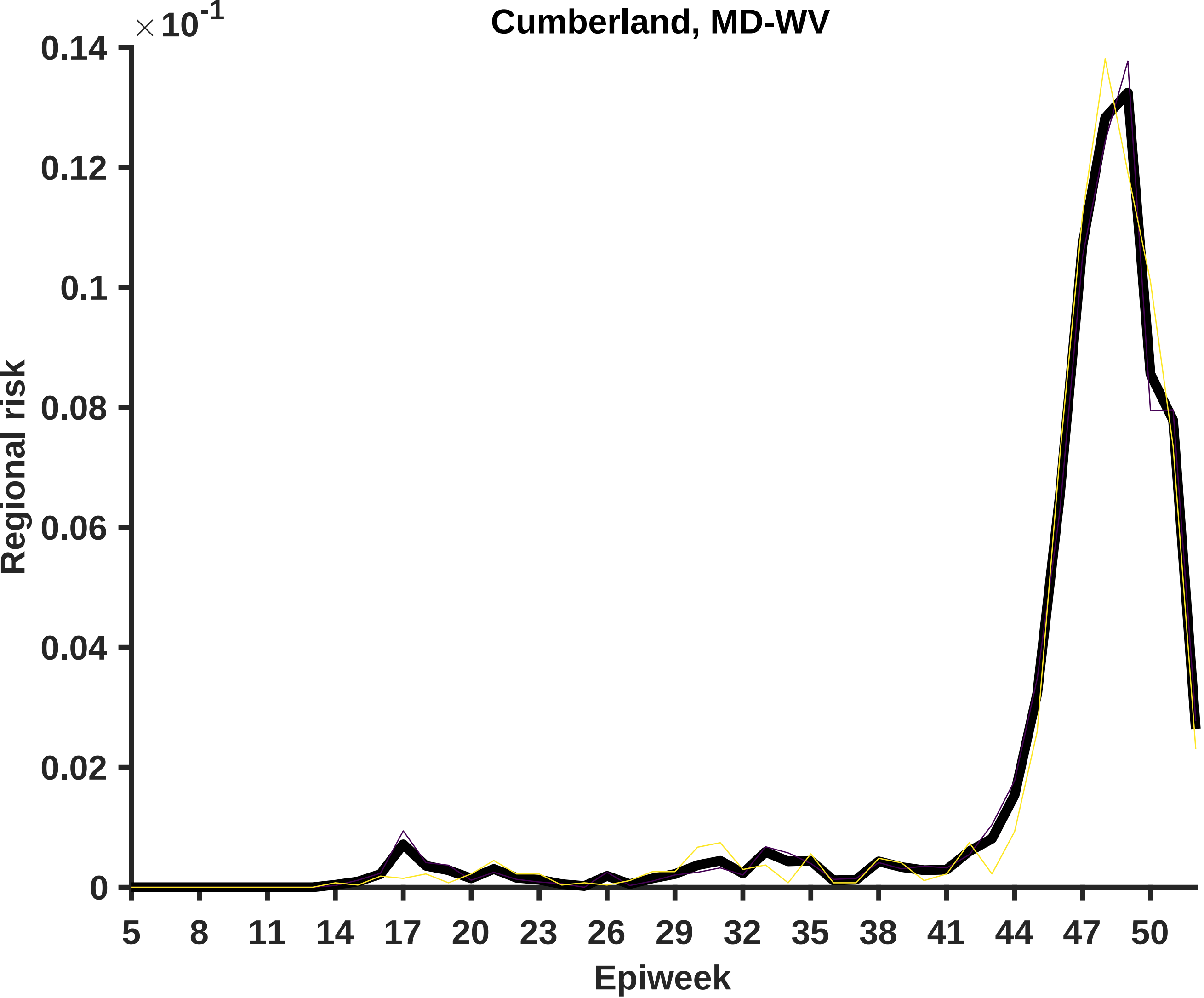
Cumberland, MD-WV
×10^-1
0.14
0.12
0.1
0.08
0.06
0.04
0.02
0
Regional risk
5
8
11
14
17
20
23
26
29
32
35
38
41
44
47
50
Epiweek

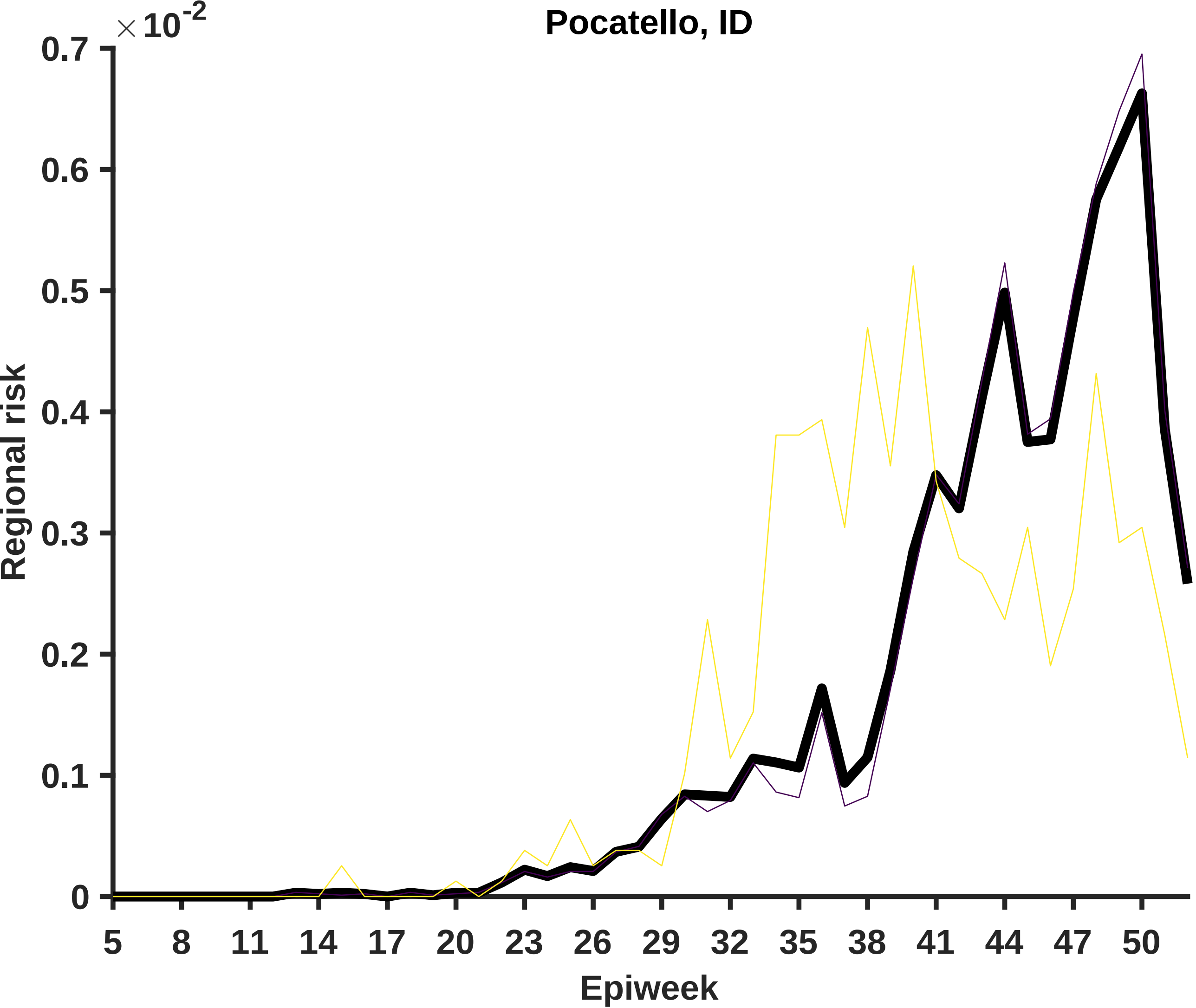
Pocatello, ID
×10^-2
0.7
0.6
0.5
0.4
0.3
0.2
0.1
0
Regional risk
5
8
11
14
17
20
23
26
29
32
35
38
41
44
47
50
Epiweek

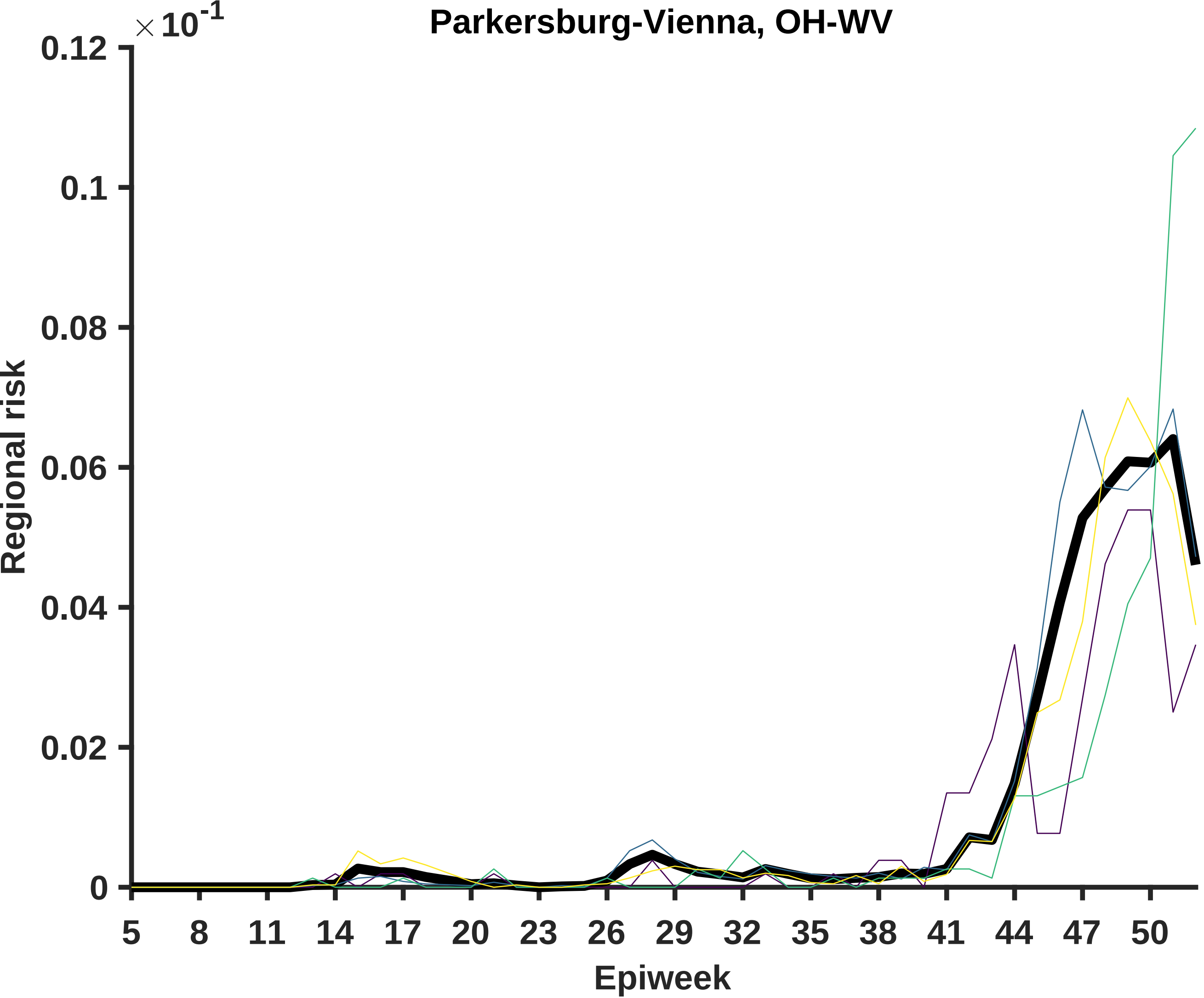

Parkersburg-Vienna, OH-WV
× 10^-1
0.12
0.1
0.08
0.06
0.04
0.02
0
Regional risk
5
8
11
14
17
20
23
26
29
32
35
38
41
44
47
50
Epiweek

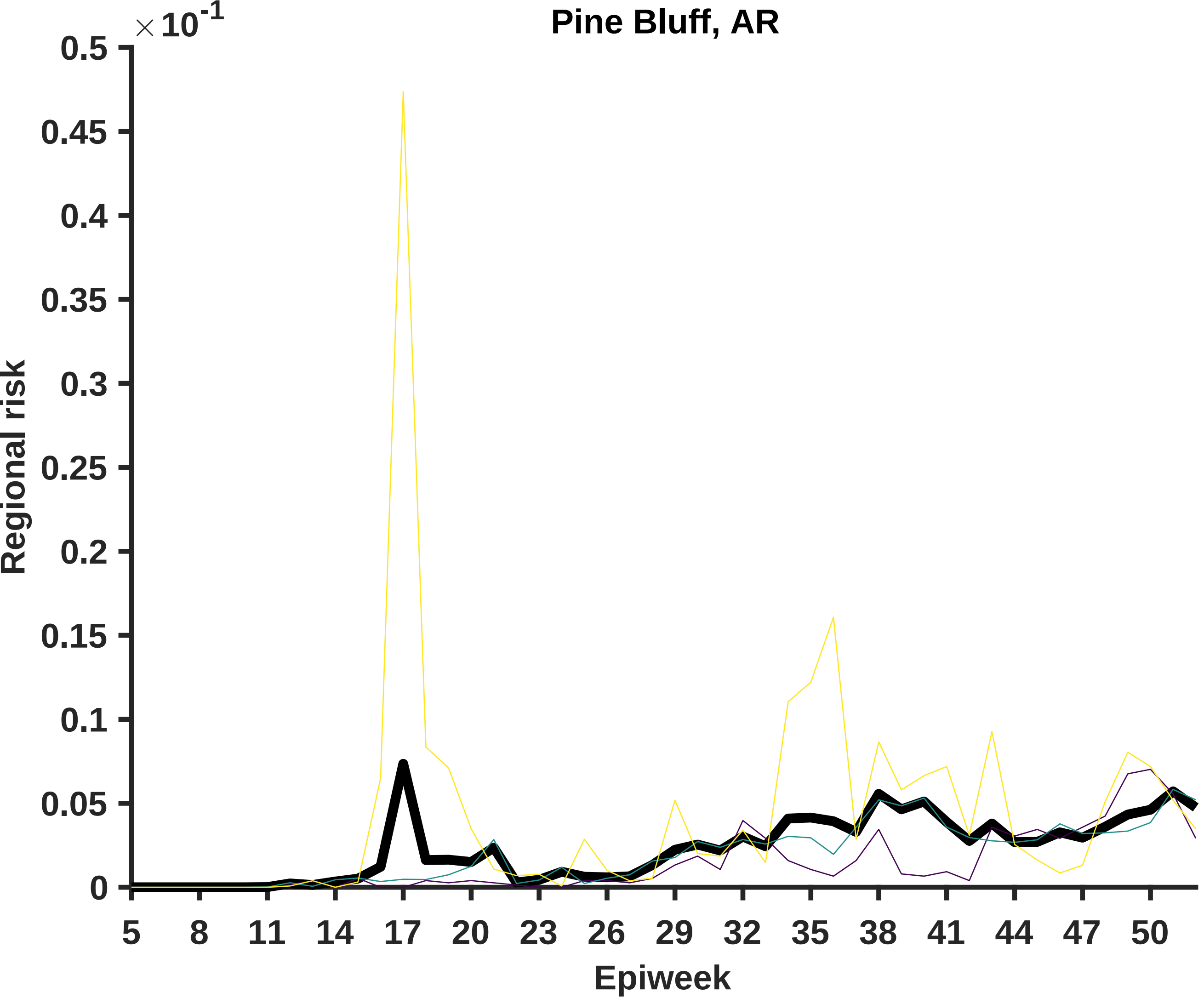

Pine Bluff, AR
×10^-1
Regional risk
0.5
0.45
0.4
0.35
0.3
0.25
0.2
0.15
0.1
0.05
0
5
8
11
14
17
20
23
26
29
32
35
38
41
44
47
50
Epiweek

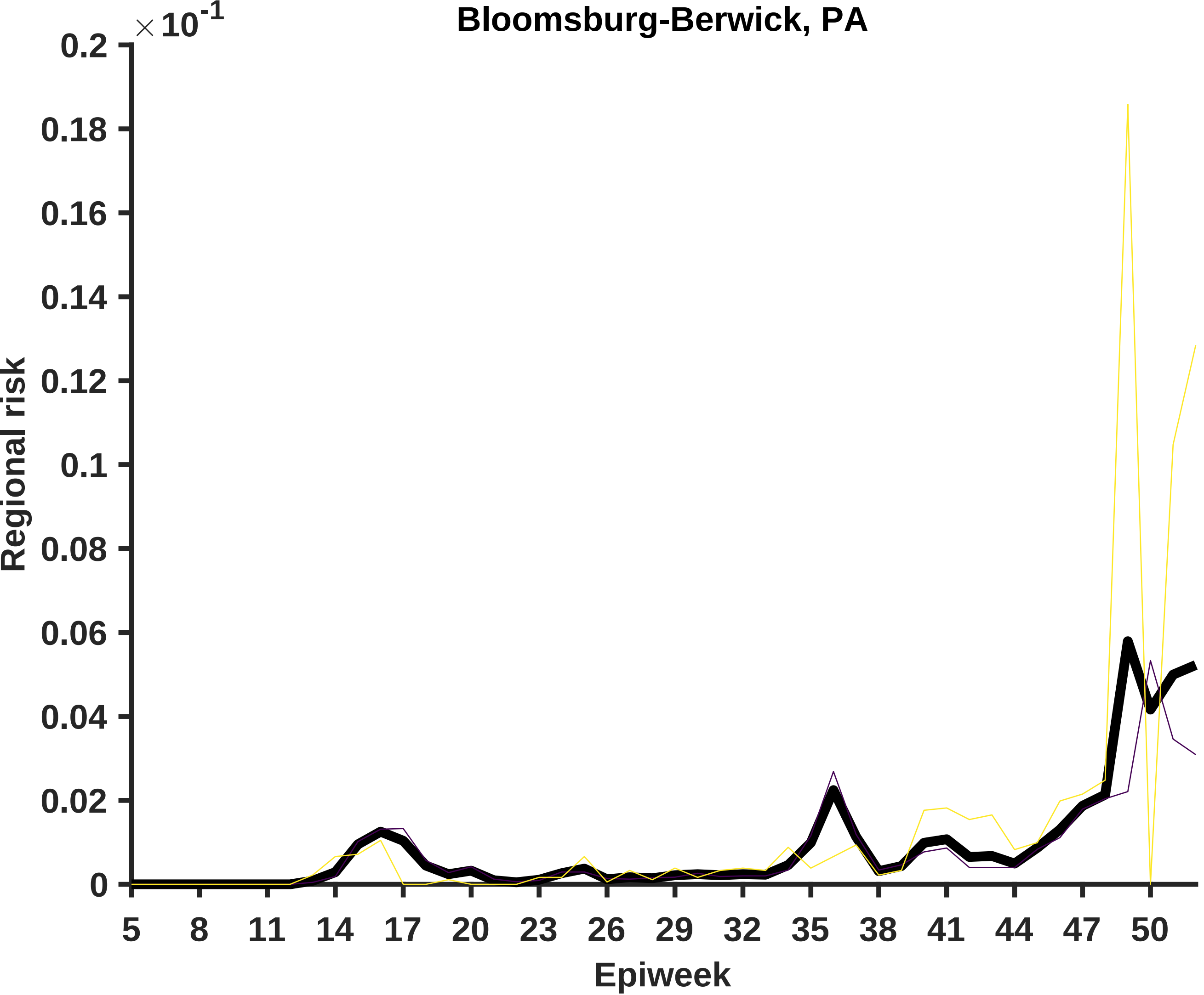
Bloomsburg-Berwick, PA
×10-1
0.2
0.18
0.16
0.14
0.12
0.1
0.08
0.06
0.04
0.02
0
Regional risk
5
8
11
14
17
20
23
26
29
32
35
38
41
44
47
50
Epiweek

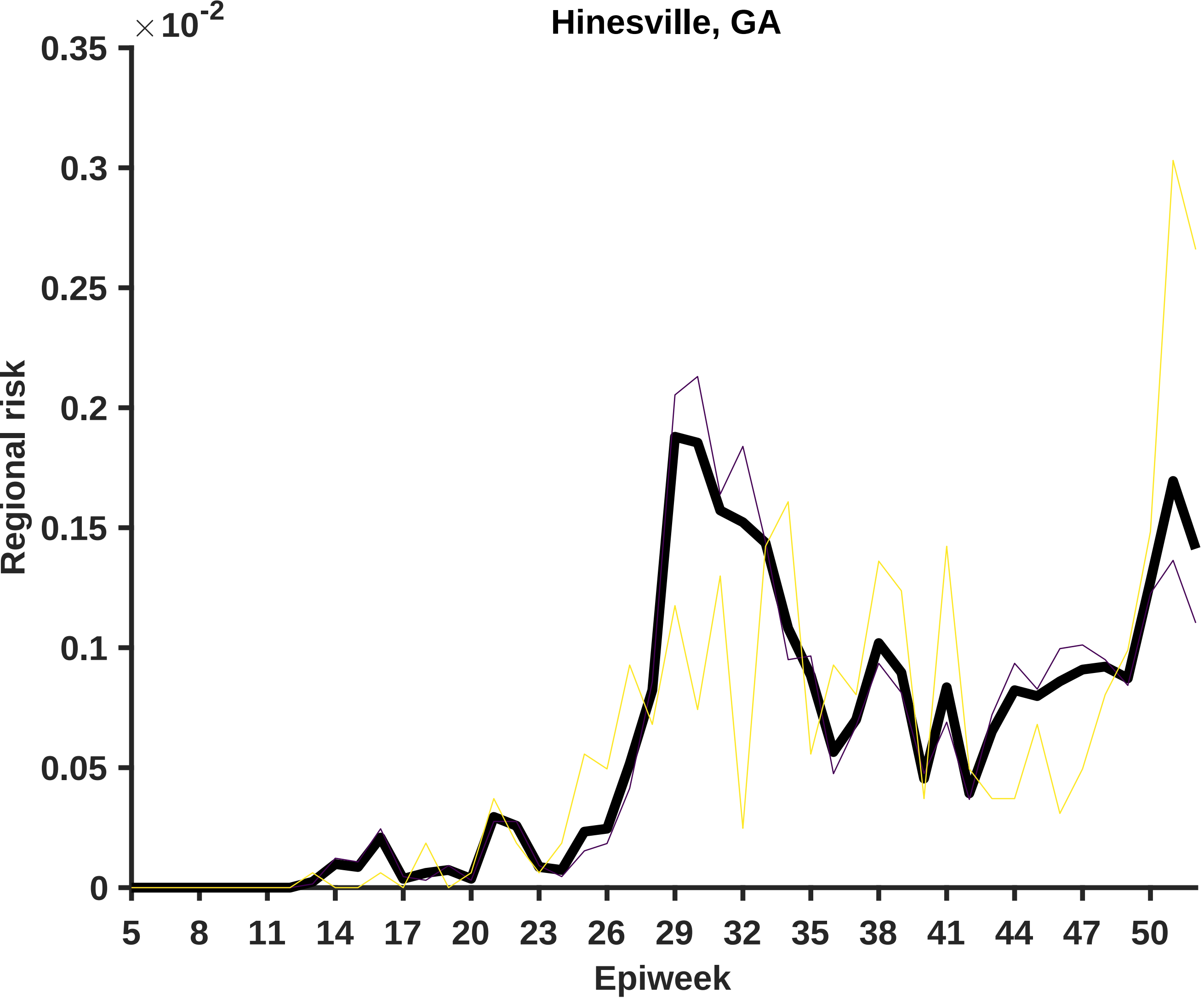

Hinesville, GA
×10^-2
0.35
0.3
0.25
0.2
0.15
0.1
0.05
0
Regional risk
5
8
11
14
17
20
23
26
29
32
35
38
41
44
47
50
Epiweek

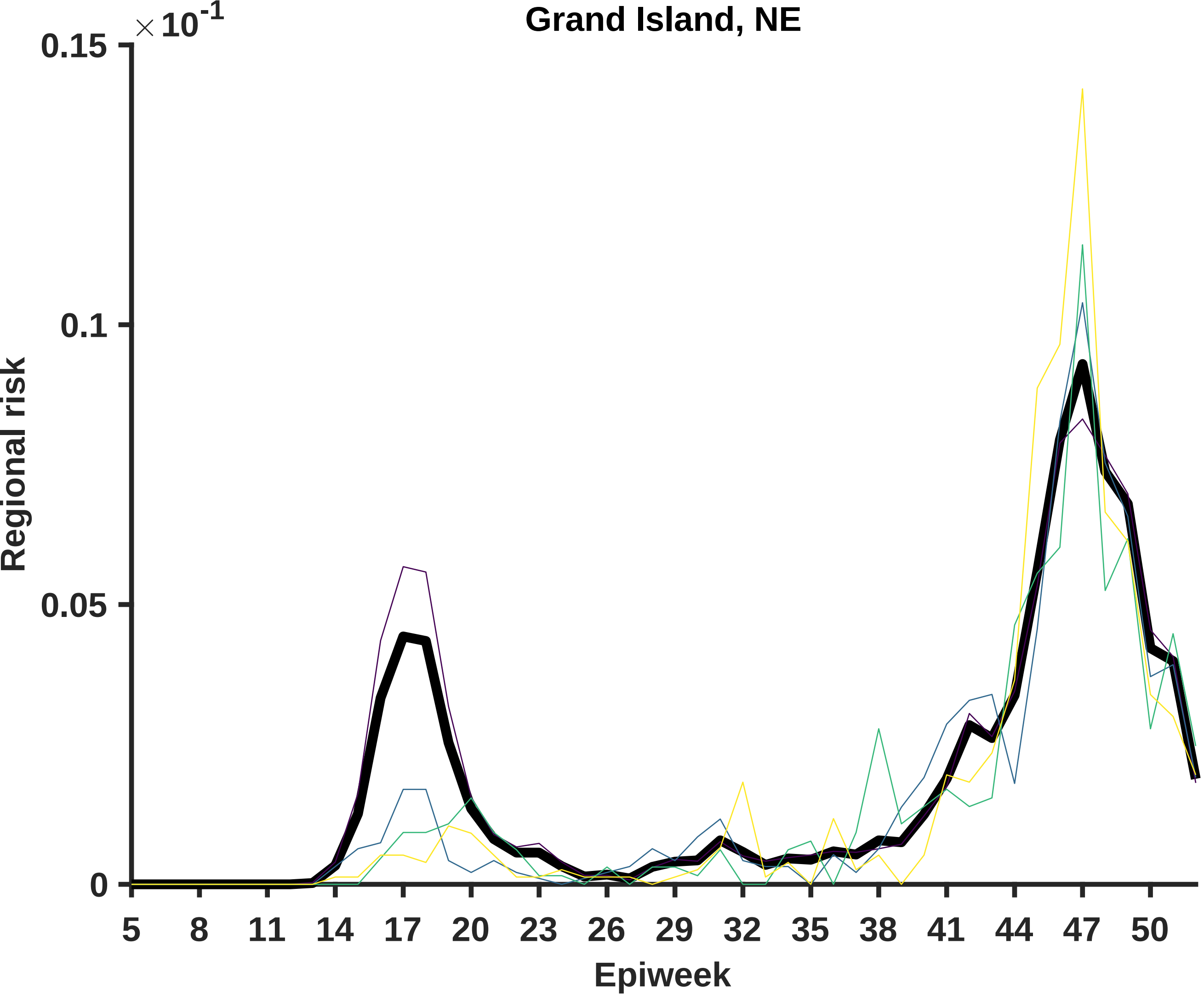
Grand Island, NE
×10-1
0.15
0.1
0.05
0
Regional risk
5
8
11
14
17
20
23
26
29
32
35
38
41
44
47
50
Epiweek

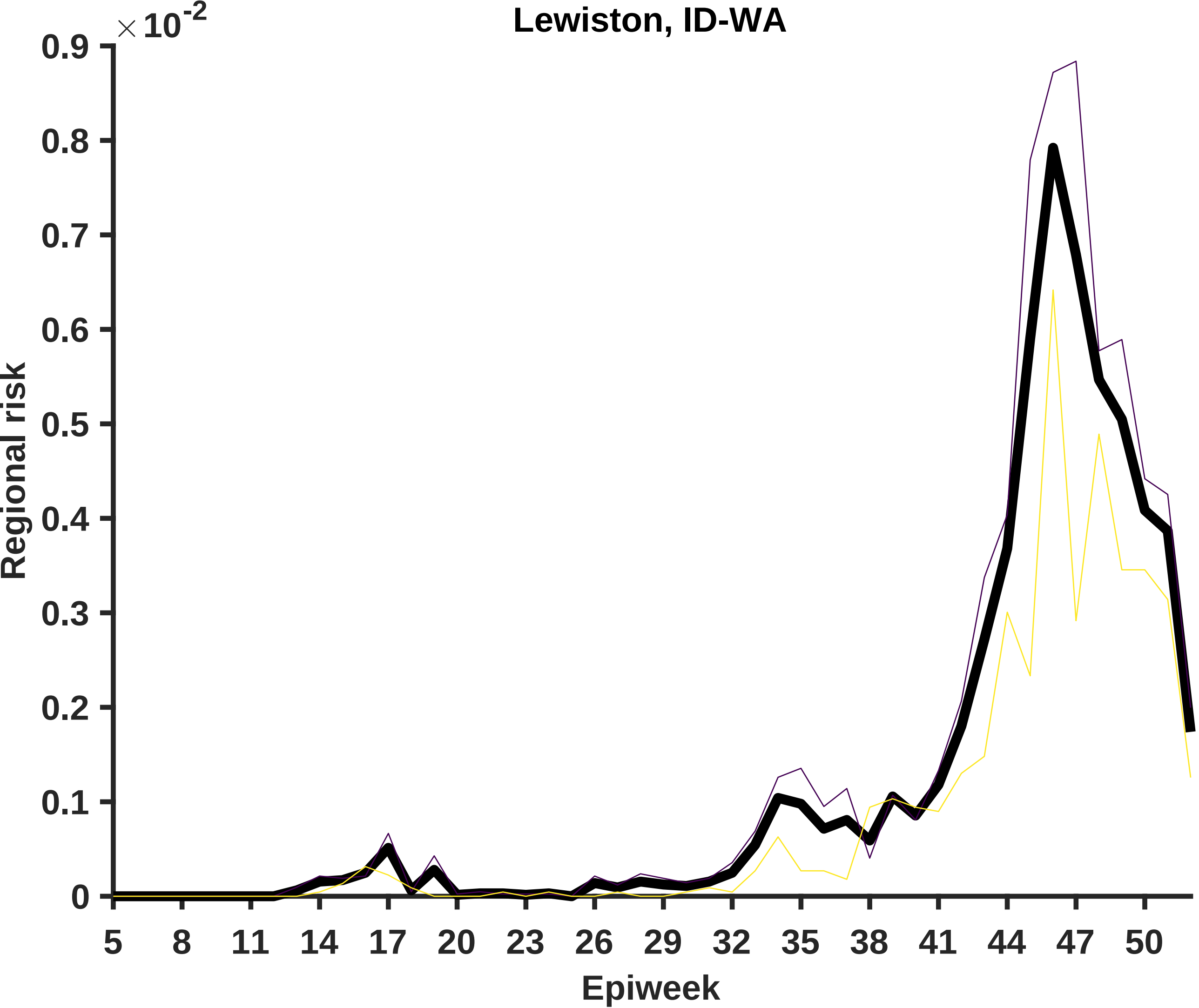

Lewiston, ID-WA
×10-2
0.9
0.8
0.7
0.6
0.5
0.4
0.3
0.2
0.1
0
Regional risk
5
8
11
14
17
20
23
26
29
32
35
38
41
44
47
50
Epiweek
