## Supplementary material for "Differential contagiousness of respiratory disease across the United States": Figure S3

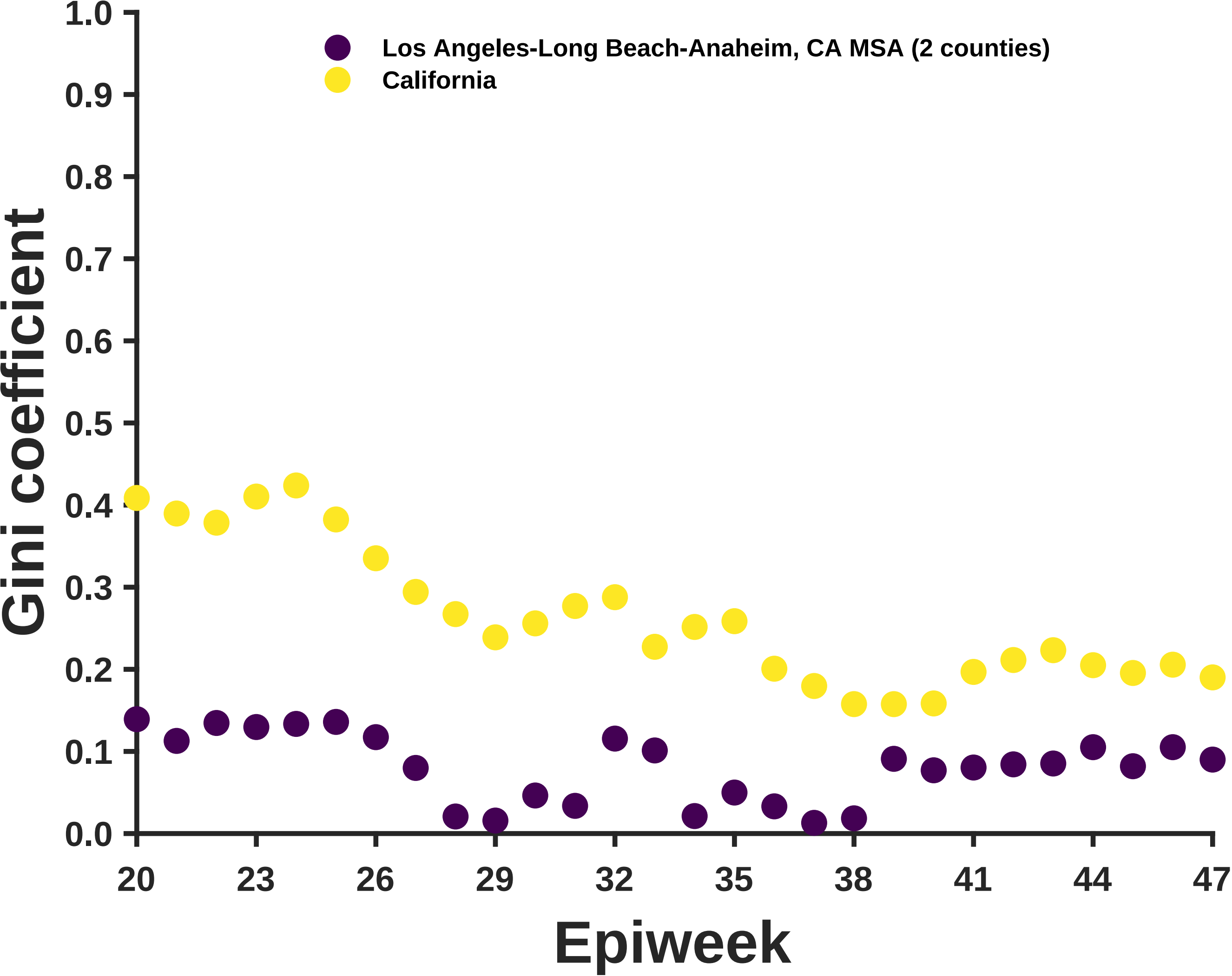
Los Angeles-Long Beach-Anaheim, CA MSA (2 counties)
California
Gini coefficient
1.0
0.9
0.8
0.7
0.6
0.5
0.4
0.3
0.2
0.1
0.0
20
23
26
29
32
35
38
41
44
47
Epiweek

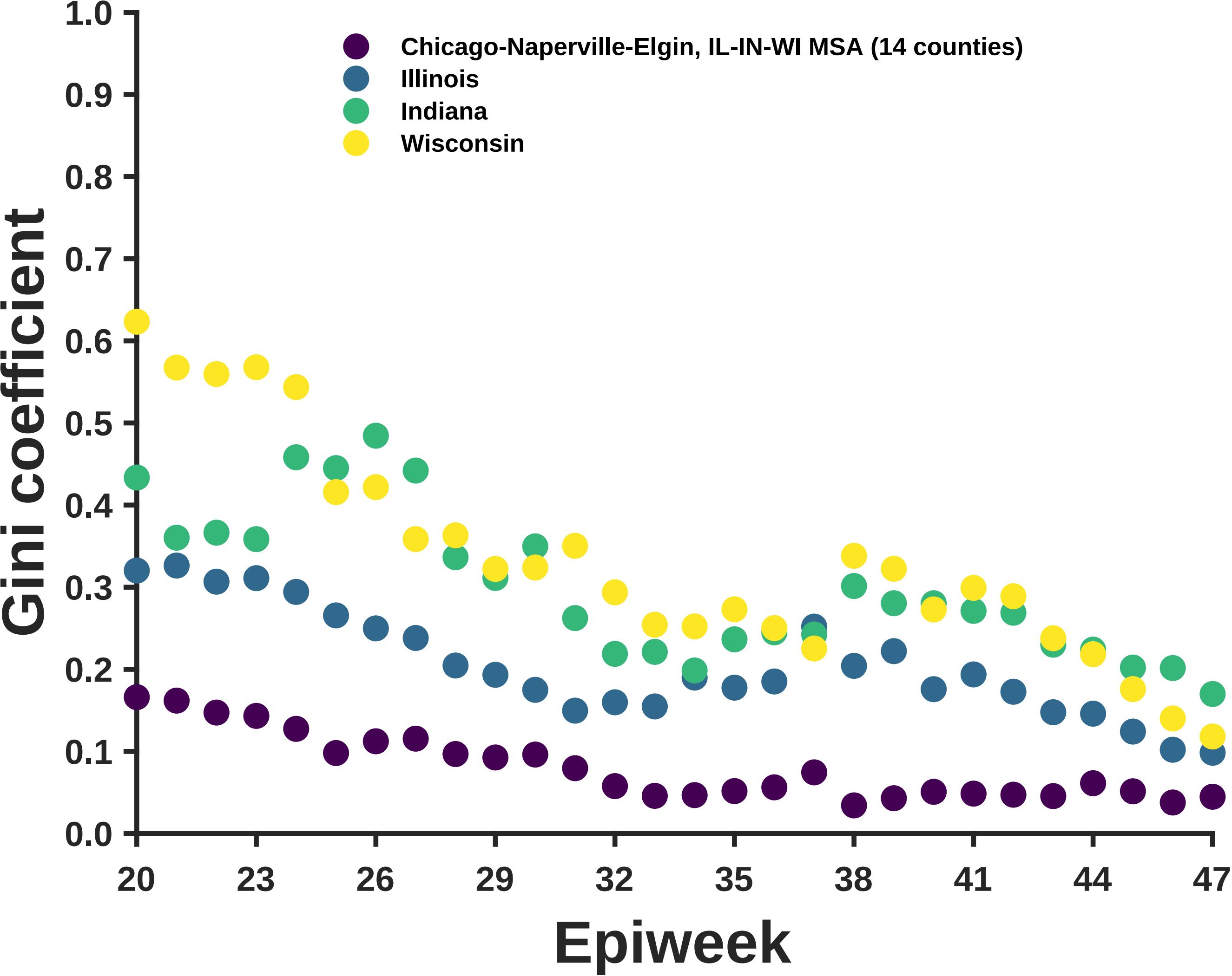
Chicago-Naperville-Elgin, IL-IN-WI MSA (14 counties)
Illinois
Indiana
Wisconsin
Gini coefficient
1.0
0.9
0.8
0.7
0.6
0.5
0.4
0.3
0.2
0.1
0.0
20
23
26
29
32
35
38
41
44
47
Epiweek

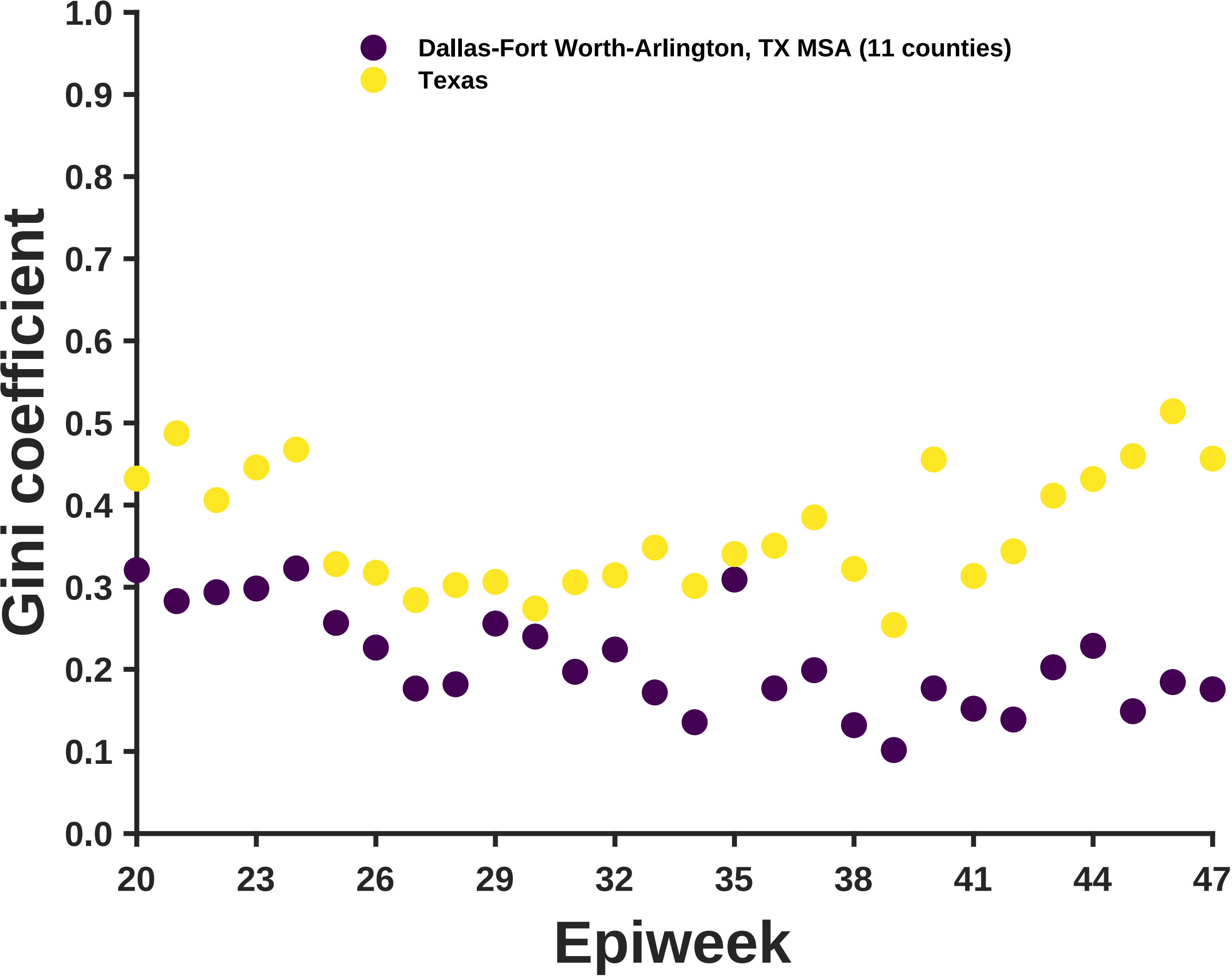

Dallas-Fort Worth-Arlington, TX MSA (11 counties)
Texas
Gini coefficient
1.0
0.9
0.8
0.7
0.6
0.5
0.4
0.3
0.2
0.1
0.0
20
23
26
29
32
35
38
41
44
47
Epiweek

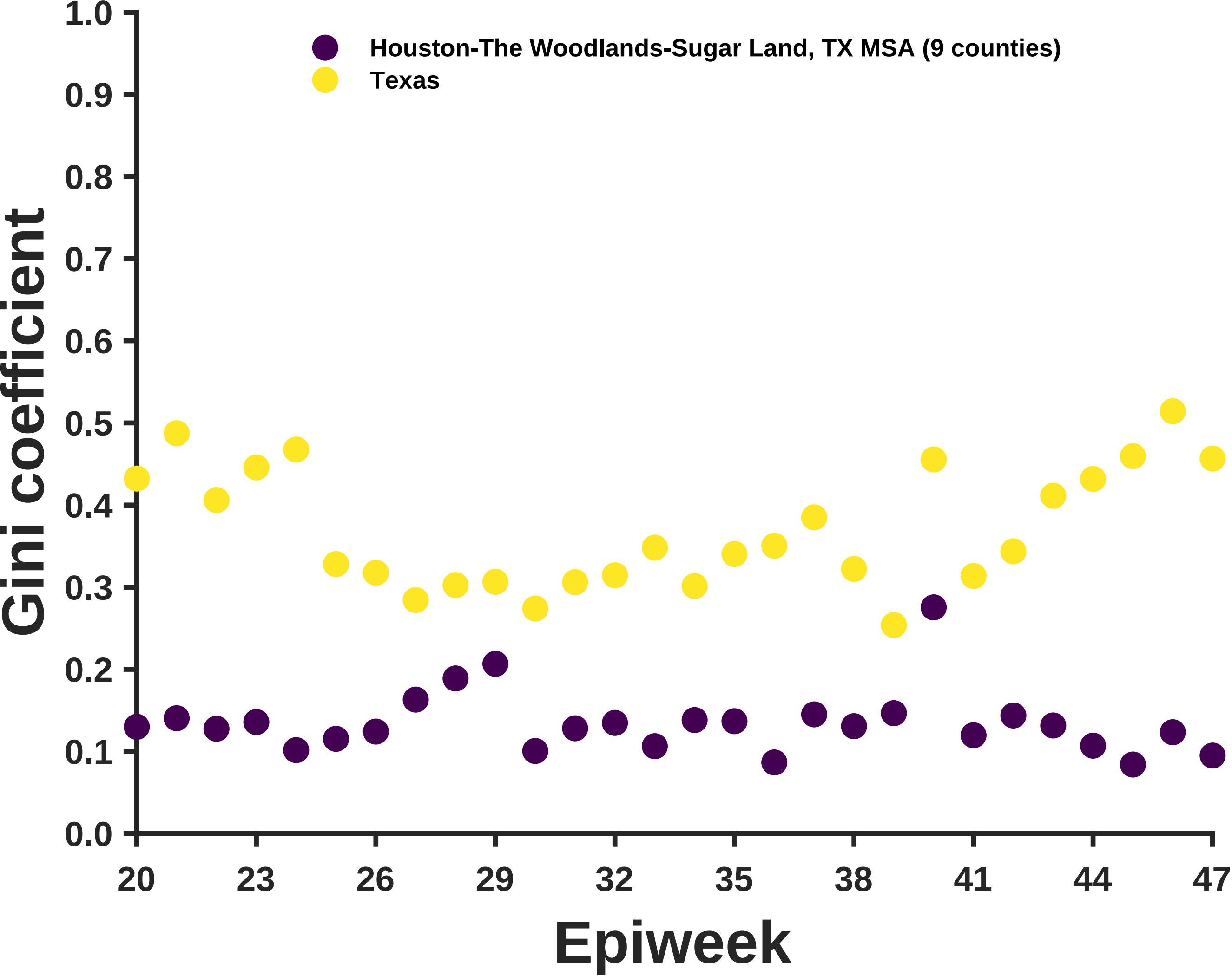

Houston-The Woodlands-Sugar Land, TX MSA (9 counties)
Texas
Gini coefficient
1.0
0.9
0.8
0.7
0.6
0.5
0.4
0.3
0.2
0.1
0.0
20
23
26
29
32
35
38
41
44
47
Epiweek

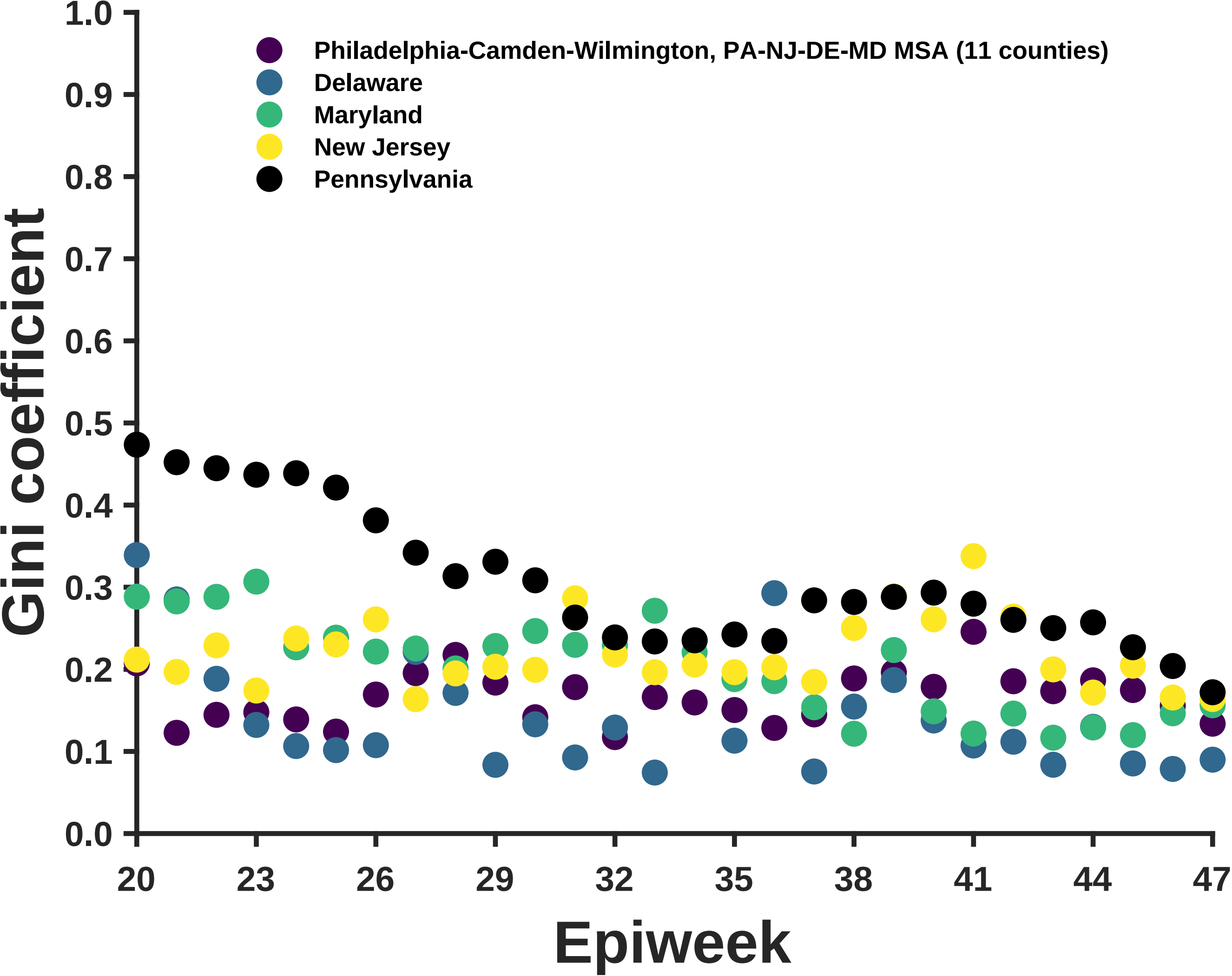
Philadelphia-Camden-Wilmington, PA-NJ-DE-MD MSA (11 counties)
Delaware
Maryland
New Jersey
Pennsylvania
Gini coefficient
1.0
0.9
0.8
0.7
0.6
0.5
0.4
0.3
0.2
0.1
0.0
20
23
26
29
32
35
38
41
44
47
Epiweek

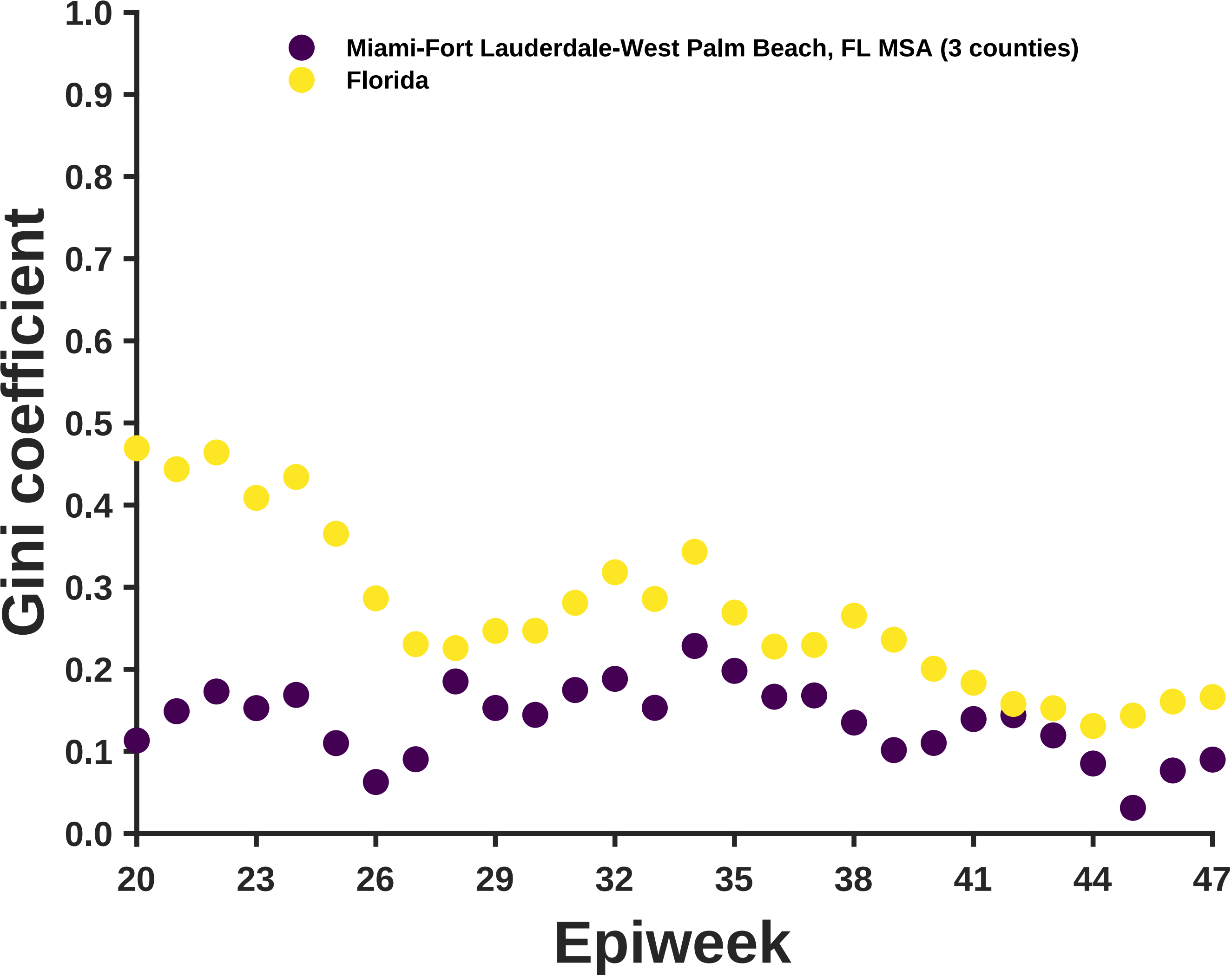

Miami-Fort Lauderdale-West Palm Beach, FL MSA (3 counties)
Florida
Gini coefficient
1.0
0.9
0.8
0.7
0.6
0.5
0.4
0.3
0.2
0.1
0.0
20
23
26
29
32
35
38
41
44
47
Epiweek

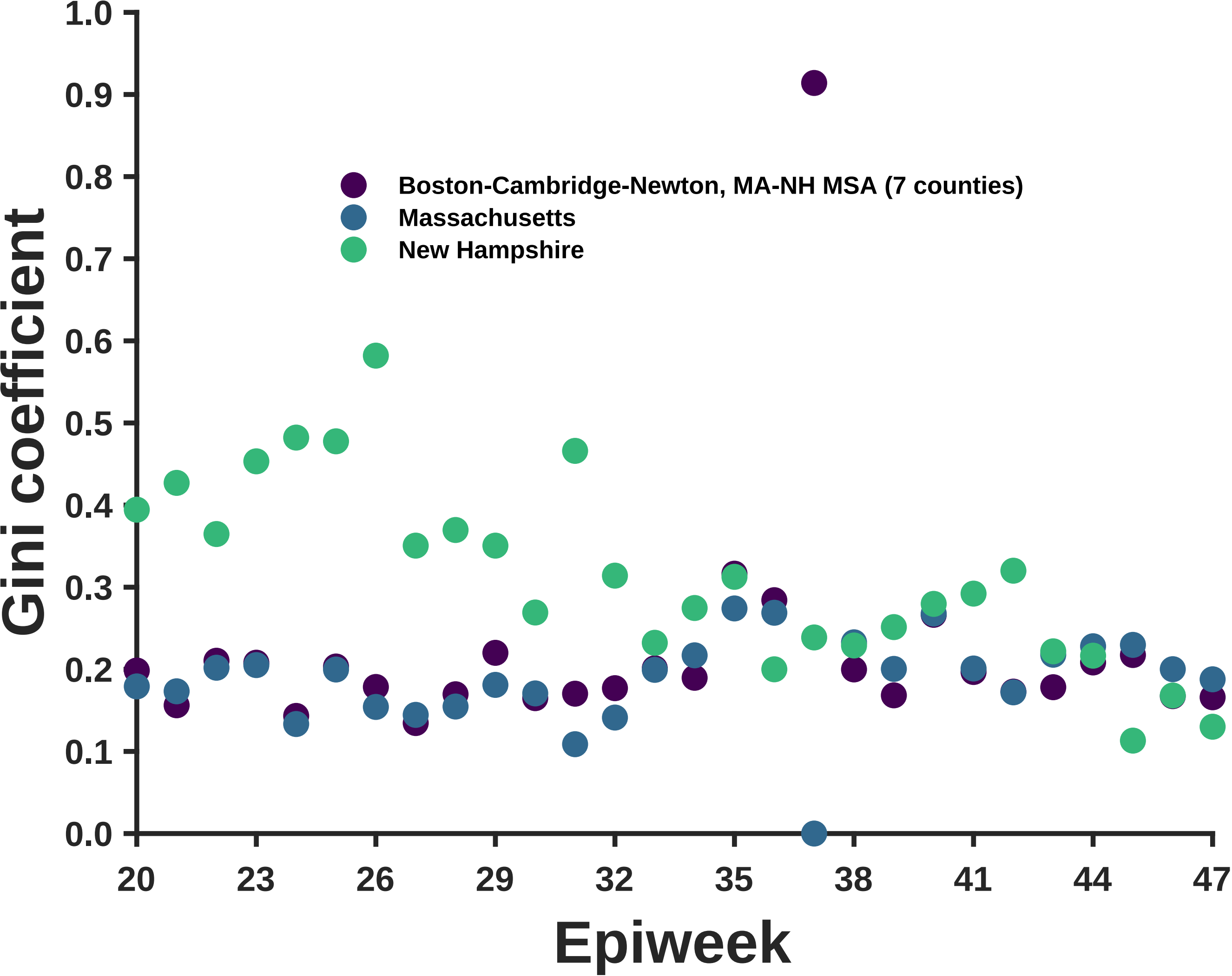
Boston-Cambridge-Newton, MA-NH MSA (7 counties)
Massachusetts
New Hampshire
Gini coefficient
1.0
0.9
0.8
0.7
0.6
0.5
0.4
0.3
0.2
0.1
0.0
20
23
26
29
32
35
38
41
44
47
Epiweek

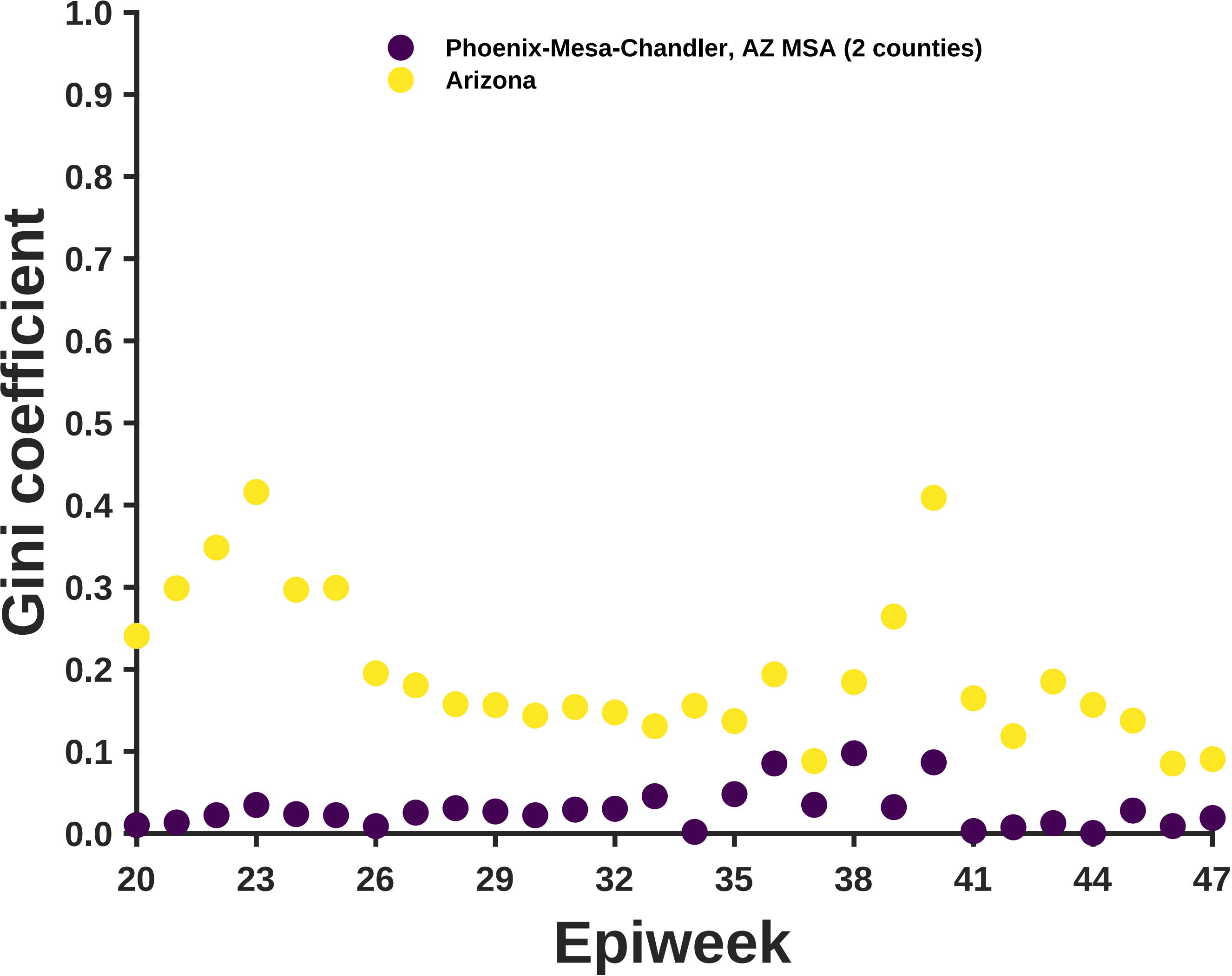
Phoenix-Mesa-Chandler, AZ MSA (2 counties)
Arizona
Gini coefficient
Epiweek
1.0
0.9
0.8
0.7
0.6
0.5
0.4
0.3
0.2
0.1
0.0
20
23
26
29
32
35
38
41
44
47

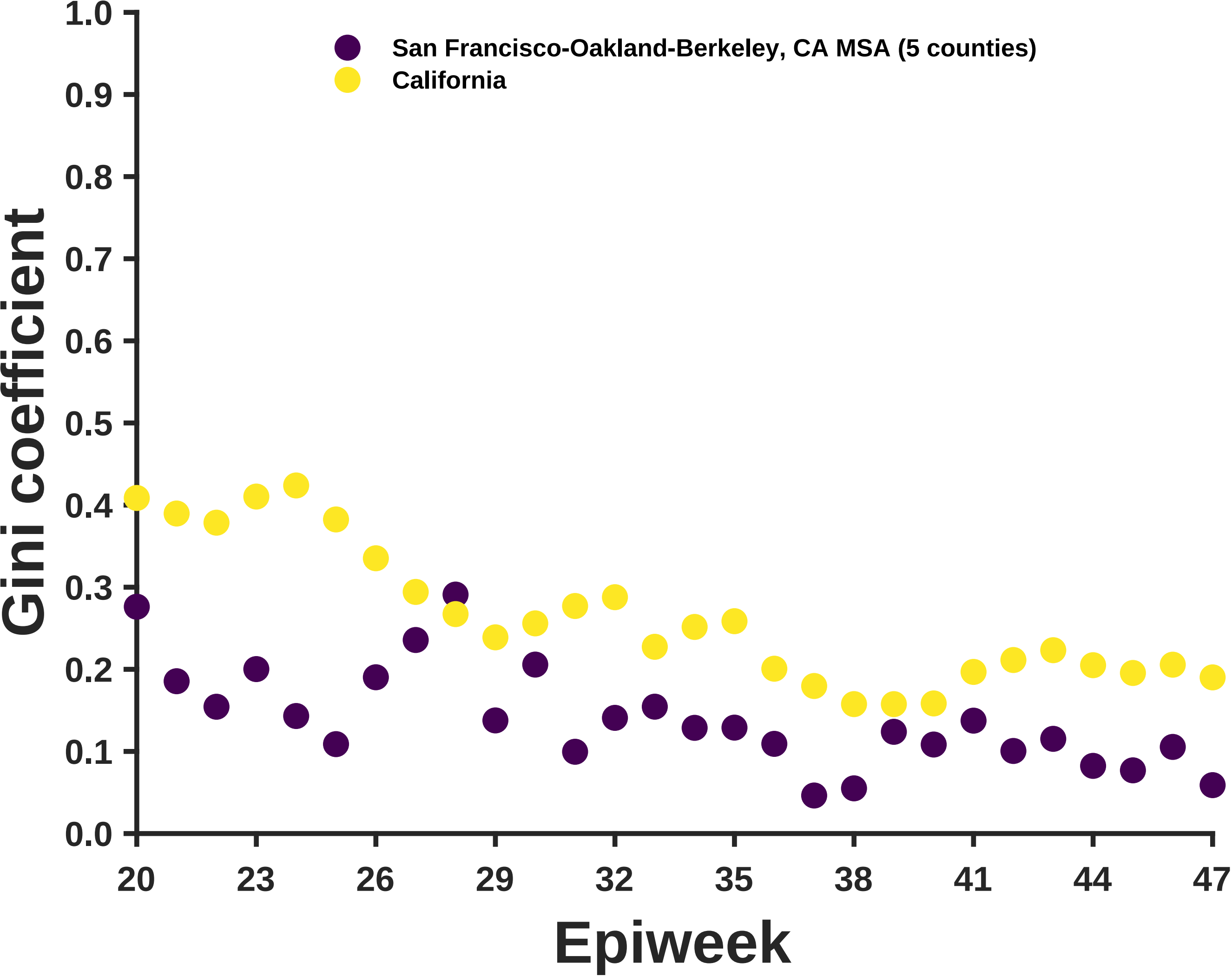
San Francisco-Oakland-Berkeley, CA MSA (5 counties)
California
Gini coefficient
0.0
0.1
0.2
0.3
0.4
0.5
0.6
0.7
0.8
0.9
1.0
20
23
26
29
32
35
38
41
44
47
Epiweek

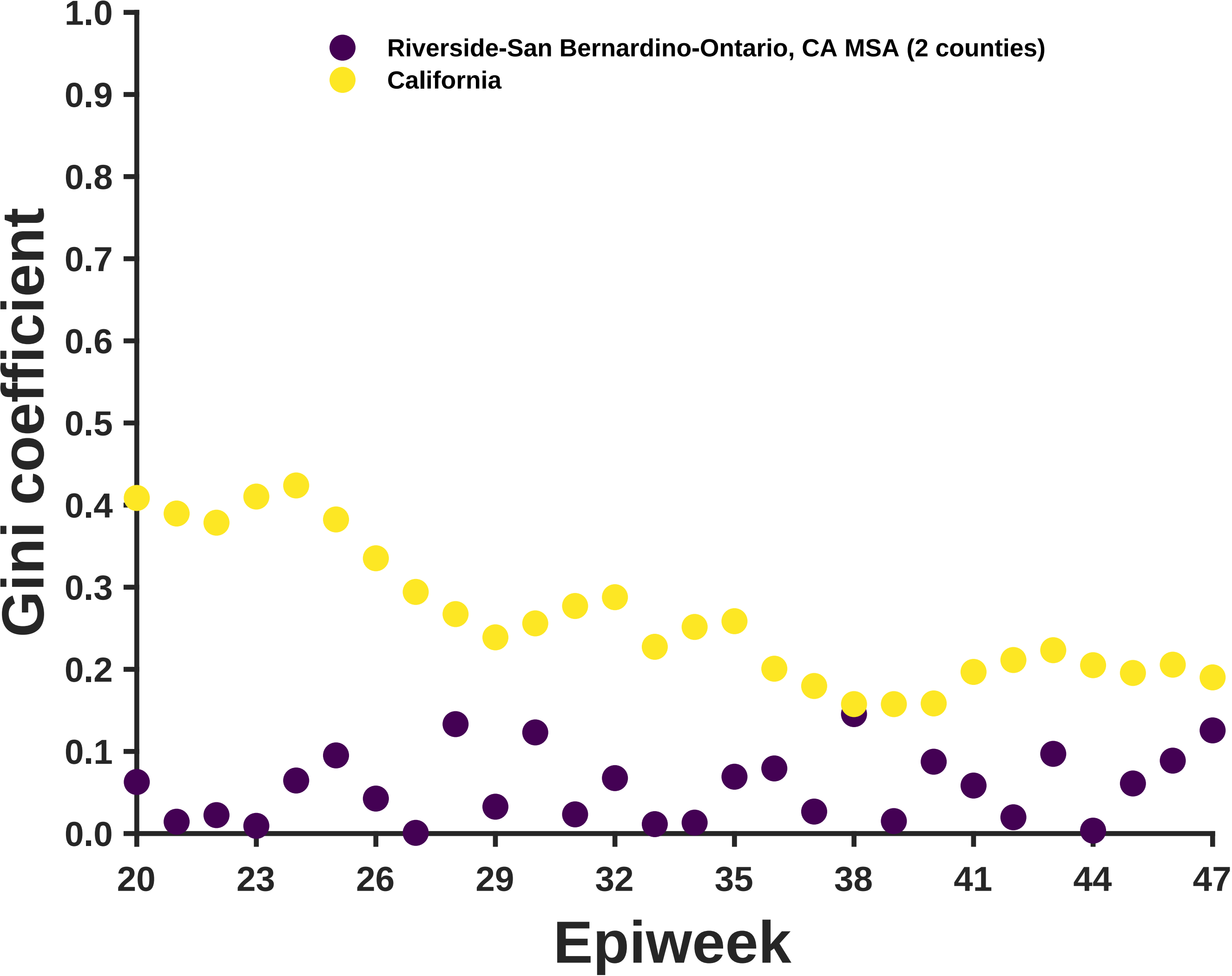

Riverside-San Bernardino-Ontario, CA MSA (2 counties)
California
Gini coefficient
1.0
0.9
0.8
0.7
0.6
0.5
0.4
0.3
0.2
0.1
0.0
20
23
26
29
32
35
38
41
44
47
Epiweek

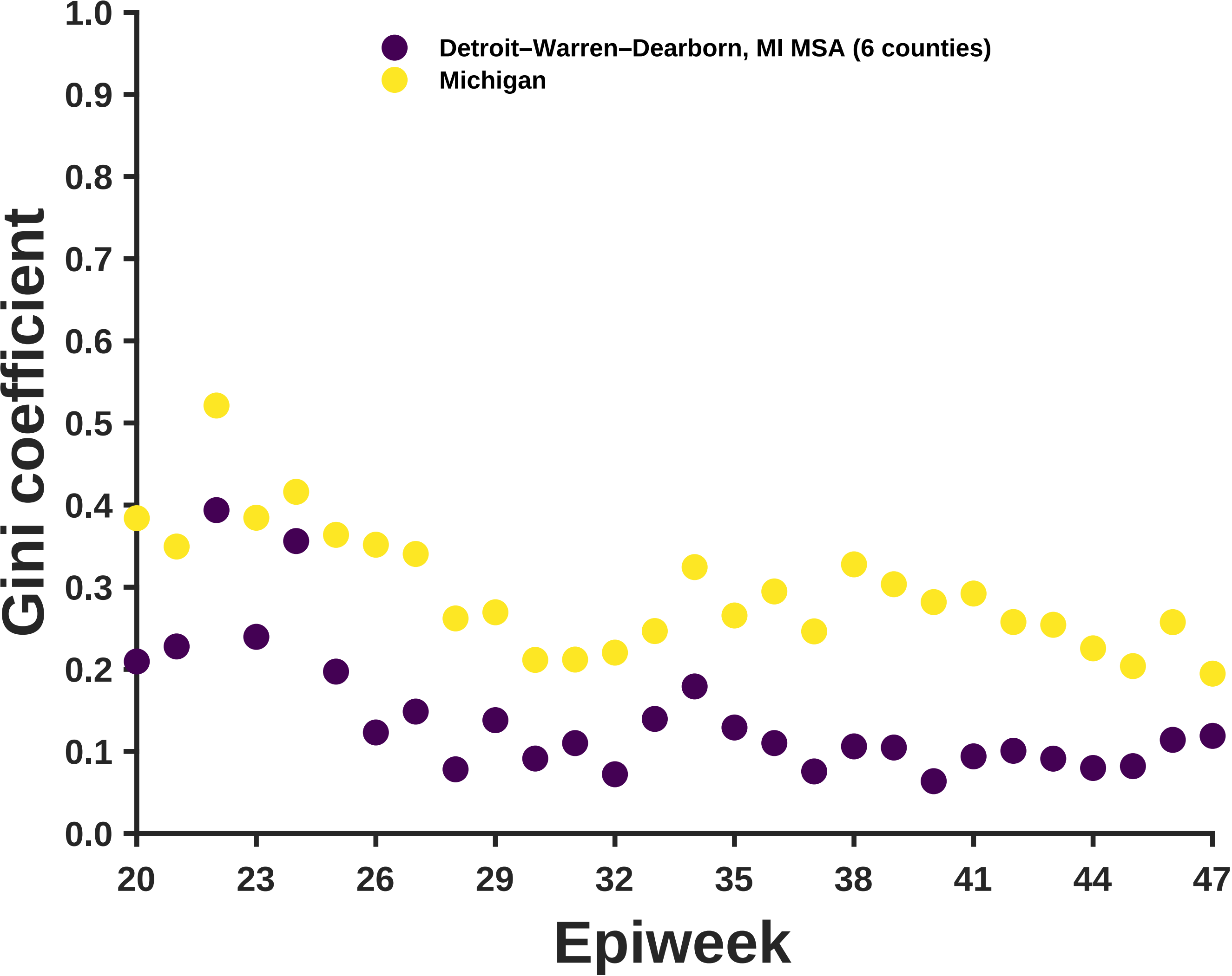

Detroit–Warren–Dearborn, MI MSA (6 counties)
Michigan
Gini coefficient
Epiweek
1.0
0.9
0.8
0.7
0.6
0.5
0.4
0.3
0.2
0.1
0.0
20
23
26
29
32
35
38
41
44
47

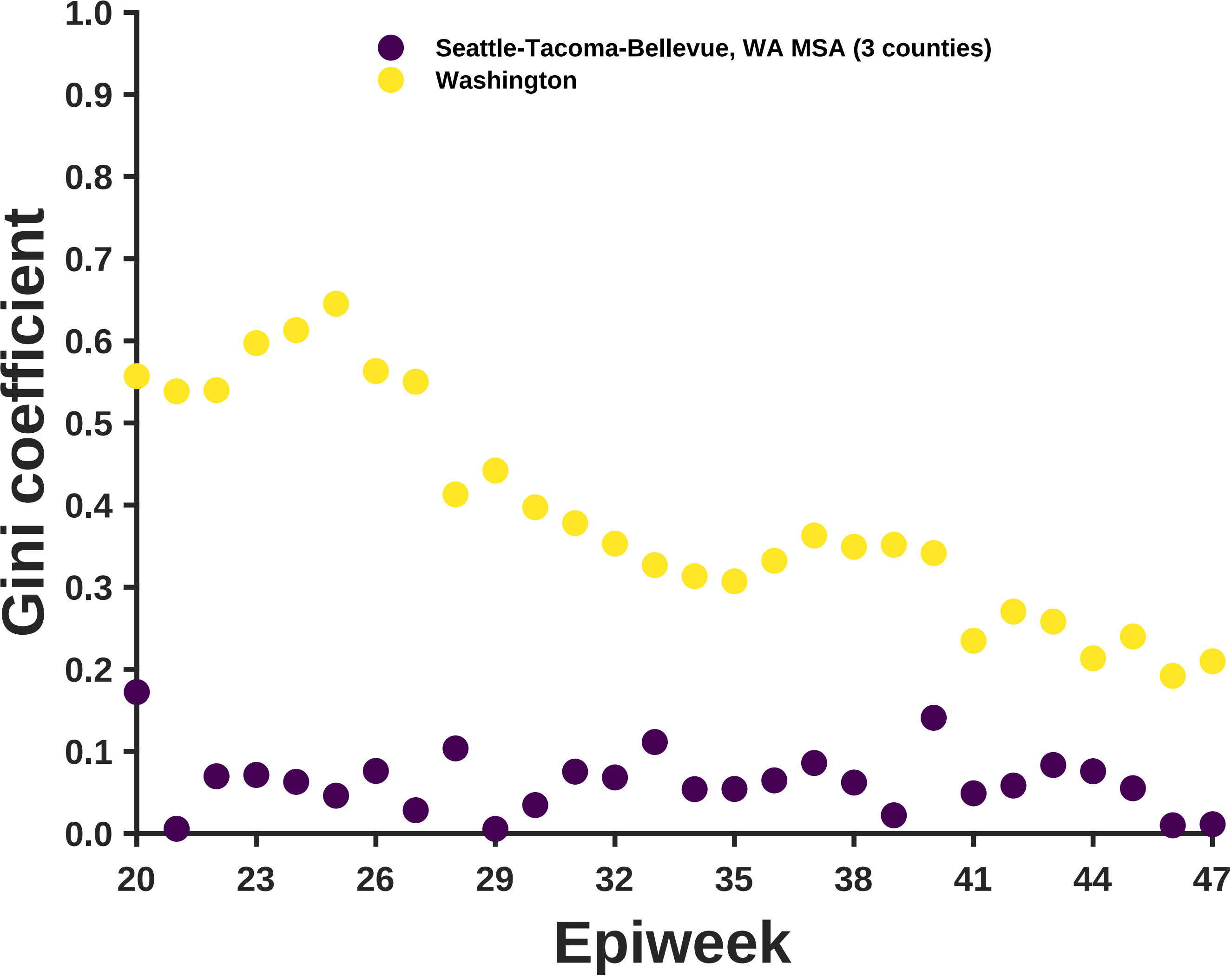

Seattle-Tacoma-Bellevue, WA MSA (3 counties)
Washington
Gini coefficient
Epiweek
0.0
0.1
0.2
0.3
0.4
0.5
0.6
0.7
0.8
0.9
1.0
20
23
26
29
32
35
38
41
44
47

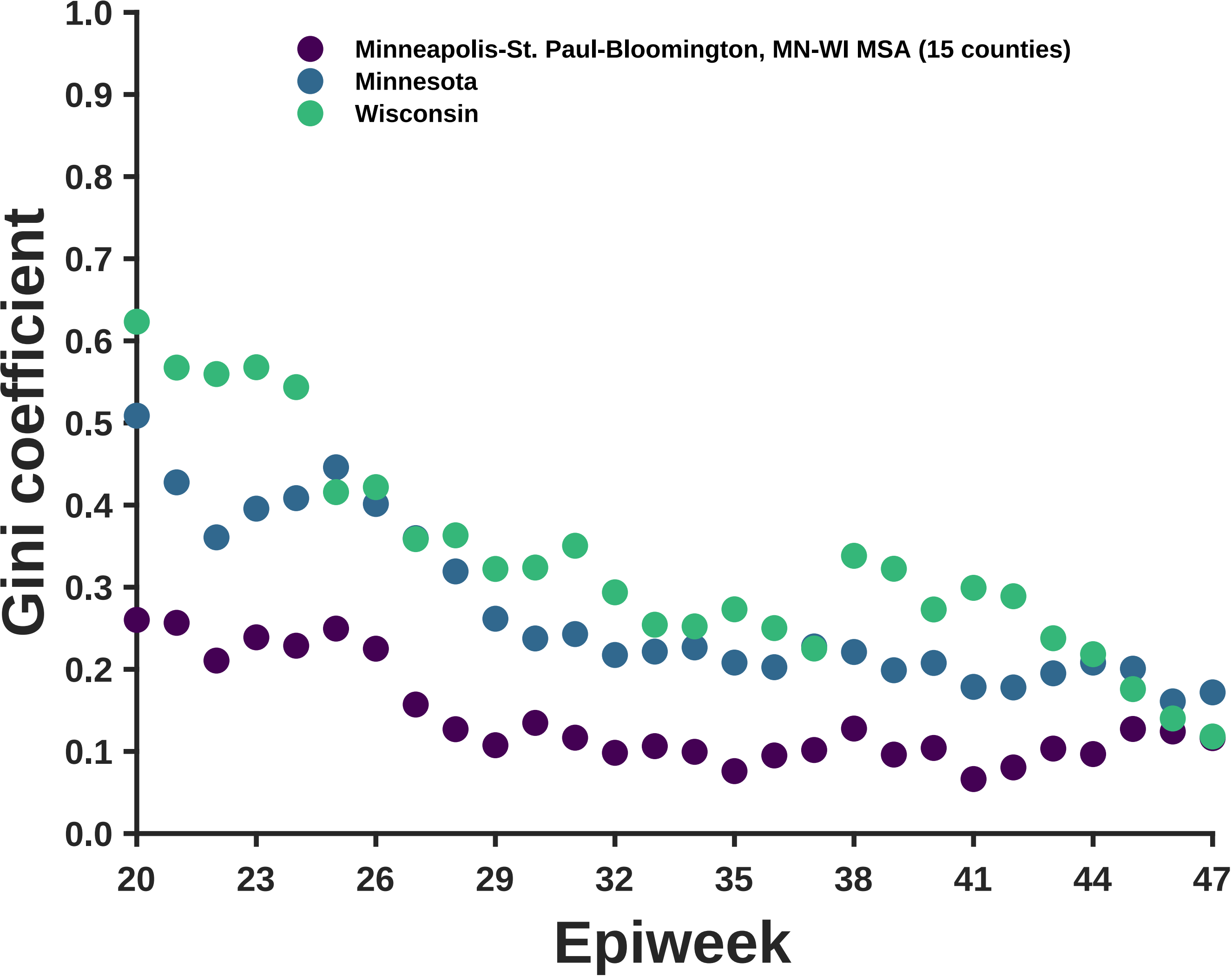
Minneapolis-St. Paul-Bloomington, MN-WI MSA (15 counties)
Minnesota
Wisconsin
Gini coefficient
Epiweek
0.0
0.1
0.2
0.3
0.4
0.5
0.6
0.7
0.8
0.9
1.0
20
23
26
29
32
35
38
41
44
47

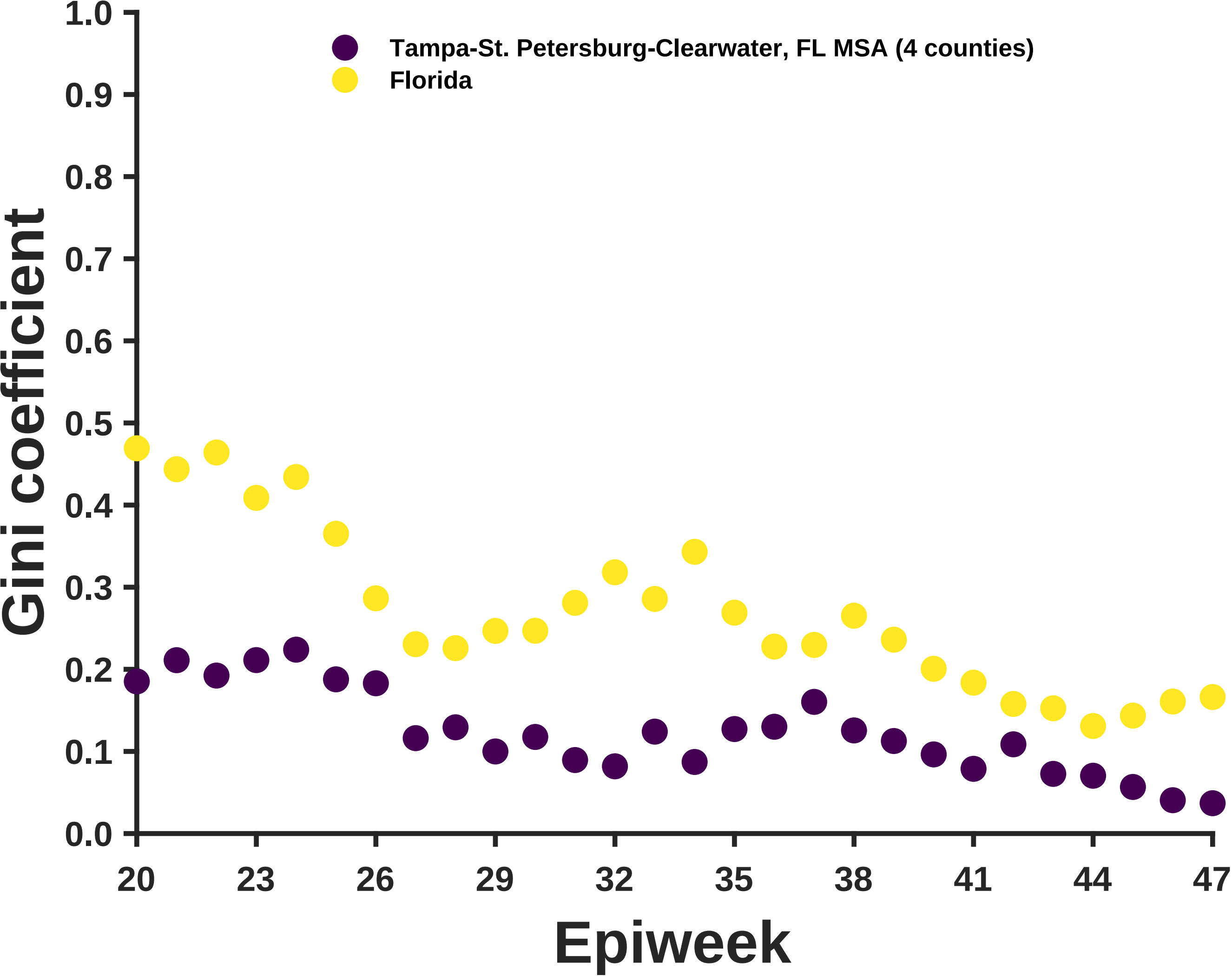

Tampa-St. Petersburg-Clearwater, FL MSA (4 counties)
Florida
Gini coefficient
1.0
0.9
0.8
0.7
0.6
0.5
0.4
0.3
0.2
0.1
0.0
20
23
26
29
32
35
38
41
44
47
Epiweek

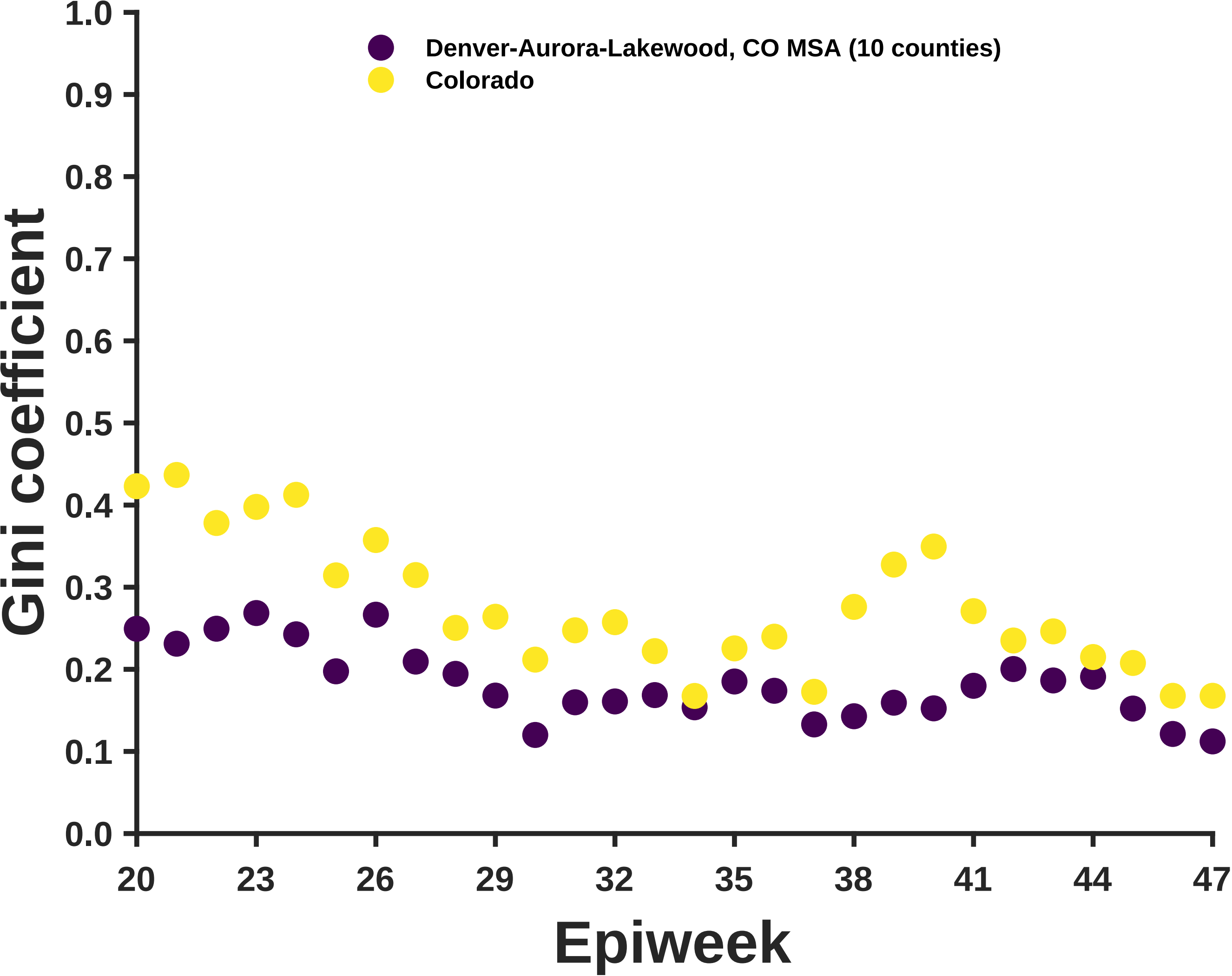

Denver-Aurora-Lakewood, CO MSA (10 counties)
Colorado
Gini coefficient
1.0
0.9
0.8
0.7
0.6
0.5
0.4
0.3
0.2
0.1
0.0
20
23
26
29
32
35
38
41
44
47
Epiweek

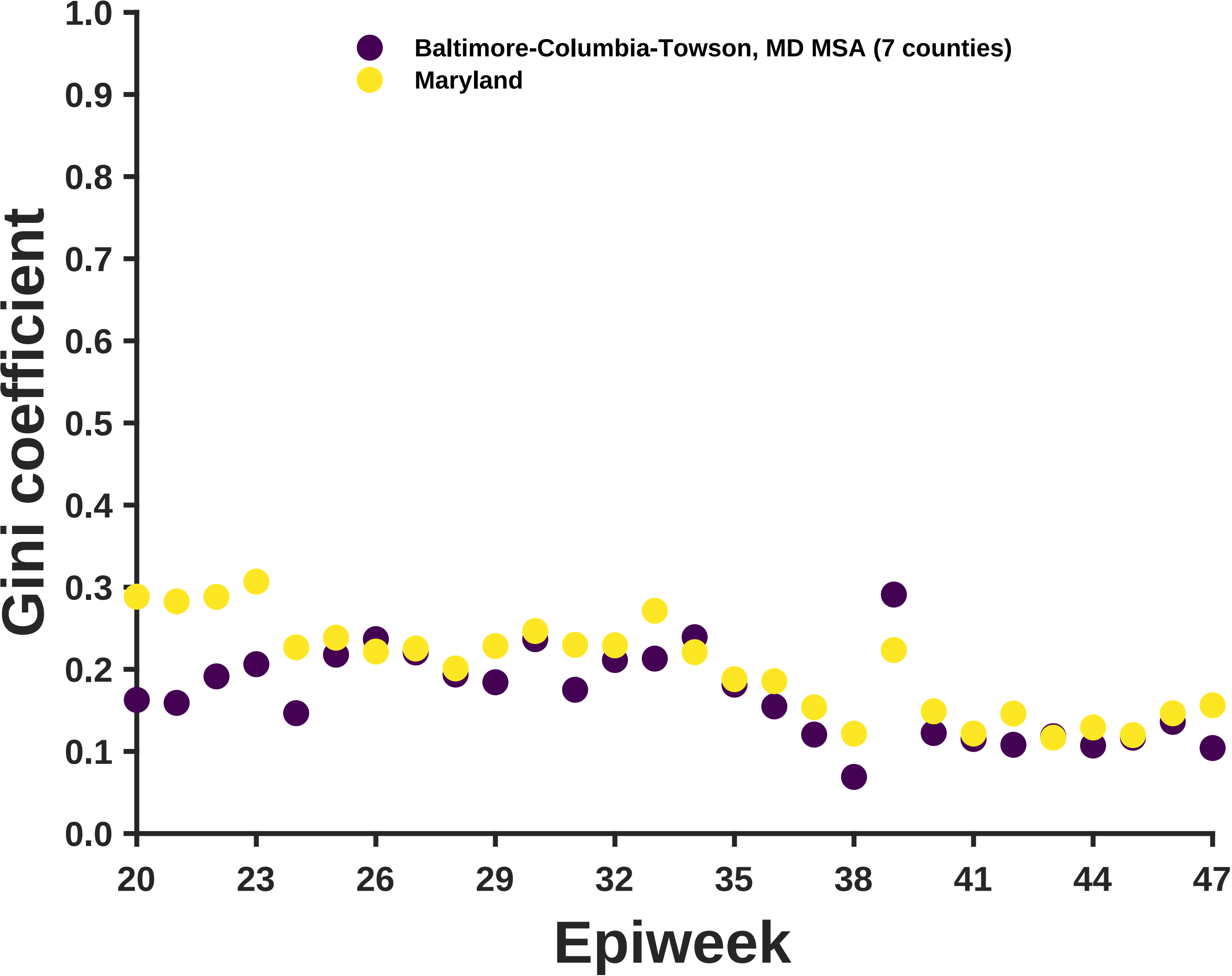
Baltimore-Columbia-Towson, MD MSA (7 counties)
Maryland
Gini coefficient
1.0
0.9
0.8
0.7
0.6
0.5
0.4
0.3
0.2
0.1
0.0
20
23
26
29
32
35
38
41
44
47
Epiweek

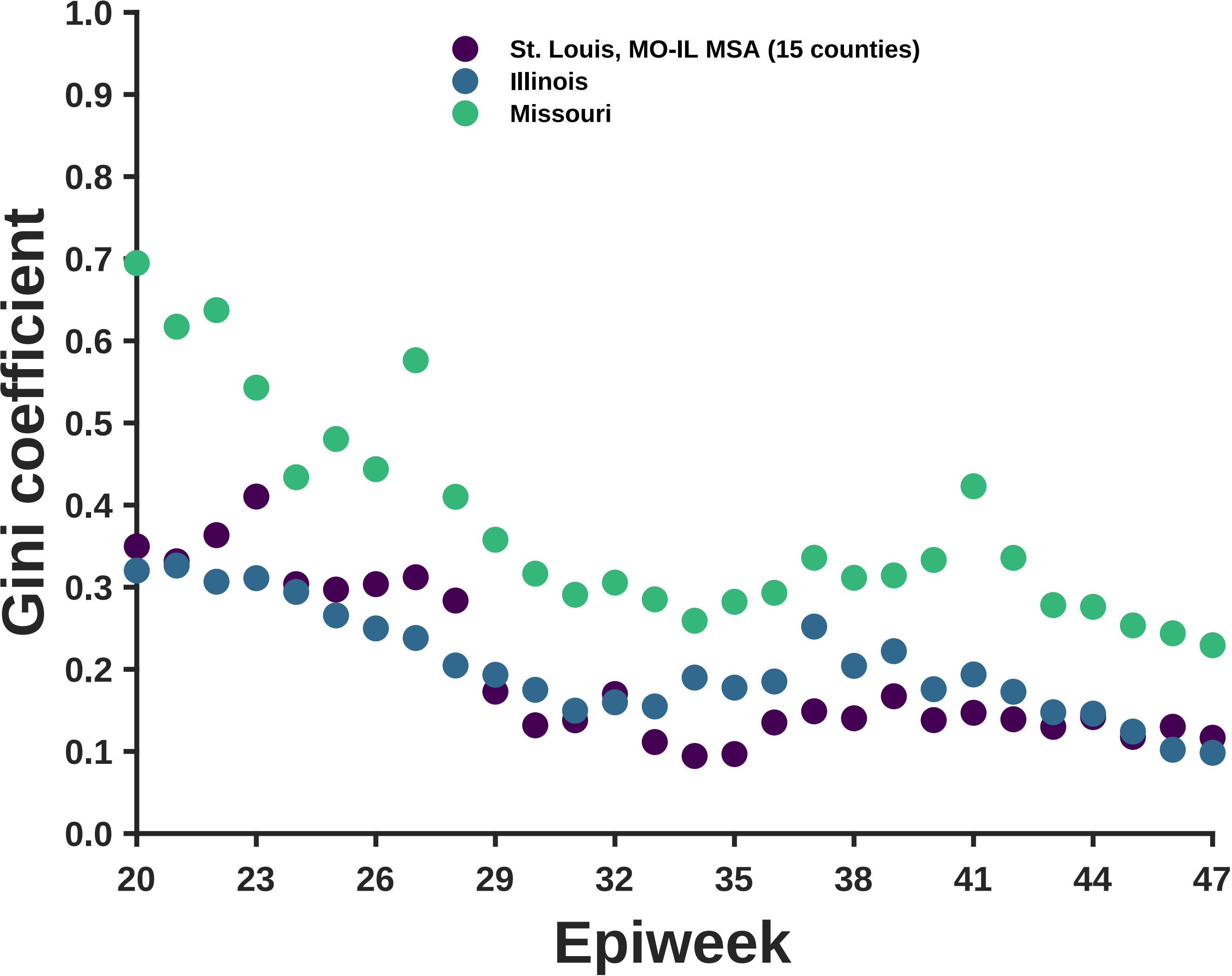

St. Louis, MO-IL MSA (15 counties)
Illinois
Missouri
Gini coefficient
1.0
0.9
0.8
0.7
0.6
0.5
0.4
0.3
0.2
0.1
0.0
20
23
26
29
32
35
38
41
44
47
Epiweek

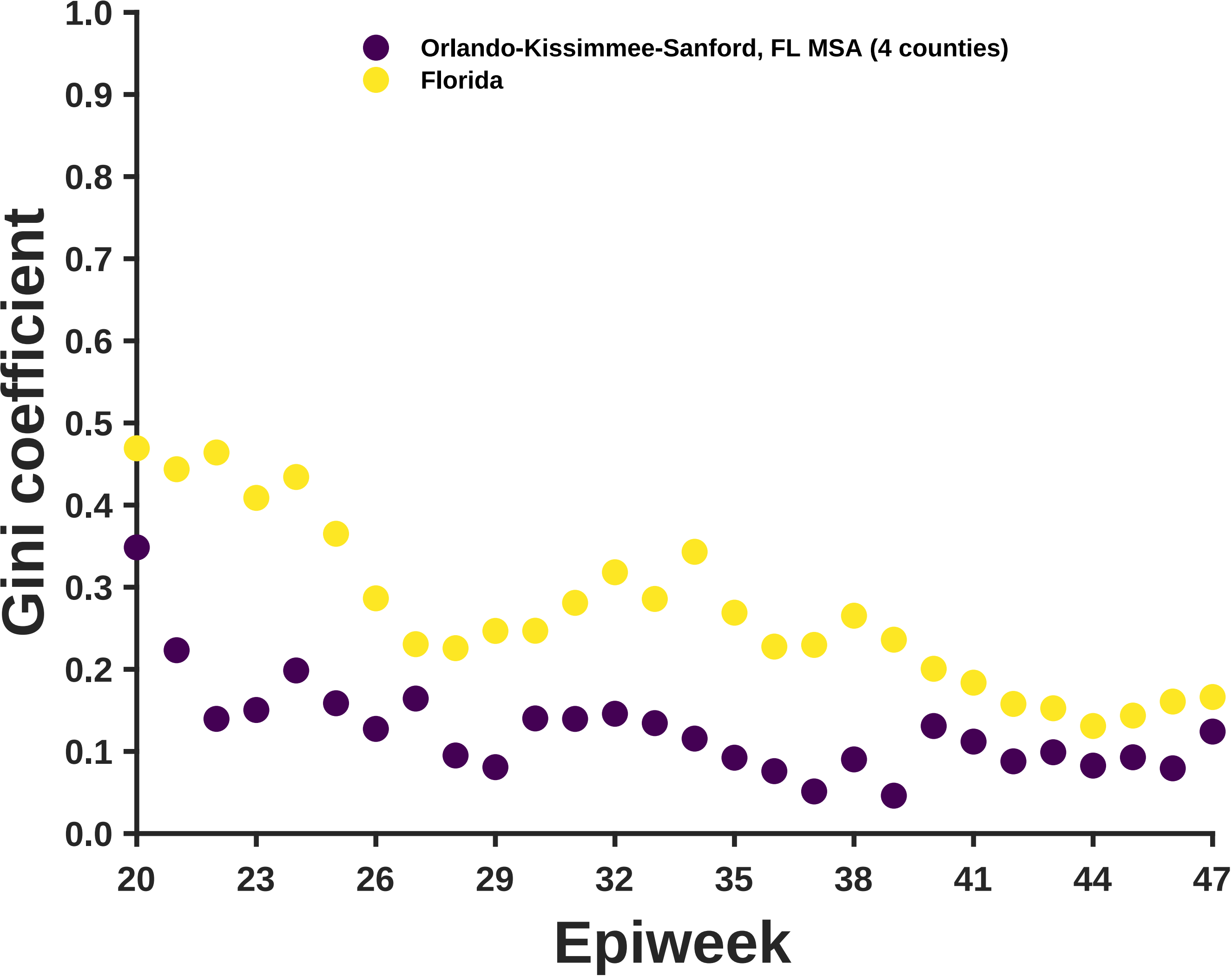

Orlando-Kissimmee-Sanford, FL MSA (4 counties)
Florida
Gini coefficient
1.0
0.9
0.8
0.7
0.6
0.5
0.4
0.3
0.2
0.1
0.0
20
23
26
29
32
35
38
41
44
47
Epiweek

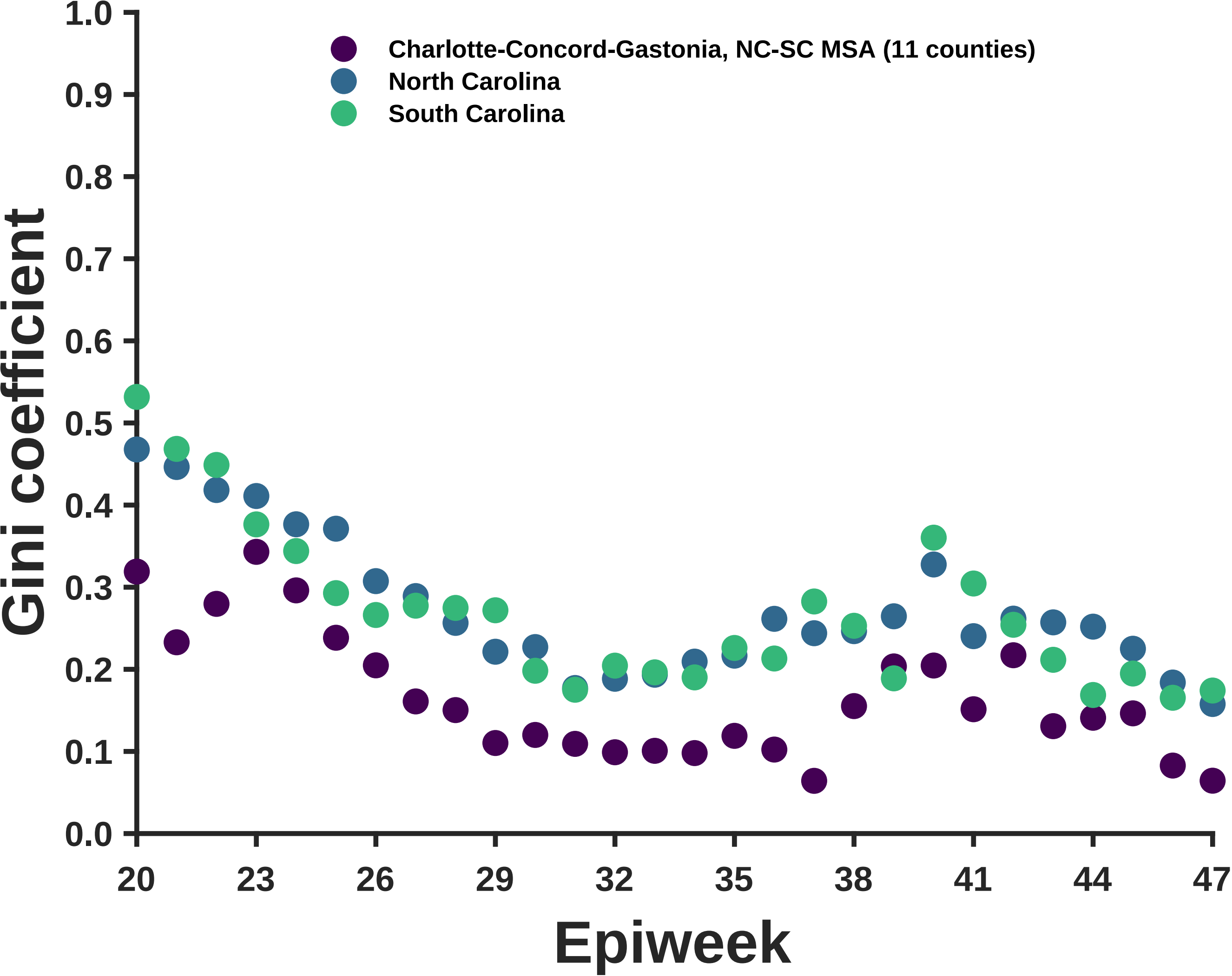
Charlotte-Concord-Gastonia, NC-SC MSA (11 counties)
North Carolina
South Carolina
Gini coefficient
1.0
0.9
0.8
0.7
0.6
0.5
0.4
0.3
0.2
0.1
0.0
20
23
26
29
32
35
38
41
44
47
Epiweek

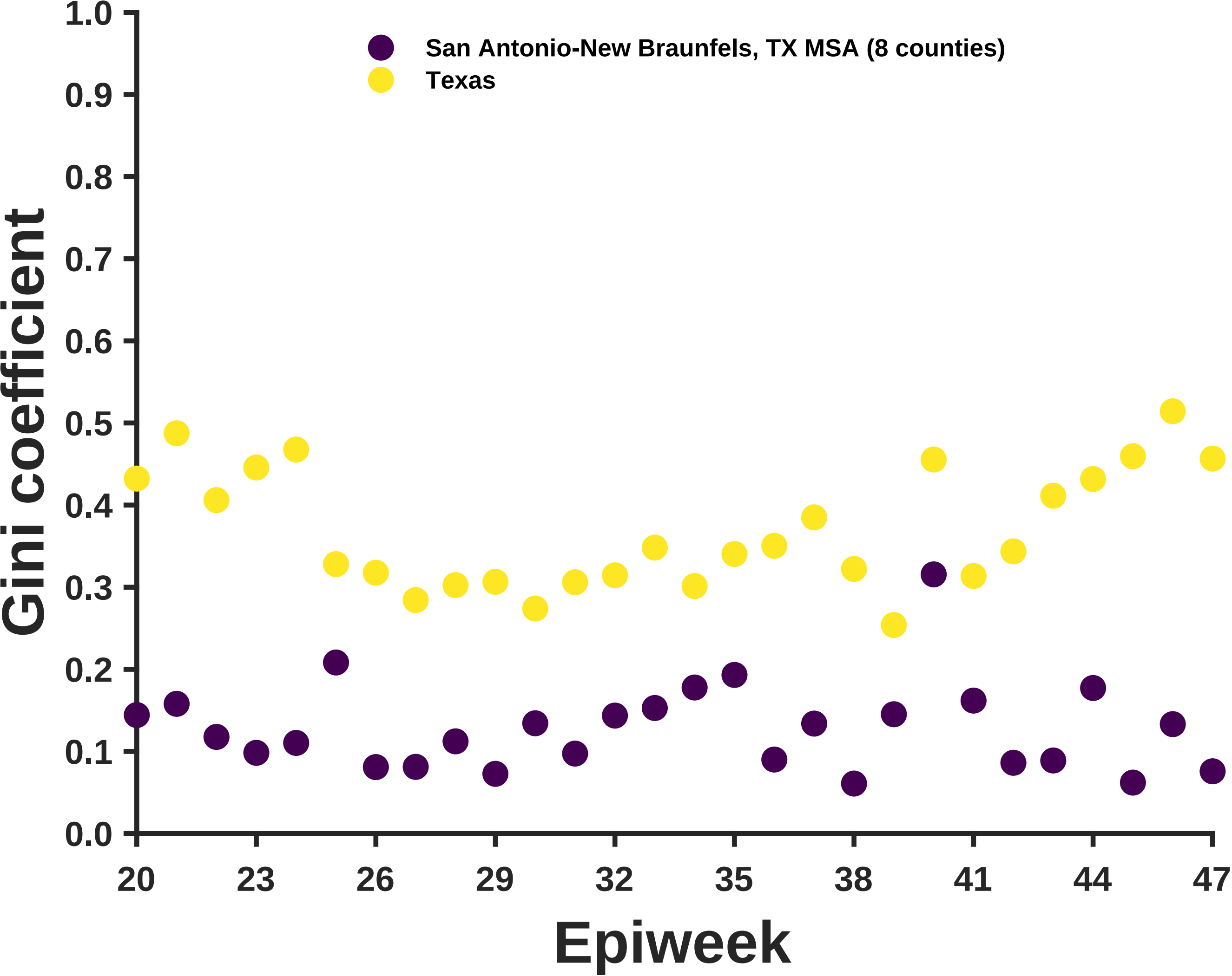
San Antonio-New Braunfels, TX MSA (8 counties)
Texas
Gini coefficient
1.0
0.9
0.8
0.7
0.6
0.5
0.4
0.3
0.2
0.1
0.0
20
23
26
29
32
35
38
41
44
47
Epiweek

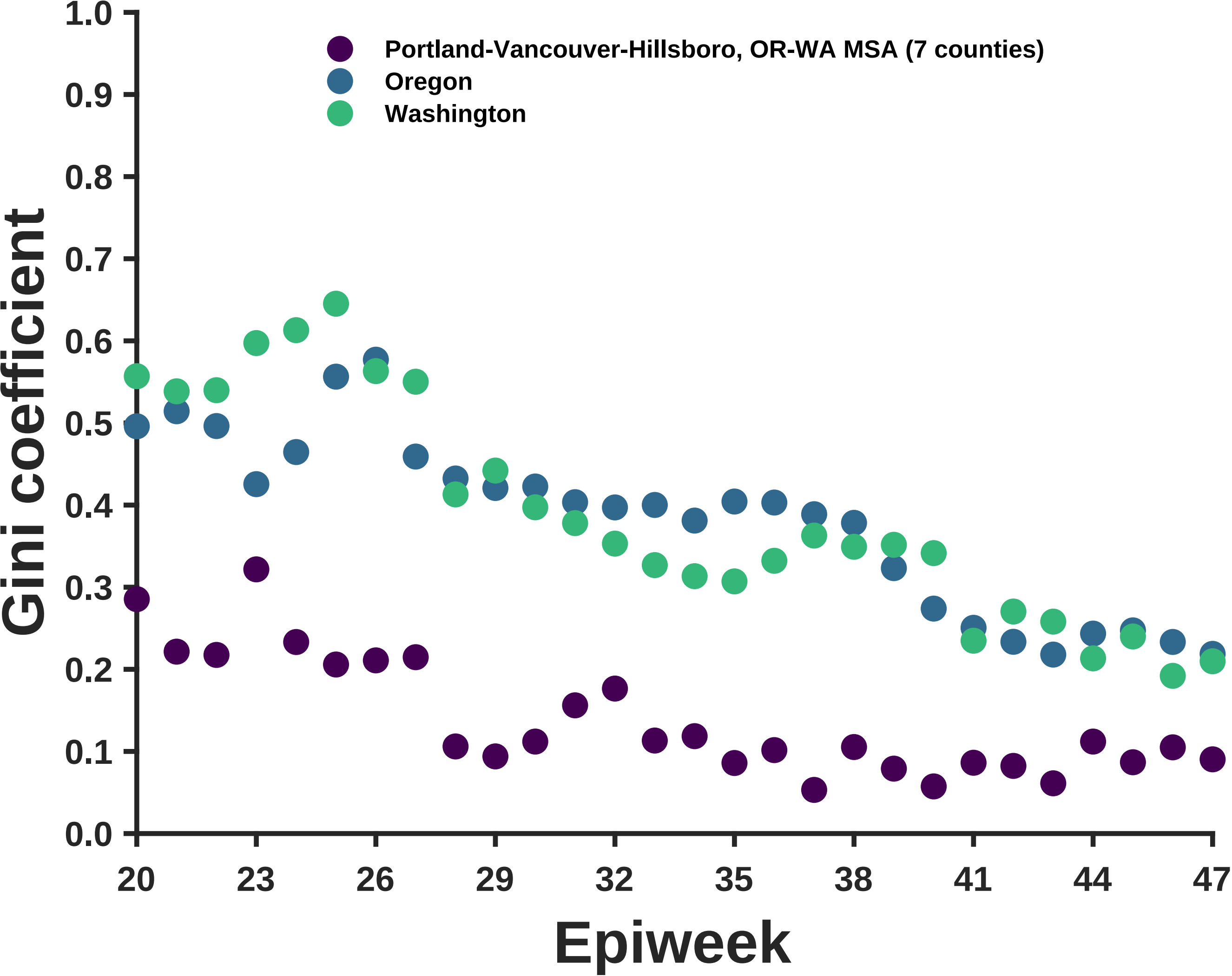

Portland-Vancouver-Hillsboro, OR-WA MSA (7 counties)
Oregon
Washington
Gini coefficient
Epiweek
0.0
0.1
0.2
0.3
0.4
0.5
0.6
0.7
0.8
0.9
1.0
20
23
26
29
32
35
38
41
44
47

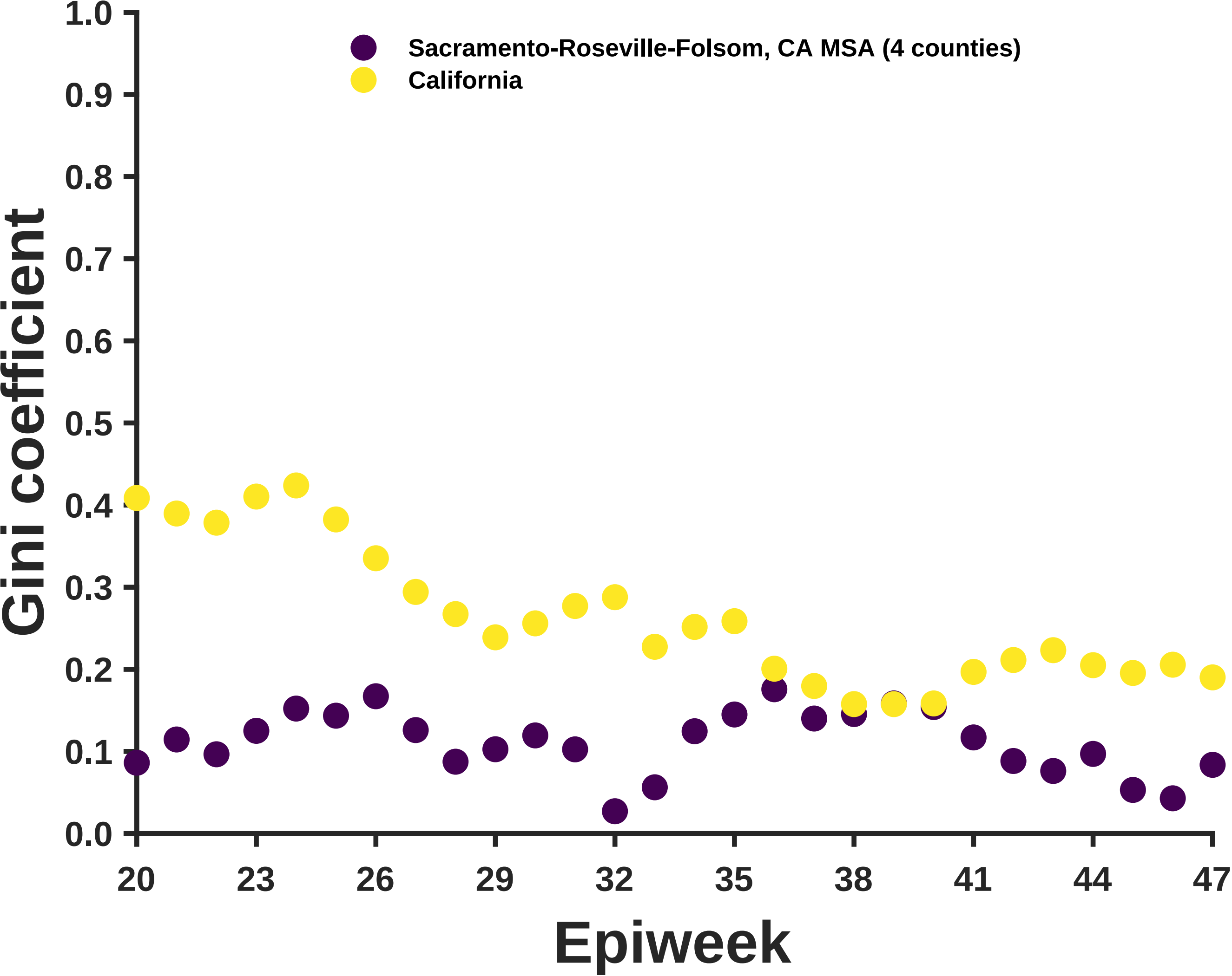
Sacramento-Roseville-Folsom, CA MSA (4 counties)
California
Gini coefficient
Epiweek
1.0
0.9
0.8
0.7
0.6
0.5
0.4
0.3
0.2
0.1
0.0
20
23
26
29
32
35
38
41
44
47

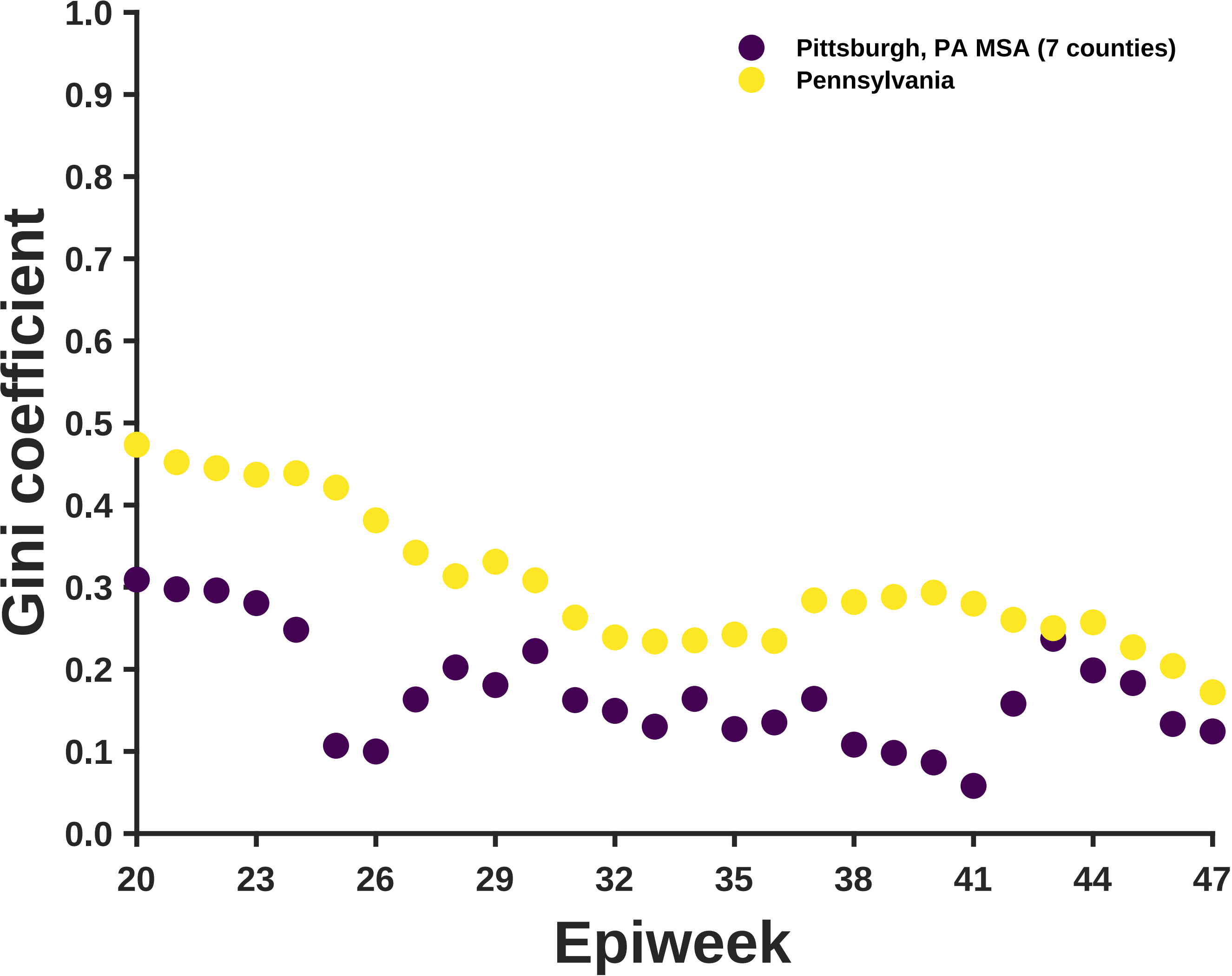

Pittsburgh, PA MSA (7 counties)
Pennsylvania
Gini coefficient
1.0
0.9
0.8
0.7
0.6
0.5
0.4
0.3
0.2
0.1
0.0
20
23
26
29
32
35
38
41
44
47
Epiweek

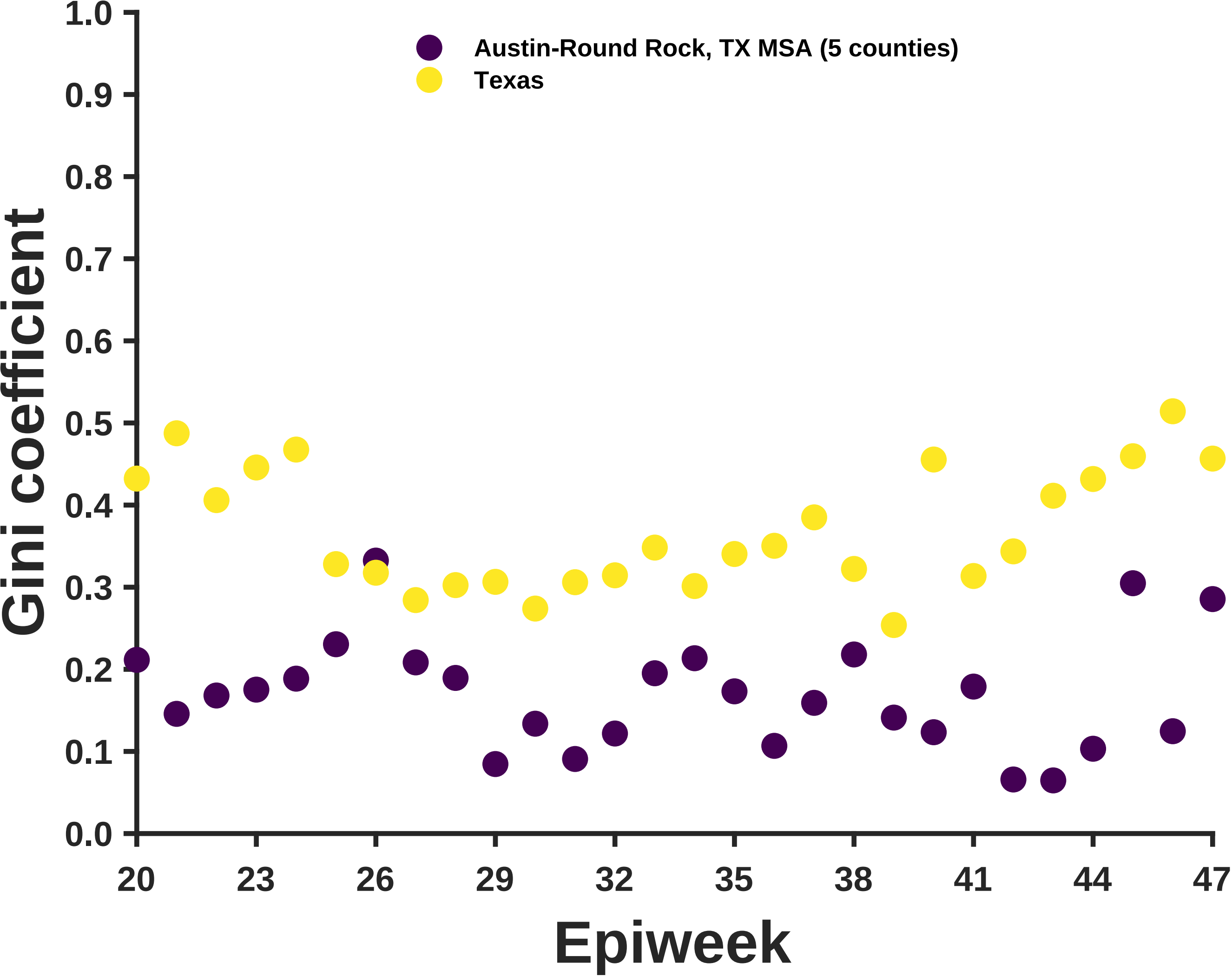

Austin-Round Rock, TX MSA (5 counties)
Texas
Gini coefficient
1.0
0.9
0.8
0.7
0.6
0.5
0.4
0.3
0.2
0.1
0.0
20
23
26
29
32
35
38
41
44
47
Epiweek

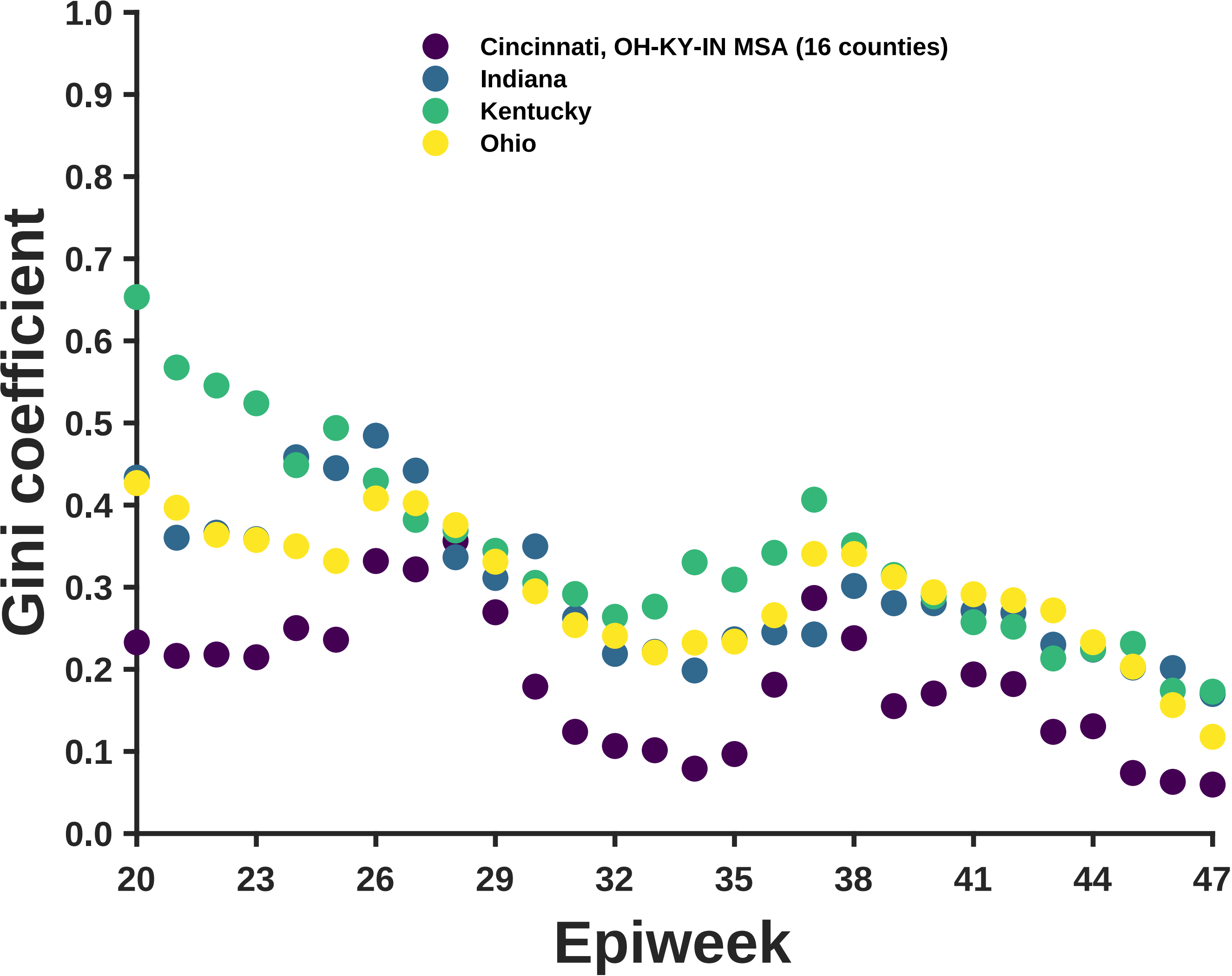

Cincinnati, OH-KY-IN MSA (16 counties)
Indiana
Kentucky
Ohio
Gini coefficient
Epiweek
1.0
0.9
0.8
0.7
0.6
0.5
0.4
0.3
0.2
0.1
0.0
20
23
26
29
32
35
38
41
44
47

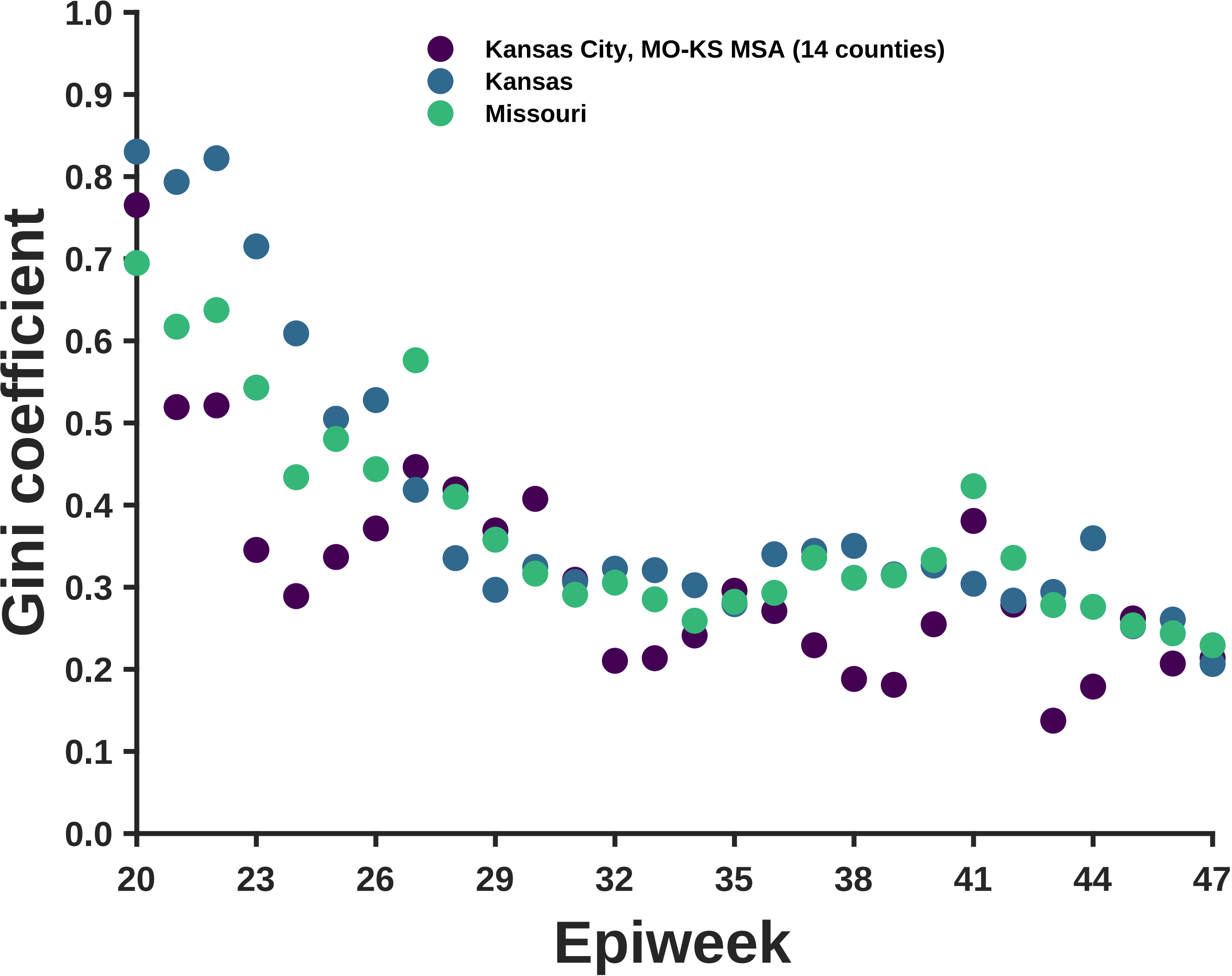

Kansas City, MO-KS MSA (14 counties)
Kansas
Missouri
Gini coefficient
1.0
0.9
0.8
0.7
0.6
0.5
0.4
0.3
0.2
0.1
0.0
20
23
26
29
32
35
38
41
44
47
Epiweek

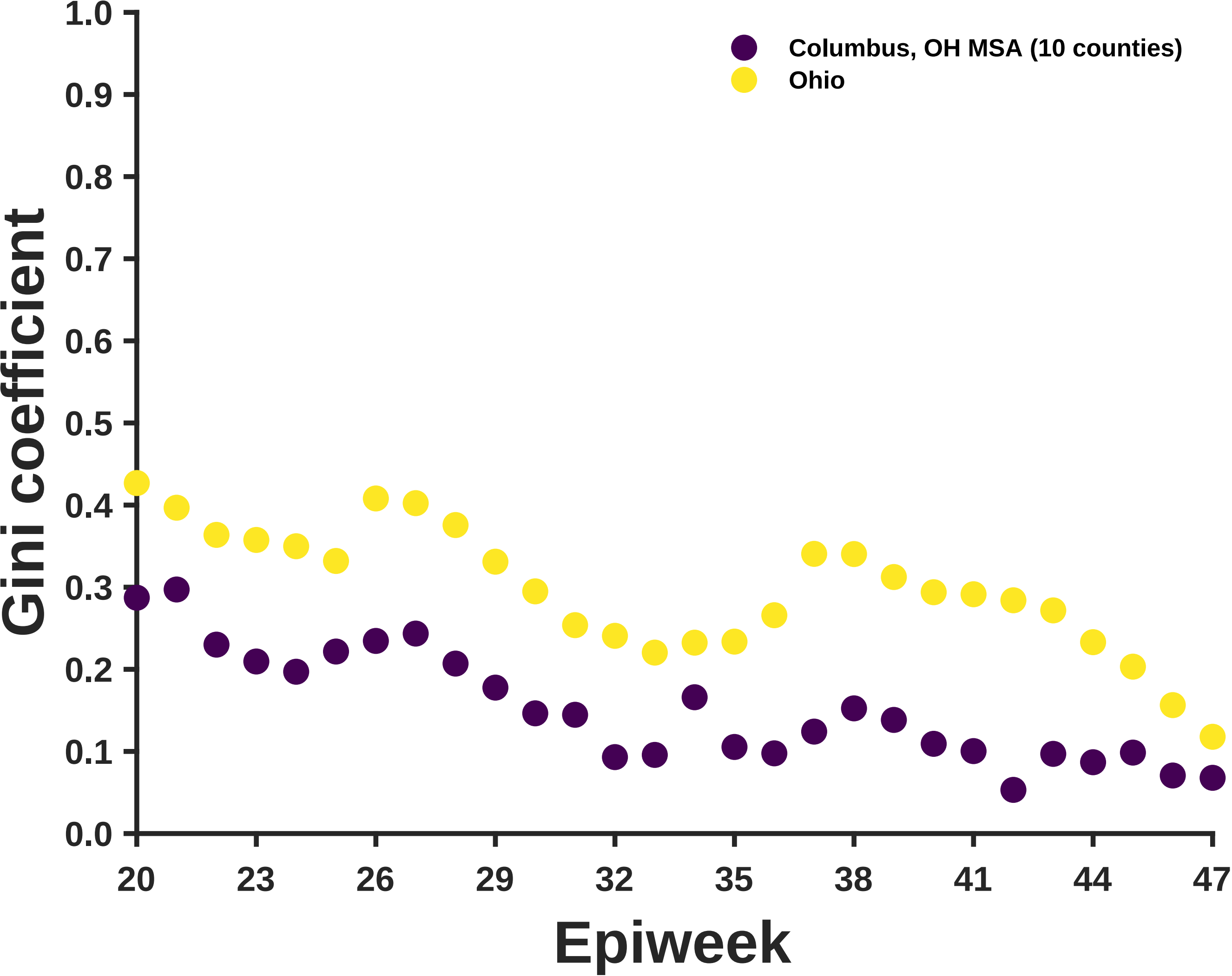
Columbus, OH MSA (10 counties)
Ohio
Gini coefficient
1.0
0.9
0.8
0.7
0.6
0.5
0.4
0.3
0.2
0.1
0.0
20
23
26
29
32
35
38
41
44
47
Epiweek

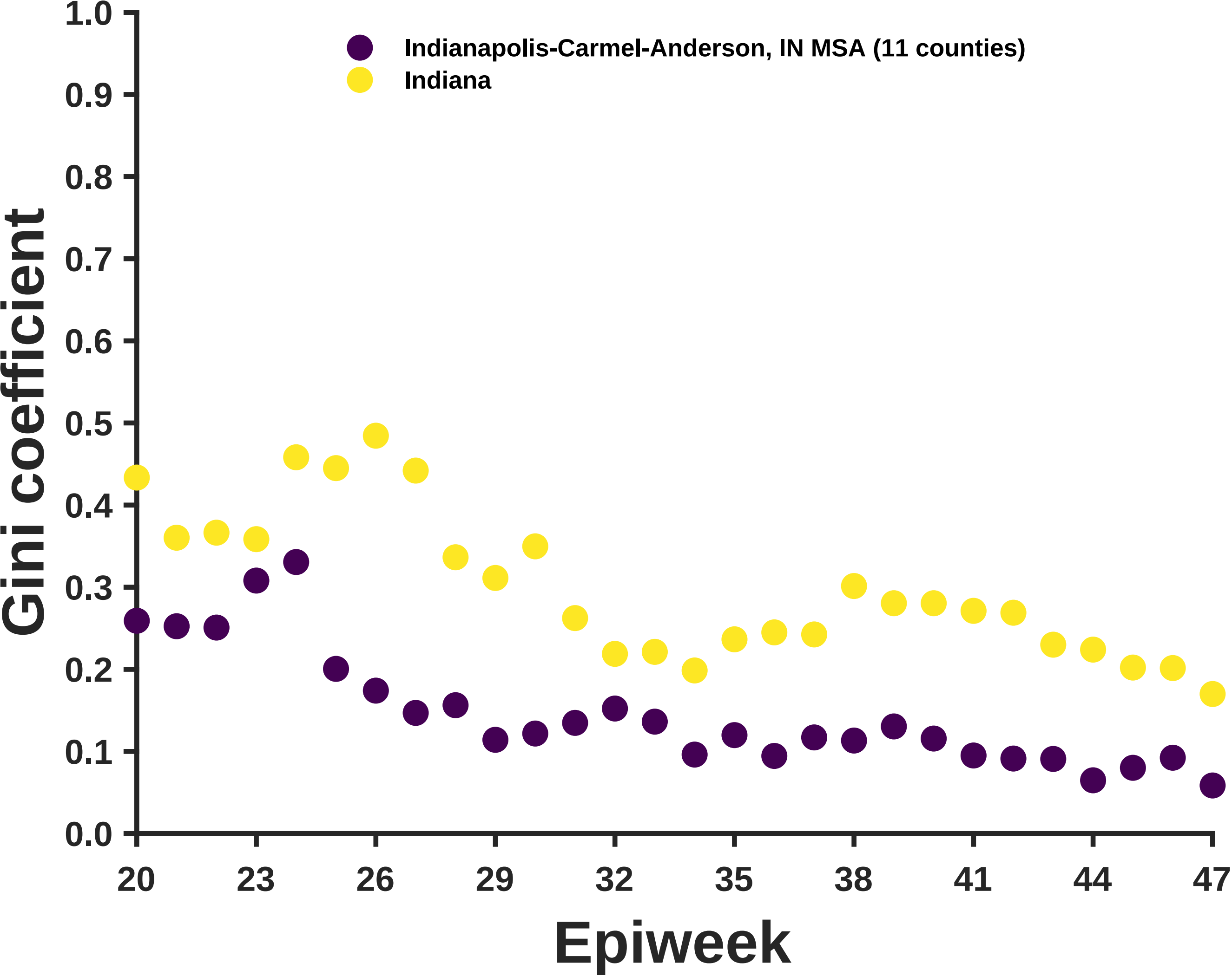

Indianapolis-Carmel-Anderson, IN MSA (11 counties)
Indiana
Gini coefficient
1.0
0.9
0.8
0.7
0.6
0.5
0.4
0.3
0.2
0.1
0.0
20
23
26
29
32
35
38
41
44
47
Epiweek

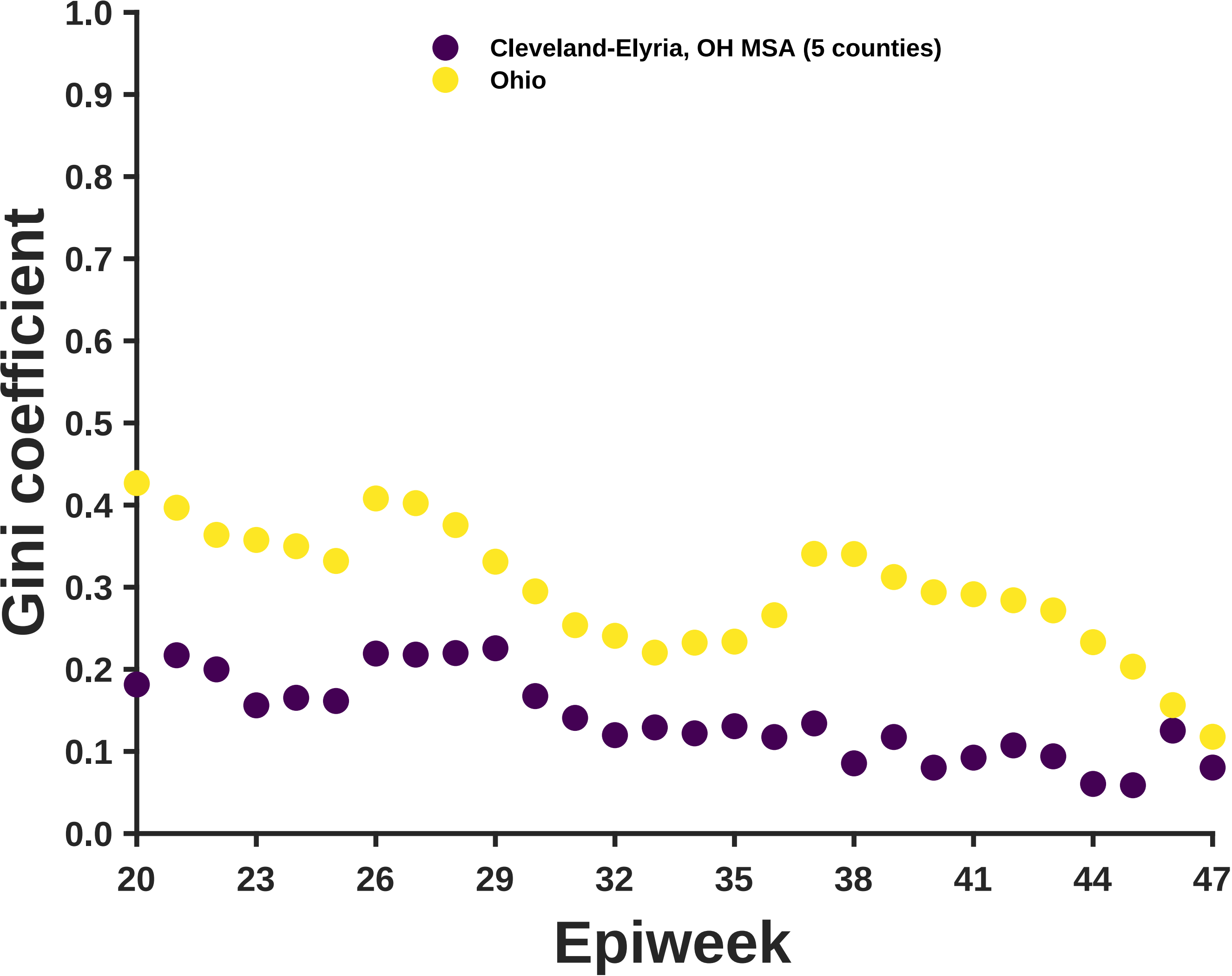

Cleveland-Elyria, OH MSA (5 counties)
Ohio
Gini coefficient
1.0
0.9
0.8
0.7
0.6
0.5
0.4
0.3
0.2
0.1
0.0
20
23
26
29
32
35
38
41
44
47
Epiweek

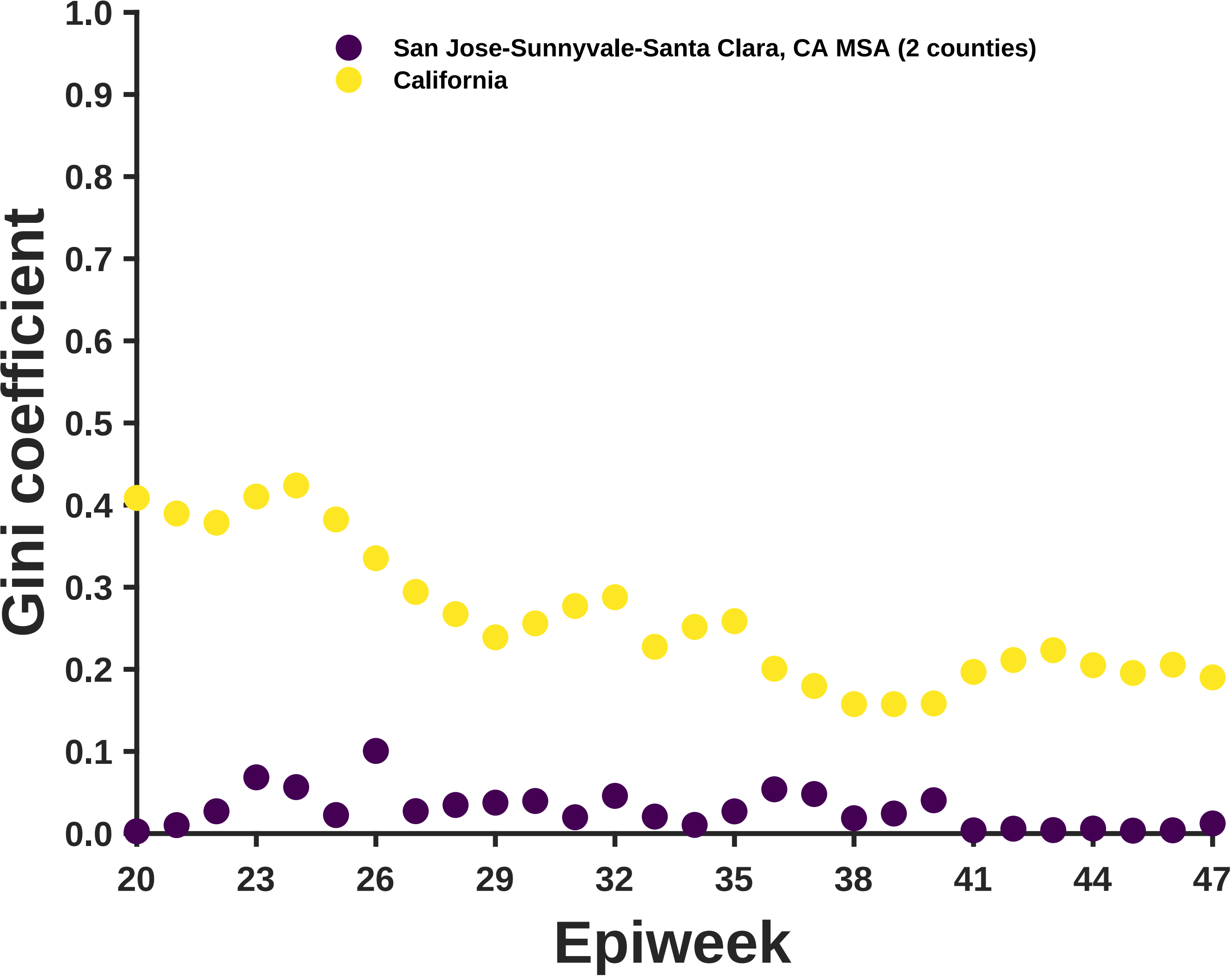
San Jose-Sunnyvale-Santa Clara, CA MSA (2 counties)
California
Gini coefficient
1.0
0.9
0.8
0.7
0.6
0.5
0.4
0.3
0.2
0.1
0.0
20
23
26
29
32
35
38
41
44
47
Epiweek

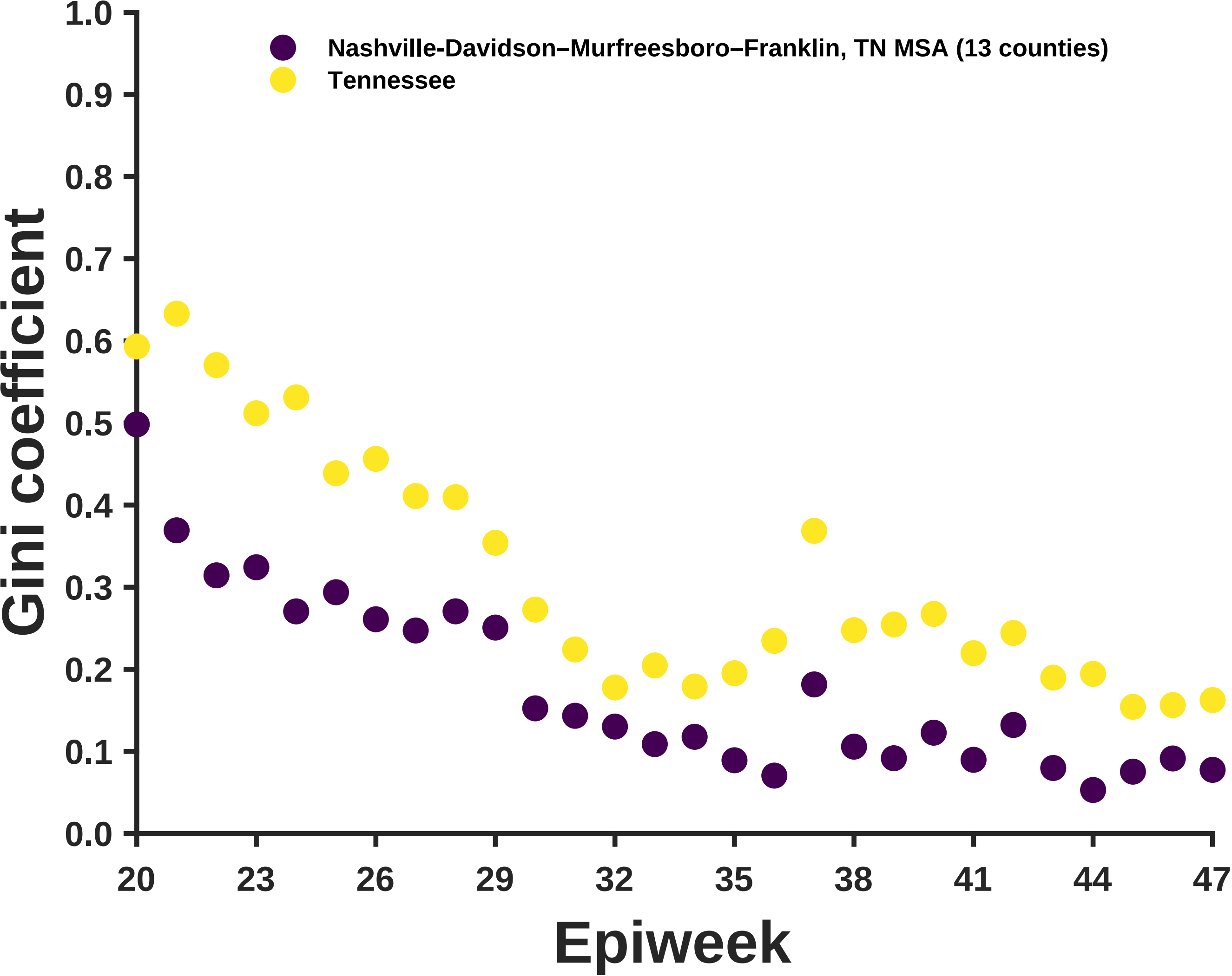

Nashville-Davidson–Murfreesboro–Franklin, TN MSA (13 counties)
Tennessee
Gini coefficient
0.0
0.1
0.2
0.3
0.4
0.5
0.6
0.7
0.8
0.9
1.0
20
23
26
29
32
35
38
41
44
47
Epiweek

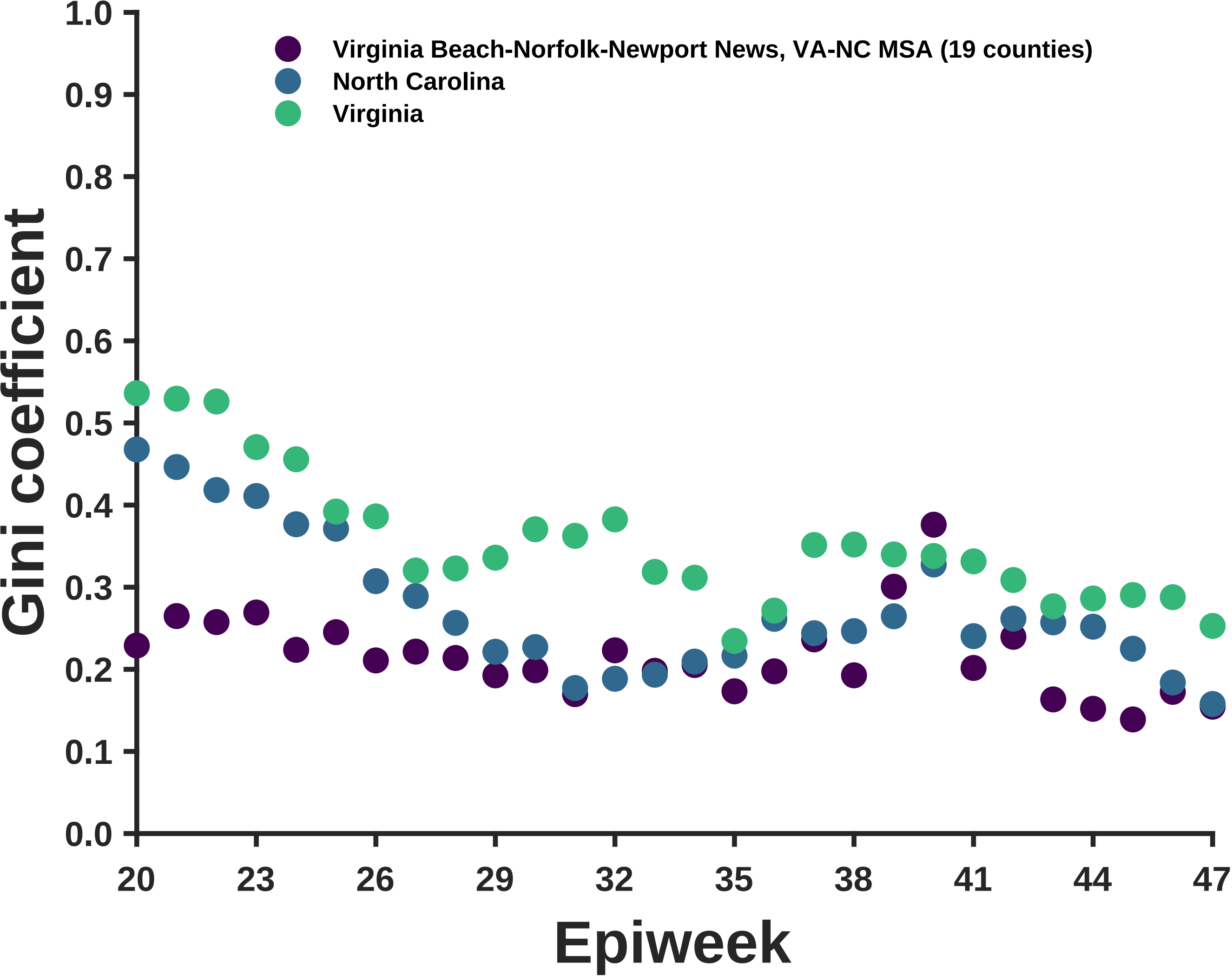

Virginia Beach-Norfolk-Newport News, VA-NC MSA (19 counties)
North Carolina
Virginia
Gini coefficient
0.0
0.1
0.2
0.3
0.4
0.5
0.6
0.7
0.8
0.9
1.0
20
23
26
29
32
35
38
41
44
47
Epiweek

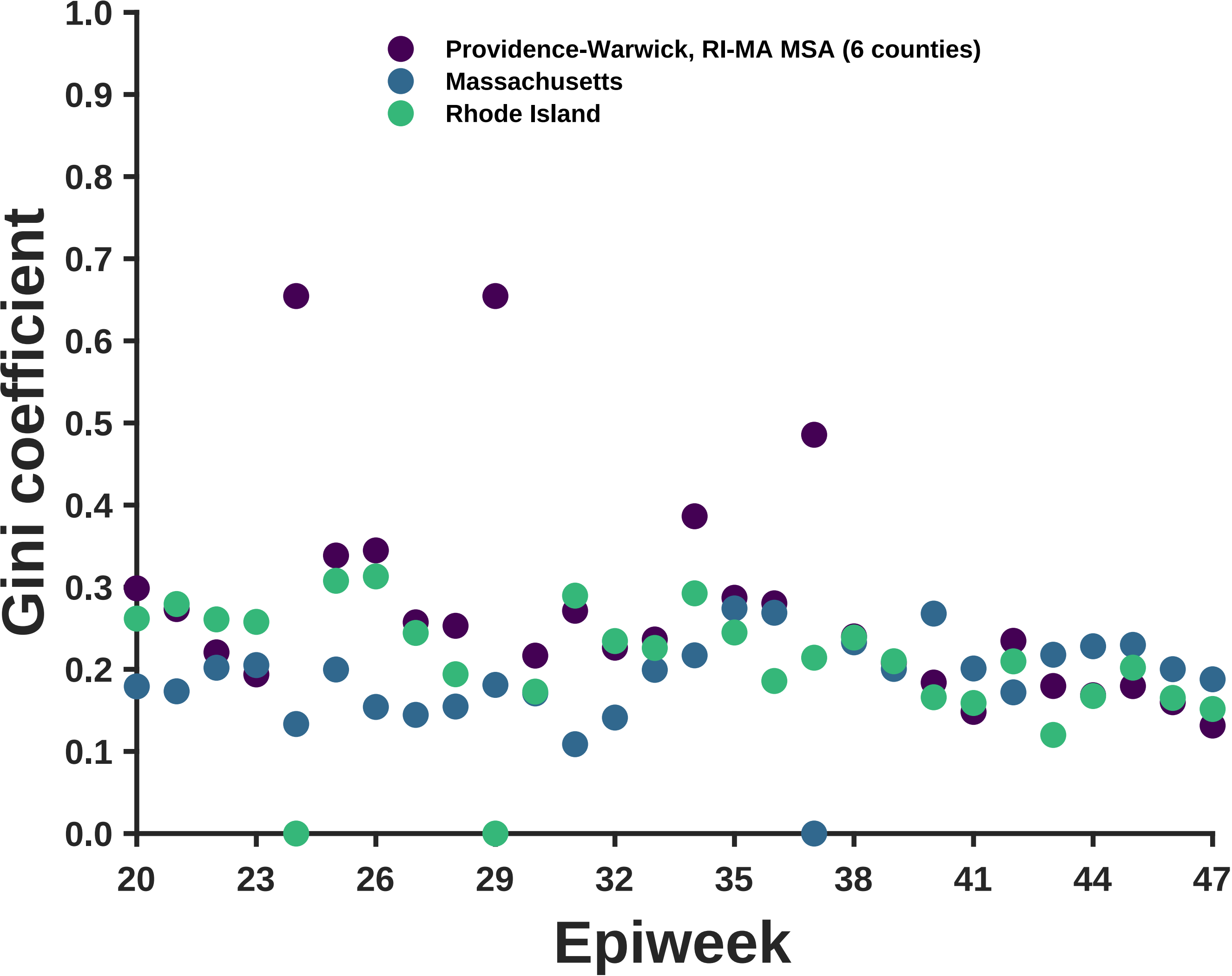

Providence-Warwick, RI-MA MSA (6 counties)
Massachusetts
Rhode Island
Gini coefficient
1.0
0.9
0.8
0.7
0.6
0.5
0.4
0.3
0.2
0.1
0.0
20
23
26
29
32
35
38
41
44
47
Epiweek

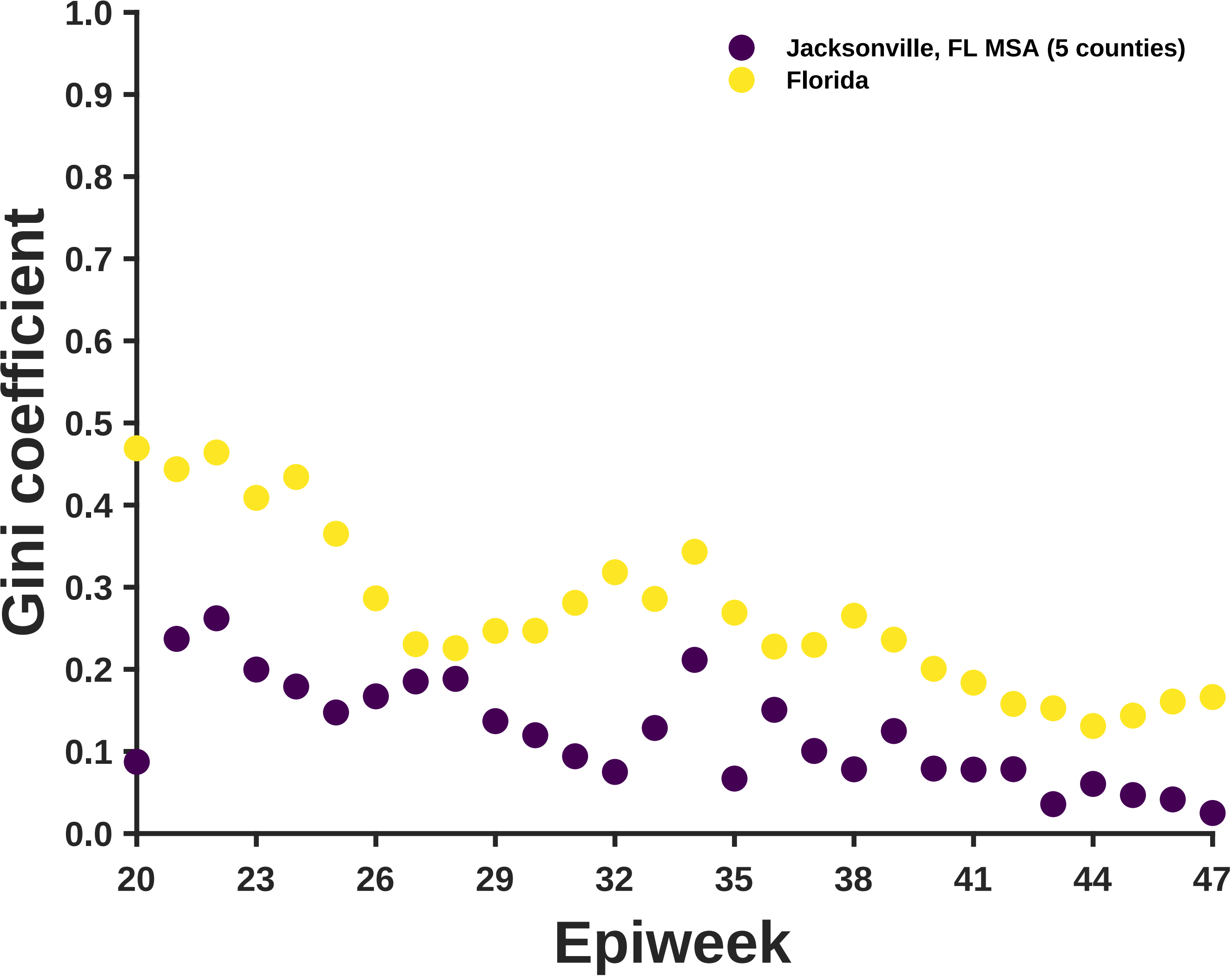
Jacksonville, FL MSA (5 counties)
Florida
Gini coefficient
Epiweek
1.0
0.9
0.8
0.7
0.6
0.5
0.4
0.3
0.2
0.1
0.0
20
23
26
29
32
35
38
41
44
47

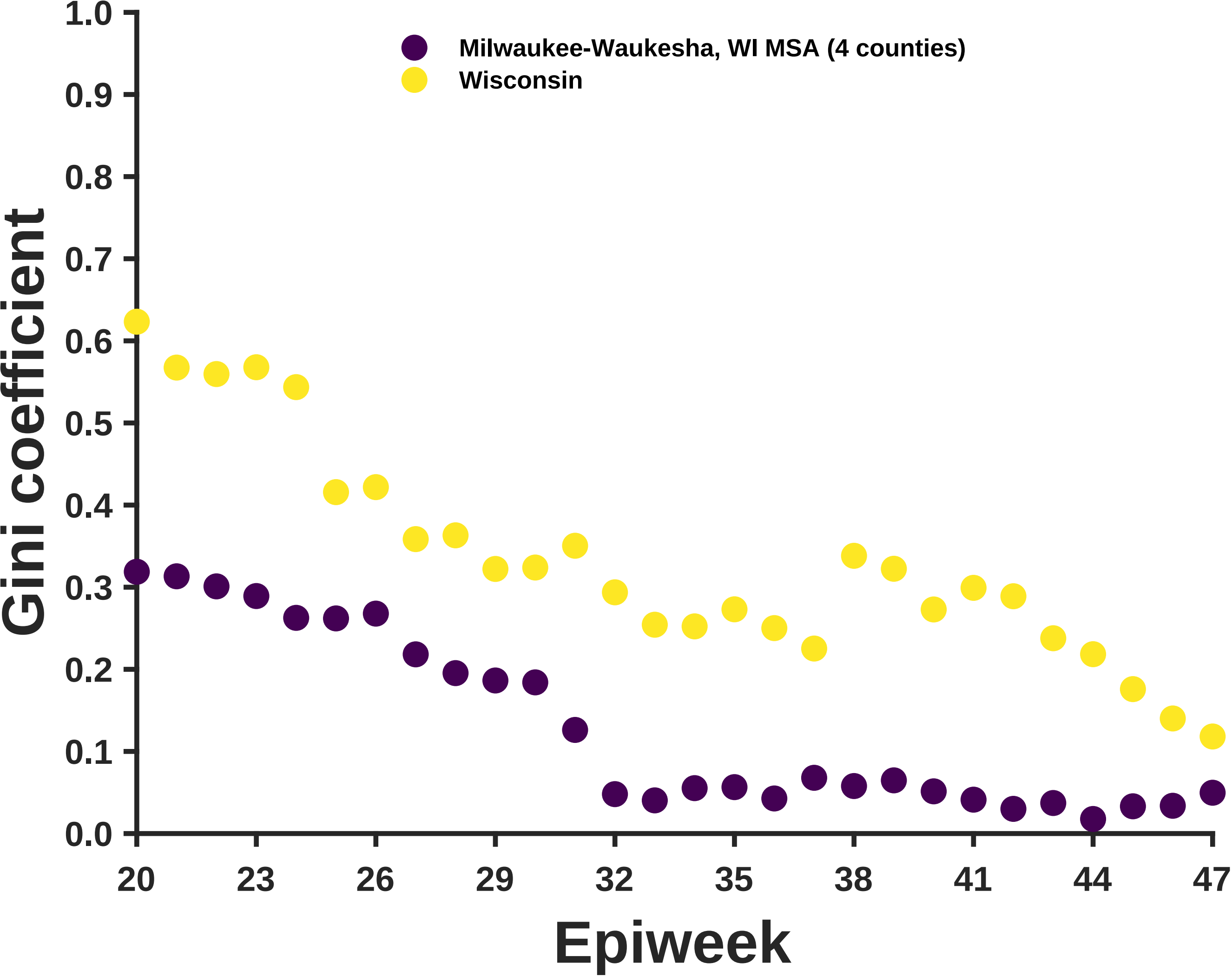

Milwaukee-Waukesha, WI MSA (4 counties)
Wisconsin
Gini coefficient
1.0
0.9
0.8
0.7
0.6
0.5
0.4
0.3
0.2
0.1
0.0
20
23
26
29
32
35
38
41
44
47
Epiweek

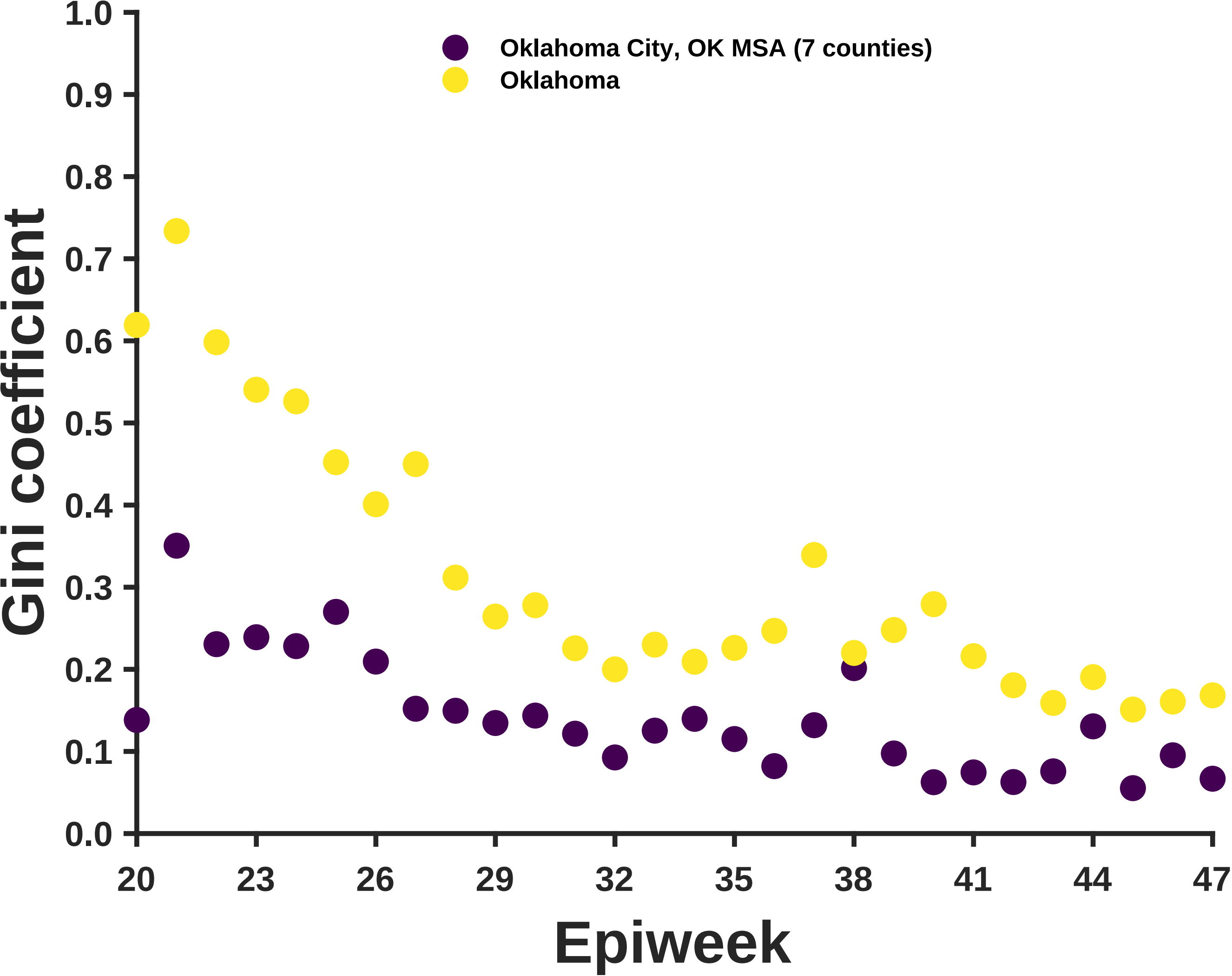

Oklahoma City, OK MSA (7 counties)
Oklahoma
Gini coefficient
1.0
0.9
0.8
0.7
0.6
0.5
0.4
0.3
0.2
0.1
0.0
20
23
26
29
32
35
38
41
44
47
Epiweek

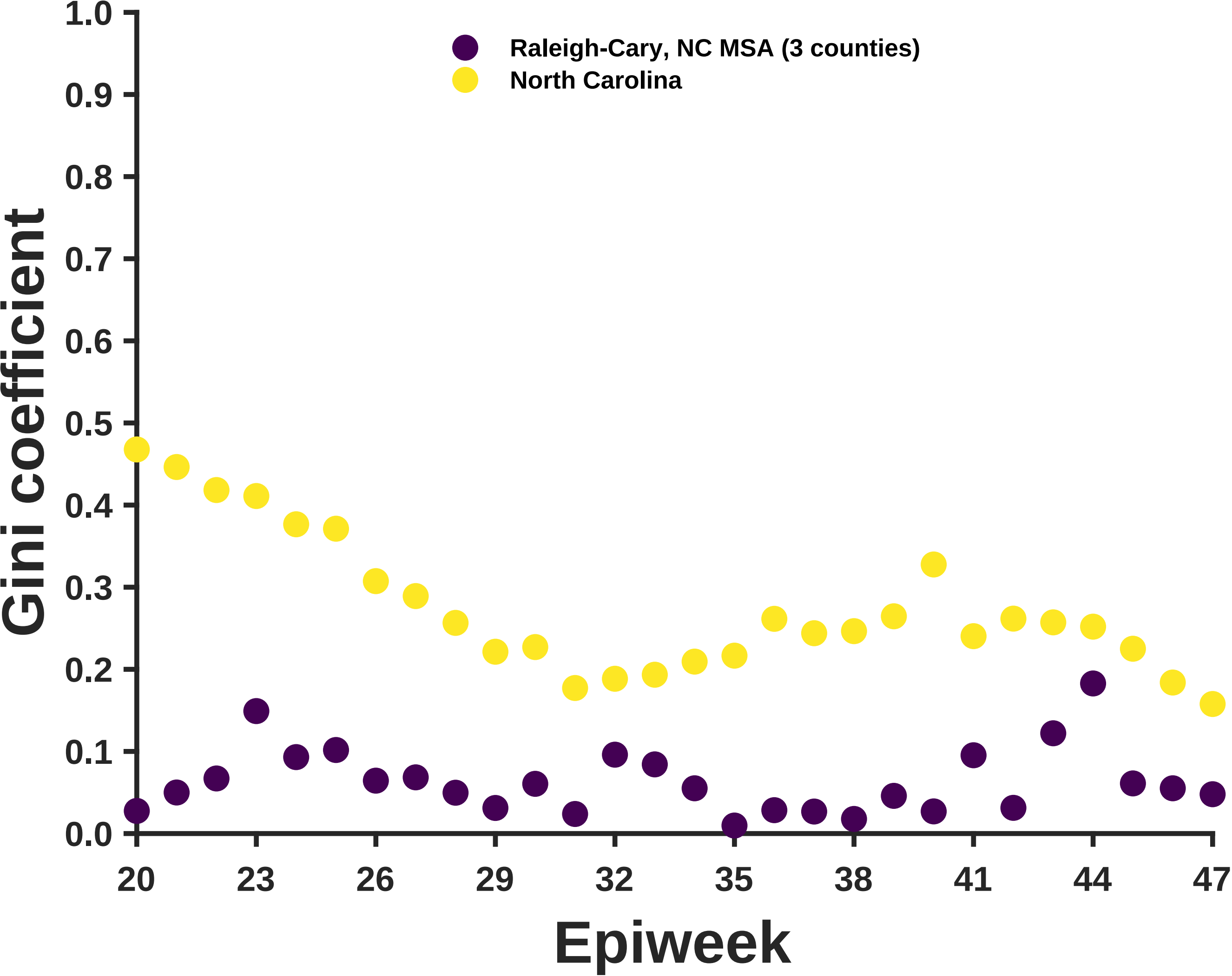
Raleigh-Cary, NC MSA (3 counties)
North Carolina
Gini coefficient
1.0
0.9
0.8
0.7
0.6
0.5
0.4
0.3
0.2
0.1
0.0
20
23
26
29
32
35
38
41
44
47
Epiweek

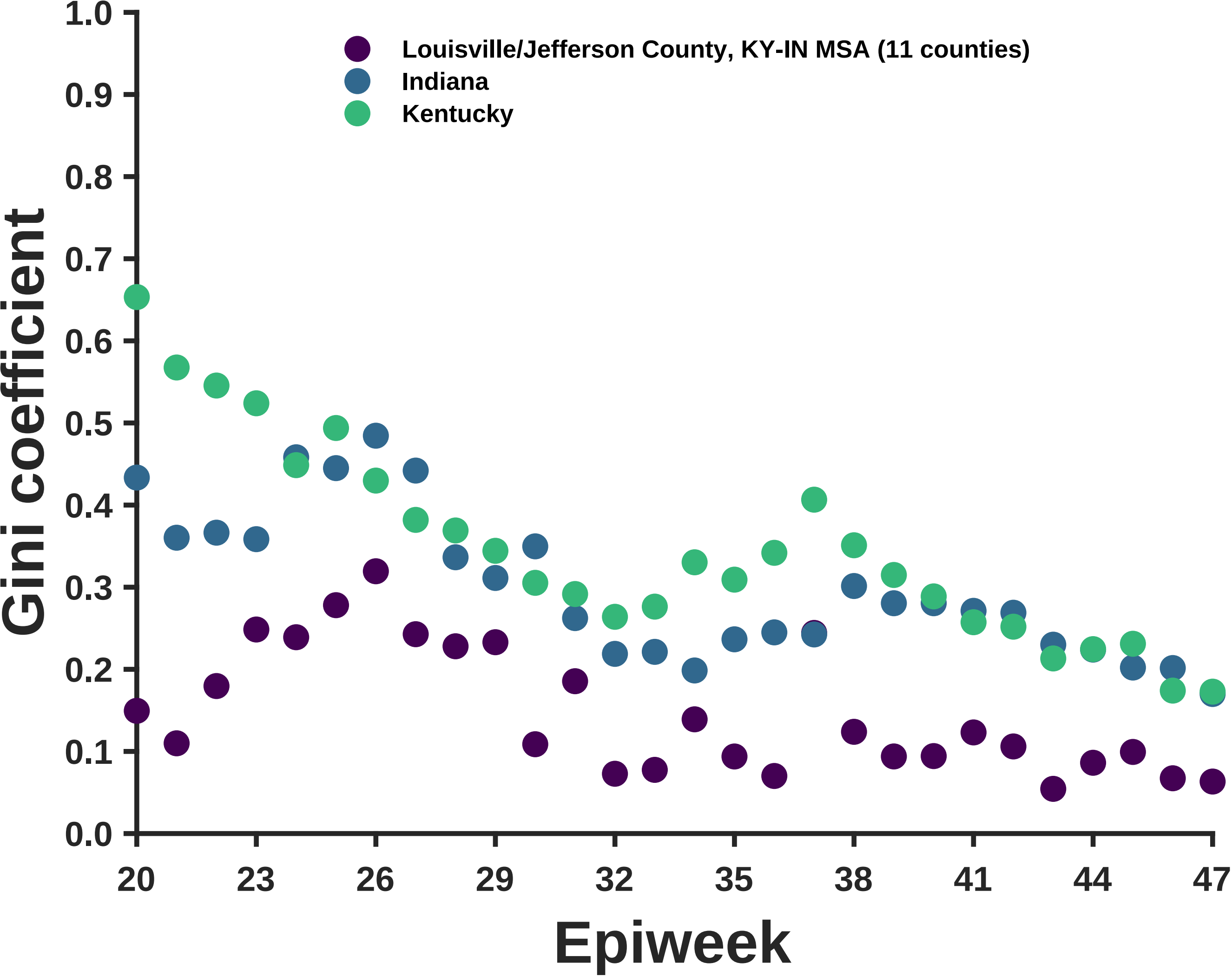

Louisville/Jefferson County, KY-IN MSA (11 counties)
Indiana
Kentucky
Gini coefficient
0.0
0.1
0.2
0.3
0.4
0.5
0.6
0.7
0.8
0.9
1.0
20
23
26
29
32
35
38
41
44
47
Epiweek

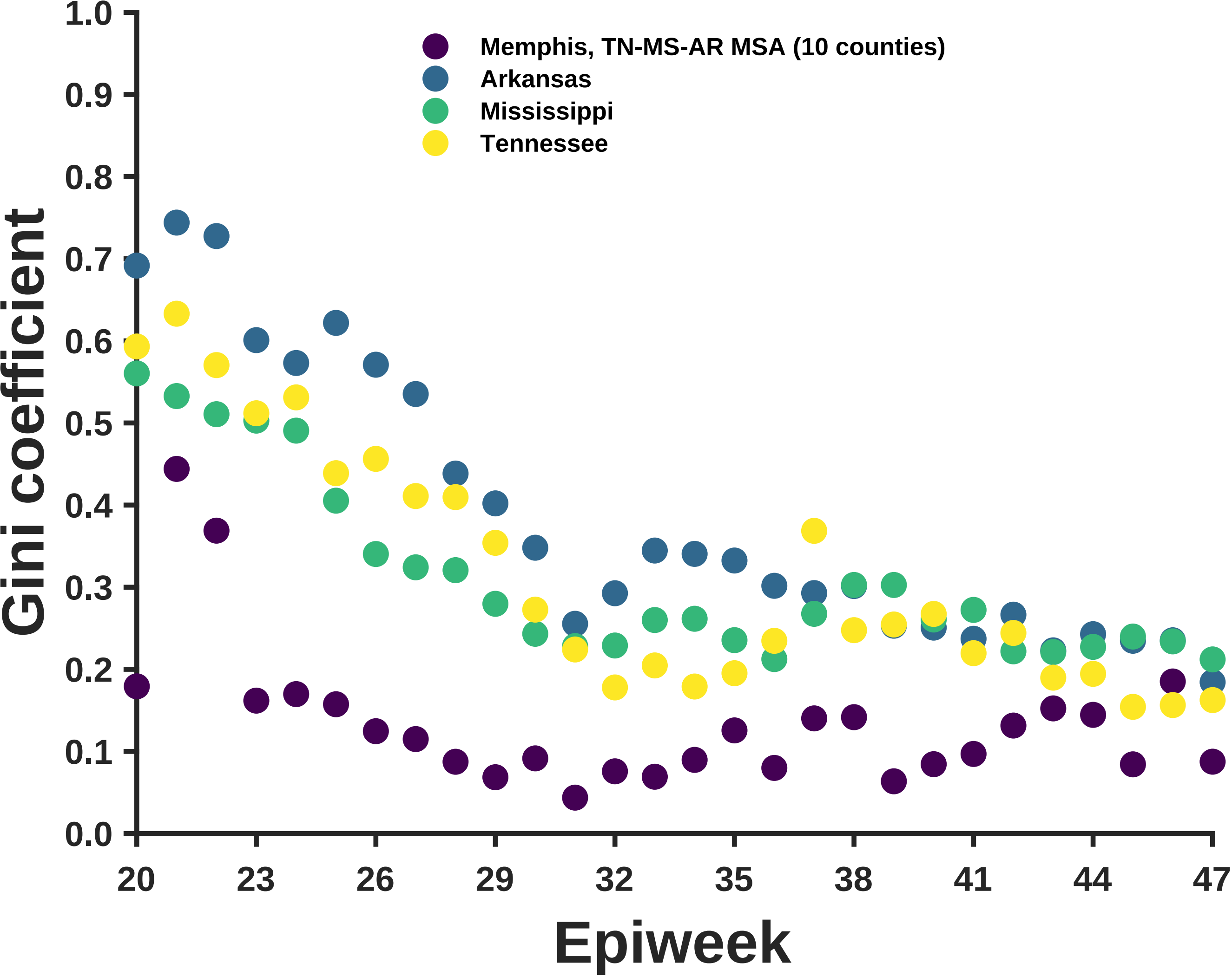
Memphis, TN-MS-AR MSA (10 counties)
Arkansas
Mississippi
Tennessee
Gini coefficient
Epiweek
1.0
0.9
0.8
0.7
0.6
0.5
0.4
0.3
0.2
0.1
0.0
20
23
26
29
32
35
38
41
44
47

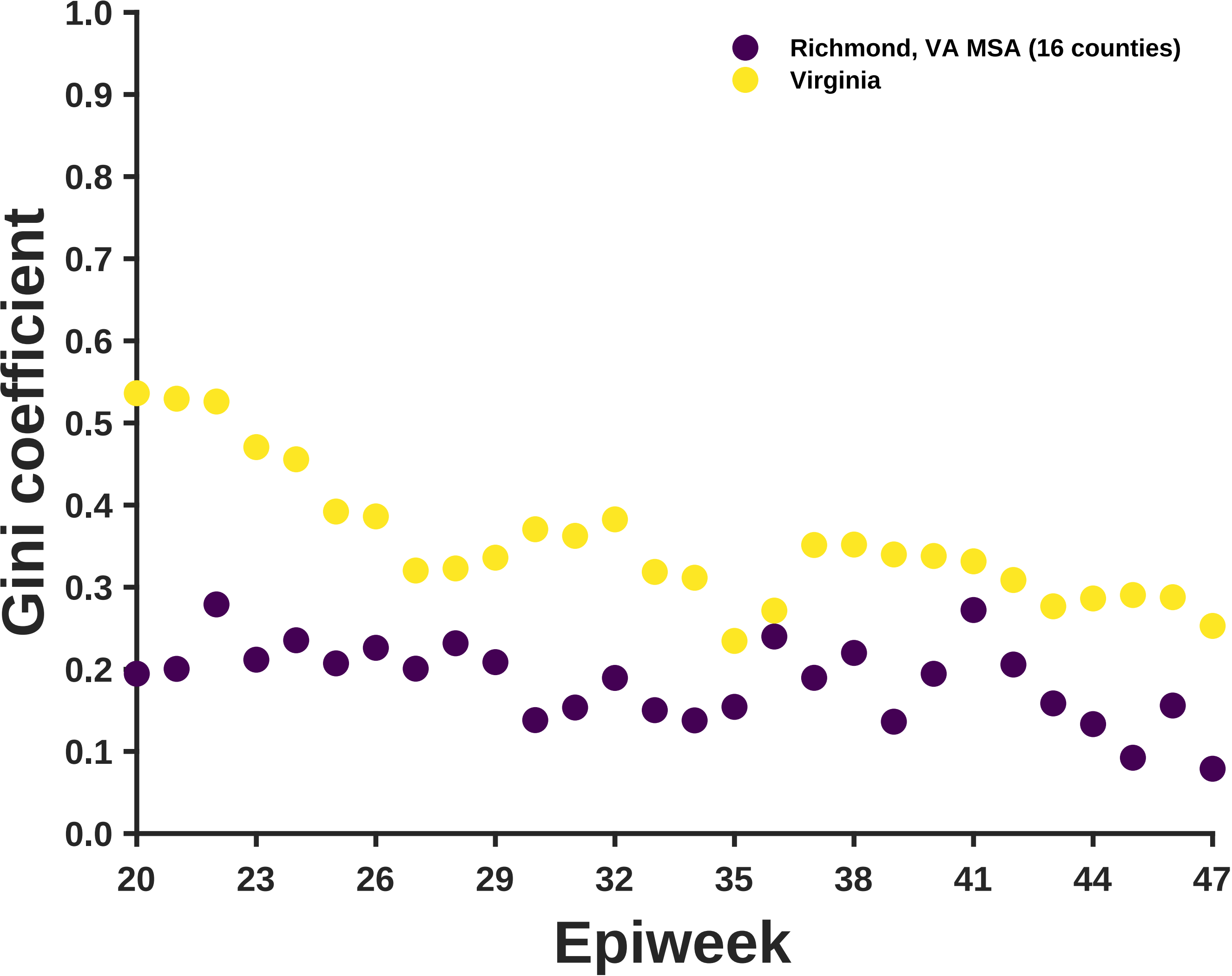

Richmond, VA MSA (16 counties)
Virginia
Gini coefficient
1.0
0.9
0.8
0.7
0.6
0.5
0.4
0.3
0.2
0.1
0.0
20
23
26
29
32
35
38
41
44
47
Epiweek

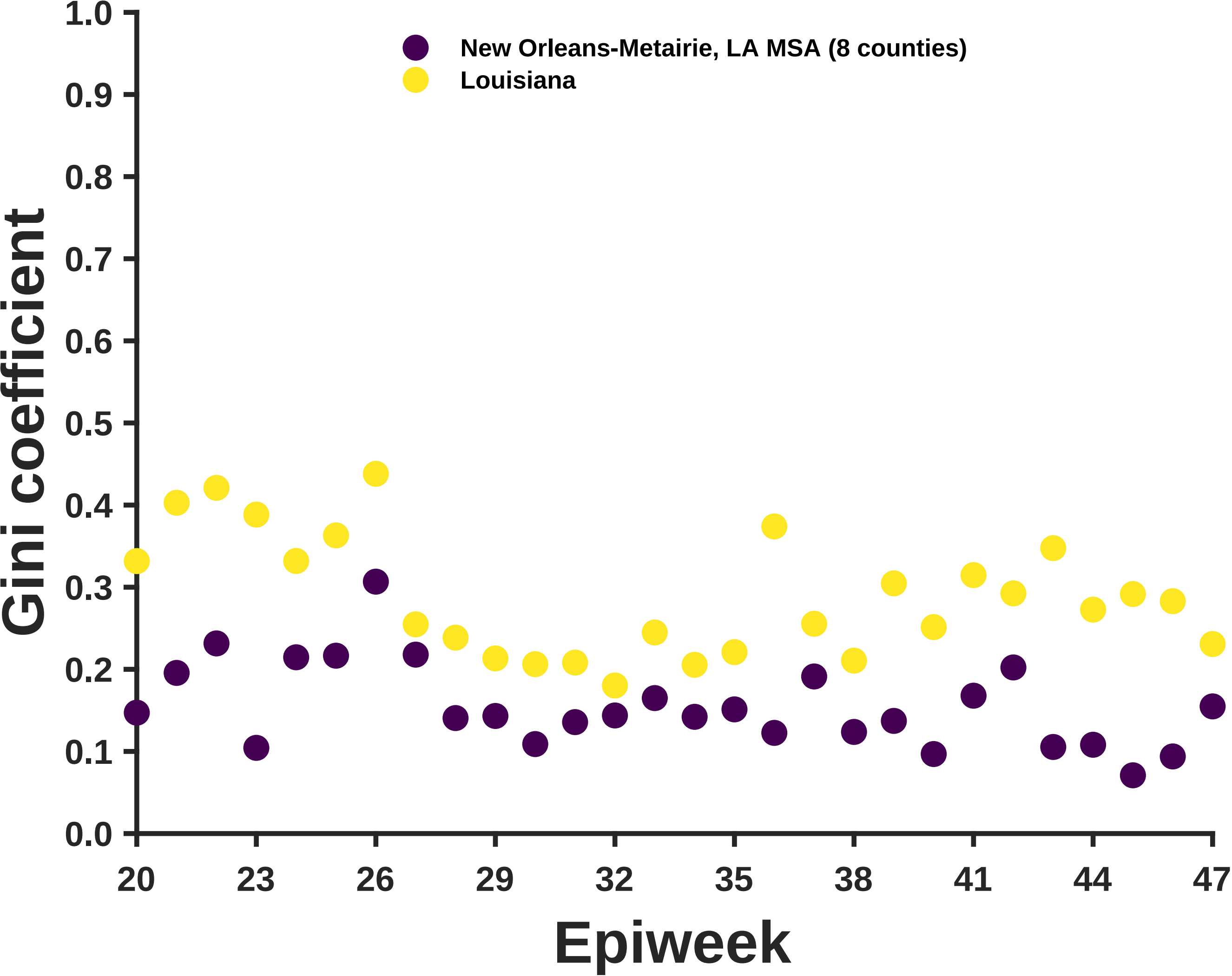

New Orleans-Metairie, LA MSA (8 counties)
Louisiana
Gini coefficient
Epiweek
1.0
0.9
0.8
0.7
0.6
0.5
0.4
0.3
0.2
0.1
0.0
20
23
26
29
32
35
38
41
44
47

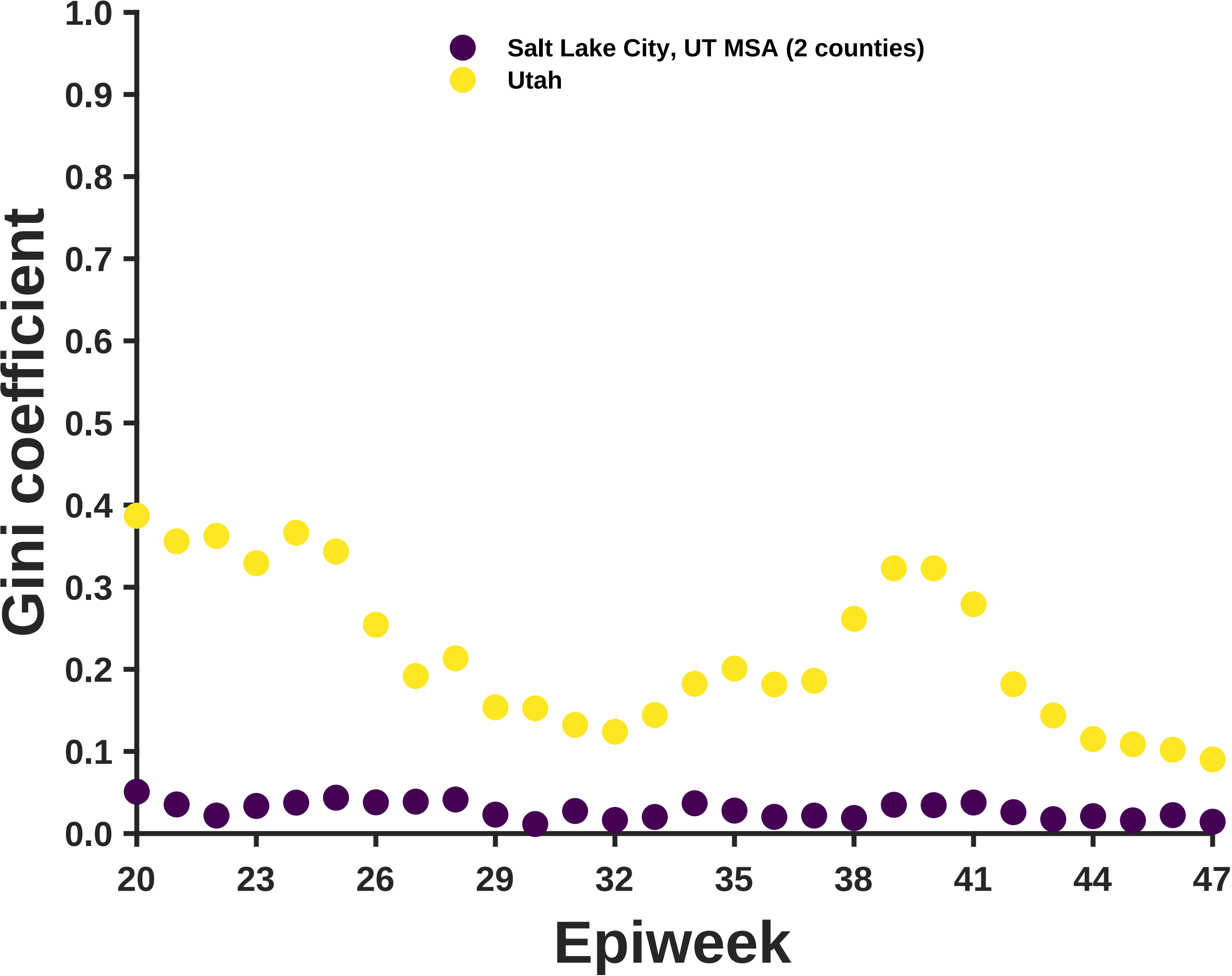

Salt Lake City, UT MSA (2 counties)
Utah
Gini coefficient
1.0
0.9
0.8
0.7
0.6
0.5
0.4
0.3
0.2
0.1
0.0
20
23
26
29
32
35
38
41
44
47
Epiweek

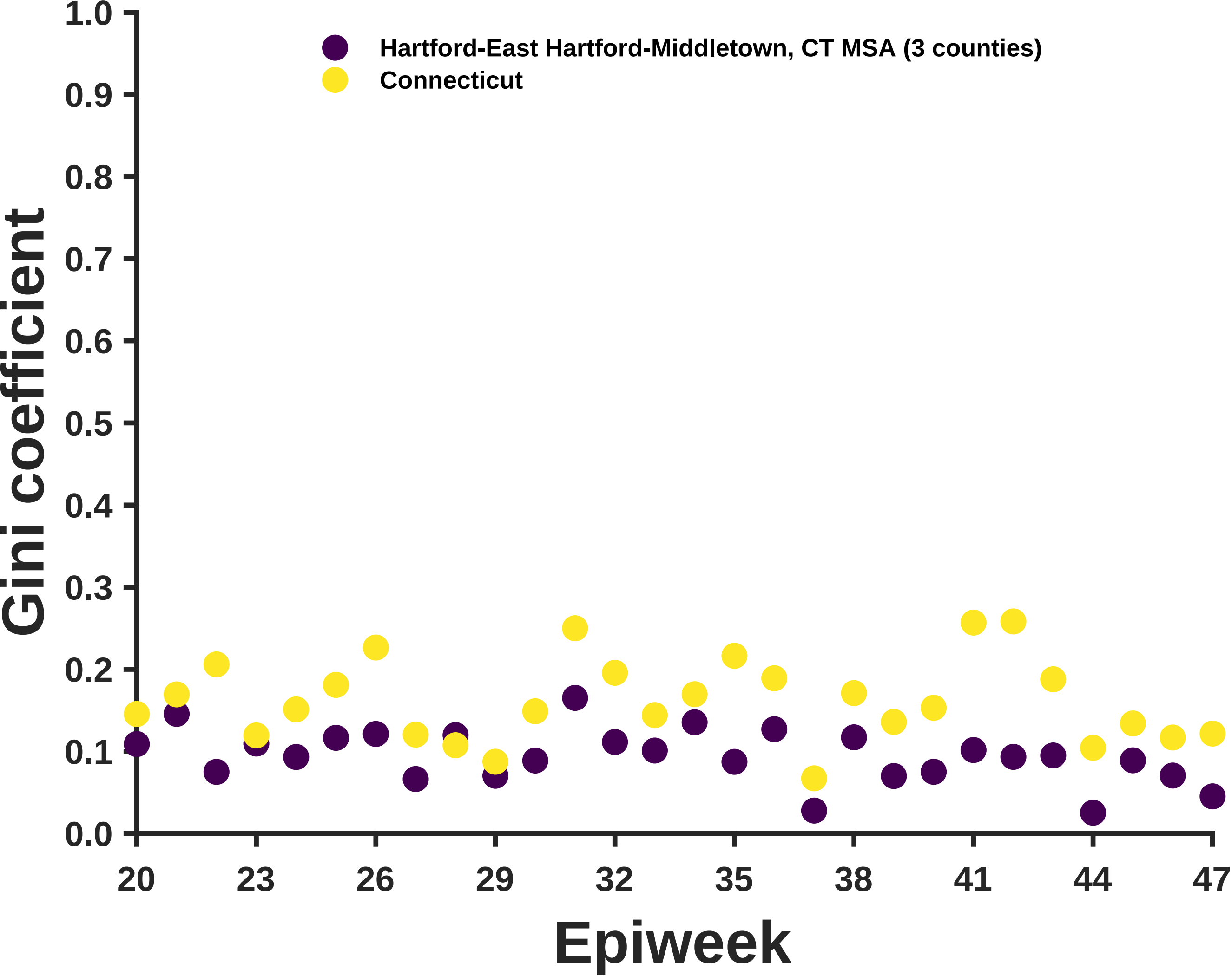

Hartford-East Hartford-Middletown, CT MSA (3 counties)
Connecticut
Gini coefficient
1.0
0.9
0.8
0.7
0.6
0.5
0.4
0.3
0.2
0.1
0.0
20
23
26
29
32
35
38
41
44
47
Epiweek

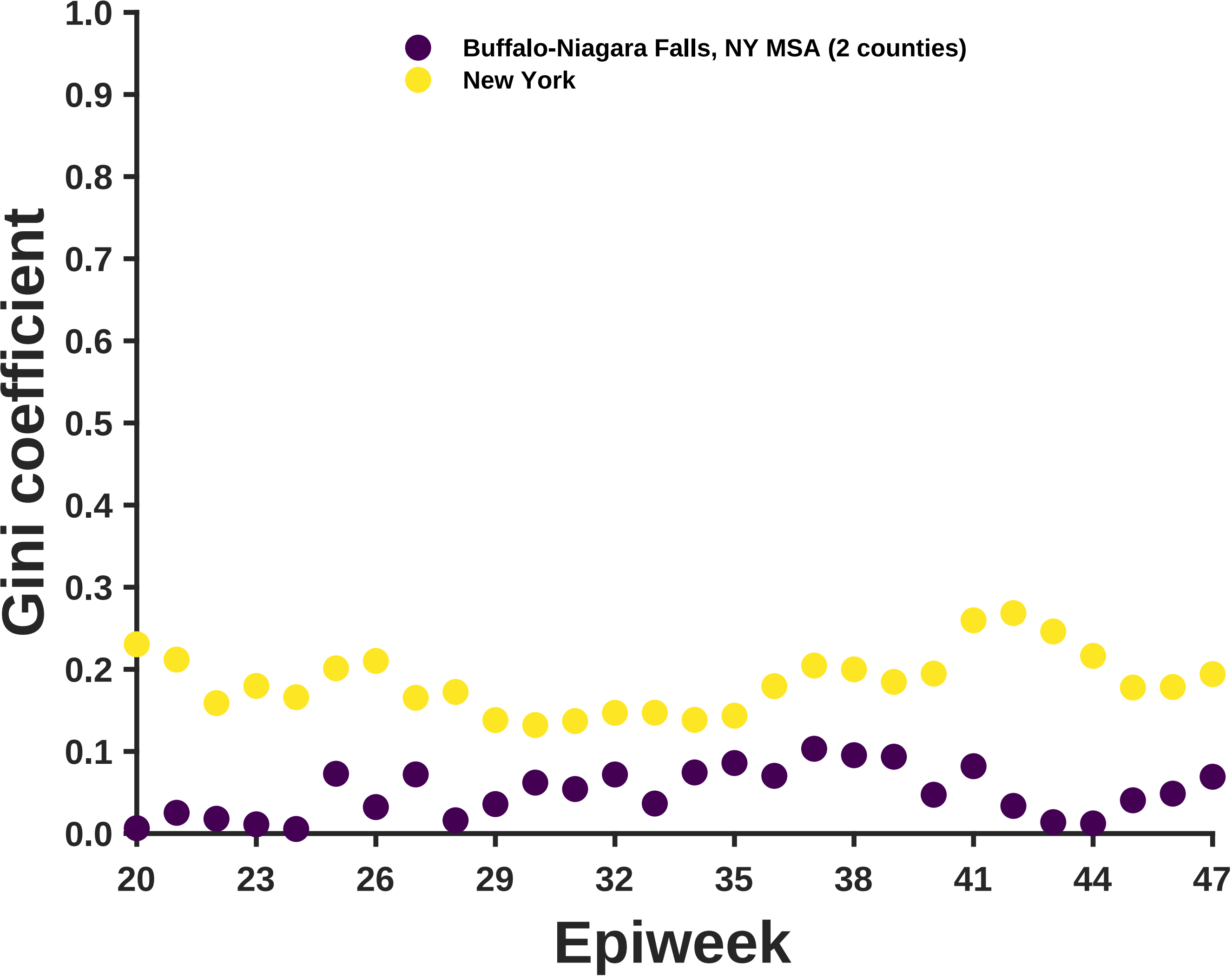

Buffalo-Niagara Falls, NY MSA (2 counties)
New York
Gini coefficient
1.0
0.9
0.8
0.7
0.6
0.5
0.4
0.3
0.2
0.1
0.0
20
23
26
29
32
35
38
41
44
47
Epiweek

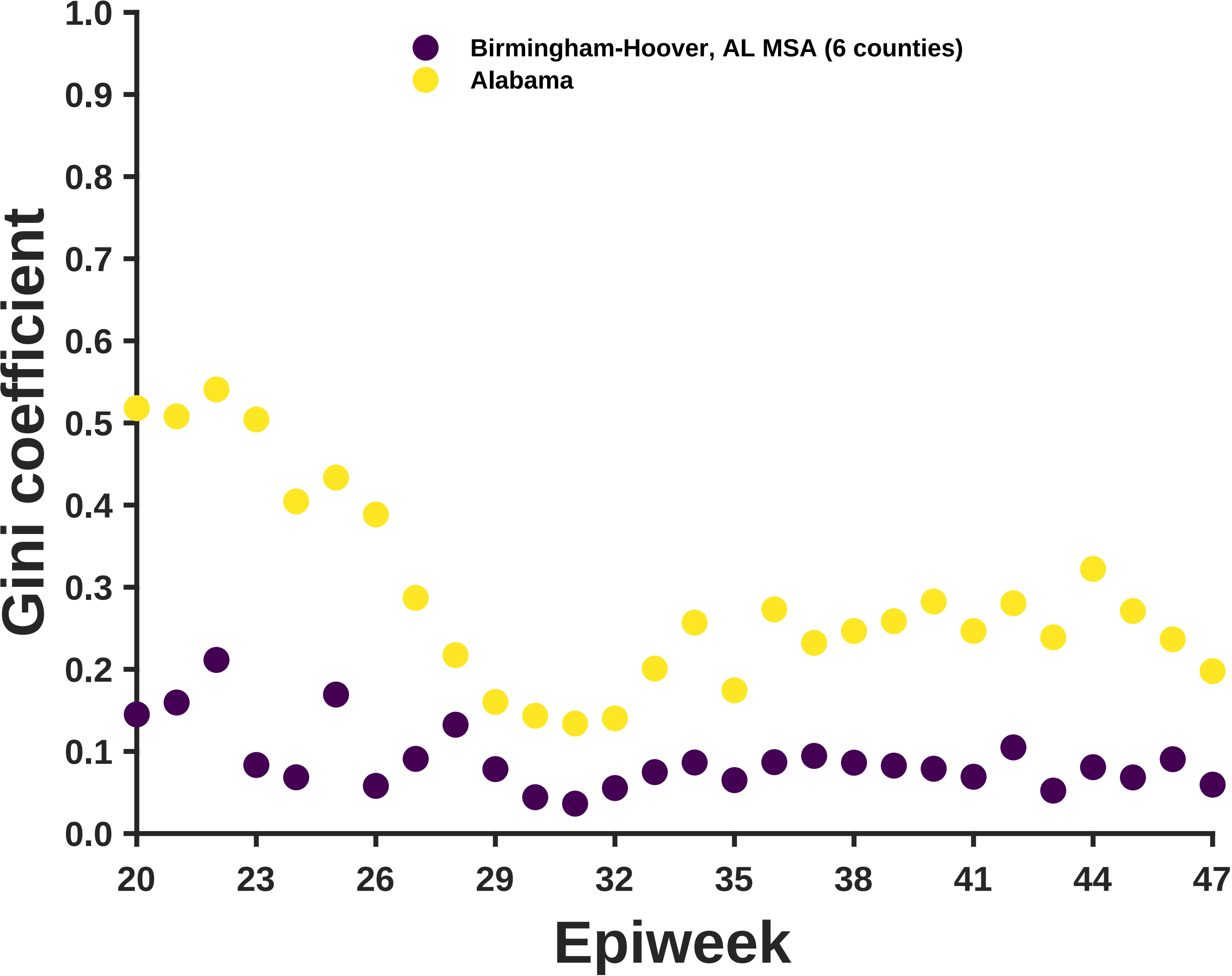

Birmingham-Hoover, AL MSA (6 counties)
Alabama
Gini coefficient
Epiweek
1.0
0.9
0.8
0.7
0.6
0.5
0.4
0.3
0.2
0.1
0.0
20
23
26
29
32
35
38
41
44
47

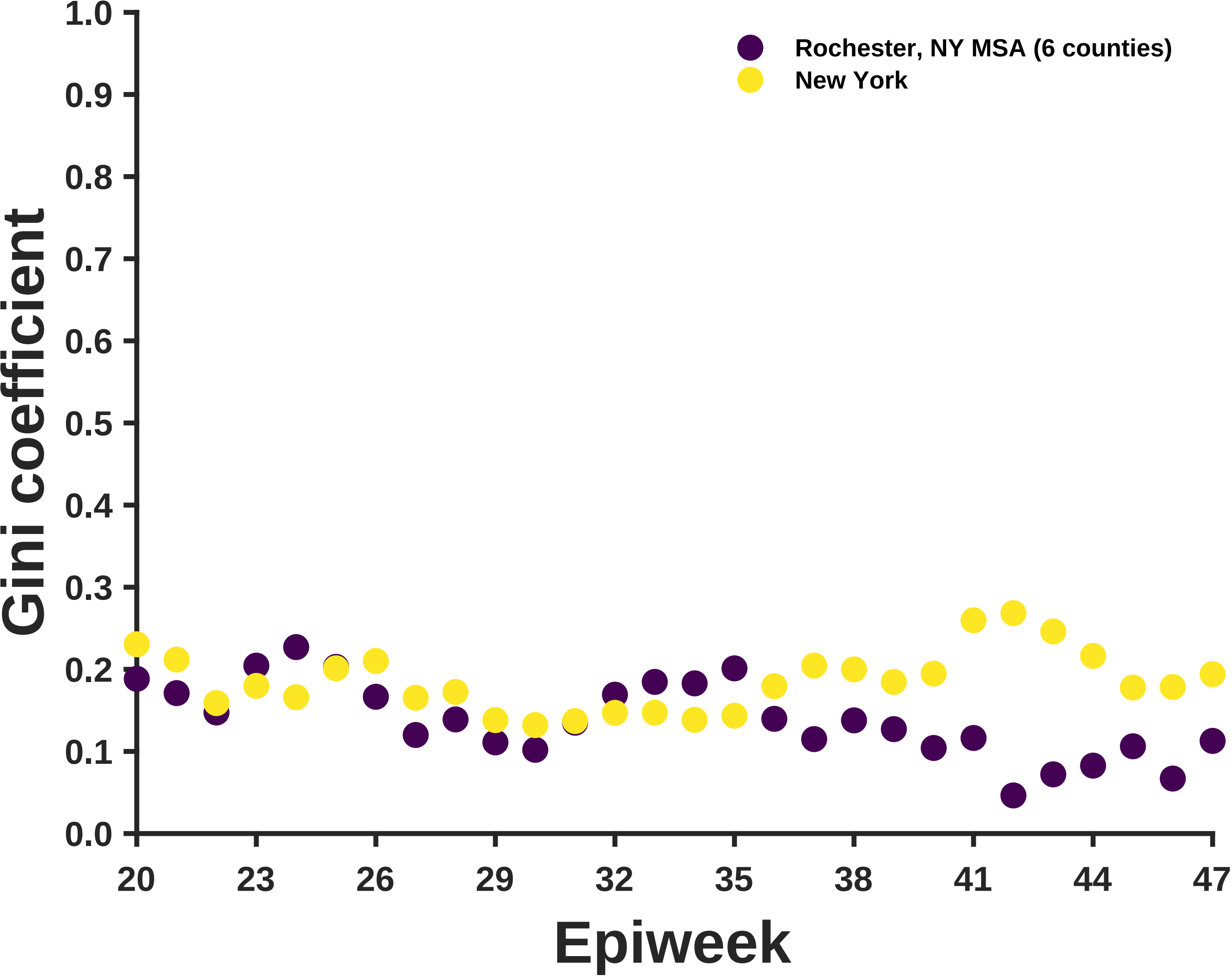

Rochester, NY MSA (6 counties)
New York
Gini coefficient
Epiweek
1.0
0.9
0.8
0.7
0.6
0.5
0.4
0.3
0.2
0.1
0.0
20
23
26
29
32
35
38
41
44
47

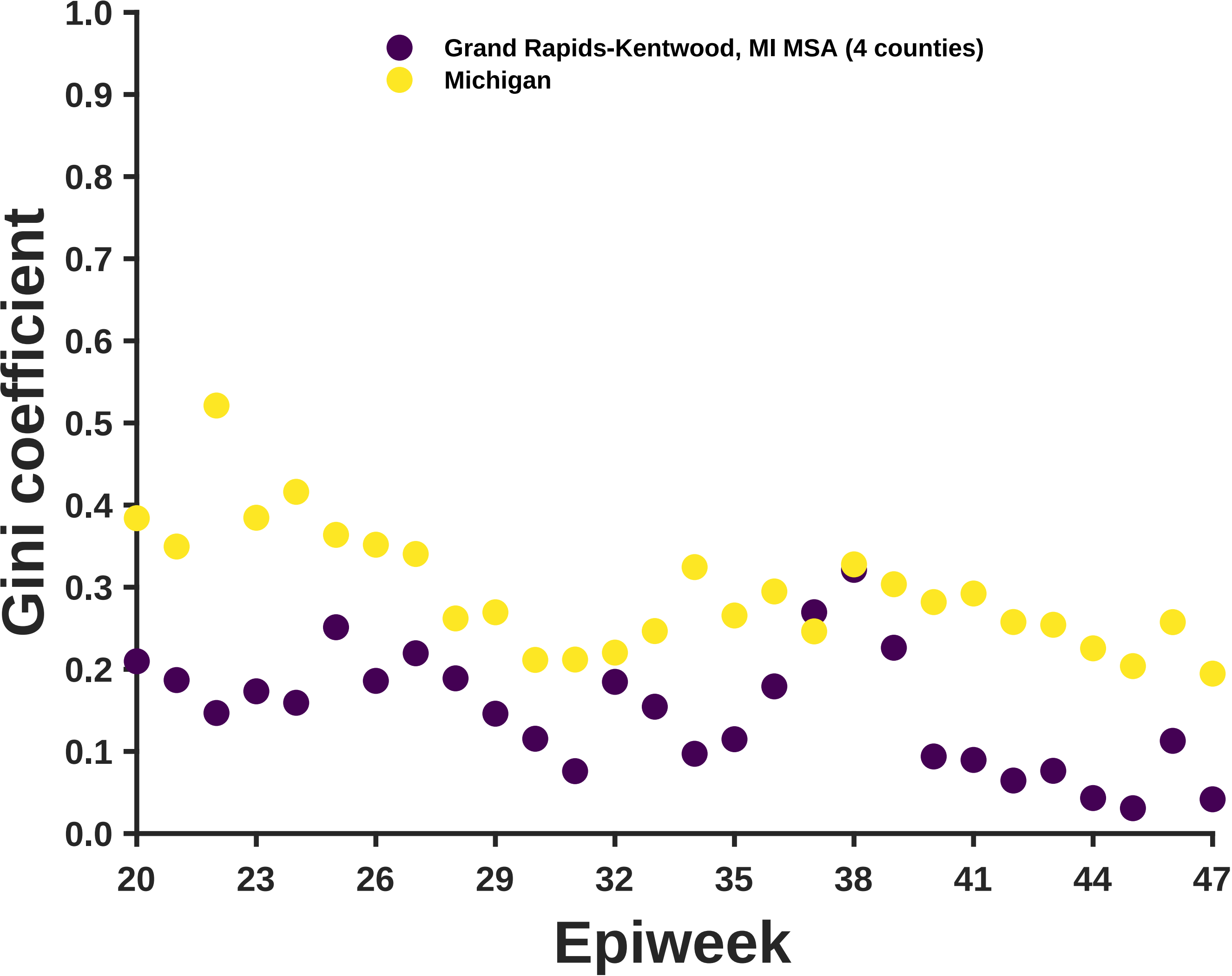

Grand Rapids-Kentwood, MI MSA (4 counties)
Michigan
Gini coefficient
1.0
0.9
0.8
0.7
0.6
0.5
0.4
0.3
0.2
0.1
0.0
20
23
26
29
32
35
38
41
44
47
Epiweek

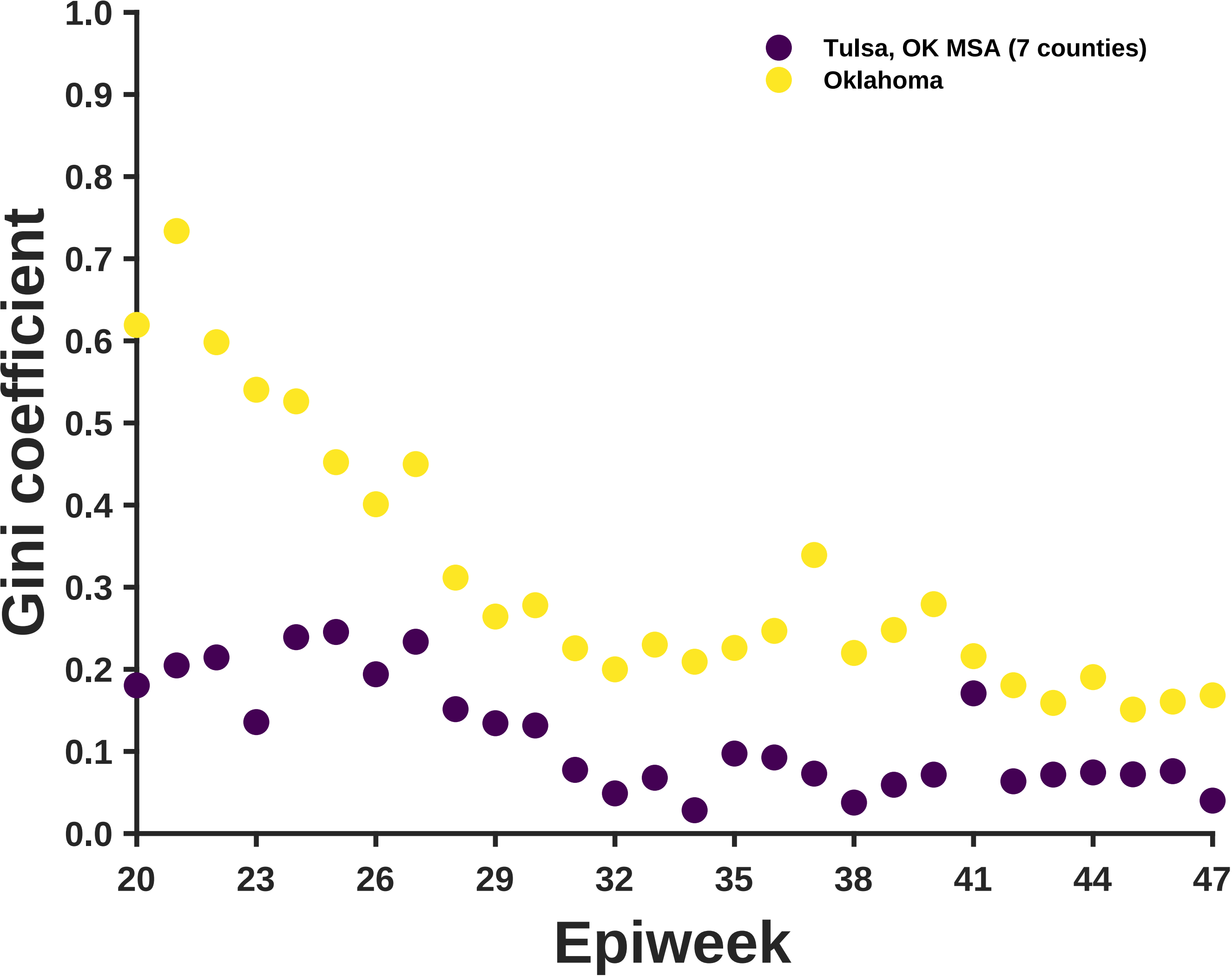
Tulsa, OK MSA (7 counties)
Oklahoma
Gini coefficient
1.0
0.9
0.8
0.7
0.6
0.5
0.4
0.3
0.2
0.1
0.0
20
23
26
29
32
35
38
41
44
47
Epiweek

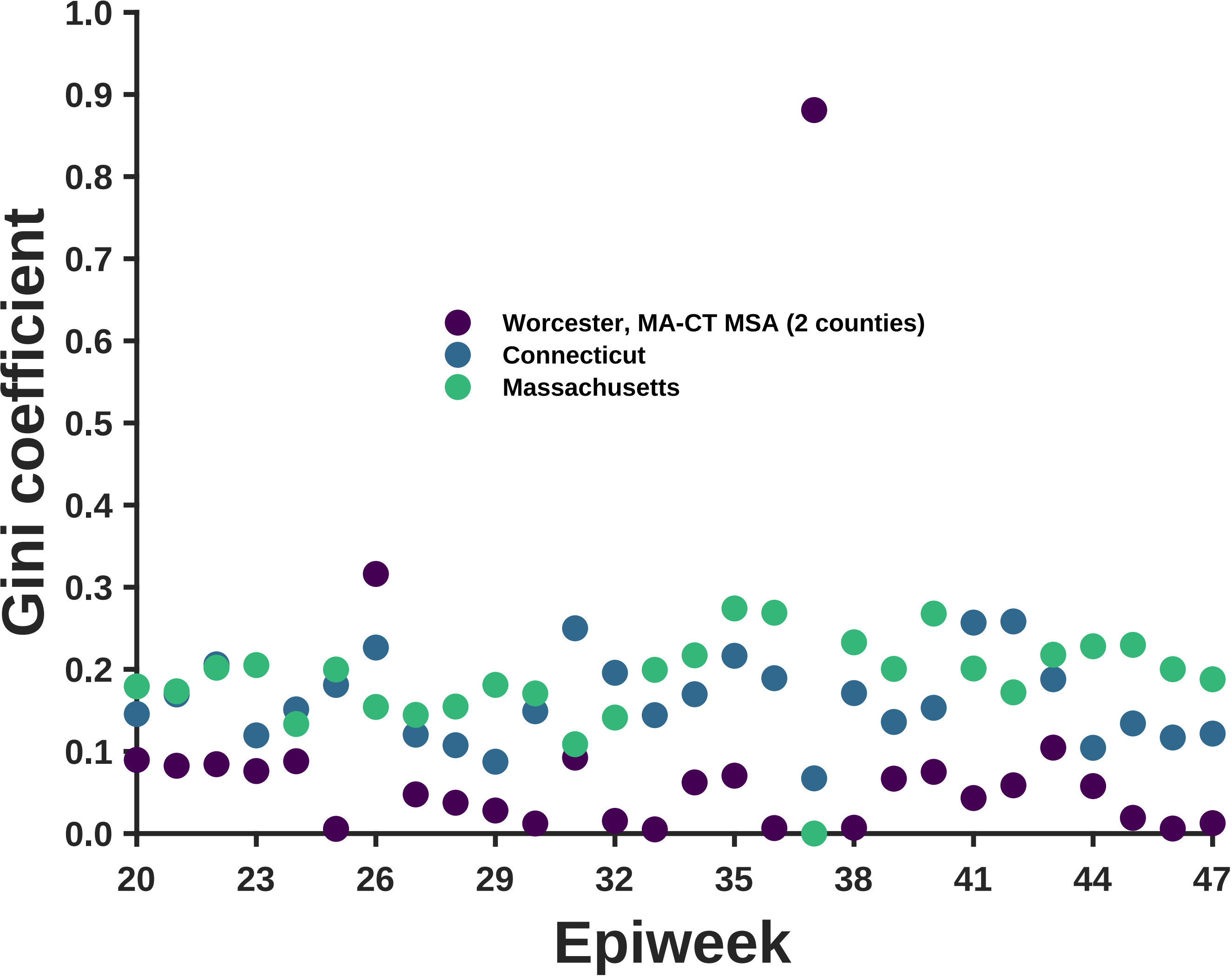

Gini coefficient
1.0
0.9
0.8
0.7
0.6
0.5
0.4
0.3
0.2
0.1
0.0
Worcester, MA-CT MSA (2 counties)
Connecticut
Massachusetts
20
23
26
29
32
35
38
41
44
47
Epiweek

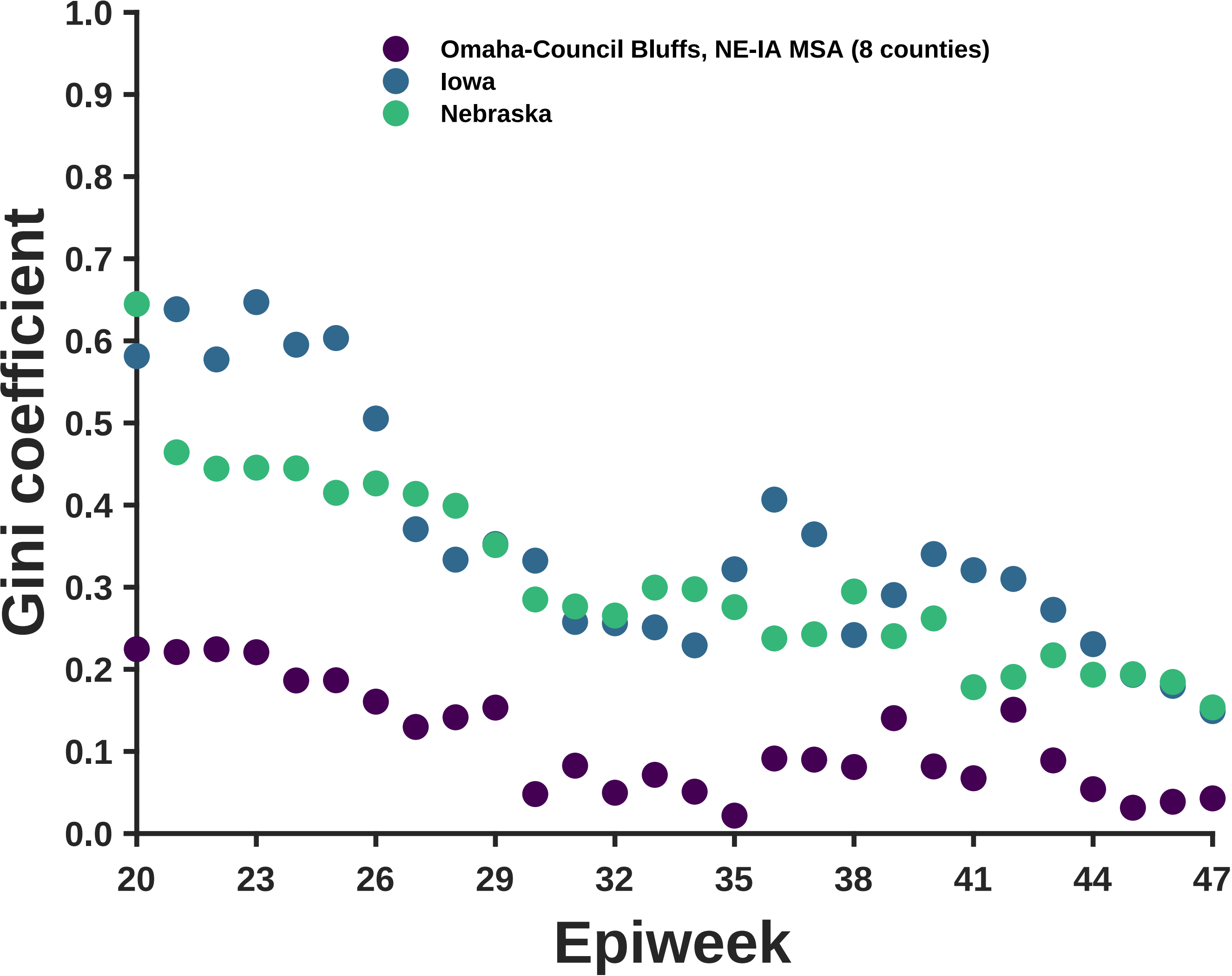
Omaha-Council Bluffs, NE-IA MSA (8 counties)
Iowa
Nebraska
Gini coefficient
Epiweek
0.0
0.1
0.2
0.3
0.4
0.5
0.6
0.7
0.8
0.9
1.0
20
23
26
29
32
35
38
41
44
47

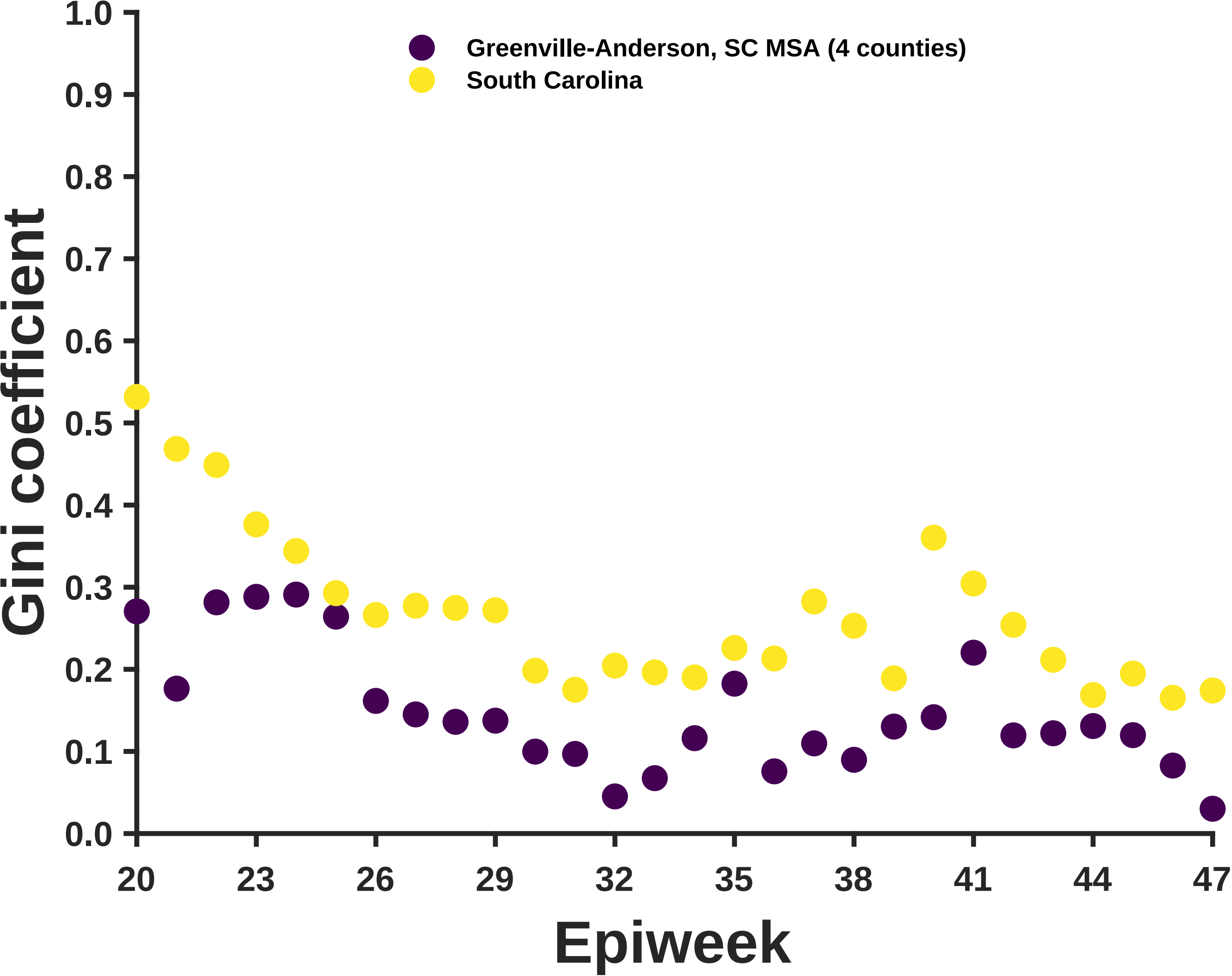

Gini coefficient
1.0
0.9
0.8
0.7
0.6
0.5
0.4
0.3
0.2
0.1
0.0
Greenville-Anderson, SC MSA (4 counties)
South Carolina
20
23
26
29
32
35
38
41
44
47
Epiweek

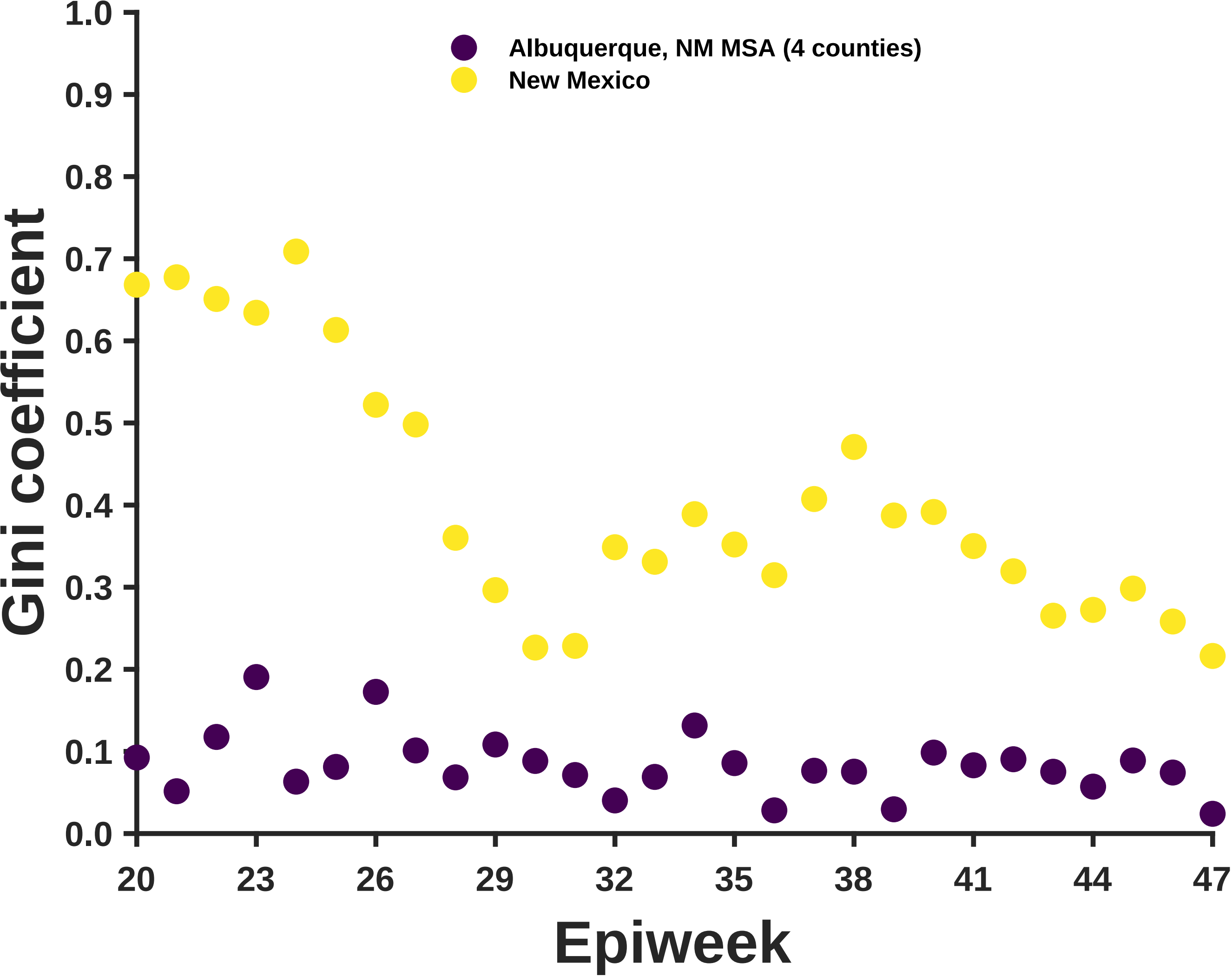

Albuquerque, NM MSA (4 counties)
New Mexico
Gini coefficient
0.0
0.1
0.2
0.3
0.4
0.5
0.6
0.7
0.8
0.9
1.0
20
23
26
29
32
35
38
41
44
47
Epiweek

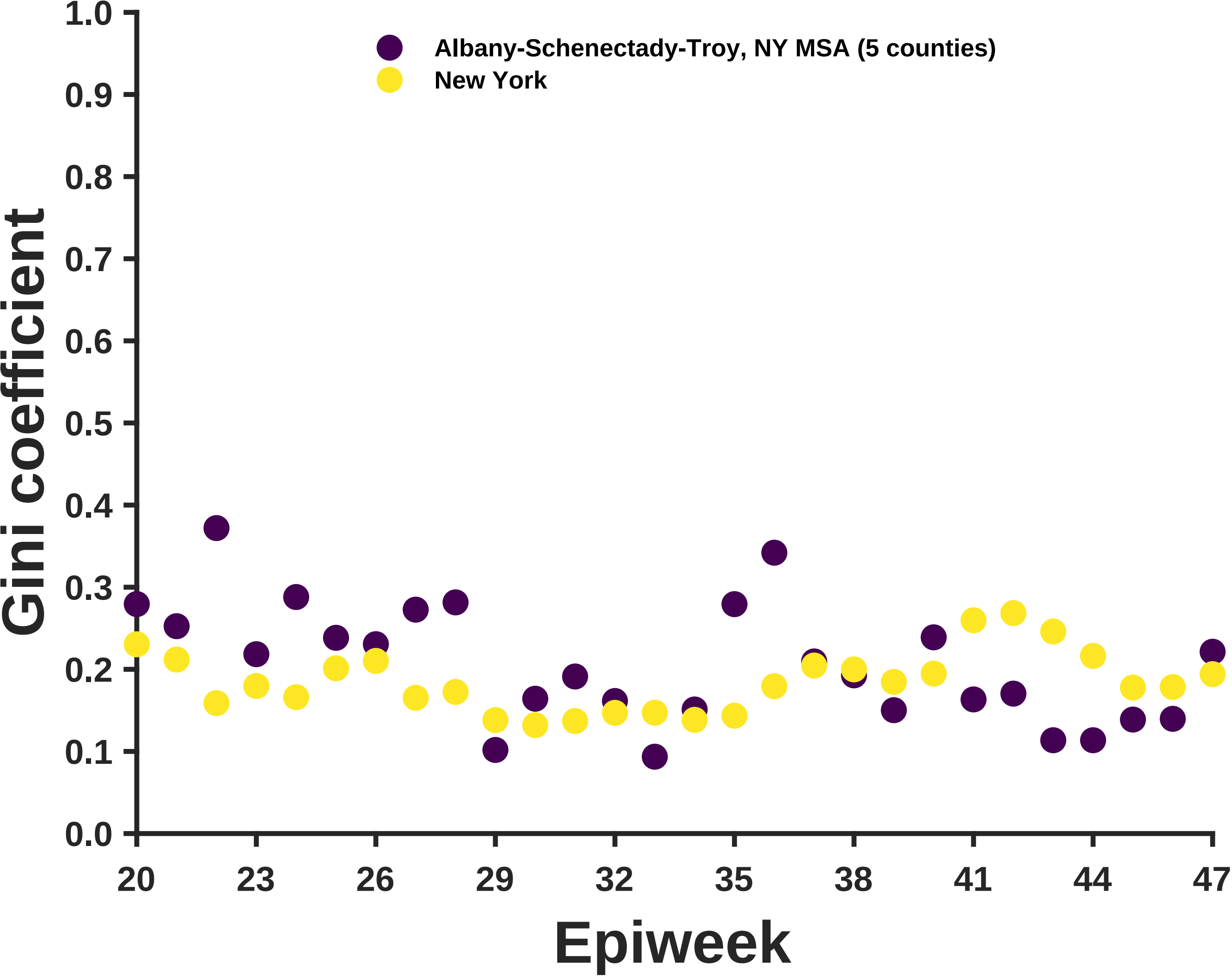
Albany-Schenectady-Troy, NY MSA (5 counties)
New York
Gini coefficient
Epiweek
1.0
0.9
0.8
0.7
0.6
0.5
0.4
0.3
0.2
0.1
0.0
20
23
26
29
32
35
38
41
44
47

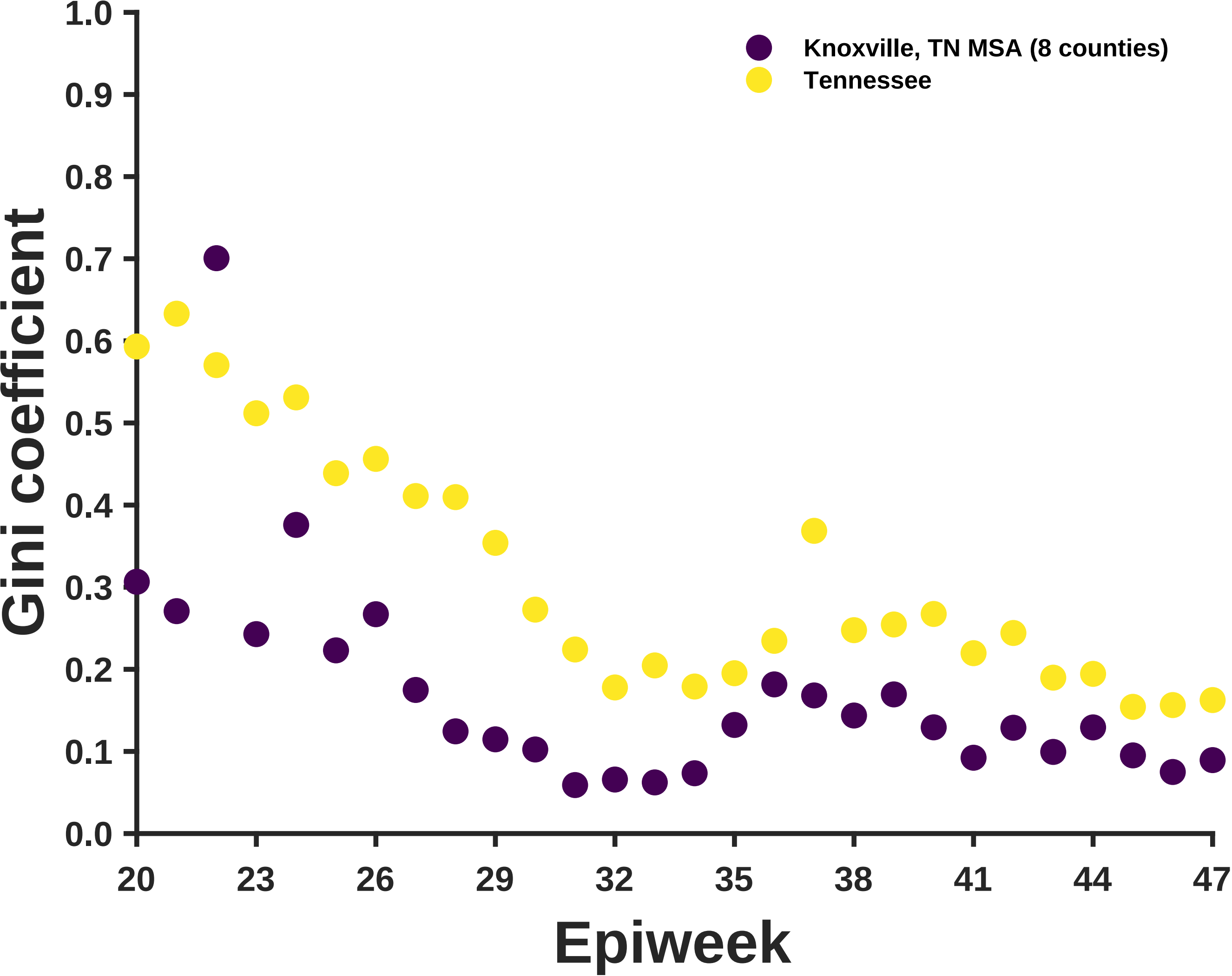

Knoxville, TN MSA (8 counties)
Tennessee
Gini coefficient
Epiweek
1.0
0.9
0.8
0.7
0.6
0.5
0.4
0.3
0.2
0.1
0.0
20
23
26
29
32
35
38
41
44
47

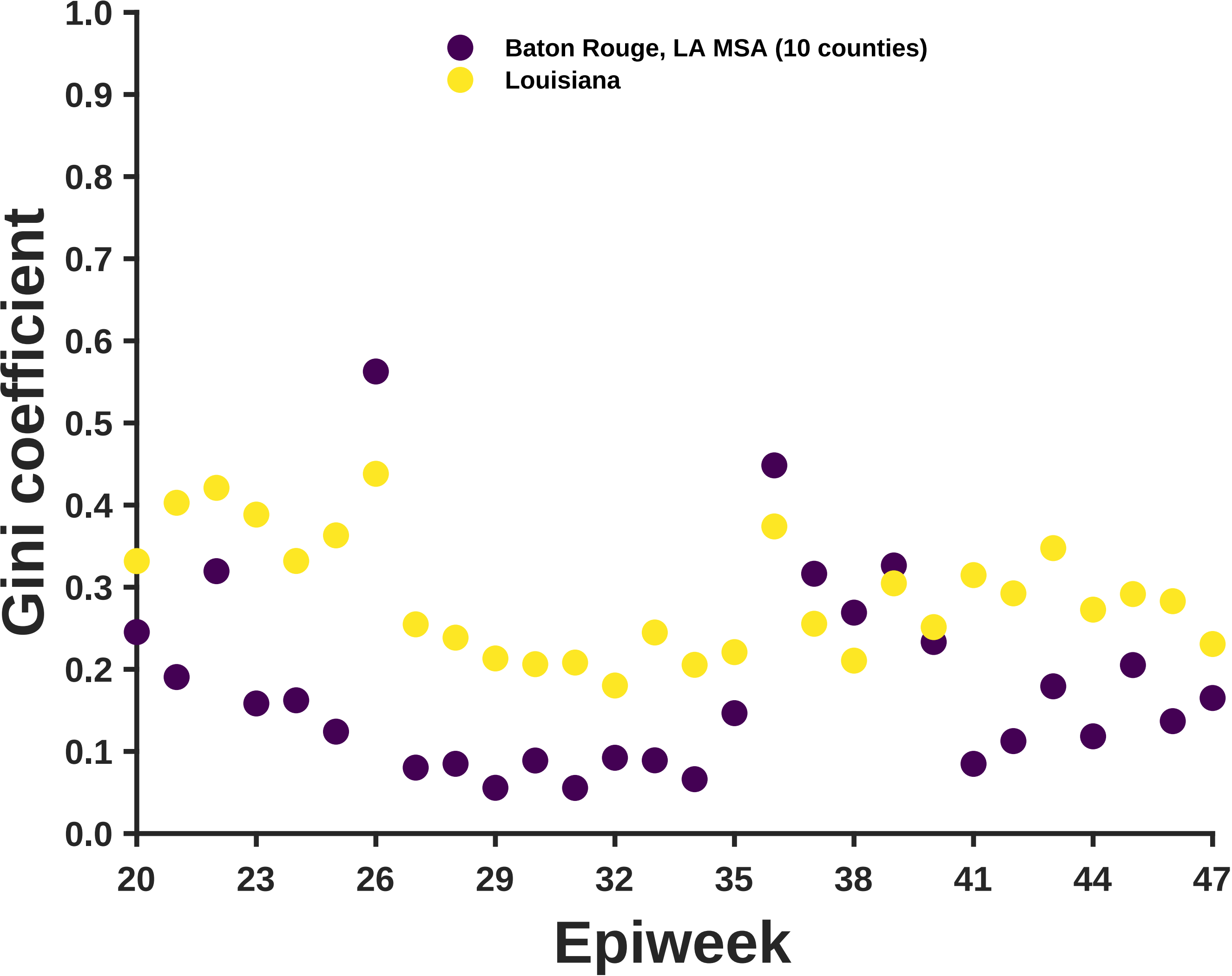

Baton Rouge, LA MSA (10 counties)
Louisiana
Gini coefficient
1.0
0.9
0.8
0.7
0.6
0.5
0.4
0.3
0.2
0.1
0.0
20
23
26
29
32
35
38
41
44
47
Epiweek

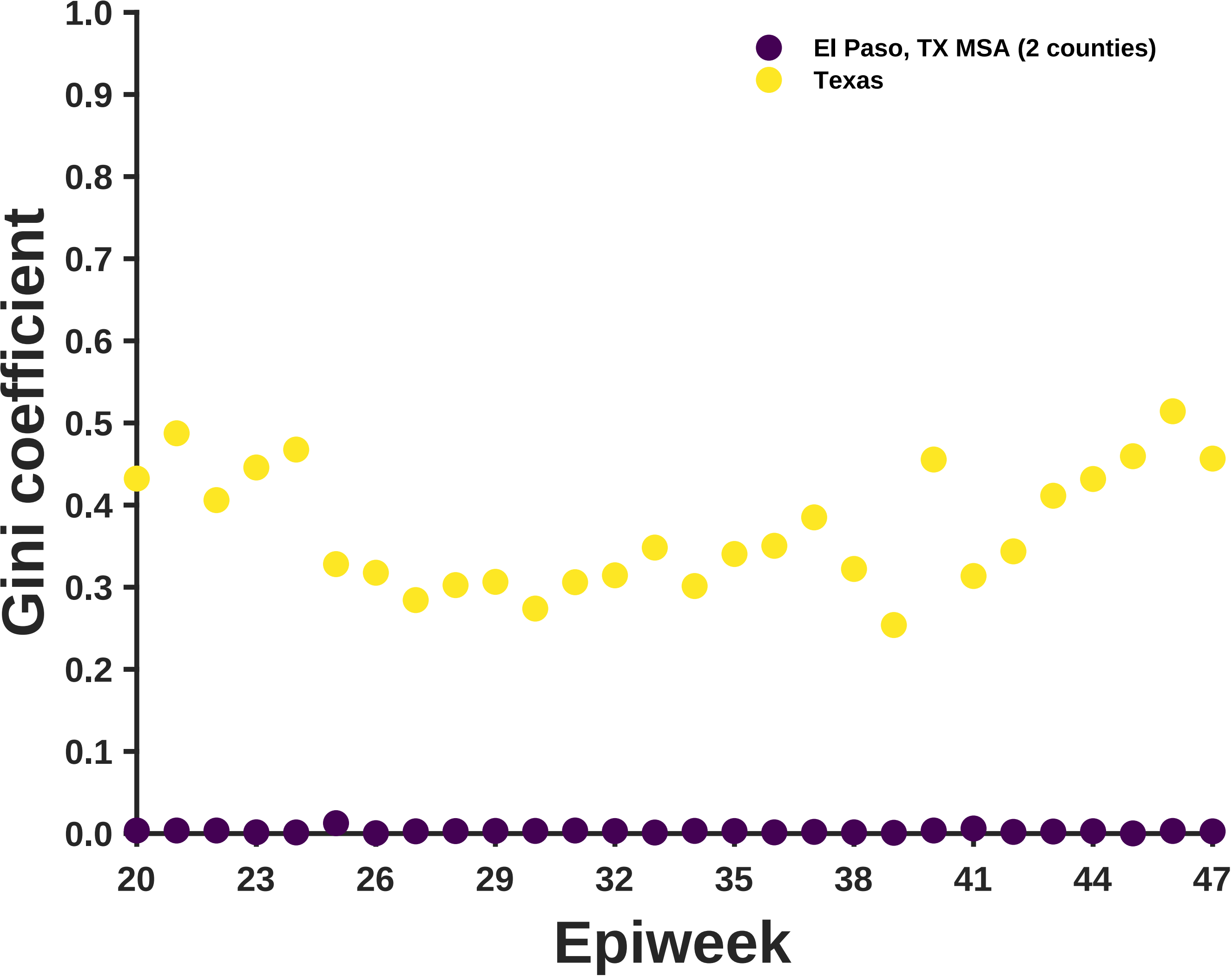
El Paso, TX MSA (2 counties)
Texas
Gini coefficient
1.0
0.9
0.8
0.7
0.6
0.5
0.4
0.3
0.2
0.1
0.0
20
23
26
29
32
35
38
41
44
47
Epiweek

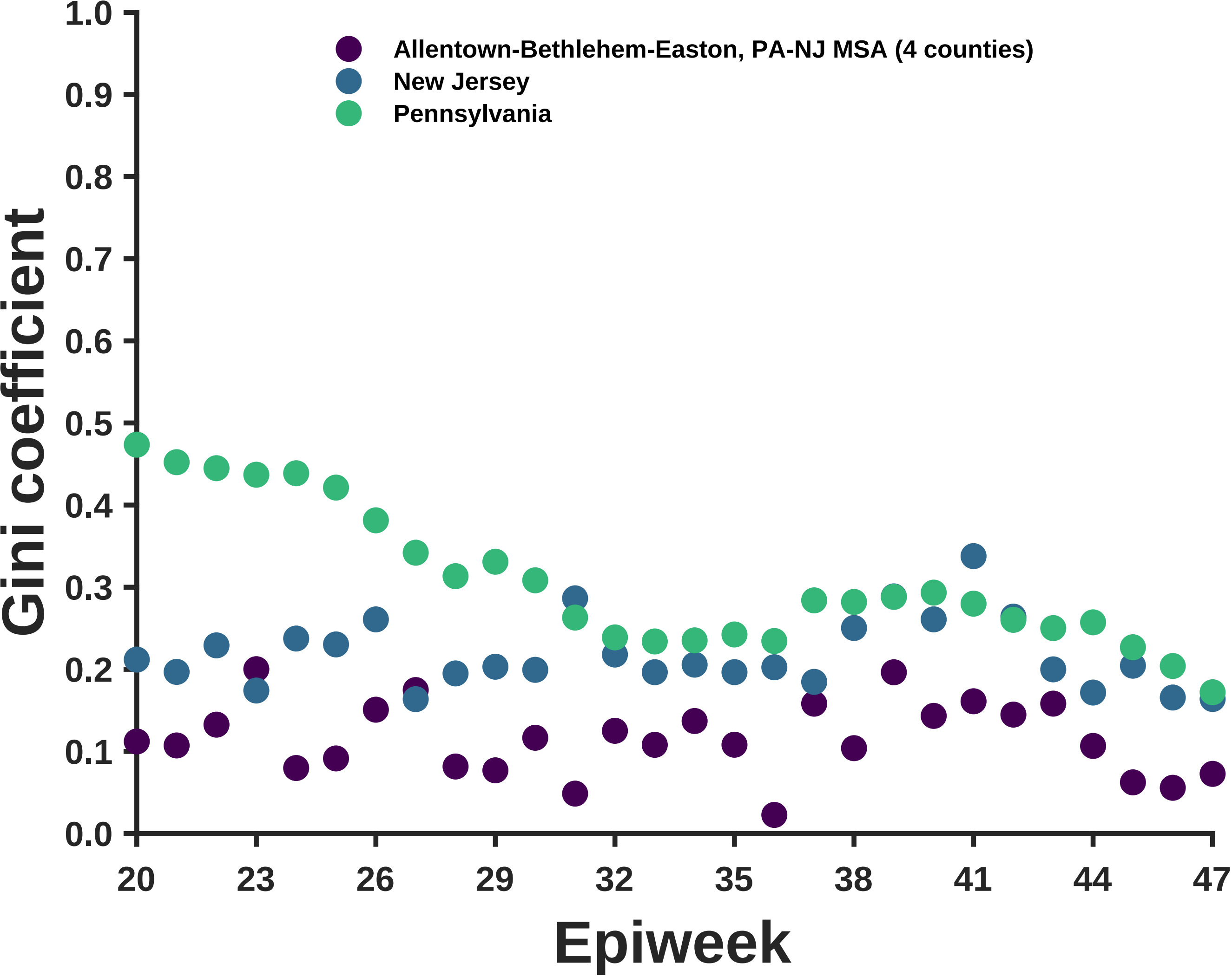

Allentown-Bethlehem-Easton, PA-NJ MSA (4 counties)
New Jersey
Pennsylvania
Gini coefficient
1.0
0.9
0.8
0.7
0.6
0.5
0.4
0.3
0.2
0.1
0.0
20
23
26
29
32
35
38
41
44
47
Epiweek

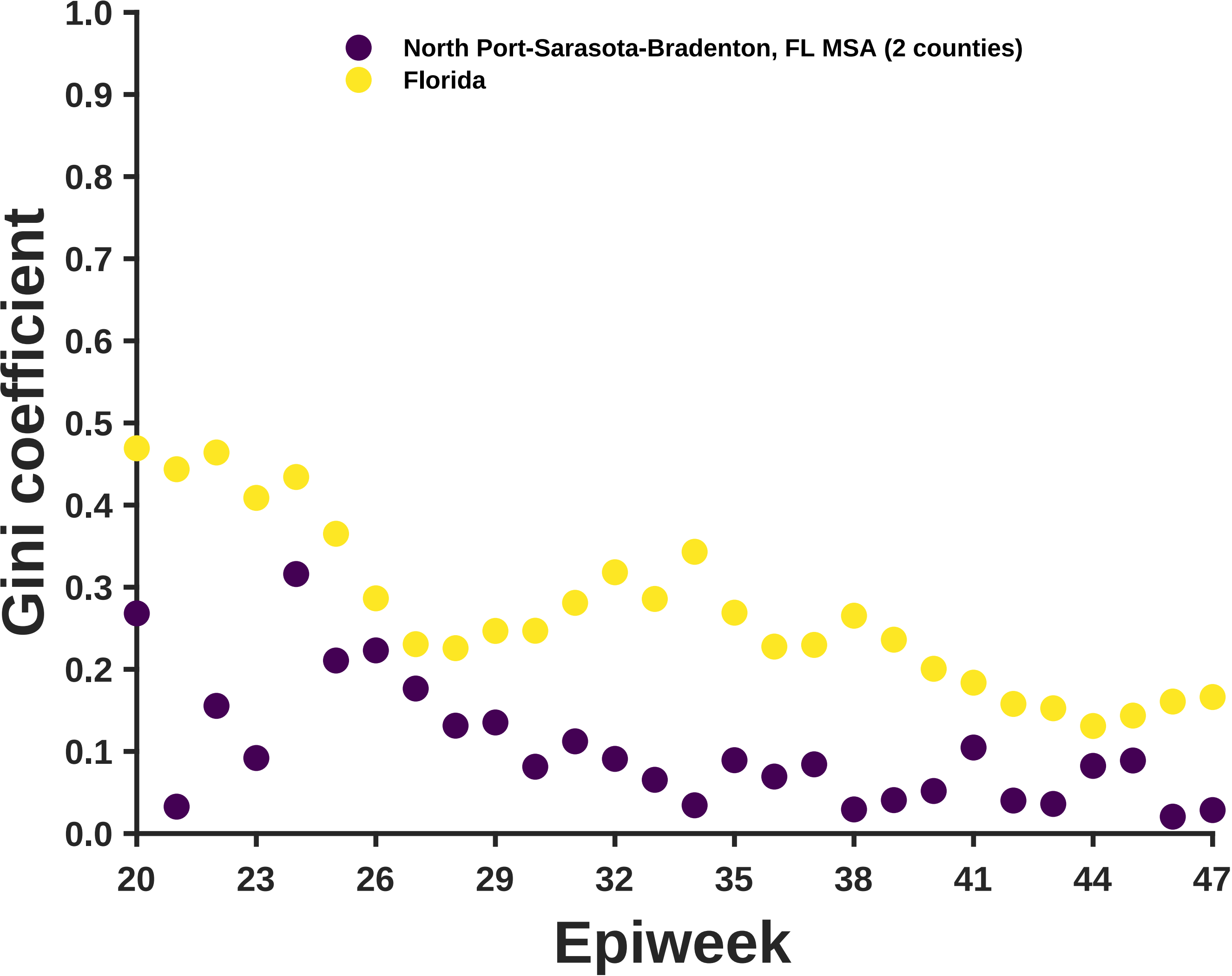
North Port-Sarasota-Bradenton, FL MSA (2 counties)
Florida
Gini coefficient
1.0
0.9
0.8
0.7
0.6
0.5
0.4
0.3
0.2
0.1
0.0
20
23
26
29
32
35
38
41
44
47
Epiweek

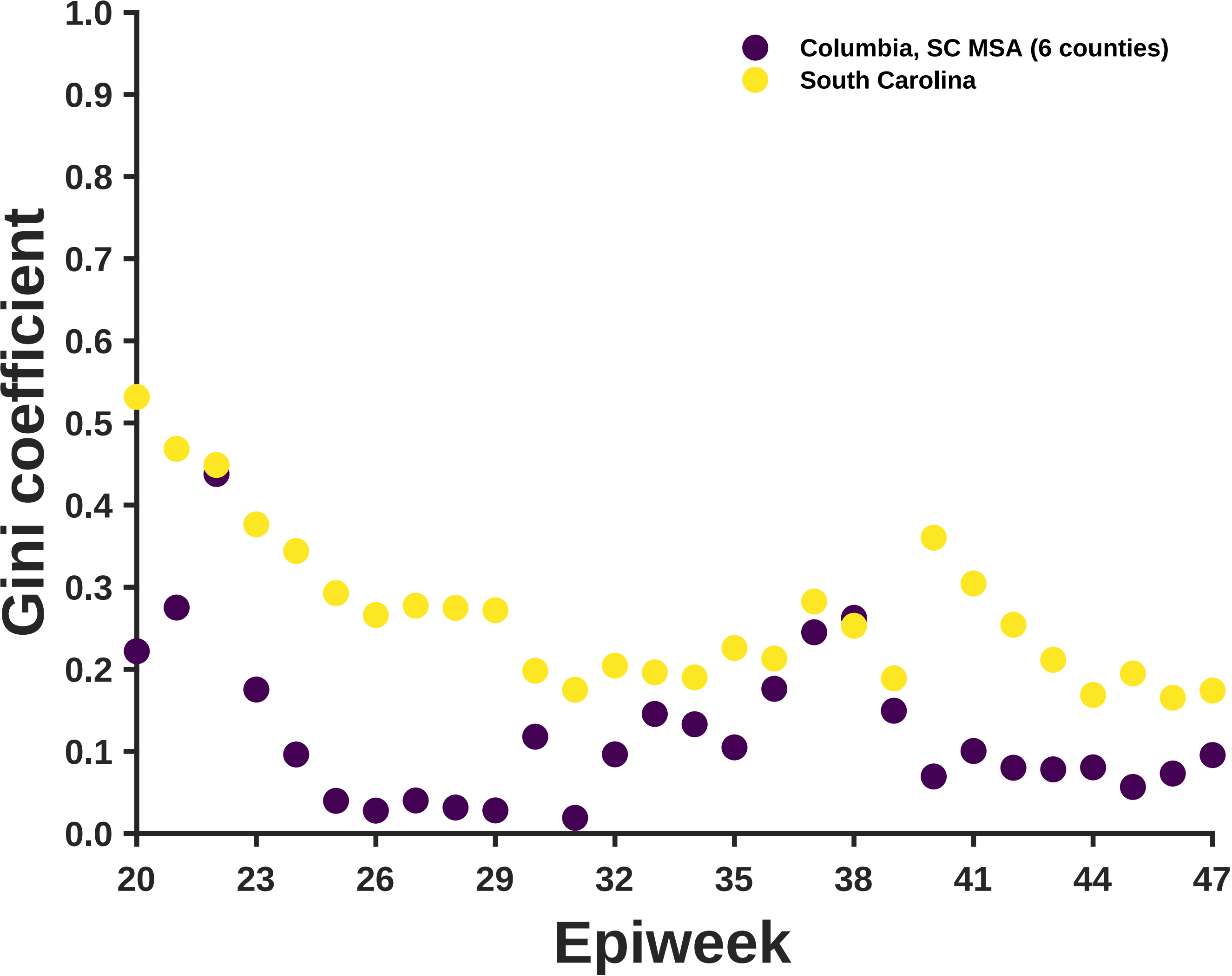

Columbia, SC MSA (6 counties)
South Carolina
Gini coefficient
1.0
0.9
0.8
0.7
0.6
0.5
0.4
0.3
0.2
0.1
0.0
20
23
26
29
32
35
38
41
44
47
Epiweek

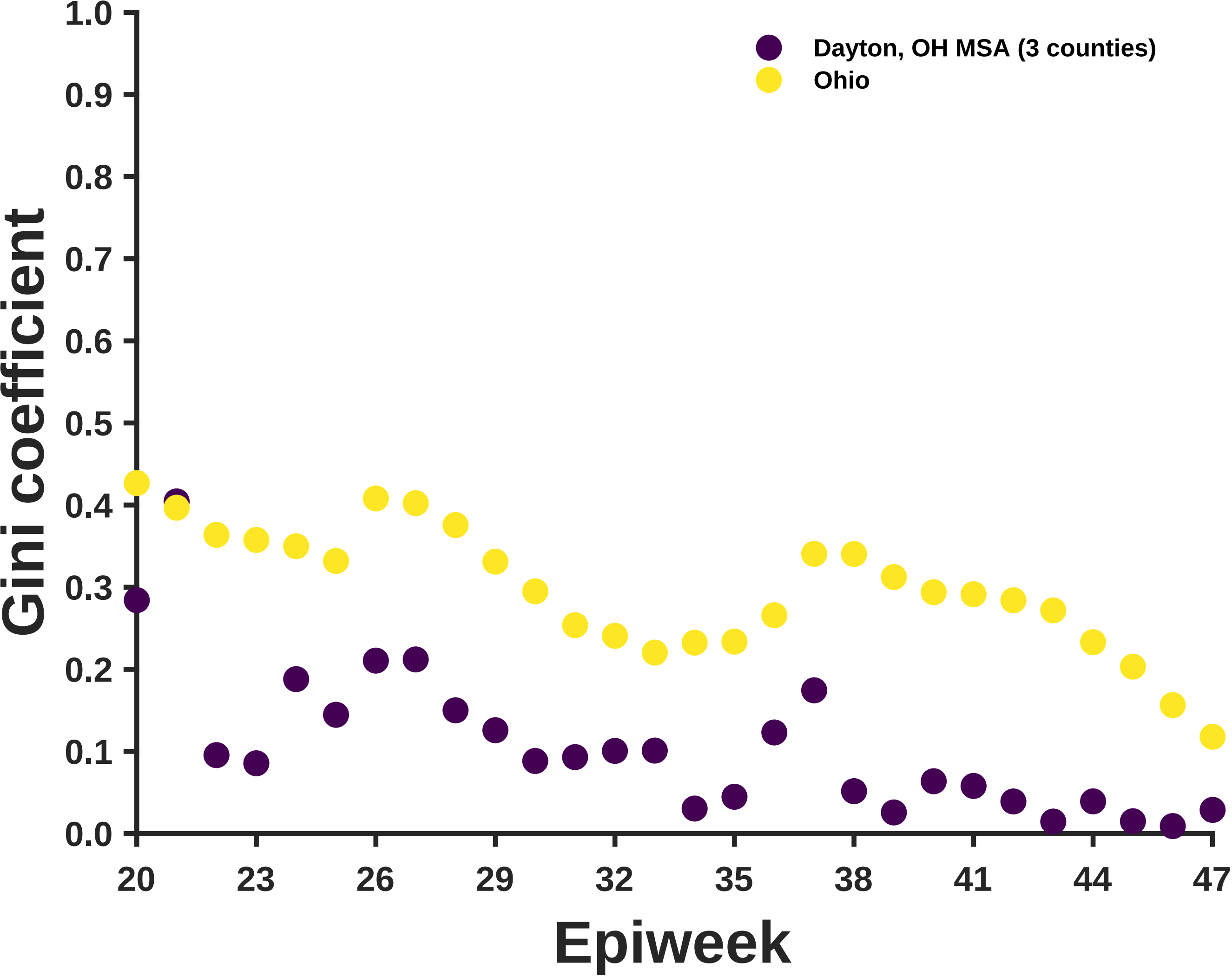

Dayton, OH MSA (3 counties)
Ohio
Gini coefficient
Epiweek
1.0
0.9
0.8
0.7
0.6
0.5
0.4
0.3
0.2
0.1
0.0
20
23
26
29
32
35
38
41
44
47

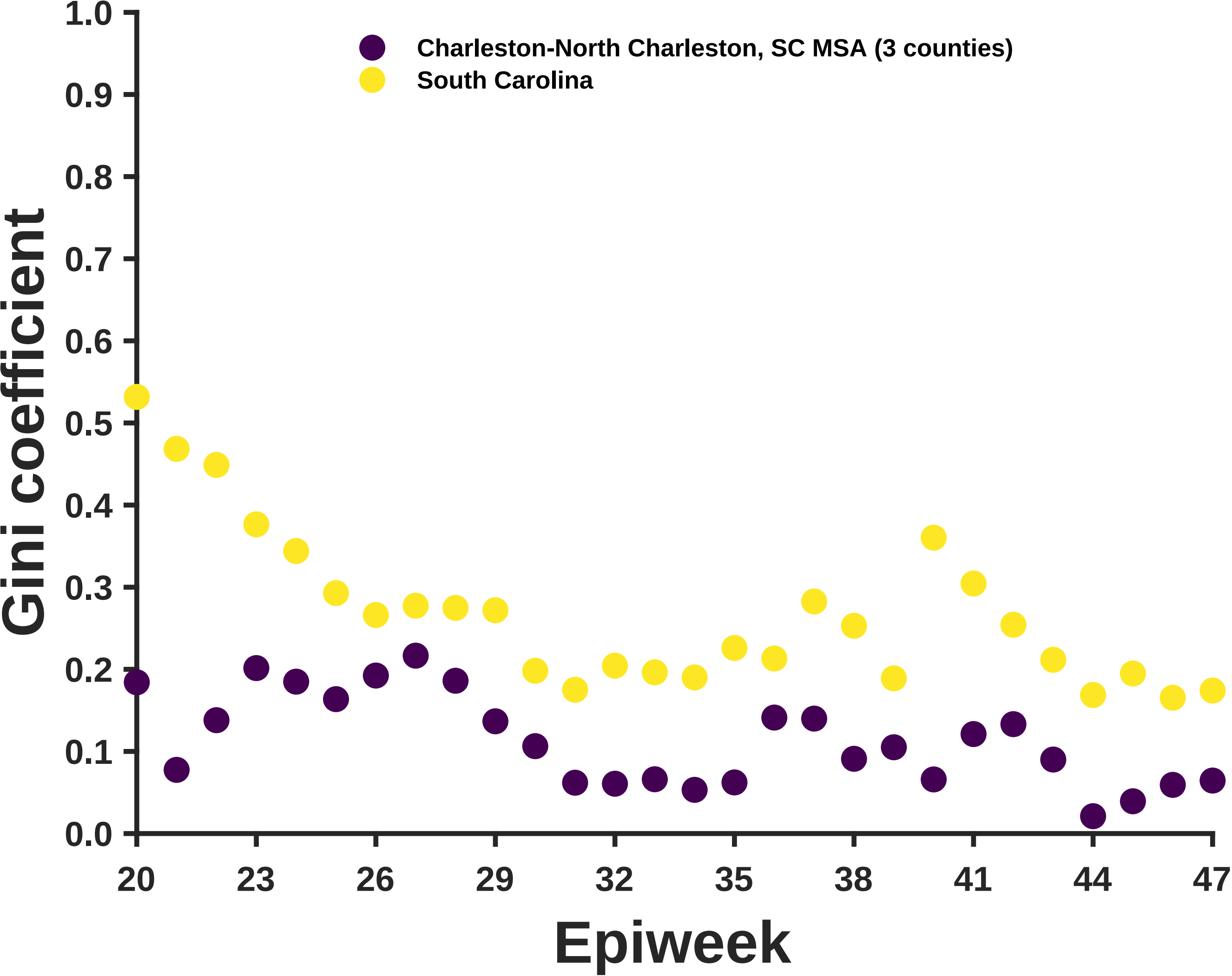

Charleston-North Charleston, SC MSA (3 counties)
South Carolina
Gini coefficient
0.0
0.1
0.2
0.3
0.4
0.5
0.6
0.7
0.8
0.9
1.0
20
23
26
29
32
35
38
41
44
47
Epiweek

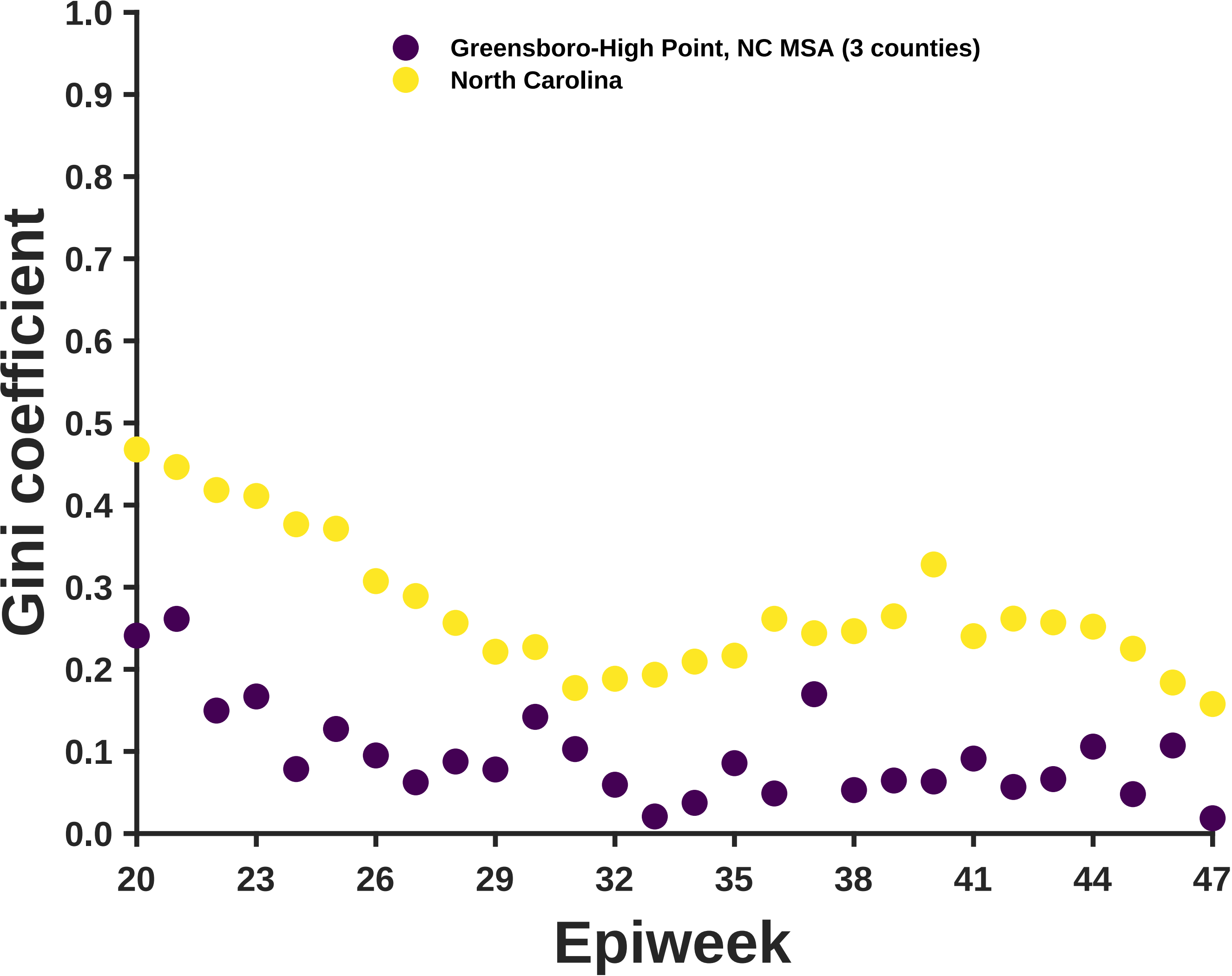
Greensboro-High Point, NC MSA (3 counties)
North Carolina
Gini coefficient
1.0
0.9
0.8
0.7
0.6
0.5
0.4
0.3
0.2
0.1
0.0
20
23
26
29
32
35
38
41
44
47
Epiweek

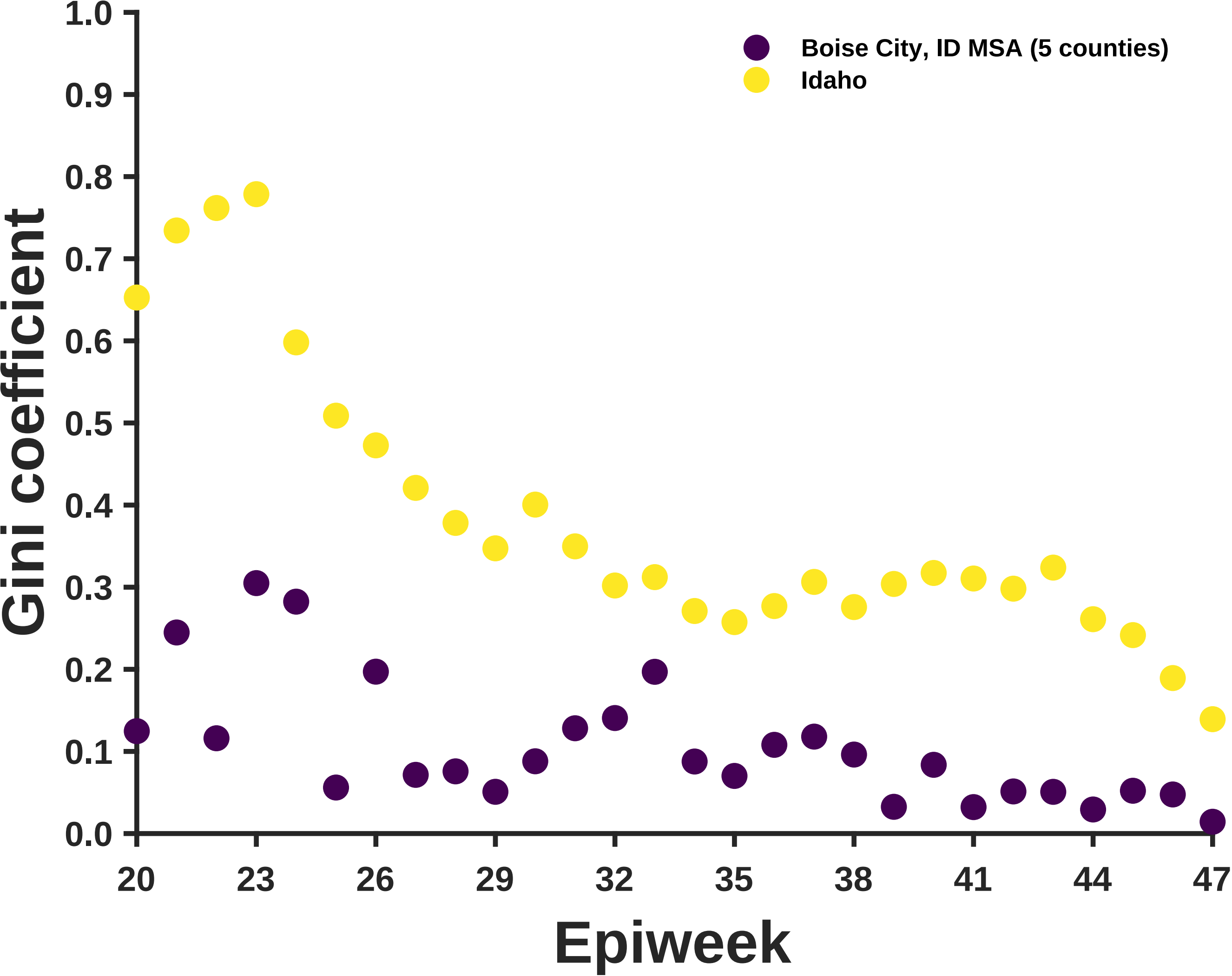
Boise City, ID MSA (5 counties)
Idaho
Gini coefficient
Epiweek
0.0
0.1
0.2
0.3
0.4
0.5
0.6
0.7
0.8
0.9
1.0
20
23
26
29
32
35
38
41
44
47

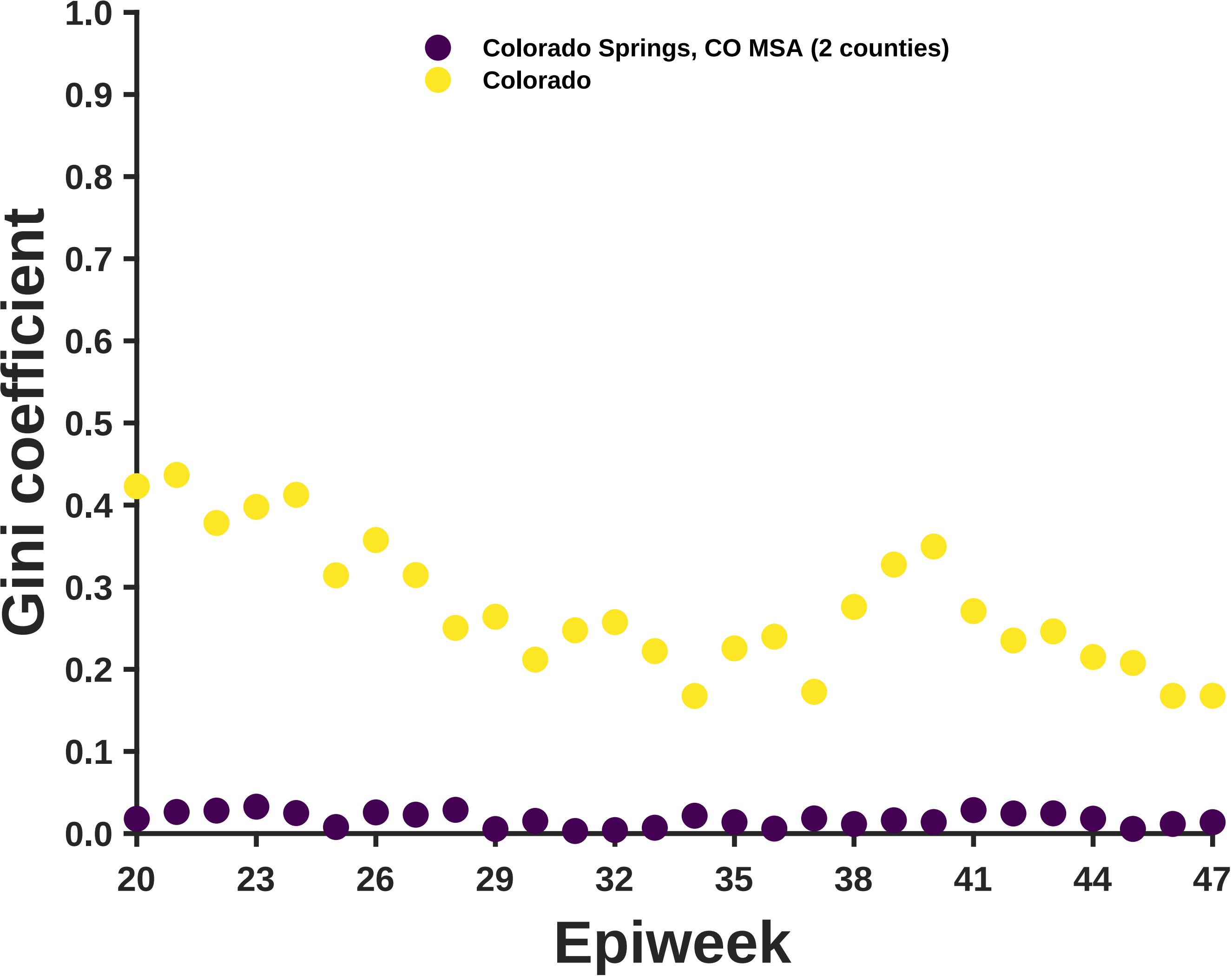
Colorado Springs, CO MSA (2 counties)
Colorado
Gini coefficient
1.0
0.9
0.8
0.7
0.6
0.5
0.4
0.3
0.2
0.1
0.0
20
23
26
29
32
35
38
41
44
47
Epiweek

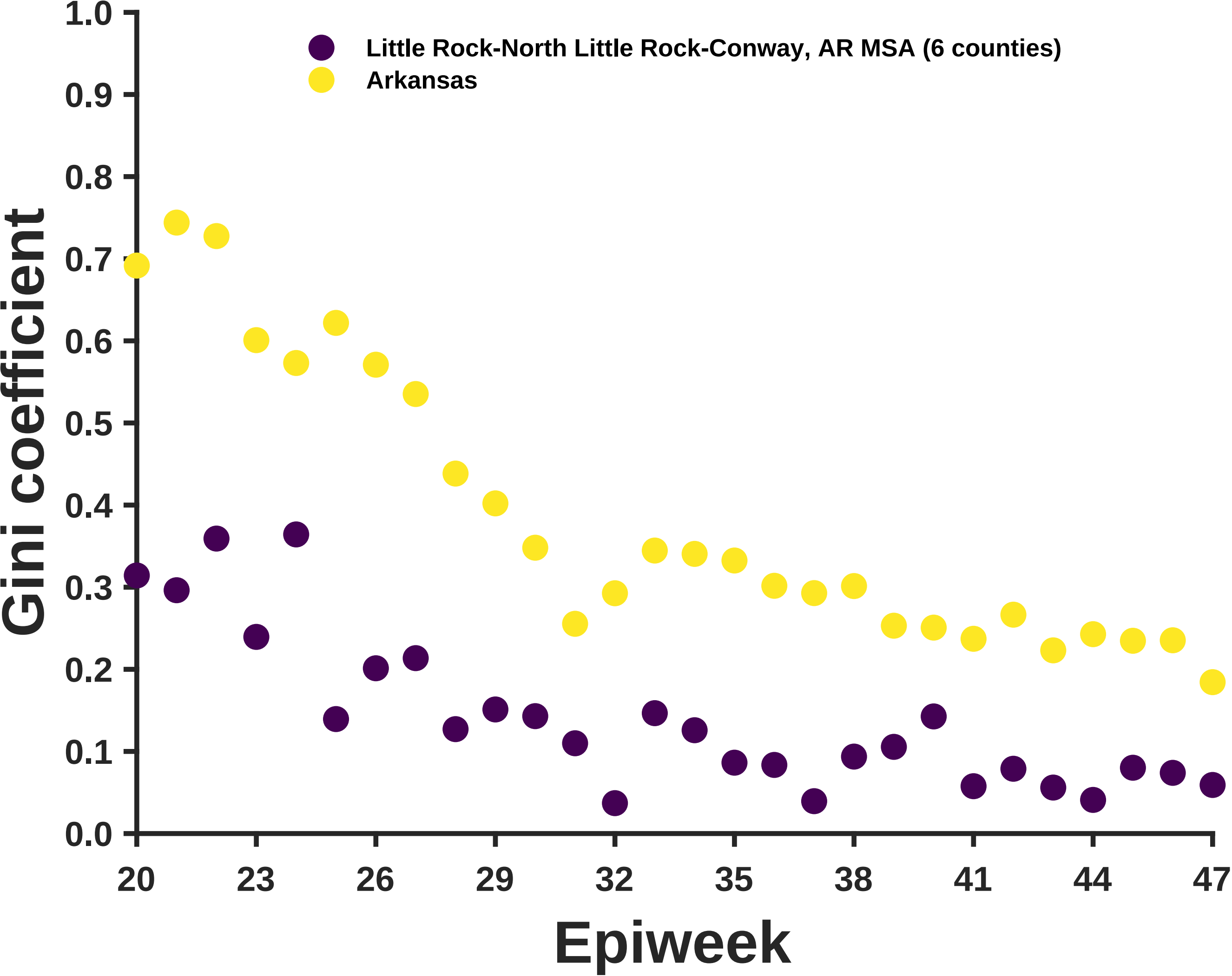

Little Rock-North Little Rock-Conway, AR MSA (6 counties)
Arkansas
Gini coefficient
Epiweek
1.0
0.9
0.8
0.7
0.6
0.5
0.4
0.3
0.2
0.1
0.0
20
23
26
29
32
35
38
41
44
47

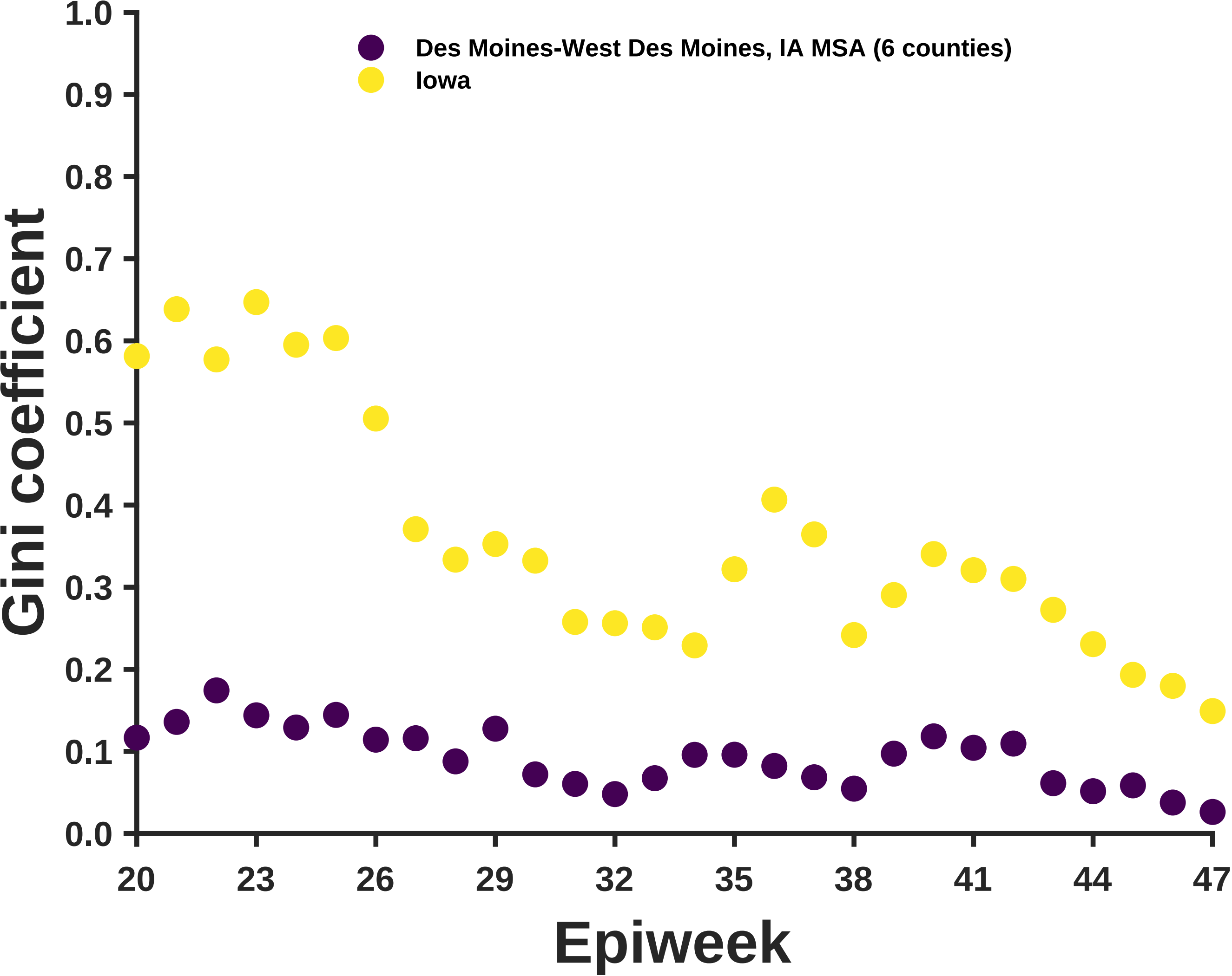

Des Moines-West Des Moines, IA MSA (6 counties)
Iowa
Gini coefficient
1.0
0.9
0.8
0.7
0.6
0.5
0.4
0.3
0.2
0.1
0.0
20
23
26
29
32
35
38
41
44
47
Epiweek

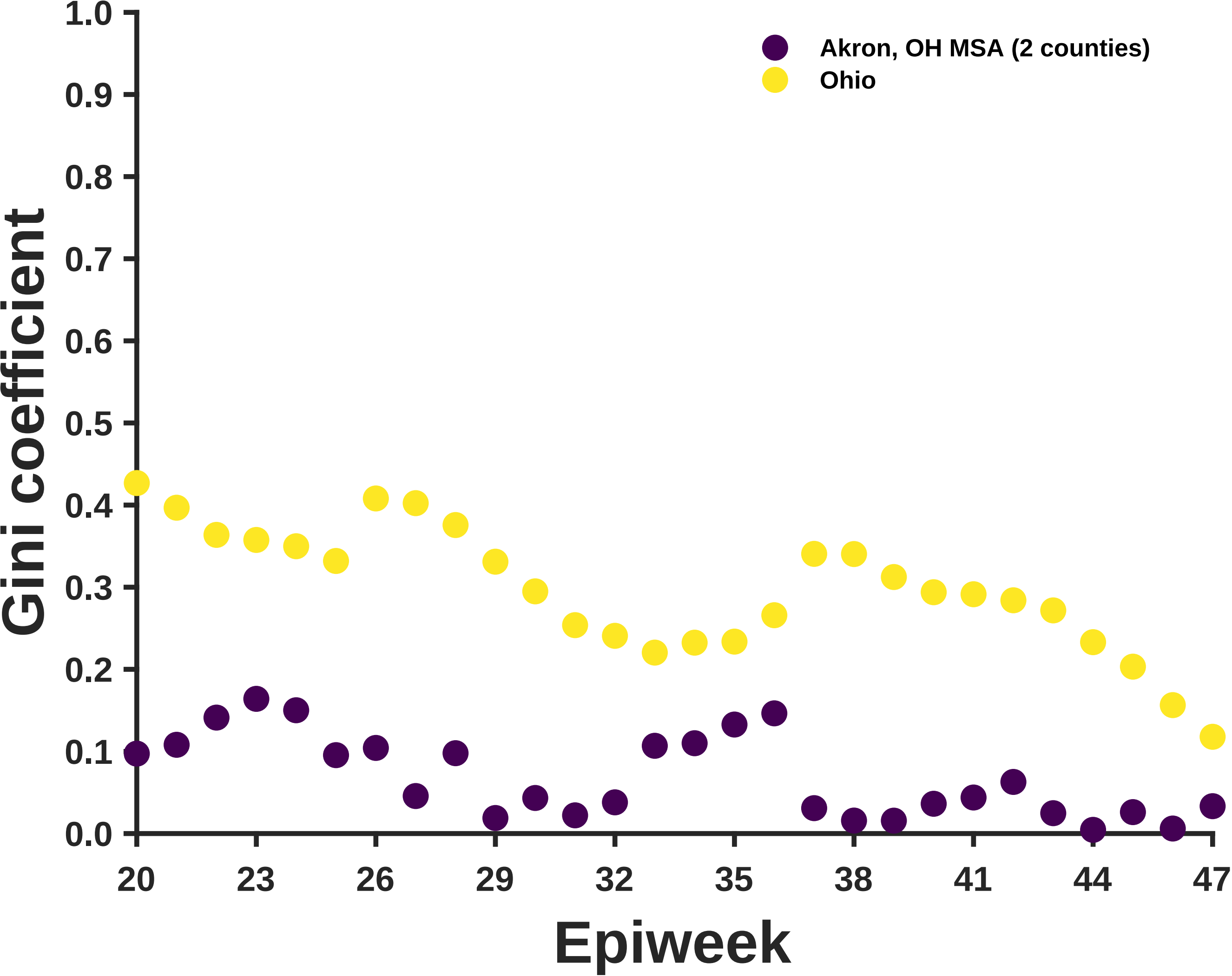

Akron, OH MSA (2 counties)
Ohio
Gini coefficient
0.0
0.1
0.2
0.3
0.4
0.5
0.6
0.7
0.8
0.9
1.0
20
23
26
29
32
35
38
41
44
47
Epiweek

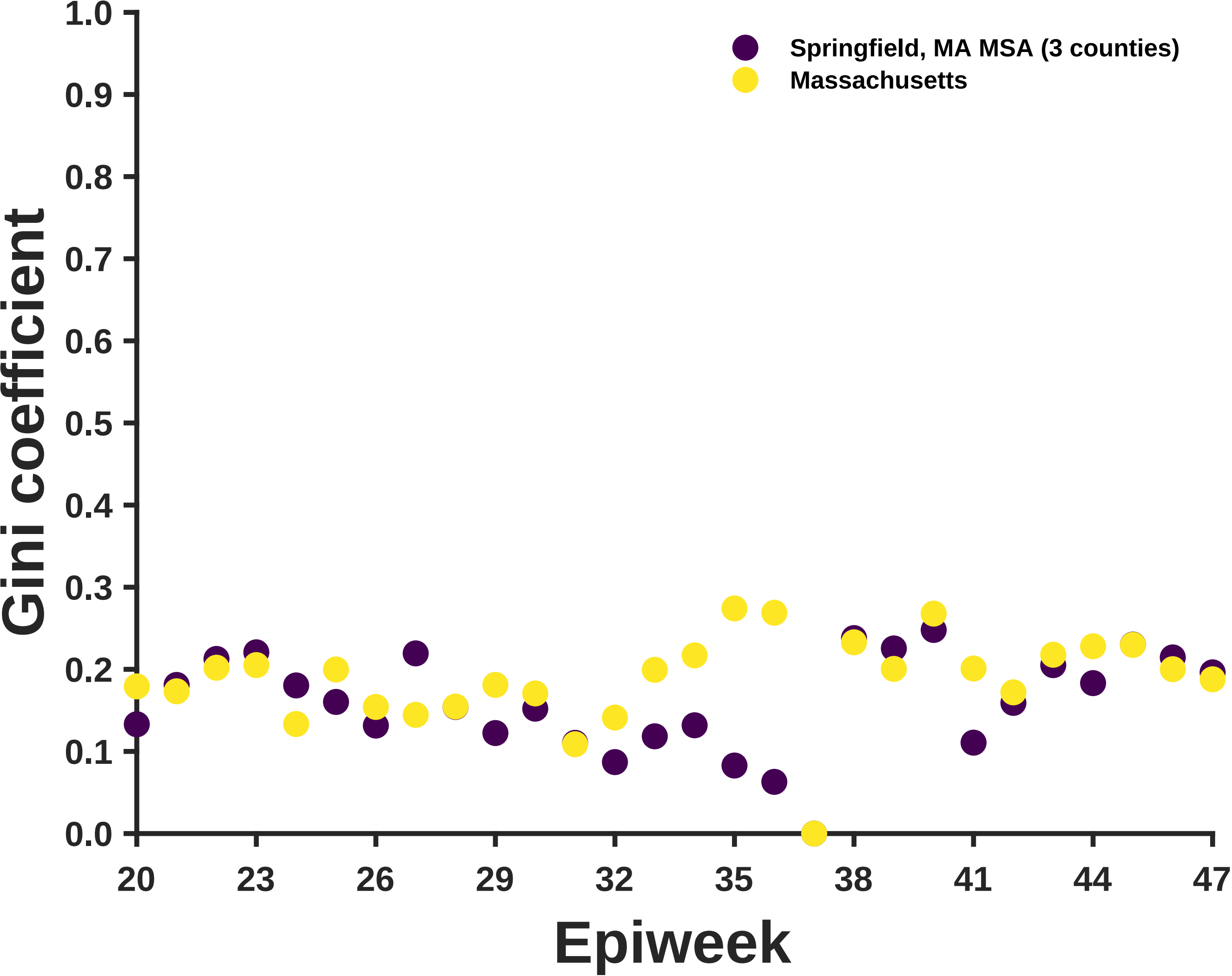

Springfield, MA MSA (3 counties)
Massachusetts
Gini coefficient
Epiweek
0.0
0.1
0.2
0.3
0.4
0.5
0.6
0.7
0.8
0.9
1.0
20
23
26
29
32
35
38
41
44
47

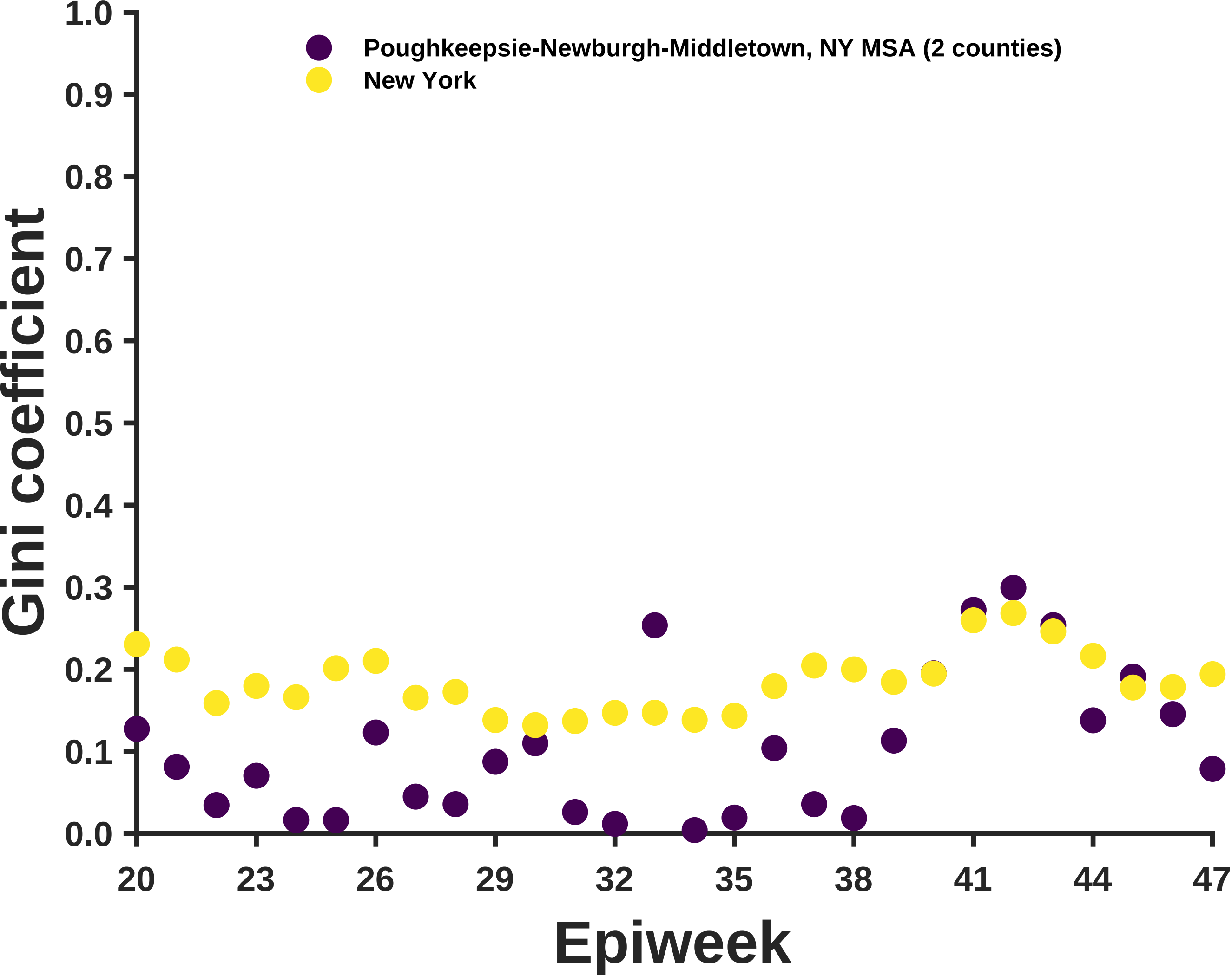
Poughkeepsie-Newburgh-Middletown, NY MSA (2 counties)
New York
Gini coefficient
1.0
0.9
0.8
0.7
0.6
0.5
0.4
0.3
0.2
0.1
0.0
20
23
26
29
32
35
38
41
44
47
Epiweek

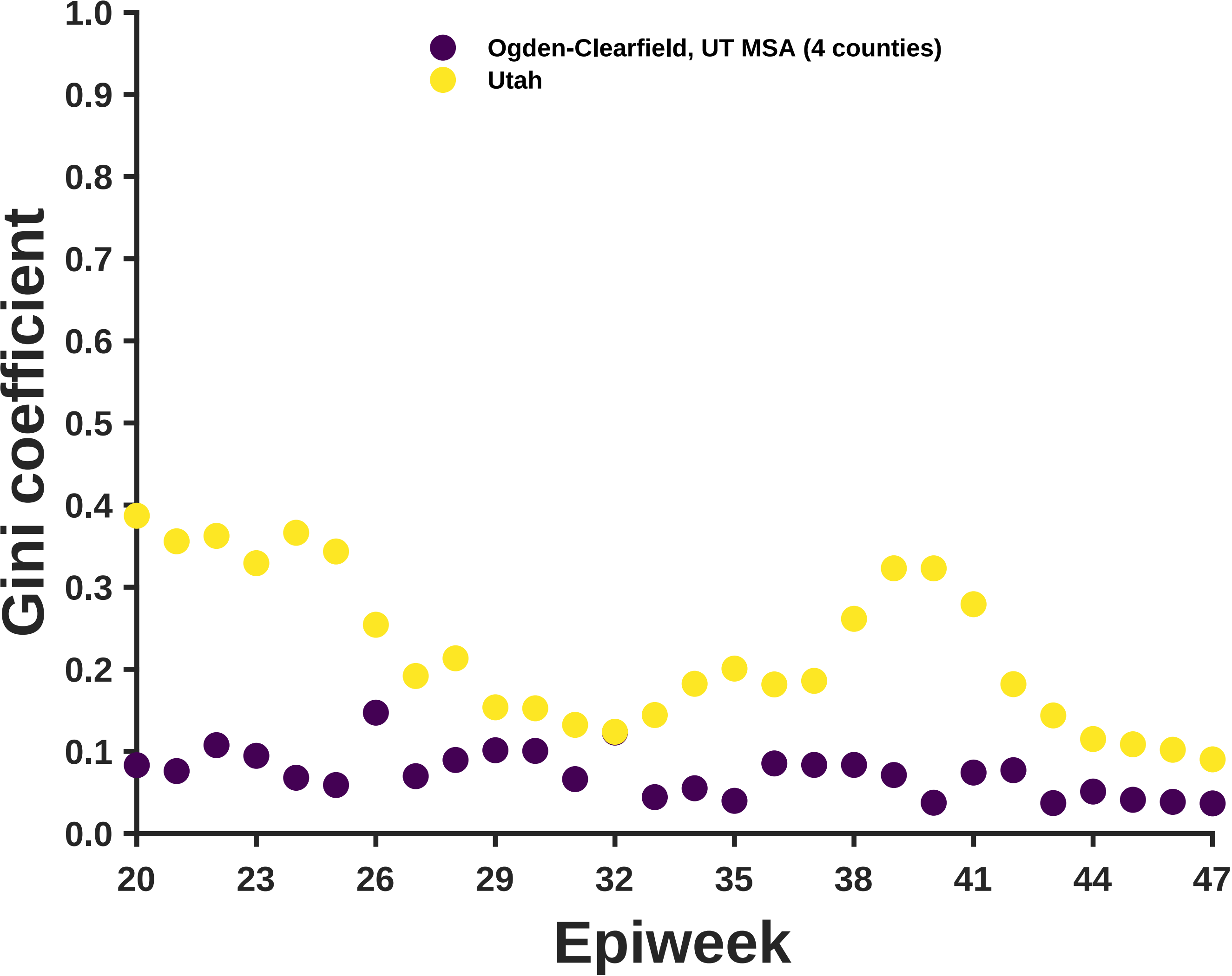

Ogden-Clearfield, UT MSA (4 counties)
Utah
Gini coefficient
Epiweek
1.0
0.9
0.8
0.7
0.6
0.5
0.4
0.3
0.2
0.1
0.0
20
23
26
29
32
35
38
41
44
47

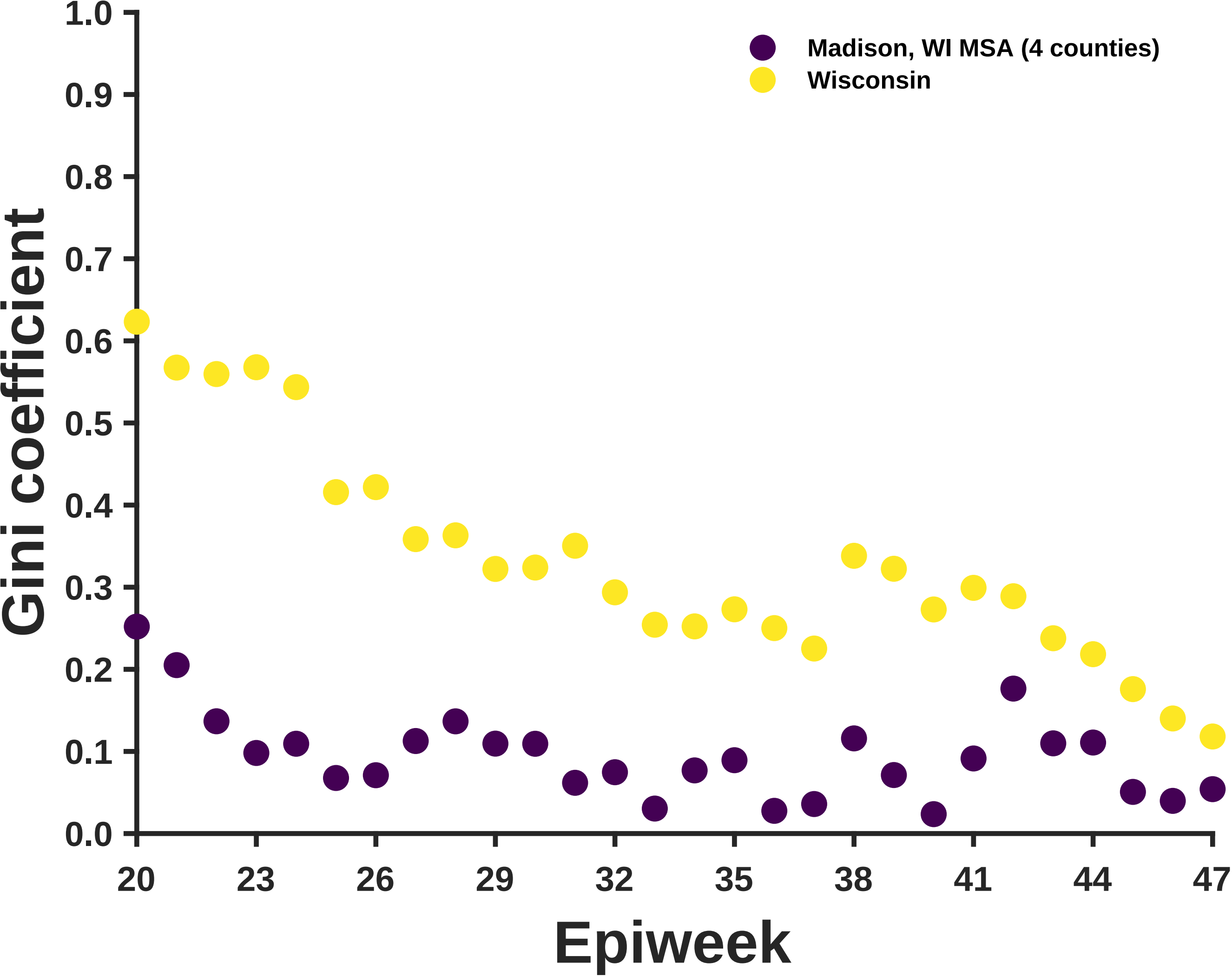
Madison, WI MSA (4 counties)
Wisconsin
Gini coefficient
Epiweek
1.0
0.9
0.8
0.7
0.6
0.5
0.4
0.3
0.2
0.1
0.0
20
23
26
29
32
35
38
41
44
47

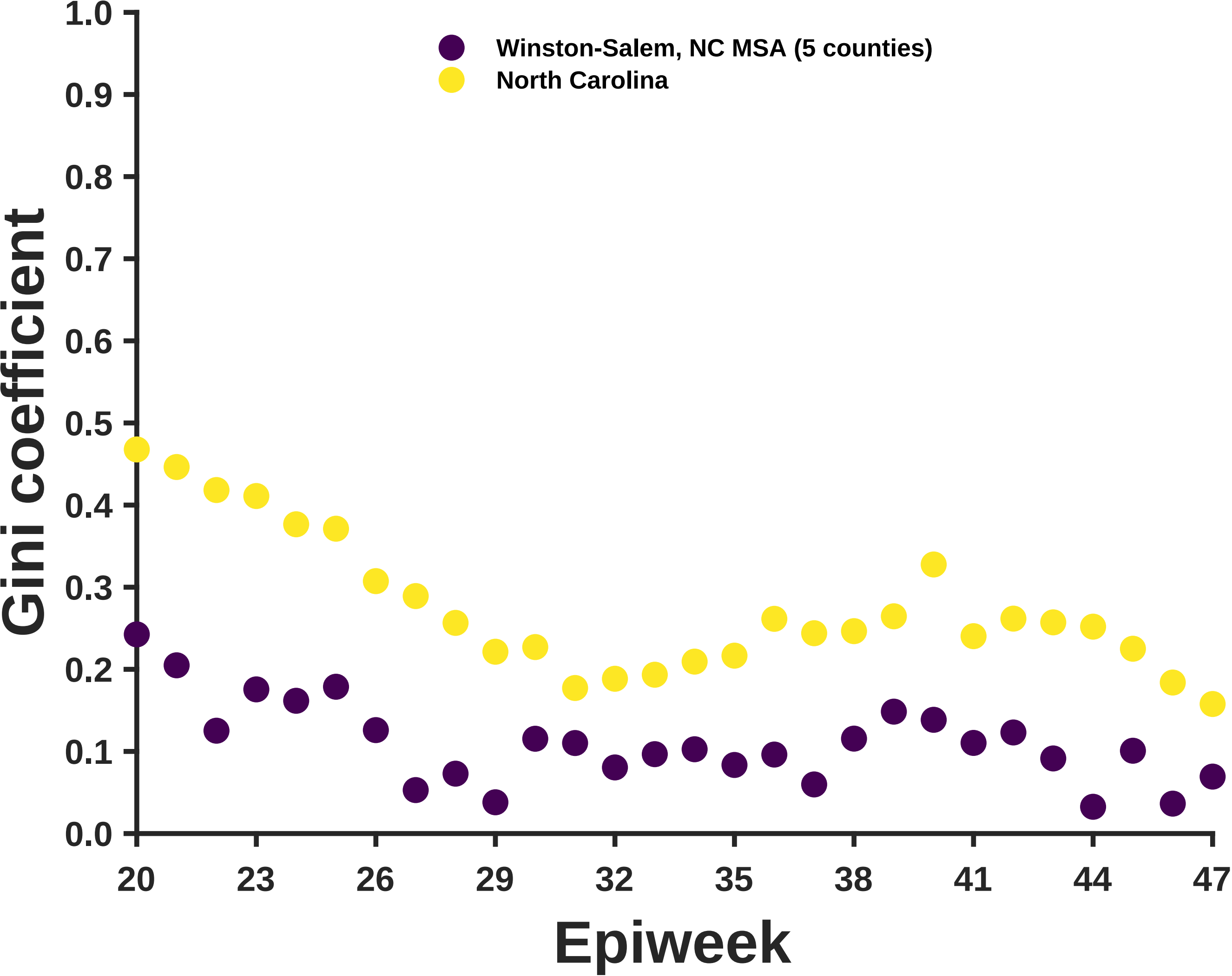
Winston-Salem, NC MSA (5 counties)
North Carolina
Gini coefficient
1.0
0.9
0.8
0.7
0.6
0.5
0.4
0.3
0.2
0.1
0.0
20
23
26
29
32
35
38
41
44
47
Epiweek

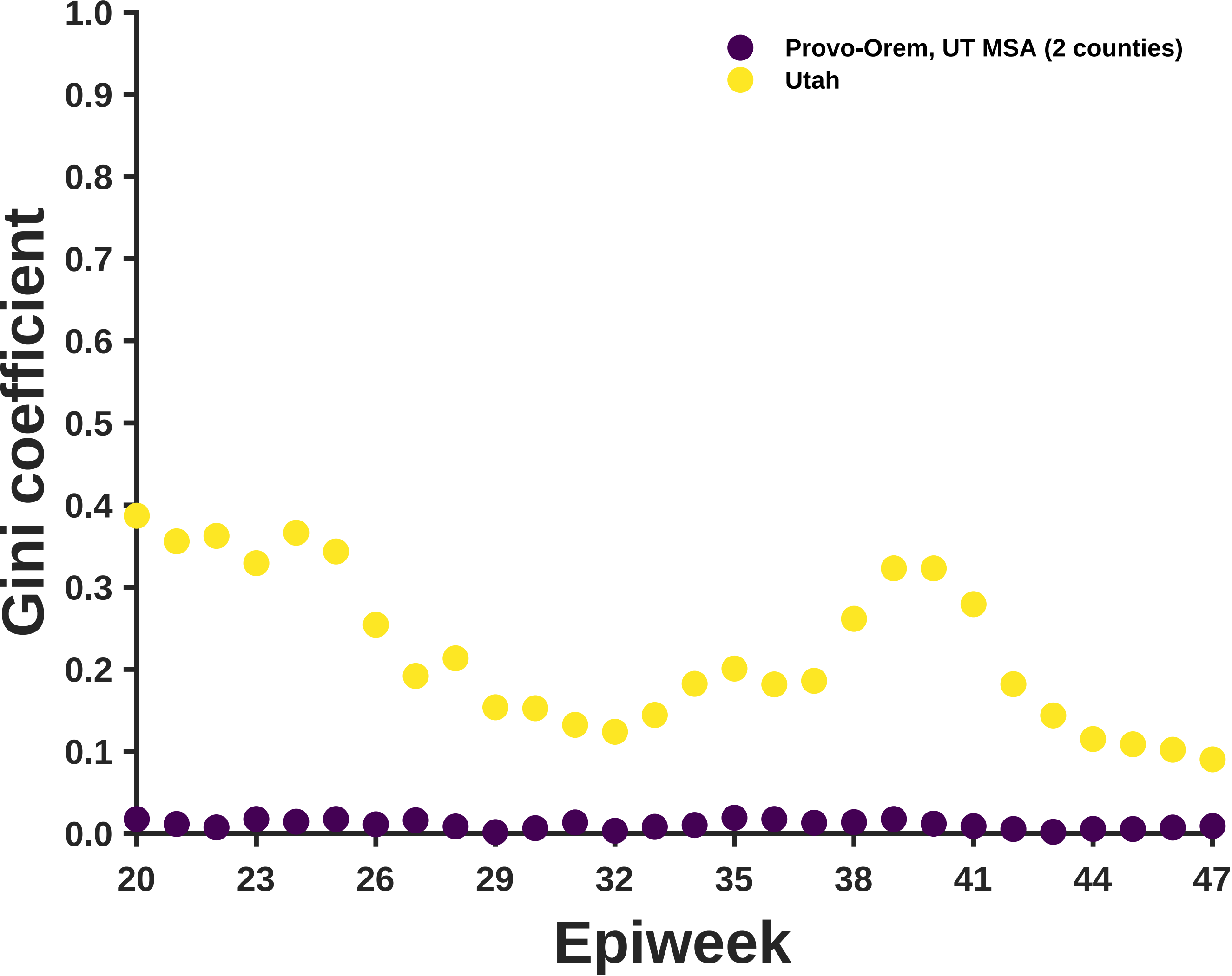

Provo-Orem, UT MSA (2 counties)
Utah
Gini coefficient
1.0
0.9
0.8
0.7
0.6
0.5
0.4
0.3
0.2
0.1
0.0
20
23
26
29
32
35
38
41
44
47
Epiweek

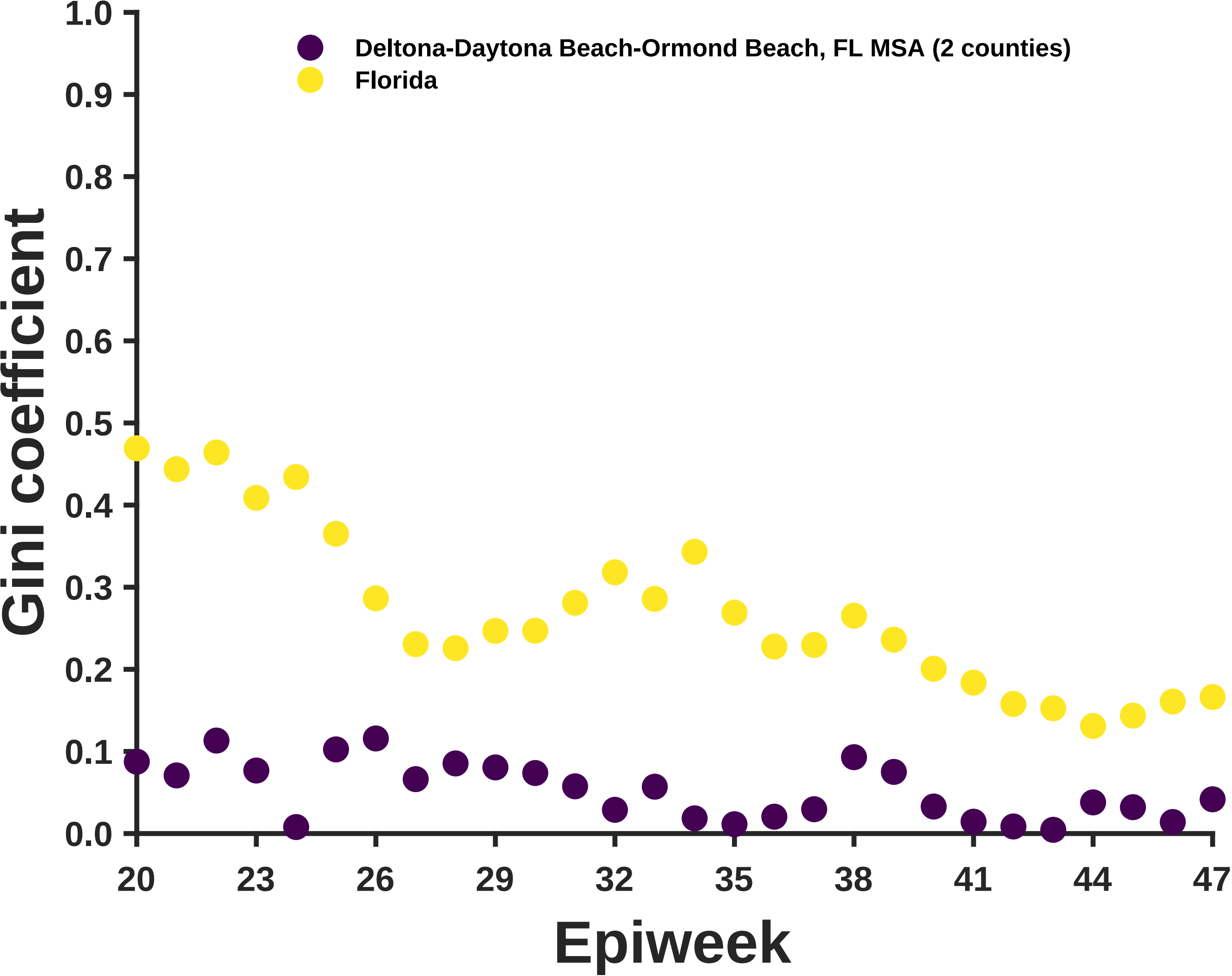
Deltona-Daytona Beach-Ormond Beach, FL MSA (2 counties)
Florida
Gini coefficient
Epiweek
1.0
0.9
0.8
0.7
0.6
0.5
0.4
0.3
0.2
0.1
0.0
20
23
26
29
32
35
38
41
44
47

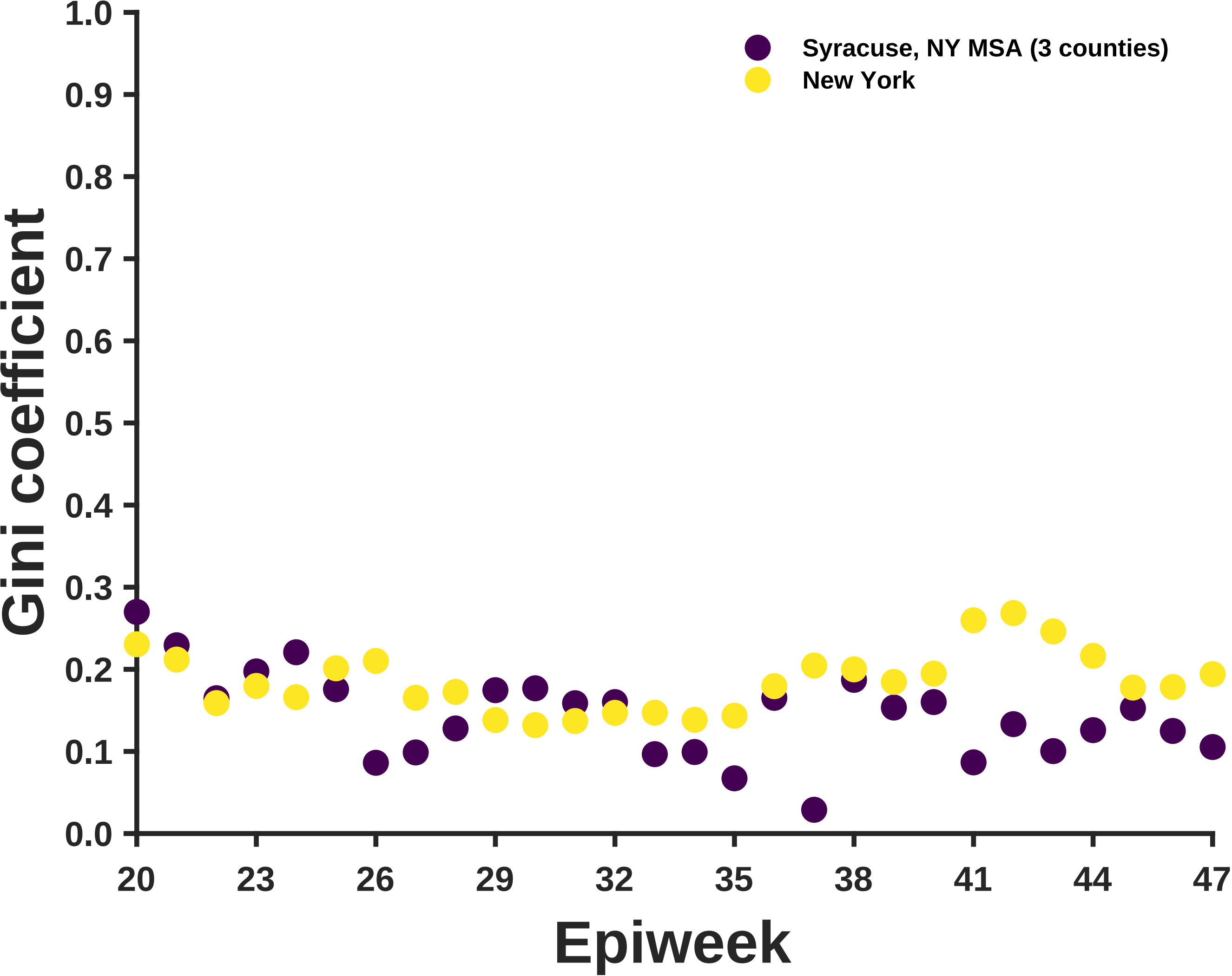

Syracuse, NY MSA (3 counties)
New York
Gini coefficient
1.0
0.9
0.8
0.7
0.6
0.5
0.4
0.3
0.2
0.1
0.0
20
23
26
29
32
35
38
41
44
47
Epiweek

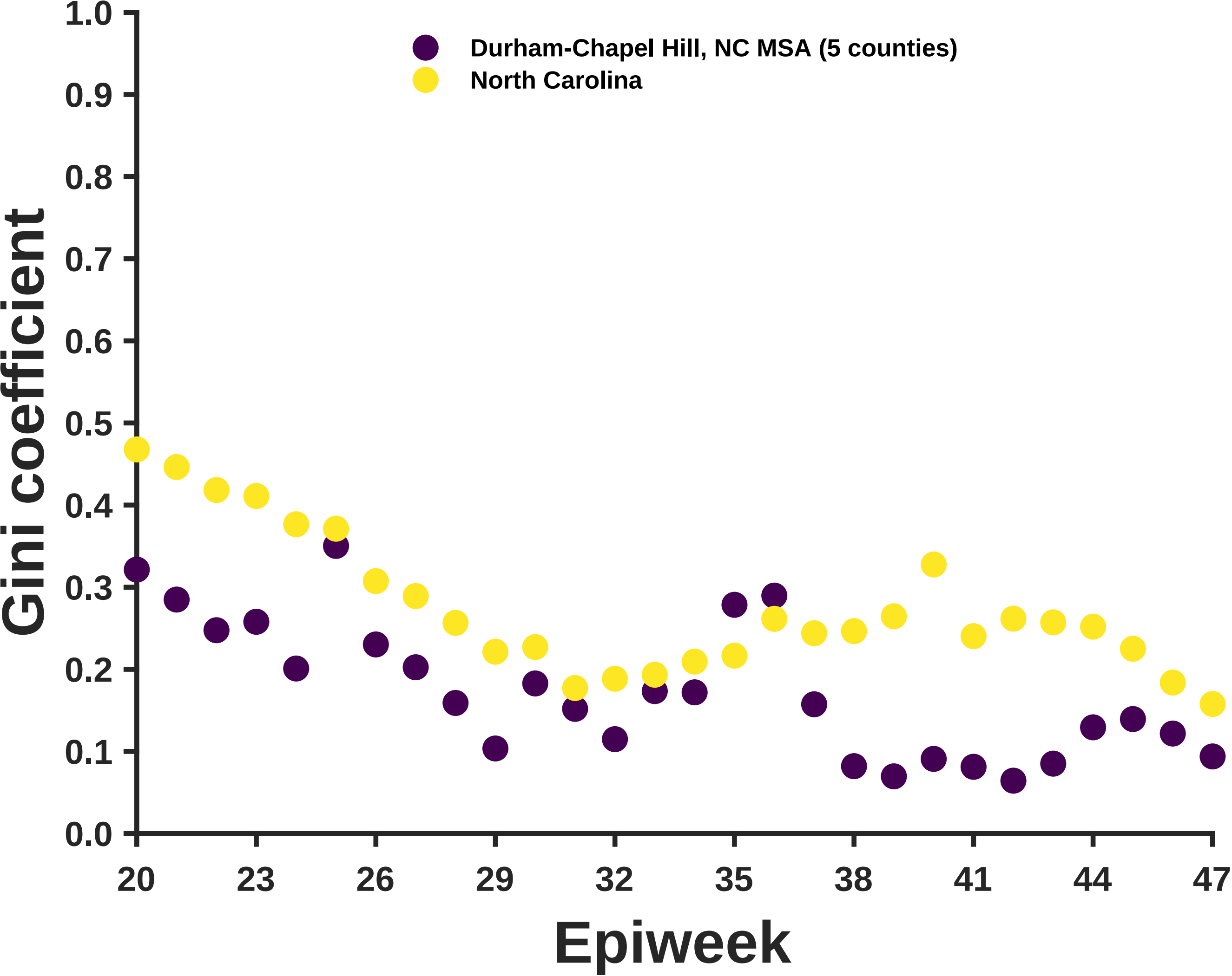
Durham-Chapel Hill, NC MSA (5 counties)
North Carolina
Gini coefficient
1.0
0.9
0.8
0.7
0.6
0.5
0.4
0.3
0.2
0.1
0.0
20
23
26
29
32
35
38
41
44
47
Epiweek

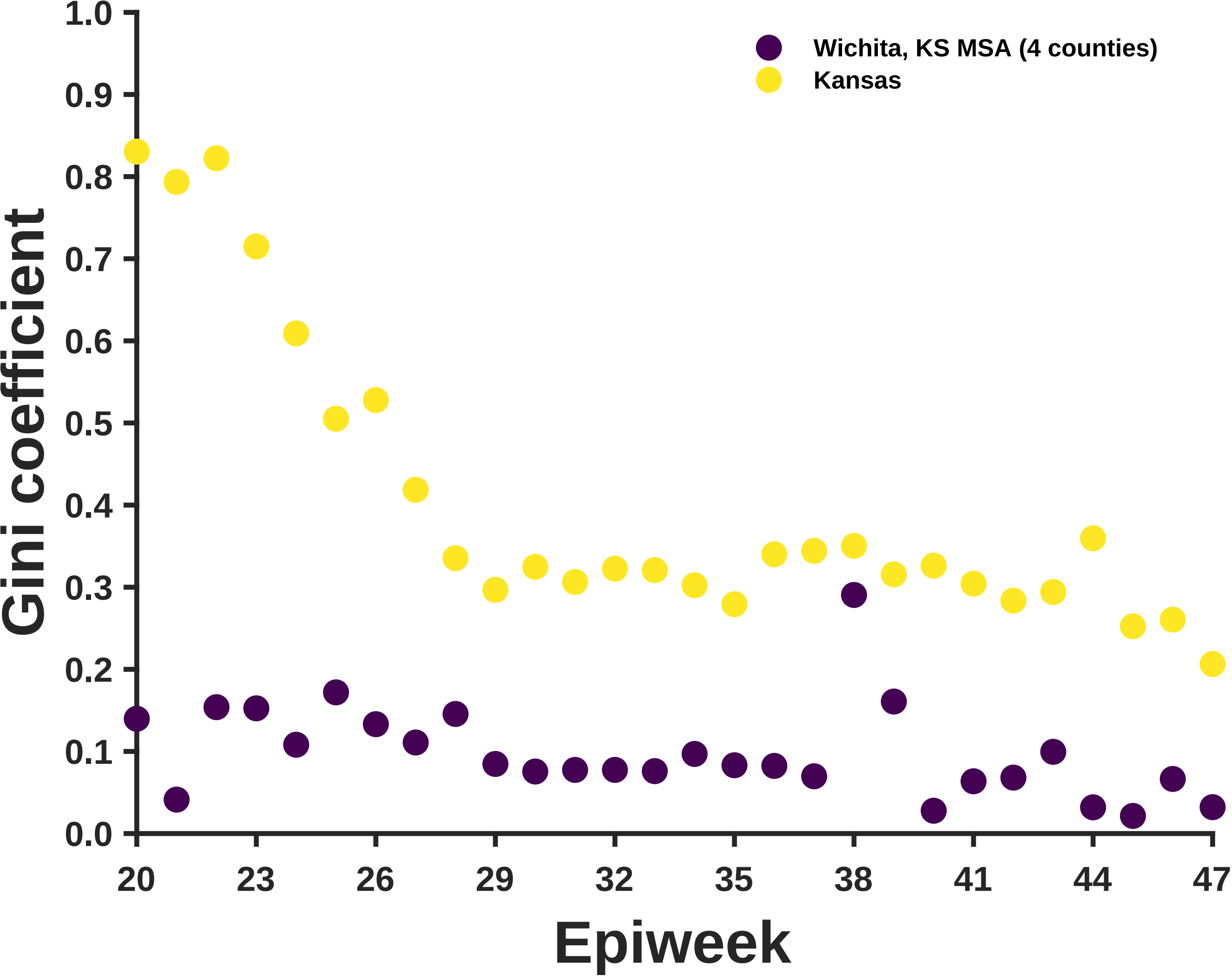
Wichita, KS MSA (4 counties)
Kansas
Gini coefficient
1.0
0.9
0.8
0.7
0.6
0.5
0.4
0.3
0.2
0.1
0.0
20
23
26
29
32
35
38
41
44
47
Epiweek

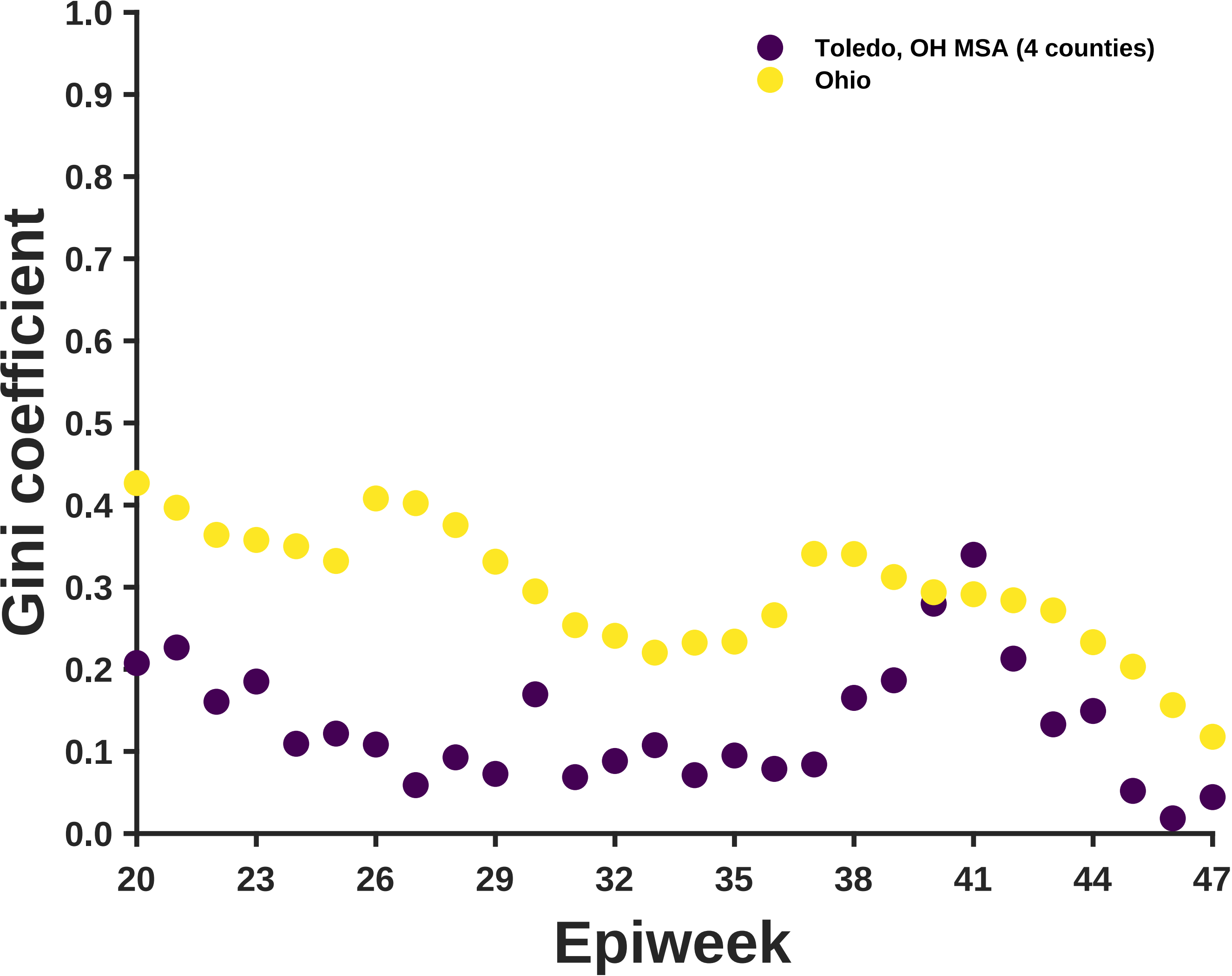

Toledo, OH MSA (4 counties)
Ohio
Gini coefficient
1.0
0.9
0.8
0.7
0.6
0.5
0.4
0.3
0.2
0.1
0.0
20
23
26
29
32
35
38
41
44
47
Epiweek

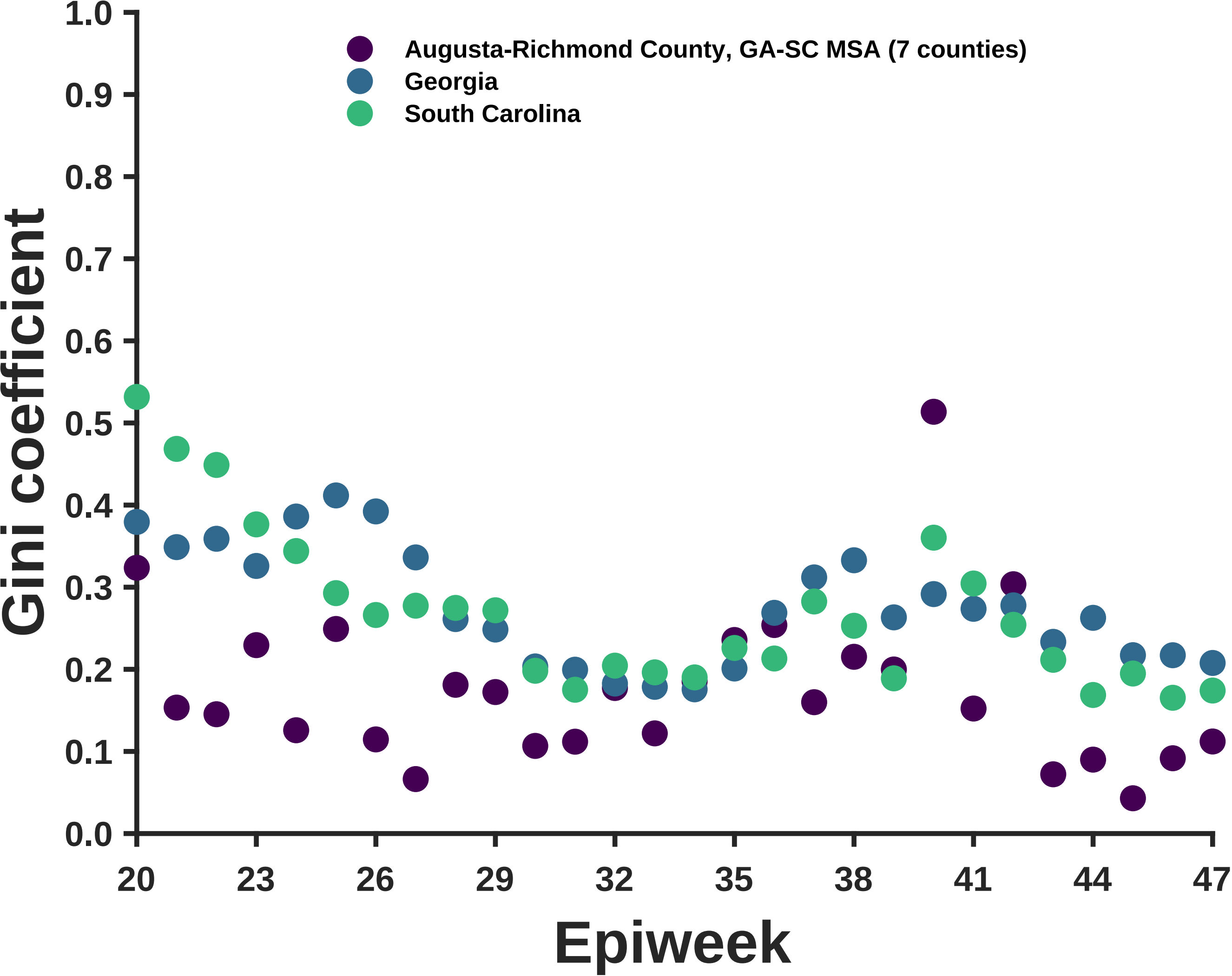
Augusta-Richmond County, GA-SC MSA (7 counties)
Georgia
South Carolina
Gini coefficient
Epiweek
1.0
0.9
0.8
0.7
0.6
0.5
0.4
0.3
0.2
0.1
0.0
20
23
26
29
32
35
38
41
44
47

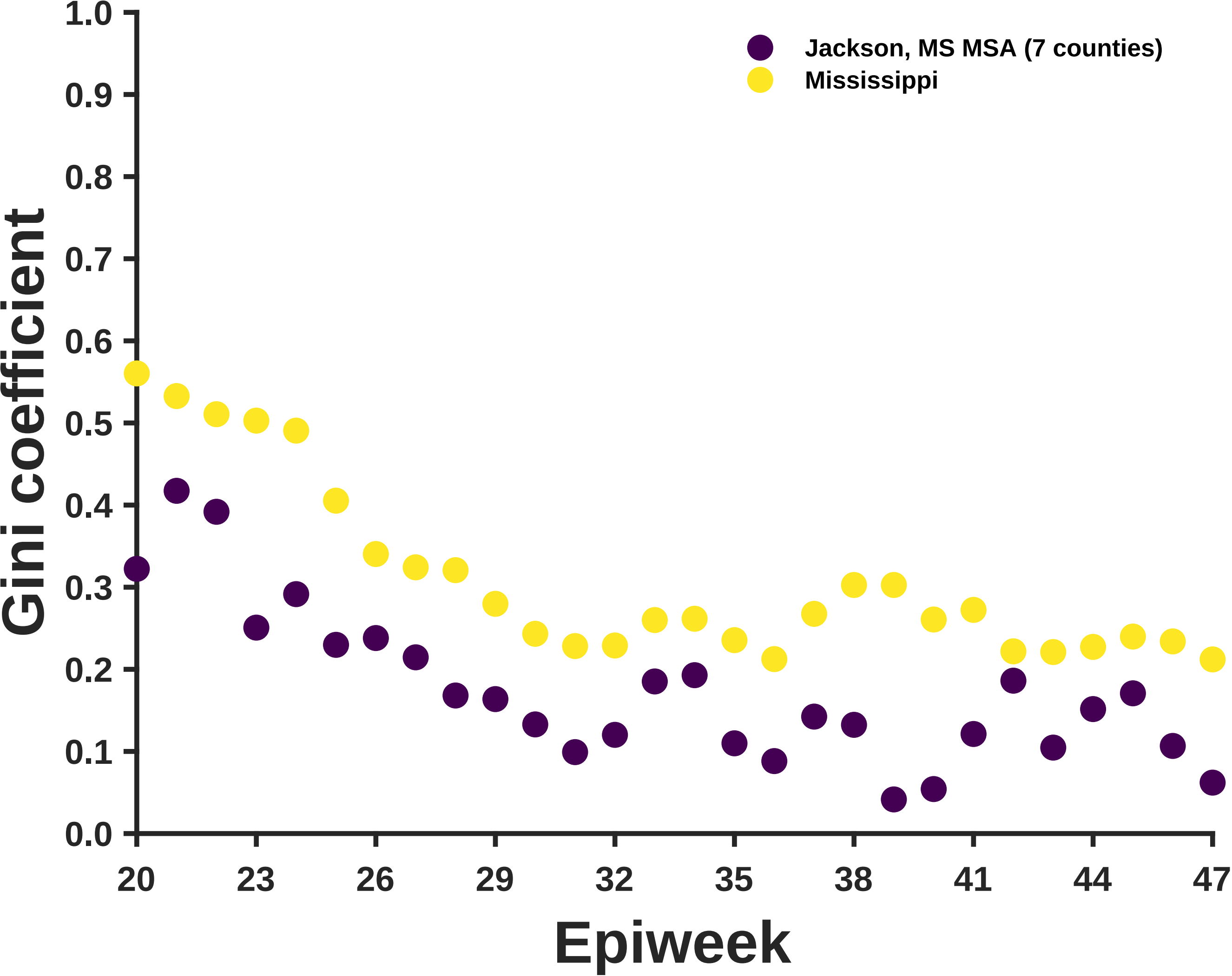
Jackson, MS MSA (7 counties)
Mississippi
Gini coefficient
Epiweek
1.0
0.9
0.8
0.7
0.6
0.5
0.4
0.3
0.2
0.1
0.0
20
23
26
29
32
35
38
41
44
47

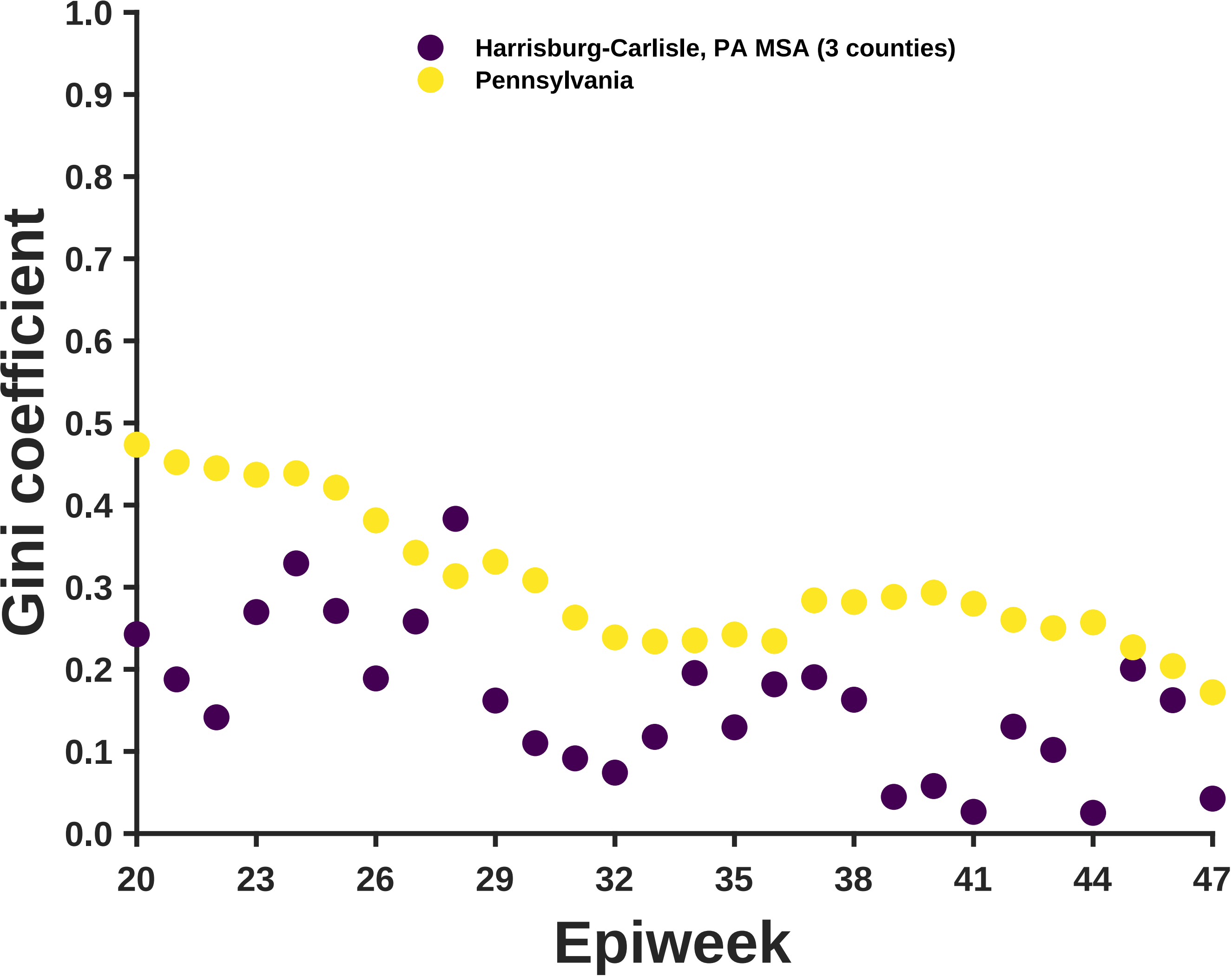
Harrisburg-Carlisle, PA MSA (3 counties)
Pennsylvania
Gini coefficient
Epiweek
1.0
0.9
0.8
0.7
0.6
0.5
0.4
0.3
0.2
0.1
0.0
20
23
26
29
32
35
38
41
44
47

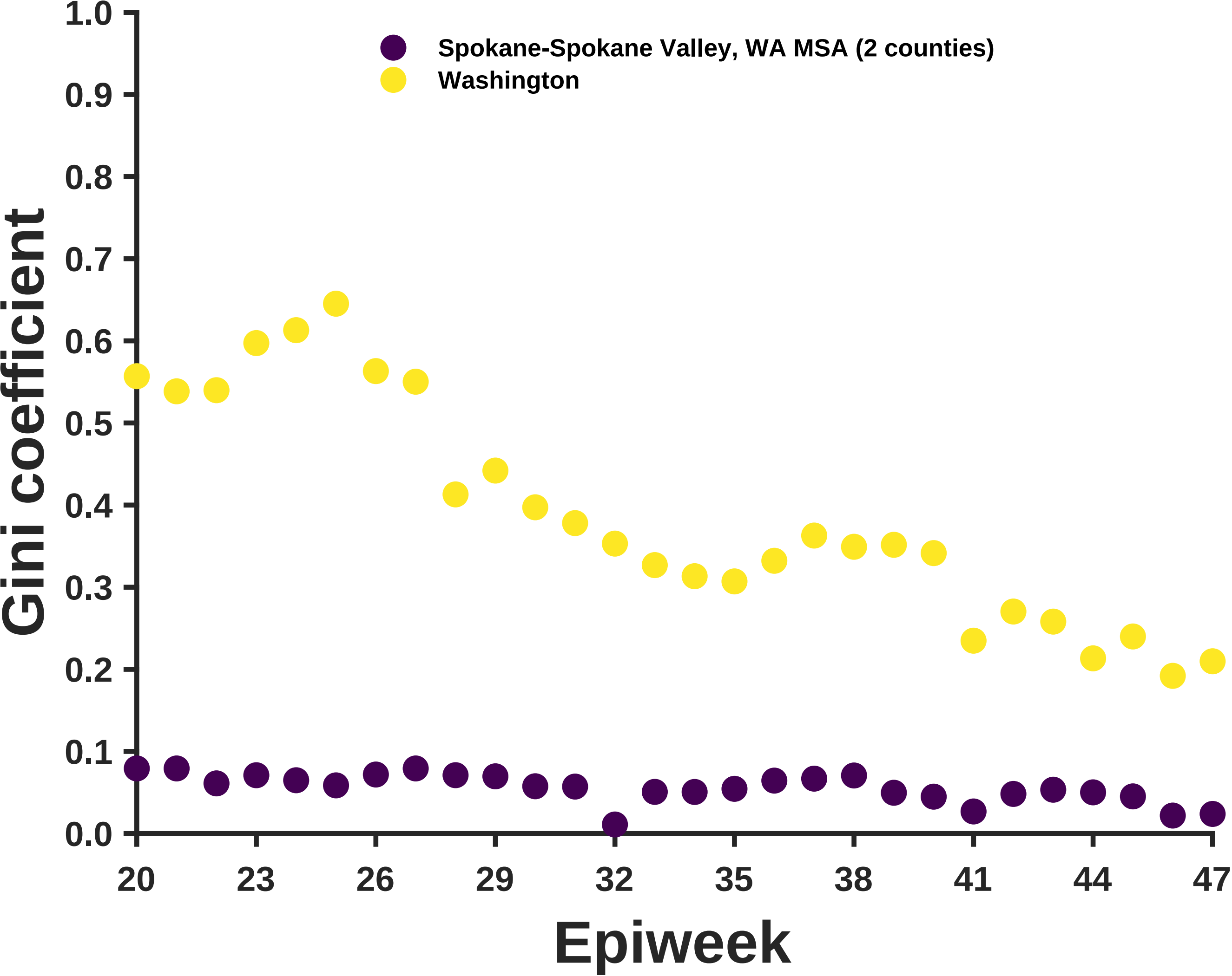

Spokane-Spokane Valley, WA MSA (2 counties)
Washington
Gini coefficient
1.0
0.9
0.8
0.7
0.6
0.5
0.4
0.3
0.2
0.1
0.0
20
23
26
29
32
35
38
41
44
47
Epiweek

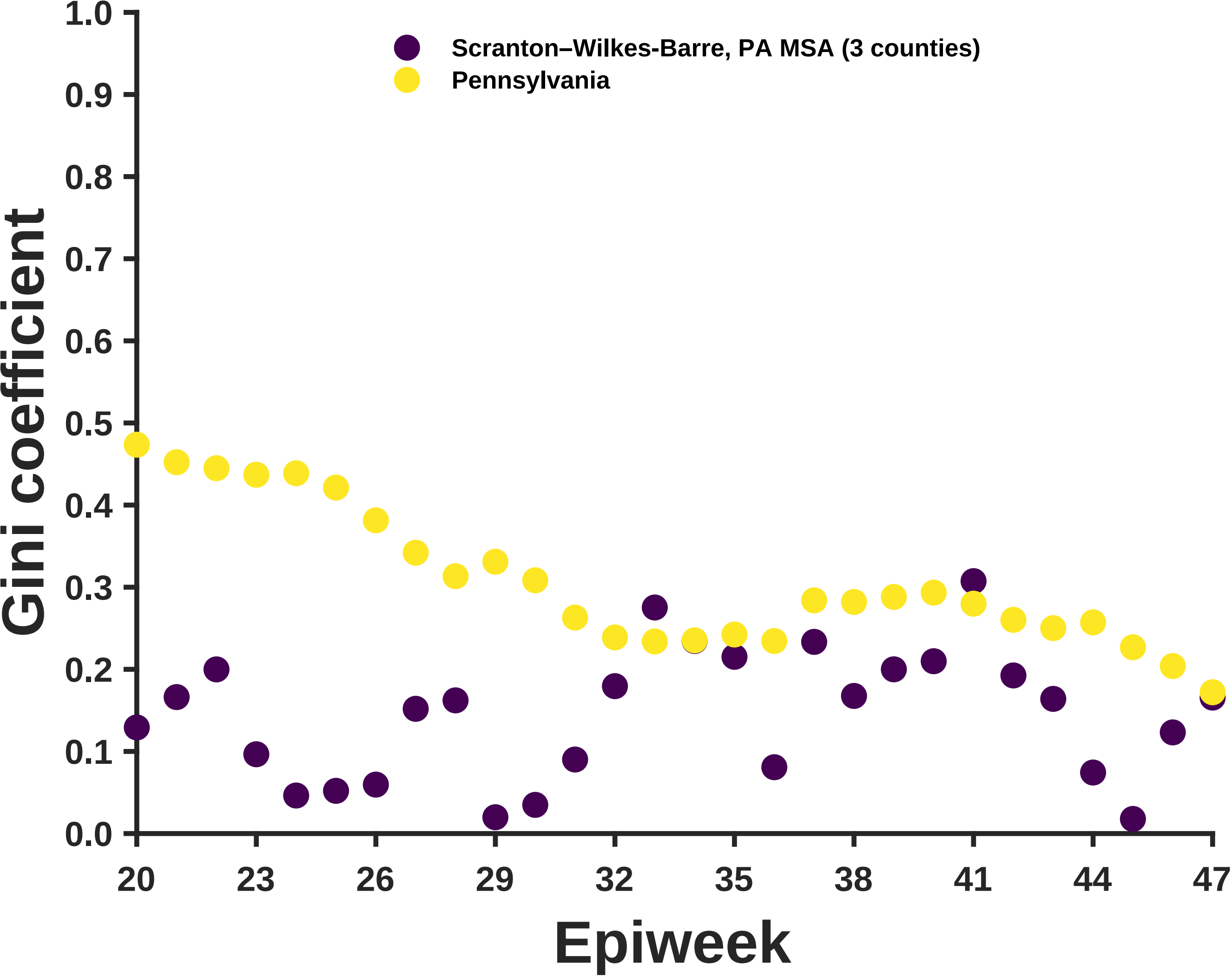

Scranton–Wilkes-Barre, PA MSA (3 counties)
Pennsylvania
Gini coefficient
1.0
0.9
0.8
0.7
0.6
0.5
0.4
0.3
0.2
0.1
0.0
20
23
26
29
32
35
38
41
44
47
Epiweek

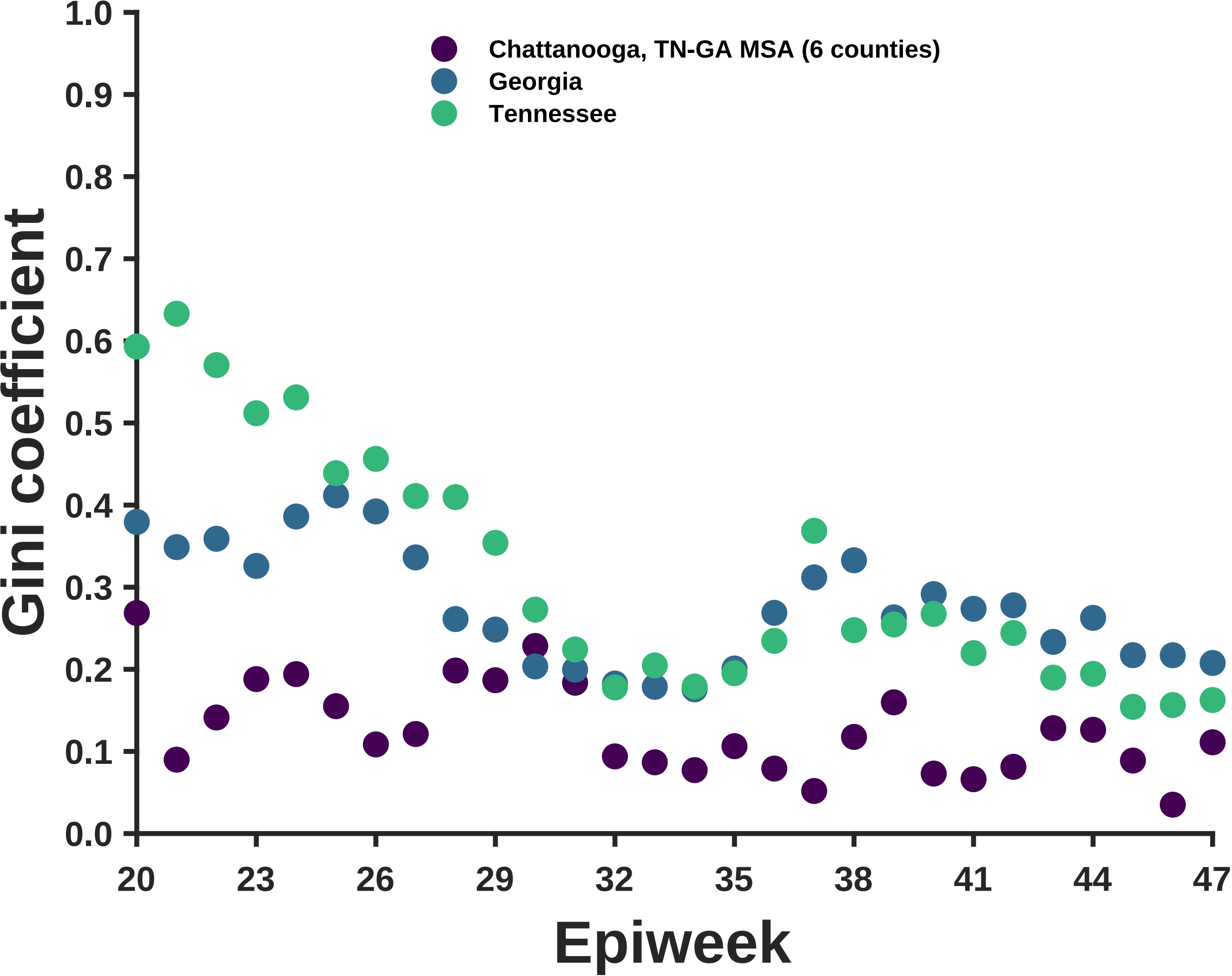
Chattanooga, TN-GA MSA (6 counties)
Georgia
Tennessee
Gini coefficient
1.0
0.9
0.8
0.7
0.6
0.5
0.4
0.3
0.2
0.1
0.0
20
23
26
29
32
35
38
41
44
47
Epiweek

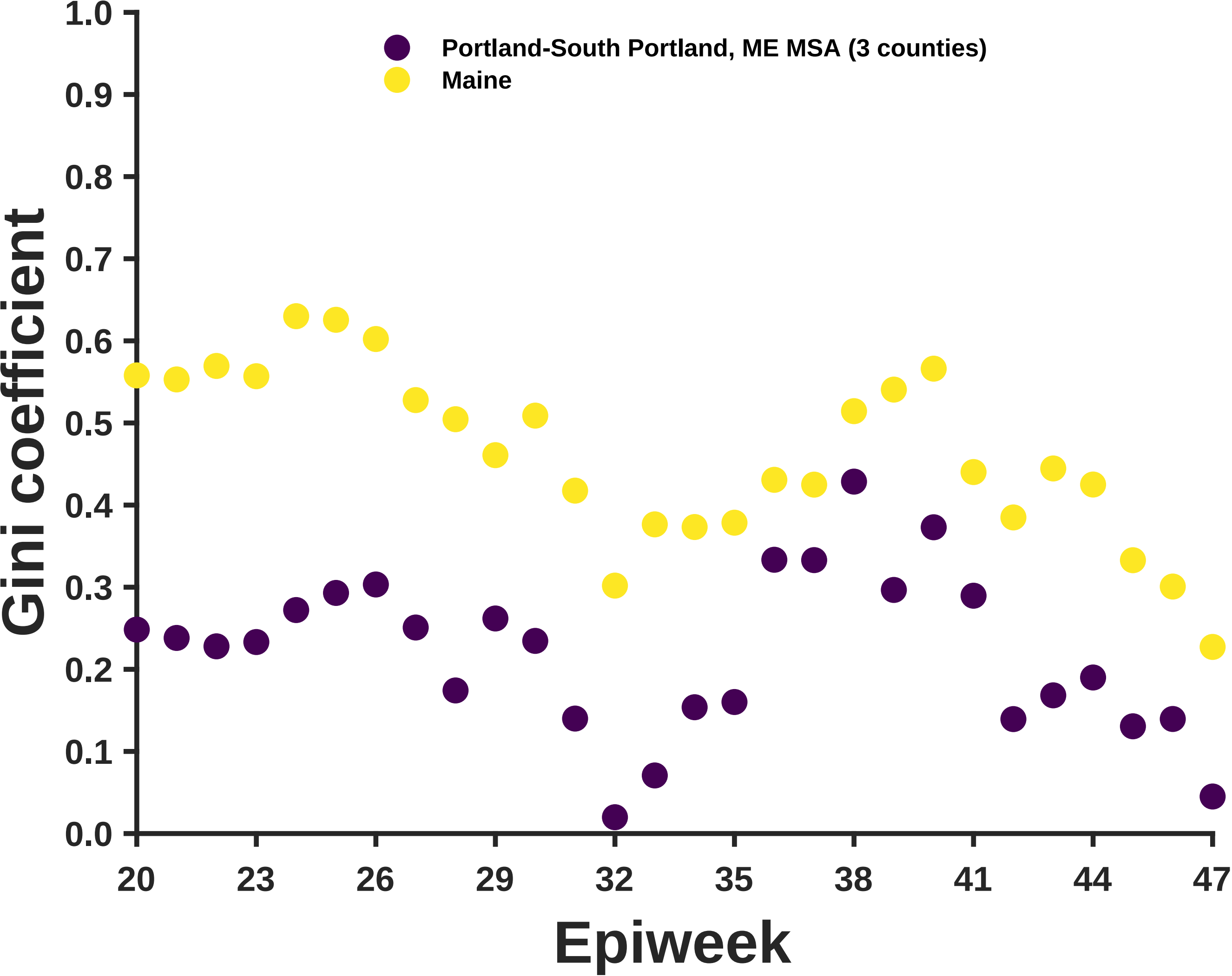
Portland-South Portland, ME MSA (3 counties)
Maine
Gini coefficient
Epiweek
1.0
0.9
0.8
0.7
0.6
0.5
0.4
0.3
0.2
0.1
0.0
20
23
26
29
32
35
38
41
44
47

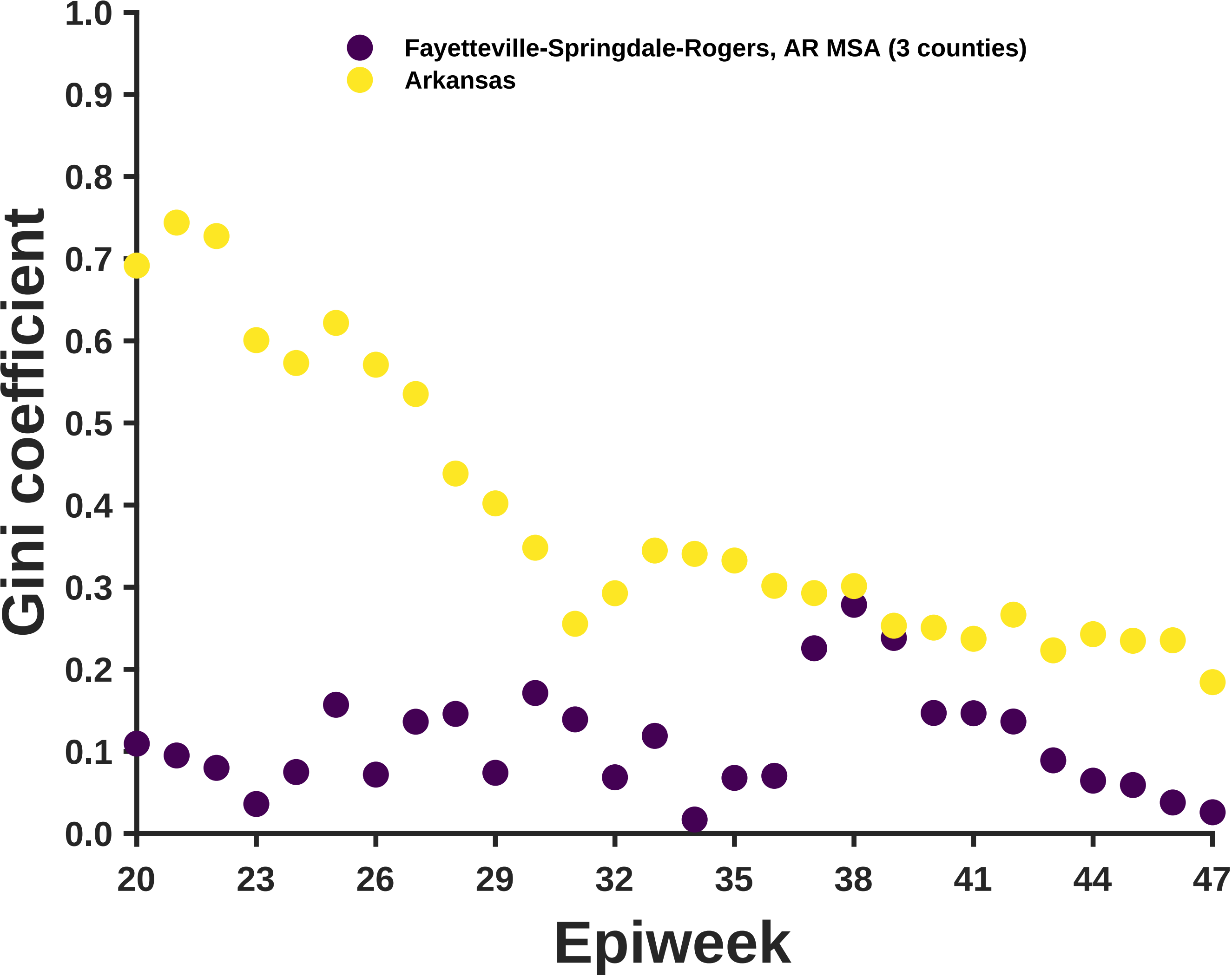
Fayetteville-Springdale-Rogers, AR MSA (3 counties)
Arkansas
Gini coefficient
Epiweek
1.0
0.9
0.8
0.7
0.6
0.5
0.4
0.3
0.2
0.1
0.0
20
23
26
29
32
35
38
41
44
47

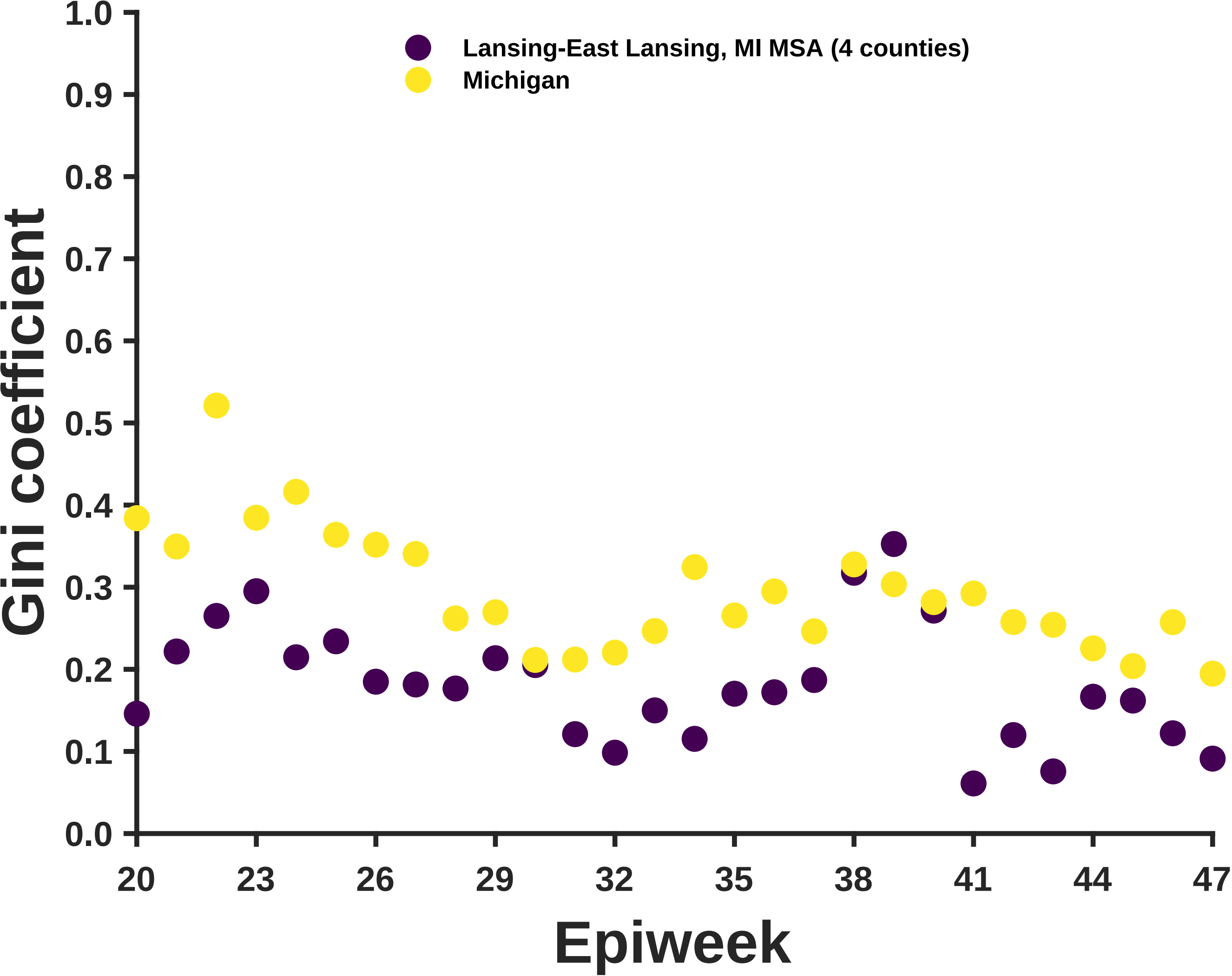

Lansing-East Lansing, MI MSA (4 counties)
Michigan
Gini coefficient
1.0
0.9
0.8
0.7
0.6
0.5
0.4
0.3
0.2
0.1
0.0
20
23
26
29
32
35
38
41
44
47
Epiweek

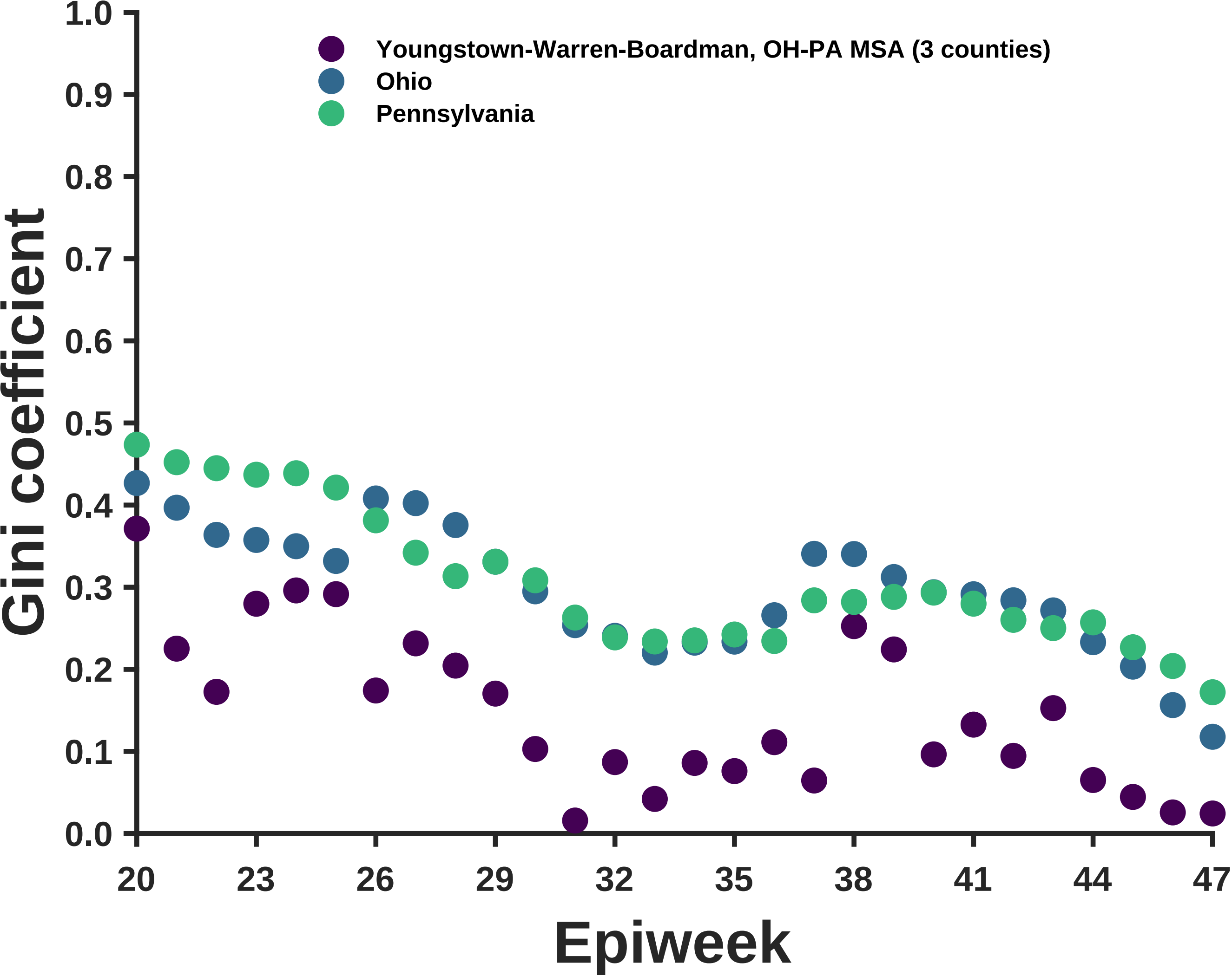

Youngstown-Warren-Boardman, OH-PA MSA (3 counties)
Ohio
Pennsylvania
Gini coefficient
1.0
0.9
0.8
0.7
0.6
0.5
0.4
0.3
0.2
0.1
0.0
20
23
26
29
32
35
38
41
44
47
Epiweek

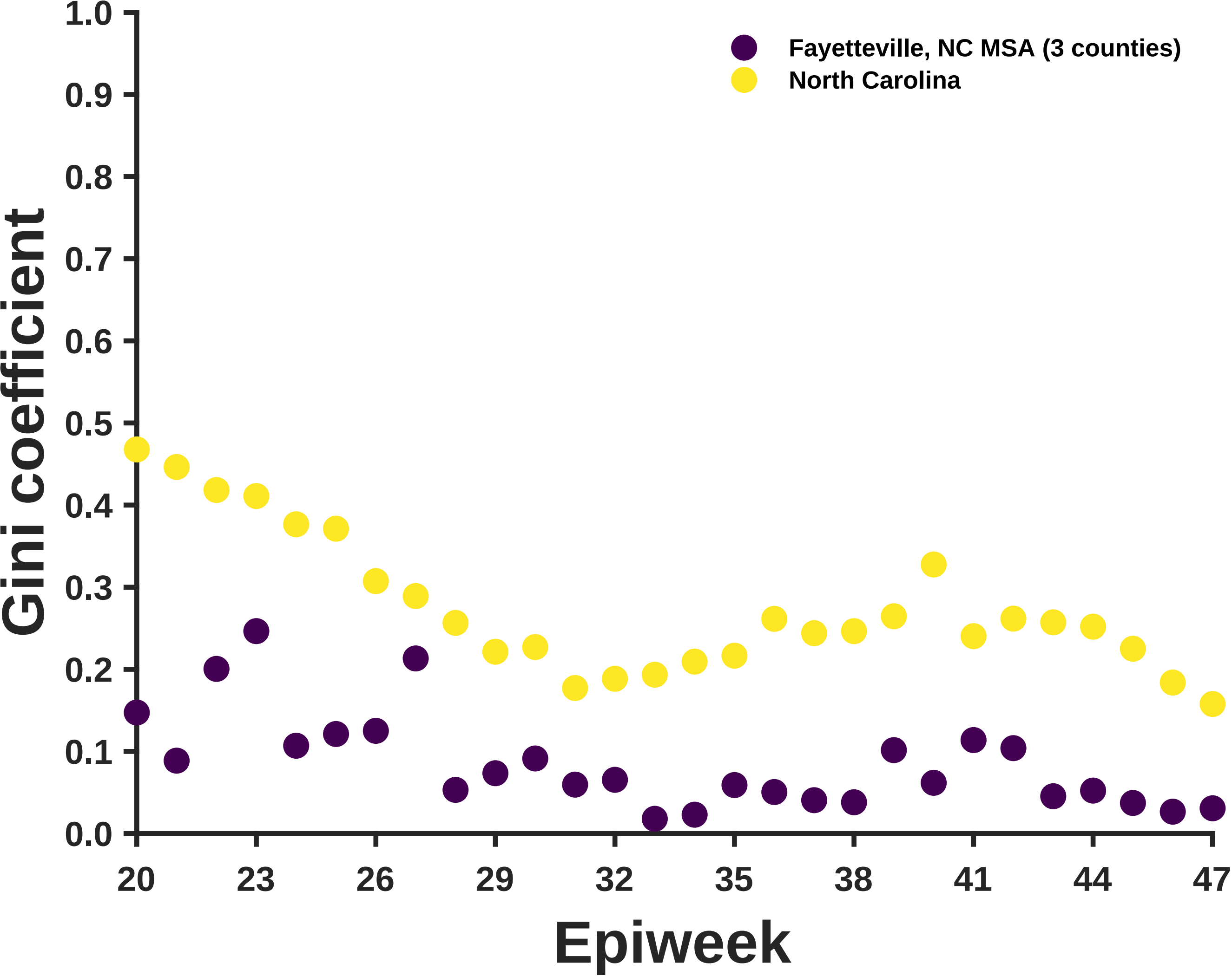

Fayetteville, NC MSA (3 counties)
North Carolina
Gini coefficient
Epiweek
1.0
0.9
0.8
0.7
0.6
0.5
0.4
0.3
0.2
0.1
0.0
20
23
26
29
32
35
38
41
44
47

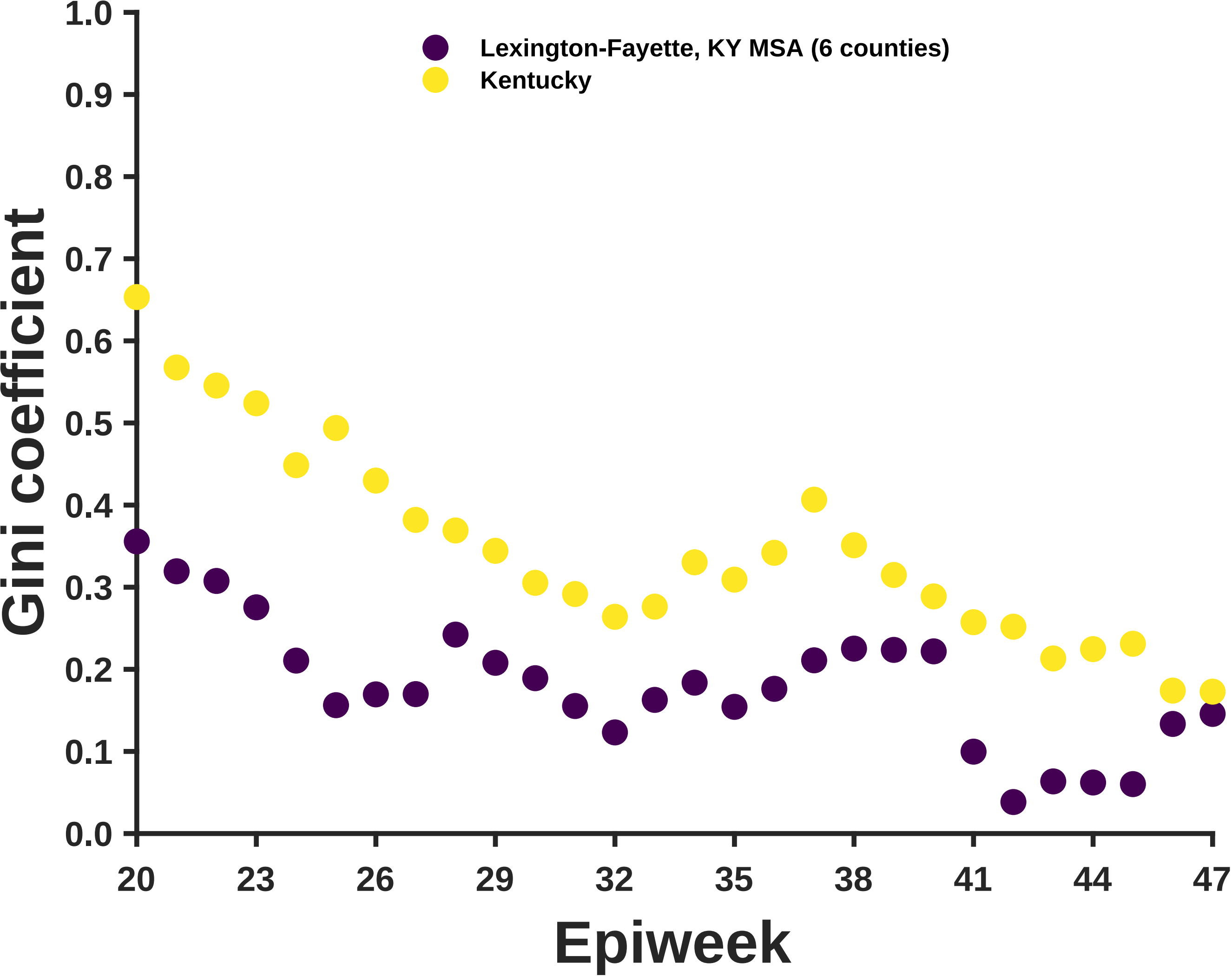
Lexington-Fayette, KY MSA (6 counties)
Kentucky
Gini coefficient
1.0
0.9
0.8
0.7
0.6
0.5
0.4
0.3
0.2
0.1
0.0
20
23
26
29
32
35
38
41
44
47
Epiweek

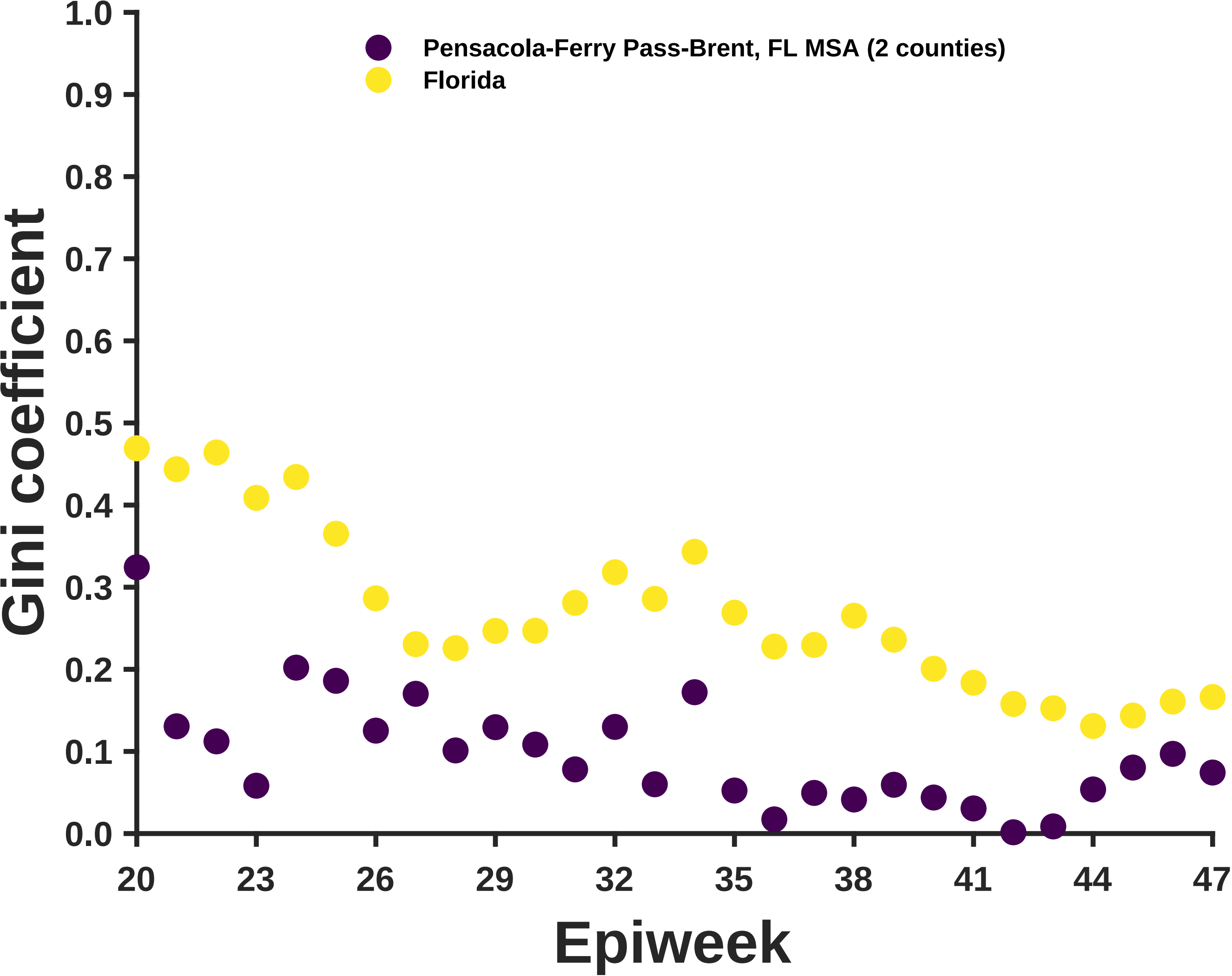

Pensacola-Ferry Pass-Brent, FL MSA (2 counties)
Florida
Gini coefficient
Epiweek
1.0
0.9
0.8
0.7
0.6
0.5
0.4
0.3
0.2
0.1
0.0
20
23
26
29
32
35
38
41
44
47

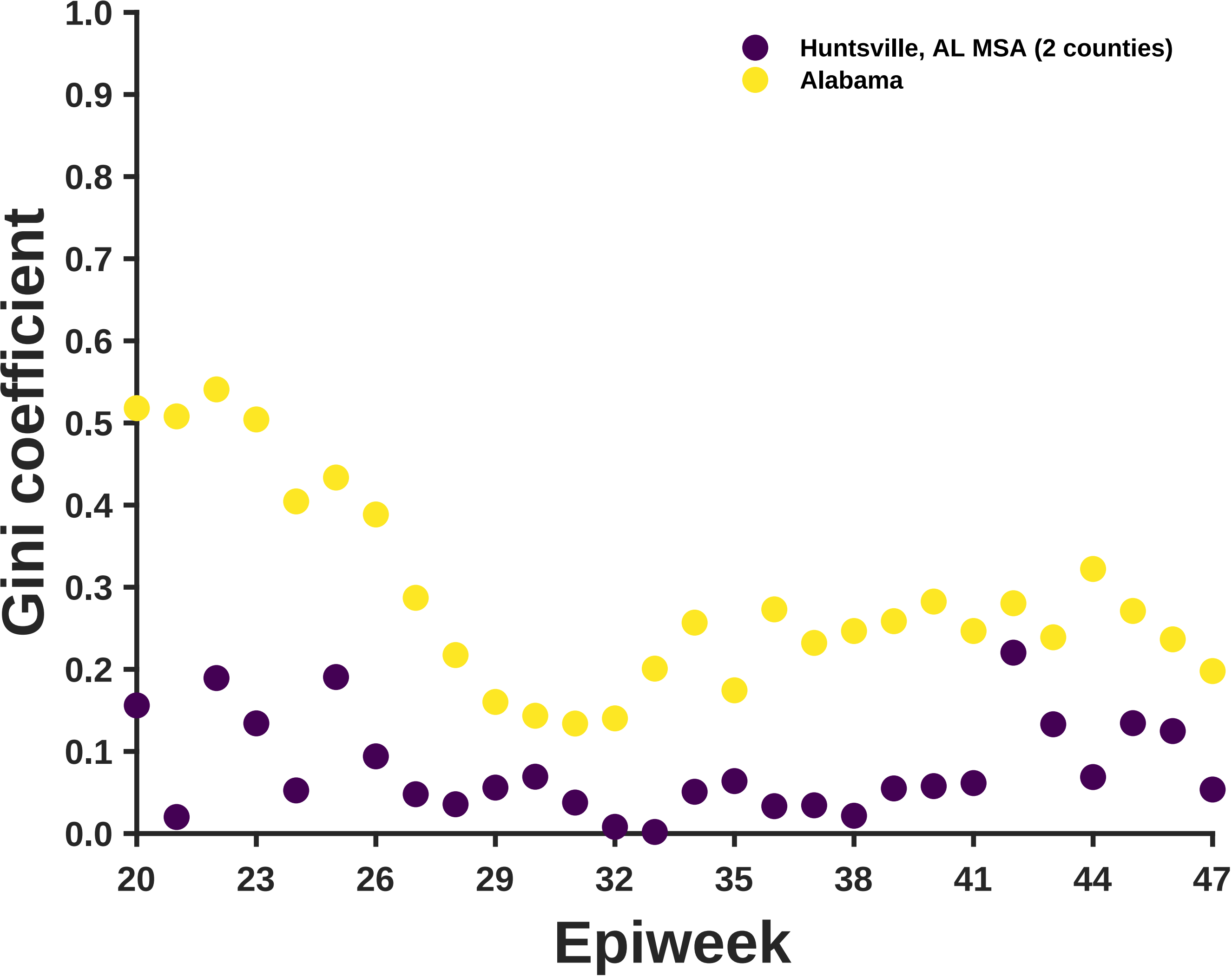

Huntsville, AL MSA (2 counties)
Alabama
Gini coefficient
1.0
0.9
0.8
0.7
0.6
0.5
0.4
0.3
0.2
0.1
0.0
20
23
26
29
32
35
38
41
44
47
Epiweek

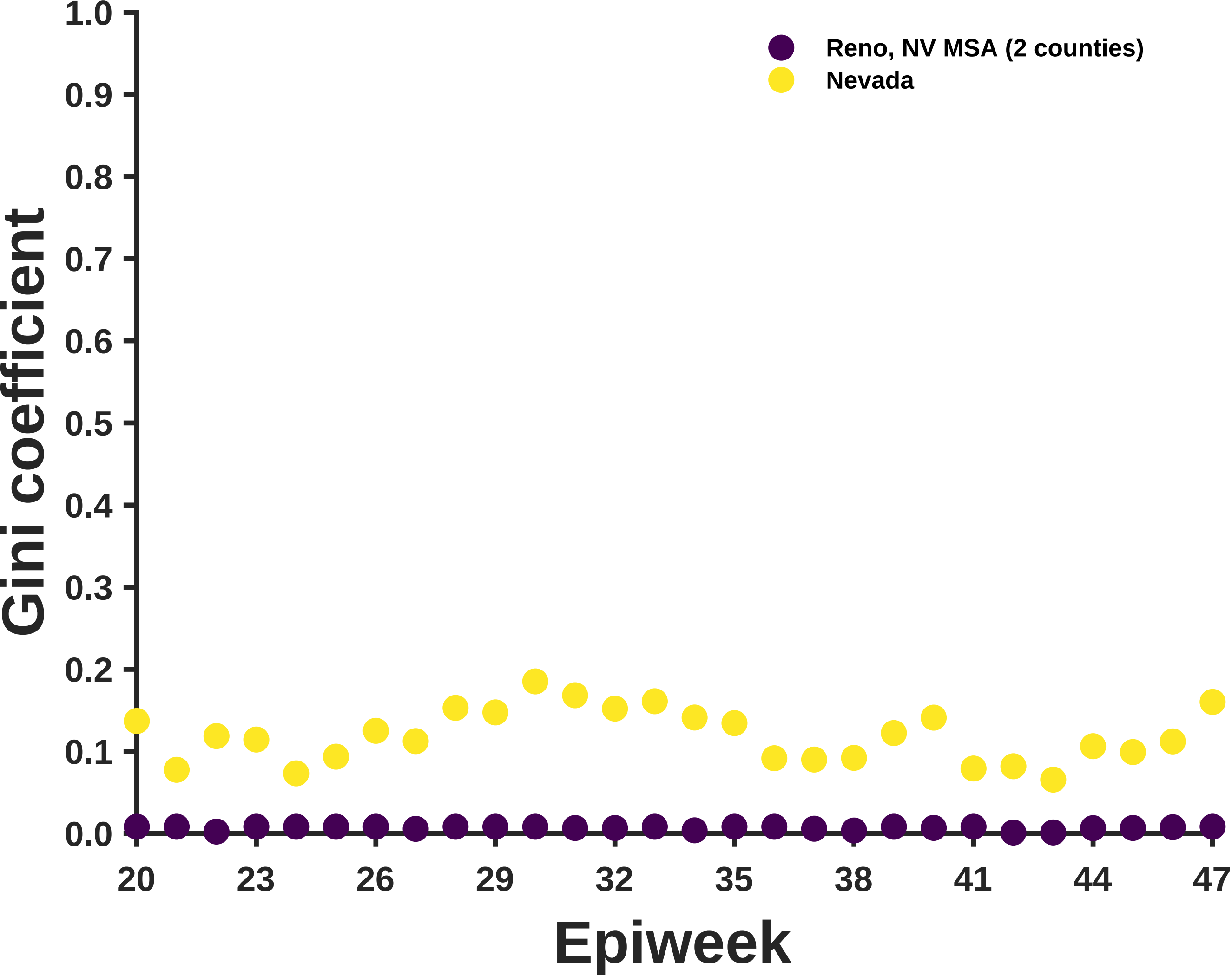

Reno, NV MSA (2 counties)
Nevada
Gini coefficient
1.0
0.9
0.8
0.7
0.6
0.5
0.4
0.3
0.2
0.1
0.0
20
23
26
29
32
35
38
41
44
47
Epiweek

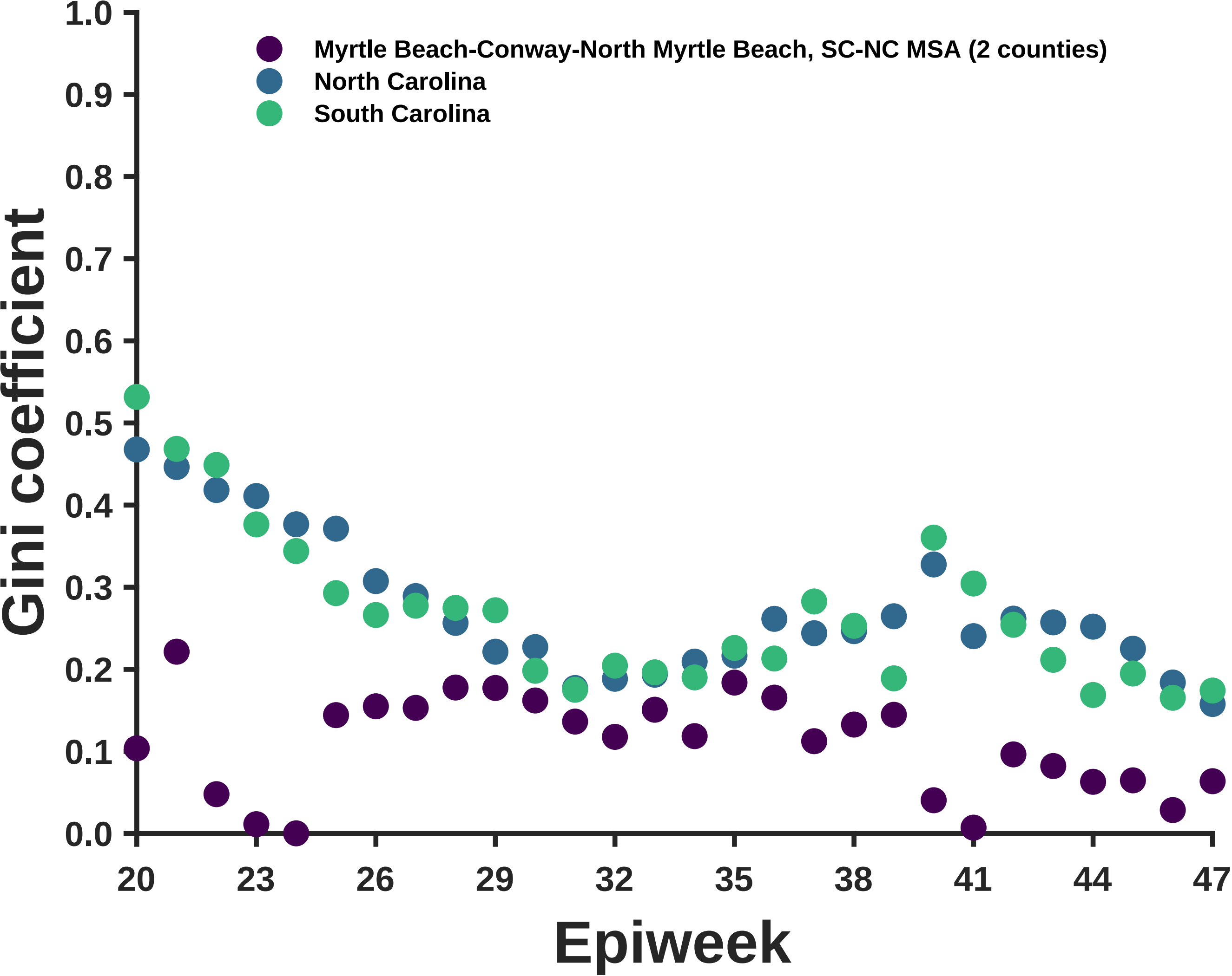

Myrtle Beach-Conway-North Myrtle Beach, SC-NC MSA (2 counties)
North Carolina
South Carolina
Gini coefficient
1.0
0.9
0.8
0.7
0.6
0.5
0.4
0.3
0.2
0.1
0.0
20
23
26
29
32
35
38
41
44
47
Epiweek

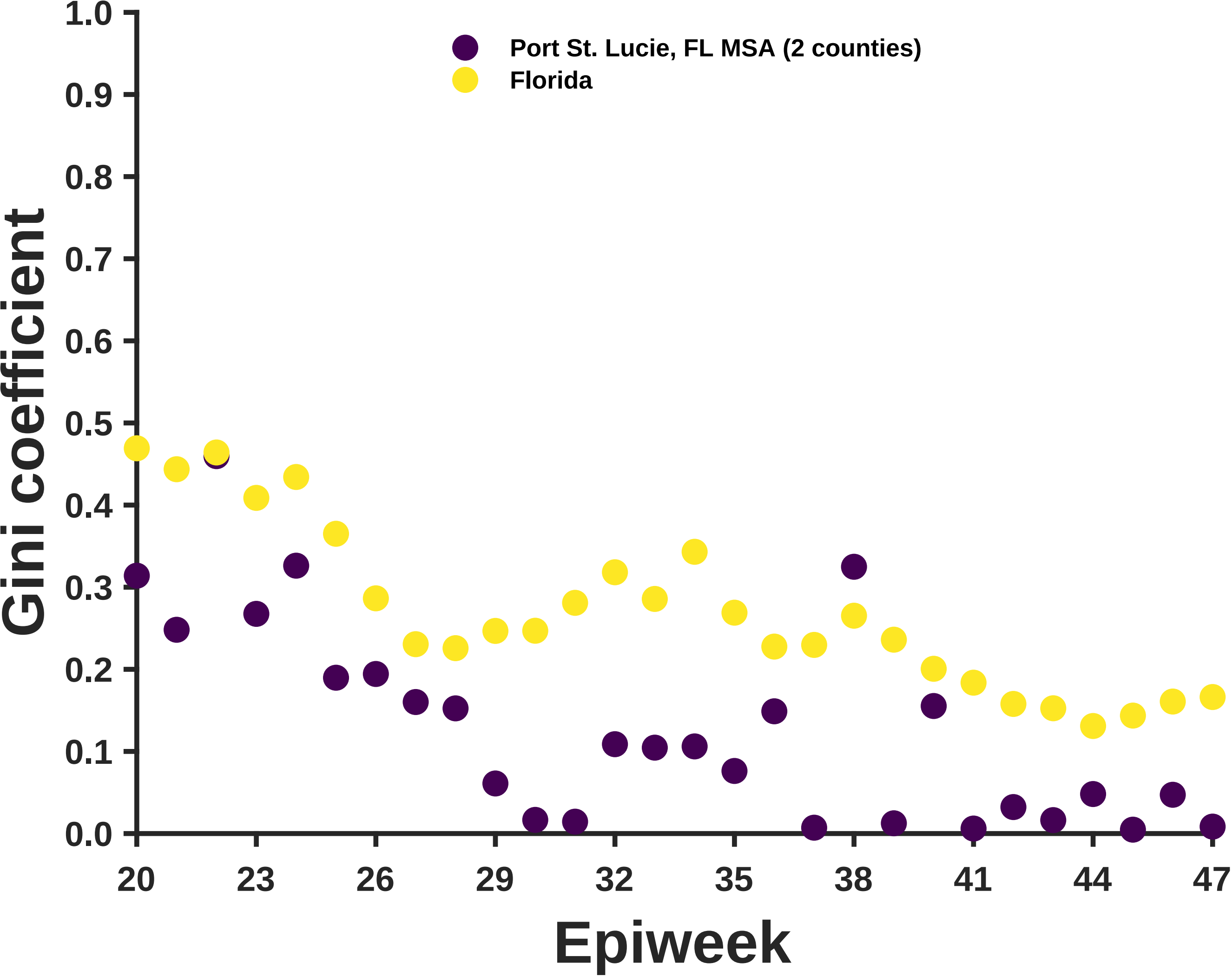

Port St. Lucie, FL MSA (2 counties)
Florida
Gini coefficient
Epiweek
1.0
0.9
0.8
0.7
0.6
0.5
0.4
0.3
0.2
0.1
0.0
20
23
26
29
32
35
38
41
44
47

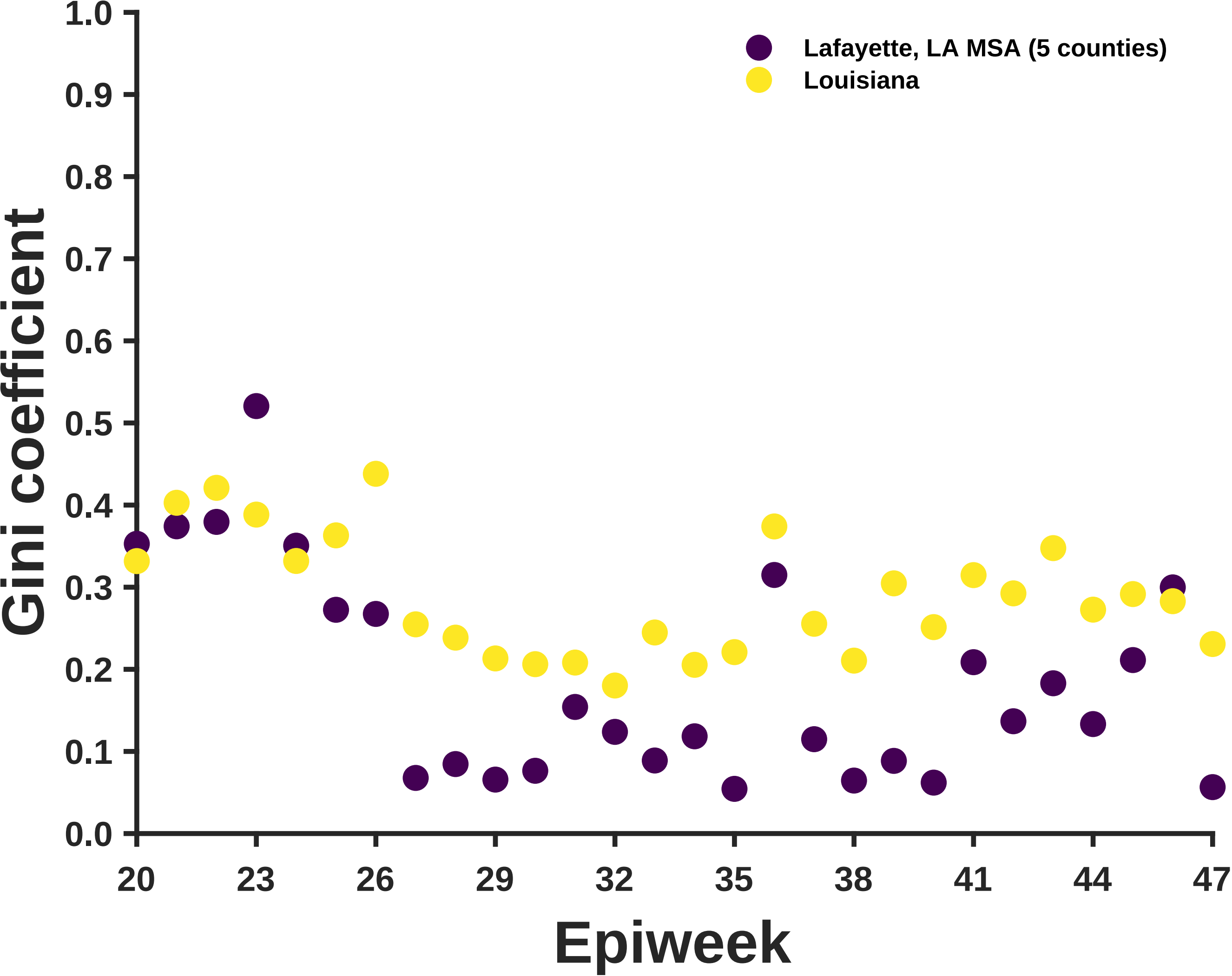

Lafayette, LA MSA (5 counties)
Louisiana
Gini coefficient
1.0
0.9
0.8
0.7
0.6
0.5
0.4
0.3
0.2
0.1
0.0
20
23
26
29
32
35
38
41
44
47
Epiweek

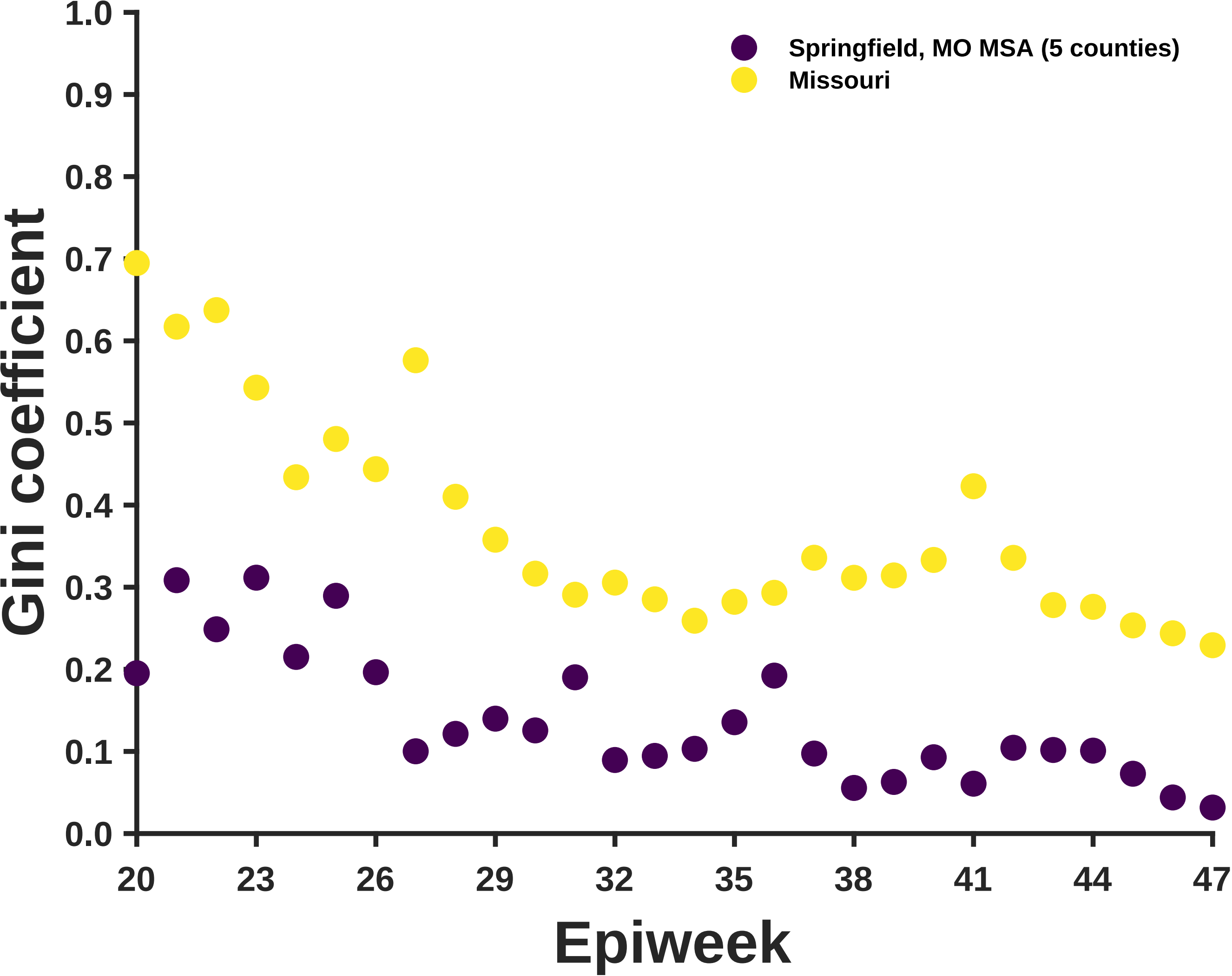

Springfield, MO MSA (5 counties)
Missouri
Gini coefficient
Epiweek
1.0
0.9
0.8
0.7
0.6
0.5
0.4
0.3
0.2
0.1
0.0
20
23
26
29
32
35
38
41
44
47

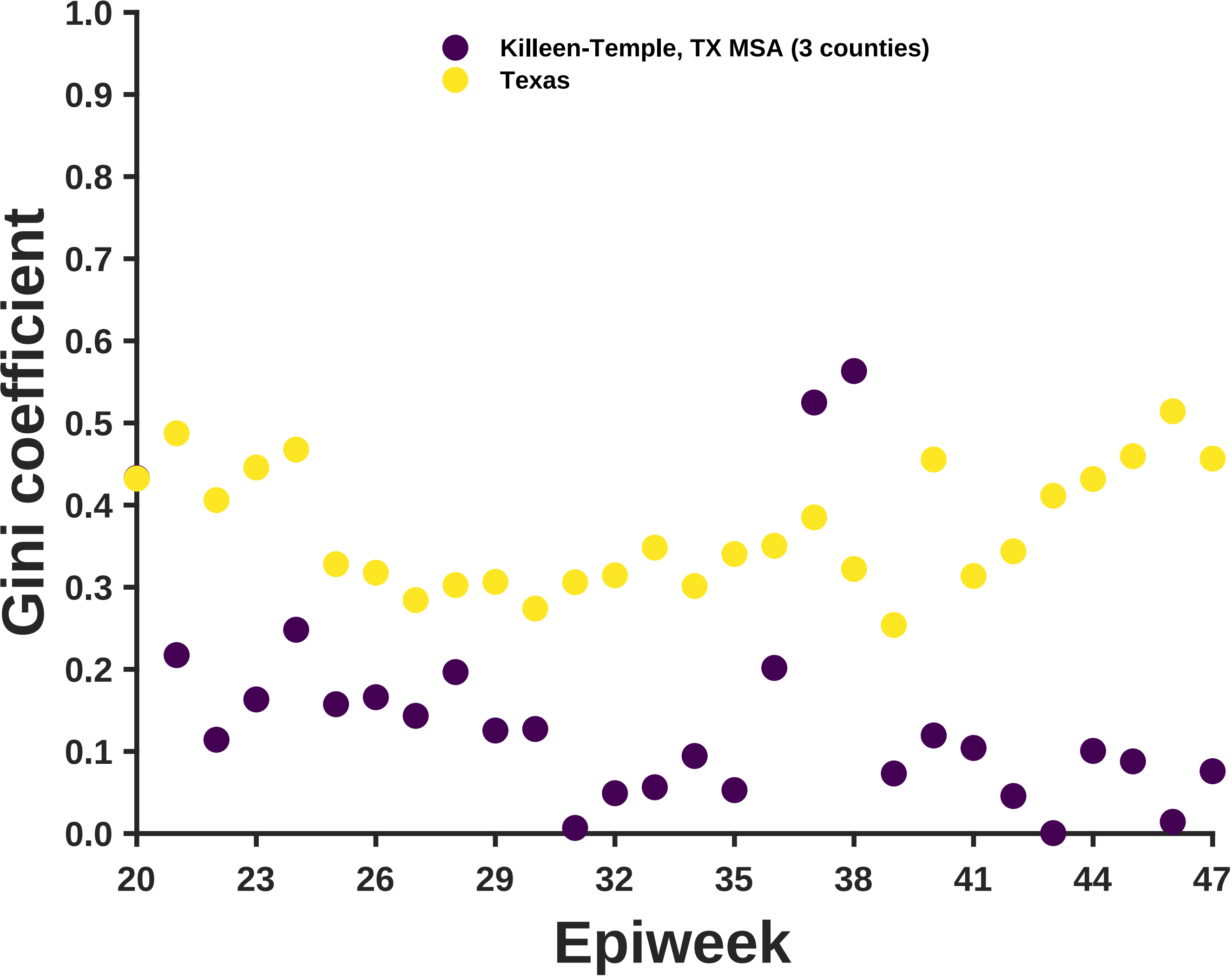
Killeen-Temple, TX MSA (3 counties)
Texas
Gini coefficient
Epiweek
1.0
0.9
0.8
0.7
0.6
0.5
0.4
0.3
0.2
0.1
0.0
20
23
26
29
32
35
38
41
44
47

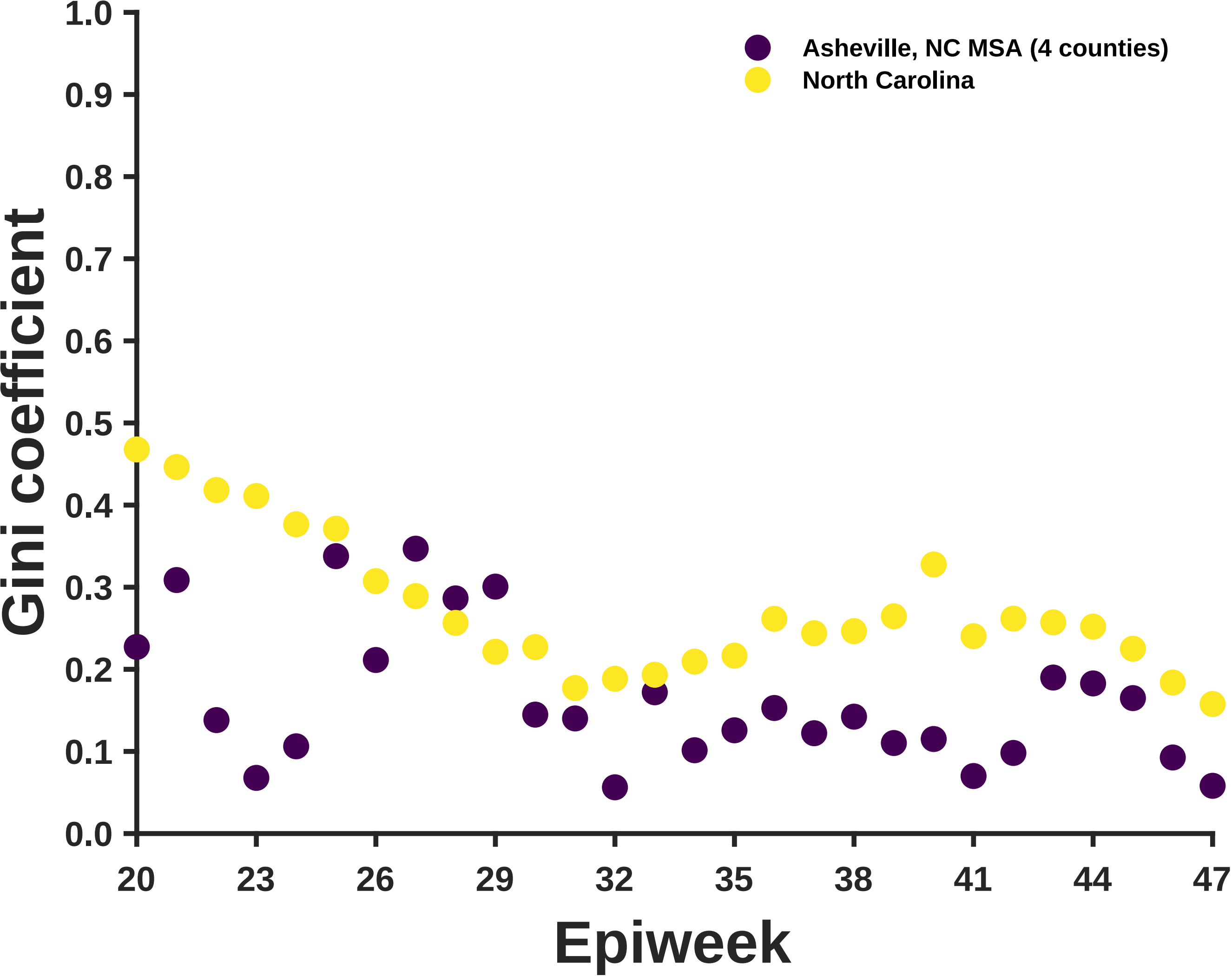
1.0
0.9
0.8
0.7
0.6
0.5
0.4
0.3
0.2
0.1
0.0
Gini coefficient
Asheville, NC MSA (4 counties)
North Carolina
20
23
26
29
32
35
38
41
44
47
Epiweek

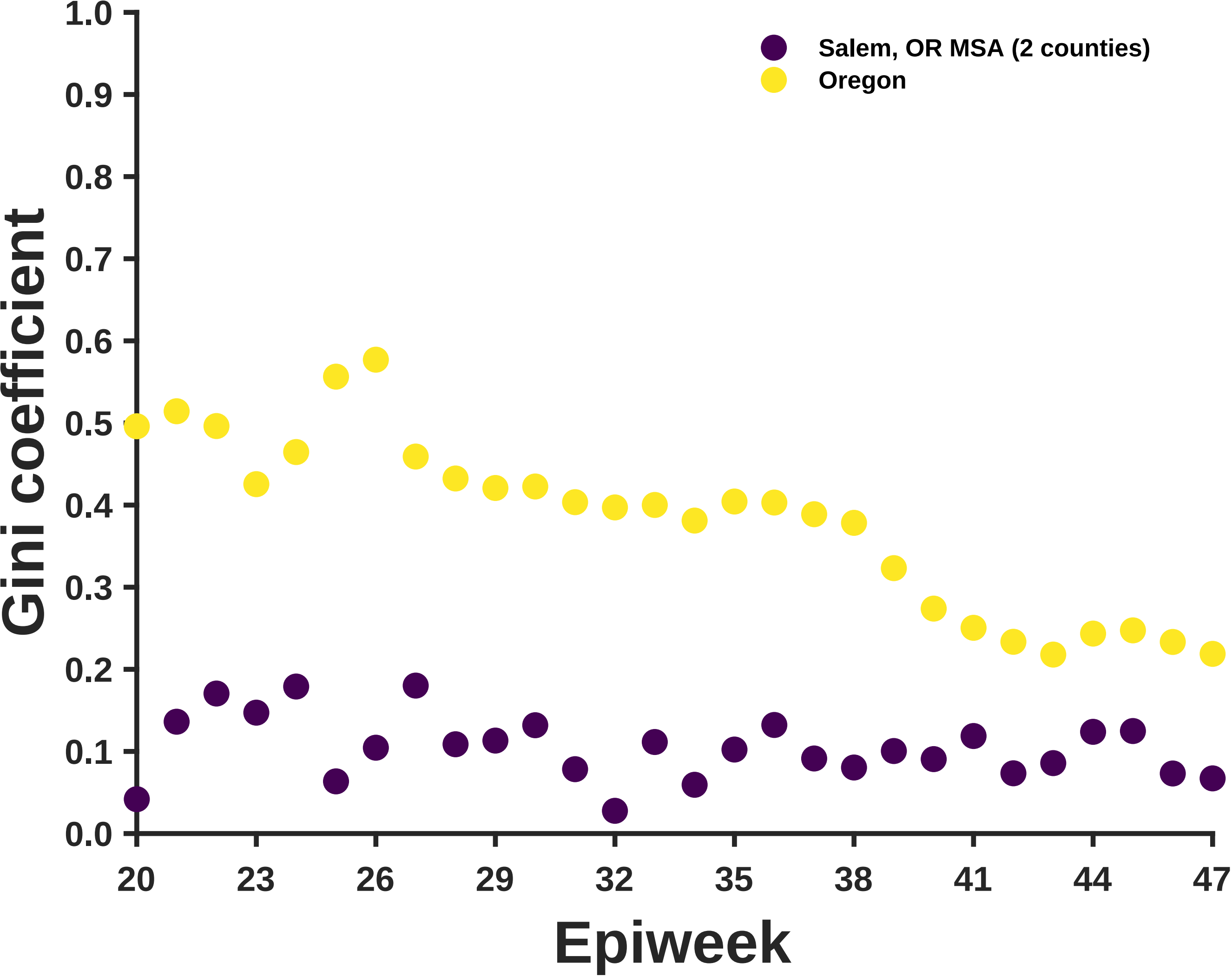
Salem, OR MSA (2 counties)
Oregon
Gini coefficient
Epiweek
1.0
0.9
0.8
0.7
0.6
0.5
0.4
0.3
0.2
0.1
0.0
20
23
26
29
32
35
38
41
44
47

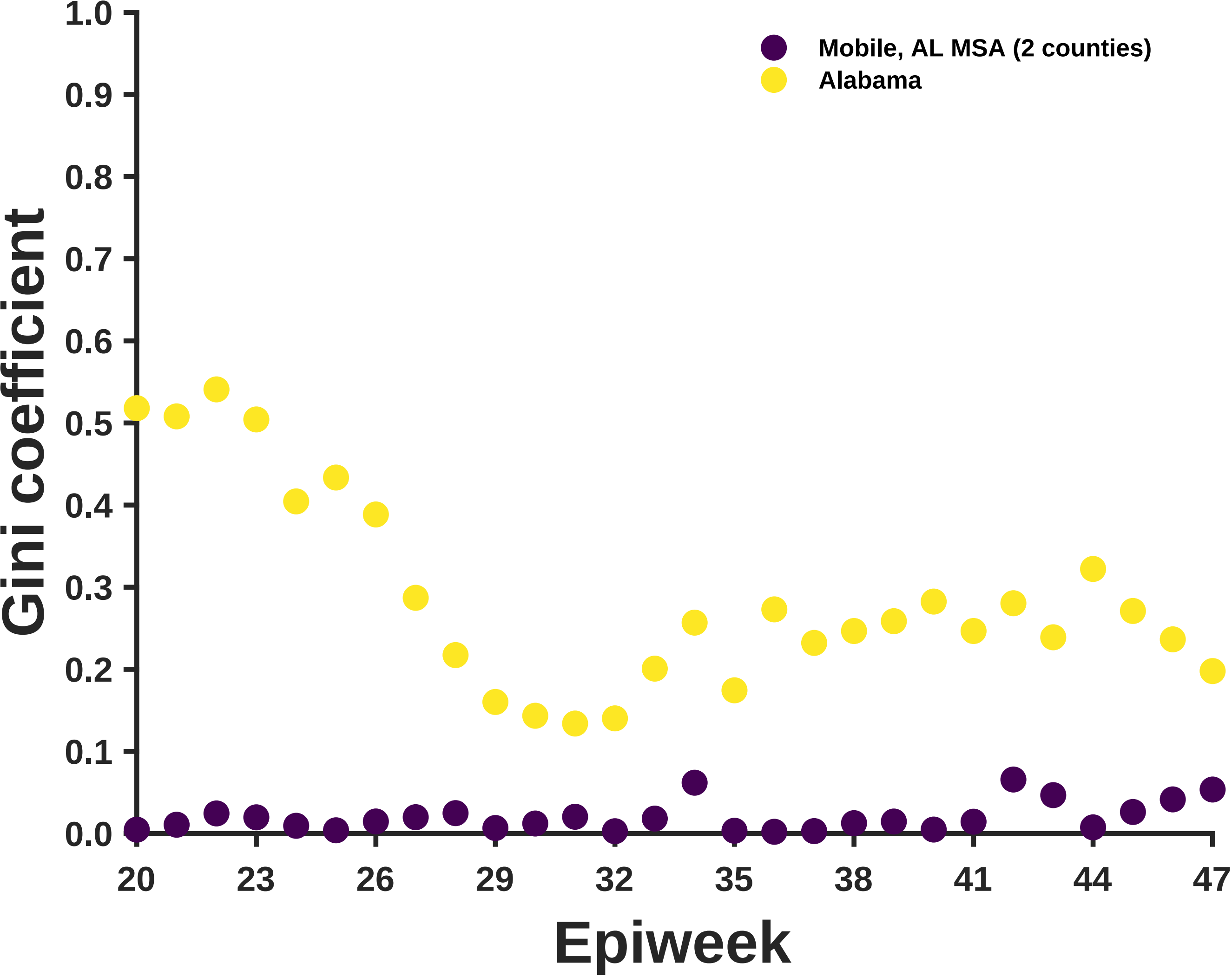

Mobile, AL MSA (2 counties)
Alabama
Gini coefficient
0.0
0.1
0.2
0.3
0.4
0.5
0.6
0.7
0.8
0.9
1.0
20
23
26
29
32
35
38
41
44
47
Epiweek

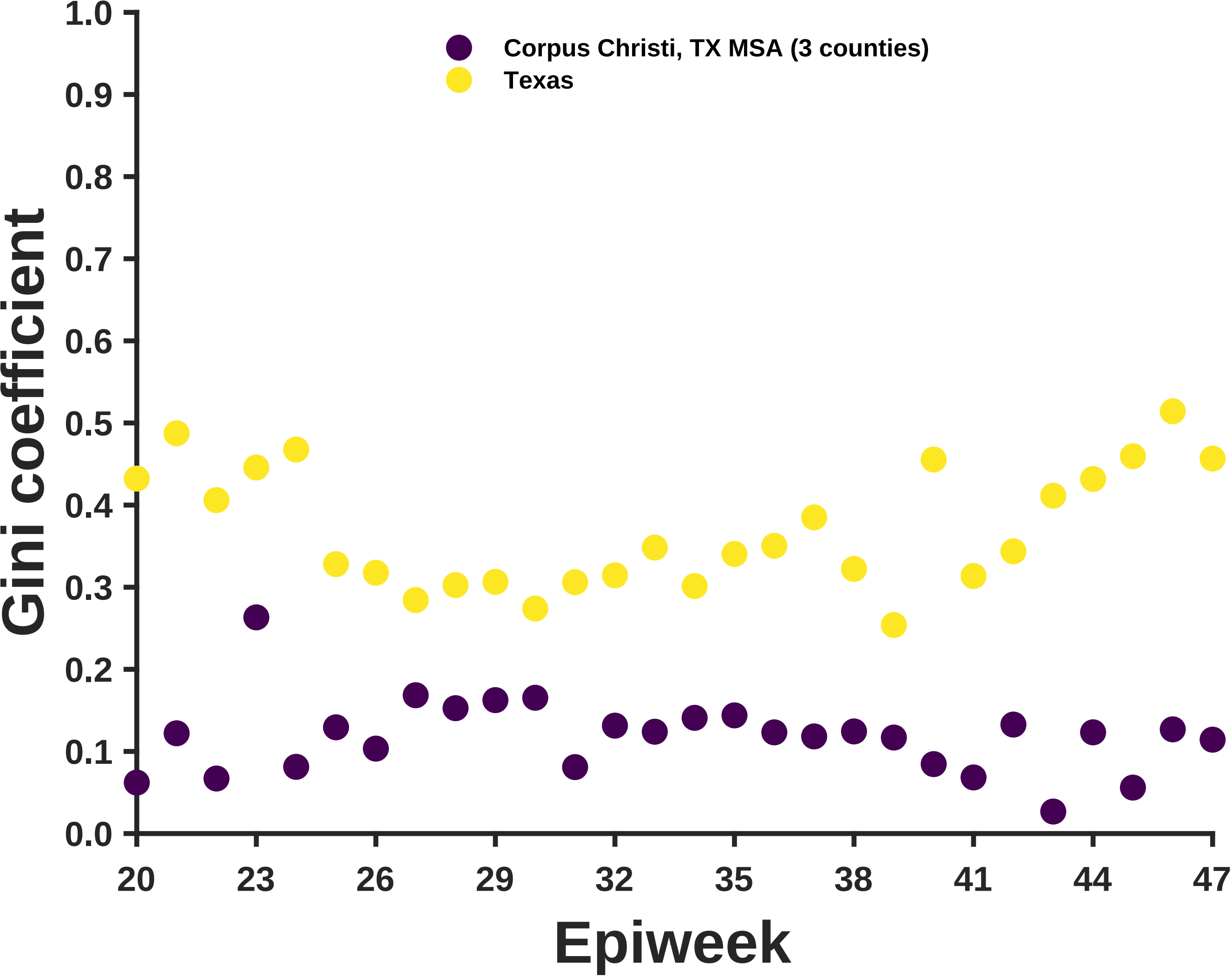

Corpus Christi, TX MSA (3 counties)
Texas
Gini coefficient
Epiweek
1.0
0.9
0.8
0.7
0.6
0.5
0.4
0.3
0.2
0.1
0.0
20
23
26
29
32
35
38
41
44
47

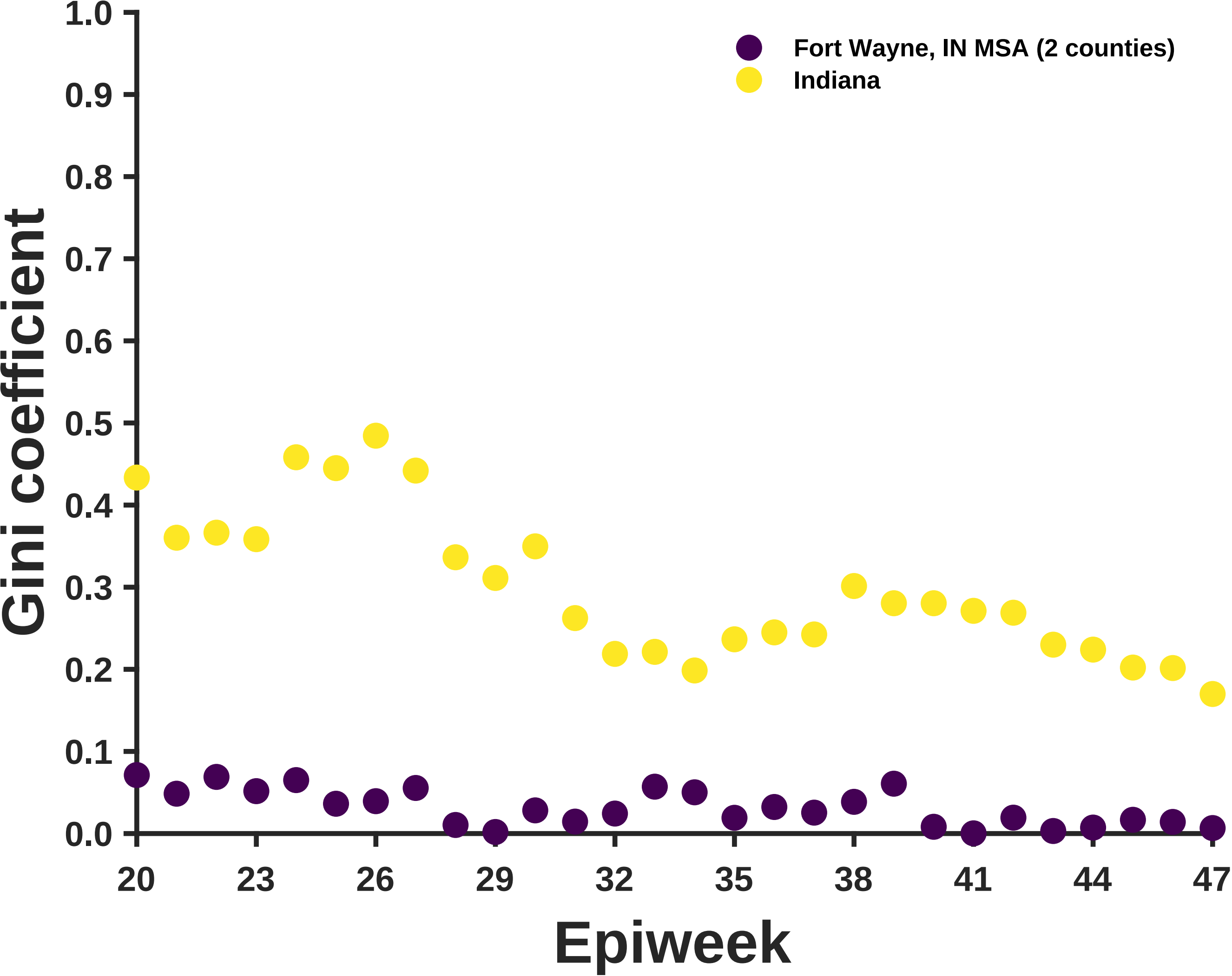

Fort Wayne, IN MSA (2 counties)
Indiana
Gini coefficient
1.0
0.9
0.8
0.7
0.6
0.5
0.4
0.3
0.2
0.1
0.0
20
23
26
29
32
35
38
41
44
47
Epiweek

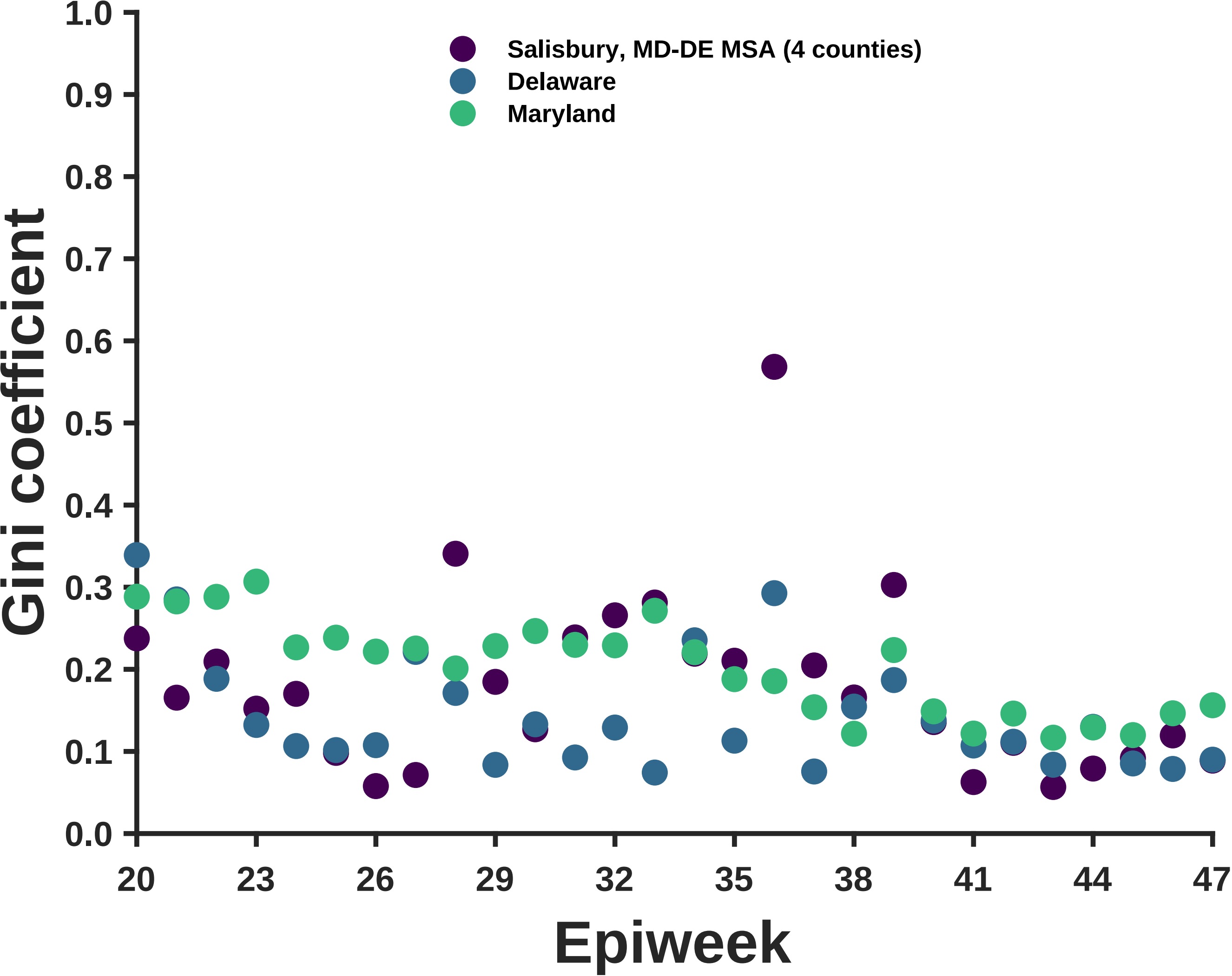
Salisbury, MD-DE MSA (4 counties)
Delaware
Maryland
Gini coefficient
1.0
0.9
0.8
0.7
0.6
0.5
0.4
0.3
0.2
0.1
0.0
20
23
26
29
32
35
38
41
44
47
Epiweek

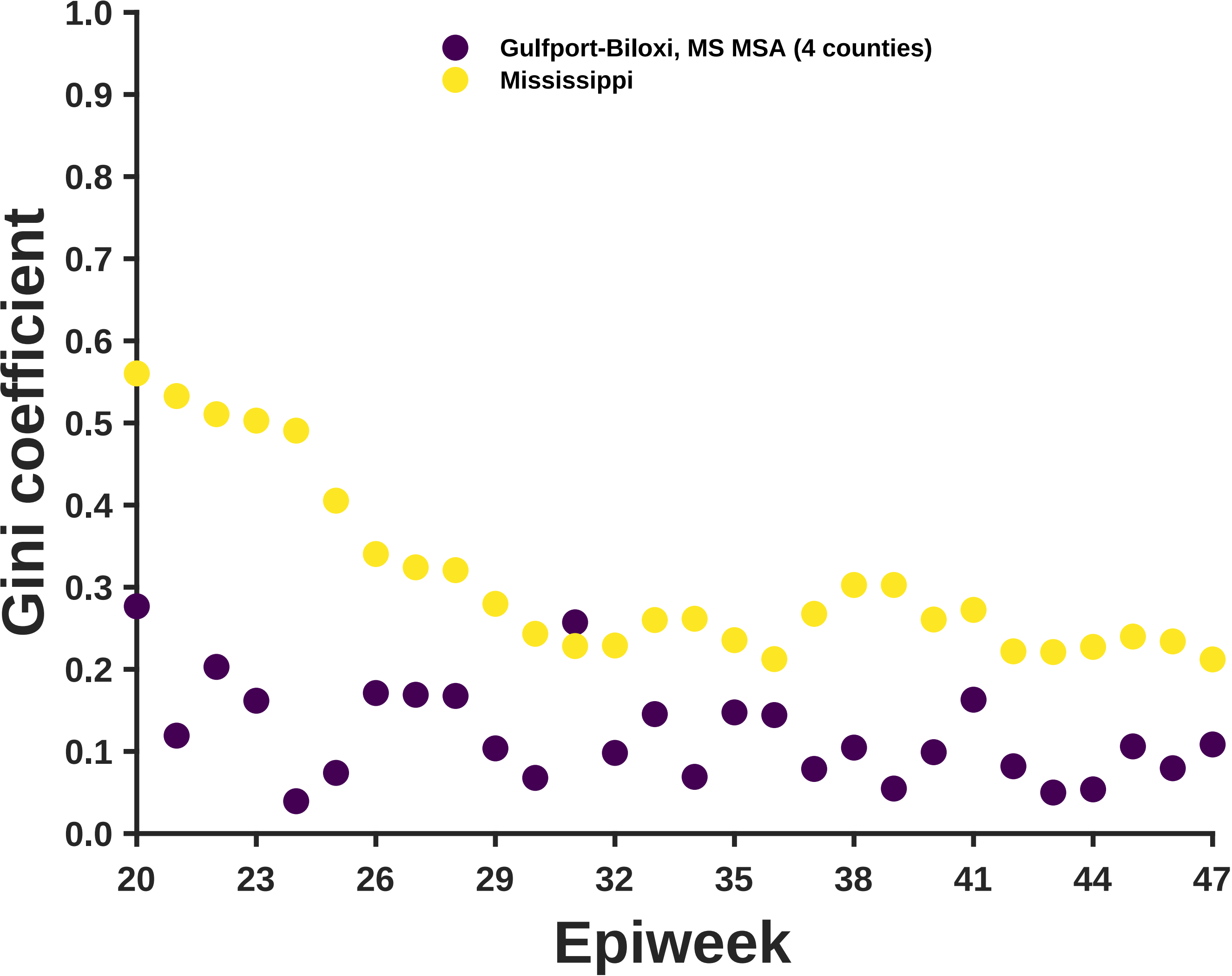

Gulfport-Biloxi, MS MSA (4 counties)
Mississippi
Gini coefficient
1.0
0.9
0.8
0.7
0.6
0.5
0.4
0.3
0.2
0.1
0.0
20
23
26
29
32
35
38
41
44
47
Epiweek

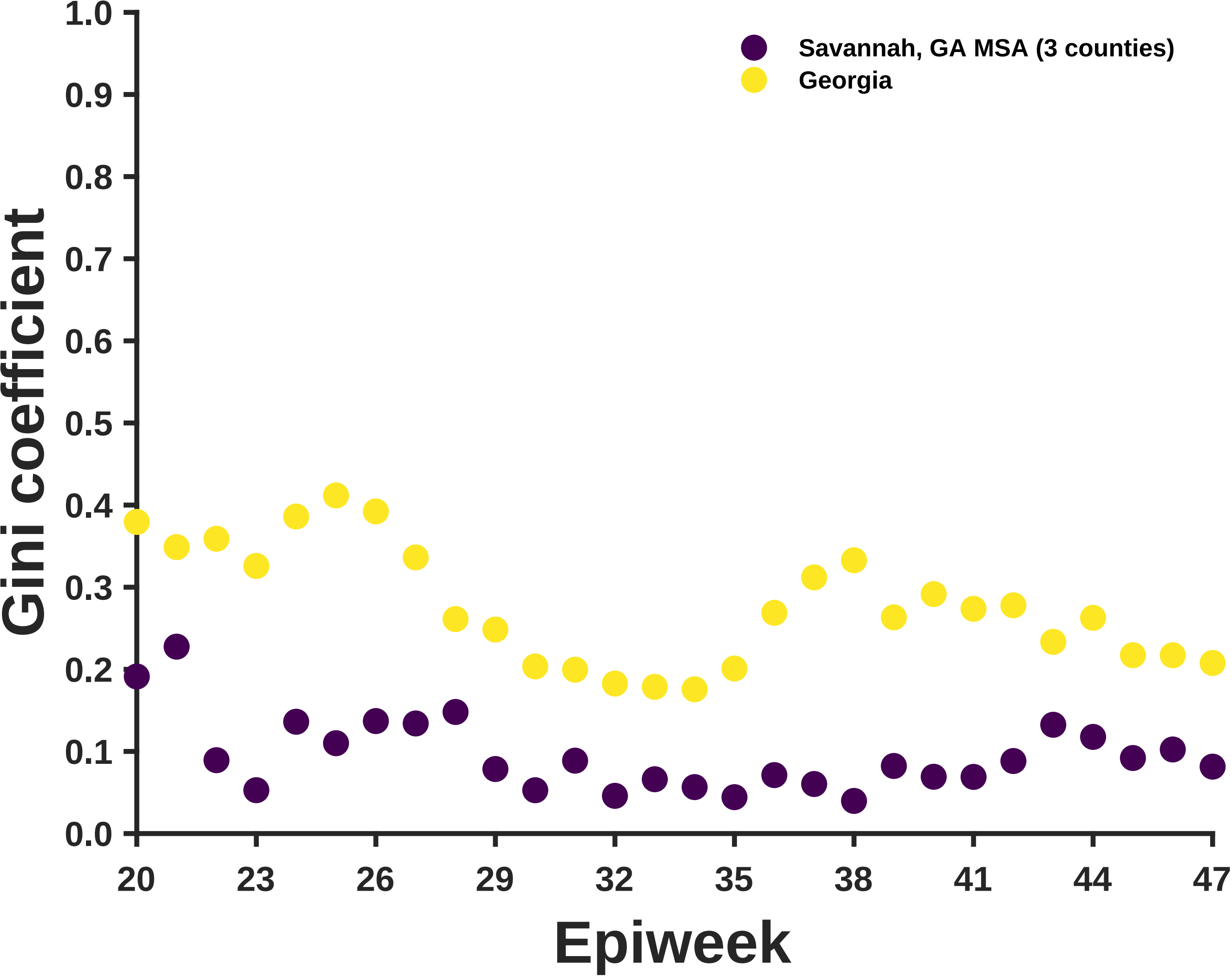

Savannah, GA MSA (3 counties)
Georgia
Gini coefficient
1.0
0.9
0.8
0.7
0.6
0.5
0.4
0.3
0.2
0.1
0.0
20
23
26
29
32
35
38
41
44
47
Epiweek

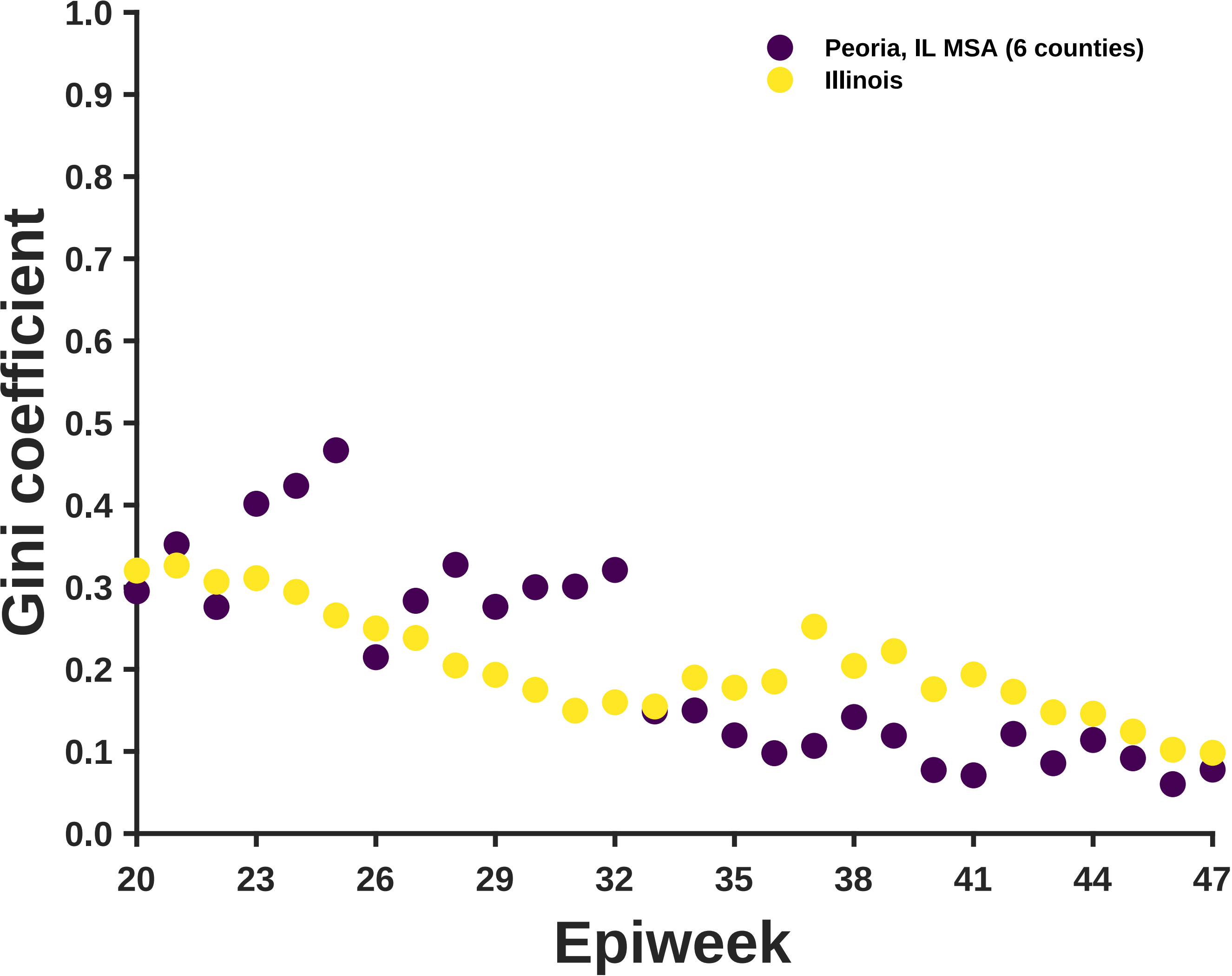

Gini coefficient
Epiweek
Peoria, IL MSA (6 counties)
Illinois
1.0
0.9
0.8
0.7
0.6
0.5
0.4
0.3
0.2
0.1
0.0
20
23
26
29
32
35
38
41
44
47

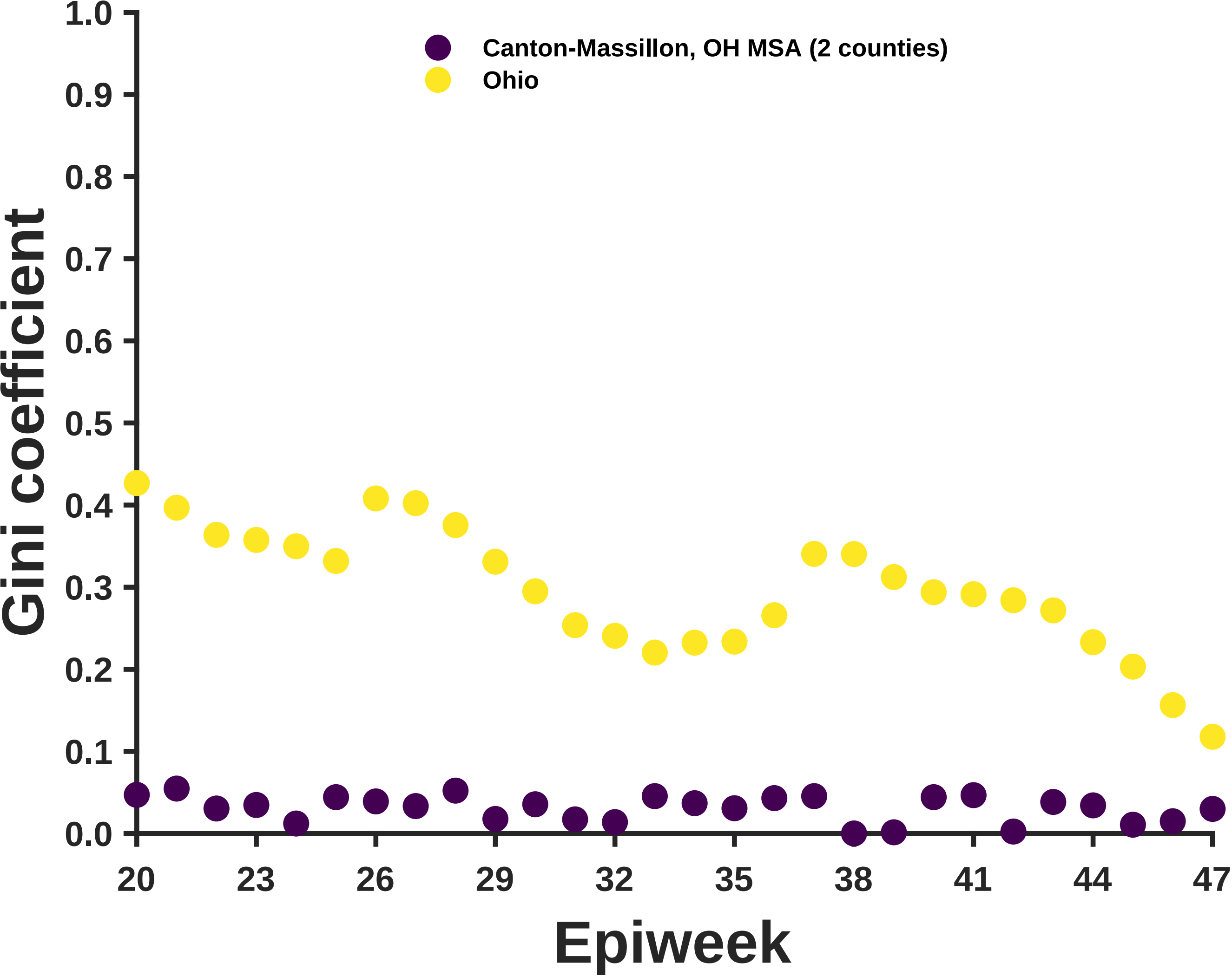
Canton-Massillon, OH MSA (2 counties)
Ohio
Gini coefficient
1.0
0.9
0.8
0.7
0.6
0.5
0.4
0.3
0.2
0.1
0.0
20
23
26
29
32
35
38
41
44
47
Epiweek

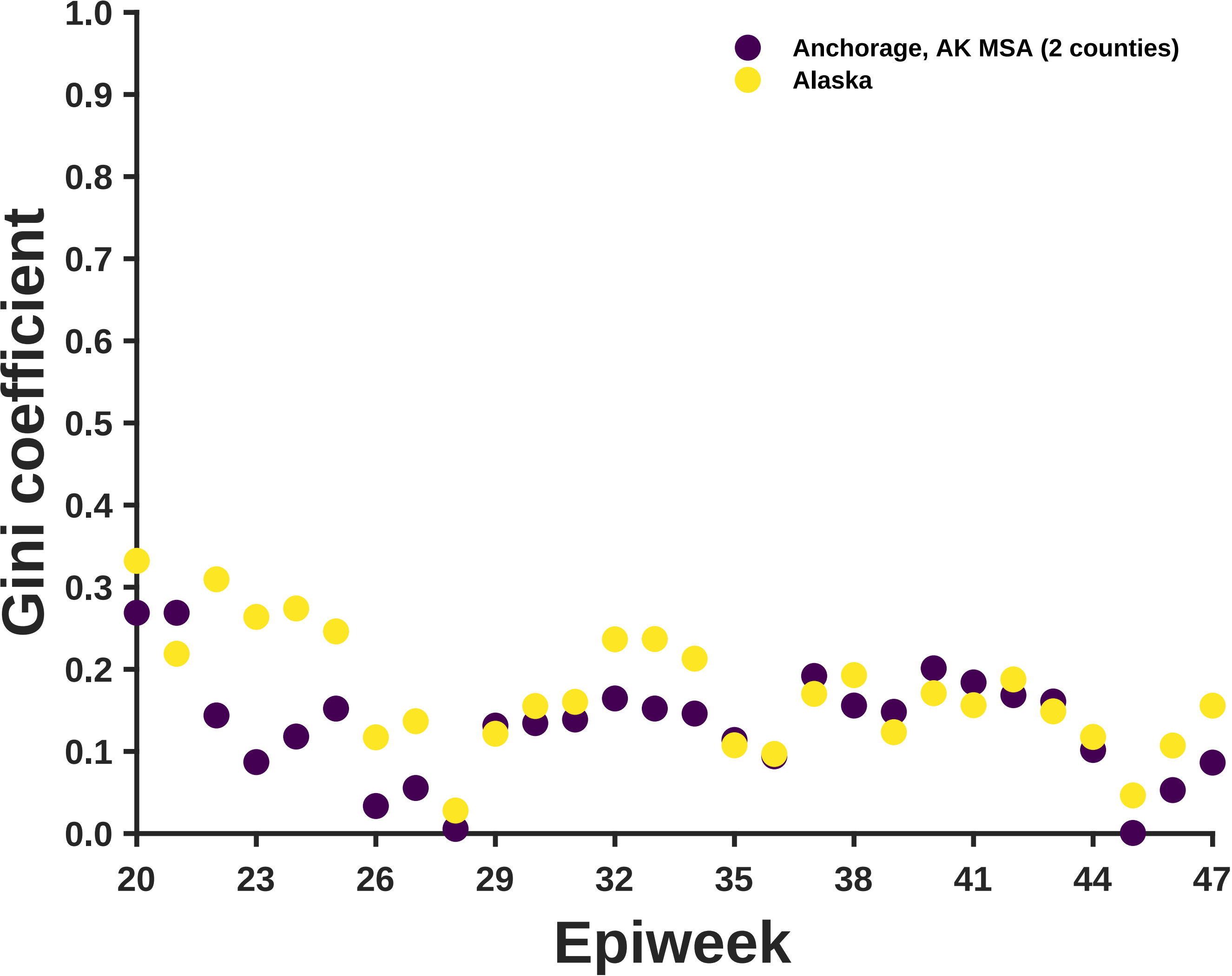

Anchorage, AK MSA (2 counties)
Alaska
Gini coefficient
1.0
0.9
0.8
0.7
0.6
0.5
0.4
0.3
0.2
0.1
0.0
20
23
26
29
32
35
38
41
44
47
Epiweek

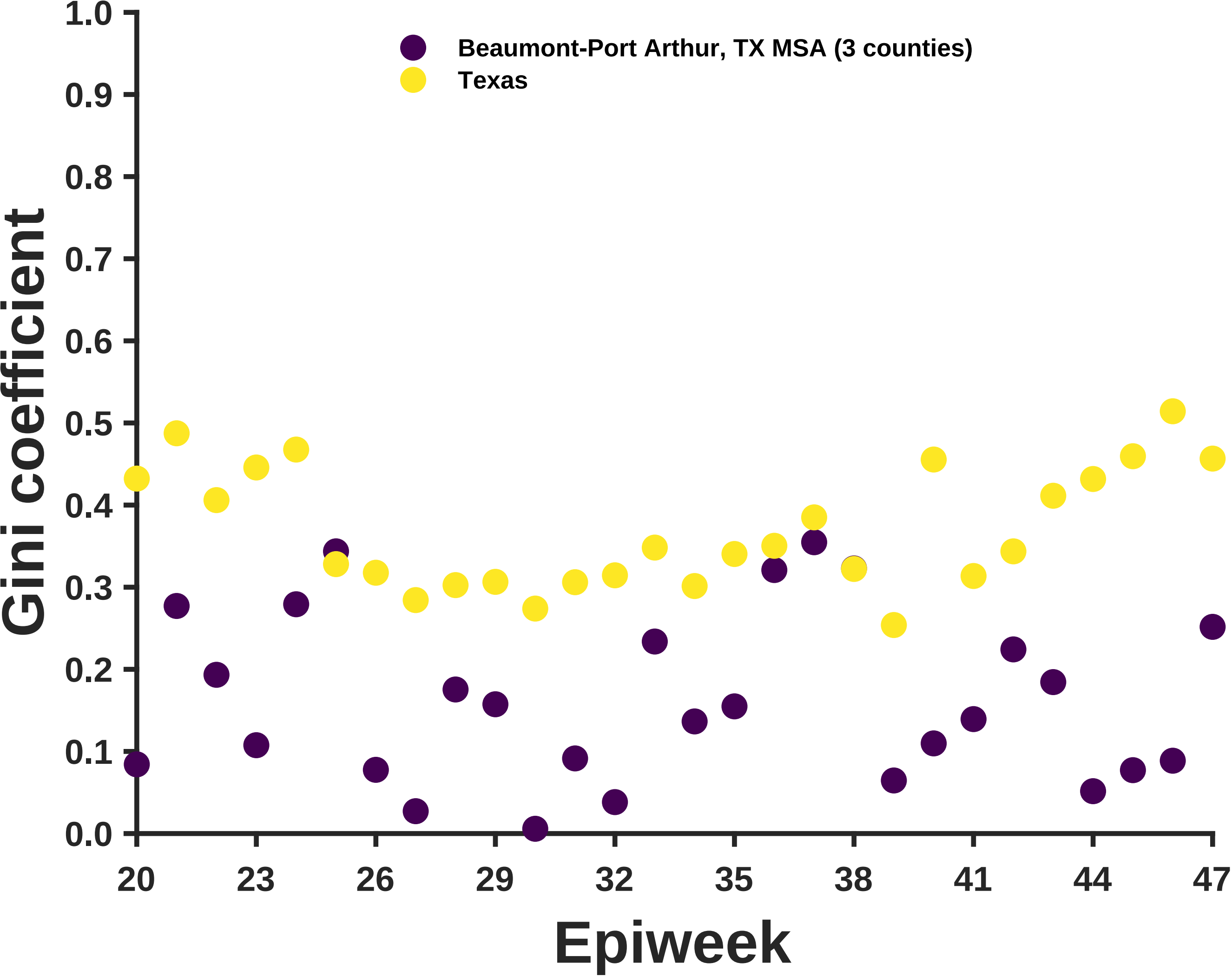
Beaumont-Port Arthur, TX MSA (3 counties)
Texas
Gini coefficient
Epiweek
0.0
0.1
0.2
0.3
0.4
0.5
0.6
0.7
0.8
0.9
1.0
20
23
26
29
32
35
38
41
44
47

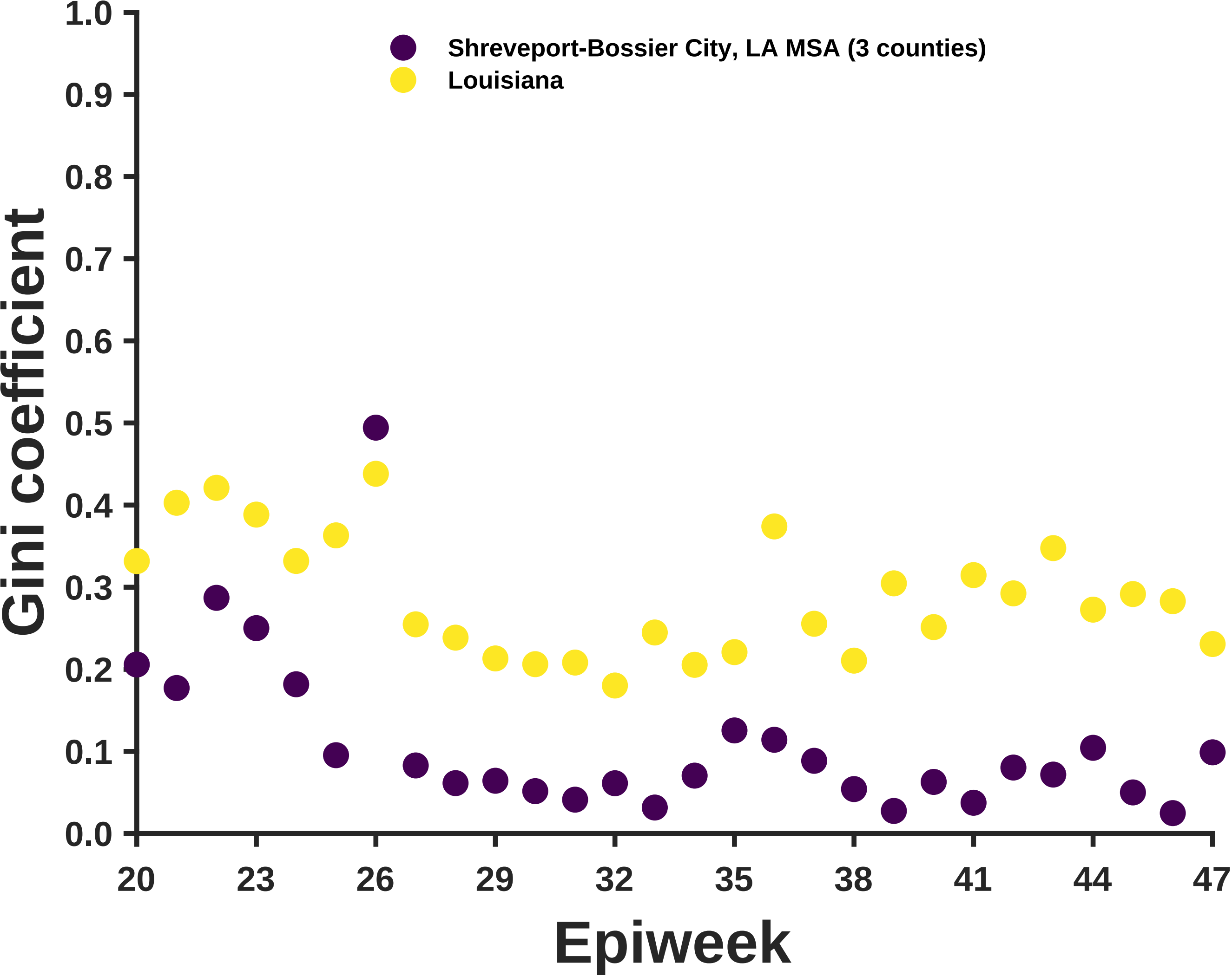
Shreveport-Bossier City, LA MSA (3 counties)
Louisiana
Gini coefficient
0.0
0.1
0.2
0.3
0.4
0.5
0.6
0.7
0.8
0.9
1.0
20
23
26
29
32
35
38
41
44
47
Epiweek

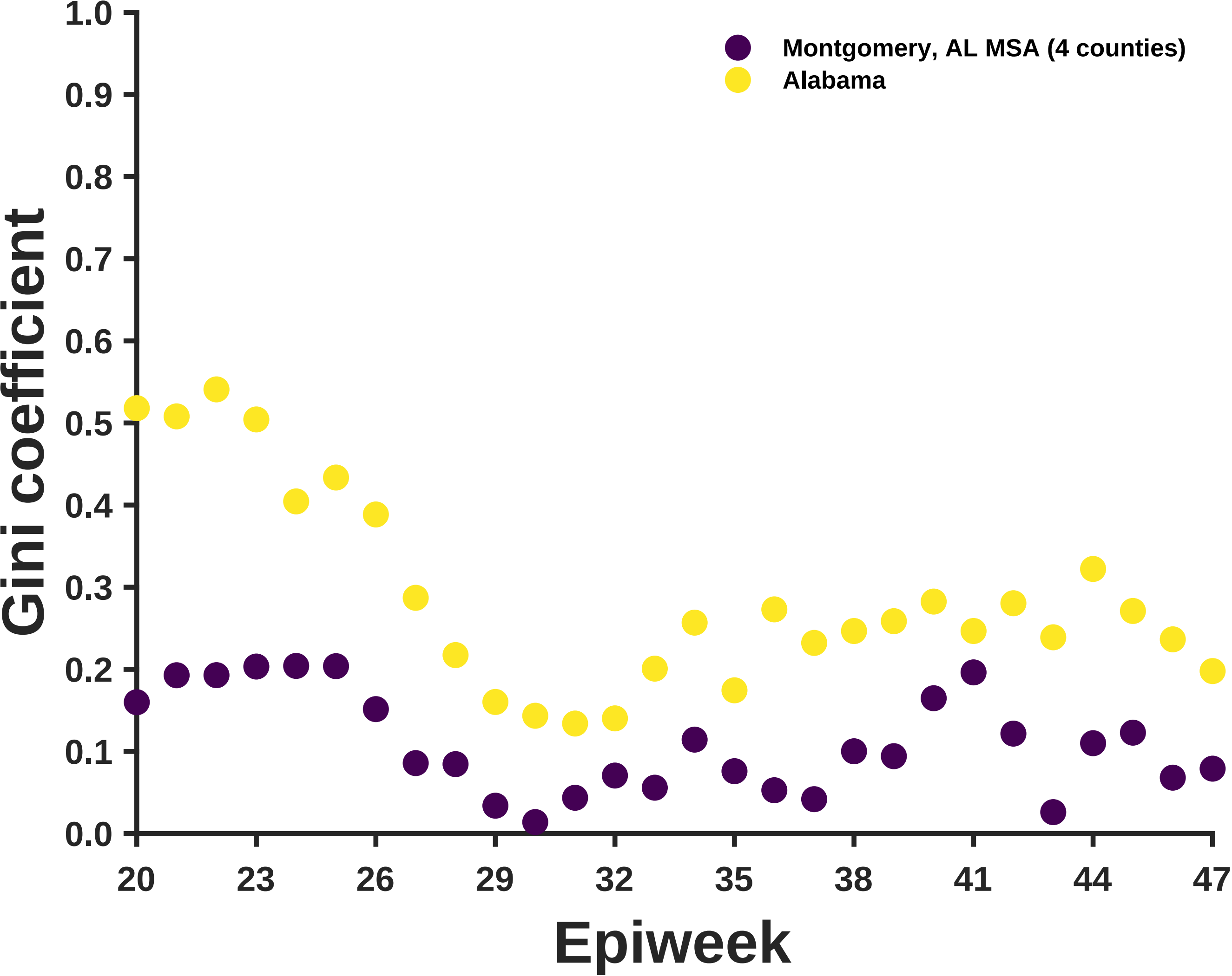

Montgomery, AL MSA (4 counties)
Alabama
Gini coefficient
1.0
0.9
0.8
0.7
0.6
0.5
0.4
0.3
0.2
0.1
0.0
20
23
26
29
32
35
38
41
44
47
Epiweek

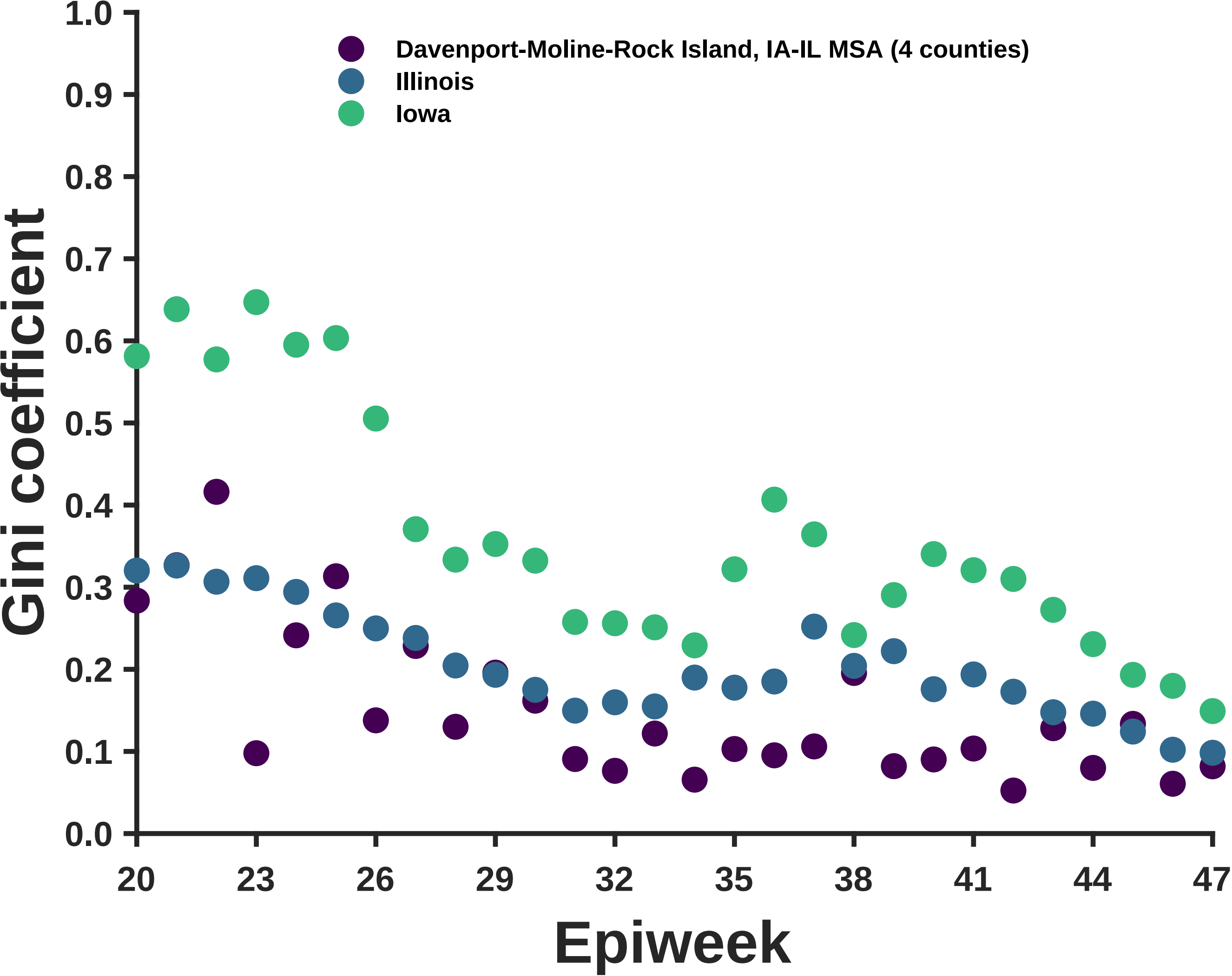

Davenport-Moline-Rock Island, IA-IL MSA (4 counties)
Illinois
Iowa
Gini coefficient
1.0
0.9
0.8
0.7
0.6
0.5
0.4
0.3
0.2
0.1
0.0
20
23
26
29
32
35
38
41
44
47
Epiweek

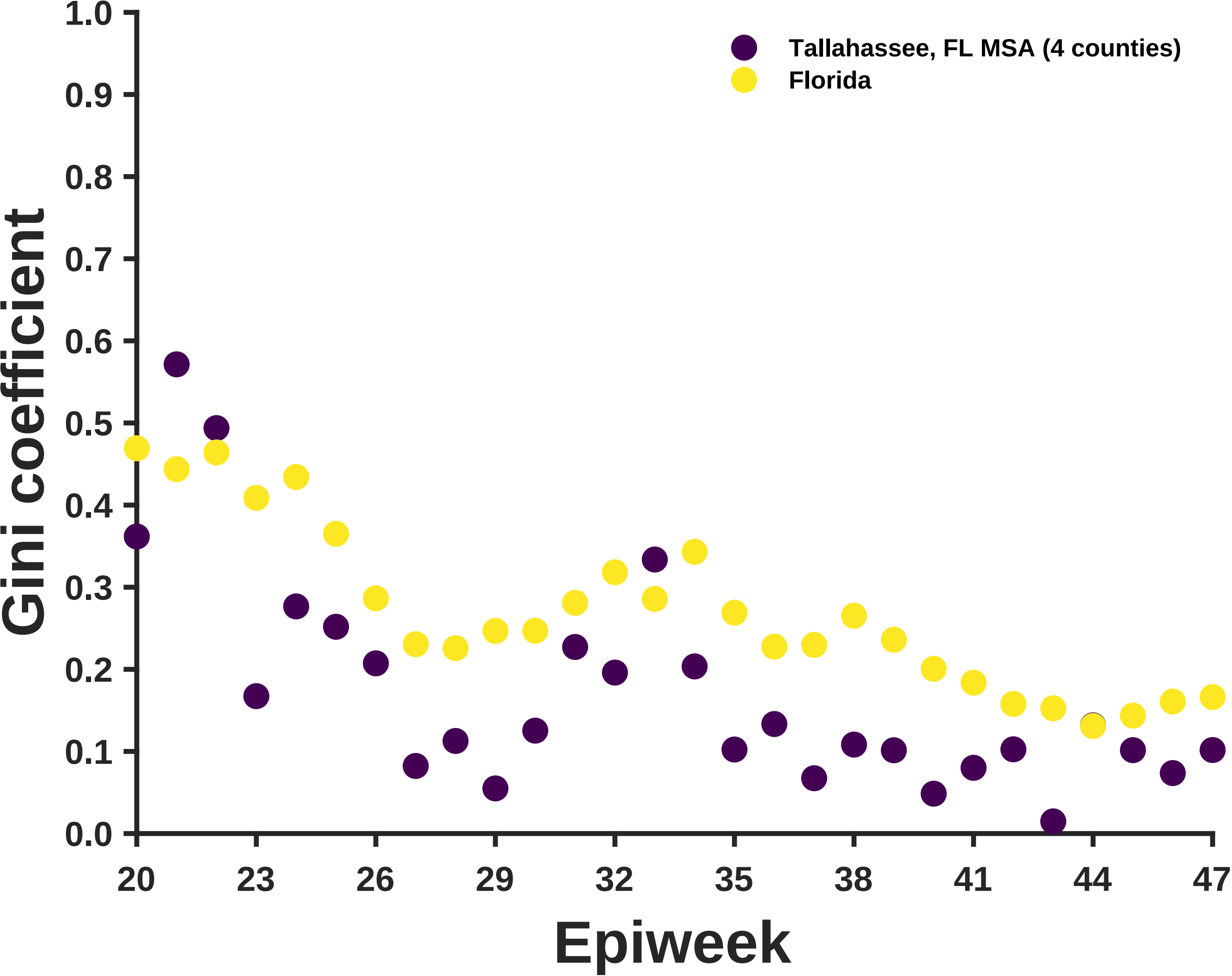

Tallahassee, FL MSA (4 counties)
Florida
Gini coefficient
1.0
0.9
0.8
0.7
0.6
0.5
0.4
0.3
0.2
0.1
0.0
20
23
26
29
32
35
38
41
44
47
Epiweek

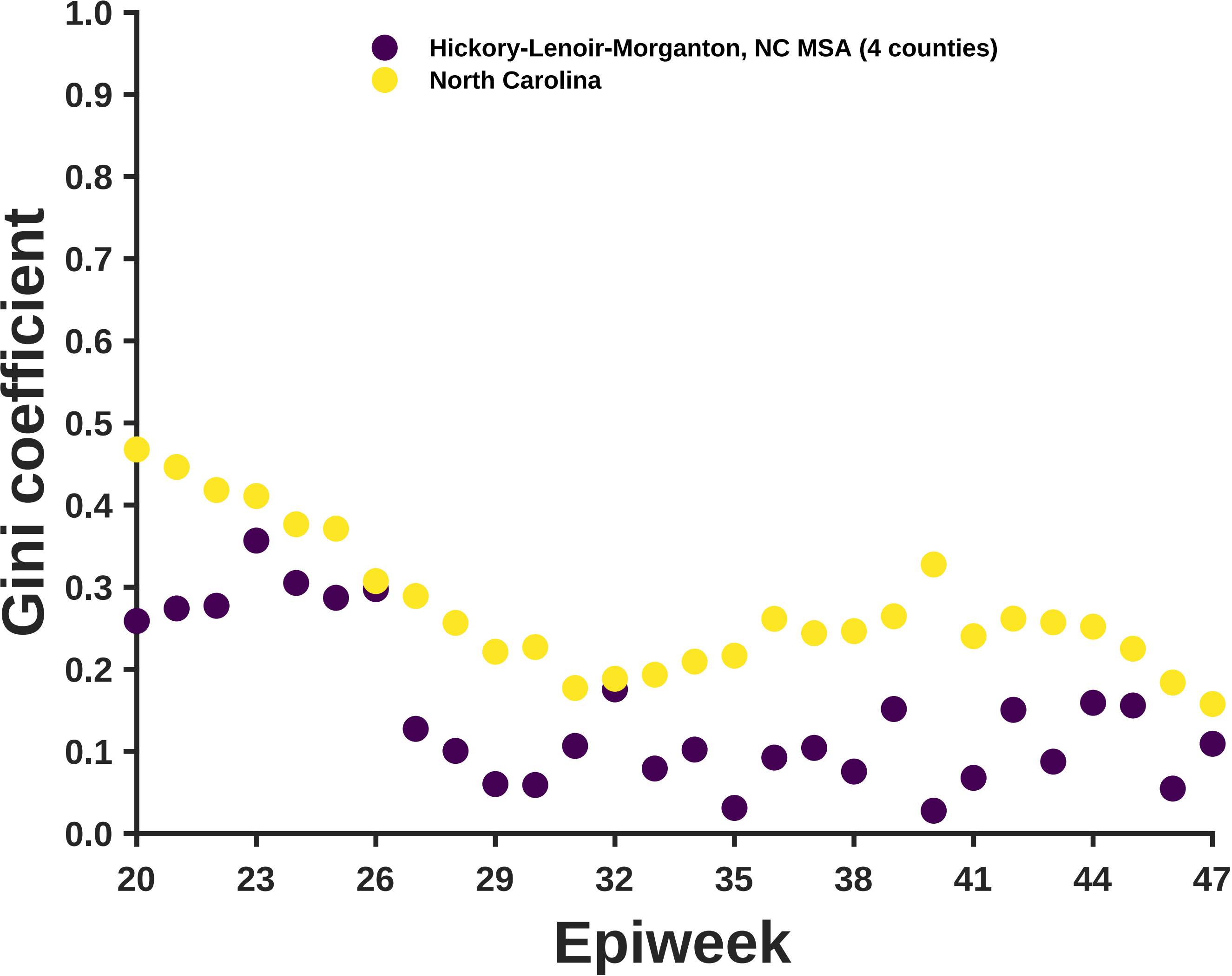

Hickory-Lenoir-Morganton, NC MSA (4 counties)
North Carolina
Gini coefficient
1.0
0.9
0.8
0.7
0.6
0.5
0.4
0.3
0.2
0.1
0.0
20
23
26
29
32
35
38
41
44
47
Epiweek

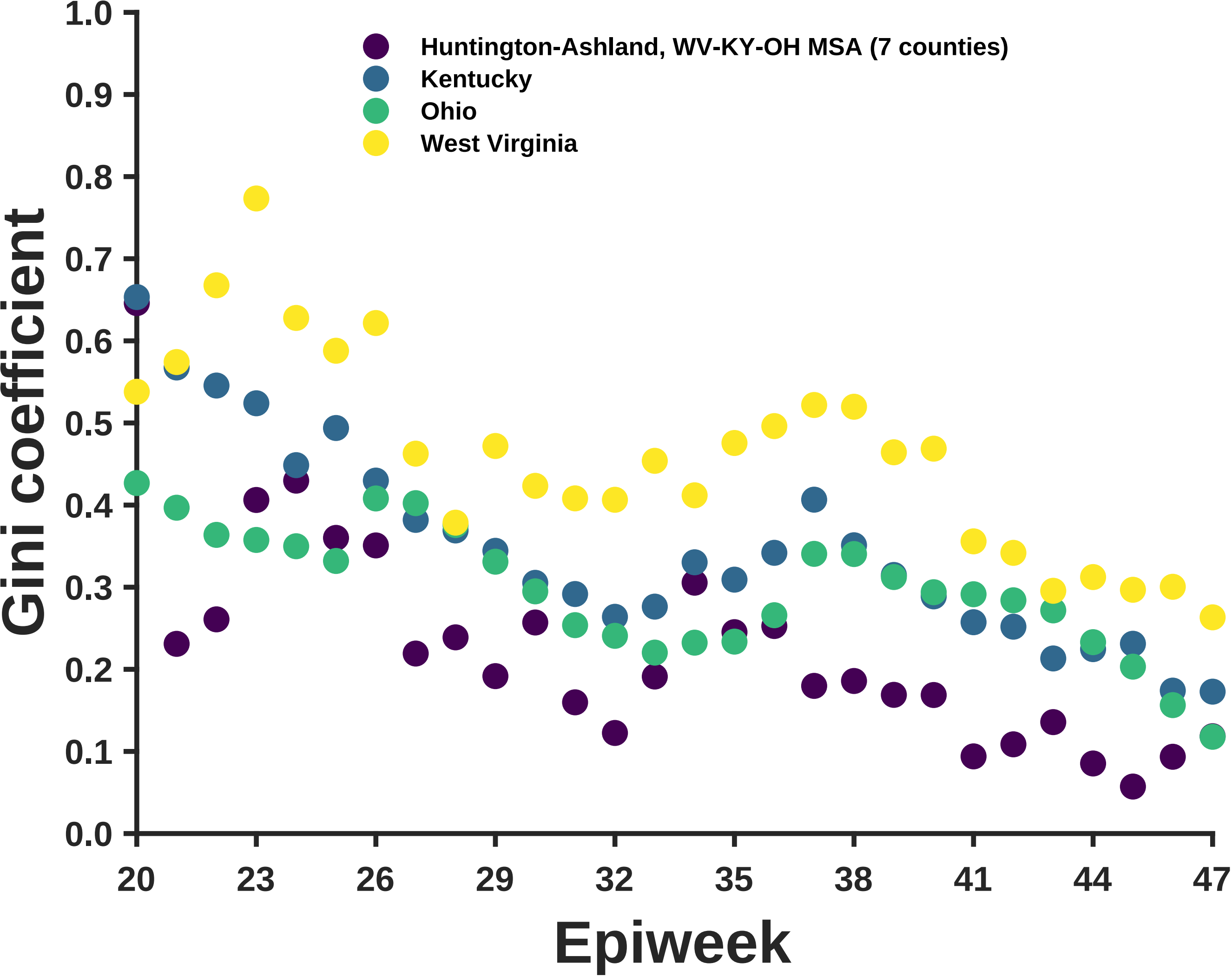

Huntington-Ashland, WV-KY-OH MSA (7 counties)
Kentucky
Ohio
West Virginia
Gini coefficient
Epiweek
0.0
0.1
0.2
0.3
0.4
0.5
0.6
0.7
0.8
0.9
1.0
20
23
26
29
32
35
38
41
44
47

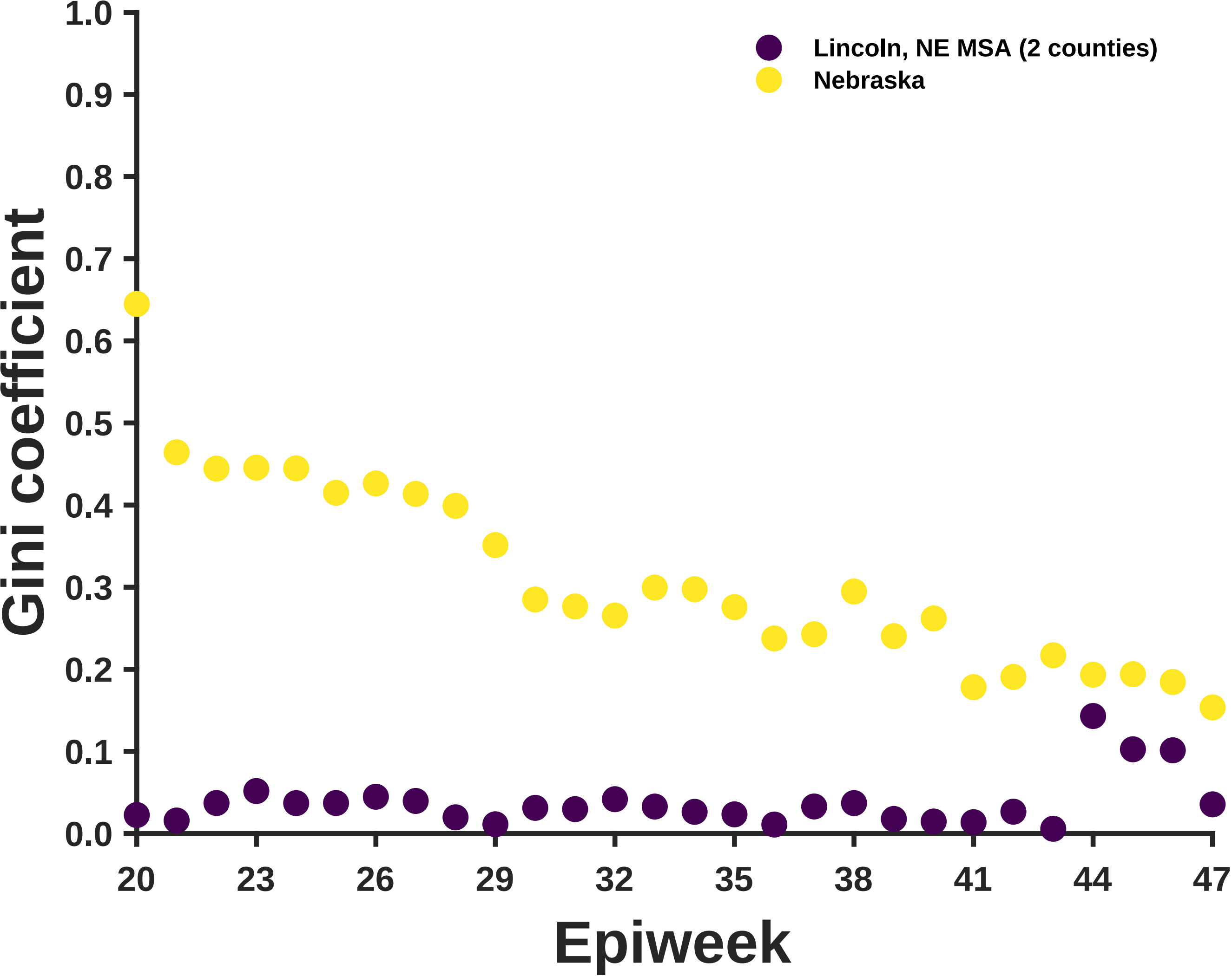

Lincoln, NE MSA (2 counties)
Nebraska
Gini coefficient
1.0
0.9
0.8
0.7
0.6
0.5
0.4
0.3
0.2
0.1
0.0
20
23
26
29
32
35
38
41
44
47
Epiweek

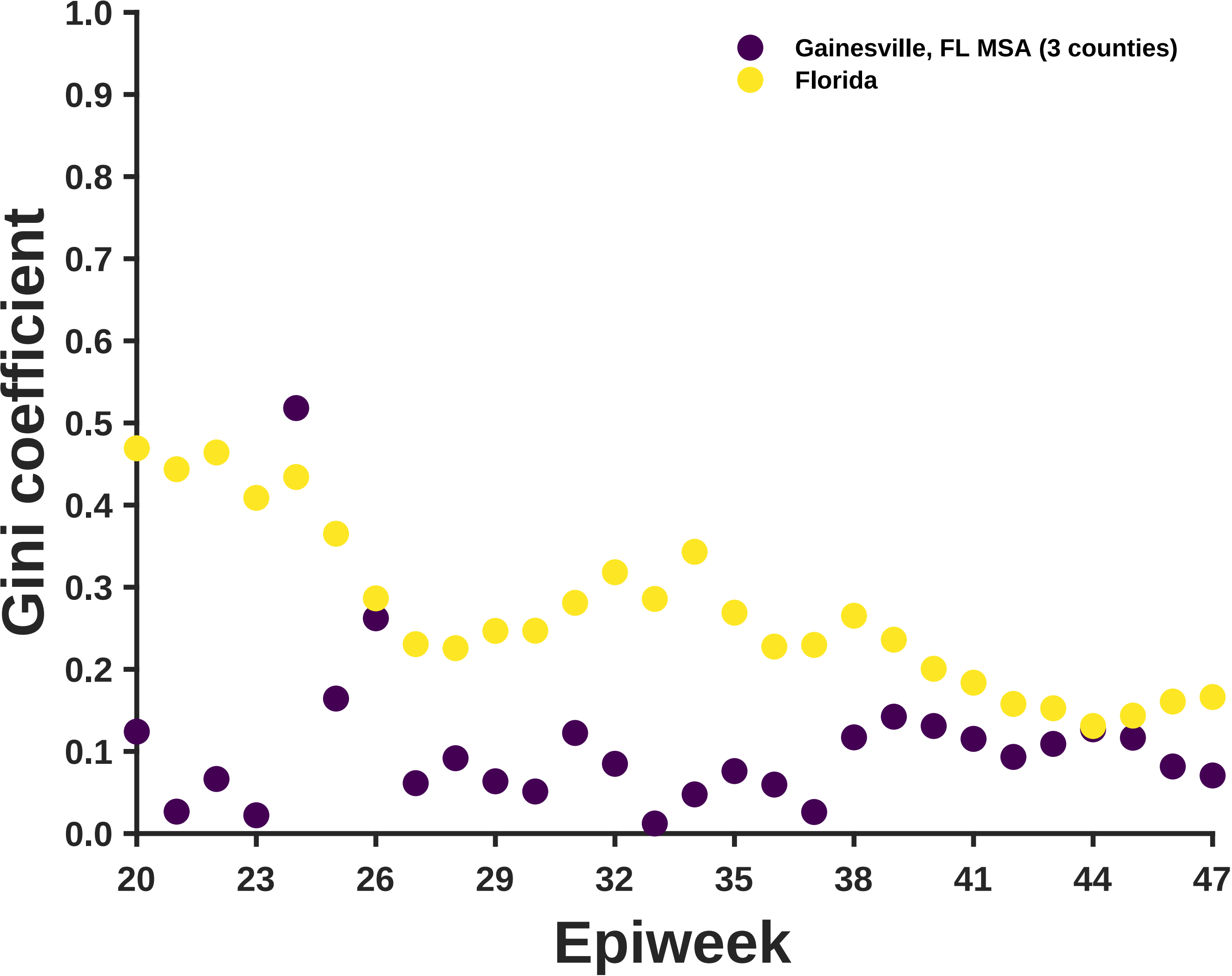

Gainesville, FL MSA (3 counties)
Florida
Gini coefficient
1.0
0.9
0.8
0.7
0.6
0.5
0.4
0.3
0.2
0.1
0.0
20
23
26
29
32
35
38
41
44
47
Epiweek

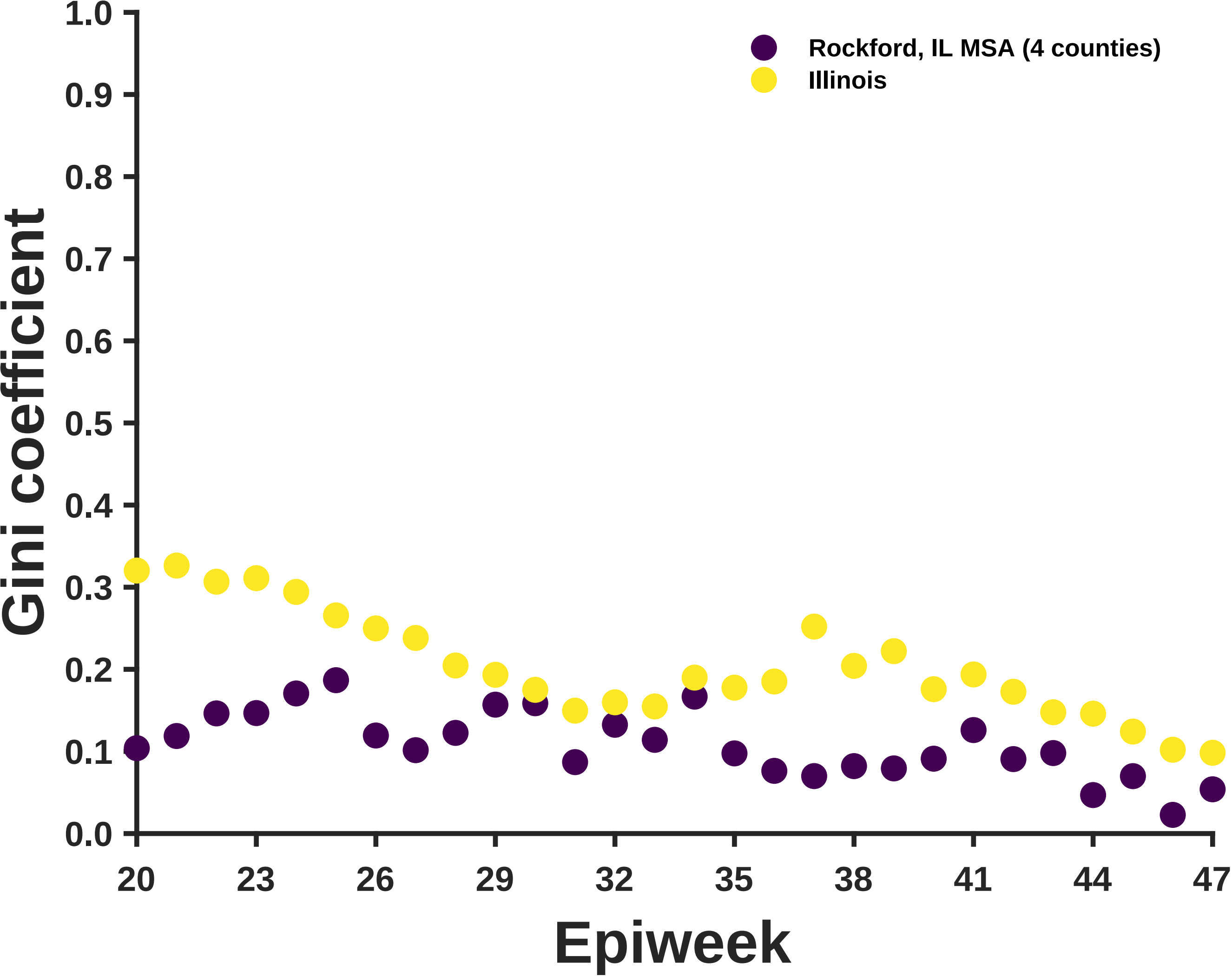

Rockford, IL MSA (4 counties)
Illinois
Gini coefficient
1.0
0.9
0.8
0.7
0.6
0.5
0.4
0.3
0.2
0.1
0.0
20
23
26
29
32
35
38
41
44
47
Epiweek

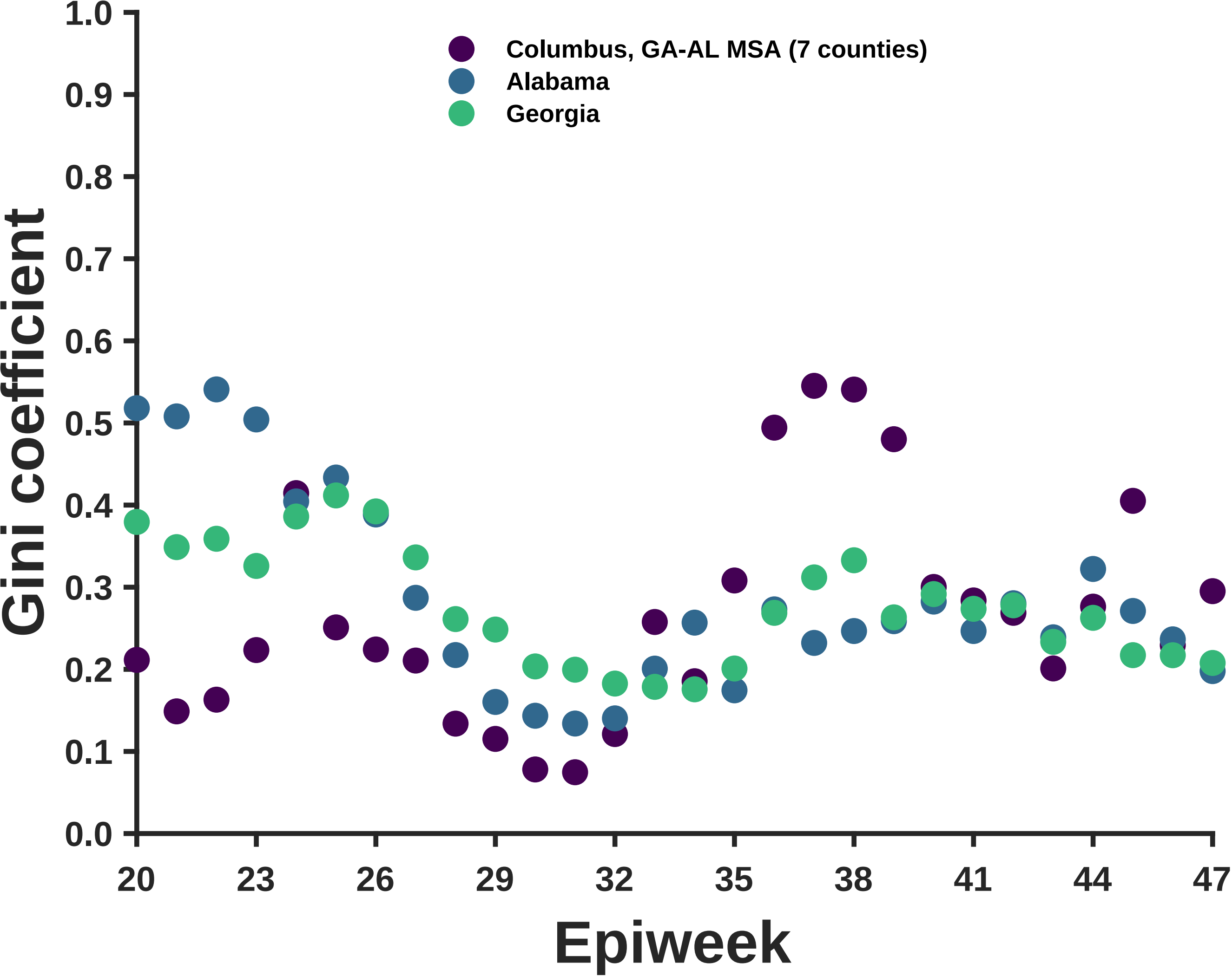

Columbus, GA-AL MSA (7 counties)
Alabama
Georgia
Gini coefficient
1.0
0.9
0.8
0.7
0.6
0.5
0.4
0.3
0.2
0.1
0.0
20
23
26
29
32
35
38
41
44
47
Epiweek

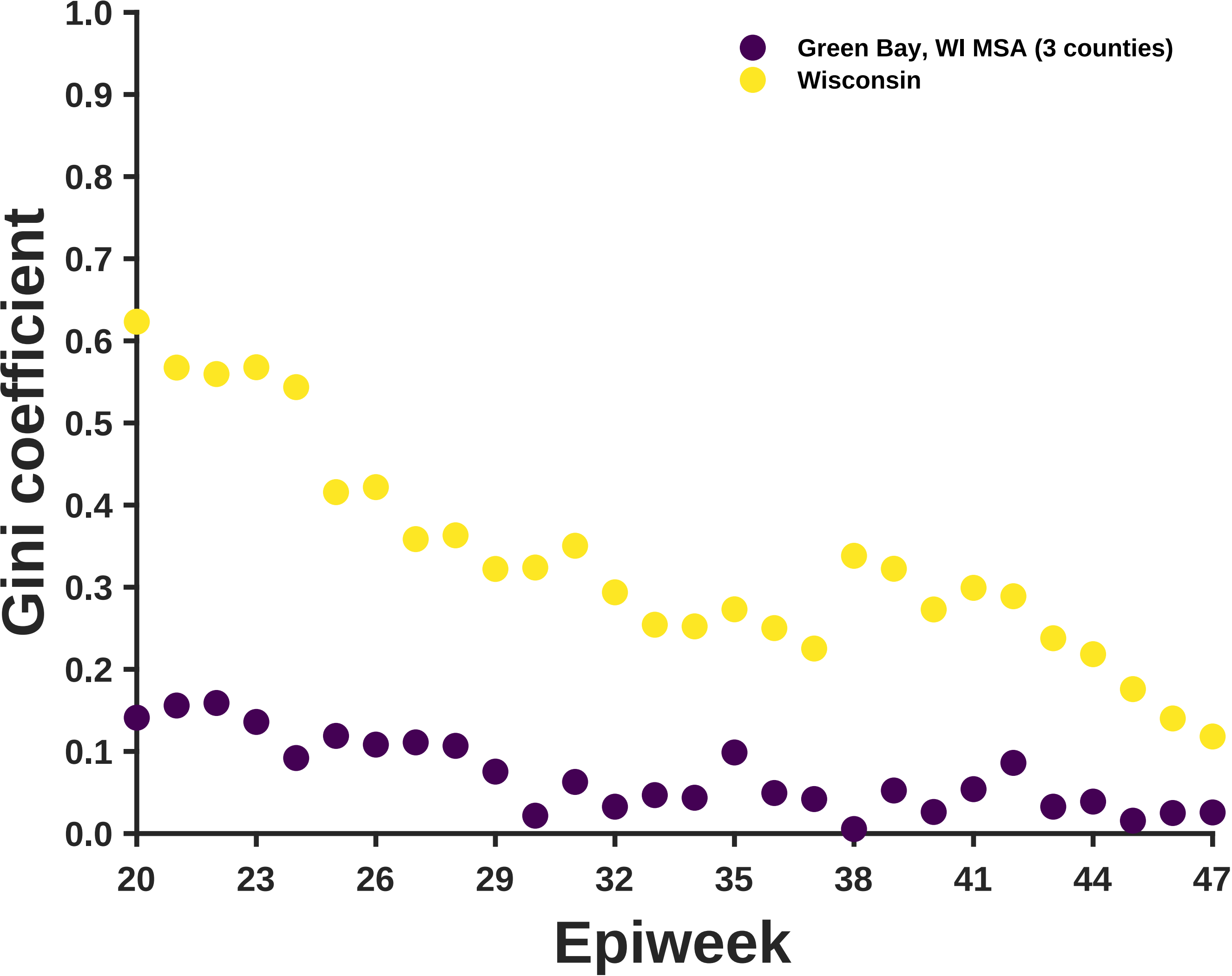
Green Bay, WI MSA (3 counties)
Wisconsin
Gini coefficient
0.0
0.1
0.2
0.3
0.4
0.5
0.6
0.7
0.8
0.9
1.0
20
23
26
29
32
35
38
41
44
47
Epiweek

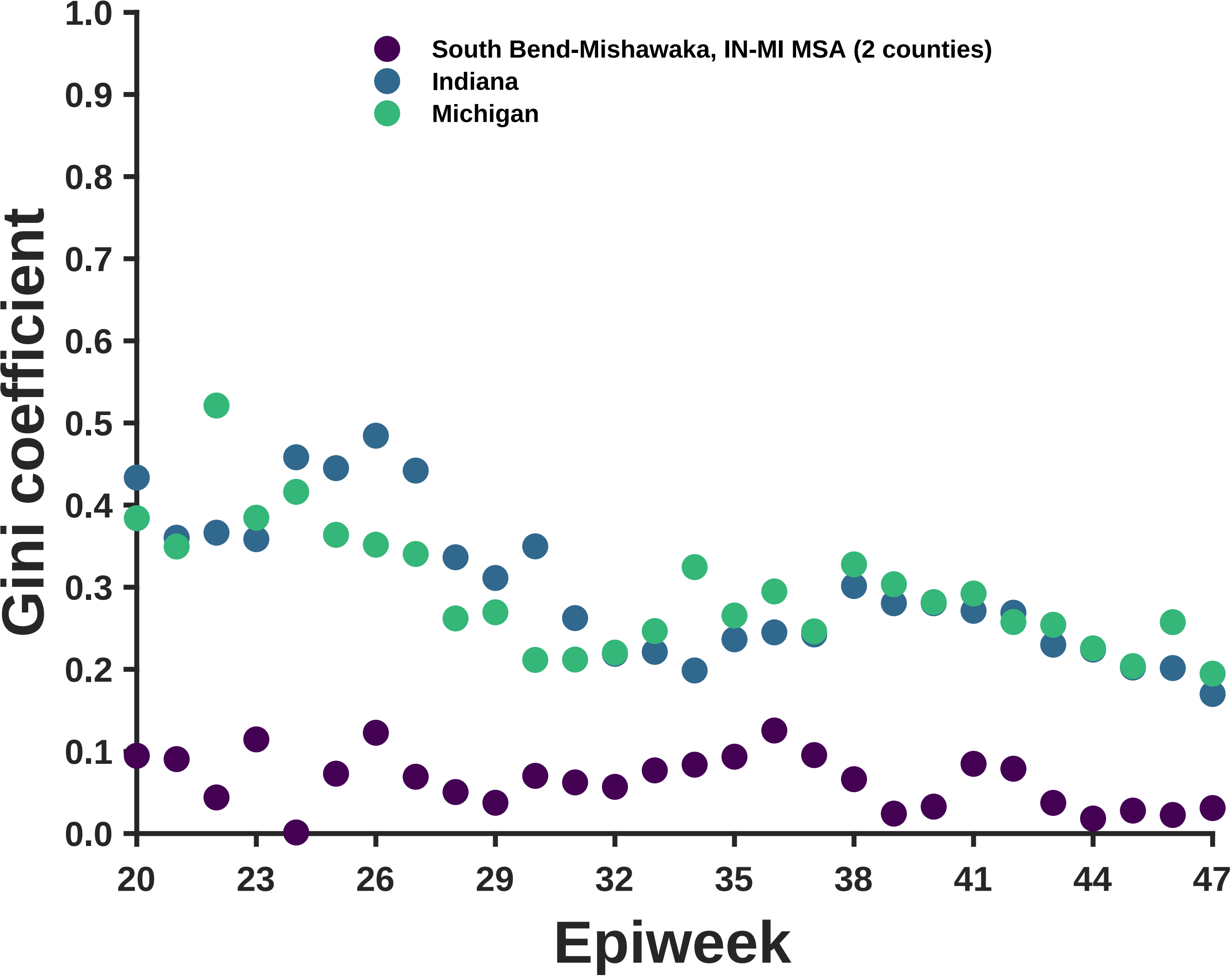

South Bend-Mishawaka, IN-MI MSA (2 counties)
Indiana
Michigan
Gini coefficient
Epiweek
1.0
0.9
0.8
0.7
0.6
0.5
0.4
0.3
0.2
0.1
0.0
20
23
26
29
32
35
38
41
44
47

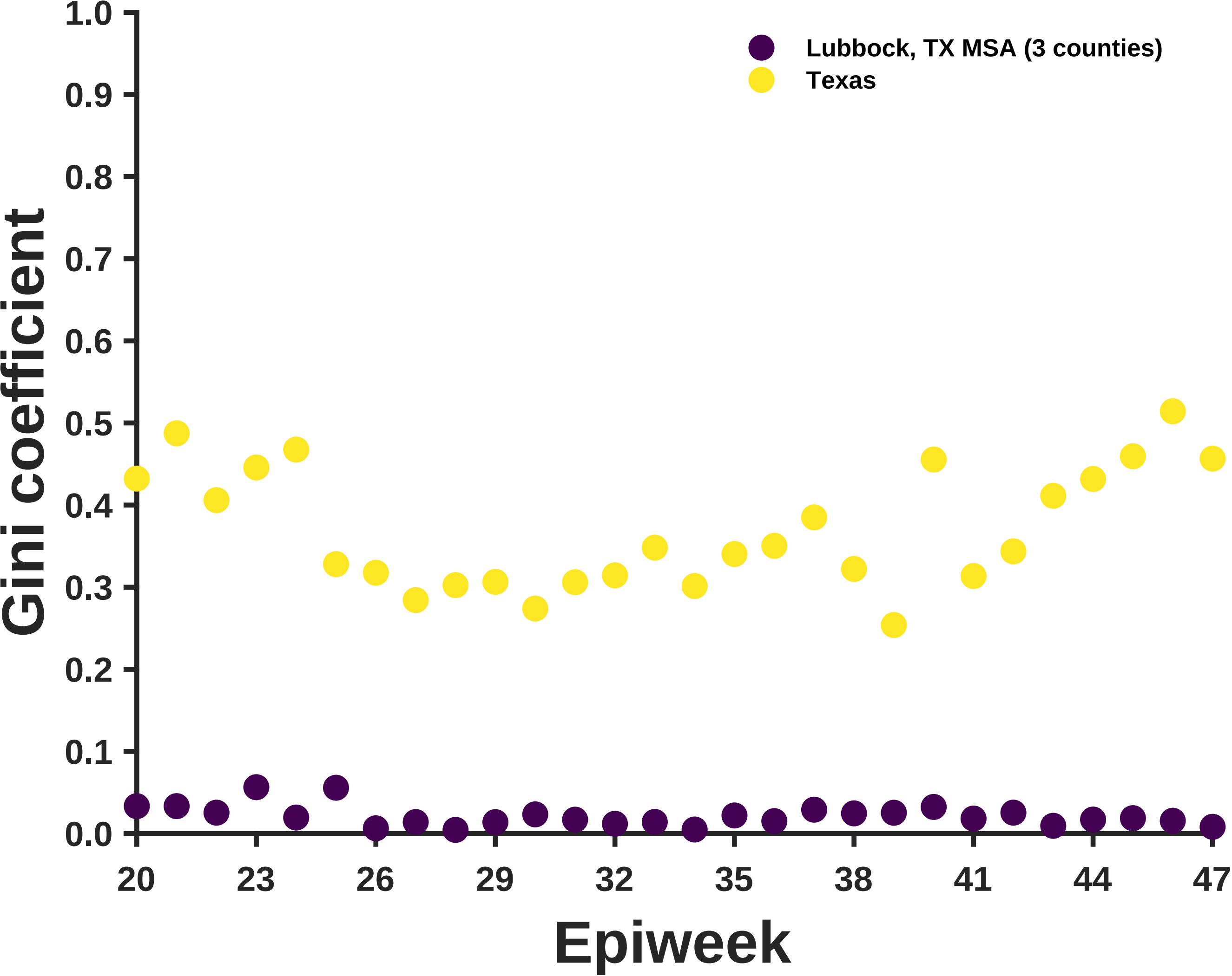

Lubbock, TX MSA (3 counties)
Texas
Gini coefficient
1.0
0.9
0.8
0.7
0.6
0.5
0.4
0.3
0.2
0.1
0.0
20
23
26
29
32
35
38
41
44
47
Epiweek

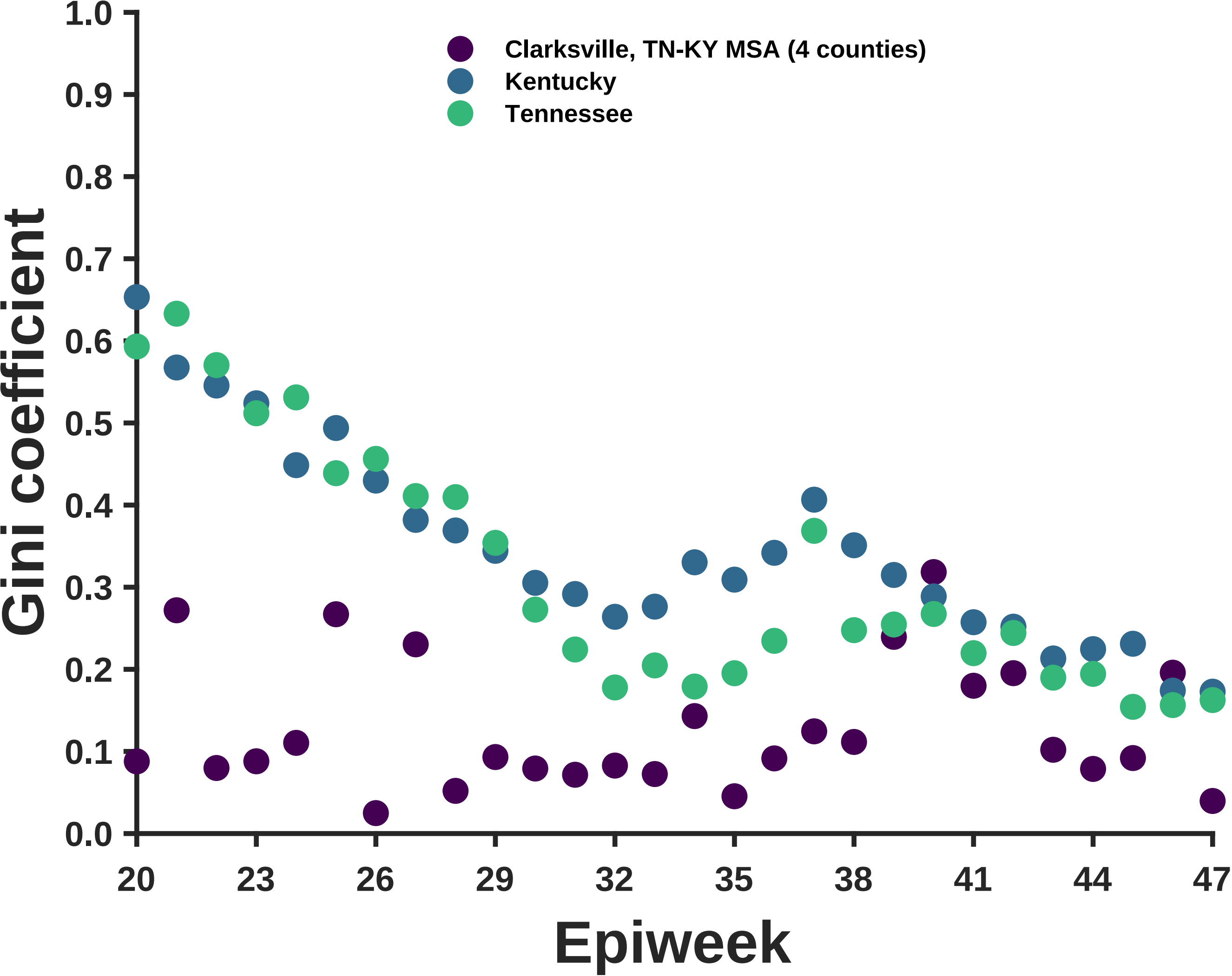
Clarksville, TN-KY MSA (4 counties)
Kentucky
Tennessee
Gini coefficient
0.0
0.1
0.2
0.3
0.4
0.5
0.6
0.7
0.8
0.9
1.0
20
23
26
29
32
35
38
41
44
47
Epiweek

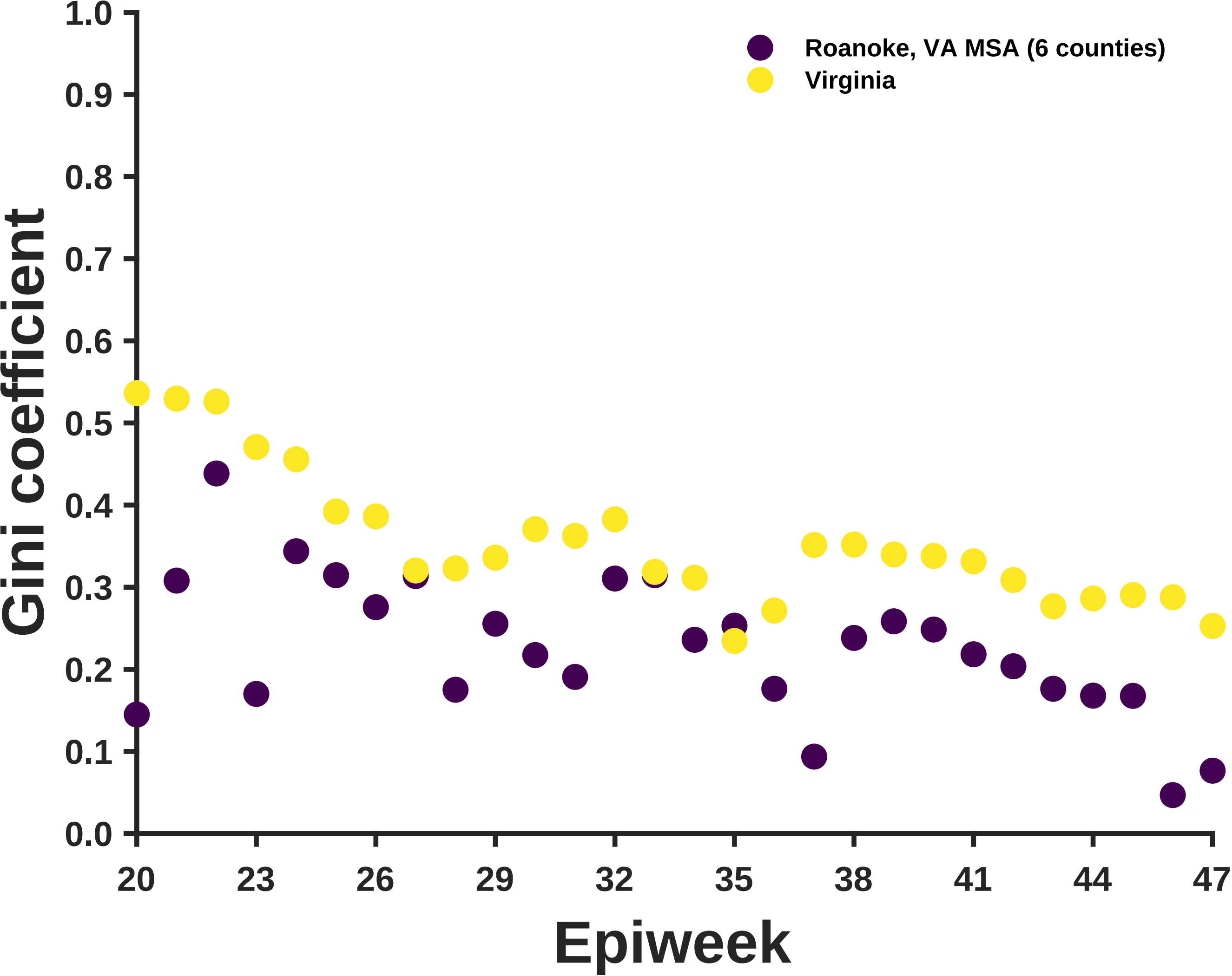
Roanoke, VA MSA (6 counties)
Virginia
Gini coefficient
1.0
0.9
0.8
0.7
0.6
0.5
0.4
0.3
0.2
0.1
0.0
20
23
26
29
32
35
38
41
44
47
Epiweek

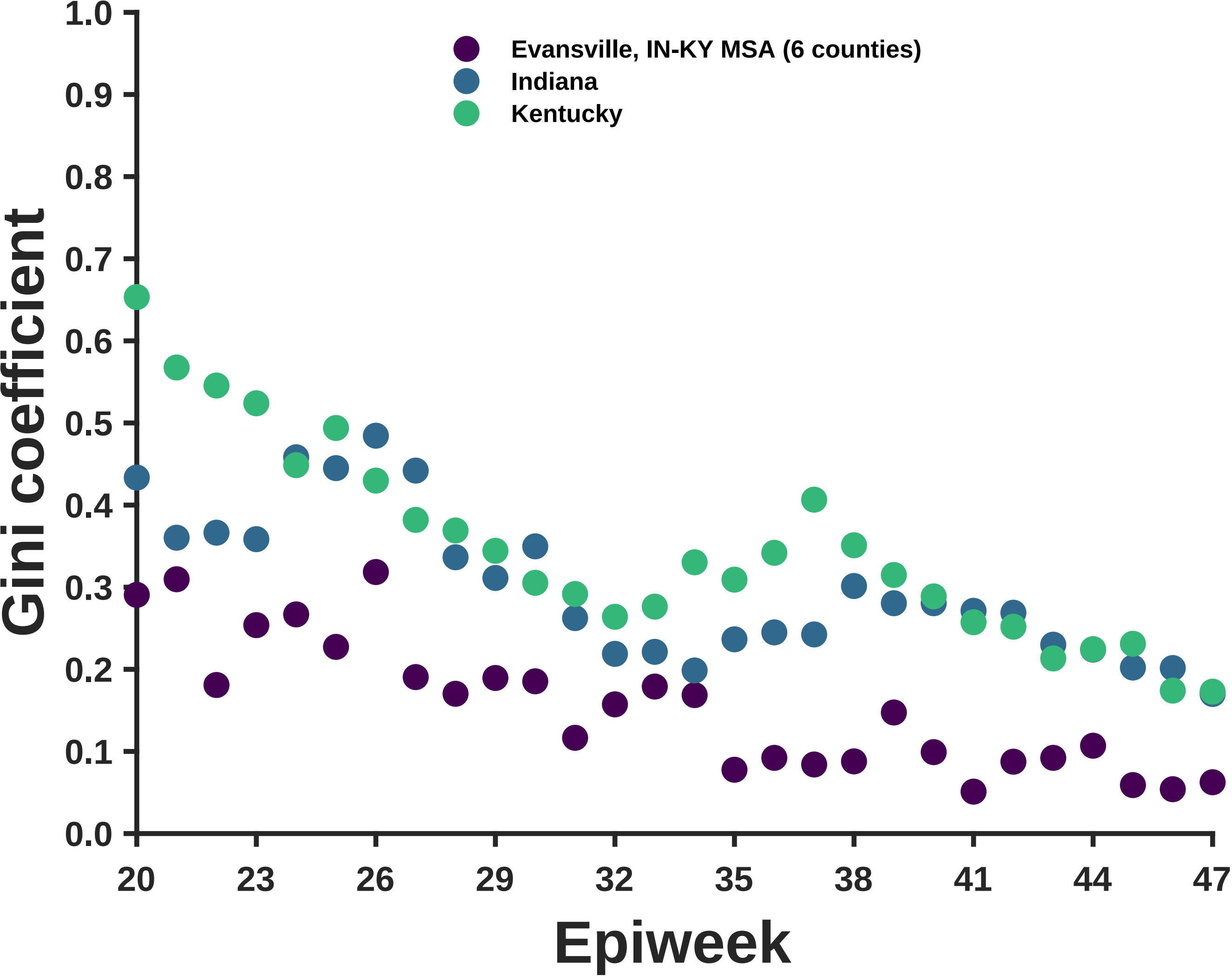

Evansville, IN-KY MSA (6 counties)
Indiana
Kentucky
Gini coefficient
1.0
0.9
0.8
0.7
0.6
0.5
0.4
0.3
0.2
0.1
0.0
20
23
26
29
32
35
38
41
44
47
Epiweek

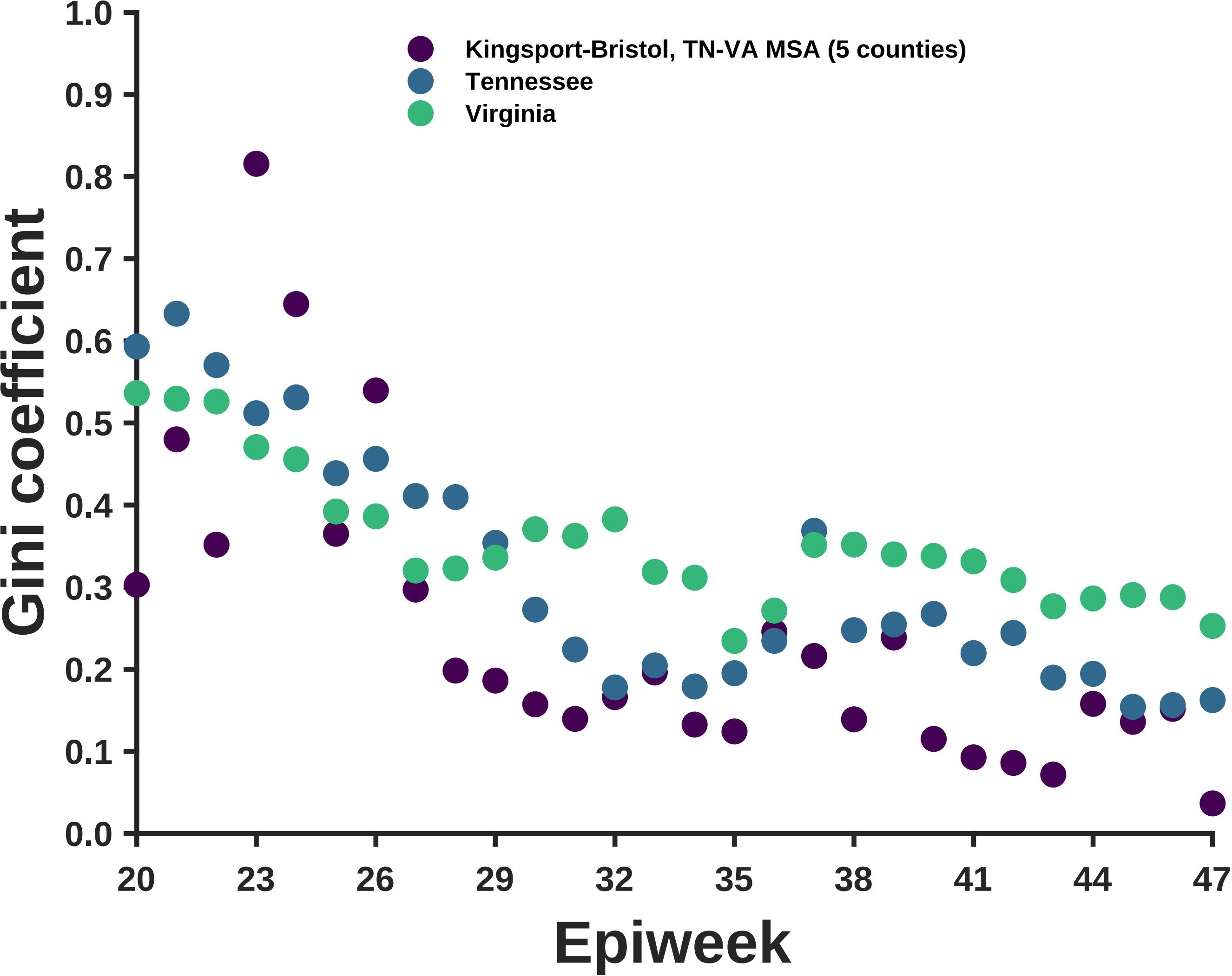

Kingsport-Bristol, TN-VA MSA (5 counties)
Tennessee
Virginia
Gini coefficient
1.0
0.9
0.8
0.7
0.6
0.5
0.4
0.3
0.2
0.1
0.0
20
23
26
29
32
35
38
41
44
47
Epiweek

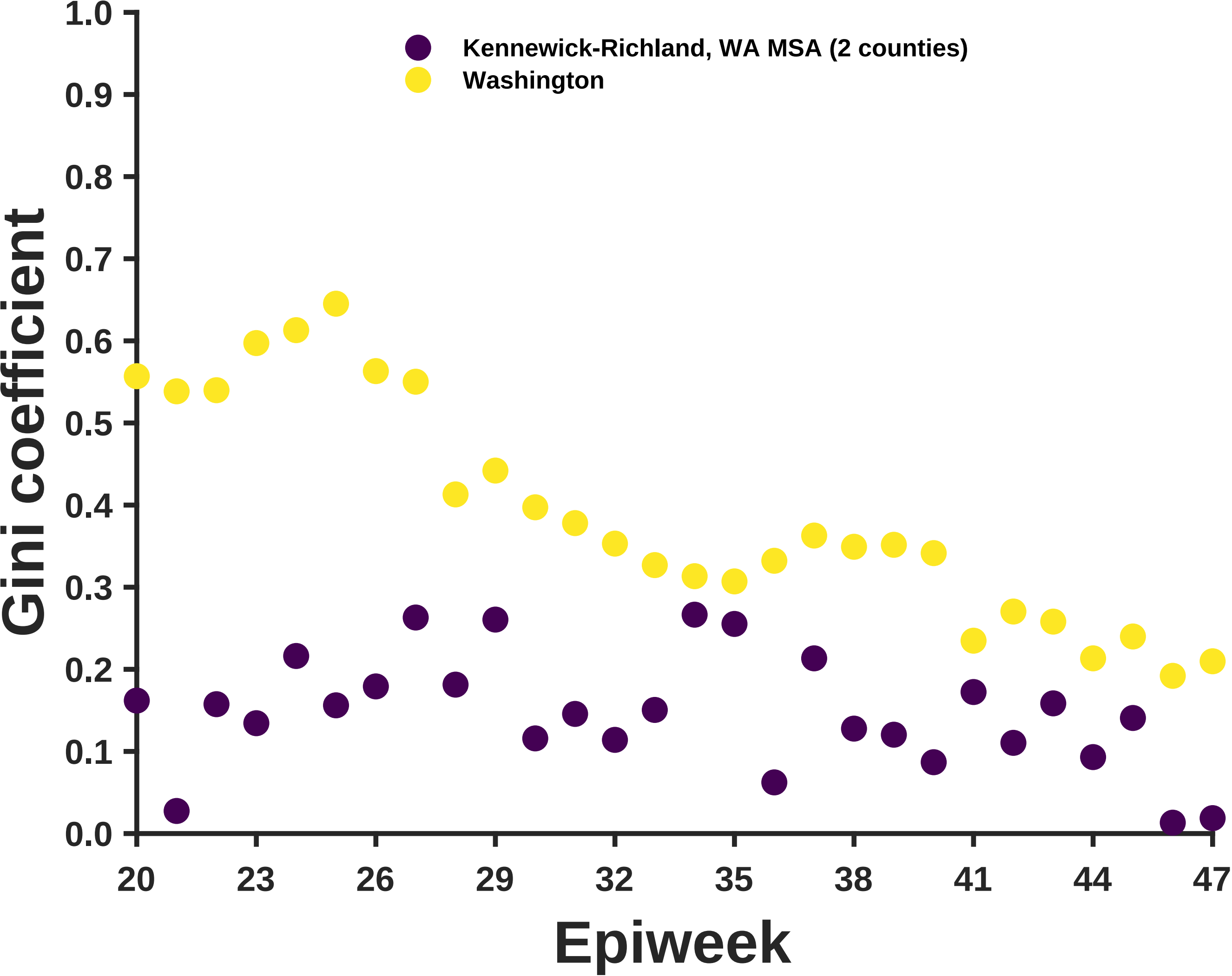

Gini coefficient
1.0
0.9
0.8
0.7
0.6
0.5
0.4
0.3
0.2
0.1
0.0
Kennewick-Richland, WA MSA (2 counties)
Washington
20
23
26
29
32
35
38
41
44
47
Epiweek

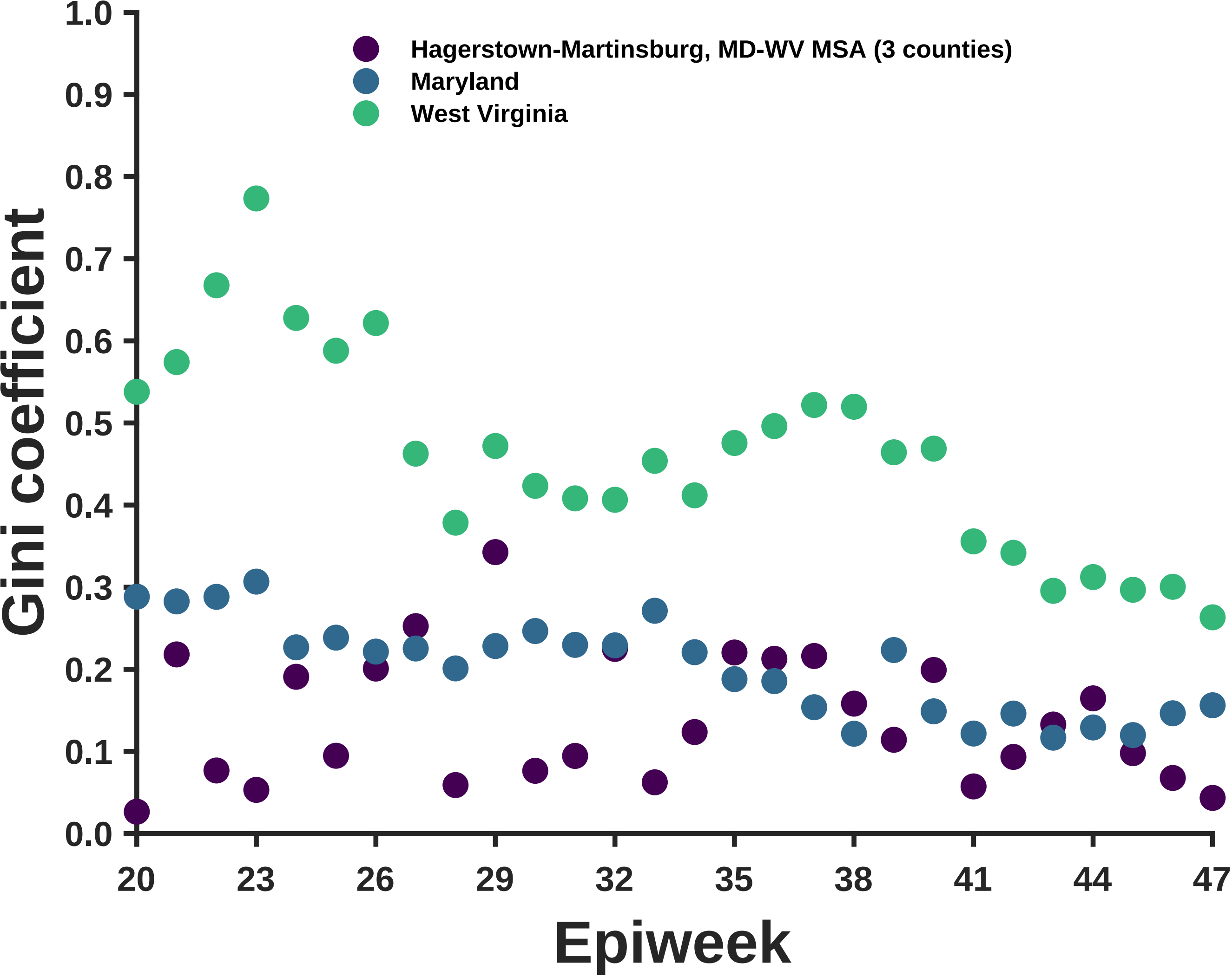
Hagerstown-Martinsburg, MD-WV MSA (3 counties)
Maryland
West Virginia
Gini coefficient
Epiweek
0.0
0.1
0.2
0.3
0.4
0.5
0.6
0.7
0.8
0.9
1.0
20
23
26
29
32
35
38
41
44
47

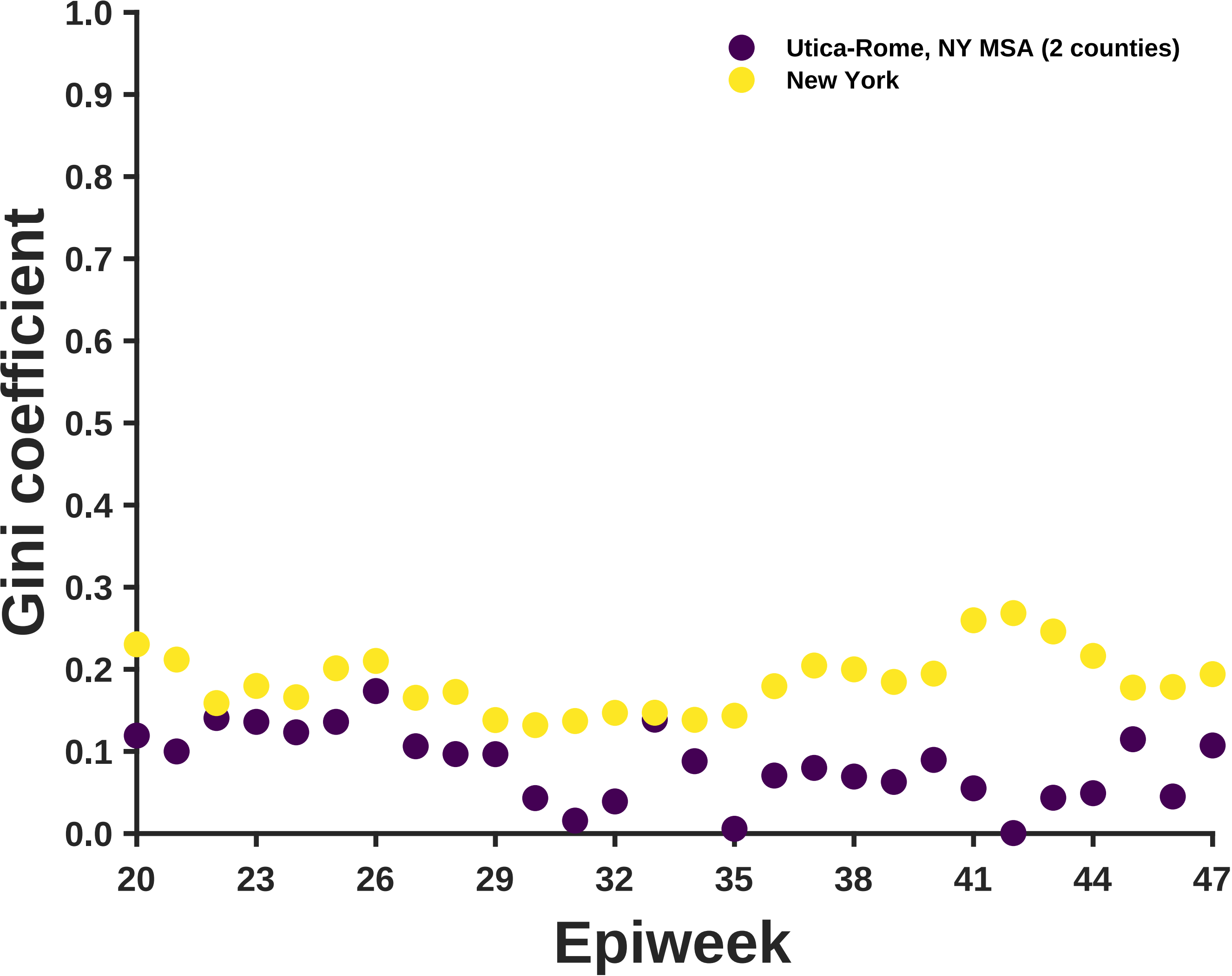

Utica-Rome, NY MSA (2 counties)
New York
Gini coefficient
1.0
0.9
0.8
0.7
0.6
0.5
0.4
0.3
0.2
0.1
0.0
20
23
26
29
32
35
38
41
44
47
Epiweek

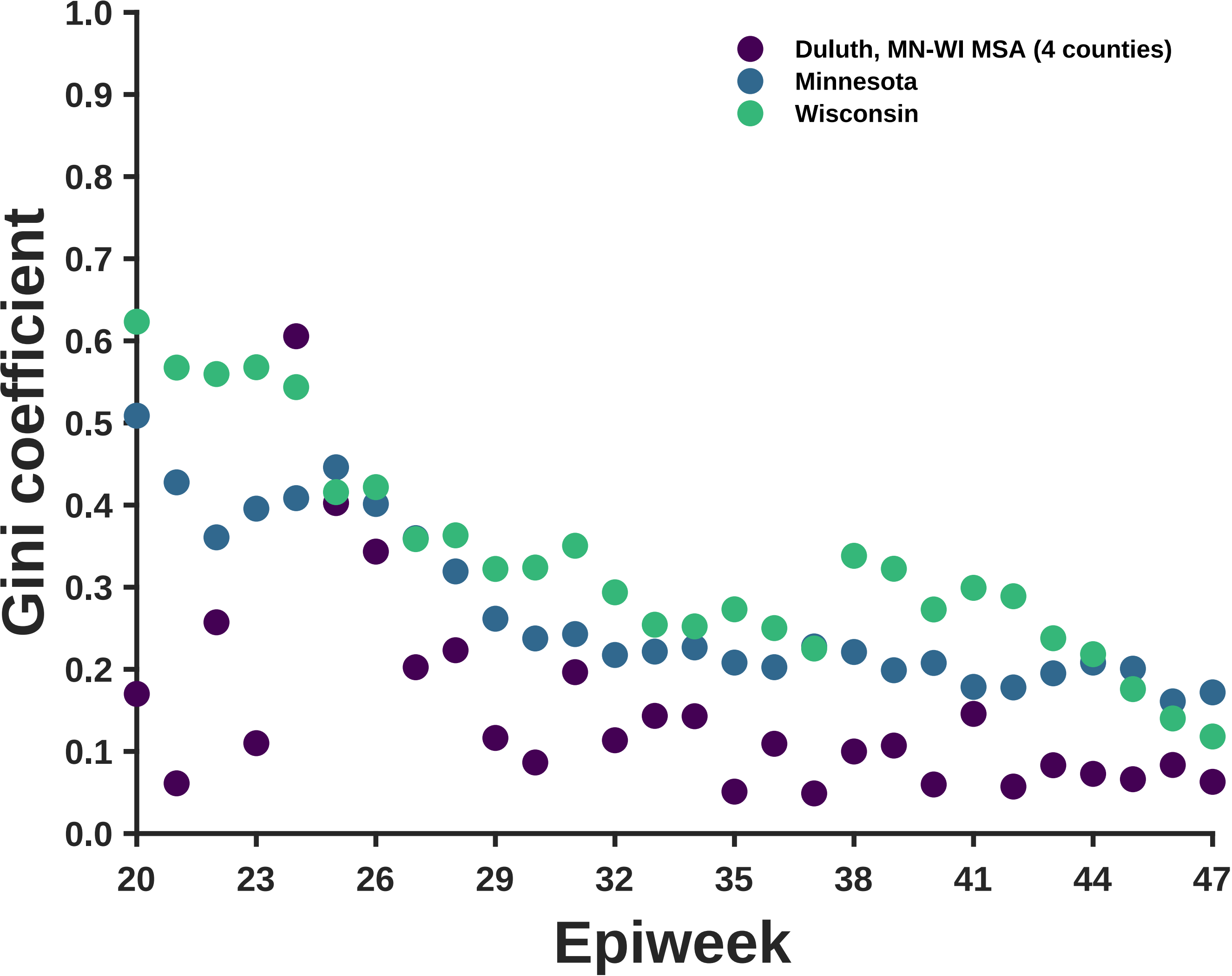

Duluth, MN-WI MSA (4 counties)
Minnesota
Wisconsin
Gini coefficient
Epiweek
1.0
0.9
0.8
0.7
0.6
0.5
0.4
0.3
0.2
0.1
0.0
20
23
26
29
32
35
38
41
44
47

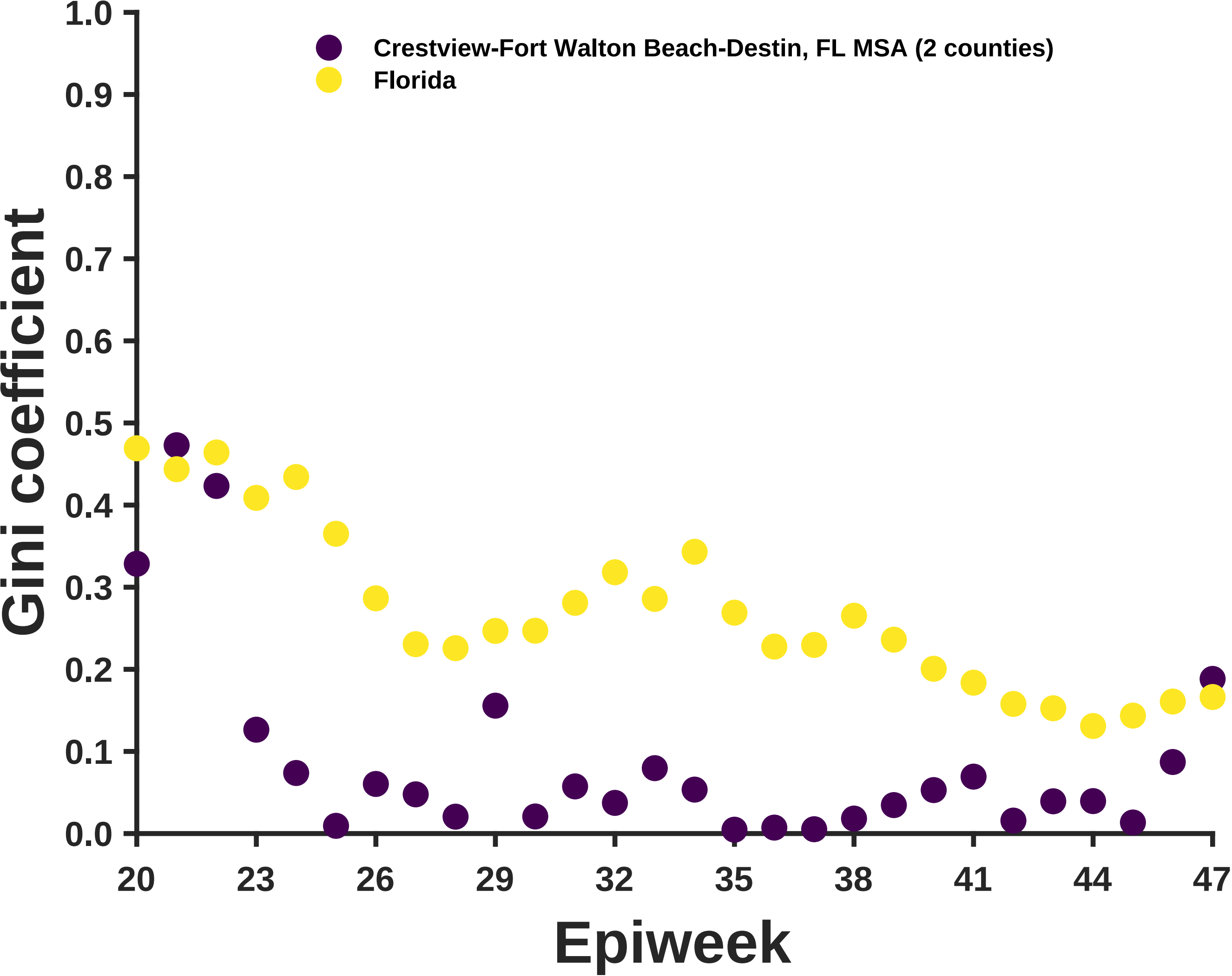

Crestview-Fort Walton Beach-Destin, FL MSA (2 counties)
Florida
Gini coefficient
0.0
0.1
0.2
0.3
0.4
0.5
0.6
0.7
0.8
0.9
1.0
20
23
26
29
32
35
38
41
44
47
Epiweek

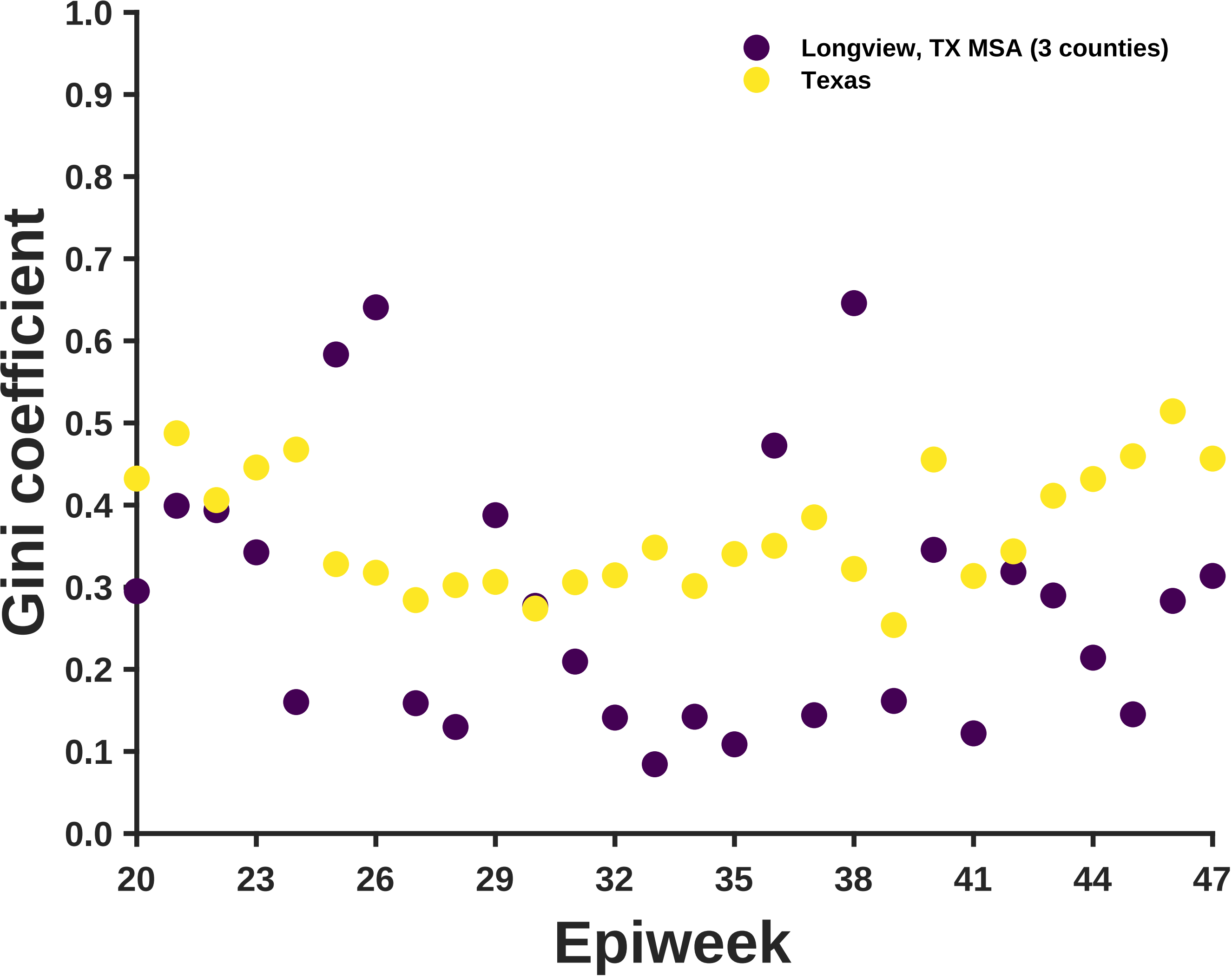

Longview, TX MSA (3 counties)
Texas
Gini coefficient
1.0
0.9
0.8
0.7
0.6
0.5
0.4
0.3
0.2
0.1
0.0
20
23
26
29
32
35
38
41
44
47
Epiweek

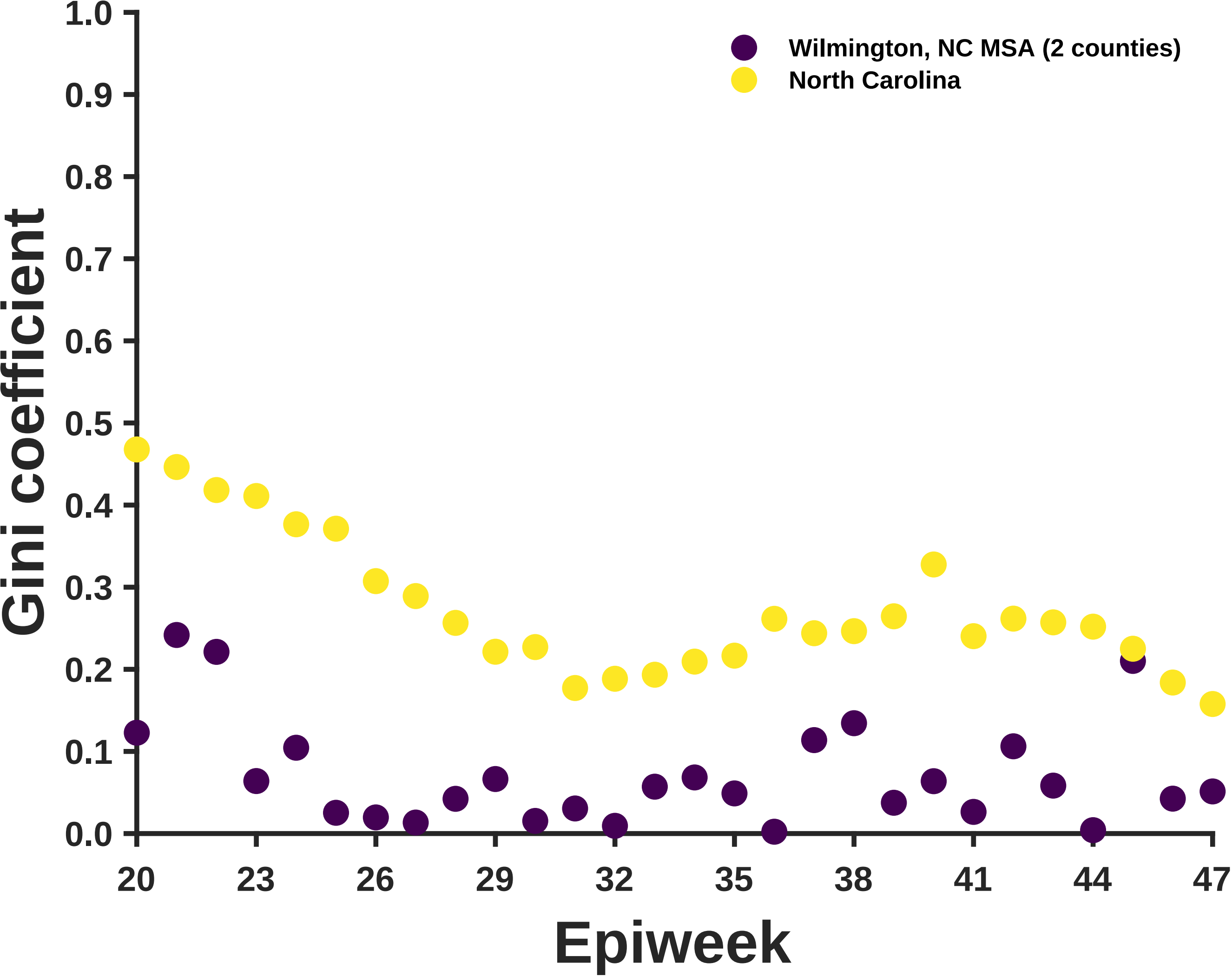

Wilmington, NC MSA (2 counties)
North Carolina
Gini coefficient
1.0
0.9
0.8
0.7
0.6
0.5
0.4
0.3
0.2
0.1
0.0
20
23
26
29
32
35
38
41
44
47
Epiweek

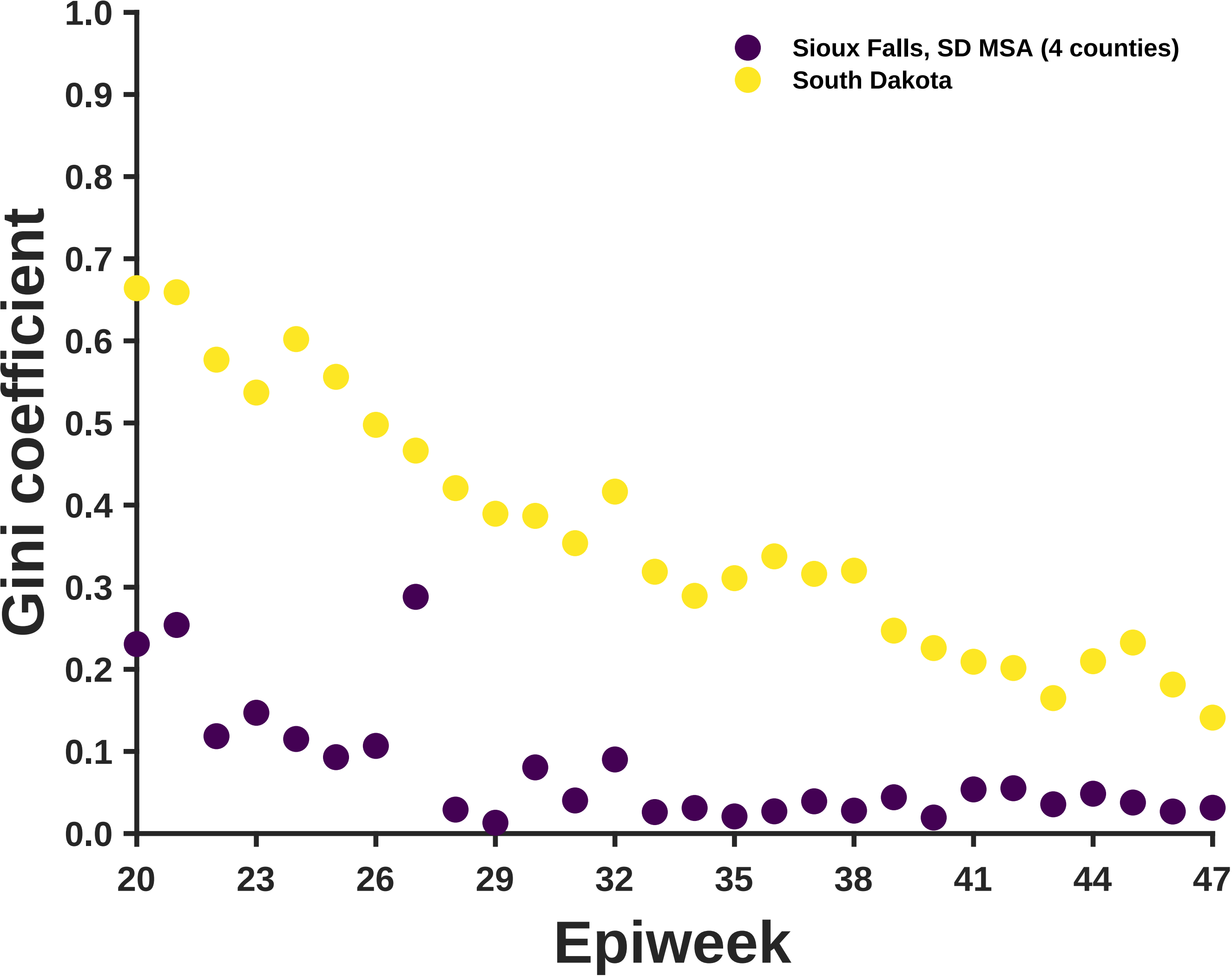

Sioux Falls, SD MSA (4 counties)
South Dakota
Gini coefficient
1.0
0.9
0.8
0.7
0.6
0.5
0.4
0.3
0.2
0.1
0.0
20
23
26
29
32
35
38
41
44
47
Epiweek

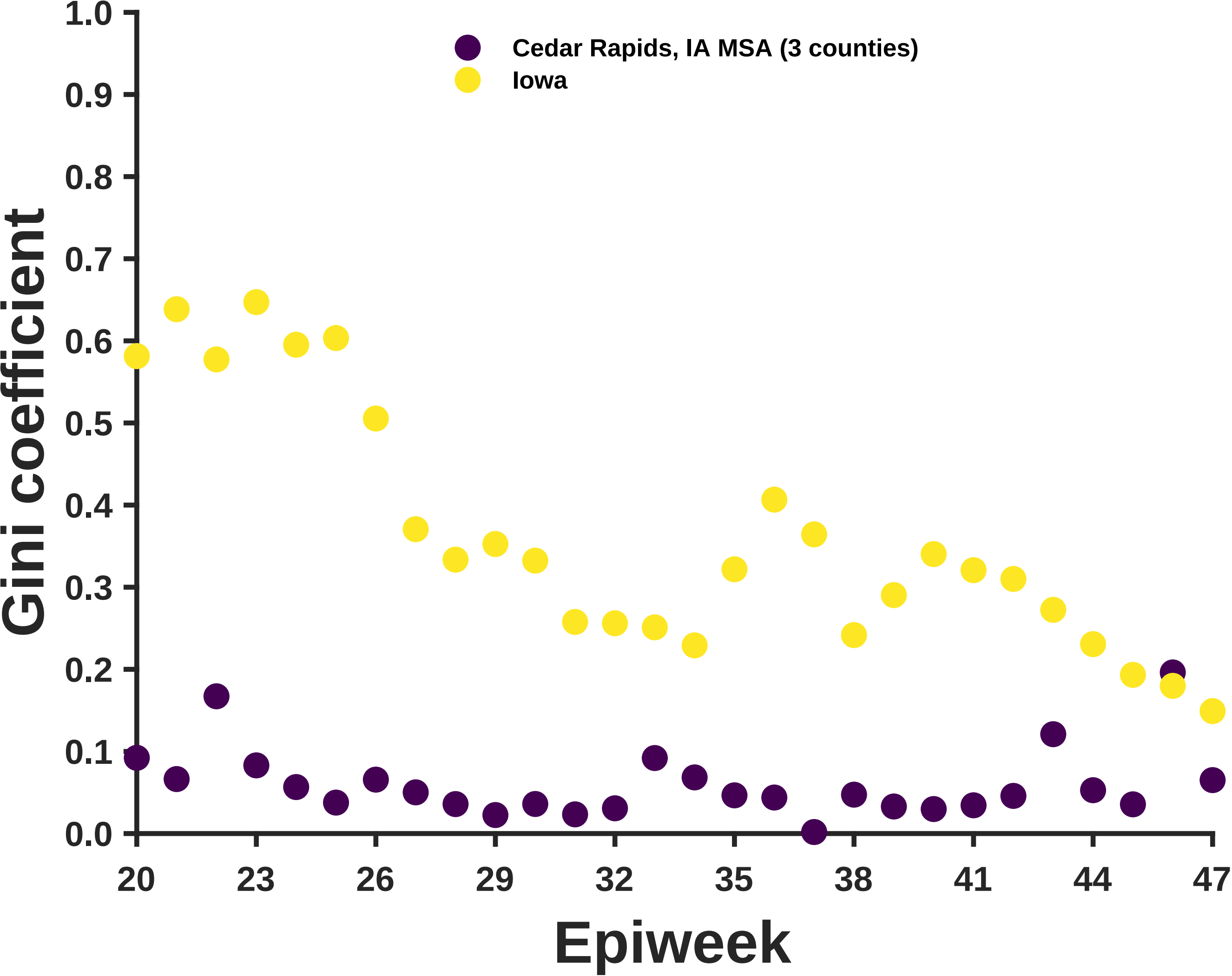
Cedar Rapids, IA MSA (3 counties)
Iowa
Gini coefficient
Epiweek
1.0
0.9
0.8
0.7
0.6
0.5
0.4
0.3
0.2
0.1
0.0
20
23
26
29
32
35
38
41
44
47

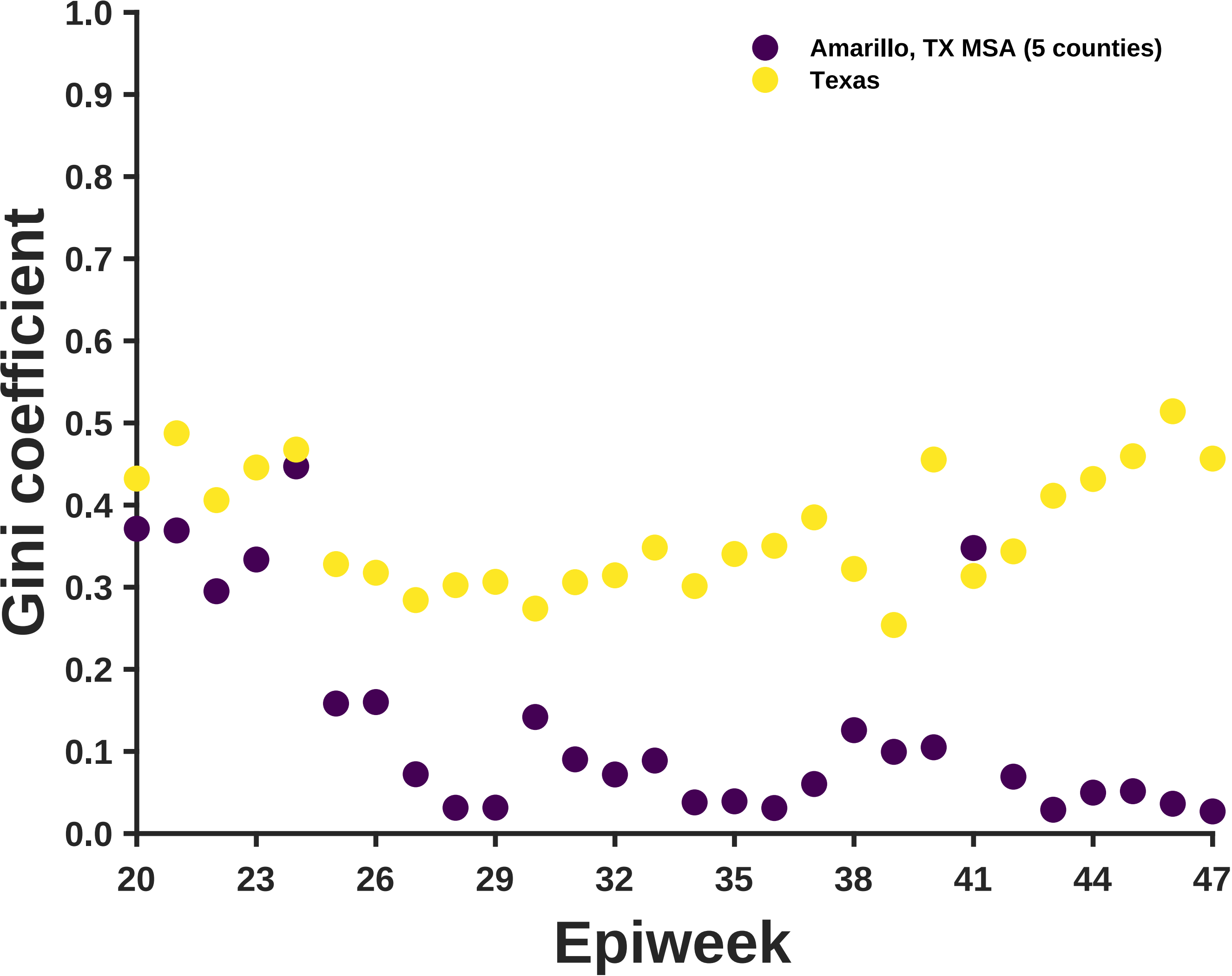

Amarillo, TX MSA (5 counties)
Texas
Gini coefficient
1.0
0.9
0.8
0.7
0.6
0.5
0.4
0.3
0.2
0.1
0.0
20
23
26
29
32
35
38
41
44
47
Epiweek

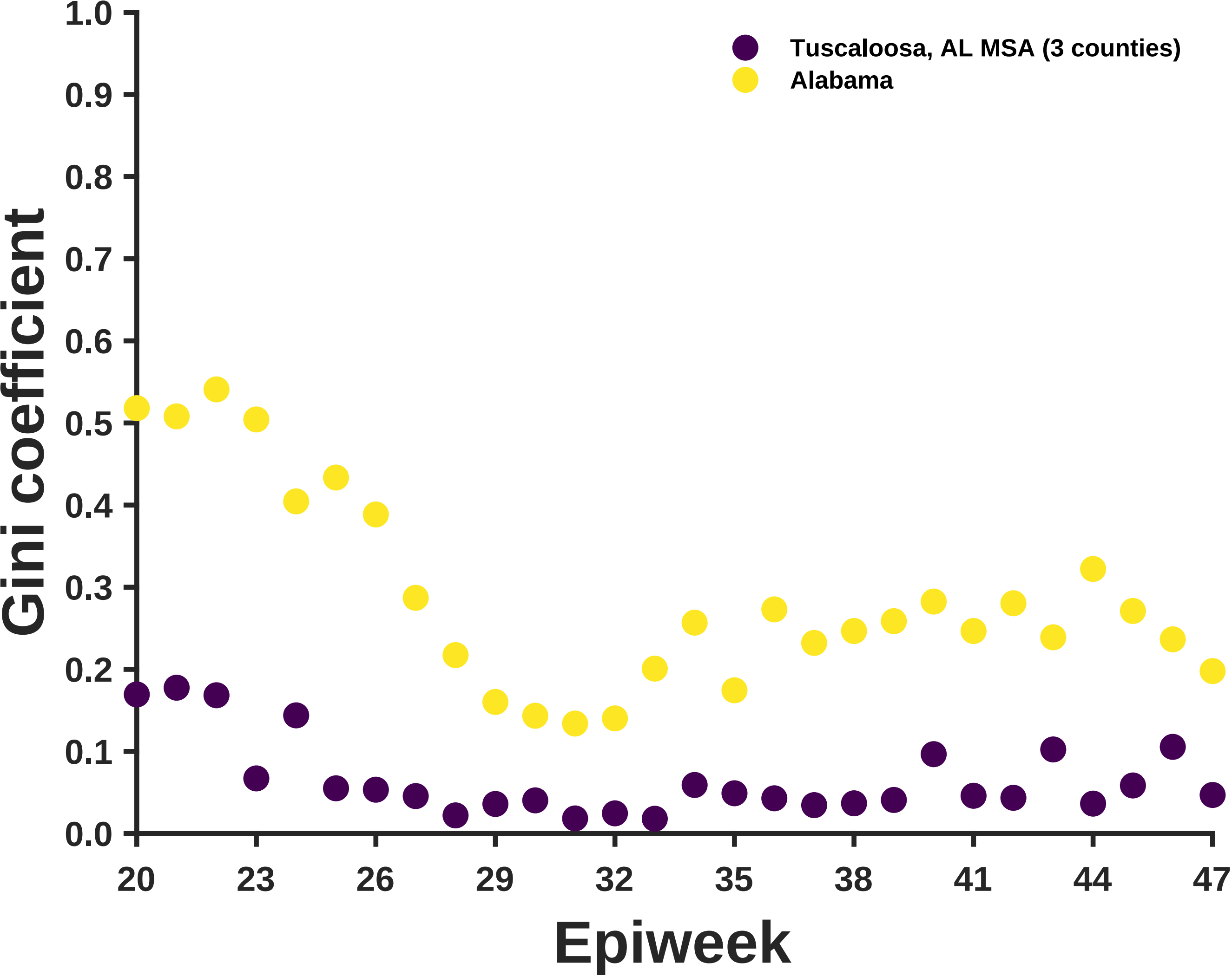

Tuscaloosa, AL MSA (3 counties)
Alabama
Gini coefficient
Epiweek
1.0
0.9
0.8
0.7
0.6
0.5
0.4
0.3
0.2
0.1
0.0
20
23
26
29
32
35
38
41
44
47

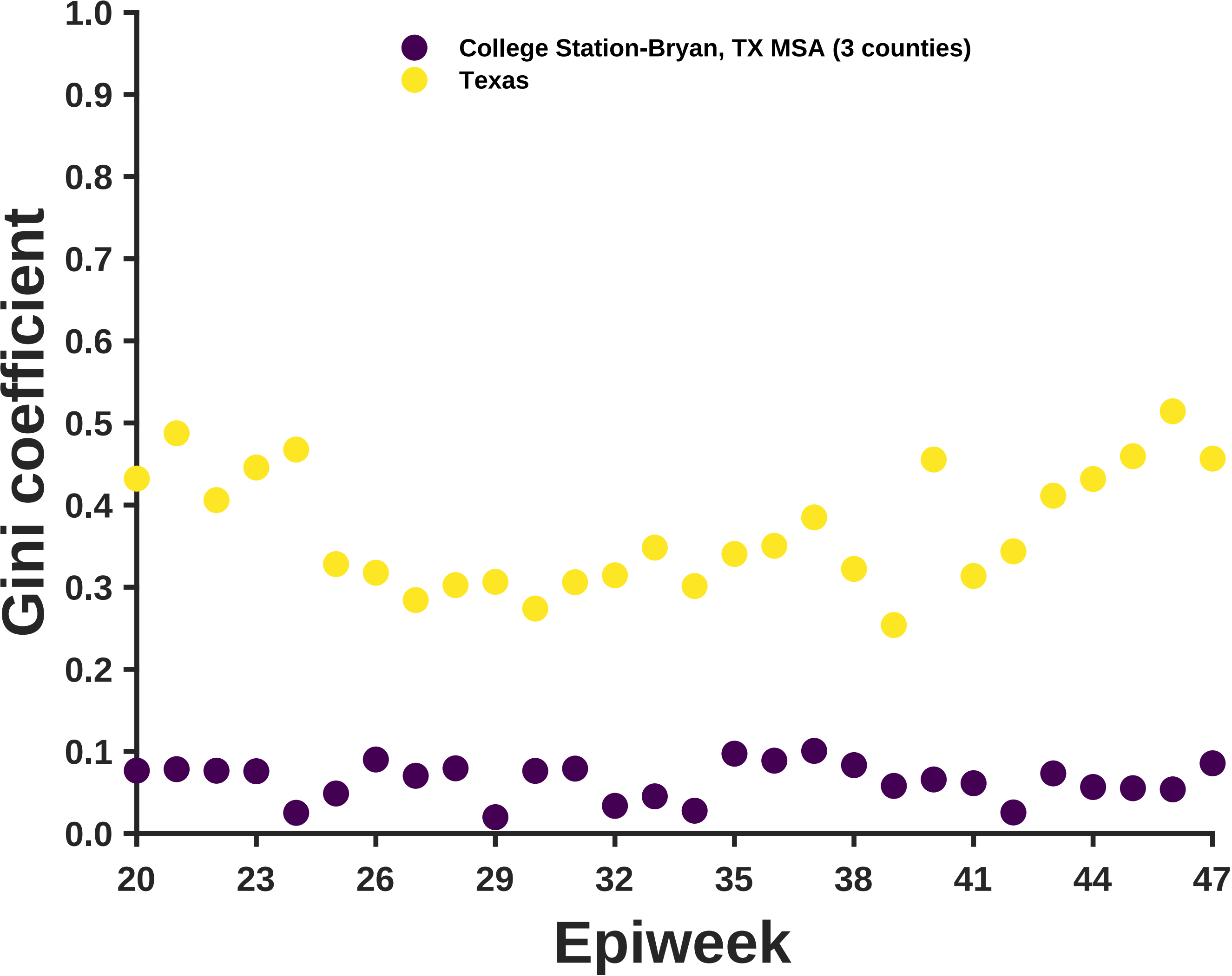

College Station-Bryan, TX MSA (3 counties)
Texas
Gini coefficient
1.0
0.9
0.8
0.7
0.6
0.5
0.4
0.3
0.2
0.1
0.0
20
23
26
29
32
35
38
41
44
47
Epiweek

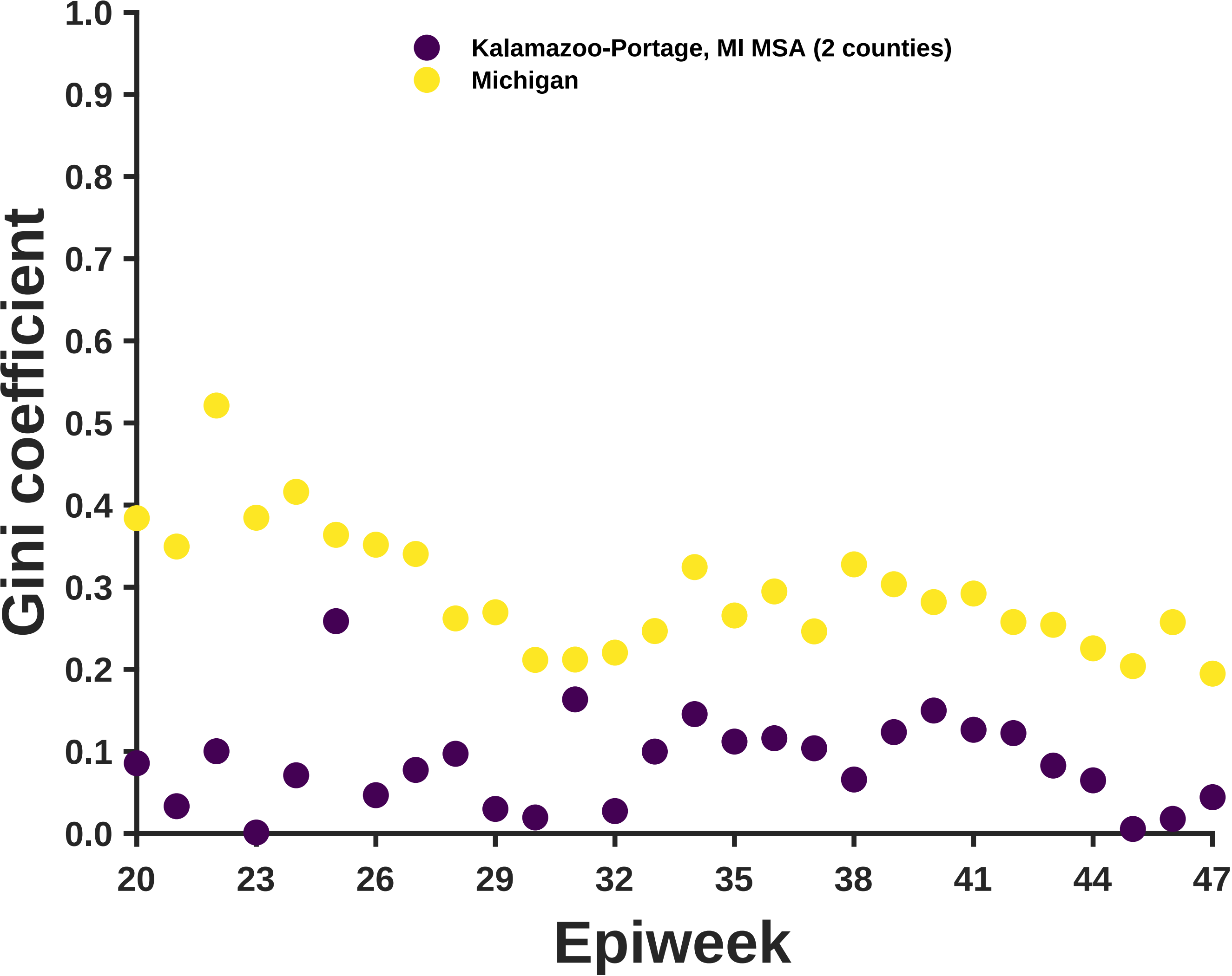
Kalamazoo-Portage, MI MSA (2 counties)
Michigan
Gini coefficient
1.0
0.9
0.8
0.7
0.6
0.5
0.4
0.3
0.2
0.1
0.0
20
23
26
29
32
35
38
41
44
47
Epiweek

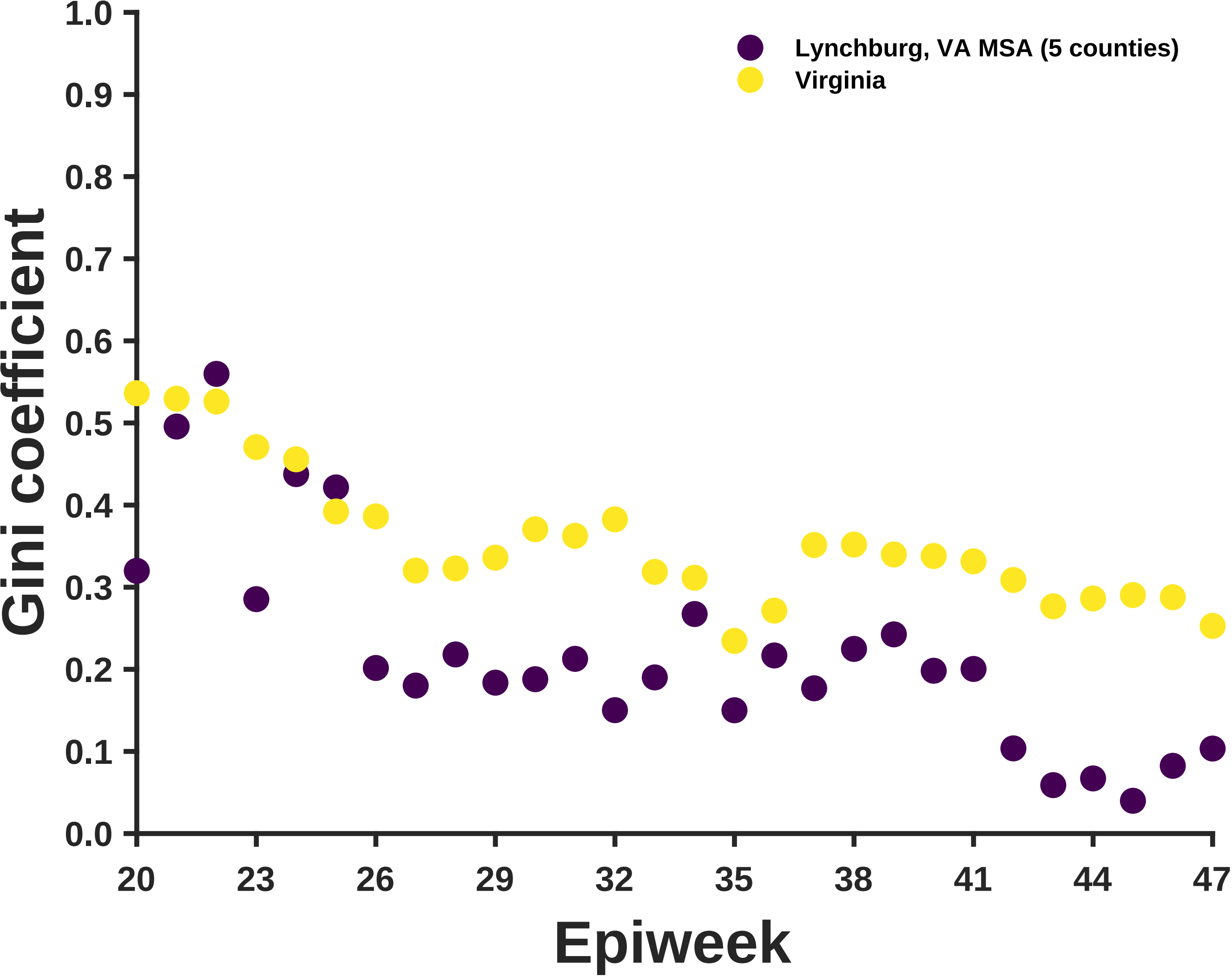

Lynchburg, VA MSA (5 counties)
Virginia
Gini coefficient
Epiweek
1.0
0.9
0.8
0.7
0.6
0.5
0.4
0.3
0.2
0.1
0.0
20
23
26
29
32
35
38
41
44
47

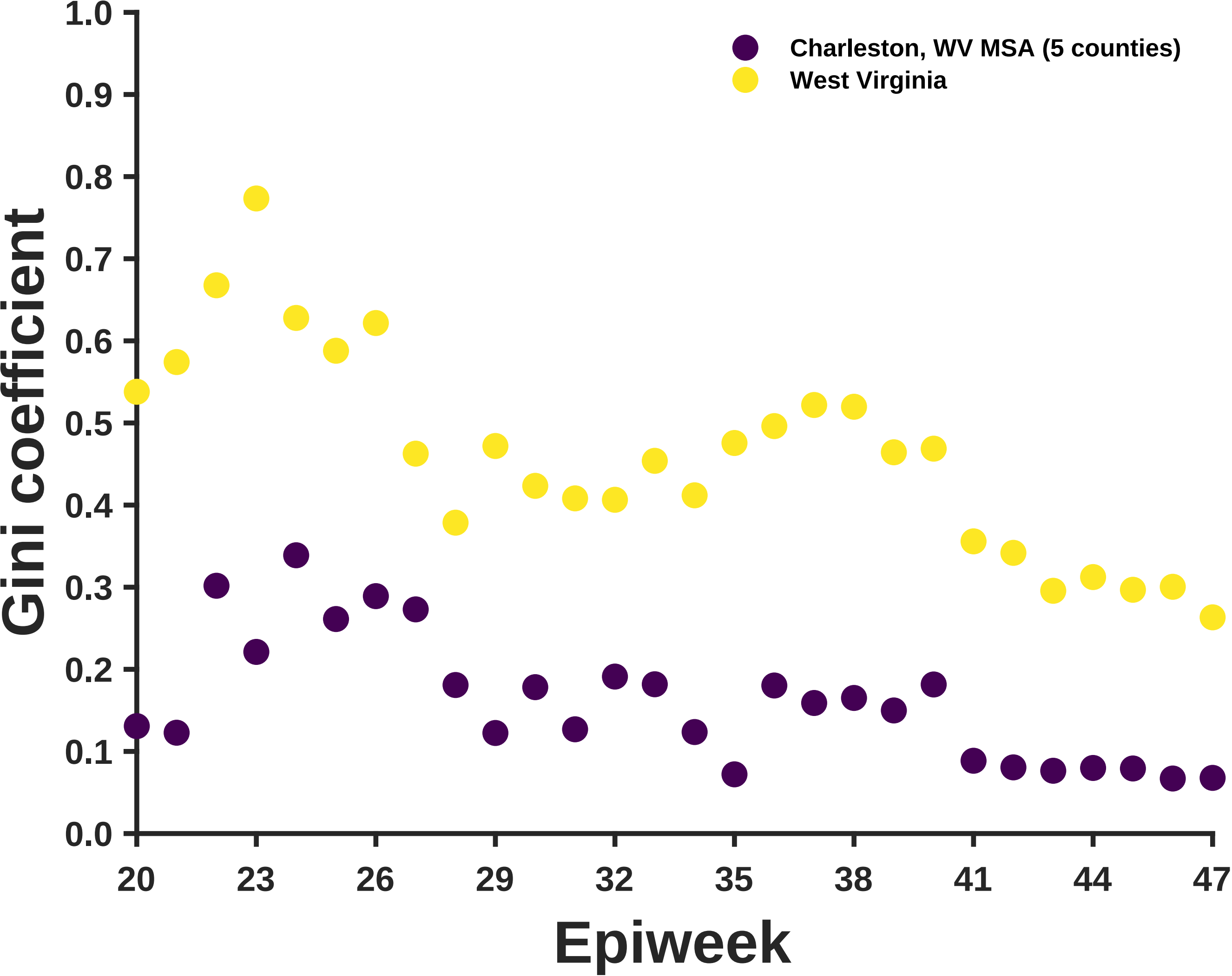

Charleston, WV MSA (5 counties)
West Virginia
Gini coefficient
Epiweek
1.0
0.9
0.8
0.7
0.6
0.5
0.4
0.3
0.2
0.1
0.0
20
23
26
29
32
35
38
41
44
47

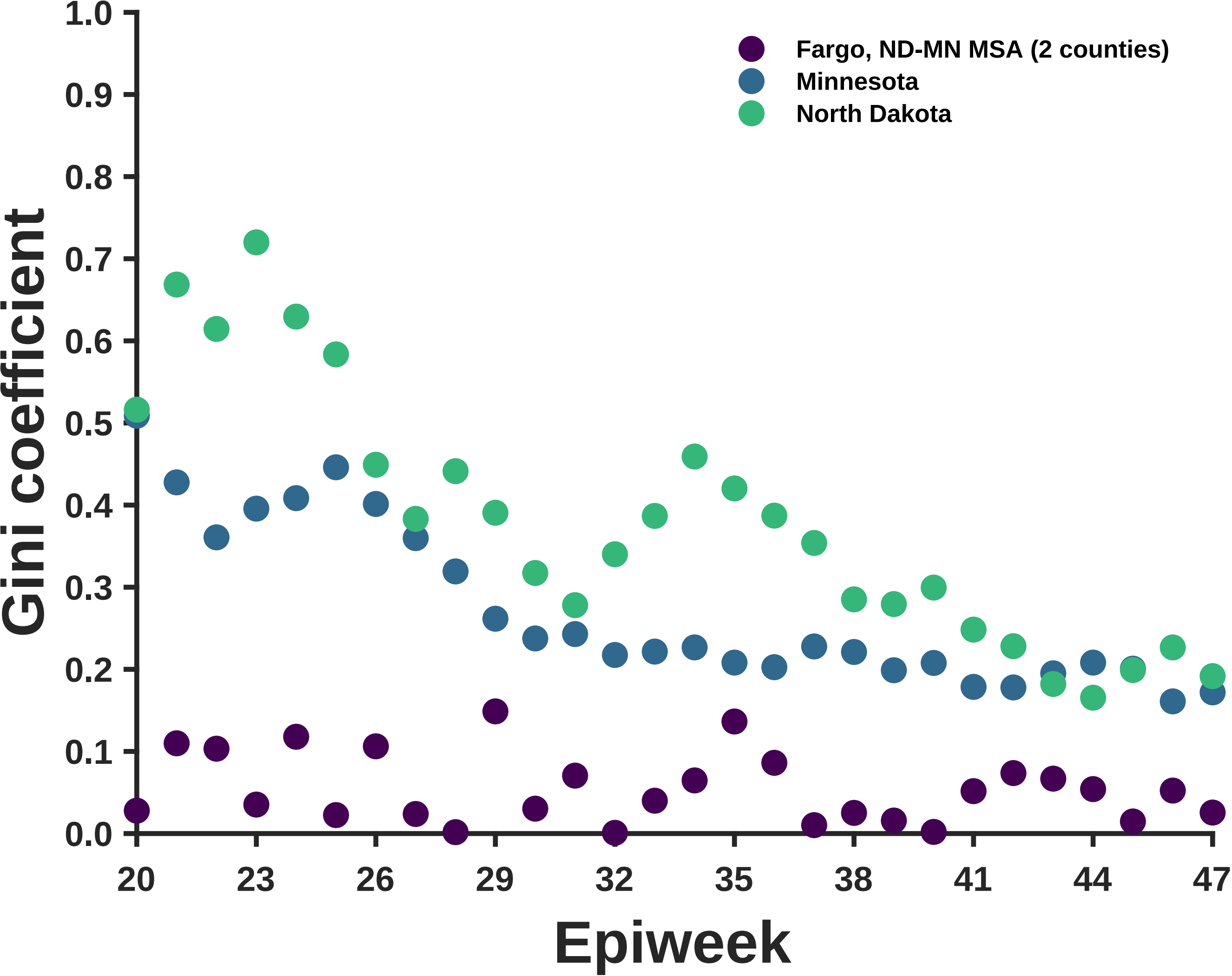
Fargo, ND-MN MSA (2 counties)
Minnesota
North Dakota
Gini coefficient
Epiweek
0.0
0.1
0.2
0.3
0.4
0.5
0.6
0.7
0.8
0.9
1.0
20
23
26
29
32
35
38
41
44
47

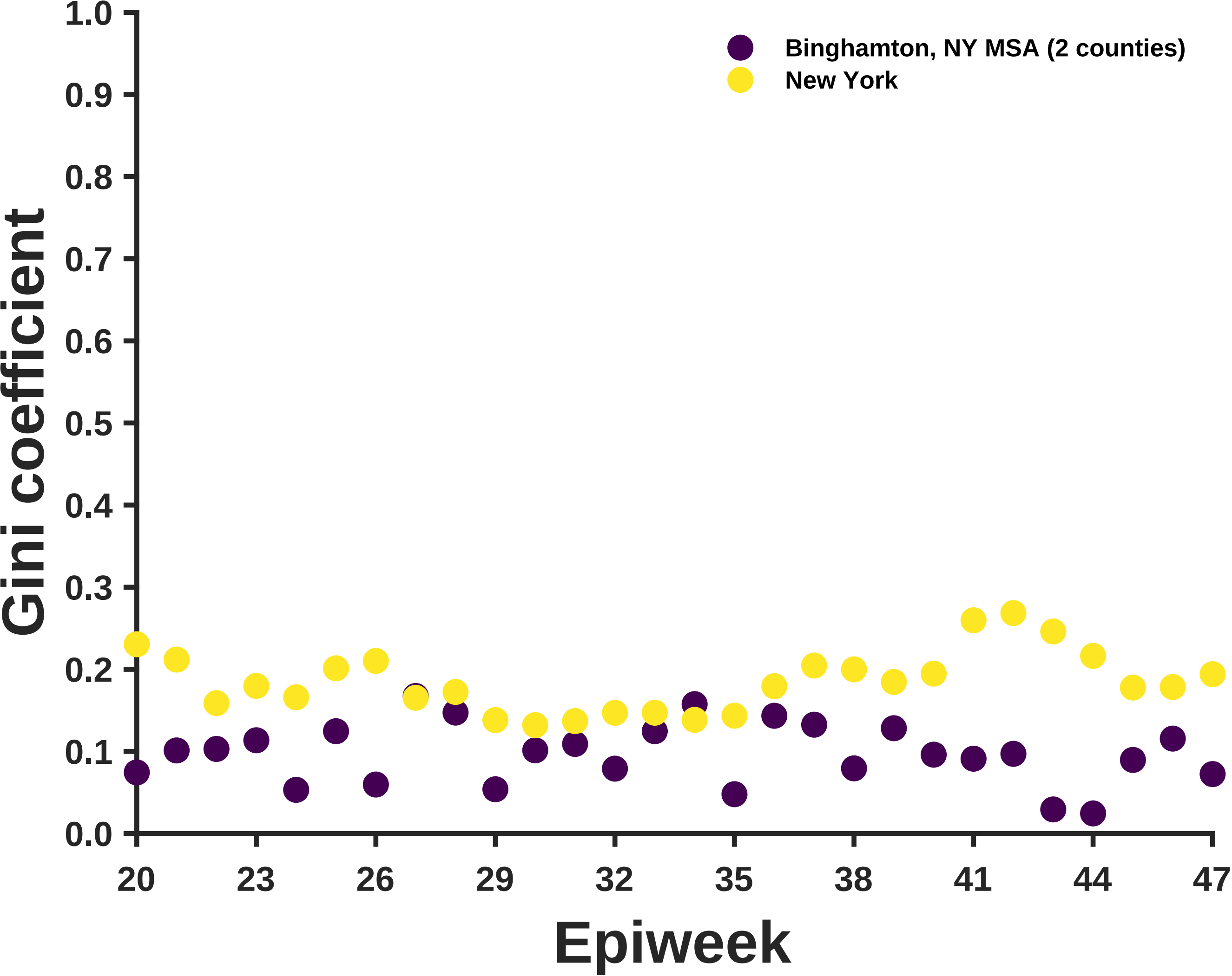
Binghamton, NY MSA (2 counties)
New York
Gini coefficient
1.0
0.9
0.8
0.7
0.6
0.5
0.4
0.3
0.2
0.1
0.0
20
23
26
29
32
35
38
41
44
47
Epiweek

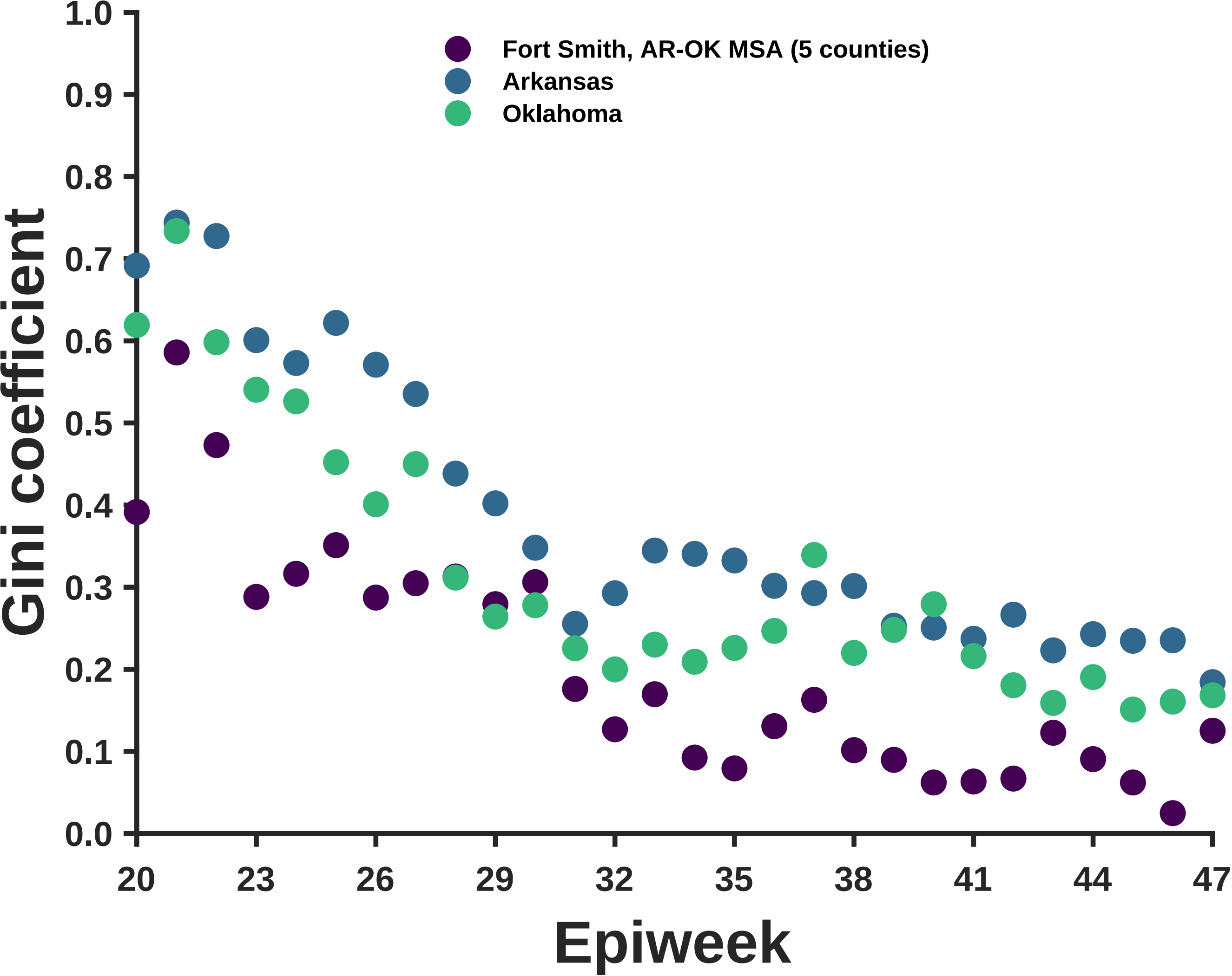

Fort Smith, AR-OK MSA (5 counties)
Arkansas
Oklahoma
Gini coefficient
Epiweek
1.0
0.9
0.8
0.7
0.6
0.5
0.4
0.3
0.2
0.1
0.0
20
23
26
29
32
35
38
41
44
47

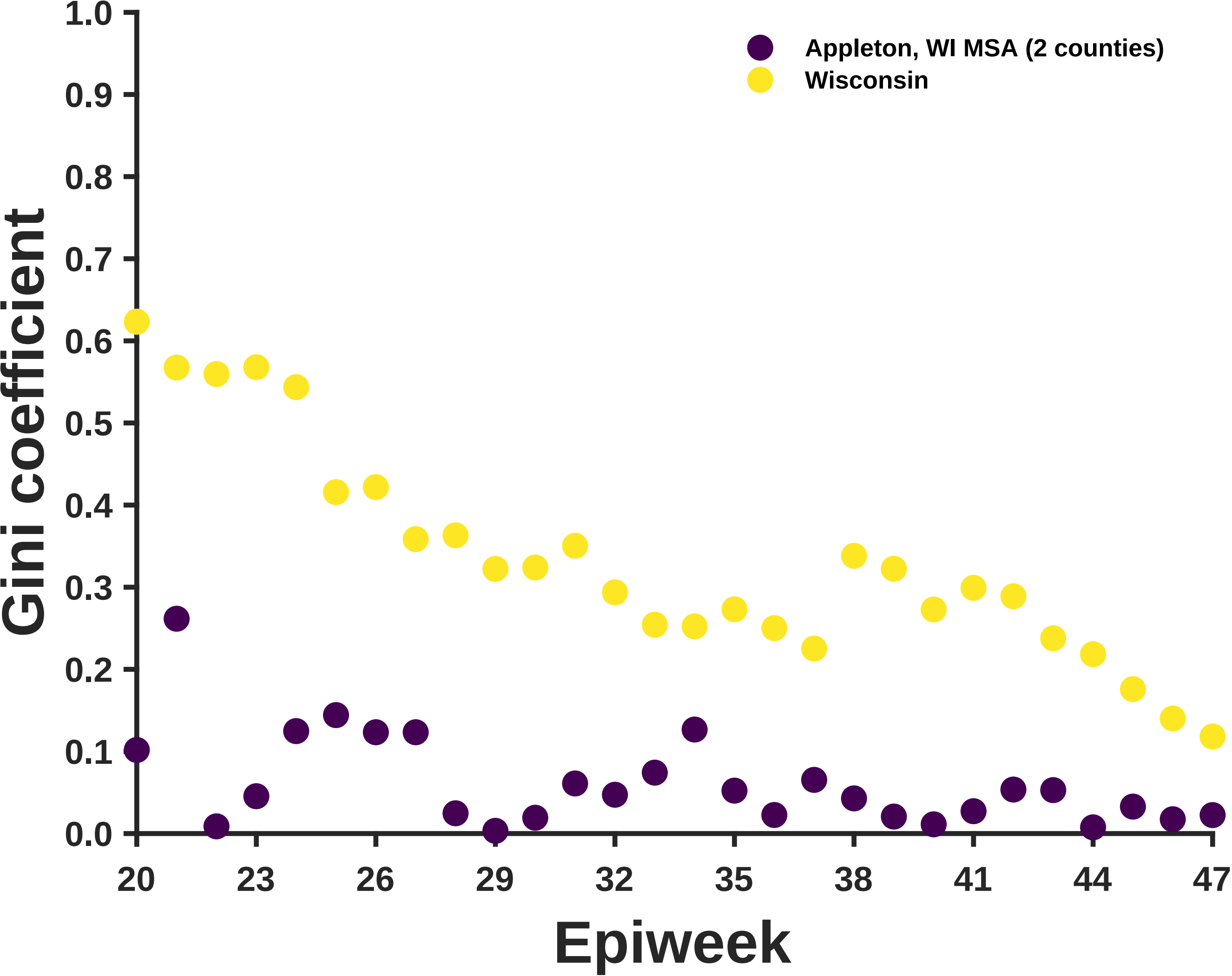
Appleton, WI MSA (2 counties)
Wisconsin
Gini coefficient
Epiweek
1.0
0.9
0.8
0.7
0.6
0.5
0.4
0.3
0.2
0.1
0.0
20
23
26
29
32
35
38
41
44
47

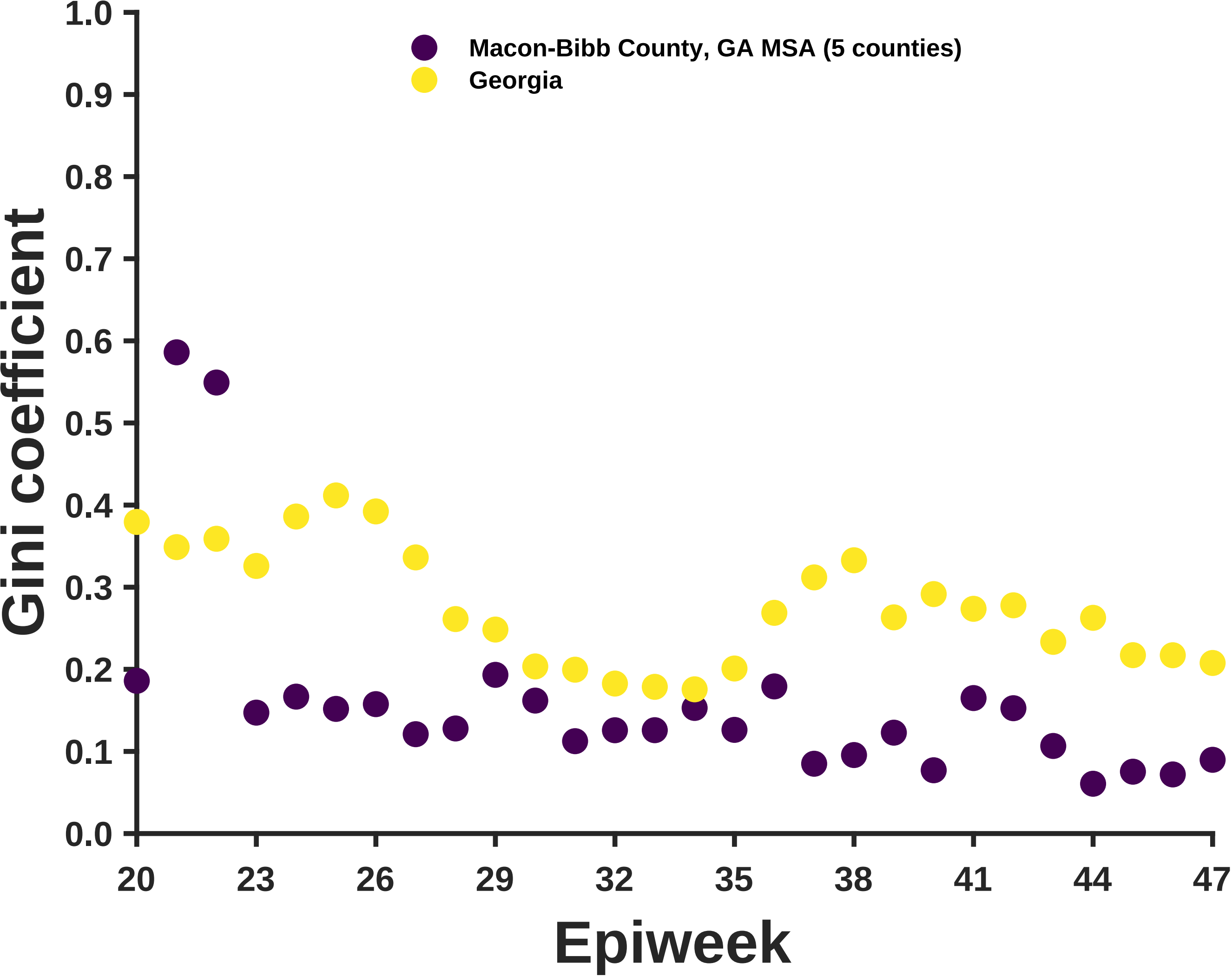

Macon-Bibb County, GA MSA (5 counties)
Georgia
Gini coefficient
1.0
0.9
0.8
0.7
0.6
0.5
0.4
0.3
0.2
0.1
0.0
20
23
26
29
32
35
38
41
44
47
Epiweek

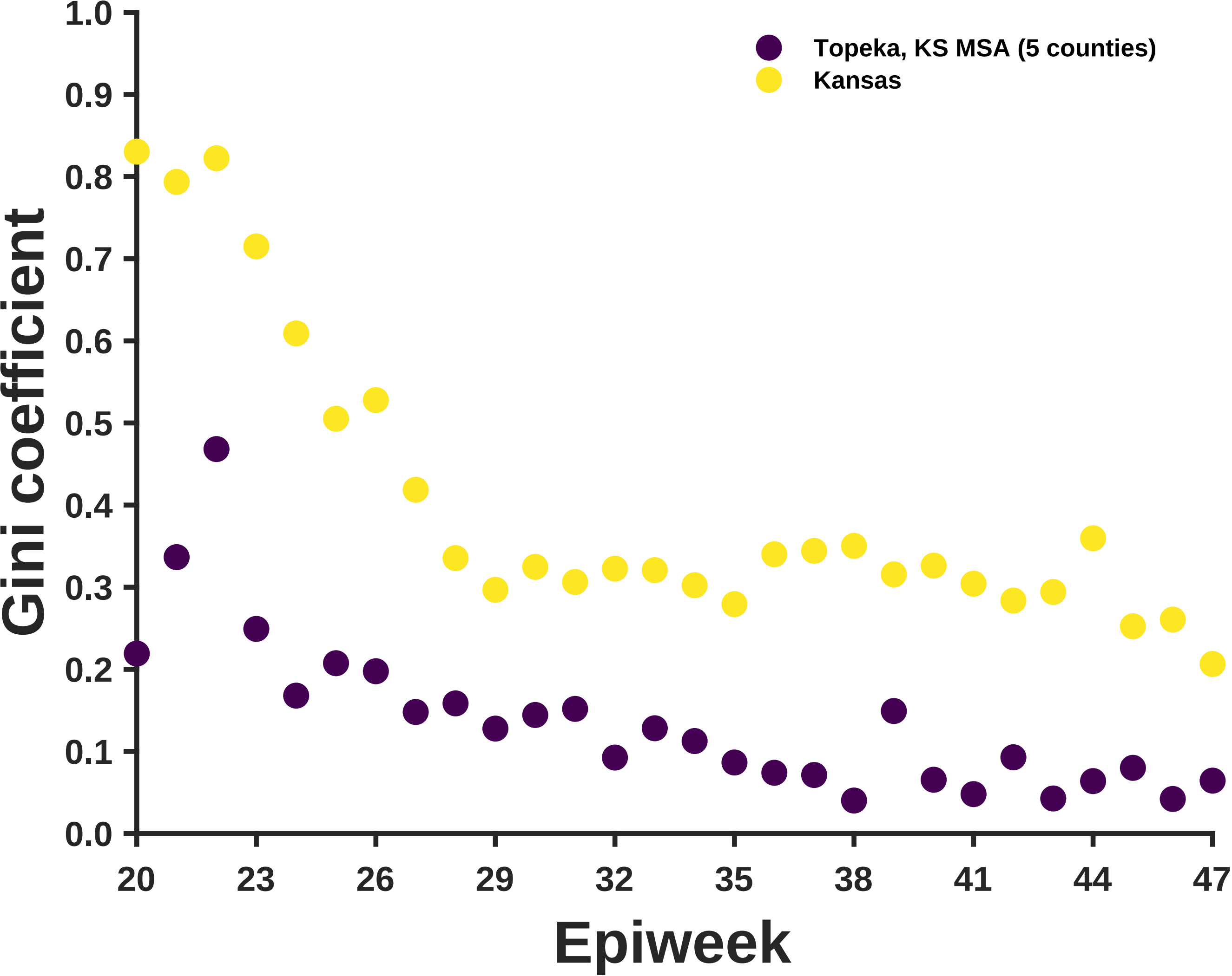

Topeka, KS MSA (5 counties)
Kansas
Gini coefficient
Epiweek
1.0
0.9
0.8
0.7
0.6
0.5
0.4
0.3
0.2
0.1
0.0
20
23
26
29
32
35
38
41
44
47

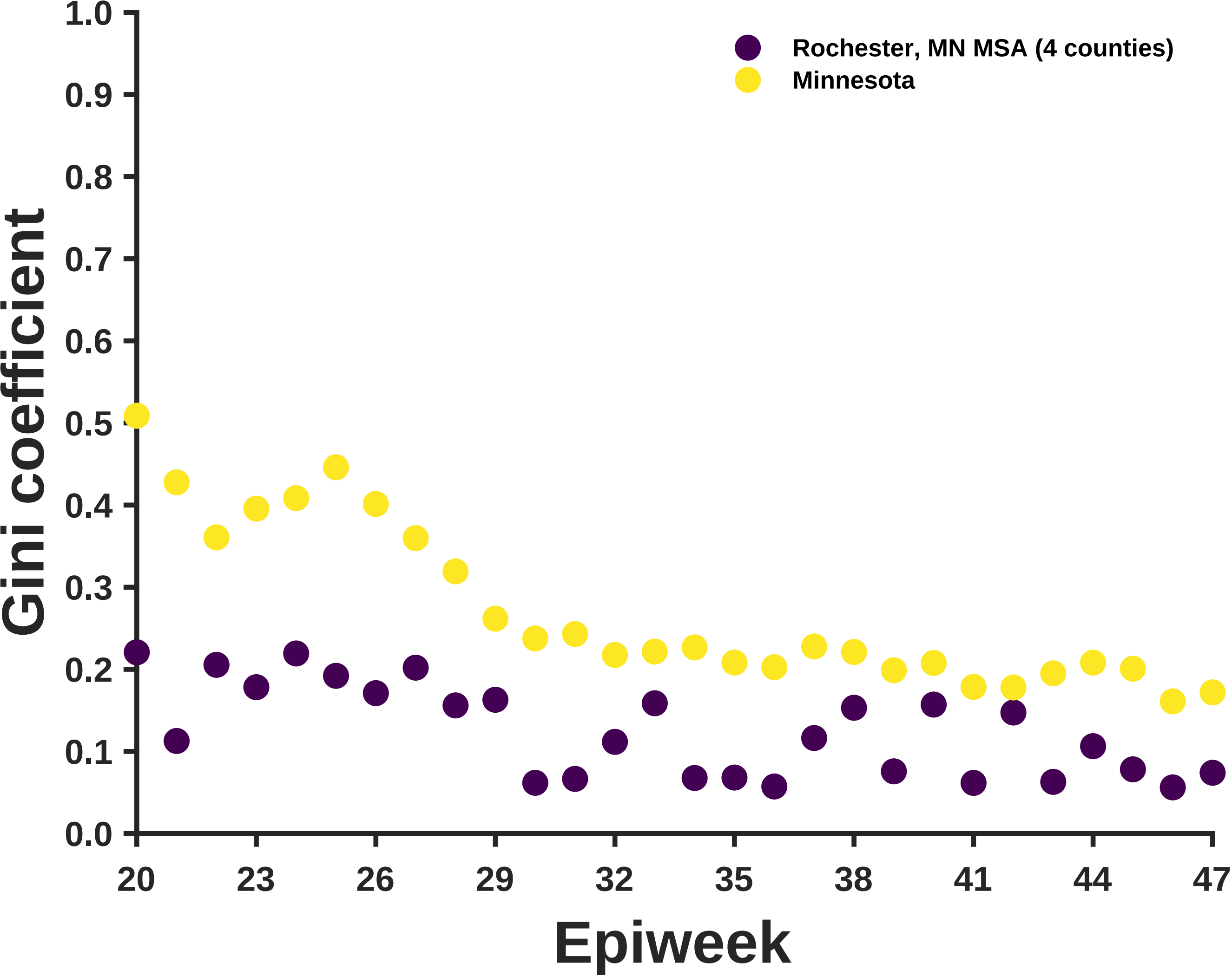
Rochester, MN MSA (4 counties)
Minnesota
Gini coefficient
1.0
0.9
0.8
0.7
0.6
0.5
0.4
0.3
0.2
0.1
0.0
20
23
26
29
32
35
38
41
44
47
Epiweek

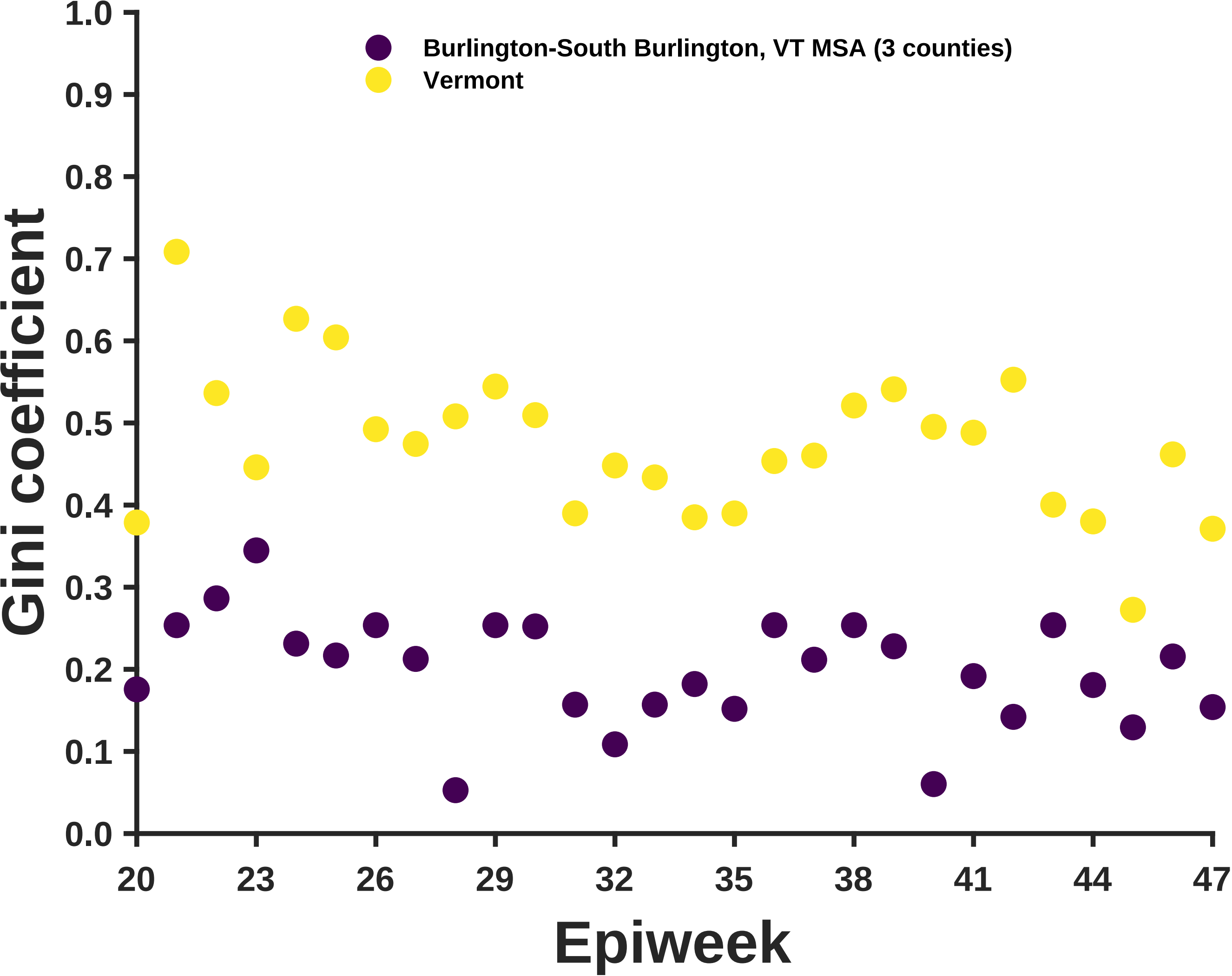

Burlington-South Burlington, VT MSA (3 counties)
Vermont
Gini coefficient
1.0
0.9
0.8
0.7
0.6
0.5
0.4
0.3
0.2
0.1
0.0
20
23
26
29
32
35
38
41
44
47
Epiweek

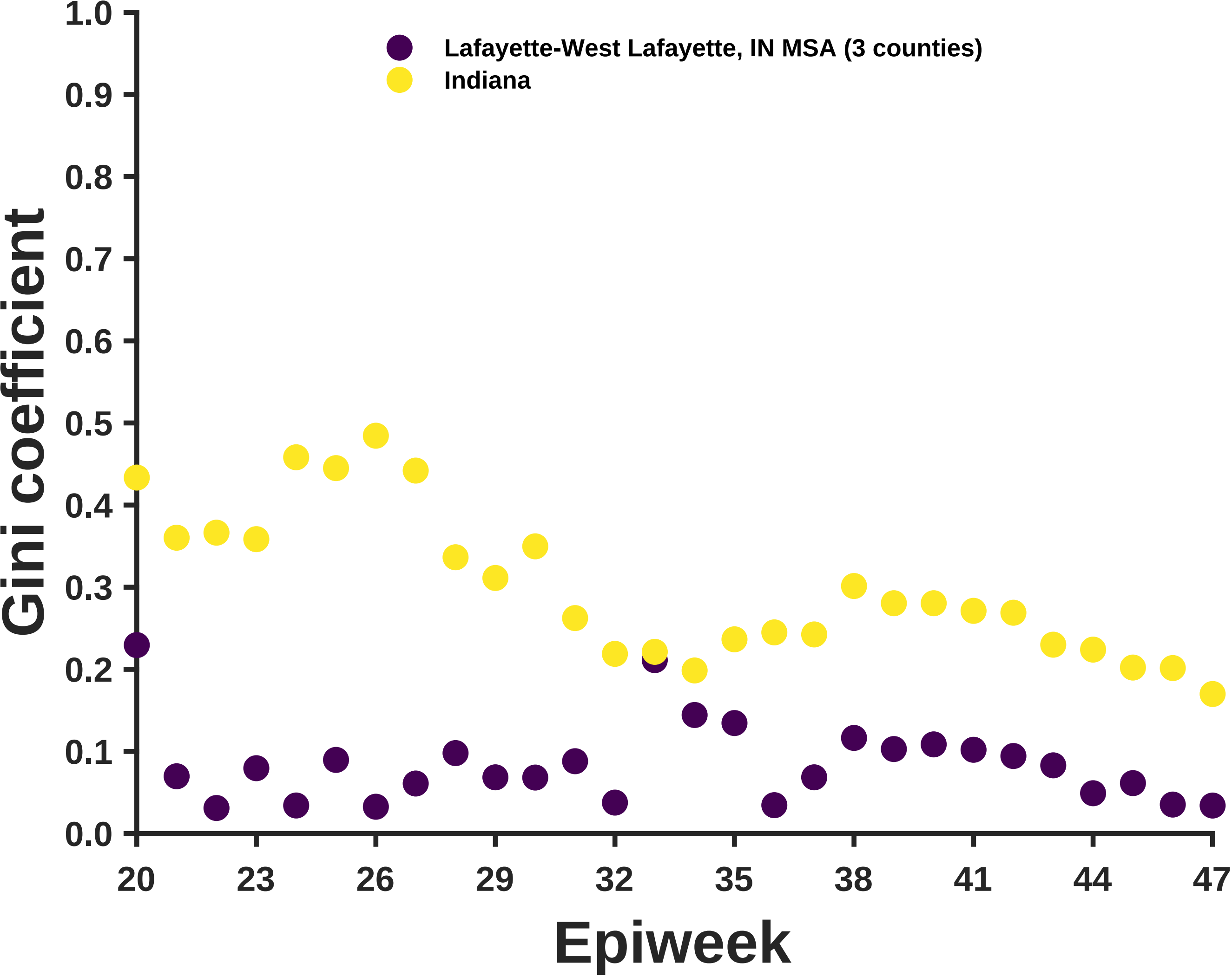

Lafayette-West Lafayette, IN MSA (3 counties)
Indiana
Gini coefficient
1.0
0.9
0.8
0.7
0.6
0.5
0.4
0.3
0.2
0.1
0.0
20
23
26
29
32
35
38
41
44
47
Epiweek

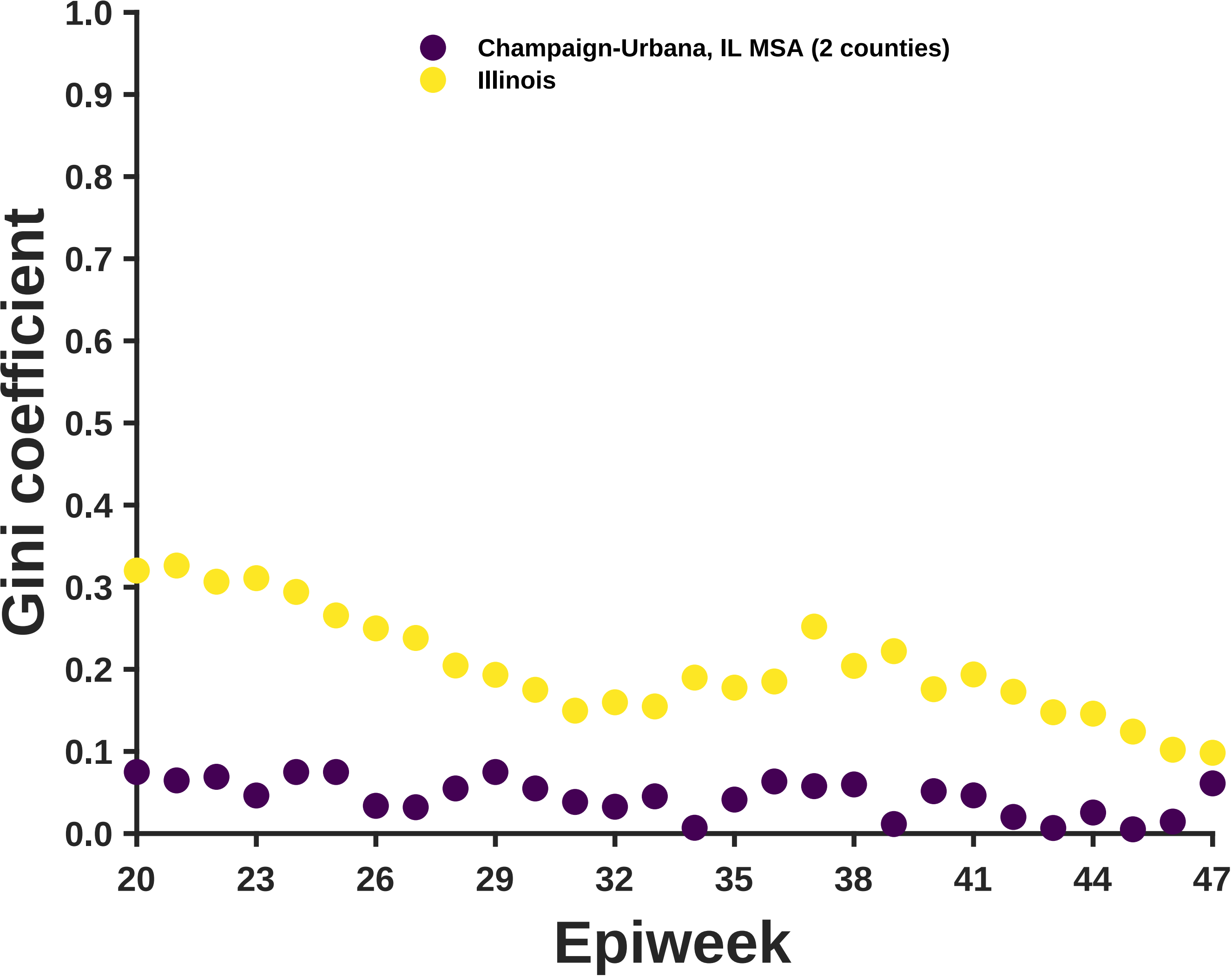
Champaign-Urbana, IL MSA (2 counties)
Illinois
Gini coefficient
1.0
0.9
0.8
0.7
0.6
0.5
0.4
0.3
0.2
0.1
0.0
20
23
26
29
32
35
38
41
44
47
Epiweek

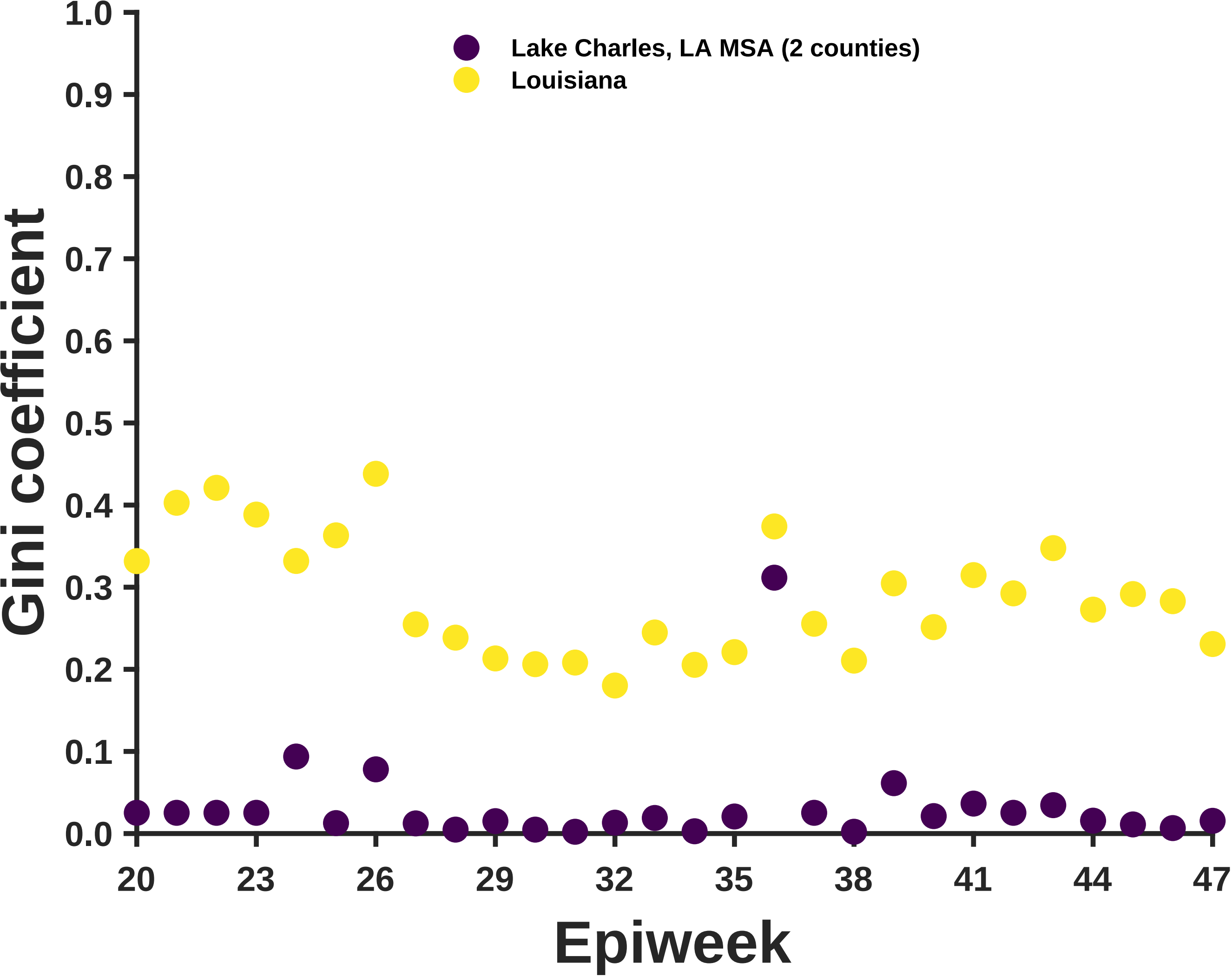

Lake Charles, LA MSA (2 counties)
Louisiana
Gini coefficient
0.0
0.1
0.2
0.3
0.4
0.5
0.6
0.7
0.8
0.9
1.0
20
23
26
29
32
35
38
41
44
47
Epiweek

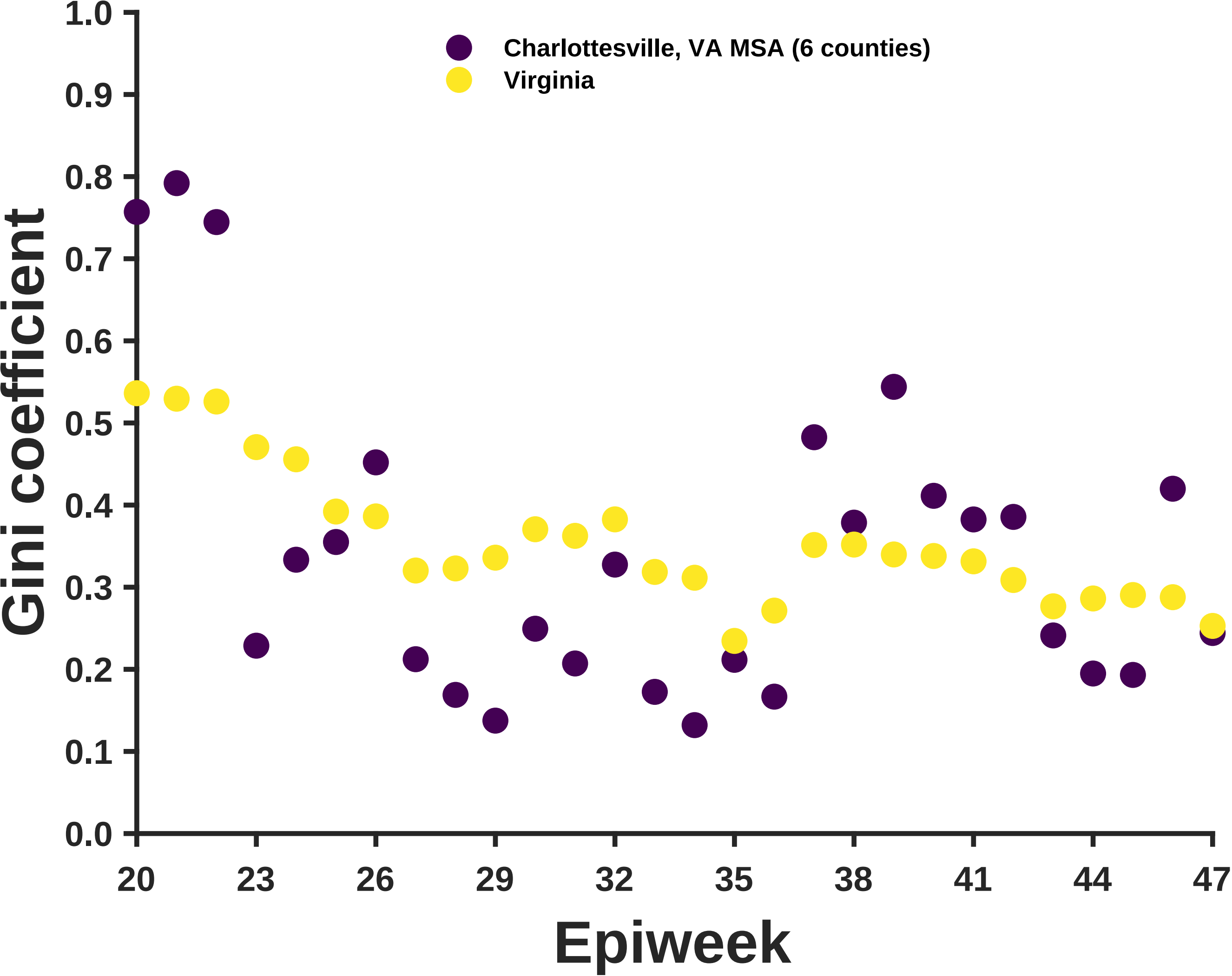
Charlottesville, VA MSA (6 counties)
Virginia
Gini coefficient
Epiweek
0.0
0.1
0.2
0.3
0.4
0.5
0.6
0.7
0.8
0.9
1.0
20
23
26
29
32
35
38
41
44
47

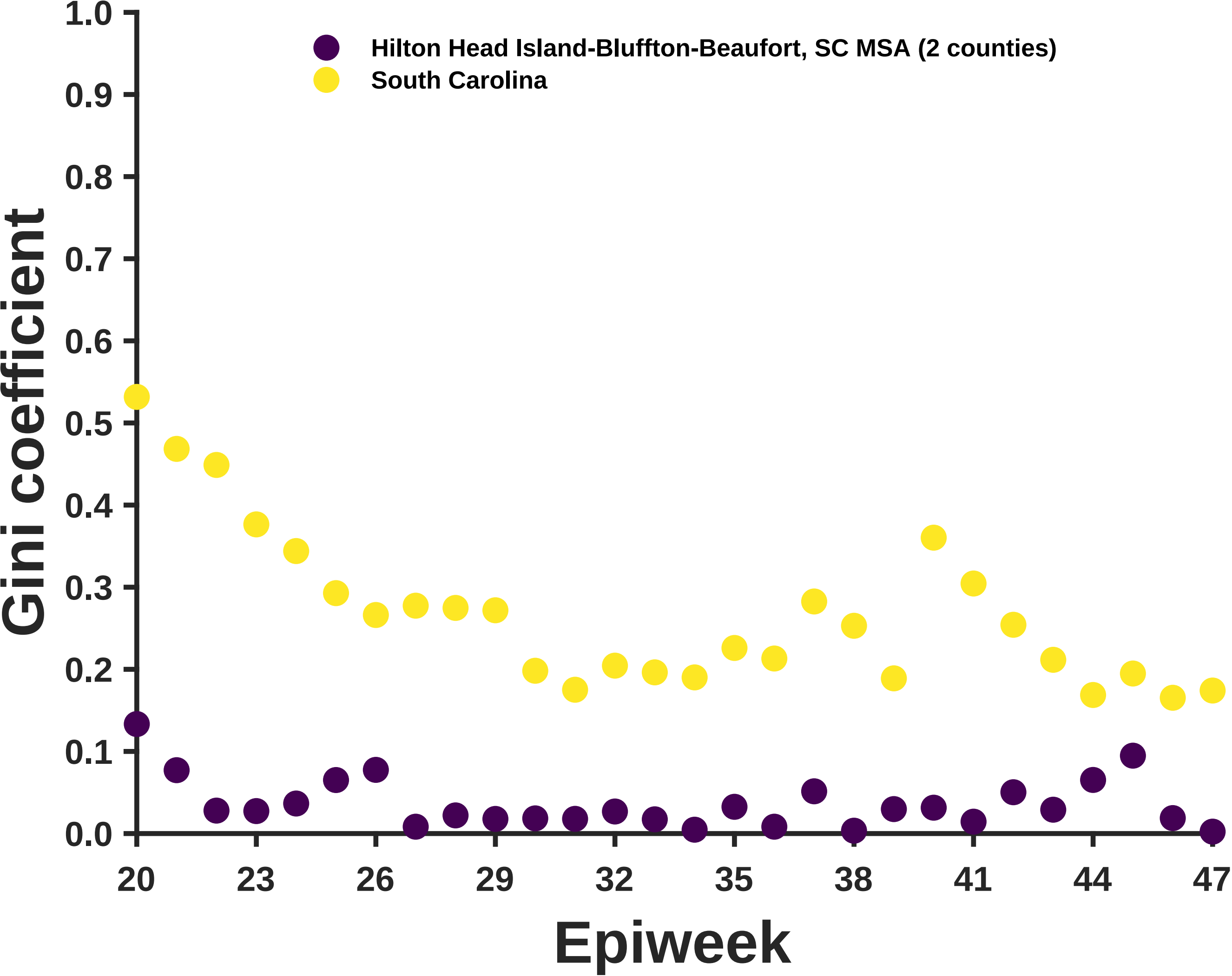
Hilton Head Island-Bluffton-Beaufort, SC MSA (2 counties)
South Carolina
Gini coefficient
1.0
0.9
0.8
0.7
0.6
0.5
0.4
0.3
0.2
0.1
0.0
20
23
26
29
32
35
38
41
44
47
Epiweek

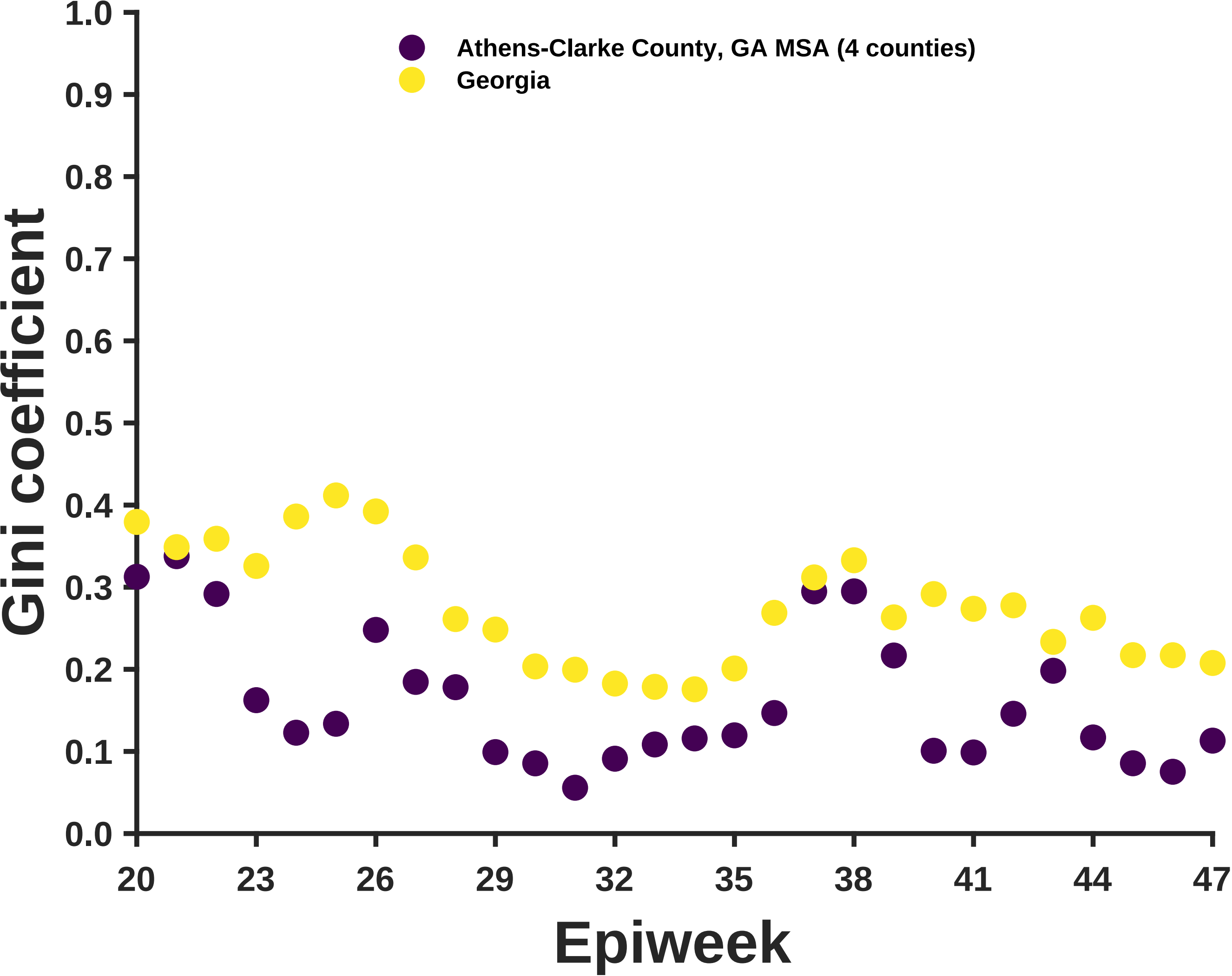

Athens-Clarke County, GA MSA (4 counties)
Georgia
Gini coefficient
Epiweek
1.0
0.9
0.8
0.7
0.6
0.5
0.4
0.3
0.2
0.1
0.0
20
23
26
29
32
35
38
41
44
47

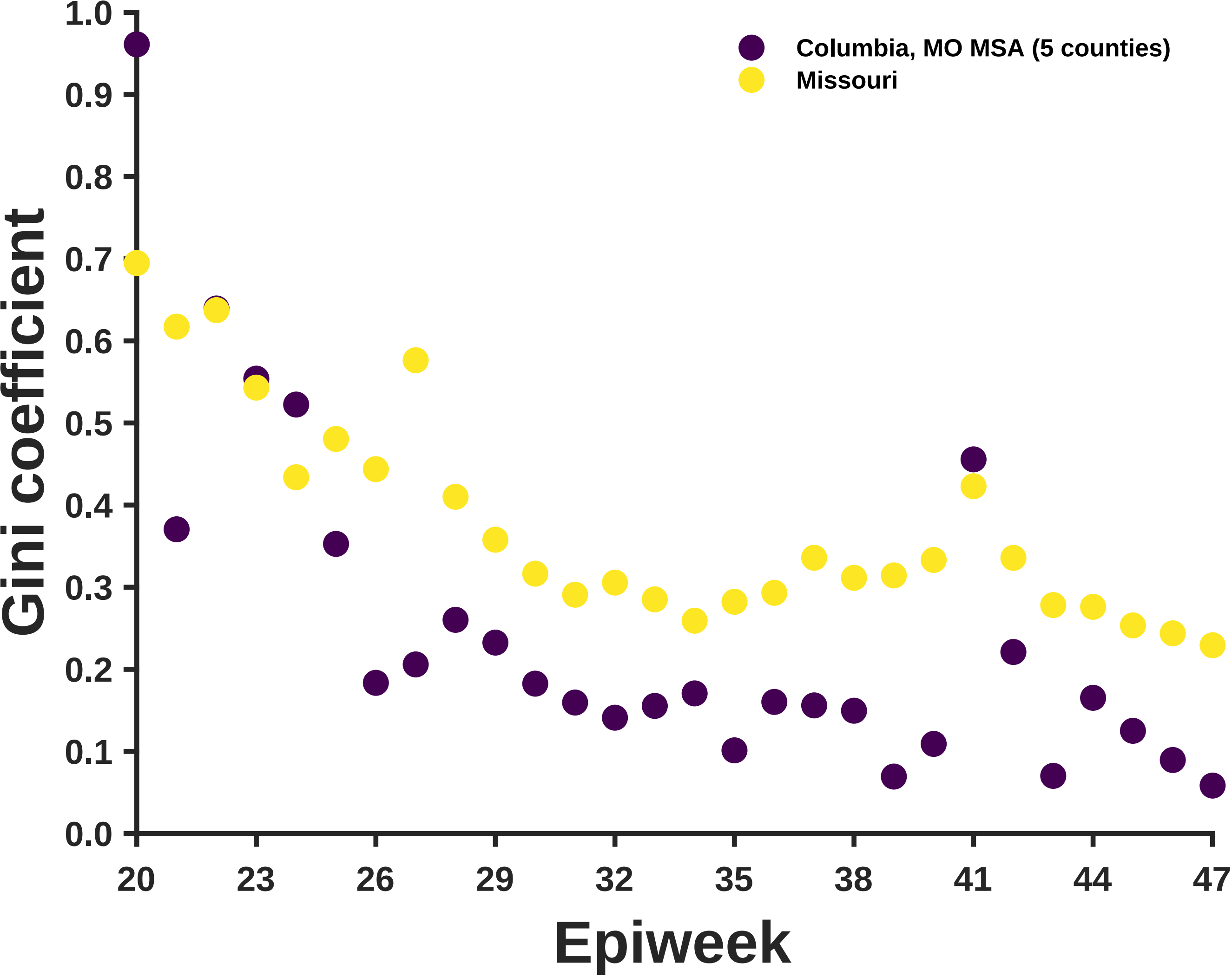

Gini coefficient
Epiweek
Columbia, MO MSA (5 counties)
Missouri
1.0
0.9
0.8
0.7
0.6
0.5
0.4
0.3
0.2
0.1
0.0
20
23
26
29
32
35
38
41
44
47

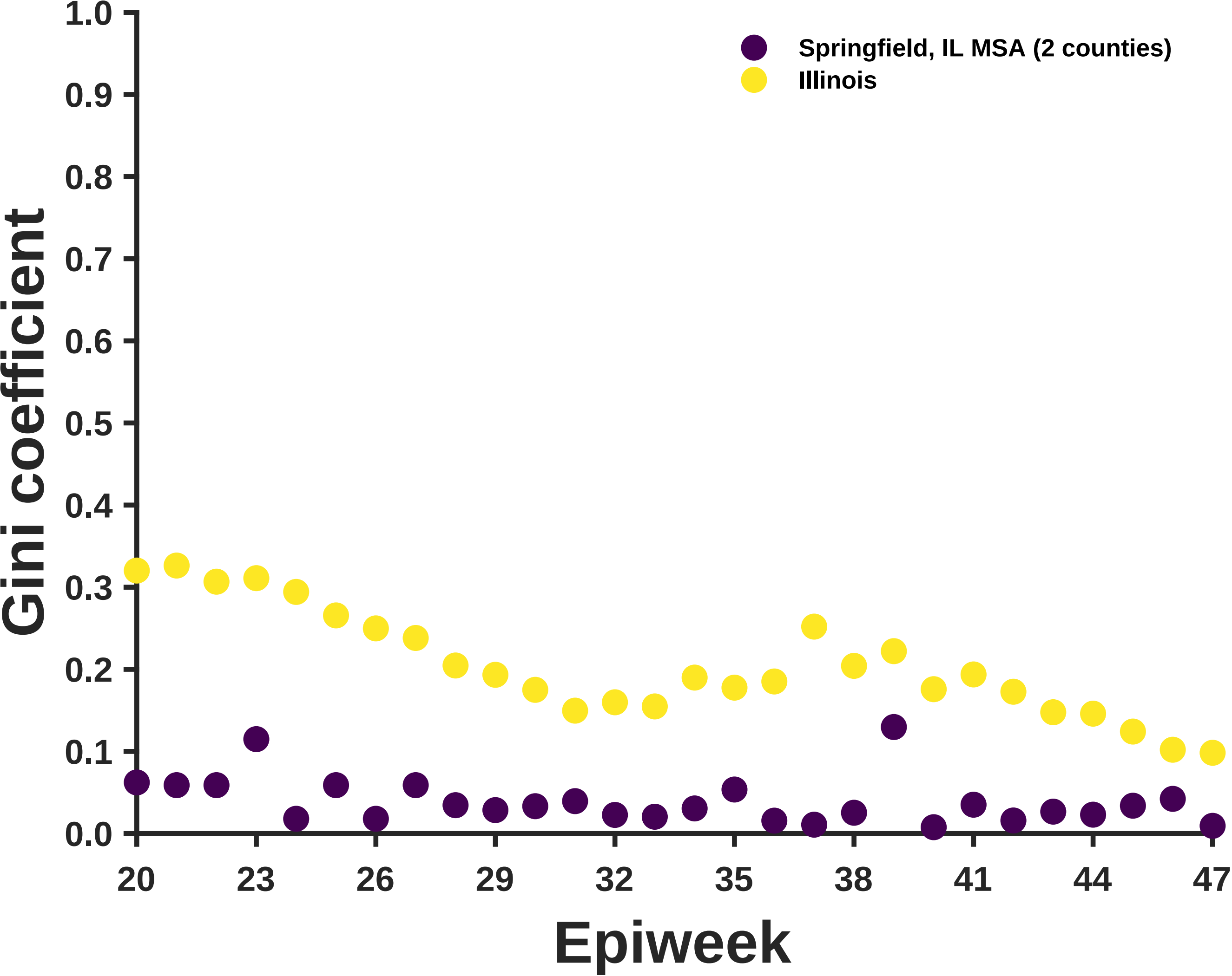

Springfield, IL MSA (2 counties)
Illinois
Gini coefficient
0.0
0.1
0.2
0.3
0.4
0.5
0.6
0.7
0.8
0.9
1.0
20
23
26
29
32
35
38
41
44
47
Epiweek

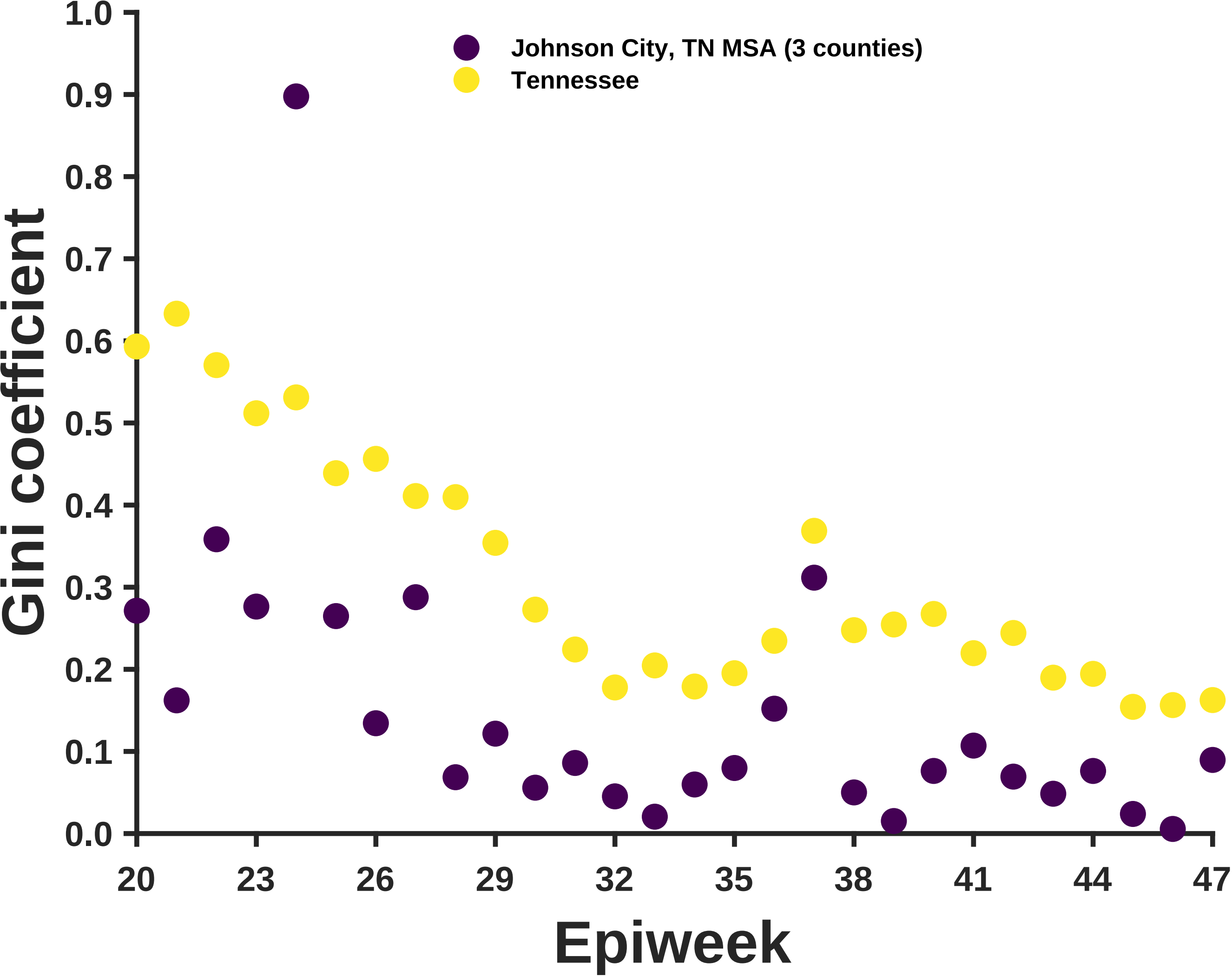

Johnson City, TN MSA (3 counties)
Tennessee
Gini coefficient
1.0
0.9
0.8
0.7
0.6
0.5
0.4
0.3
0.2
0.1
0.0
20
23
26
29
32
35
38
41
44
47
Epiweek

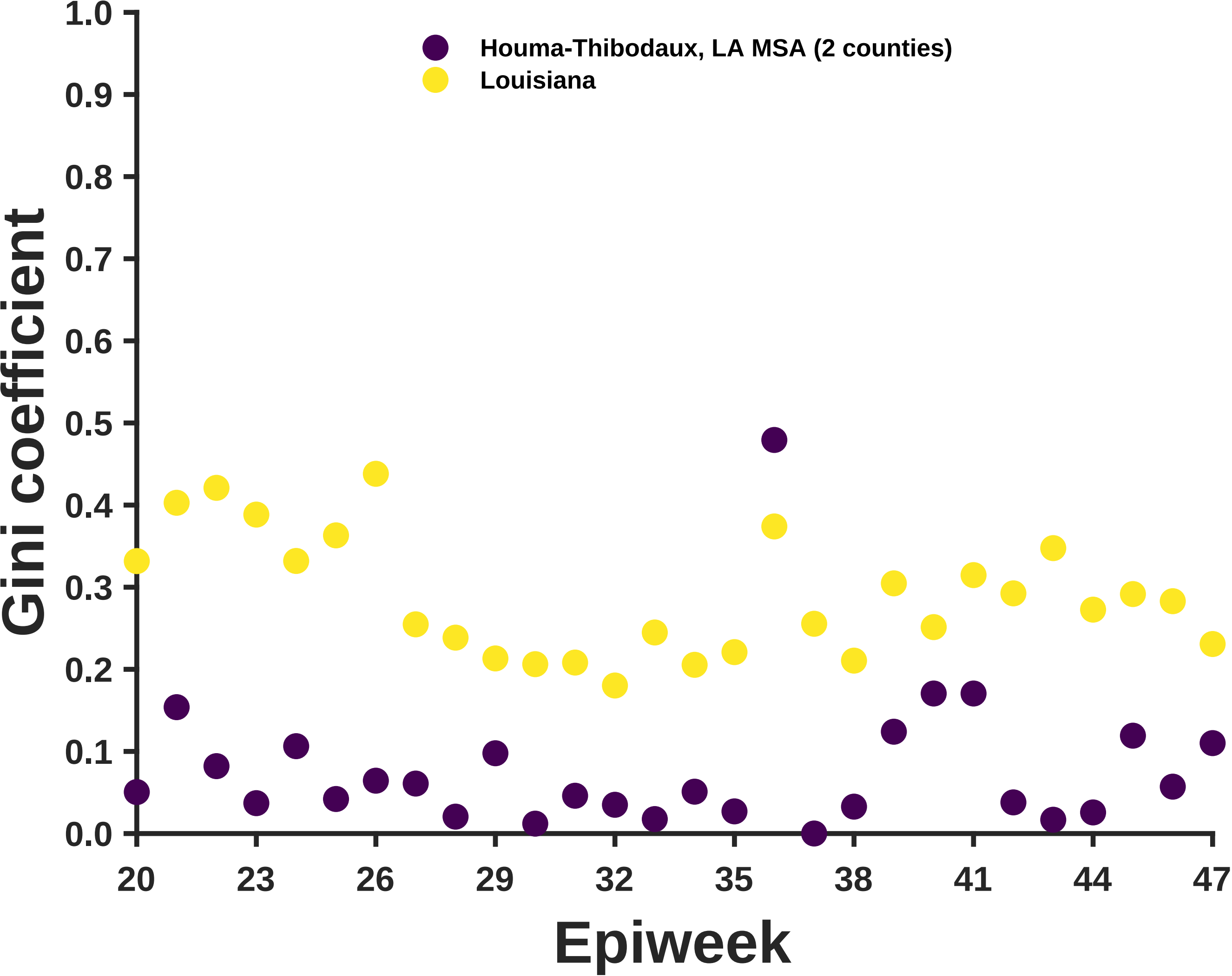

Houma-Thibodaux, LA MSA (2 counties)
Louisiana
Gini coefficient
1.0
0.9
0.8
0.7
0.6
0.5
0.4
0.3
0.2
0.1
0.0
20
23
26
29
32
35
38
41
44
47
Epiweek

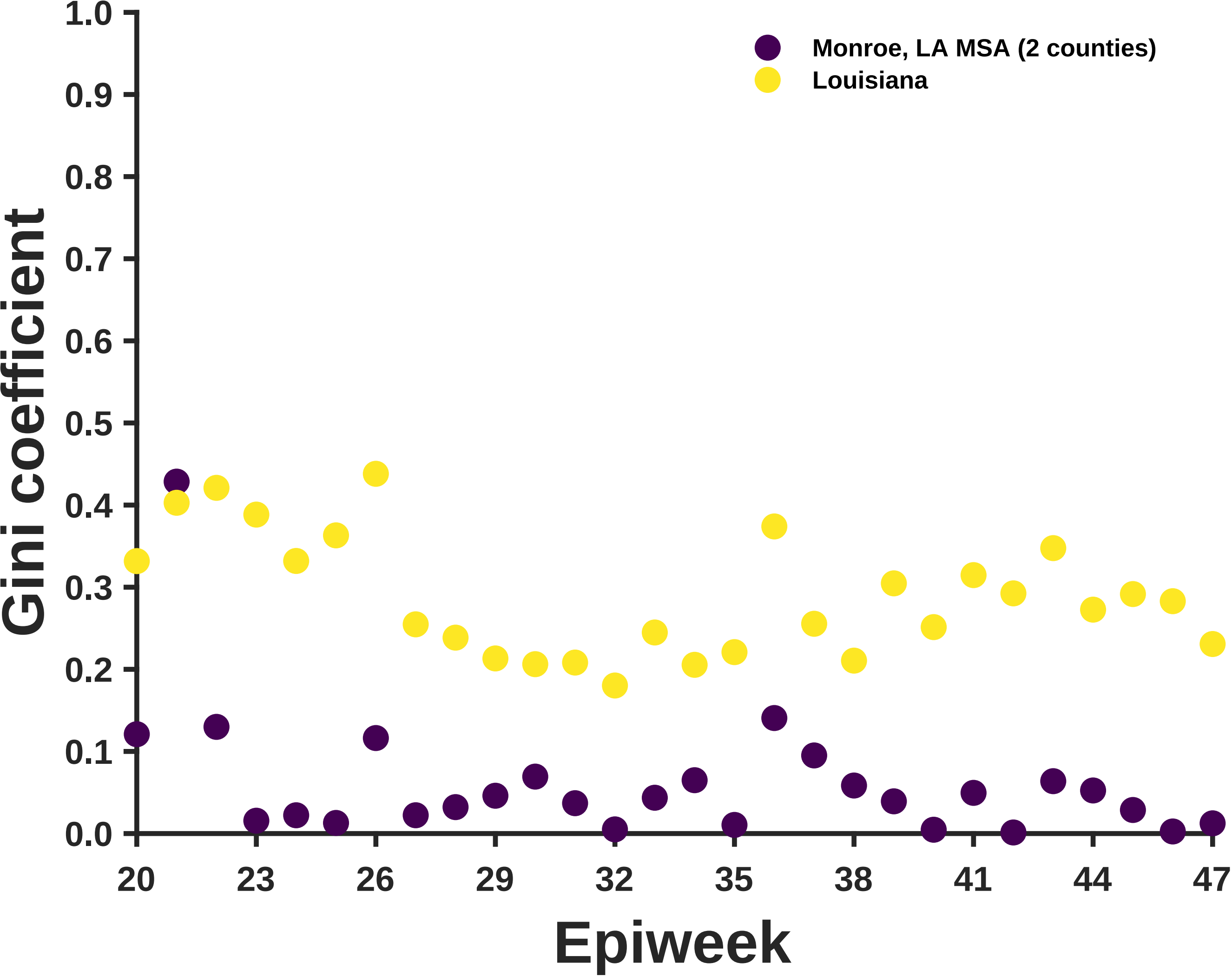

Monroe, LA MSA (2 counties)
Louisiana
Gini coefficient
Epiweek
1.0
0.9
0.8
0.7
0.6
0.5
0.4
0.3
0.2
0.1
0.0
20
23
26
29
32
35
38
41
44
47

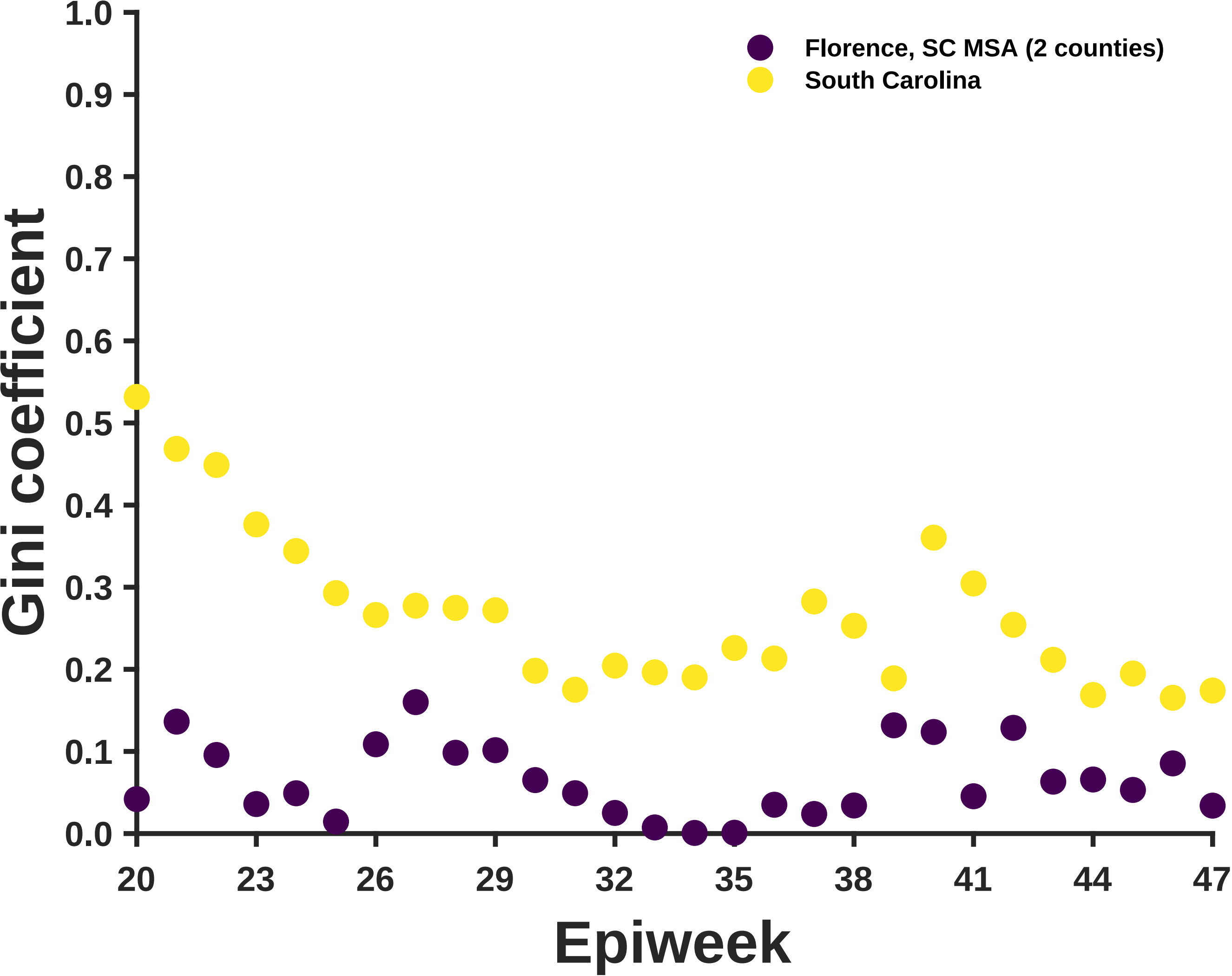

Florence, SC MSA (2 counties)
South Carolina
Gini coefficient
Epiweek
1.0
0.9
0.8
0.7
0.6
0.5
0.4
0.3
0.2
0.1
0.0
20
23
26
29
32
35
38
41
44
47

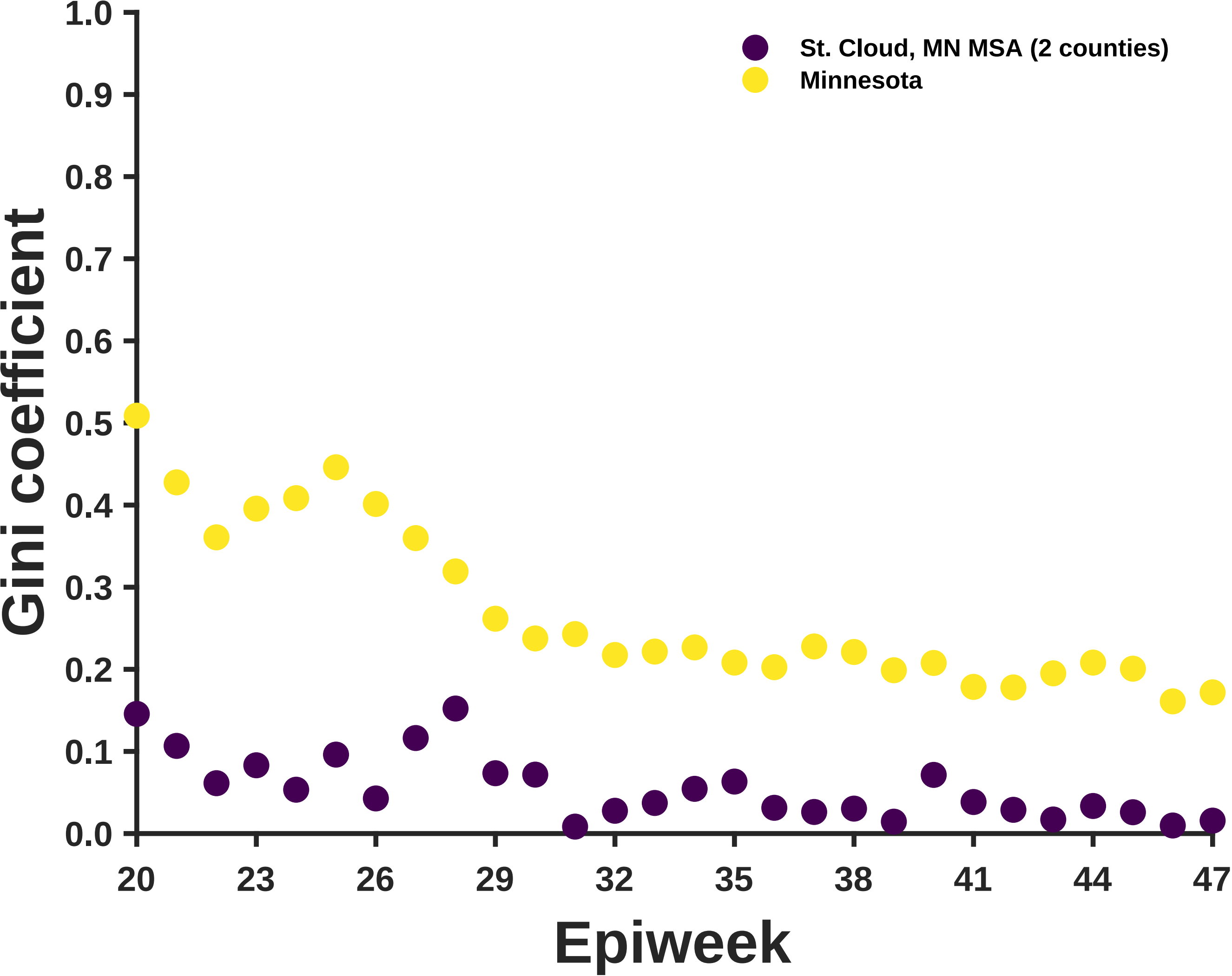

St. Cloud, MN MSA (2 counties)
Minnesota
Gini coefficient
1.0
0.9
0.8
0.7
0.6
0.5
0.4
0.3
0.2
0.1
0.0
20
23
26
29
32
35
38
41
44
47
Epiweek

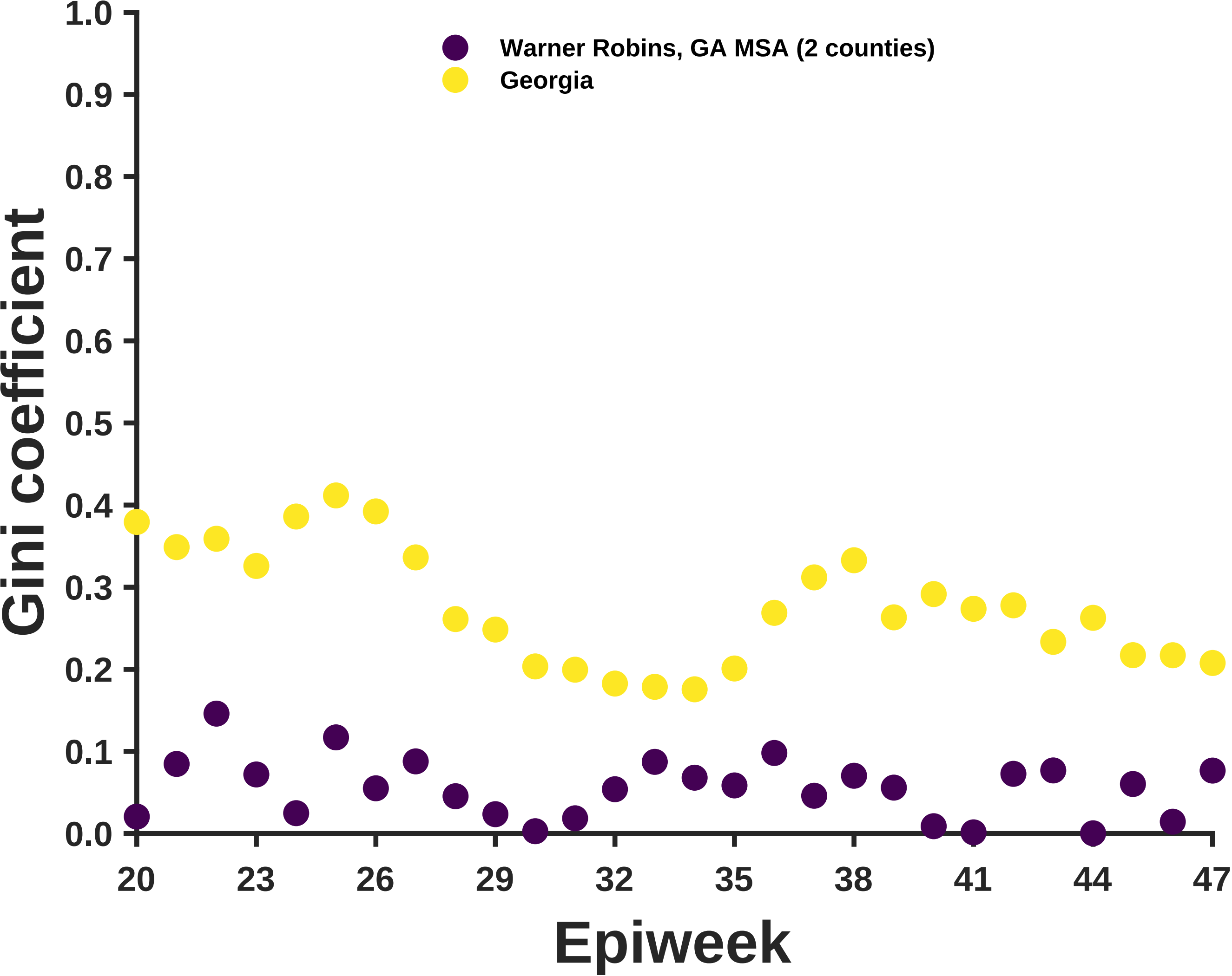

Warner Robins, GA MSA (2 counties)
Georgia
Gini coefficient
1.0
0.9
0.8
0.7
0.6
0.5
0.4
0.3
0.2
0.1
0.0
20
23
26
29
32
35
38
41
44
47
Epiweek

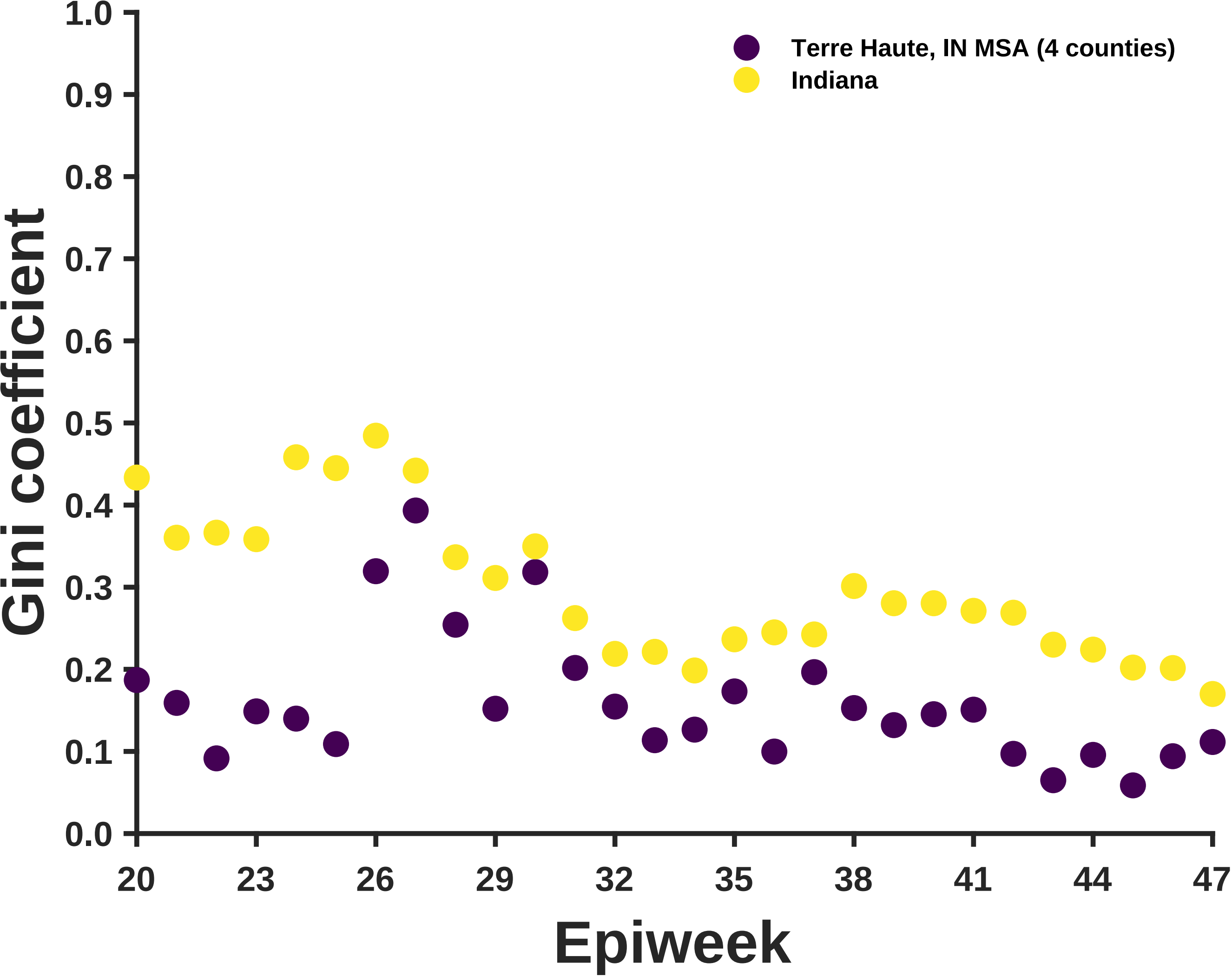

Terre Haute, IN MSA (4 counties)
Indiana
Gini coefficient
1.0
0.9
0.8
0.7
0.6
0.5
0.4
0.3
0.2
0.1
0.0
20
23
26
29
32
35
38
41
44
47
Epiweek

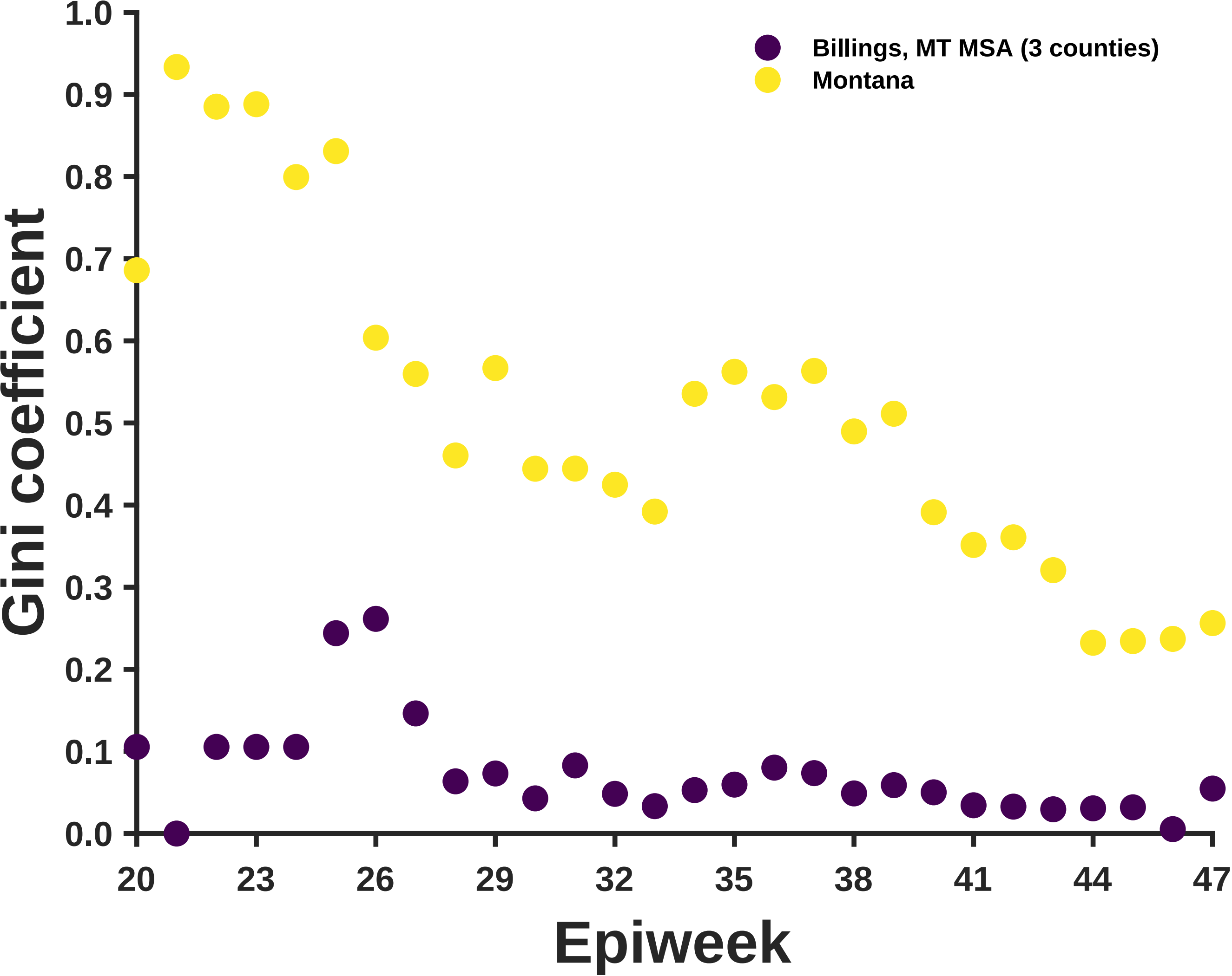

Billings, MT MSA (3 counties)
Montana
Gini coefficient
Epiweek
0.0
0.1
0.2
0.3
0.4
0.5
0.6
0.7
0.8
0.9
1.0
20
23
26
29
32
35
38
41
44
47

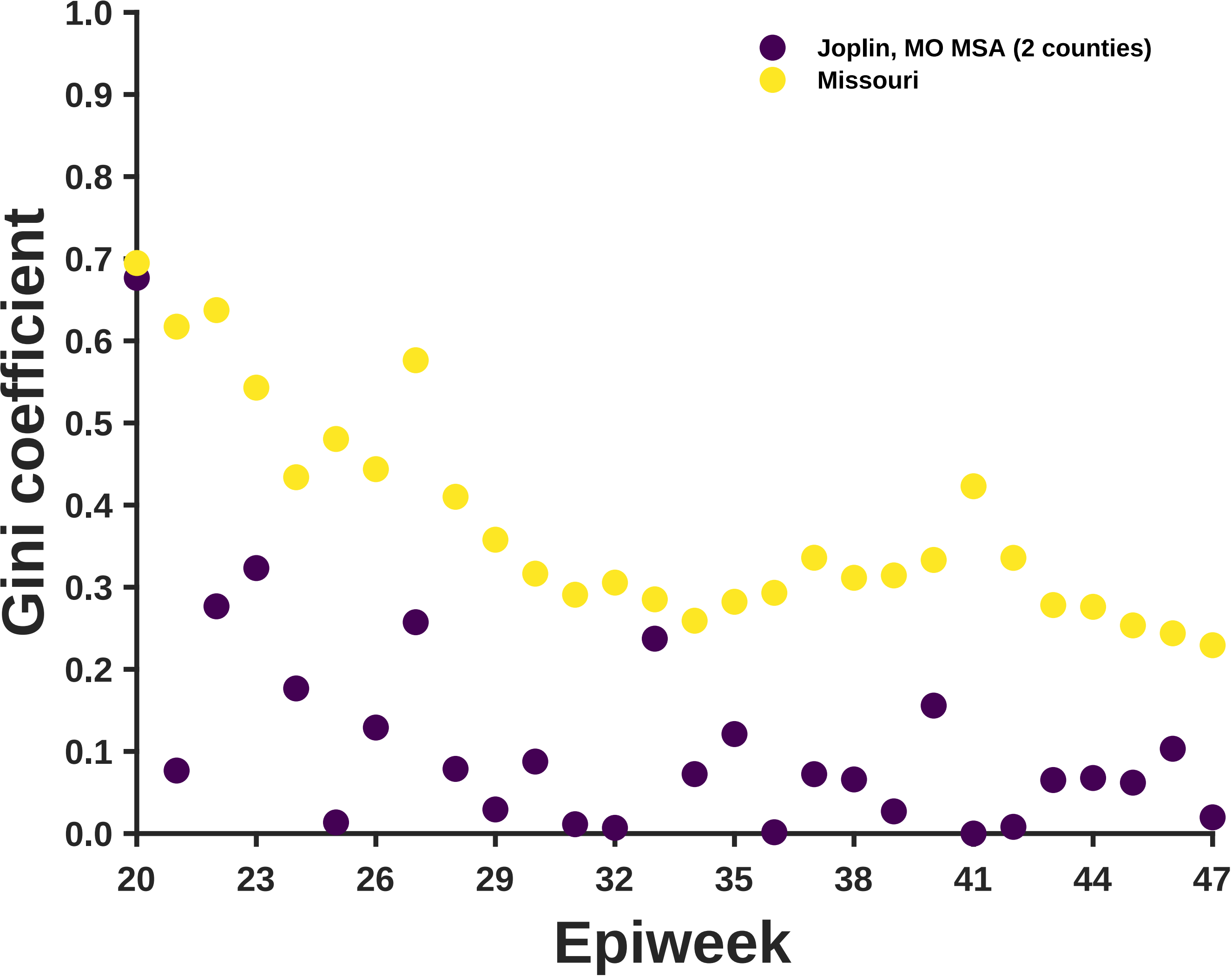

Joplin, MO MSA (2 counties)
Missouri
Gini coefficient
Epiweek
1.0
0.9
0.8
0.7
0.6
0.5
0.4
0.3
0.2
0.1
0.0
20
23
26
29
32
35
38
41
44
47

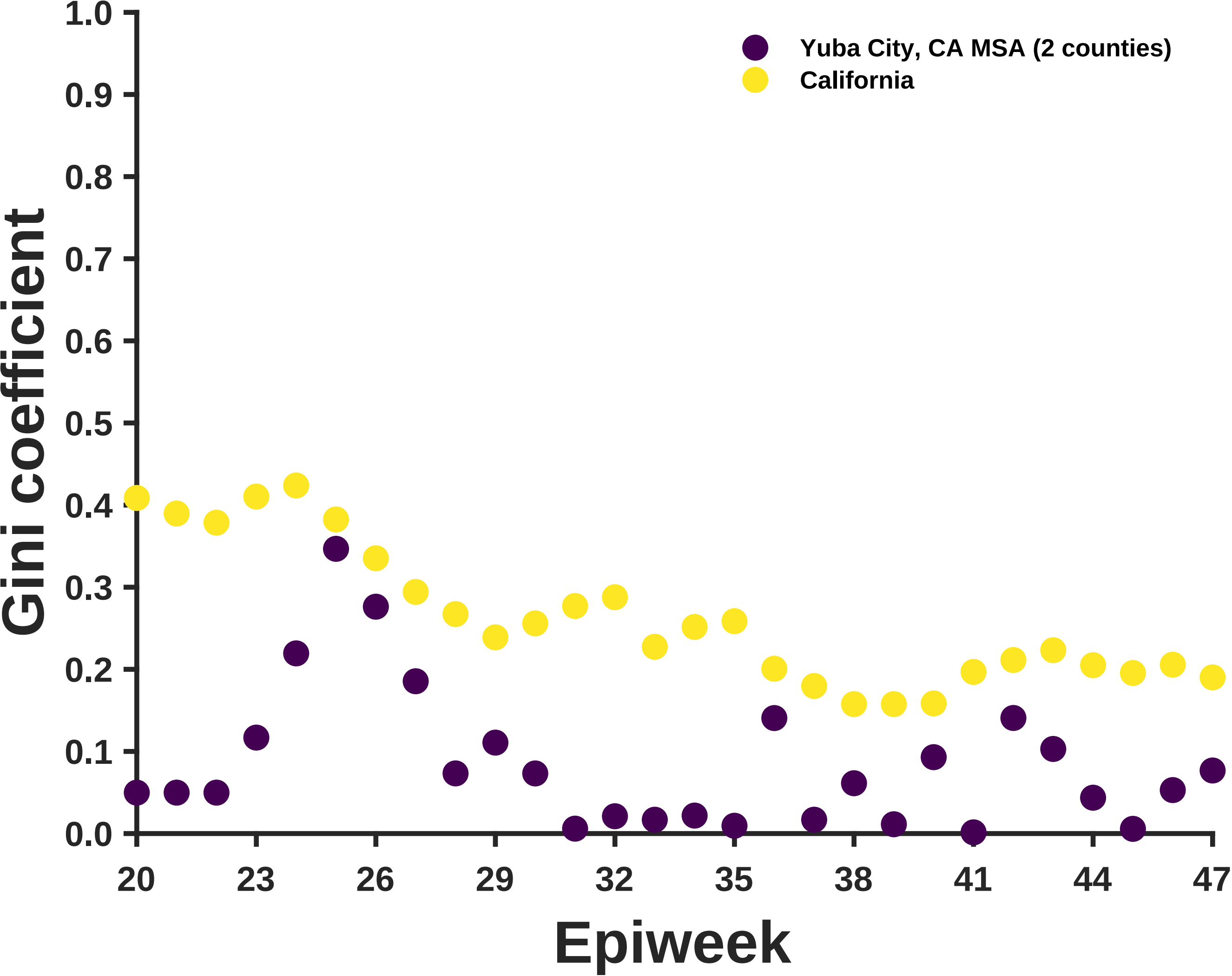
Yuba City, CA MSA (2 counties)
California
Gini coefficient
1.0
0.9
0.8
0.7
0.6
0.5
0.4
0.3
0.2
0.1
0.0
20
23
26
29
32
35
38
41
44
47
Epiweek

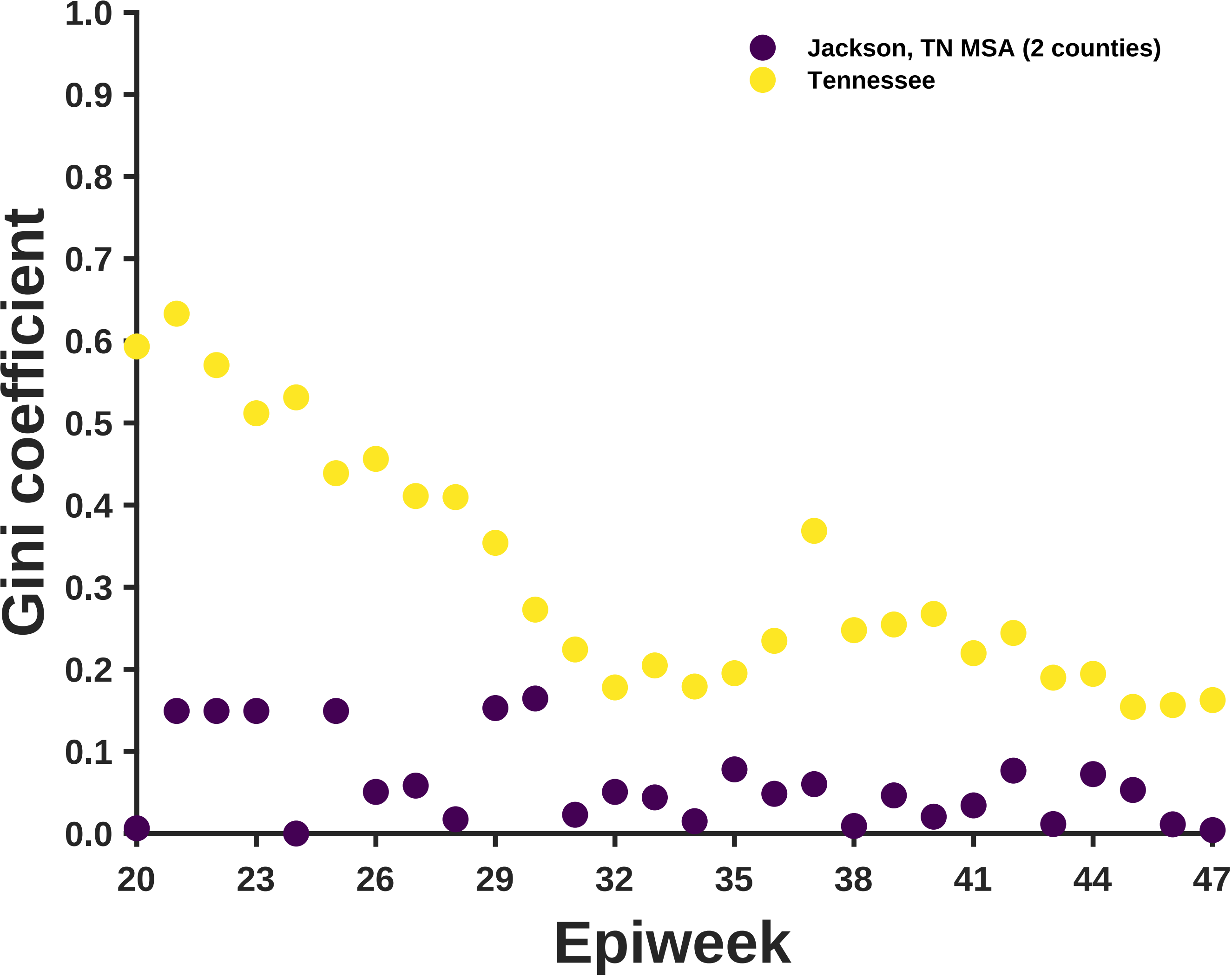

Jackson, TN MSA (2 counties)
Tennessee
Gini coefficient
Epiweek
0.0
0.1
0.2
0.3
0.4
0.5
0.6
0.7
0.8
0.9
1.0
20
23
26
29
32
35
38
41
44
47

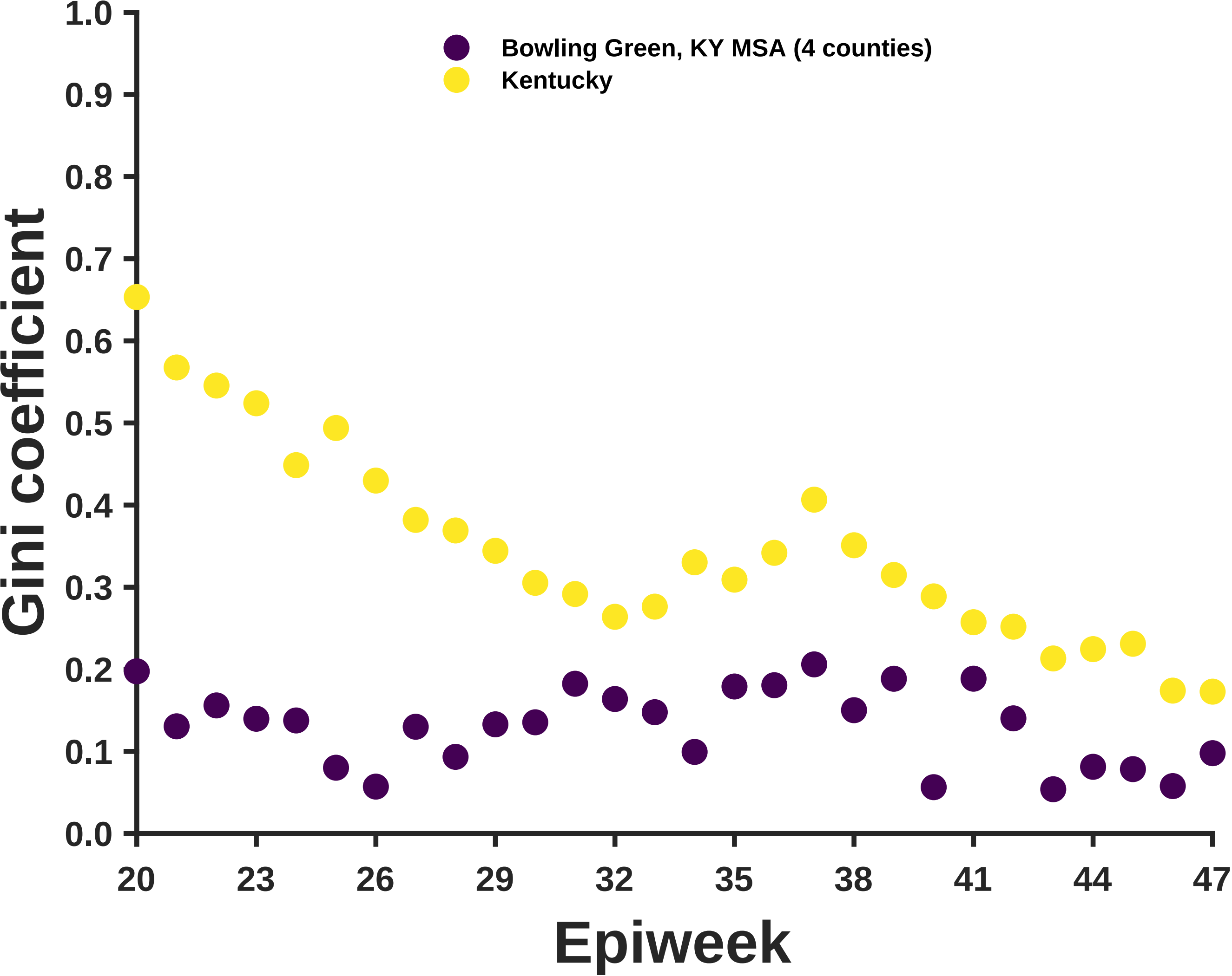

Bowling Green, KY MSA (4 counties)
Kentucky
Gini coefficient
1.0
0.9
0.8
0.7
0.6
0.5
0.4
0.3
0.2
0.1
0.0
20
23
26
29
32
35
38
41
44
47
Epiweek

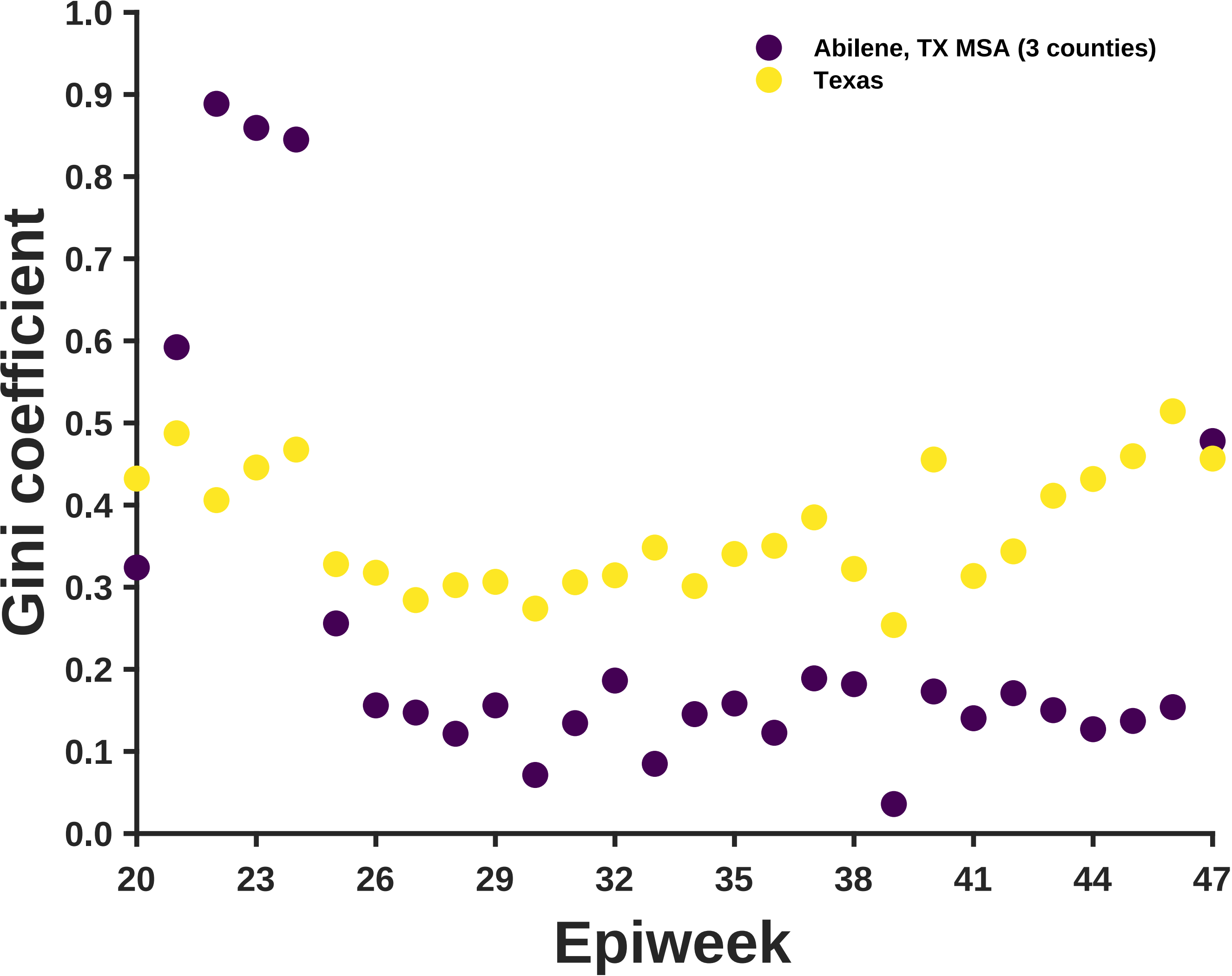

Gini coefficient
1.0
0.9
0.8
0.7
0.6
0.5
0.4
0.3
0.2
0.1
0.0
20
23
26
29
32
35
38
41
44
47
Epiweek
Abilene, TX MSA (3 counties)
Texas

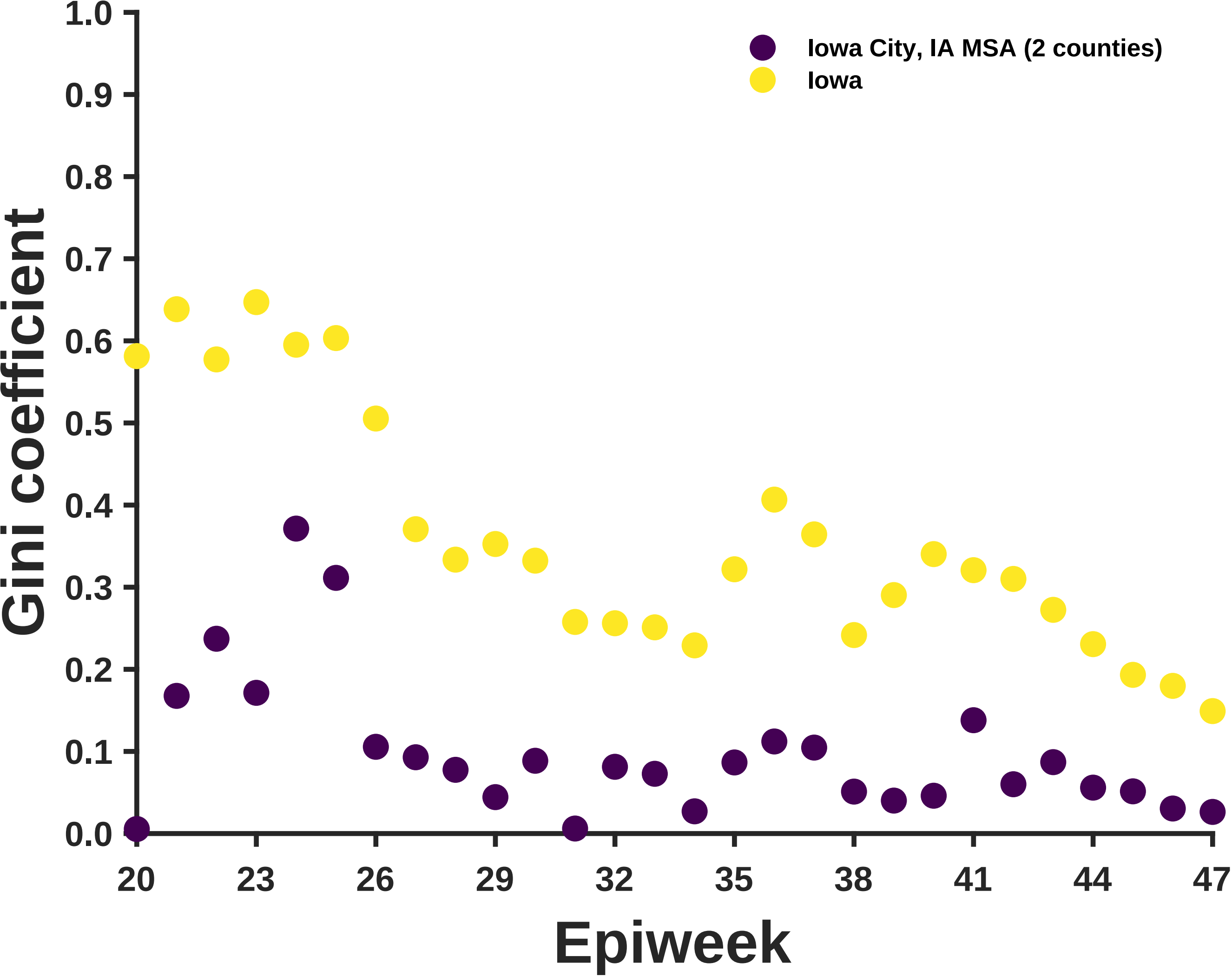
Iowa City, IA MSA (2 counties)
Iowa
Gini coefficient
1.0
0.9
0.8
0.7
0.6
0.5
0.4
0.3
0.2
0.1
0.0
20
23
26
29
32
35
38
41
44
47
Epiweek

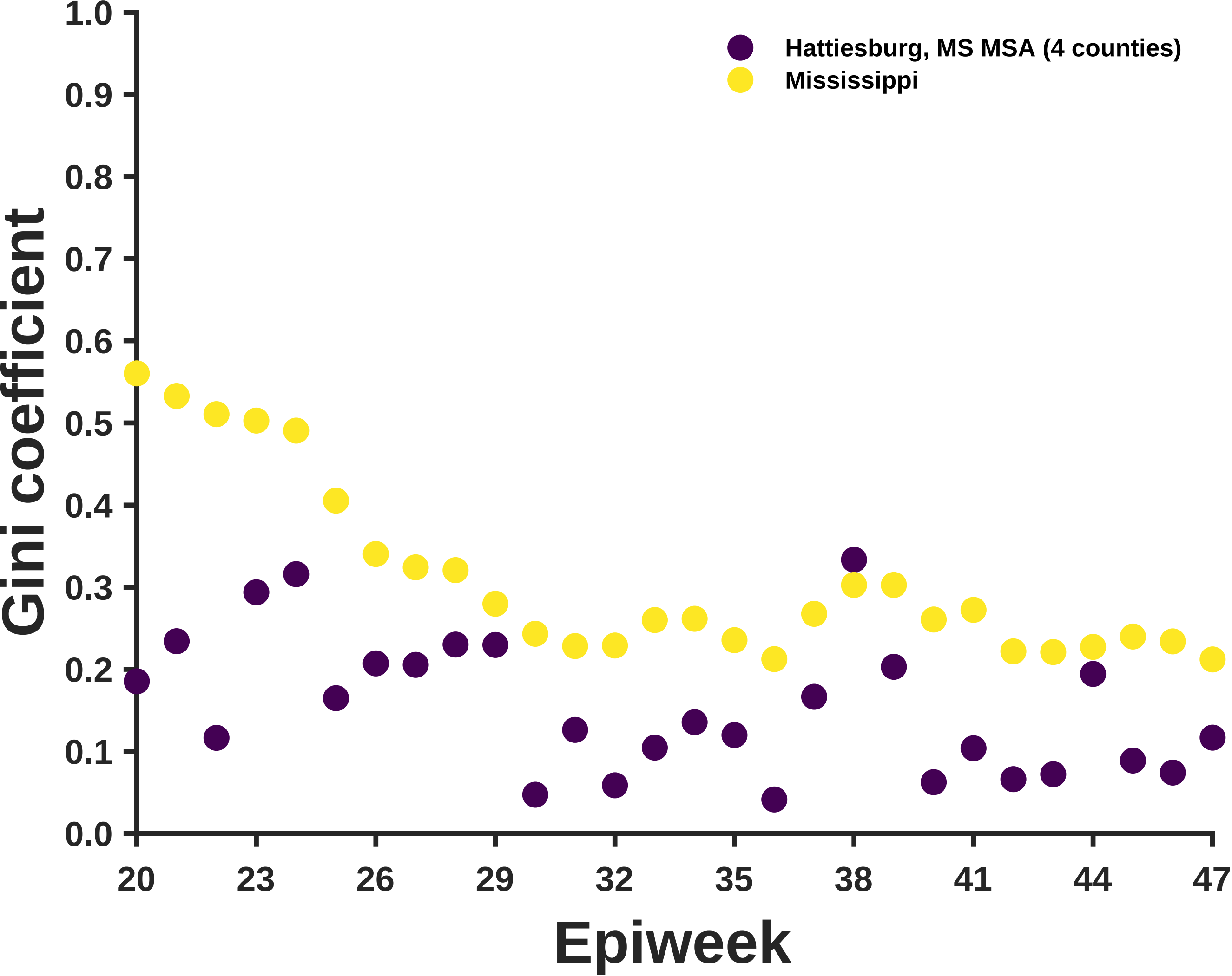

Hattiesburg, MS MSA (4 counties)
Mississippi
Gini coefficient
1.0
0.9
0.8
0.7
0.6
0.5
0.4
0.3
0.2
0.1
0.0
20
23
26
29
32
35
38
41
44
47
Epiweek

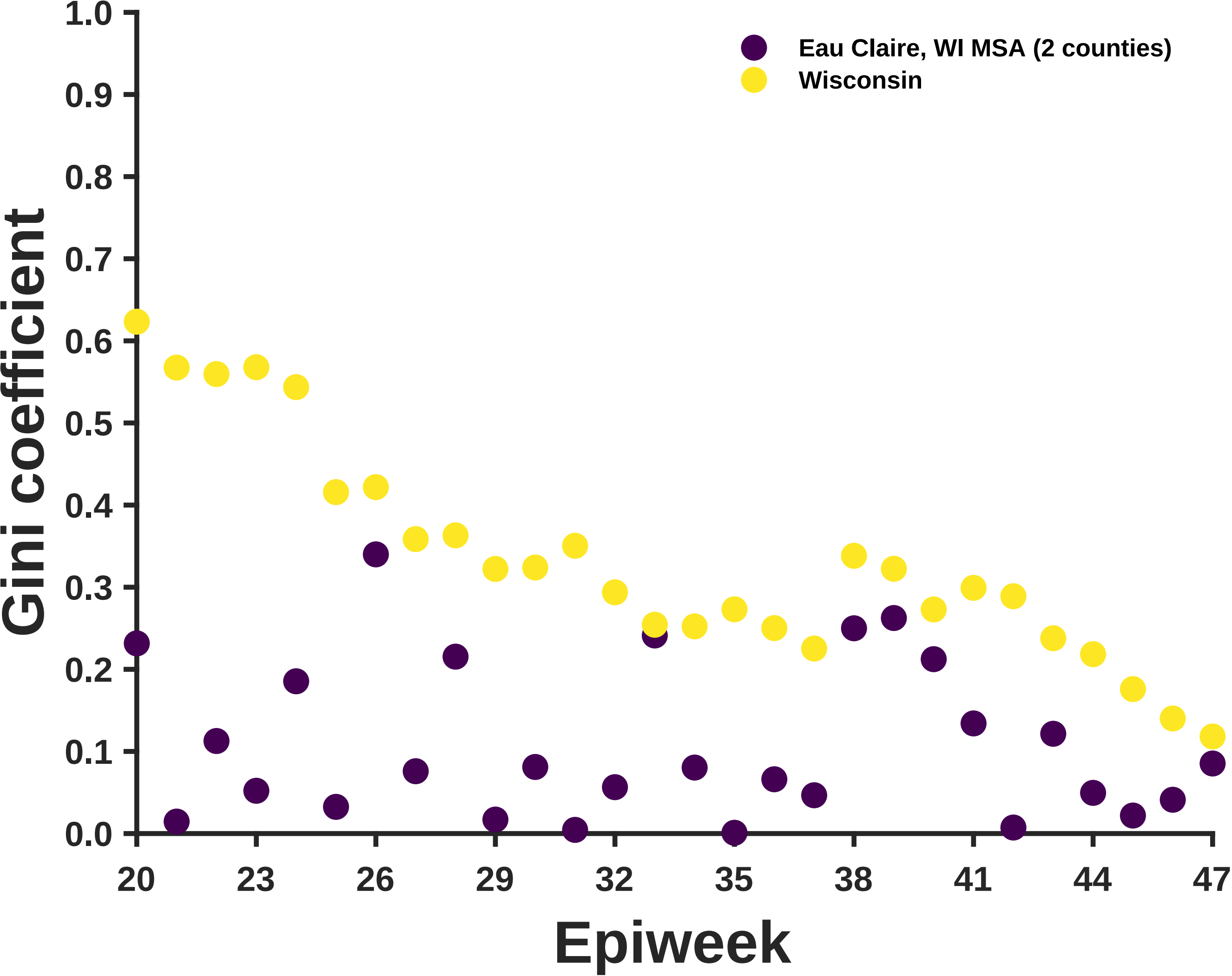

Eau Claire, WI MSA (2 counties)
Wisconsin
Gini coefficient
1.0
0.9
0.8
0.7
0.6
0.5
0.4
0.3
0.2
0.1
0.0
20
23
26
29
32
35
38
41
44
47
Epiweek

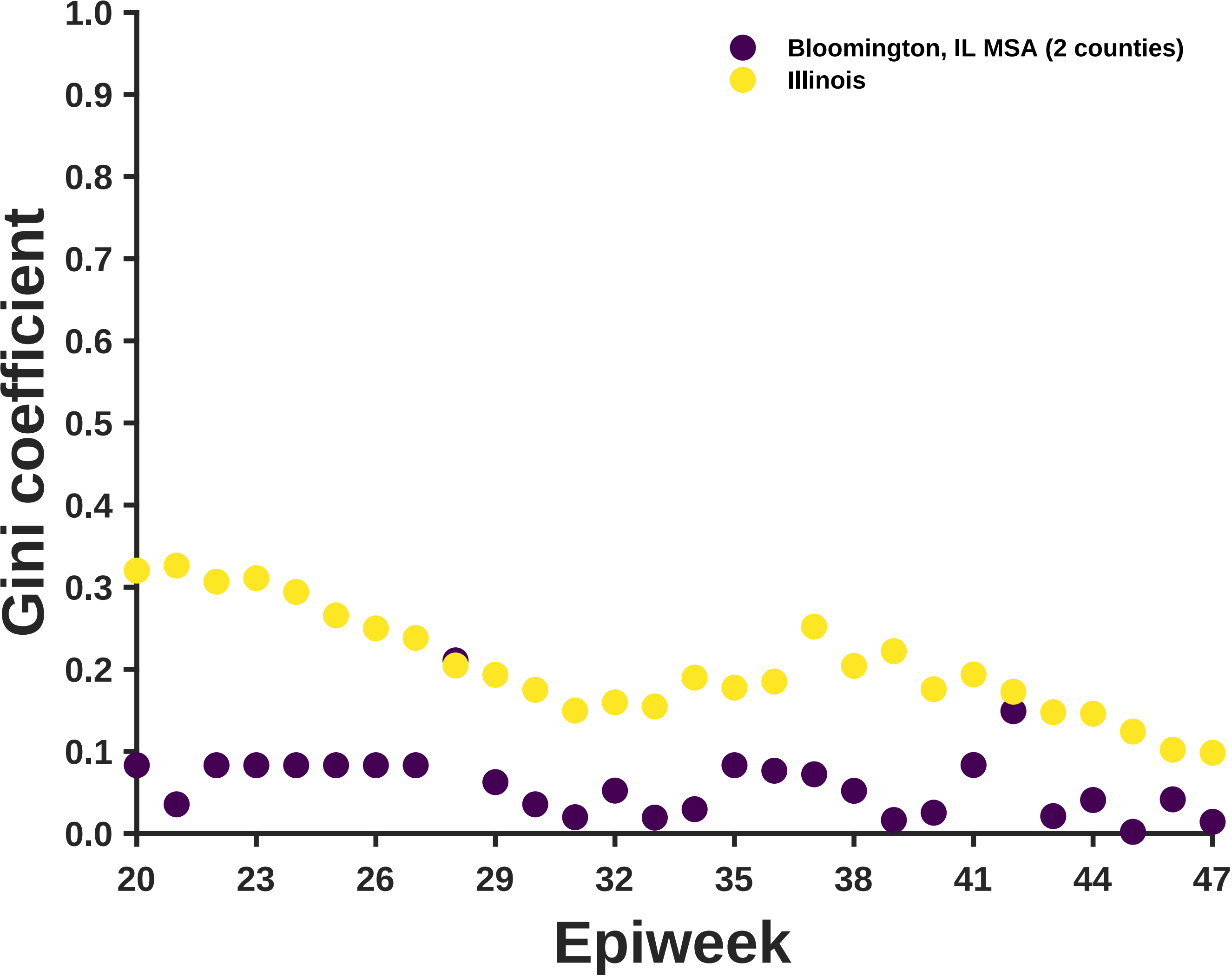

Bloomington, IL MSA (2 counties)
Illinois
Gini coefficient
1.0
0.9
0.8
0.7
0.6
0.5
0.4
0.3
0.2
0.1
0.0
20
23
26
29
32
35
38
41
44
47
Epiweek

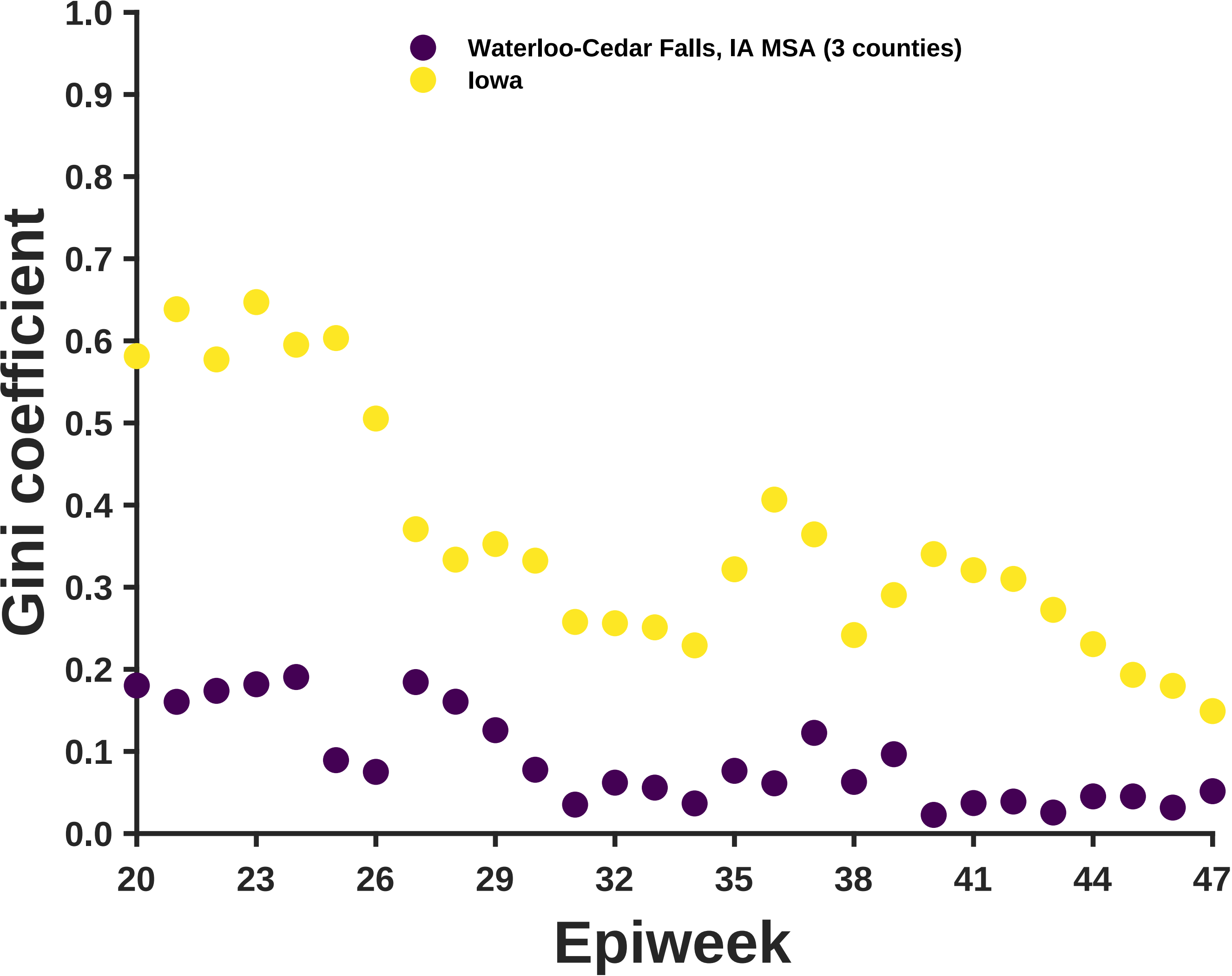

Waterloo-Cedar Falls, IA MSA (3 counties)
Iowa
Gini coefficient
1.0
0.9
0.8
0.7
0.6
0.5
0.4
0.3
0.2
0.1
0.0
20
23
26
29
32
35
38
41
44
47
Epiweek

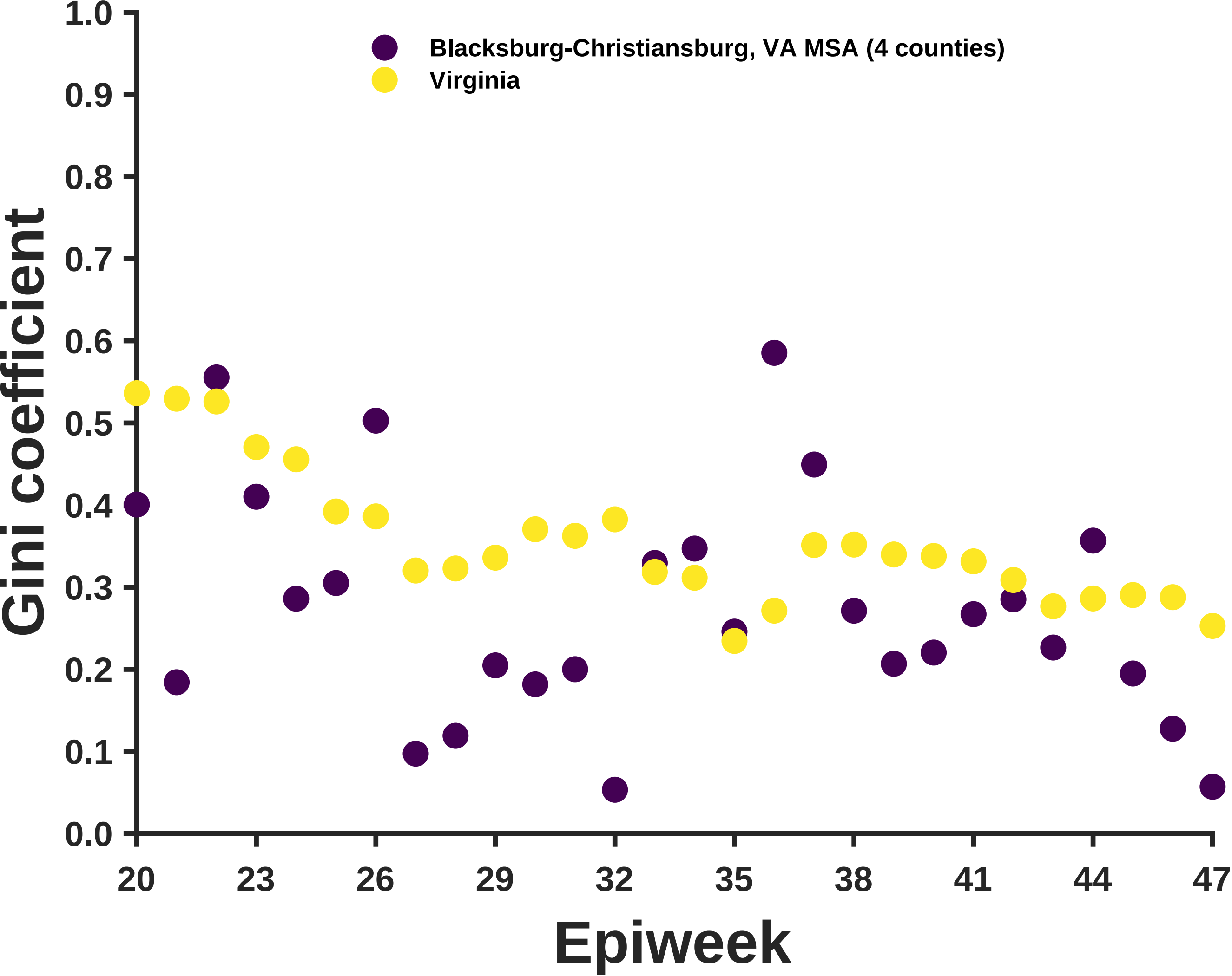

Blacksburg-Christiansburg, VA MSA (4 counties)
Virginia
Gini coefficient
Epiweek
1.0
0.9
0.8
0.7
0.6
0.5
0.4
0.3
0.2
0.1
0.0
20
23
26
29
32
35
38
41
44
47

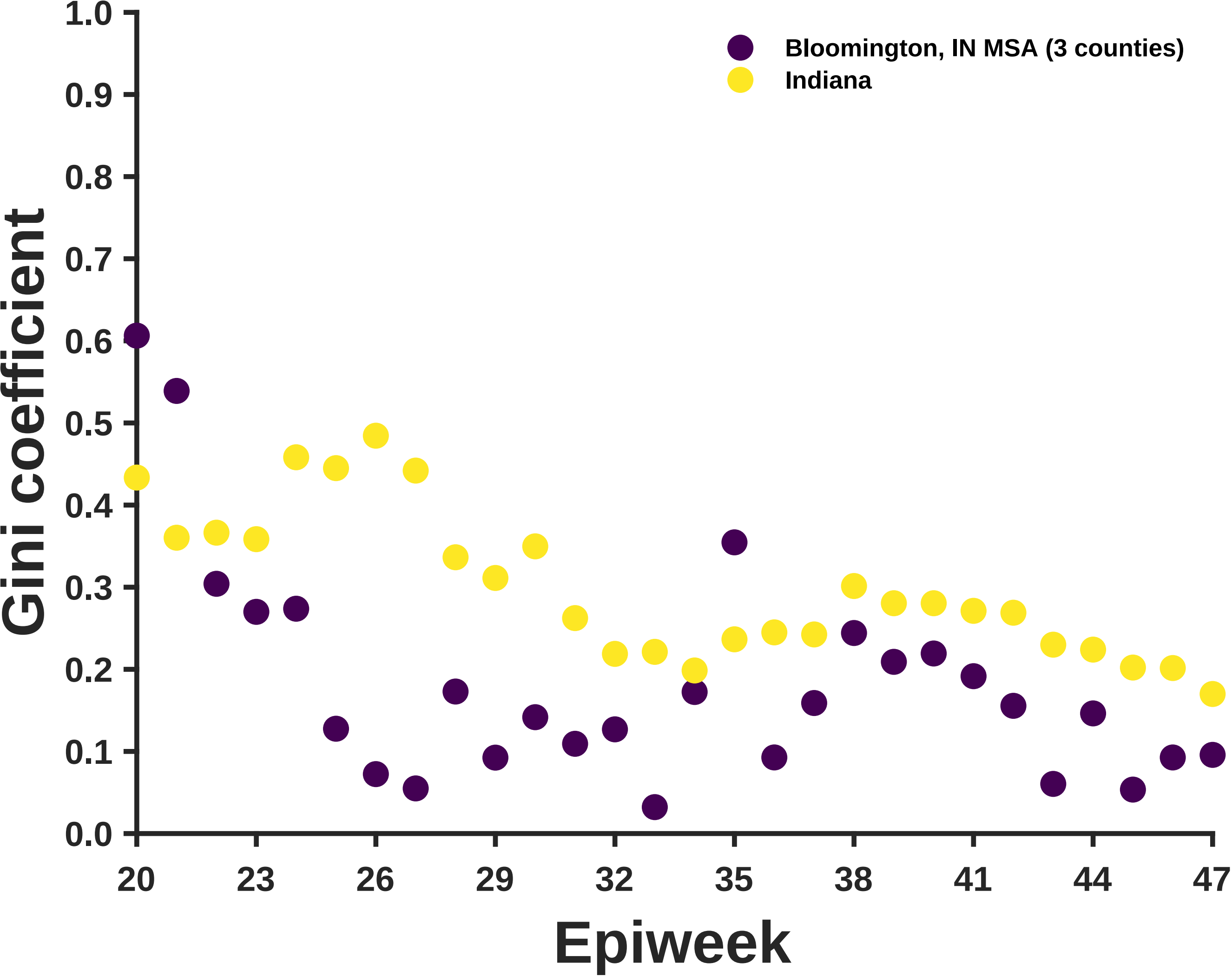

Bloomington, IN MSA (3 counties)
Indiana
Gini coefficient
1.0
0.9
0.8
0.7
0.6
0.5
0.4
0.3
0.2
0.1
0.0
20
23
26
29
32
35
38
41
44
47
Epiweek

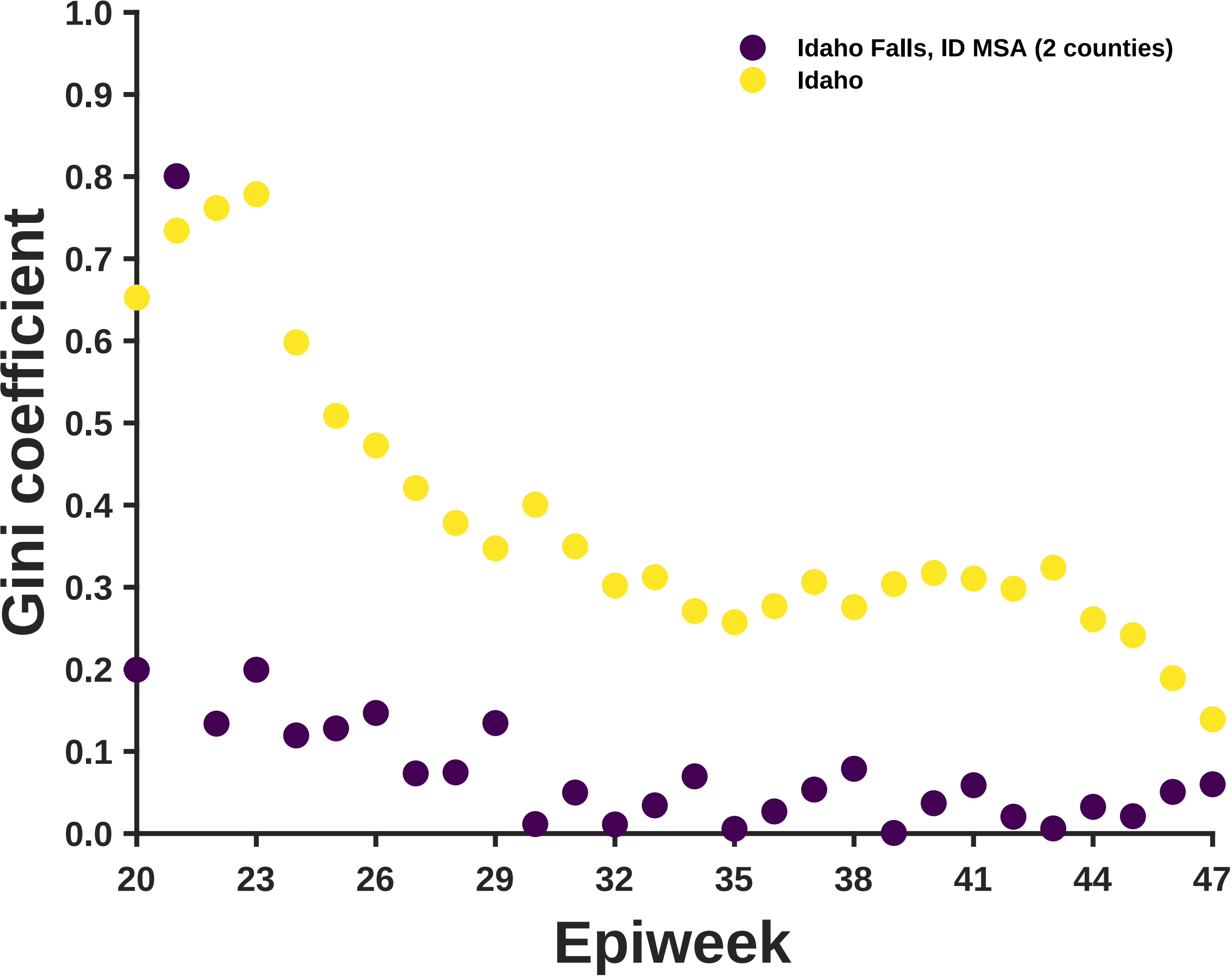
Idaho Falls, ID MSA (2 counties)
Idaho
Gini coefficient
1.0
0.9
0.8
0.7
0.6
0.5
0.4
0.3
0.2
0.1
0.0
20
23
26
29
32
35
38
41
44
47
Epiweek

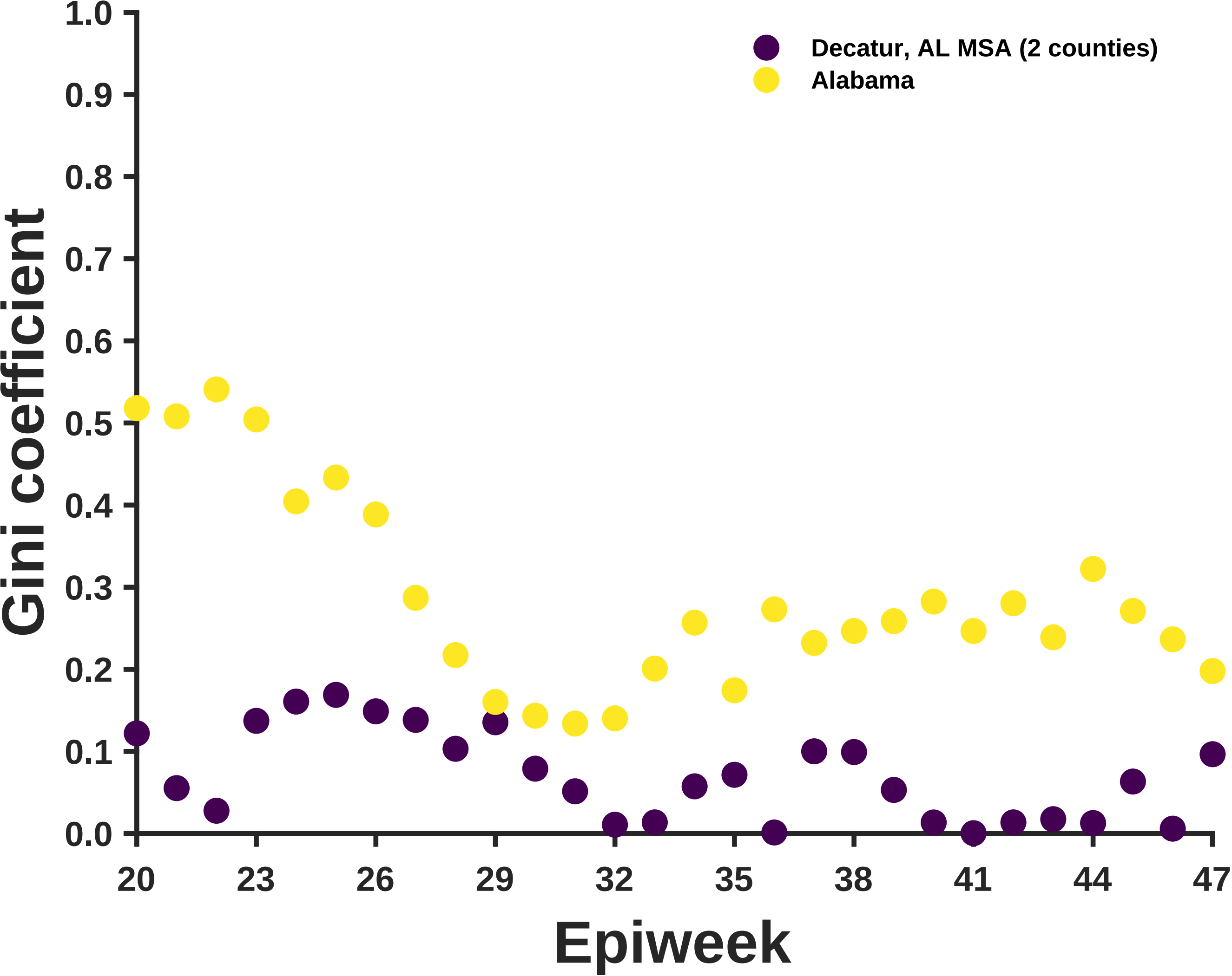
Decatur, AL MSA (2 counties)
Alabama
Gini coefficient
Epiweek
0.0
0.1
0.2
0.3
0.4
0.5
0.6
0.7
0.8
0.9
1.0
20
23
26
29
32
35
38
41
44
47

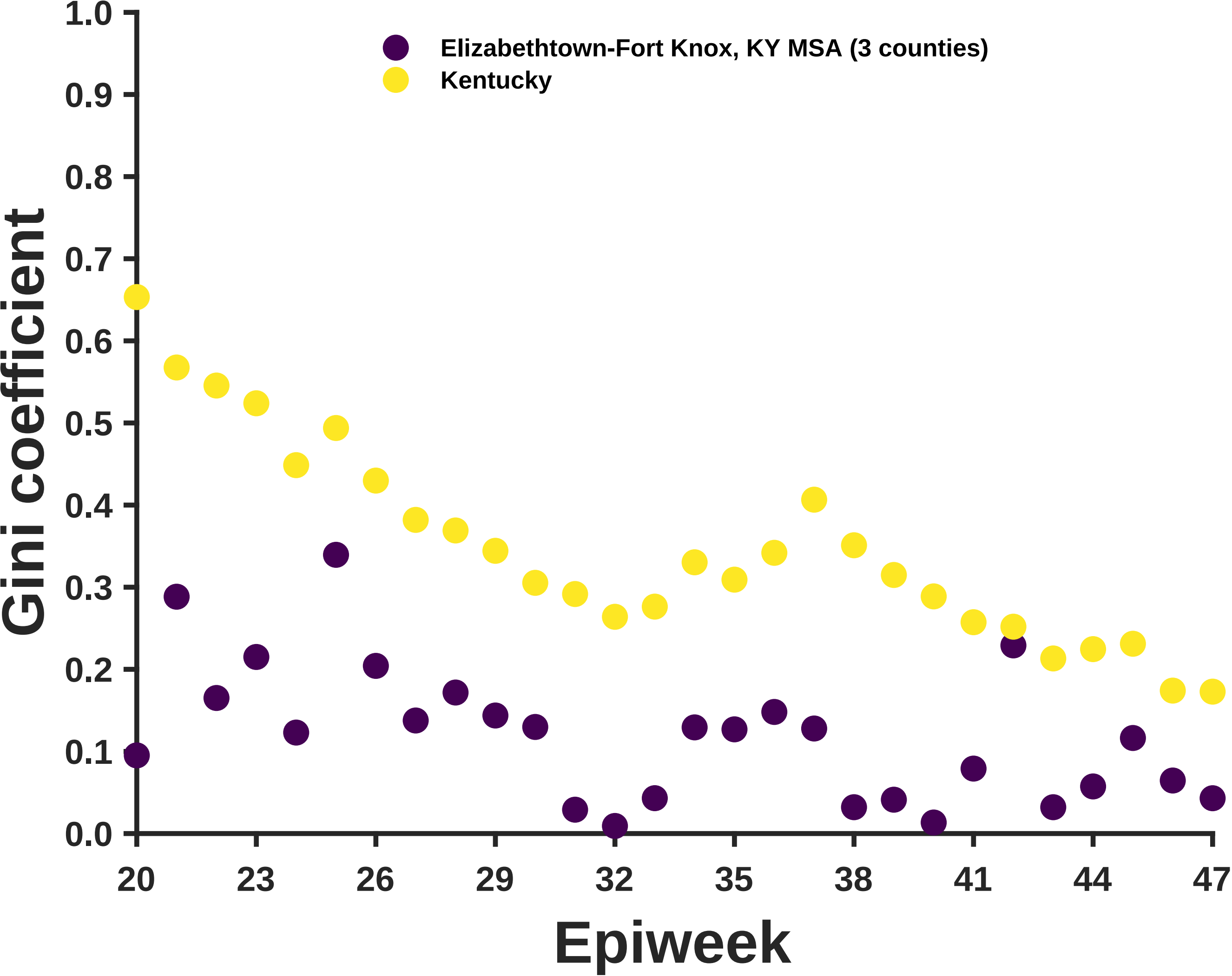

Elizabethtown-Fort Knox, KY MSA (3 counties)
Kentucky
Gini coefficient
Epiweek
0.0
0.1
0.2
0.3
0.4
0.5
0.6
0.7
0.8
0.9
1.0
20
23
26
29
32
35
38
41
44
47

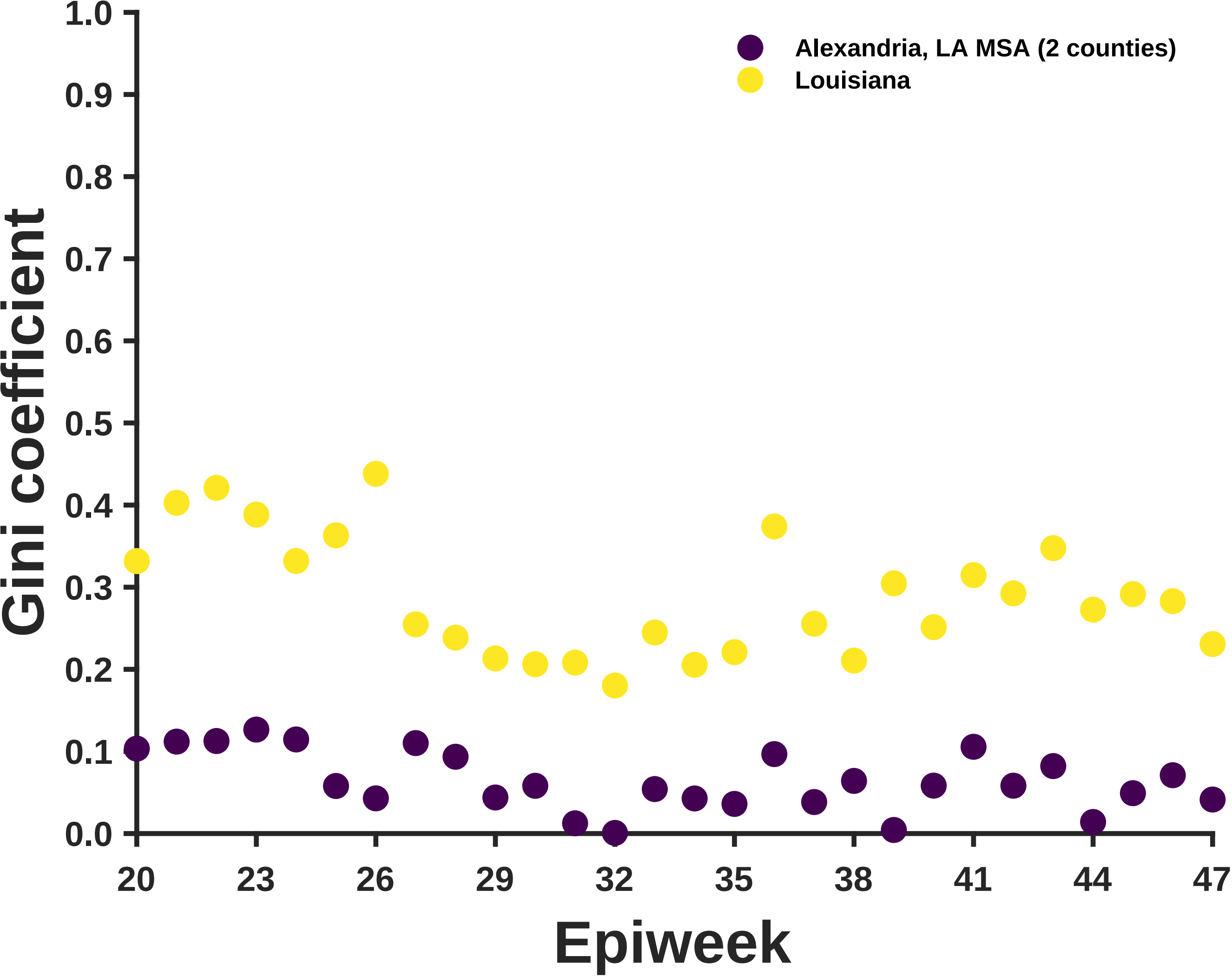

Alexandria, LA MSA (2 counties)
Louisiana
Gini coefficient
Epiweek
1.0
0.9
0.8
0.7
0.6
0.5
0.4
0.3
0.2
0.1
0.0
20
23
26
29
32
35
38
41
44
47

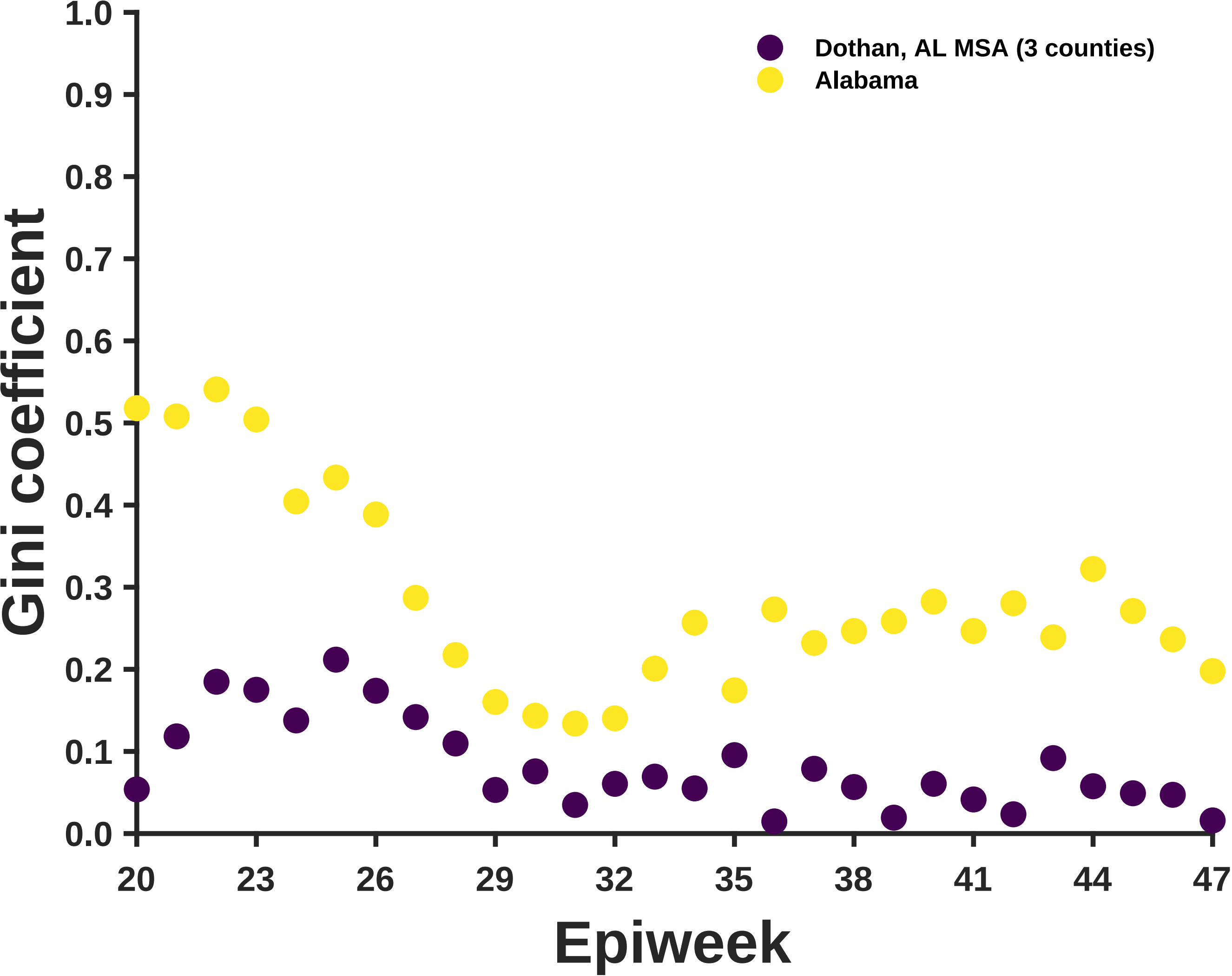

Dothan, AL MSA (3 counties)
Alabama
Gini coefficient
0.0
0.1
0.2
0.3
0.4
0.5
0.6
0.7
0.8
0.9
1.0
20
23
26
29
32
35
38
41
44
47
Epiweek

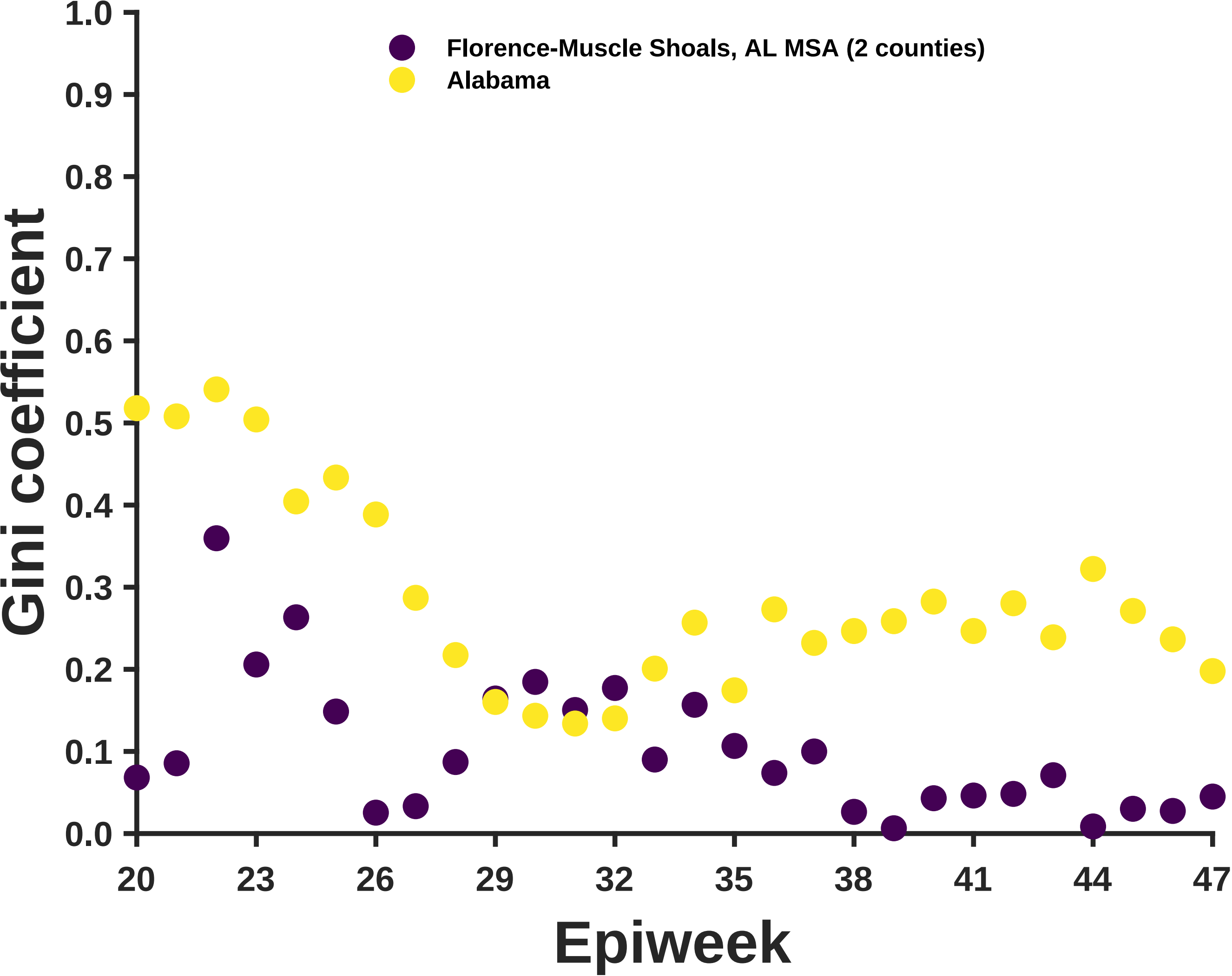

Florence-Muscle Shoals, AL MSA (2 counties)
Alabama
Gini coefficient
1.0
0.9
0.8
0.7
0.6
0.5
0.4
0.3
0.2
0.1
0.0
20
23
26
29
32
35
38
41
44
47
Epiweek

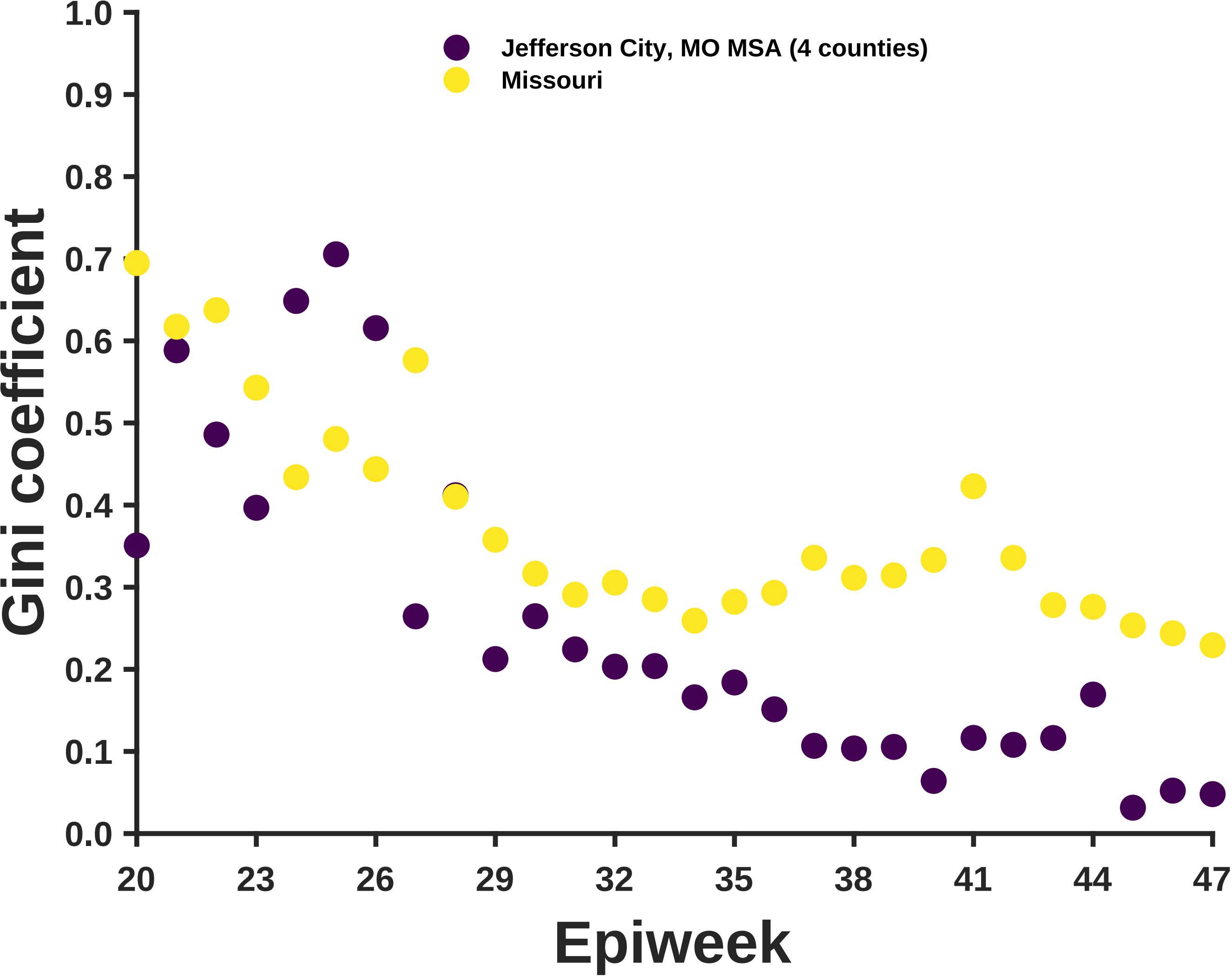

Jefferson City, MO MSA (4 counties)
Missouri
Gini coefficient
Epiweek
1.0
0.9
0.8
0.7
0.6
0.5
0.4
0.3
0.2
0.1
0.0
20
23
26
29
32
35
38
41
44
47

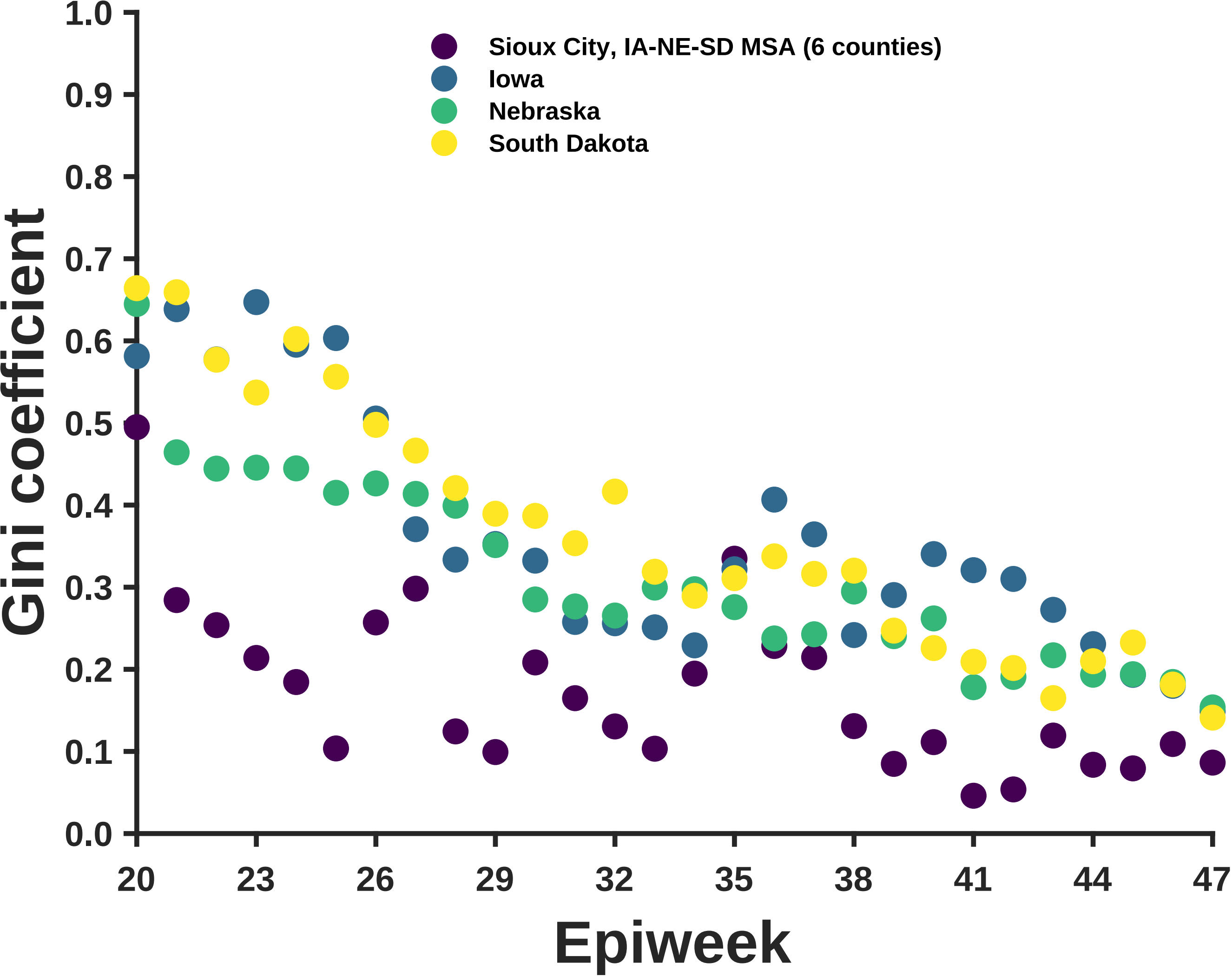
Sioux City, IA-NE-SD MSA (6 counties)
Iowa
Nebraska
South Dakota
Gini coefficient
1.0
0.9
0.8
0.7
0.6
0.5
0.4
0.3
0.2
0.1
0.0
20
23
26
29
32
35
38
41
44
47
Epiweek

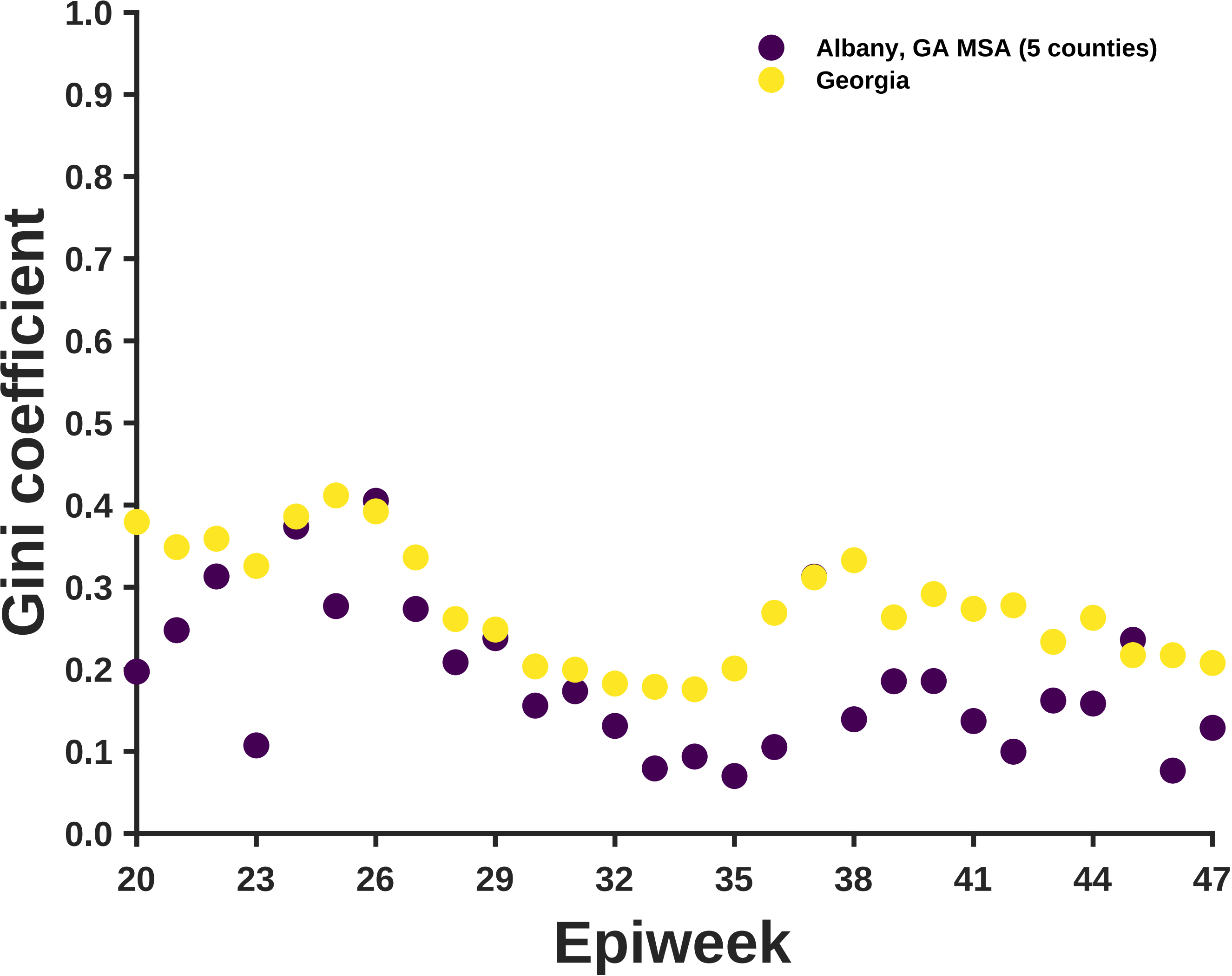

Albany, GA MSA (5 counties)
Georgia
Gini coefficient
1.0
0.9
0.8
0.7
0.6
0.5
0.4
0.3
0.2
0.1
0.0
20
23
26
29
32
35
38
41
44
47
Epiweek

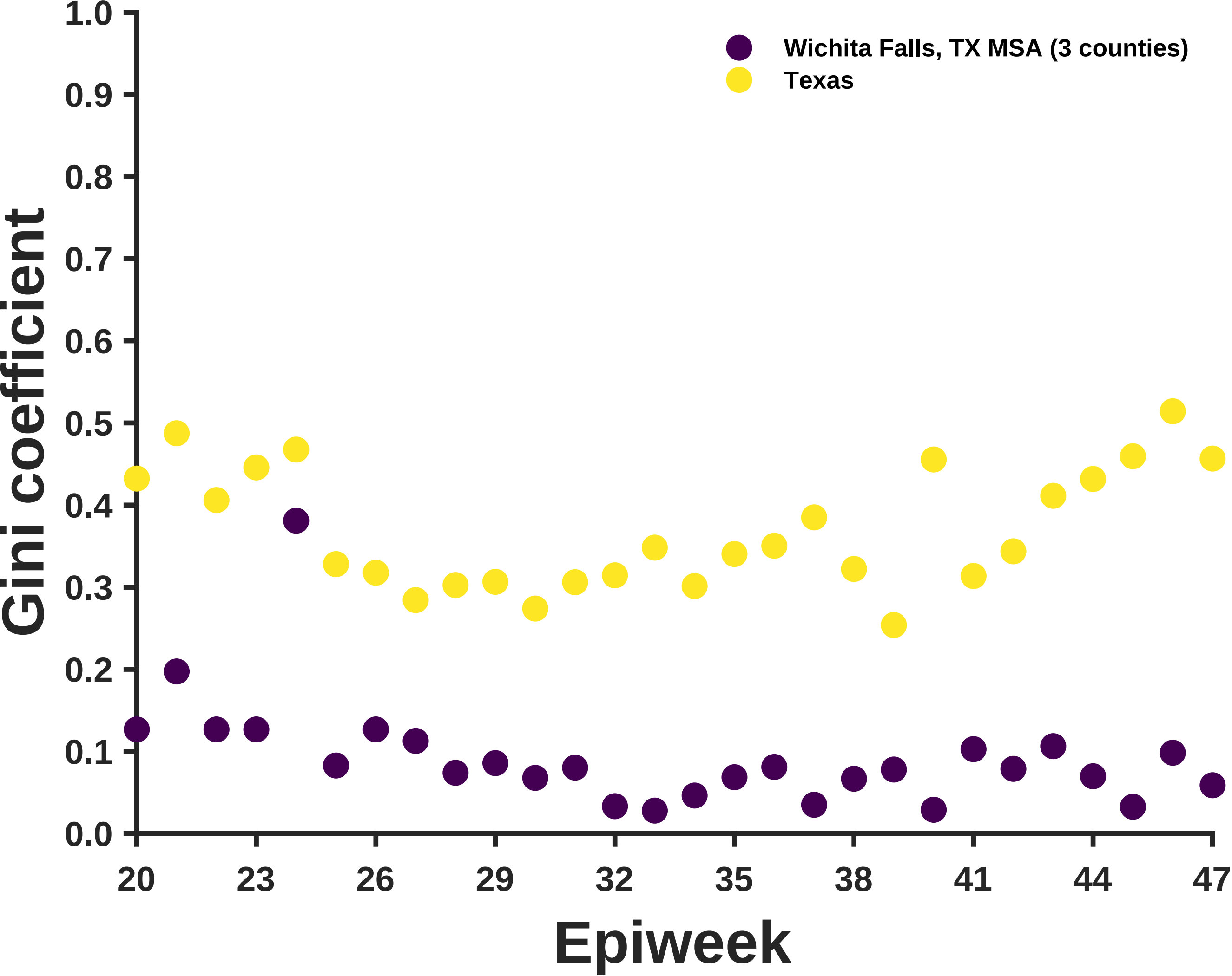
Wichita Falls, TX MSA (3 counties)
Texas
Gini coefficient
Epiweek
0.0
0.1
0.2
0.3
0.4
0.5
0.6
0.7
0.8
0.9
1.0
20
23
26
29
32
35
38
41
44
47

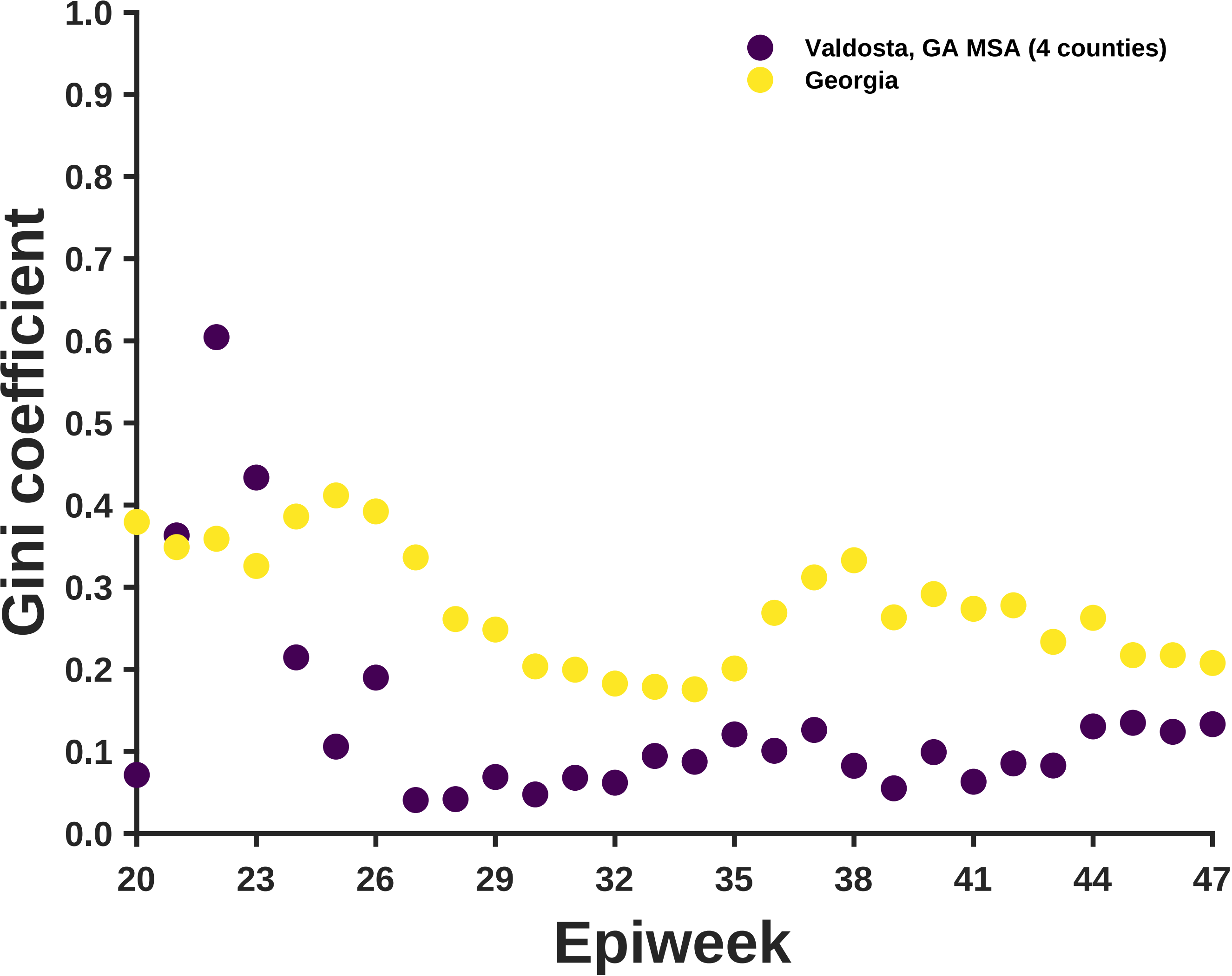

Valdosta, GA MSA (4 counties)
Georgia
Gini coefficient
1.0
0.9
0.8
0.7
0.6
0.5
0.4
0.3
0.2
0.1
0.0
20
23
26
29
32
35
38
41
44
47
Epiweek

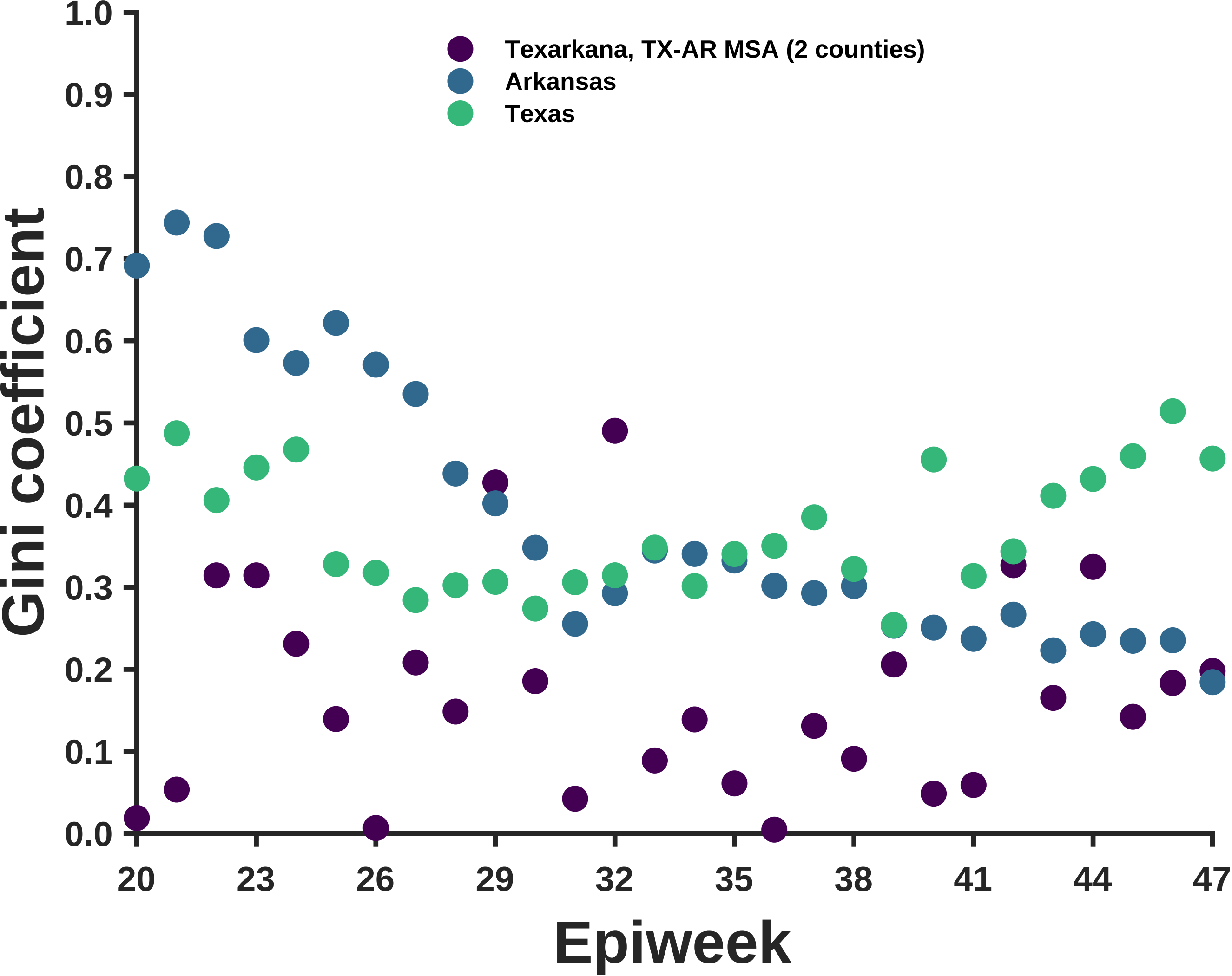

Texarkana, TX-AR MSA (2 counties)
Arkansas
Texas
Gini coefficient
Epiweek
1.0
0.9
0.8
0.7
0.6
0.5
0.4
0.3
0.2
0.1
0.0
20
23
26
29
32
35
38
41
44
47

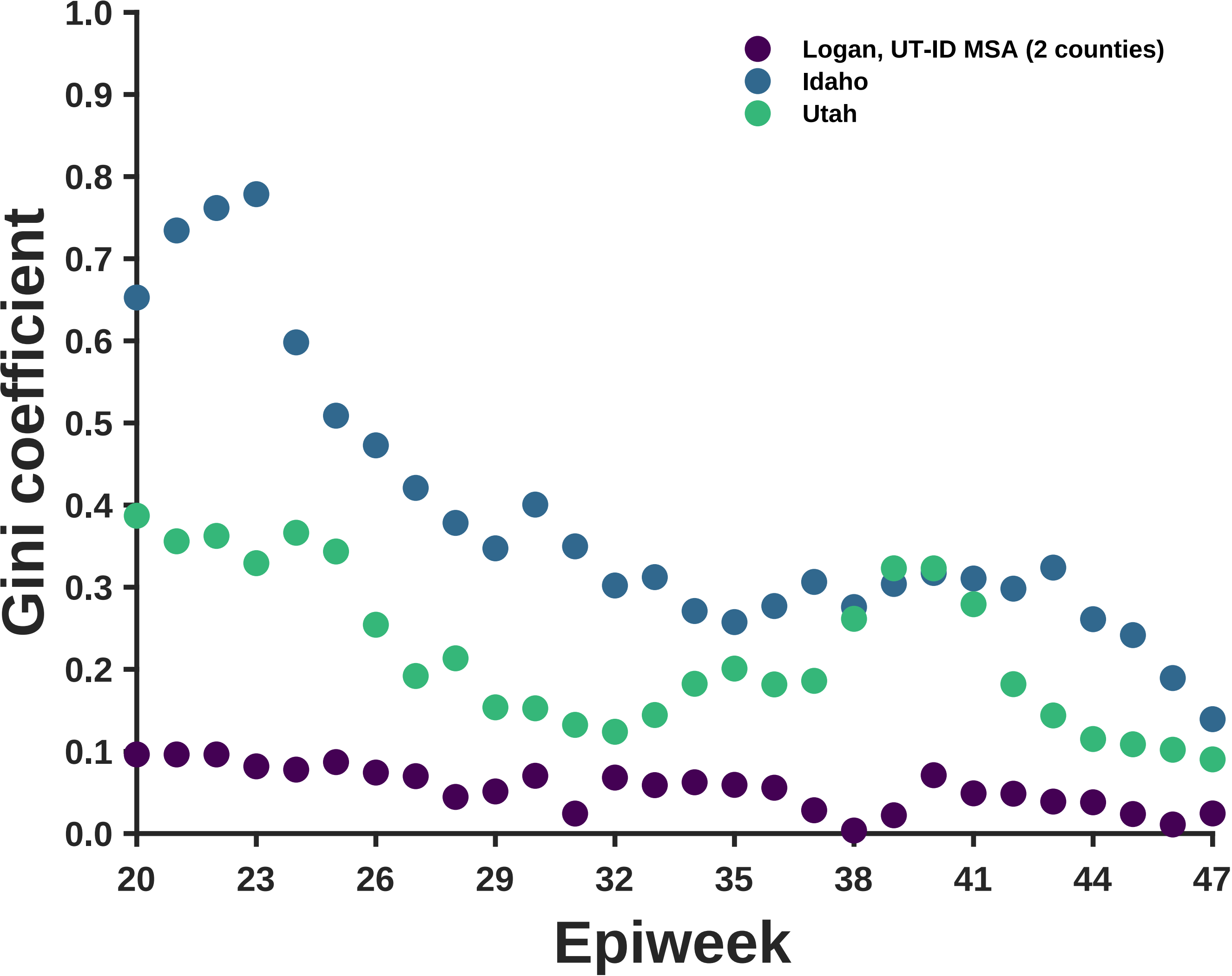

Logan, UT-ID MSA (2 counties)
Idaho
Utah
Gini coefficient
Epiweek
0.0
0.1
0.2
0.3
0.4
0.5
0.6
0.7
0.8
0.9
1.0
20
23
26
29
32
35
38
41
44
47

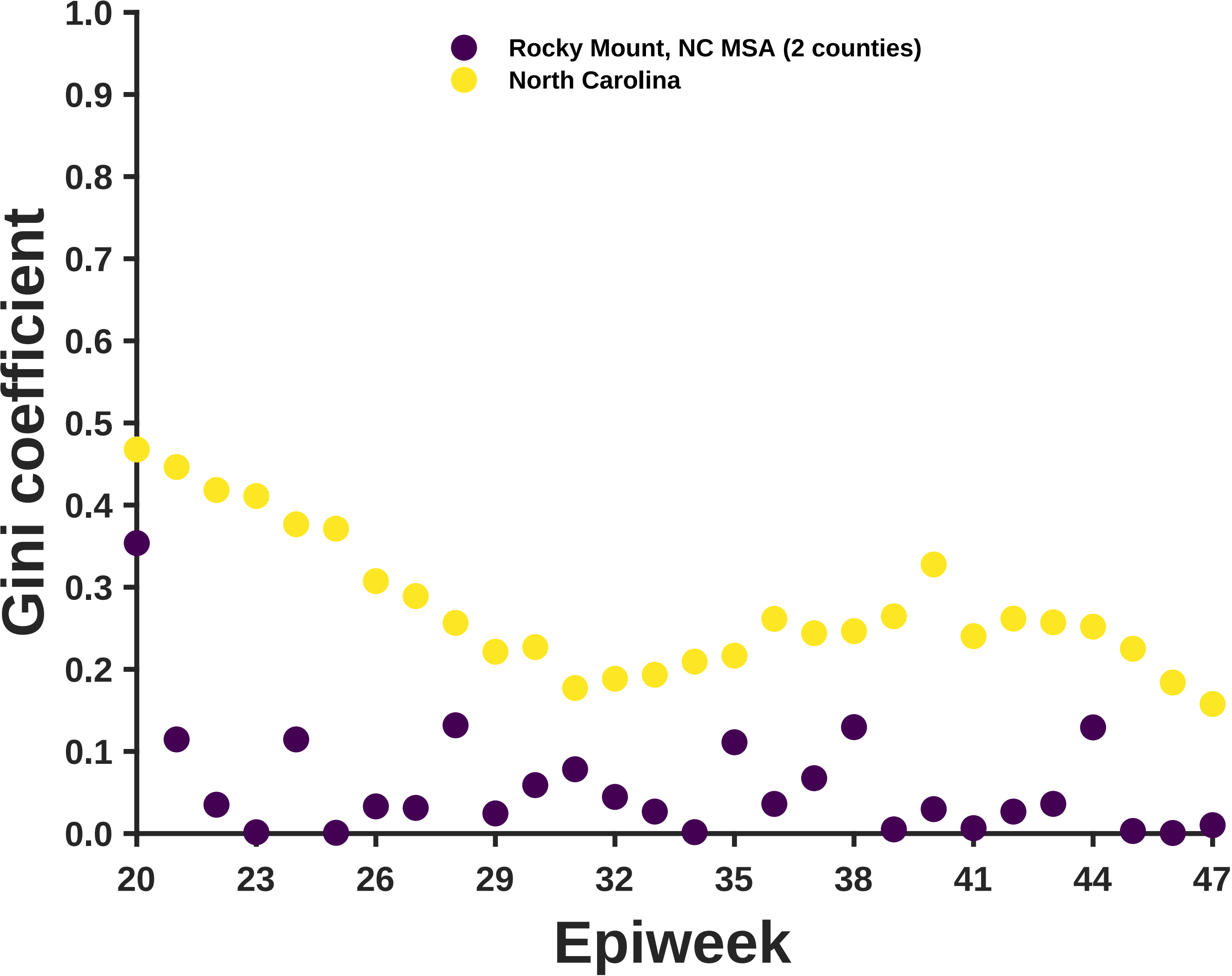

Rocky Mount, NC MSA (2 counties)
North Carolina
Gini coefficient
1.0
0.9
0.8
0.7
0.6
0.5
0.4
0.3
0.2
0.1
0.0
20
23
26
29
32
35
38
41
44
47
Epiweek

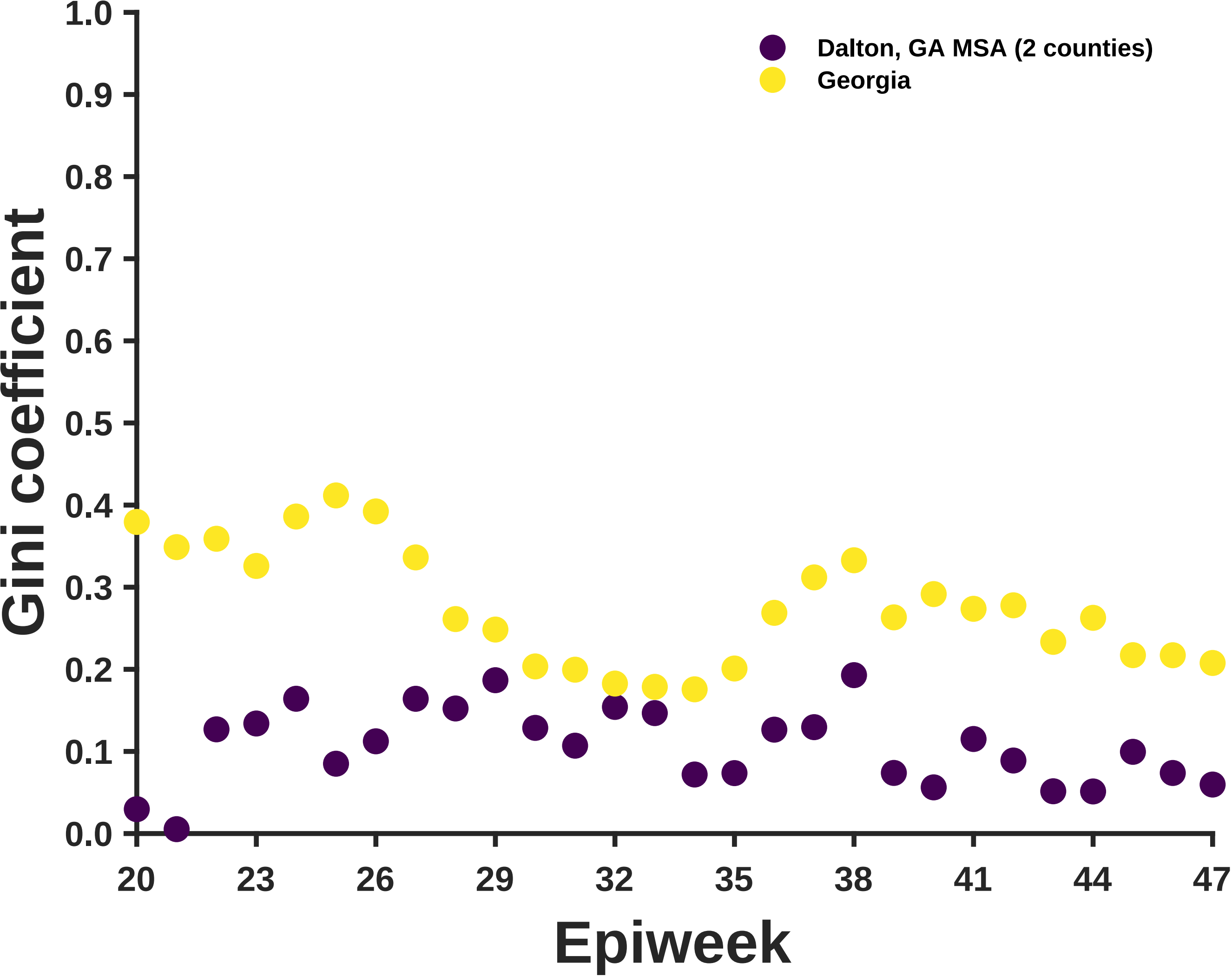

Dalton, GA MSA (2 counties)
Georgia
Gini coefficient
1.0
0.9
0.8
0.7
0.6
0.5
0.4
0.3
0.2
0.1
0.0
20
23
26
29
32
35
38
41
44
47
Epiweek

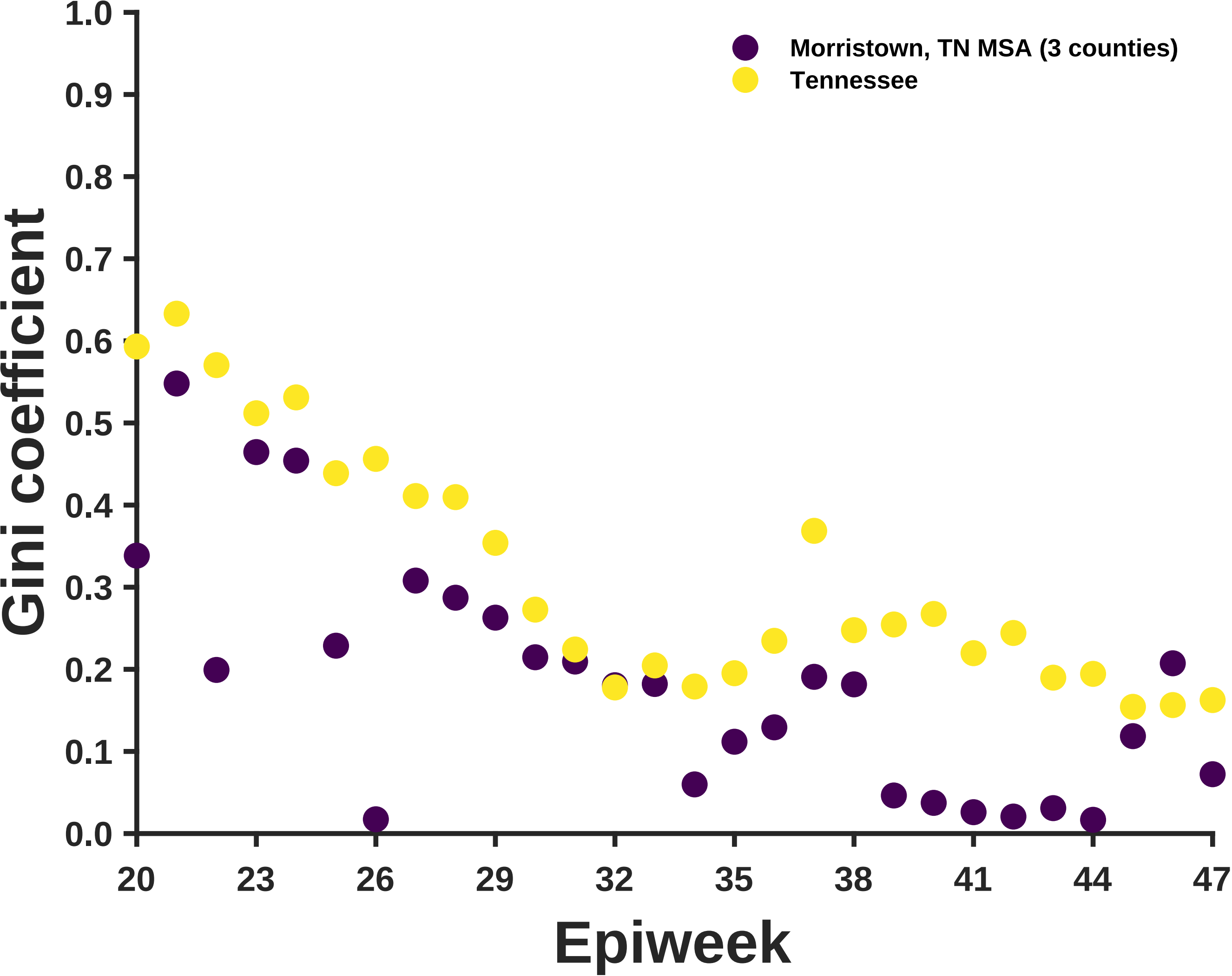

Morristown, TN MSA (3 counties)
Tennessee
Gini coefficient
1.0
0.9
0.8
0.7
0.6
0.5
0.4
0.3
0.2
0.1
0.0
20
23
26
29
32
35
38
41
44
47
Epiweek

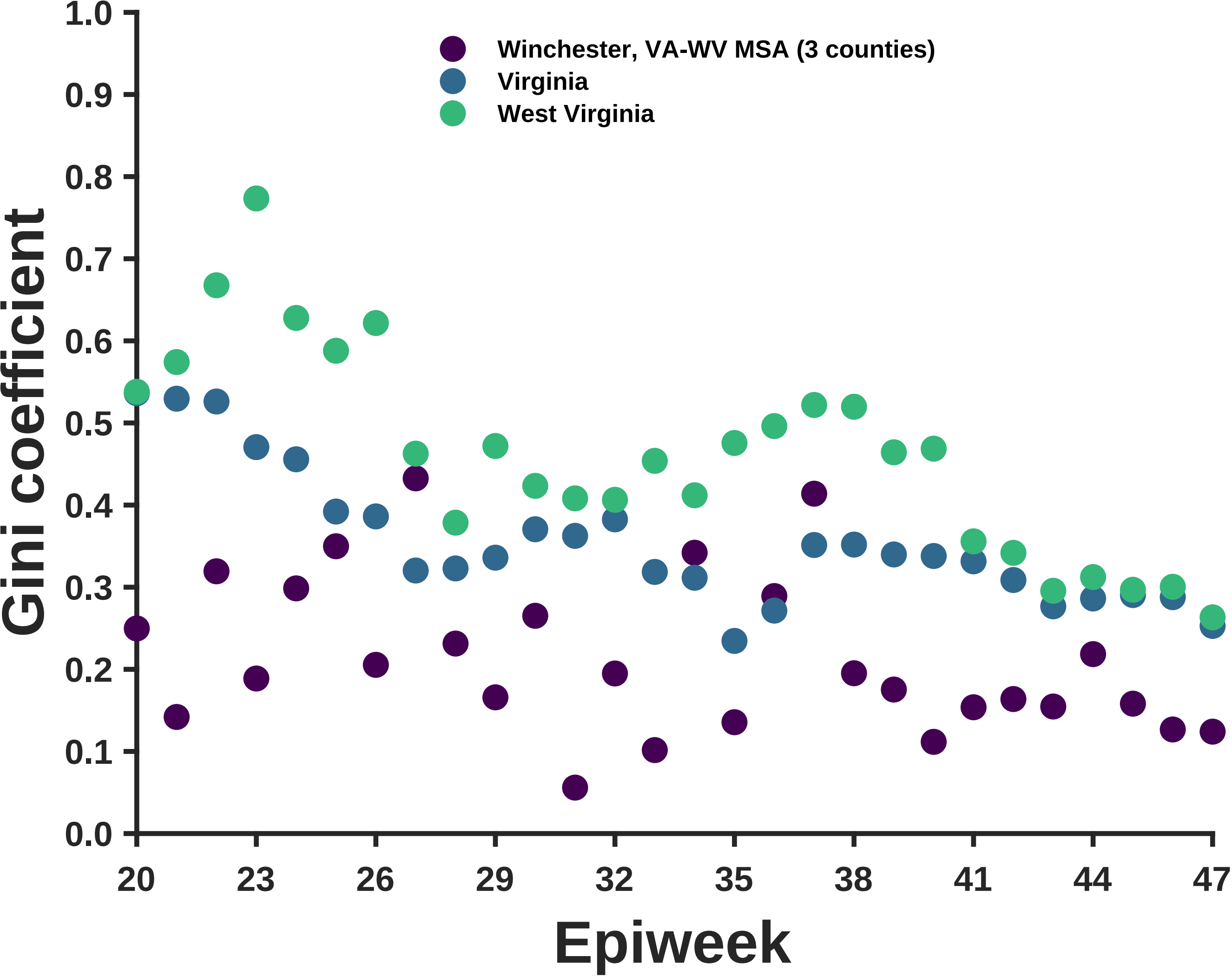
Winchester, VA-WV MSA (3 counties)
Virginia
West Virginia
Gini coefficient
0.0
0.1
0.2
0.3
0.4
0.5
0.6
0.7
0.8
0.9
1.0
20
23
26
29
32
35
38
41
44
47
Epiweek

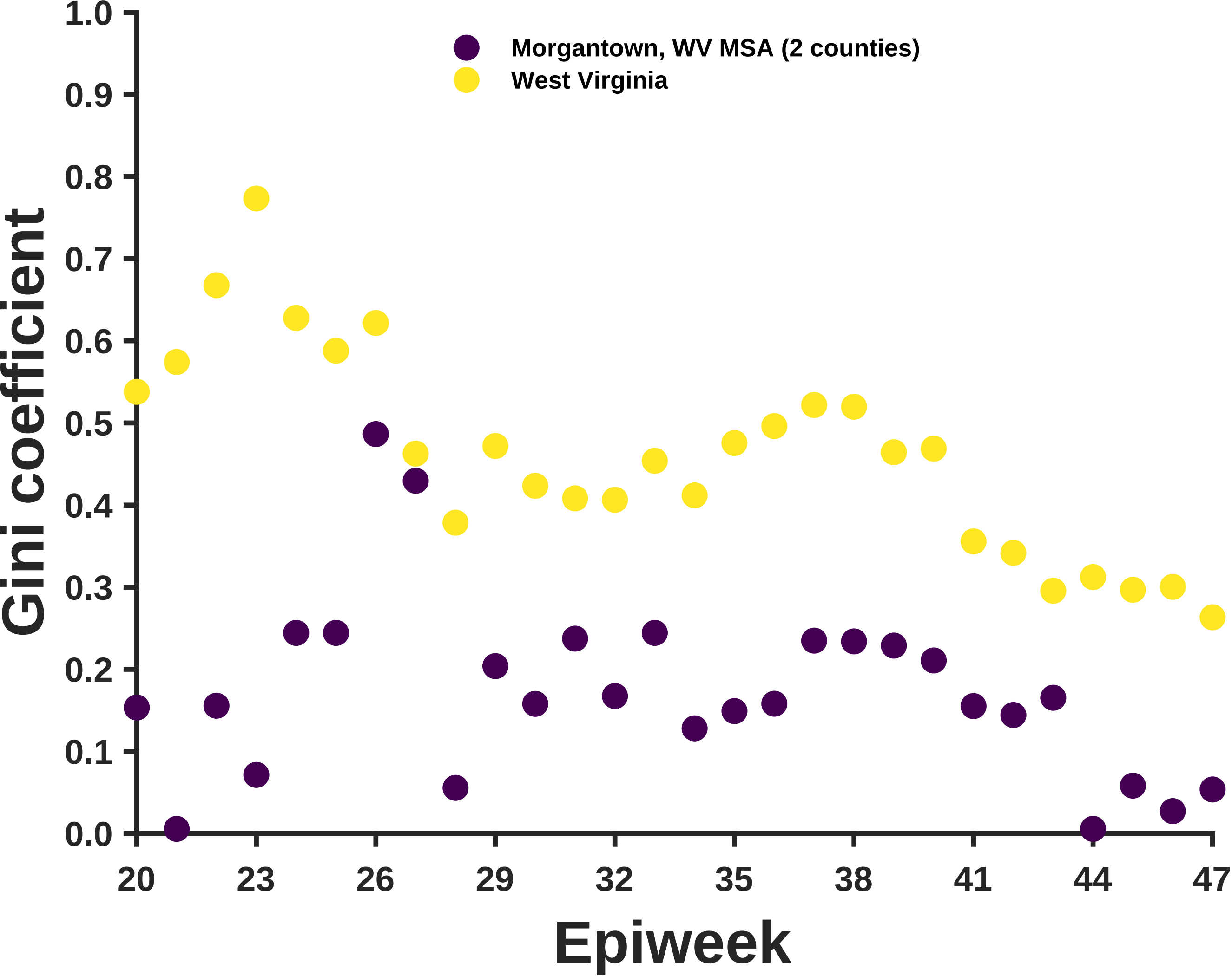

Morgantown, WV MSA (2 counties)
West Virginia
Gini coefficient
Epiweek
1.0
0.9
0.8
0.7
0.6
0.5
0.4
0.3
0.2
0.1
0.0
20
23
26
29
32
35
38
41
44
47

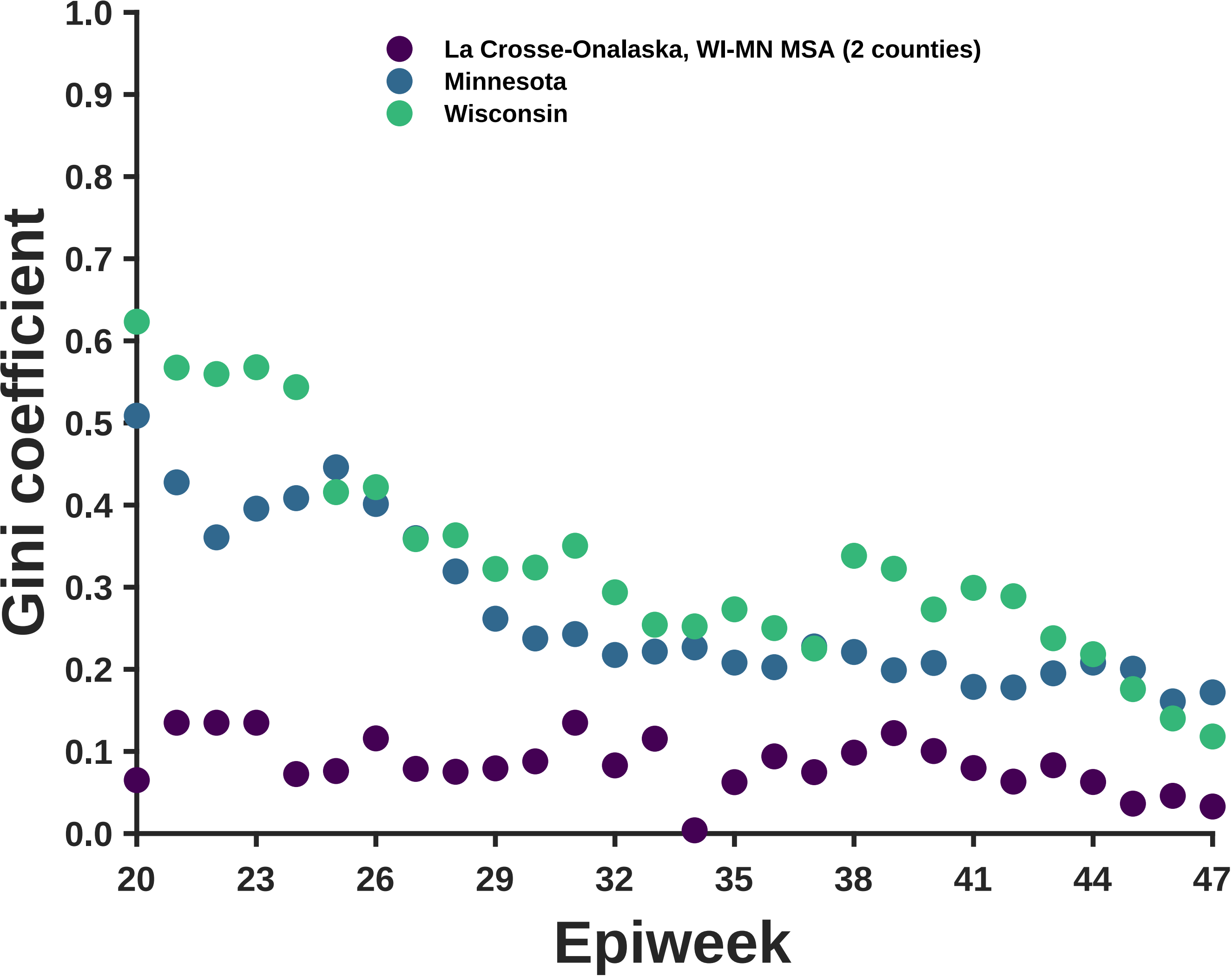

La Crosse-Onalaska, WI-MN MSA (2 counties)
Minnesota
Wisconsin
Gini coefficient
0.0
0.1
0.2
0.3
0.4
0.5
0.6
0.7
0.8
0.9
1.0
20
23
26
29
32
35
38
41
44
47
Epiweek

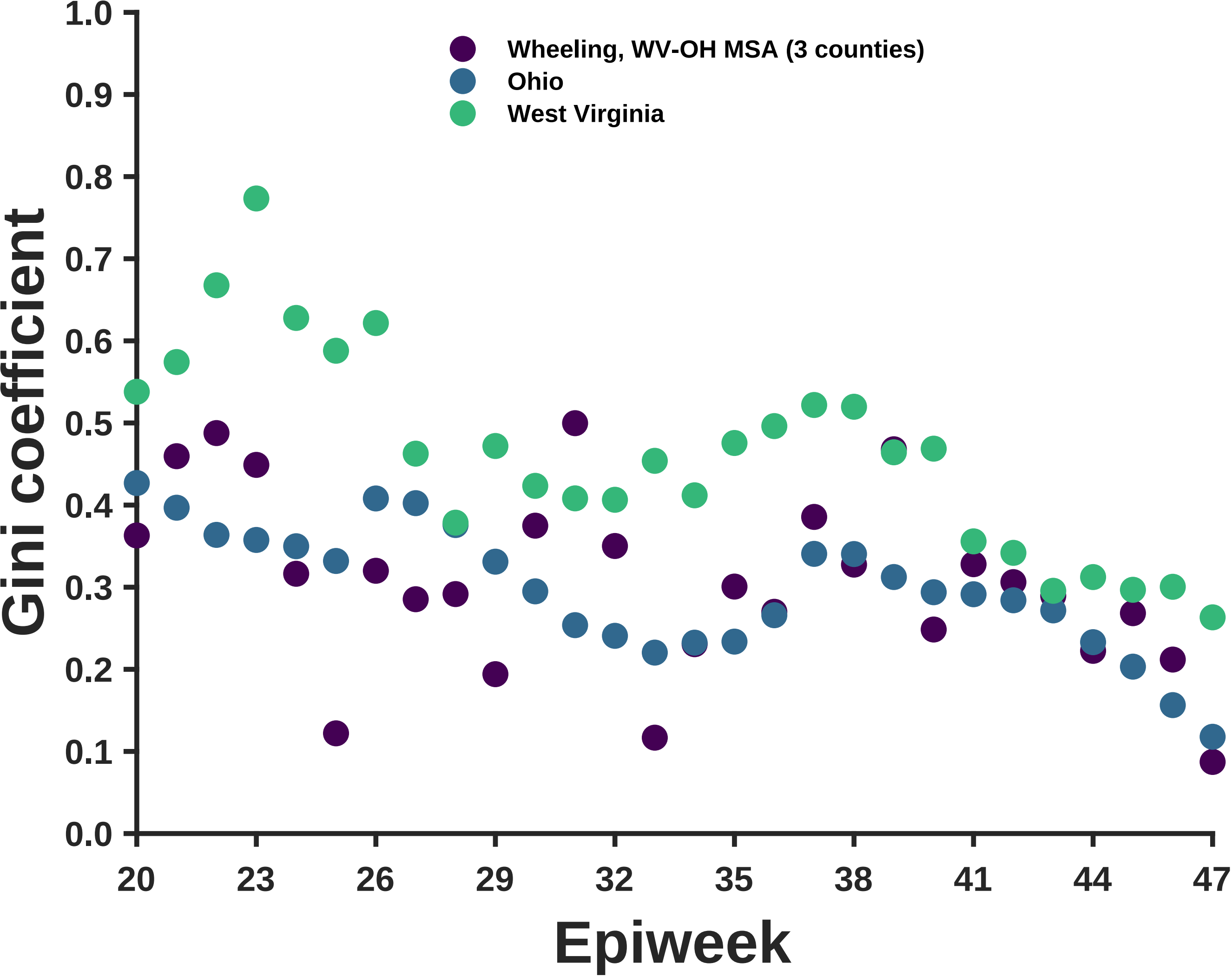

Wheeling, WV-OH MSA (3 counties)
Ohio
West Virginia
Gini coefficient
1.0
0.9
0.8
0.7
0.6
0.5
0.4
0.3
0.2
0.1
0.0
20
23
26
29
32
35
38
41
44
47
Epiweek

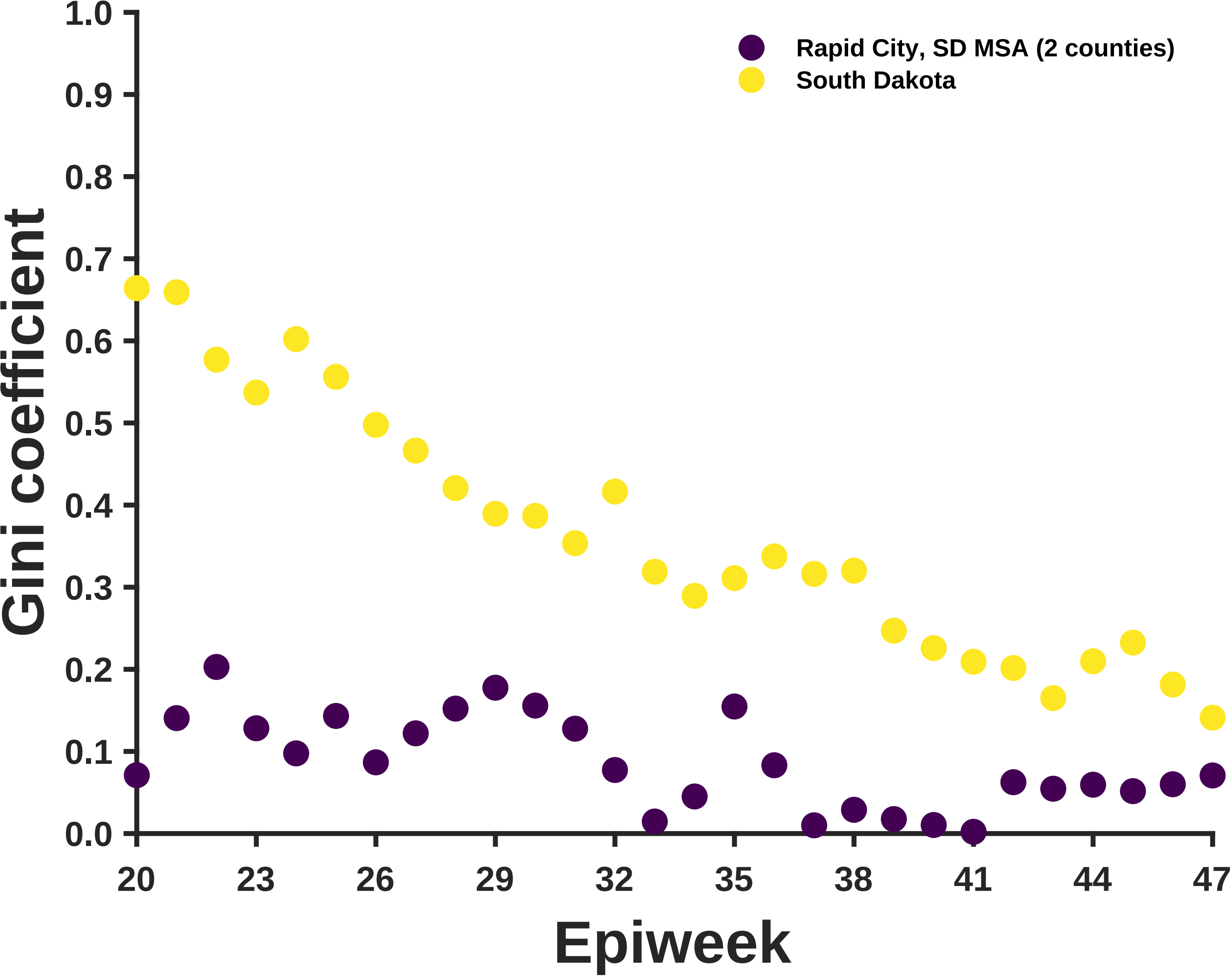

Rapid City, SD MSA (2 counties)
South Dakota
Gini coefficient
Epiweek
1.0
0.9
0.8
0.7
0.6
0.5
0.4
0.3
0.2
0.1
0.0
20
23
26
29
32
35
38
41
44
47

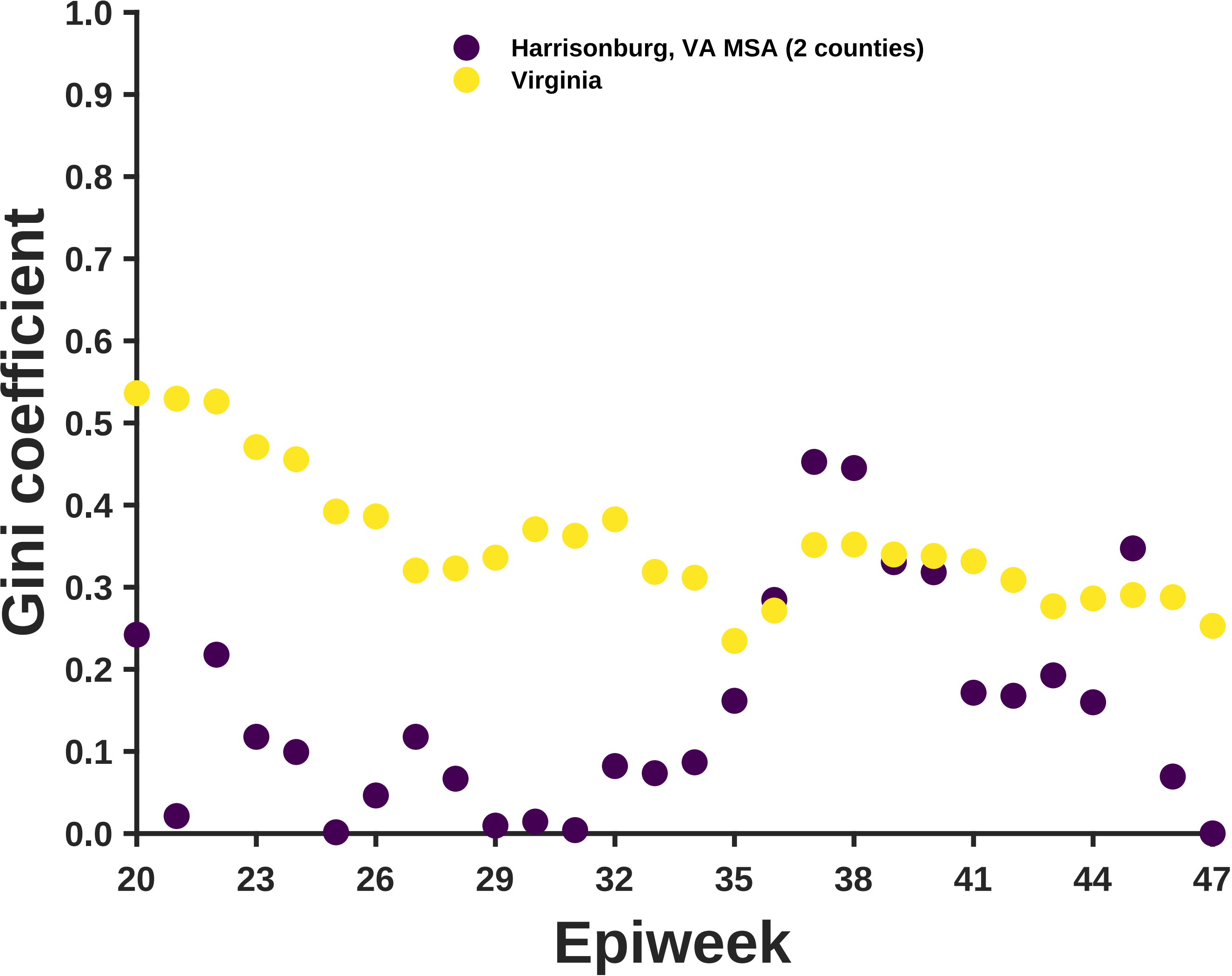
Harrisonburg, VA MSA (2 counties)
Virginia
Gini coefficient
Epiweek
0.0
0.1
0.2
0.3
0.4
0.5
0.6
0.7
0.8
0.9
1.0
20
23
26
29
32
35
38
41
44
47

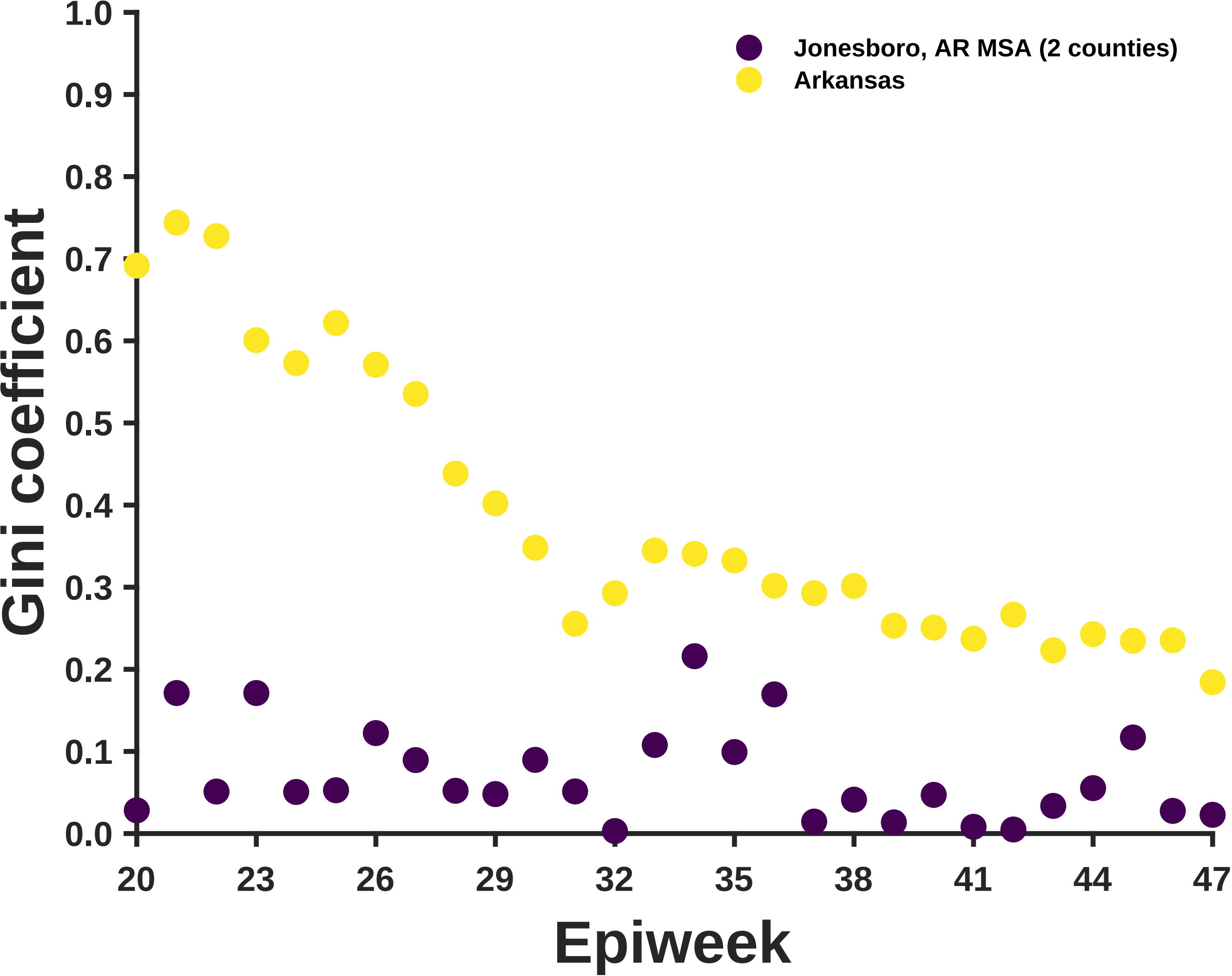
Jonesboro, AR MSA (2 counties)
Arkansas
Gini coefficient
0.0
0.1
0.2
0.3
0.4
0.5
0.6
0.7
0.8
0.9
1.0
20
23
26
29
32
35
38
41
44
47
Epiweek

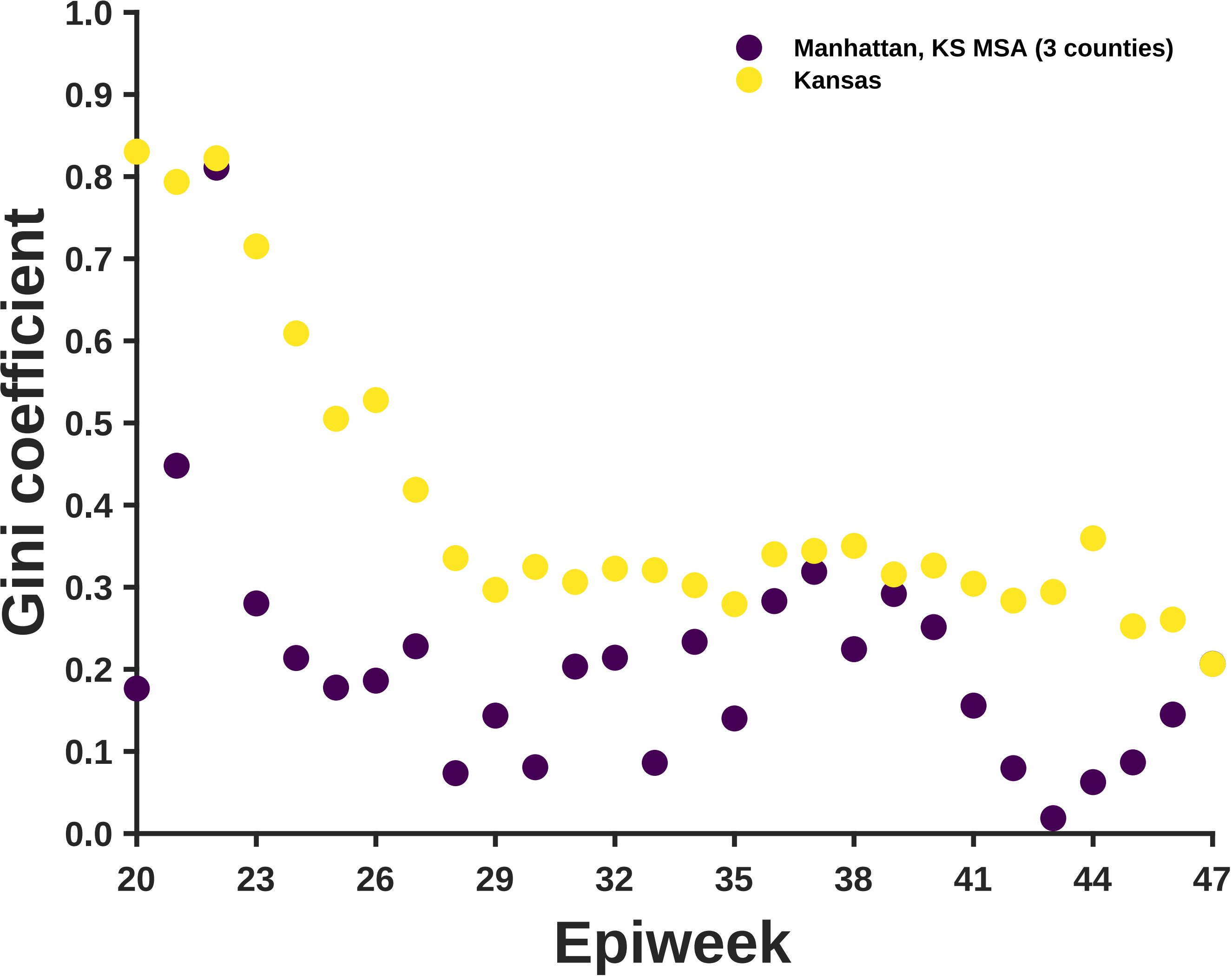
Manhattan, KS MSA (3 counties)
Kansas
Gini coefficient
1.0
0.9
0.8
0.7
0.6
0.5
0.4
0.3
0.2
0.1
0.0
20
23
26
29
32
35
38
41
44
47
Epiweek

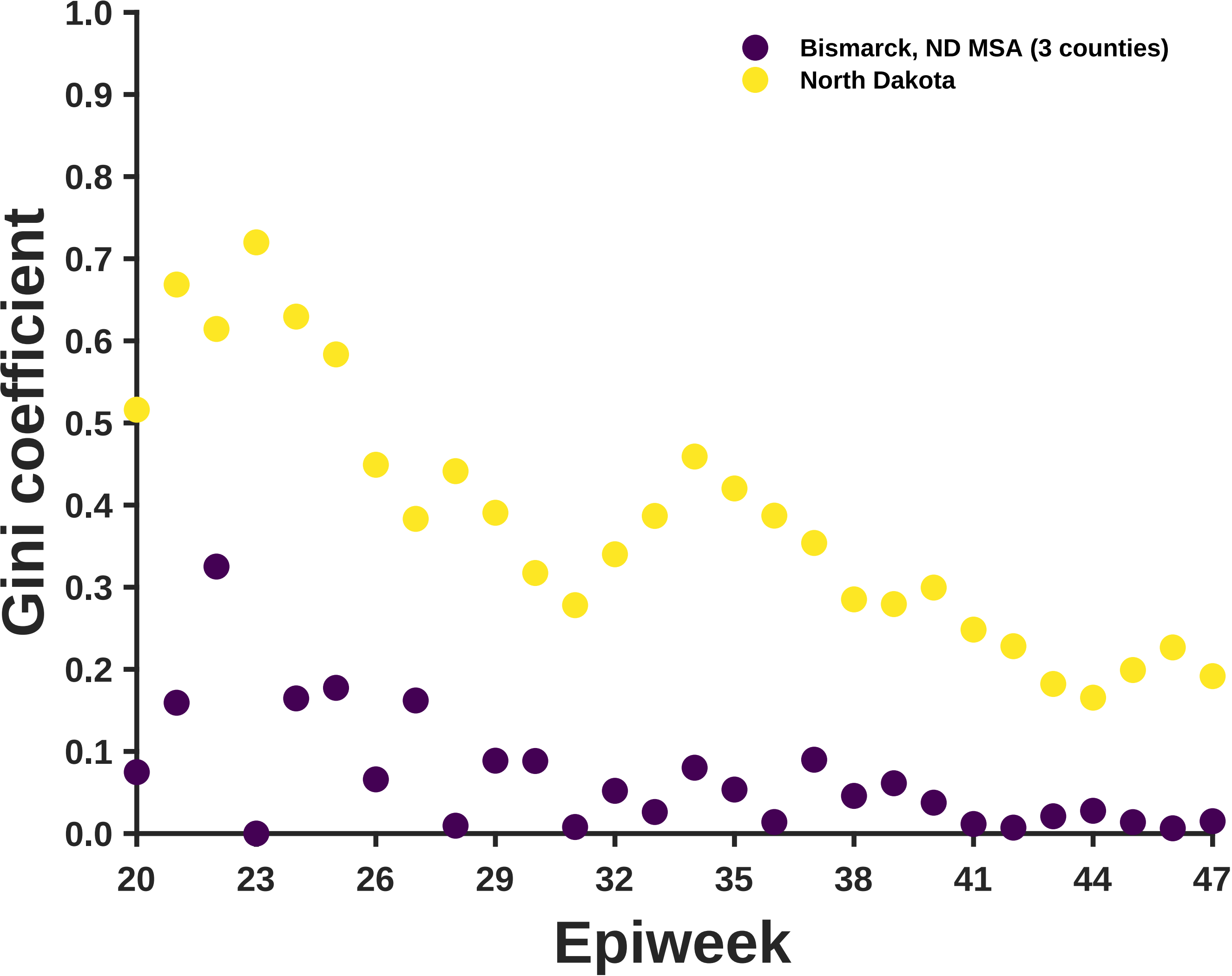

Gini coefficient
1.0
0.9
0.8
0.7
0.6
0.5
0.4
0.3
0.2
0.1
0.0
Bismarck, ND MSA (3 counties)
North Dakota
20
23
26
29
32
35
38
41
44
47
Epiweek

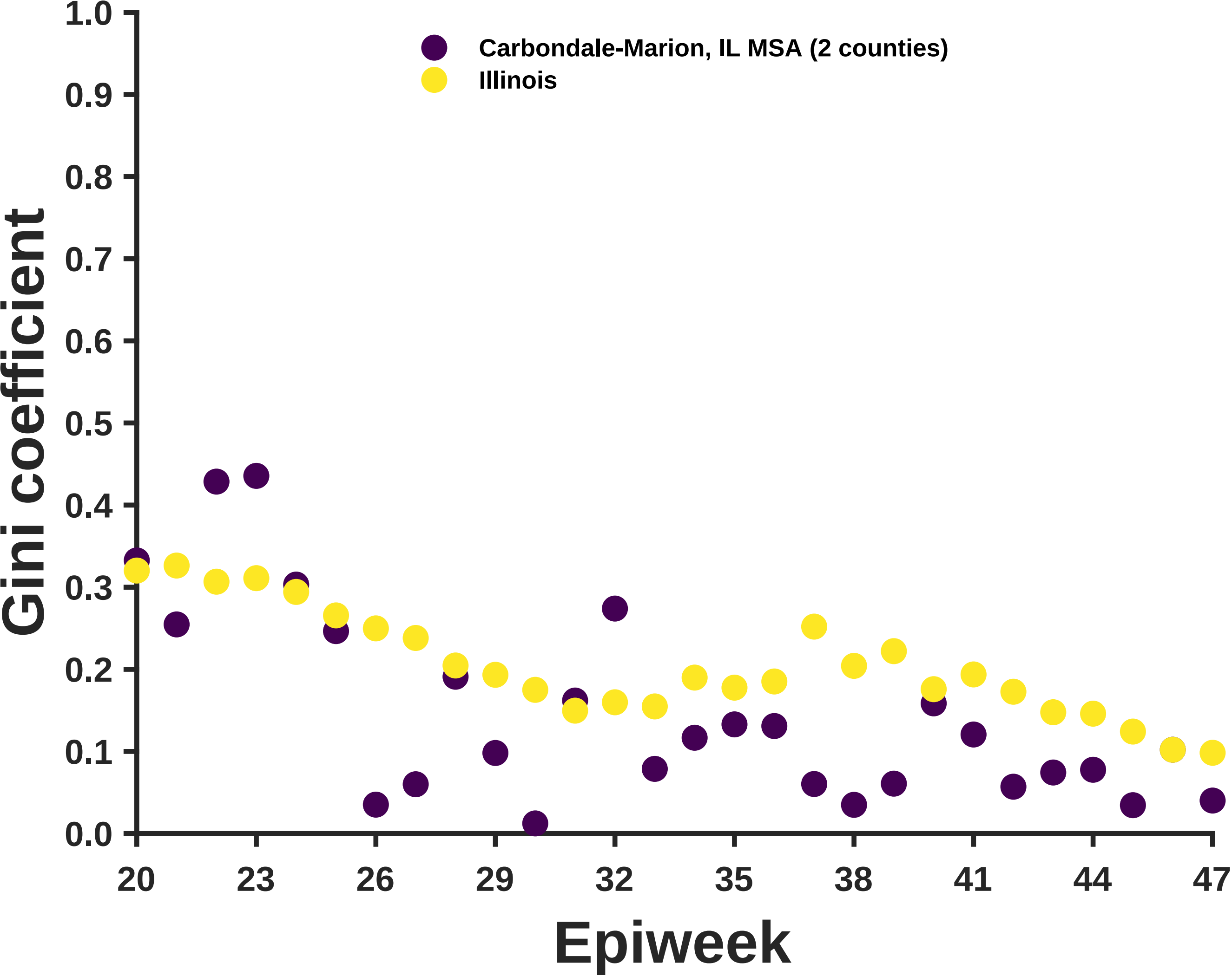

Carbondale-Marion, IL MSA (2 counties)
Illinois
Gini coefficient
Epiweek
1.0
0.9
0.8
0.7
0.6
0.5
0.4
0.3
0.2
0.1
0.0
20
23
26
29
32
35
38
41
44
47

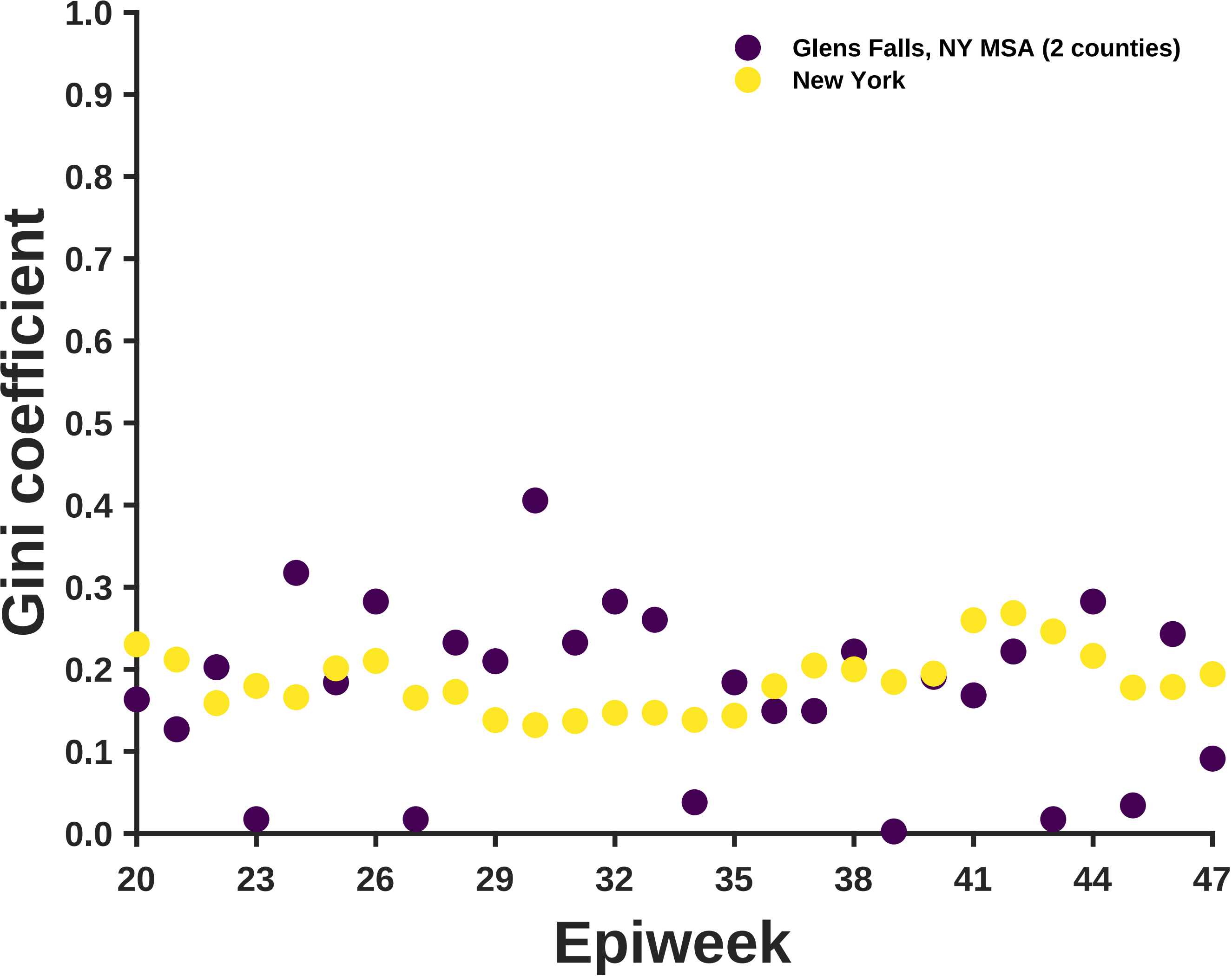

Glens Falls, NY MSA (2 counties)
New York
Gini coefficient
1.0
0.9
0.8
0.7
0.6
0.5
0.4
0.3
0.2
0.1
0.0
20
23
26
29
32
35
38
41
44
47
Epiweek

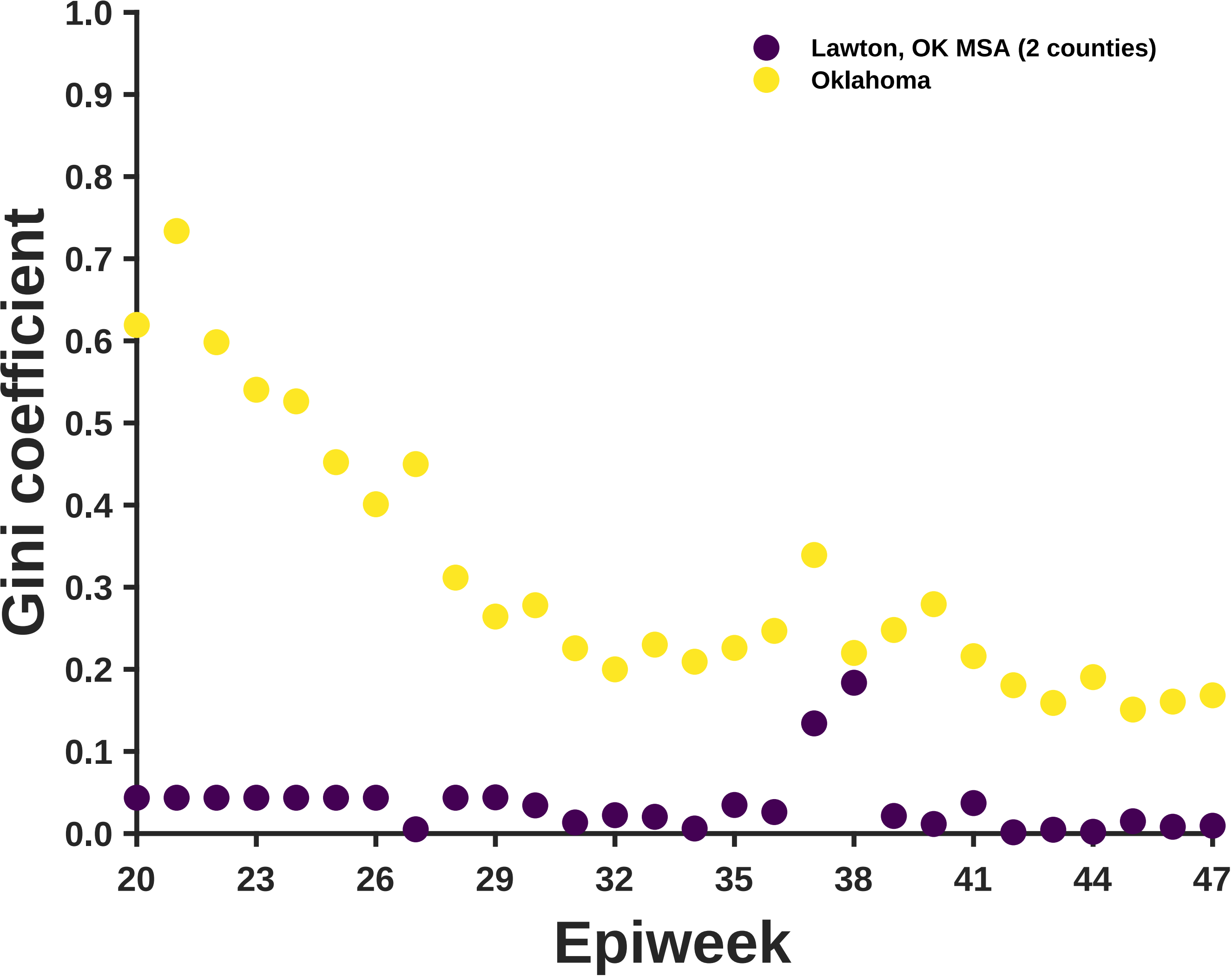

Lawton, OK MSA (2 counties)
Oklahoma
Gini coefficient
1.0
0.9
0.8
0.7
0.6
0.5
0.4
0.3
0.2
0.1
0.0
20
23
26
29
32
35
38
41
44
47
Epiweek

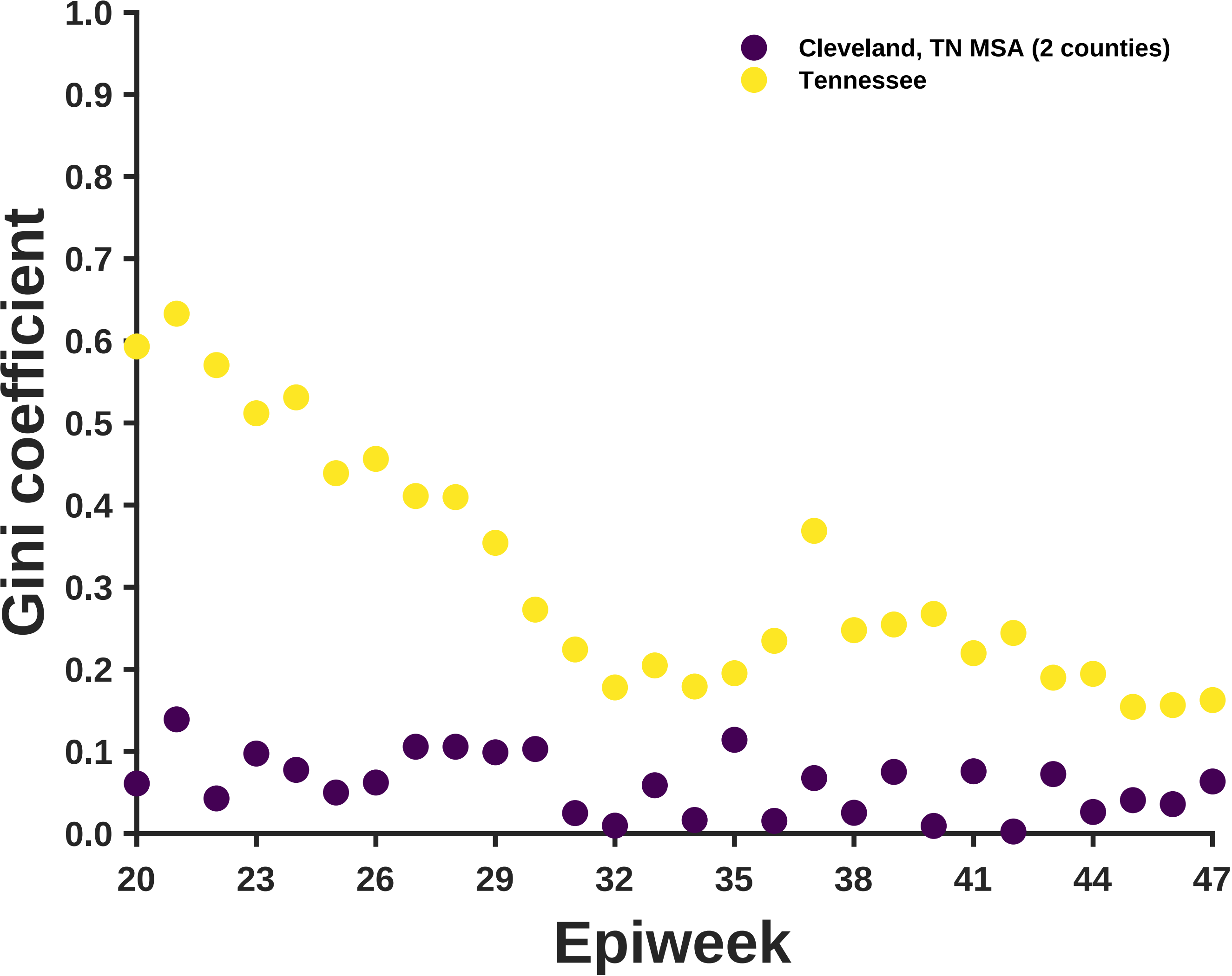
Cleveland, TN MSA (2 counties)
Tennessee
Gini coefficient
1.0
0.9
0.8
0.7
0.6
0.5
0.4
0.3
0.2
0.1
0.0
20
23
26
29
32
35
38
41
44
47
Epiweek

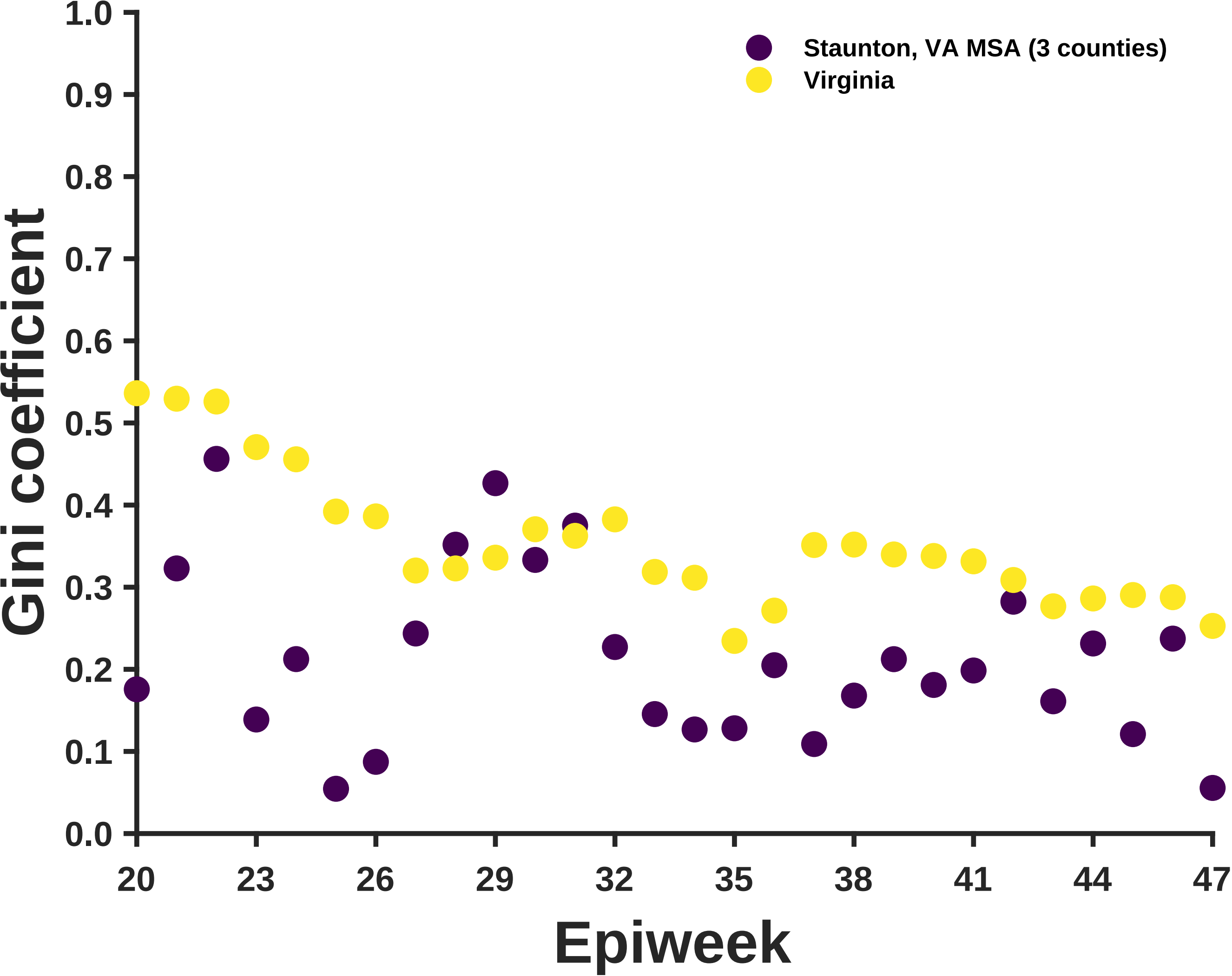

Staunton, VA MSA (3 counties)
Virginia
Gini coefficient
1.0
0.9
0.8
0.7
0.6
0.5
0.4
0.3
0.2
0.1
0.0
20
23
26
29
32
35
38
41
44
47
Epiweek

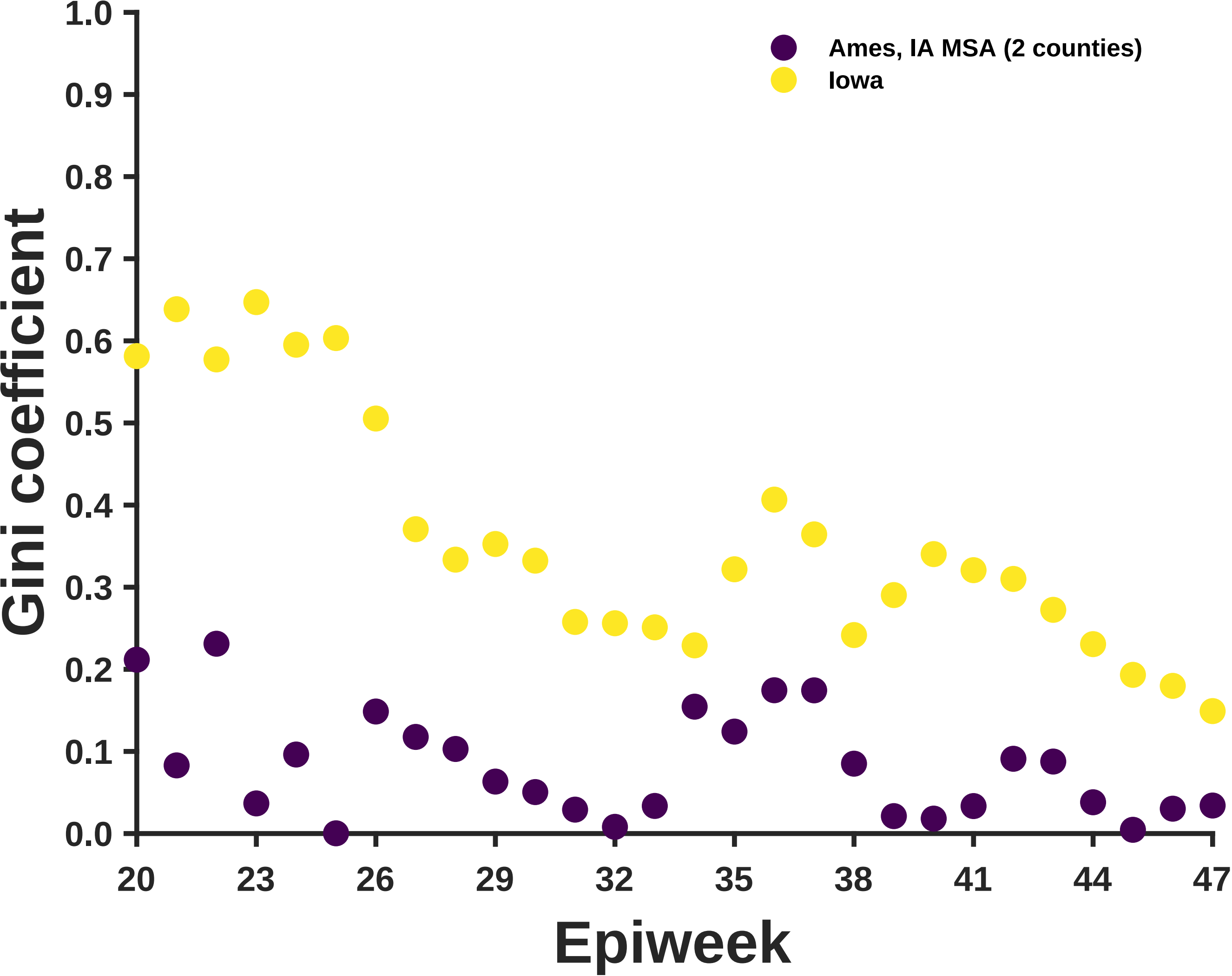
Ames, IA MSA (2 counties)
Iowa
Gini coefficient
Epiweek
0.0
0.1
0.2
0.3
0.4
0.5
0.6
0.7
0.8
0.9
1.0
20
23
26
29
32
35
38
41
44
47

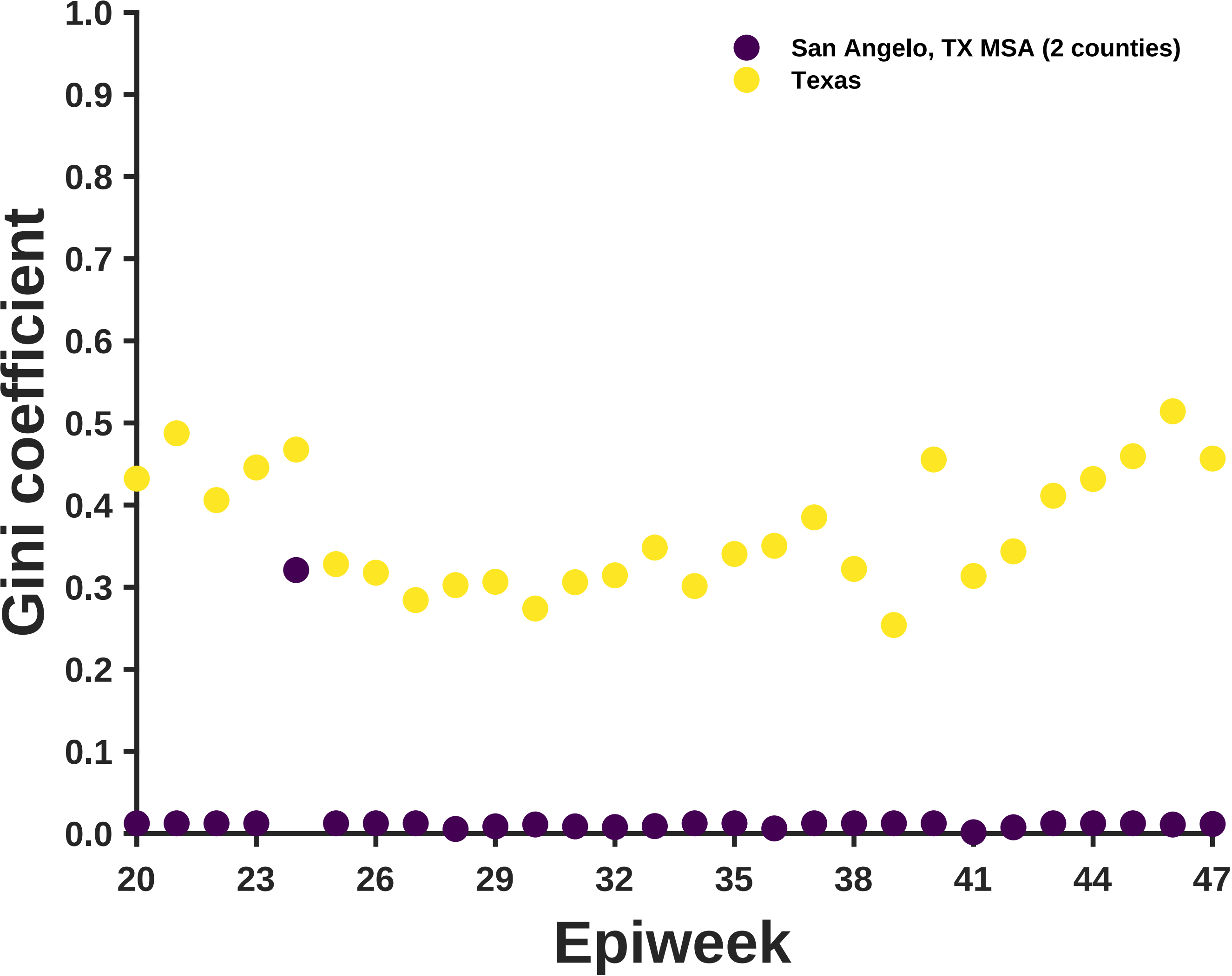

San Angelo, TX MSA (2 counties)
Texas
Gini coefficient
1.0
0.9
0.8
0.7
0.6
0.5
0.4
0.3
0.2
0.1
0.0
20
23
26
29
32
35
38
41
44
47
Epiweek

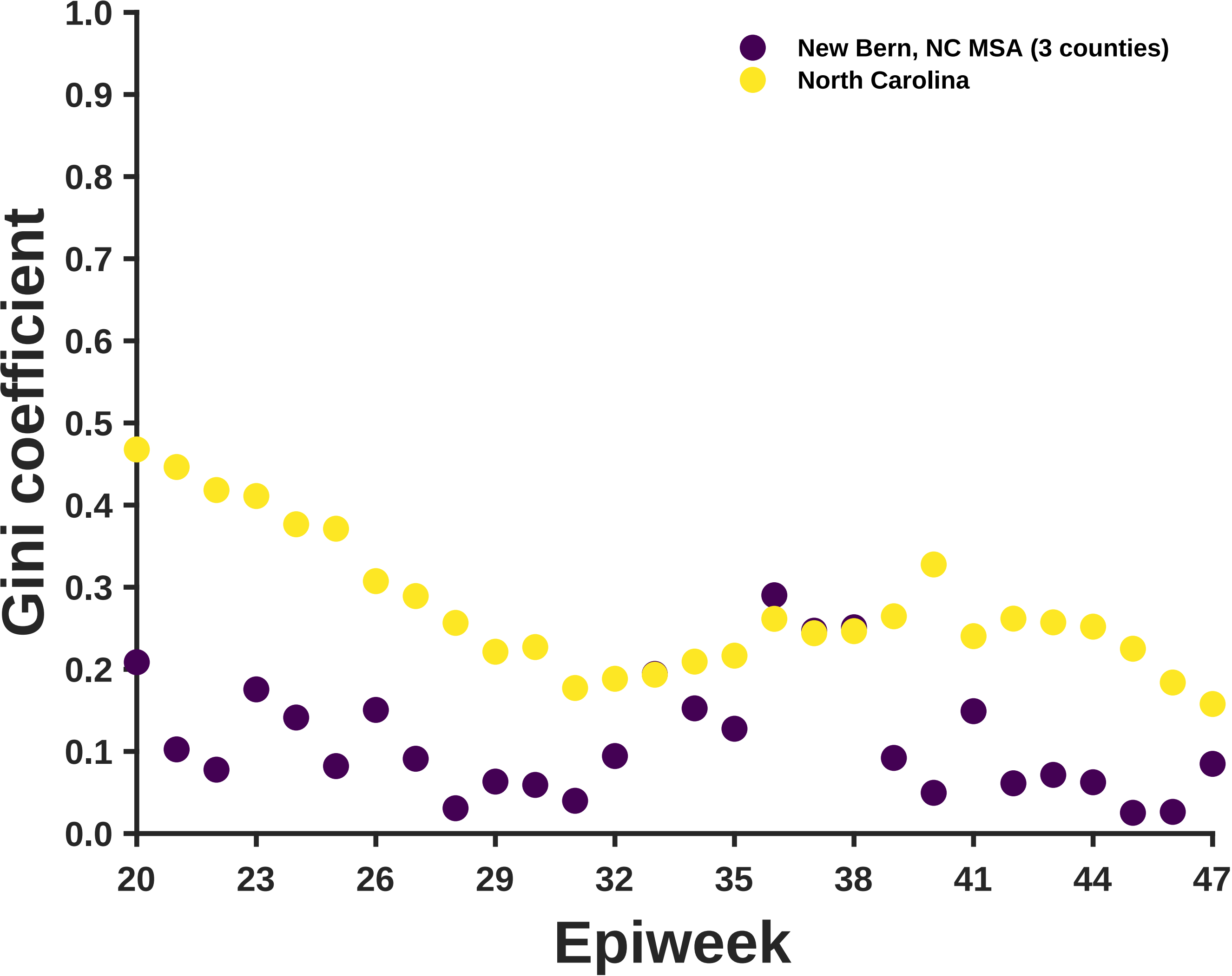

New Bern, NC MSA (3 counties)
North Carolina
Gini coefficient
Epiweek
1.0
0.9
0.8
0.7
0.6
0.5
0.4
0.3
0.2
0.1
0.0
20
23
26
29
32
35
38
41
44
47

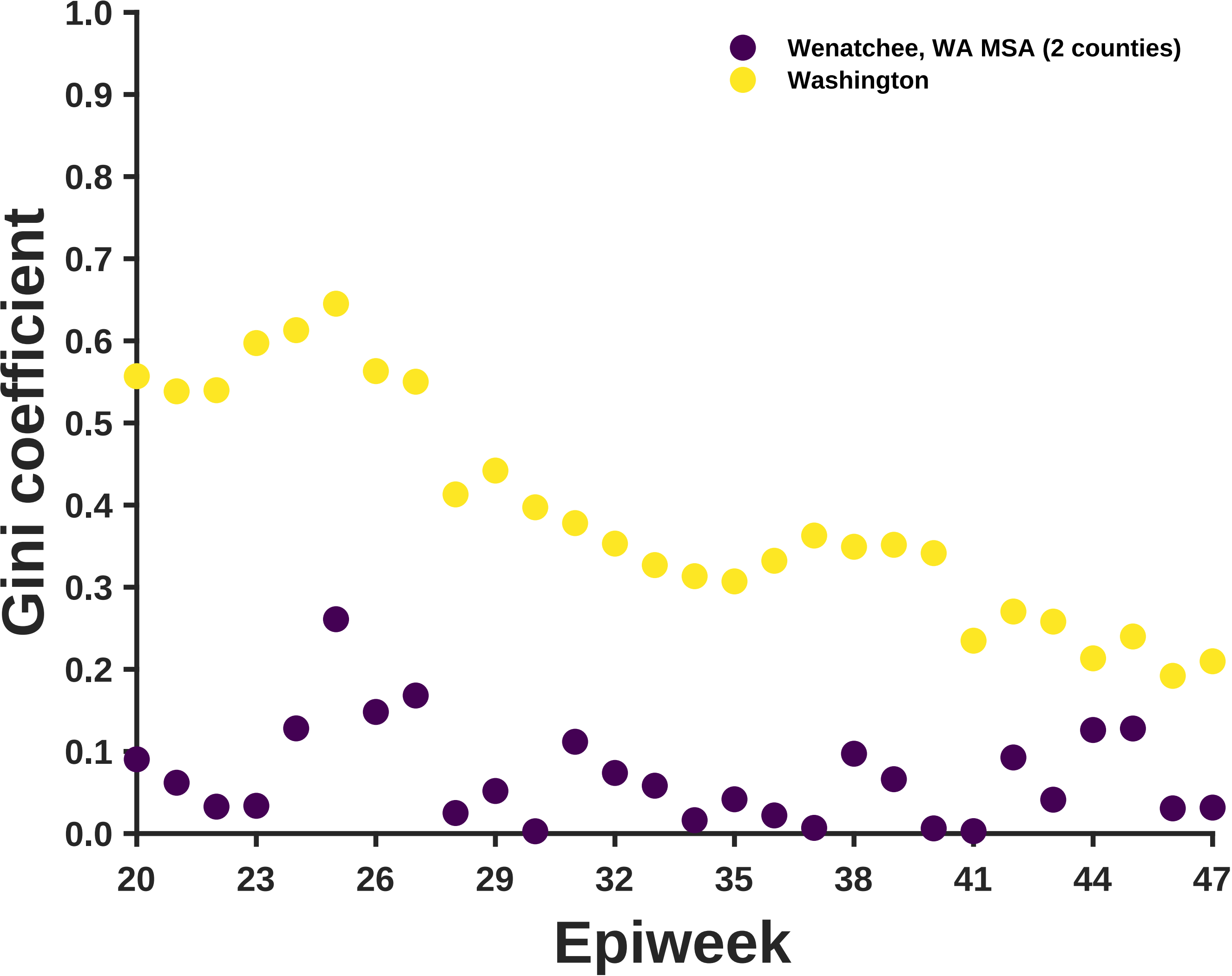
Wenatchee, WA MSA (2 counties)
Washington
Gini coefficient
1.0
0.9
0.8
0.7
0.6
0.5
0.4
0.3
0.2
0.1
0.0
20
23
26
29
32
35
38
41
44
47
Epiweek

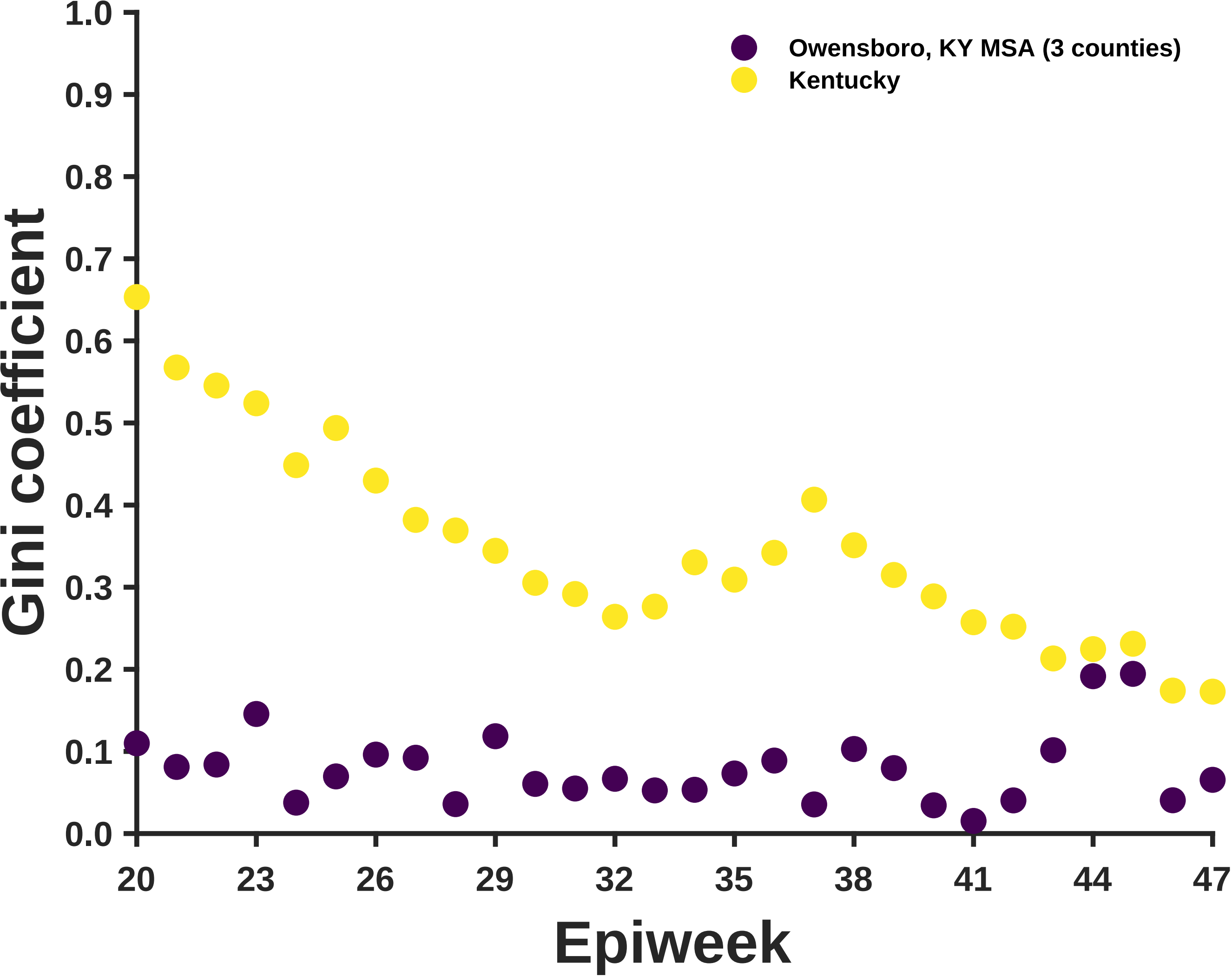

Owensboro, KY MSA (3 counties)
Kentucky
Gini coefficient
0.0
0.1
0.2
0.3
0.4
0.5
0.6
0.7
0.8
0.9
1.0
20
23
26
29
32
35
38
41
44
47
Epiweek

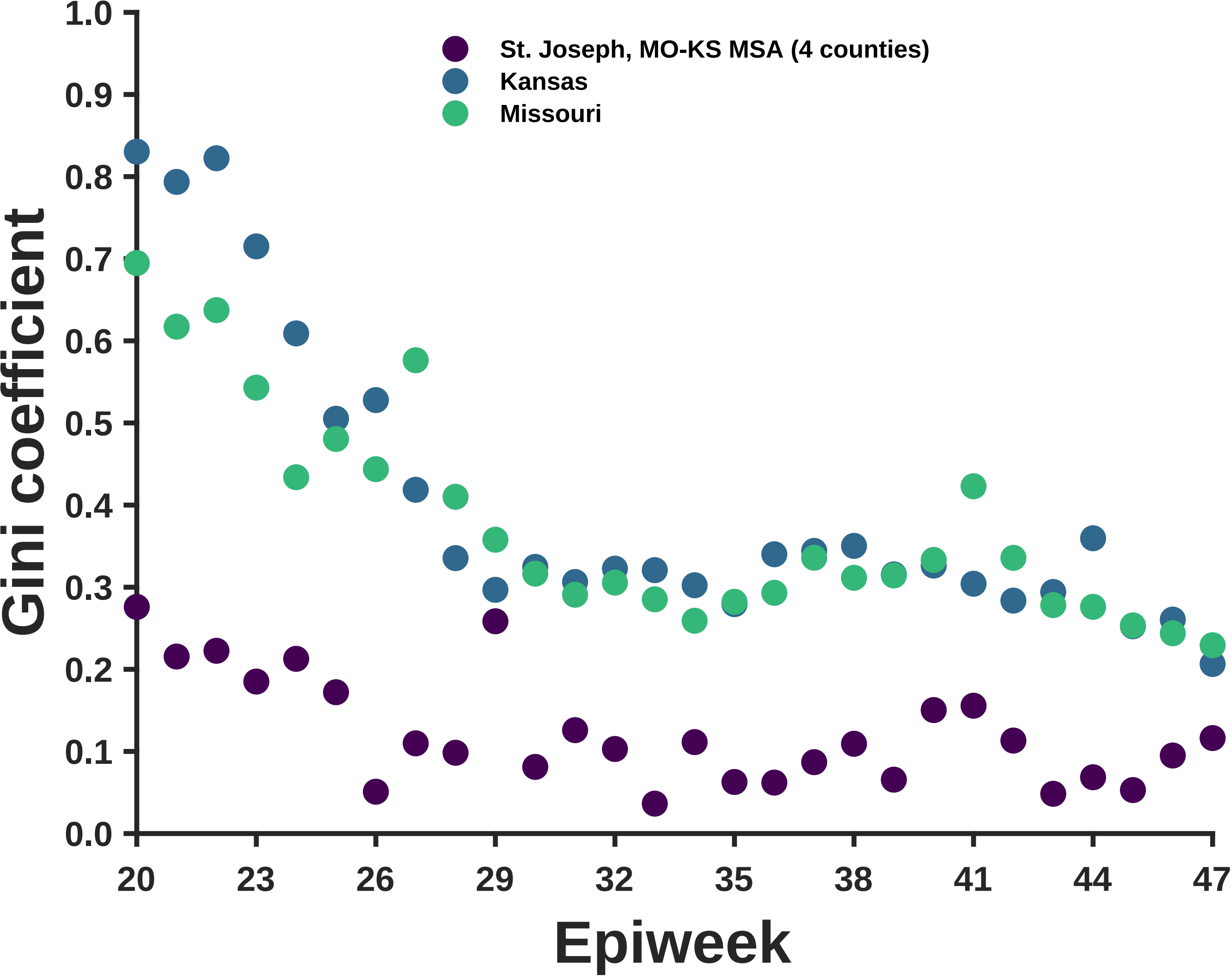
St. Joseph, MO-KS MSA (4 counties)
Kansas
Missouri
Gini coefficient
Epiweek
1.0
0.9
0.8
0.7
0.6
0.5
0.4
0.3
0.2
0.1
0.0
20
23
26
29
32
35
38
41
44
47

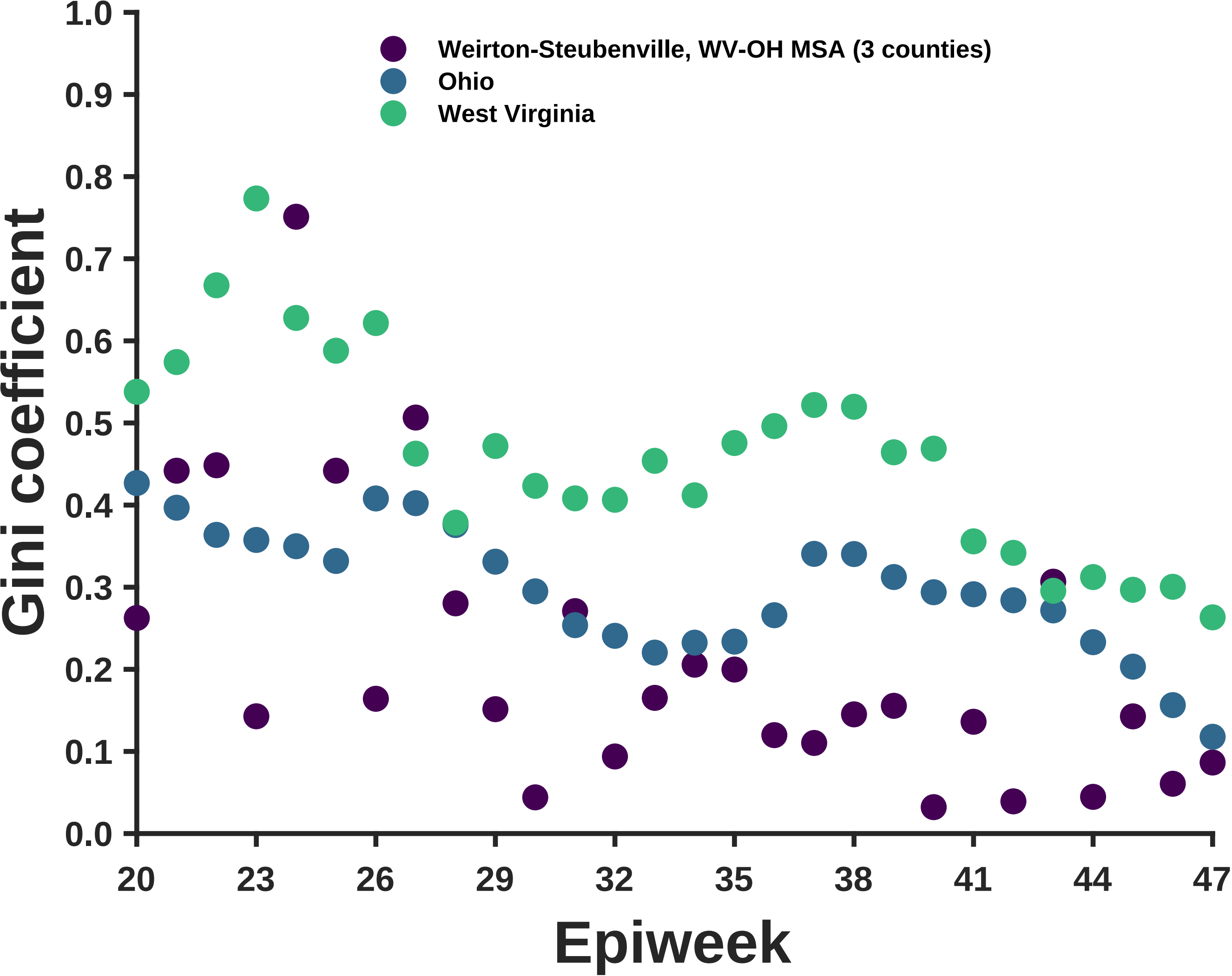

Weirton-Steubenville, WV-OH MSA (3 counties)
Ohio
West Virginia
Gini coefficient
Epiweek
1.0
0.9
0.8
0.7
0.6
0.5
0.4
0.3
0.2
0.1
0.0
20
23
26
29
32
35
38
41
44
47

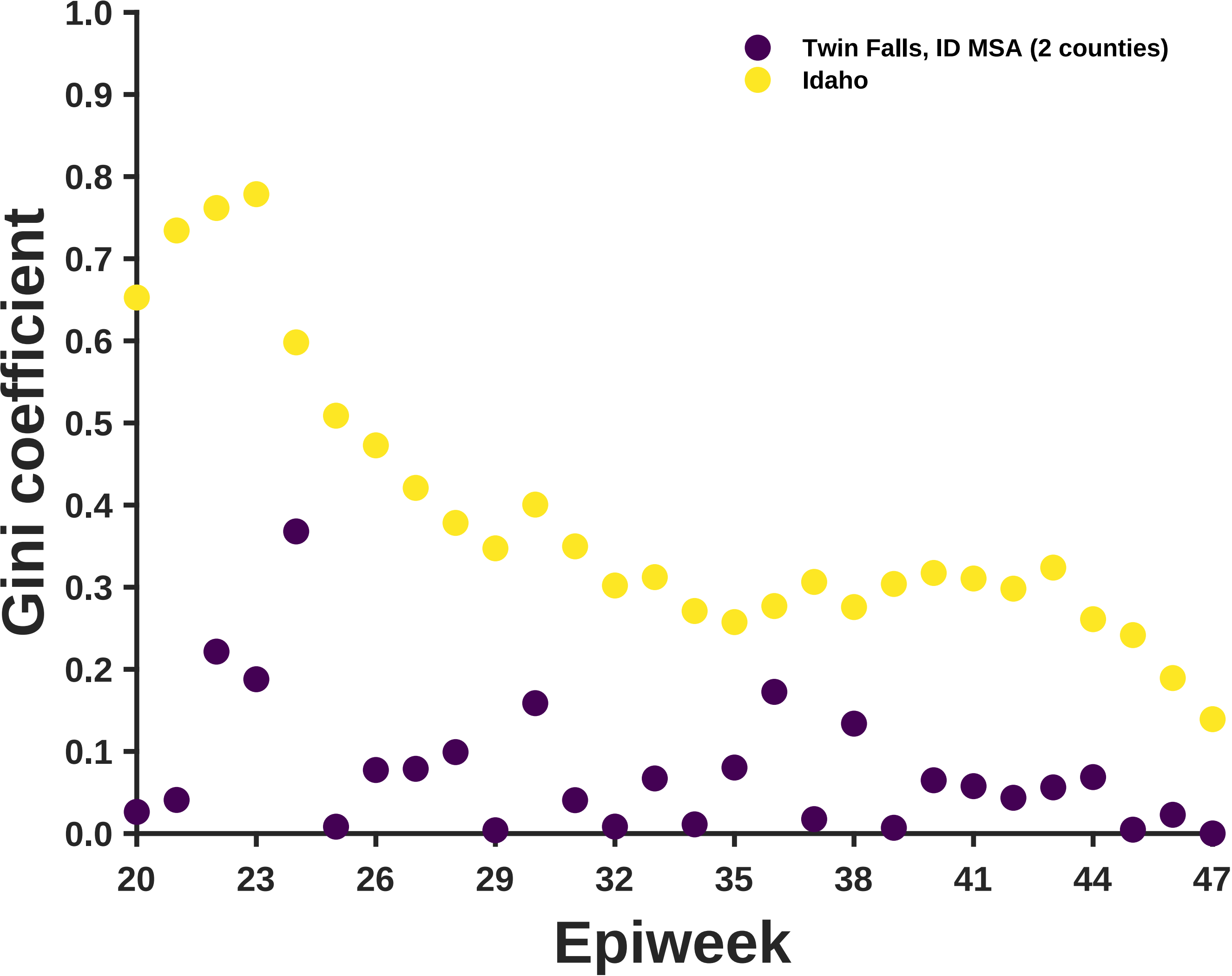

Gini coefficient
1.0
0.9
0.8
0.7
0.6
0.5
0.4
0.3
0.2
0.1
0.0
20
23
26
29
32
35
38
41
44
47
Epiweek
Twin Falls, ID MSA (2 counties)
Idaho

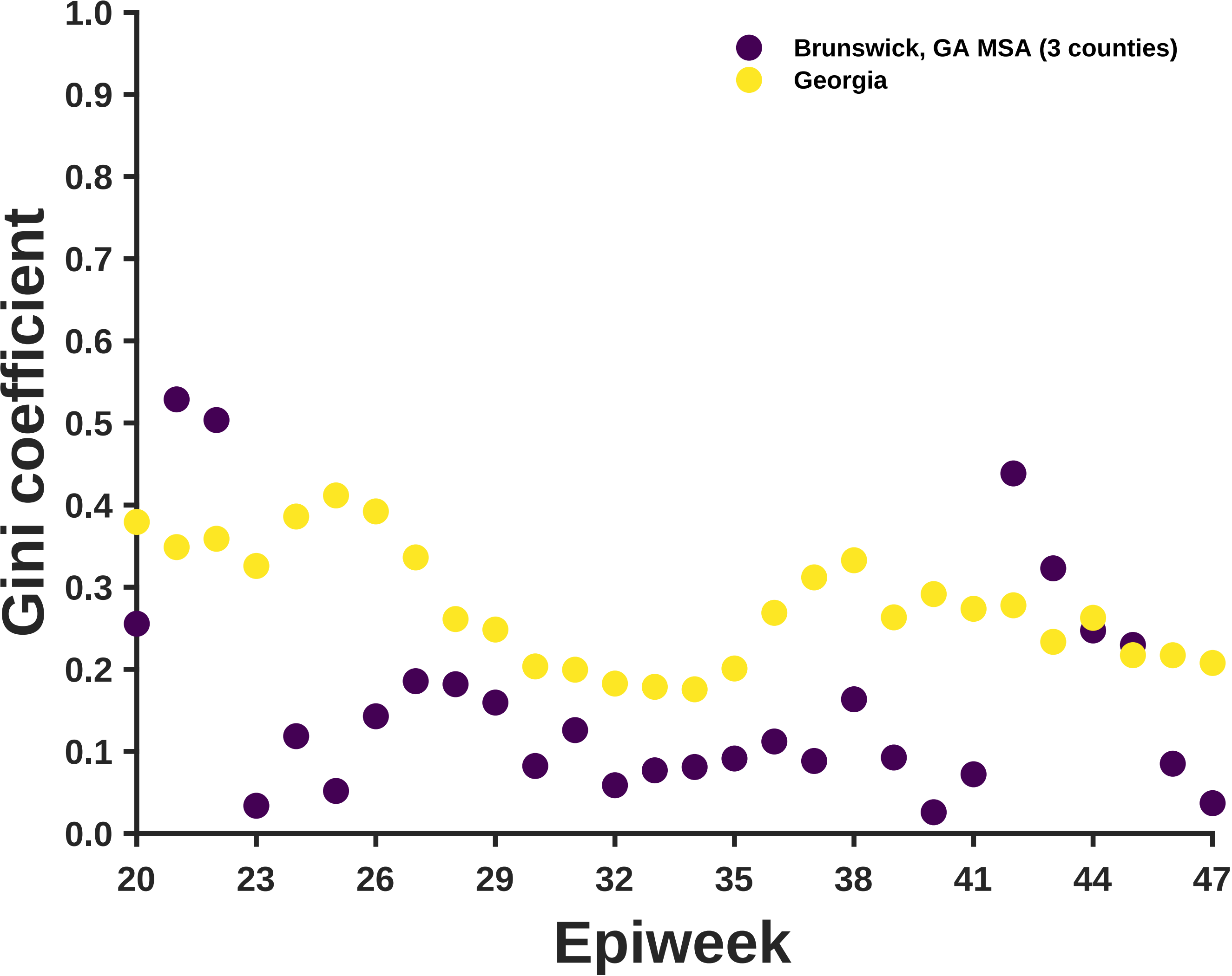
Brunswick, GA MSA (3 counties)
Georgia
Gini coefficient
Epiweek
1.0
0.9
0.8
0.7
0.6
0.5
0.4
0.3
0.2
0.1
0.0
20
23
26
29
32
35
38
41
44
47

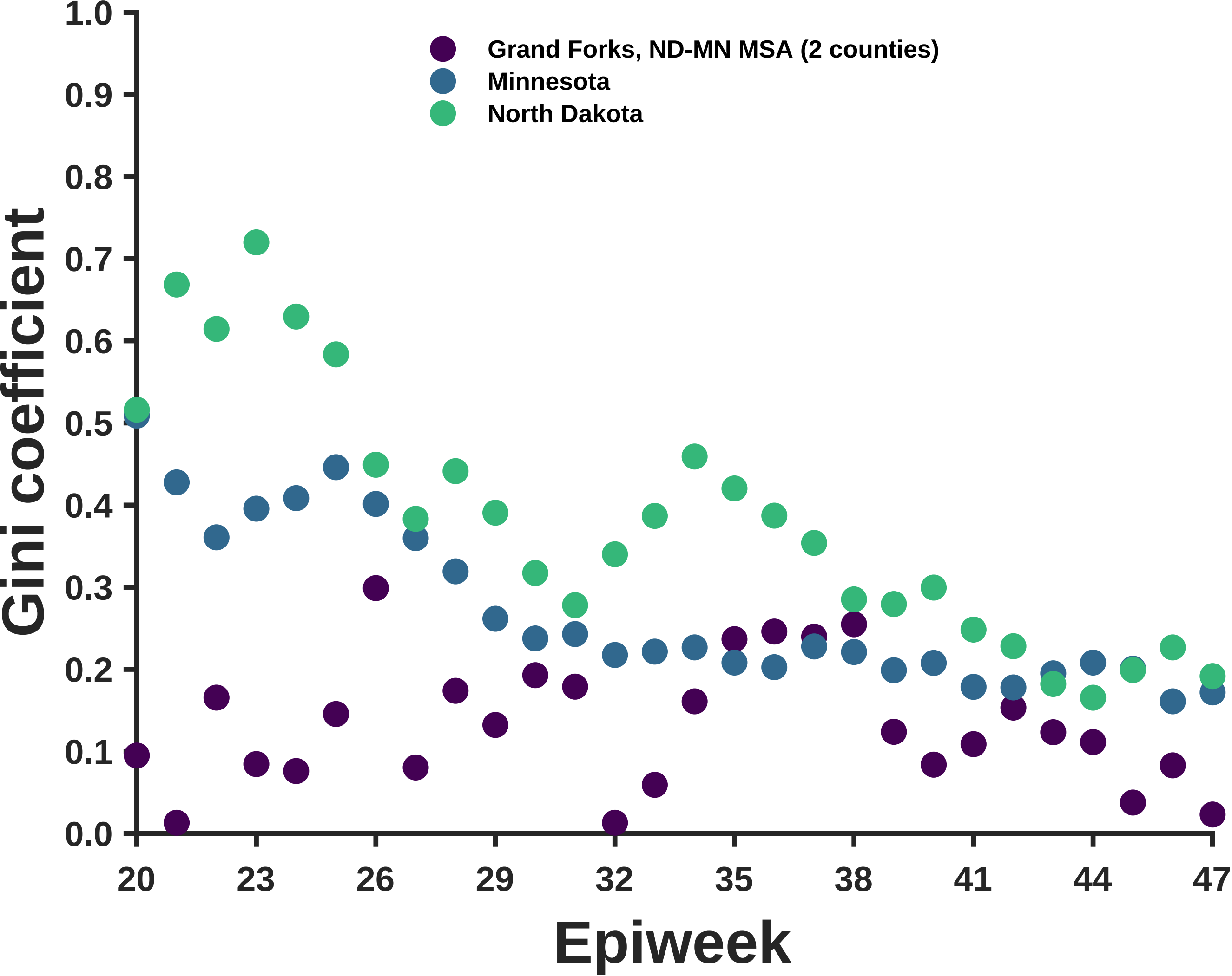

Grand Forks, ND-MN MSA (2 counties)
Minnesota
North Dakota
Gini coefficient
0.0
0.1
0.2
0.3
0.4
0.5
0.6
0.7
0.8
0.9
1.0
20
23
26
29
32
35
38
41
44
47
Epiweek

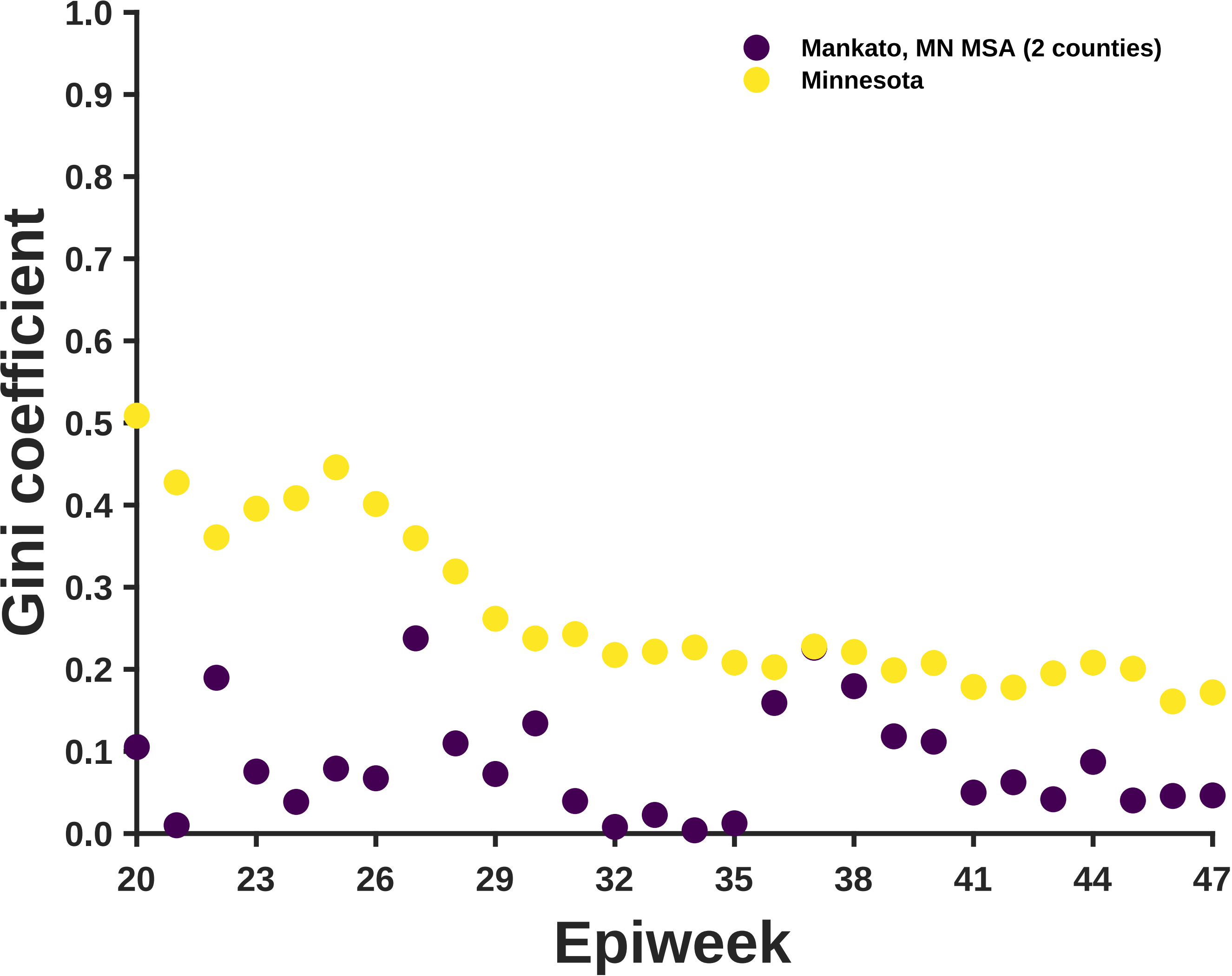

Mankato, MN MSA (2 counties)
Minnesota
Gini coefficient
Epiweek
1.0
0.9
0.8
0.7
0.6
0.5
0.4
0.3
0.2
0.1
0.0
20
23
26
29
32
35
38
41
44
47

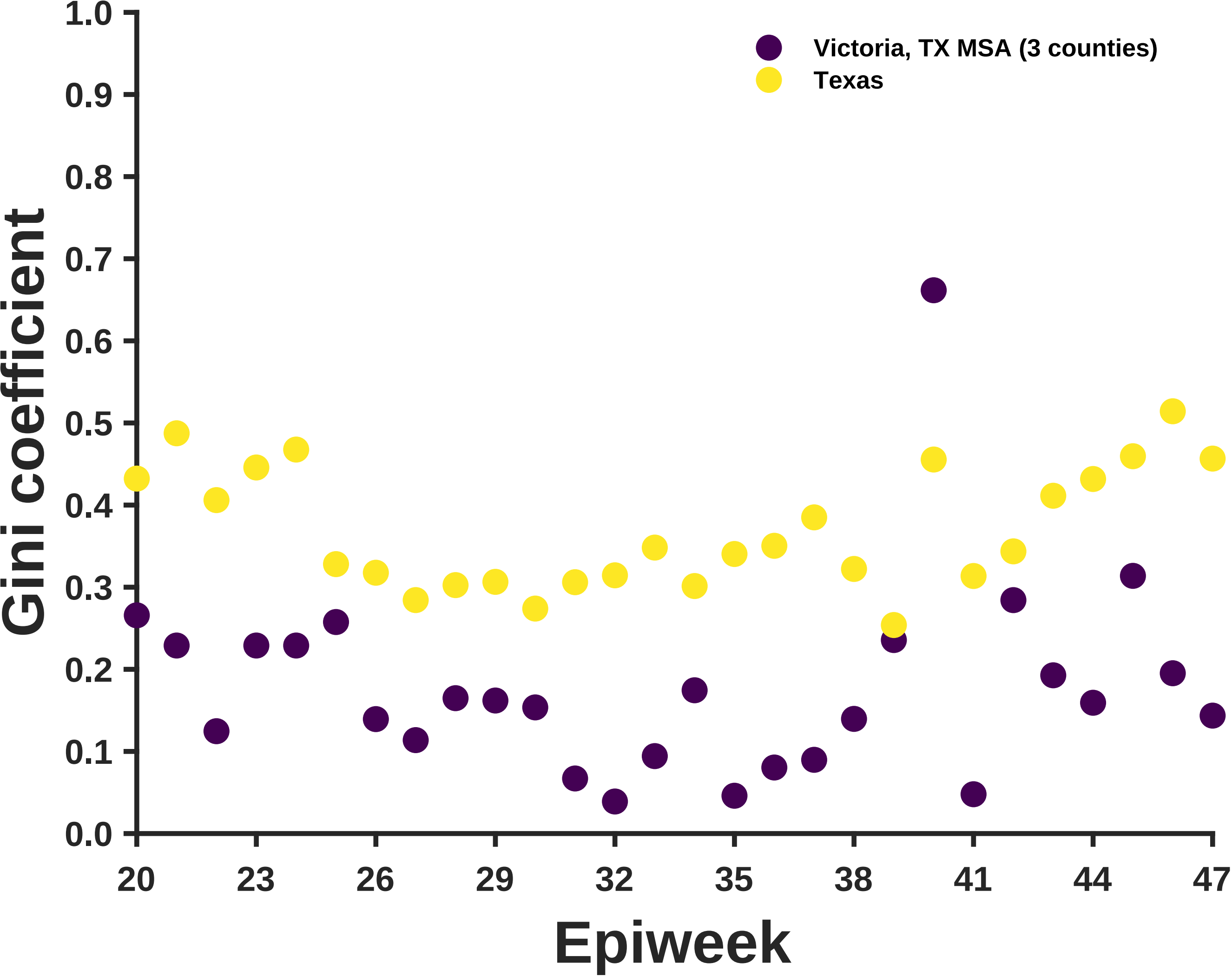
Victoria, TX MSA (3 counties)
Texas
Gini coefficient
1.0
0.9
0.8
0.7
0.6
0.5
0.4
0.3
0.2
0.1
0.0
20
23
26
29
32
35
38
41
44
47
Epiweek

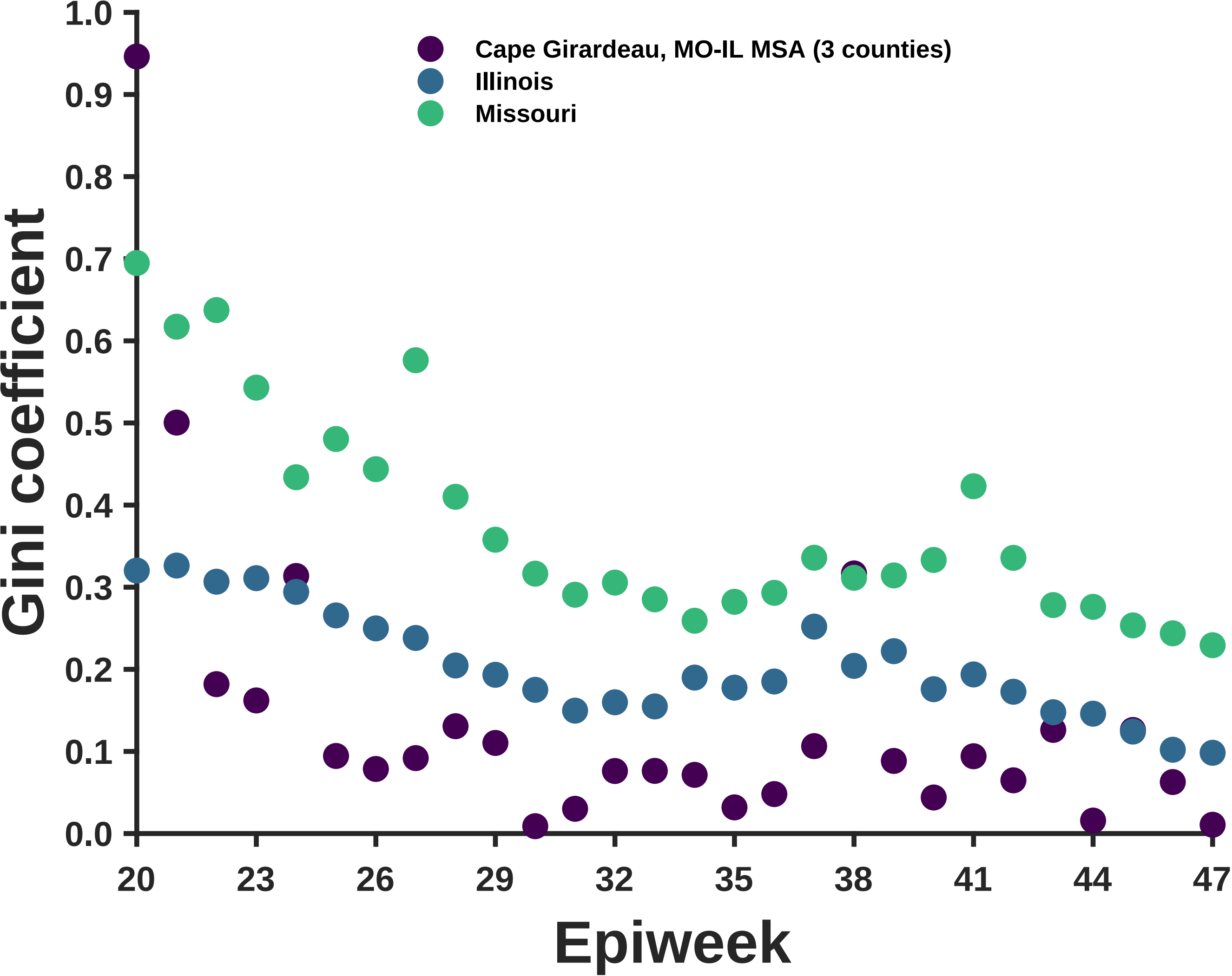

Cape Girardeau, MO-IL MSA (3 counties)
Illinois
Missouri
Gini coefficient
Epiweek
0.0
0.1
0.2
0.3
0.4
0.5
0.6
0.7
0.8
0.9
1.0
20
23
26
29
32
35
38
41
44
47

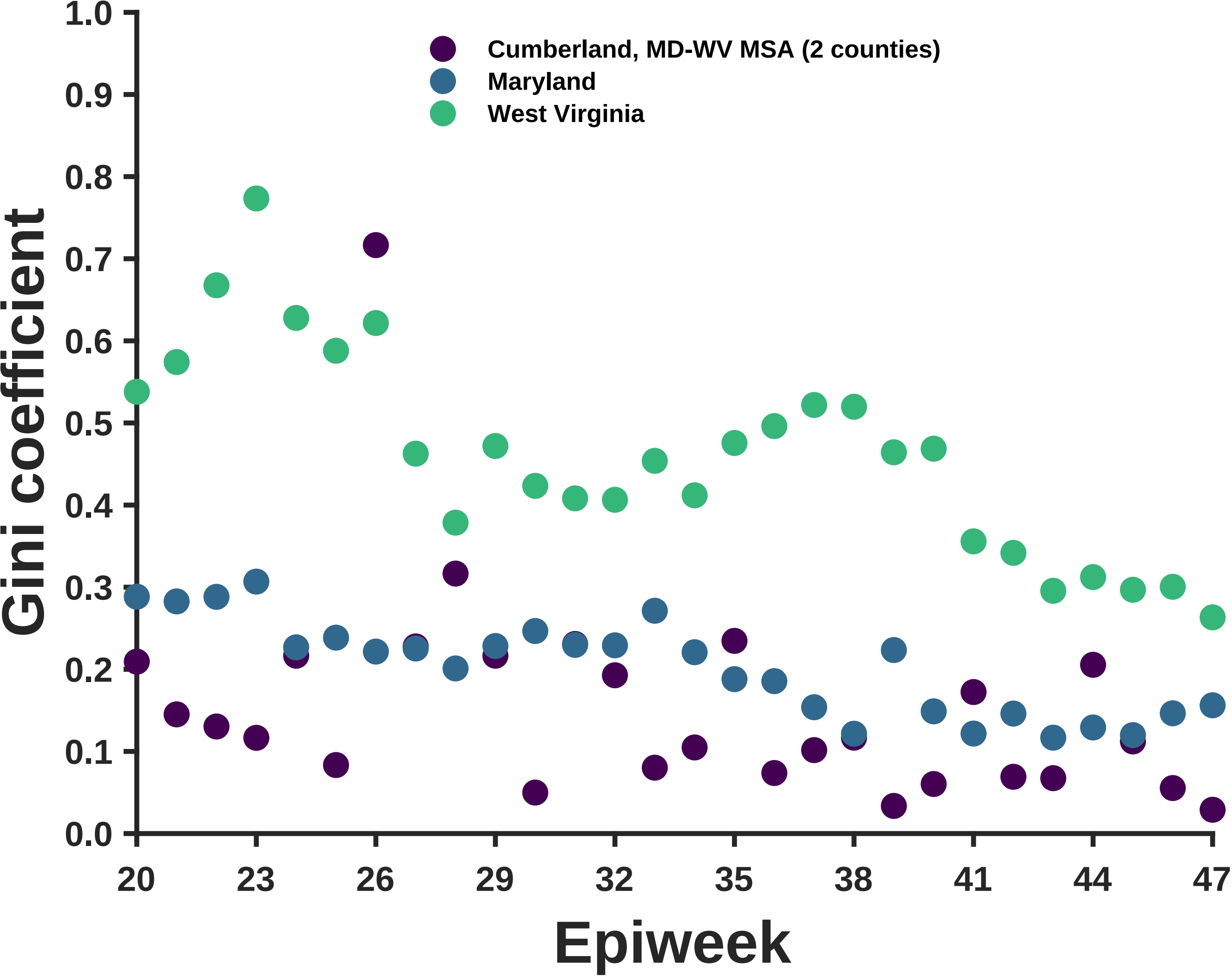

Cumberland, MD-WV MSA (2 counties)
Maryland
West Virginia
Gini coefficient
Epiweek
1.0
0.9
0.8
0.7
0.6
0.5
0.4
0.3
0.2
0.1
0.0
20
23
26
29
32
35
38
41
44
47

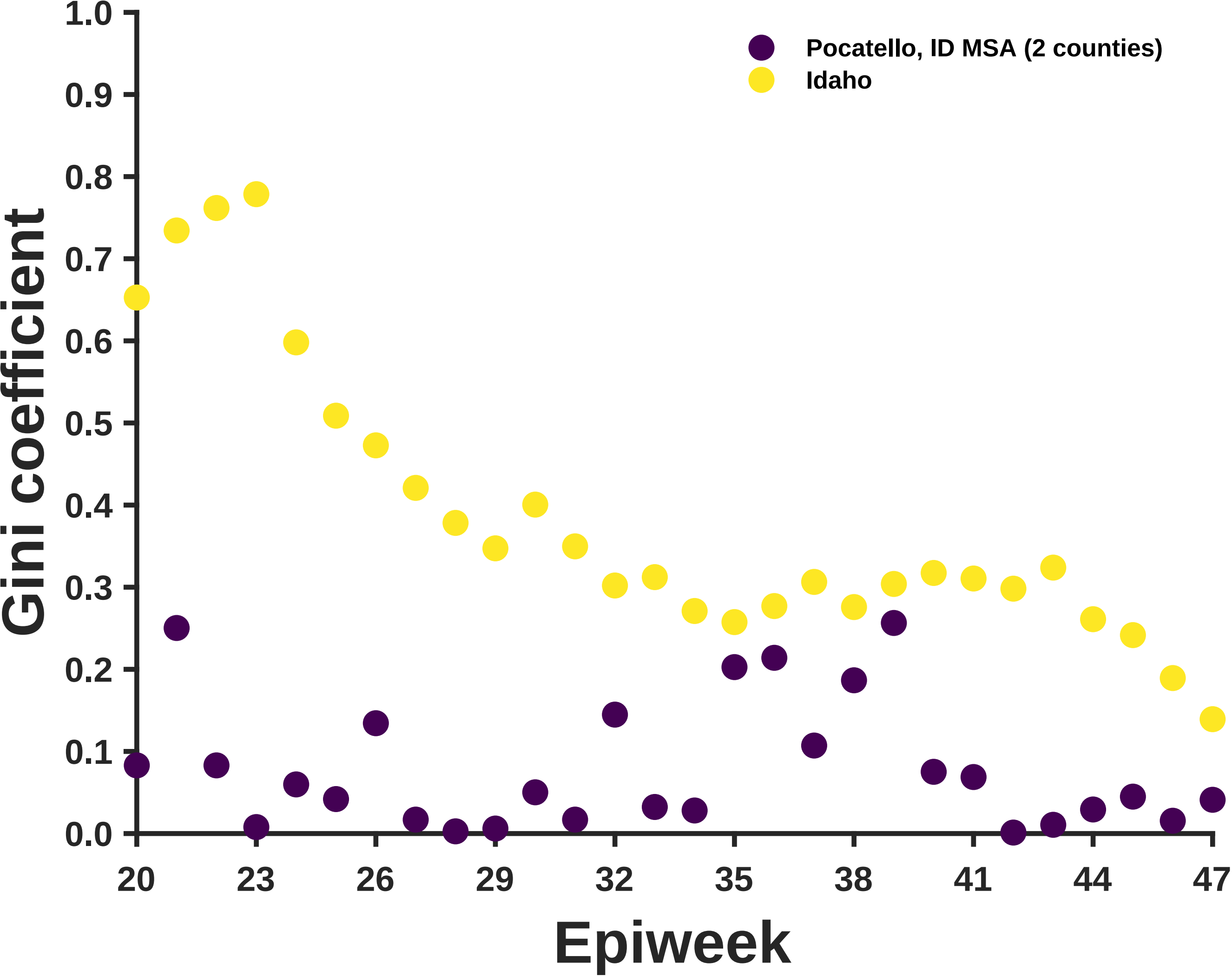
Pocatello, ID MSA (2 counties)
Idaho
Gini coefficient
Epiweek
1.0
0.9
0.8
0.7
0.6
0.5
0.4
0.3
0.2
0.1
0.0
20
23
26
29
32
35
38
41
44
47

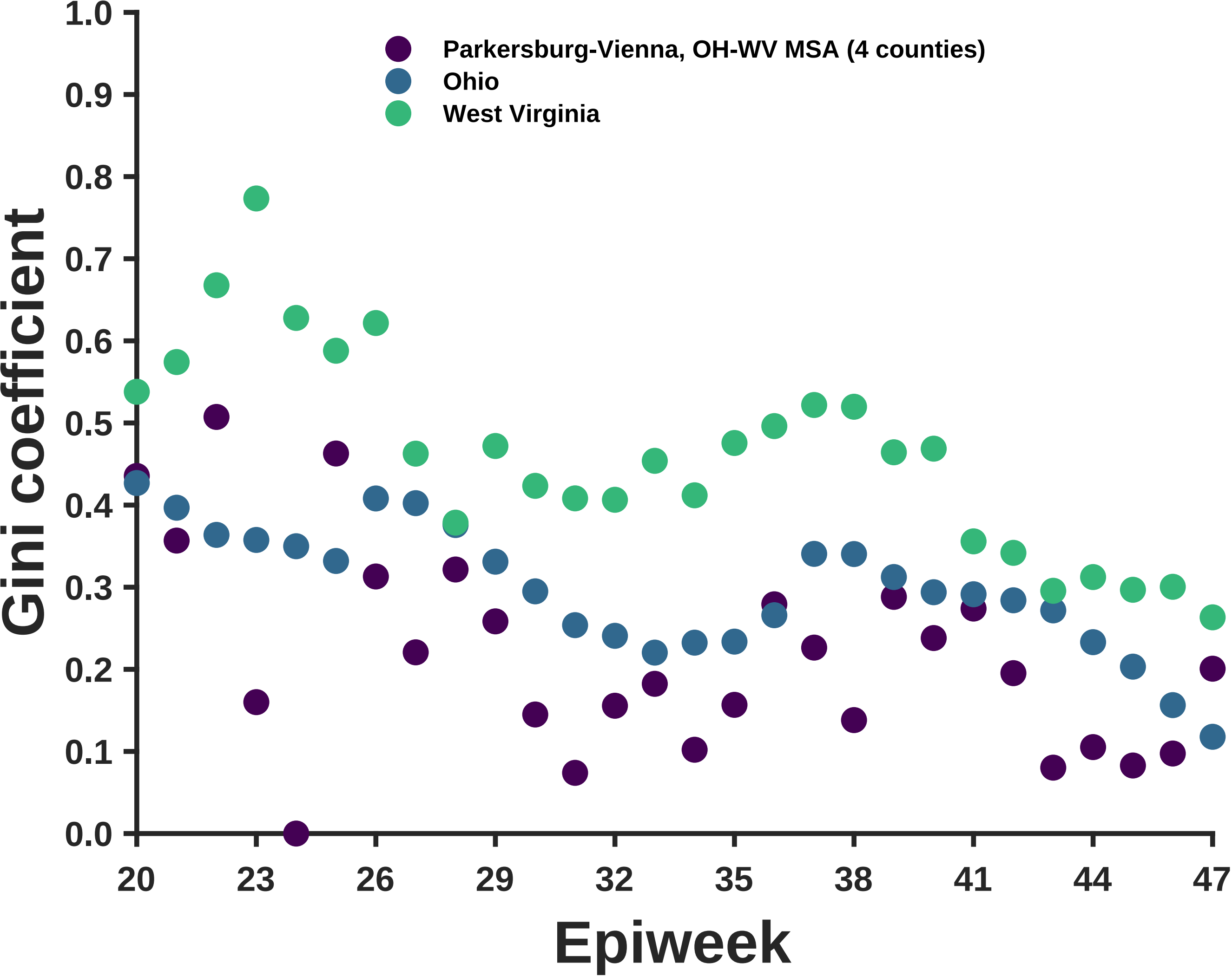
Parkersburg-Vienna, OH-WV MSA (4 counties)
Ohio
West Virginia
Gini coefficient
0.0
0.1
0.2
0.3
0.4
0.5
0.6
0.7
0.8
0.9
1.0
20
23
26
29
32
35
38
41
44
47
Epiweek

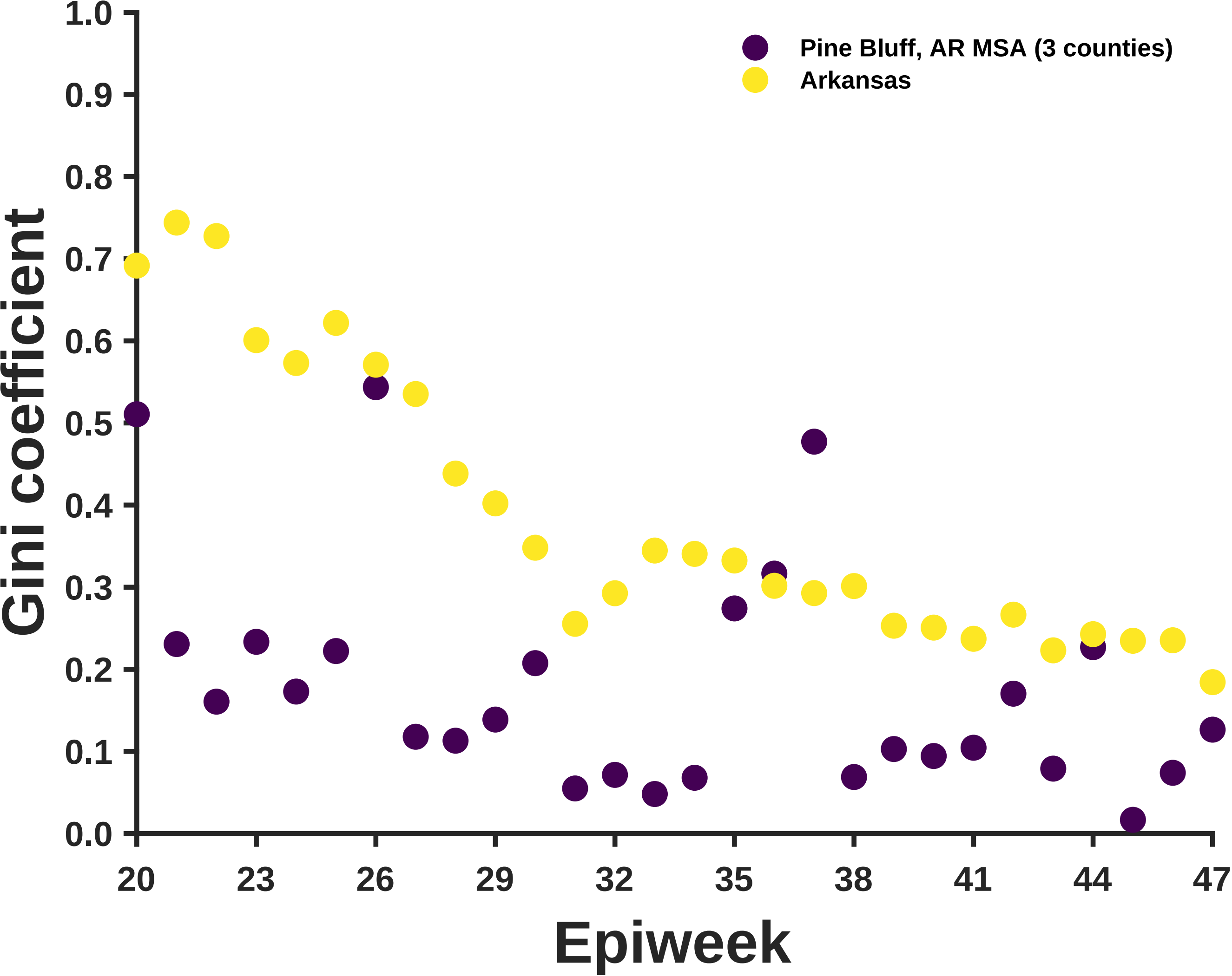
Pine Bluff, AR MSA (3 counties)
Arkansas
Gini coefficient
Epiweek
0.0
0.1
0.2
0.3
0.4
0.5
0.6
0.7
0.8
0.9
1.0
20
23
26
29
32
35
38
41
44
47

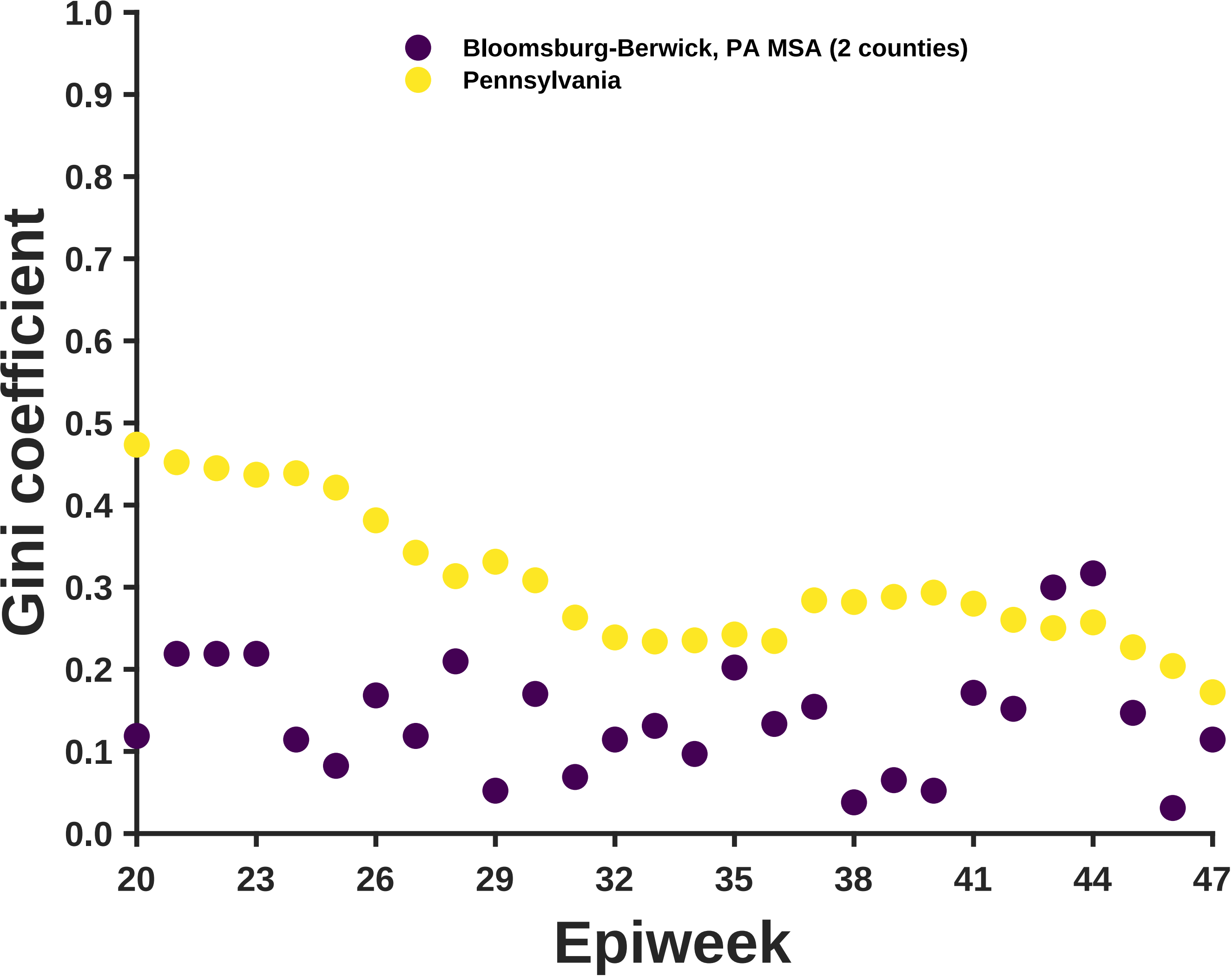
Bloomsburg-Berwick, PA MSA (2 counties)
Pennsylvania
Gini coefficient
1.0
0.9
0.8
0.7
0.6
0.5
0.4
0.3
0.2
0.1
0.0
20
23
26
29
32
35
38
41
44
47
Epiweek

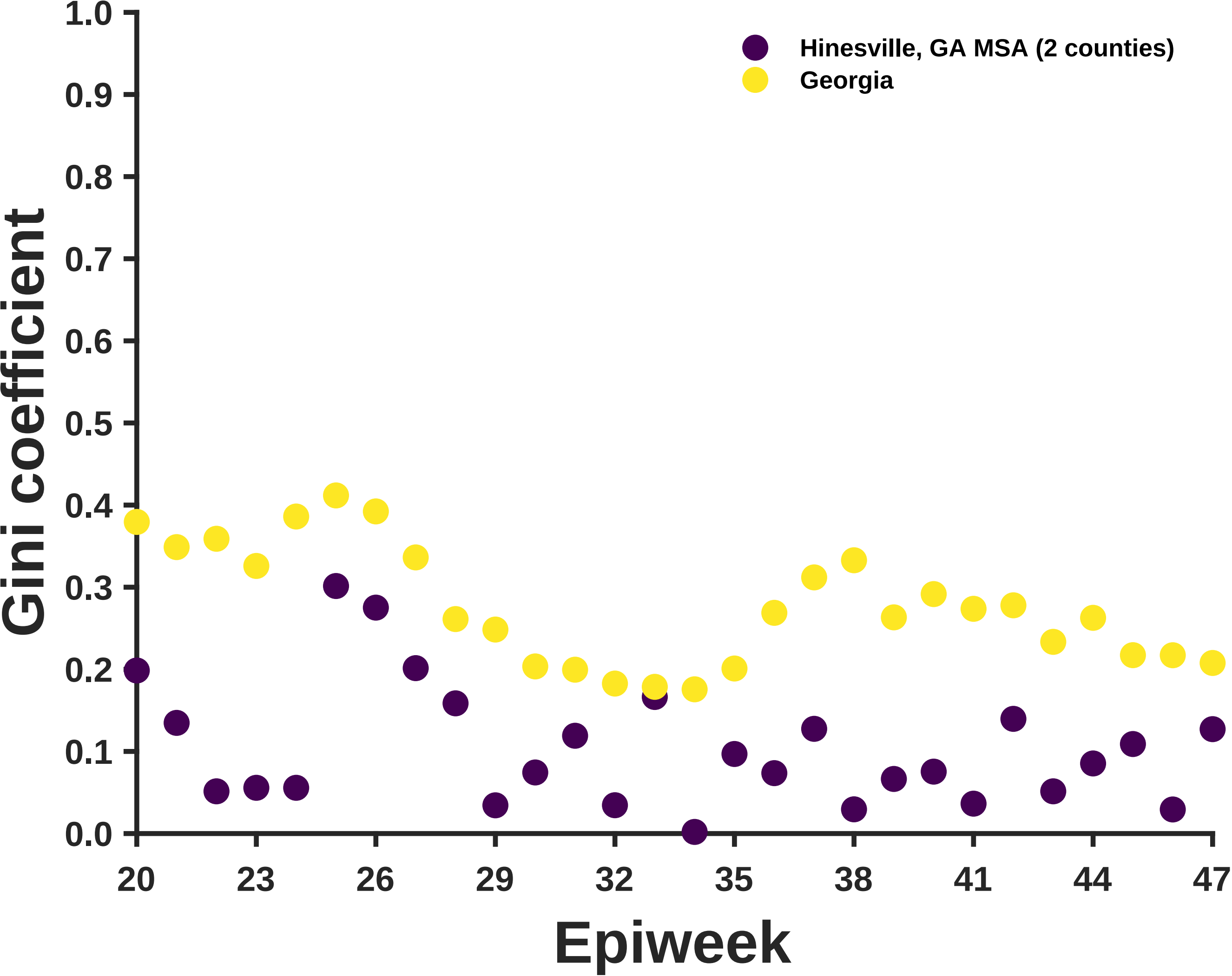

Hinesville, GA MSA (2 counties)
Georgia
Gini coefficient
Epiweek
1.0
0.9
0.8
0.7
0.6
0.5
0.4
0.3
0.2
0.1
0.0
20
23
26
29
32
35
38
41
44
47

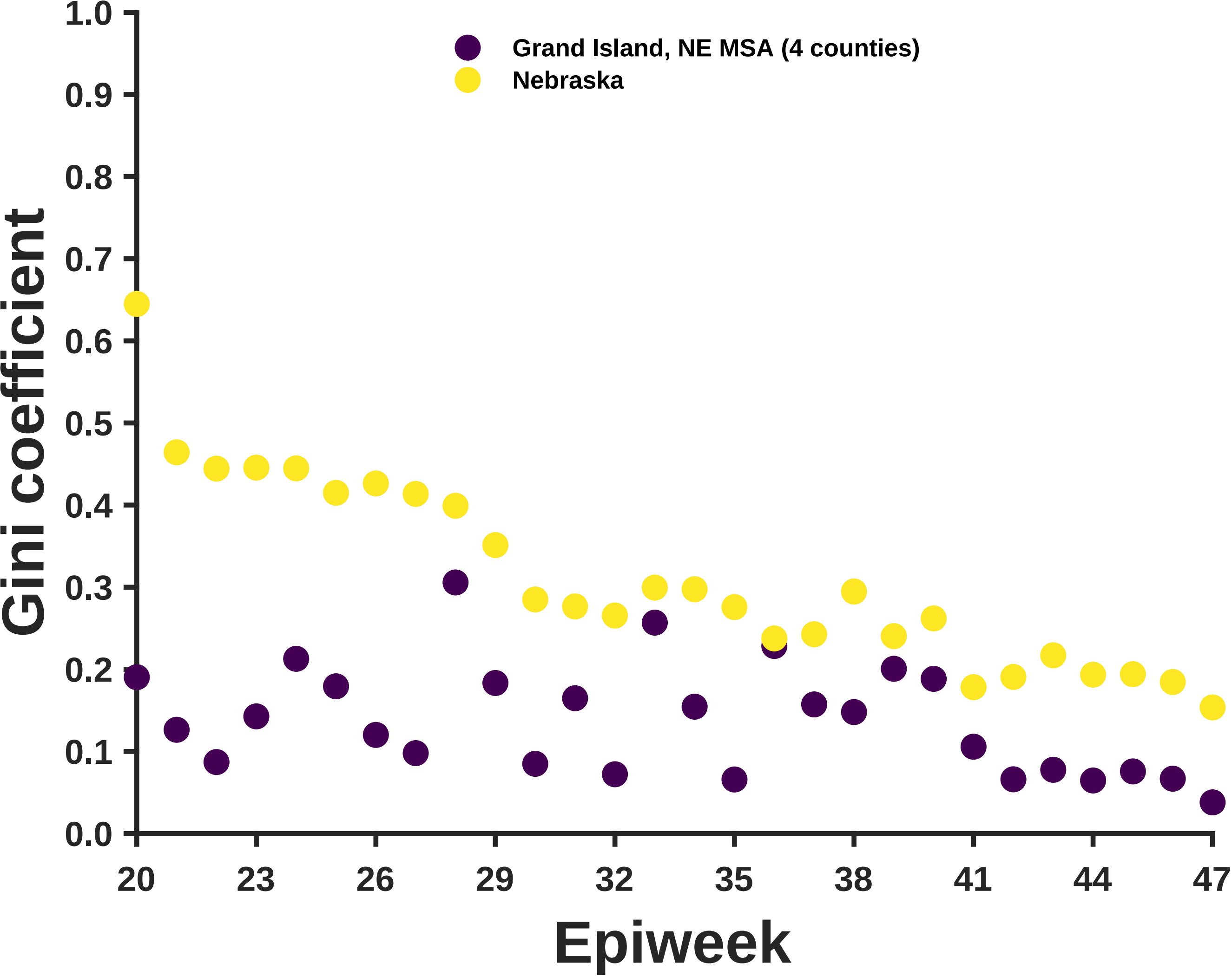
Grand Island, NE MSA (4 counties)
Nebraska
Gini coefficient
1.0
0.9
0.8
0.7
0.6
0.5
0.4
0.3
0.2
0.1
0.0
20
23
26
29
32
35
38
41
44
47
Epiweek

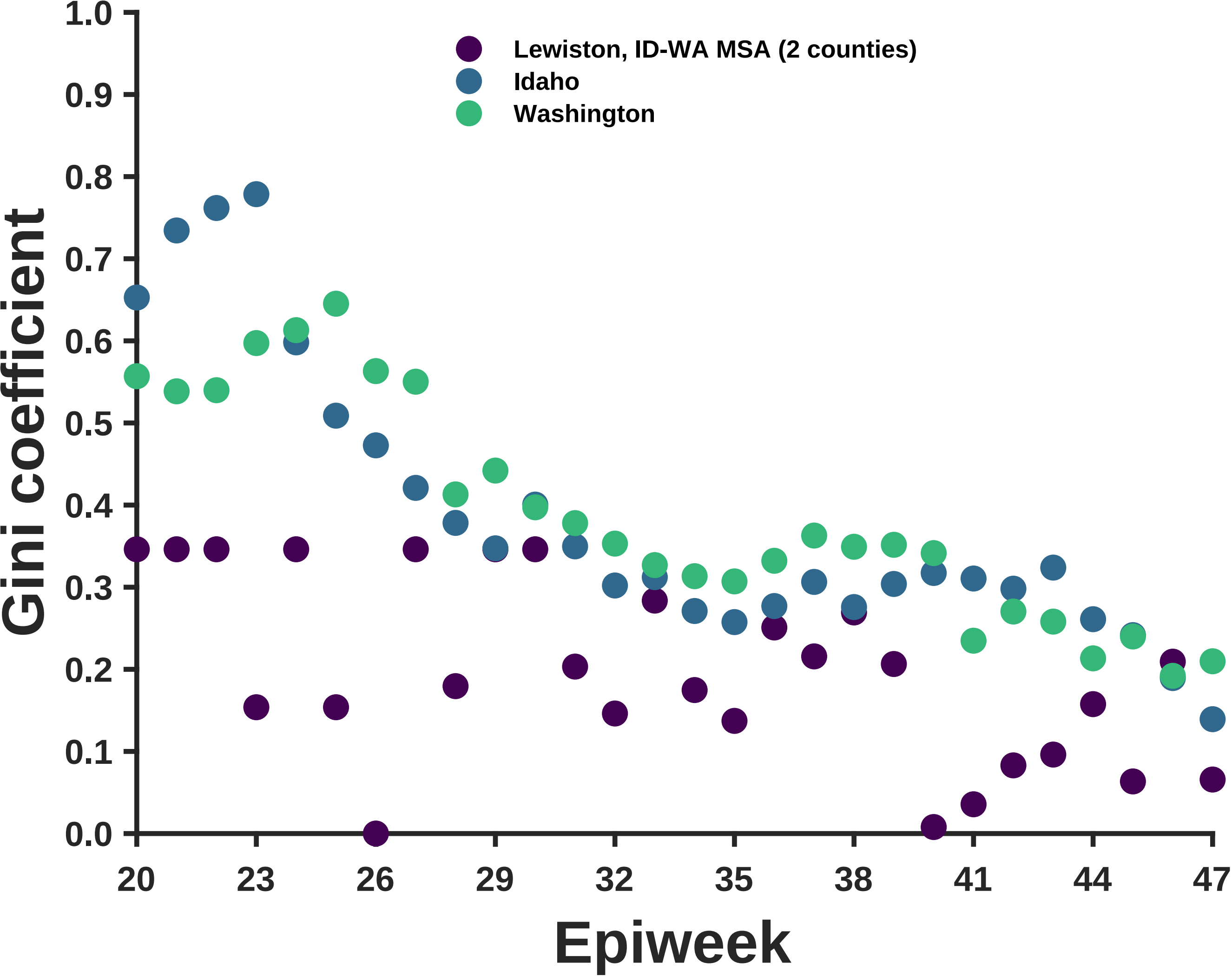

Lewiston, ID-WA MSA (2 counties)
Idaho
Washington
Gini coefficient
1.0
0.9
0.8
0.7
0.6
0.5
0.4
0.3
0.2
0.1
0.0
20
23
26
29
32
35
38
41
44
47
Epiweek
