## Supplementary material for "Differential contagiousness of respiratory disease across the United States": Figure S4

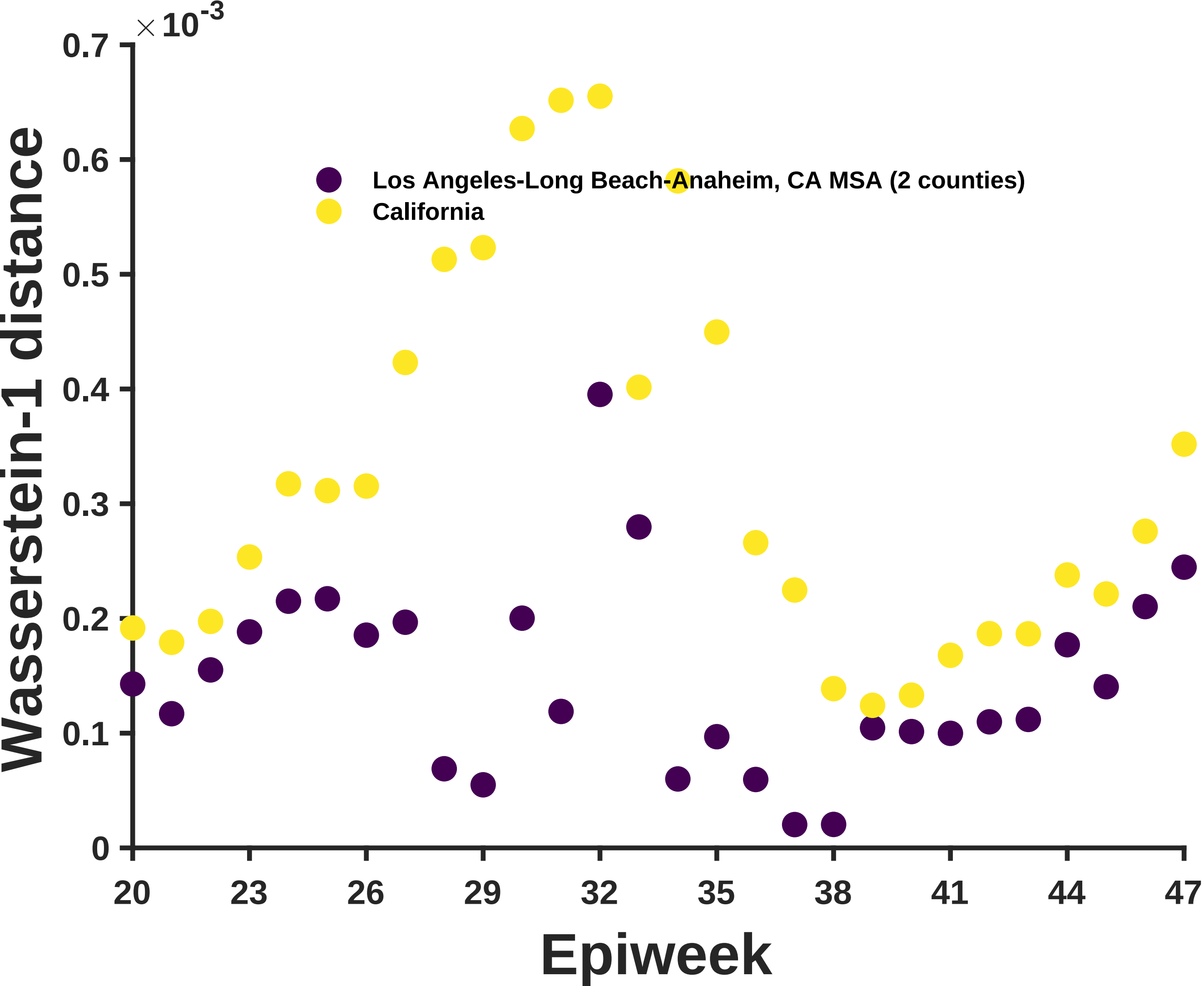

×10^-3
0.7
0.6
0.5
0.4
0.3
0.2
0.1
0
Wasserstein-1 distance
Los Angeles-Long Beach-Anaheim, CA MSA (2 counties)
California
20
23
26
29
32
35
38
41
44
47
Epiweek

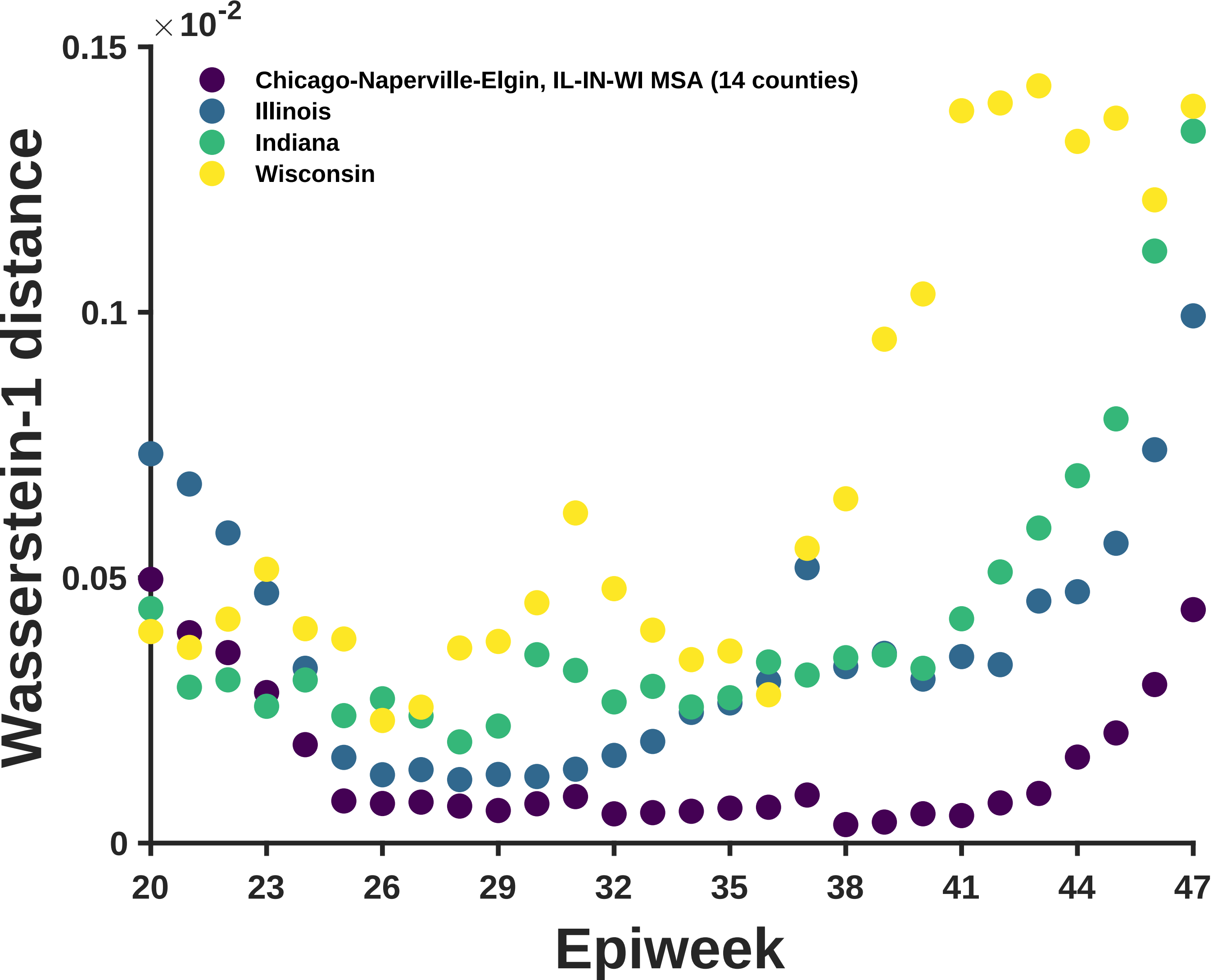

×10^-2
0.15
0.1
0.05
0
Wasserstein-1 distance
Chicago-Naperville-Elgin, IL-IN-WI MSA (14 counties)
Illinois
Indiana
Wisconsin
20
23
26
29
32
35
38
41
44
47
Epiweek

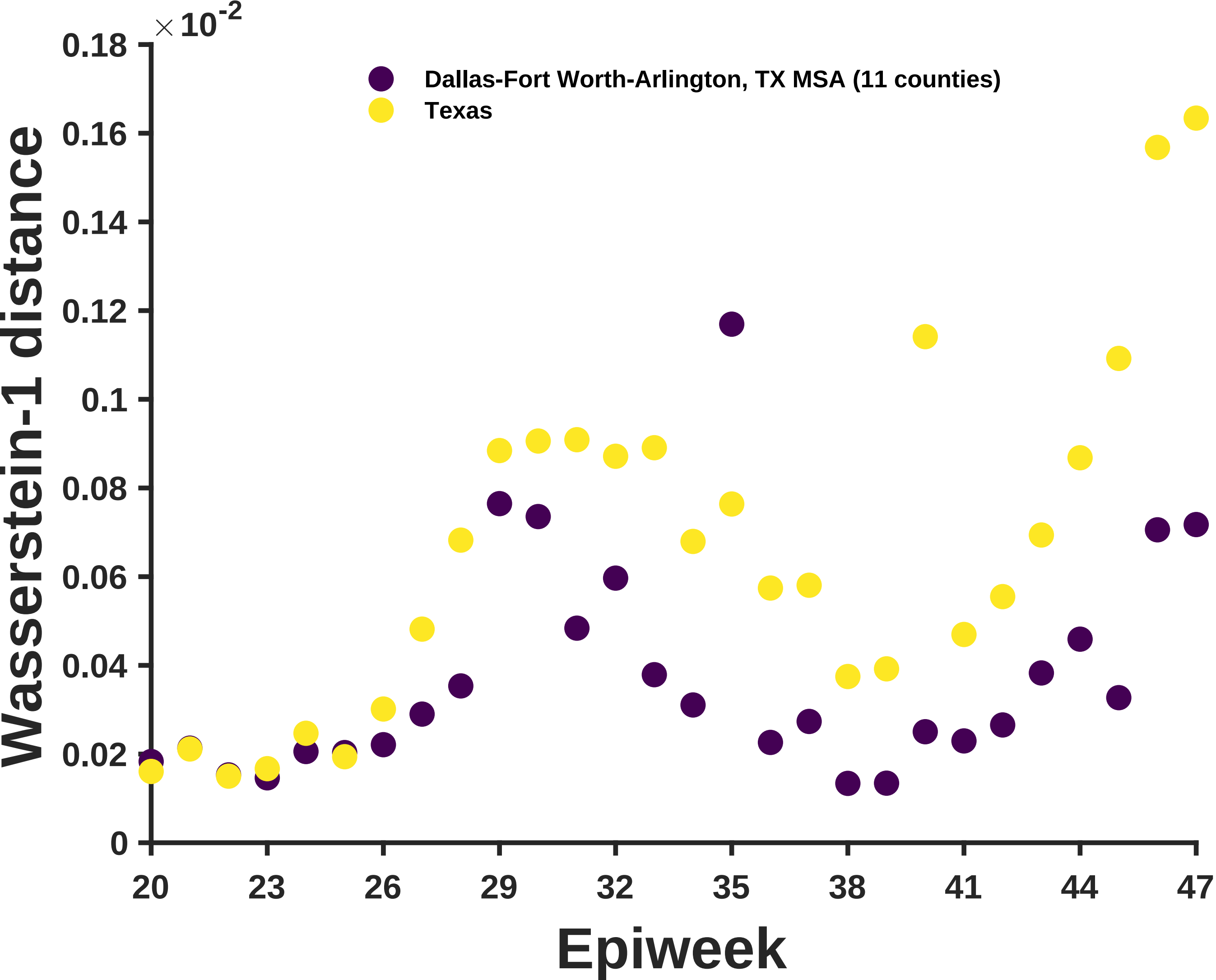
×10^-2
0.18
0.16
0.14
0.12
0.1
0.08
0.06
0.04
0.02
0
Wasserstein-1 distance
Dallas-Fort Worth-Arlington, TX MSA (11 counties)
Texas
20
23
26
29
32
35
38
41
44
47
Epiweek

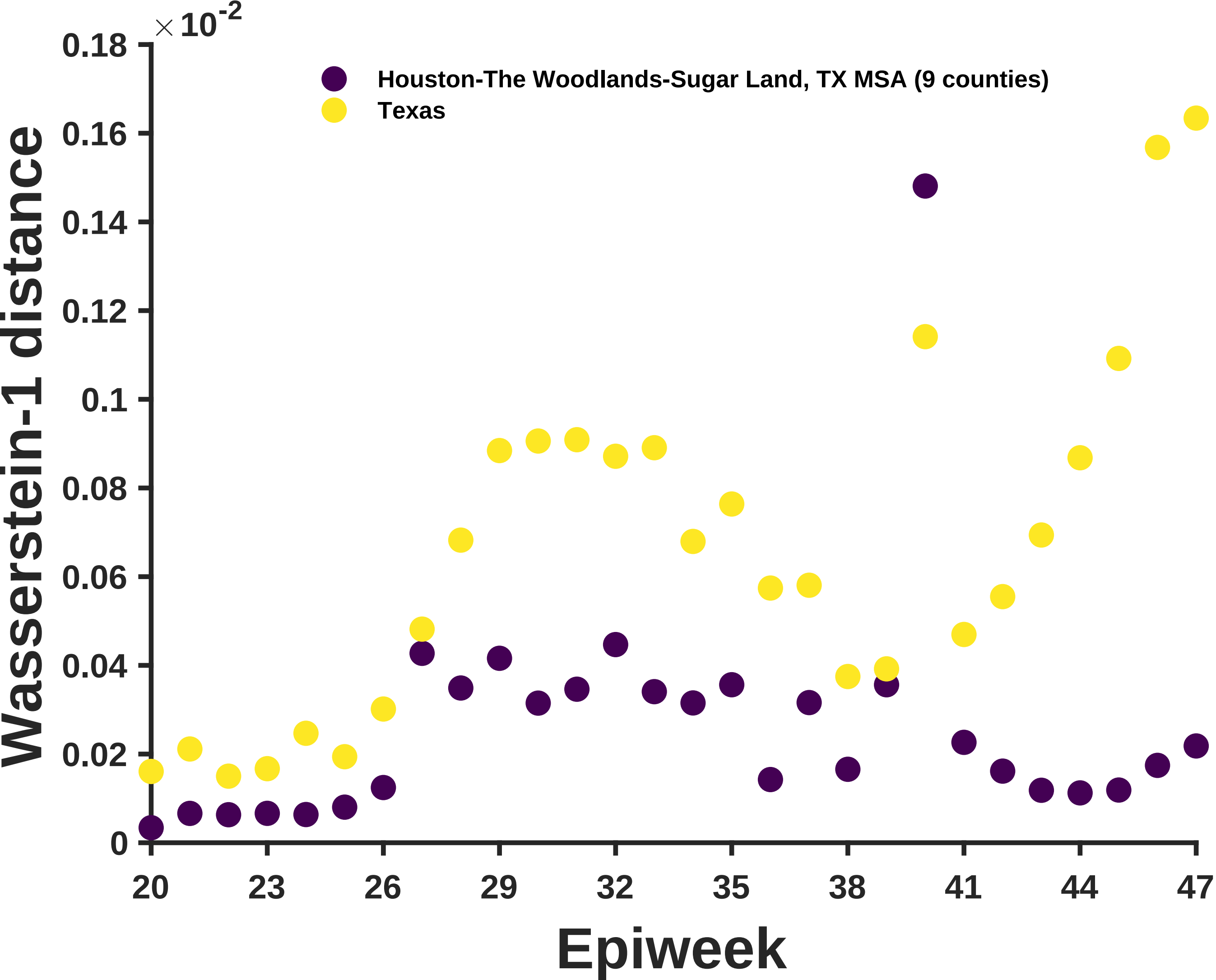

×10^-2
0.18
0.16
0.14
0.12
0.1
0.08
0.06
0.04
0.02
0
Wasserstein-1 distance
Houston-The Woodlands-Sugar Land, TX MSA (9 counties)
Texas
20
23
26
29
32
35
38
41
44
47
Epiweek

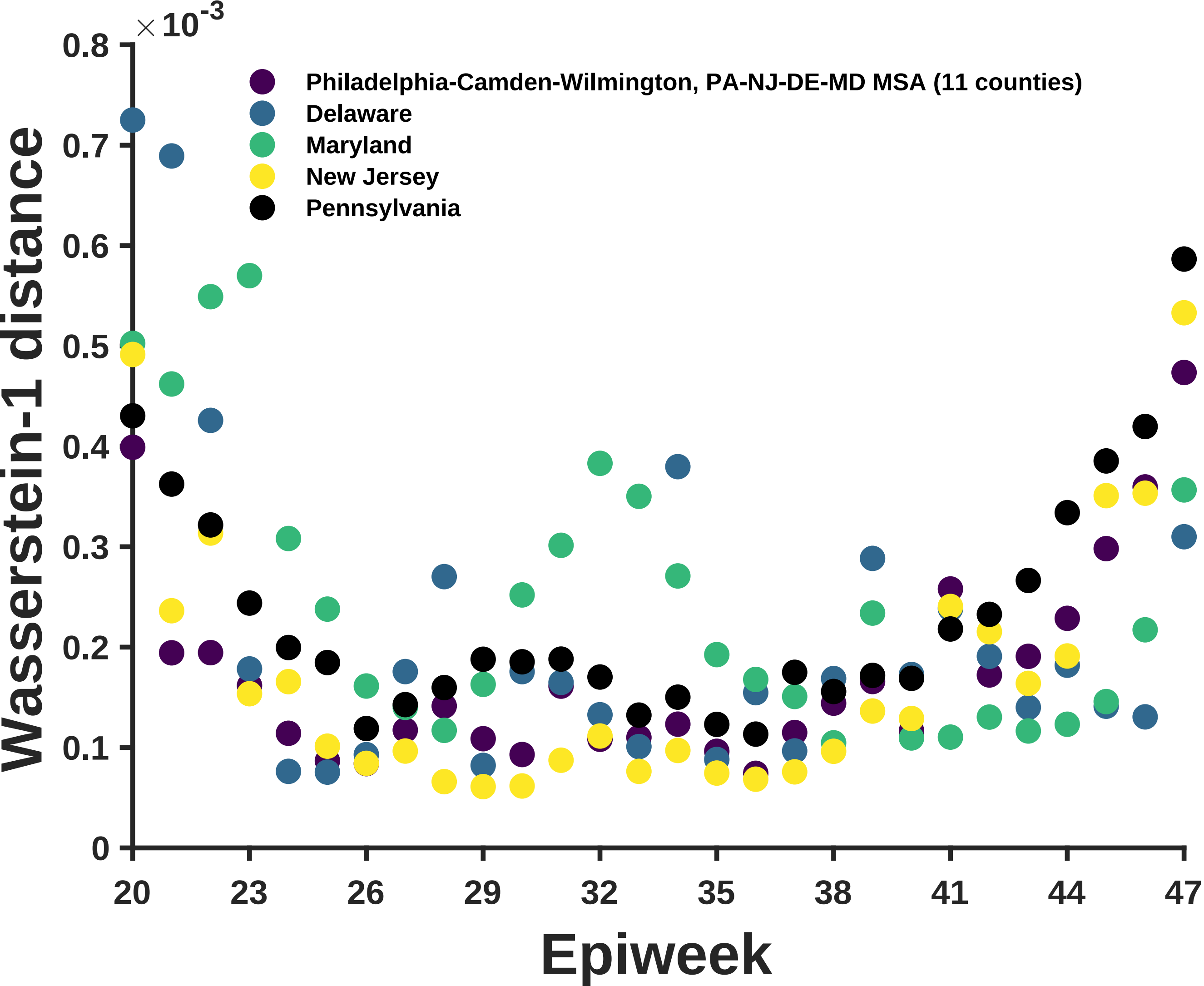

×10^-3
0.8
0.7
0.6
0.5
0.4
0.3
0.2
0.1
0
Wasserstein-1 distance
Philadelphia-Camden-Wilmington, PA-NJ-DE-MD MSA (11 counties)
Delaware
Maryland
New Jersey
Pennsylvania
20
23
26
29
32
35
38
41
44
47
Epiweek

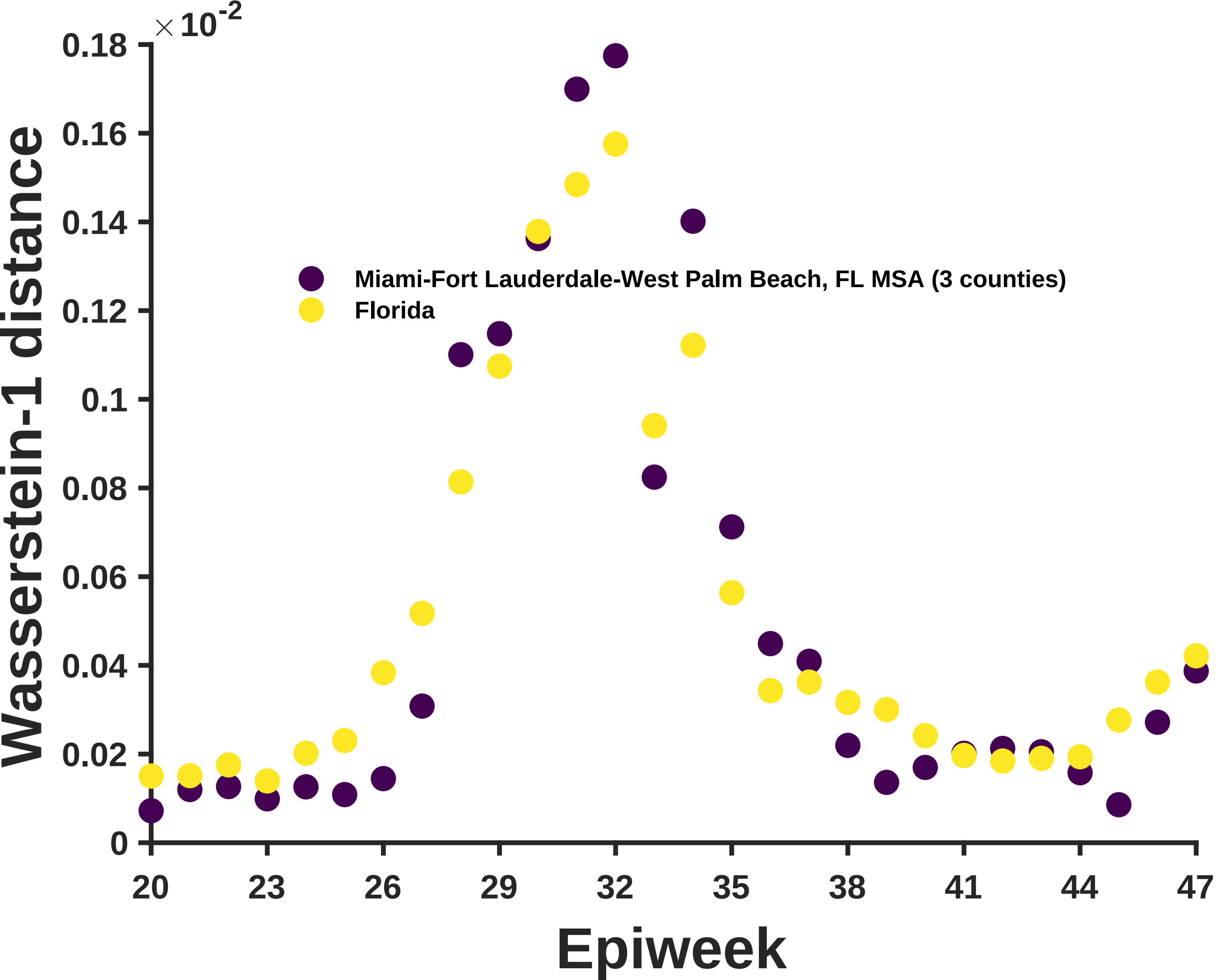

×10^-2
0.18
0.16
0.14
0.12
0.1
0.08
0.06
0.04
0.02
0
Wasserstein-1 distance
Miami-Fort Lauderdale-West Palm Beach, FL MSA (3 counties)
Florida
20
23
26
29
32
35
38
41
44
47
Epiweek

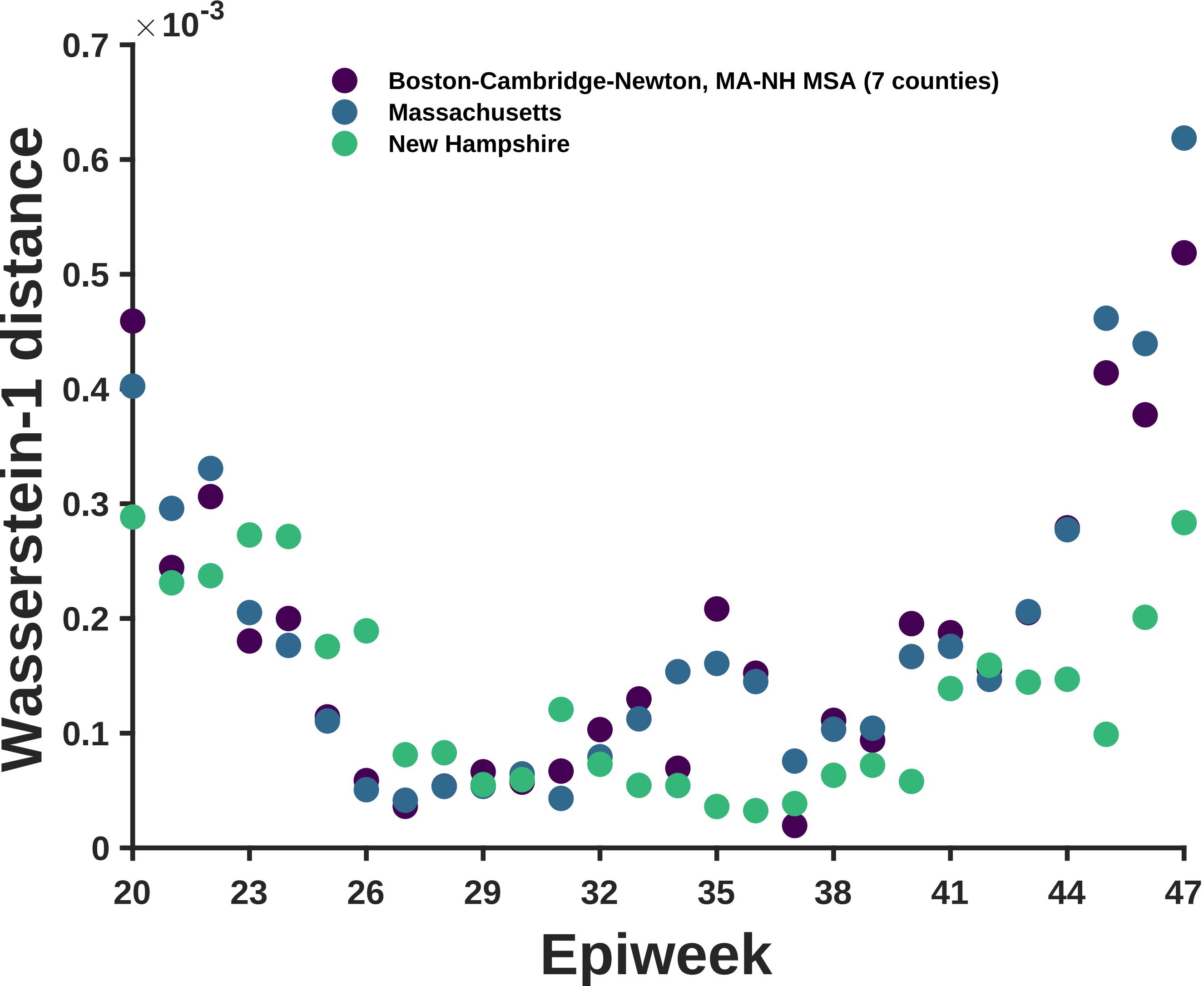

×10^-3
0.7
0.6
0.5
0.4
0.3
0.2
0.1
0
Wasserstein-1 distance
Boston-Cambridge-Newton, MA-NH MSA (7 counties)
Massachusetts
New Hampshire
20
23
26
29
32
35
38
41
44
47
Epiweek

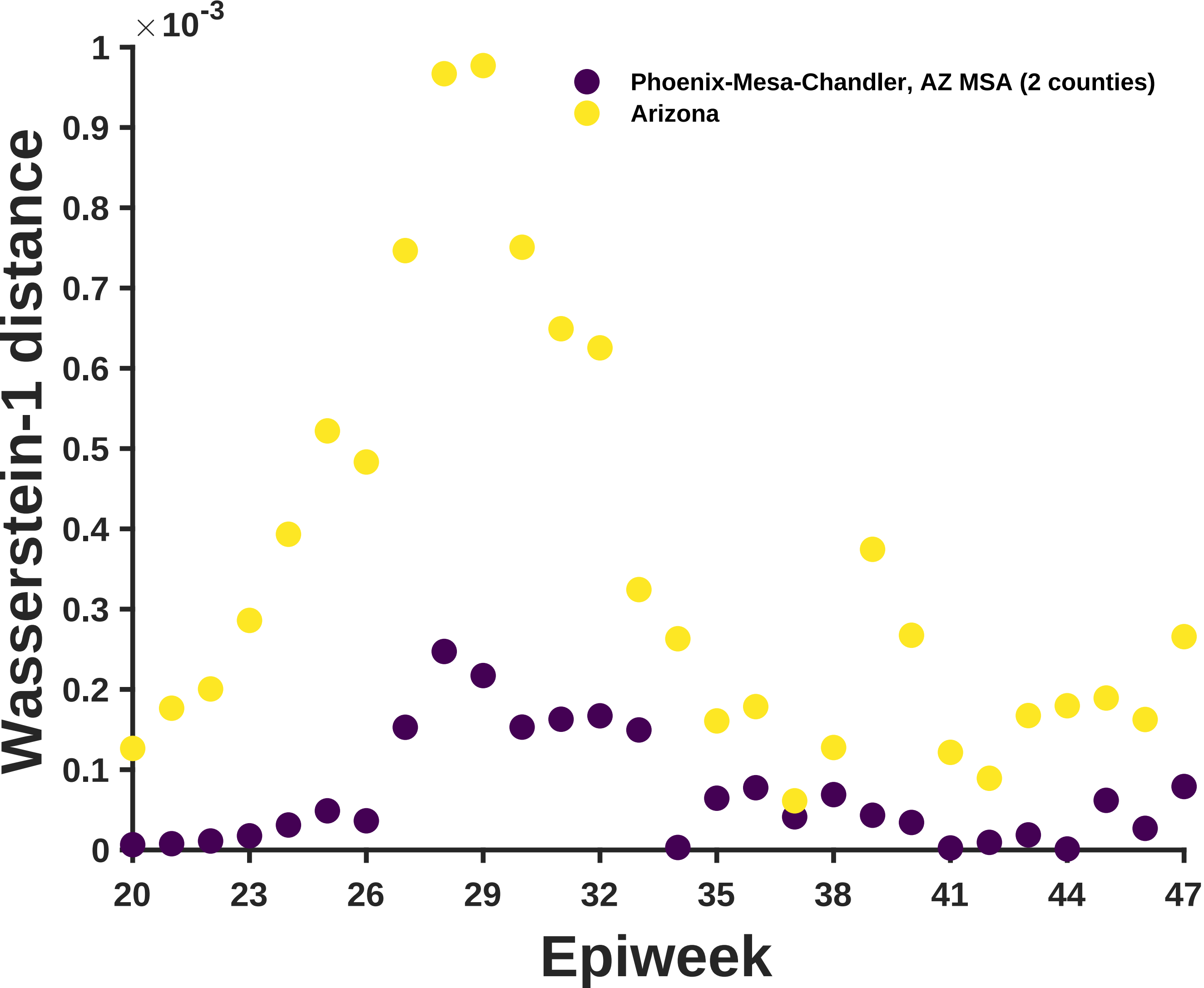

×10^-3
1
0.9
0.8
0.7
0.6
0.5
0.4
0.3
0.2
0.1
0
Wasserstein-1 distance
20
23
26
29
32
35
38
41
44
47
Epiweek
Phoenix-Mesa-Chandler, AZ MSA (2 counties)
Arizona

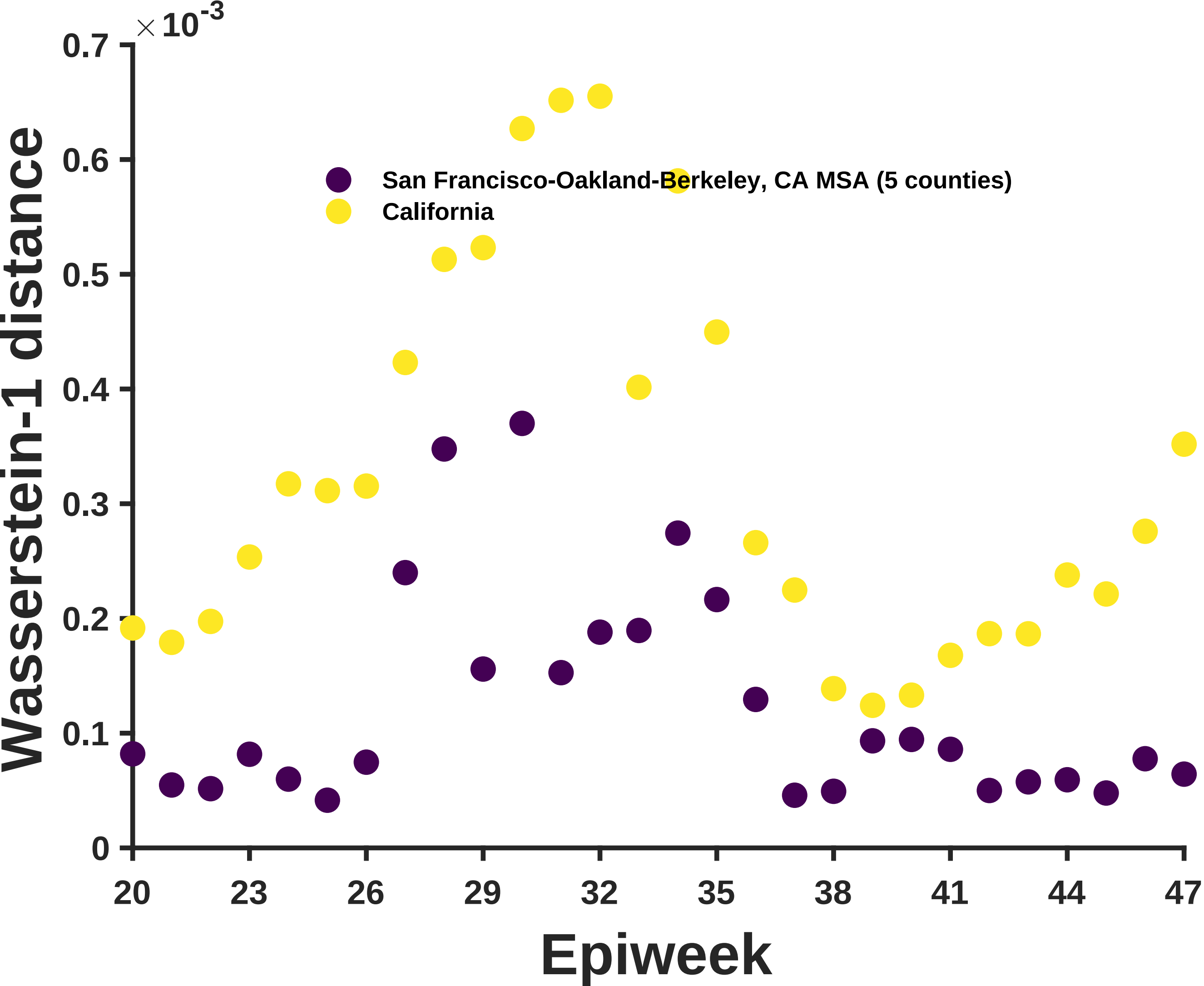

×10^-3
San Francisco-Oakland-Berkeley, CA MSA (5 counties)
California
Wasserstein-1 distance
Epiweek
0
0.1
0.2
0.3
0.4
0.5
0.6
0.7
20
23
26
29
32
35
38
41
44
47

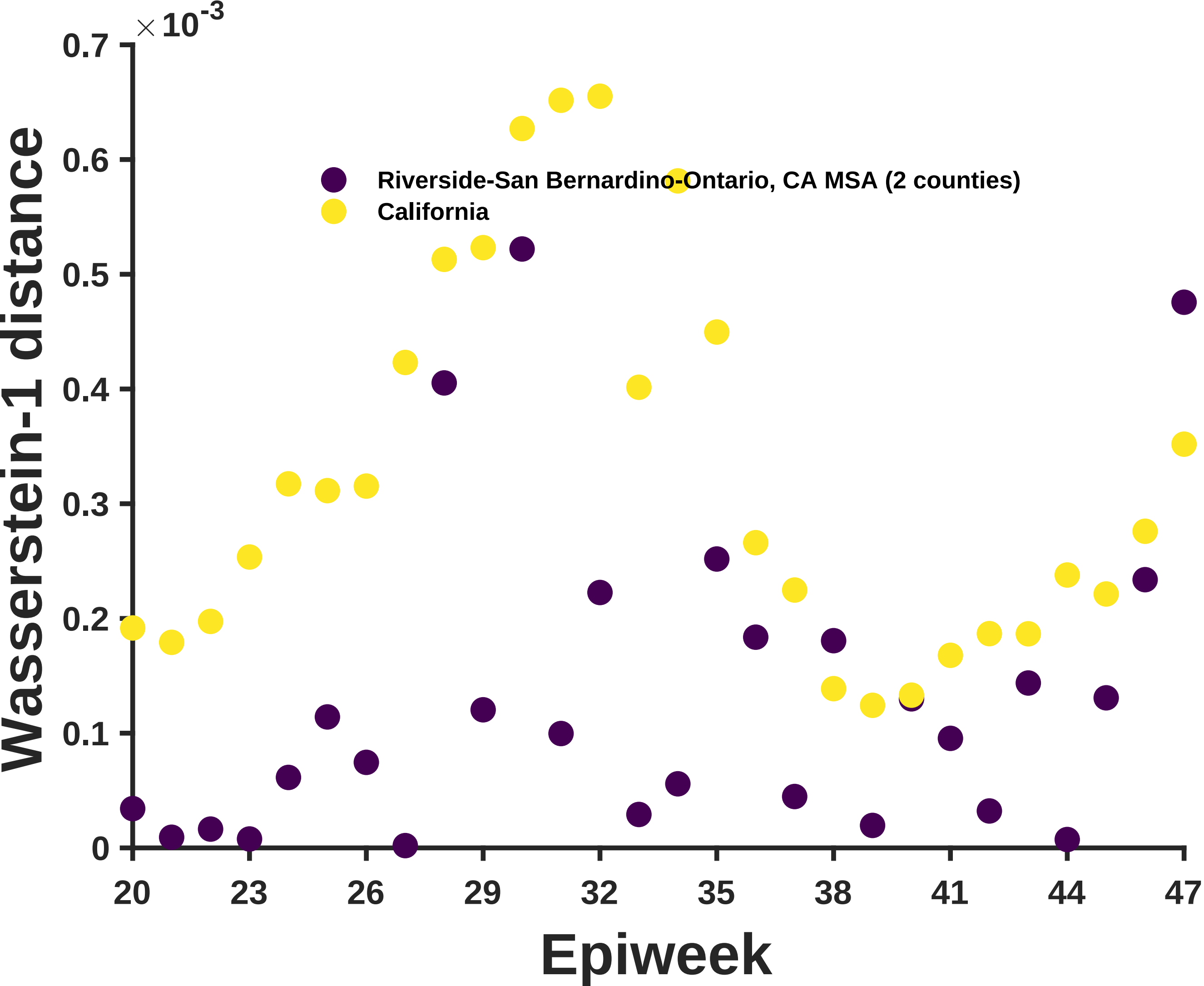

×10^-3
0.7
0.6
0.5
0.4
0.3
0.2
0.1
0
Wasserstein-1 distance
Riverside-San Bernardino-Ontario, CA MSA (2 counties)
California
20
23
26
29
32
35
38
41
44
47
Epiweek

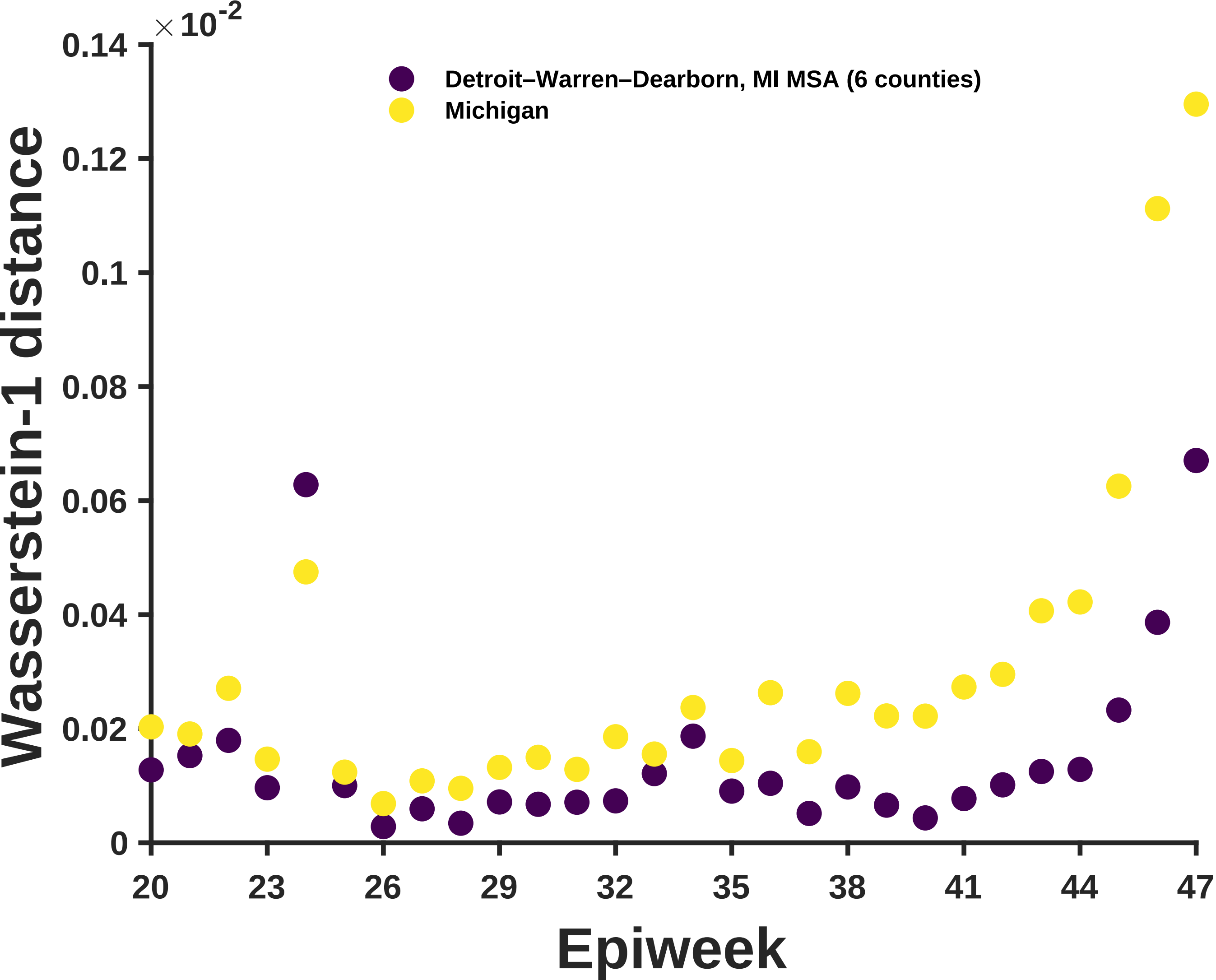

×10^-2
0.14
0.12
0.1
0.08
0.06
0.04
0.02
0
Wasserstein-1 distance
Detroit–Warren–Dearborn, MI MSA (6 counties)
Michigan
20
23
26
29
32
35
38
41
44
47
Epiweek

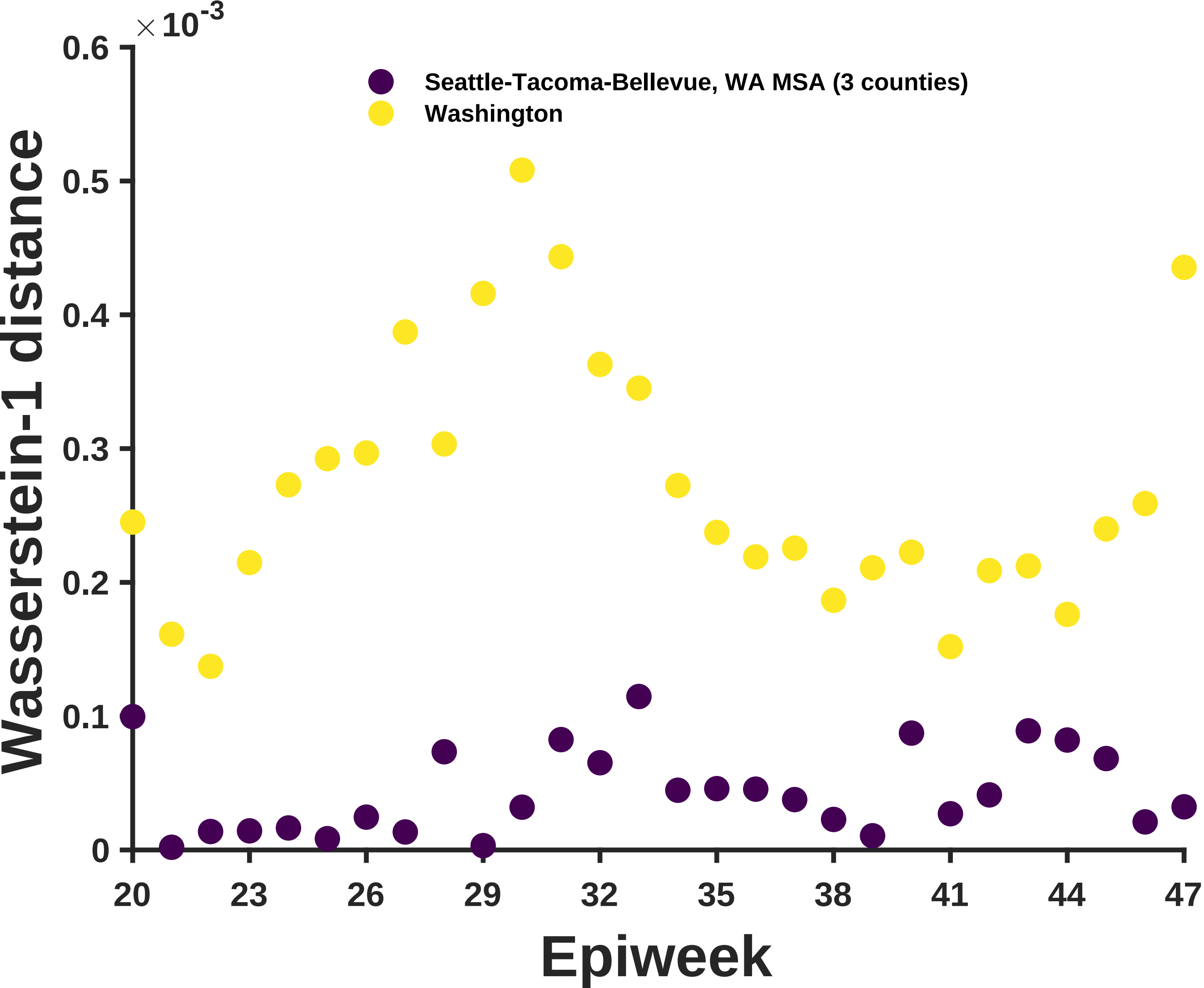
×10^-3
0.6
0.5
0.4
0.3
0.2
0.1
0
Wasserstein-1 distance
Seattle-Tacoma-Bellevue, WA MSA (3 counties)
Washington
20
23
26
29
32
35
38
41
44
47
Epiweek

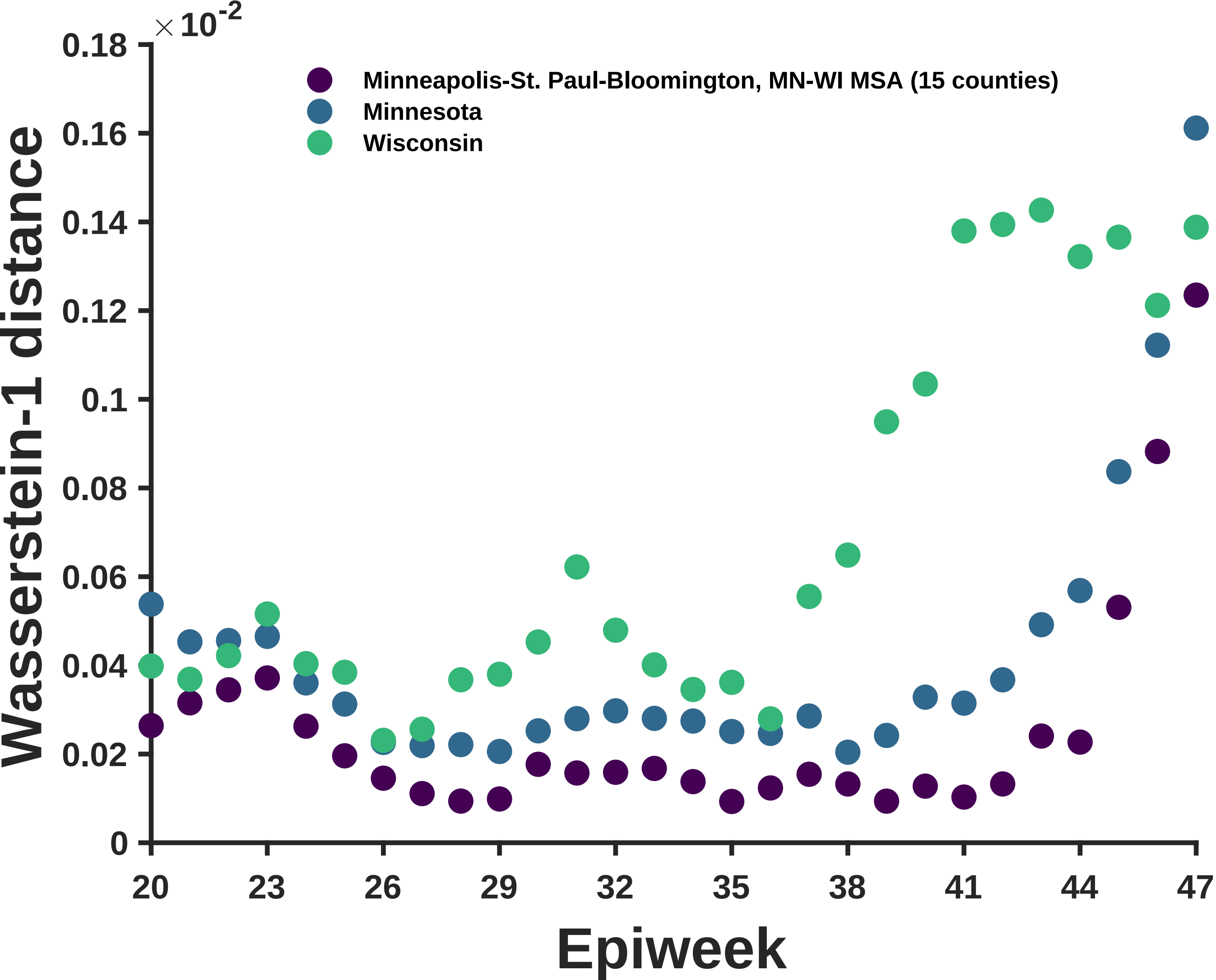

×10^-2
0.18
0.16
0.14
0.12
0.1
0.08
0.06
0.04
0.02
0
Wasserstein-1 distance
Minneapolis-St. Paul-Bloomington, MN-WI MSA (15 counties)
Minnesota
Wisconsin
20
23
26
29
32
35
38
41
44
47
Epiweek

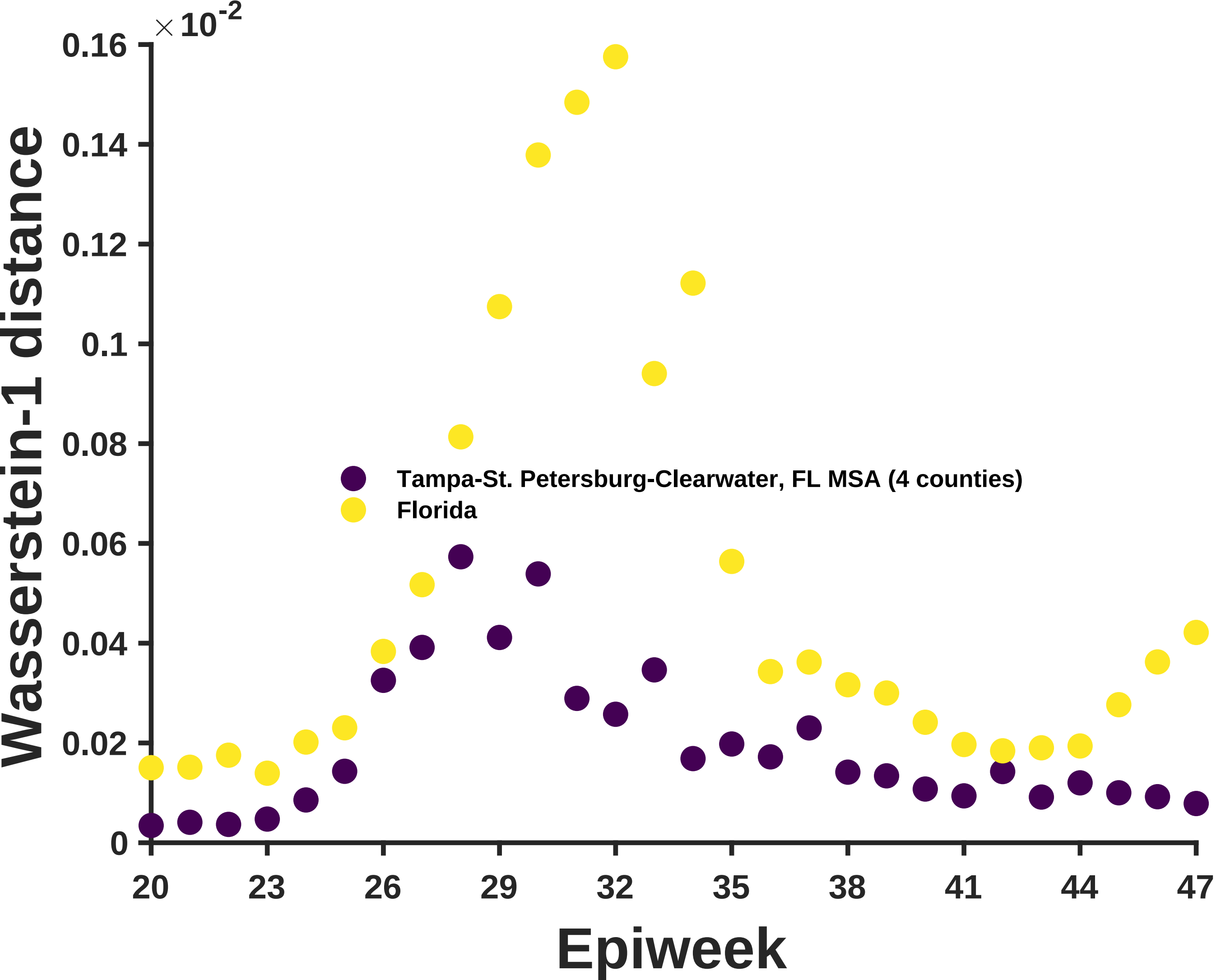

×10^-2
0.16
0.14
0.12
0.1
0.08
0.06
0.04
0.02
0
Wasserstein-1 distance
Tampa-St. Petersburg-Clearwater, FL MSA (4 counties)
Florida
20
23
26
29
32
35
38
41
44
47
Epiweek

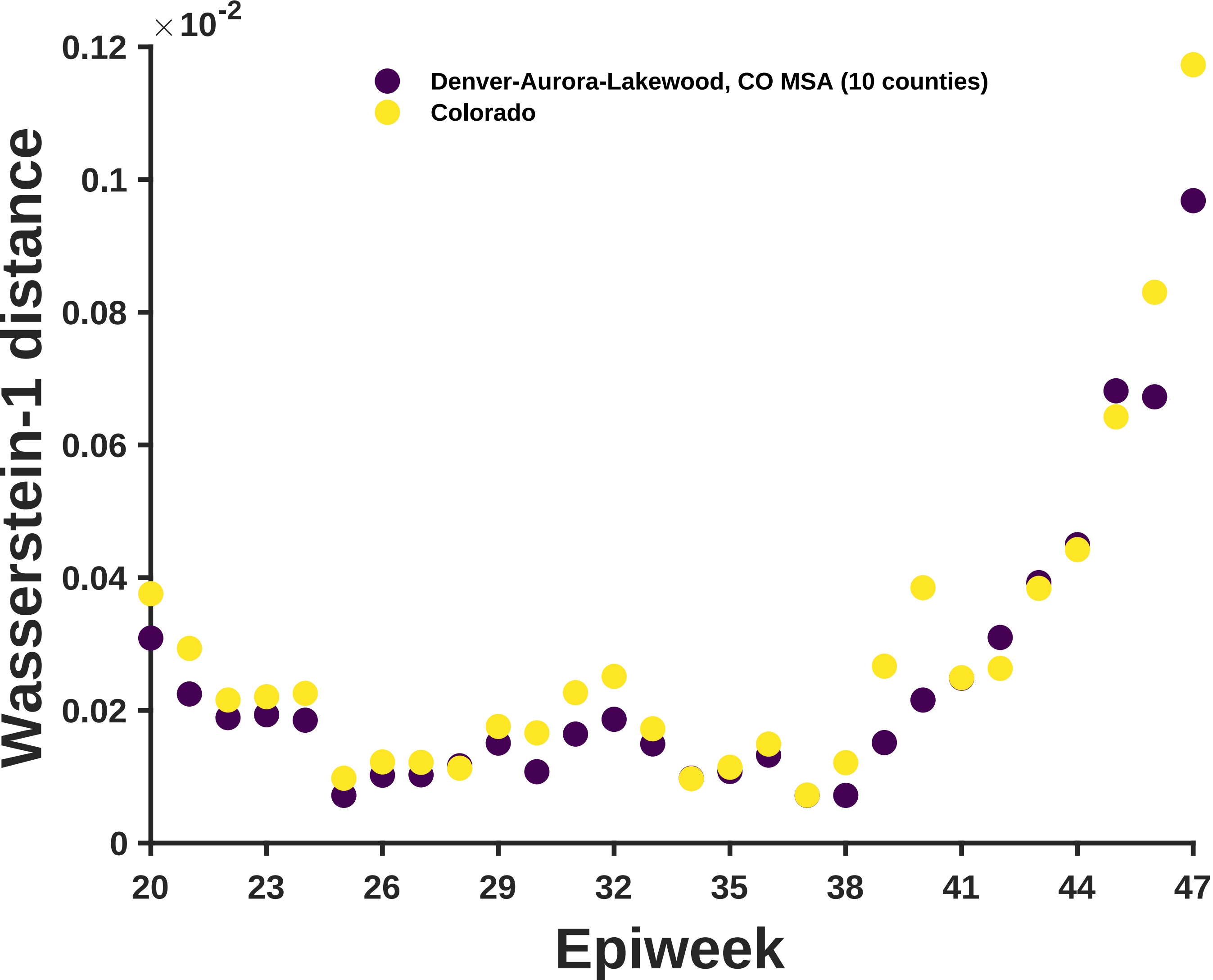

×10^-2
Denver-Aurora-Lakewood, CO MSA (10 counties)
Colorado
Wasserstein-1 distance
0.12
0.1
0.08
0.06
0.04
0.02
0
20
23
26
29
32
35
38
41
44
47
Epiweek

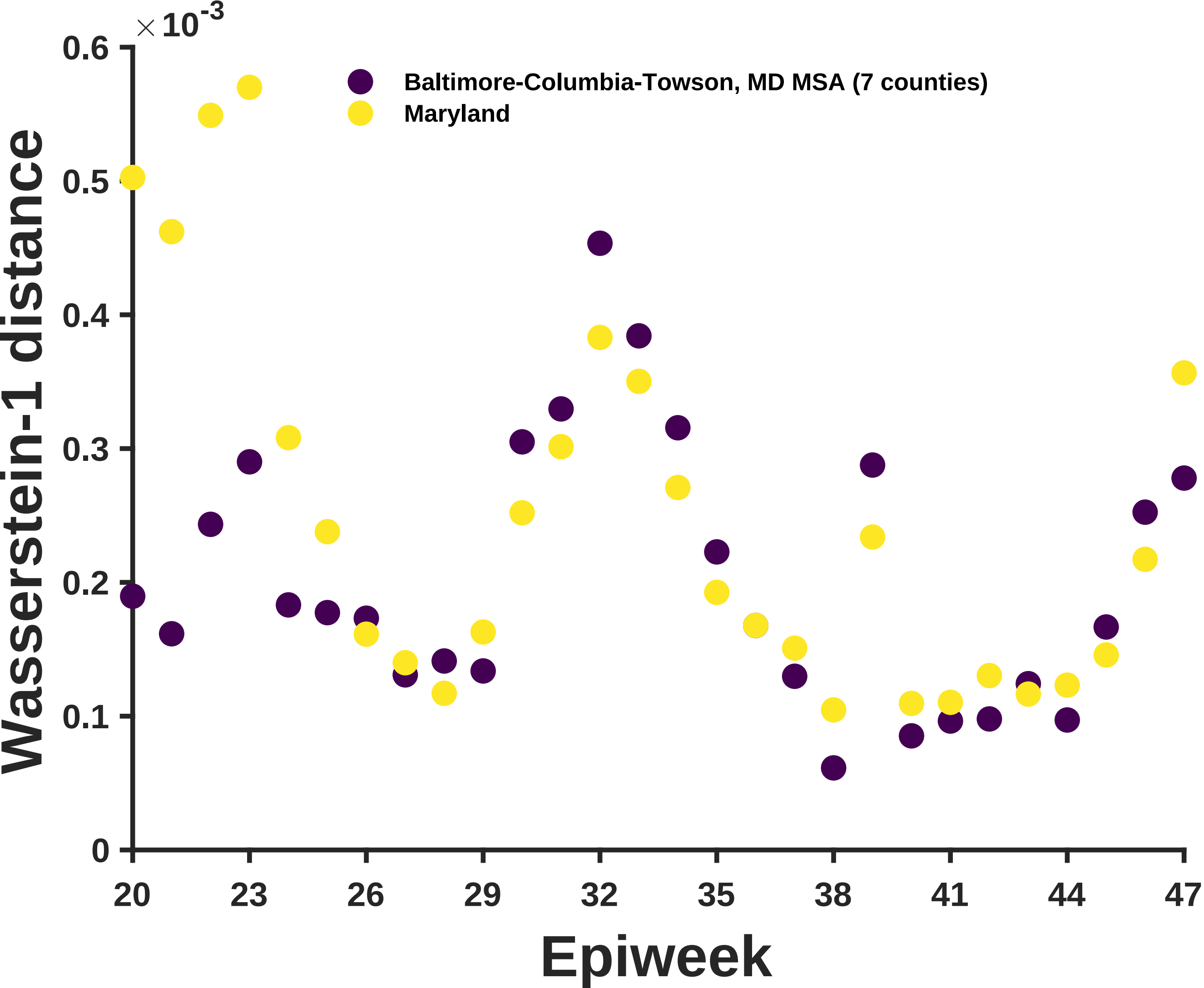

×10^-3
0.6
0.5
0.4
0.3
0.2
0.1
0
Wasserstein-1 distance
Baltimore-Columbia-Towson, MD MSA (7 counties)
Maryland
20
23
26
29
32
35
38
41
44
47
Epiweek

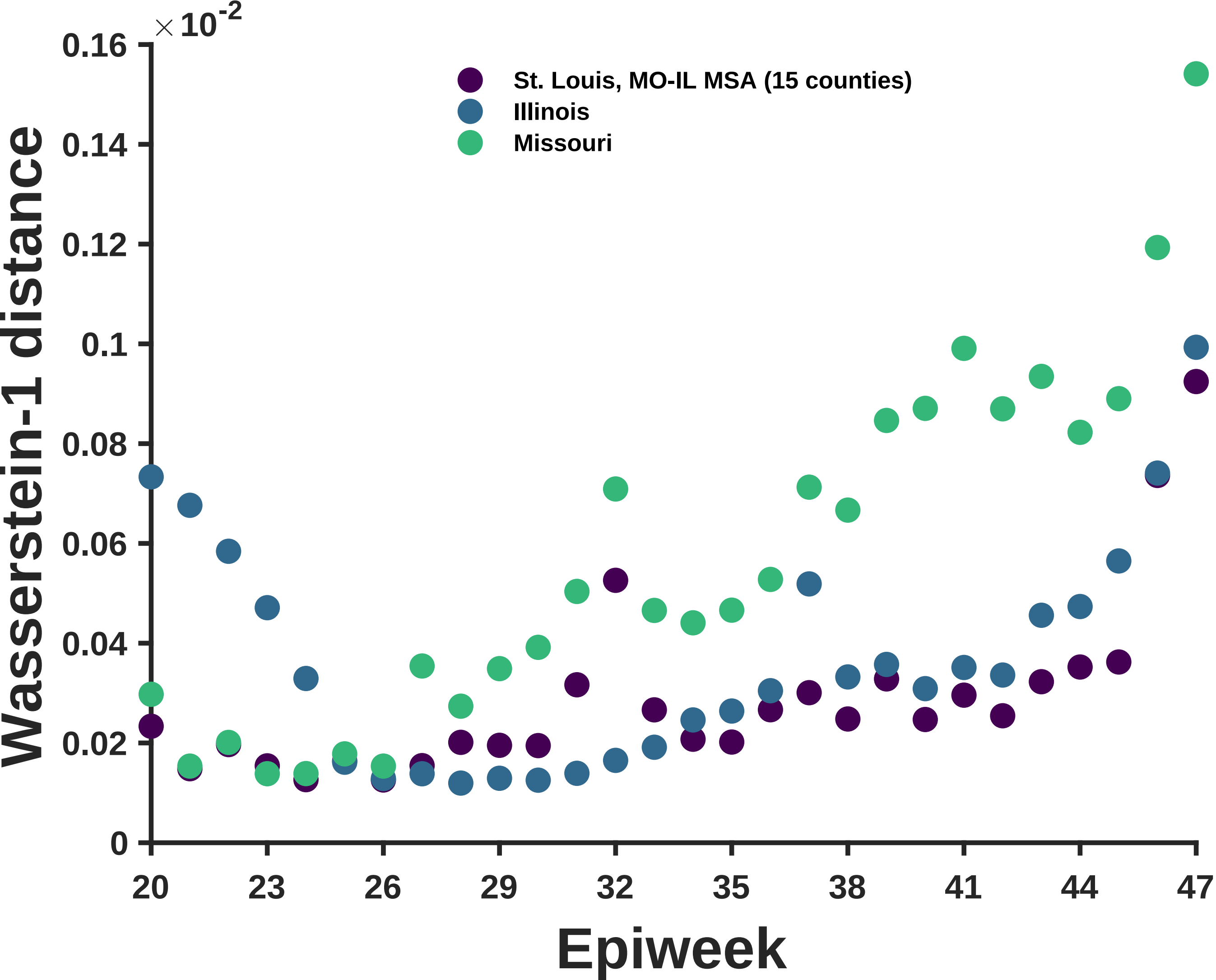

× 10^-2
0.16
0.14
0.12
0.1
0.08
0.06
0.04
0.02
0
Wasserstein-1 distance
St. Louis, MO-IL MSA (15 counties)
Illinois
Missouri
20
23
26
29
32
35
38
41
44
47
Epiweek

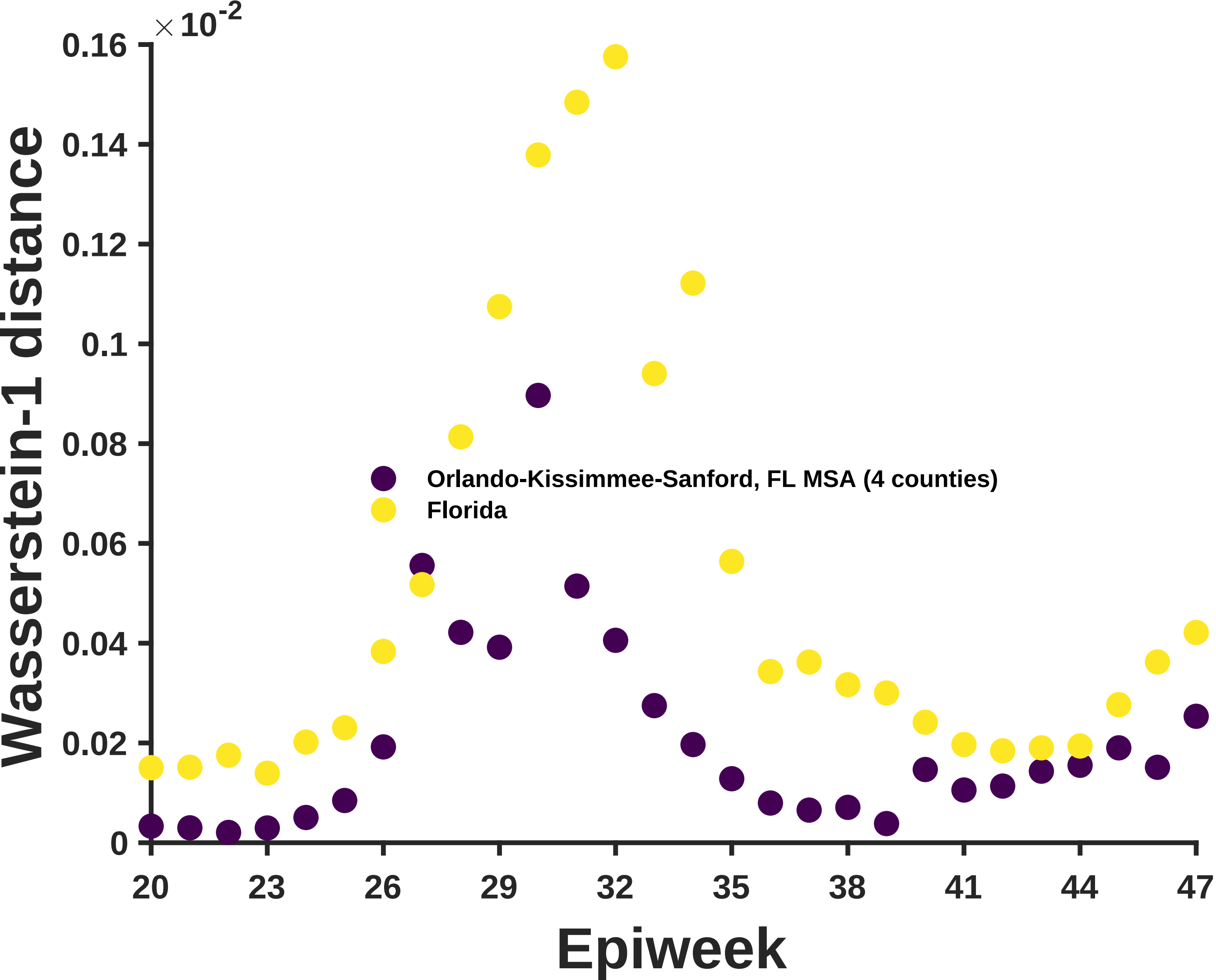

×10^-2
0.16
0.14
0.12
0.1
0.08
0.06
0.04
0.02
0
Wasserstein-1 distance
Orlando-Kissimmee-Sanford, FL MSA (4 counties)
Florida
20
23
26
29
32
35
38
41
44
47
Epiweek

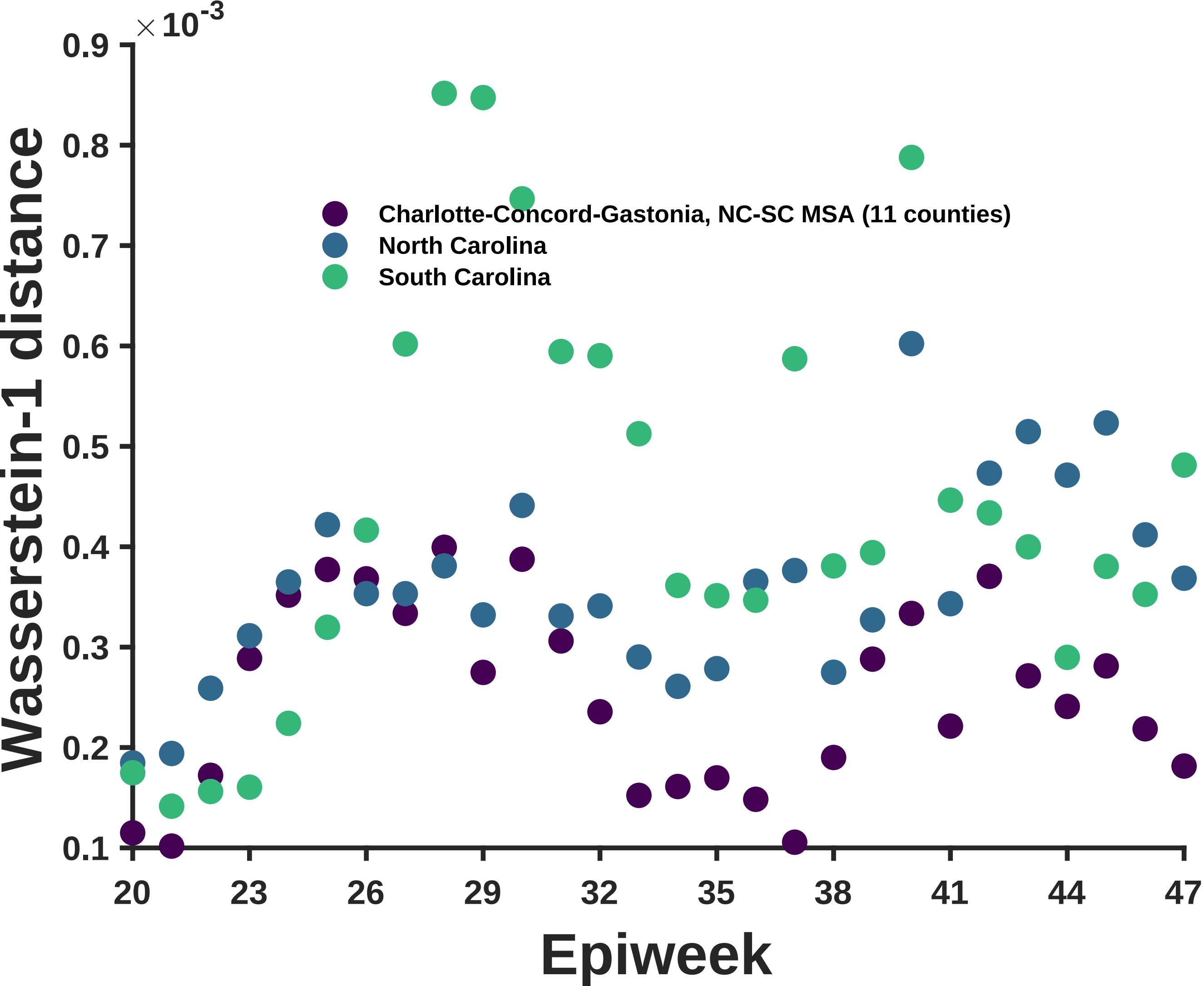

×10^-3
0.9
0.8
0.7
0.6
0.5
0.4
0.3
0.2
0.1
Wasserstein-1 distance
Charlotte-Concord-Gastonia, NC-SC MSA (11 counties)
North Carolina
South Carolina
20
23
26
29
32
35
38
41
44
47
Epiweek

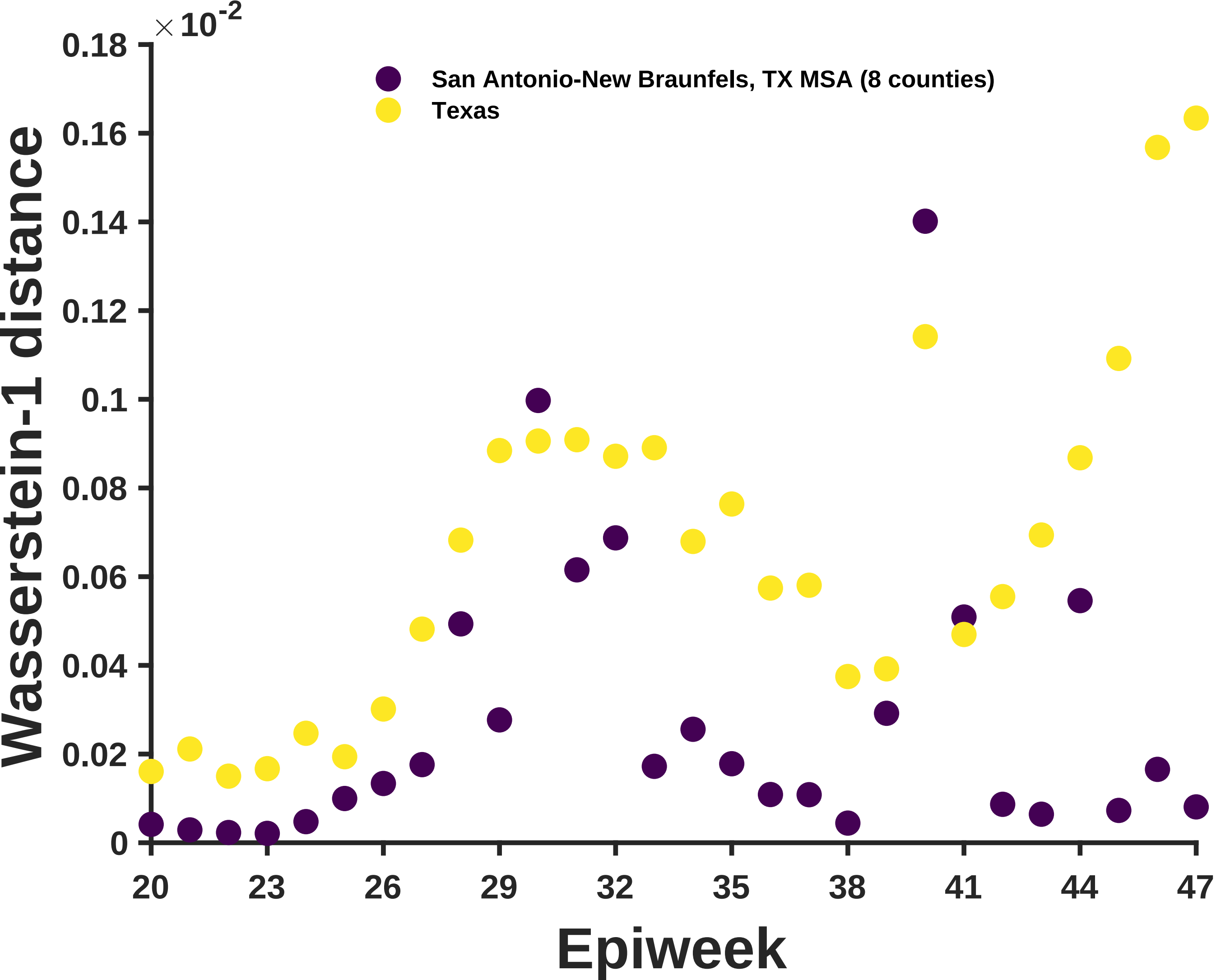

×10^-2
0.18
0.16
0.14
0.12
0.1
0.08
0.06
0.04
0.02
0
Wasserstein-1 distance
San Antonio-New Braunfels, TX MSA (8 counties)
Texas
20
23
26
29
32
35
38
41
44
47
Epiweek

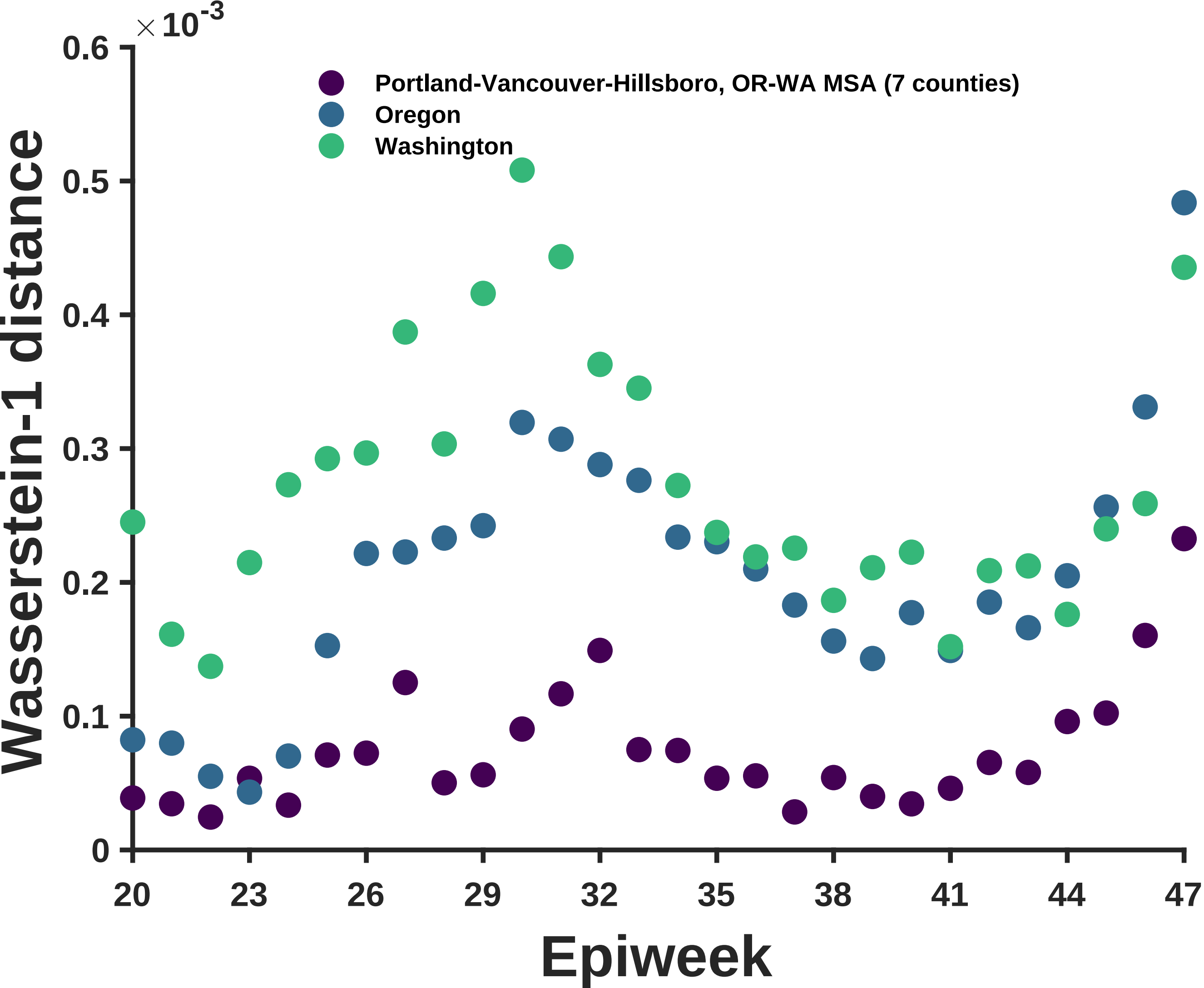

×10^-3
0.6
0.5
0.4
0.3
0.2
0.1
0
Wasserstein-1 distance
Portland-Vancouver-Hillsboro, OR-WA MSA (7 counties)
Oregon
Washington
20
23
26
29
32
35
38
41
44
47
Epiweek

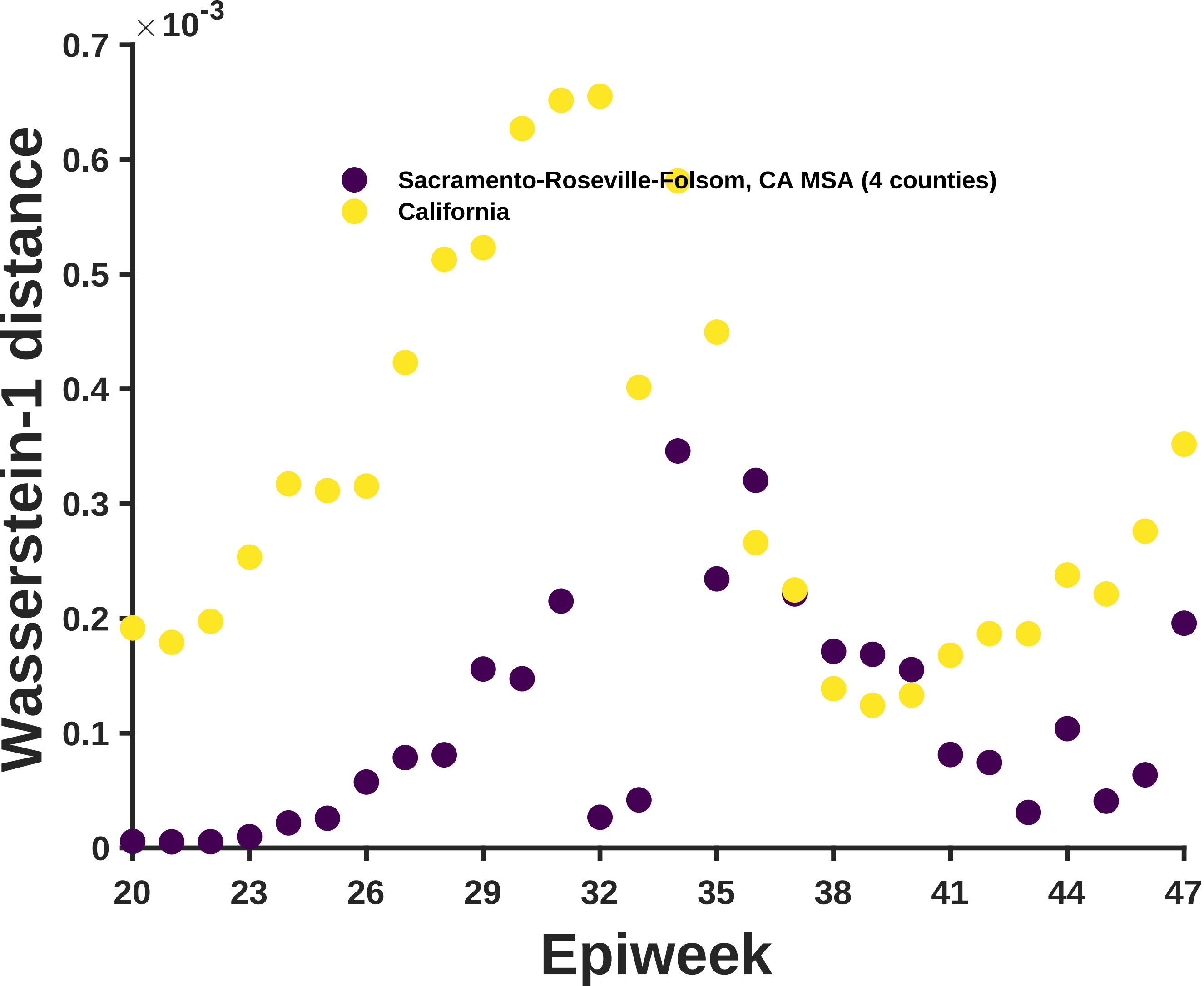

×10-3
0.7
0.6
0.5
0.4
0.3
0.2
0.1
0
Wasserstein-1 distance
Sacramento-Roseville-Folsom, CA MSA (4 counties)
California
20
23
26
29
32
35
38
41
44
47
Epiweek

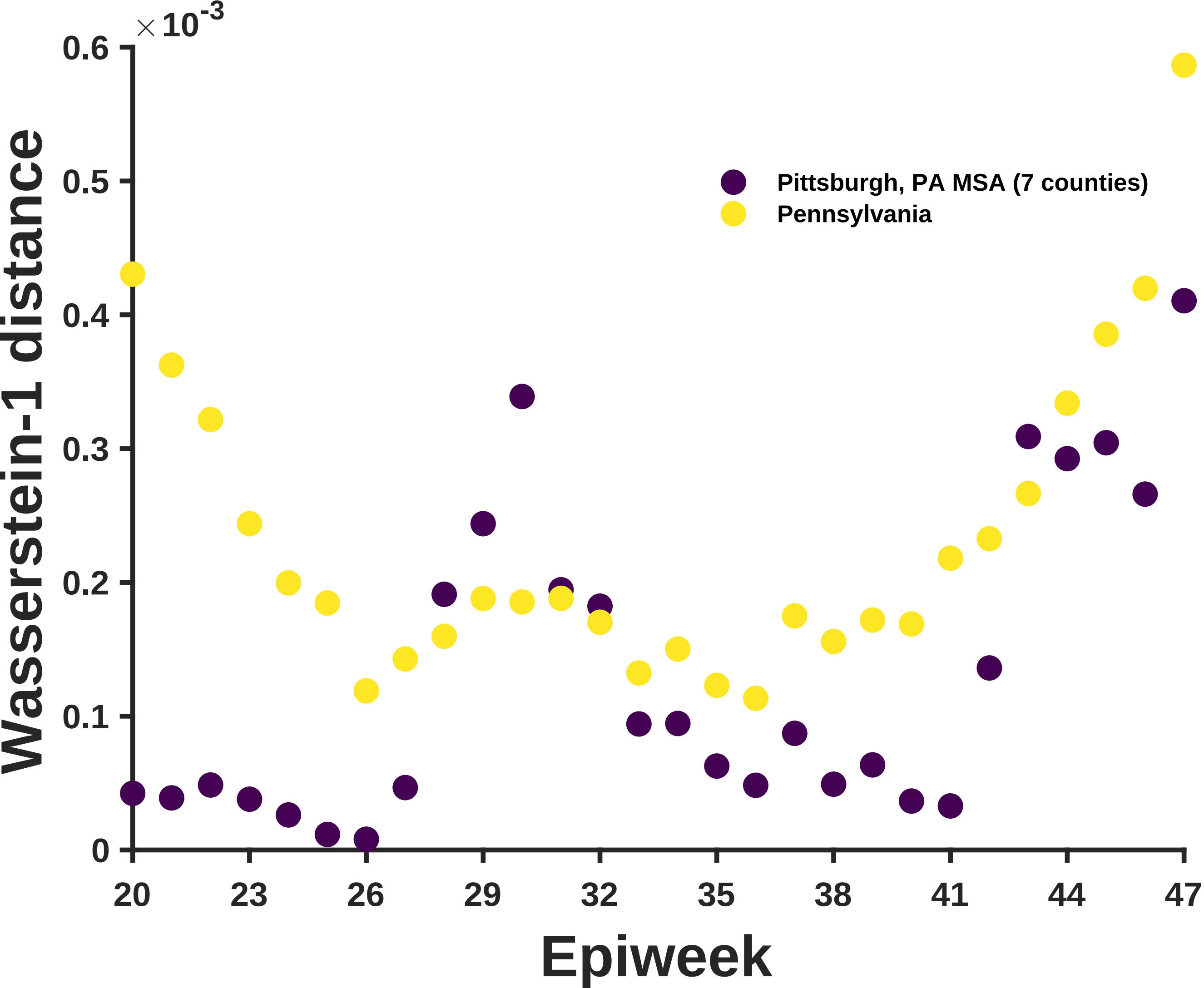

×10^-3
0.6
0.5
0.4
0.3
0.2
0.1
0
Wasserstein-1 distance
20
23
26
29
32
35
38
41
44
47
Epiweek
Pittsburgh, PA MSA (7 counties)
Pennsylvania

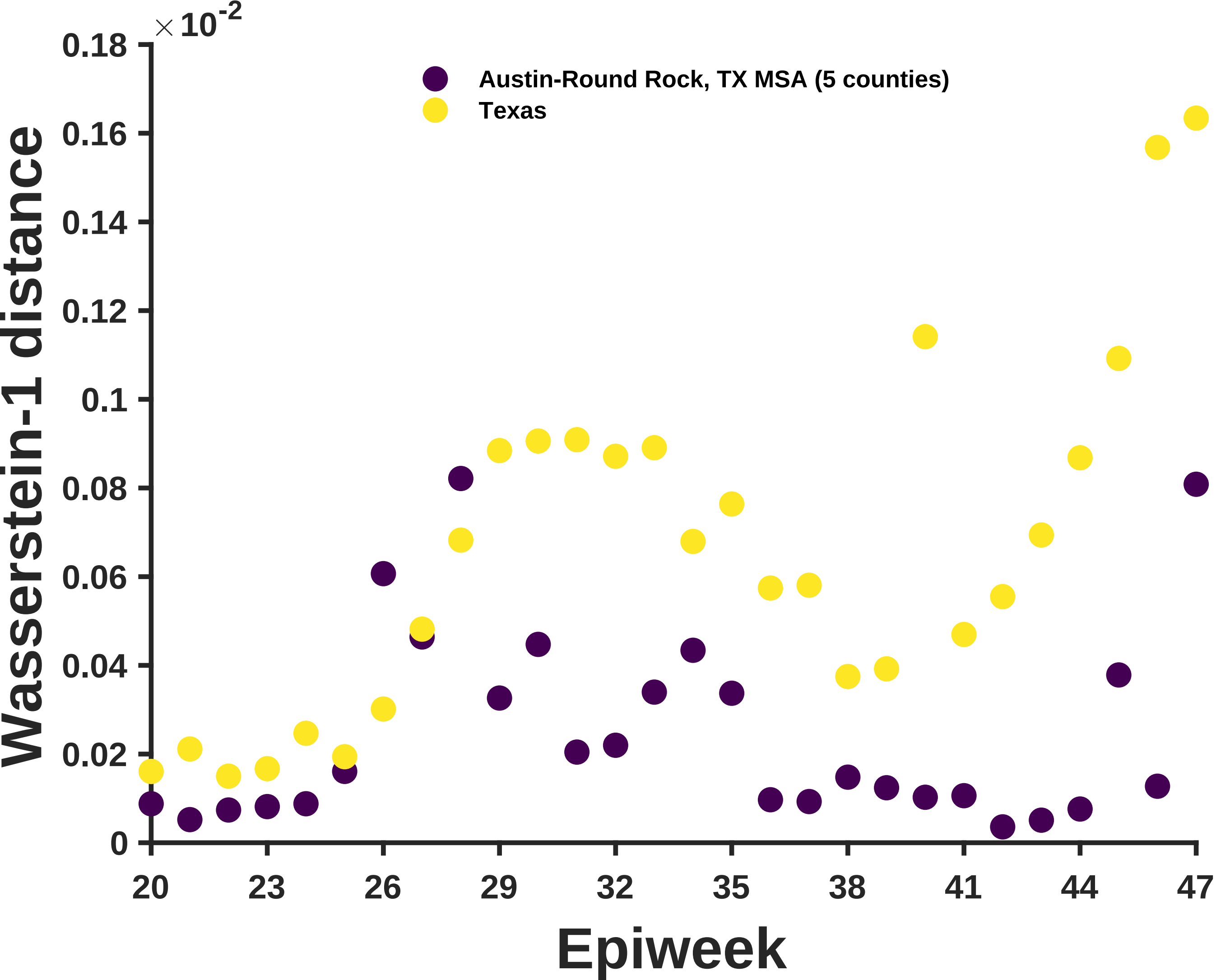

×10^-2
0.18
0.16
0.14
0.12
0.1
0.08
0.06
0.04
0.02
0
Wasserstein-1 distance
Austin-Round Rock, TX MSA (5 counties)
Texas
20
23
26
29
32
35
38
41
44
47
Epiweek

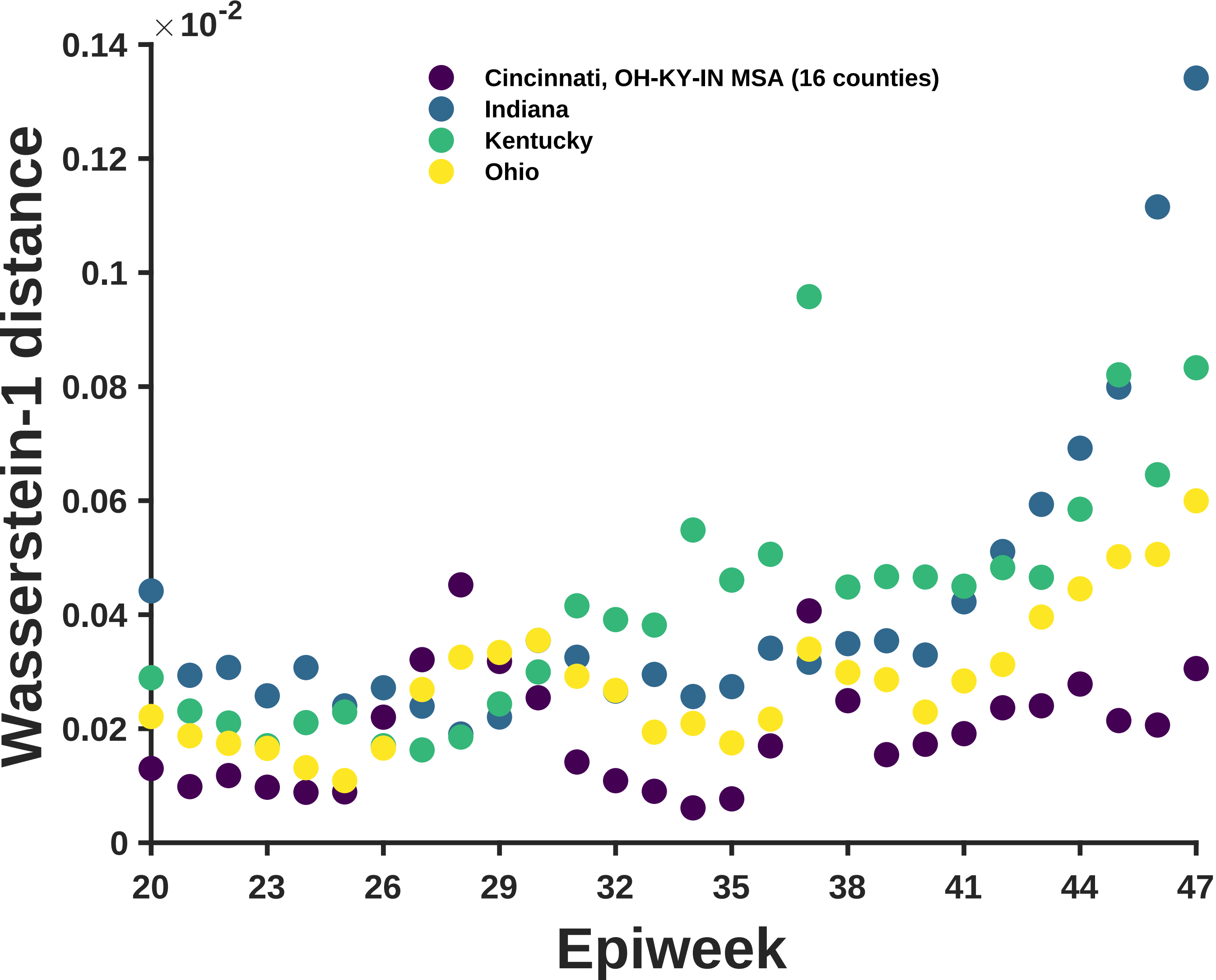

×10^-2
0.14
0.12
0.1
0.08
0.06
0.04
0.02
0
Wasserstein-1 distance
Cincinnati, OH-KY-IN MSA (16 counties)
Indiana
Kentucky
Ohio
20
23
26
29
32
35
38
41
44
47
Epiweek

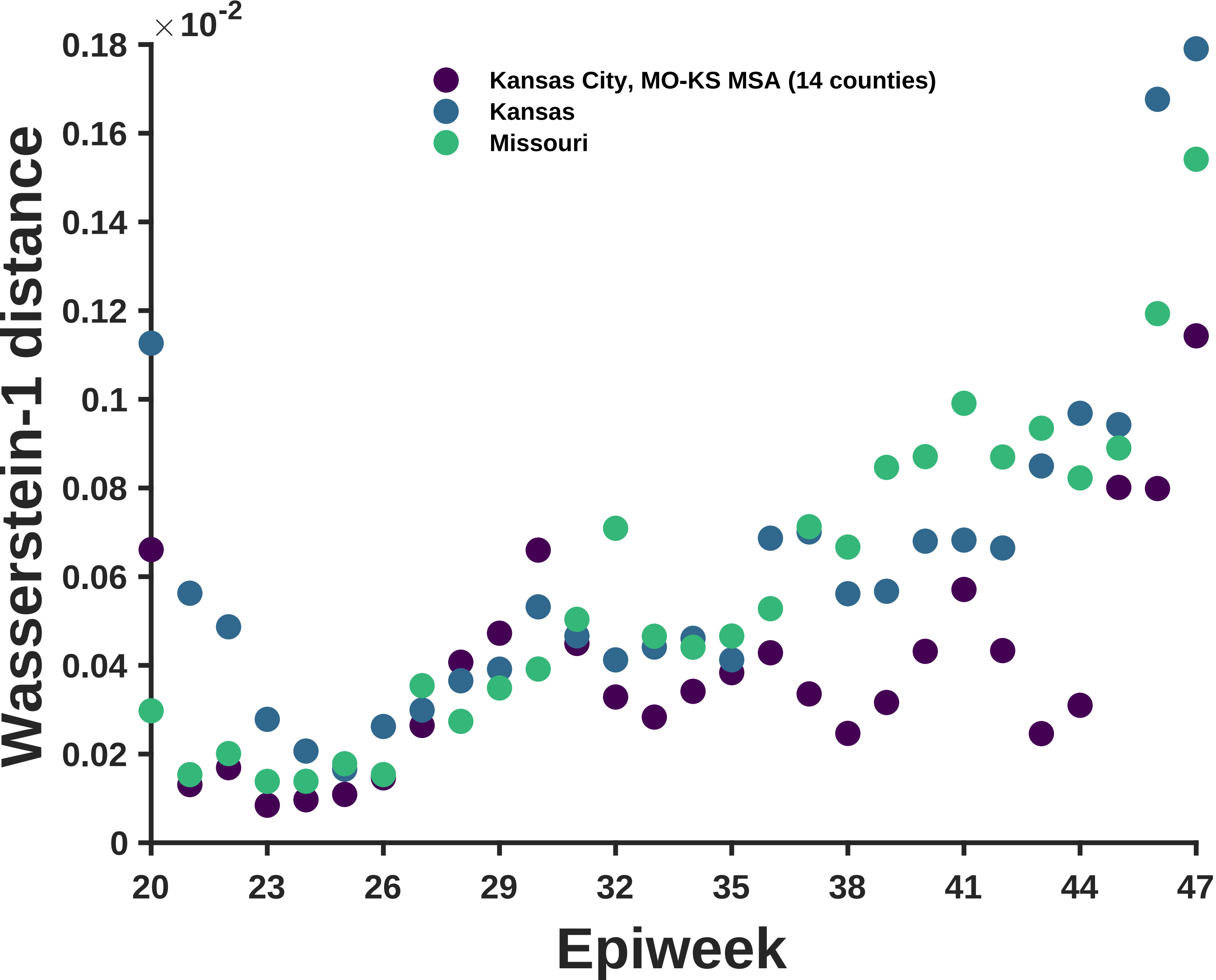
×10^-2
0.18
0.16
0.14
0.12
0.1
0.08
0.06
0.04
0.02
0
Wasserstein-1 distance
Kansas City, MO-KS MSA (14 counties)
Kansas
Missouri
20
23
26
29
32
35
38
41
44
47
Epiweek

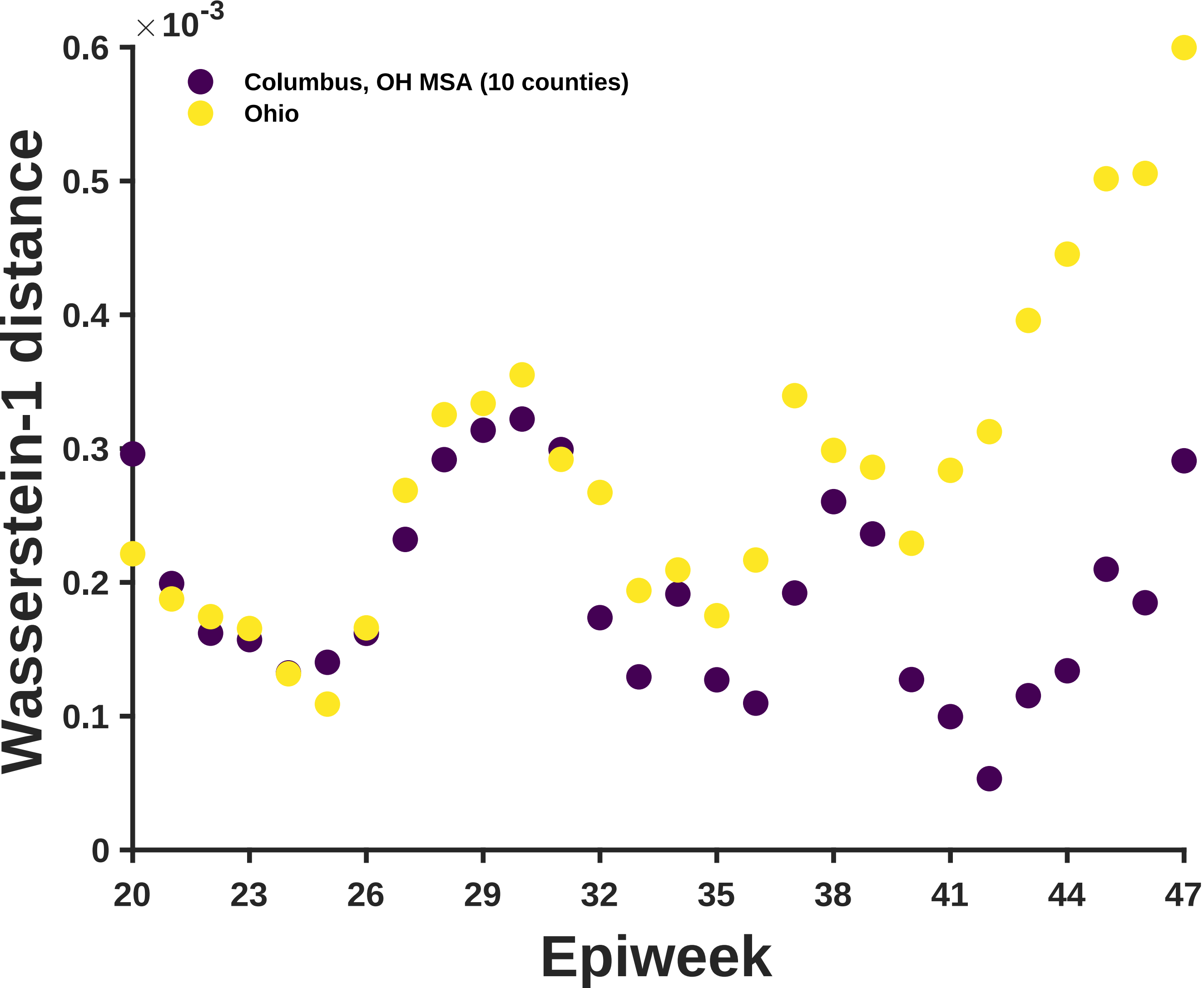

×10^-3
0.6
0.5
0.4
0.3
0.2
0.1
0
Columbus, OH MSA (10 counties)
Ohio
Wasserstein-1 distance
20
23
26
29
32
35
38
41
44
47
Epiweek

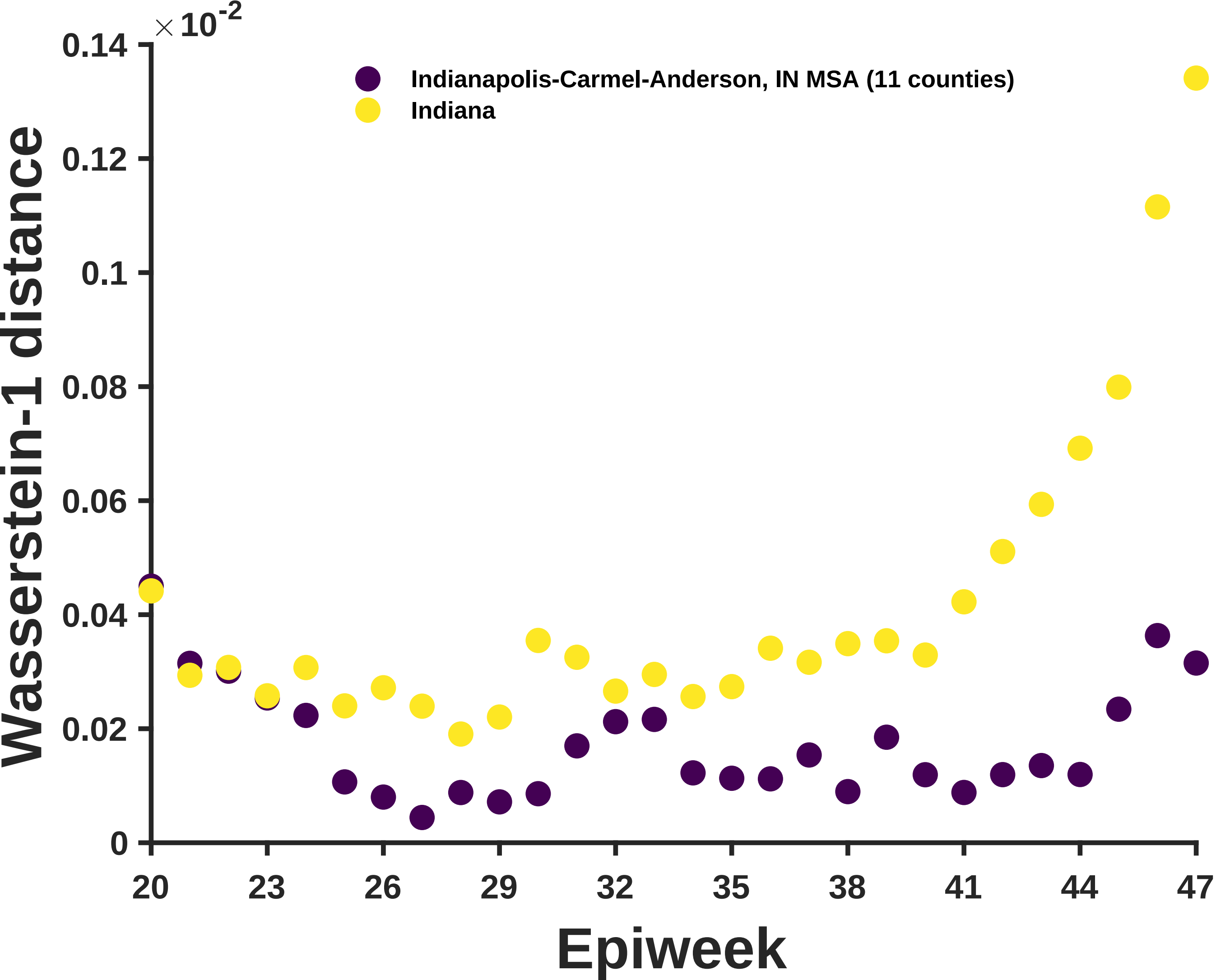

×10^-2
0.14
0.12
0.1
0.08
0.06
0.04
0.02
0
Wasserstein-1 distance
Indianapolis-Carmel-Anderson, IN MSA (11 counties)
Indiana
20
23
26
29
32
35
38
41
44
47
Epiweek

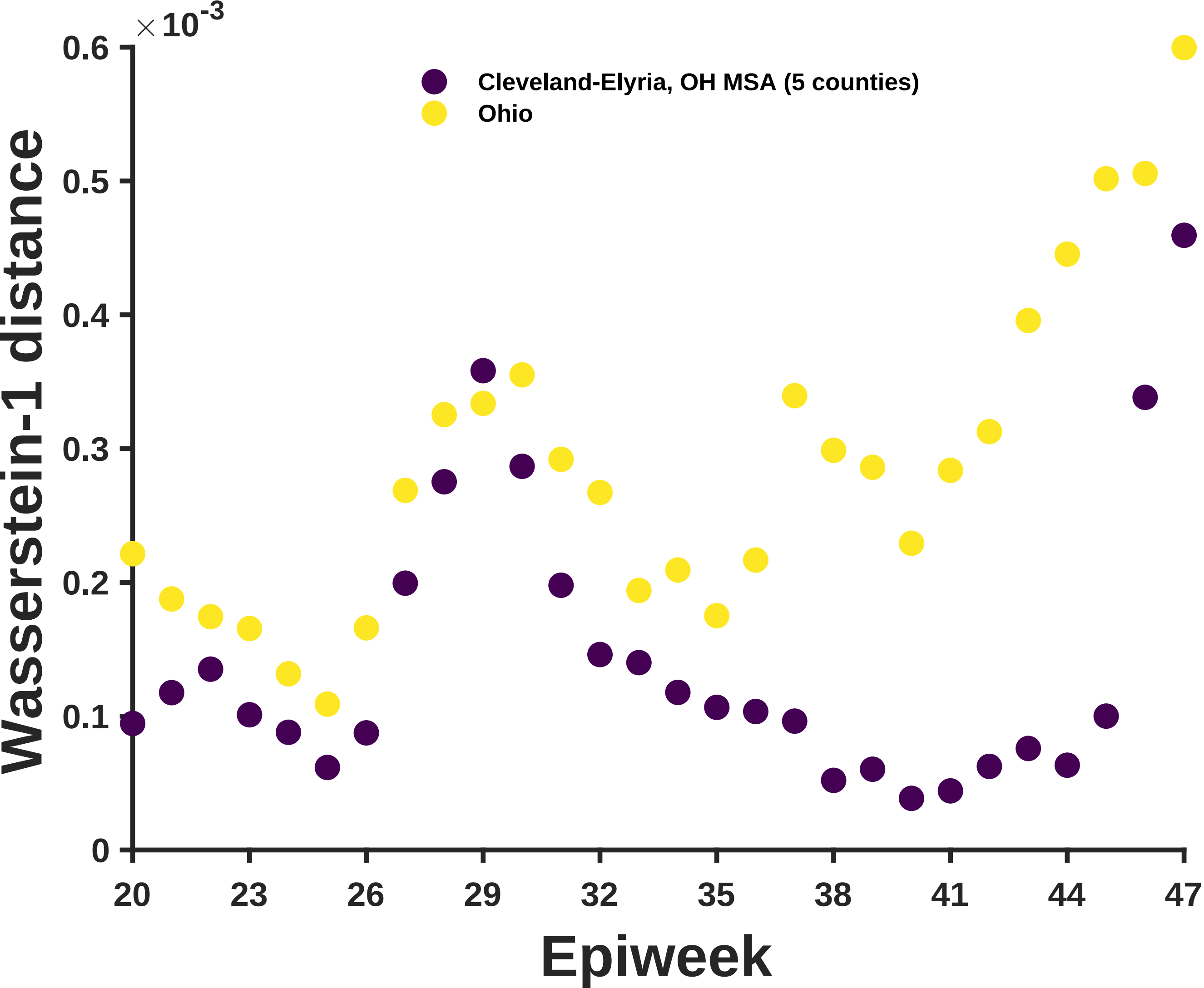

×10^-3
0.6
0.5
0.4
0.3
0.2
0.1
0
Cleveland-Elyria, OH MSA (5 counties)
Ohio
Wasserstein-1 distance
20
23
26
29
32
35
38
41
44
47
Epiweek

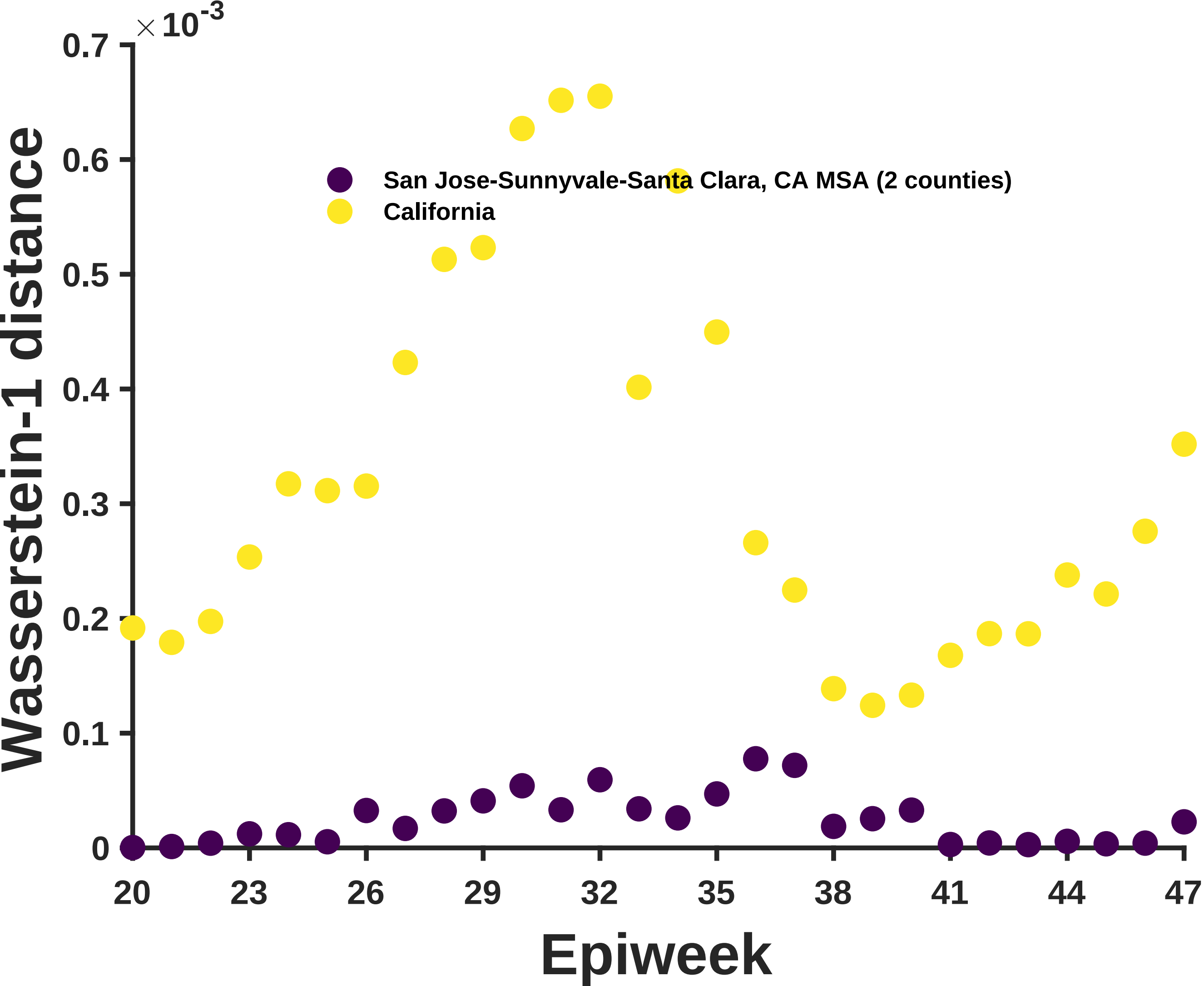
×10^-3
0.7
0.6
0.5
0.4
0.3
0.2
0.1
0
Wasserstein-1 distance
San Jose-Sunnyvale-Santa Clara, CA MSA (2 counties)
California
20
23
26
29
32
35
38
41
44
47
Epiweek

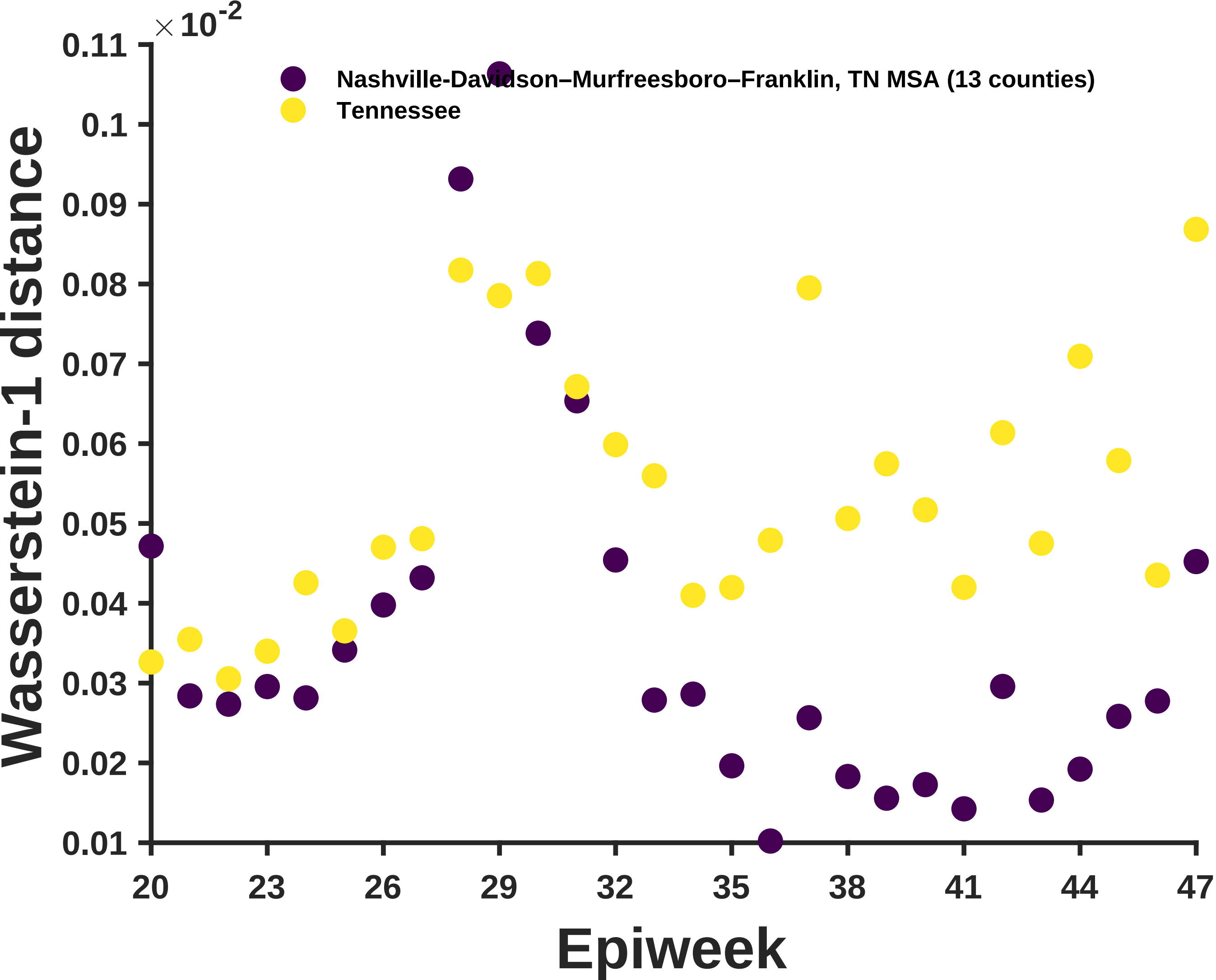

$\times 10^{-2}$
0.11
0.1
0.09
0.08
0.07
0.06
0.05
0.04
0.03
0.02
0.01
Wasserstein-1 distance
Nashville-Davidson–Murfreesboro–Franklin, TN MSA (13 counties)
Tennessee
20
23
26
29
32
35
38
41
44
47
Epiweek

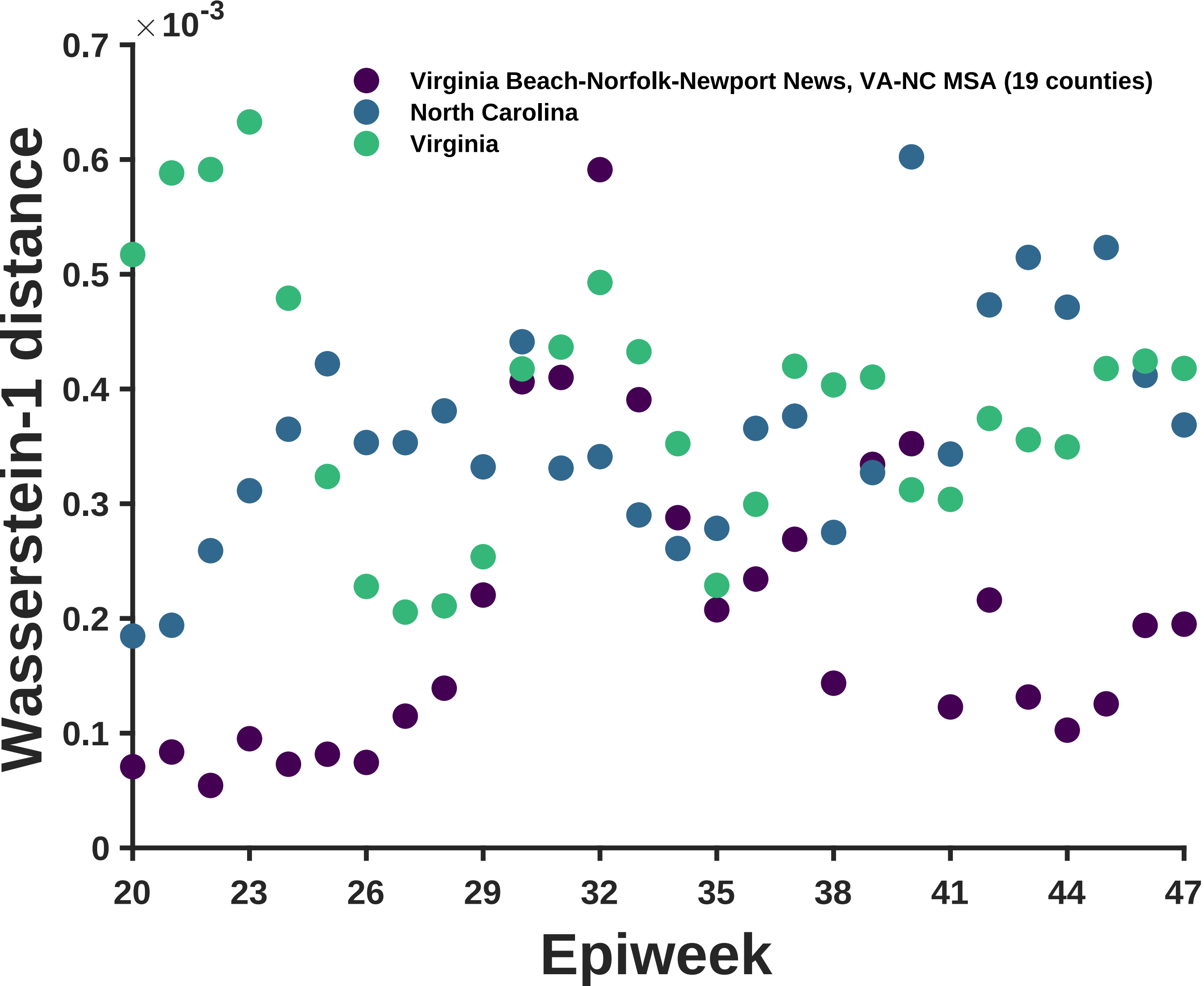

×10^-3
0.7
0.6
0.5
0.4
0.3
0.2
0.1
0
Wasserstein-1 distance
Virginia Beach-Norfolk-Newport News, VA-NC MSA (19 counties)
North Carolina
Virginia
20
23
26
29
32
35
38
41
44
47
Epiweek

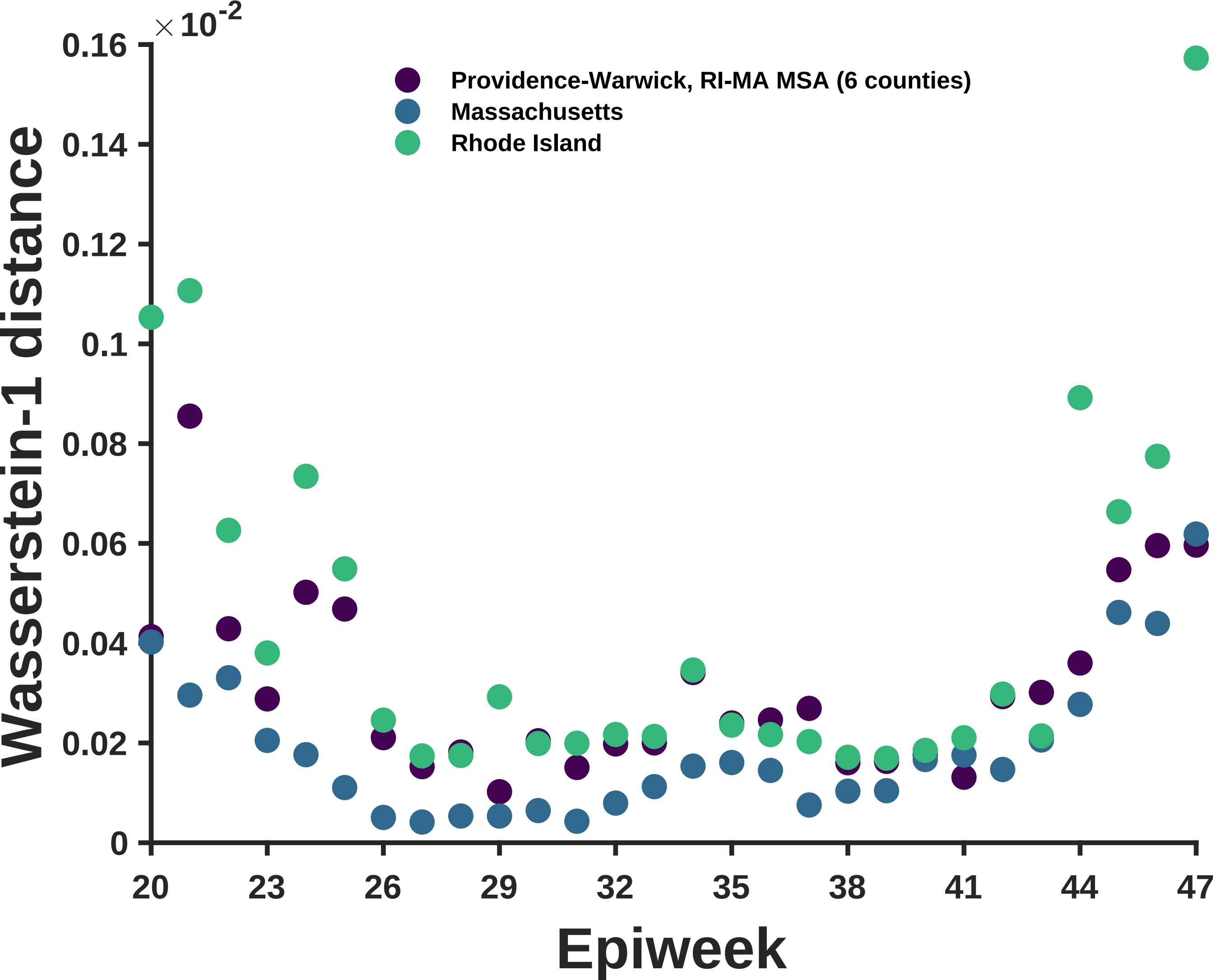

×10^-2
0.16
0.14
0.12
0.1
0.08
0.06
0.04
0.02
0
Wasserstein-1 distance
Providence-Warwick, RI-MA MSA (6 counties)
Massachusetts
Rhode Island
20
23
26
29
32
35
38
41
44
47
Epiweek

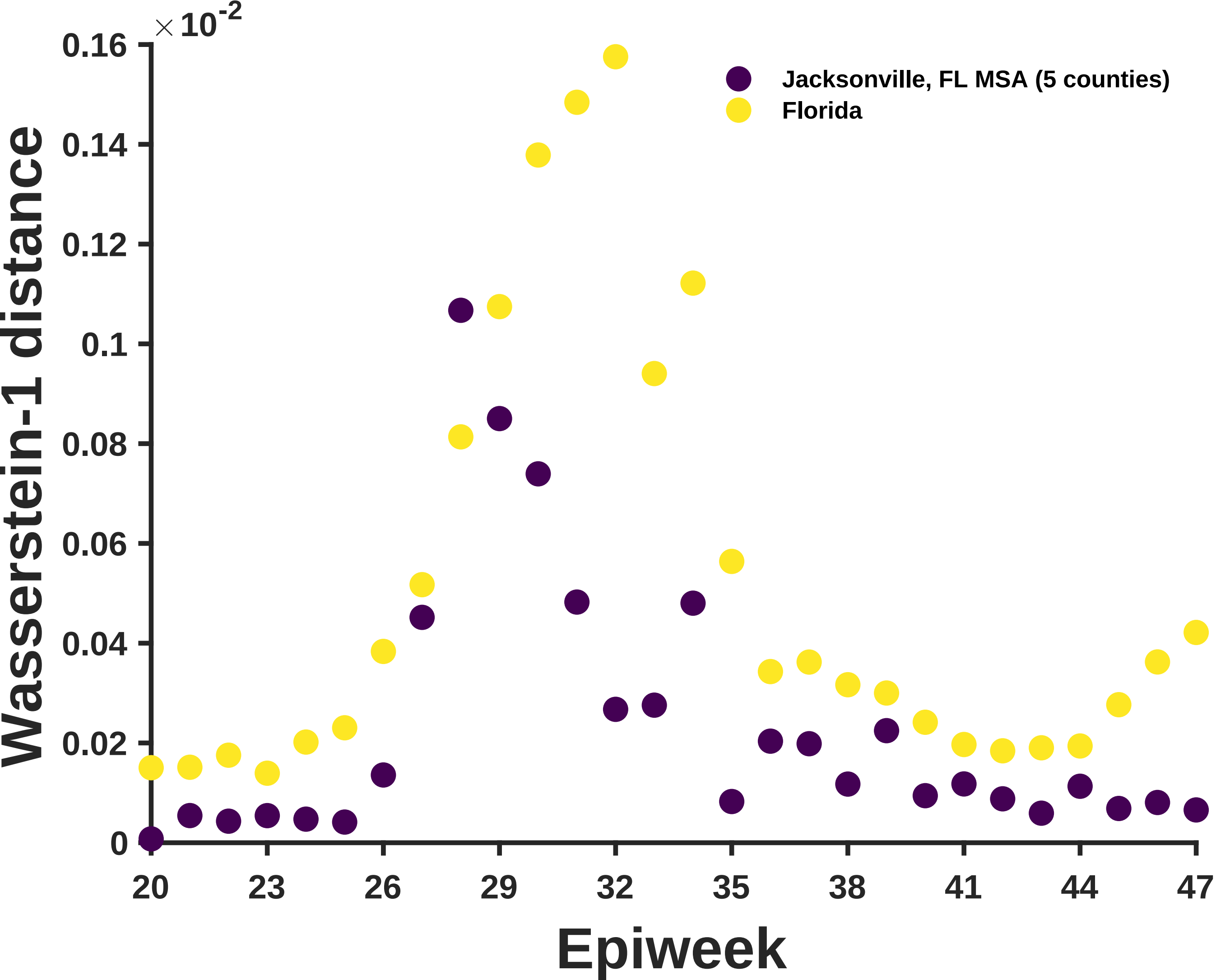
×10^-2
0.16
0.14
0.12
0.1
0.08
0.06
0.04
0.02
0
Wasserstein-1 distance
20
23
26
29
32
35
38
41
44
47
Epiweek
Jacksonville, FL MSA (5 counties)
Florida

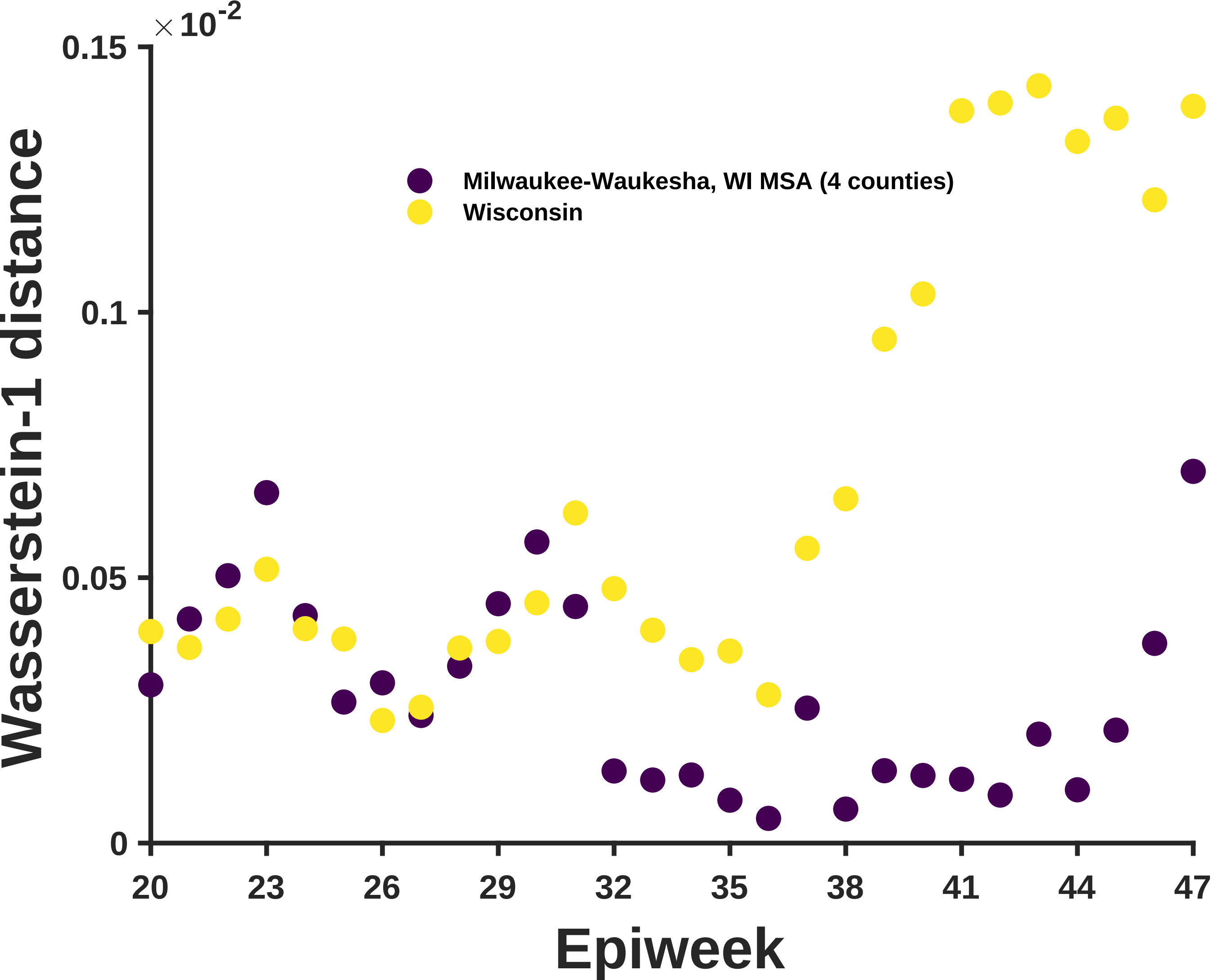

×10^-2
0.15
0.1
0.05
0
Wasserstein-1 distance
Milwaukee-Waukesha, WI MSA (4 counties)
Wisconsin
20
23
26
29
32
35
38
41
44
47
Epiweek

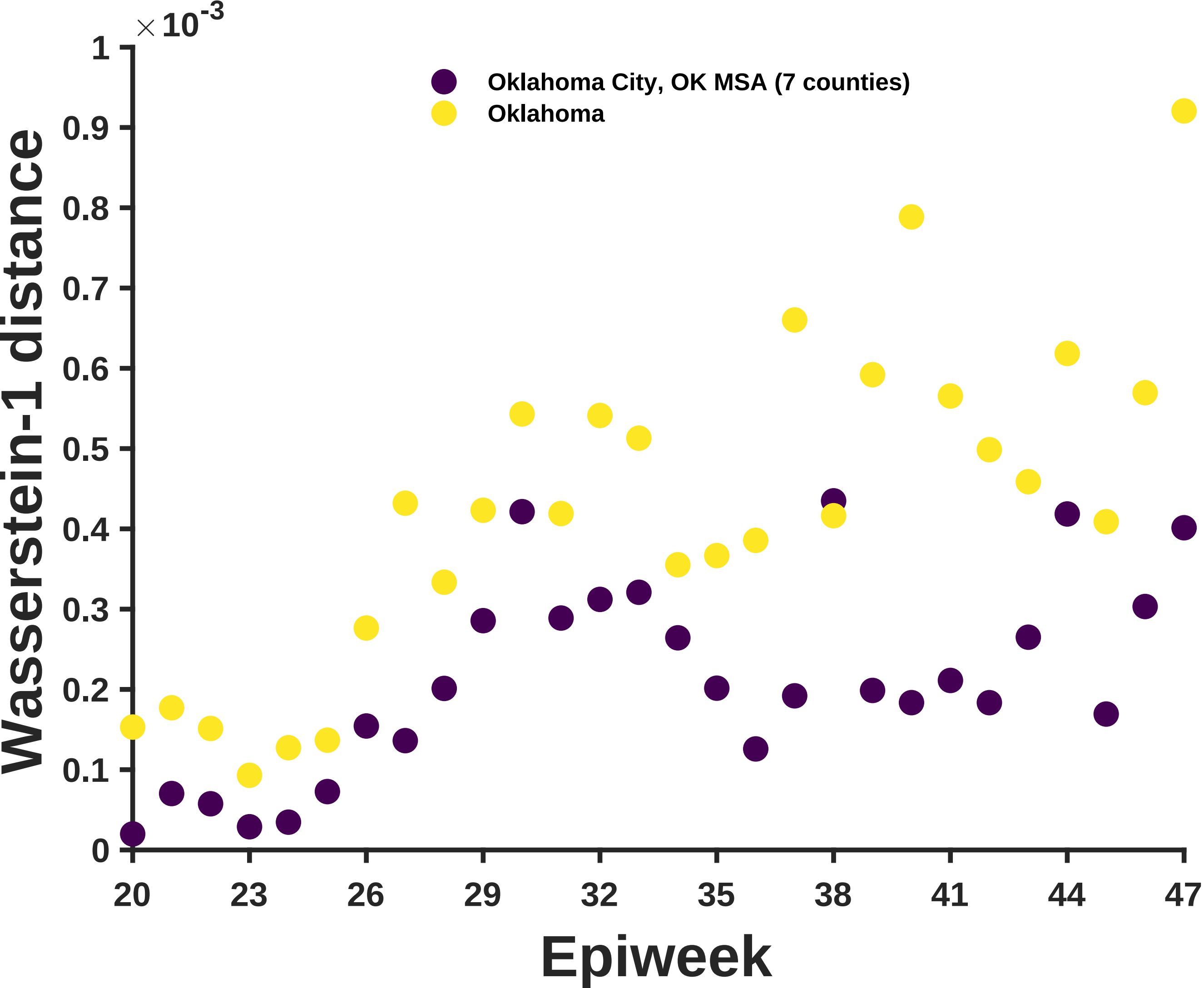

× 10^-3
1
0.9
0.8
0.7
0.6
0.5
0.4
0.3
0.2
0.1
0
Wasserstein-1 distance
Oklahoma City, OK MSA (7 counties)
Oklahoma
20
23
26
29
32
35
38
41
44
47
Epiweek

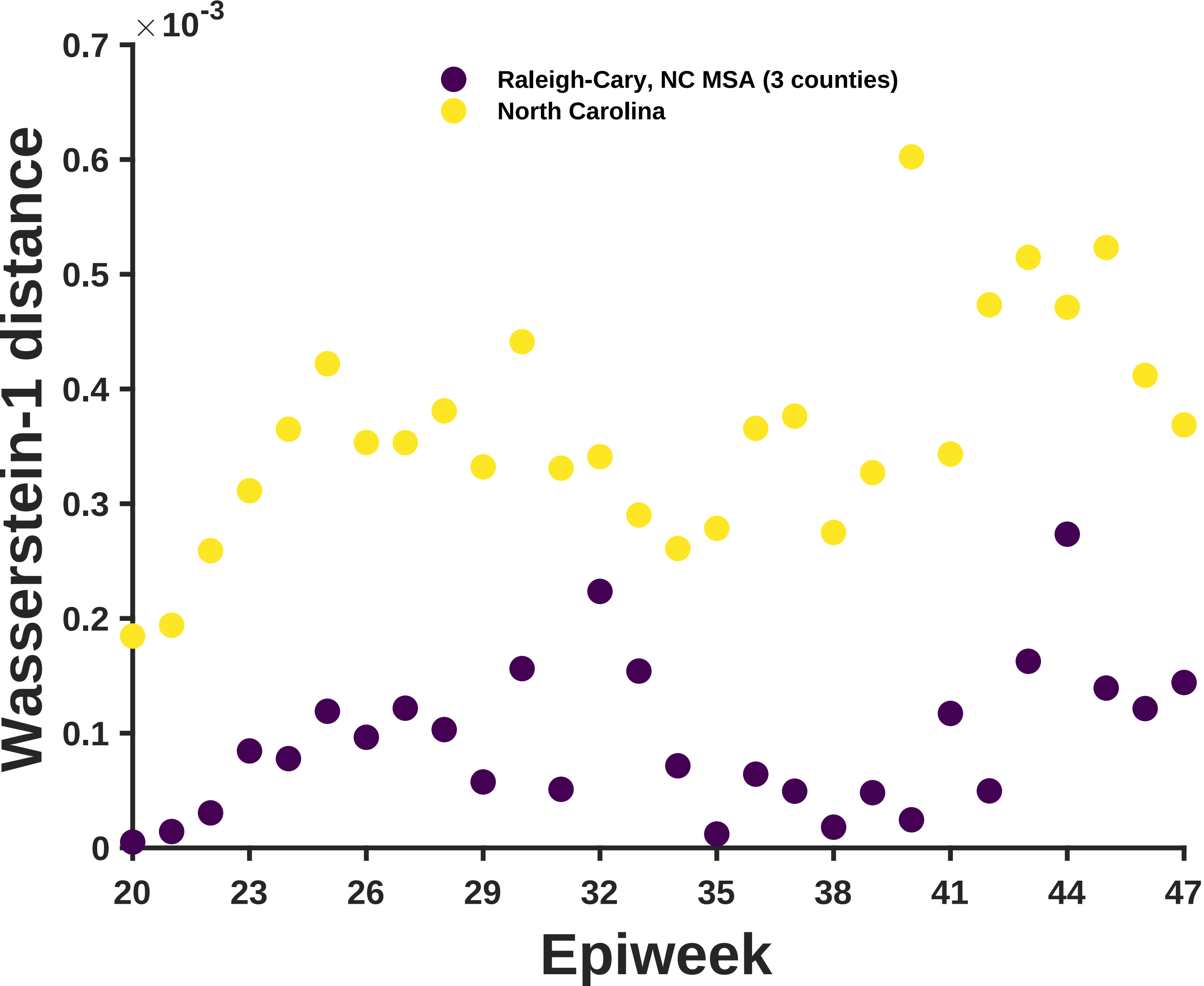

×10^-3
0.7
0.6
0.5
0.4
0.3
0.2
0.1
0
Wasserstein-1 distance
Raleigh-Cary, NC MSA (3 counties)
North Carolina
20
23
26
29
32
35
38
41
44
47
Epiweek

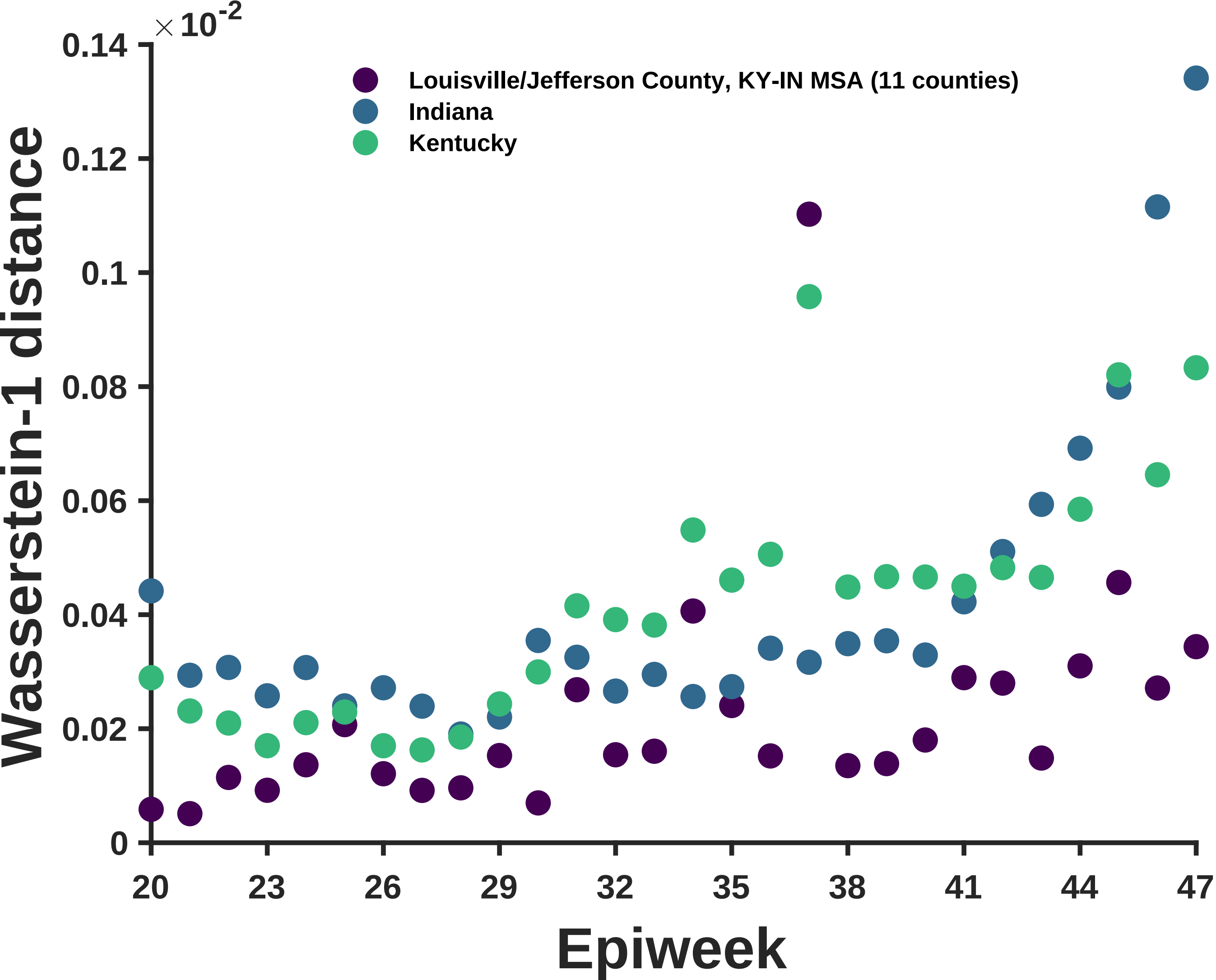

×10^-2
0.14
0.12
0.1
0.08
0.06
0.04
0.02
0
Wasserstein-1 distance
Louisville/Jefferson County, KY-IN MSA (11 counties)
Indiana
Kentucky
20
23
26
29
32
35
38
41
44
47
Epiweek

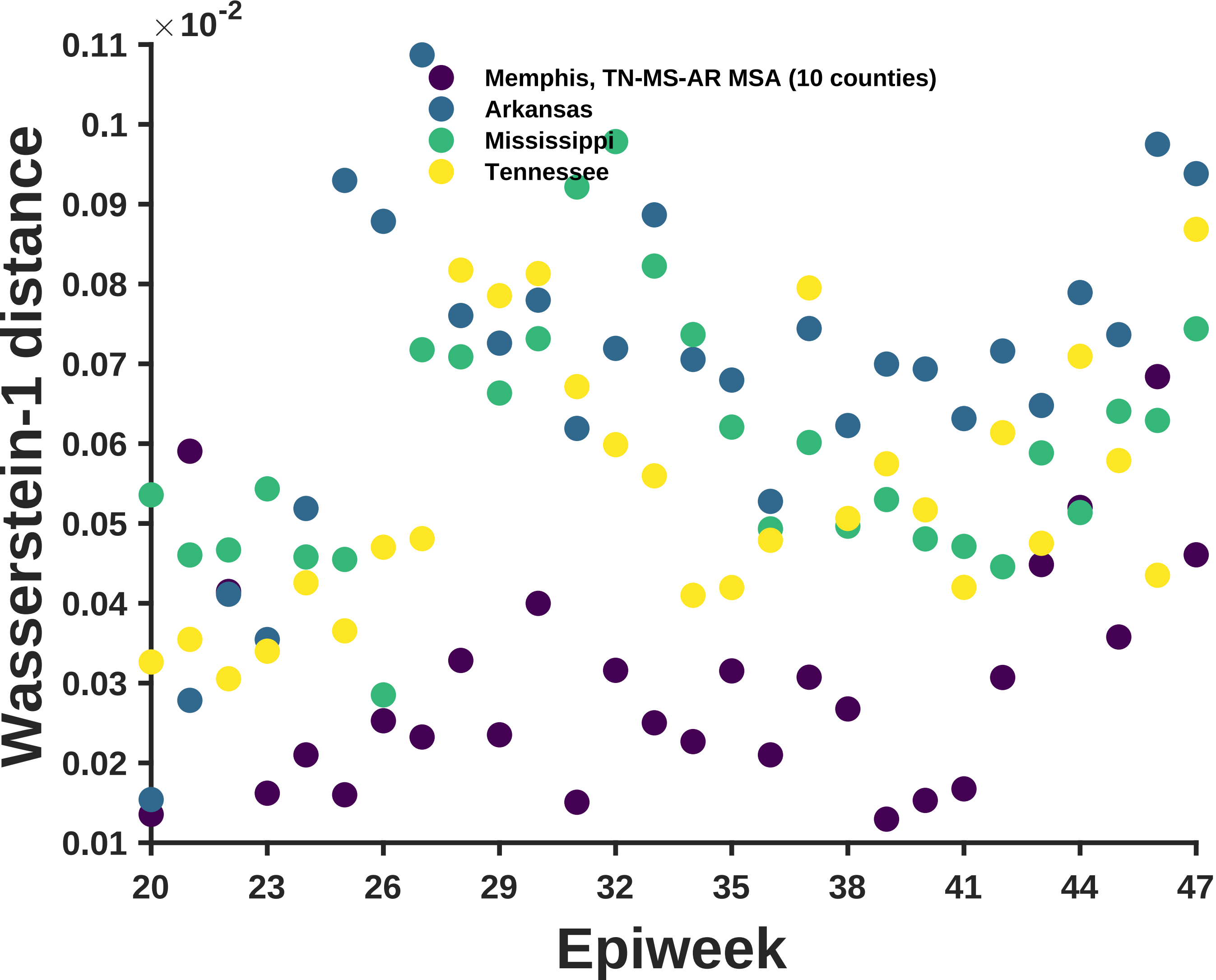

×10^-2
0.11
0.1
0.09
0.08
0.07
0.06
0.05
0.04
0.03
0.02
0.01
Wasserstein-1 distance
Memphis, TN-MS-AR MSA (10 counties)
Arkansas
Mississippi
Tennessee
20
23
26
29
32
35
38
41
44
47
Epiweek

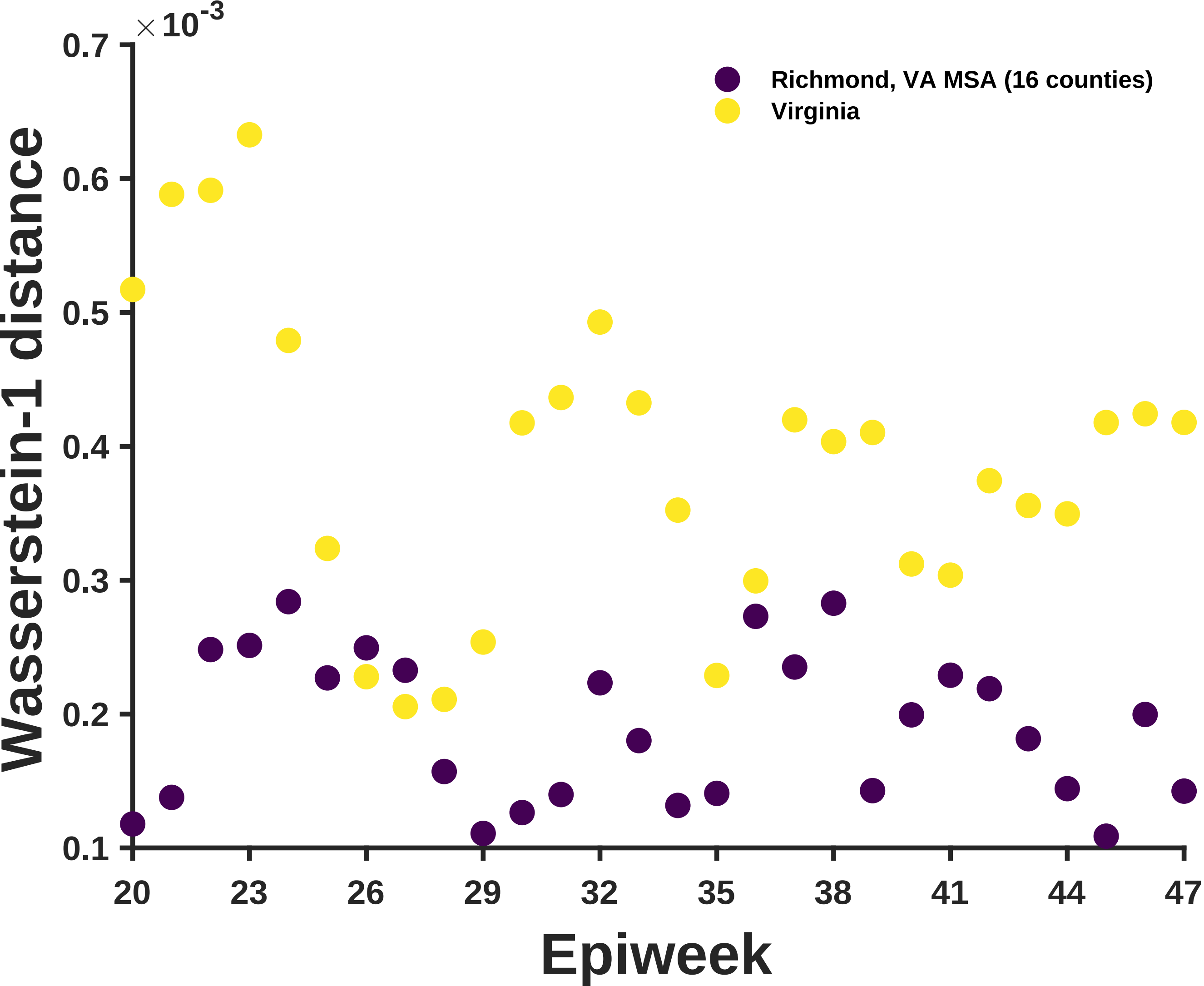

$\times 10^{-3}$
0.7
0.6
0.5
0.4
0.3
0.2
0.1
Wasserstein-1 distance
Richmond, VA MSA (16 counties)
Virginia
20
23
26
29
32
35
38
41
44
47
Epiweek

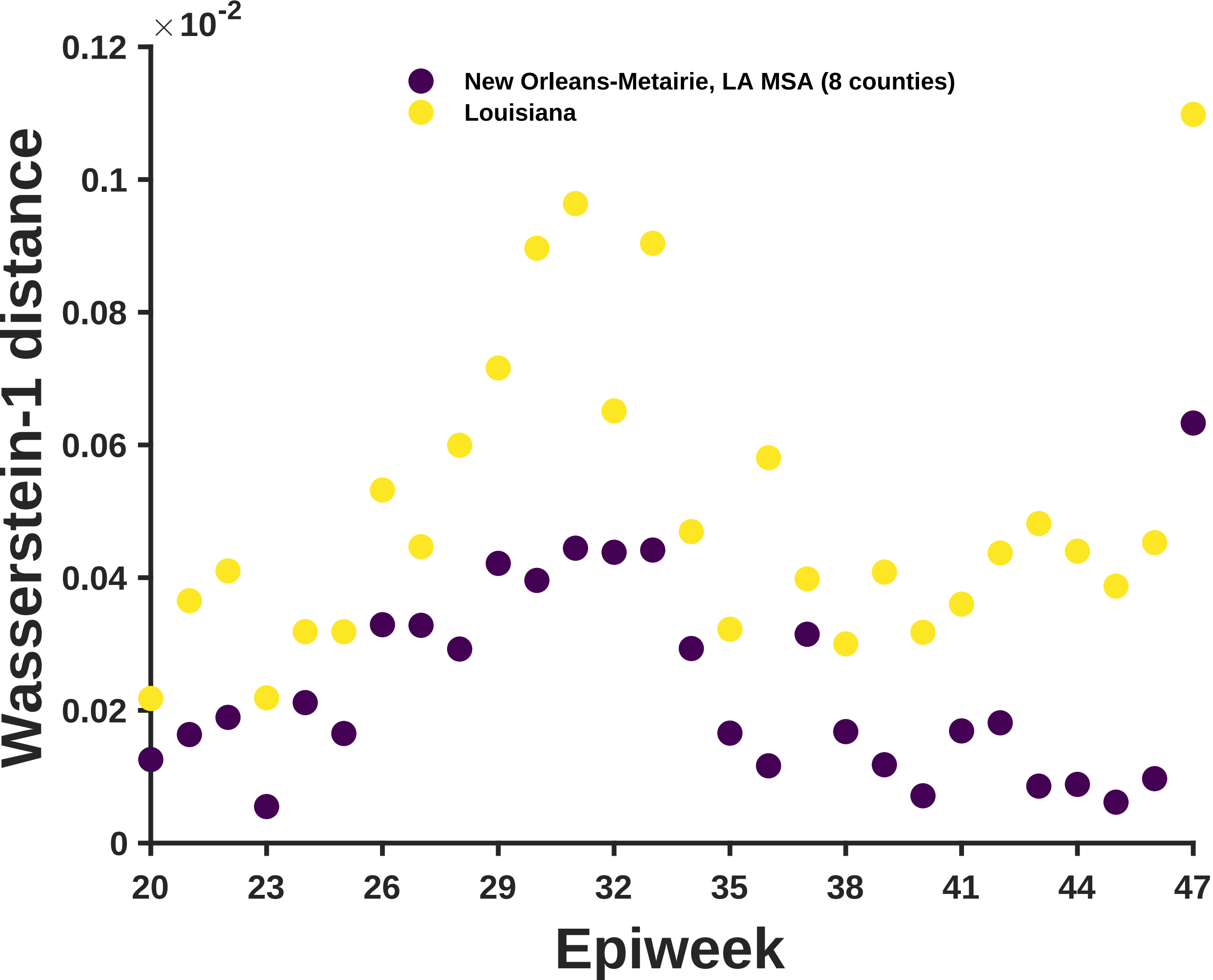

×10^-2
0.12
0.1
0.08
0.06
0.04
0.02
0
Wasserstein-1 distance
New Orleans-Metairie, LA MSA (8 counties)
Louisiana
20
23
26
29
32
35
38
41
44
47
Epiweek

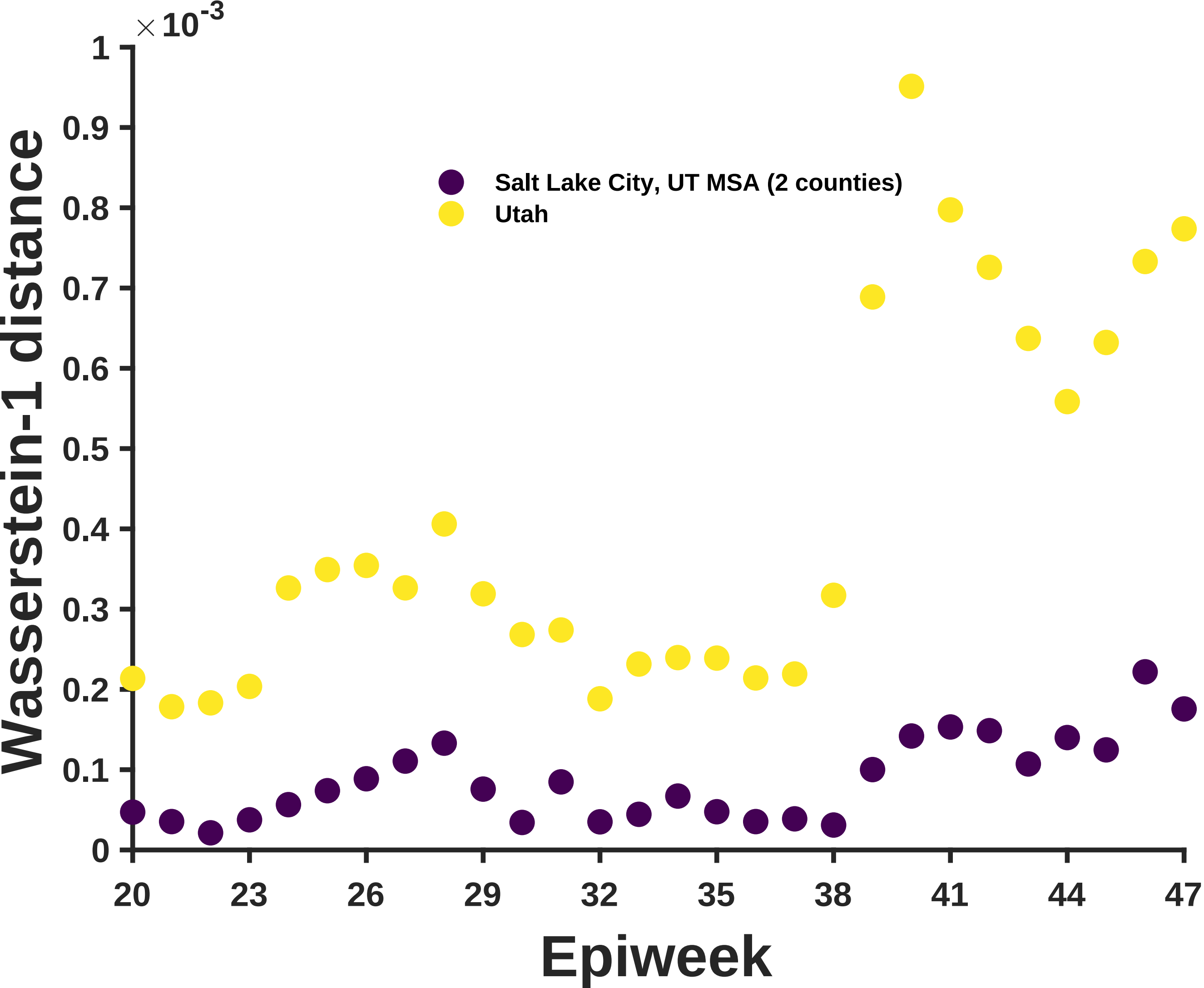

×10^-3
Wasserstein-1 distance
Epiweek
Salt Lake City, UT MSA (2 counties)
Utah
0
0.1
0.2
0.3
0.4
0.5
0.6
0.7
0.8
0.9
1
20
23
26
29
32
35
38
41
44
47

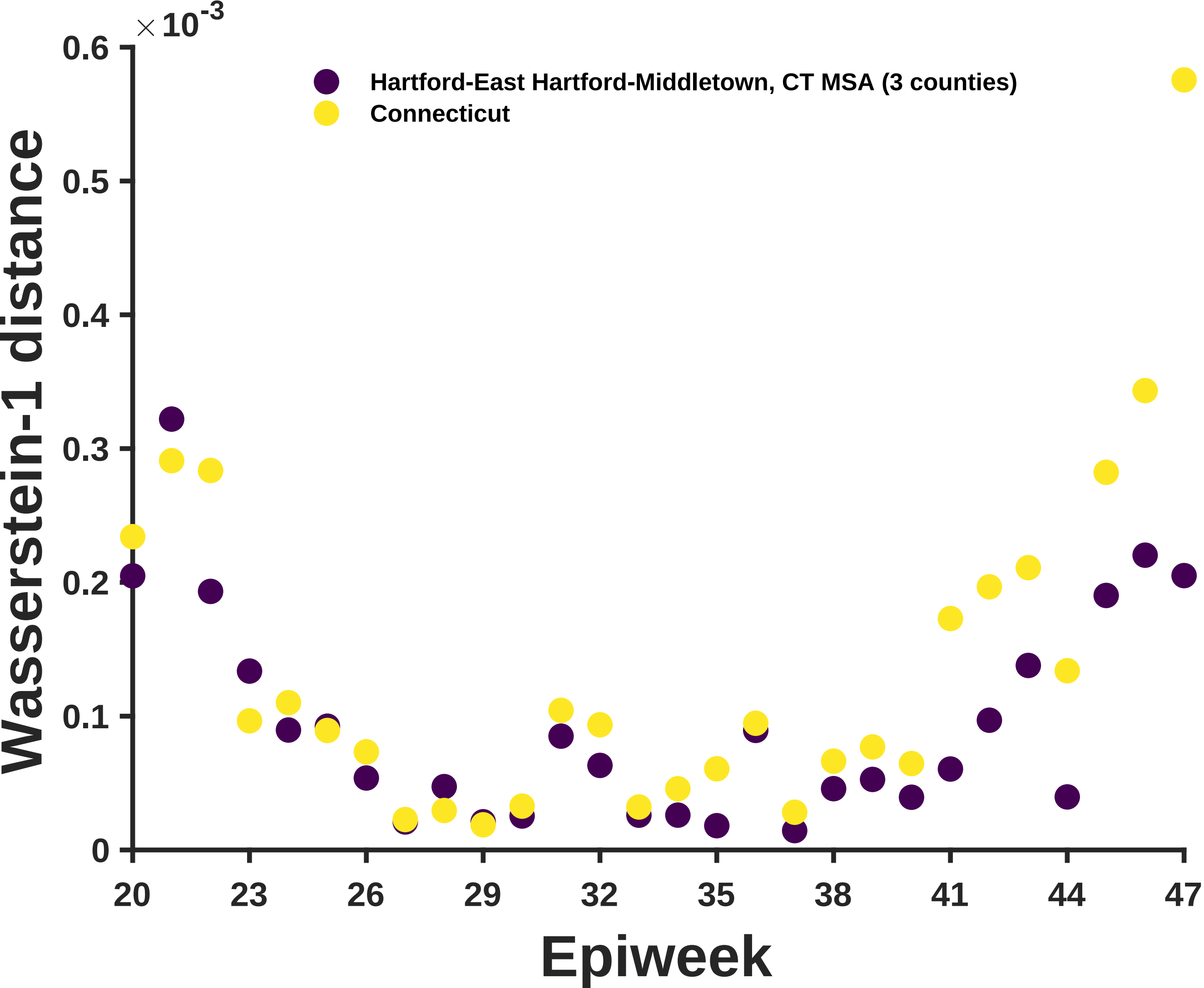

×10^-3
0.6
0.5
0.4
0.3
0.2
0.1
0
Wasserstein-1 distance
Hartford-East Hartford-Middletown, CT MSA (3 counties)
Connecticut
20
23
26
29
32
35
38
41
44
47
Epiweek

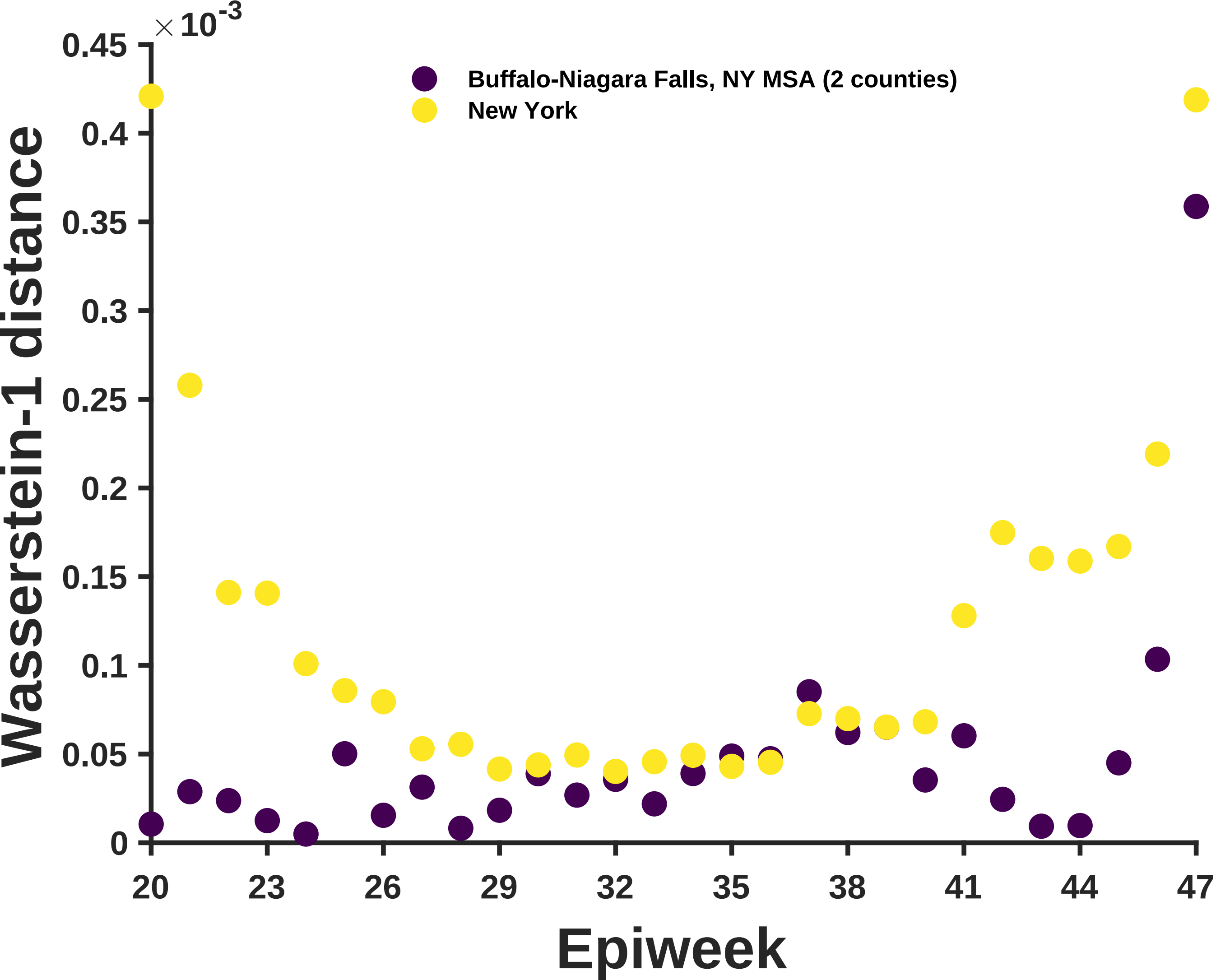

×10^-3
0.45
0.4
0.35
0.3
0.25
0.2
0.15
0.1
0.05
0
Wasserstein-1 distance
Buffalo-Niagara Falls, NY MSA (2 counties)
New York
20
23
26
29
32
35
38
41
44
47
Epiweek

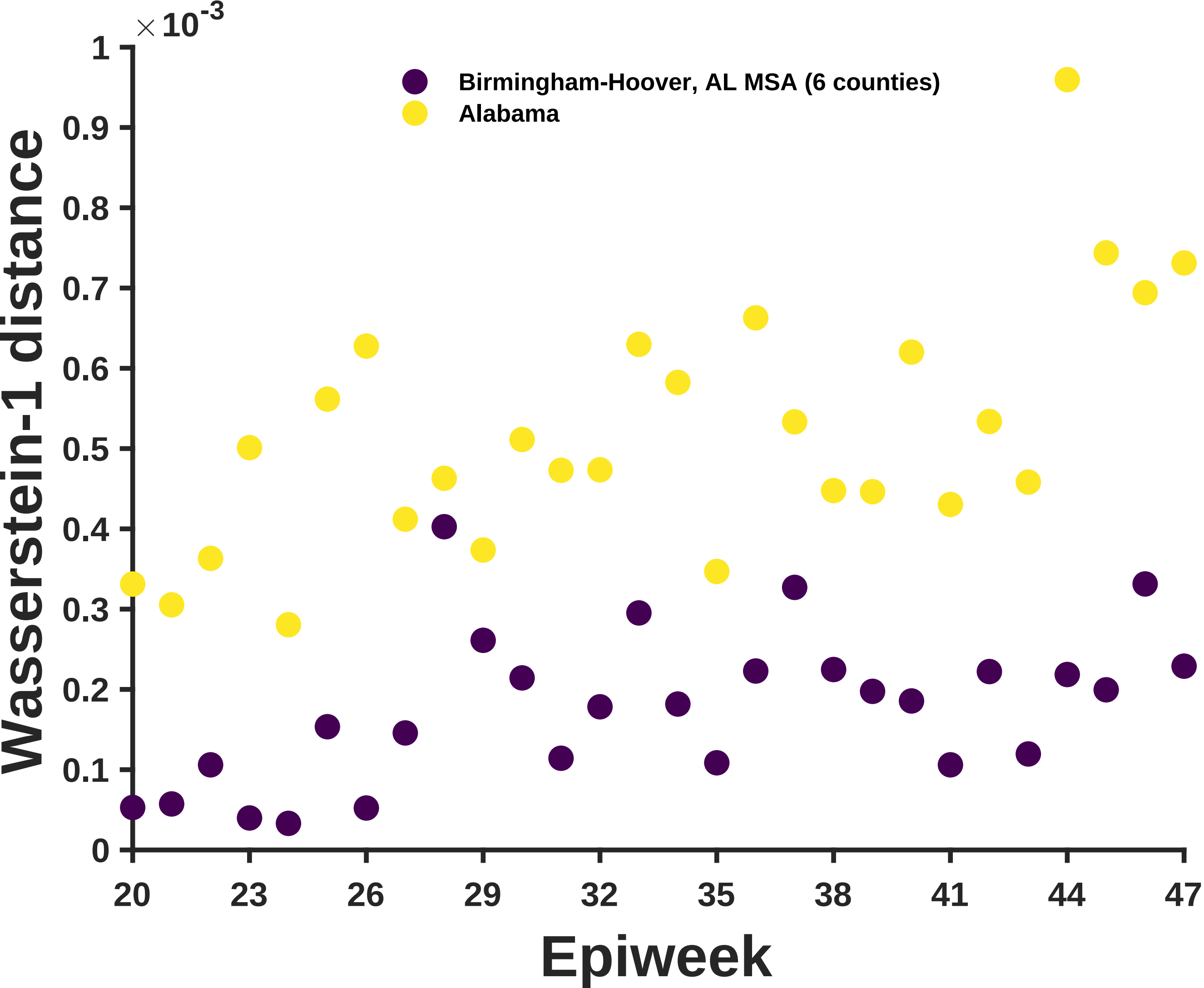

×10^-3
1
0.9
0.8
0.7
0.6
0.5
0.4
0.3
0.2
0.1
0
Wasserstein-1 distance
Birmingham-Hoover, AL MSA (6 counties)
Alabama
20
23
26
29
32
35
38
41
44
47
Epiweek

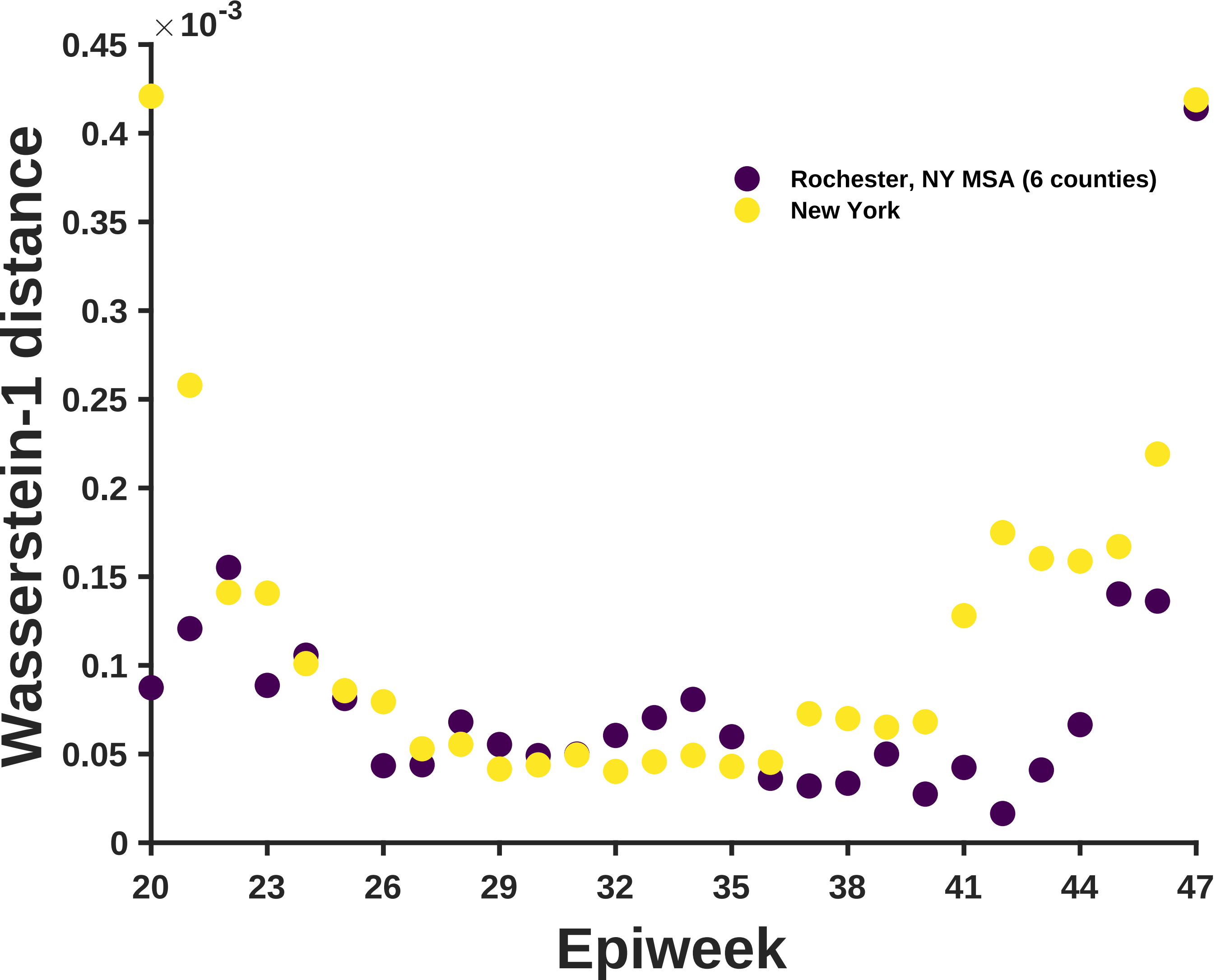
×10^-3
0.45
0.4
0.35
0.3
0.25
0.2
0.15
0.1
0.05
0
Wasserstein-1 distance
20
23
26
29
32
35
38
41
44
47
Epiweek
Rochester, NY MSA (6 counties)
New York

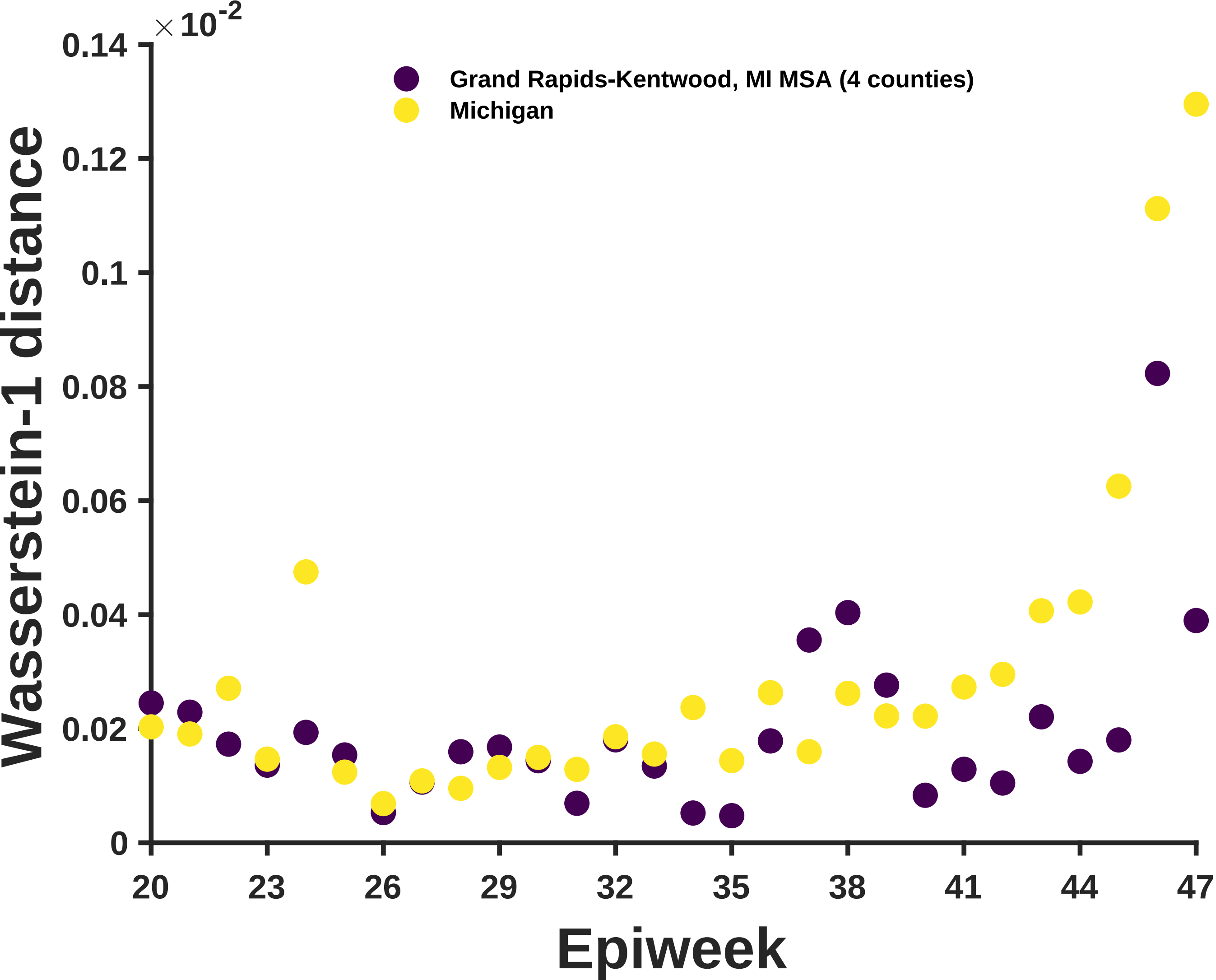

×10^-2
0.14
0.12
0.1
0.08
0.06
0.04
0.02
0
Wasserstein-1 distance
Grand Rapids-Kentwood, MI MSA (4 counties)
Michigan
20
23
26
29
32
35
38
41
44
47
Epiweek

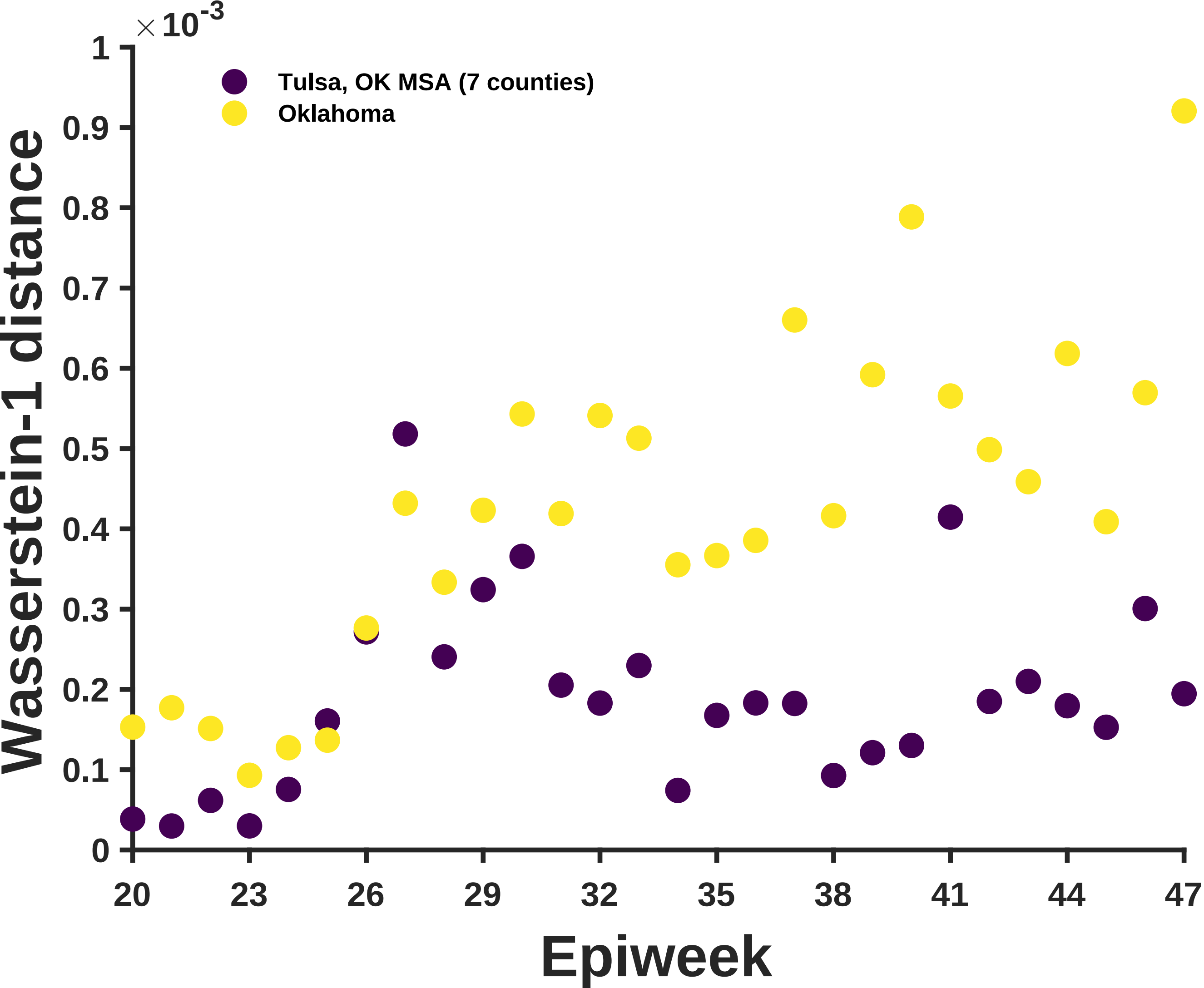

×10^-3
Tulsa, OK MSA (7 counties)
Oklahoma
Wasserstein-1 distance
0
0.1
0.2
0.3
0.4
0.5
0.6
0.7
0.8
0.9
1
20
23
26
29
32
35
38
41
44
47
Epiweek

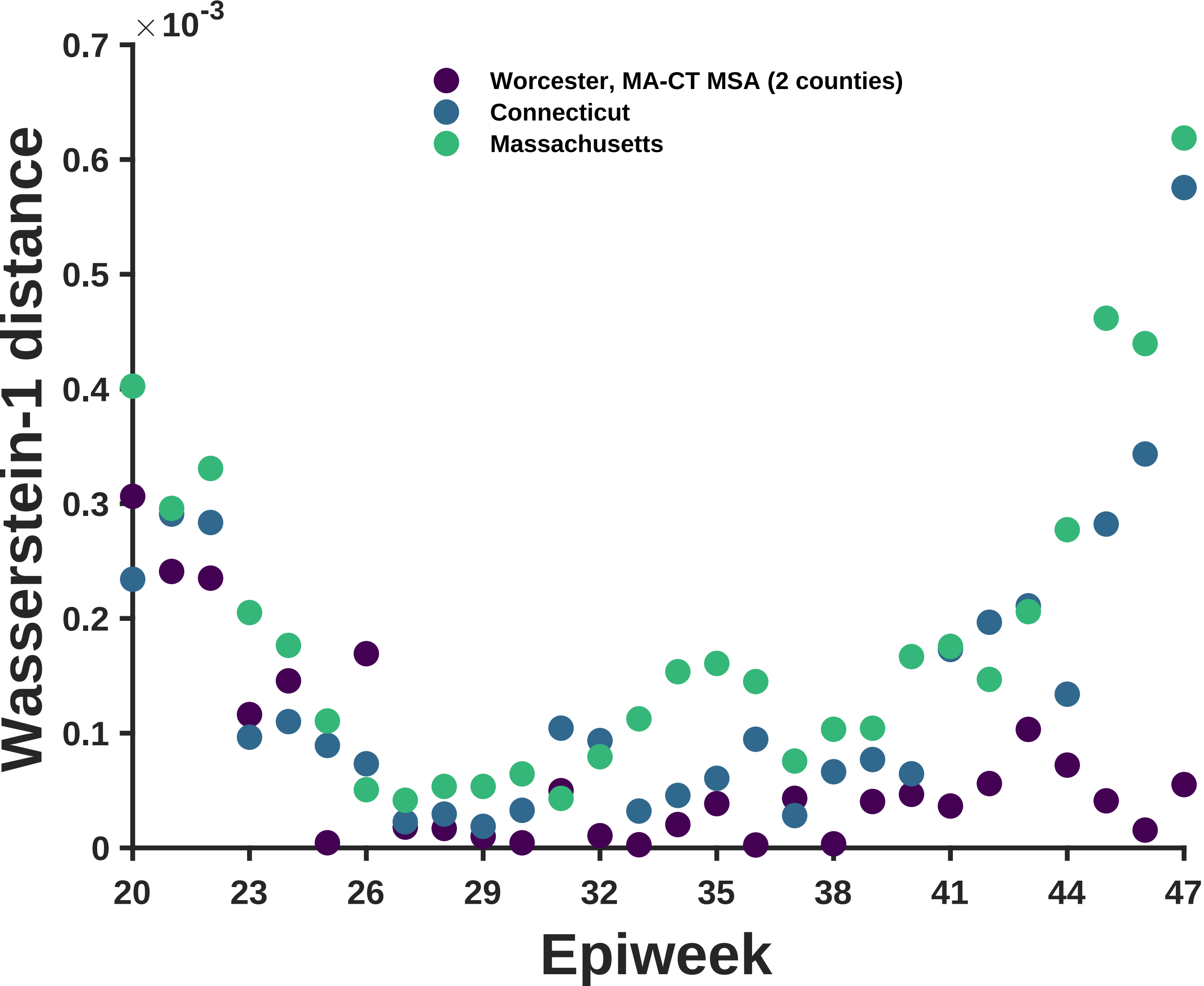

× 10^-3
0.7
0.6
0.5
0.4
0.3
0.2
0.1
0
Wasserstein-1 distance
Worcester, MA-CT MSA (2 counties)
Connecticut
Massachusetts
20
23
26
29
32
35
38
41
44
47
Epiweek

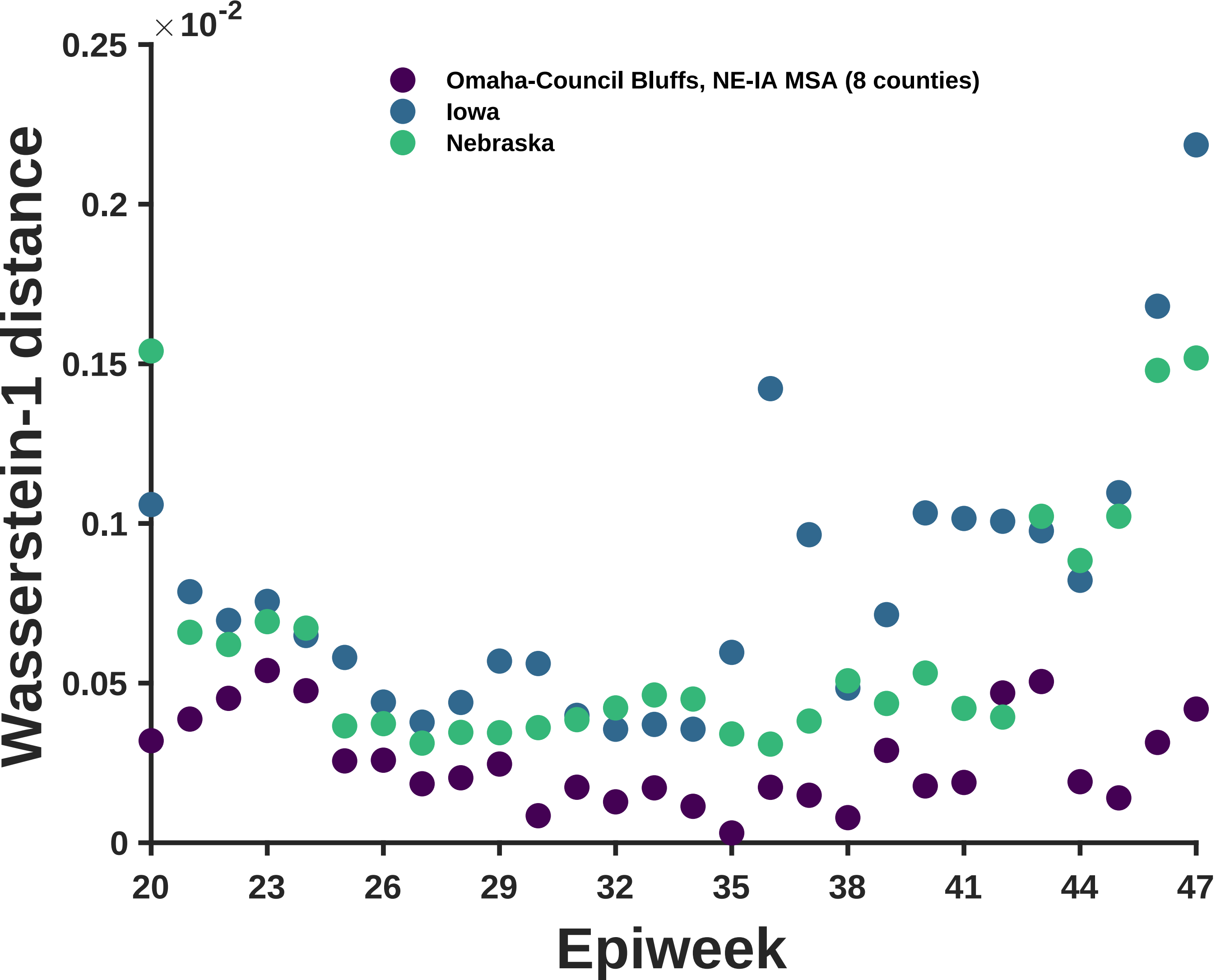

×10-2
0.25
0.2
0.15
0.1
0.05
0
Wasserstein-1 distance
Omaha-Council Bluffs, NE-IA MSA (8 counties)
Iowa
Nebraska
20
23
26
29
32
35
38
41
44
47
Epiweek

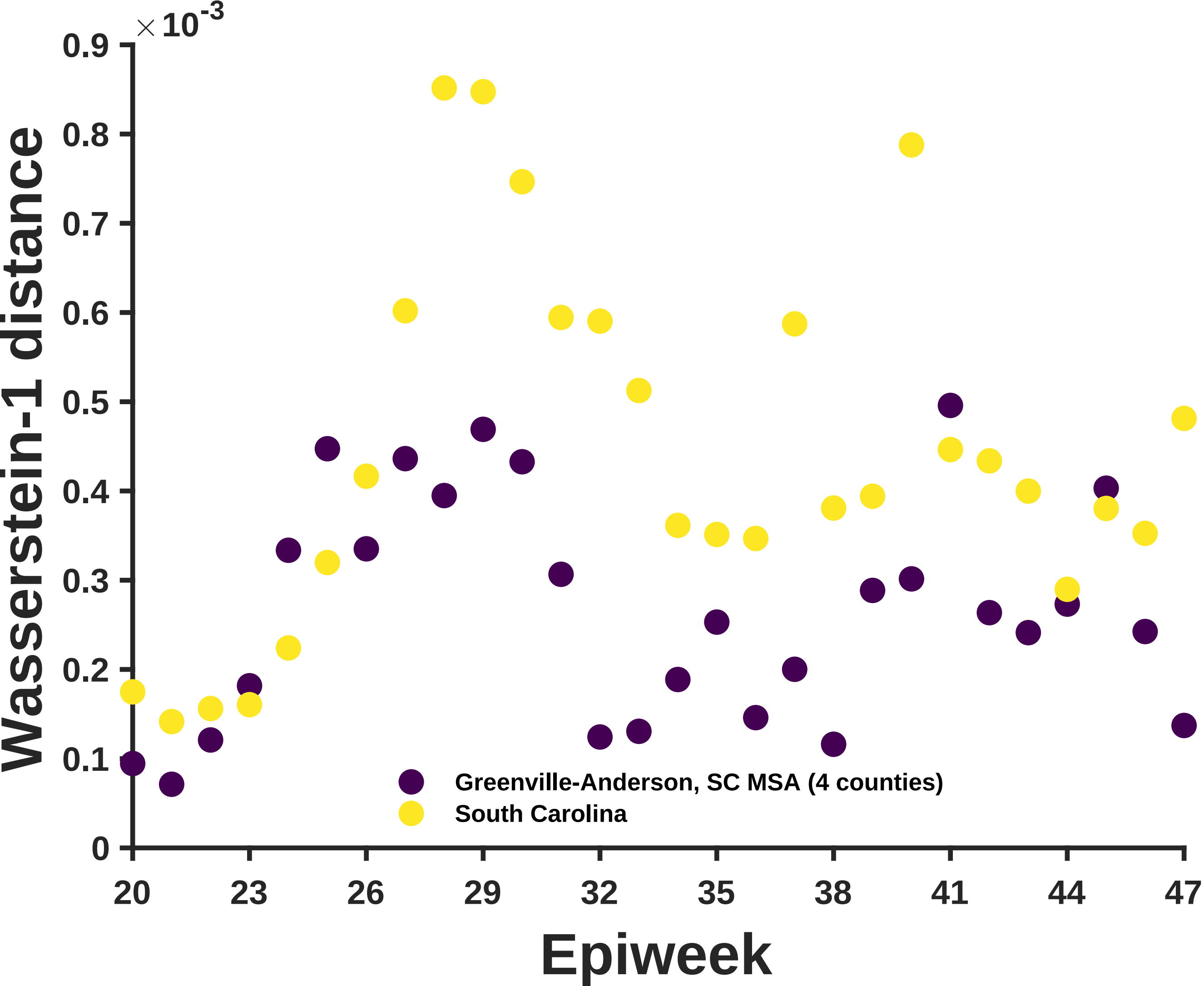

×10^-3
0.9
0.8
0.7
0.6
0.5
0.4
0.3
0.2
0.1
0
Wasserstein-1 distance
20
23
26
29
32
35
38
41
44
47
Epiweek
Greenville-Anderson, SC MSA (4 counties)
South Carolina

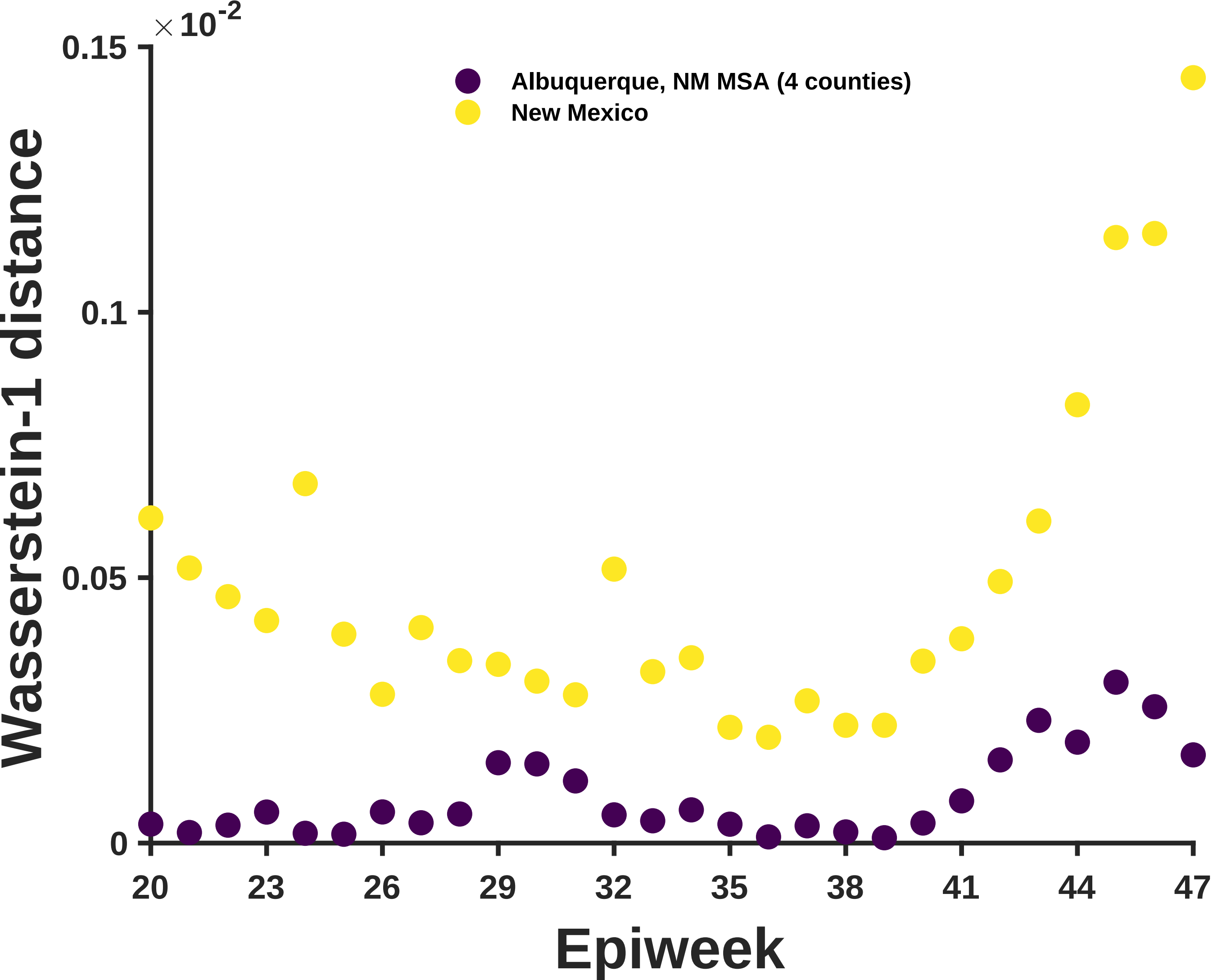

×10^-2
0.15
0.1
0.05
0
Albuquerque, NM MSA (4 counties)
New Mexico
Wasserstein-1 distance
20
23
26
29
32
35
38
41
44
47
Epiweek

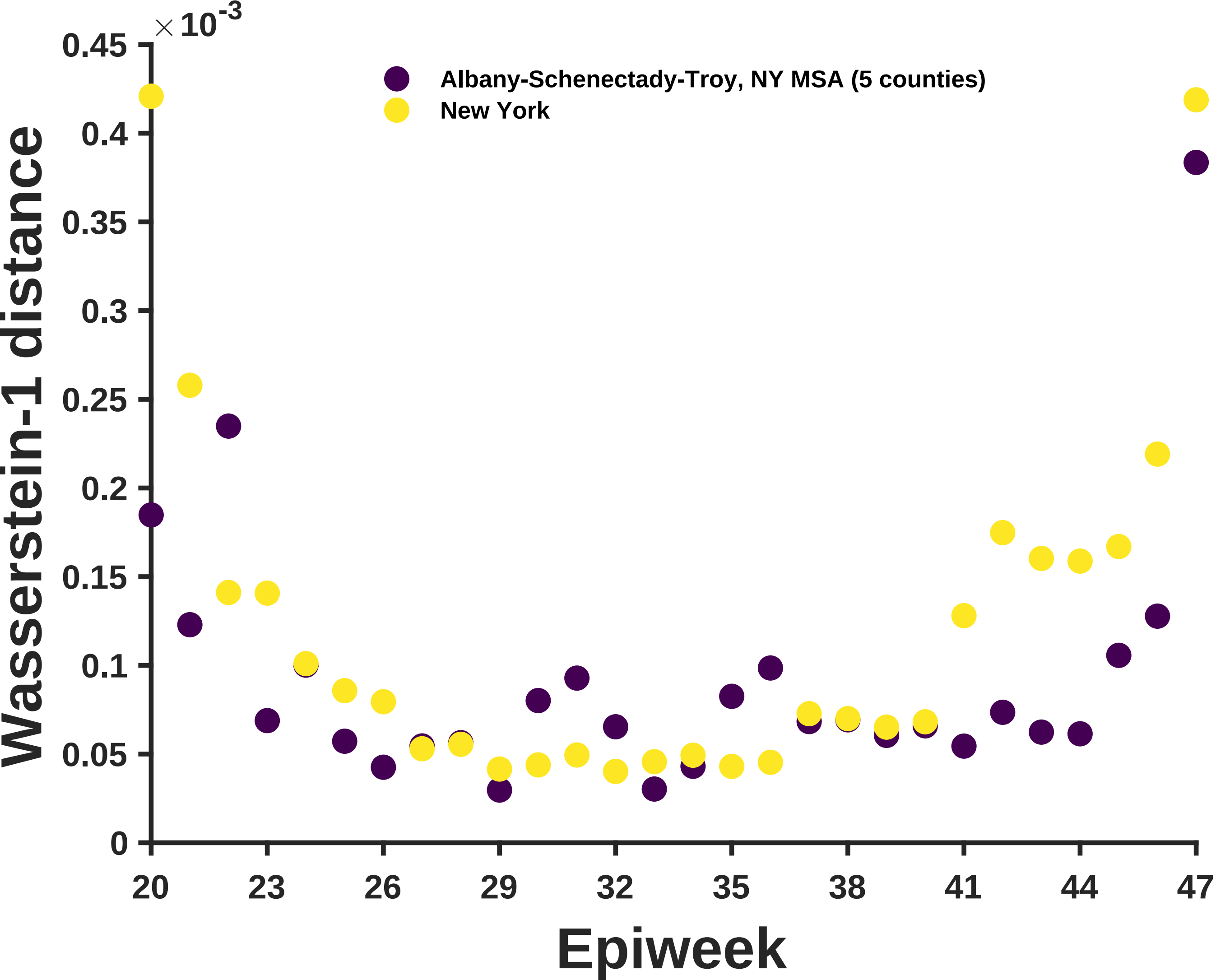

×10^-3
0.45
0.4
0.35
0.3
0.25
0.2
0.15
0.1
0.05
0
Wasserstein-1 distance
Albany-Schenectady-Troy, NY MSA (5 counties)
New York
20
23
26
29
32
35
38
41
44
47
Epiweek

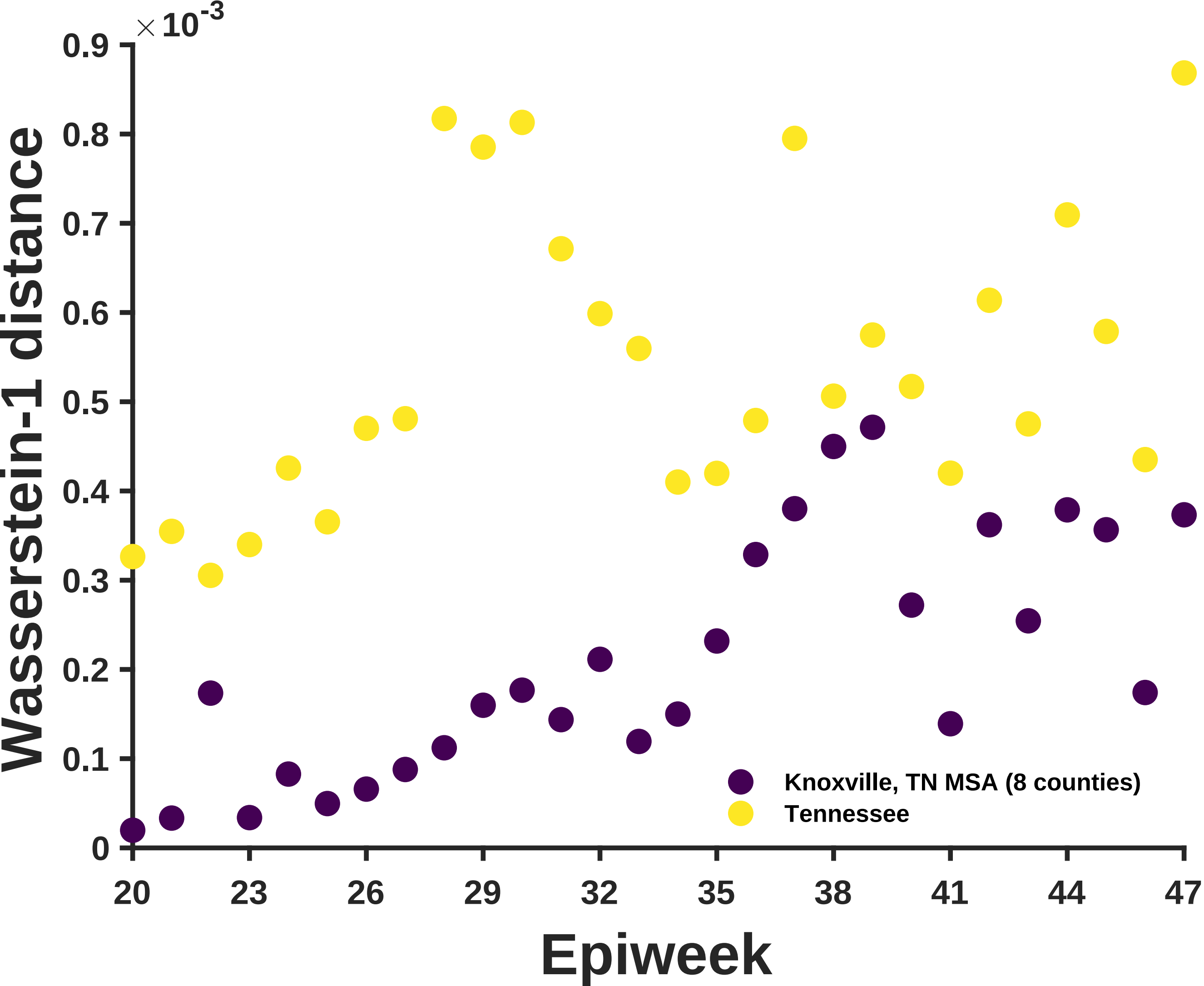

×10^-3
0.9
0.8
0.7
0.6
0.5
0.4
0.3
0.2
0.1
0
Wasserstein-1 distance
20
23
26
29
32
35
38
41
44
47
Epiweek
Knoxville, TN MSA (8 counties)
Tennessee

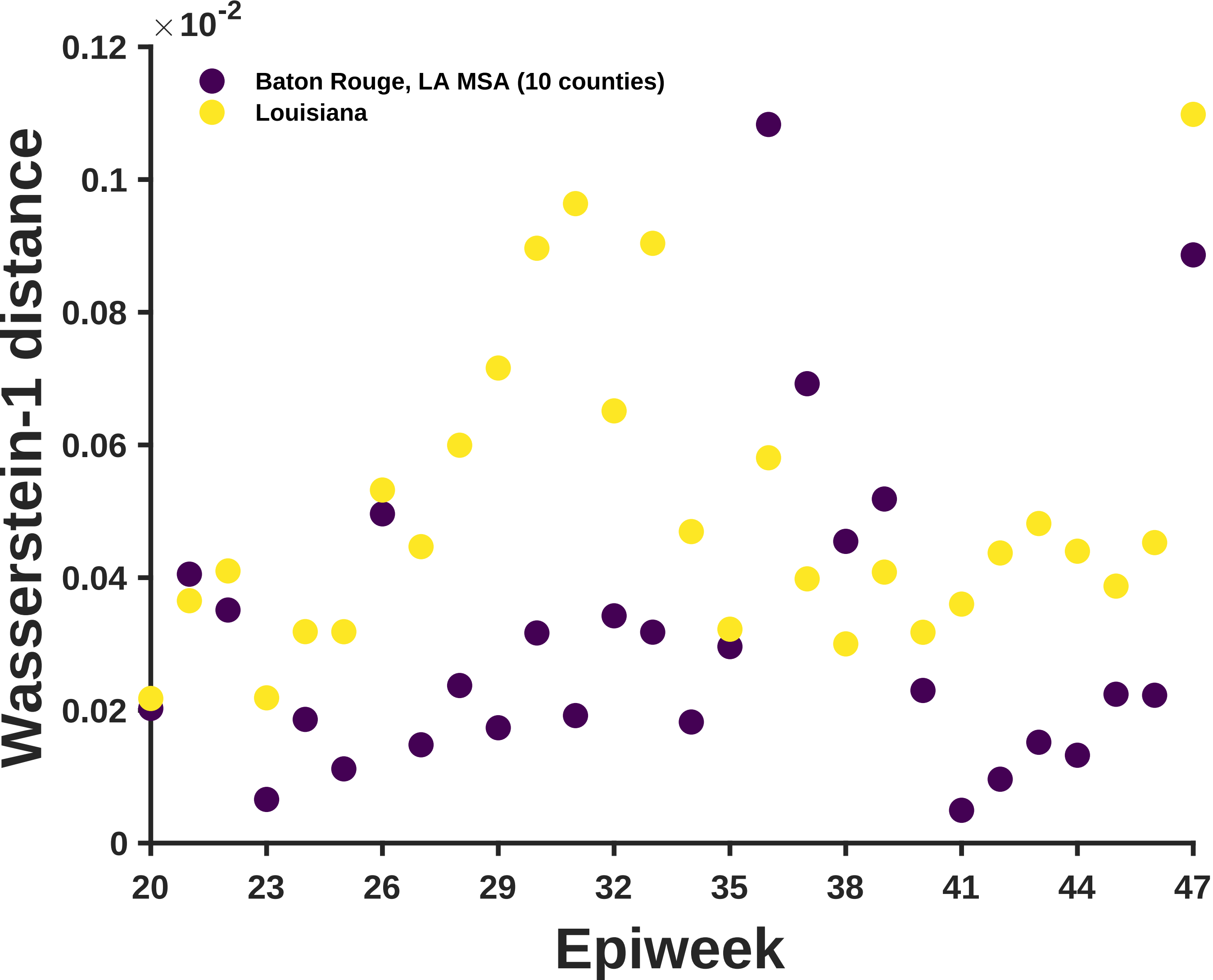

$\times 10^{-2}$
0.12
0.1
0.08
0.06
0.04
0.02
0
Wasserstein-1 distance
Baton Rouge, LA MSA (10 counties)
Louisiana
20
23
26
29
32
35
38
41
44
47
Epiweek

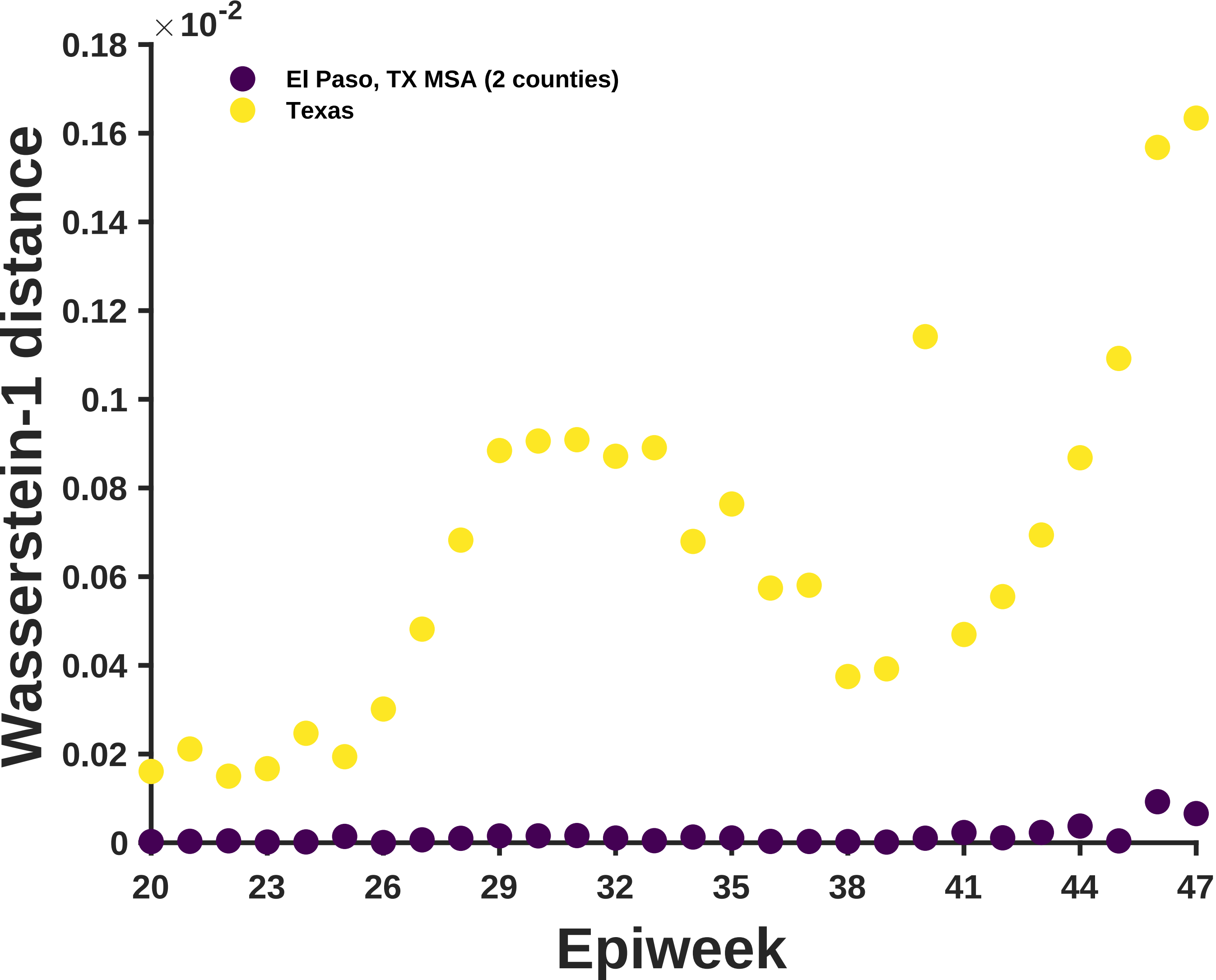

×10^-2
0.18
0.16
0.14
0.12
0.1
0.08
0.06
0.04
0.02
0
Wasserstein-1 distance
El Paso, TX MSA (2 counties)
Texas
20
23
26
29
32
35
38
41
44
47
Epiweek

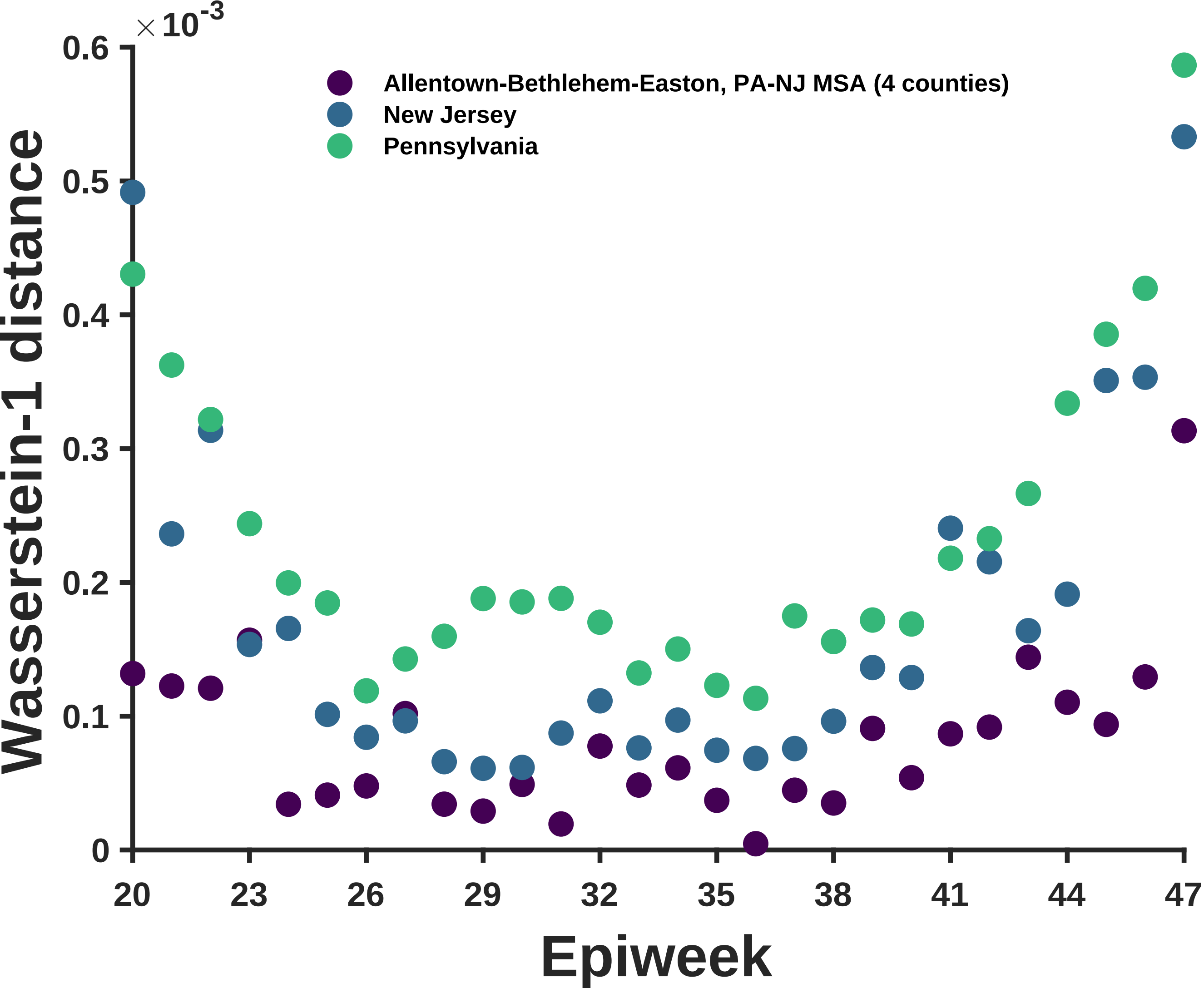

×10^-3
0.6
0.5
0.4
0.3
0.2
0.1
0
Wasserstein-1 distance
Allentown-Bethlehem-Easton, PA-NJ MSA (4 counties)
New Jersey
Pennsylvania
20
23
26
29
32
35
38
41
44
47
Epiweek

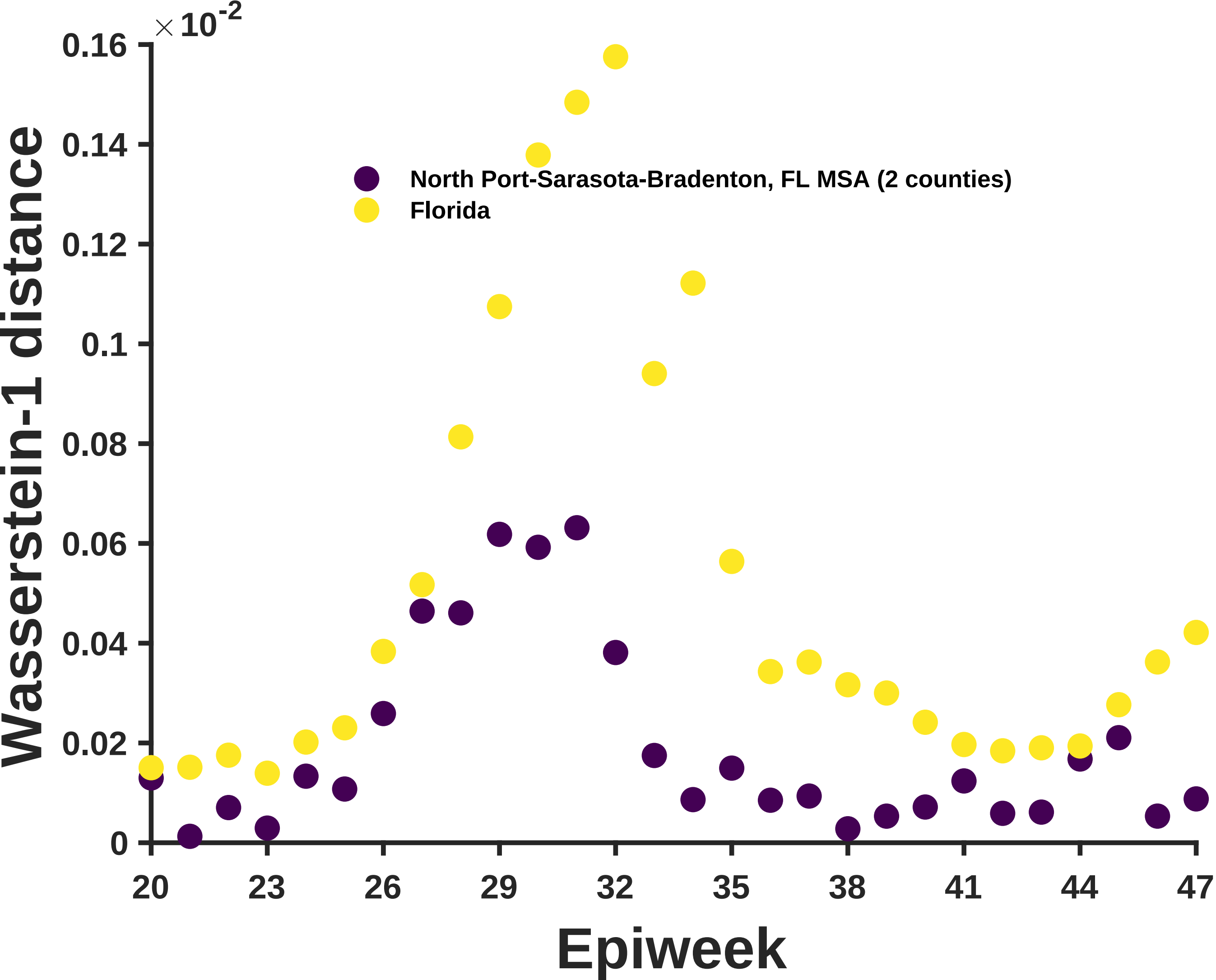

×10^-2
0.16
0.14
0.12
0.1
0.08
0.06
0.04
0.02
0
Wasserstein-1 distance
North Port-Sarasota-Bradenton, FL MSA (2 counties)
Florida
20
23
26
29
32
35
38
41
44
47
Epiweek

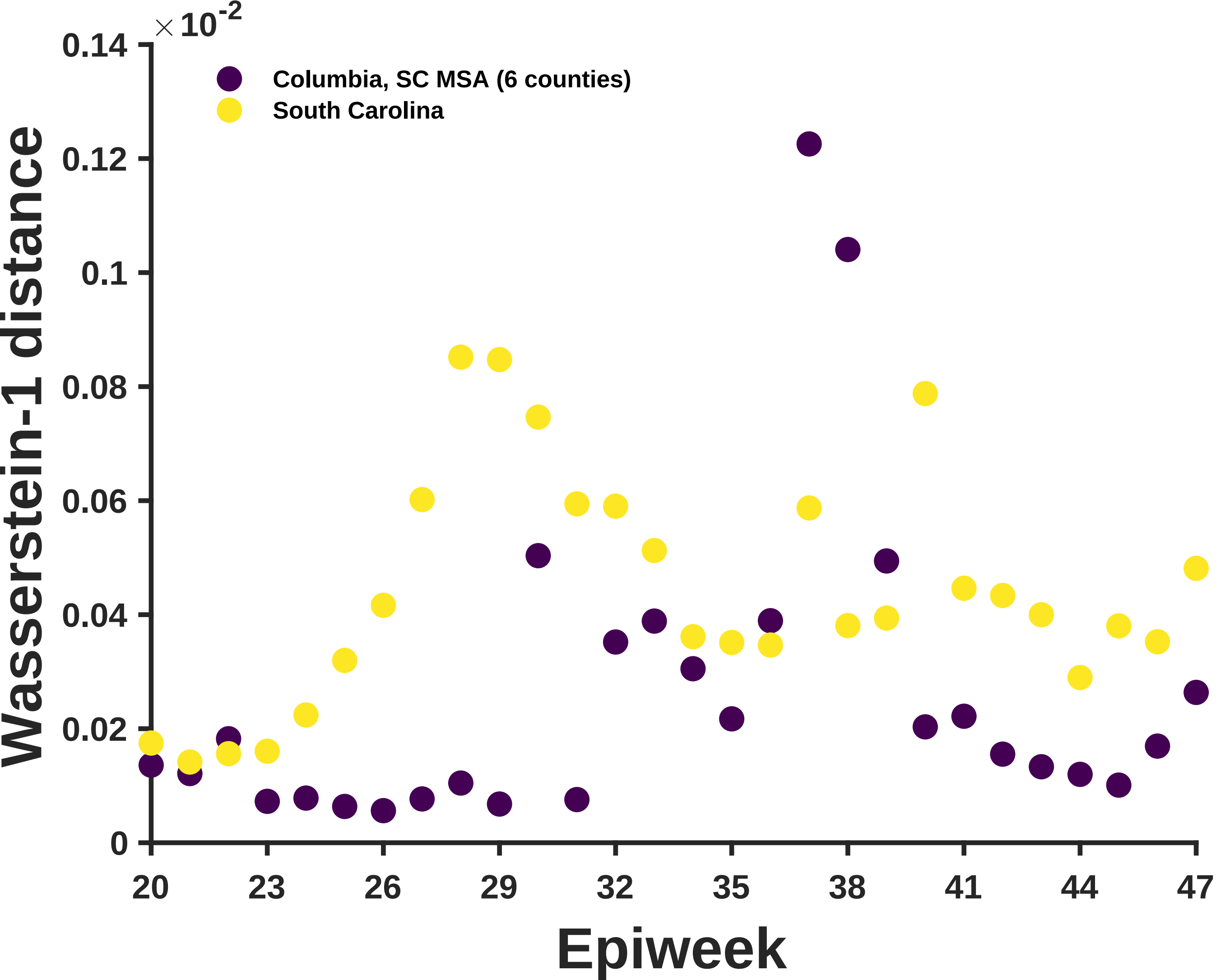

×10-2
0.14
0.12
0.1
0.08
0.06
0.04
0.02
0
Wasserstein-1 distance
Columbia, SC MSA (6 counties)
South Carolina
20
23
26
29
32
35
38
41
44
47
Epiweek

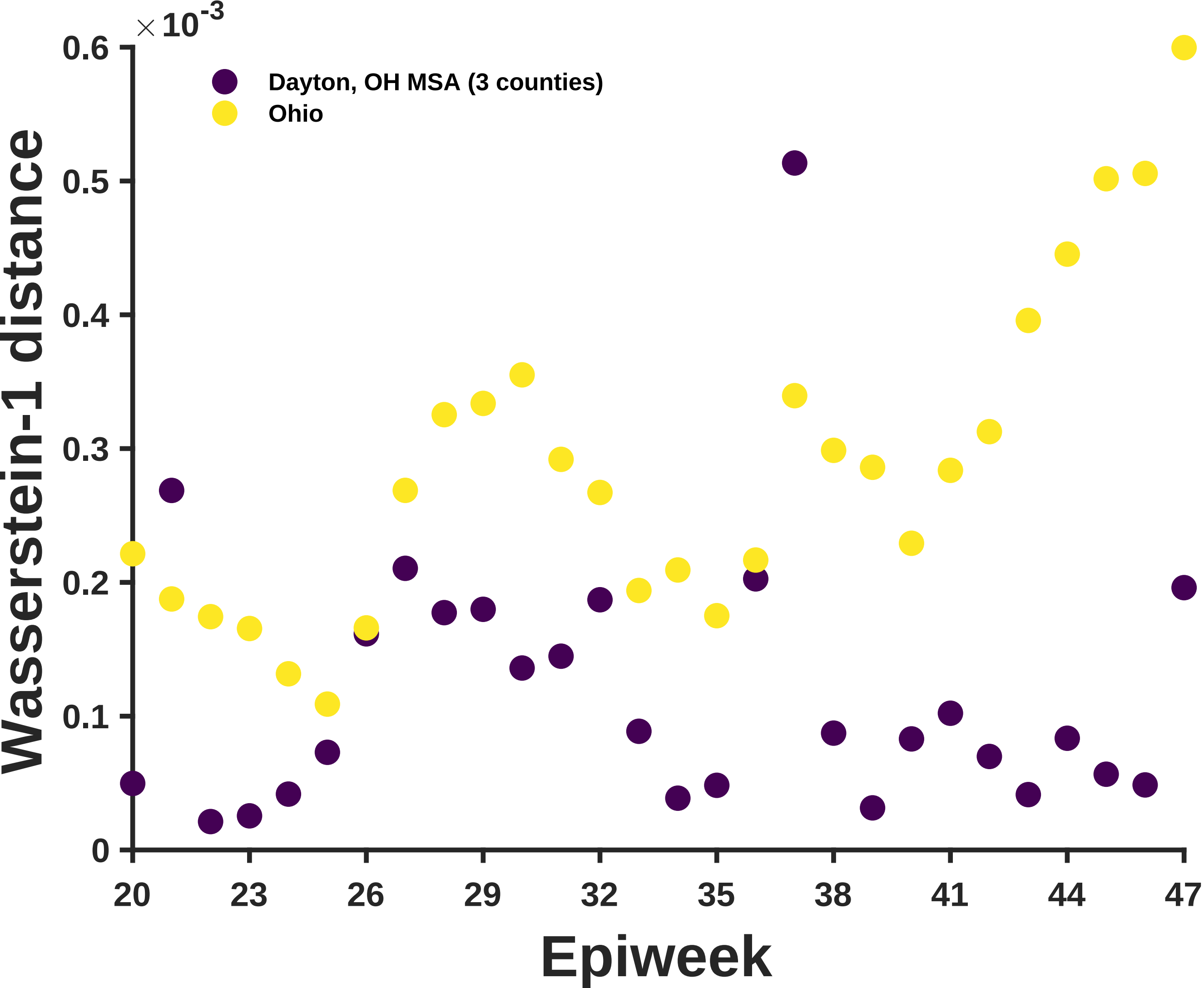
×10^-3
0.6
0.5
0.4
0.3
0.2
0.1
0
Wasserstein-1 distance
Dayton, OH MSA (3 counties)
Ohio
20
23
26
29
32
35
38
41
44
47
Epiweek

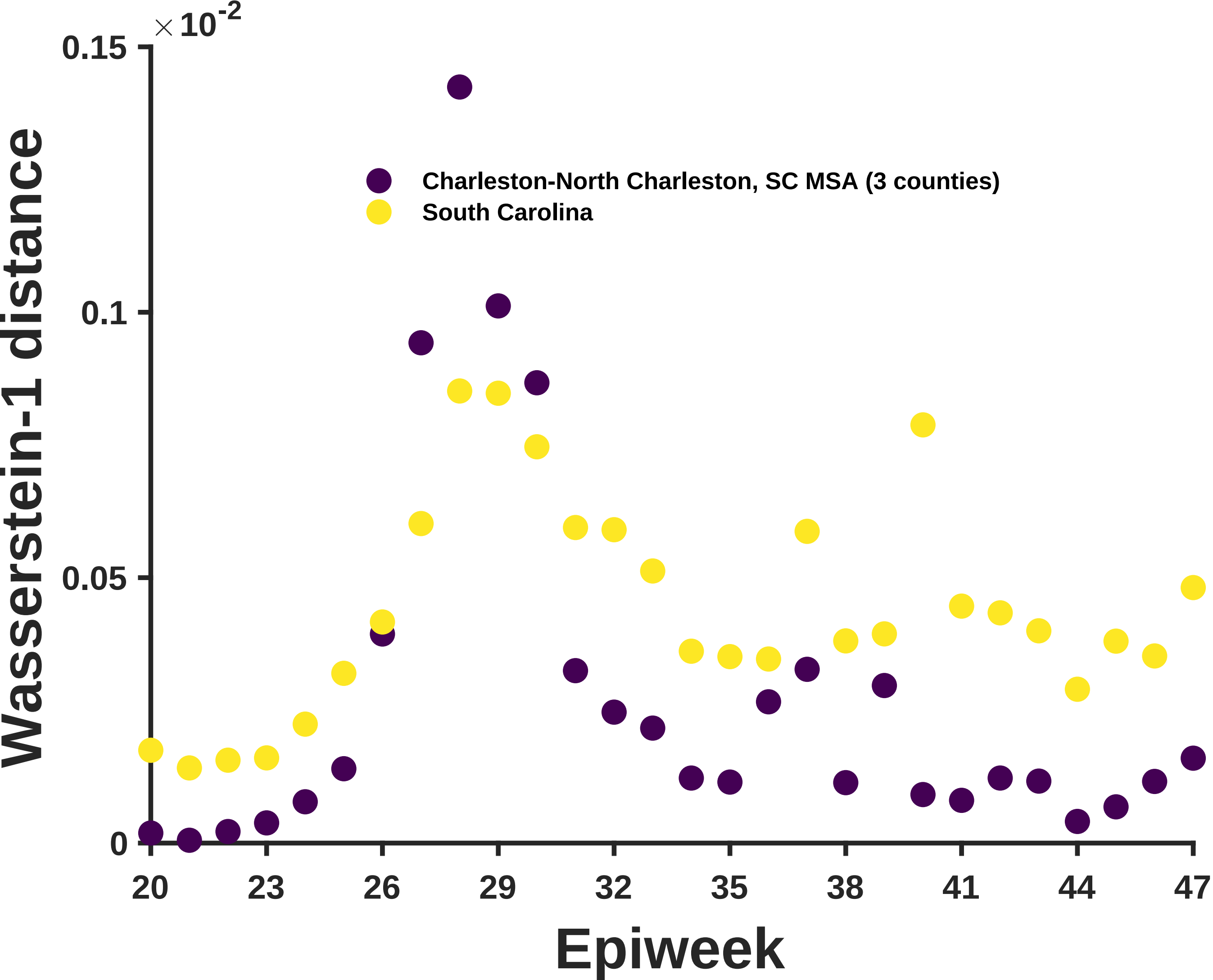

×10-2
0.15
0.1
0.05
0
Wasserstein-1 distance
Charleston-North Charleston, SC MSA (3 counties)
South Carolina
20
23
26
29
32
35
38
41
44
47
Epiweek

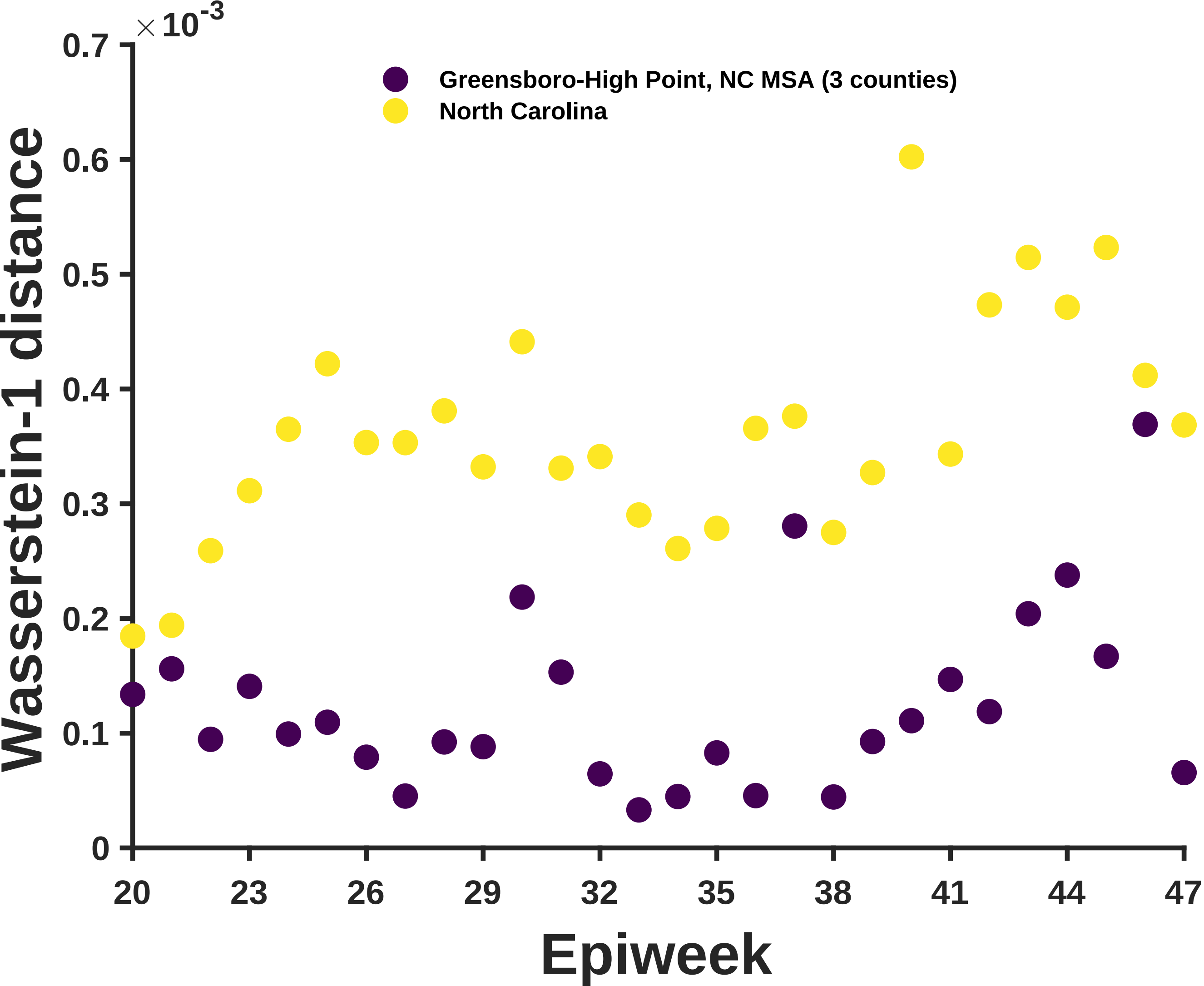

×10^-3
0.7
0.6
0.5
0.4
0.3
0.2
0.1
0
Wasserstein-1 distance
Greensboro-High Point, NC MSA (3 counties)
North Carolina
20
23
26
29
32
35
38
41
44
47
Epiweek

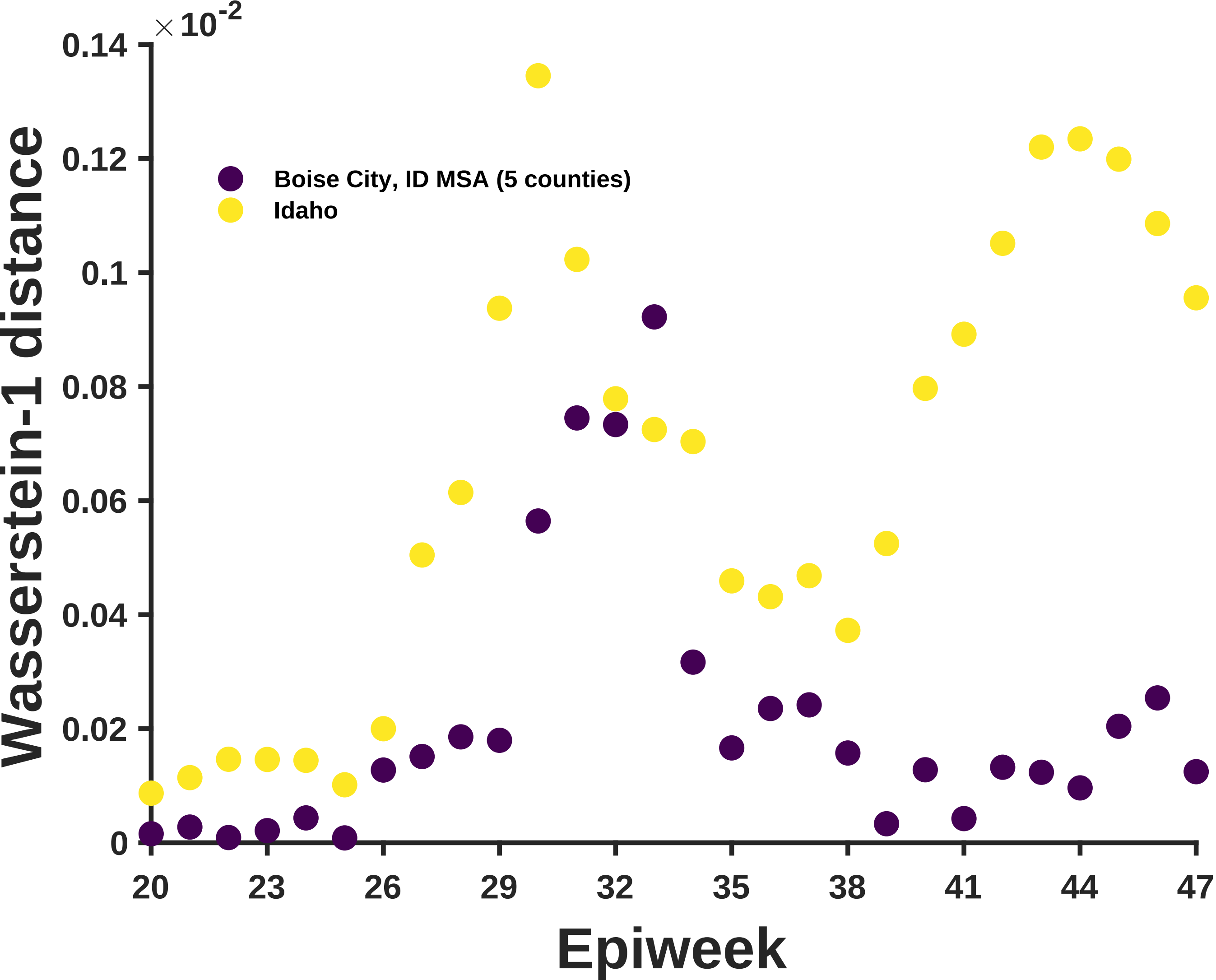

×10^-2
0.14
0.12
0.1
0.08
0.06
0.04
0.02
0
Wasserstein-1 distance
Boise City, ID MSA (5 counties)
Idaho
20
23
26
29
32
35
38
41
44
47
Epiweek

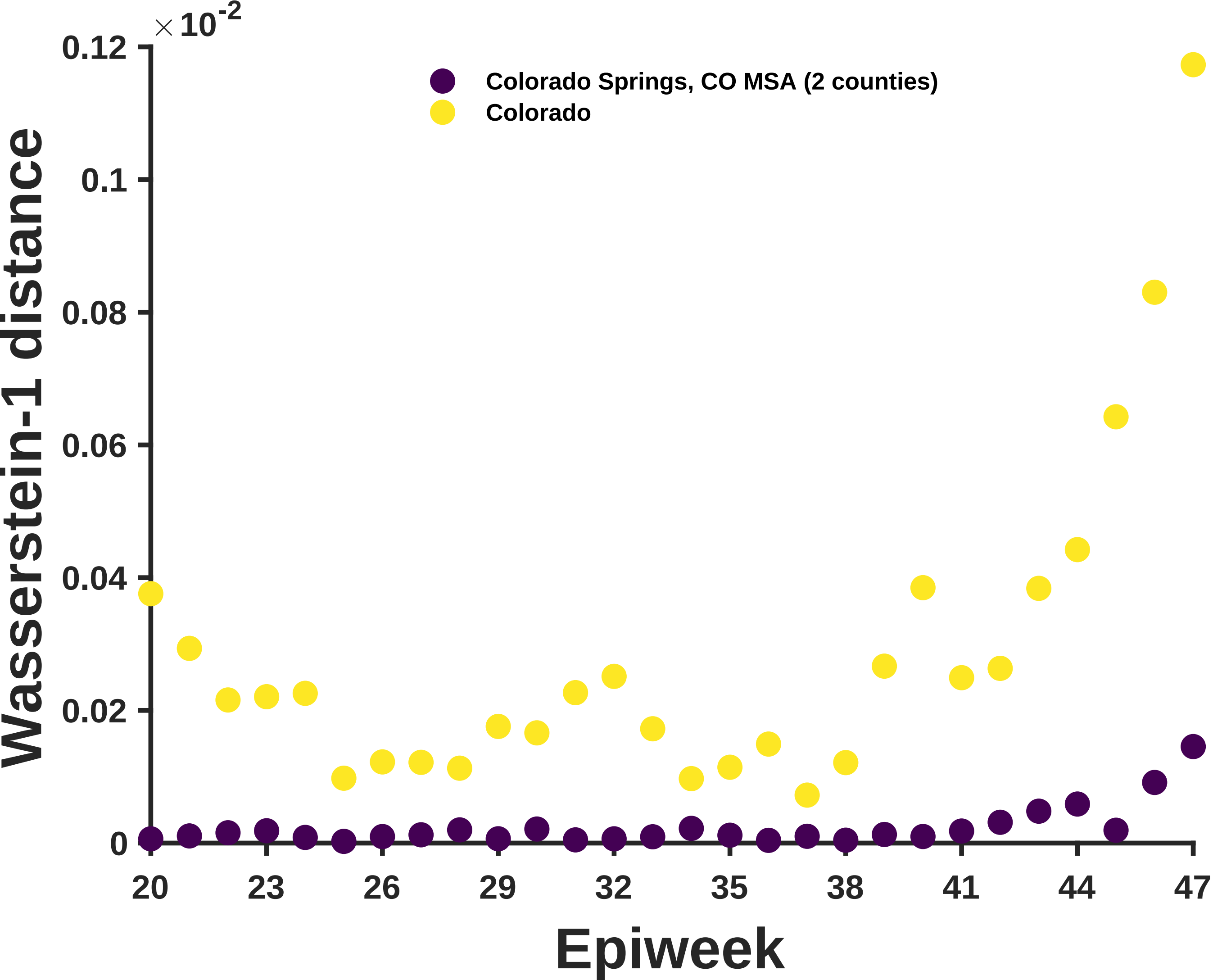

×10^-2
0.12
0.1
0.08
0.06
0.04
0.02
0
Wasserstein-1 distance
Colorado Springs, CO MSA (2 counties)
Colorado
20
23
26
29
32
35
38
41
44
47
Epiweek

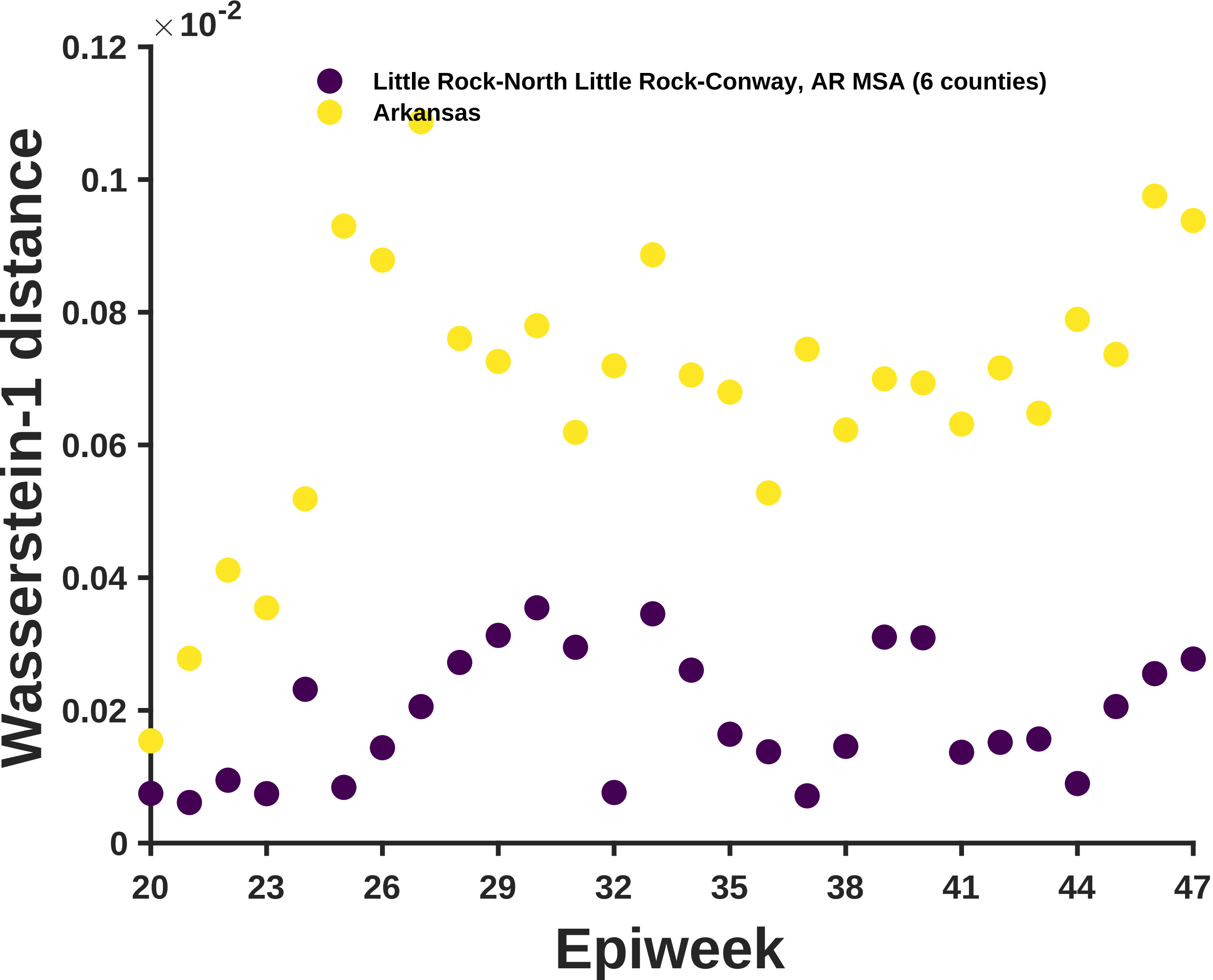

×10^-2
0.12
0.1
0.08
0.06
0.04
0.02
0
Wasserstein-1 distance
Little Rock-North Little Rock-Conway, AR MSA (6 counties)
Arkansas
20
23
26
29
32
35
38
41
44
47
Epiweek

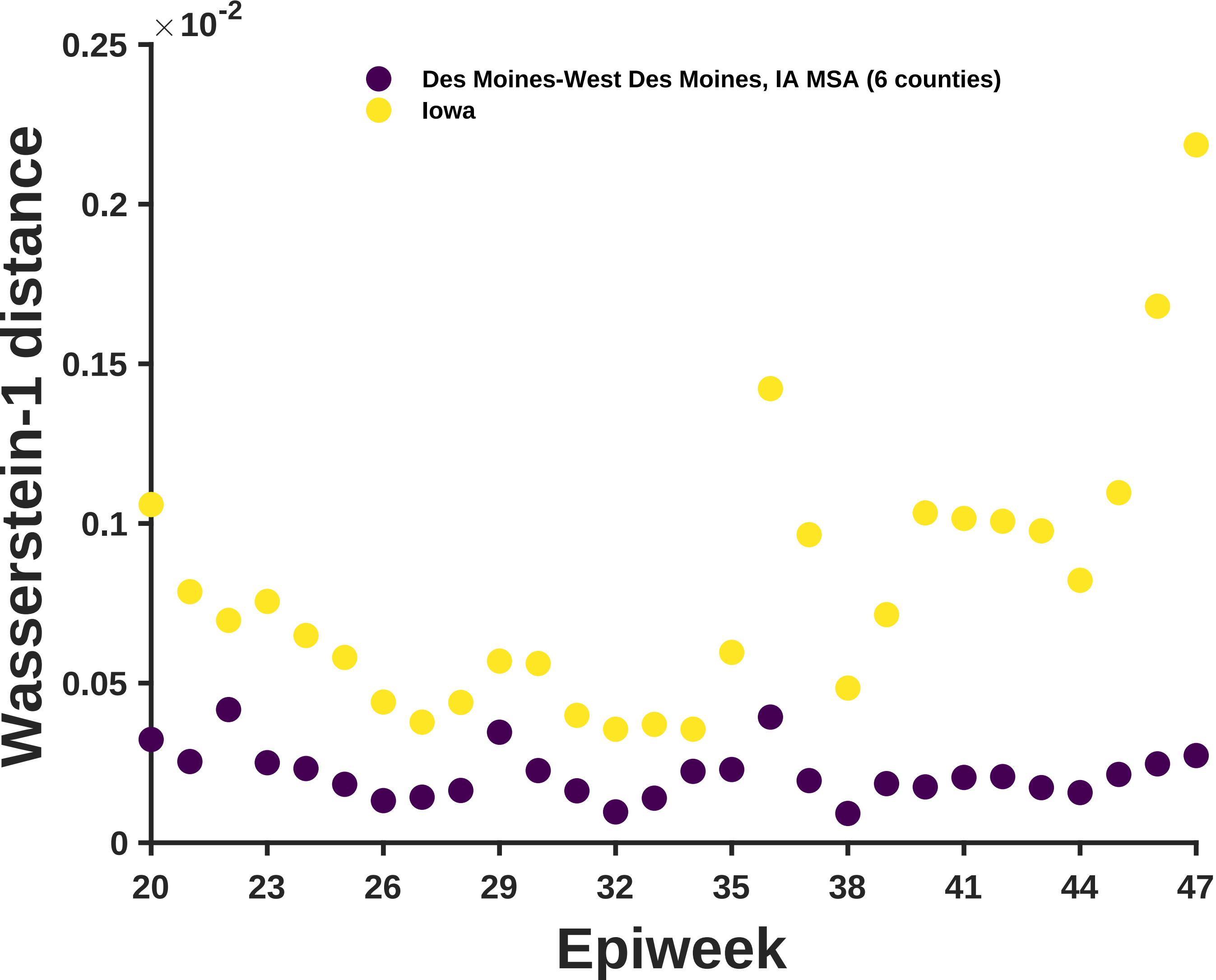
×10-2
0.25
0.2
0.15
0.1
0.05
0
Des Moines-West Des Moines, IA MSA (6 counties)
Iowa
Wasserstein-1 distance
20
23
26
29
32
35
38
41
44
47
Epiweek

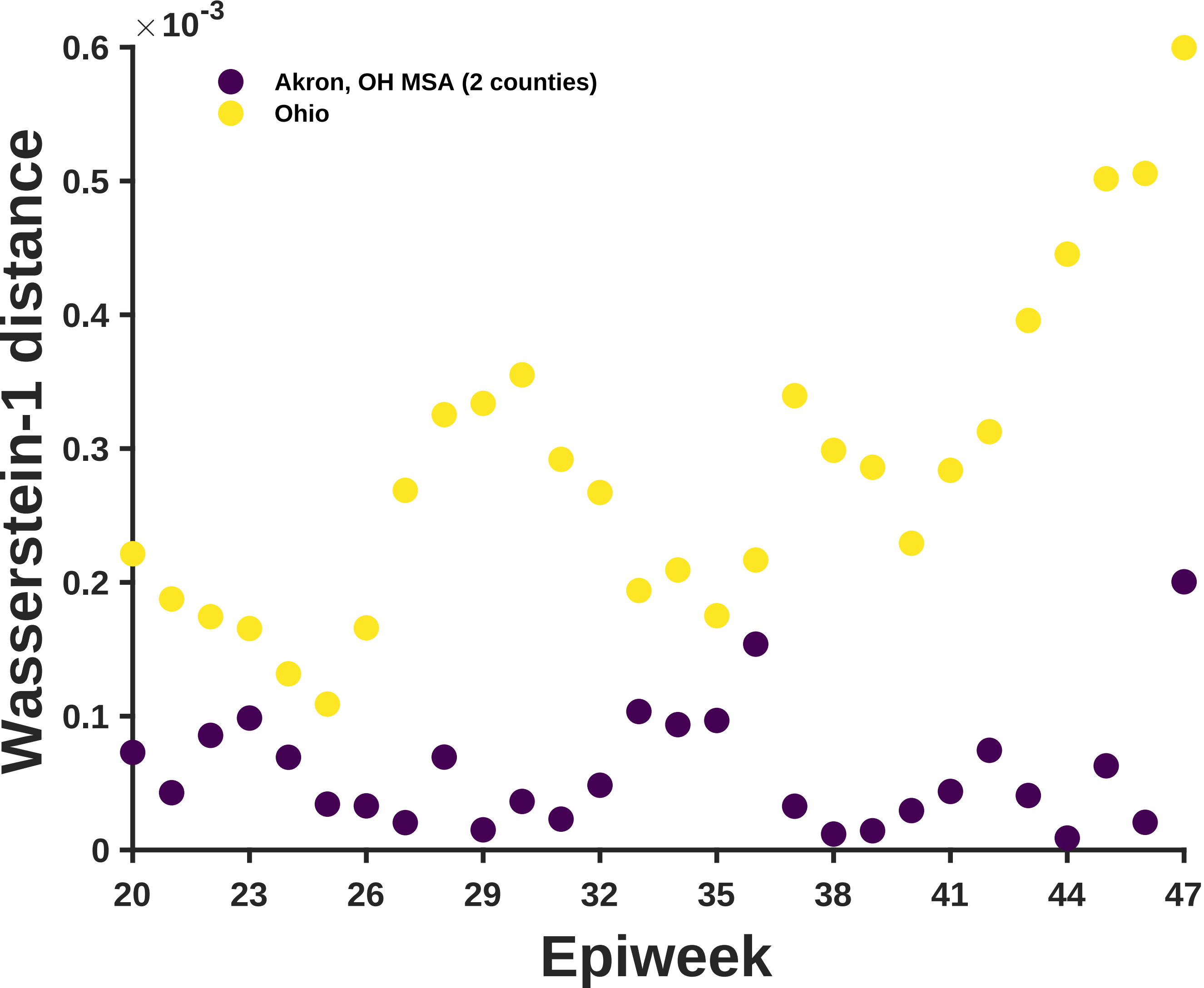

×10^-3
0.6
0.5
0.4
0.3
0.2
0.1
0
Akron, OH MSA (2 counties)
Ohio
Wasserstein-1 distance
20
23
26
29
32
35
38
41
44
47
Epiweek

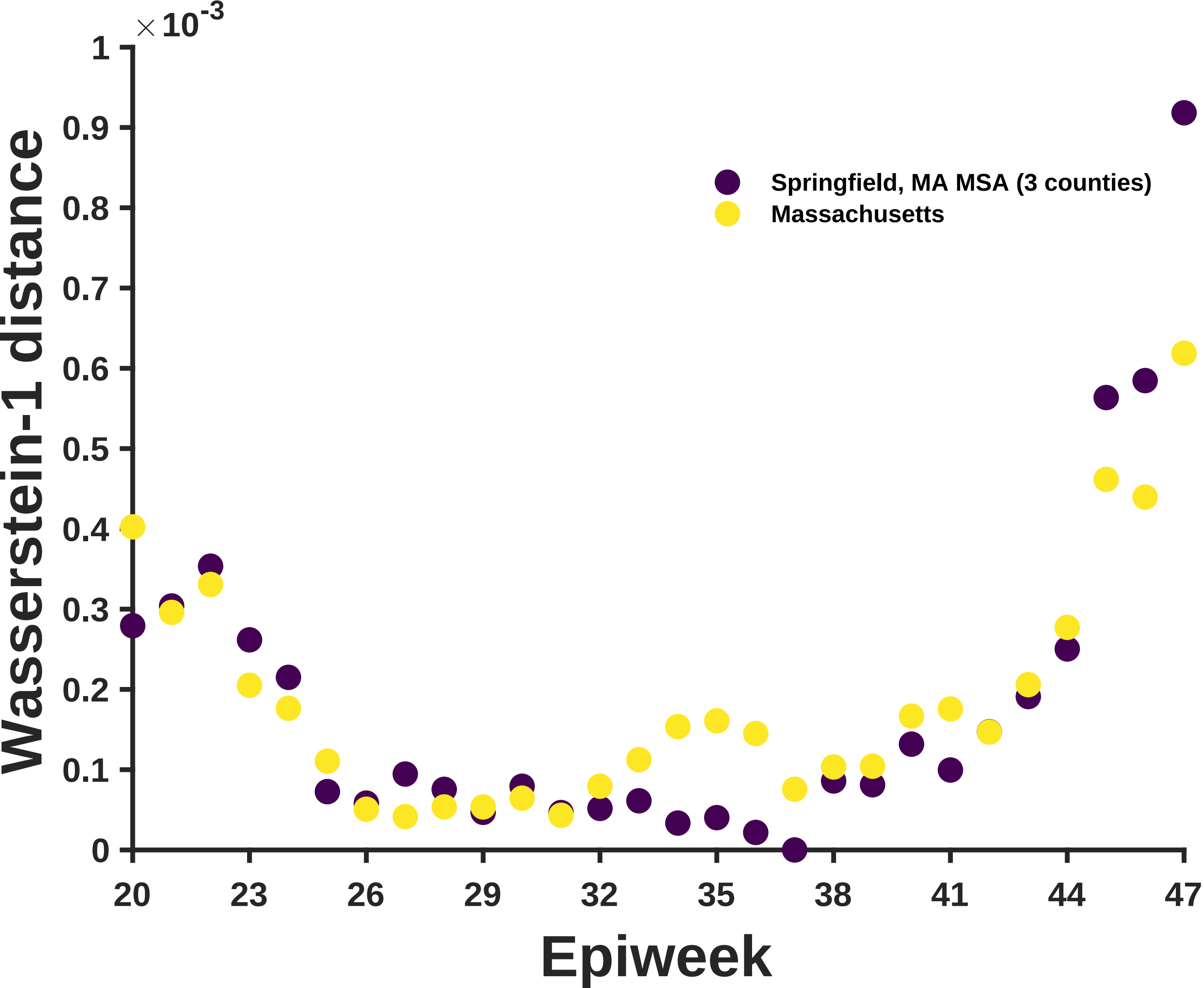

×10^-3
1
0.9
0.8
0.7
0.6
0.5
0.4
0.3
0.2
0.1
0
Wasserstein-1 distance
20
23
26
29
32
35
38
41
44
47
Epiweek
Springfield, MA MSA (3 counties)
Massachusetts

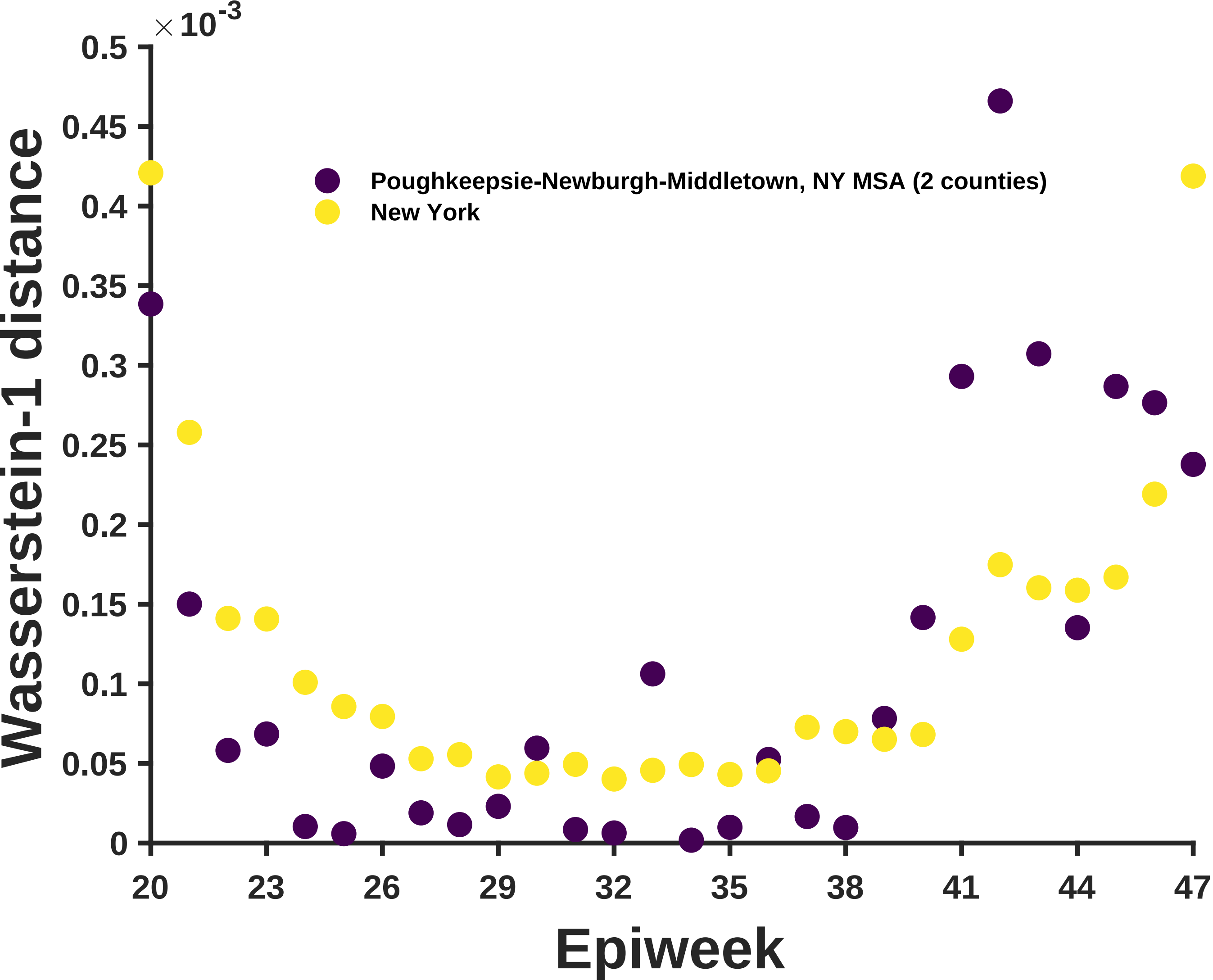

×10^-3
0.5
0.45
0.4
0.35
0.3
0.25
0.2
0.15
0.1
0.05
0
Wasserstein-1 distance
Poughkeepsie-Newburgh-Middletown, NY MSA (2 counties)
New York
20
23
26
29
32
35
38
41
44
47
Epiweek

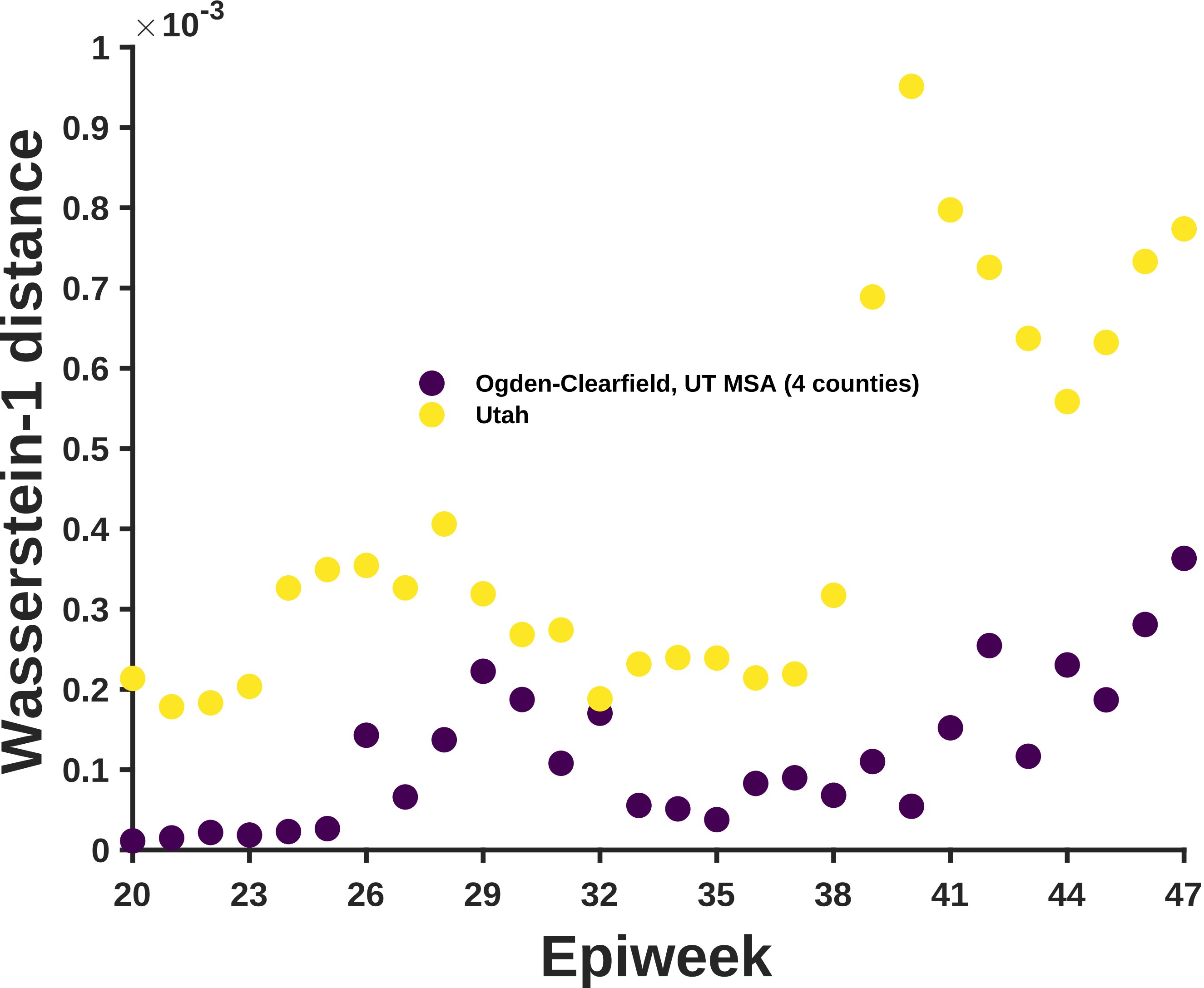
×10^-3
Wasserstein-1 distance
0
0.1
0.2
0.3
0.4
0.5
0.6
0.7
0.8
0.9
1
Ogden-Clearfield, UT MSA (4 counties)
Utah
20
23
26
29
32
35
38
41
44
47
Epiweek

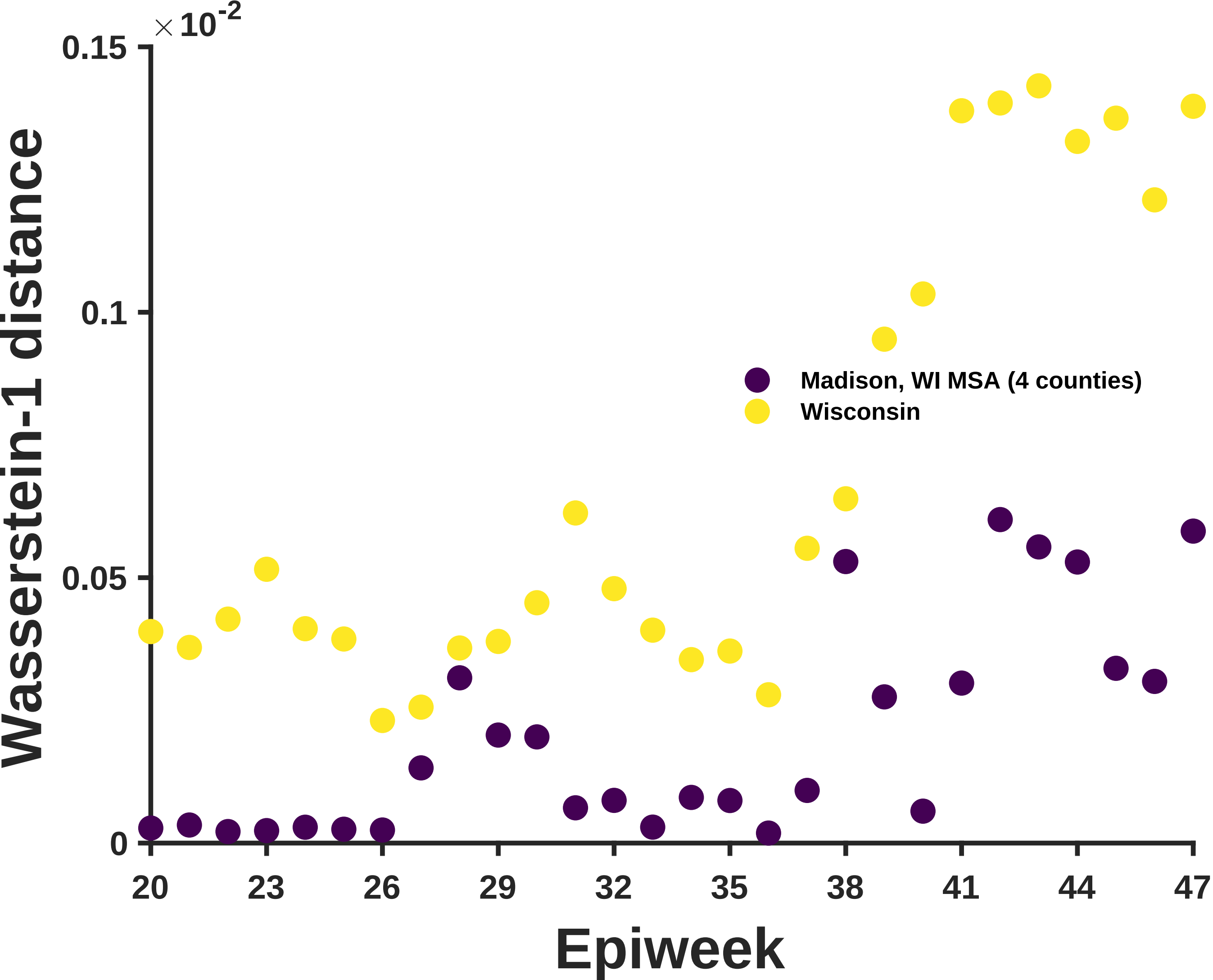

$\times 10^{-2}$
0.15
0.1
0.05
0
Wasserstein-1 distance
20
23
26
29
32
35
38
41
44
47
Epiweek
Madison, WI MSA (4 counties)
Wisconsin

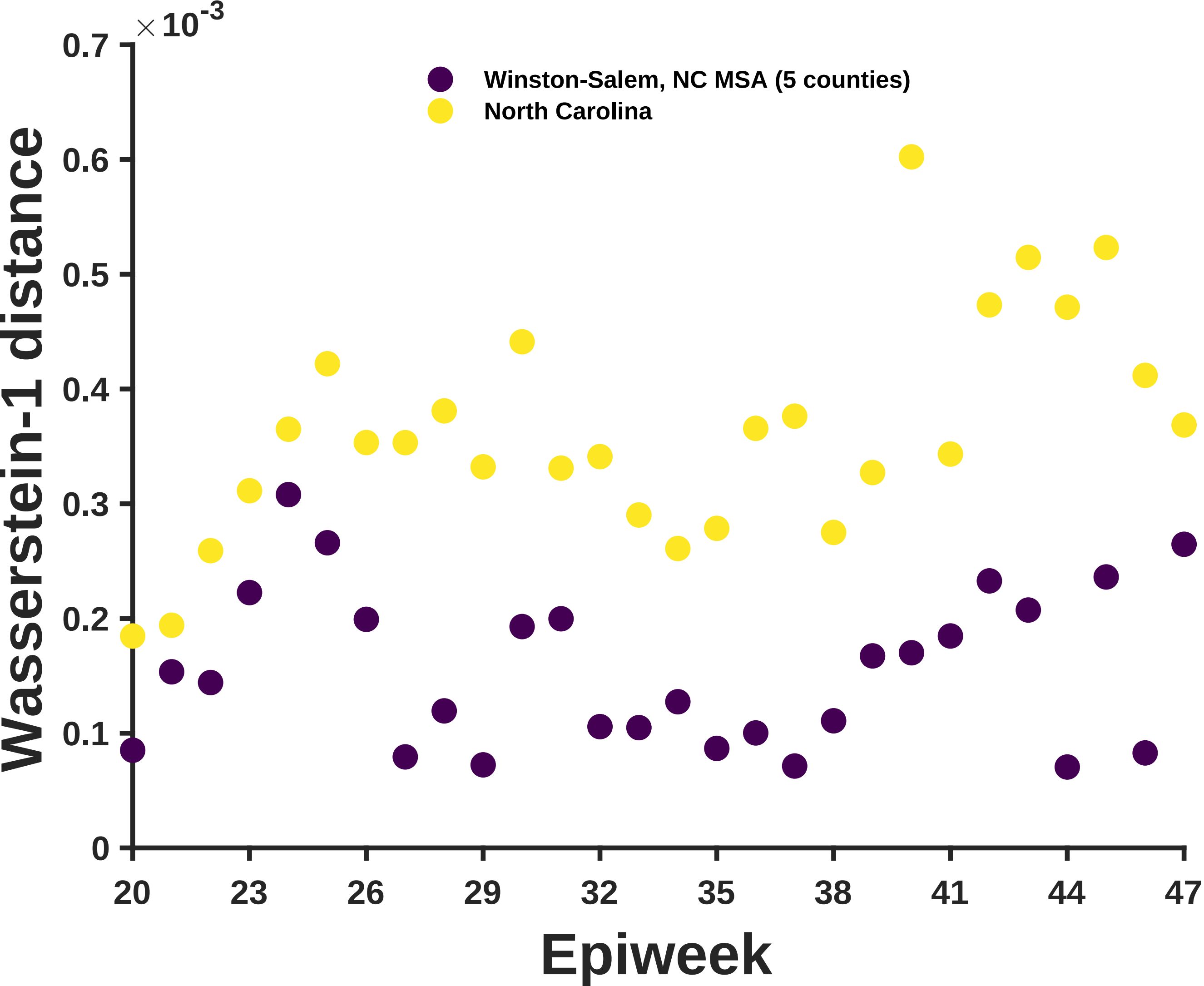

×10^-3
0.7
0.6
0.5
0.4
0.3
0.2
0.1
0
Winston-Salem, NC MSA (5 counties)
North Carolina
Wasserstein-1 distance
20
23
26
29
32
35
38
41
44
47
Epiweek

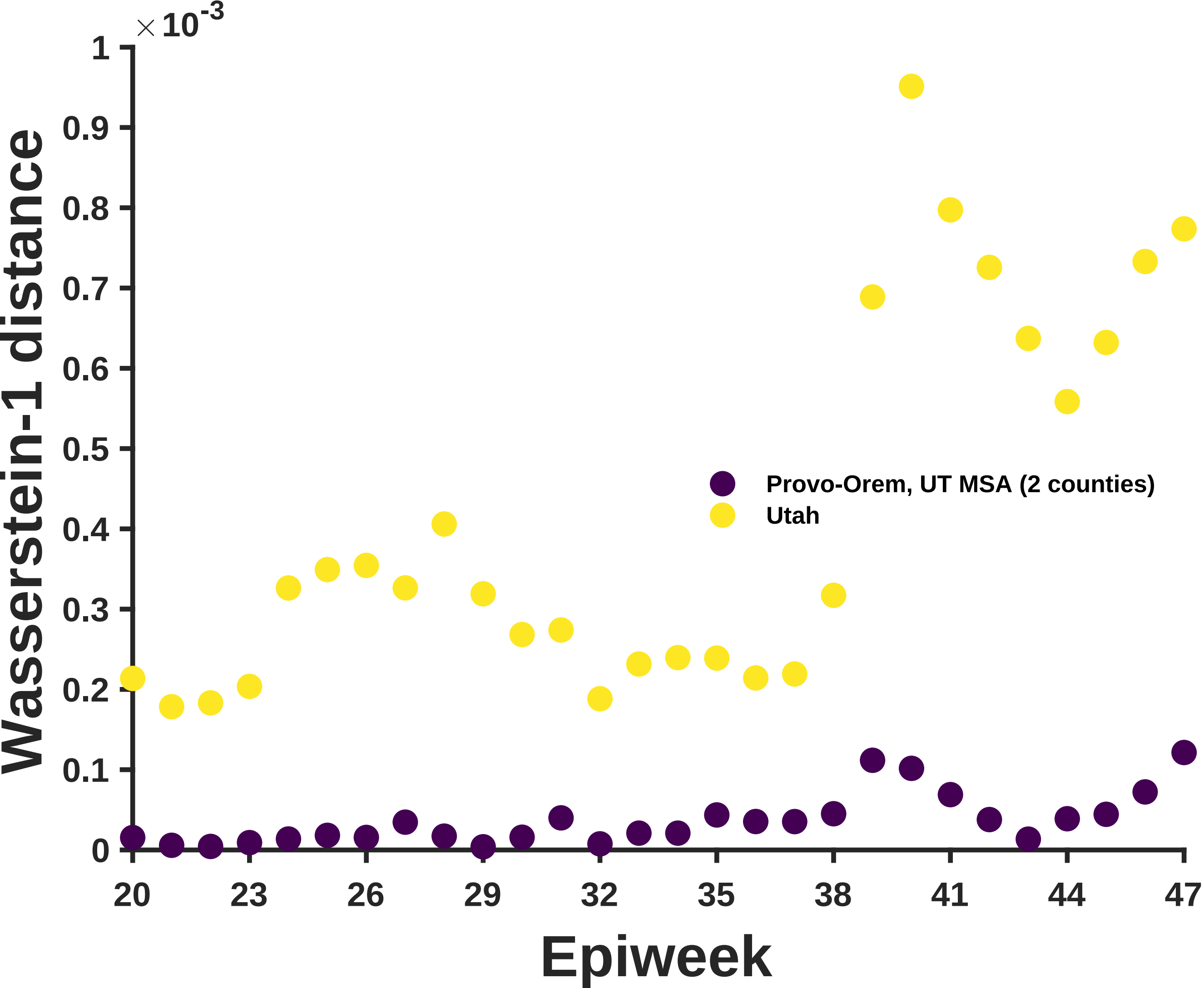

$\times 10^{-3}$
1
0.9
0.8
0.7
0.6
0.5
0.4
0.3
0.2
0.1
0
Wasserstein-1 distance
20
23
26
29
32
35
38
41
44
47
Epiweek
Provo-Orem, UT MSA (2 counties)
Utah

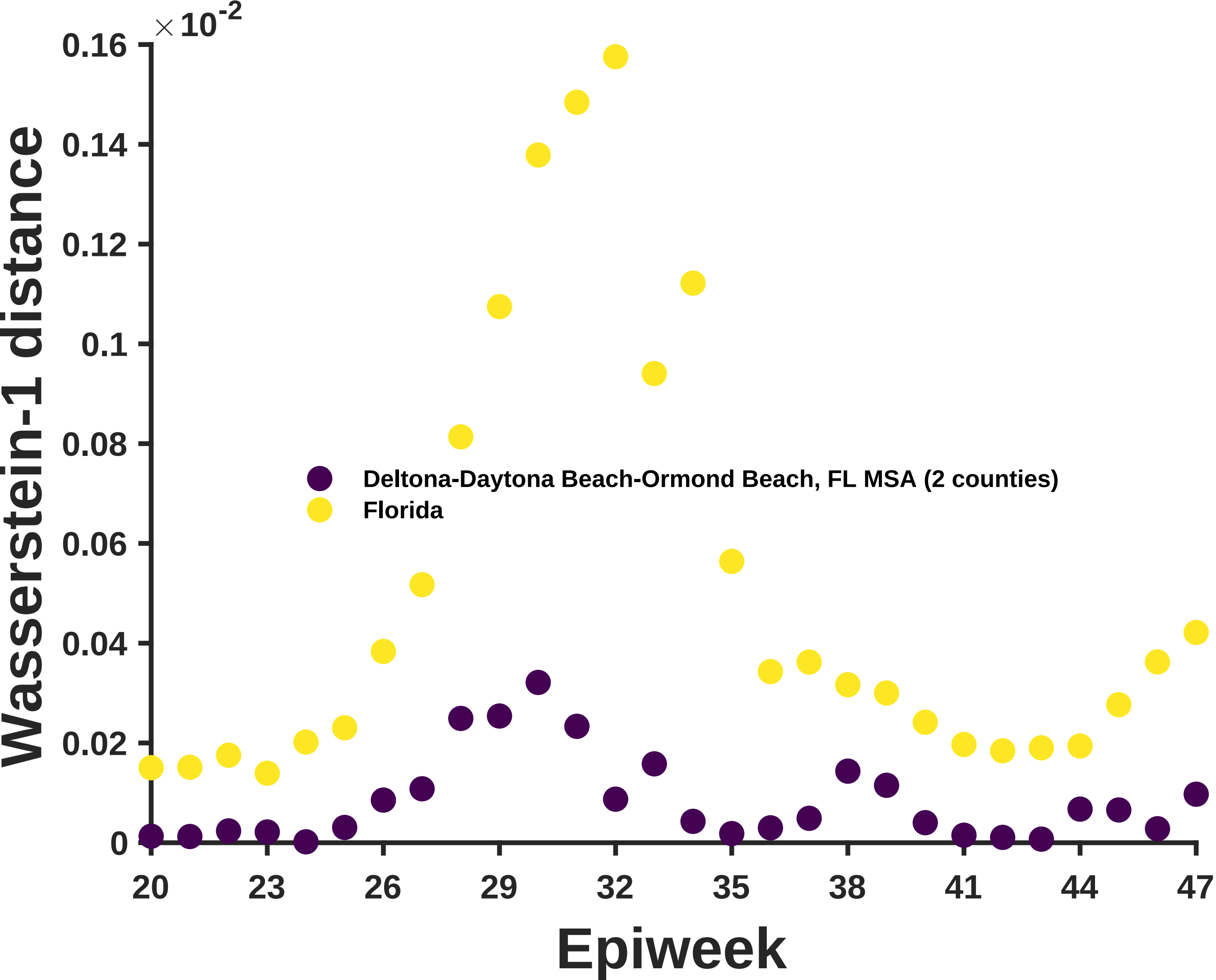

×10^-2
0.16
0.14
0.12
0.1
0.08
0.06
0.04
0.02
0
Wasserstein-1 distance
20
23
26
29
32
35
38
41
44
47
Epiweek
Deltona-Daytona Beach-Ormond Beach, FL MSA (2 counties)
Florida

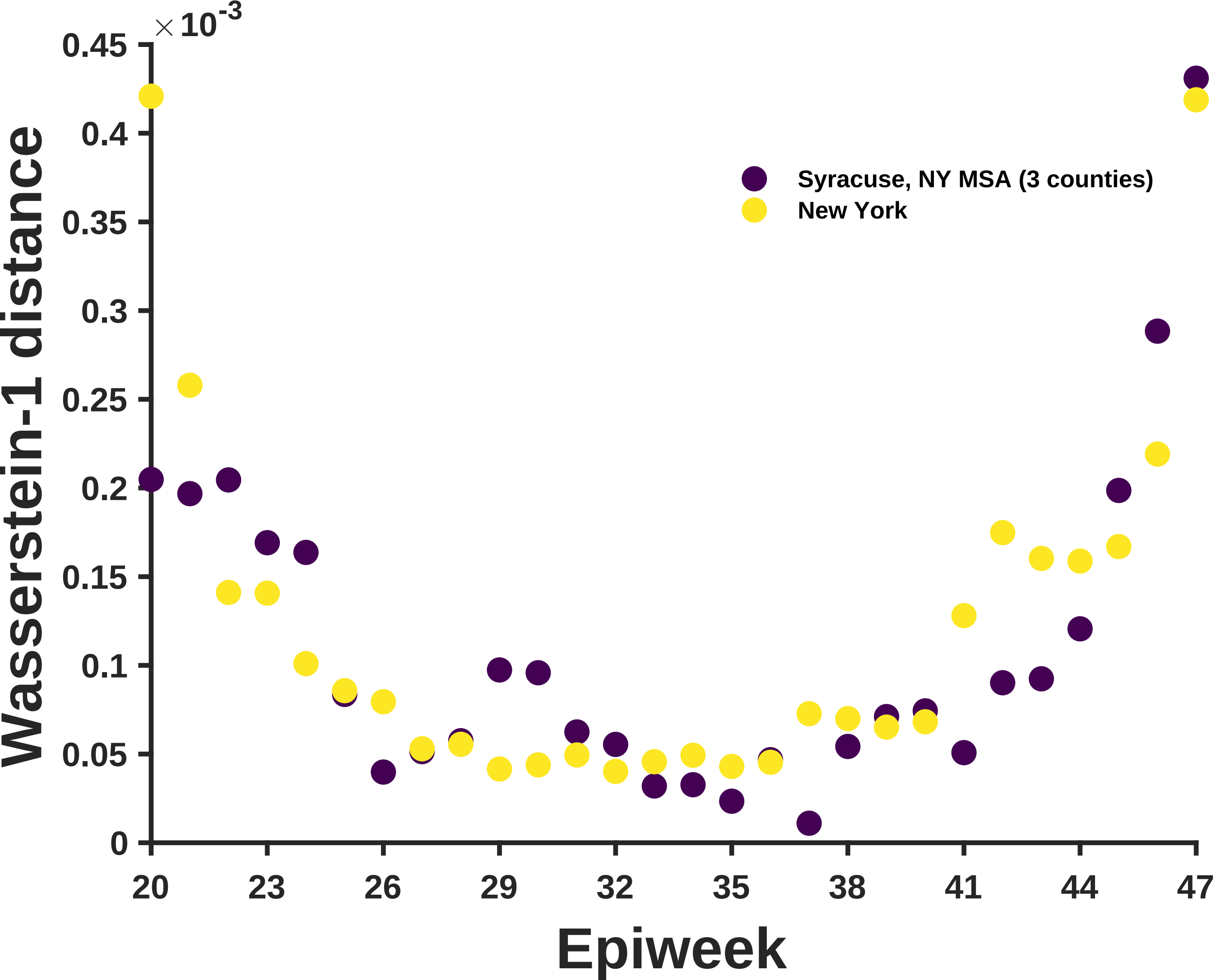

×10^-3
0.45
0.4
0.35
0.3
0.25
0.2
0.15
0.1
0.05
0
Wasserstein-1 distance
20
23
26
29
32
35
38
41
44
47
Epiweek
Syracuse, NY MSA (3 counties)
New York

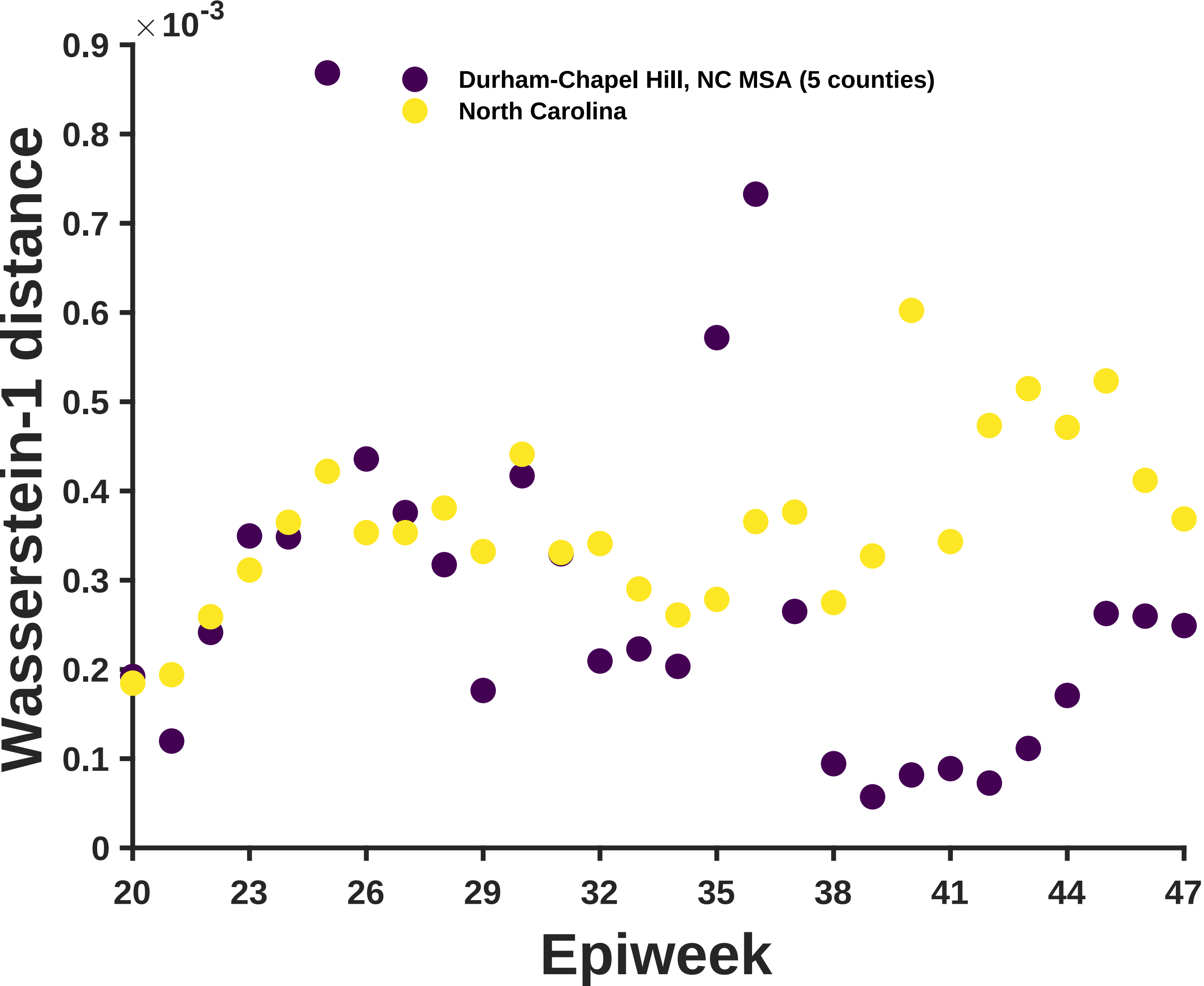
×10^-3
Durham-Chapel Hill, NC MSA (5 counties)
North Carolina
Wasserstein-1 distance
Epiweek
0
0.1
0.2
0.3
0.4
0.5
0.6
0.7
0.8
0.9
20
23
26
29
32
35
38
41
44
47

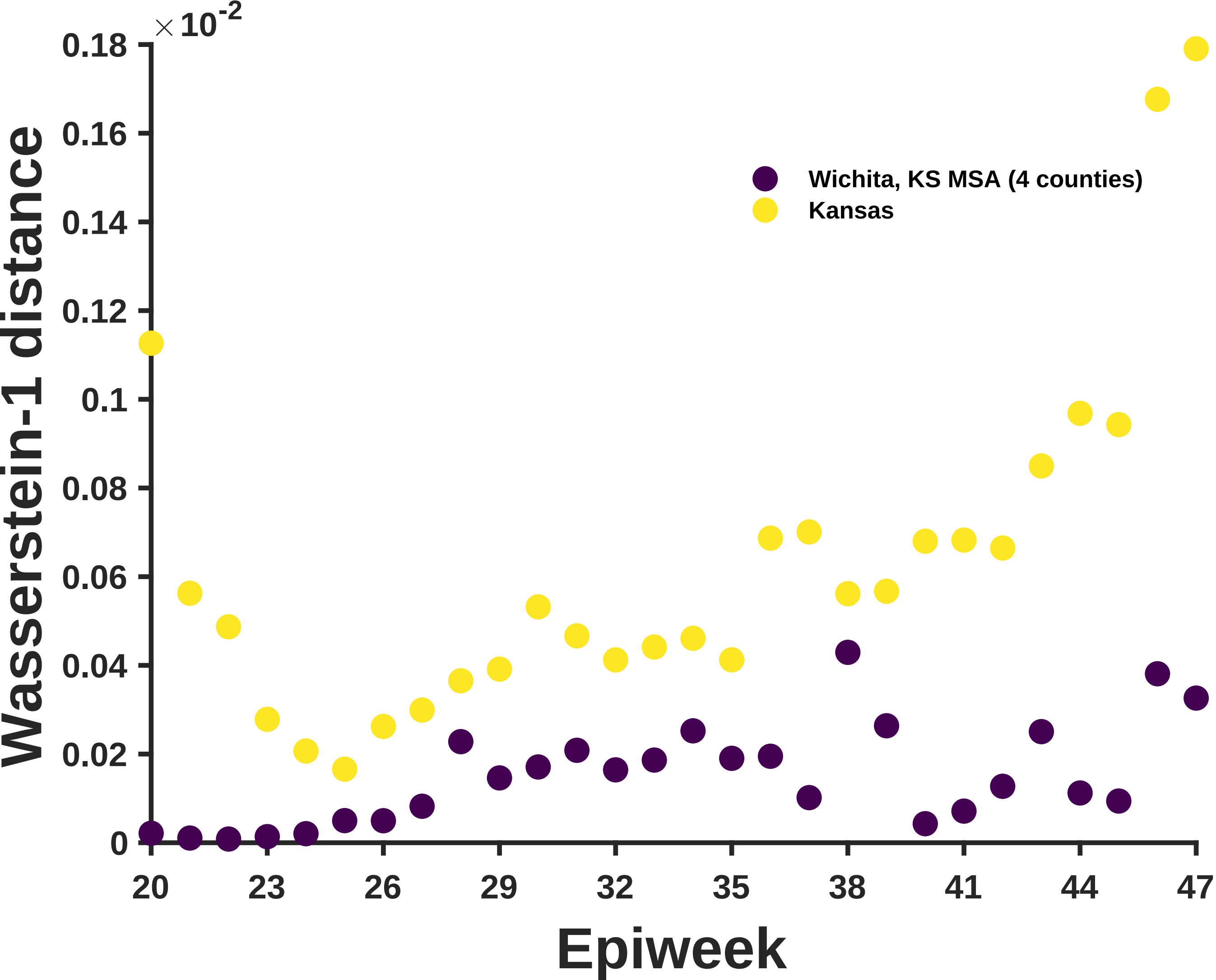

×10^-2
0.18
0.16
0.14
0.12
0.1
0.08
0.06
0.04
0.02
0
Wasserstein-1 distance
Wichita, KS MSA (4 counties)
Kansas
20
23
26
29
32
35
38
41
44
47
Epiweek

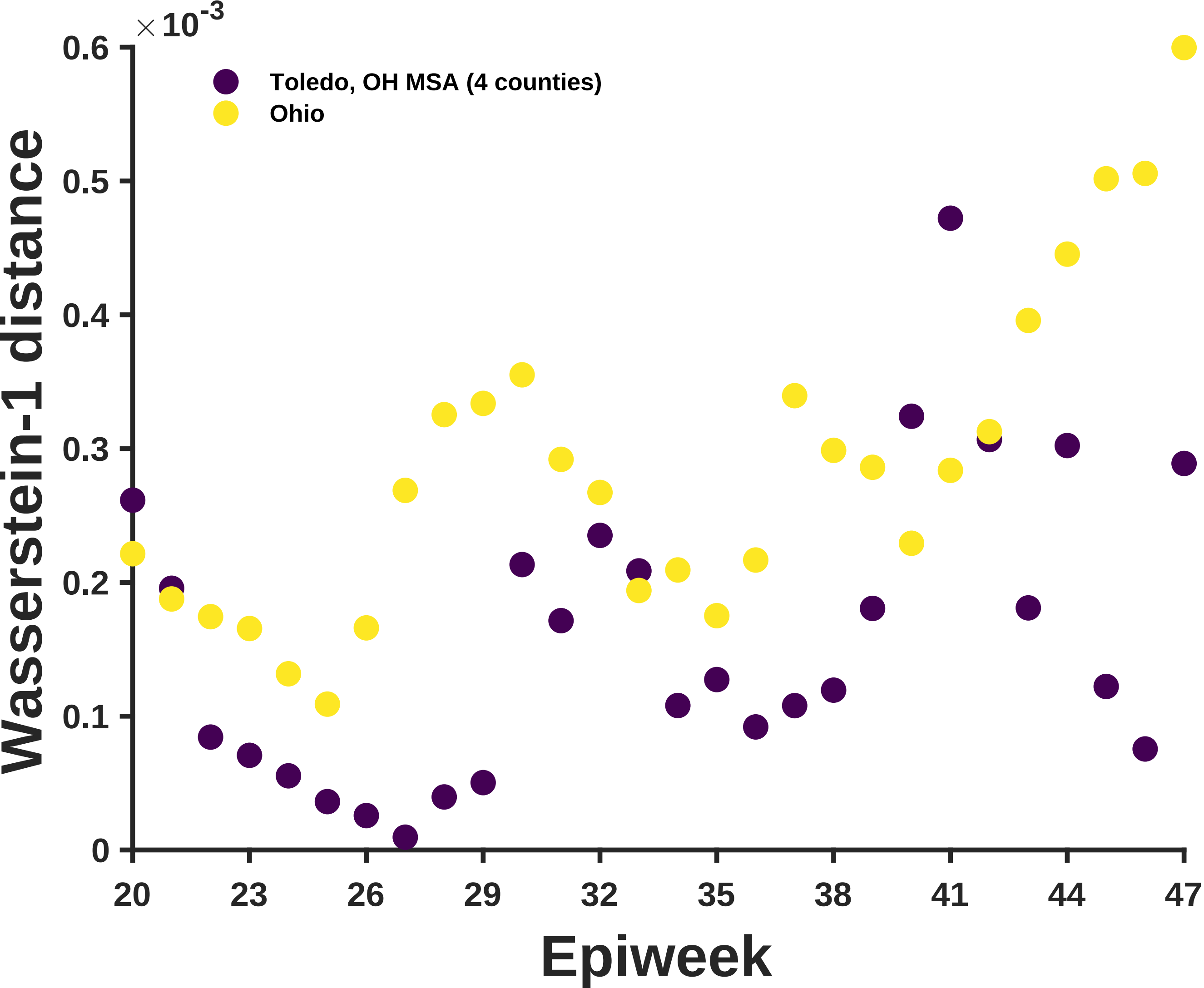

$\times 10^{-3}$
0.6
0.5
0.4
0.3
0.2
0.1
0
Toledo, OH MSA (4 counties)
Ohio
Wasserstein-1 distance
20
23
26
29
32
35
38
41
44
47
Epiweek

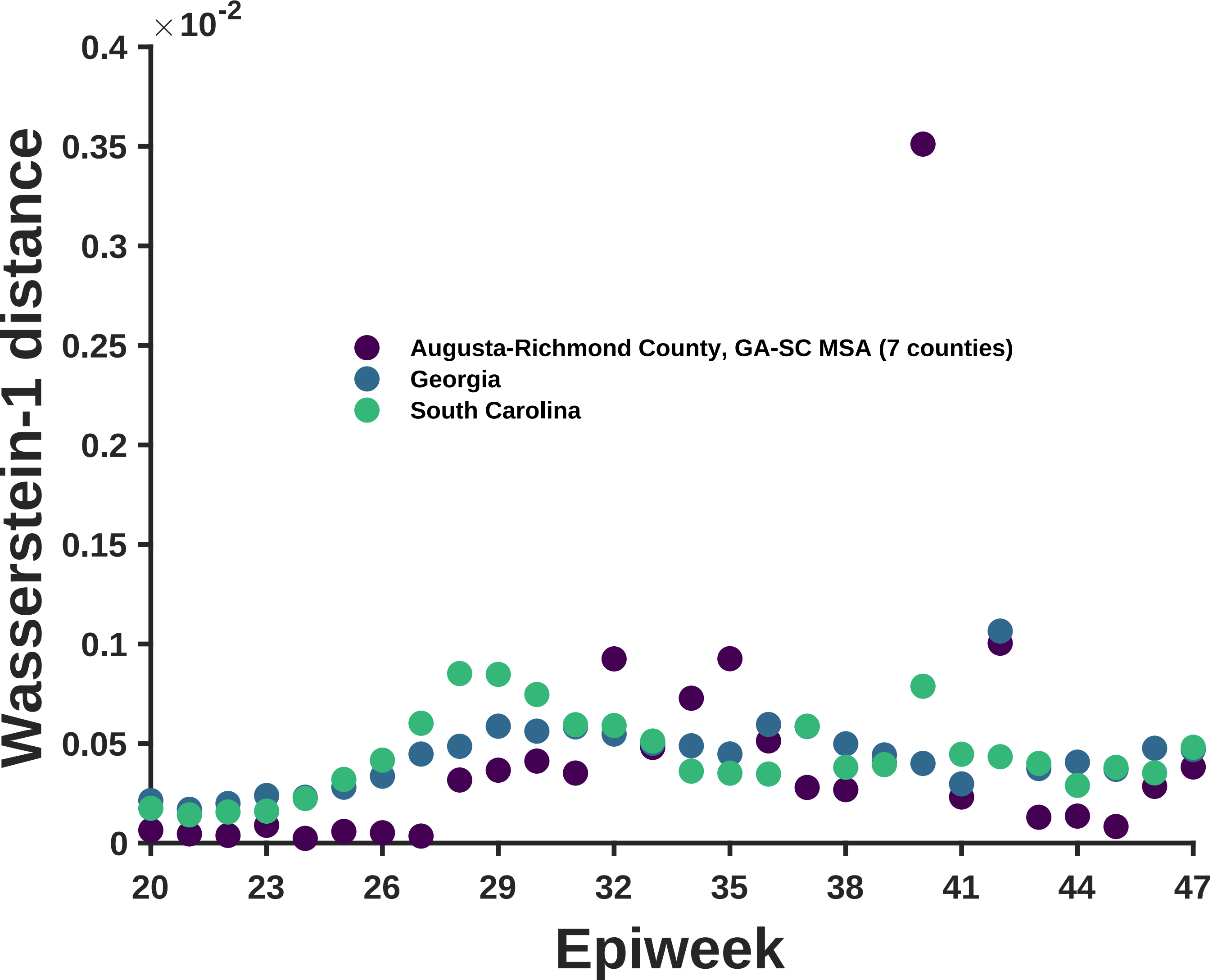

×10^-2
0.4
0.35
0.3
0.25
0.2
0.15
0.1
0.05
0
Wasserstein-1 distance
Augusta-Richmond County, GA-SC MSA (7 counties)
Georgia
South Carolina
20
23
26
29
32
35
38
41
44
47
Epiweek

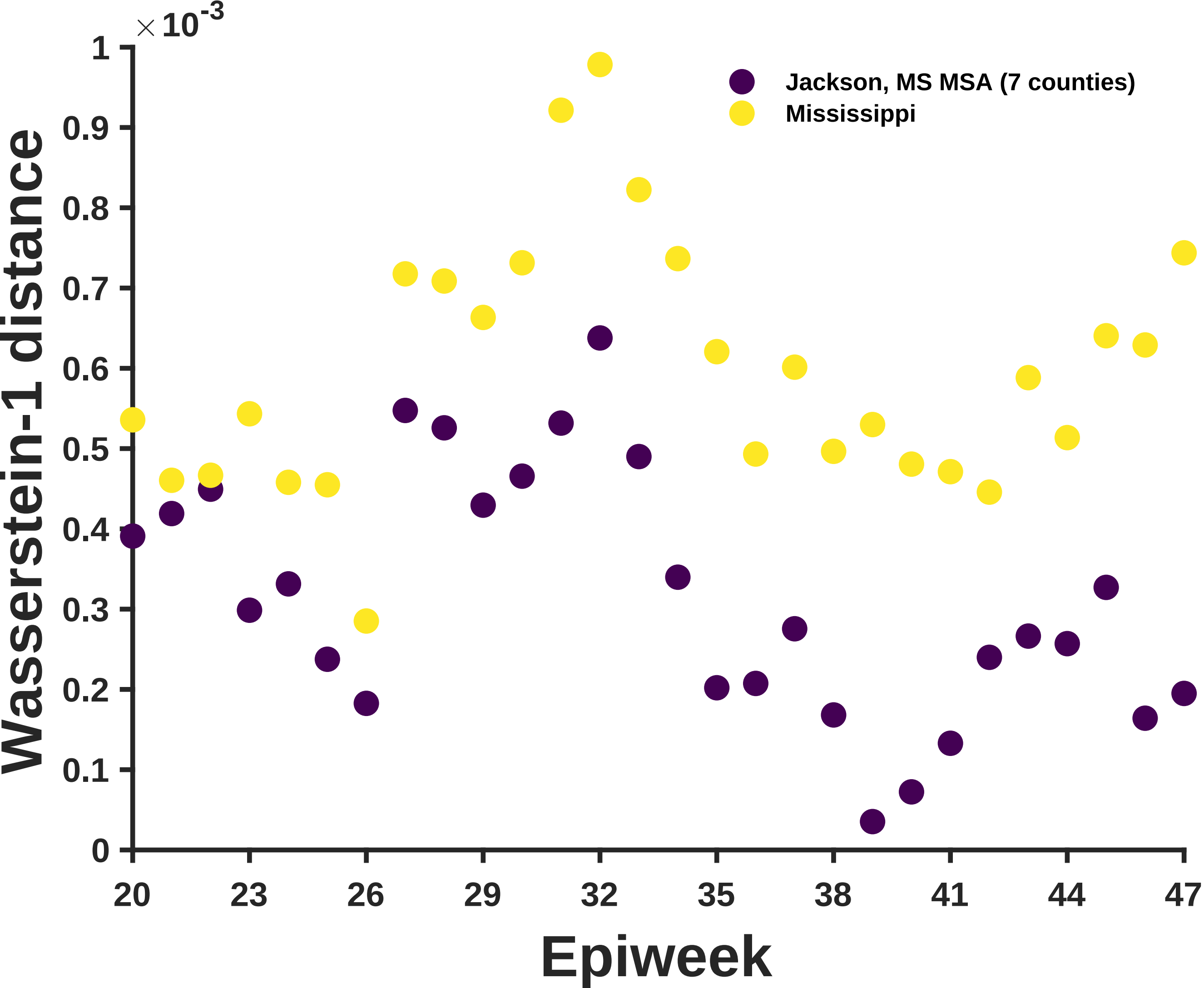

×10^-3
1
0.9
0.8
0.7
0.6
0.5
0.4
0.3
0.2
0.1
0
Wasserstein-1 distance
20
23
26
29
32
35
38
41
44
47
Epiweek
Jackson, MS MSA (7 counties)
Mississippi

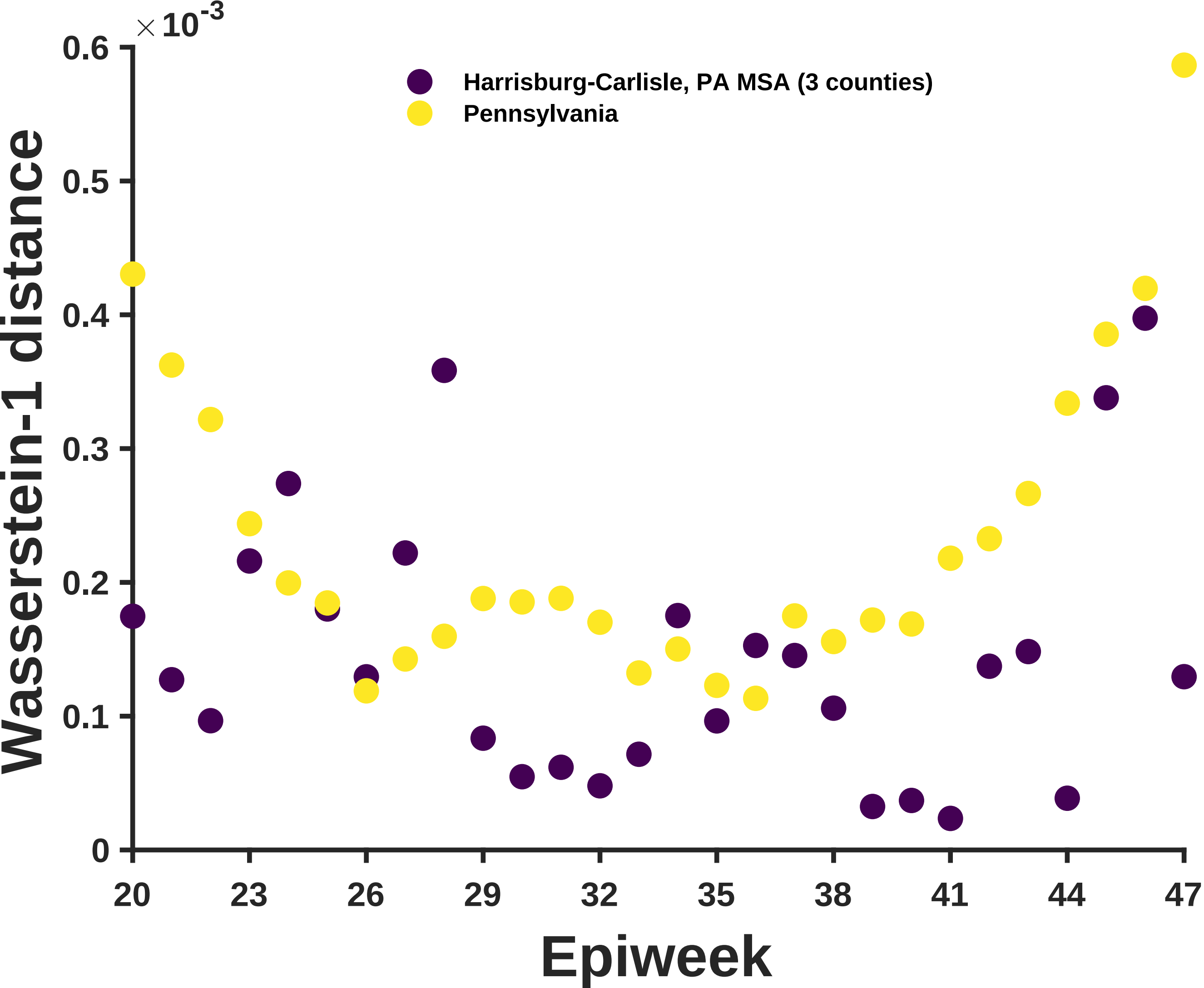

×10^-3
0.6
0.5
0.4
0.3
0.2
0.1
0
Wasserstein-1 distance
Harrisburg-Carlisle, PA MSA (3 counties)
Pennsylvania
20
23
26
29
32
35
38
41
44
47
Epiweek

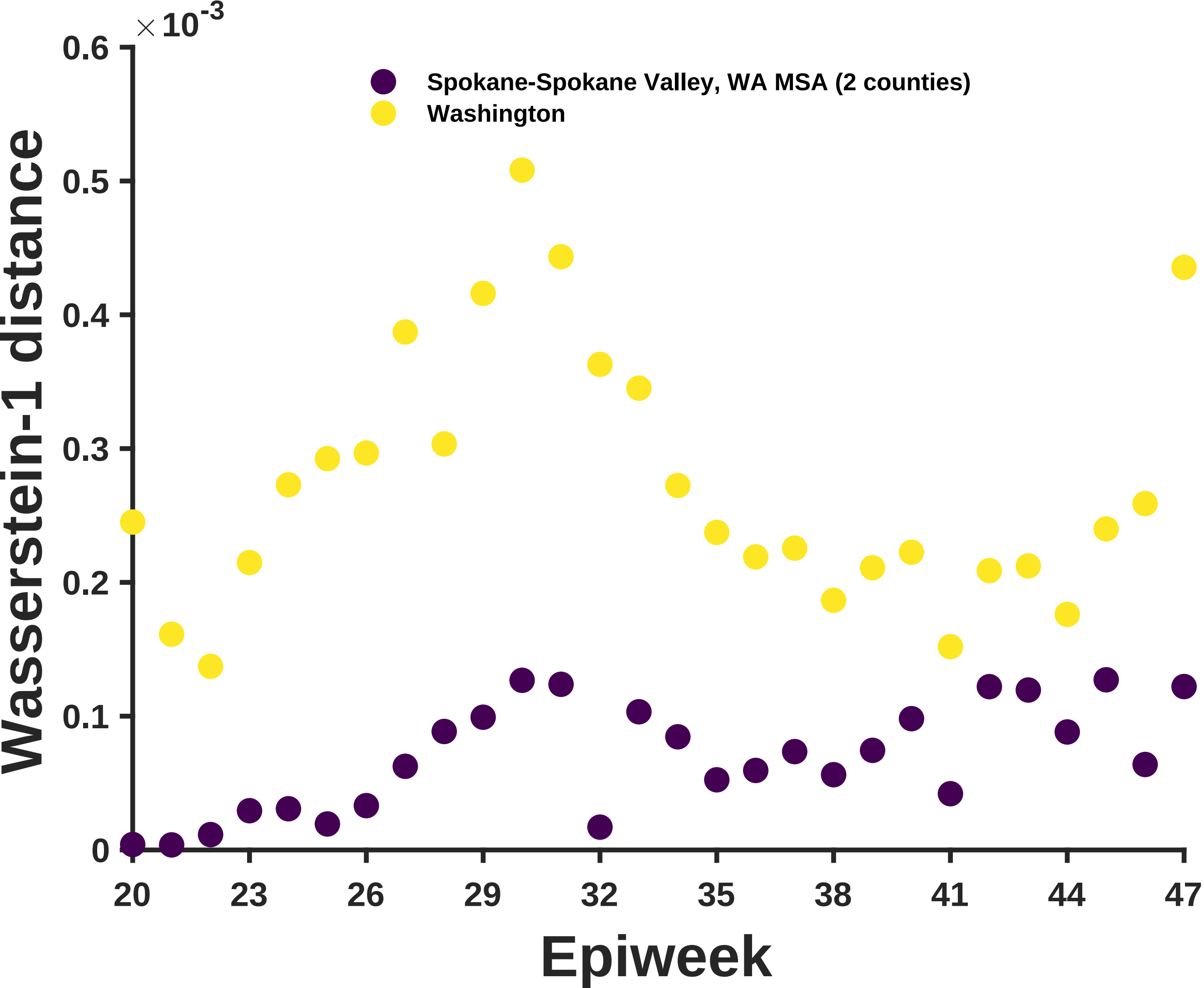

×10^-3
0.6
0.5
0.4
0.3
0.2
0.1
0
Wasserstein-1 distance
Spokane-Spokane Valley, WA MSA (2 counties)
Washington
20
23
26
29
32
35
38
41
44
47
Epiweek

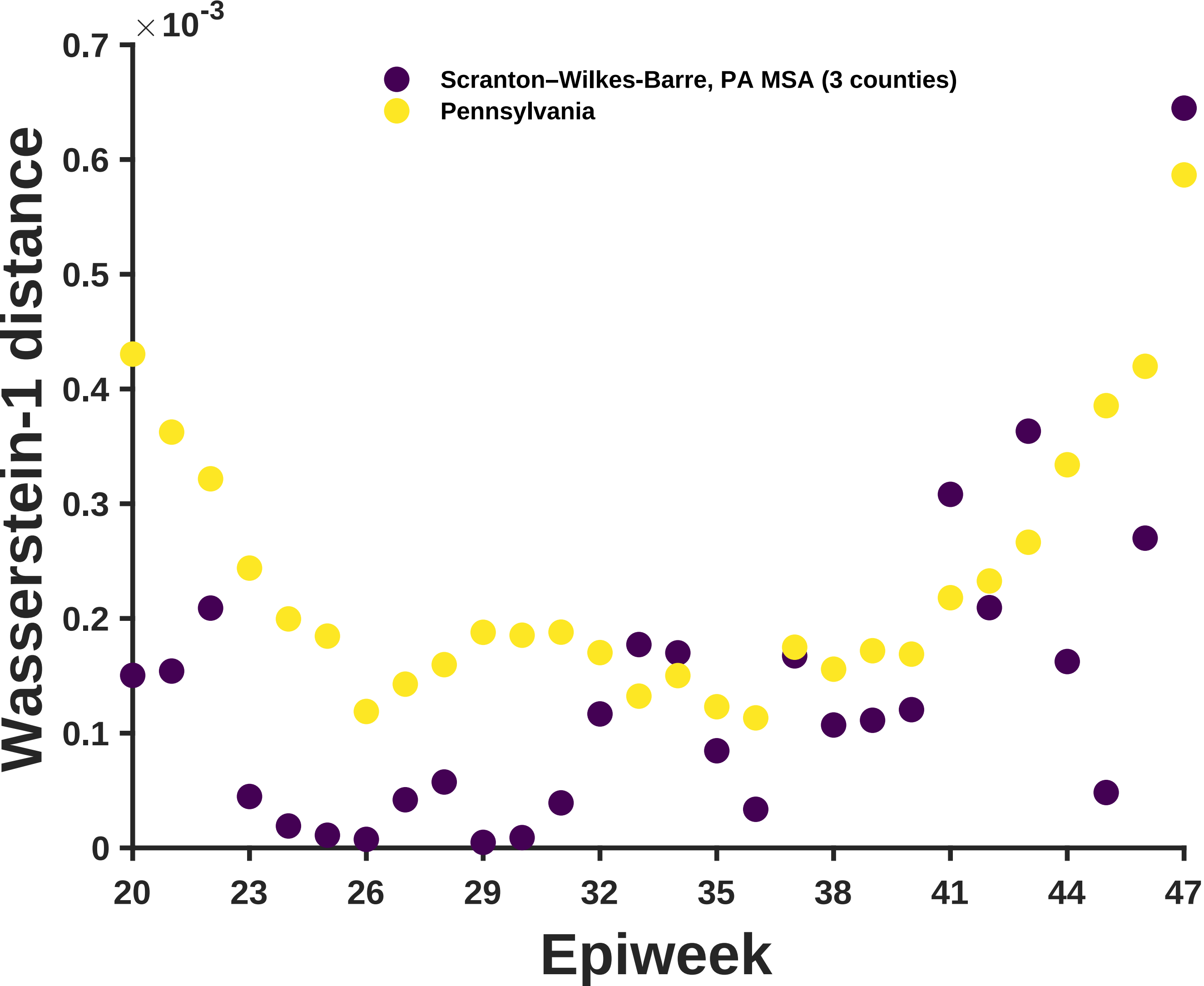

× 10^-3
0.7
0.6
0.5
0.4
0.3
0.2
0.1
0
Wasserstein-1 distance
Scranton–Wilkes-Barre, PA MSA (3 counties)
Pennsylvania
20
23
26
29
32
35
38
41
44
47
Epiweek

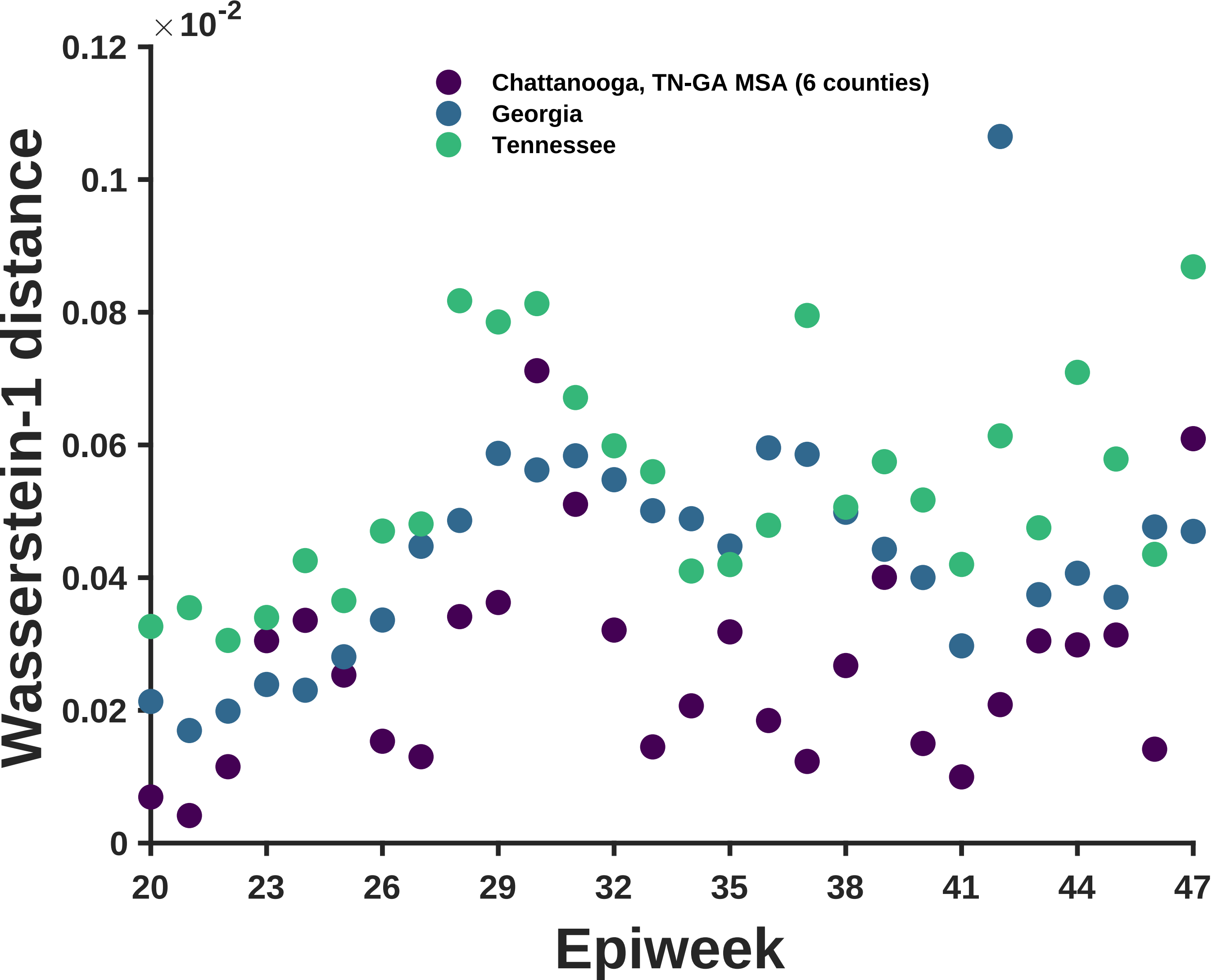

×10^-2
0.12
0.1
0.08
0.06
0.04
0.02
0
Wasserstein-1 distance
Chattanooga, TN-GA MSA (6 counties)
Georgia
Tennessee
20
23
26
29
32
35
38
41
44
47
Epiweek

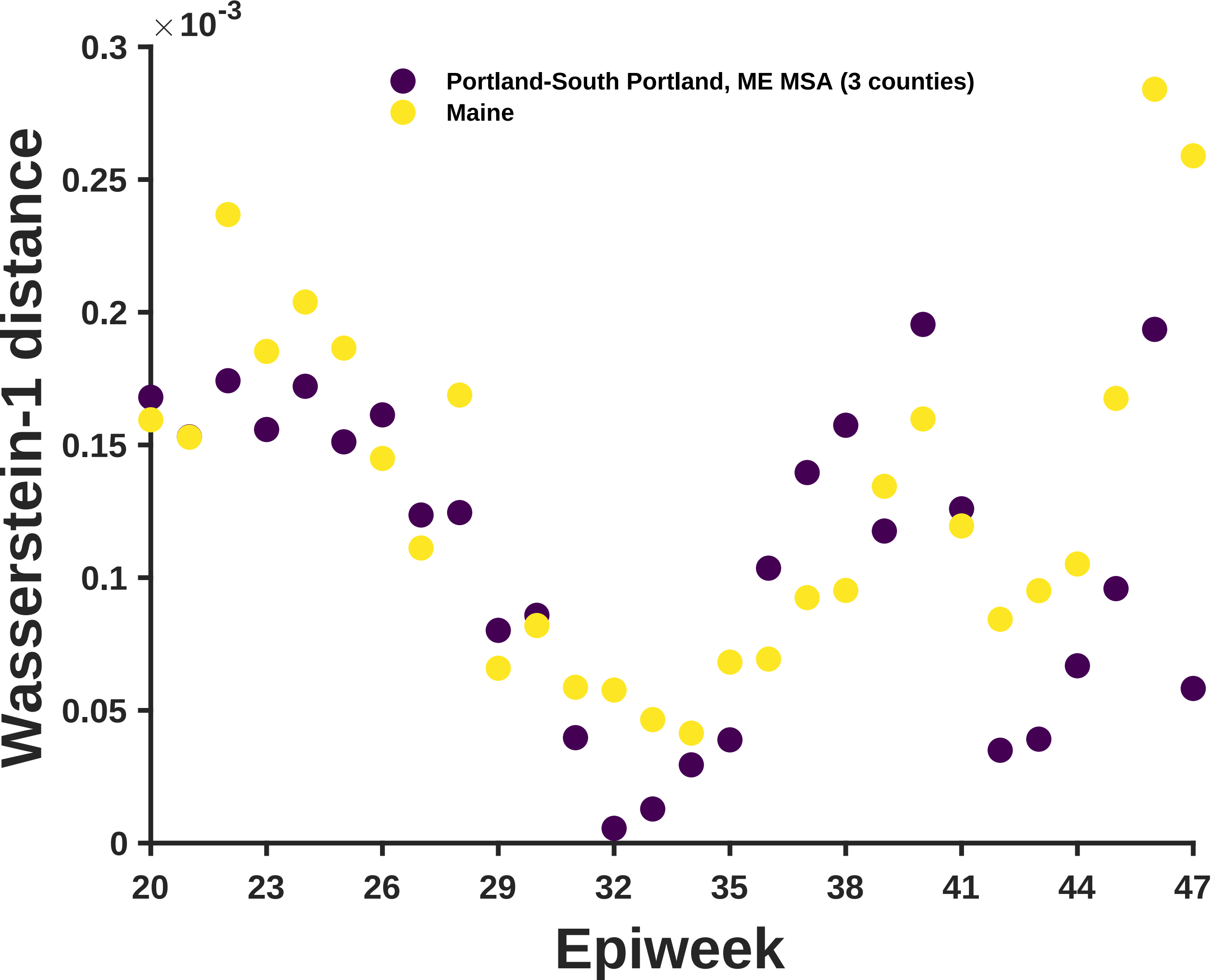

×10^-3
0.3
0.25
0.2
0.15
0.1
0.05
0
Wasserstein-1 distance
Portland-South Portland, ME MSA (3 counties)
Maine
20
23
26
29
32
35
38
41
44
47
Epiweek

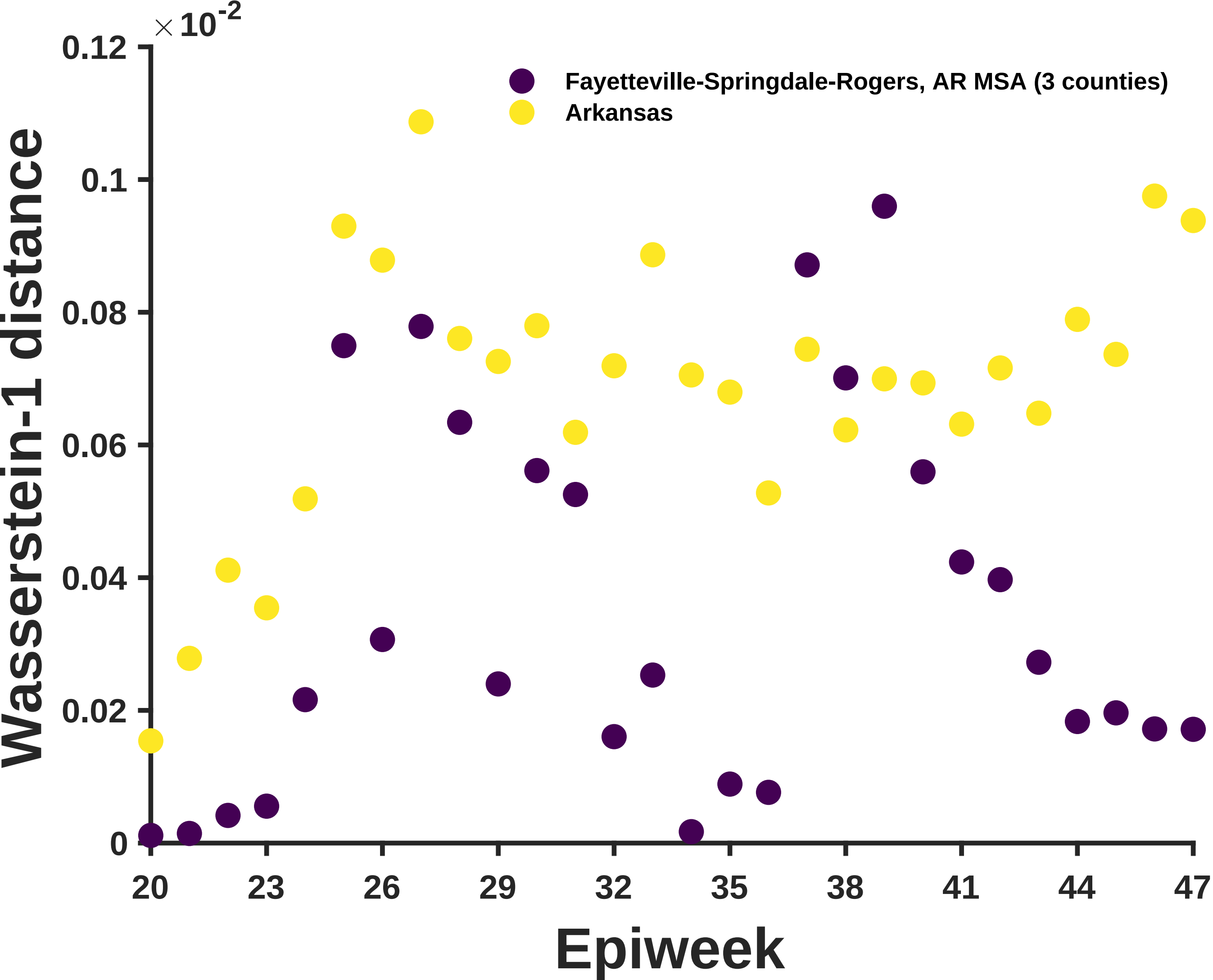

×10^-2
0.12
0.1
0.08
0.06
0.04
0.02
0
Wasserstein-1 distance
Fayetteville-Springdale-Rogers, AR MSA (3 counties)
Arkansas
20
23
26
29
32
35
38
41
44
47
Epiweek

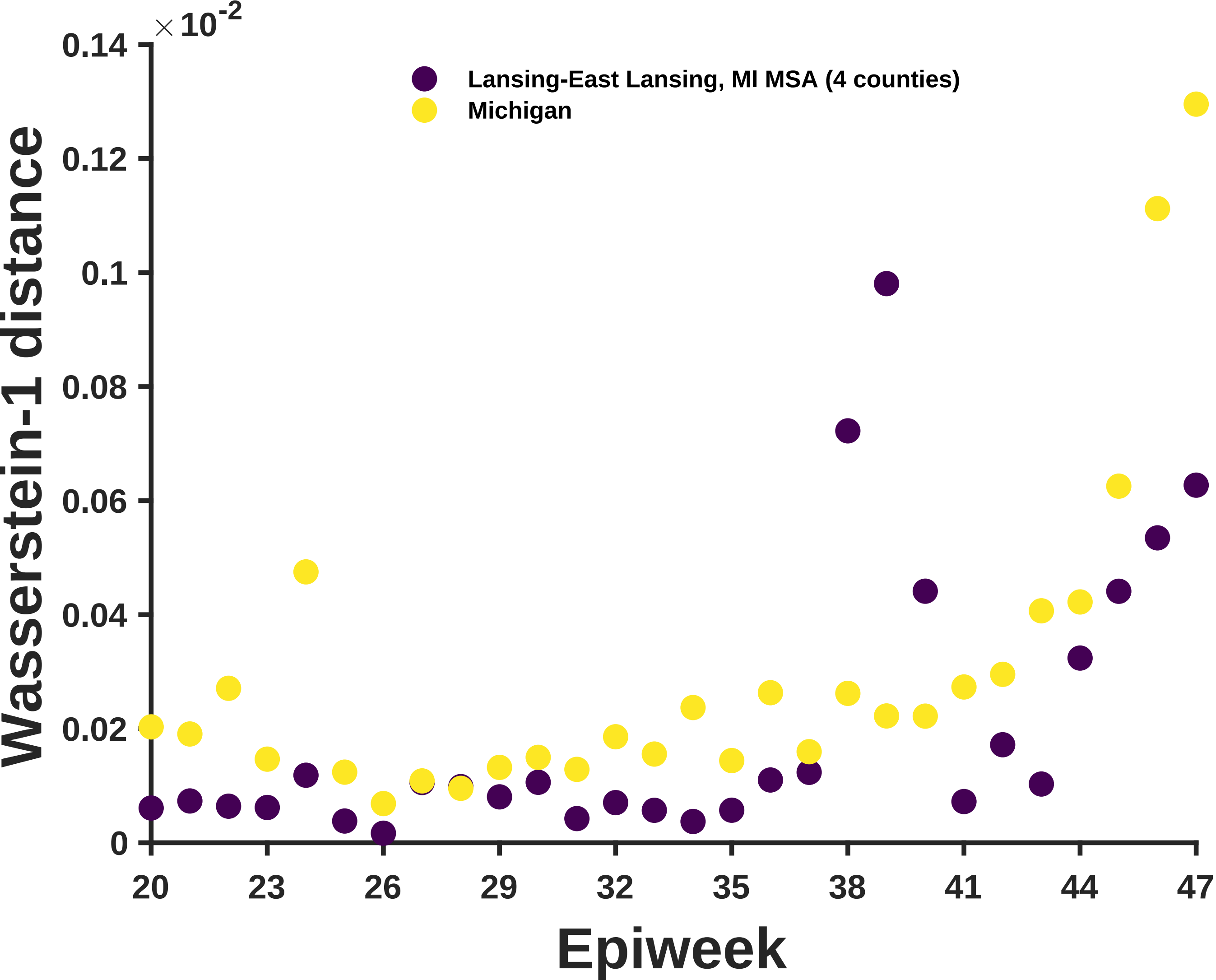

×10^-2
0.14
0.12
0.1
0.08
0.06
0.04
0.02
0
Wasserstein-1 distance
Lansing-East Lansing, MI MSA (4 counties)
Michigan
20
23
26
29
32
35
38
41
44
47
Epiweek

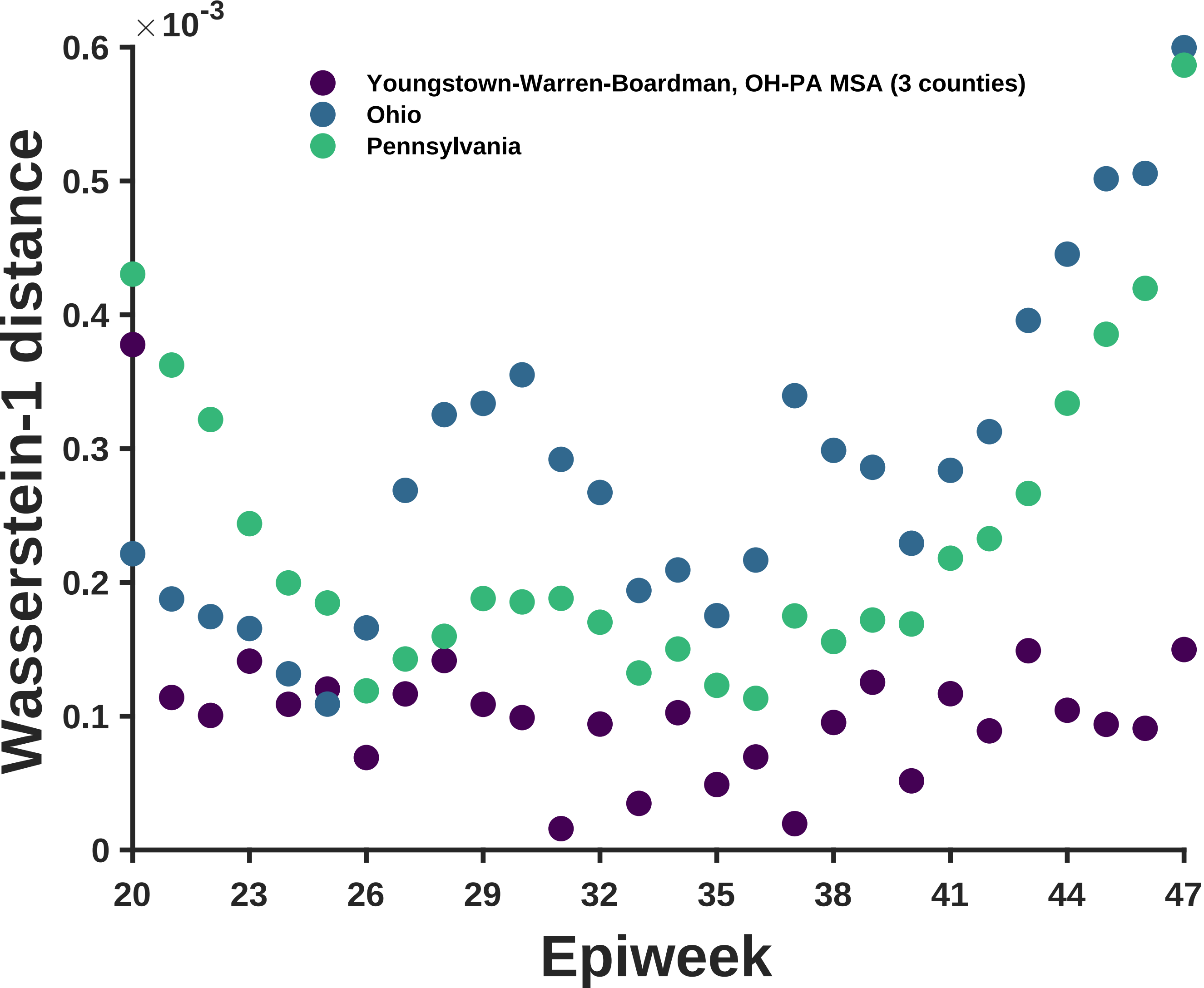

×10-3
0.6
0.5
0.4
0.3
0.2
0.1
0
Wasserstein-1 distance
Youngstown-Warren-Boardman, OH-PA MSA (3 counties)
Ohio
Pennsylvania
20
23
26
29
32
35
38
41
44
47
Epiweek

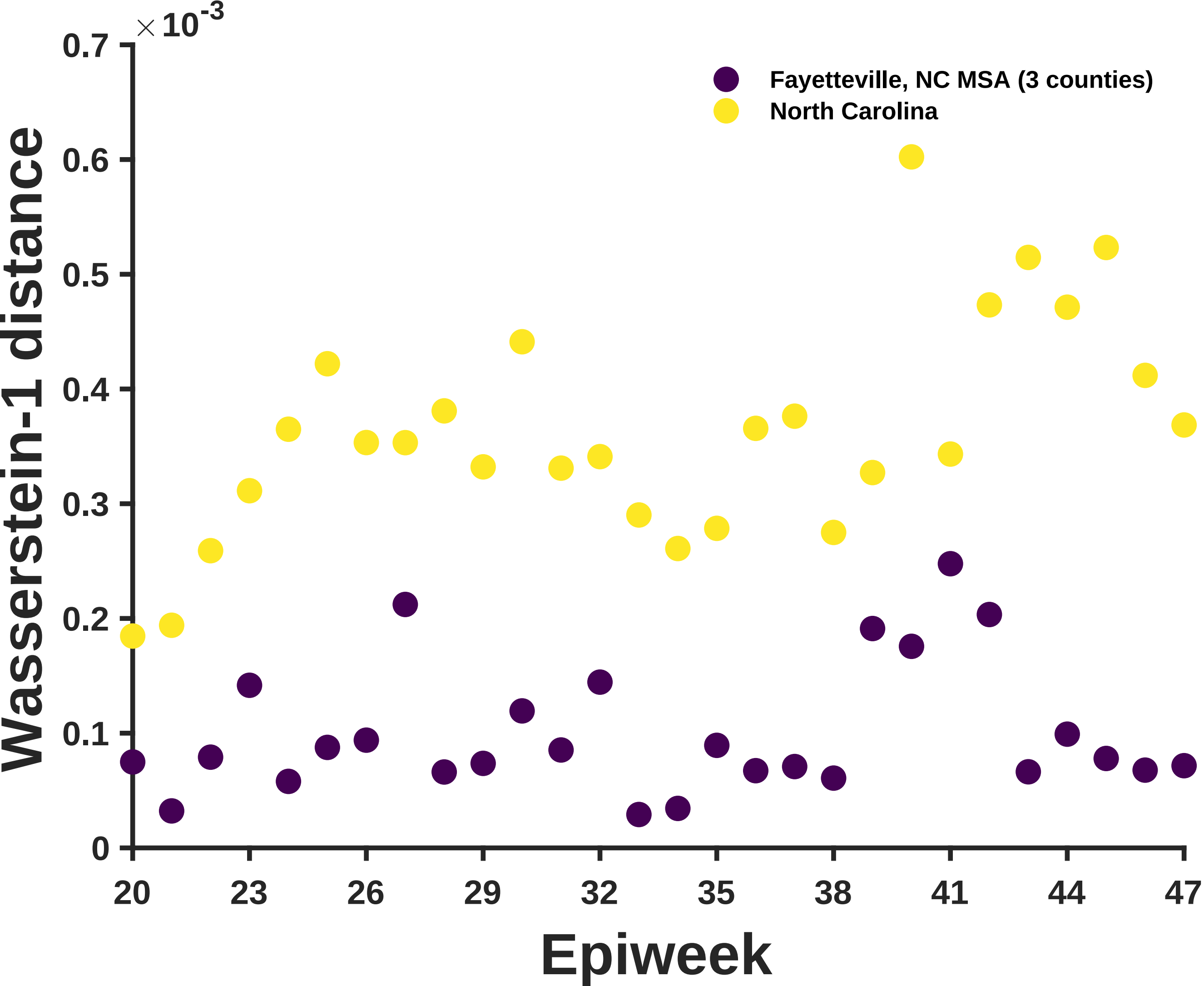

$\times 10^{-3}$
0.7
0.6
0.5
0.4
0.3
0.2
0.1
0
Wasserstein-1 distance
Fayetteville, NC MSA (3 counties)
North Carolina
20
23
26
29
32
35
38
41
44
47
Epiweek

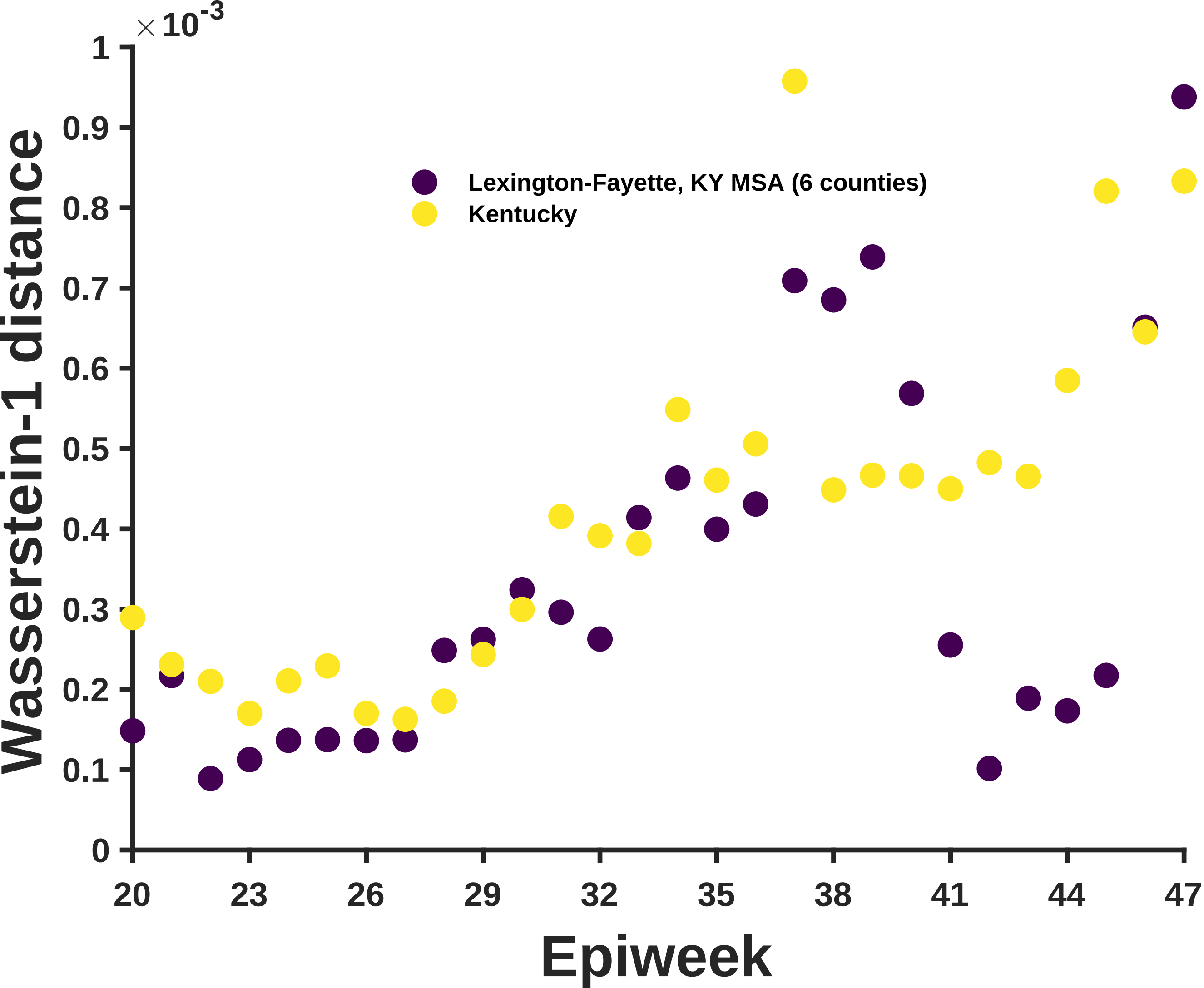

×10^-3
1
0.9
0.8
0.7
0.6
0.5
0.4
0.3
0.2
0.1
0
Wasserstein-1 distance
Lexington-Fayette, KY MSA (6 counties)
Kentucky
20
23
26
29
32
35
38
41
44
47
Epiweek

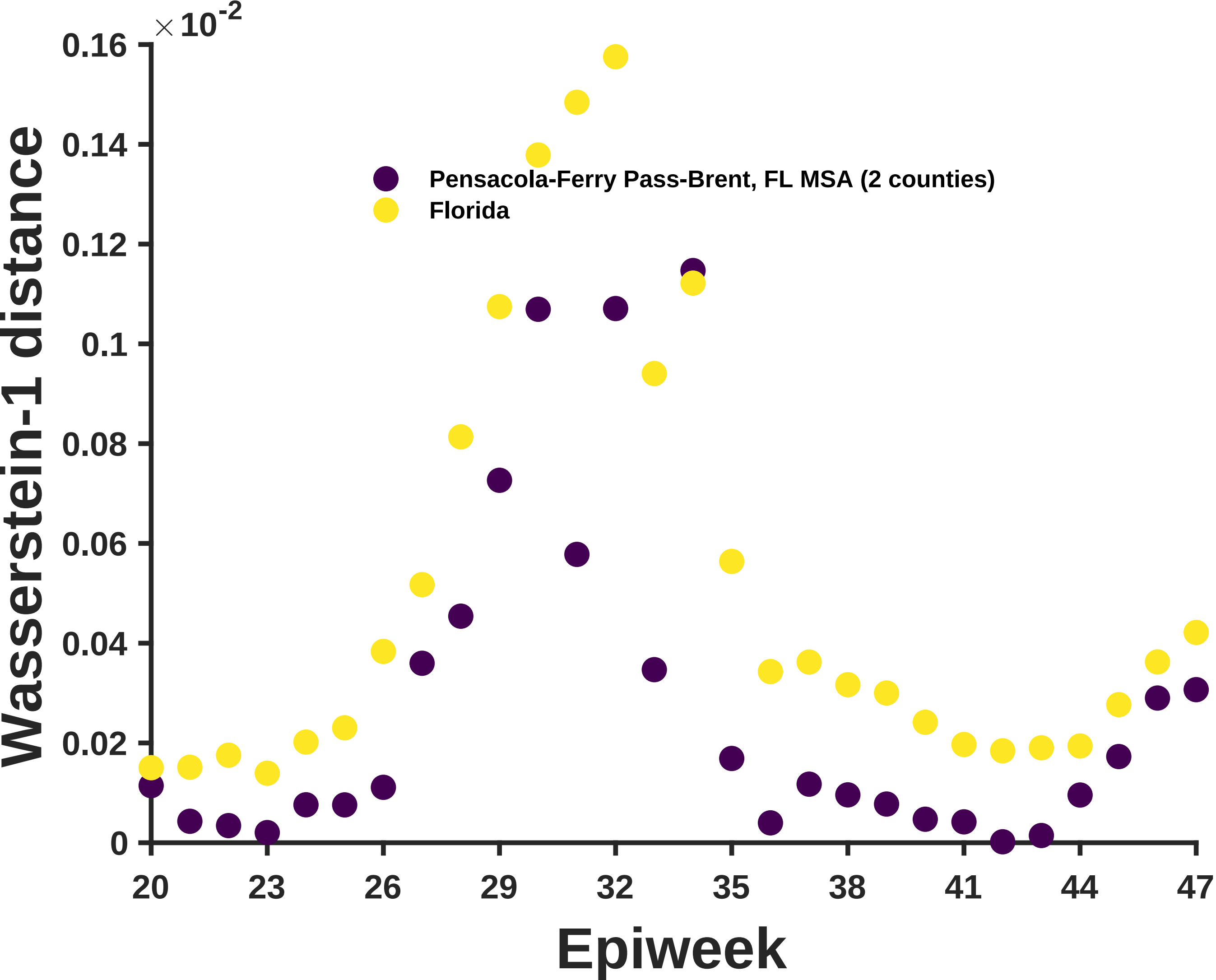

×10^-2
0.16
0.14
0.12
0.1
0.08
0.06
0.04
0.02
0
Wasserstein-1 distance
Pensacola-Ferry Pass-Brent, FL MSA (2 counties)
Florida
20
23
26
29
32
35
38
41
44
47
Epiweek

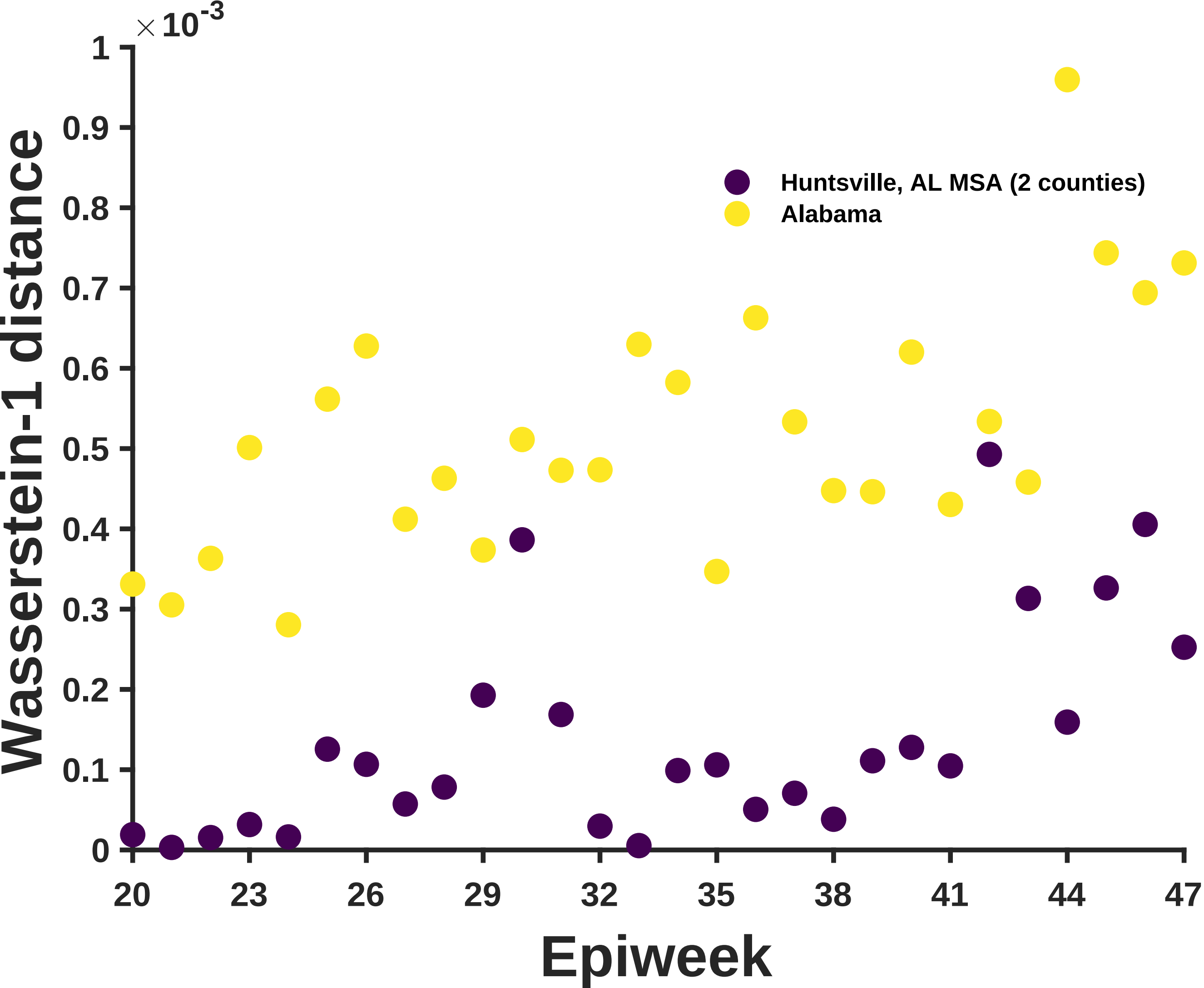

×10^-3
Wasserstein-1 distance
0
0.1
0.2
0.3
0.4
0.5
0.6
0.7
0.8
0.9
1
20
23
26
29
32
35
38
41
44
47
Epiweek
Huntsville, AL MSA (2 counties)
Alabama

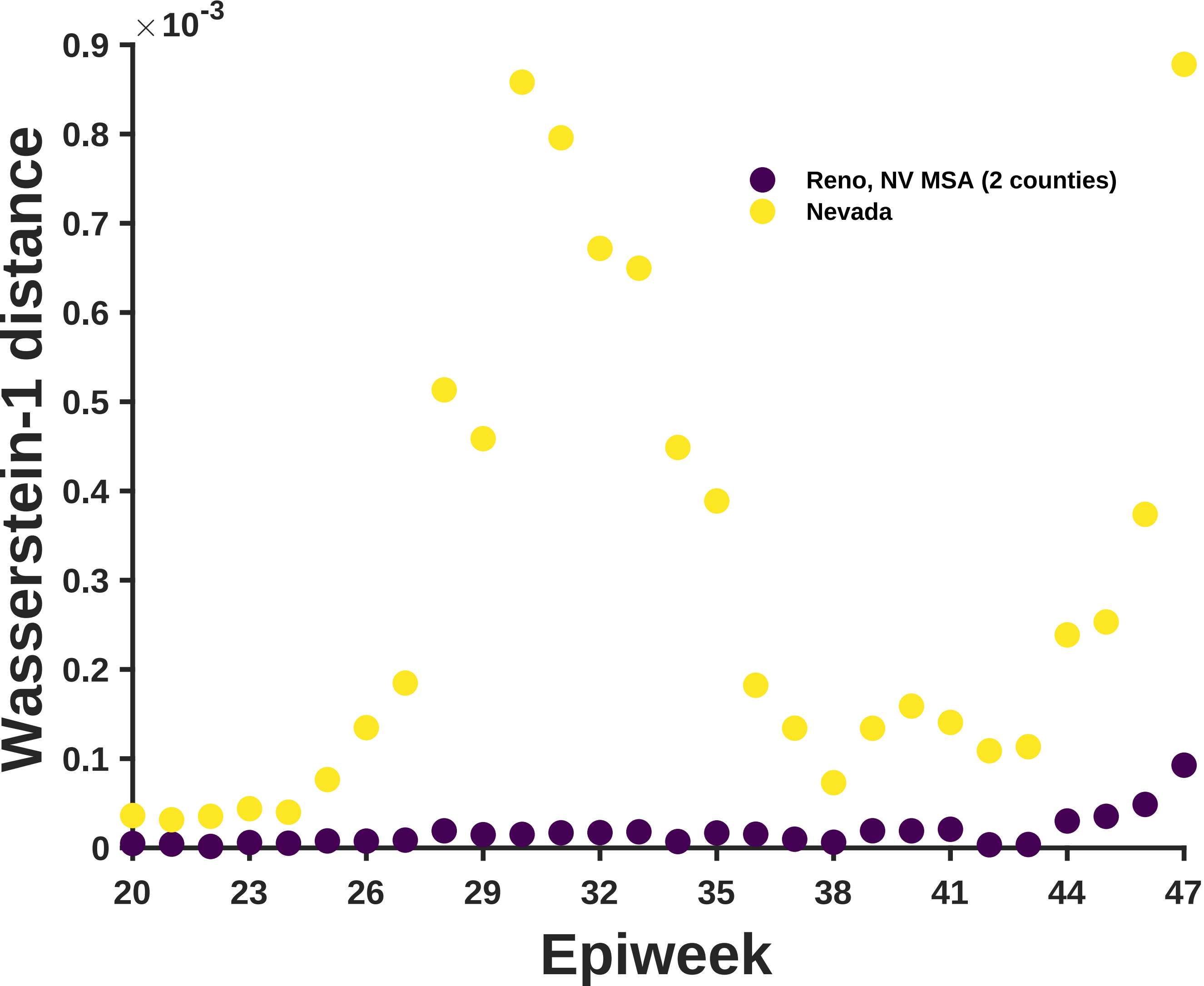

×10-3
0.9
0.8
0.7
0.6
0.5
0.4
0.3
0.2
0.1
0
Wasserstein-1 distance
Reno, NV MSA (2 counties)
Nevada
20
23
26
29
32
35
38
41
44
47
Epiweek

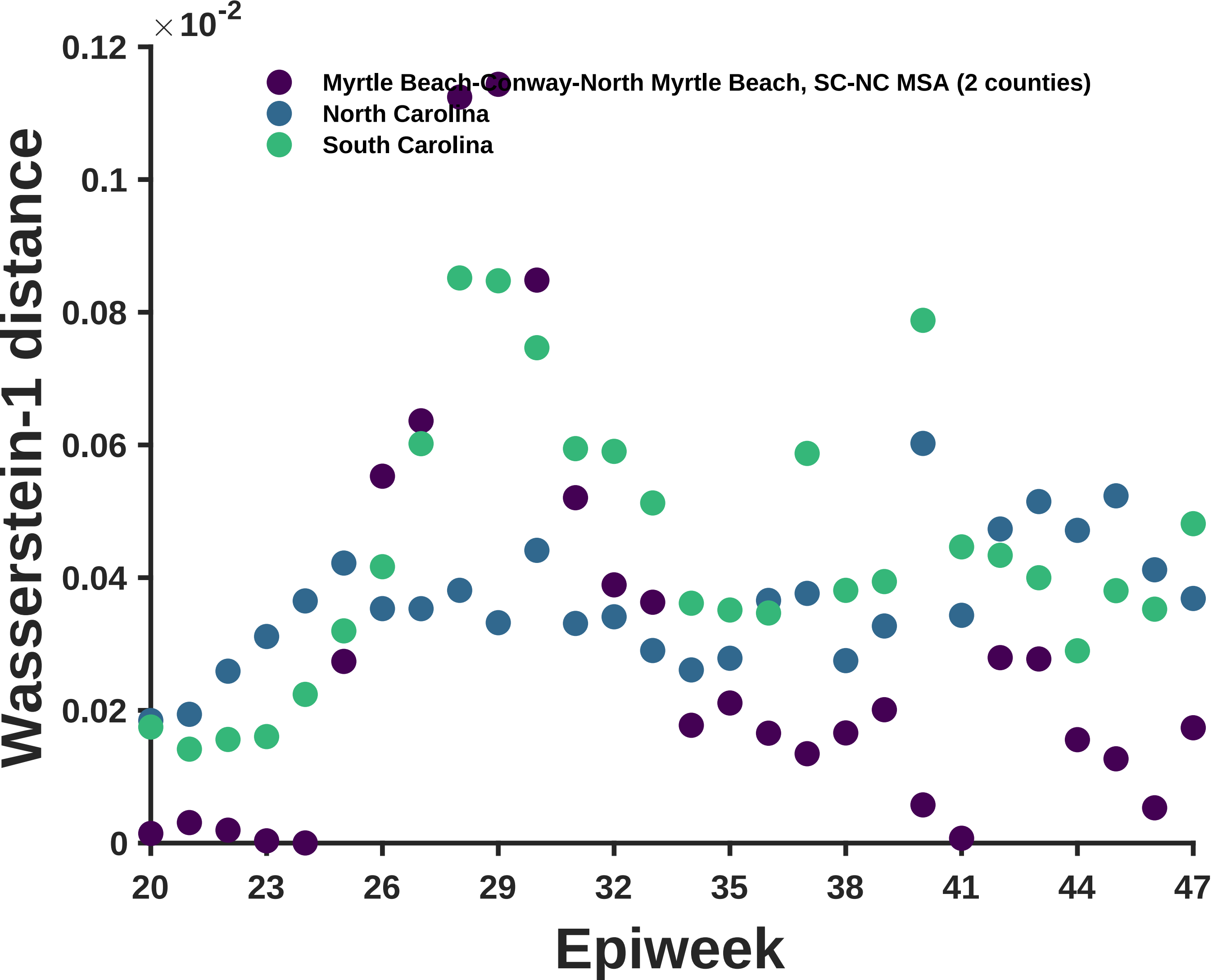

×10^-2
0.12
0.1
0.08
0.06
0.04
0.02
0
Wasserstein-1 distance
Myrtle Beach-Conway-North Myrtle Beach, SC-NC MSA (2 counties)
North Carolina
South Carolina
20
23
26
29
32
35
38
41
44
47
Epiweek

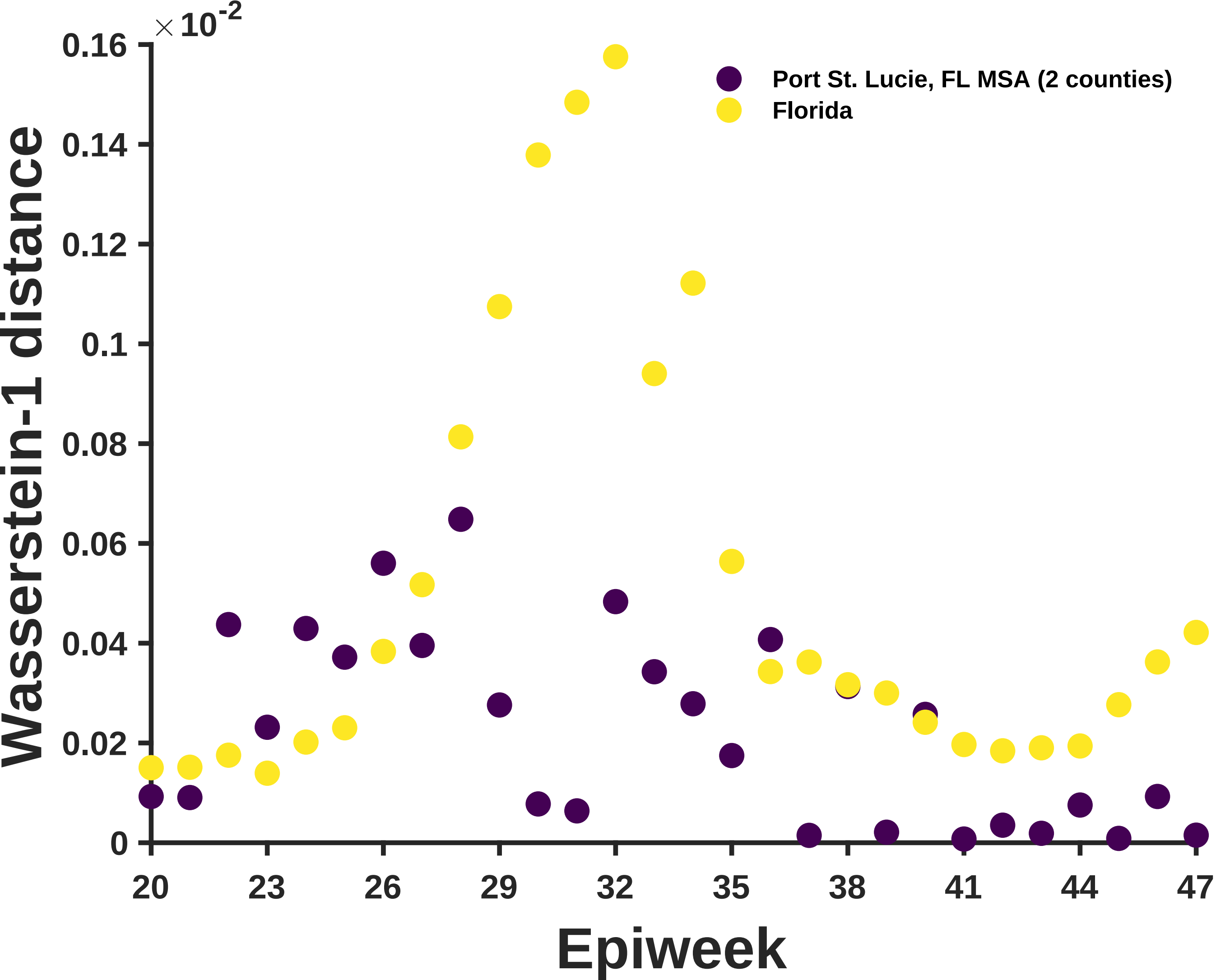

$\times 10^{-2}$
0.16
0.14
0.12
0.1
0.08
0.06
0.04
0.02
0
Wasserstein-1 distance
20
23
26
29
32
35
38
41
44
47
Epiweek
Port St. Lucie, FL MSA (2 counties)
Florida

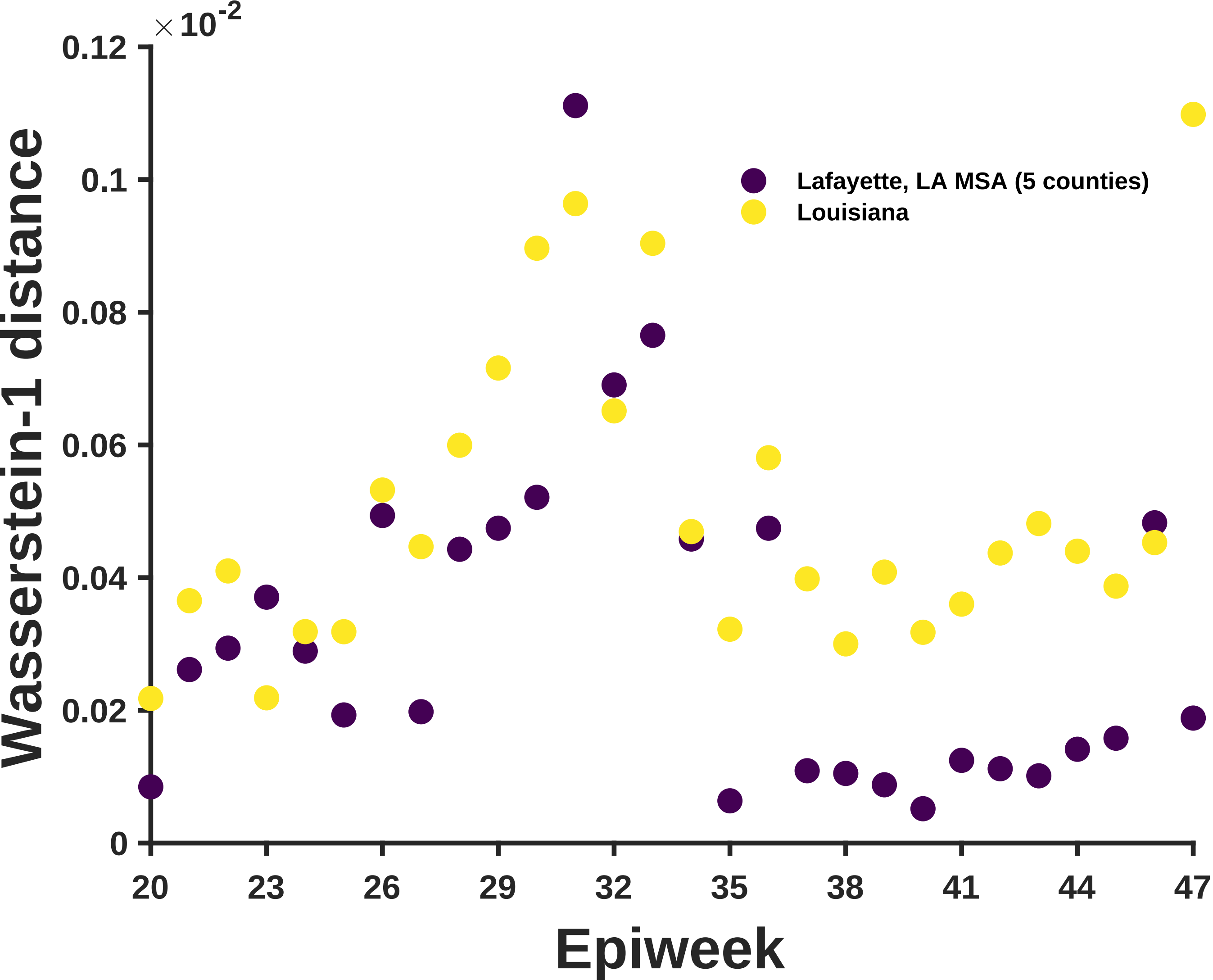
×10^-2
0.12
0.1
0.08
0.06
0.04
0.02
0
Wasserstein-1 distance
Lafayette, LA MSA (5 counties)
Louisiana
20
23
26
29
32
35
38
41
44
47
Epiweek

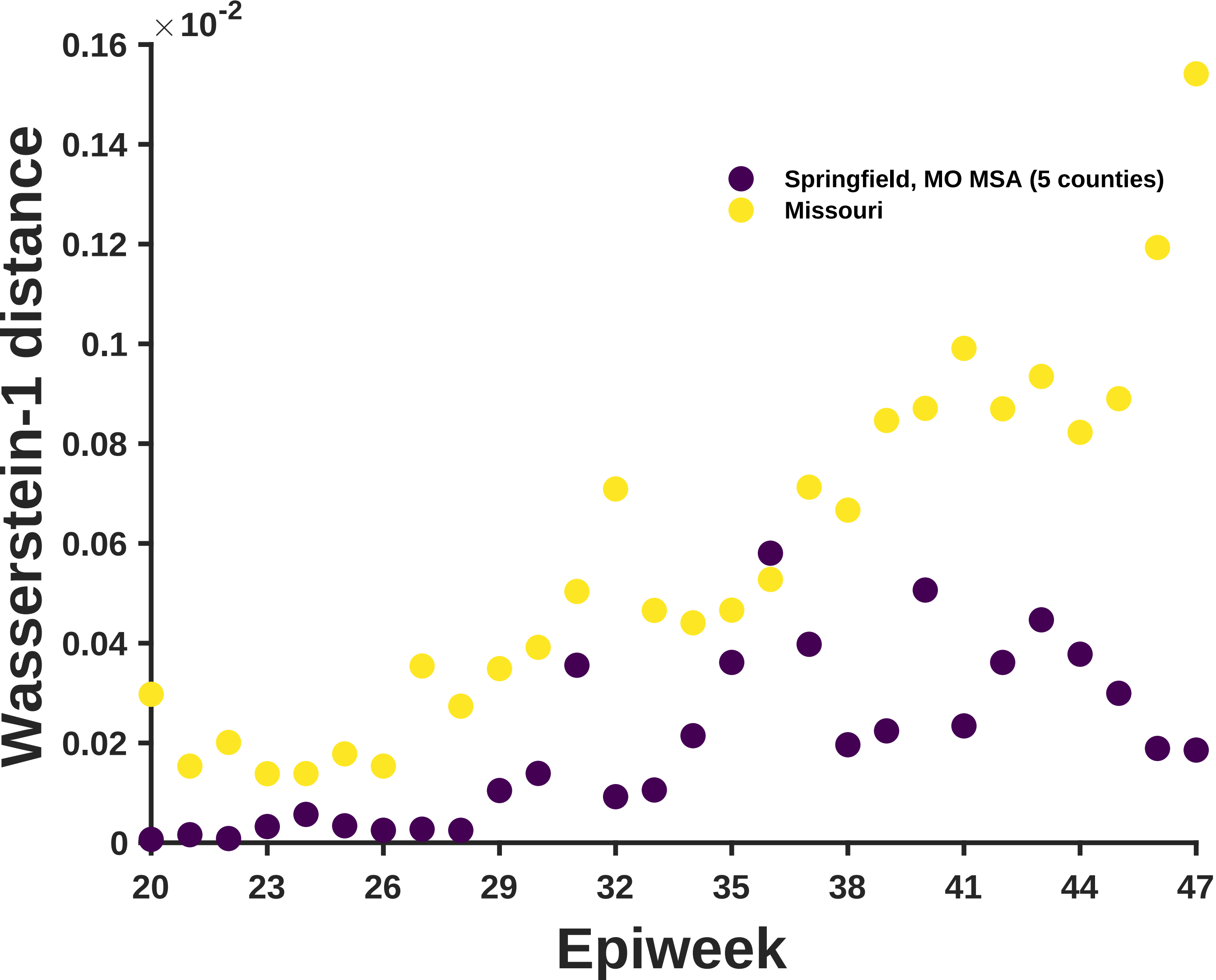

×10^-2
0.16
0.14
0.12
0.1
0.08
0.06
0.04
0.02
0
Wasserstein-1 distance
Springfield, MO MSA (5 counties)
Missouri
20
23
26
29
32
35
38
41
44
47
Epiweek

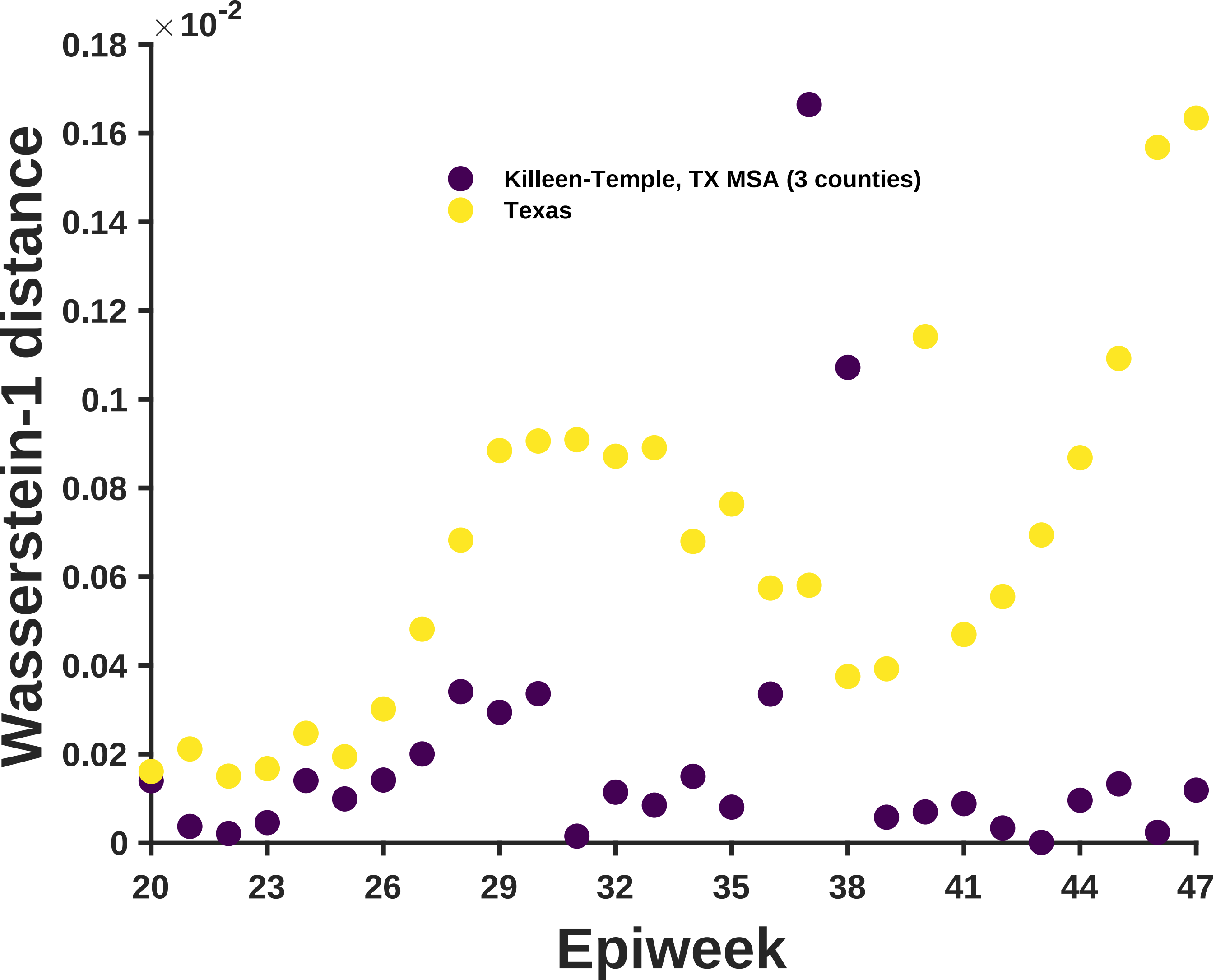

×10^-2
0.18
0.16
0.14
0.12
0.1
0.08
0.06
0.04
0.02
0
Wasserstein-1 distance
Killeen-Temple, TX MSA (3 counties)
Texas
20
23
26
29
32
35
38
41
44
47
Epiweek

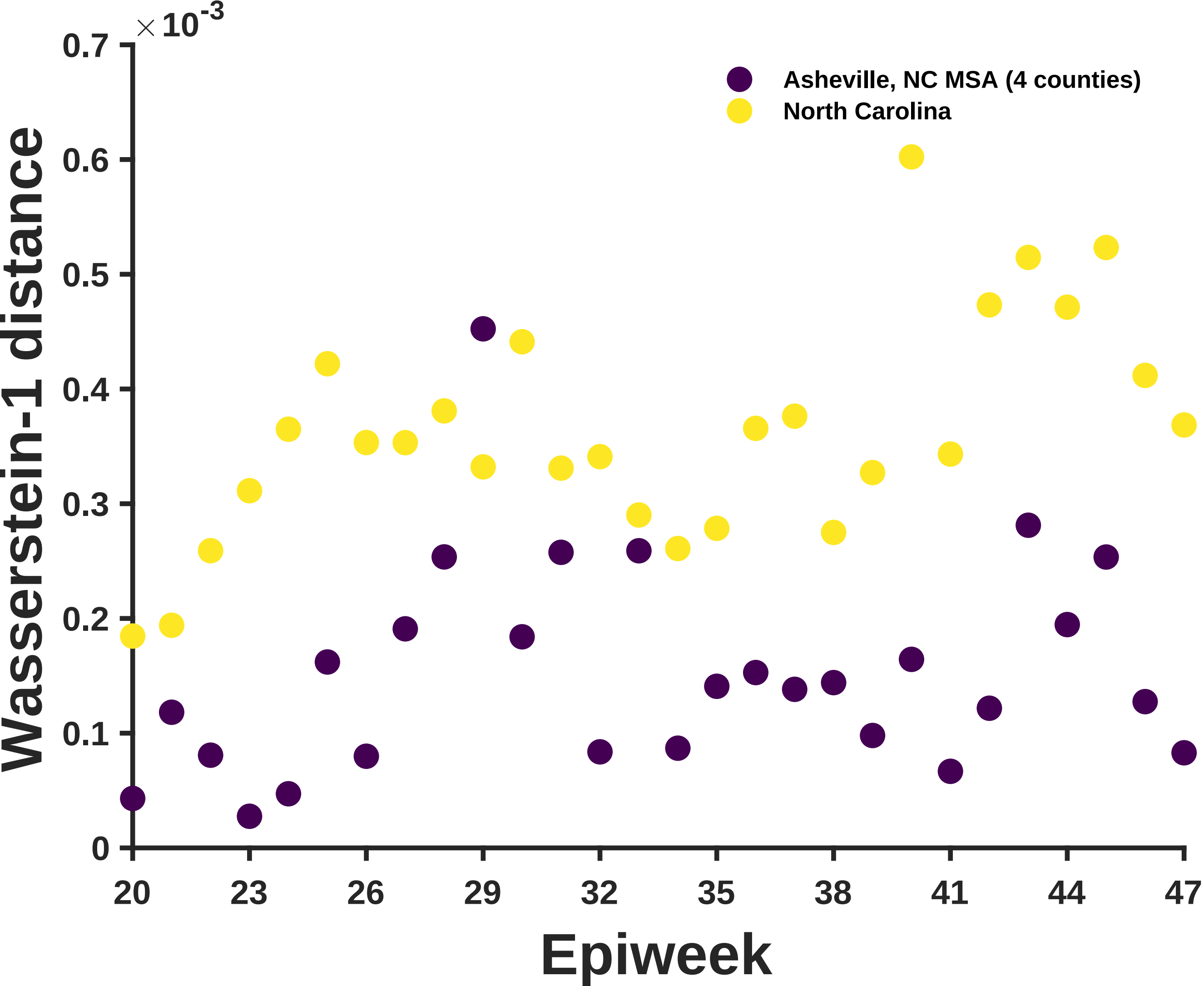

×10^-3
0.7
0.6
0.5
0.4
0.3
0.2
0.1
0
Wasserstein-1 distance
Asheville, NC MSA (4 counties)
North Carolina
20
23
26
29
32
35
38
41
44
47
Epiweek

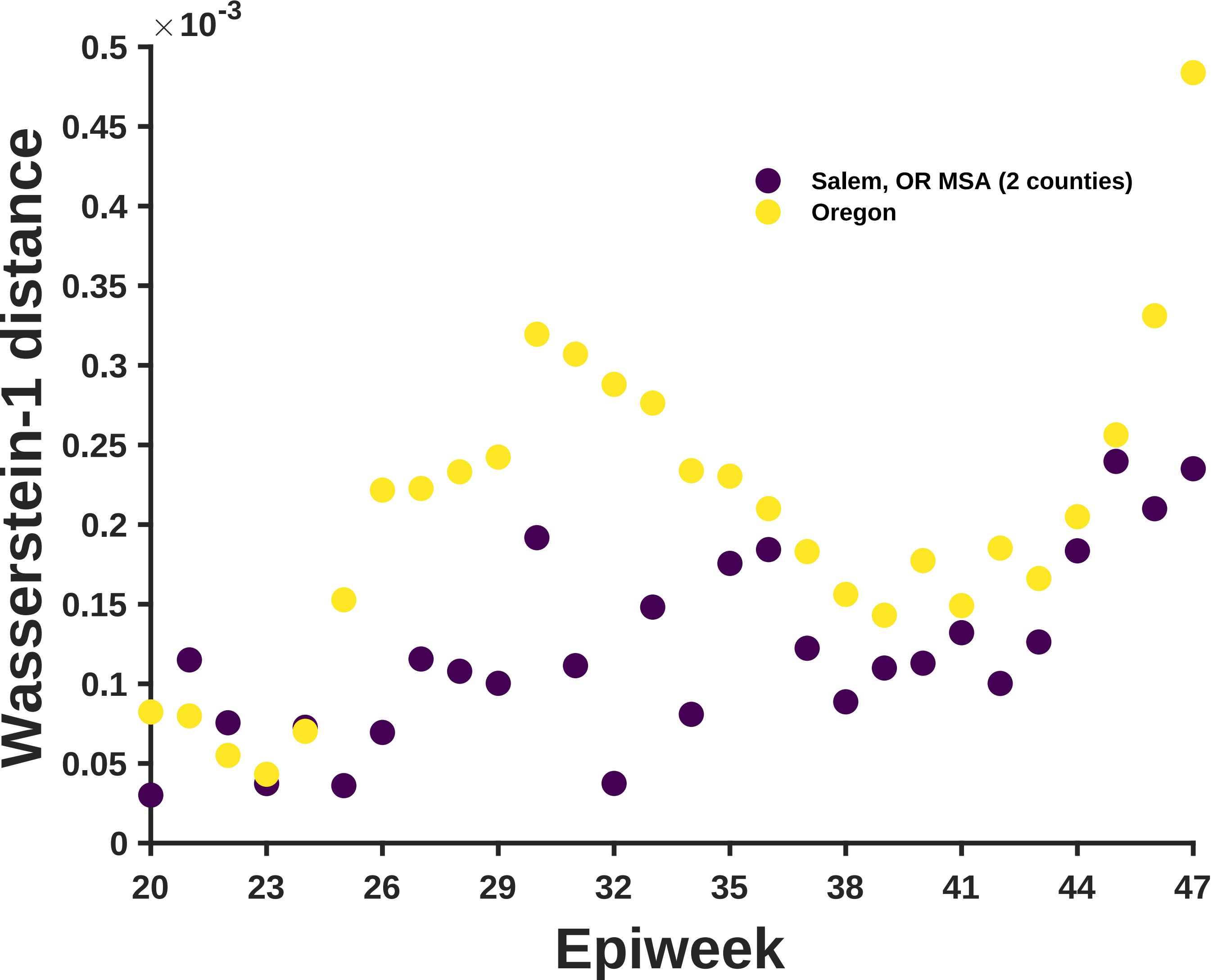

×10^-3
0.5
0.45
0.4
0.35
0.3
0.25
0.2
0.15
0.1
0.05
0
Wasserstein-1 distance
Salem, OR MSA (2 counties)
Oregon
20
23
26
29
32
35
38
41
44
47
Epiweek

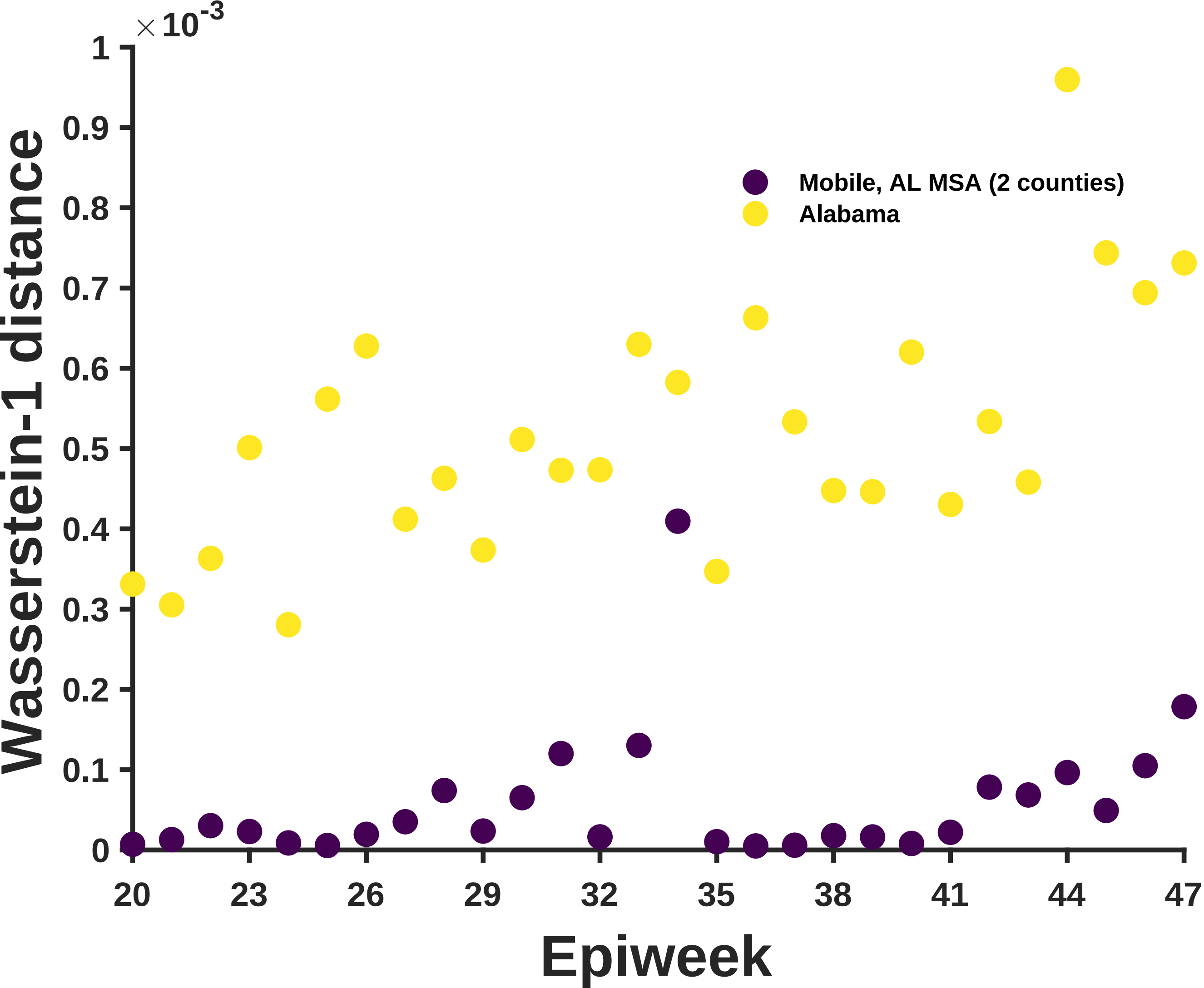

×10^-3
Wasserstein-1 distance
Epiweek
Mobile, AL MSA (2 counties)
Alabama
0
0.1
0.2
0.3
0.4
0.5
0.6
0.7
0.8
0.9
1
20
23
26
29
32
35
38
41
44
47

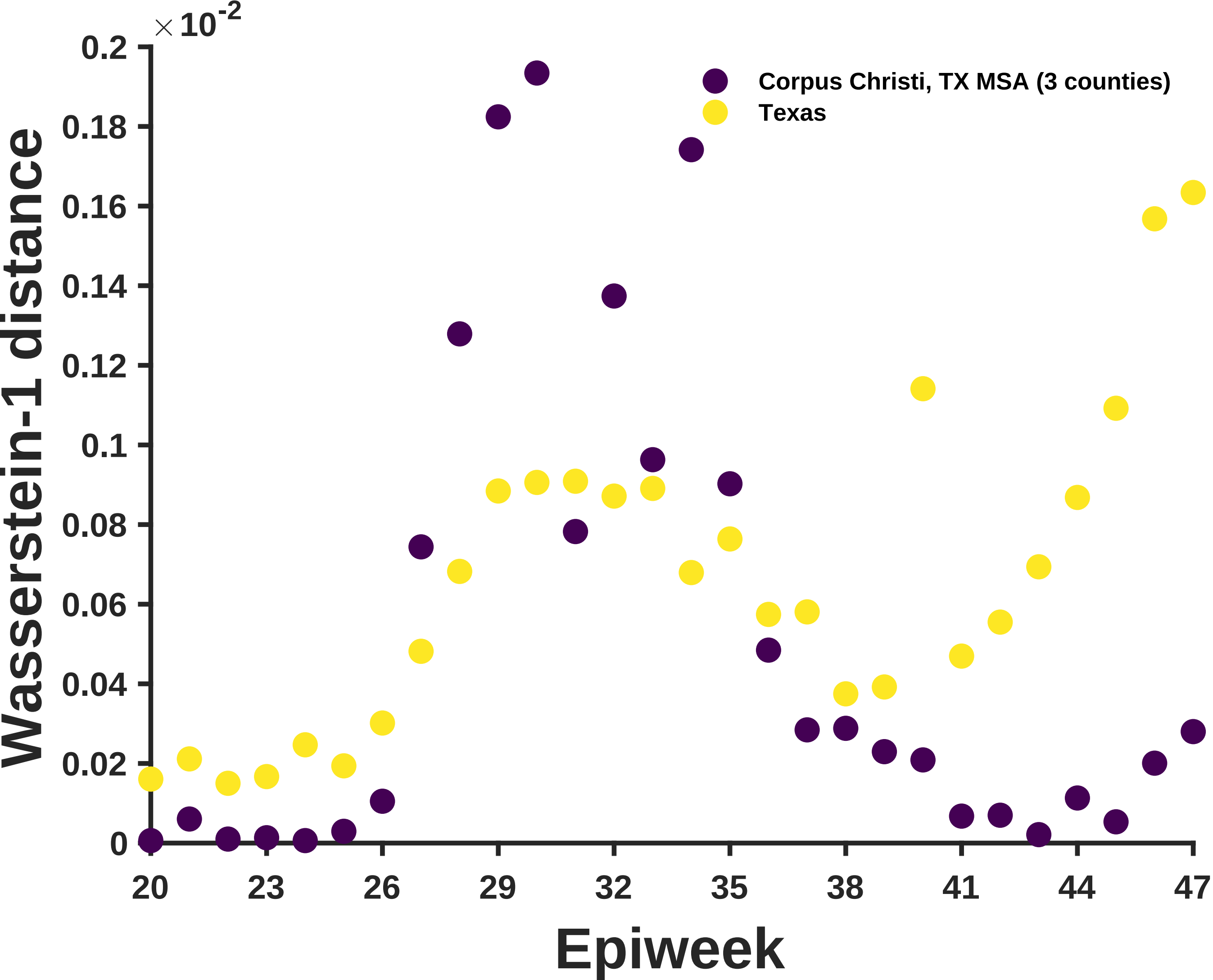

×10^-2
0.2
0.18
0.16
0.14
0.12
0.1
0.08
0.06
0.04
0.02
0
Wasserstein-1 distance
Corpus Christi, TX MSA (3 counties)
Texas
20
23
26
29
32
35
38
41
44
47
Epiweek

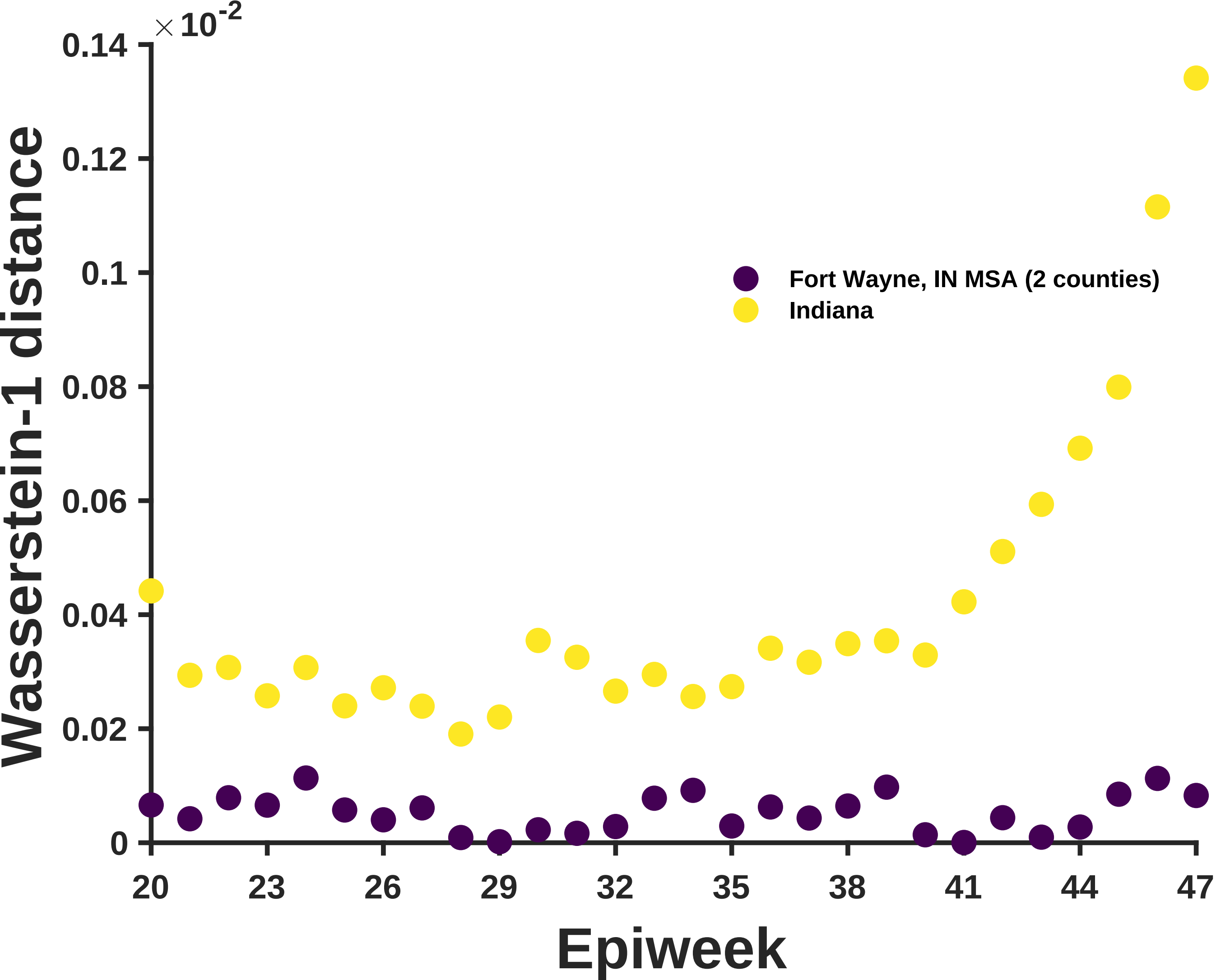

×10^-2
0.14
0.12
0.1
0.08
0.06
0.04
0.02
0
Wasserstein-1 distance
Fort Wayne, IN MSA (2 counties)
Indiana
20
23
26
29
32
35
38
41
44
47
Epiweek

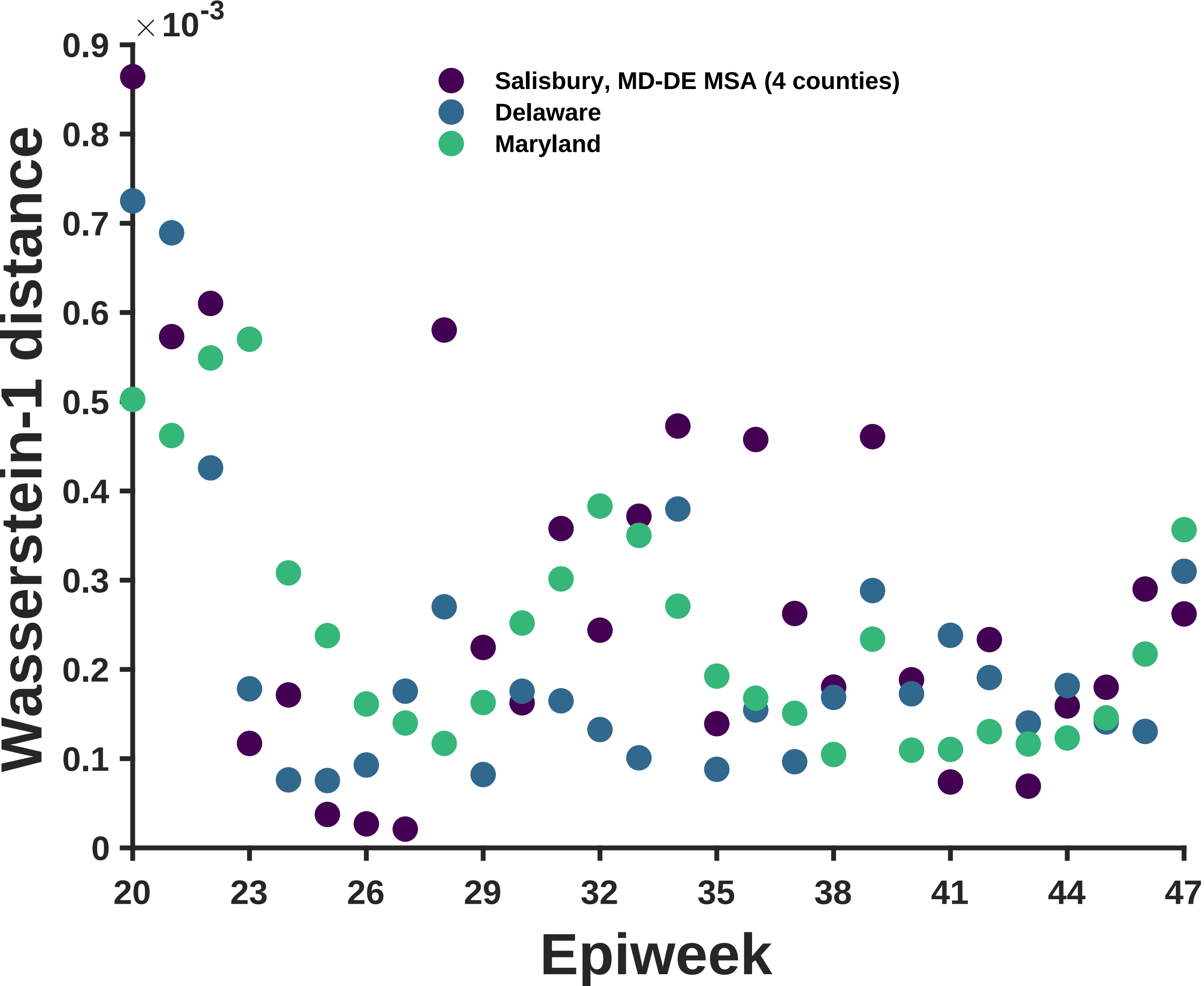

$\times 10^{-3}$
0.9
0.8
0.7
0.6
0.5
0.4
0.3
0.2
0.1
0
Wasserstein-1 distance
Salisbury, MD-DE MSA (4 counties)
Delaware
Maryland
20
23
26
29
32
35
38
41
44
47
Epiweek

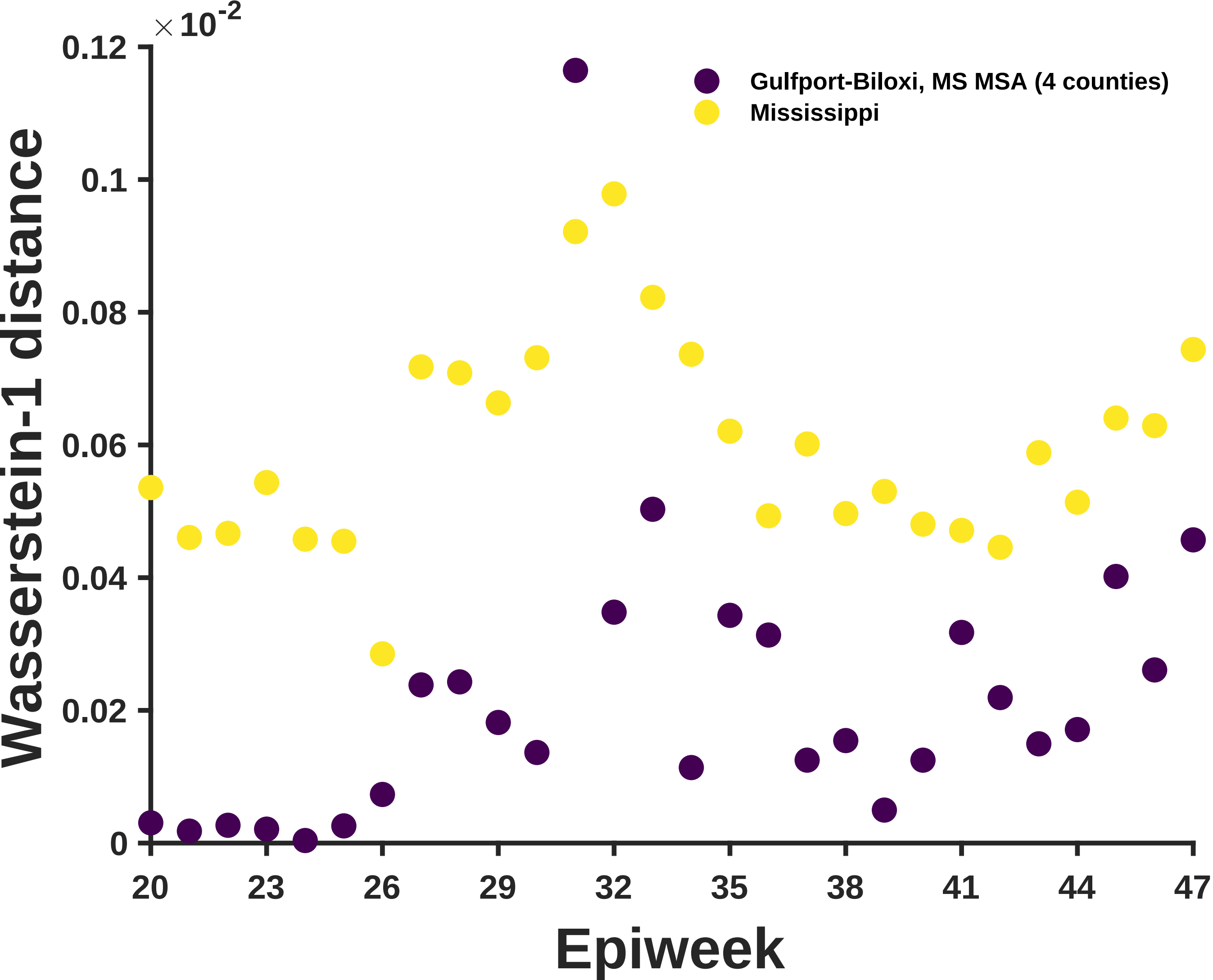

×10^-2
0.12
0.1
0.08
0.06
0.04
0.02
0
Wasserstein-1 distance
Gulfport-Biloxi, MS MSA (4 counties)
Mississippi
20
23
26
29
32
35
38
41
44
47
Epiweek

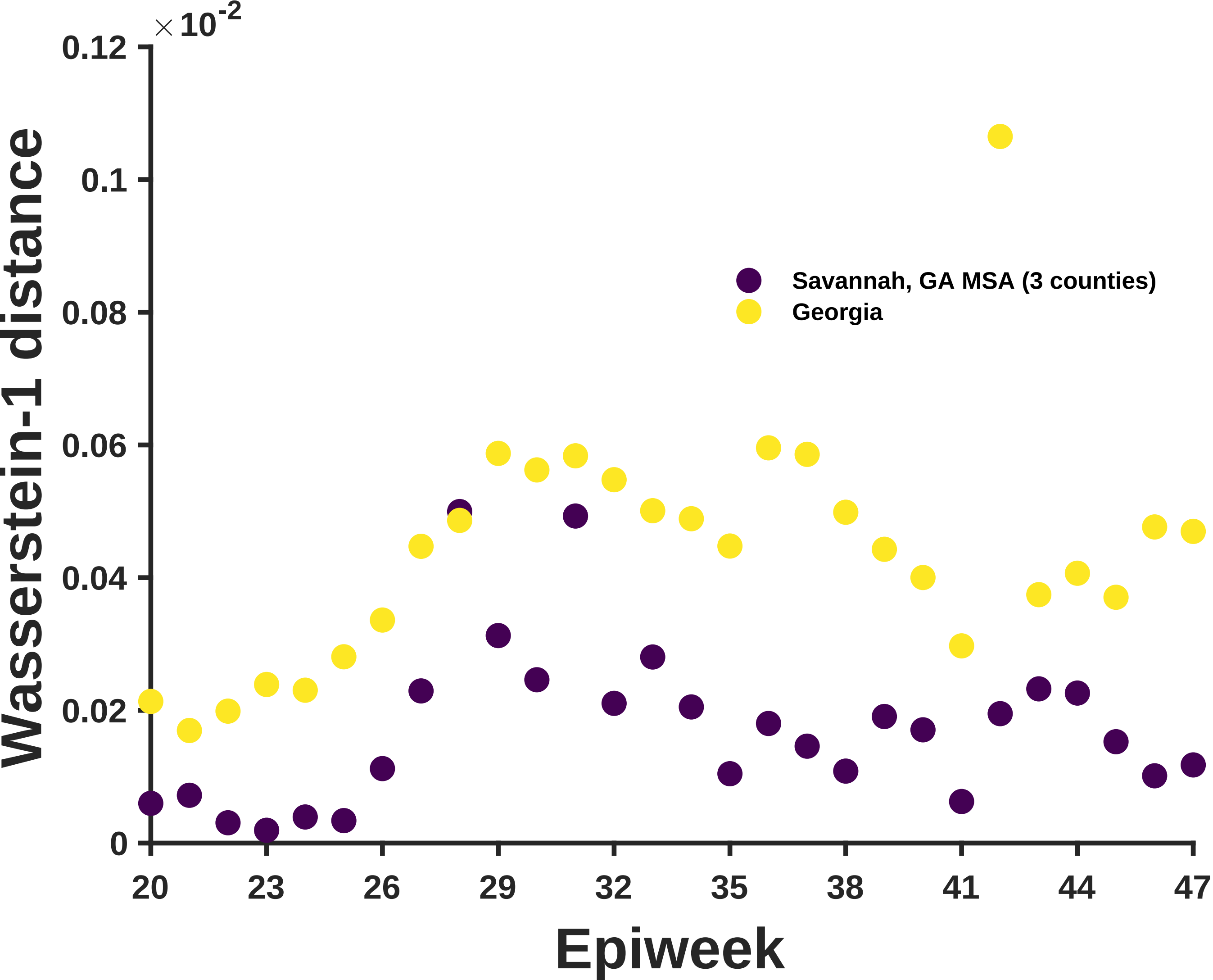

×10^-2
0.12
0.1
0.08
0.06
0.04
0.02
0
Wasserstein-1 distance
Savannah, GA MSA (3 counties)
Georgia
20
23
26
29
32
35
38
41
44
47
Epiweek

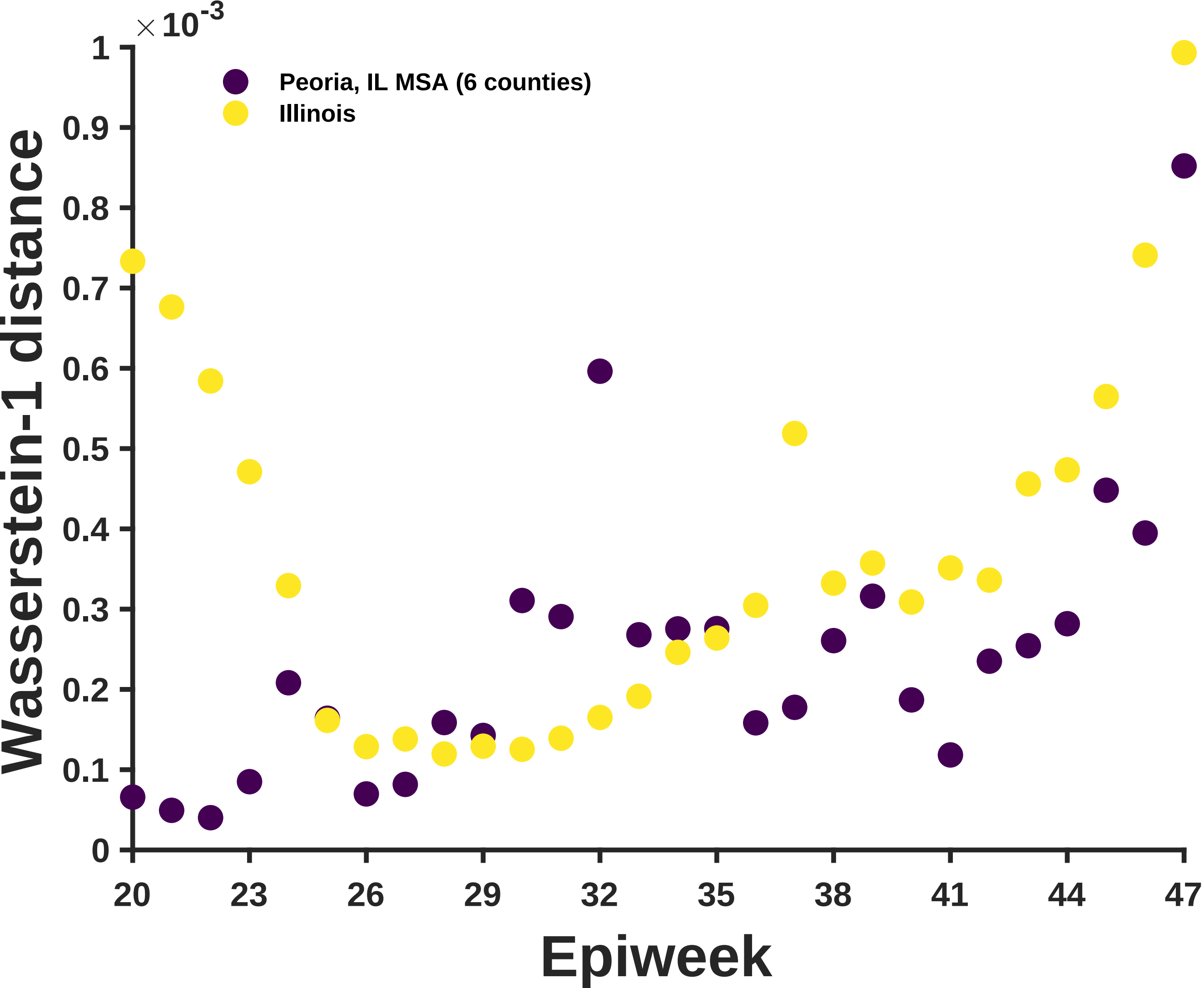

×10^-3
1
0.9
0.8
0.7
0.6
0.5
0.4
0.3
0.2
0.1
0
Wasserstein-1 distance
Peoria, IL MSA (6 counties)
Illinois
20
23
26
29
32
35
38
41
44
47
Epiweek

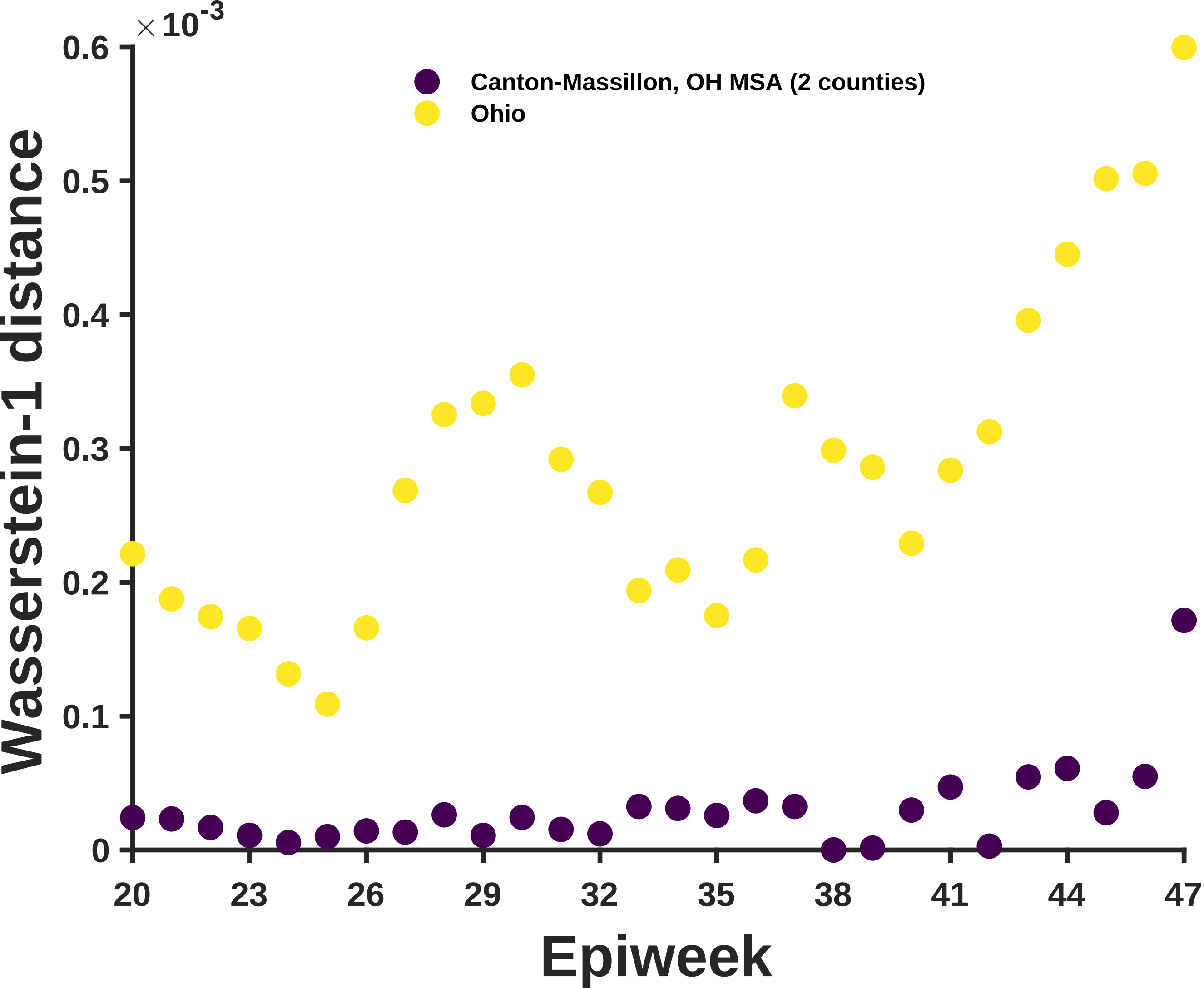

×10^-3
0.6
0.5
0.4
0.3
0.2
0.1
0
Wasserstein-1 distance
Canton-Massillon, OH MSA (2 counties)
Ohio
20
23
26
29
32
35
38
41
44
47
Epiweek

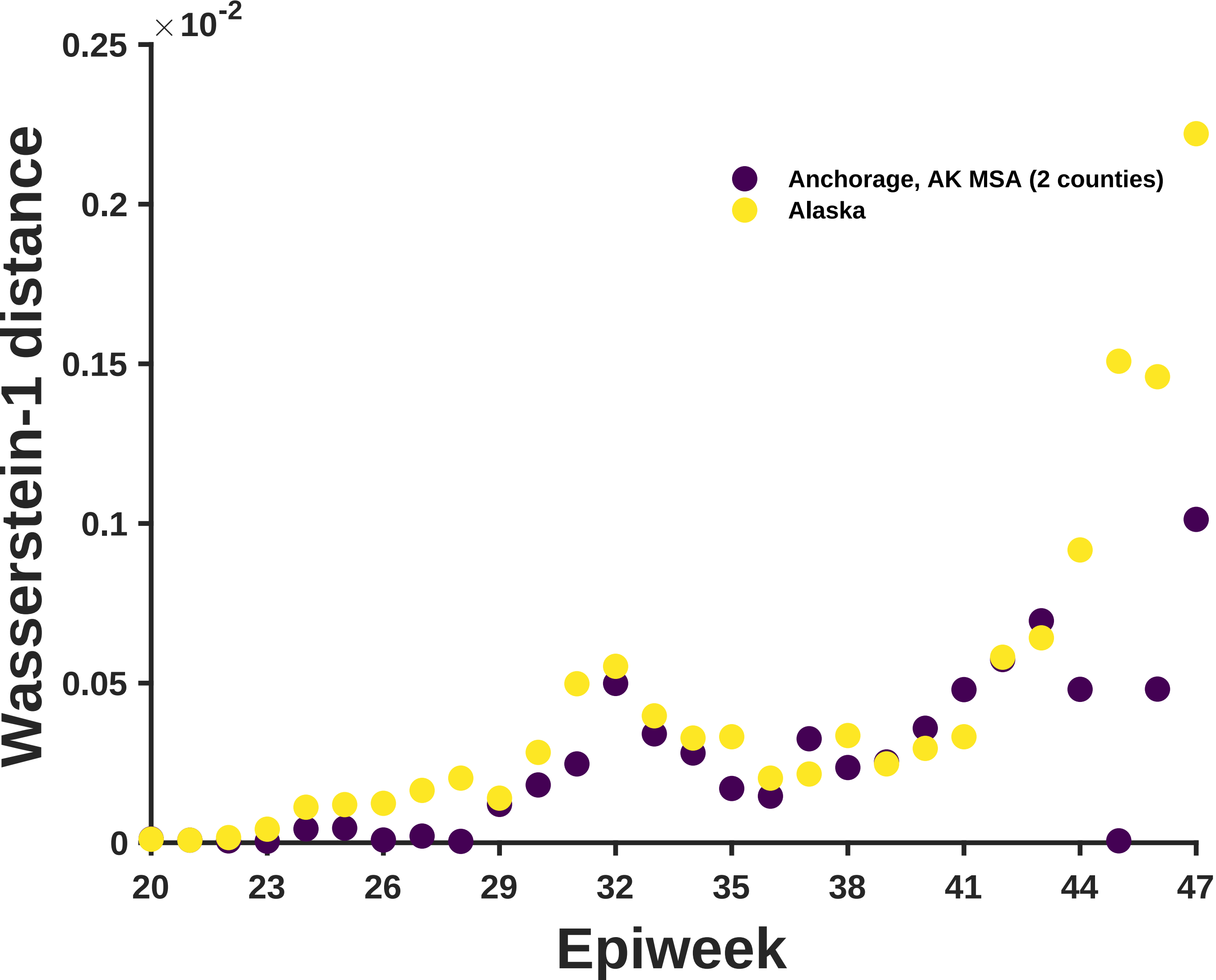

×10^-2
0.25
0.2
0.15
0.1
0.05
0
Wasserstein-1 distance
20
23
26
29
32
35
38
41
44
47
Epiweek
Anchorage, AK MSA (2 counties)
Alaska

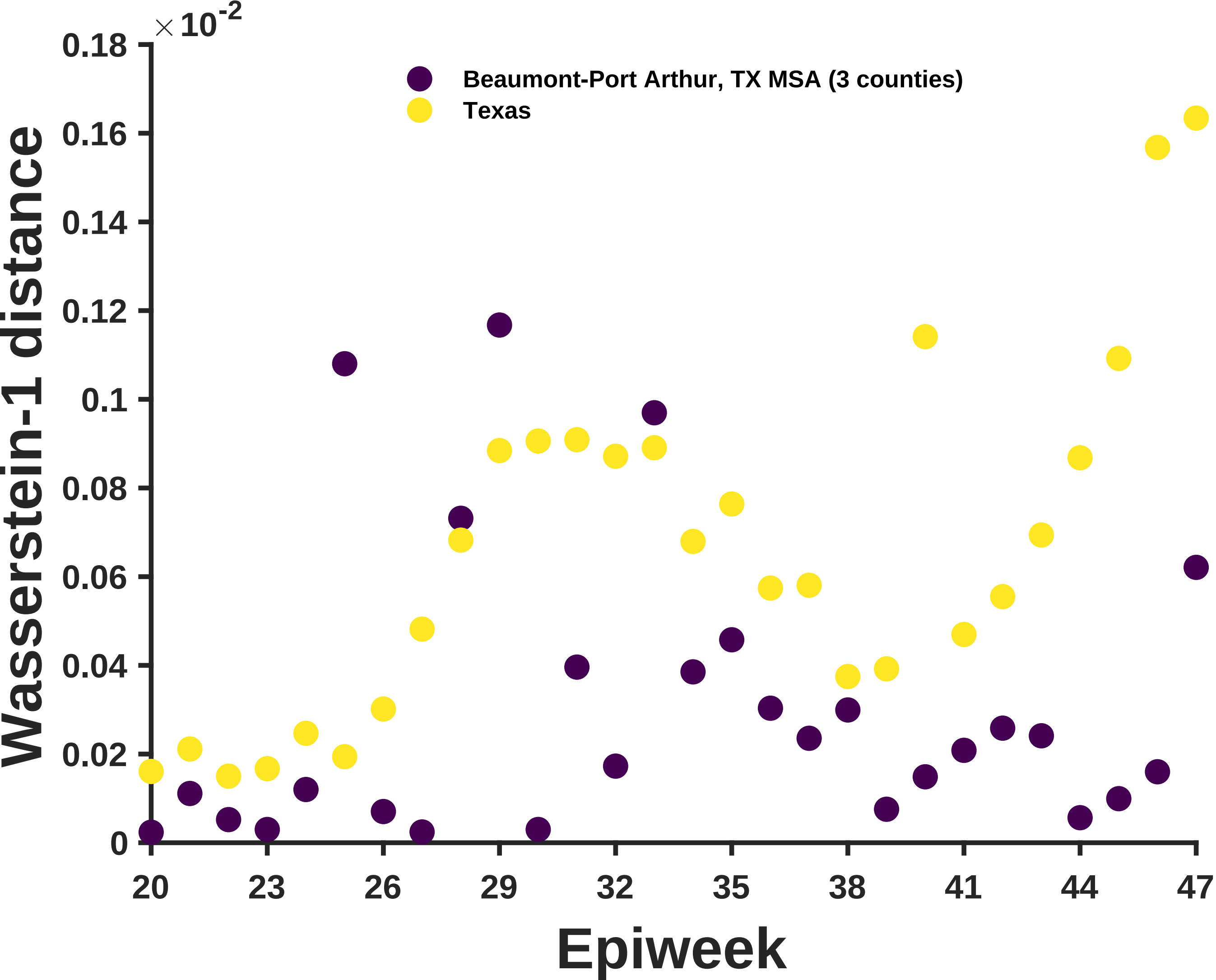

×10^-2
0.18
0.16
0.14
0.12
0.1
0.08
0.06
0.04
0.02
0
Wasserstein-1 distance
Beaumont-Port Arthur, TX MSA (3 counties)
Texas
20
23
26
29
32
35
38
41
44
47
Epiweek

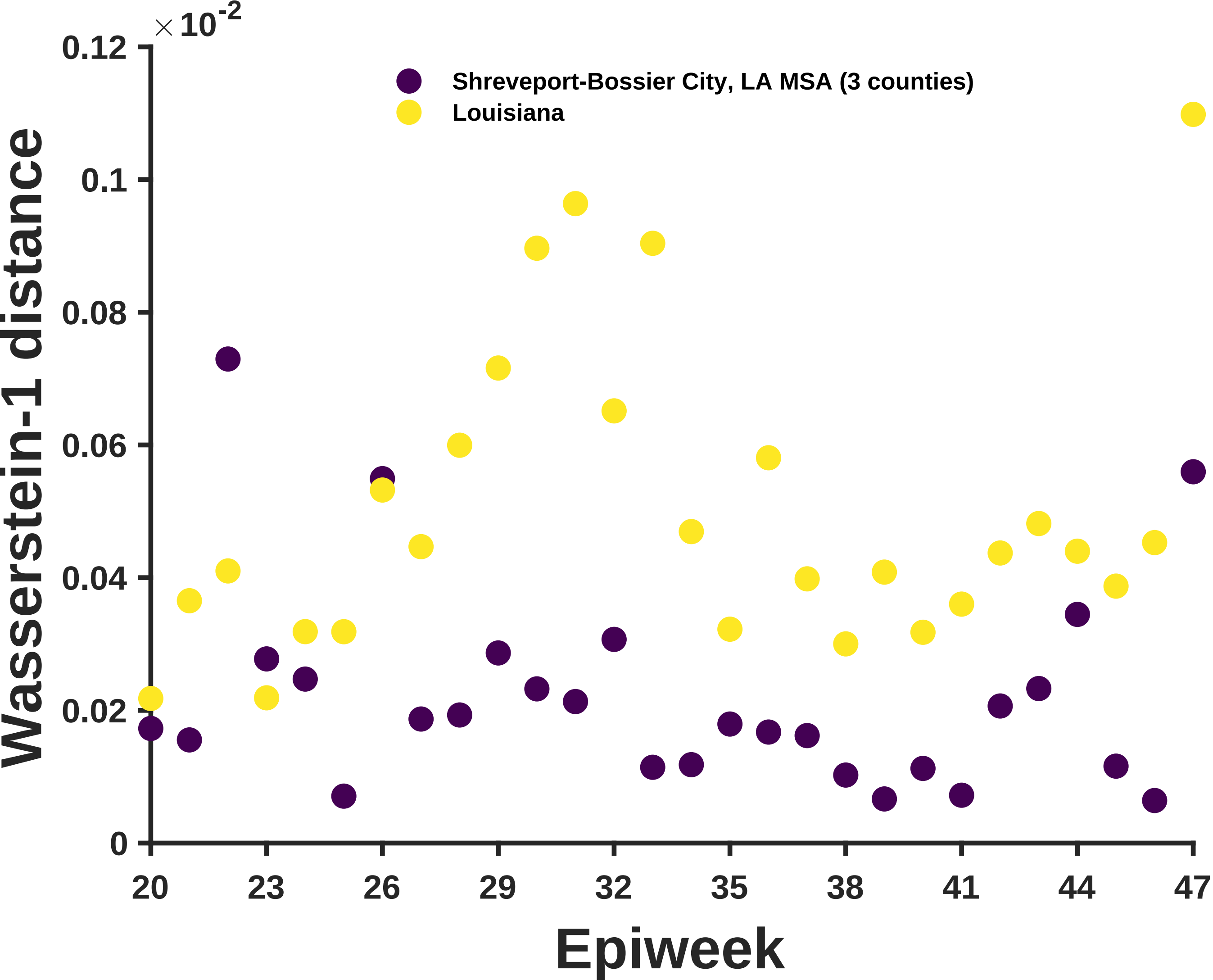

×10^-2
0.12
0.1
0.08
0.06
0.04
0.02
0
Wasserstein-1 distance
Shreveport-Bossier City, LA MSA (3 counties)
Louisiana
20
23
26
29
32
35
38
41
44
47
Epiweek

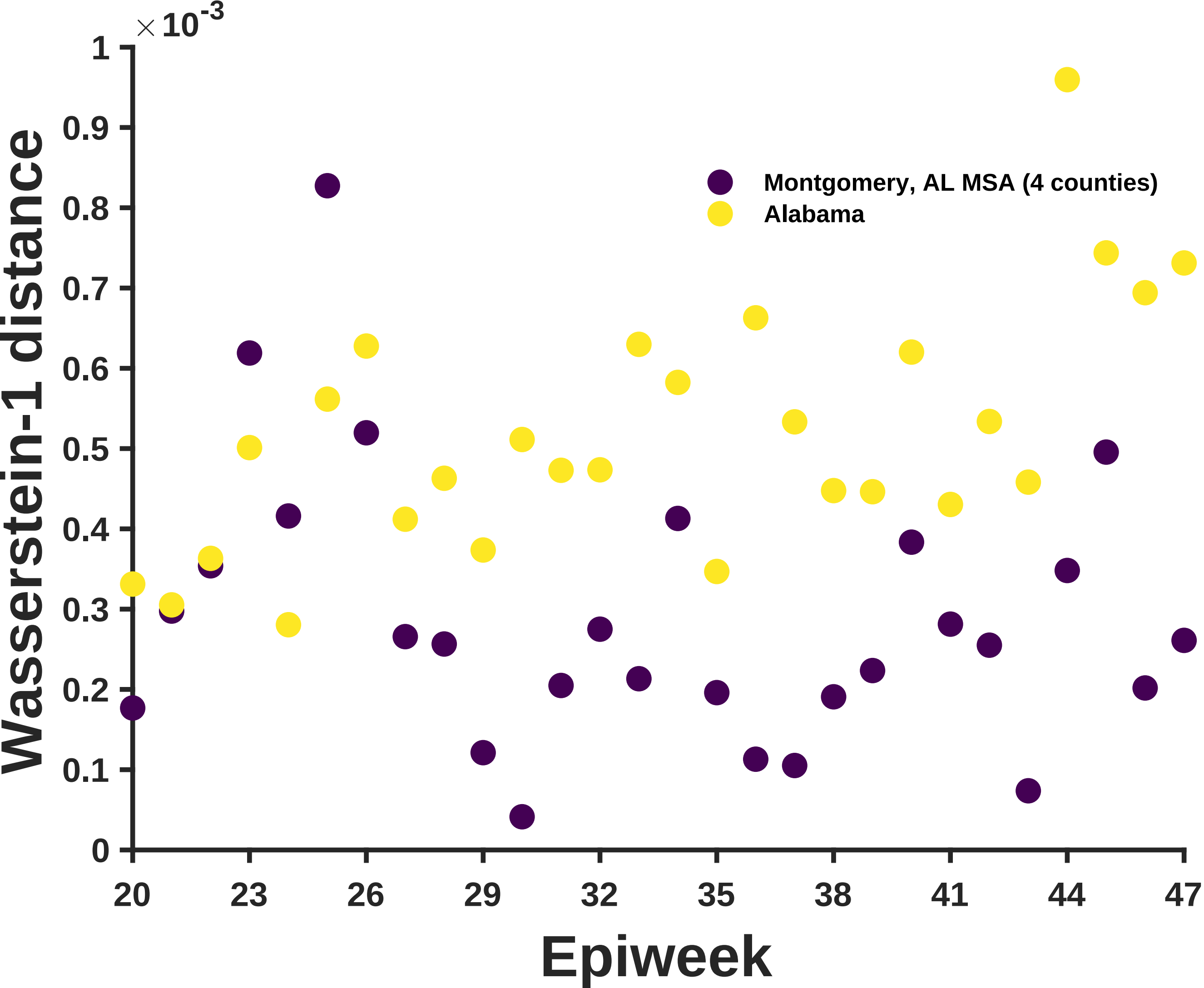

×10^-3
1
0.9
0.8
0.7
0.6
0.5
0.4
0.3
0.2
0.1
0
Wasserstein-1 distance
Montgomery, AL MSA (4 counties)
Alabama
20
23
26
29
32
35
38
41
44
47
Epiweek

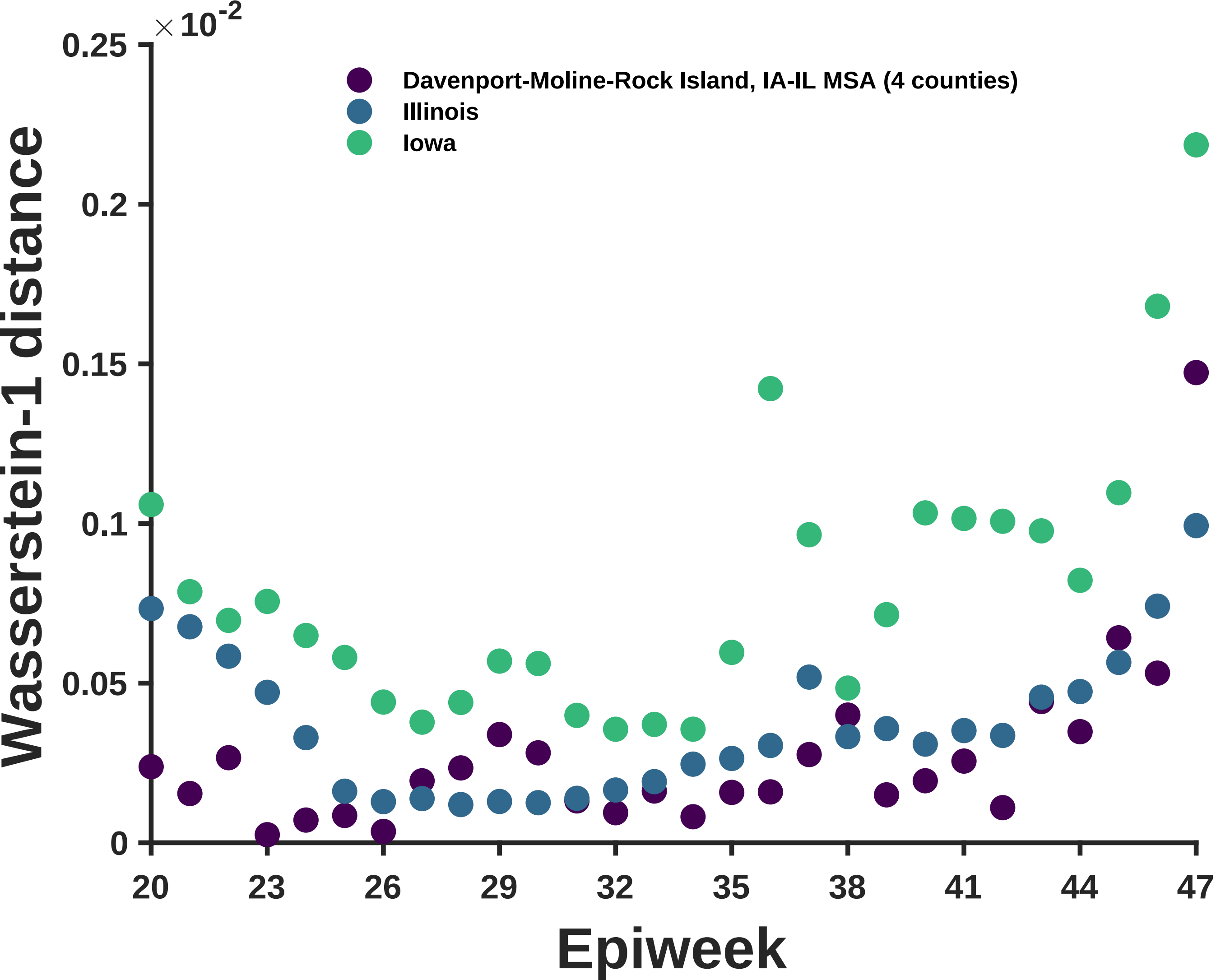

×10^-2
0.25
0.2
0.15
0.1
0.05
0
Davenport-Moline-Rock Island, IA-IL MSA (4 counties)
Illinois
Iowa
Wasserstein-1 distance
20
23
26
29
32
35
38
41
44
47
Epiweek

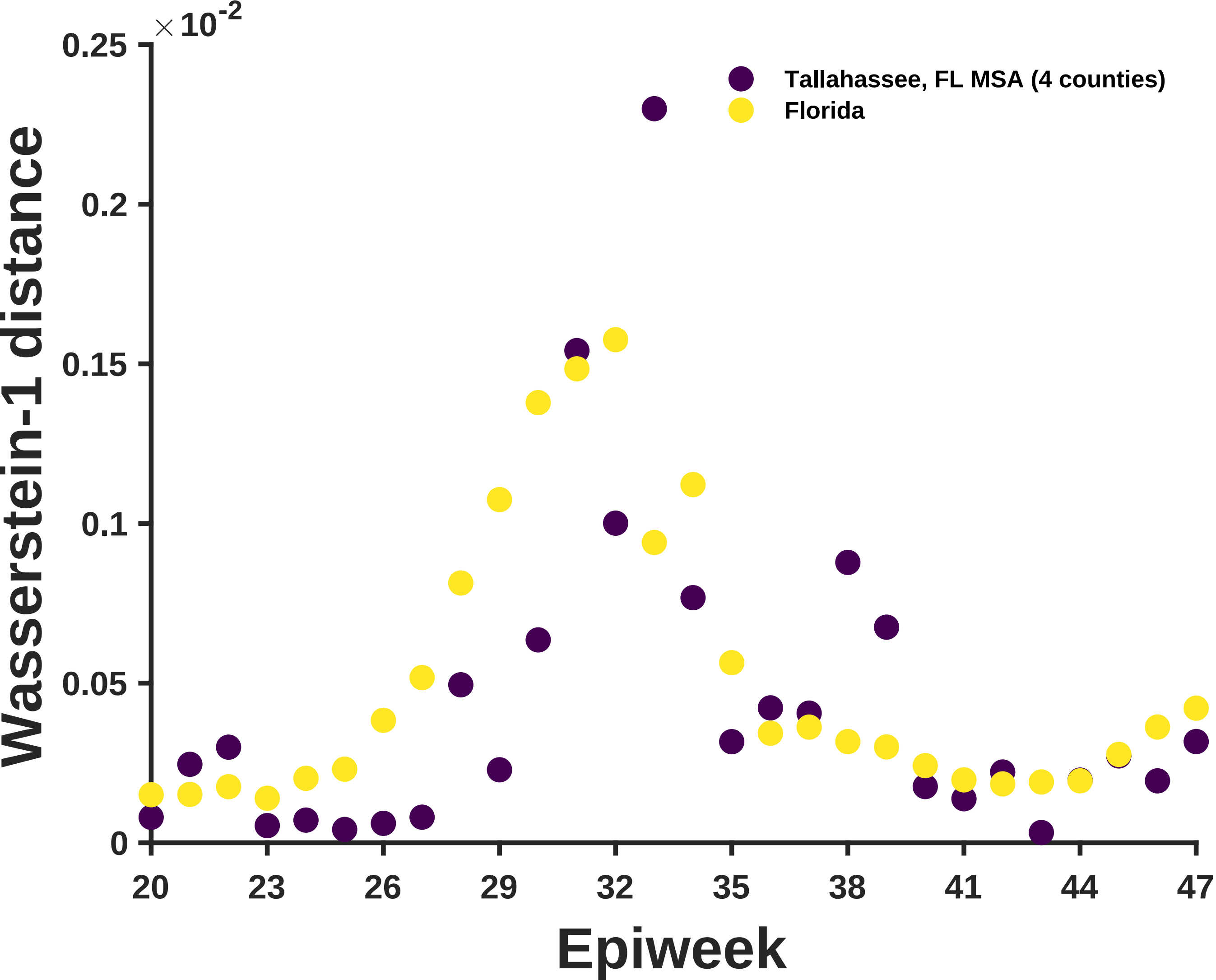
×10^-2
0.25
0.2
0.15
0.1
0.05
0
Wasserstein-1 distance
Tallahassee, FL MSA (4 counties)
Florida
20
23
26
29
32
35
38
41
44
47
Epiweek

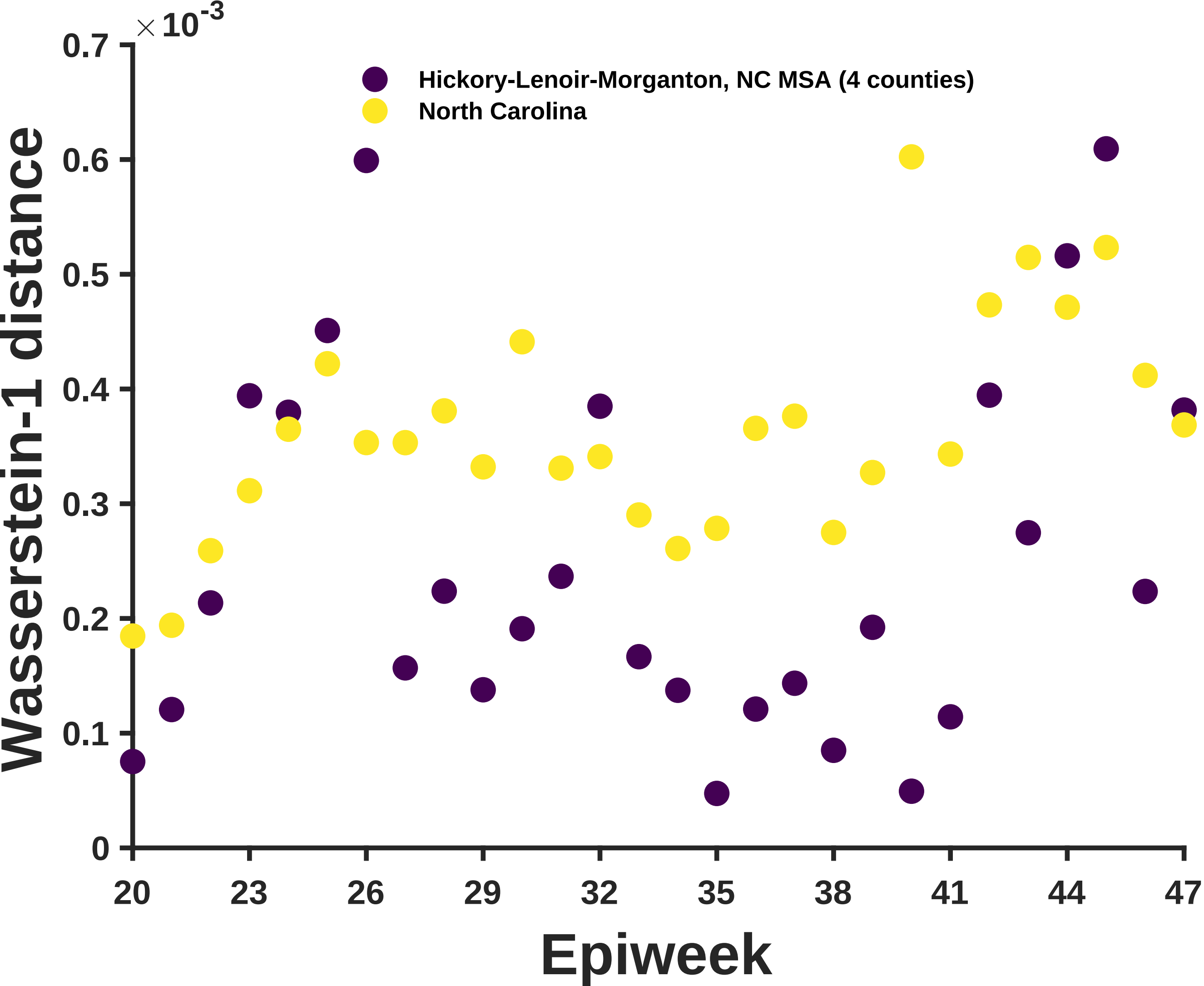

×10^-3
0.7
0.6
0.5
0.4
0.3
0.2
0.1
0
Wasserstein-1 distance
Hickory-Lenoir-Morganton, NC MSA (4 counties)
North Carolina
20
23
26
29
32
35
38
41
44
47
Epiweek

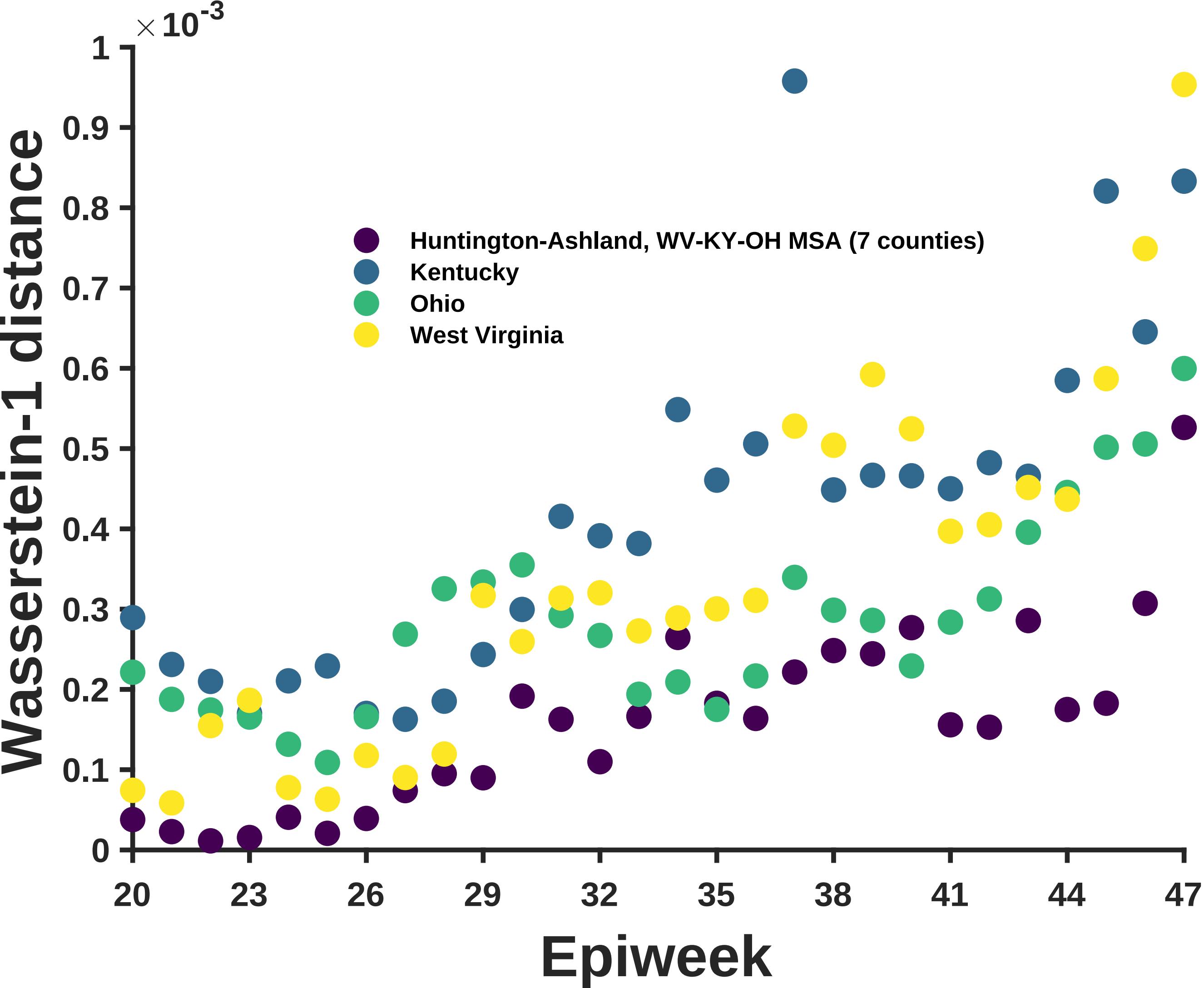

×10-3
1
0.9
0.8
0.7
0.6
0.5
0.4
0.3
0.2
0.1
0
Wasserstein-1 distance
Huntington-Ashland, WV-KY-OH MSA (7 counties)
Kentucky
Ohio
West Virginia
20
23
26
29
32
35
38
41
44
47
Epiweek

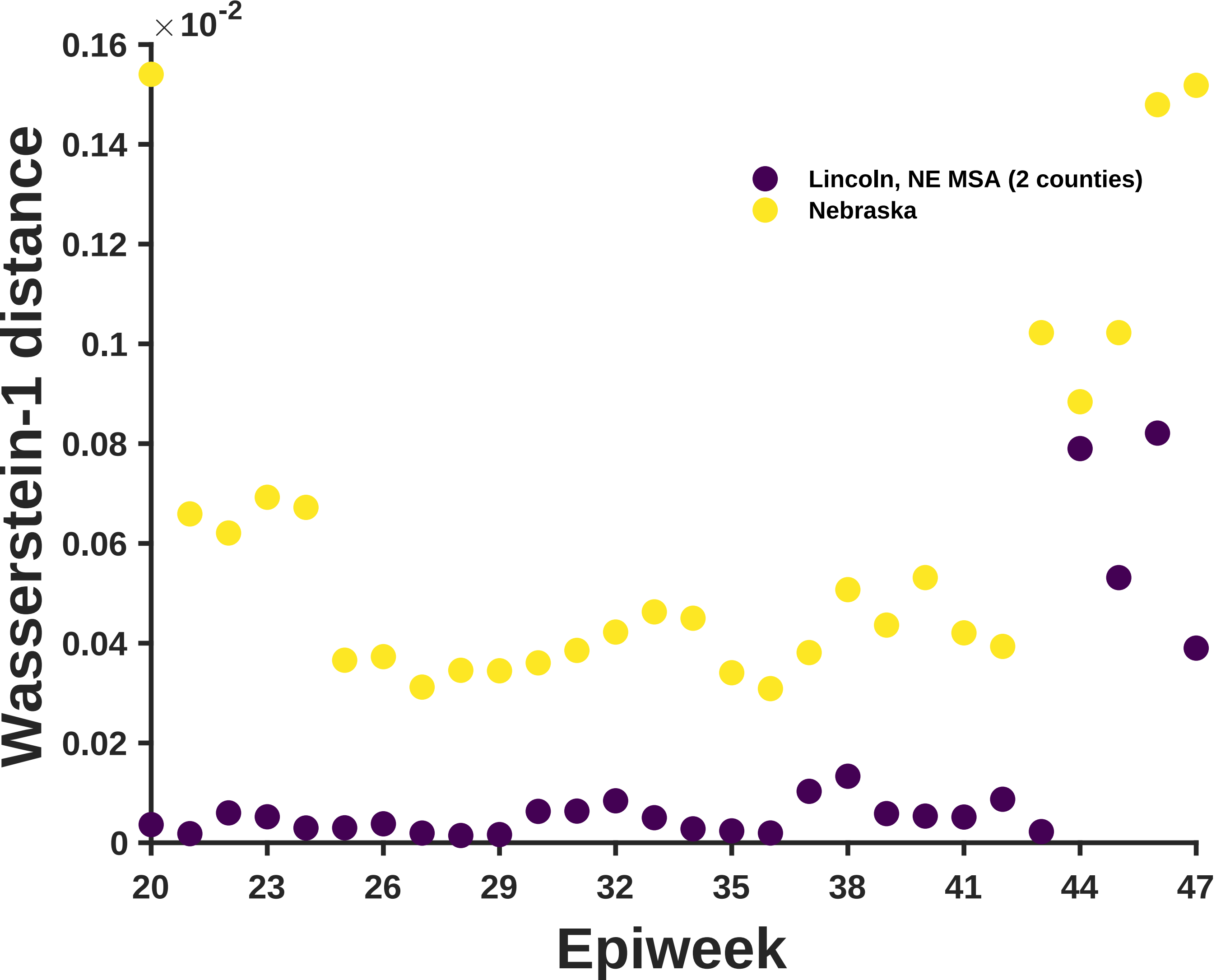

×10^-2
0.16
0.14
0.12
0.1
0.08
0.06
0.04
0.02
0
Wasserstein-1 distance
Lincoln, NE MSA (2 counties)
Nebraska
20
23
26
29
32
35
38
41
44
47
Epiweek

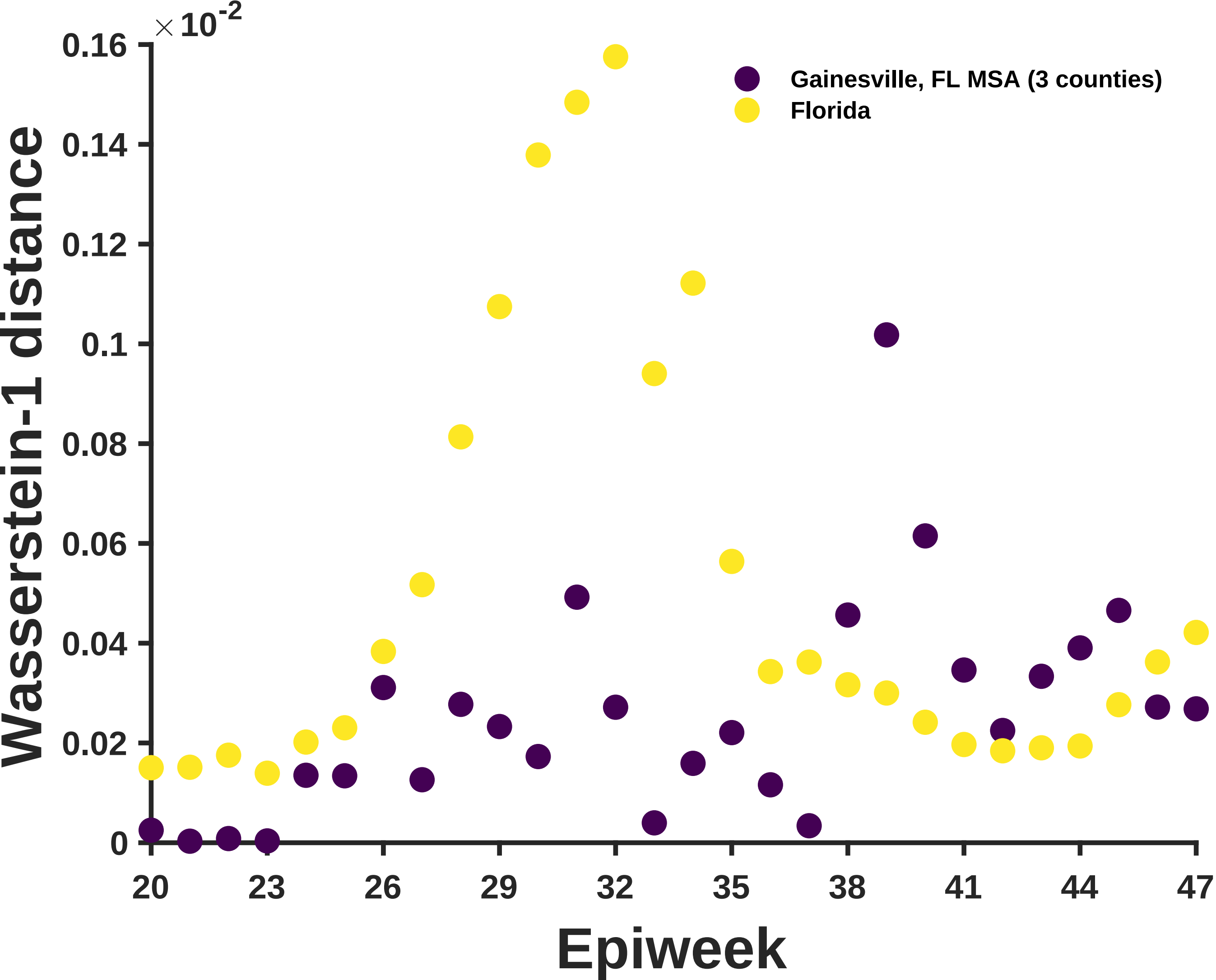

×10^-2
0.16
0.14
0.12
0.1
0.08
0.06
0.04
0.02
0
Wasserstein-1 distance
Gainesville, FL MSA (3 counties)
Florida
20
23
26
29
32
35
38
41
44
47
Epiweek

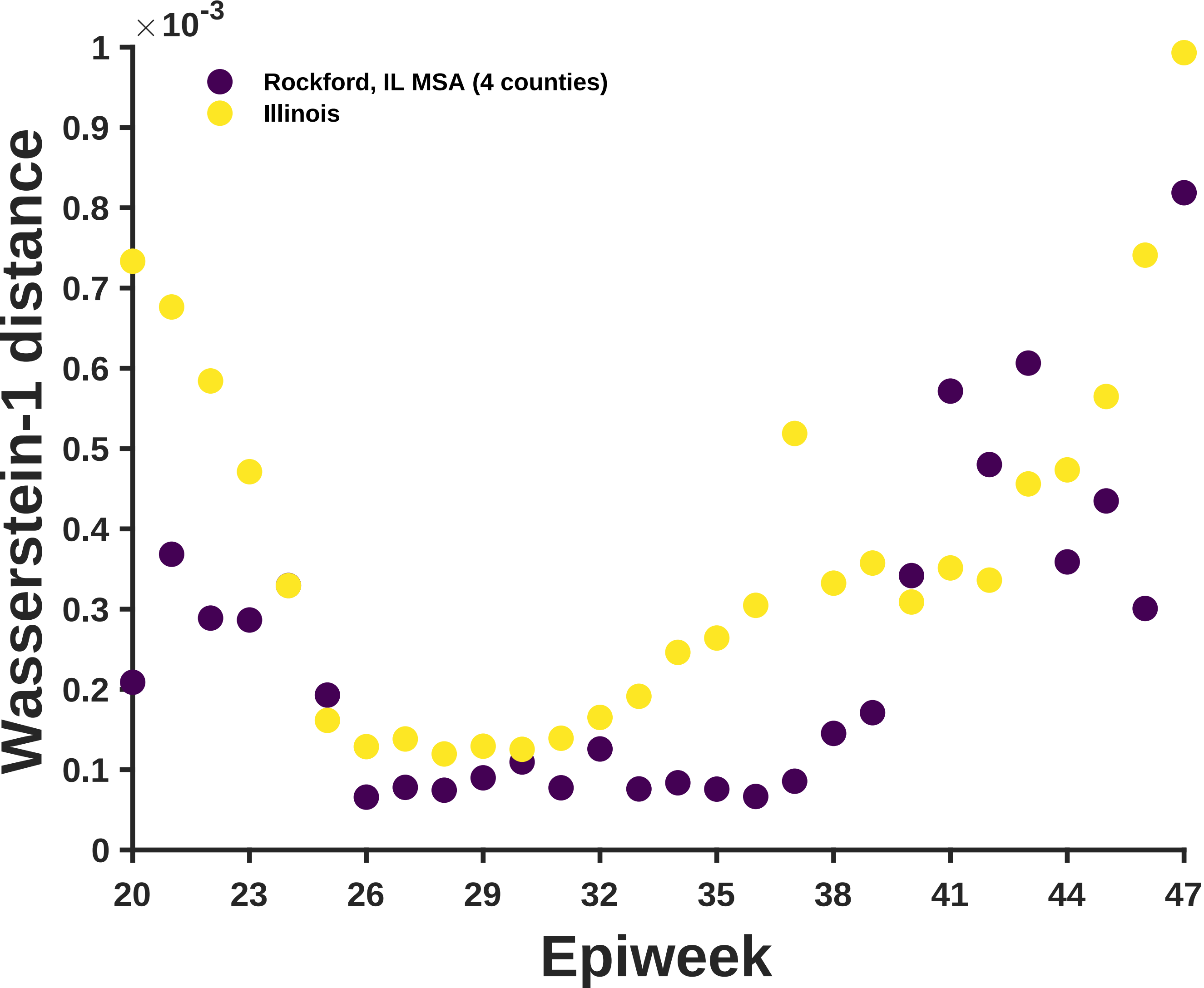

Rockford, IL MSA (4 counties)
Illinois
Wasserstein-1 distance
Epiweek
$\times 10^{-3}$
0
0.1
0.2
0.3
0.4
0.5
0.6
0.7
0.8
0.9
1
20
23
26
29
32
35
38
41
44
47

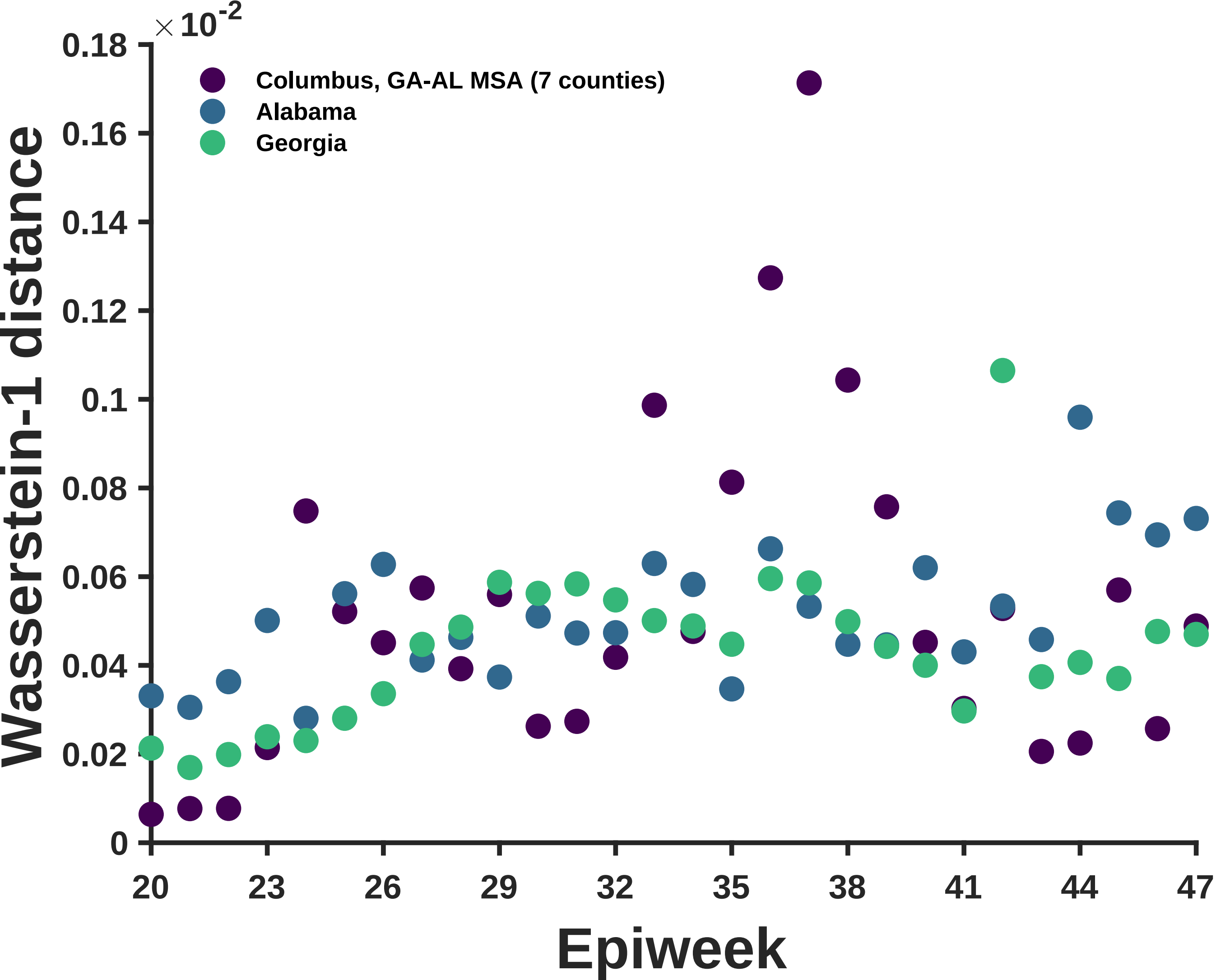

×10^-2
0.18
0.16
0.14
0.12
0.1
0.08
0.06
0.04
0.02
0
Wasserstein-1 distance
Columbus, GA-AL MSA (7 counties)
Alabama
Georgia
20
23
26
29
32
35
38
41
44
47
Epiweek

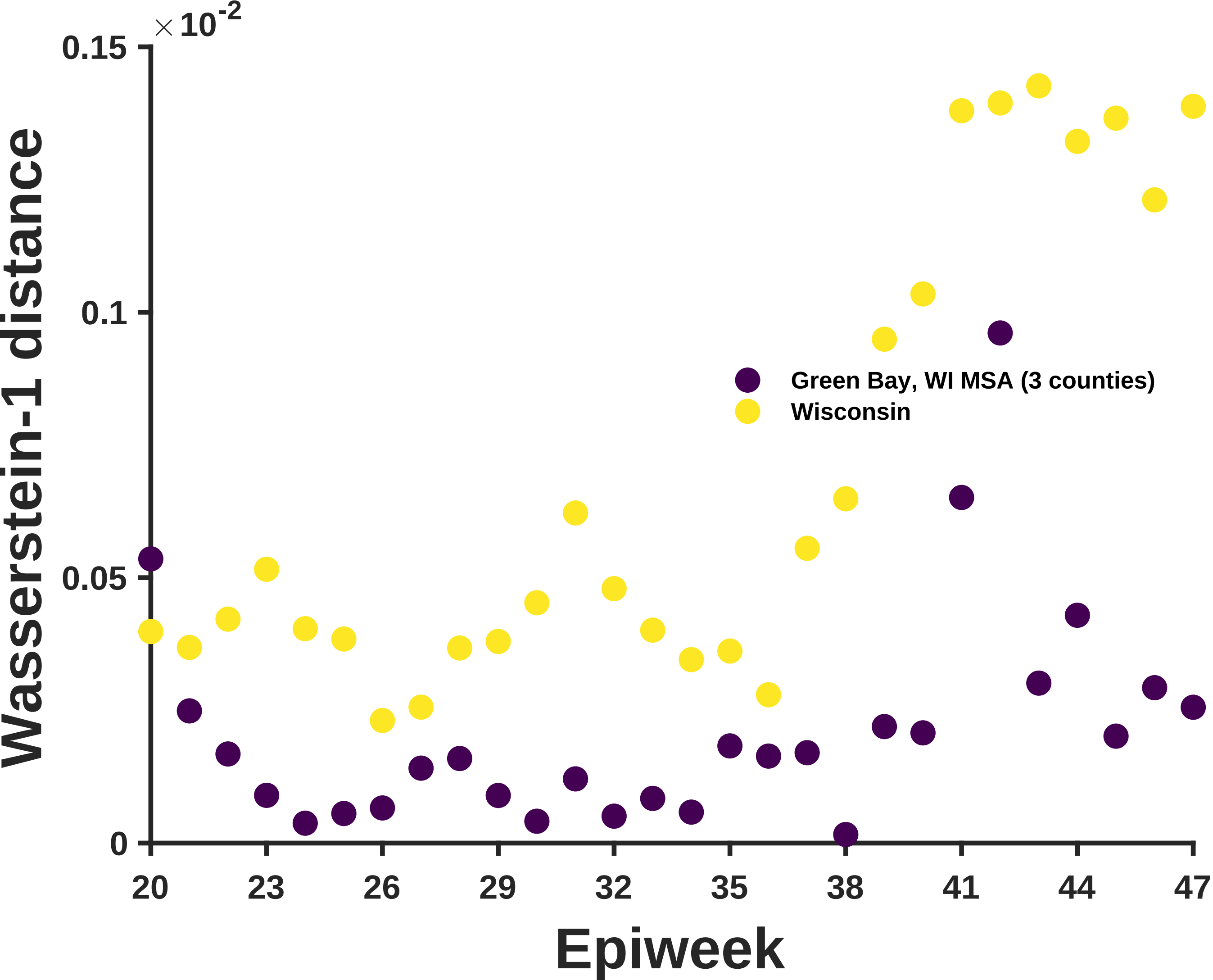

×10^-2
0.15
0.1
0.05
0
Wasserstein-1 distance
20
23
26
29
32
35
38
41
44
47
Epiweek
Green Bay, WI MSA (3 counties)
Wisconsin

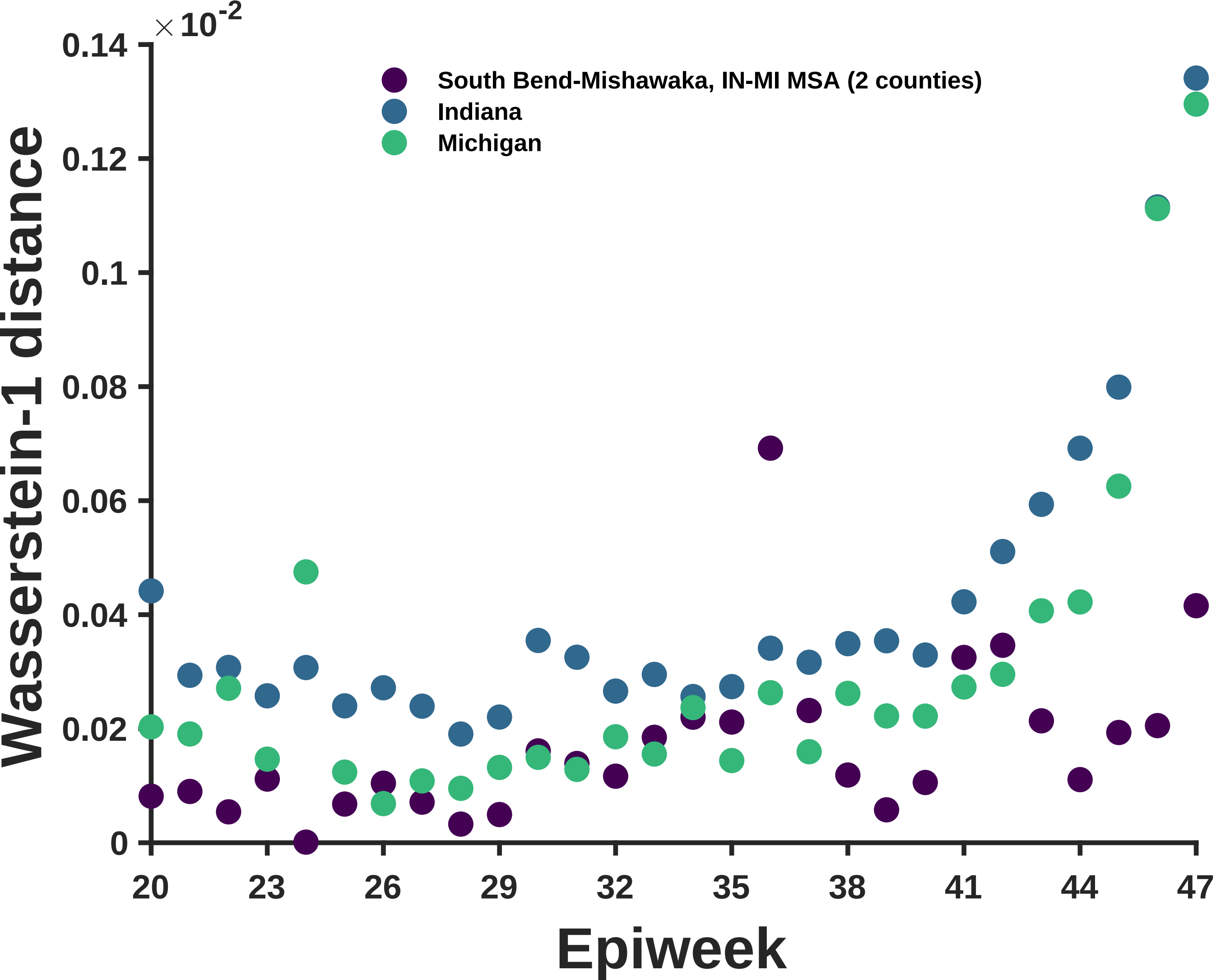

×10^-2
0.14
0.12
0.1
0.08
0.06
0.04
0.02
0
Wasserstein-1 distance
South Bend-Mishawaka, IN-MI MSA (2 counties)
Indiana
Michigan
20
23
26
29
32
35
38
41
44
47
Epiweek

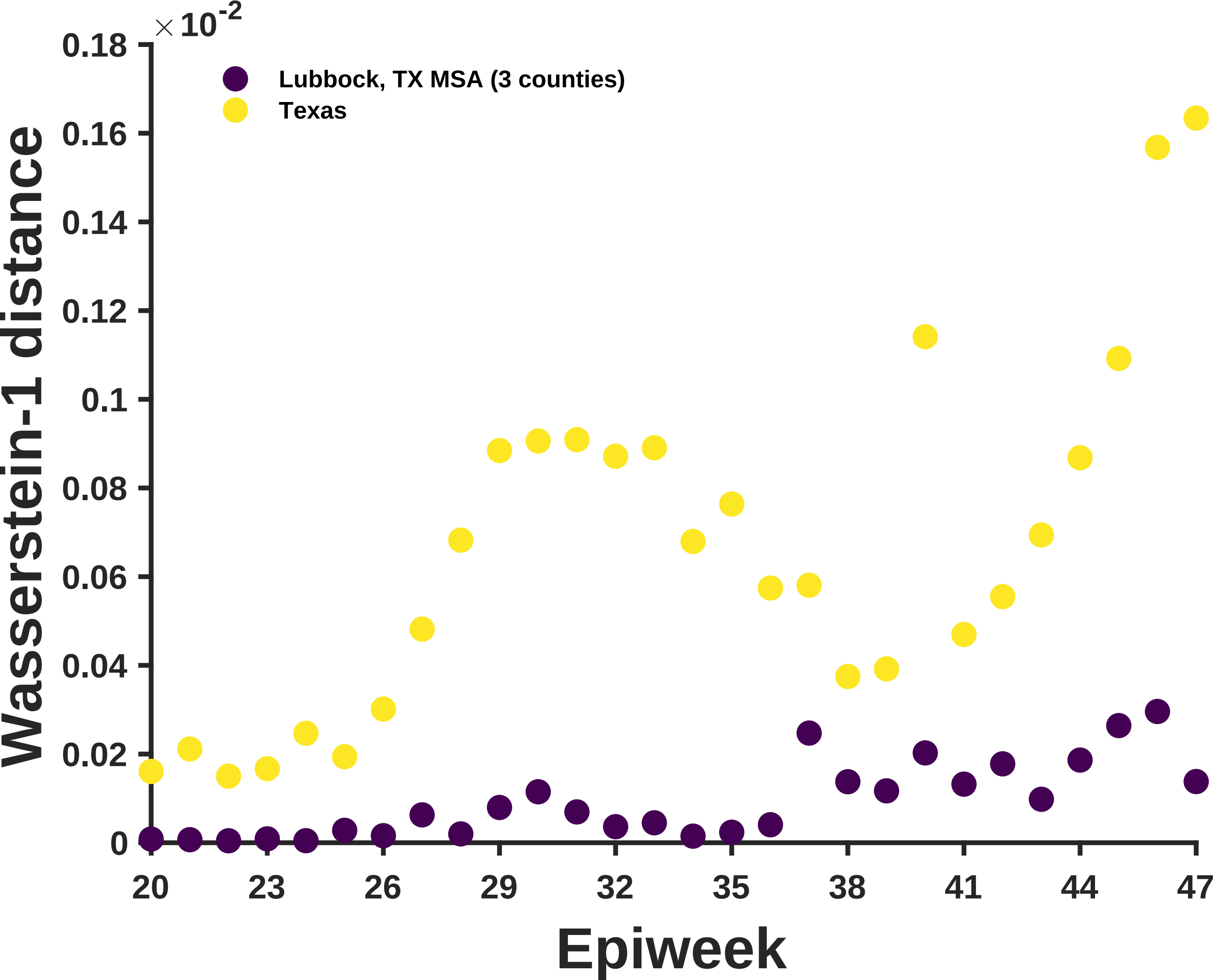

×10^-2
0.18
0.16
0.14
0.12
0.1
0.08
0.06
0.04
0.02
0
Wasserstein-1 distance
Lubbock, TX MSA (3 counties)
Texas
20
23
26
29
32
35
38
41
44
47
Epiweek

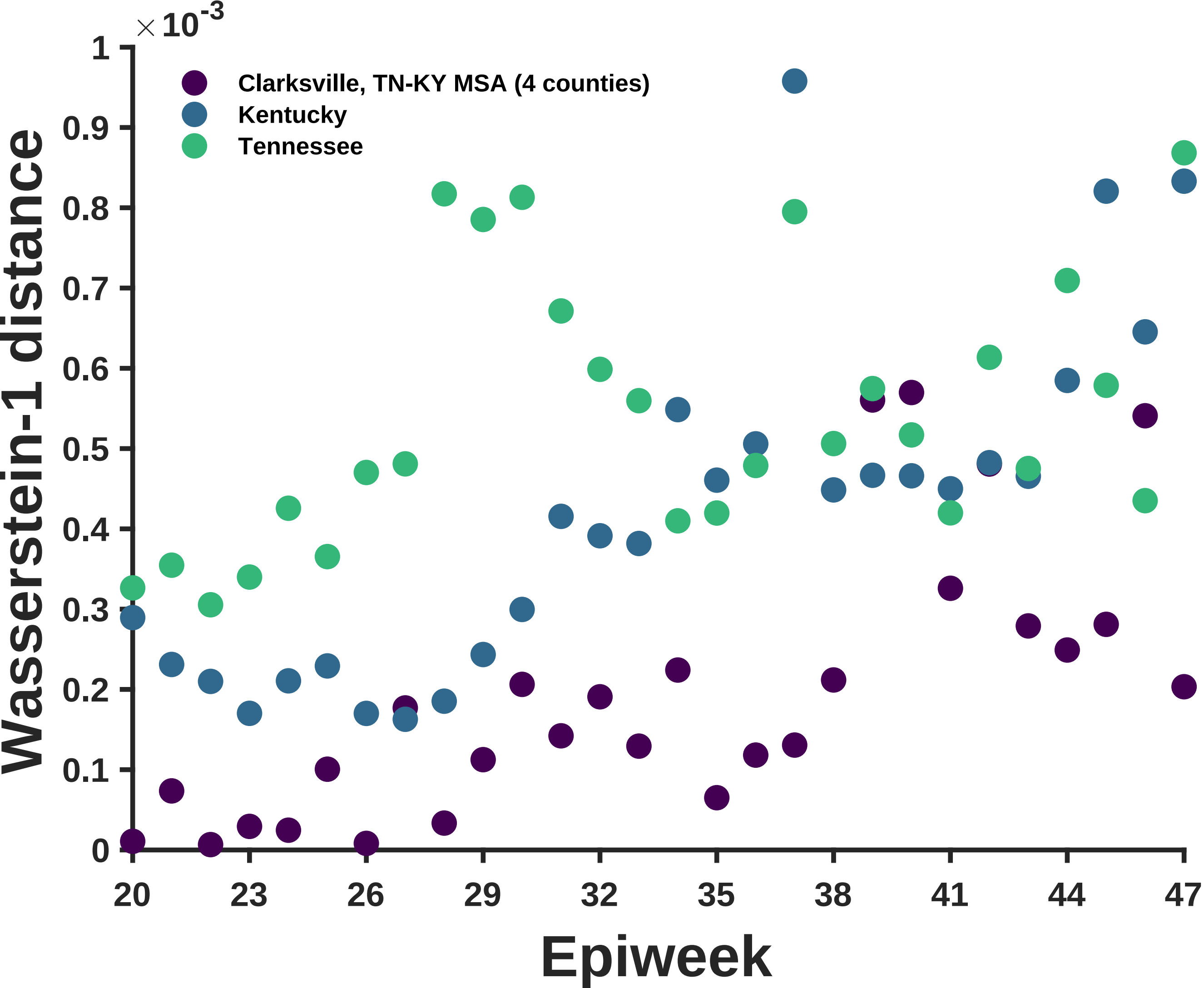

×10^-3
Clarksville, TN-KY MSA (4 counties)
Kentucky
Tennessee
Wasserstein-1 distance
Epiweek
0
0.1
0.2
0.3
0.4
0.5
0.6
0.7
0.8
0.9
1
20
23
26
29
32
35
38
41
44
47

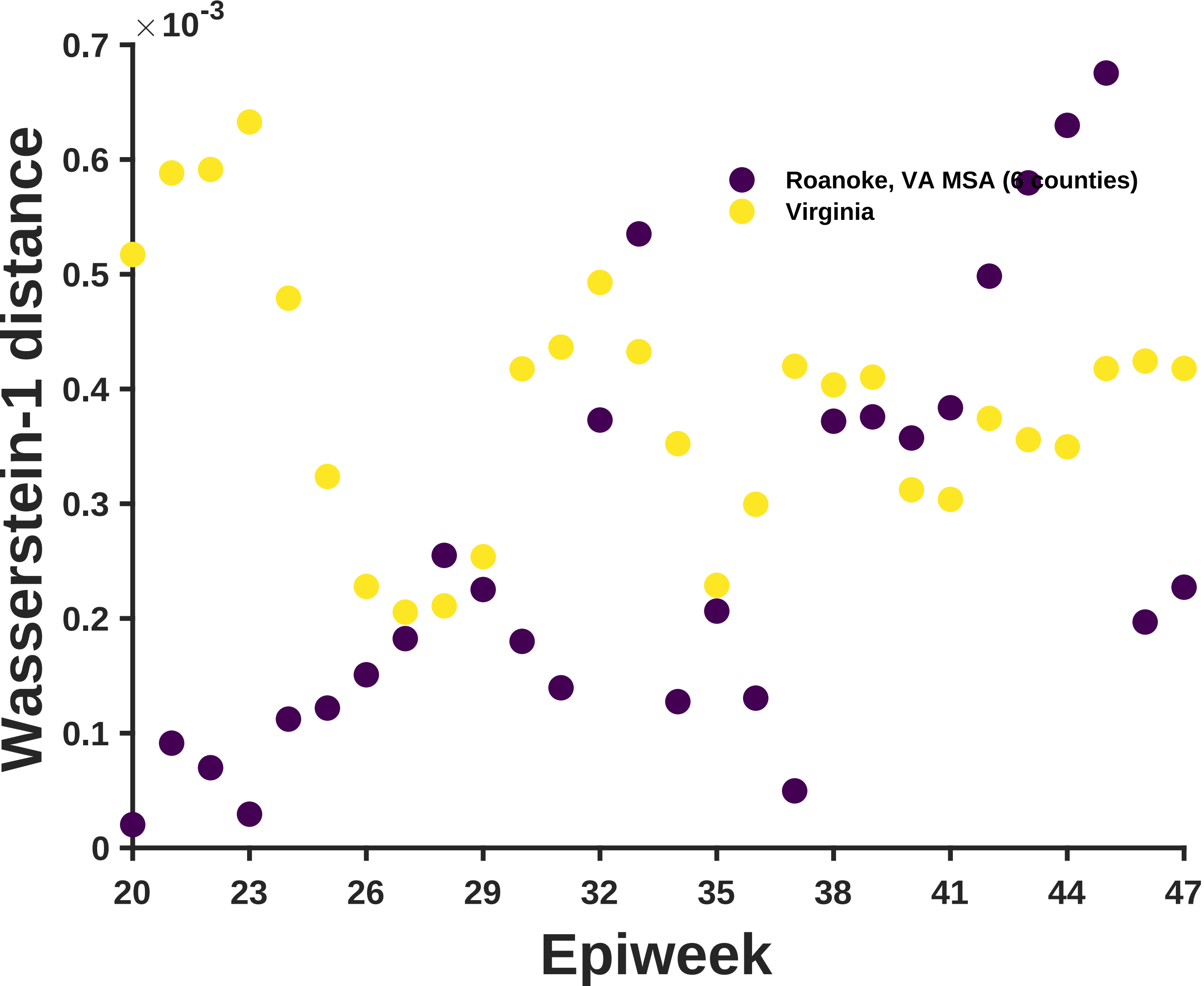

×10^-3
0.7
0.6
0.5
0.4
0.3
0.2
0.1
0
Wasserstein-1 distance
Roanoke, VA MSA (6 counties)
Virginia
20
23
26
29
32
35
38
41
44
47
Epiweek

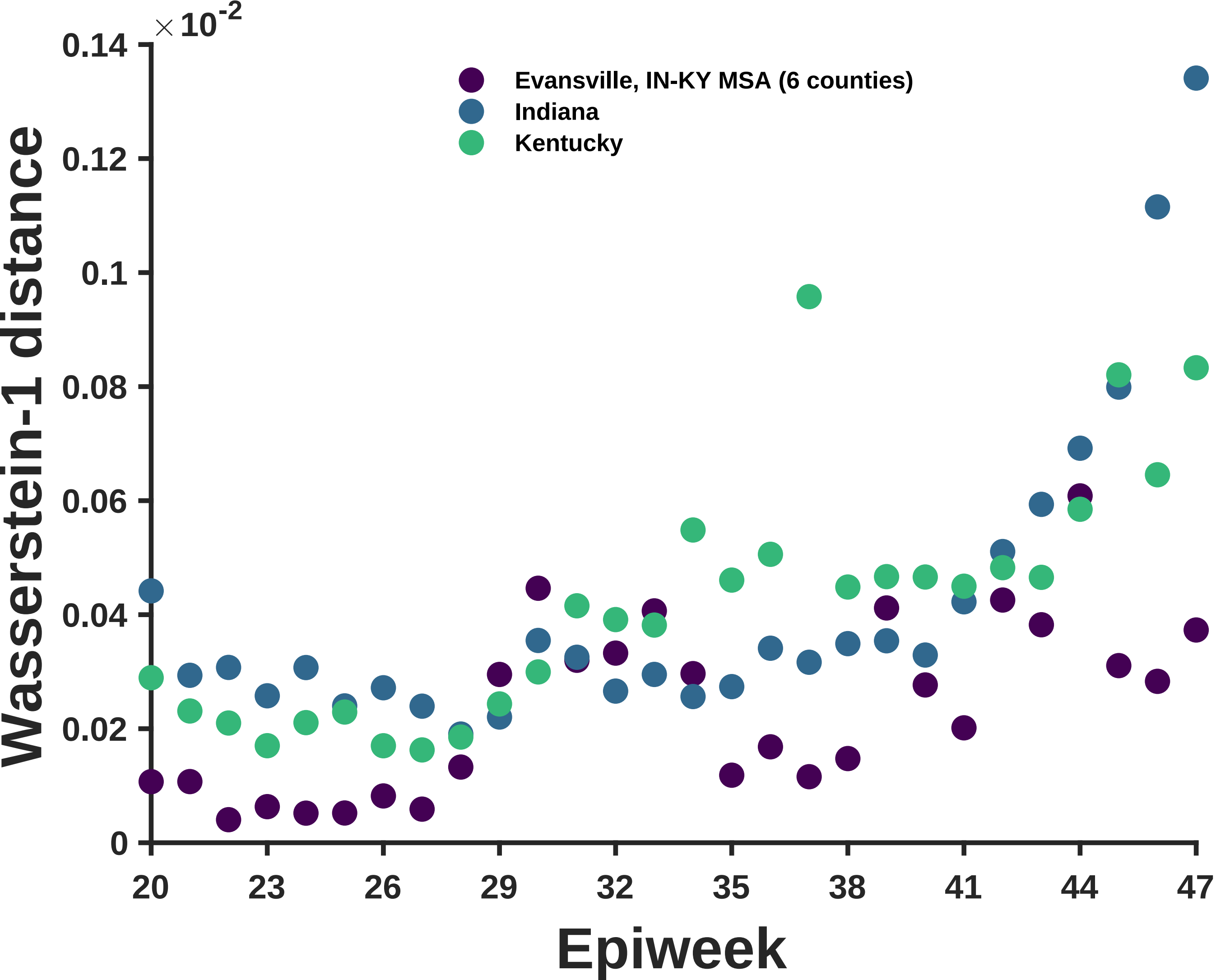

×10^-2
0.14
0.12
0.1
0.08
0.06
0.04
0.02
0
Wasserstein-1 distance
Evansville, IN-KY MSA (6 counties)
Indiana
Kentucky
20
23
26
29
32
35
38
41
44
47
Epiweek

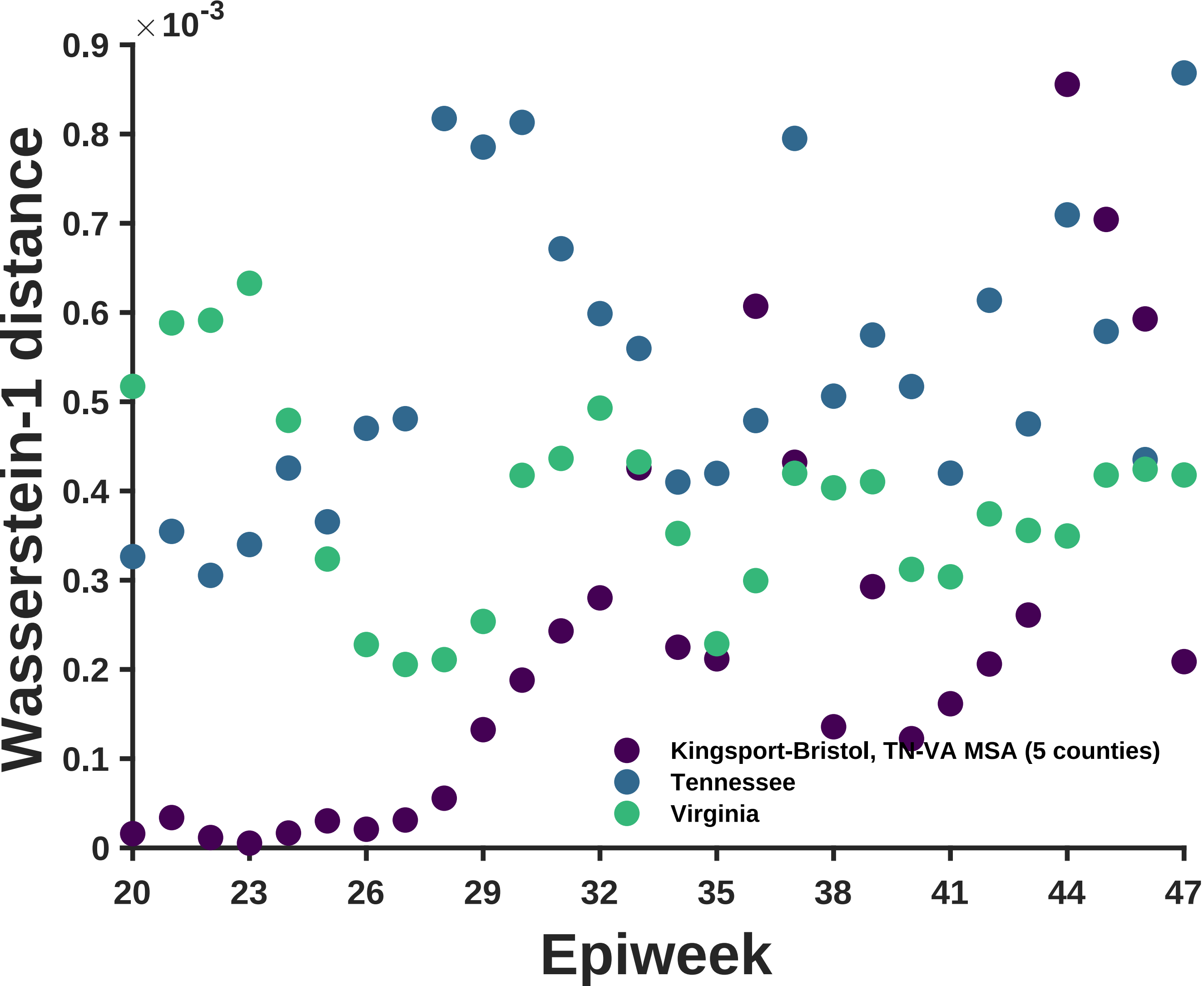
×10^-3
0.9
0.8
0.7
0.6
0.5
0.4
0.3
0.2
0.1
0
Wasserstein-1 distance
20
23
26
29
32
35
38
41
44
47
Epiweek
Kingsport-Bristol, TN-VA MSA (5 counties)
Tennessee
Virginia

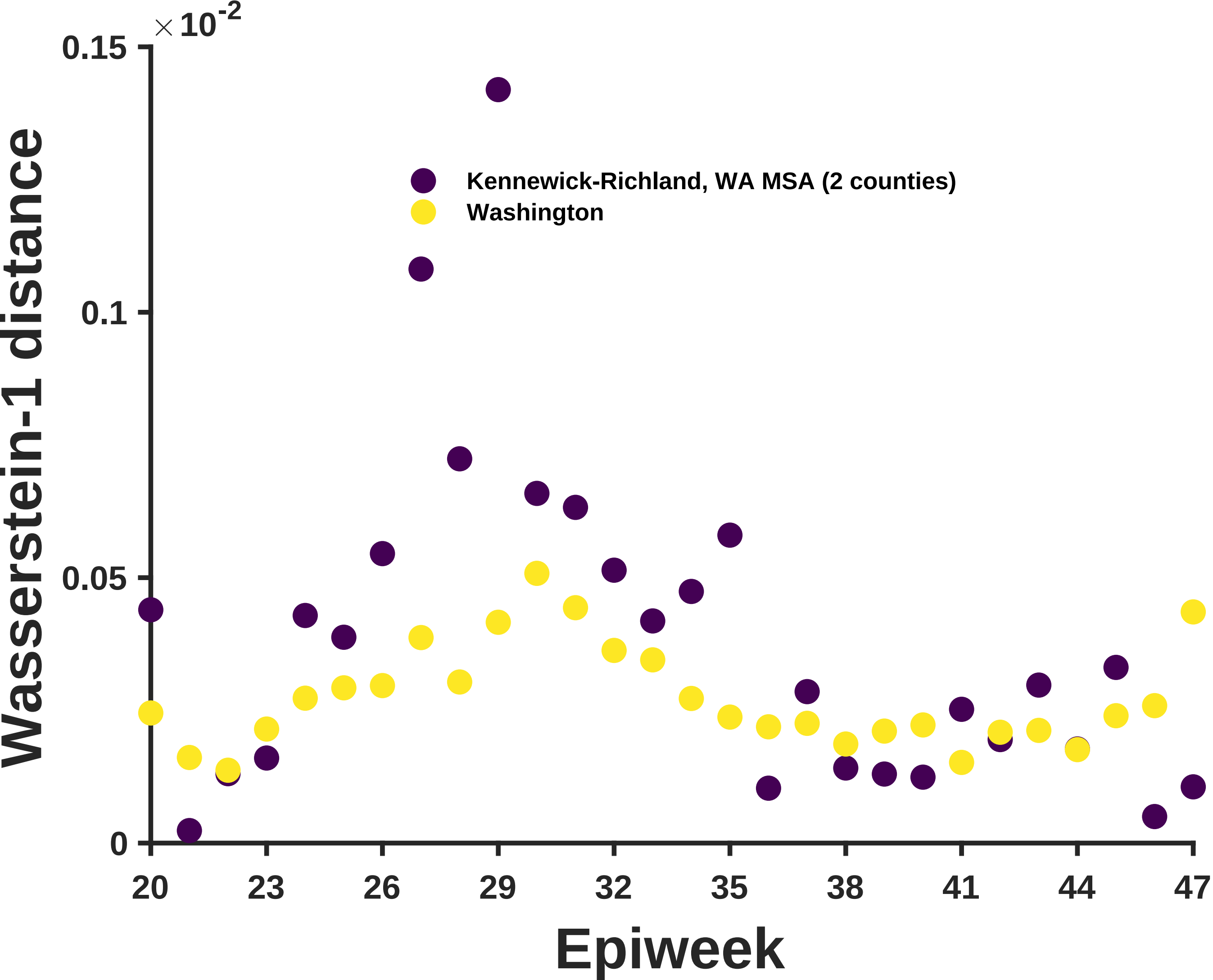

×10^-2
0.15
0.1
0.05
0
Wasserstein-1 distance
Kennewick-Richland, WA MSA (2 counties)
Washington
20
23
26
29
32
35
38
41
44
47
Epiweek

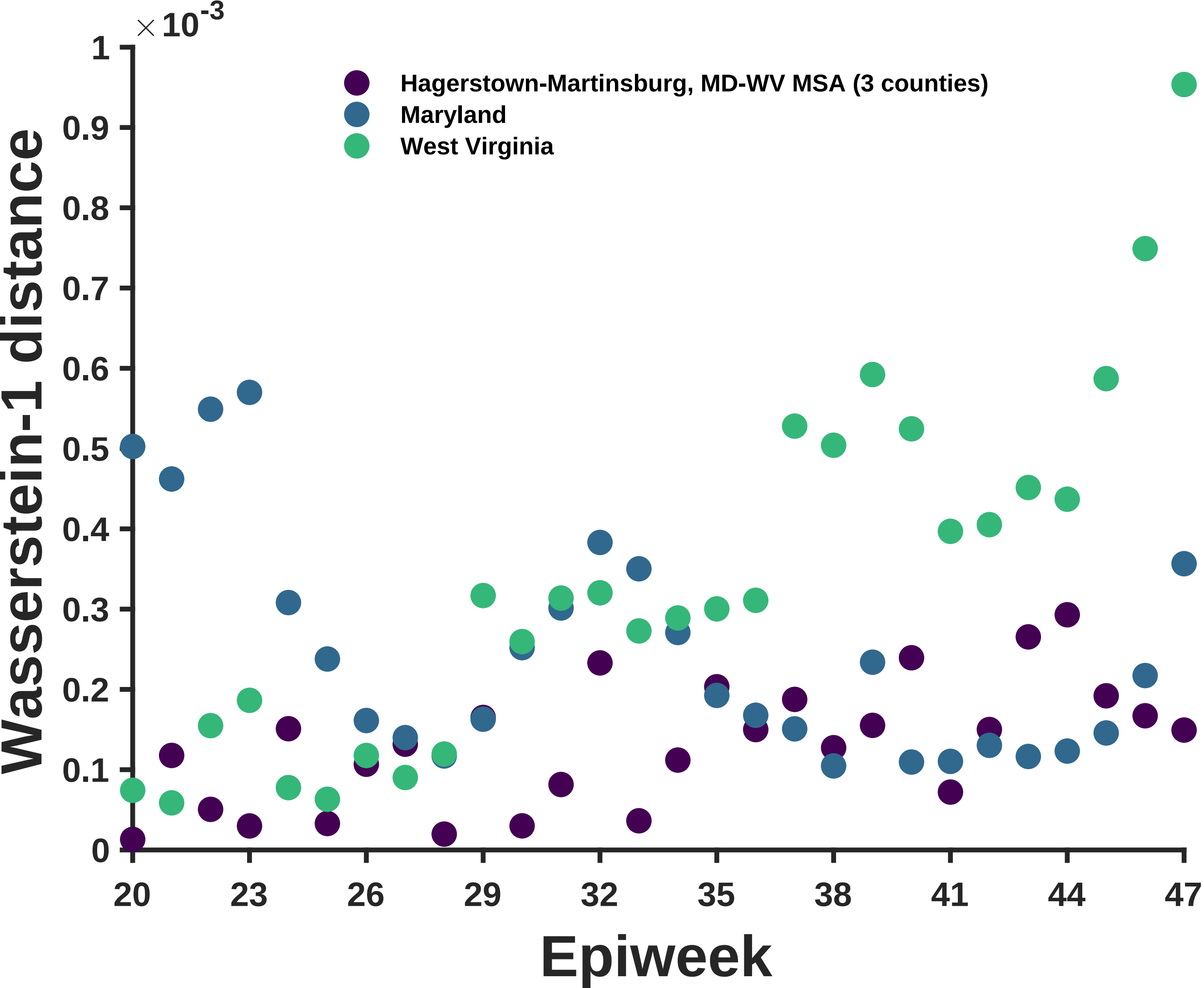

×10-3
Hagerstown-Martinsburg, MD-WV MSA (3 counties)
Maryland
West Virginia
Wasserstein-1 distance
Epiweek
0
0.1
0.2
0.3
0.4
0.5
0.6
0.7
0.8
0.9
1
20
23
26
29
32
35
38
41
44
47

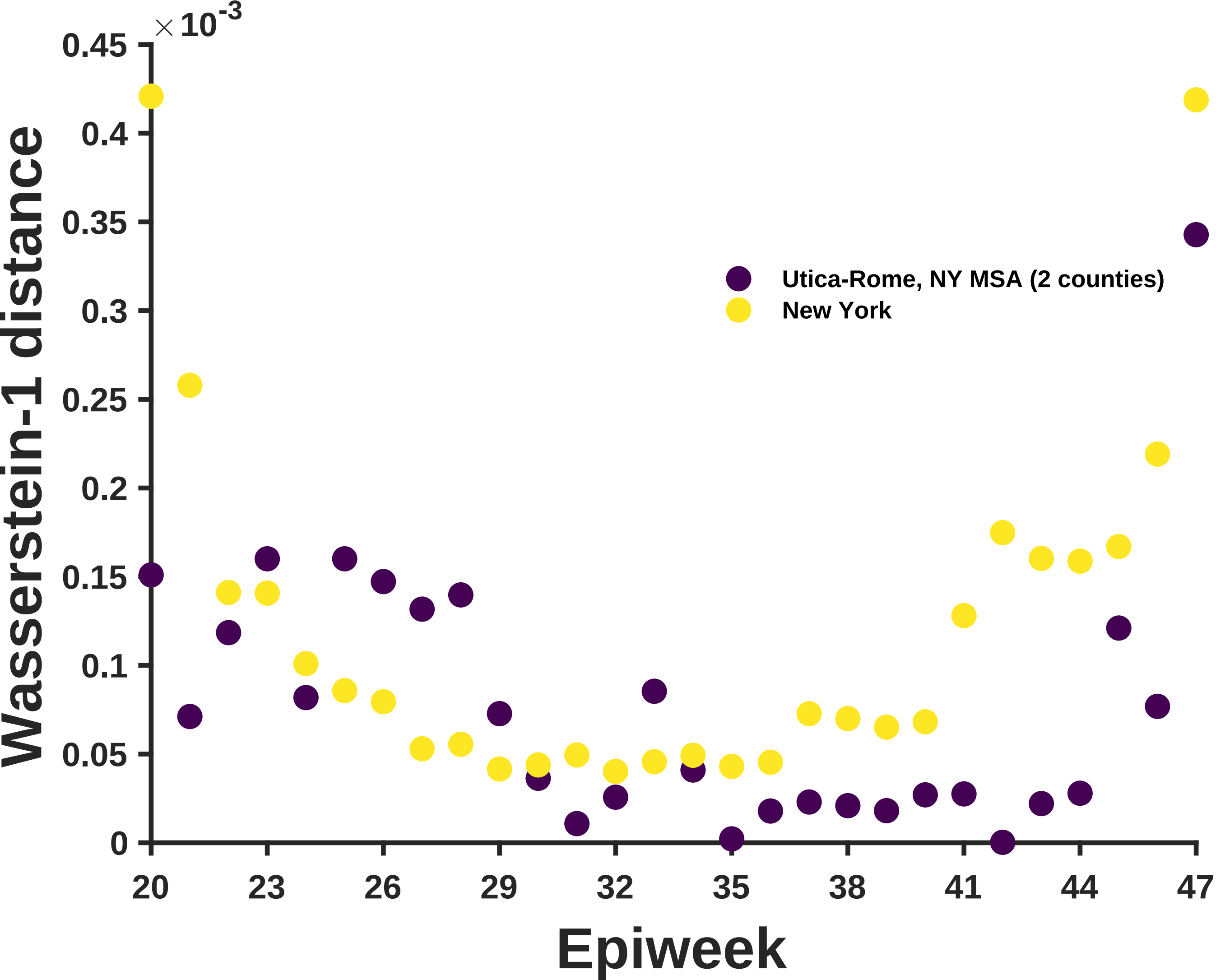
×10^-3
0.45
0.4
0.35
0.3
0.25
0.2
0.15
0.1
0.05
0
Wasserstein-1 distance
20
23
26
29
32
35
38
41
44
47
Epiweek
Utica-Rome, NY MSA (2 counties)
New York

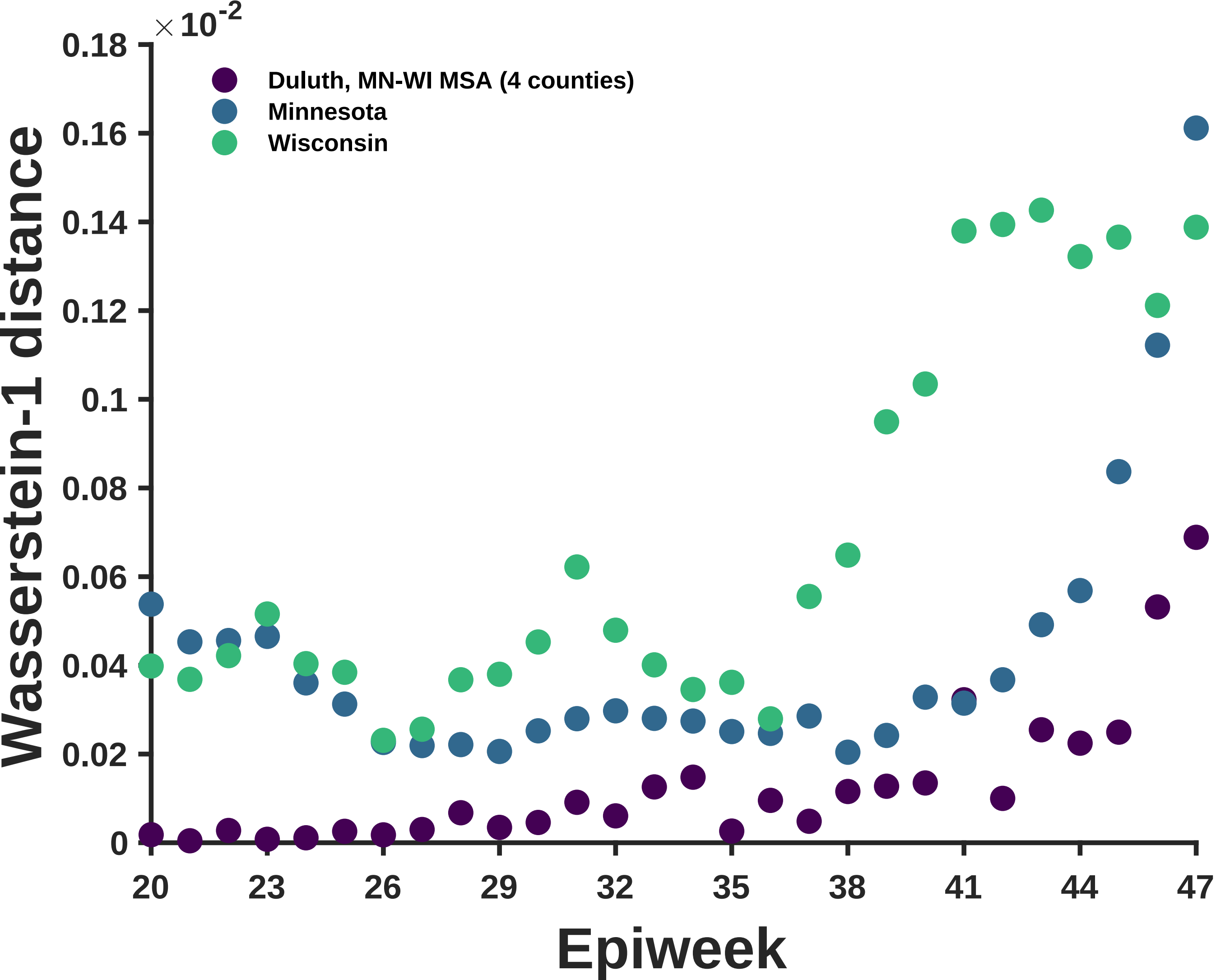

×10^-2
0.18
0.16
0.14
0.12
0.1
0.08
0.06
0.04
0.02
0
Wasserstein-1 distance
Duluth, MN-WI MSA (4 counties)
Minnesota
Wisconsin
20
23
26
29
32
35
38
41
44
47
Epiweek

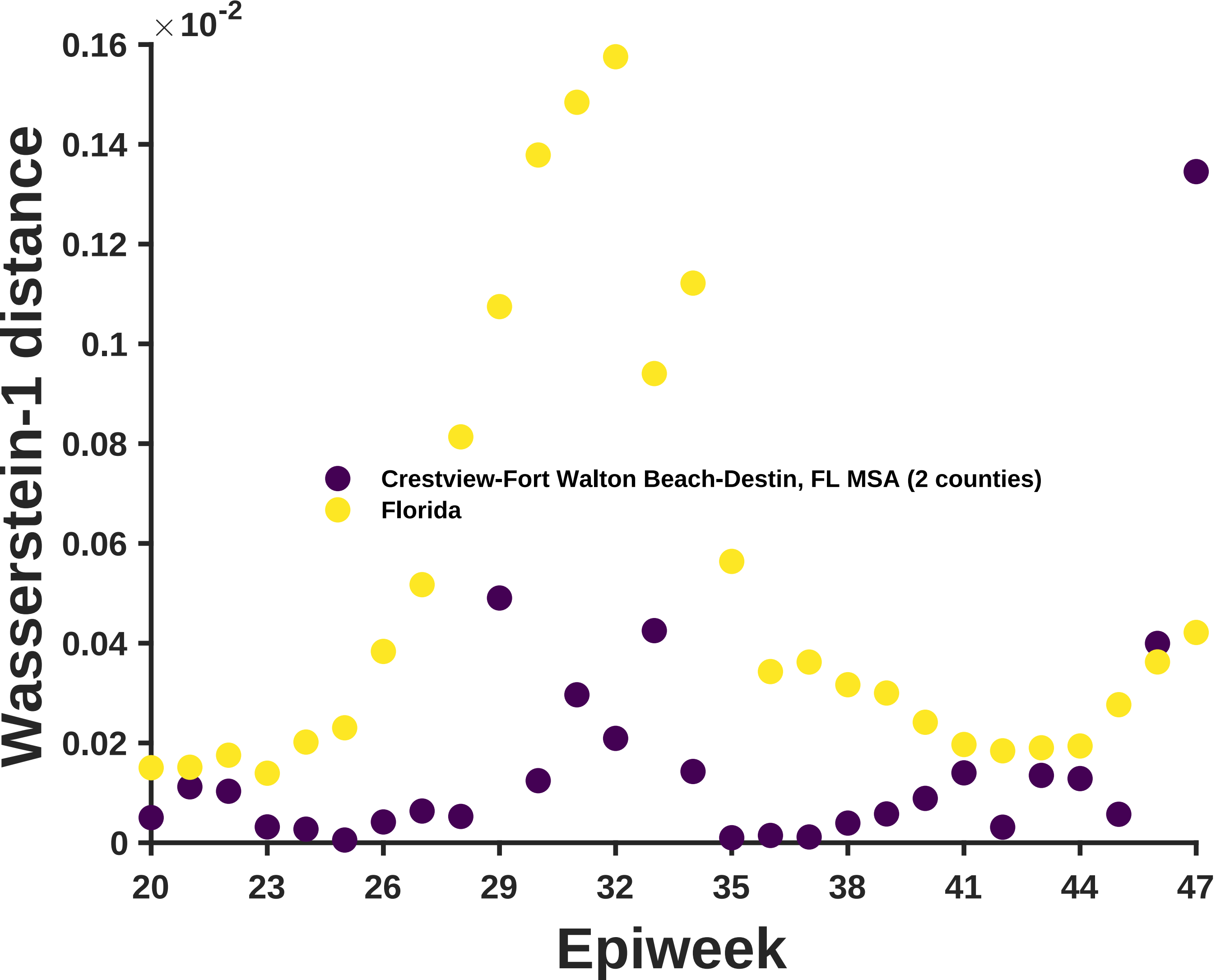

×10-2
0.16
0.14
0.12
0.1
0.08
0.06
0.04
0.02
0
Wasserstein-1 distance
Crestview-Fort Walton Beach-Destin, FL MSA (2 counties)
Florida
20
23
26
29
32
35
38
41
44
47
Epiweek

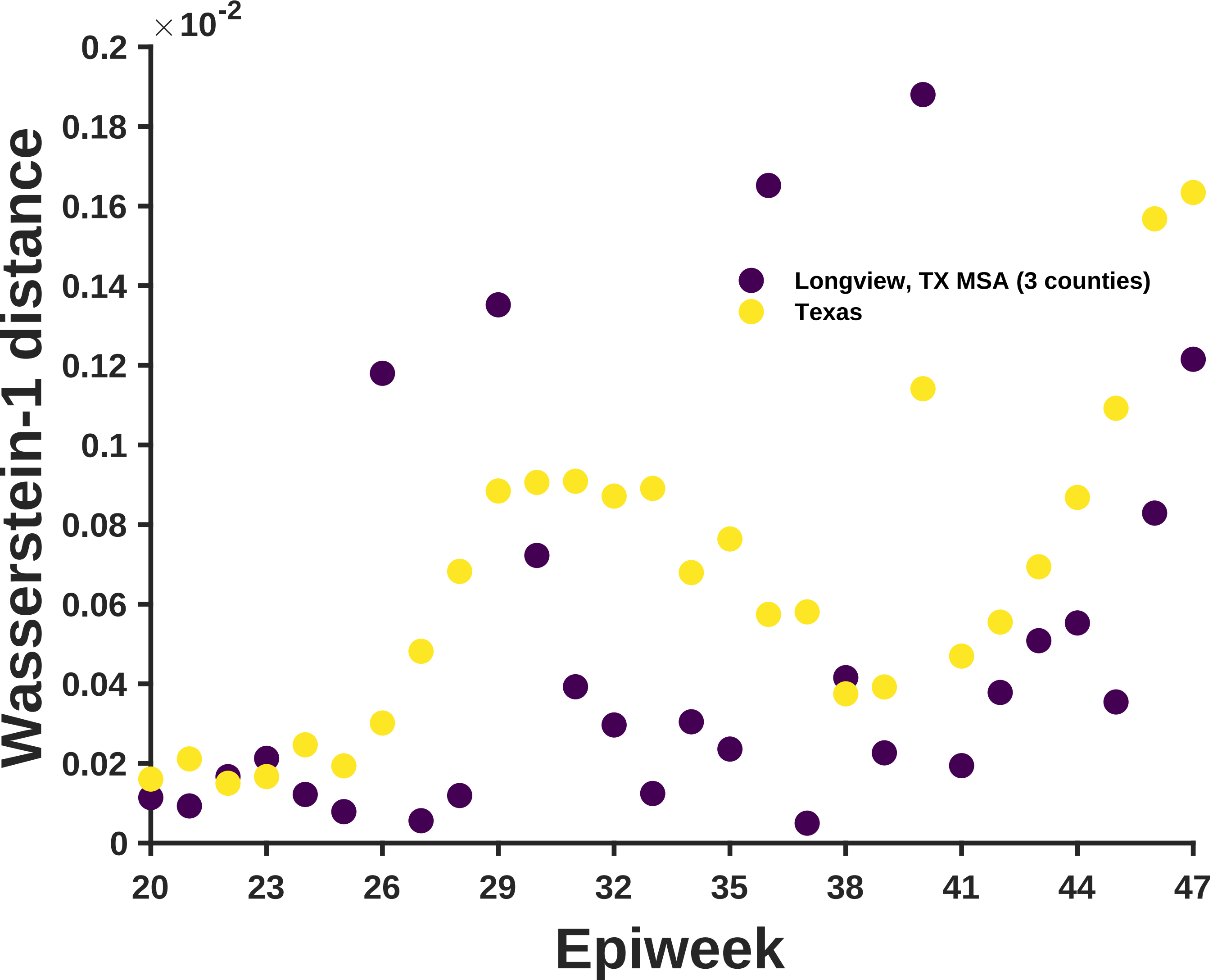

×10^-2
0.2
0.18
0.16
0.14
0.12
0.1
0.08
0.06
0.04
0.02
0
Wasserstein-1 distance
Longview, TX MSA (3 counties)
Texas
20
23
26
29
32
35
38
41
44
47
Epiweek

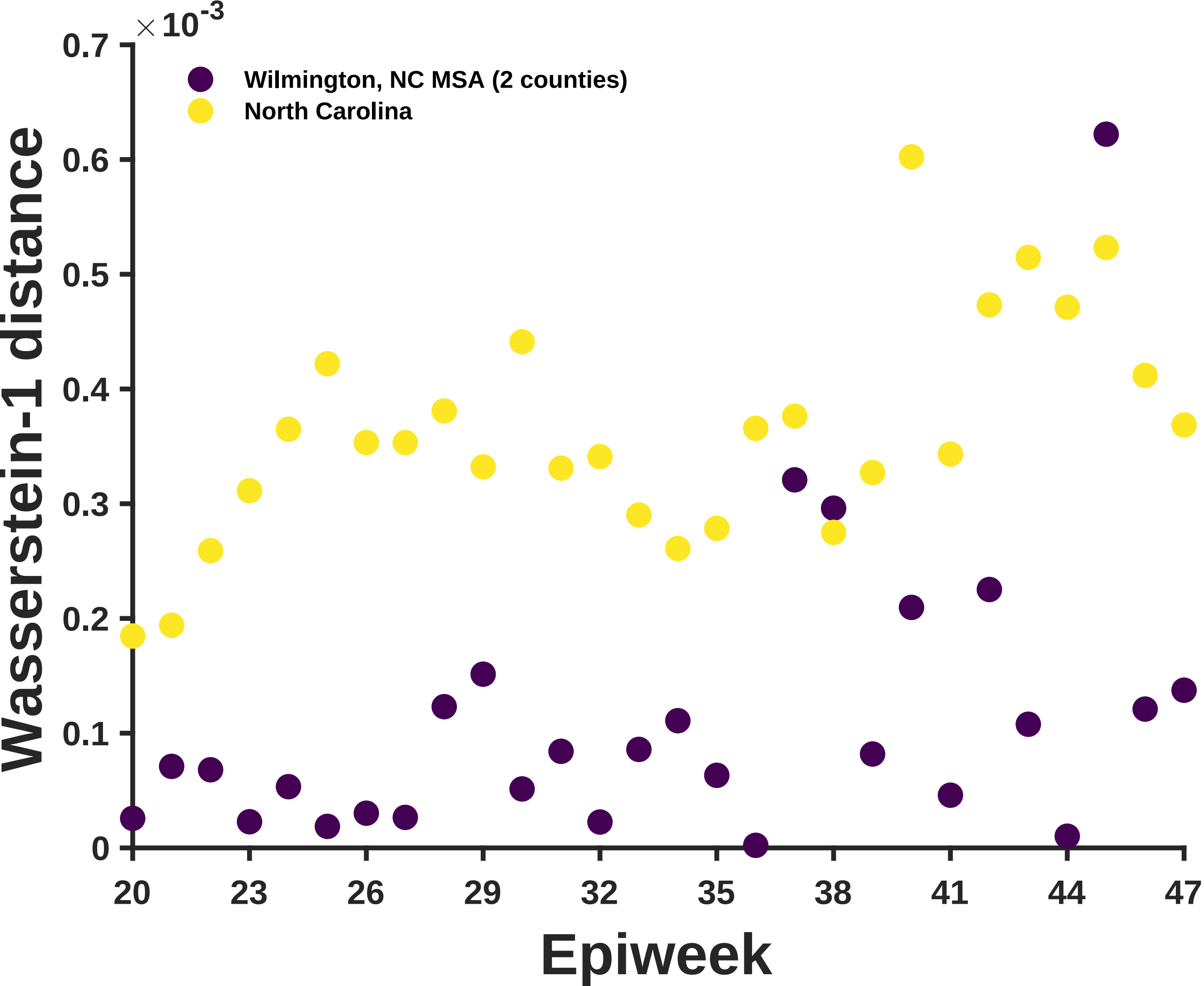

×10^-3
0.7
0.6
0.5
0.4
0.3
0.2
0.1
0
Wilmington, NC MSA (2 counties)
North Carolina
Wasserstein-1 distance
20
23
26
29
32
35
38
41
44
47
Epiweek

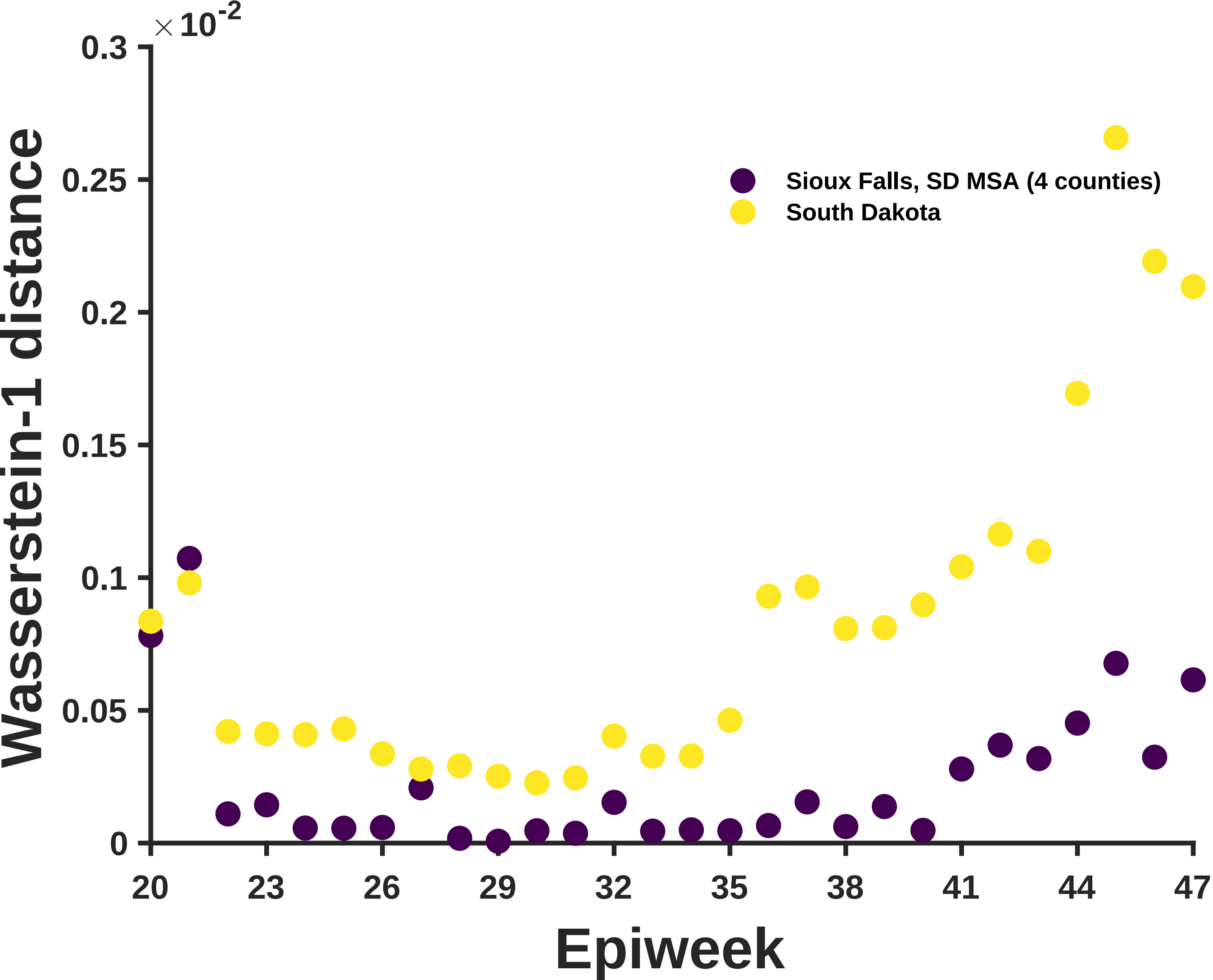

×10^-2
0.3
0.25
0.2
0.15
0.1
0.05
0
Wasserstein-1 distance
Sioux Falls, SD MSA (4 counties)
South Dakota
20
23
26
29
32
35
38
41
44
47
Epiweek

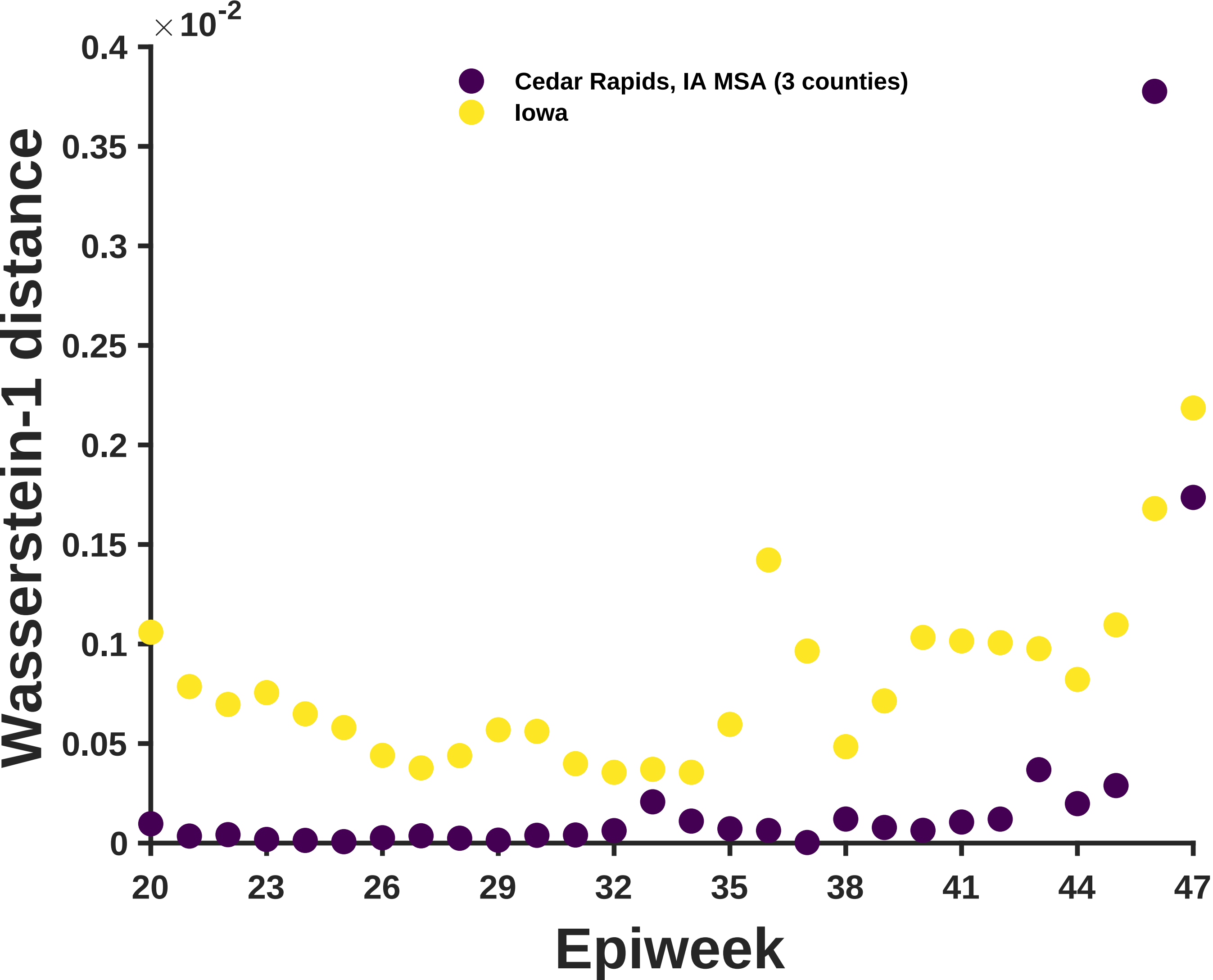

×10^-2
0.4
0.35
0.3
0.25
0.2
0.15
0.1
0.05
0
Wasserstein-1 distance
Cedar Rapids, IA MSA (3 counties)
Iowa
20
23
26
29
32
35
38
41
44
47
Epiweek

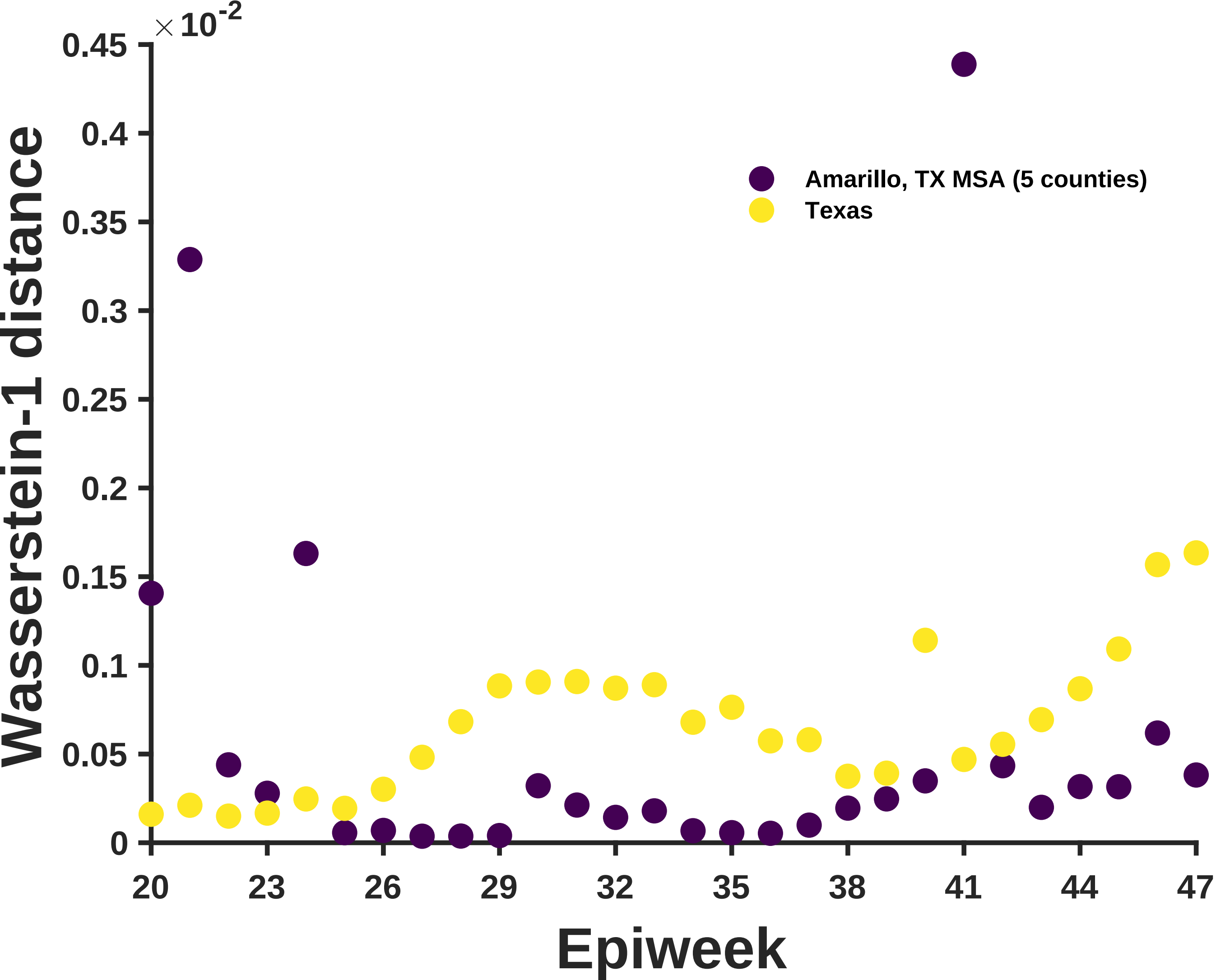
×10^-2
0.45
0.4
0.35
0.3
0.25
0.2
0.15
0.1
0.05
0
Wasserstein-1 distance
20
23
26
29
32
35
38
41
44
47
Epiweek
Amarillo, TX MSA (5 counties)
Texas

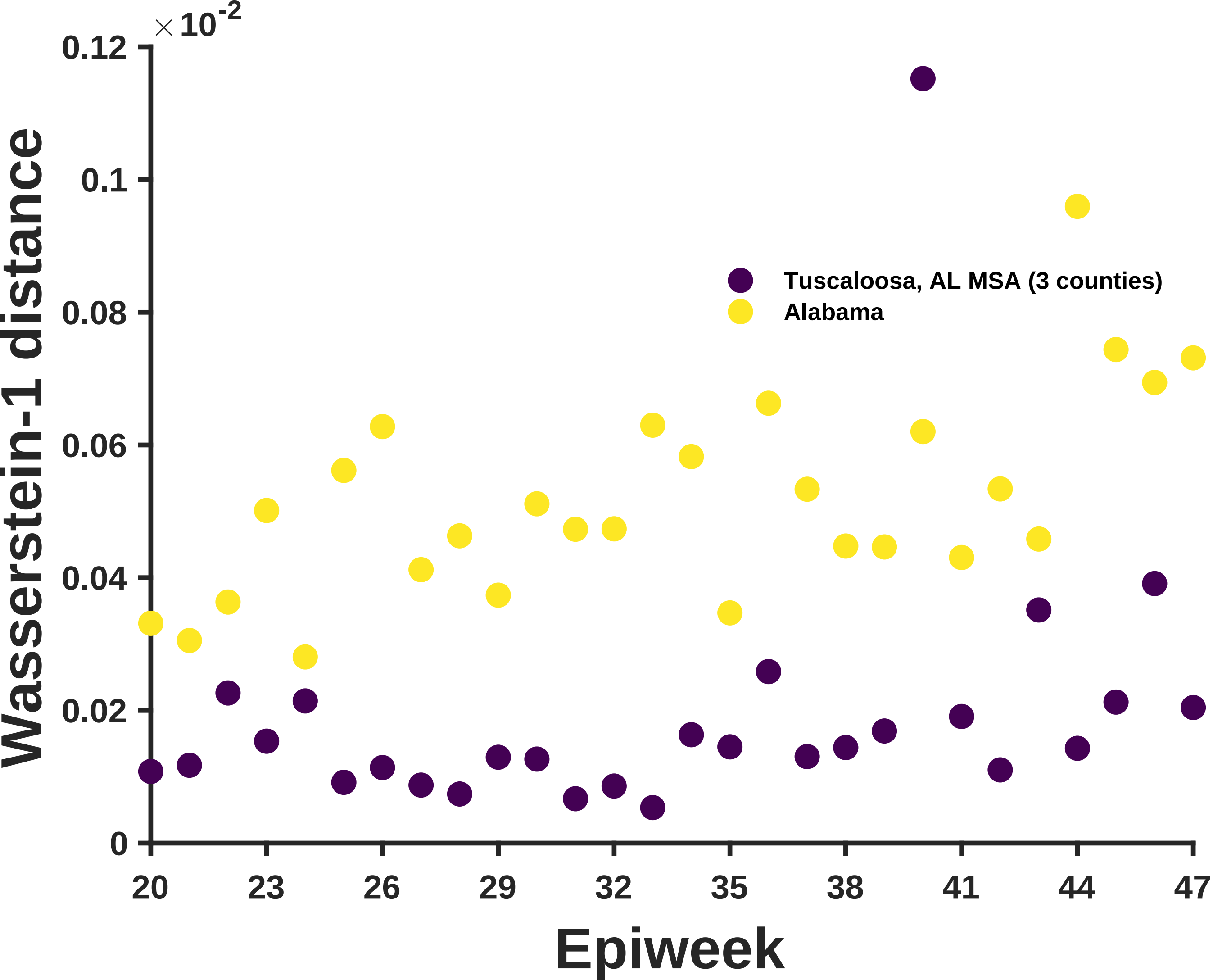
×10^-2
0.12
0.1
0.08
0.06
0.04
0.02
0
Wasserstein-1 distance
Tuscaloosa, AL MSA (3 counties)
Alabama
20
23
26
29
32
35
38
41
44
47
Epiweek

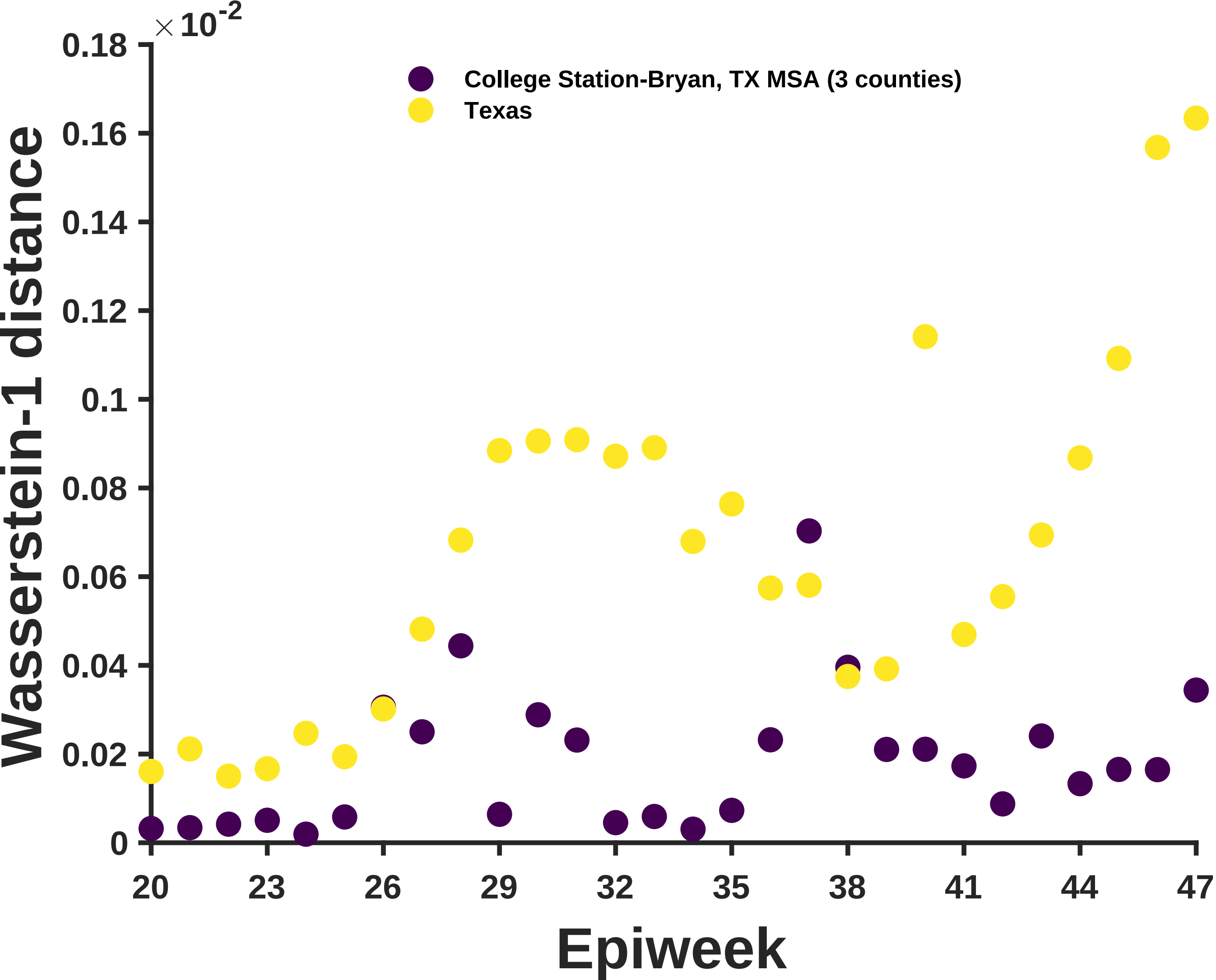
×10^-2
0.18
0.16
0.14
0.12
0.1
0.08
0.06
0.04
0.02
0
College Station-Bryan, TX MSA (3 counties)
Texas
Wasserstein-1 distance
20
23
26
29
32
35
38
41
44
47
Epiweek

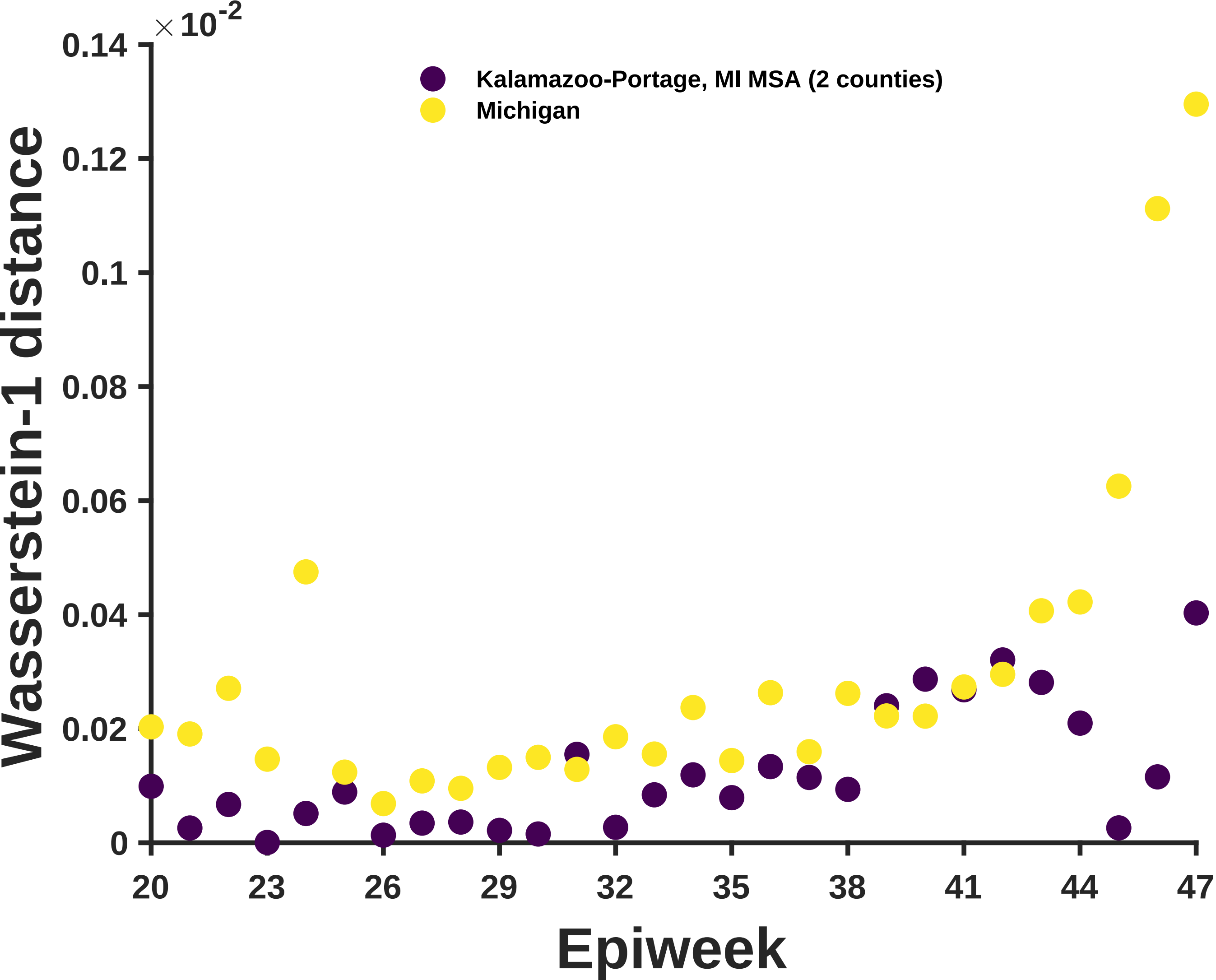

×10^-2
0.14
0.12
0.1
0.08
0.06
0.04
0.02
0
Wasserstein-1 distance
Kalamazoo-Portage, MI MSA (2 counties)
Michigan
20
23
26
29
32
35
38
41
44
47
Epiweek

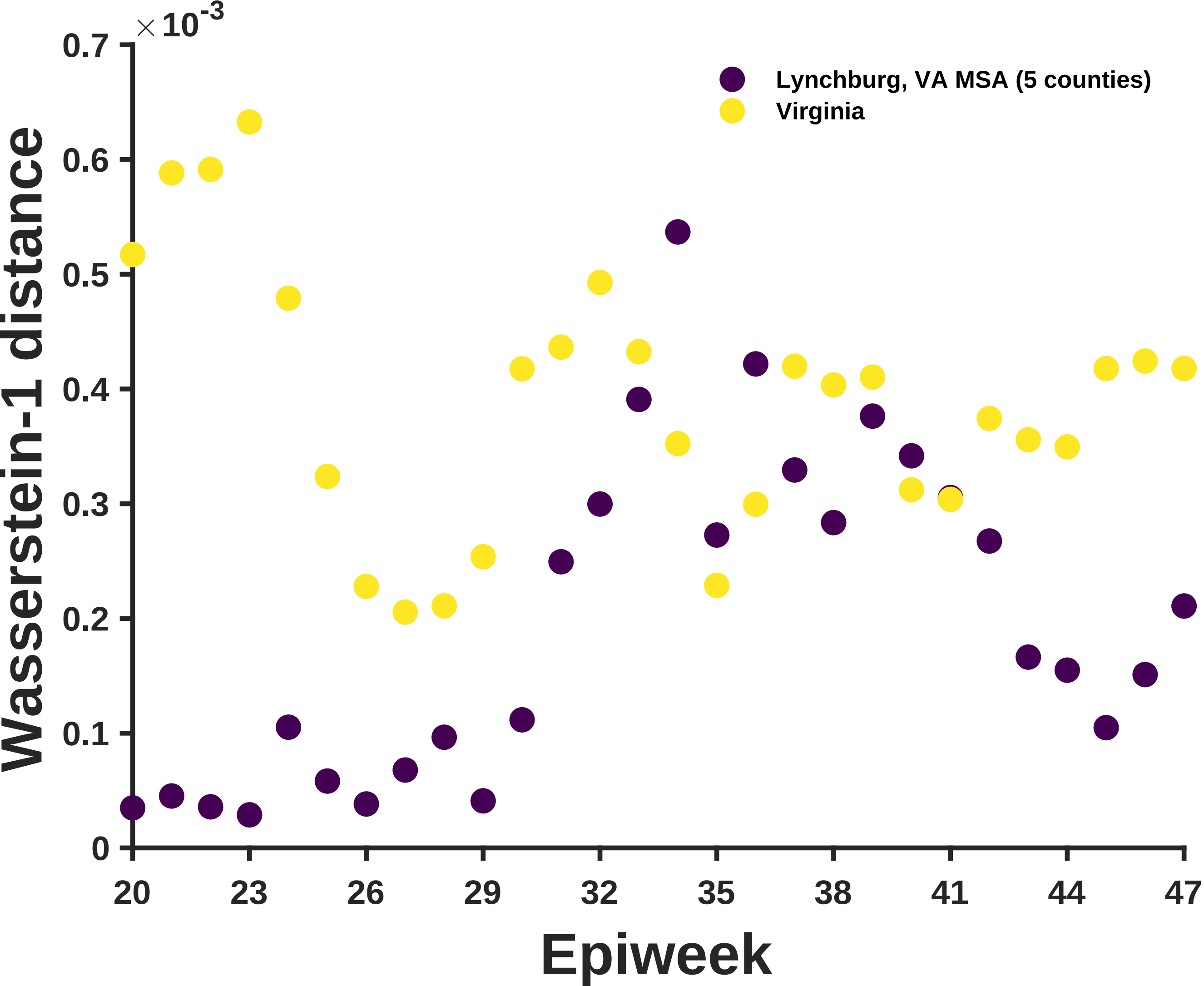

×10^-3
Wasserstein-1 distance
0
0.1
0.2
0.3
0.4
0.5
0.6
0.7
Epiweek
20
23
26
29
32
35
38
41
44
47
Lynchburg, VA MSA (5 counties)
Virginia

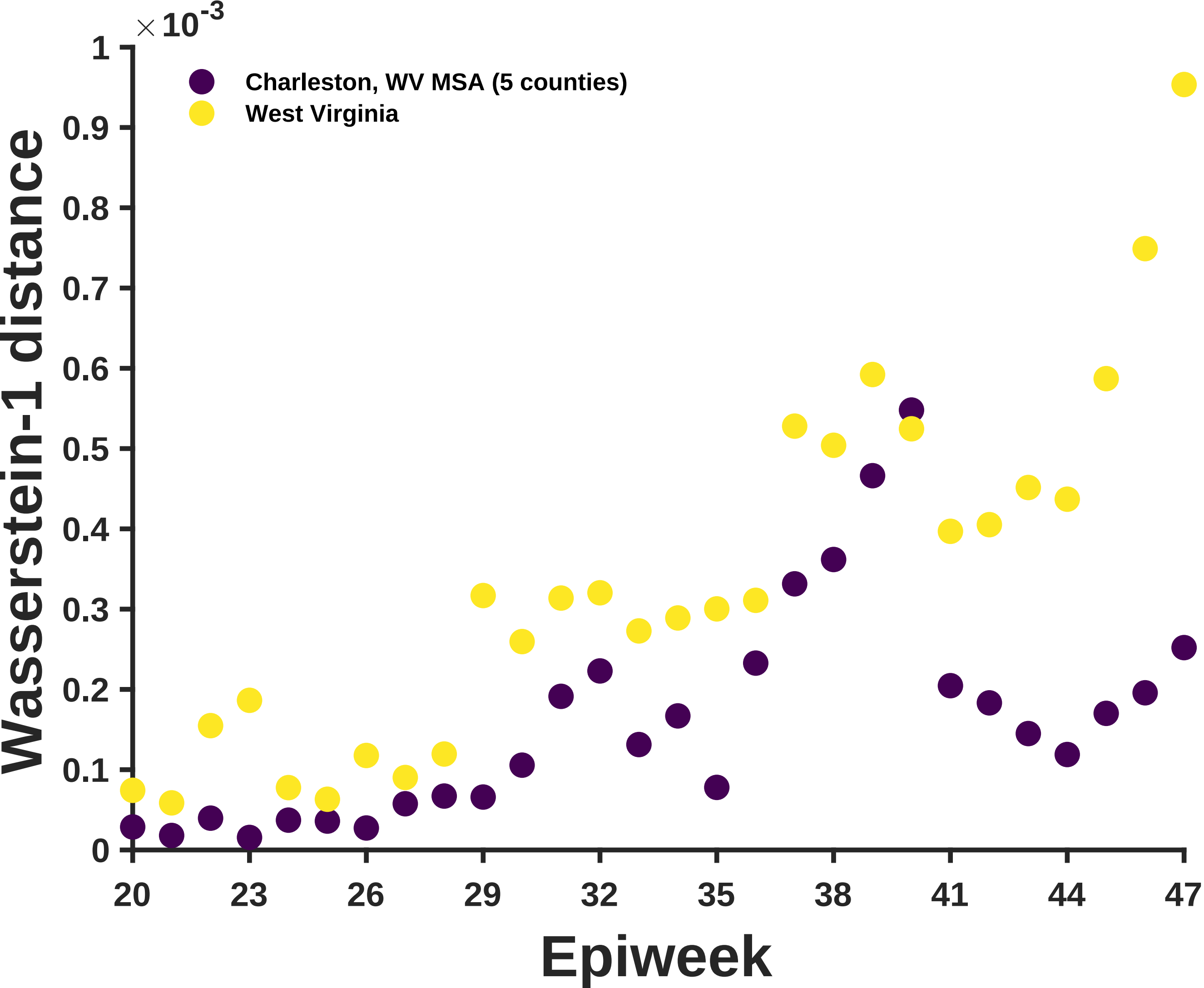

×10^-3
Charleston, WV MSA (5 counties)
West Virginia
Wasserstein-1 distance
Epiweek
0
0.1
0.2
0.3
0.4
0.5
0.6
0.7
0.8
0.9
1
20
23
26
29
32
35
38
41
44
47

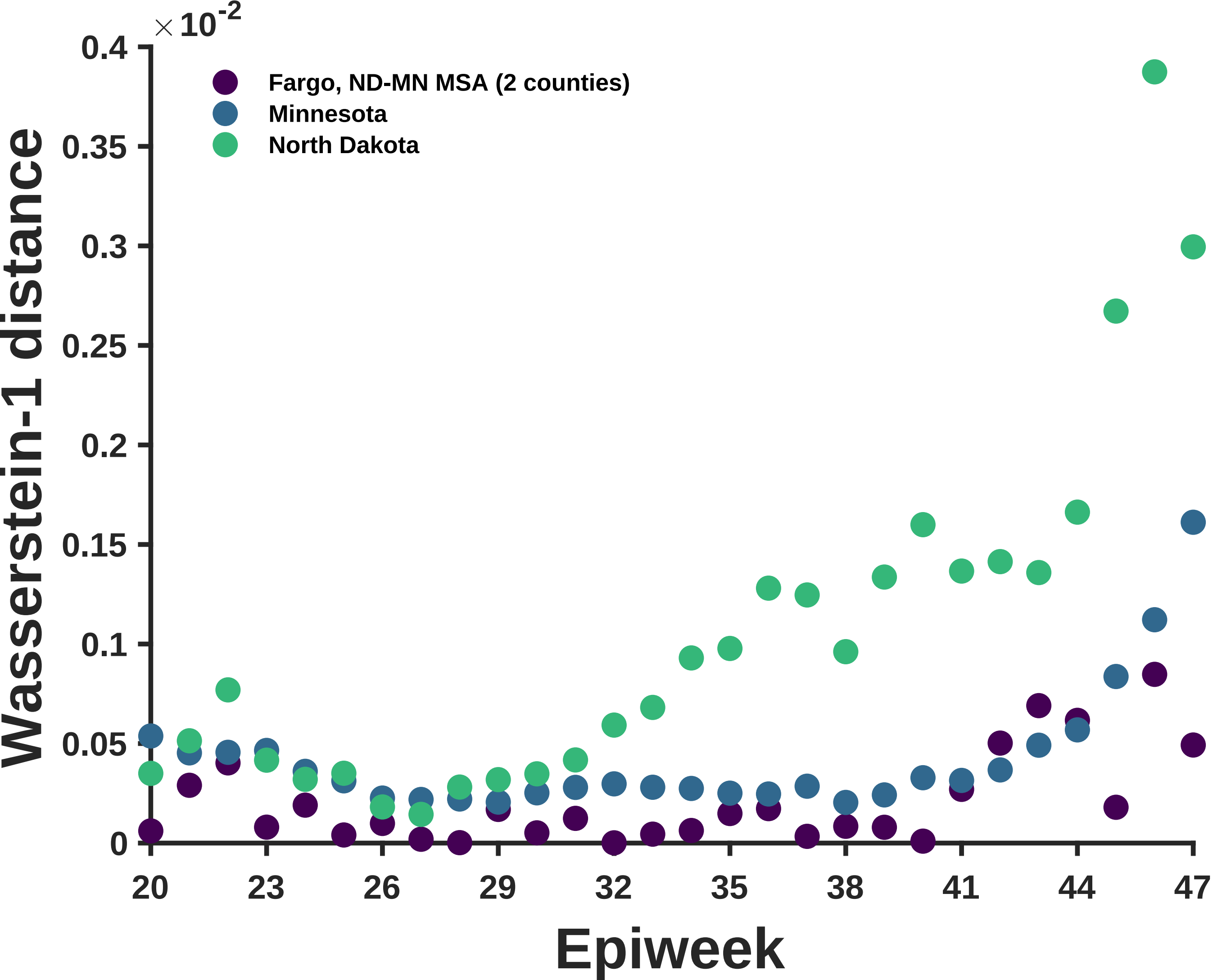

×10^-2
0.4
0.35
0.3
0.25
0.2
0.15
0.1
0.05
0
Wasserstein-1 distance
Fargo, ND-MN MSA (2 counties)
Minnesota
North Dakota
20
23
26
29
32
35
38
41
44
47
Epiweek

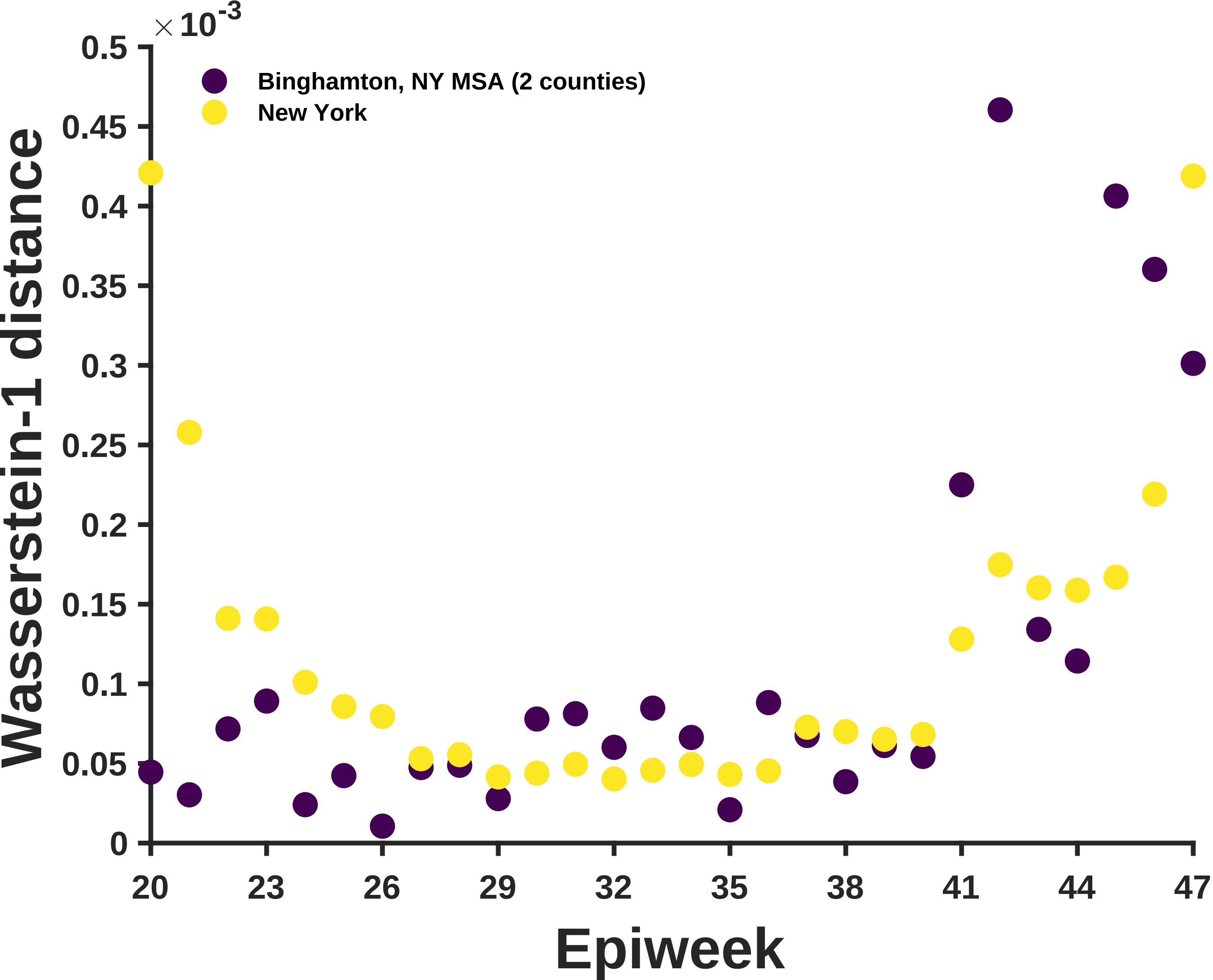

×10-3
Binghamton, NY MSA (2 counties)
New York
Wasserstein-1 distance
0.5
0.45
0.4
0.35
0.3
0.25
0.2
0.15
0.1
0.05
0
20
23
26
29
32
35
38
41
44
47
Epiweek

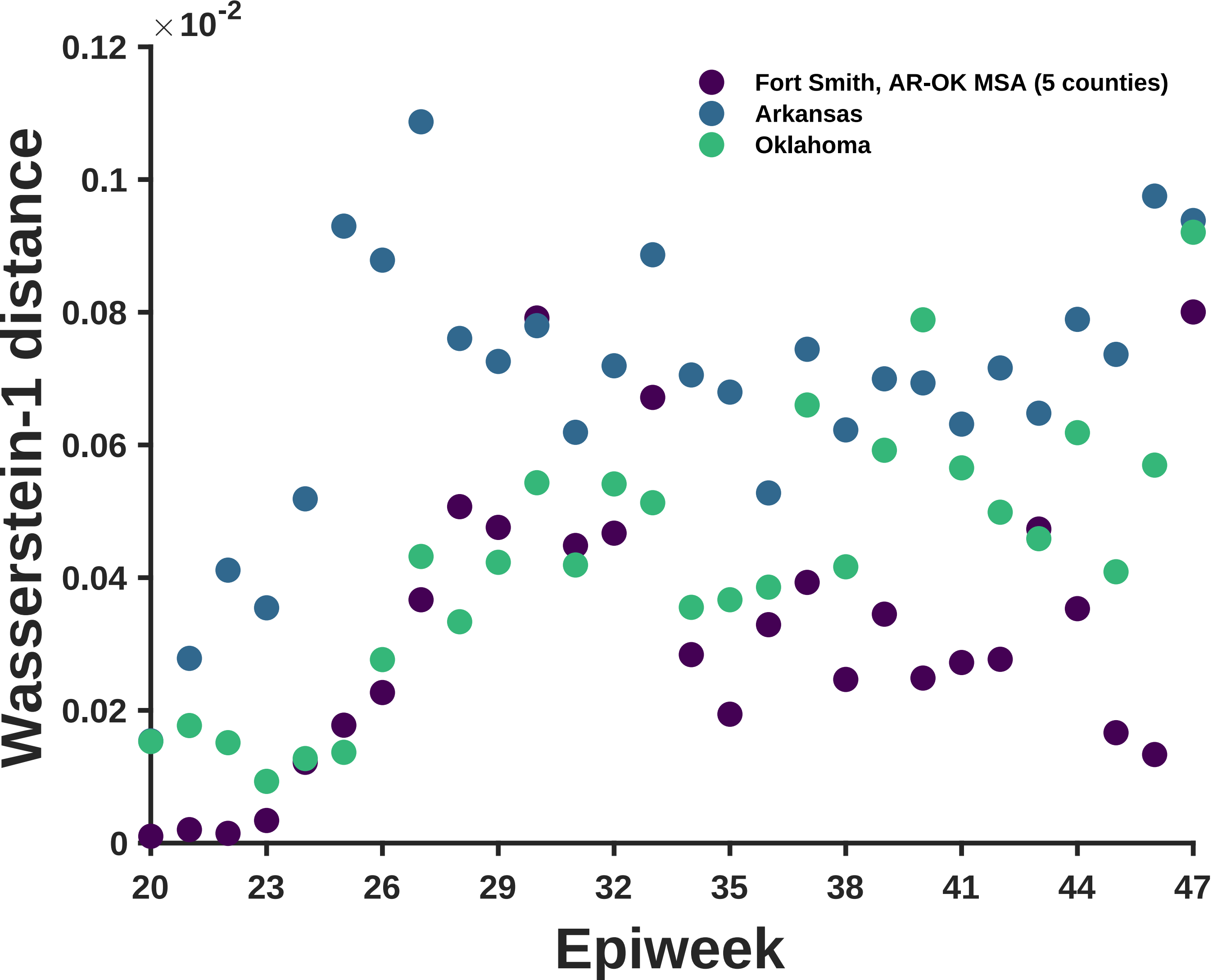
×10^-2
0.12
0.1
0.08
0.06
0.04
0.02
0
Wasserstein-1 distance
Fort Smith, AR-OK MSA (5 counties)
Arkansas
Oklahoma
20
23
26
29
32
35
38
41
44
47
Epiweek

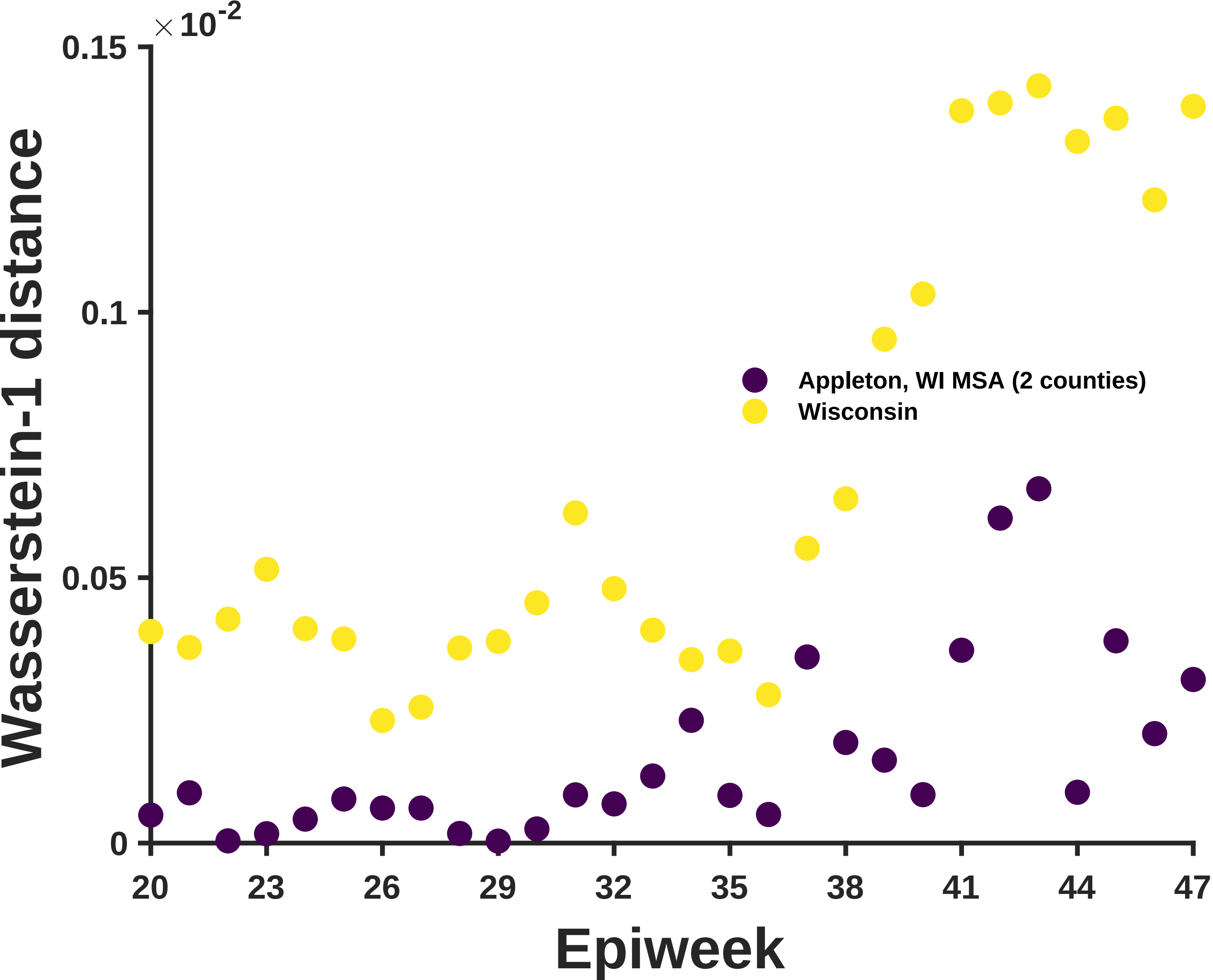
×10^-2
0.15
0.1
0.05
0
Wasserstein-1 distance
20
23
26
29
32
35
38
41
44
47
Epiweek
Appleton, WI MSA (2 counties)
Wisconsin

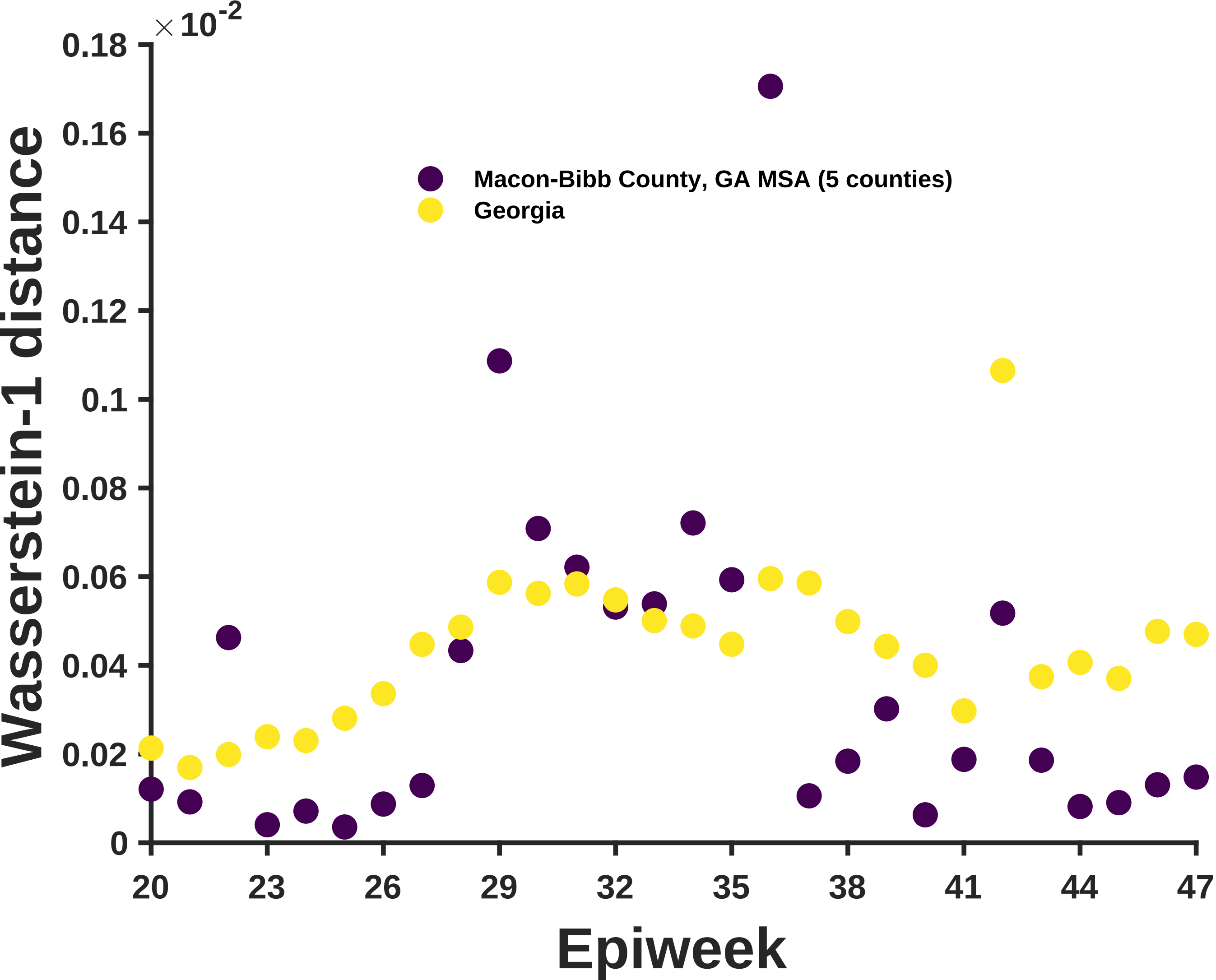
×10^-2
0.18
0.16
0.14
0.12
0.1
0.08
0.06
0.04
0.02
0
Wasserstein-1 distance
Macon-Bibb County, GA MSA (5 counties)
Georgia
20
23
26
29
32
35
38
41
44
47
Epiweek

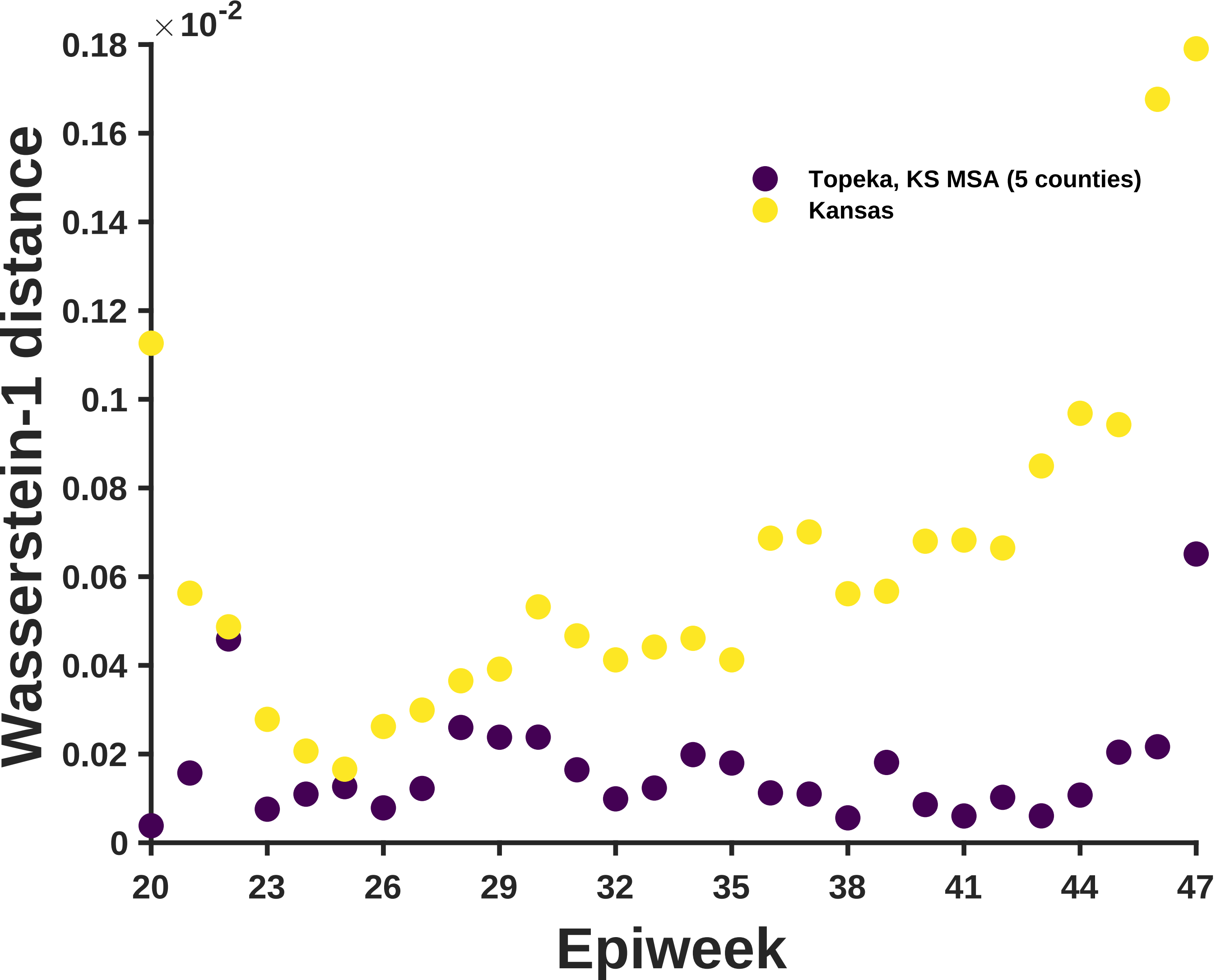

×10^-2
0.18
0.16
0.14
0.12
0.1
0.08
0.06
0.04
0.02
0
Wasserstein-1 distance
Topeka, KS MSA (5 counties)
Kansas
20
23
26
29
32
35
38
41
44
47
Epiweek

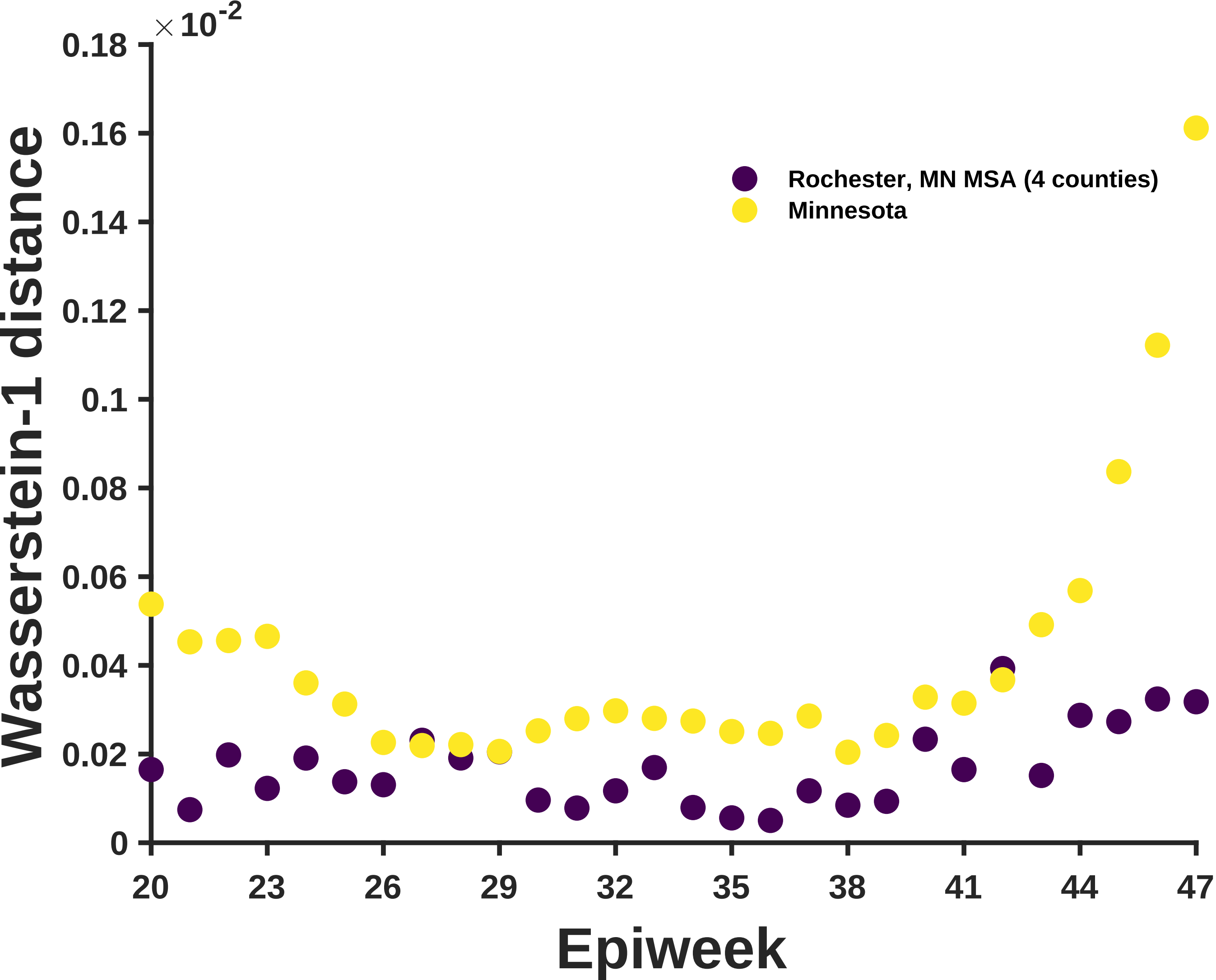

×10^-2
0.18
0.16
0.14
0.12
0.1
0.08
0.06
0.04
0.02
0
Wasserstein-1 distance
Rochester, MN MSA (4 counties)
Minnesota
20
23
26
29
32
35
38
41
44
47
Epiweek

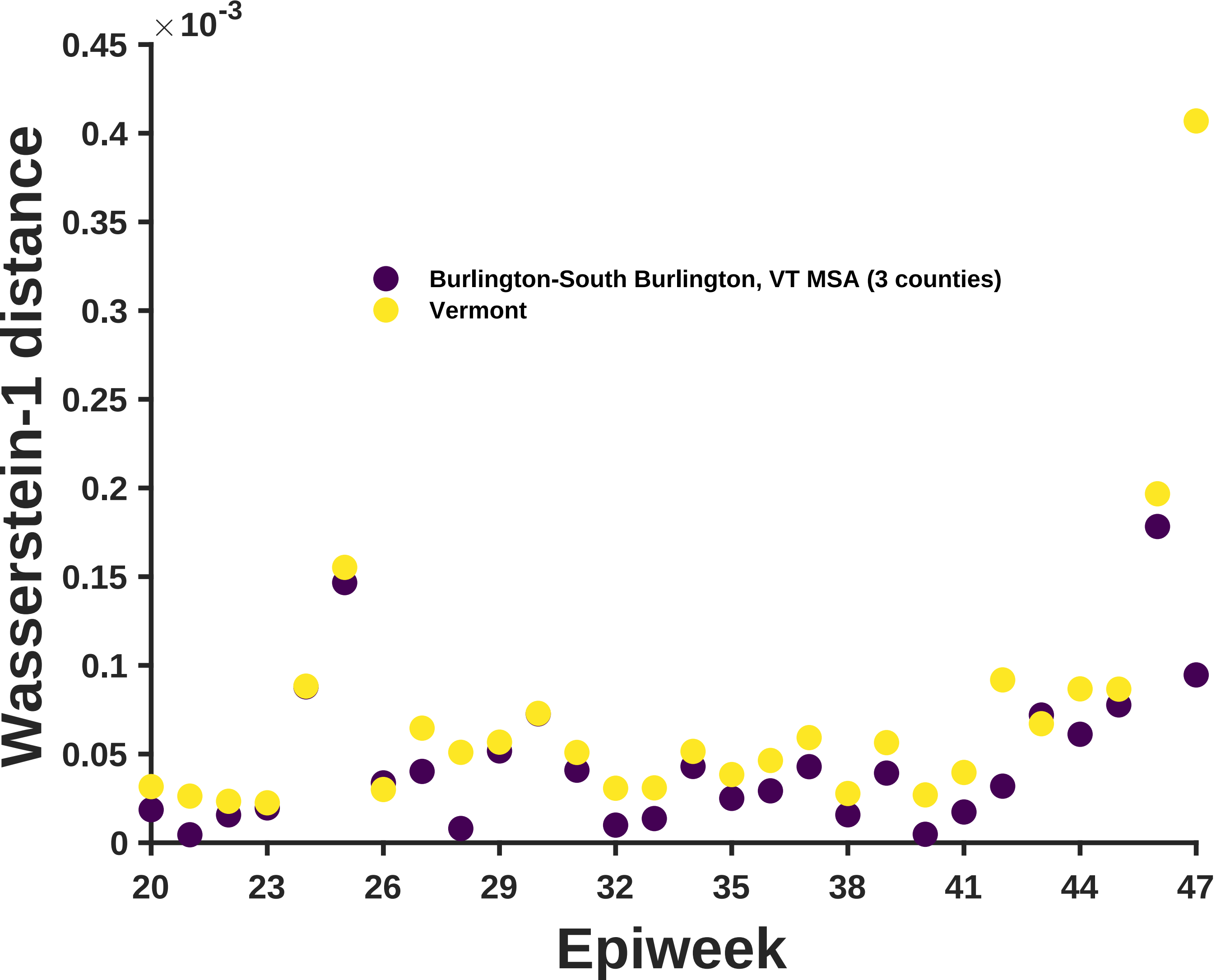

×10^-3
0.45
0.4
0.35
0.3
0.25
0.2
0.15
0.1
0.05
0
Wasserstein-1 distance
Burlington-South Burlington, VT MSA (3 counties)
Vermont
20
23
26
29
32
35
38
41
44
47
Epiweek

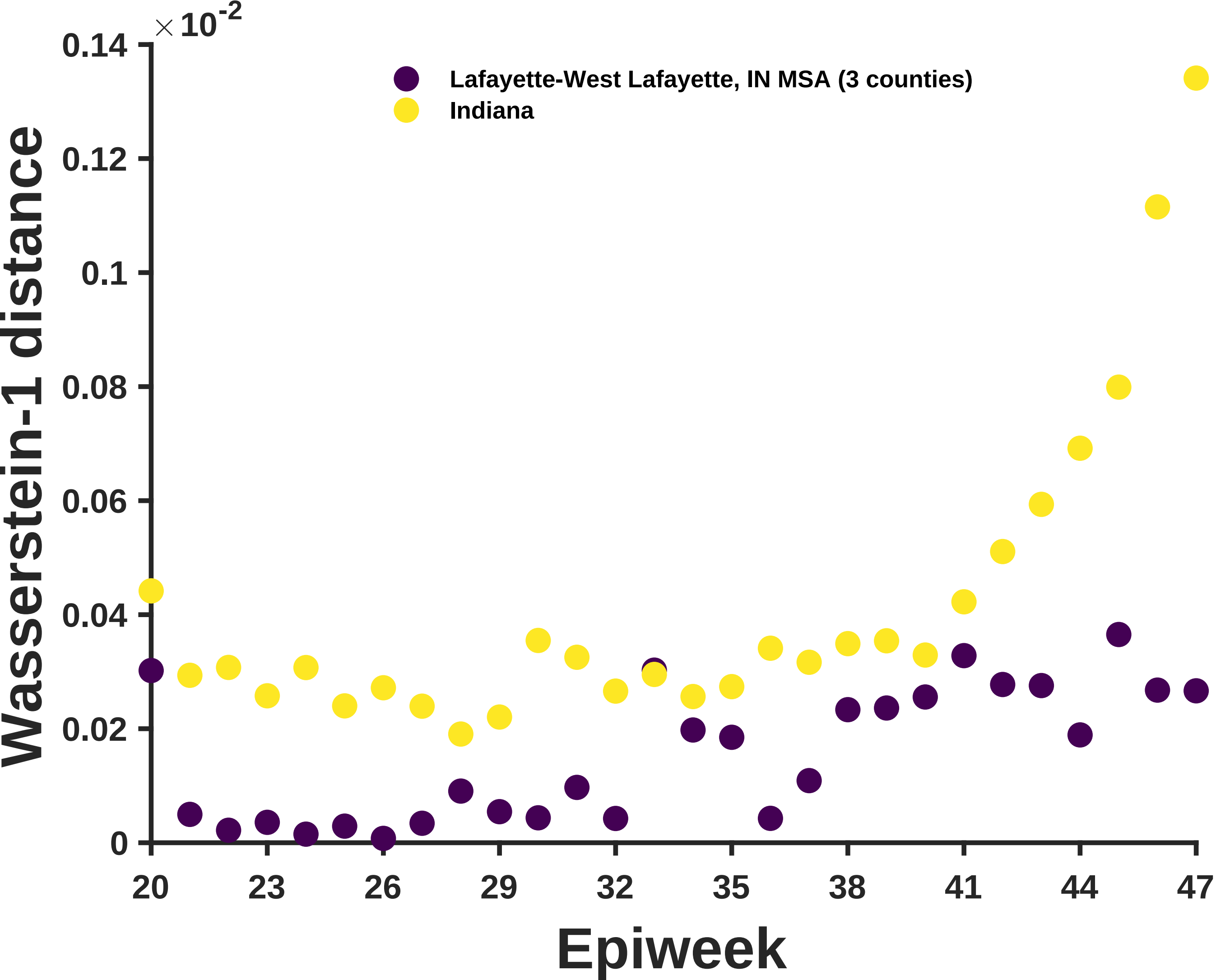

$\times 10^{-2}$
0.14
0.12
0.1
0.08
0.06
0.04
0.02
0
Wasserstein-1 distance
Lafayette-West Lafayette, IN MSA (3 counties)
Indiana
20
23
26
29
32
35
38
41
44
47
Epiweek

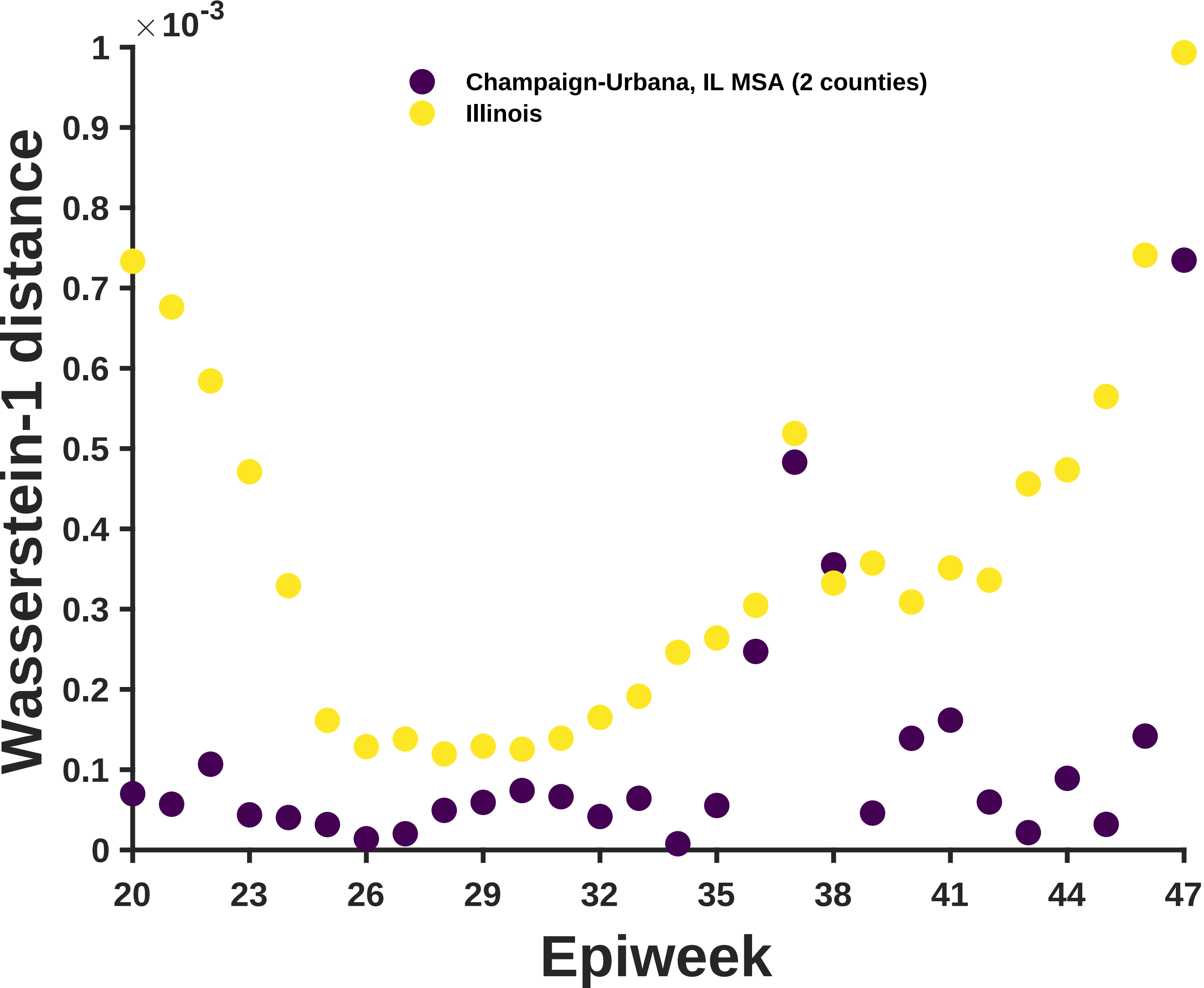

Champaign-Urbana, IL MSA (2 counties)
Illinois
Wasserstein-1 distance
Epiweek
×10^-3
0
0.1
0.2
0.3
0.4
0.5
0.6
0.7
0.8
0.9
1
20
23
26
29
32
35
38
41
44
47

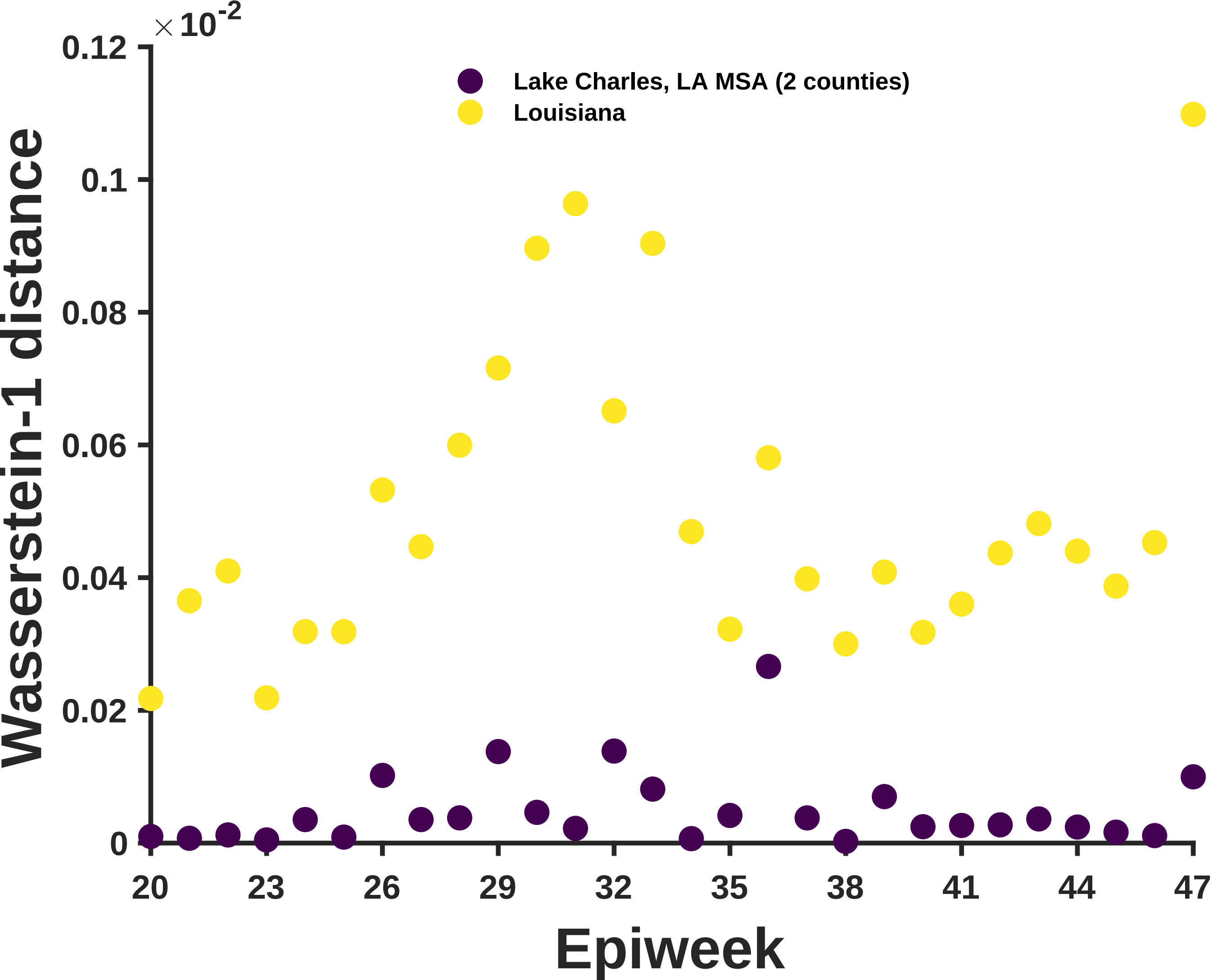

×10^-2
0.12
0.1
0.08
0.06
0.04
0.02
0
Wasserstein-1 distance
Lake Charles, LA MSA (2 counties)
Louisiana
20
23
26
29
32
35
38
41
44
47
Epiweek

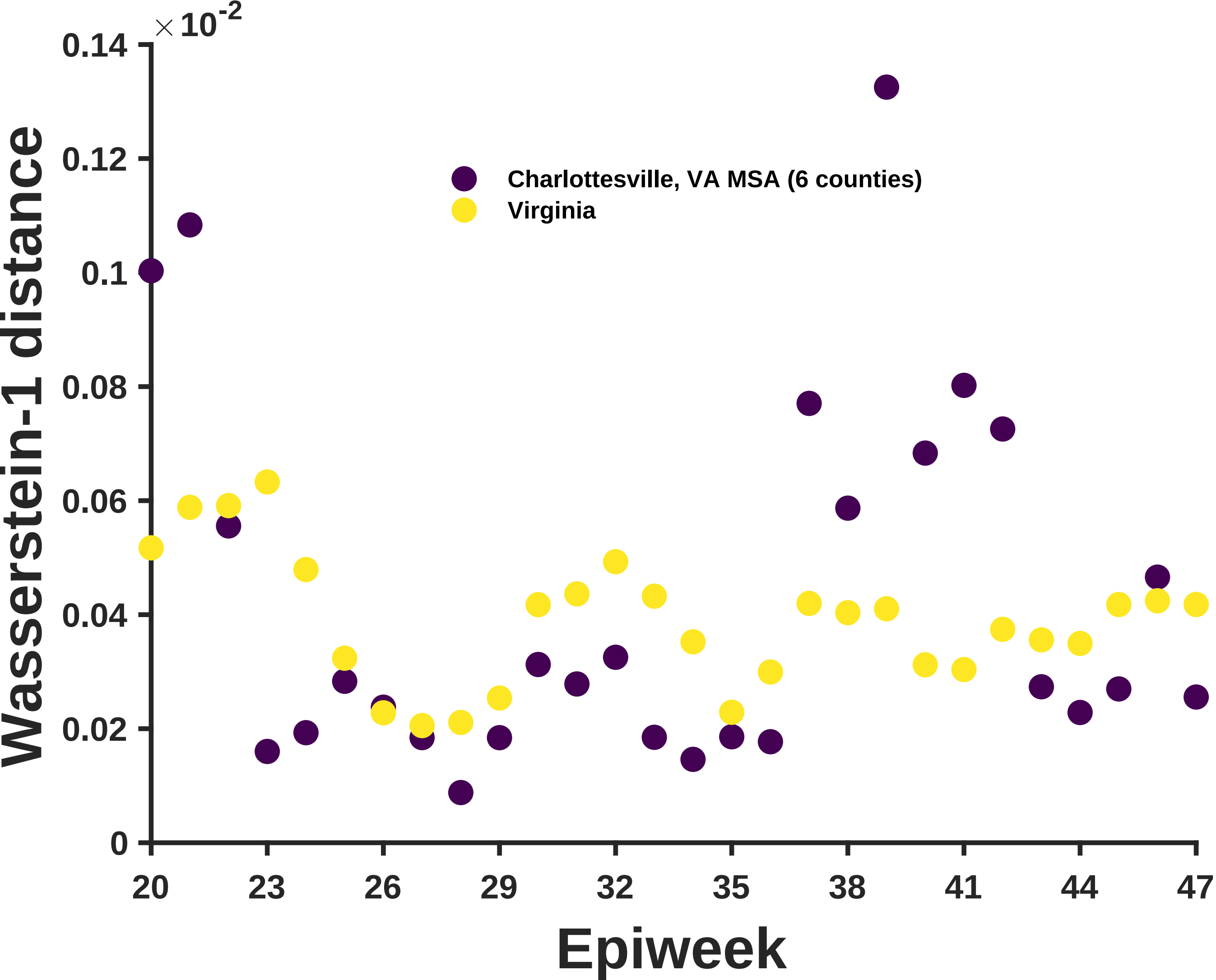

$\times 10^{-2}$
0.14
0.12
0.1
0.08
0.06
0.04
0.02
0
Wasserstein-1 distance
Charlottesville, VA MSA (6 counties)
Virginia
20
23
26
29
32
35
38
41
44
47
Epiweek

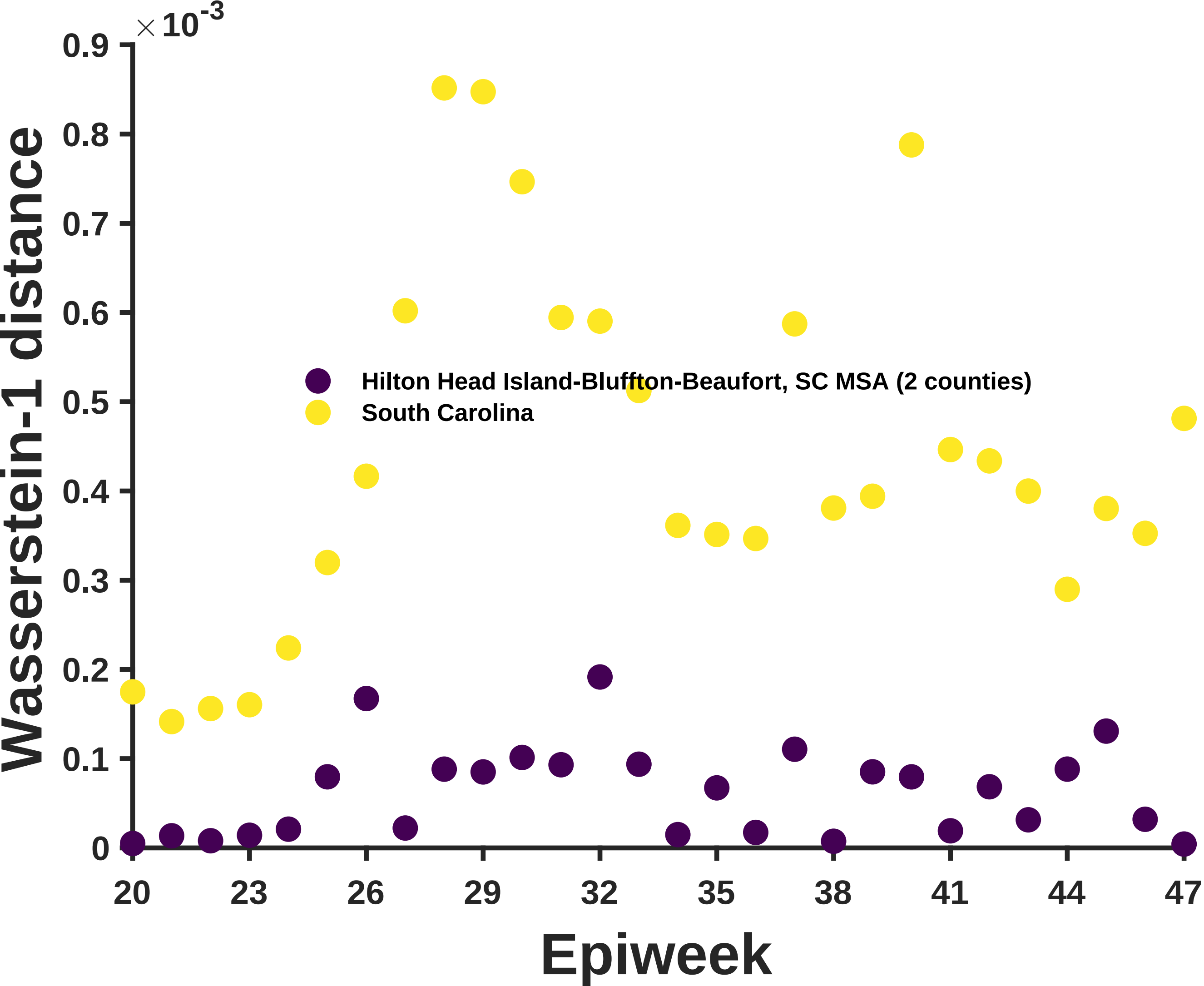

×10^-3
0.9
0.8
0.7
0.6
0.5
0.4
0.3
0.2
0.1
0
Wasserstein-1 distance
Hilton Head Island-Bluffton-Beaufort, SC MSA (2 counties)
South Carolina
20
23
26
29
32
35
38
41
44
47
Epiweek

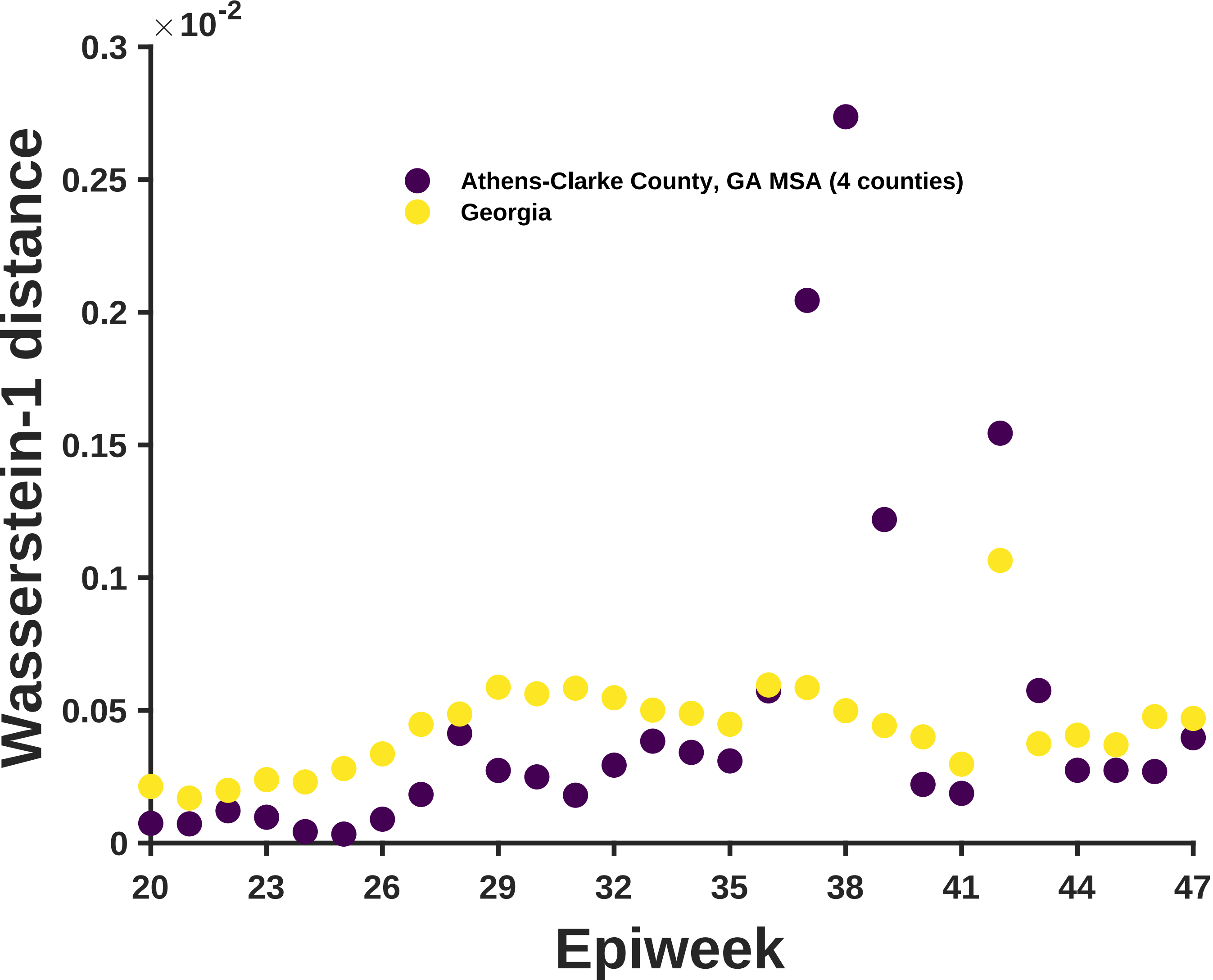

$\times 10^{-2}$
0.3
0.25
0.2
0.15
0.1
0.05
0
Wasserstein-1 distance
Athens-Clarke County, GA MSA (4 counties)
Georgia
20
23
26
29
32
35
38
41
44
47
Epiweek

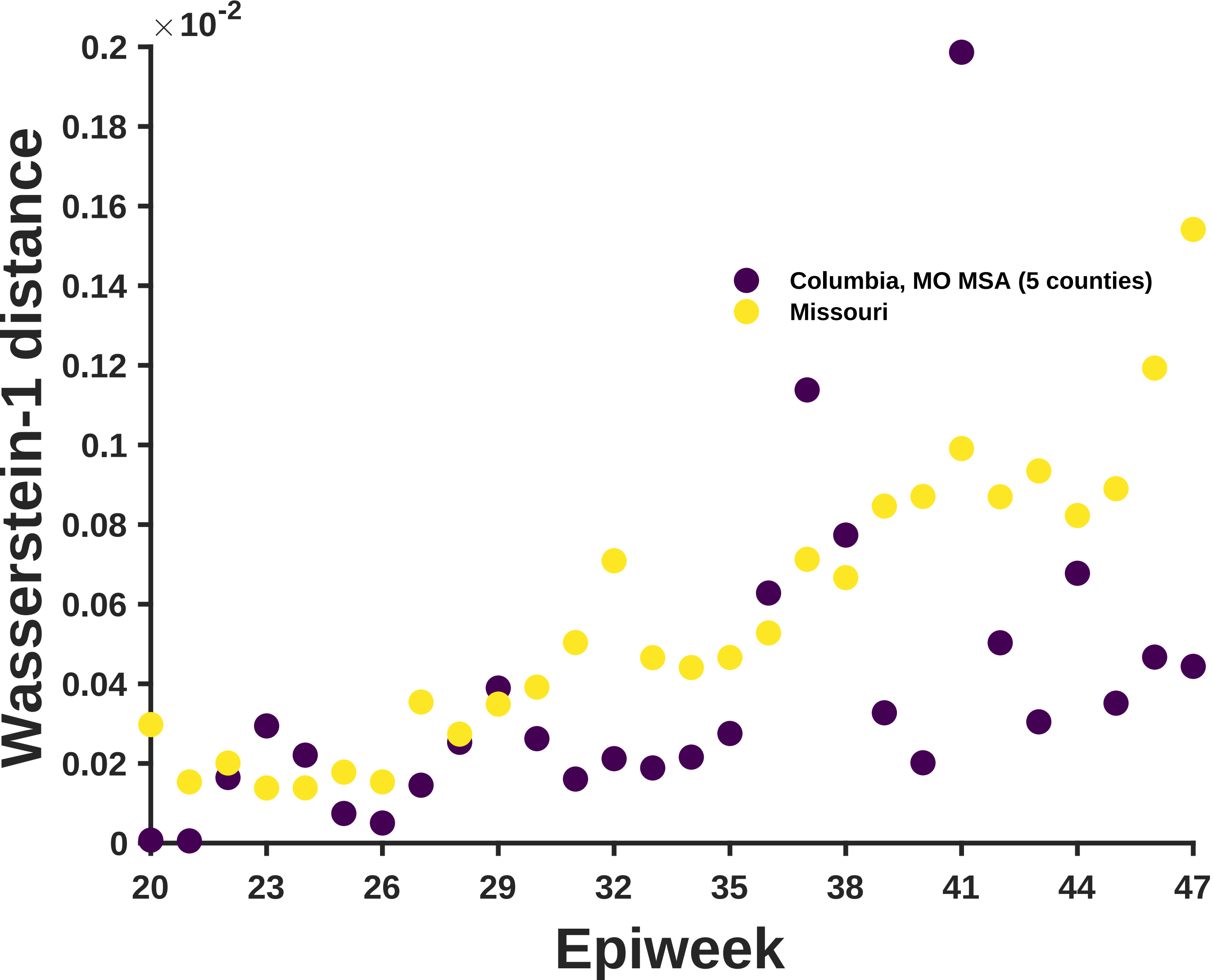
×10^-2
0.2
0.18
0.16
0.14
0.12
0.1
0.08
0.06
0.04
0.02
0
Wasserstein-1 distance
Columbia, MO MSA (5 counties)
Missouri
20
23
26
29
32
35
38
41
44
47
Epiweek

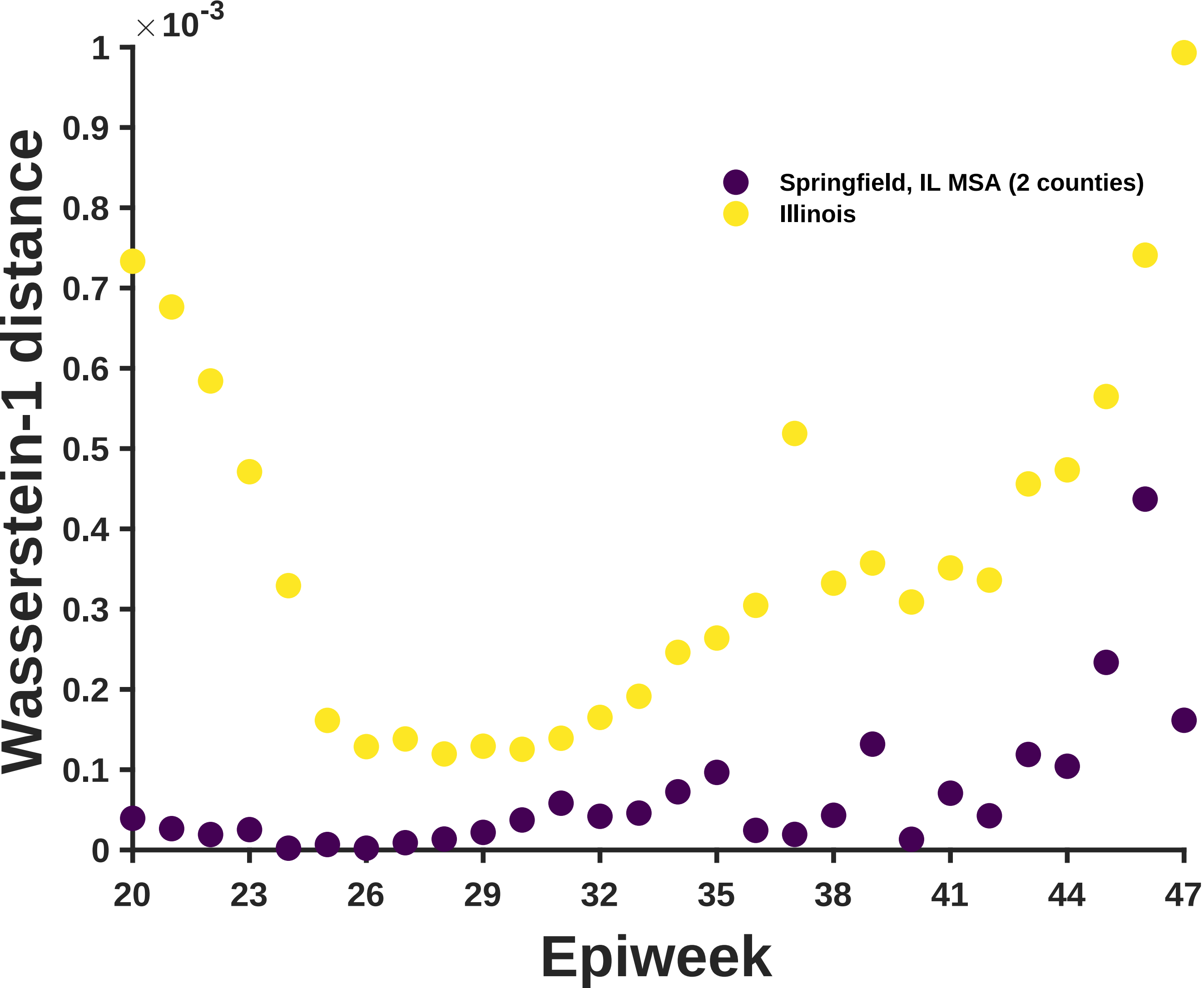
×10^-3
Wasserstein-1 distance
0
0.1
0.2
0.3
0.4
0.5
0.6
0.7
0.8
0.9
1
20
23
26
29
32
35
38
41
44
47
Epiweek
Springfield, IL MSA (2 counties)
Illinois

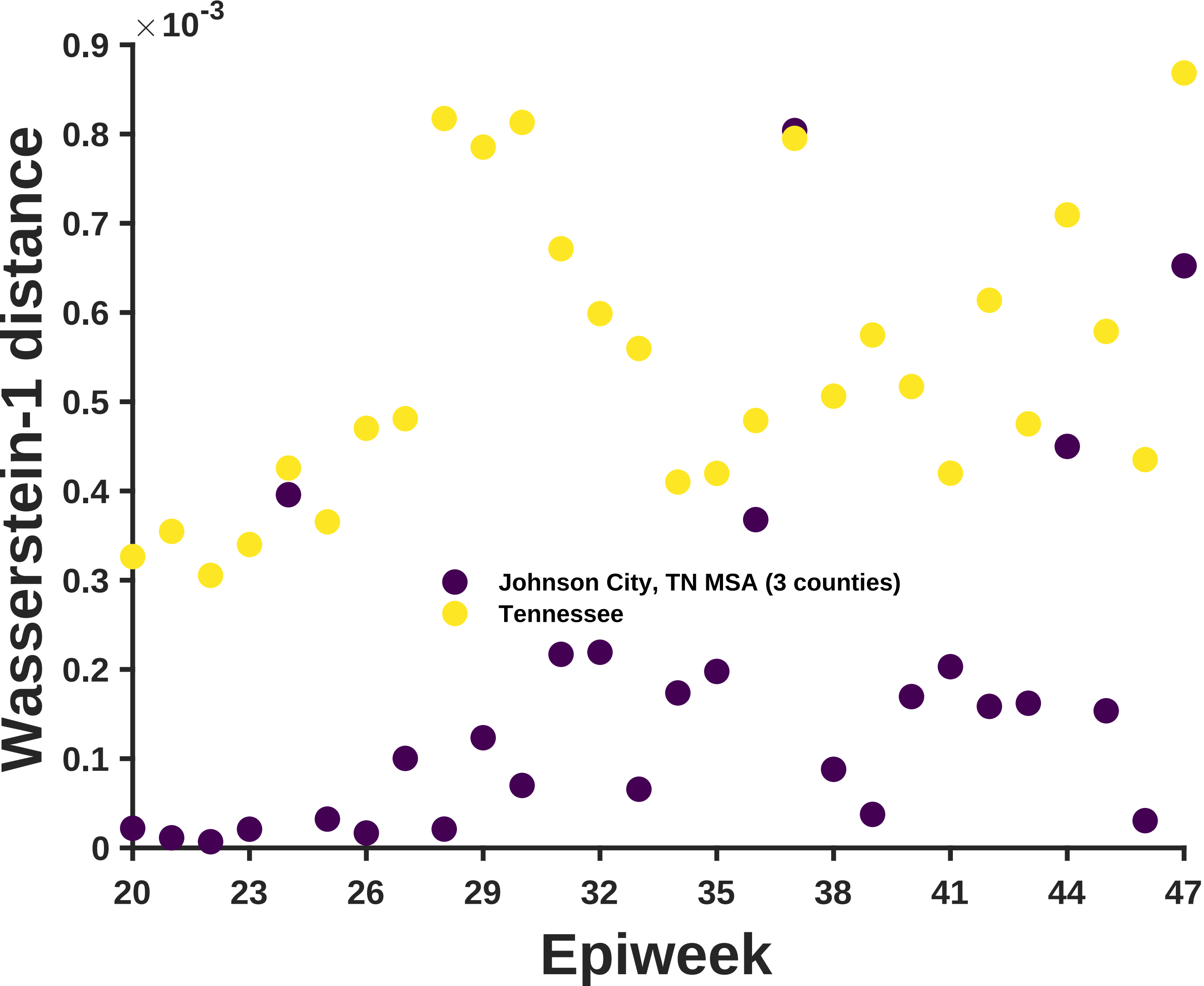

×10^-3
0.9
0.8
0.7
0.6
0.5
0.4
0.3
0.2
0.1
0
Wasserstein-1 distance
Johnson City, TN MSA (3 counties)
Tennessee
20
23
26
29
32
35
38
41
44
47
Epiweek

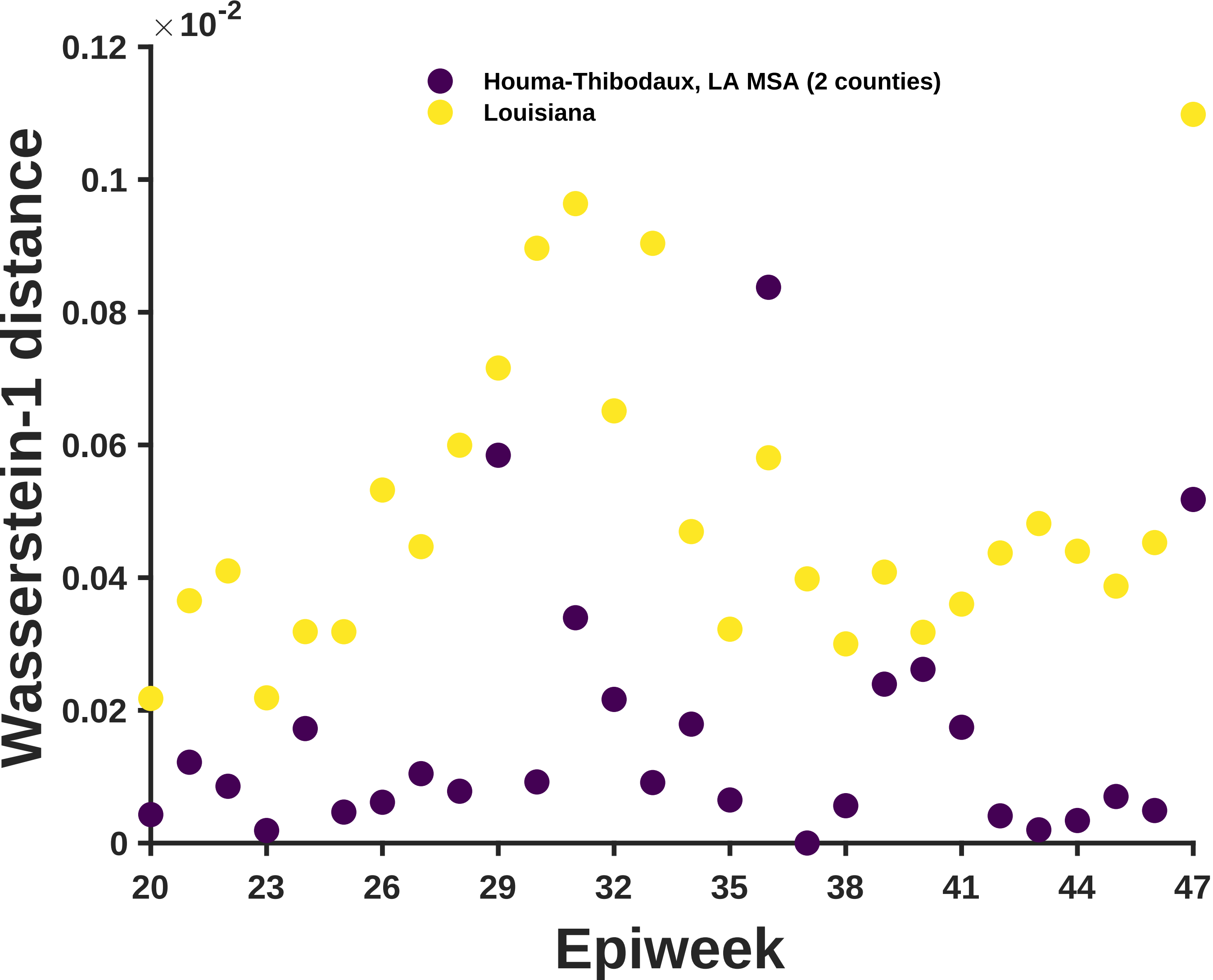

×10^-2
0.12
0.1
0.08
0.06
0.04
0.02
0
Wasserstein-1 distance
Houma-Thibodaux, LA MSA (2 counties)
Louisiana
20
23
26
29
32
35
38
41
44
47
Epiweek

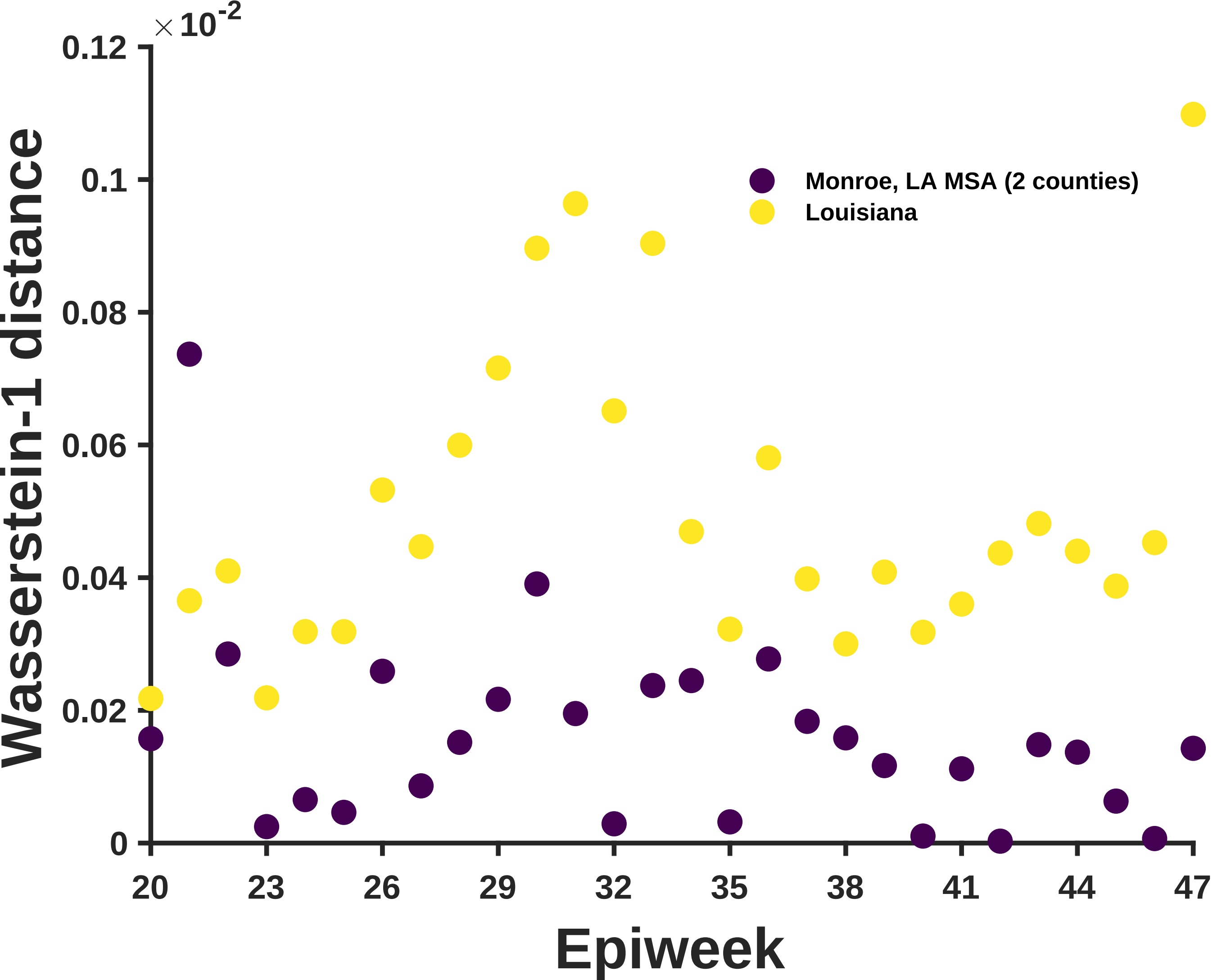

×10^-2
0.12
0.1
0.08
0.06
0.04
0.02
0
Wasserstein-1 distance
Monroe, LA MSA (2 counties)
Louisiana
20
23
26
29
32
35
38
41
44
47
Epiweek

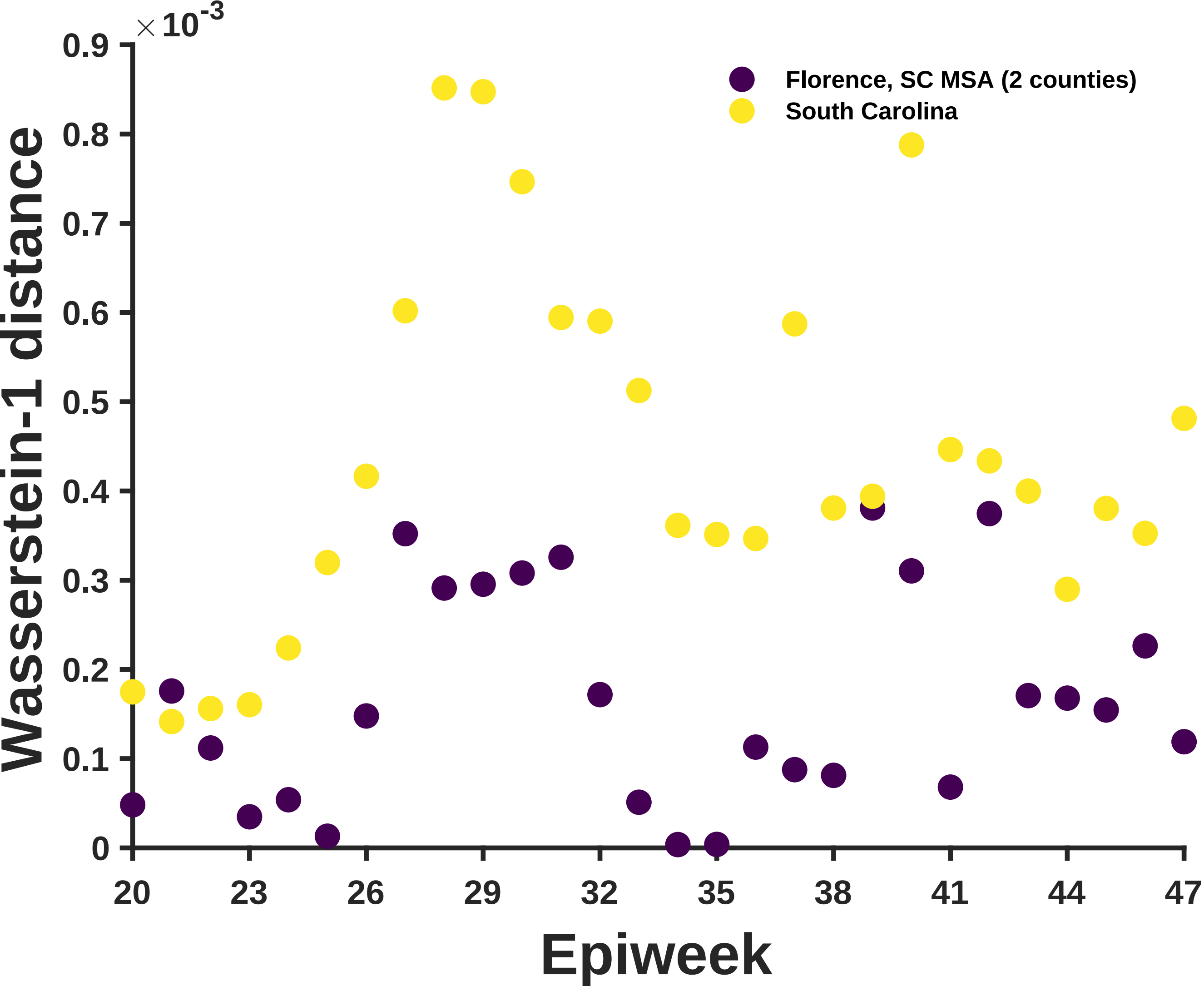

×10^-3
0.9
0.8
0.7
0.6
0.5
0.4
0.3
0.2
0.1
0
Wasserstein-1 distance
20
23
26
29
32
35
38
41
44
47
Epiweek
Florence, SC MSA (2 counties)
South Carolina

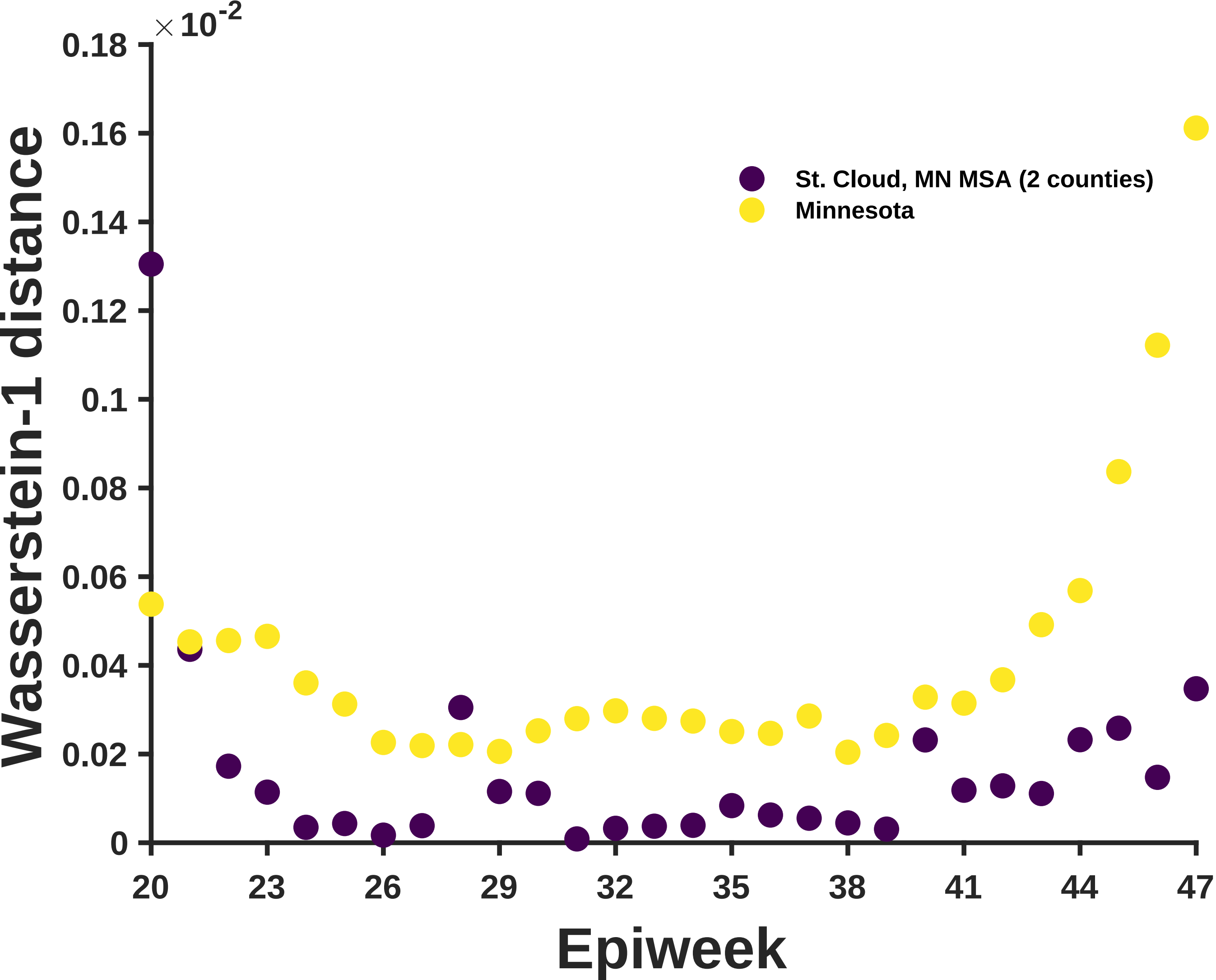

×10^-2
0.18
0.16
0.14
0.12
0.1
0.08
0.06
0.04
0.02
0
Wasserstein-1 distance
20
23
26
29
32
35
38
41
44
47
Epiweek
St. Cloud, MN MSA (2 counties)
Minnesota

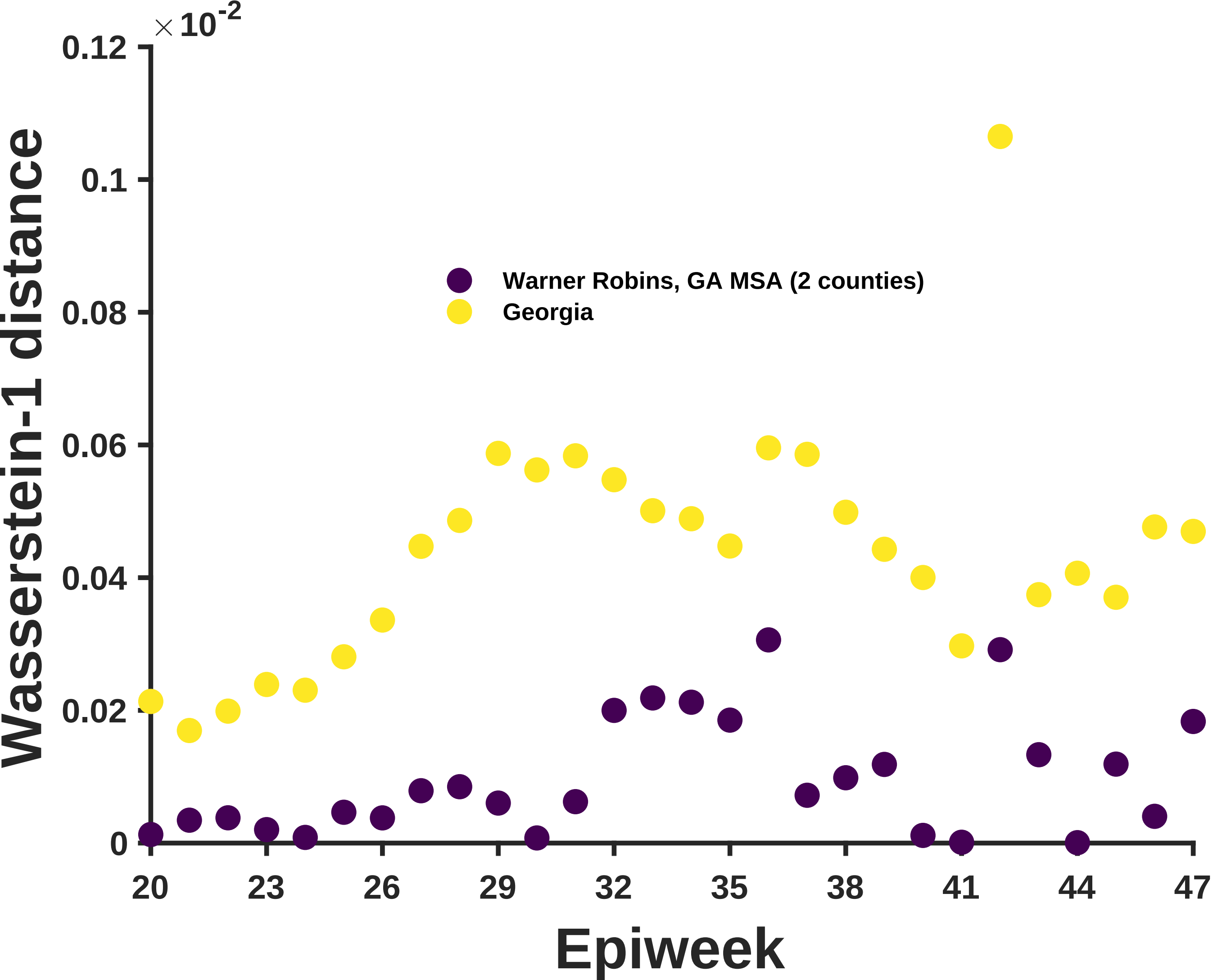

×10^-2
0.12
0.1
0.08
0.06
0.04
0.02
0
Wasserstein-1 distance
Warner Robins, GA MSA (2 counties)
Georgia
20
23
26
29
32
35
38
41
44
47
Epiweek

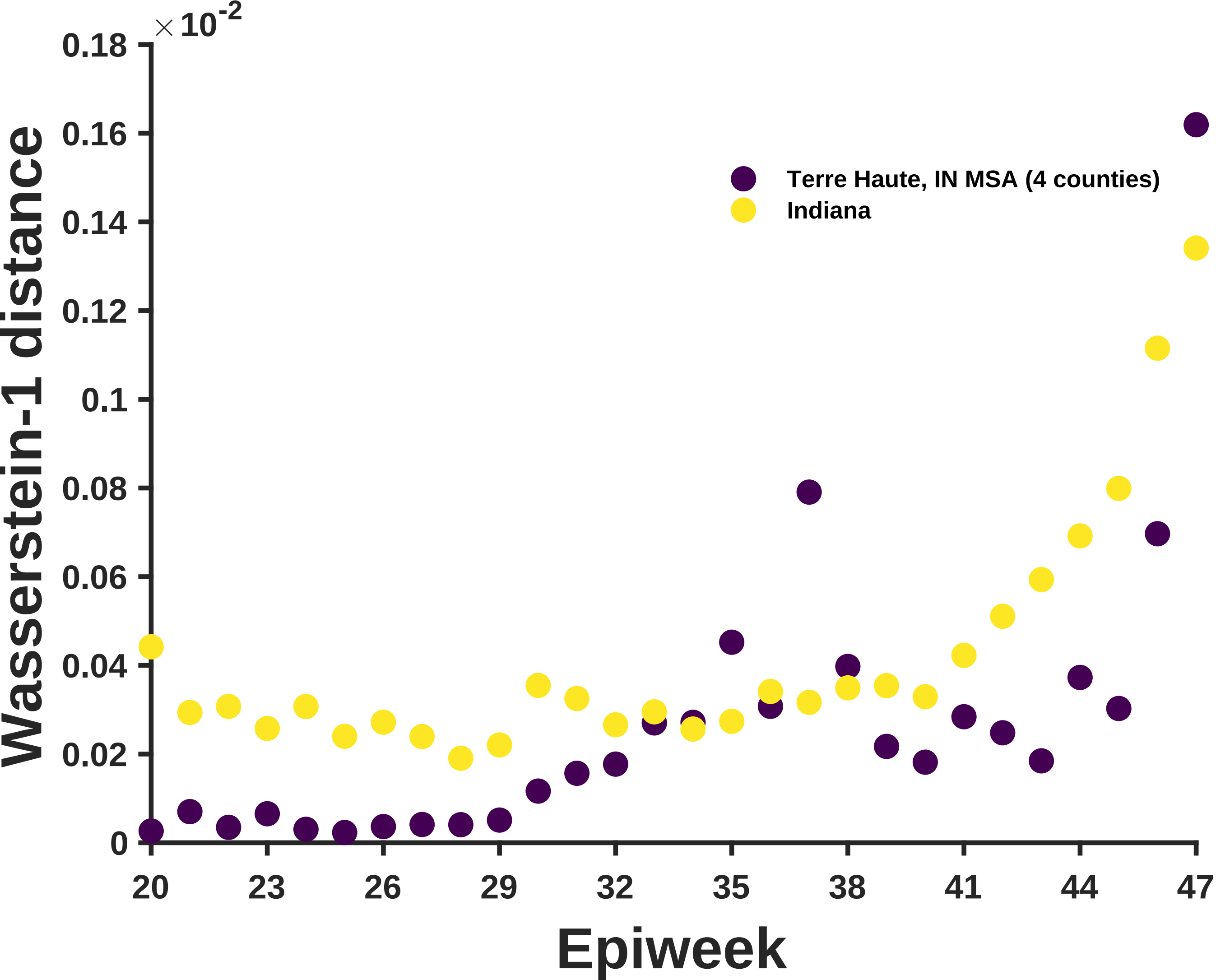

×10^-2
0.18
0.16
0.14
0.12
0.1
0.08
0.06
0.04
0.02
0
Wasserstein-1 distance
Terre Haute, IN MSA (4 counties)
Indiana
20
23
26
29
32
35
38
41
44
47
Epiweek

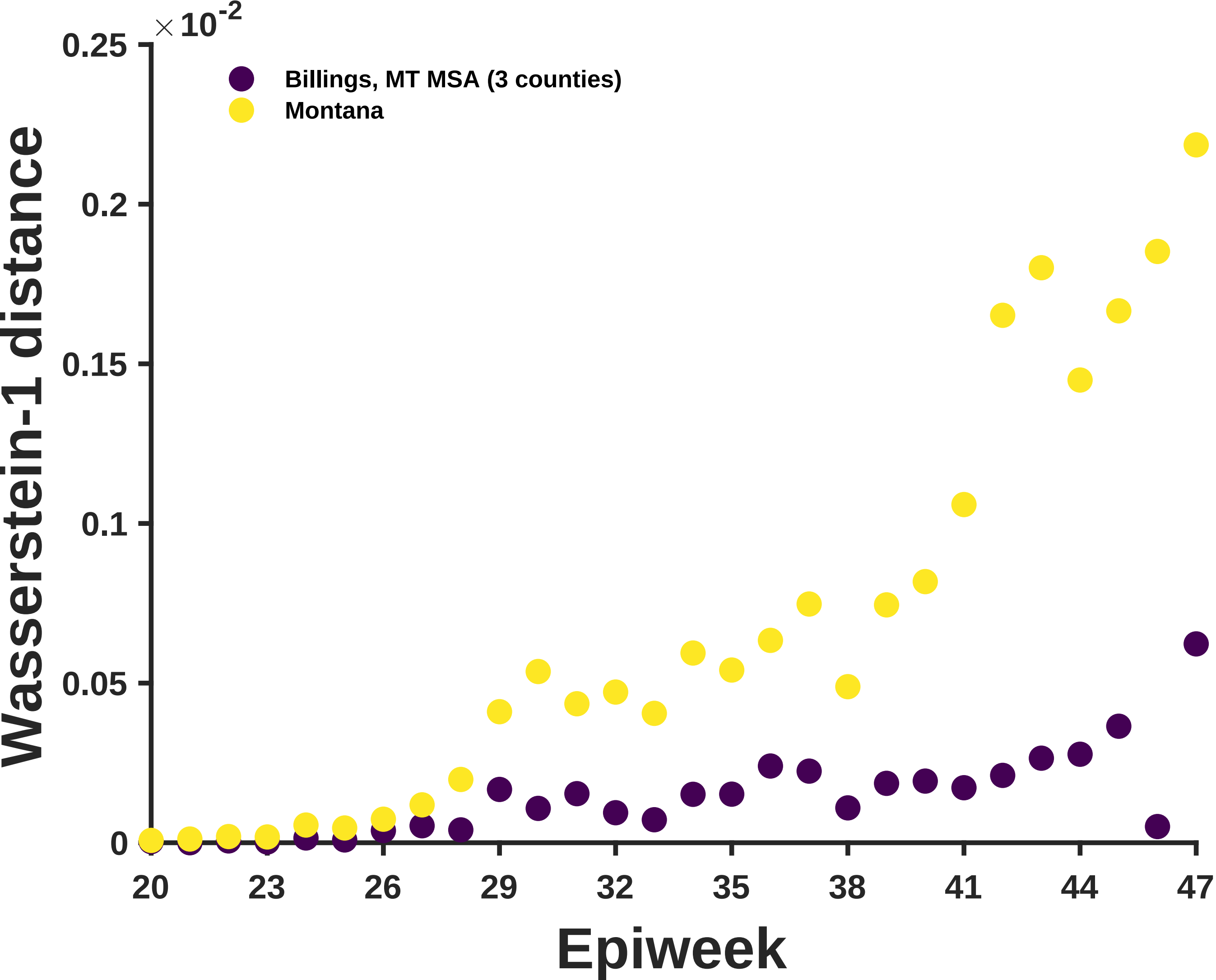

×10^-2
0.25
0.2
0.15
0.1
0.05
0
Wasserstein-1 distance
Billings, MT MSA (3 counties)
Montana
20
23
26
29
32
35
38
41
44
47
Epiweek

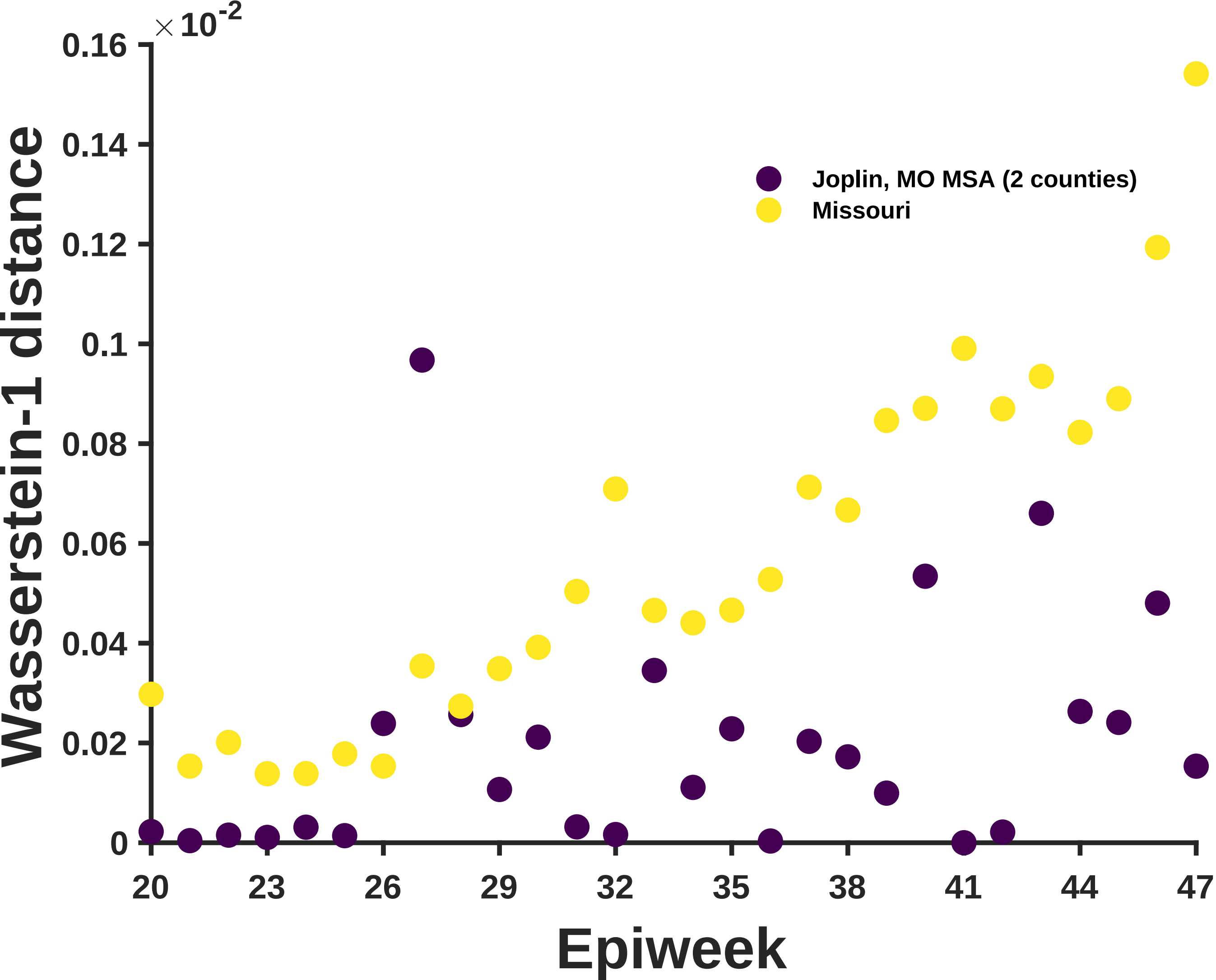

×10^-2
0.16
0.14
0.12
0.1
0.08
0.06
0.04
0.02
0
Wasserstein-1 distance
20
23
26
29
32
35
38
41
44
47
Epiweek
Joplin, MO MSA (2 counties)
Missouri

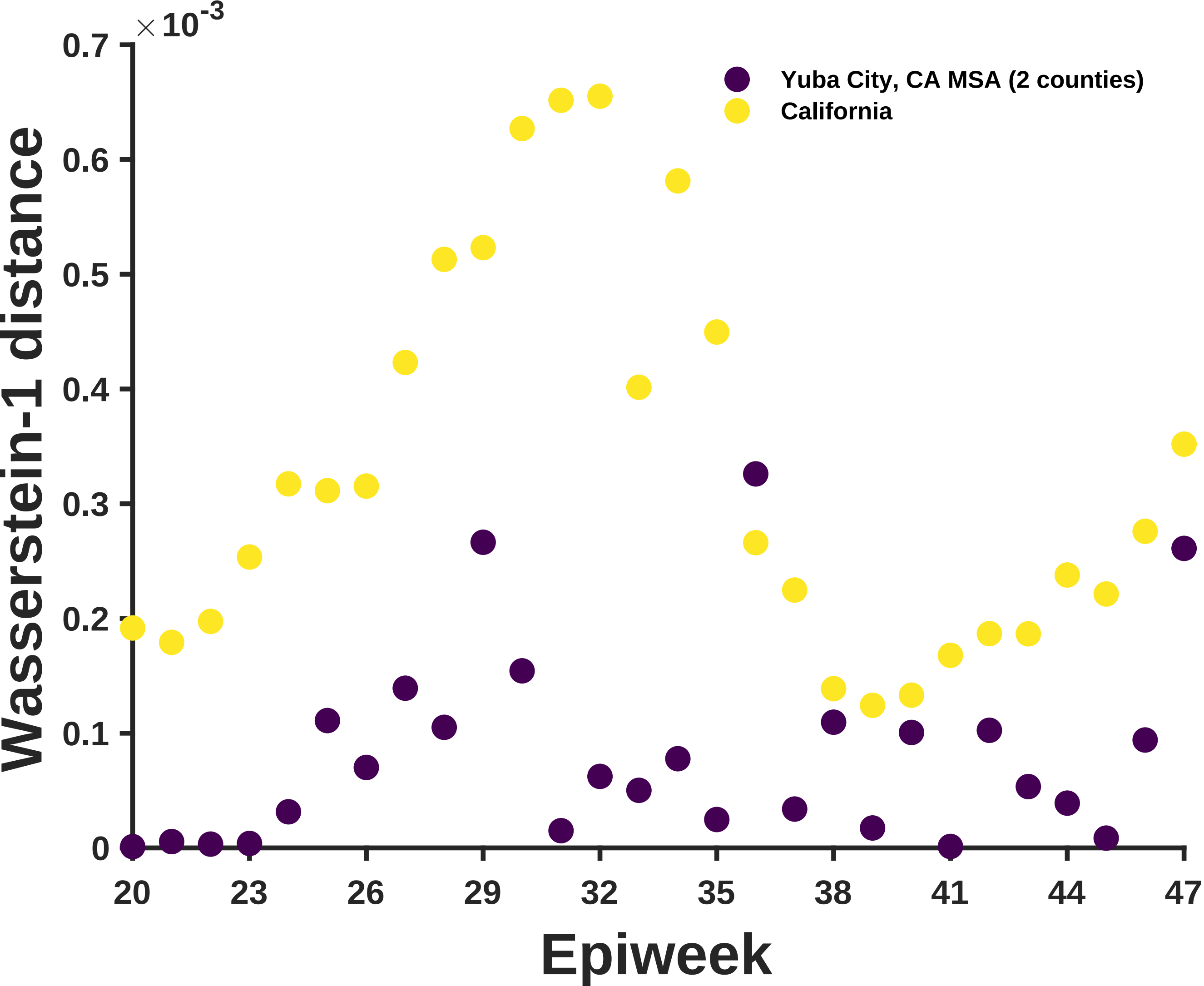

×10^-3
0.7
0.6
0.5
0.4
0.3
0.2
0.1
0
Wasserstein-1 distance
Yuba City, CA MSA (2 counties)
California
20
23
26
29
32
35
38
41
44
47
Epiweek

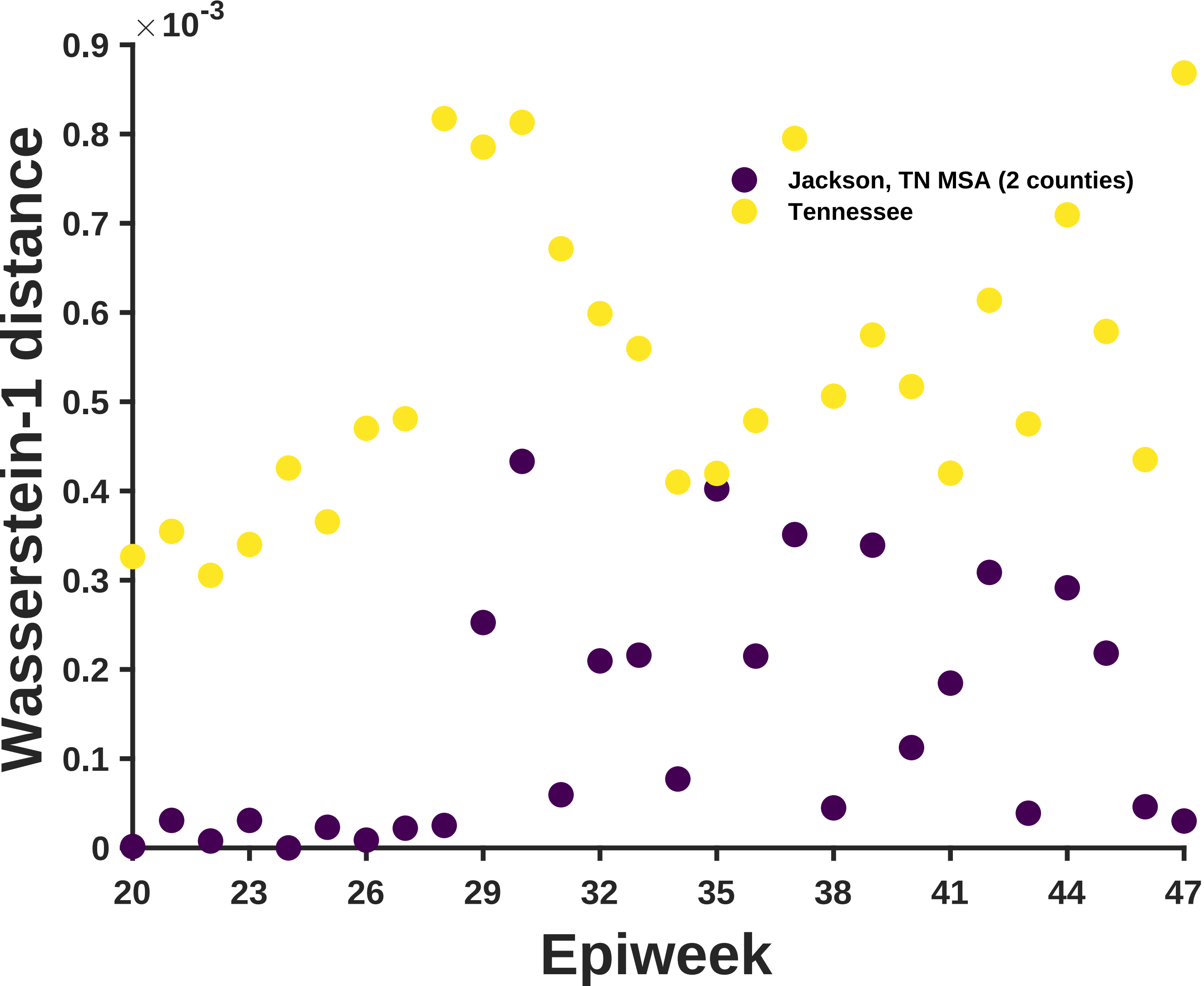

×10^-3
0.9
0.8
0.7
0.6
0.5
0.4
0.3
0.2
0.1
0
Wasserstein-1 distance
Jackson, TN MSA (2 counties)
Tennessee
20
23
26
29
32
35
38
41
44
47
Epiweek

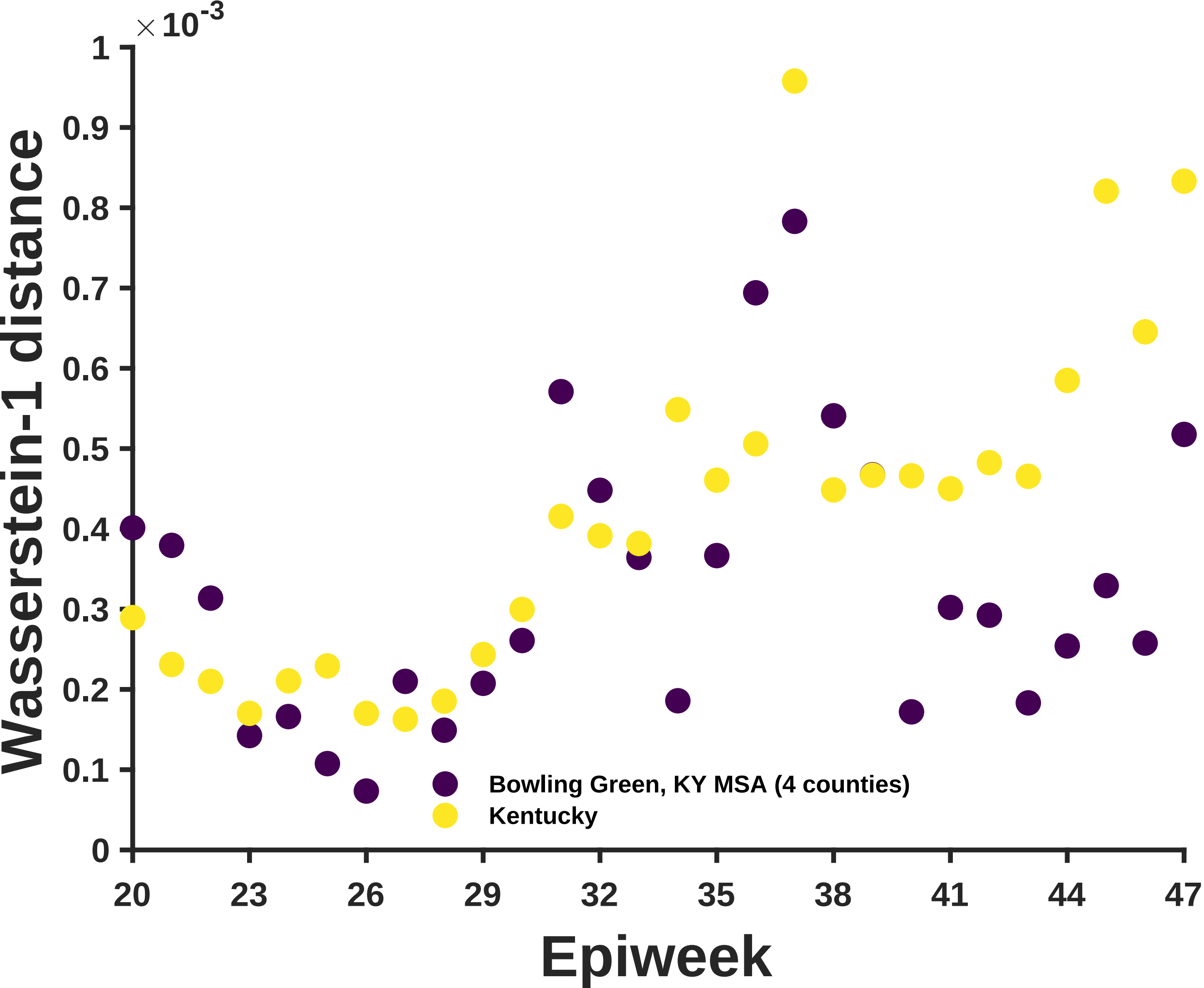

×10^-3
Wasserstein-1 distance
Epiweek
0
0.1
0.2
0.3
0.4
0.5
0.6
0.7
0.8
0.9
1
20
23
26
29
32
35
38
41
44
47
Bowling Green, KY MSA (4 counties)
Kentucky

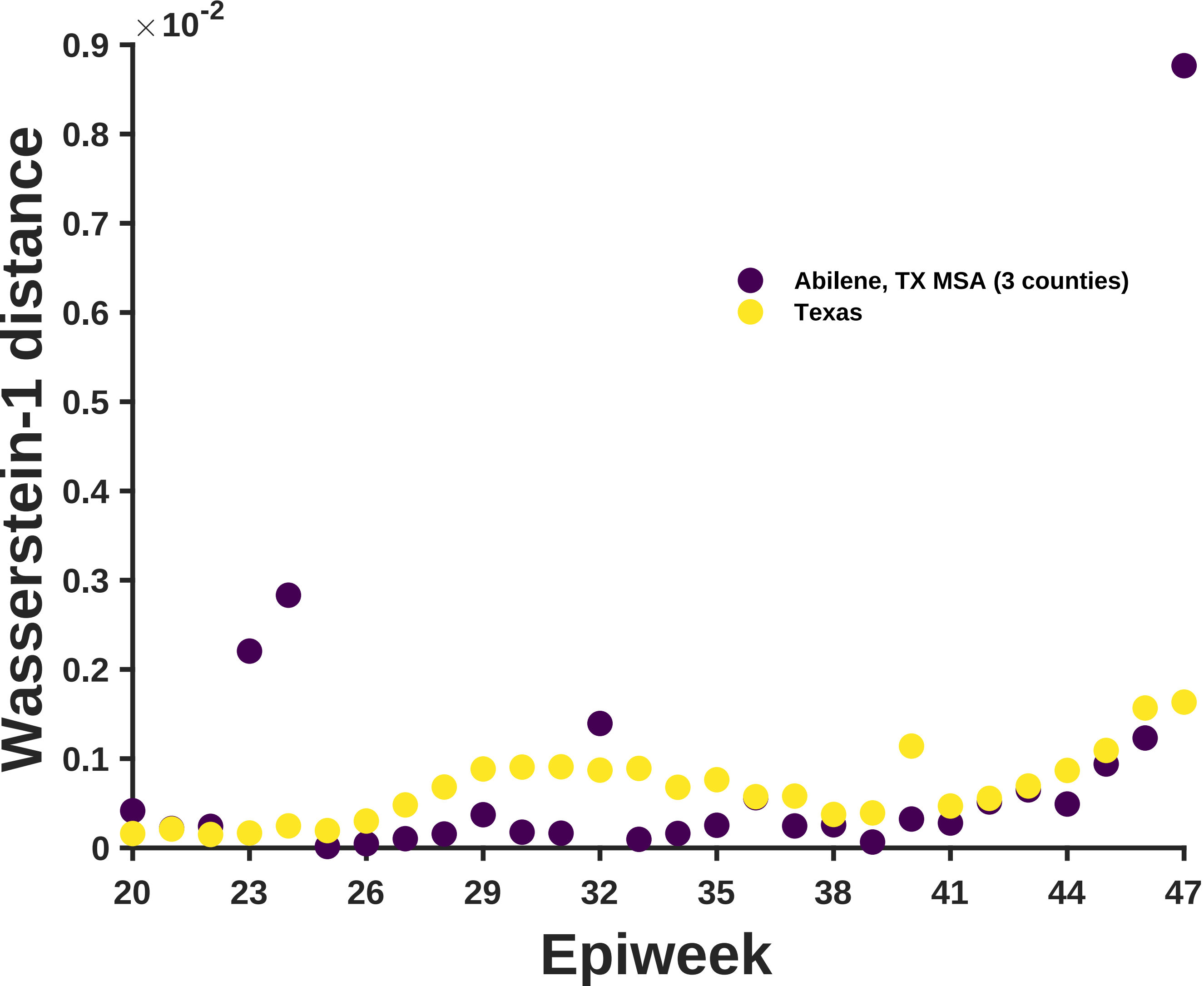

×10^-2
0.9
0.8
0.7
0.6
0.5
0.4
0.3
0.2
0.1
0
Wasserstein-1 distance
Abilene, TX MSA (3 counties)
Texas
20
23
26
29
32
35
38
41
44
47
Epiweek

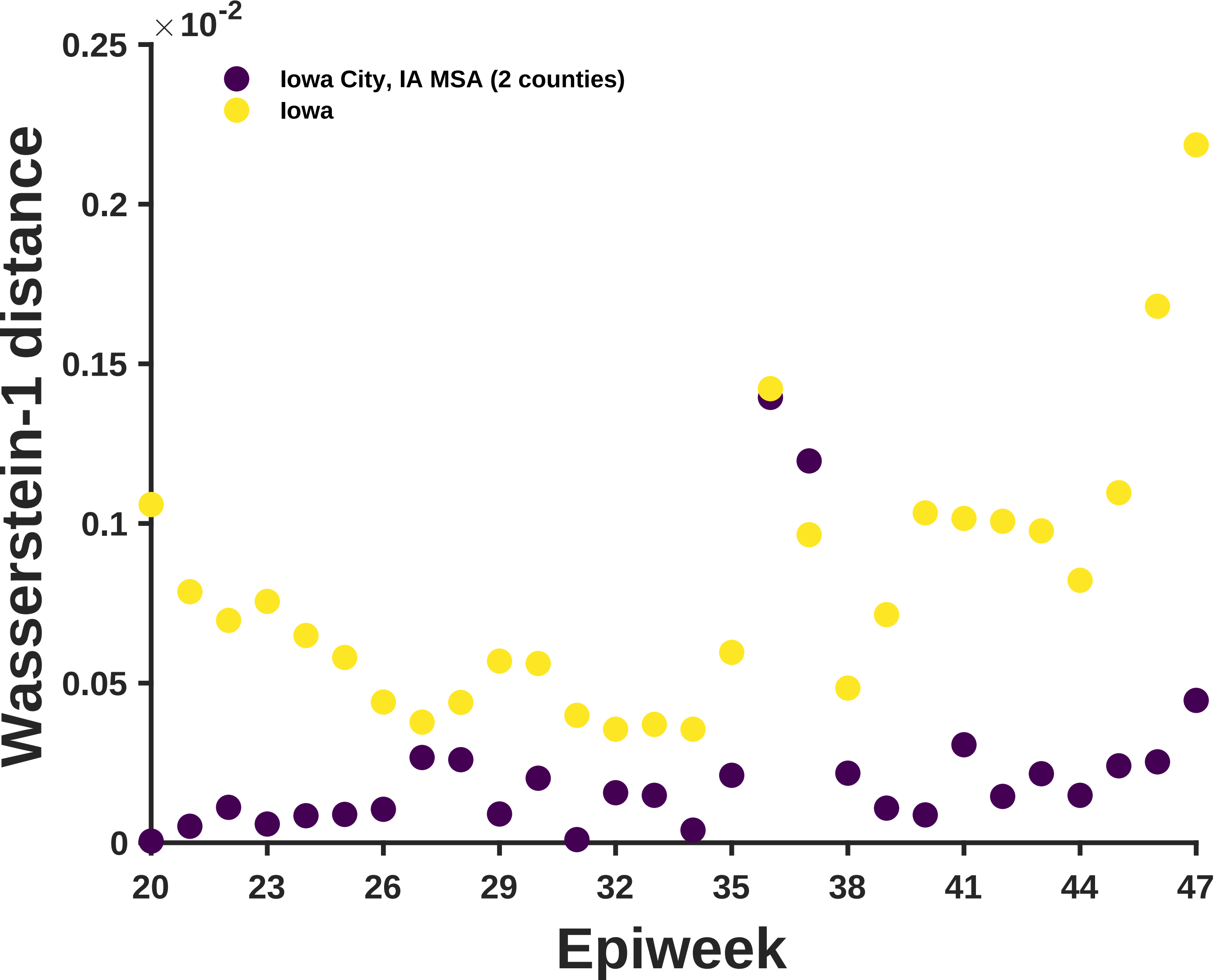

×10^-2
0.25
0.2
0.15
0.1
0.05
0
Iowa City, IA MSA (2 counties)
Iowa
Wasserstein-1 distance
20
23
26
29
32
35
38
41
44
47
Epiweek

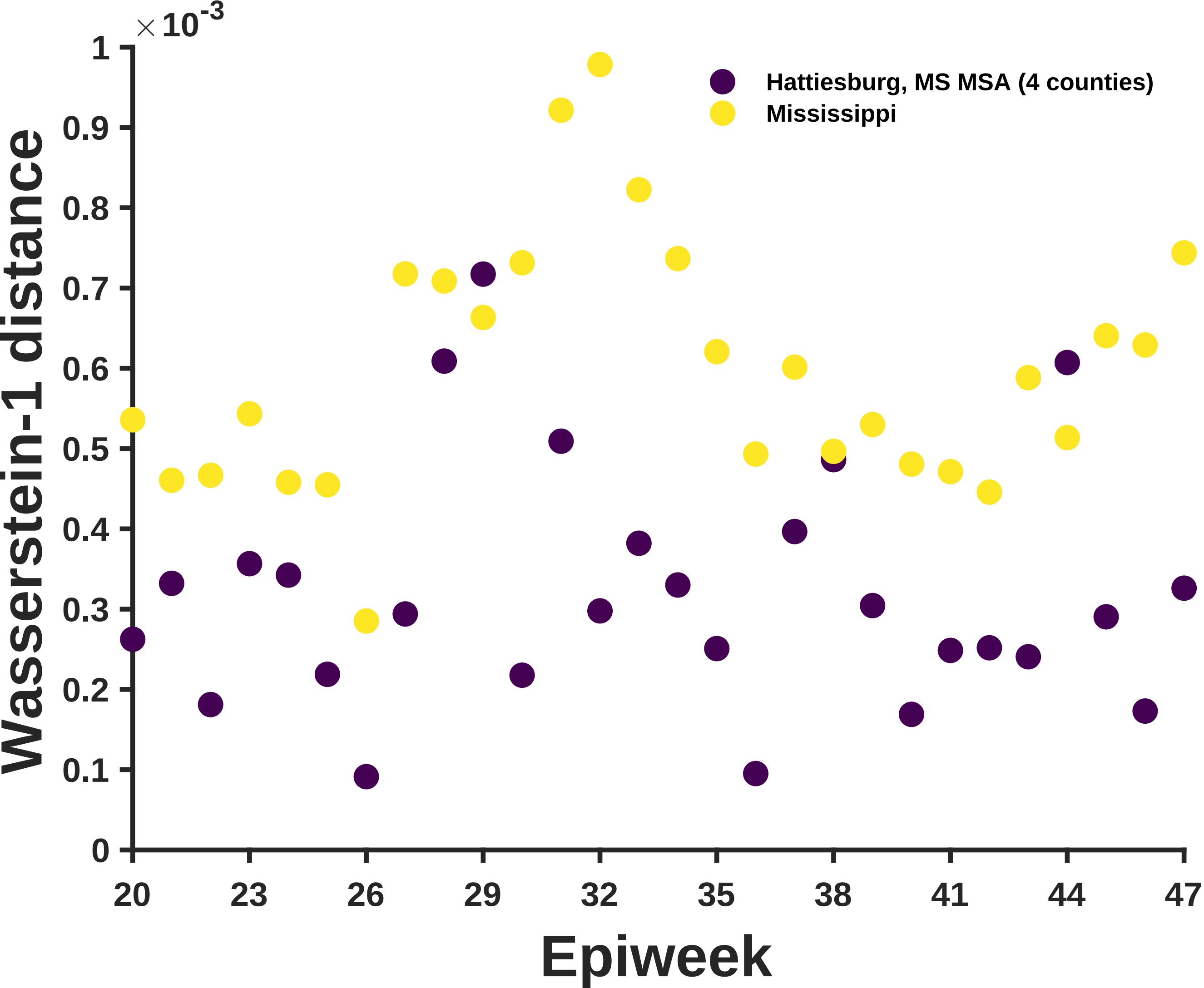

×10^-3
1
0.9
0.8
0.7
0.6
0.5
0.4
0.3
0.2
0.1
0
Wasserstein-1 distance
Hattiesburg, MS MSA (4 counties)
Mississippi
20
23
26
29
32
35
38
41
44
47
Epiweek

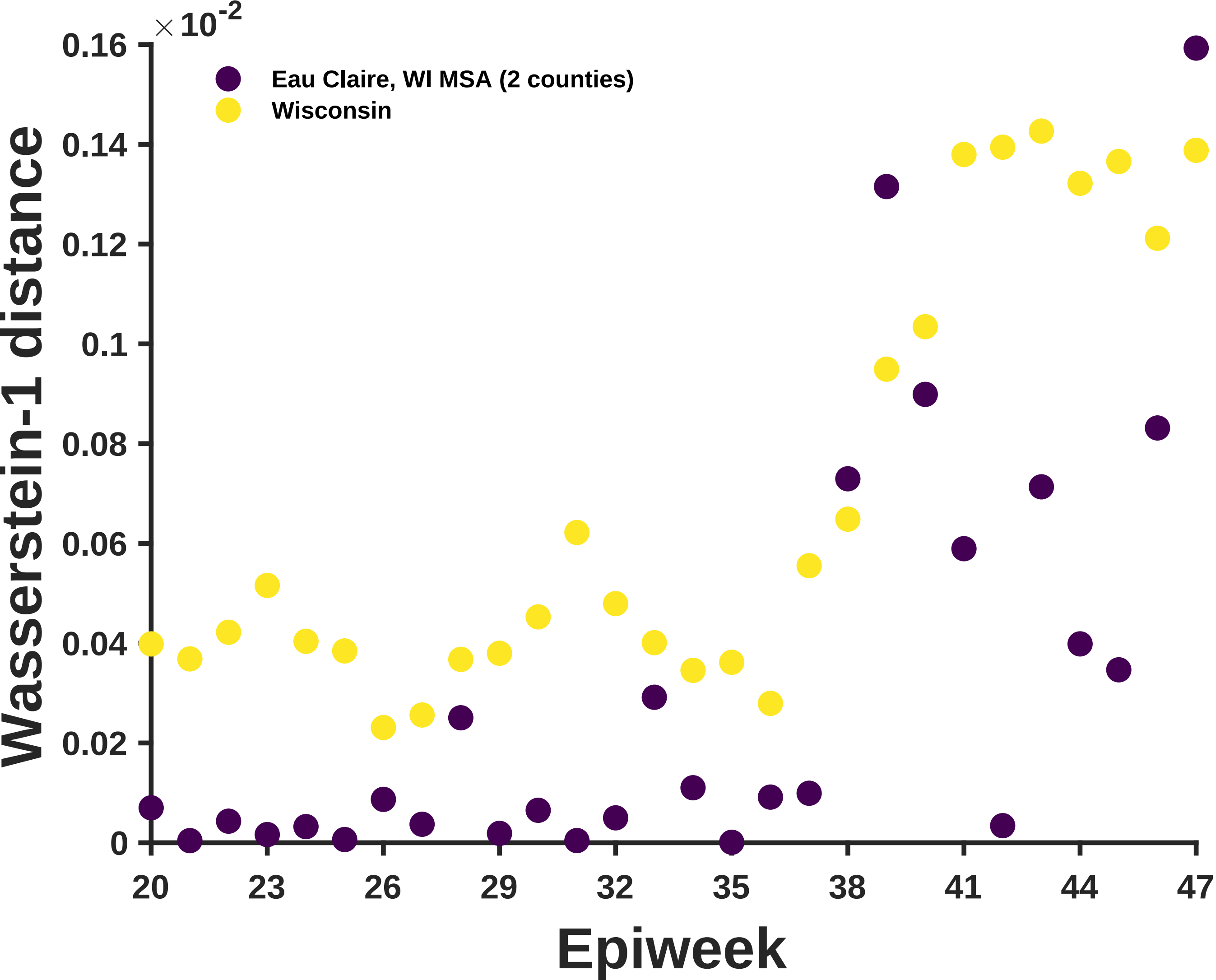

×10^-2
0.16
0.14
0.12
0.1
0.08
0.06
0.04
0.02
0
Wasserstein-1 distance
Eau Claire, WI MSA (2 counties)
Wisconsin
20
23
26
29
32
35
38
41
44
47
Epiweek

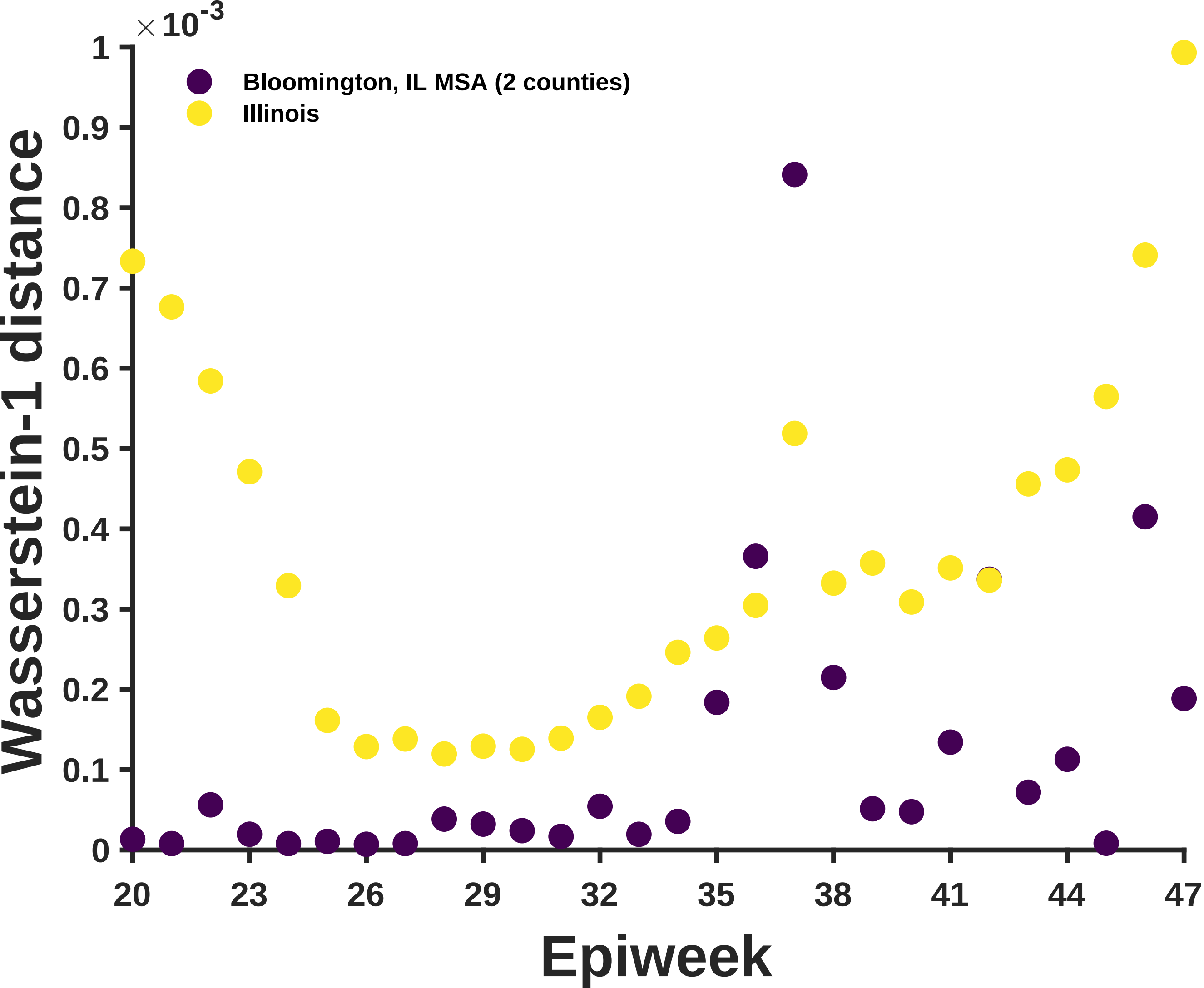

×10^-3
Bloomington, IL MSA (2 counties)
Illinois
Wasserstein-1 distance
0
0.1
0.2
0.3
0.4
0.5
0.6
0.7
0.8
0.9
1
20
23
26
29
32
35
38
41
44
47
Epiweek

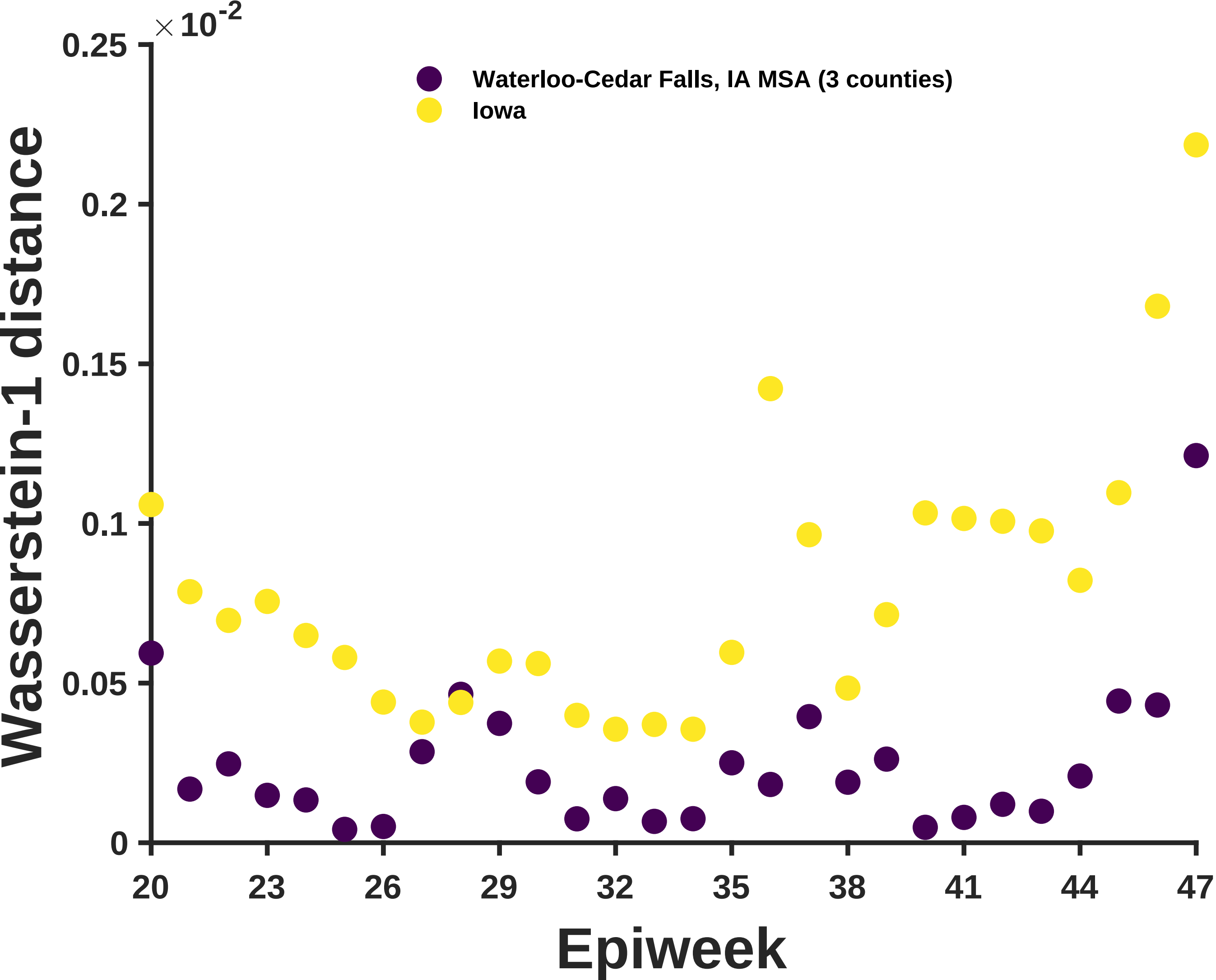

×10^-2
0.25
0.2
0.15
0.1
0.05
0
Wasserstein-1 distance
Waterloo-Cedar Falls, IA MSA (3 counties)
Iowa
20
23
26
29
32
35
38
41
44
47
Epiweek

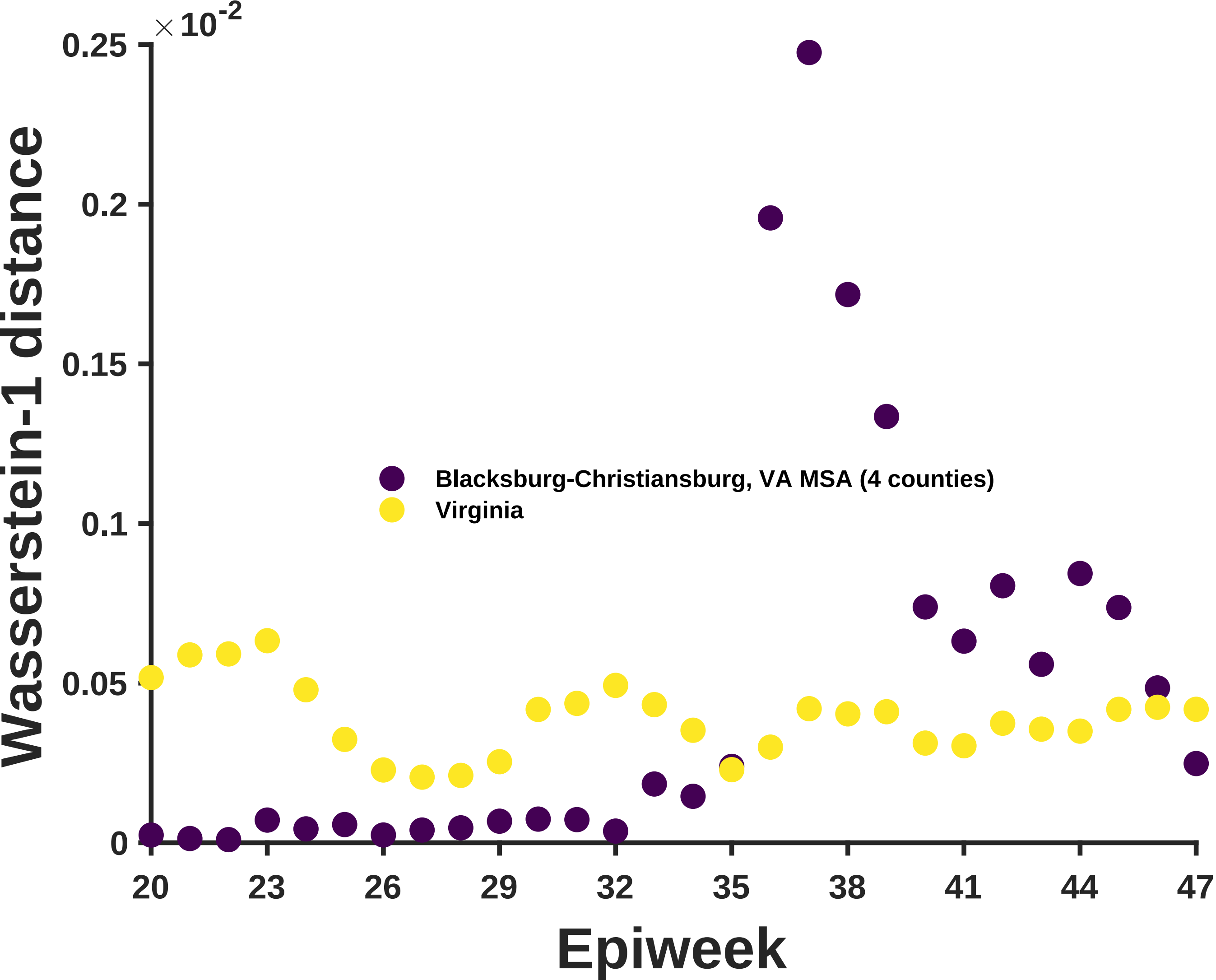

×10^-2
0.25
0.2
0.15
0.1
0.05
0
Wasserstein-1 distance
Blacksburg-Christiansburg, VA MSA (4 counties)
Virginia
20
23
26
29
32
35
38
41
44
47
Epiweek

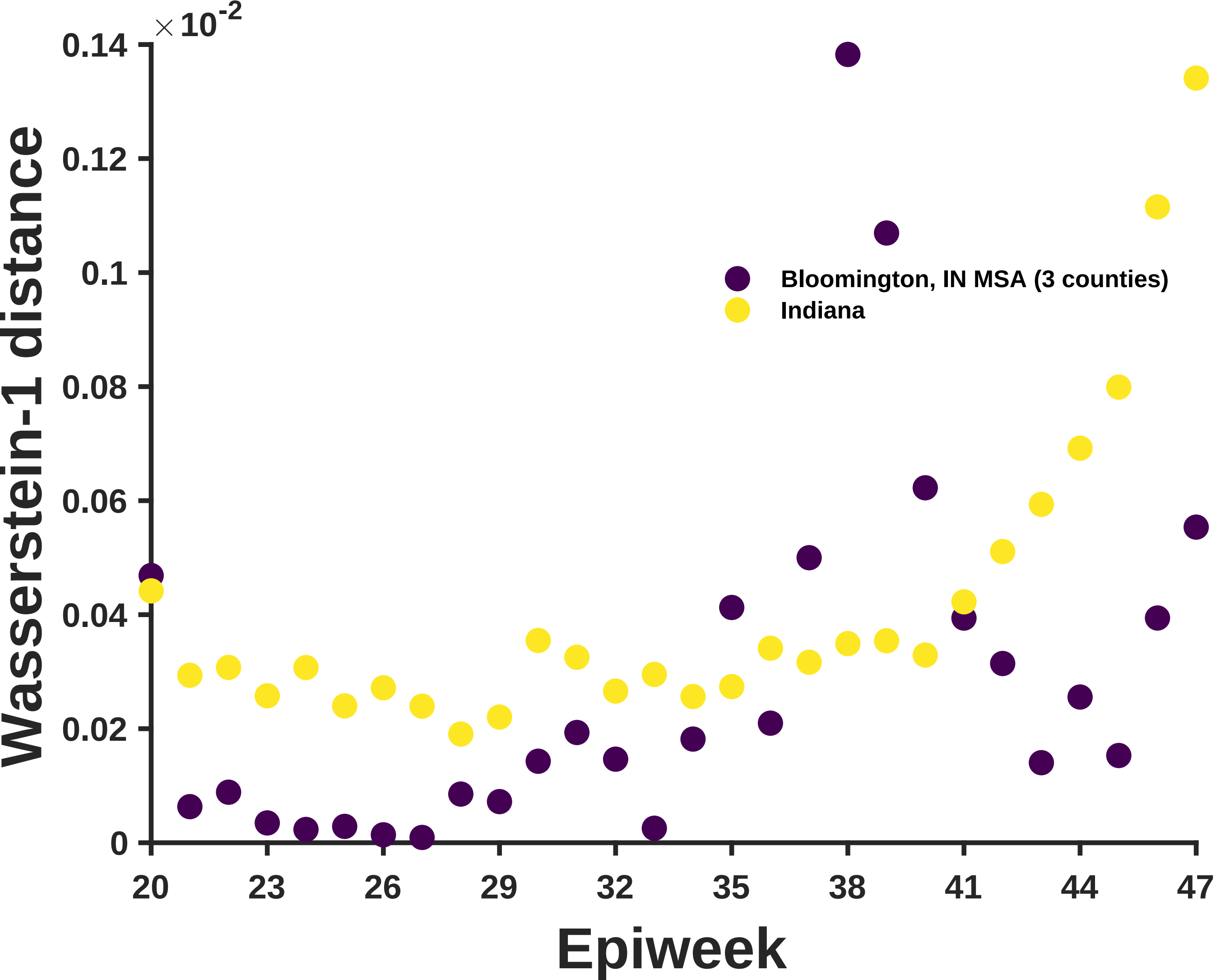

×10^-2
0.14
0.12
0.1
0.08
0.06
0.04
0.02
0
Wasserstein-1 distance
Bloomington, IN MSA (3 counties)
Indiana
20
23
26
29
32
35
38
41
44
47
Epiweek

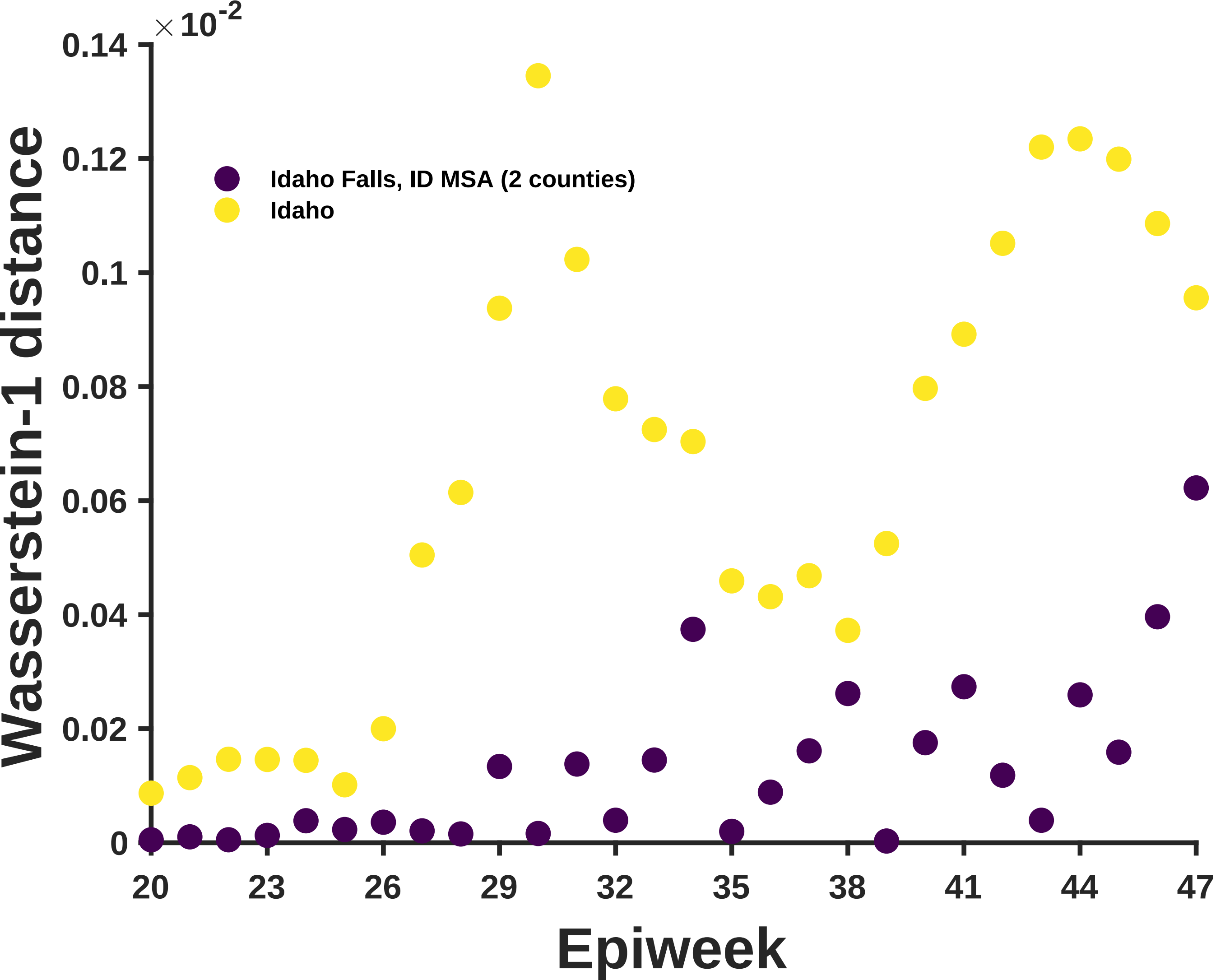

×10^-2
0.14
0.12
0.1
0.08
0.06
0.04
0.02
0
Wasserstein-1 distance
Idaho Falls, ID MSA (2 counties)
Idaho
20
23
26
29
32
35
38
41
44
47
Epiweek

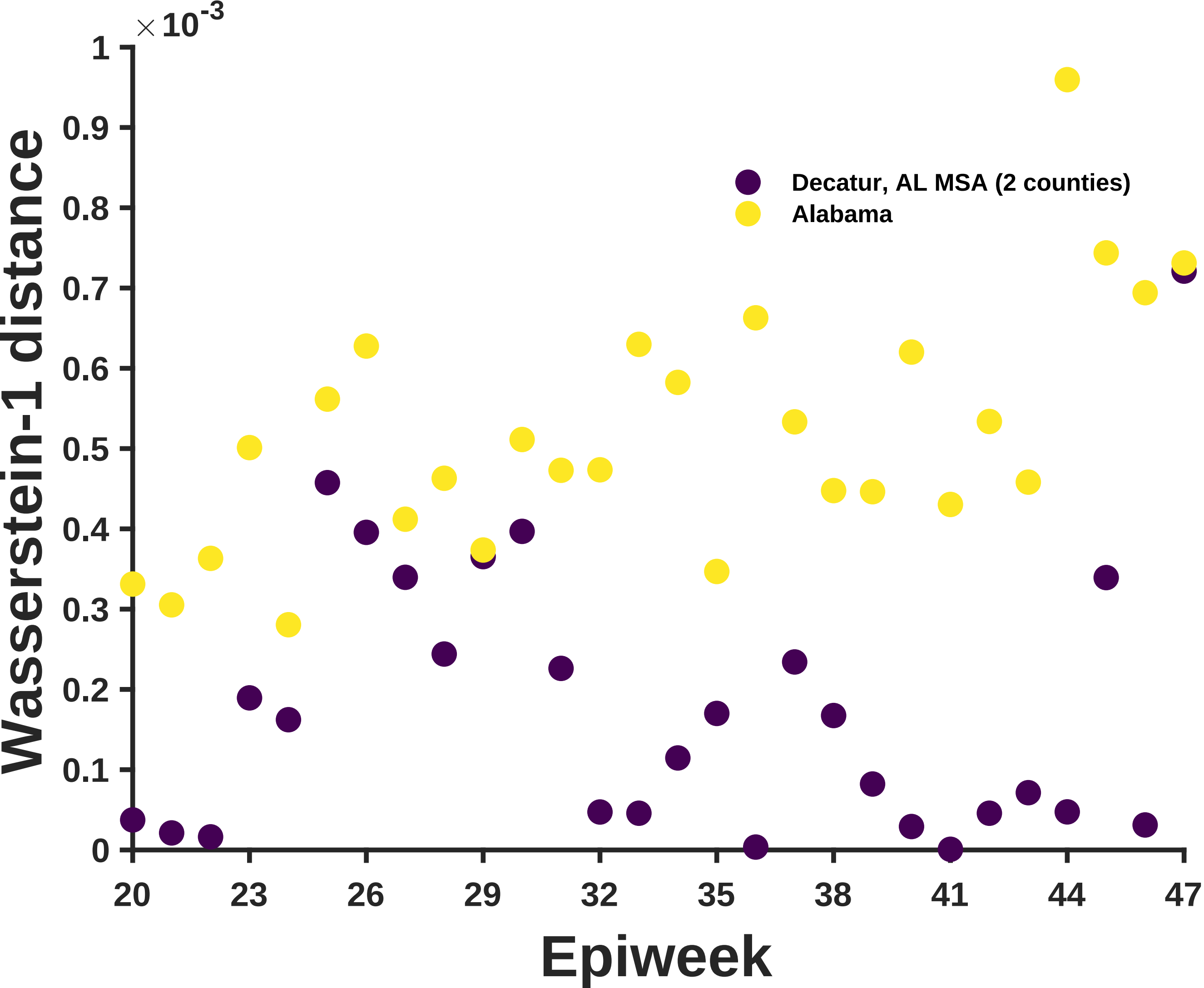

×10^-3
Wasserstein-1 distance
Epiweek
Decatur, AL MSA (2 counties)
Alabama
0
0.1
0.2
0.3
0.4
0.5
0.6
0.7
0.8
0.9
1
20
23
26
29
32
35
38
41
44
47

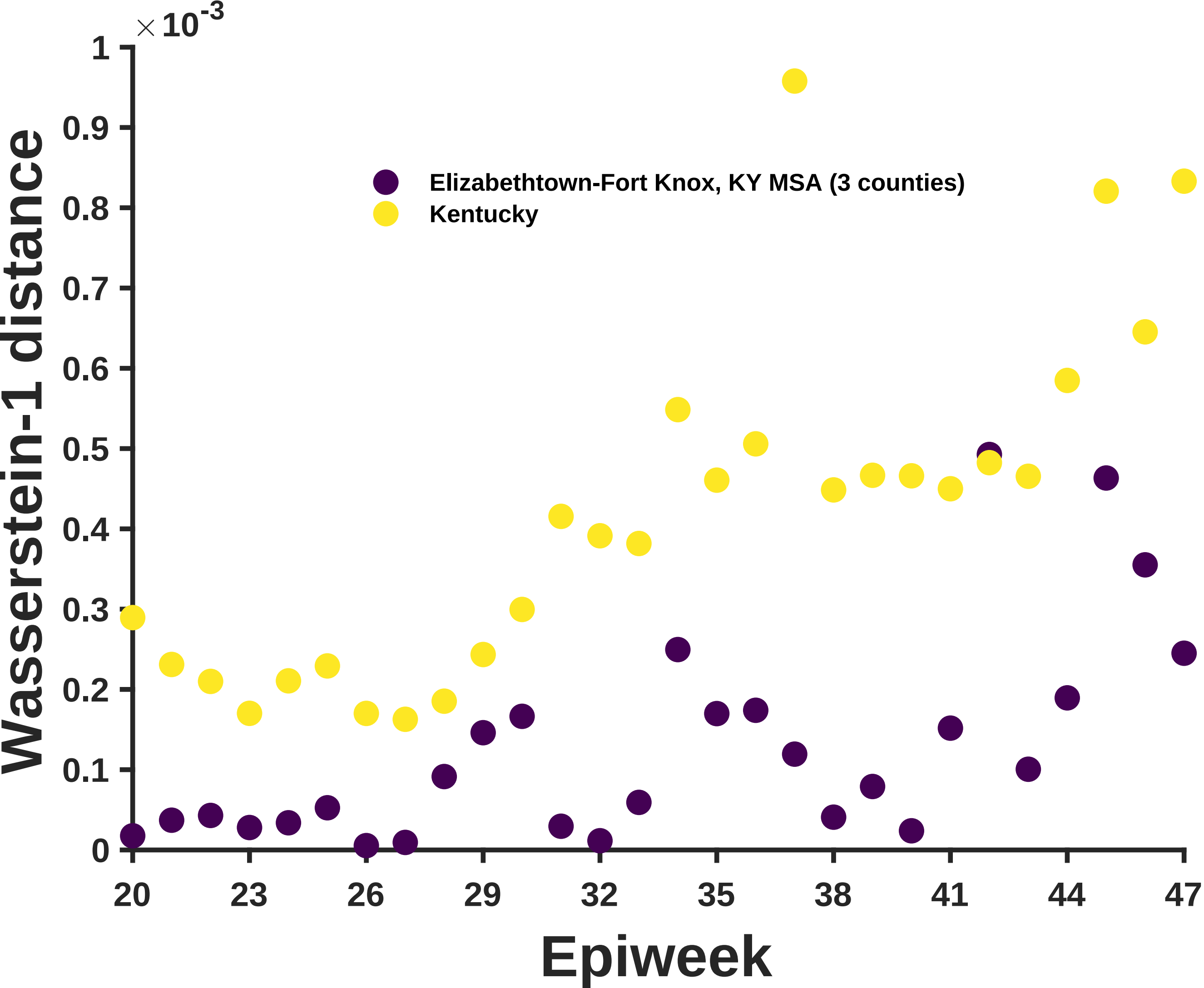

×10^-3
Wasserstein-1 distance
Elizabethtown-Fort Knox, KY MSA (3 counties)
Kentucky
Epiweek
0
0.1
0.2
0.3
0.4
0.5
0.6
0.7
0.8
0.9
1
20
23
26
29
32
35
38
41
44
47

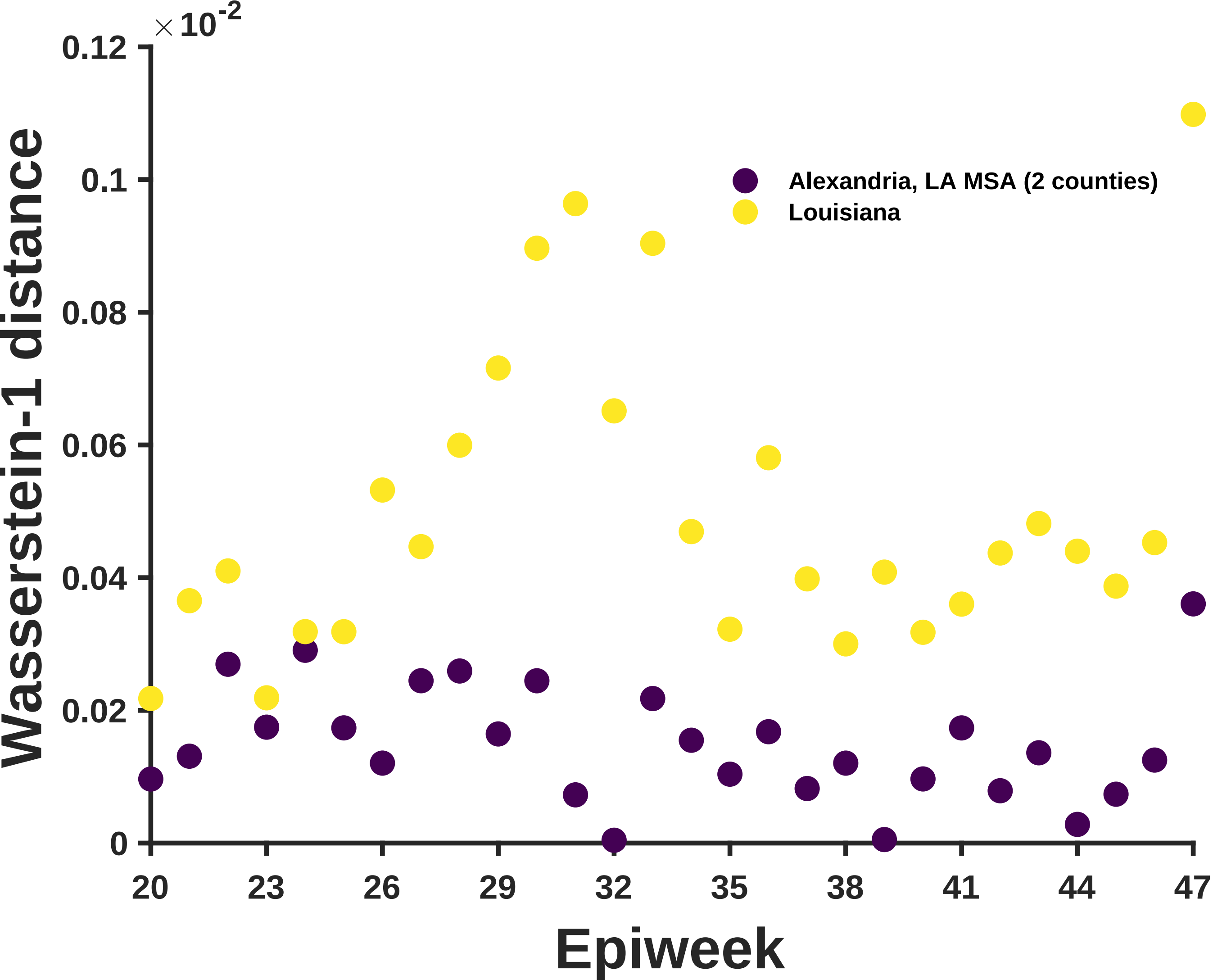

×10^-2
0.12
0.1
0.08
0.06
0.04
0.02
0
Wasserstein-1 distance
Alexandria, LA MSA (2 counties)
Louisiana
20
23
26
29
32
35
38
41
44
47
Epiweek

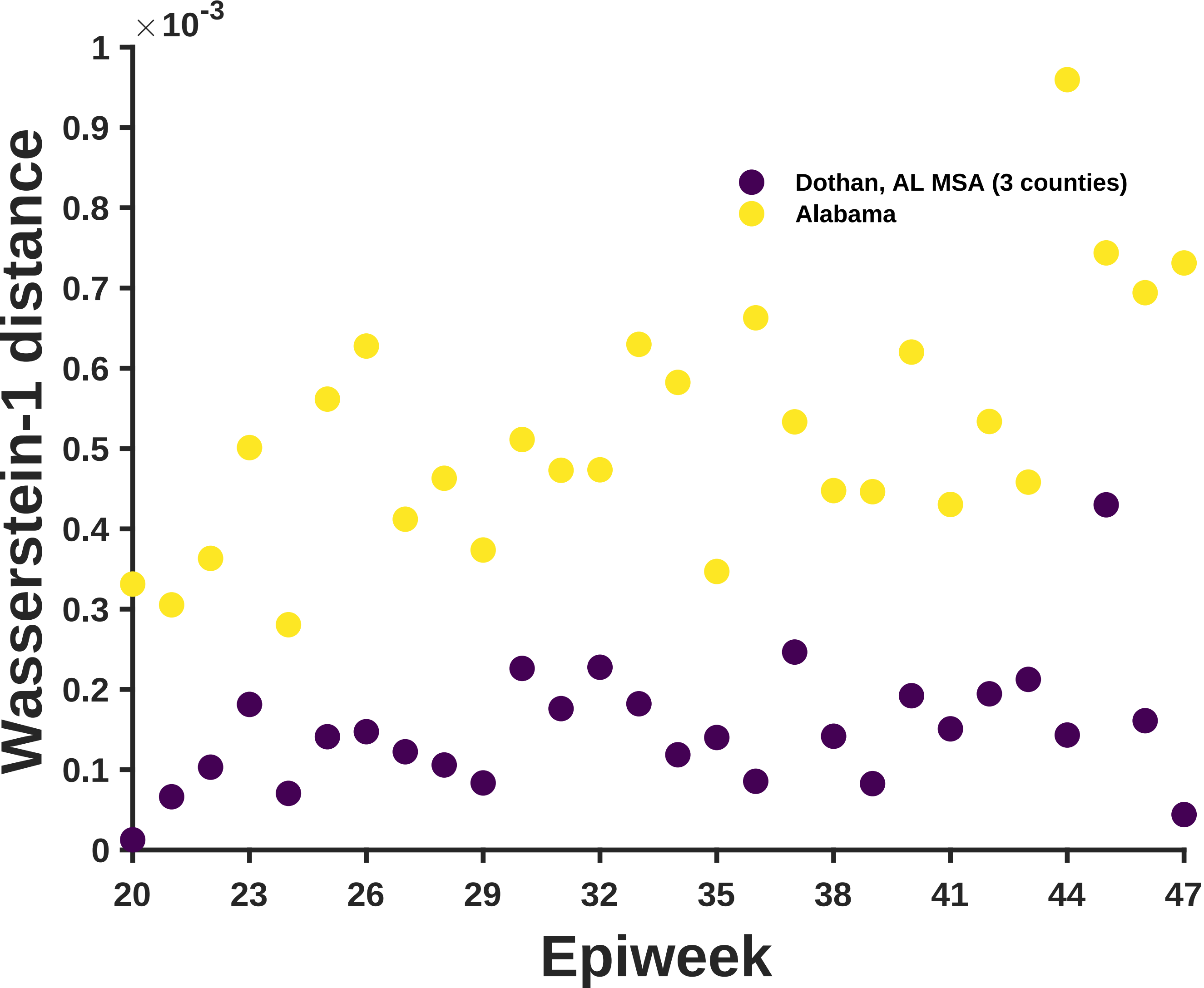

×10^-3
1
0.9
0.8
0.7
0.6
0.5
0.4
0.3
0.2
0.1
0
Wasserstein-1 distance
Dothan, AL MSA (3 counties)
Alabama
20
23
26
29
32
35
38
41
44
47
Epiweek

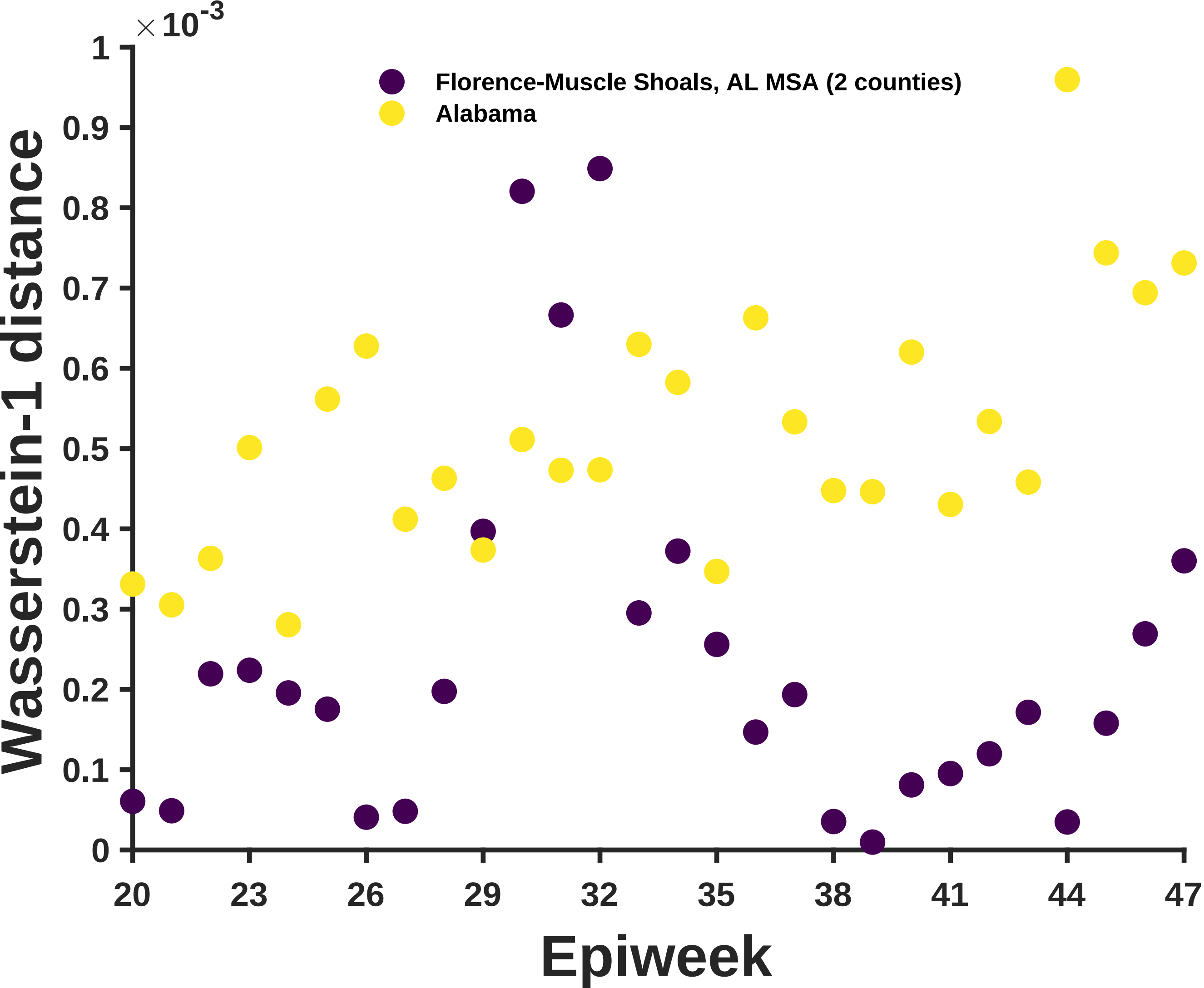
×10^-3
Florence-Muscle Shoals, AL MSA (2 counties)
Alabama
Wasserstein-1 distance
Epiweek
0
0.1
0.2
0.3
0.4
0.5
0.6
0.7
0.8
0.9
1
20
23
26
29
32
35
38
41
44
47

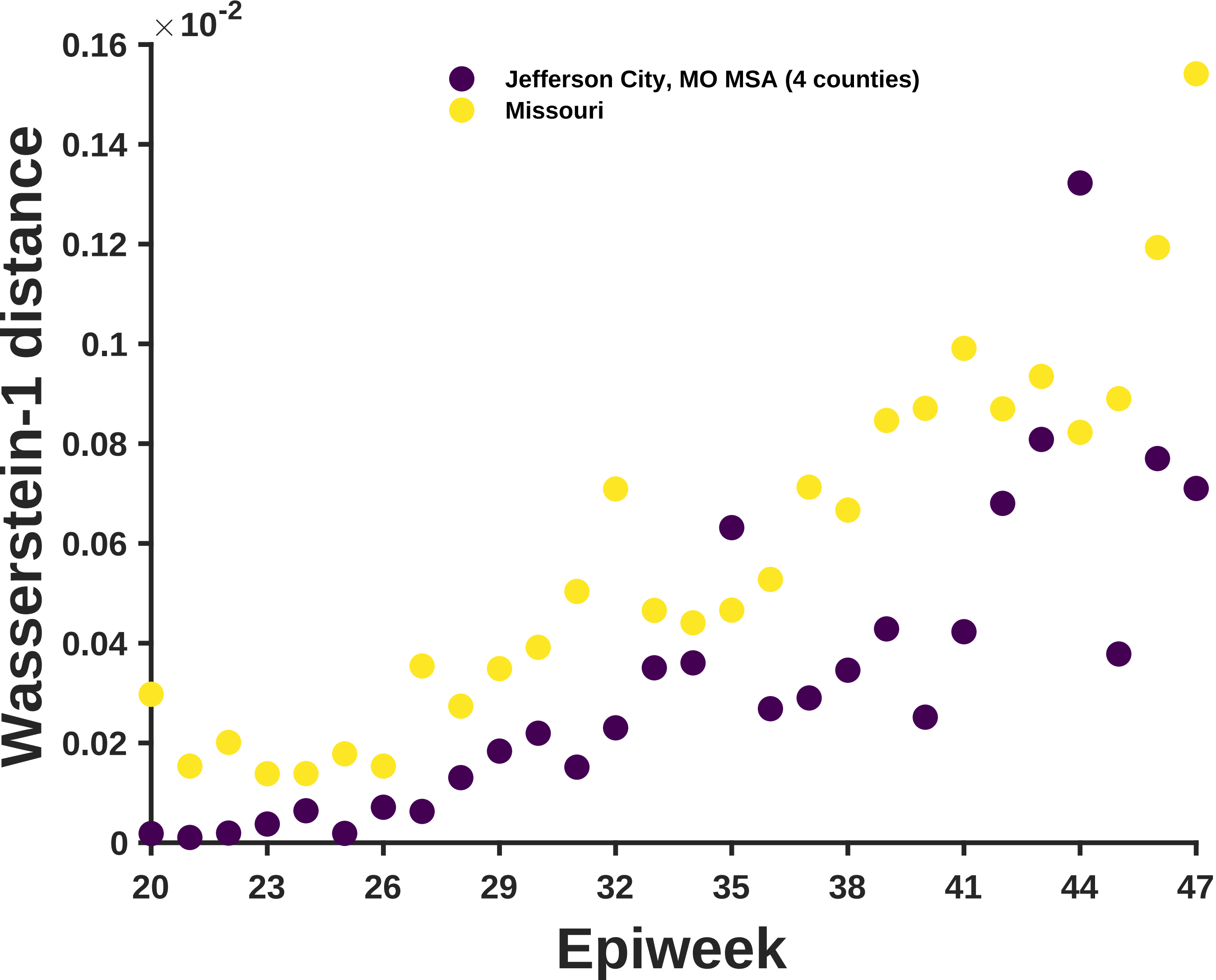

×10^-2
0.16
0.14
0.12
0.1
0.08
0.06
0.04
0.02
0
Wasserstein-1 distance
Jefferson City, MO MSA (4 counties)
Missouri
20
23
26
29
32
35
38
41
44
47
Epiweek

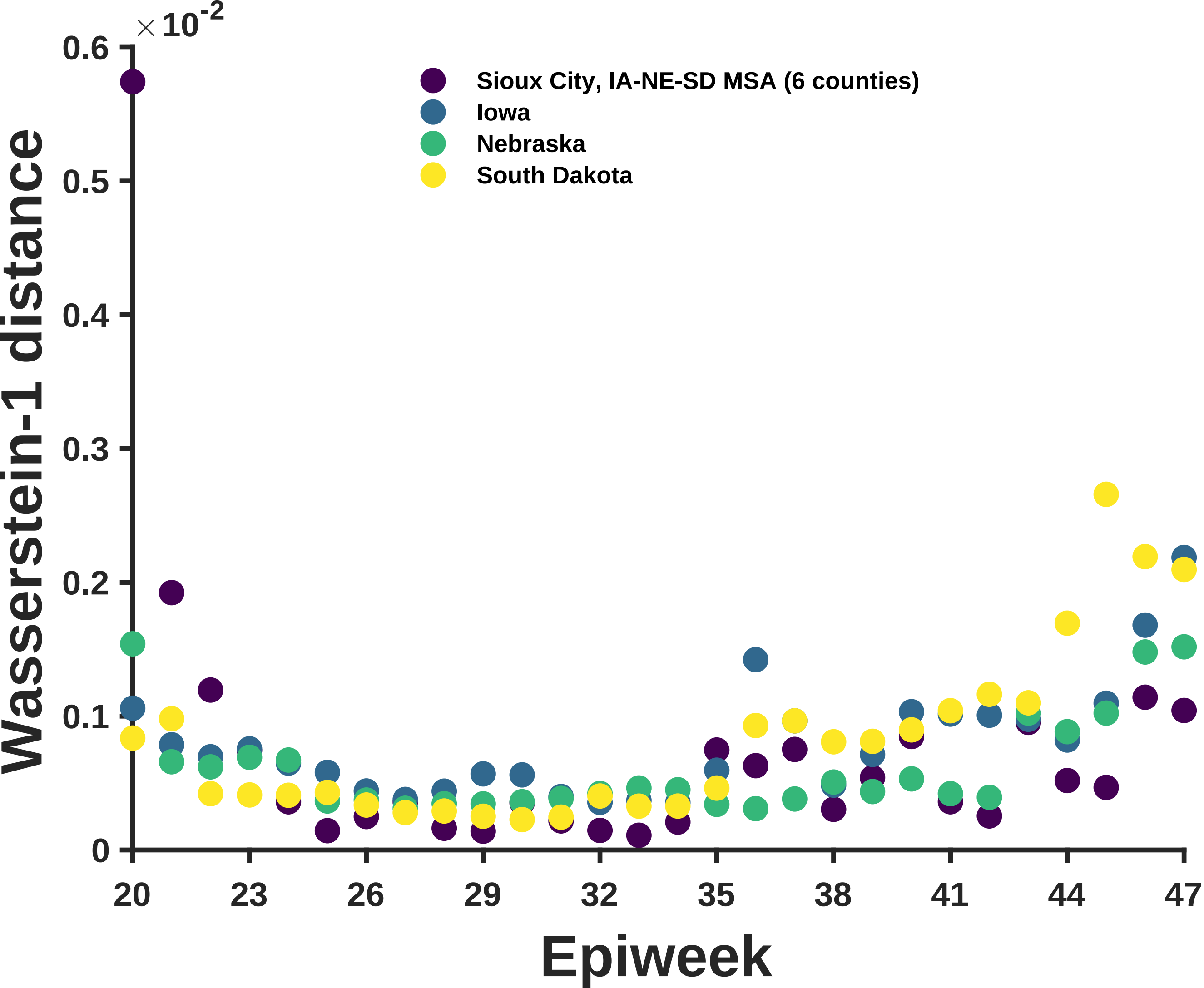

×10^-2
0.6
0.5
0.4
0.3
0.2
0.1
0
Wasserstein-1 distance
Sioux City, IA-NE-SD MSA (6 counties)
Iowa
Nebraska
South Dakota
20
23
26
29
32
35
38
41
44
47
Epiweek

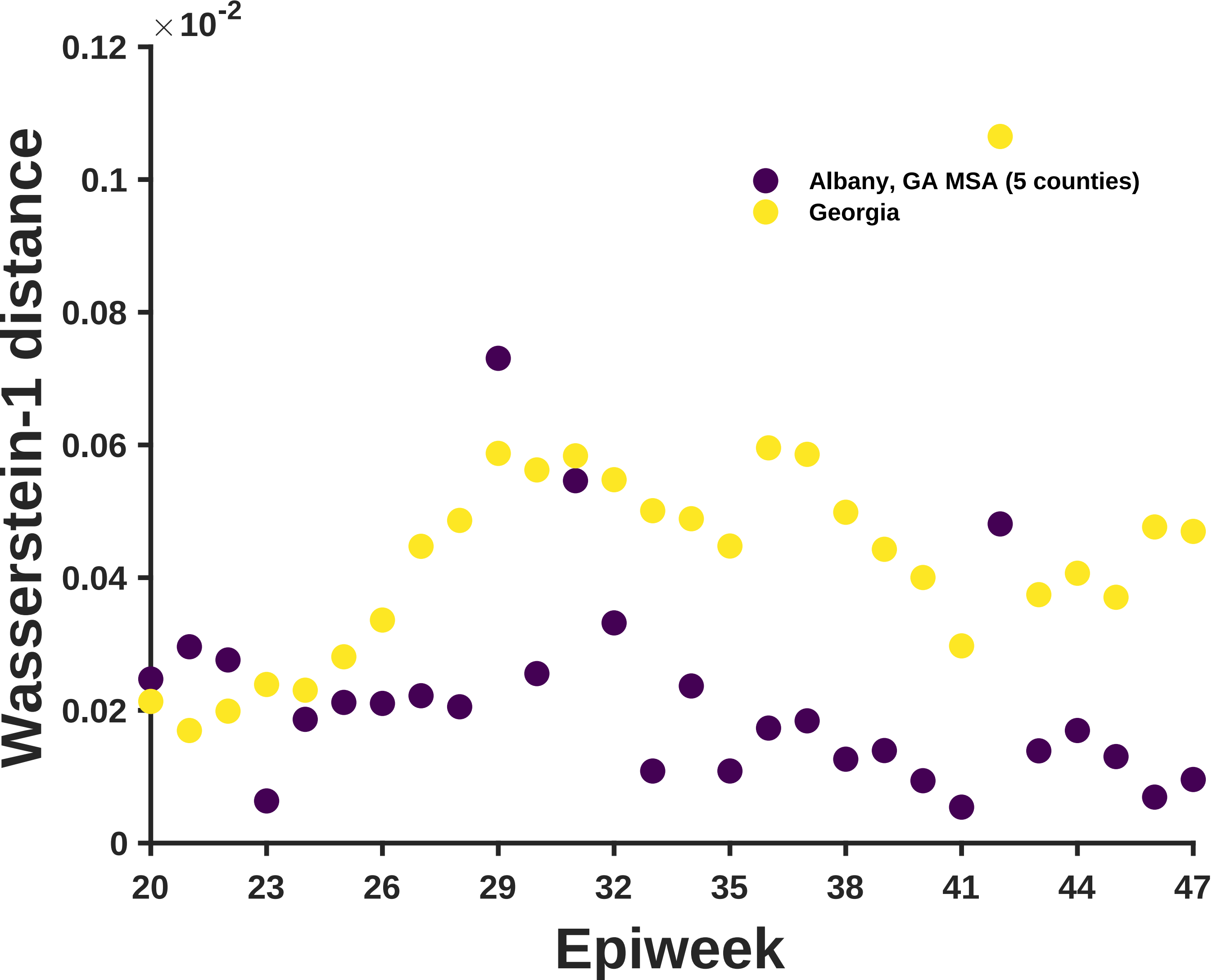

×10^-2
0.12
0.1
0.08
0.06
0.04
0.02
0
Wasserstein-1 distance
Albany, GA MSA (5 counties)
Georgia
20
23
26
29
32
35
38
41
44
47
Epiweek

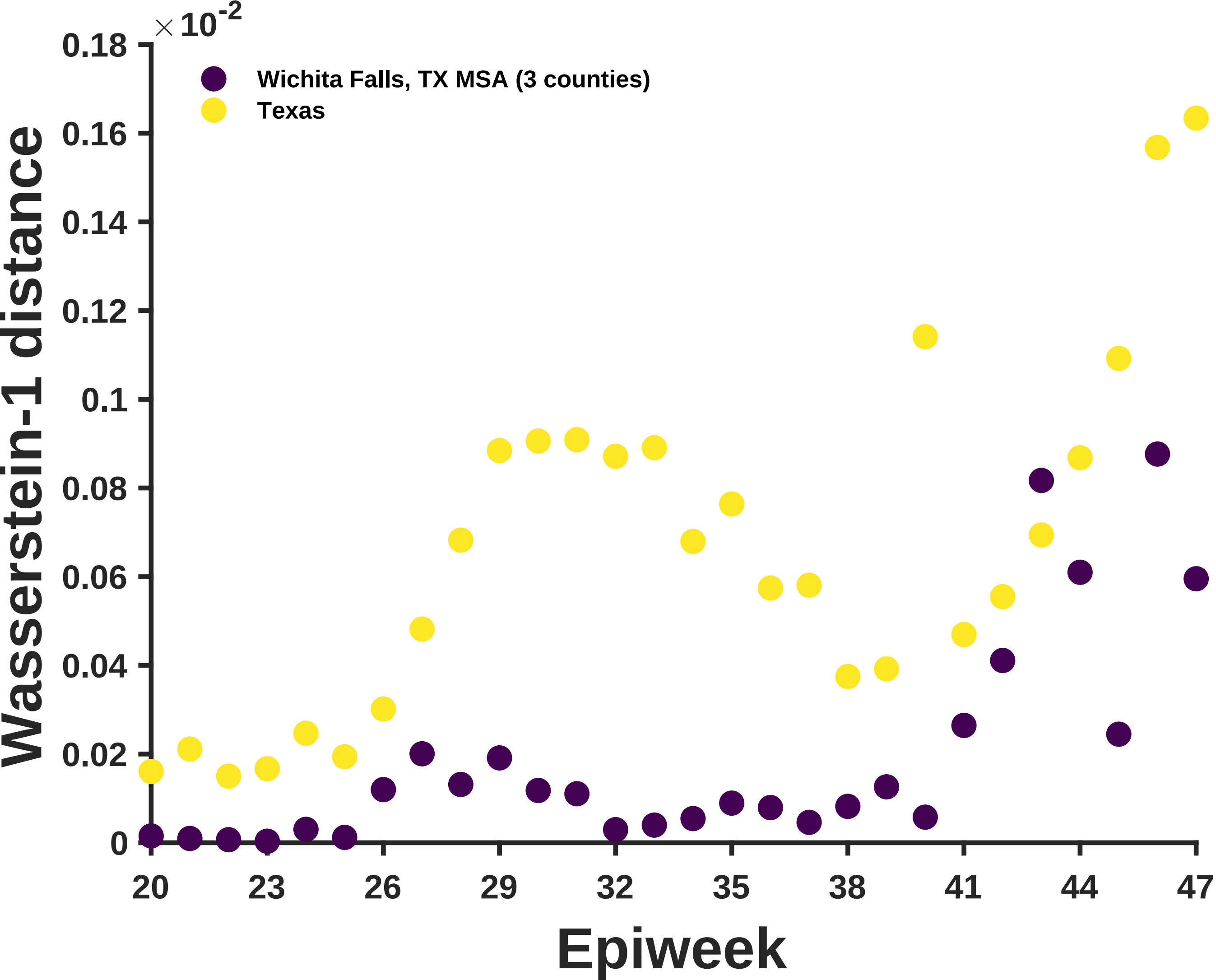

×10^-2
0.18
0.16
0.14
0.12
0.1
0.08
0.06
0.04
0.02
0
Wichita Falls, TX MSA (3 counties)
Texas
Wasserstein-1 distance
20
23
26
29
32
35
38
41
44
47
Epiweek

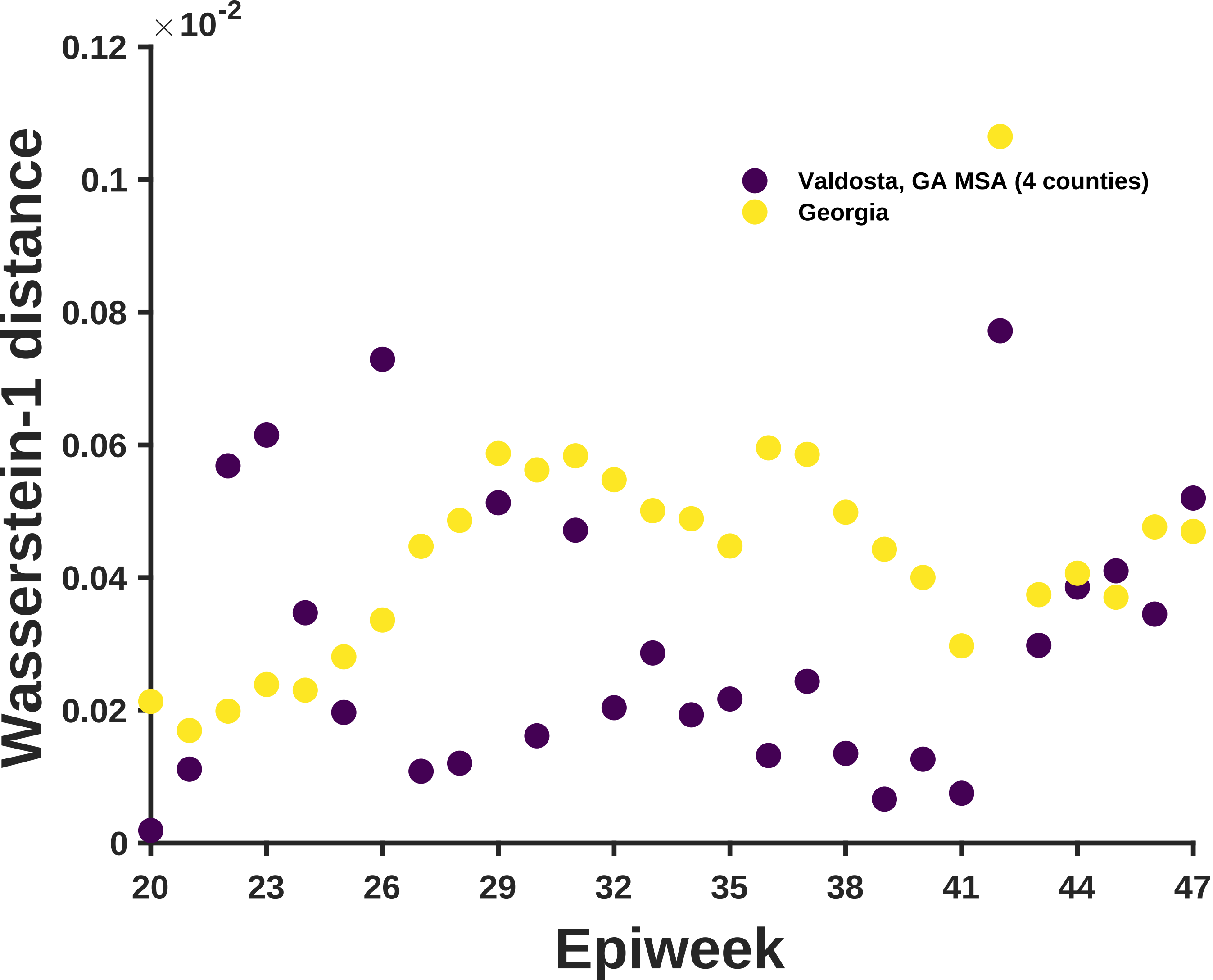

×10^-2
0.12
0.1
0.08
0.06
0.04
0.02
0
Wasserstein-1 distance
Valdosta, GA MSA (4 counties)
Georgia
20
23
26
29
32
35
38
41
44
47
Epiweek

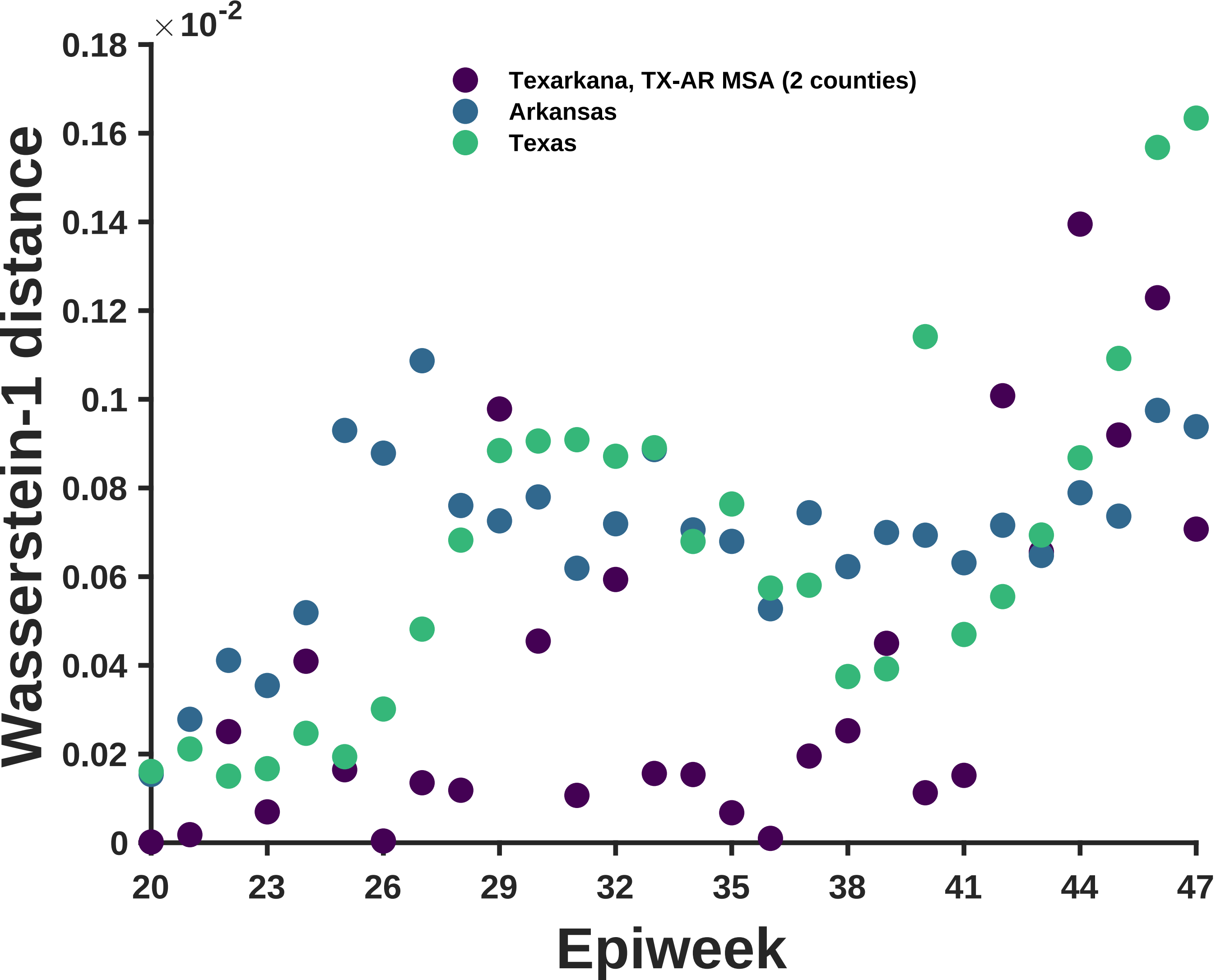

×10^-2
0.18
0.16
0.14
0.12
0.1
0.08
0.06
0.04
0.02
0
Wasserstein-1 distance
Texarkana, TX-AR MSA (2 counties)
Arkansas
Texas
20
23
26
29
32
35
38
41
44
47
Epiweek

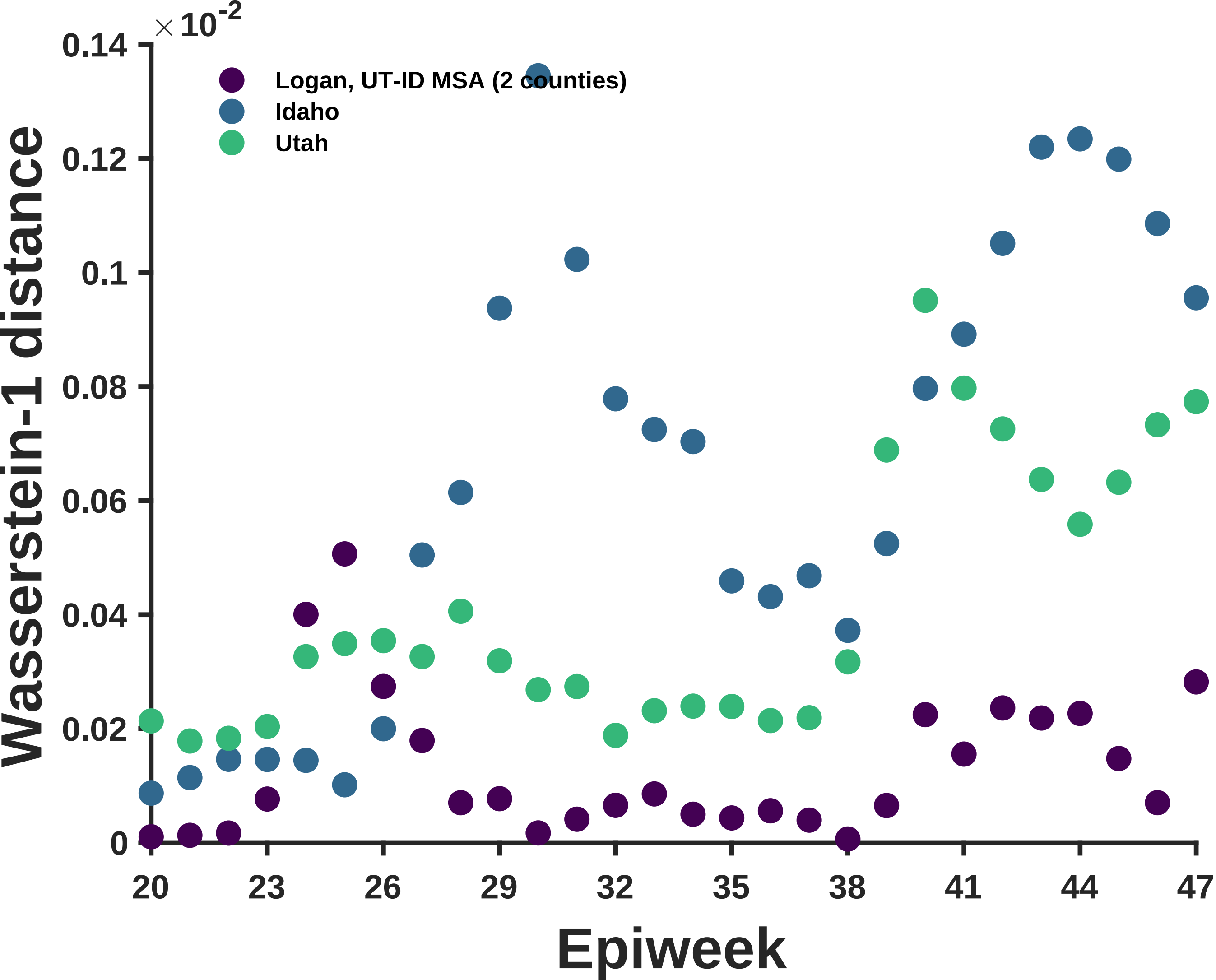

×10^-2
0.14
0.12
0.1
0.08
0.06
0.04
0.02
0
Wasserstein-1 distance
Logan, UT-ID MSA (2 counties)
Idaho
Utah
20
23
26
29
32
35
38
41
44
47
Epiweek

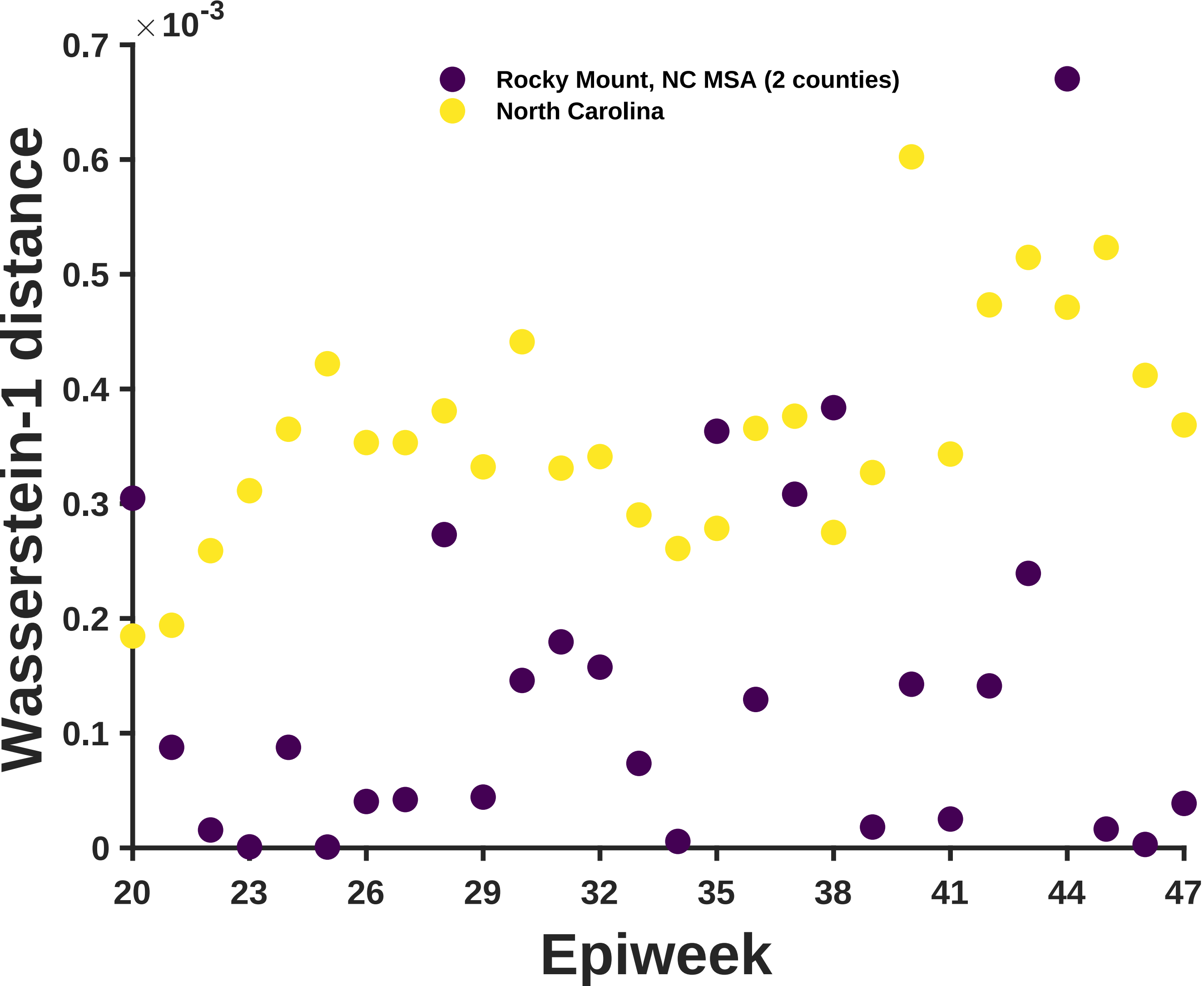

×10^-3
0.7
0.6
0.5
0.4
0.3
0.2
0.1
0
Wasserstein-1 distance
Rocky Mount, NC MSA (2 counties)
North Carolina
20
23
26
29
32
35
38
41
44
47
Epiweek

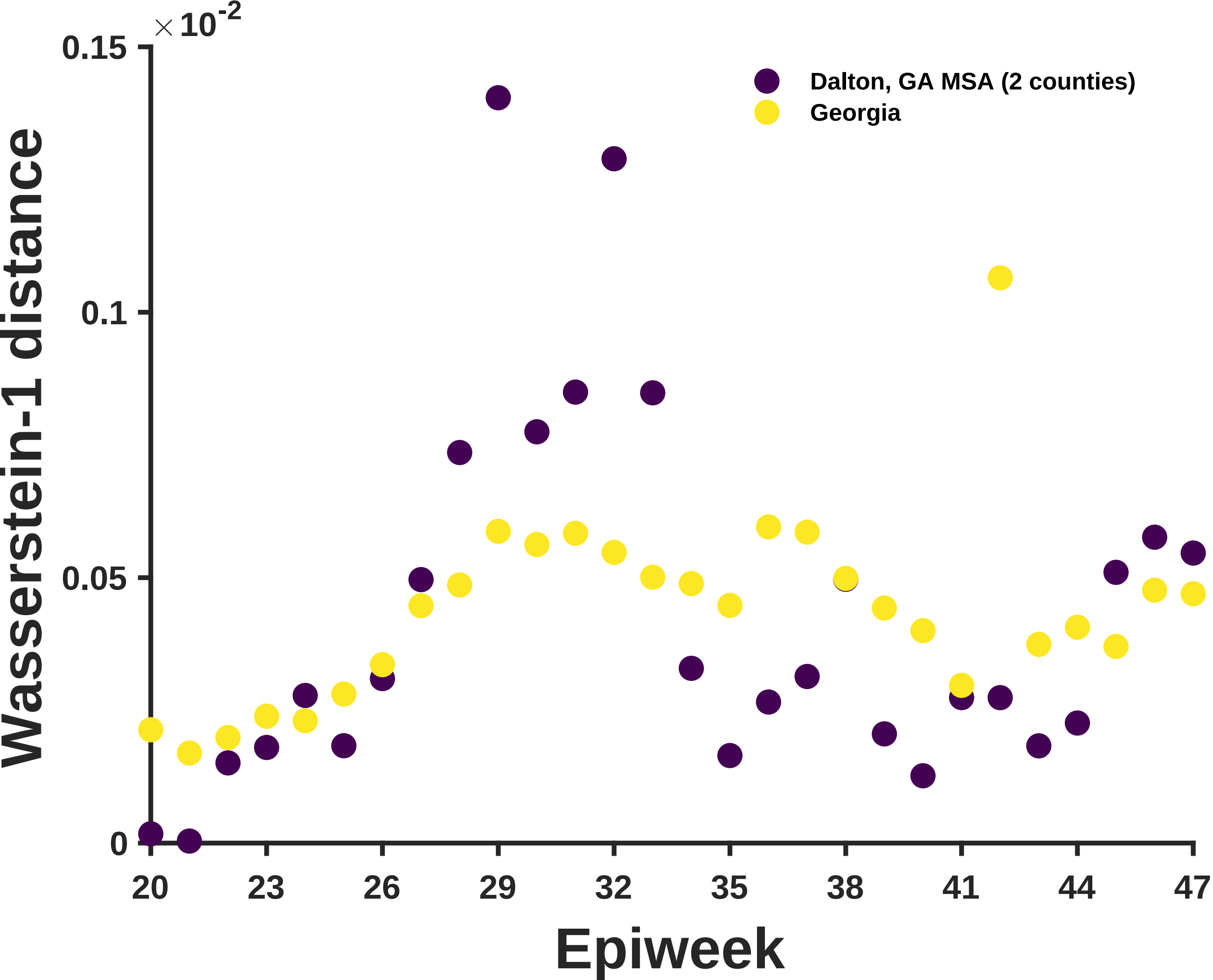

×10^-2
0.15
0.1
0.05
0
Wasserstein-1 distance
Dalton, GA MSA (2 counties)
Georgia
20
23
26
29
32
35
38
41
44
47
Epiweek

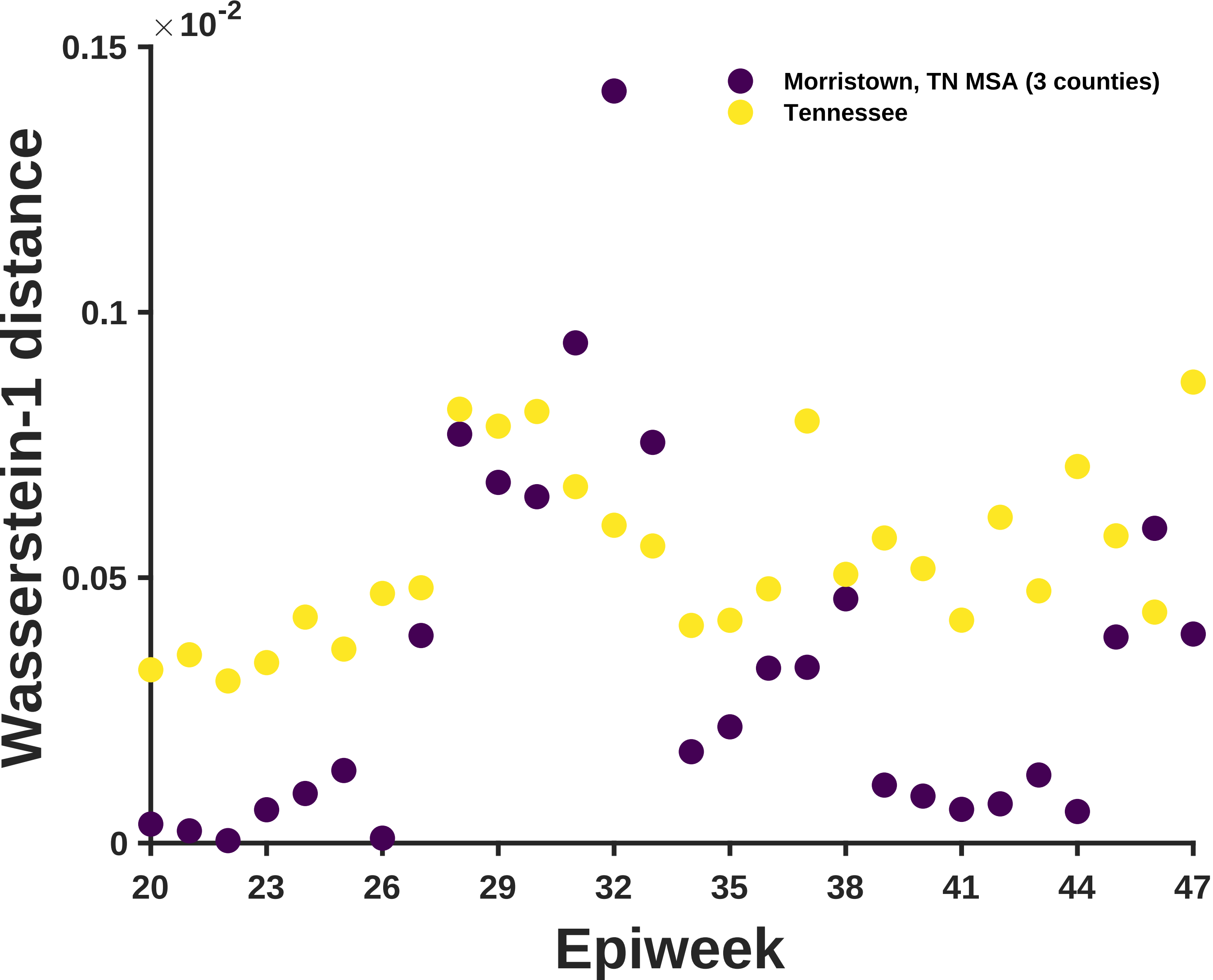

×10^-2
0.15
0.1
0.05
0
Morristown, TN MSA (3 counties)
Tennessee
Wasserstein-1 distance
20
23
26
29
32
35
38
41
44
47
Epiweek

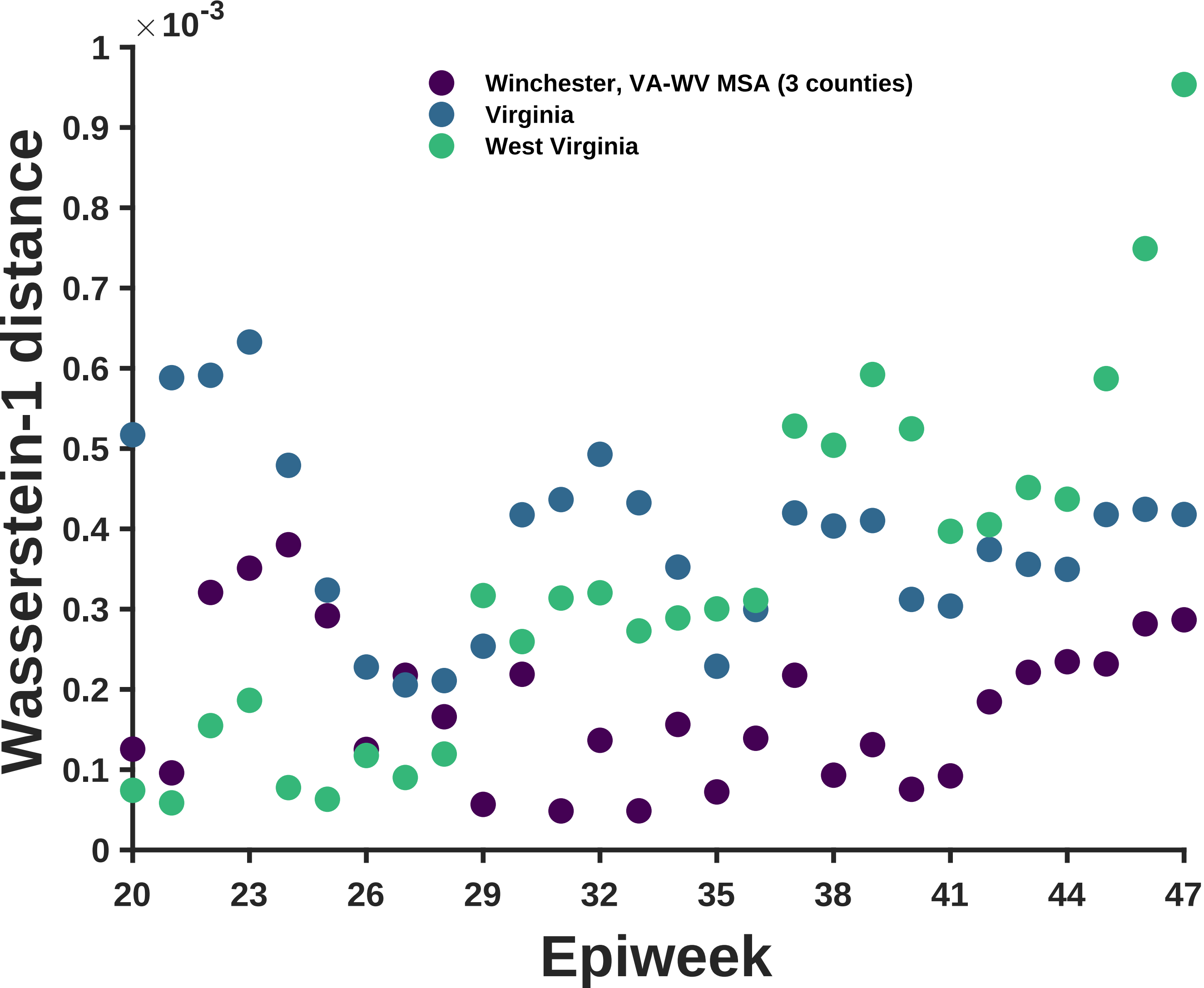

×10^-3
1
0.9
0.8
0.7
0.6
0.5
0.4
0.3
0.2
0.1
0
Wasserstein-1 distance
Winchester, VA-WV MSA (3 counties)
Virginia
West Virginia
20
23
26
29
32
35
38
41
44
47
Epiweek

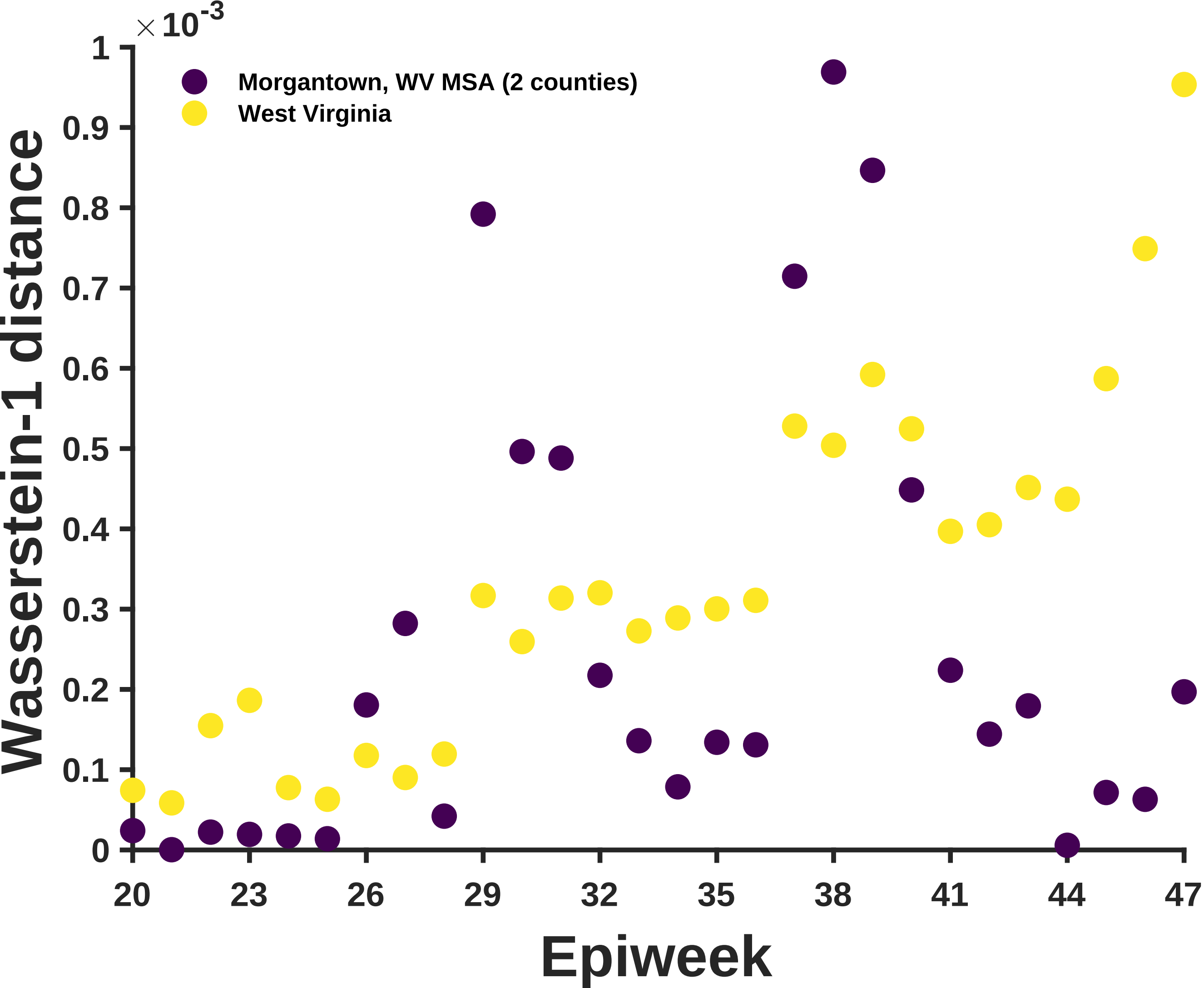
×10-3
Morgantown, WV MSA (2 counties)
West Virginia
Wasserstein-1 distance
Epiweek
0
0.1
0.2
0.3
0.4
0.5
0.6
0.7
0.8
0.9
1
20
23
26
29
32
35
38
41
44
47

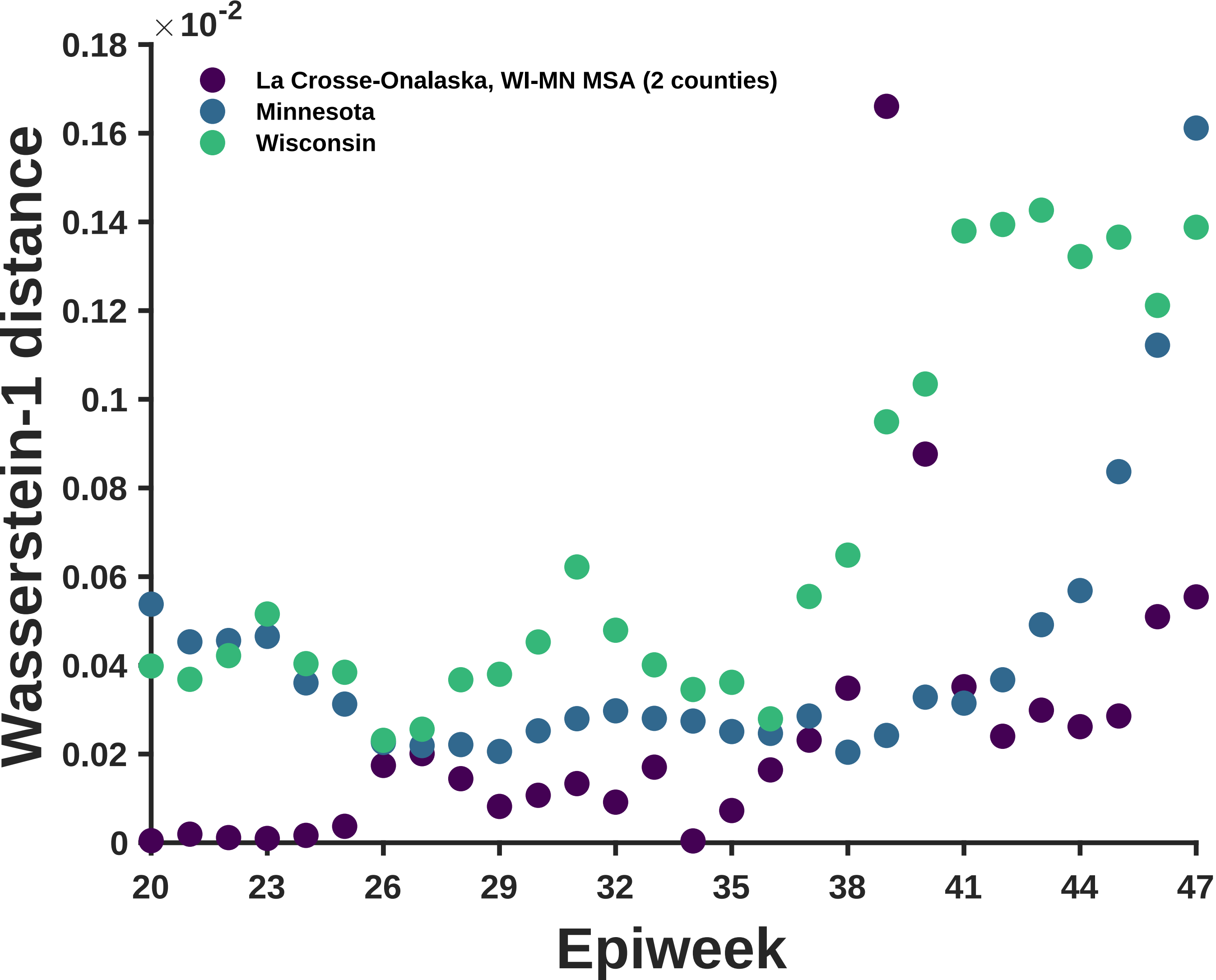

×10^-2
0.18
0.16
0.14
0.12
0.1
0.08
0.06
0.04
0.02
0
Wasserstein-1 distance
La Crosse-Onalaska, WI-MN MSA (2 counties)
Minnesota
Wisconsin
20
23
26
29
32
35
38
41
44
47
Epiweek

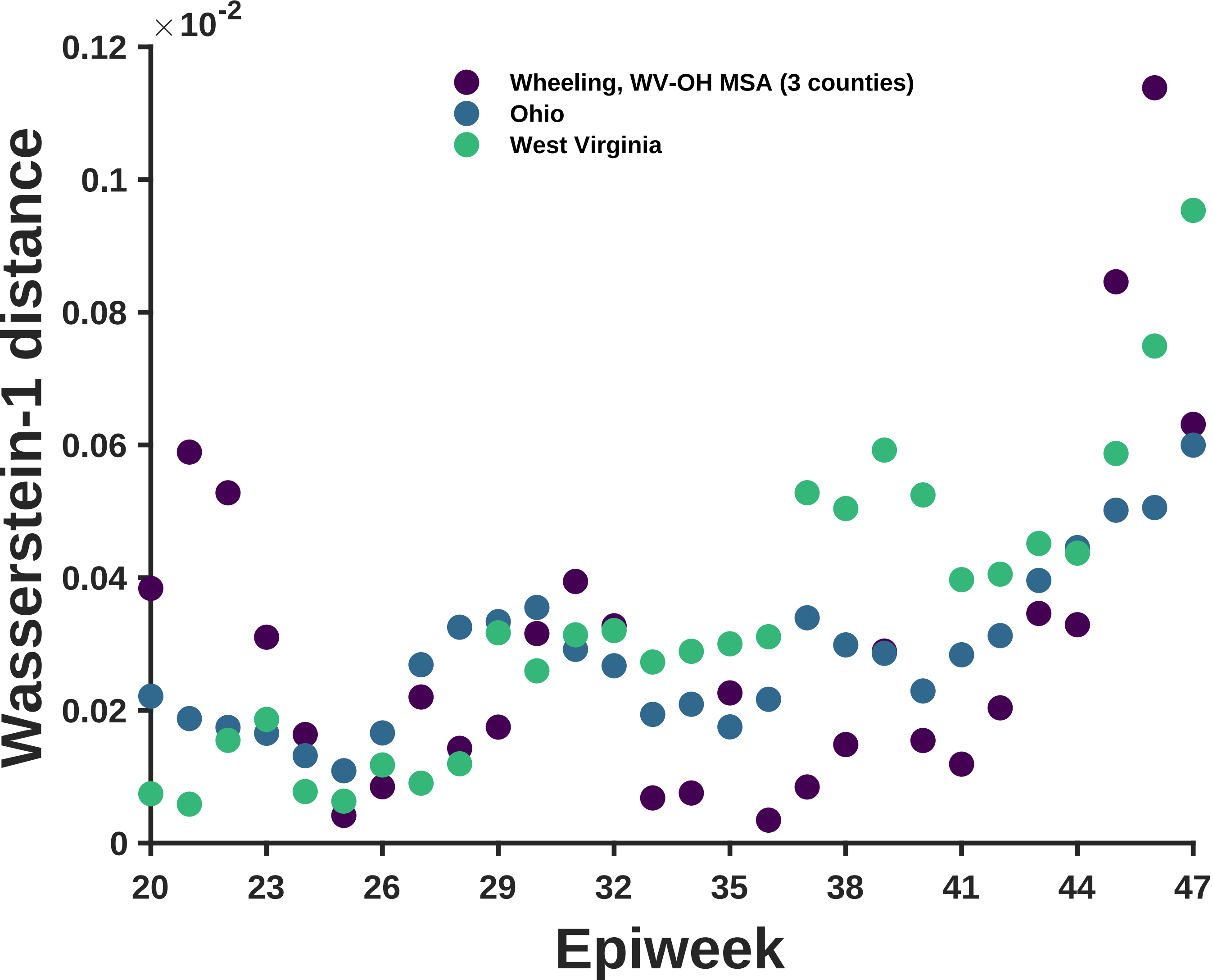

$\times 10^{-2}$
0.12
0.1
0.08
0.06
0.04
0.02
0
Wasserstein-1 distance
Wheeling, WV-OH MSA (3 counties)
Ohio
West Virginia
20
23
26
29
32
35
38
41
44
47
Epiweek

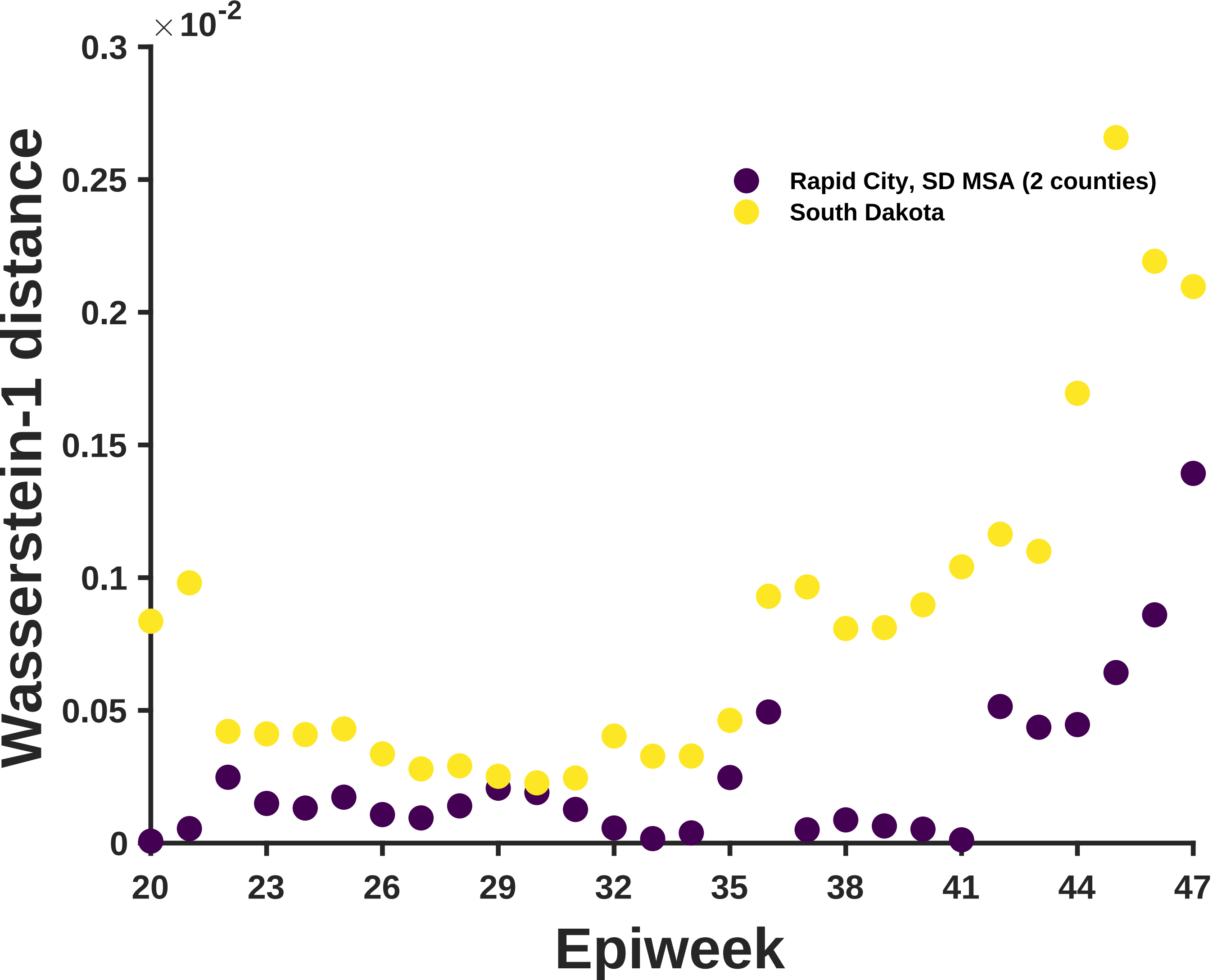

×10^-2
0.3
0.25
0.2
0.15
0.1
0.05
0
Wasserstein-1 distance
Rapid City, SD MSA (2 counties)
South Dakota
20
23
26
29
32
35
38
41
44
47
Epiweek

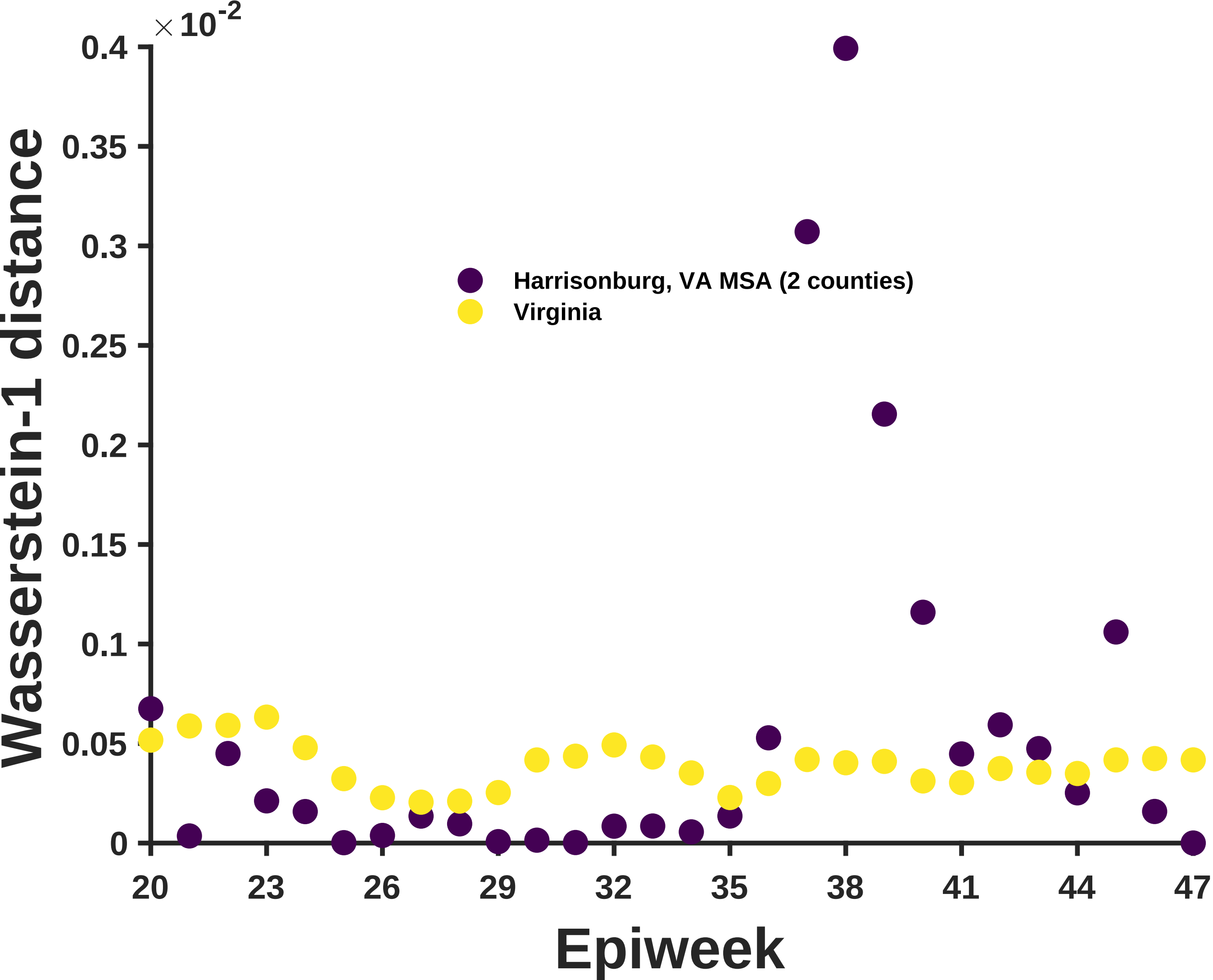
×10-2
0.4
0.35
0.3
0.25
0.2
0.15
0.1
0.05
0
Wasserstein-1 distance
Harrisonburg, VA MSA (2 counties)
Virginia
20
23
26
29
32
35
38
41
44
47
Epiweek

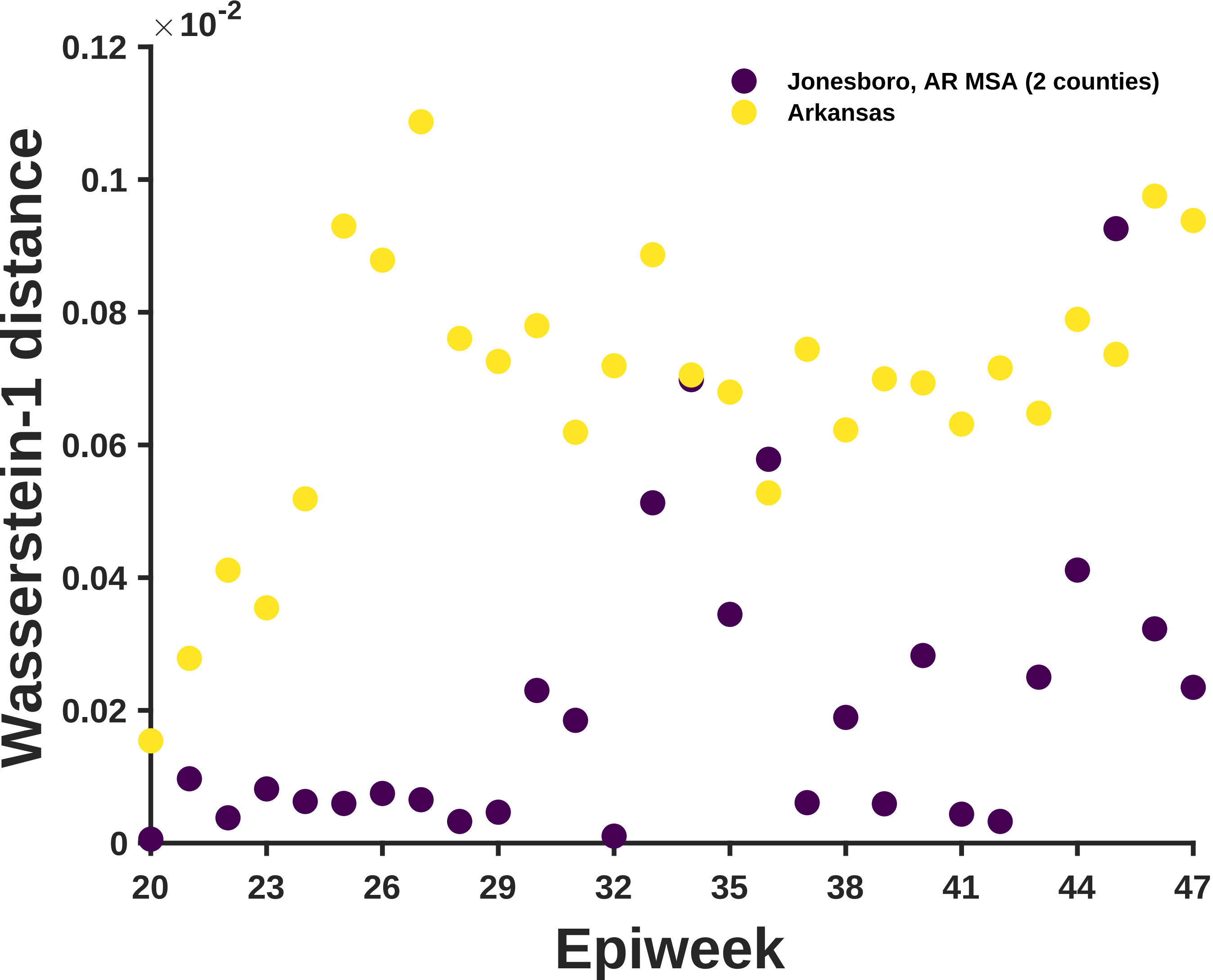

×10^-2
0.12
0.1
0.08
0.06
0.04
0.02
0
Wasserstein-1 distance
Jonesboro, AR MSA (2 counties)
Arkansas
20
23
26
29
32
35
38
41
44
47
Epiweek

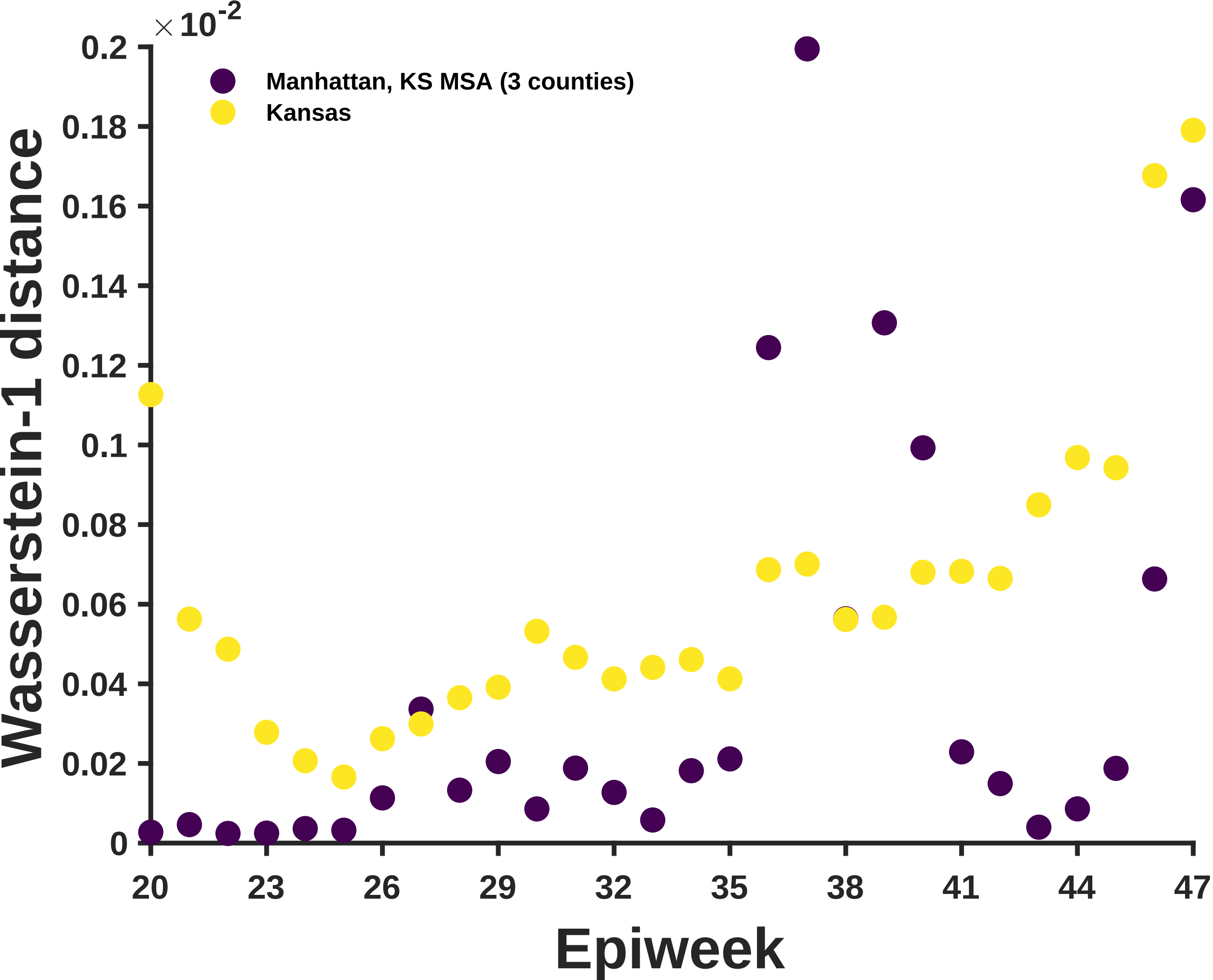

×10^-2
0.2
0.18
0.16
0.14
0.12
0.1
0.08
0.06
0.04
0.02
0
Wasserstein-1 distance
Manhattan, KS MSA (3 counties)
Kansas
20
23
26
29
32
35
38
41
44
47
Epiweek

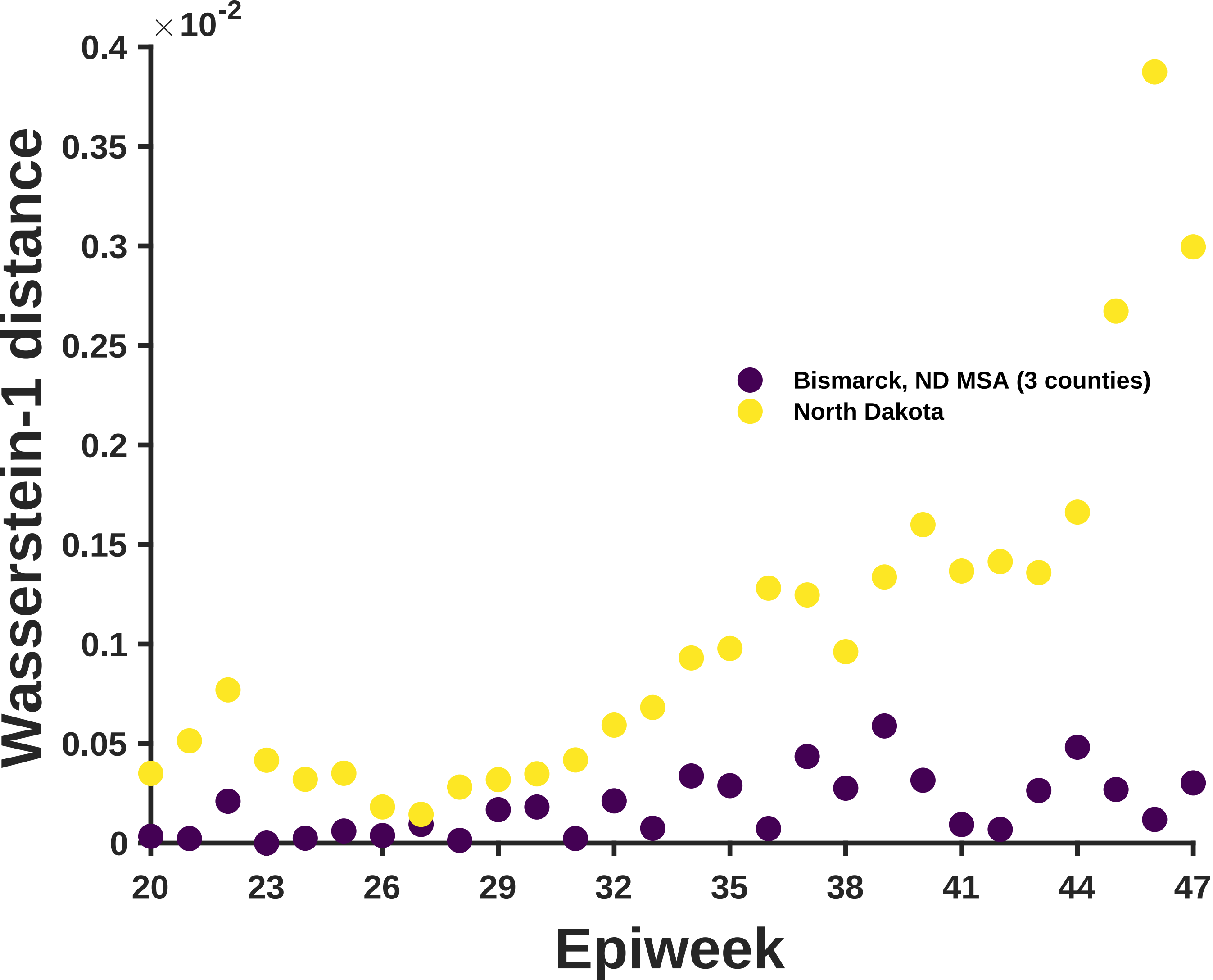

×10^-2
0.4
0.35
0.3
0.25
0.2
0.15
0.1
0.05
0
Wasserstein-1 distance
20
23
26
29
32
35
38
41
44
47
Epiweek
Bismarck, ND MSA (3 counties)
North Dakota

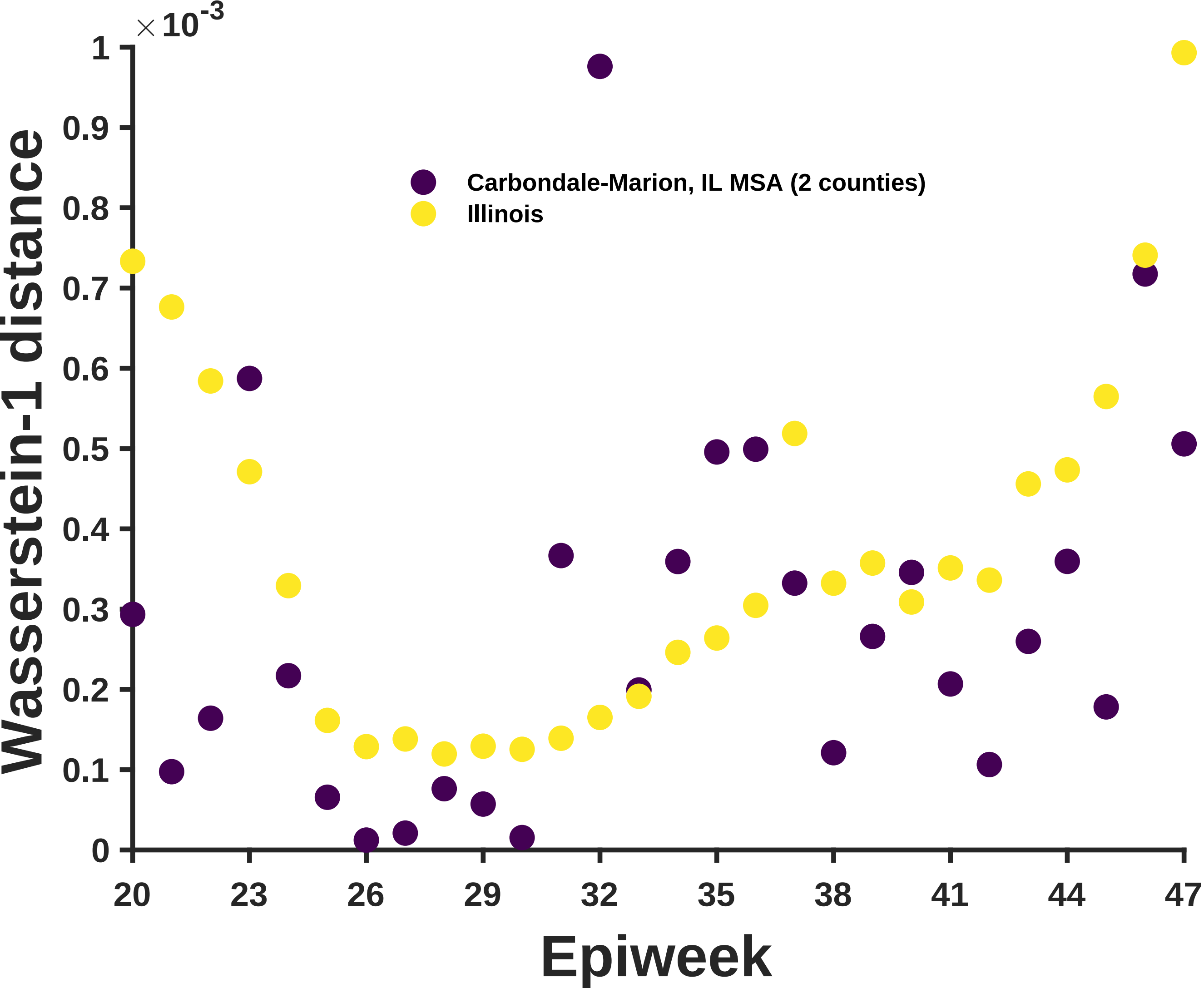

× 10^-3
1
0.9
0.8
0.7
0.6
0.5
0.4
0.3
0.2
0.1
0
Wasserstein-1 distance
Carbondale-Marion, IL MSA (2 counties)
Illinois
20
23
26
29
32
35
38
41
44
47
Epiweek

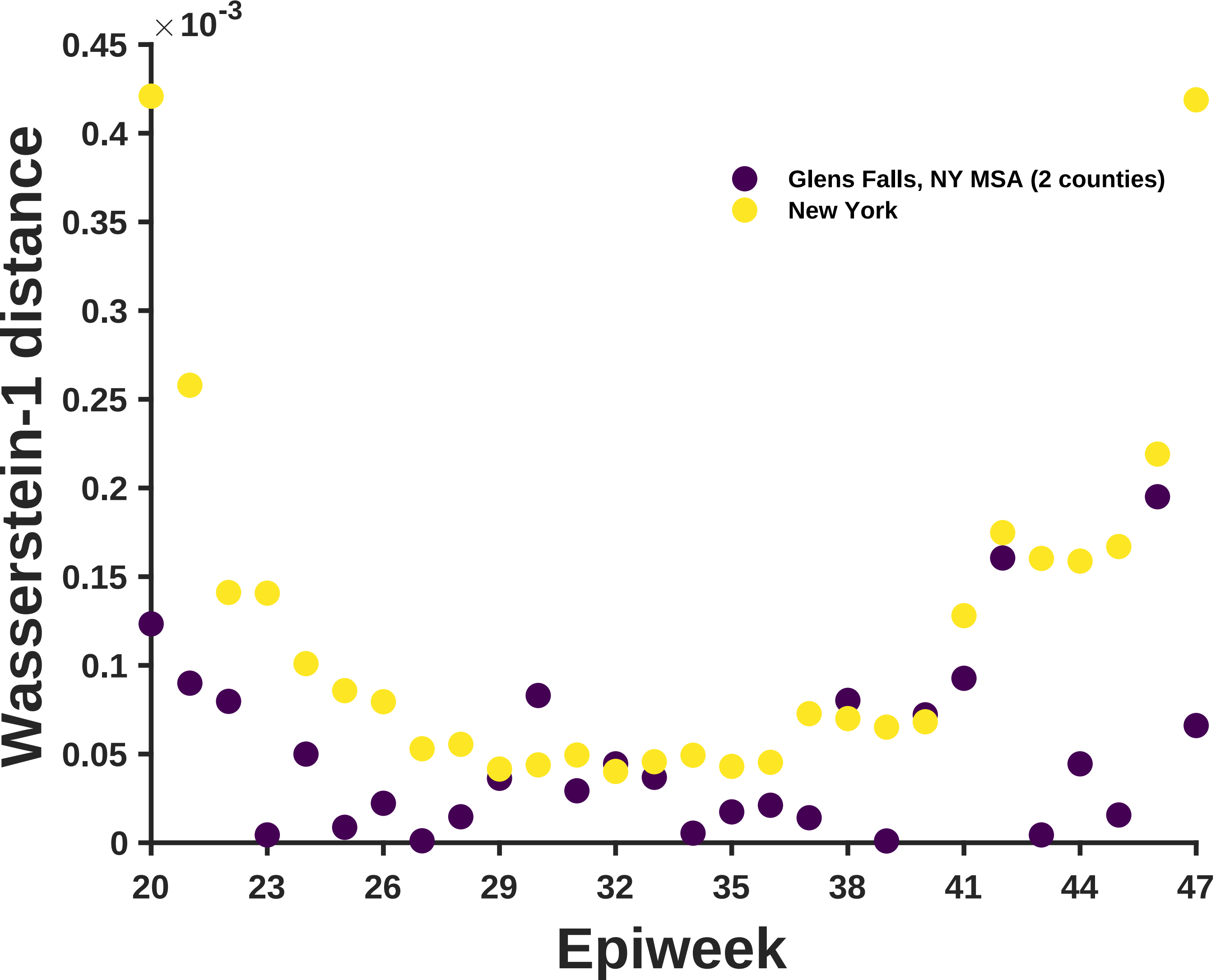
×10^-3
0.45
0.4
0.35
0.3
0.25
0.2
0.15
0.1
0.05
0
Wasserstein-1 distance
20
23
26
29
32
35
38
41
44
47
Epiweek
Glens Falls, NY MSA (2 counties)
New York

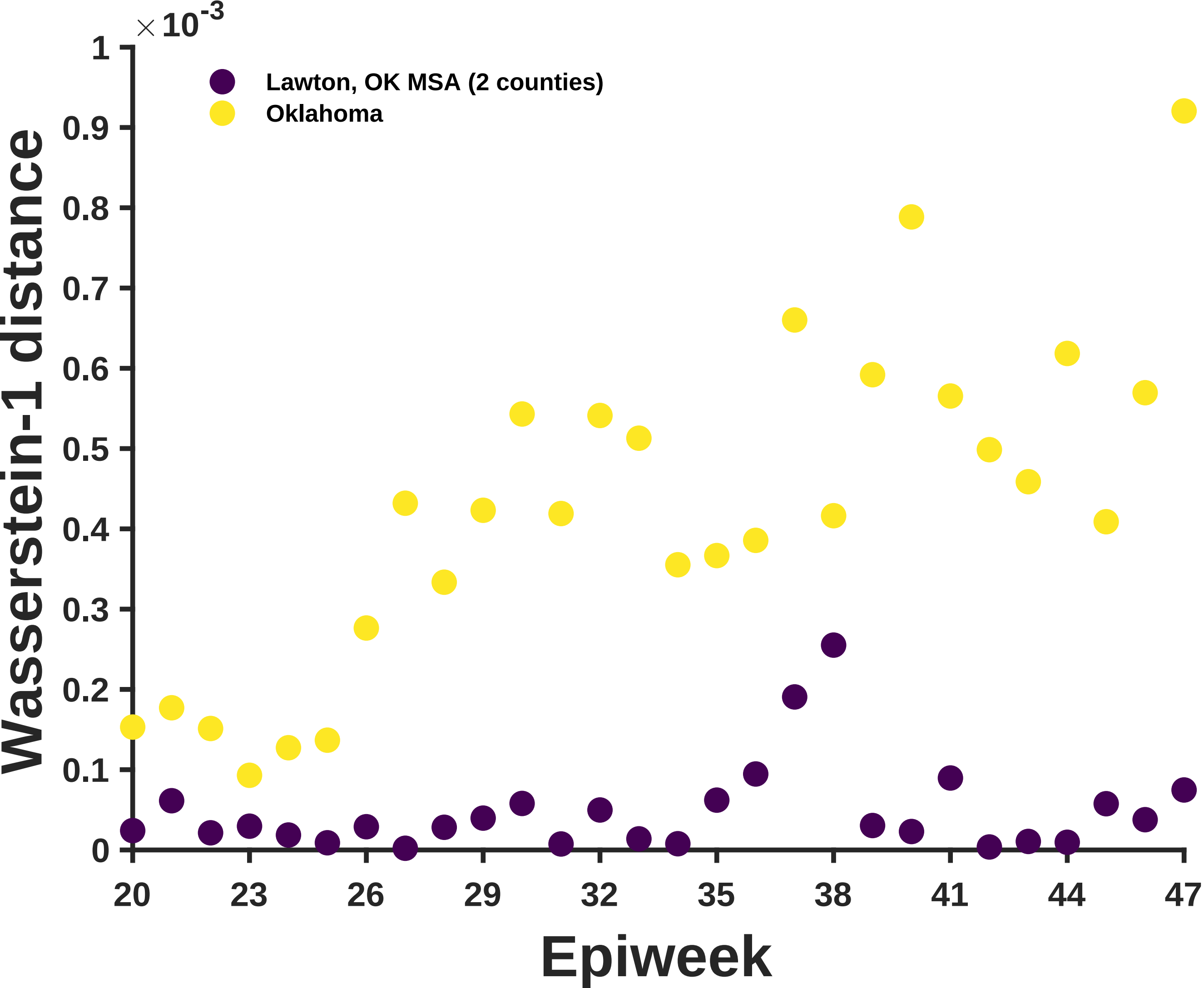

×10^-3
1
0.9
0.8
0.7
0.6
0.5
0.4
0.3
0.2
0.1
0
Wasserstein-1 distance
Lawton, OK MSA (2 counties)
Oklahoma
20
23
26
29
32
35
38
41
44
47
Epiweek

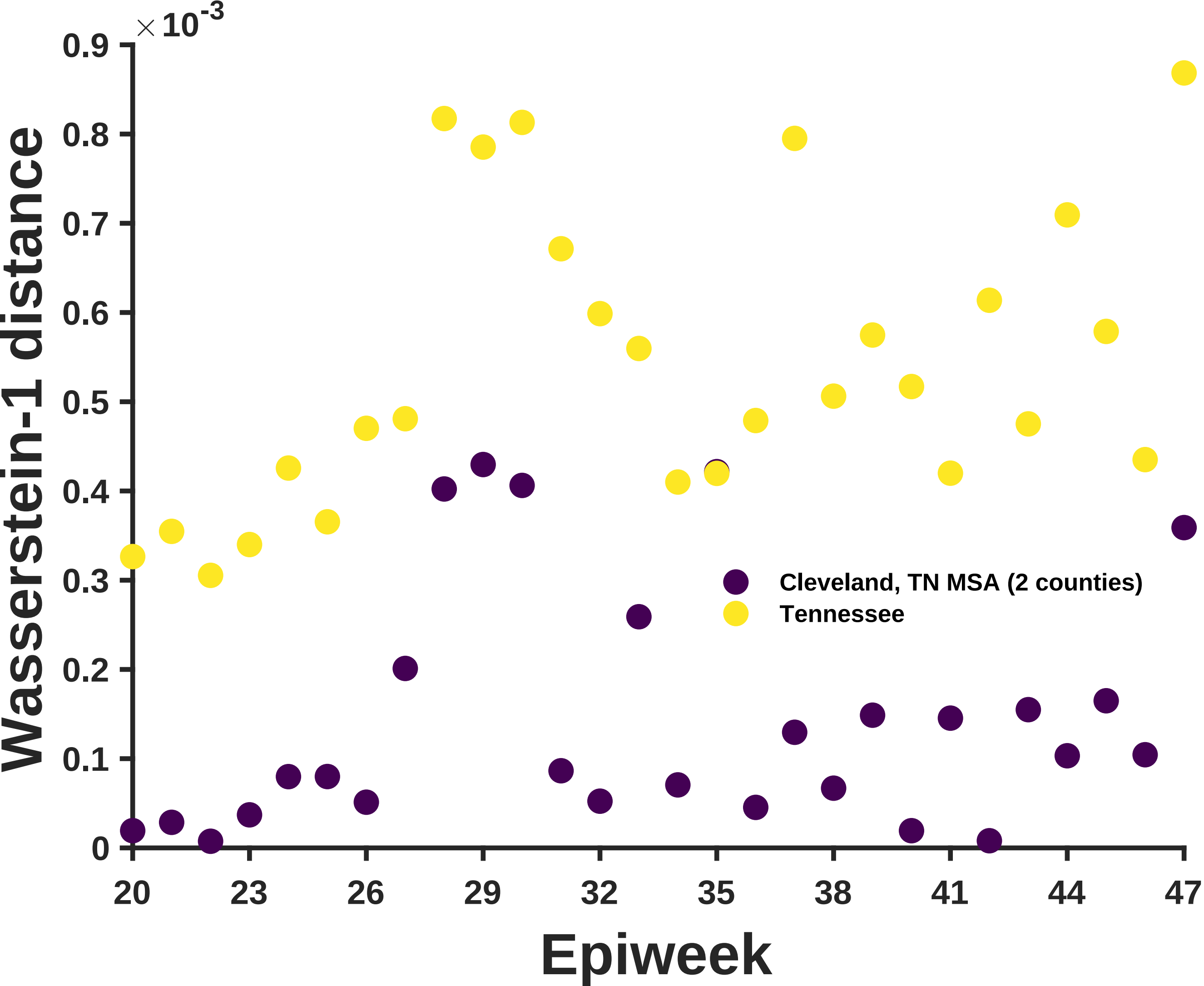

×10^-3
0.9
0.8
0.7
0.6
0.5
0.4
0.3
0.2
0.1
0
Wasserstein-1 distance
20
23
26
29
32
35
38
41
44
47
Epiweek
Cleveland, TN MSA (2 counties)
Tennessee

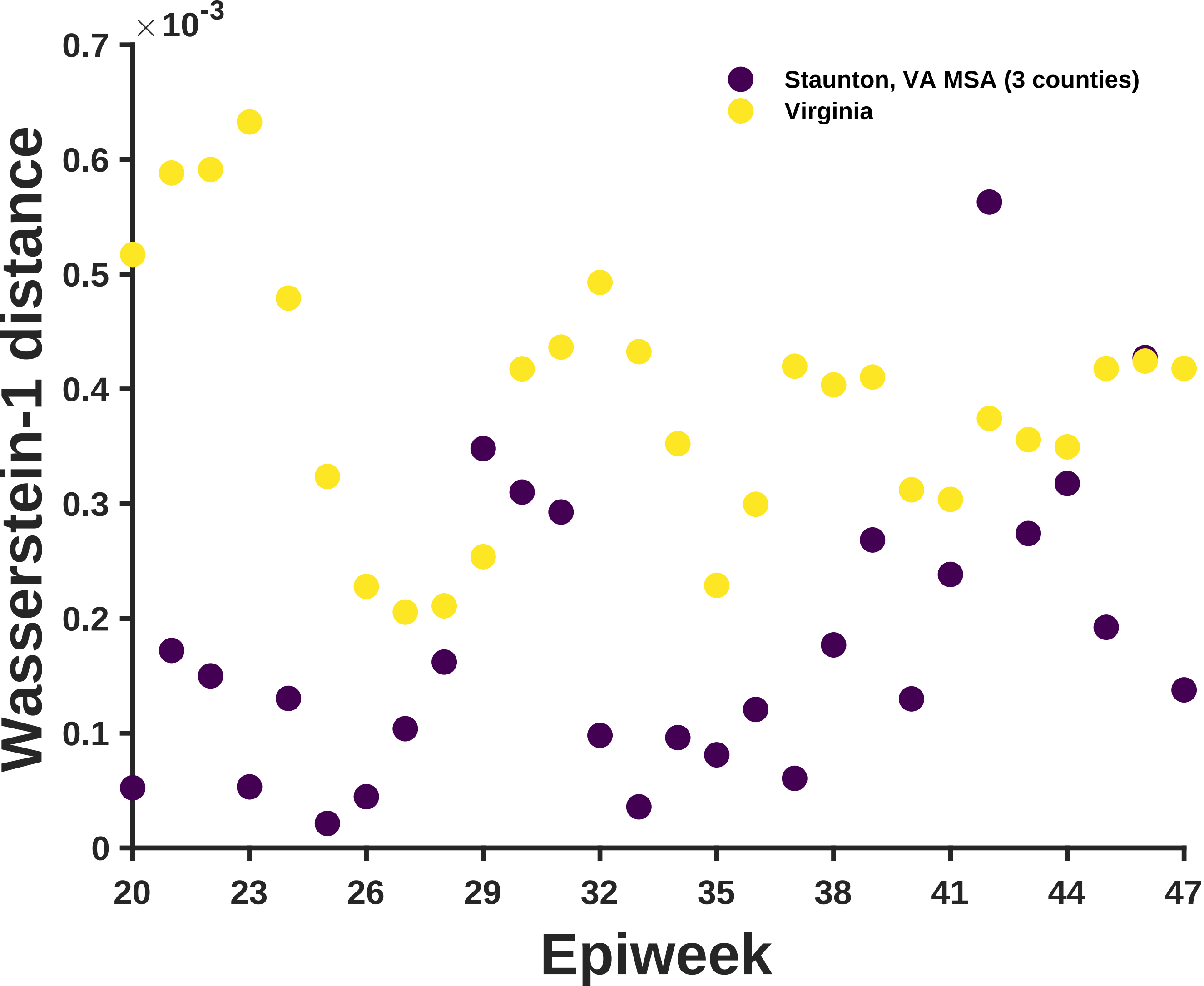

×10^-3
0.7
0.6
0.5
0.4
0.3
0.2
0.1
0
Wasserstein-1 distance
Staunton, VA MSA (3 counties)
Virginia
20
23
26
29
32
35
38
41
44
47
Epiweek

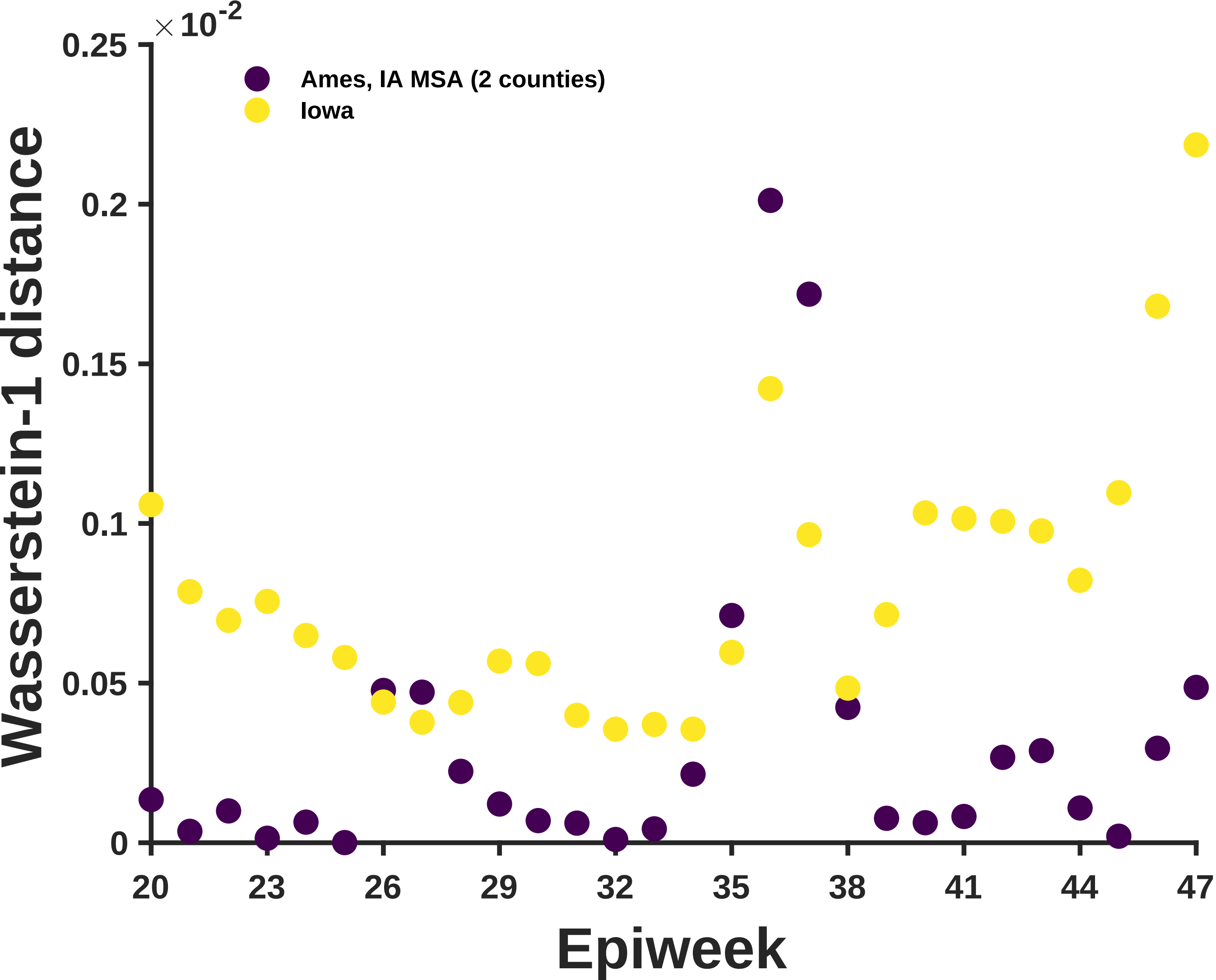

×10^-2
0.25
0.2
0.15
0.1
0.05
0
Wasserstein-1 distance
Ames, IA MSA (2 counties)
Iowa
20
23
26
29
32
35
38
41
44
47
Epiweek

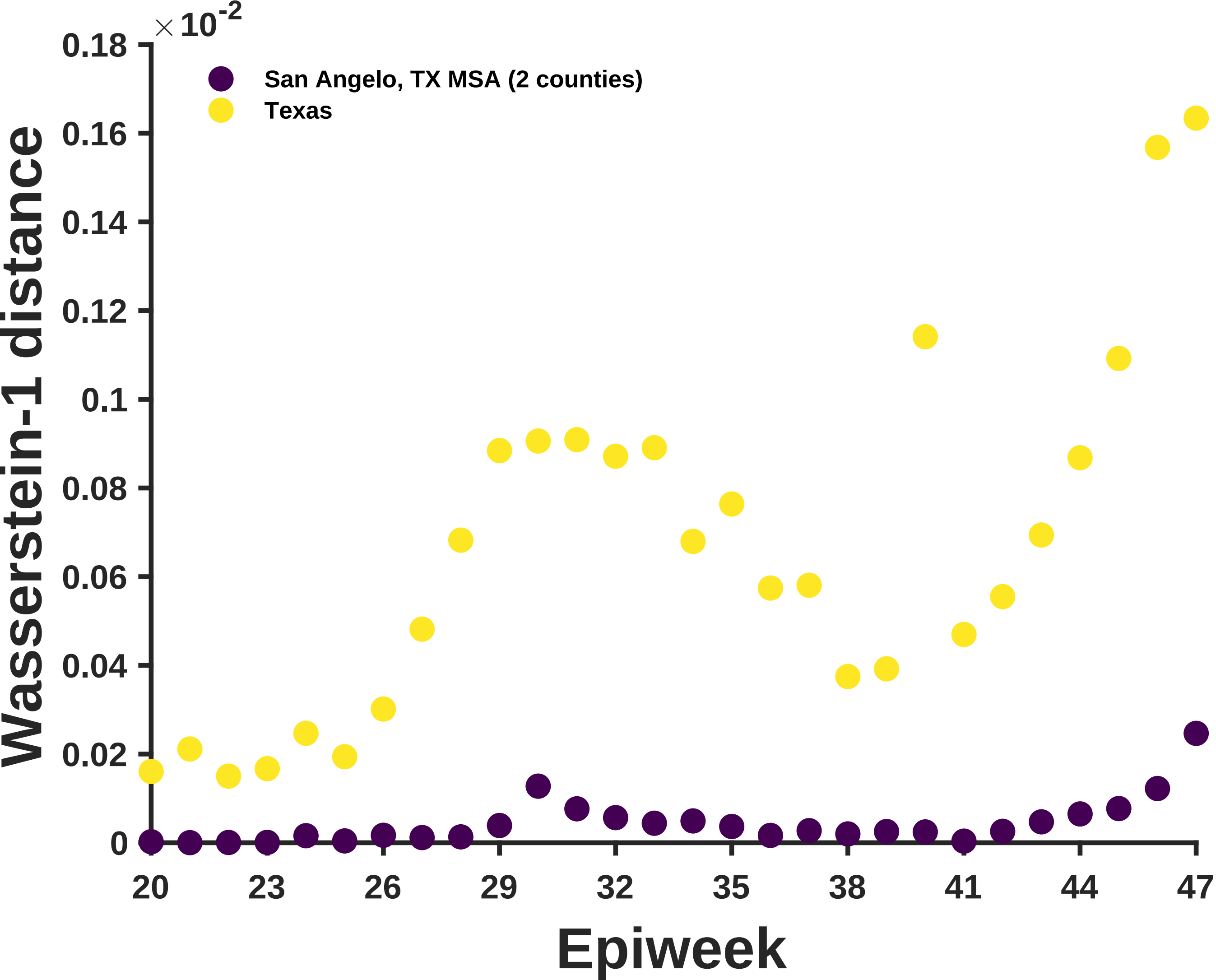

×10^-2
0.18
0.16
0.14
0.12
0.1
0.08
0.06
0.04
0.02
0
San Angelo, TX MSA (2 counties)
Texas
Wasserstein-1 distance
20
23
26
29
32
35
38
41
44
47
Epiweek

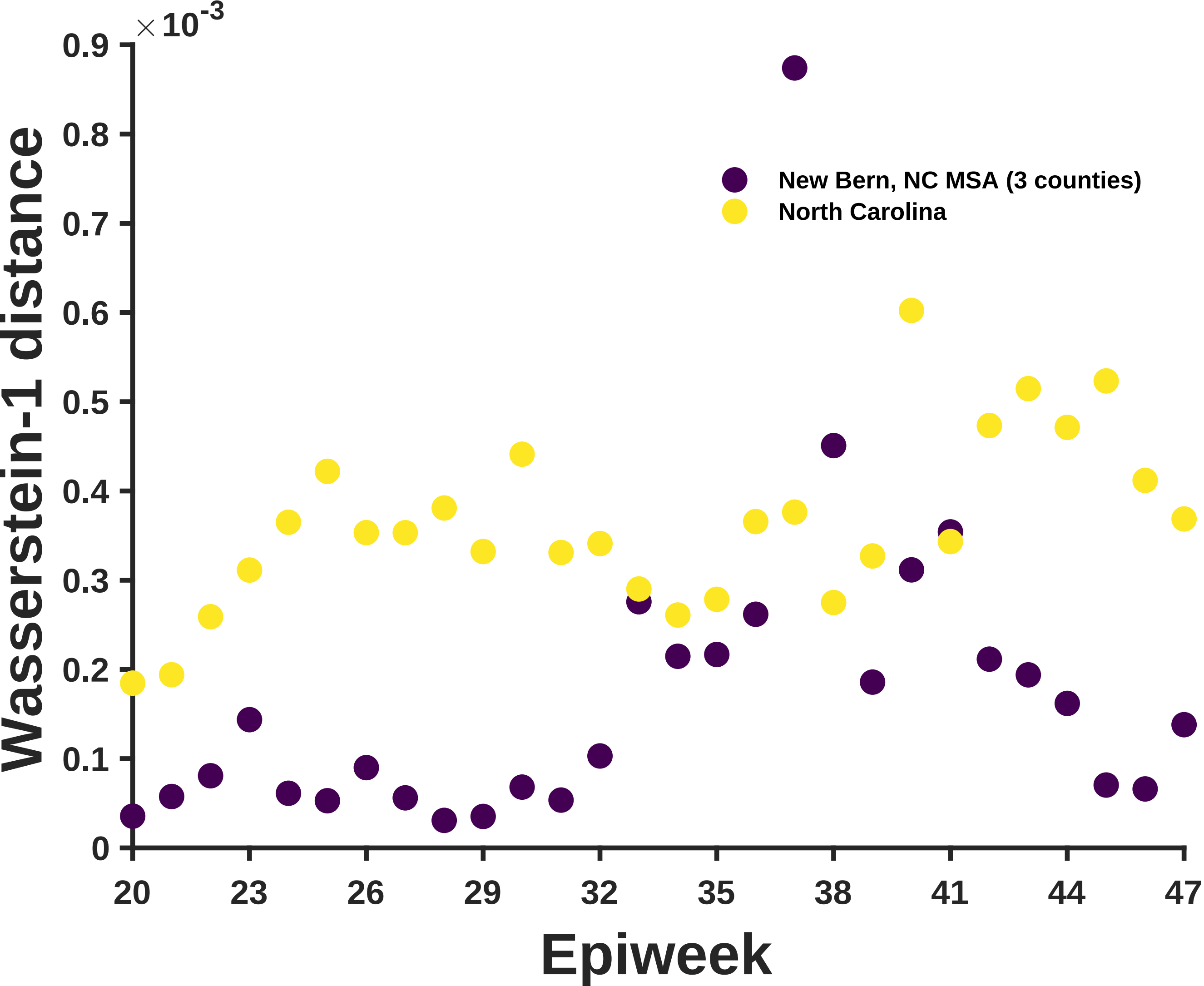
×10^-3
0.9
0.8
0.7
0.6
0.5
0.4
0.3
0.2
0.1
0
Wasserstein-1 distance
New Bern, NC MSA (3 counties)
North Carolina
20
23
26
29
32
35
38
41
44
47
Epiweek

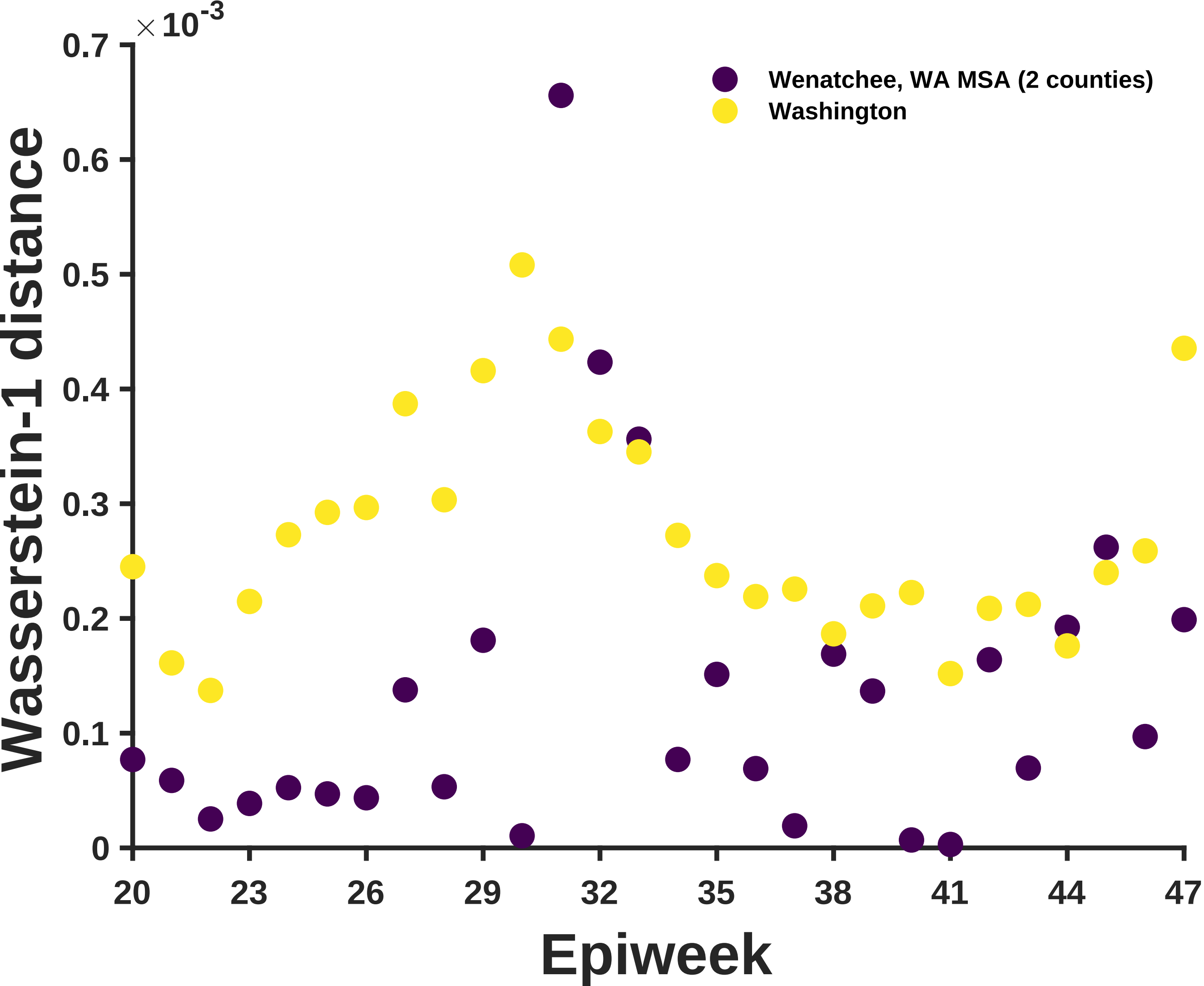

×10^-3
0.7
0.6
0.5
0.4
0.3
0.2
0.1
0
Wasserstein-1 distance
Wenatchee, WA MSA (2 counties)
Washington
20
23
26
29
32
35
38
41
44
47
Epiweek

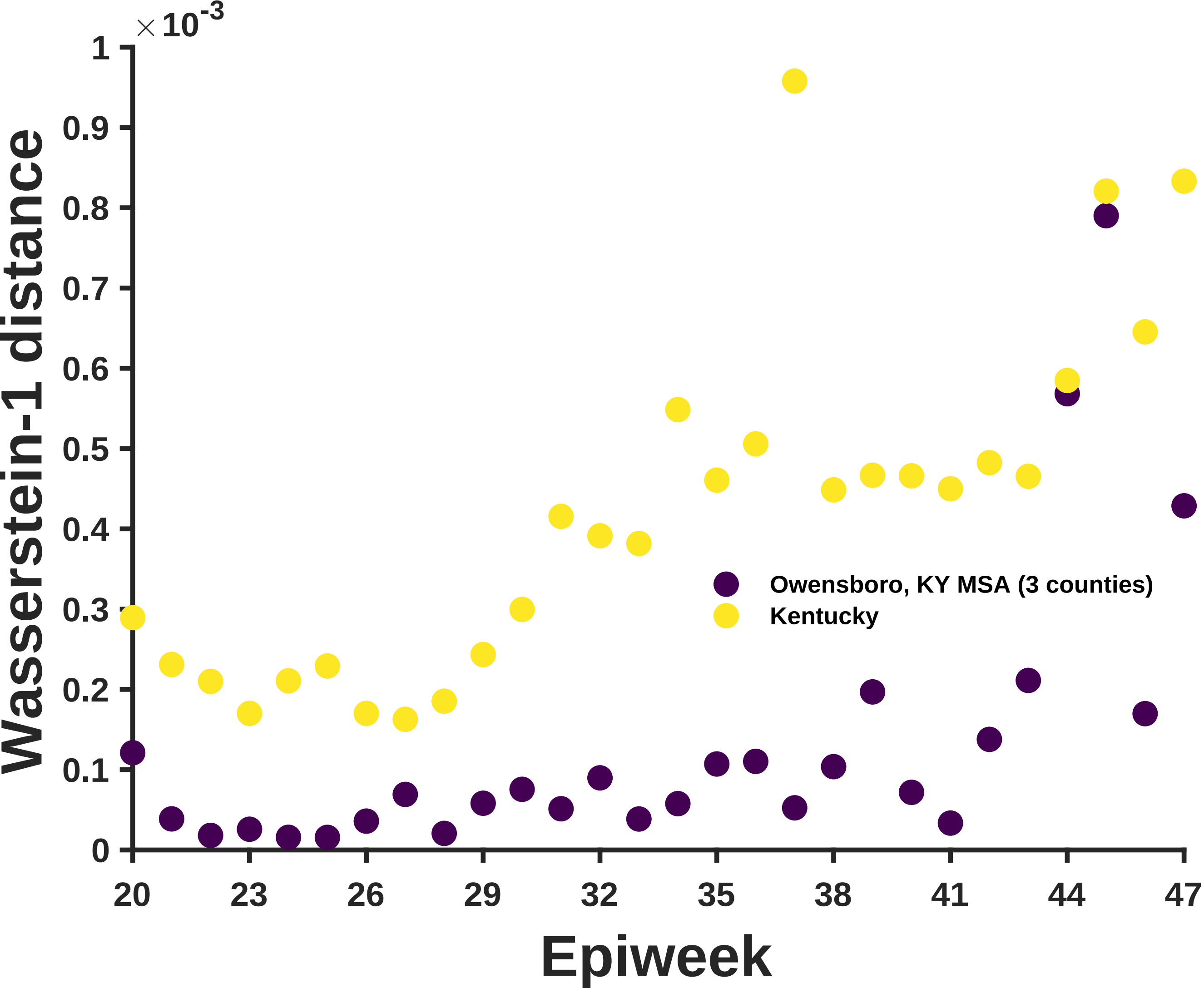

×10^-3
1
0.9
0.8
0.7
0.6
0.5
0.4
0.3
0.2
0.1
0
Wasserstein-1 distance
20
23
26
29
32
35
38
41
44
47
Epiweek
Owensboro, KY MSA (3 counties)
Kentucky

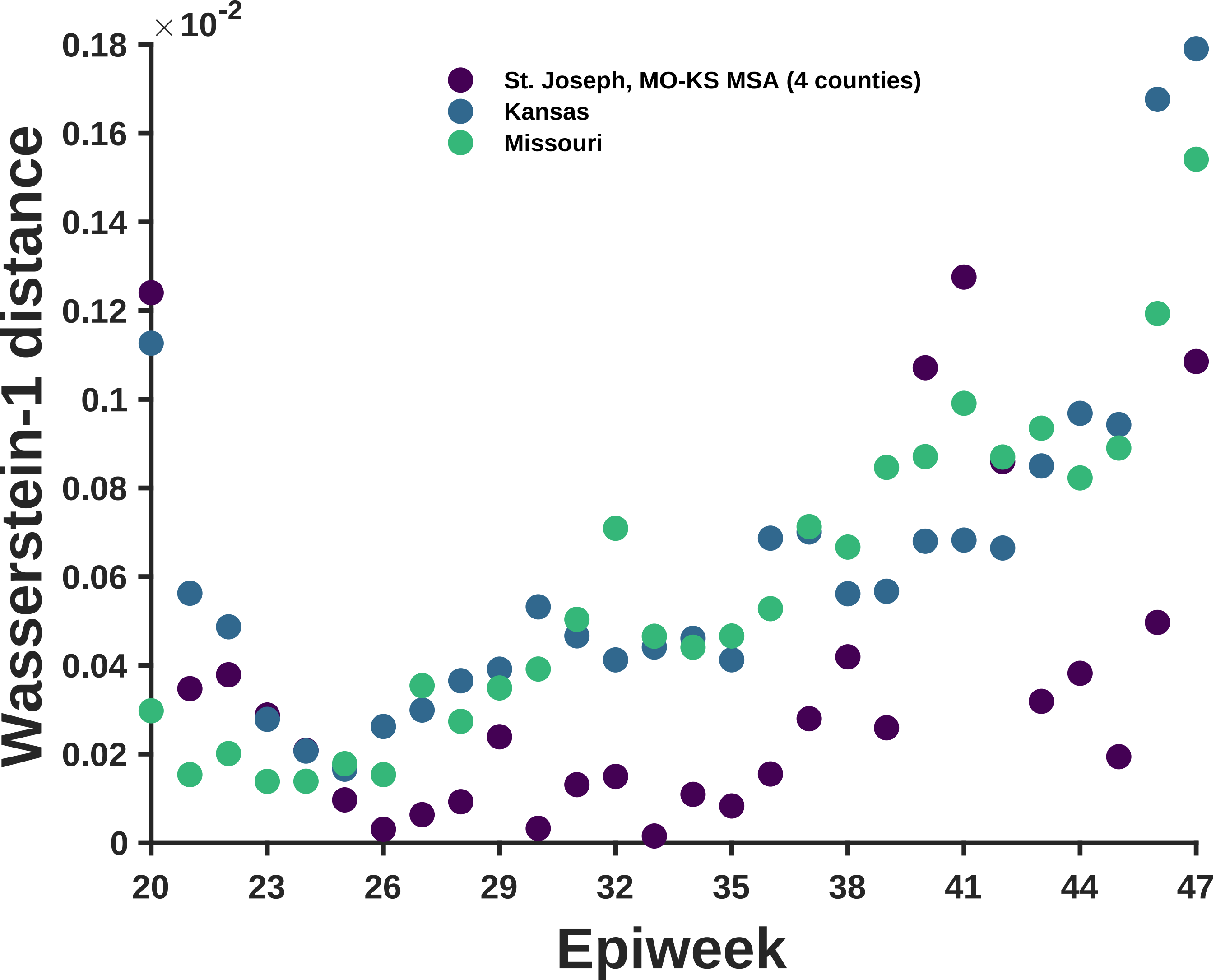

×10^-2
0.18
0.16
0.14
0.12
0.1
0.08
0.06
0.04
0.02
0
Wasserstein-1 distance
20
23
26
29
32
35
38
41
44
47
Epiweek
St. Joseph, MO-KS MSA (4 counties)
Kansas
Missouri

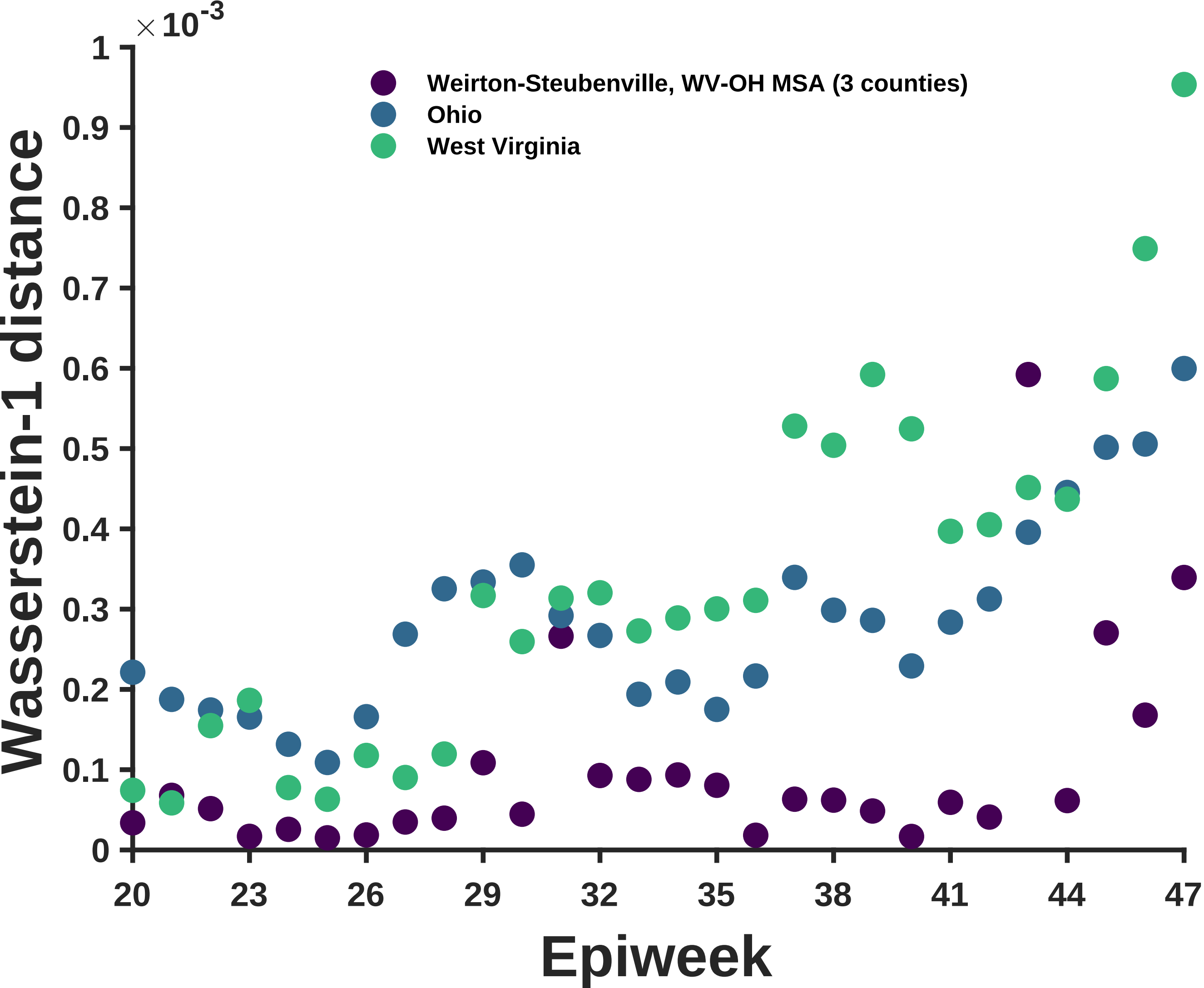

×10^-3
Weirton-Steubenville, WV-OH MSA (3 counties)
Ohio
West Virginia
Wasserstein-1 distance
Epiweek
0
0.1
0.2
0.3
0.4
0.5
0.6
0.7
0.8
0.9
1
20
23
26
29
32
35
38
41
44
47

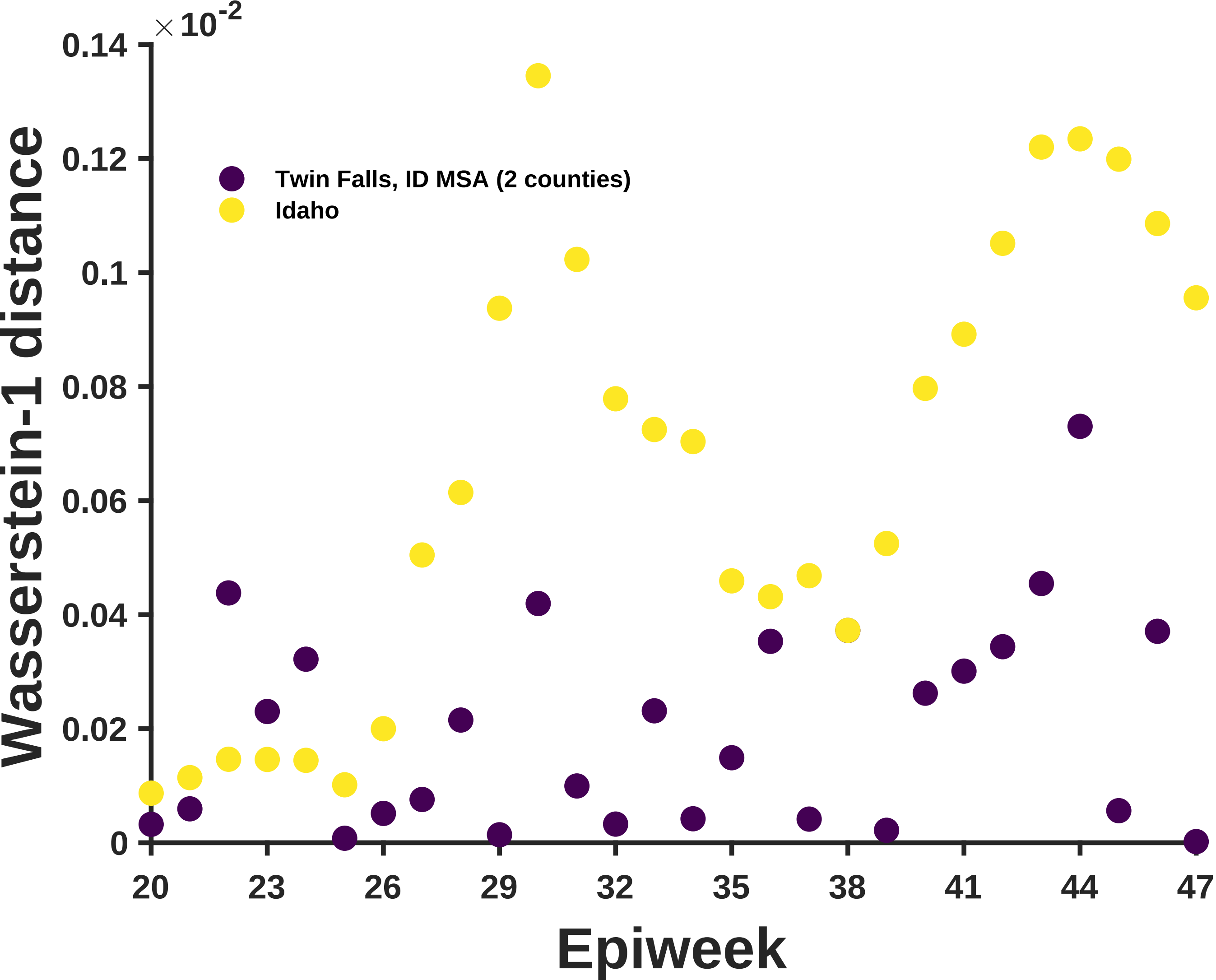

×10^-2
0.14
0.12
0.1
0.08
0.06
0.04
0.02
0
Wasserstein-1 distance
Twin Falls, ID MSA (2 counties)
Idaho
20
23
26
29
32
35
38
41
44
47
Epiweek

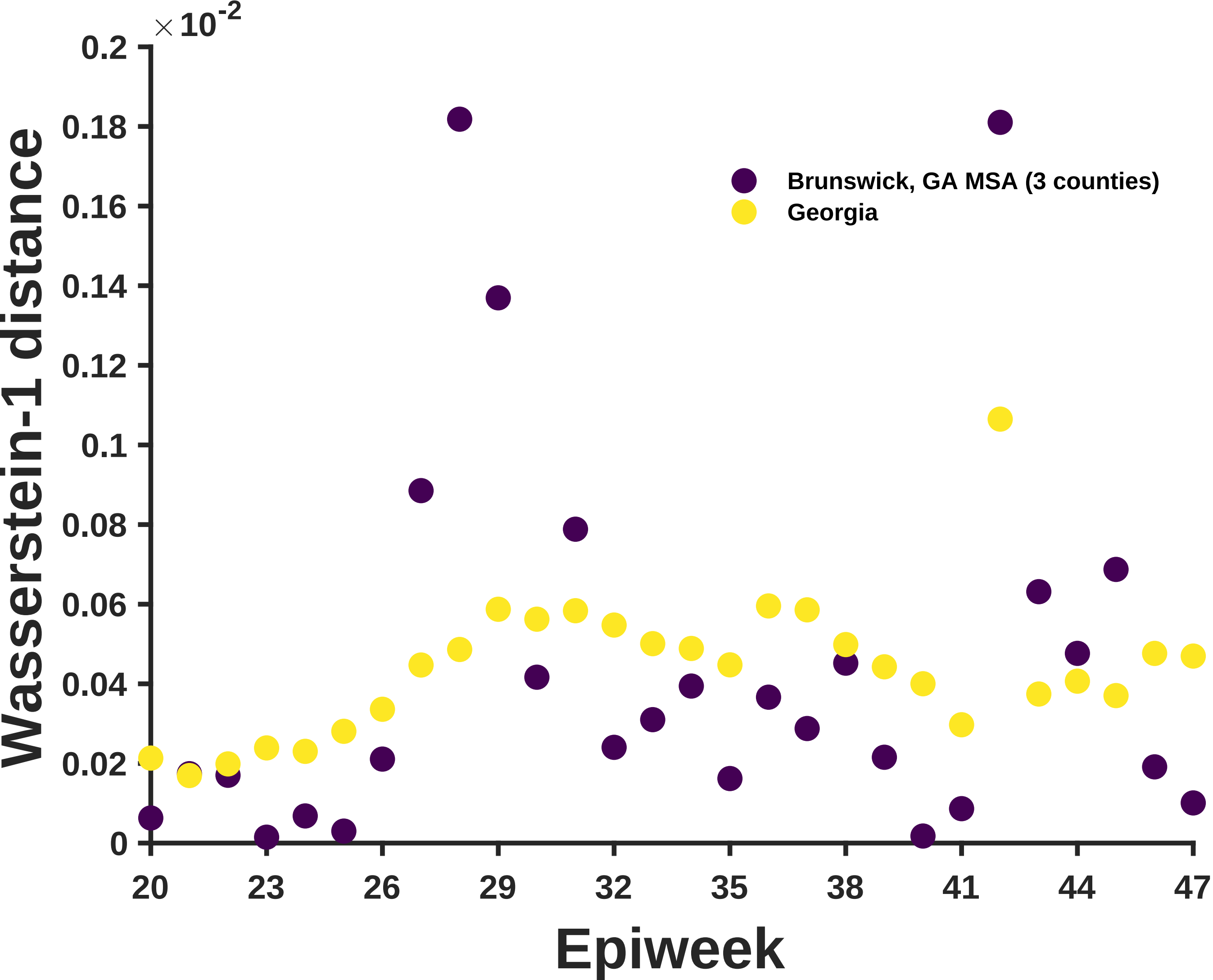

×10^-2
0.2
0.18
0.16
0.14
0.12
0.1
0.08
0.06
0.04
0.02
0
Wasserstein-1 distance
20
23
26
29
32
35
38
41
44
47
Epiweek
Brunswick, GA MSA (3 counties)
Georgia

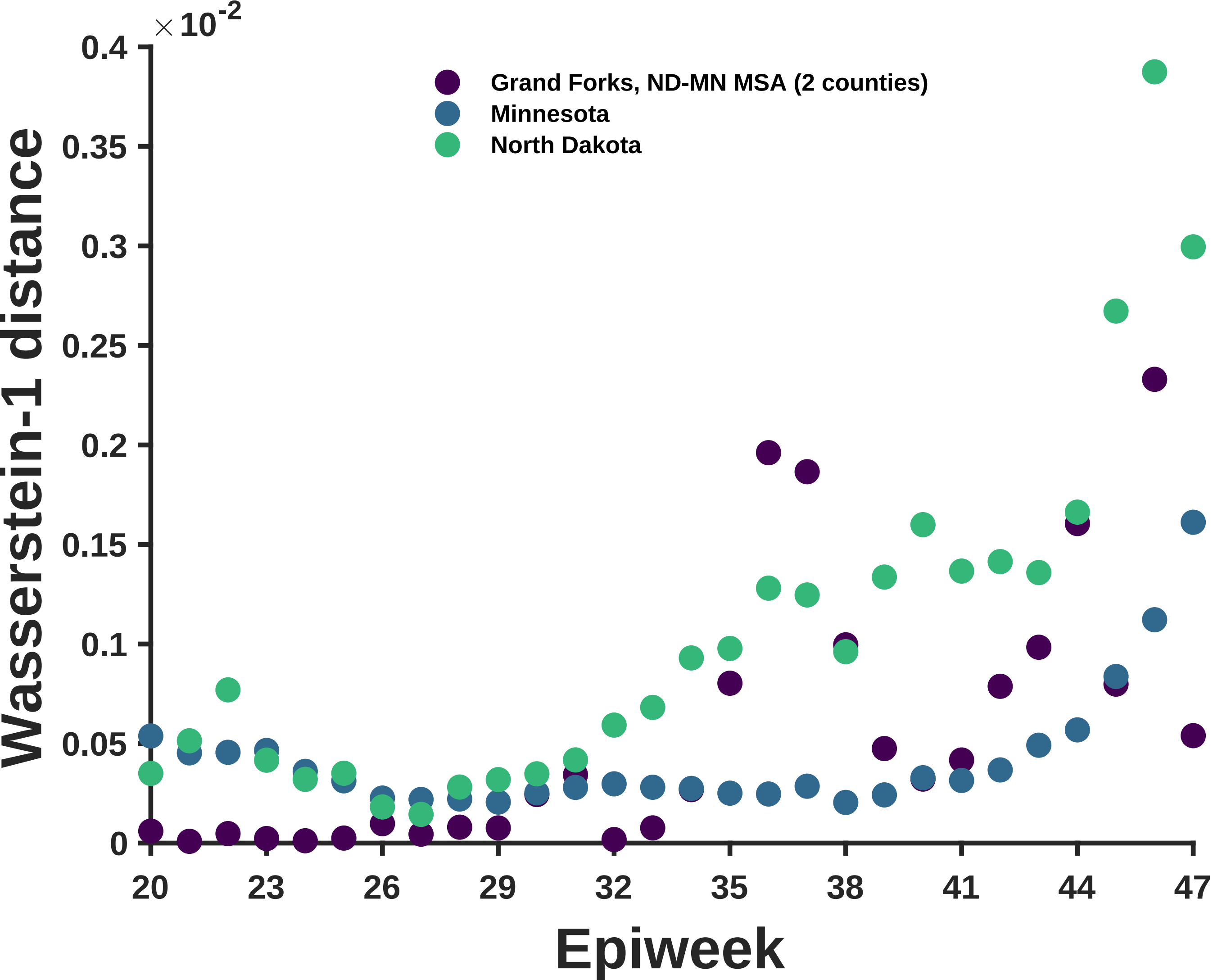

×10-2
0.4
0.35
0.3
0.25
0.2
0.15
0.1
0.05
0
Wasserstein-1 distance
Grand Forks, ND-MN MSA (2 counties)
Minnesota
North Dakota
20
23
26
29
32
35
38
41
44
47
Epiweek

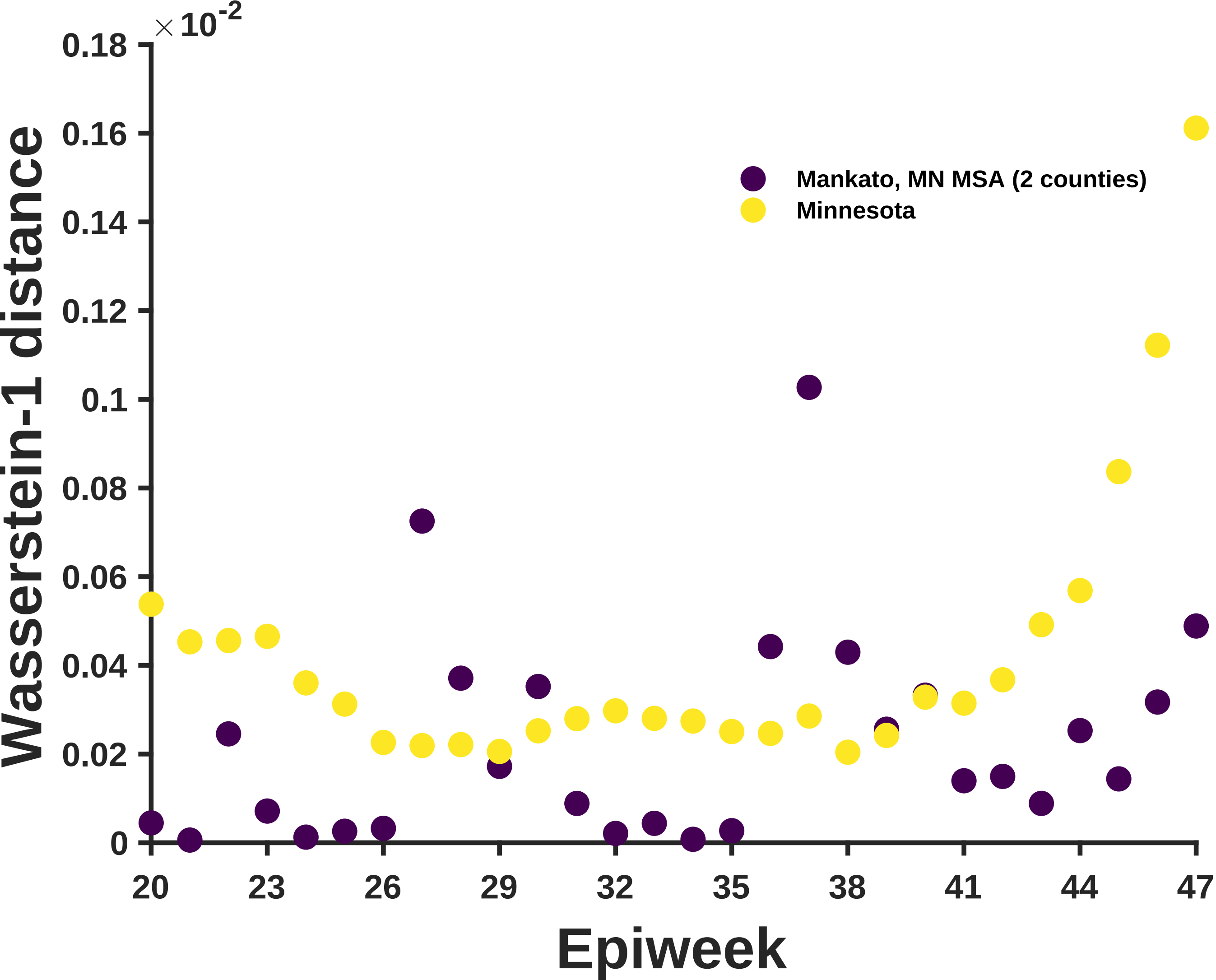

×10^-2
0.18
0.16
0.14
0.12
0.1
0.08
0.06
0.04
0.02
0
Wasserstein-1 distance
Mankato, MN MSA (2 counties)
Minnesota
20
23
26
29
32
35
38
41
44
47
Epiweek

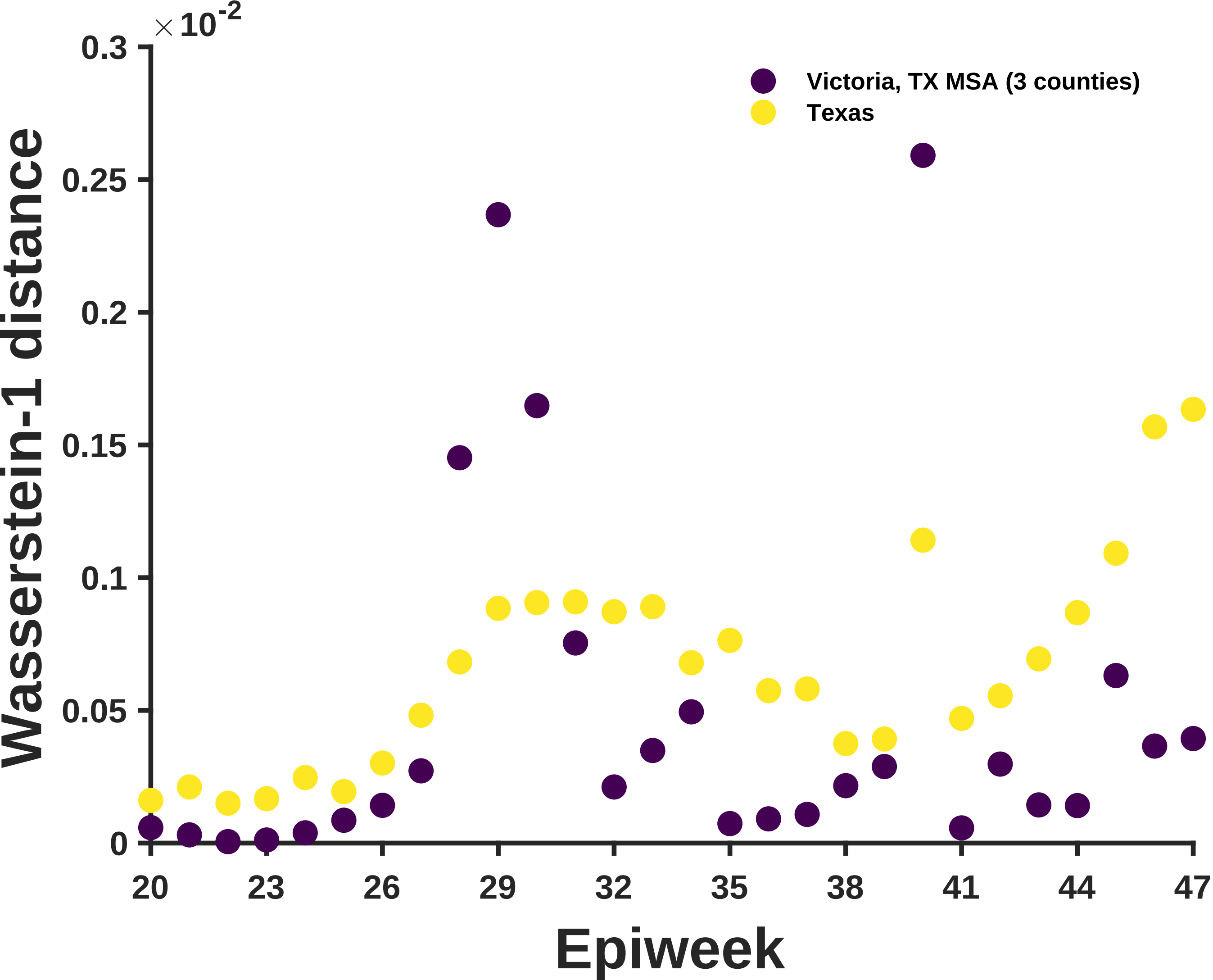

×10^-2
0.3
0.25
0.2
0.15
0.1
0.05
0
Wasserstein-1 distance
Victoria, TX MSA (3 counties)
Texas
20
23
26
29
32
35
38
41
44
47
Epiweek

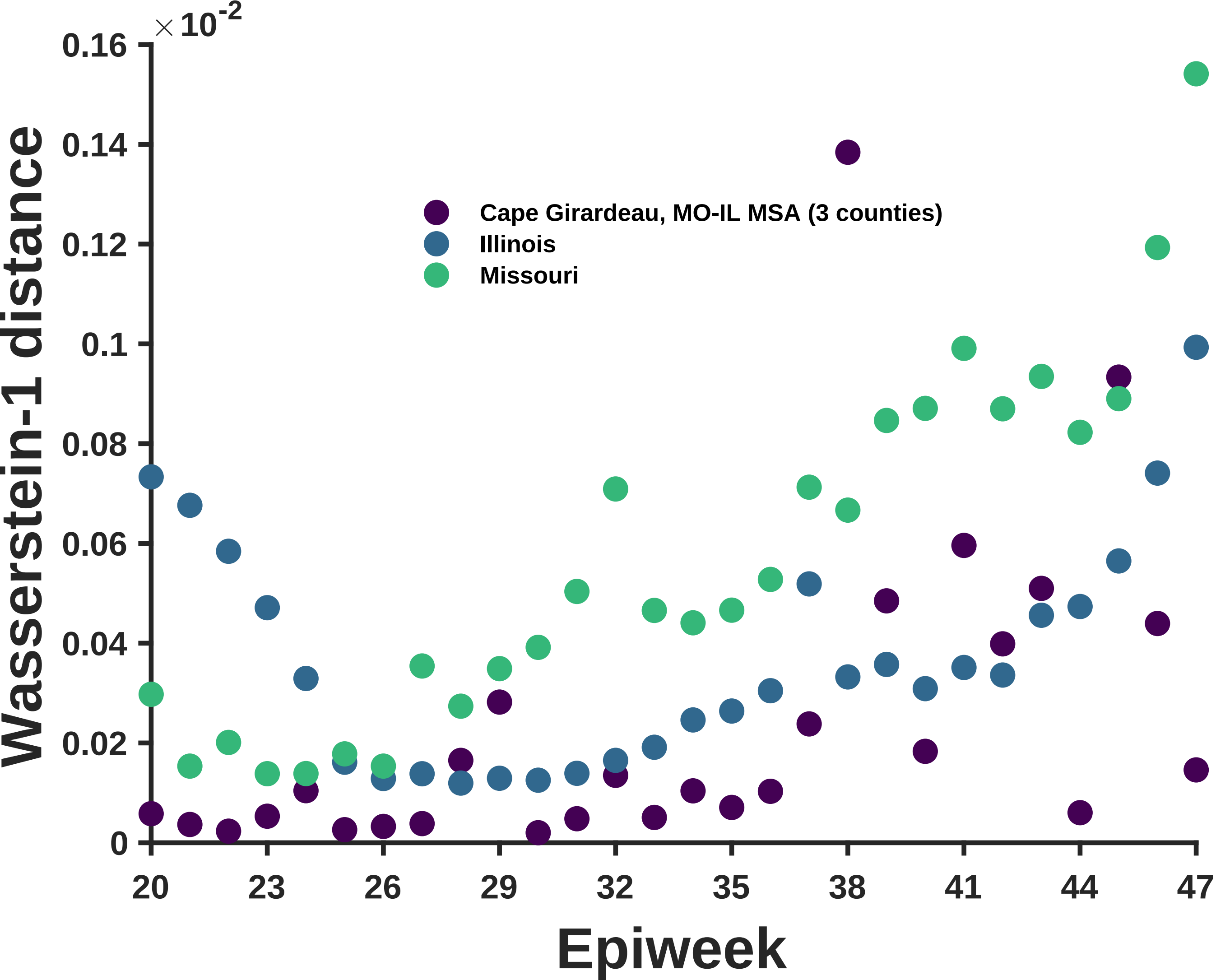

×10^-2
0.16
0.14
0.12
0.1
0.08
0.06
0.04
0.02
0
Wasserstein-1 distance
Cape Girardeau, MO-IL MSA (3 counties)
Illinois
Missouri
20
23
26
29
32
35
38
41
44
47
Epiweek

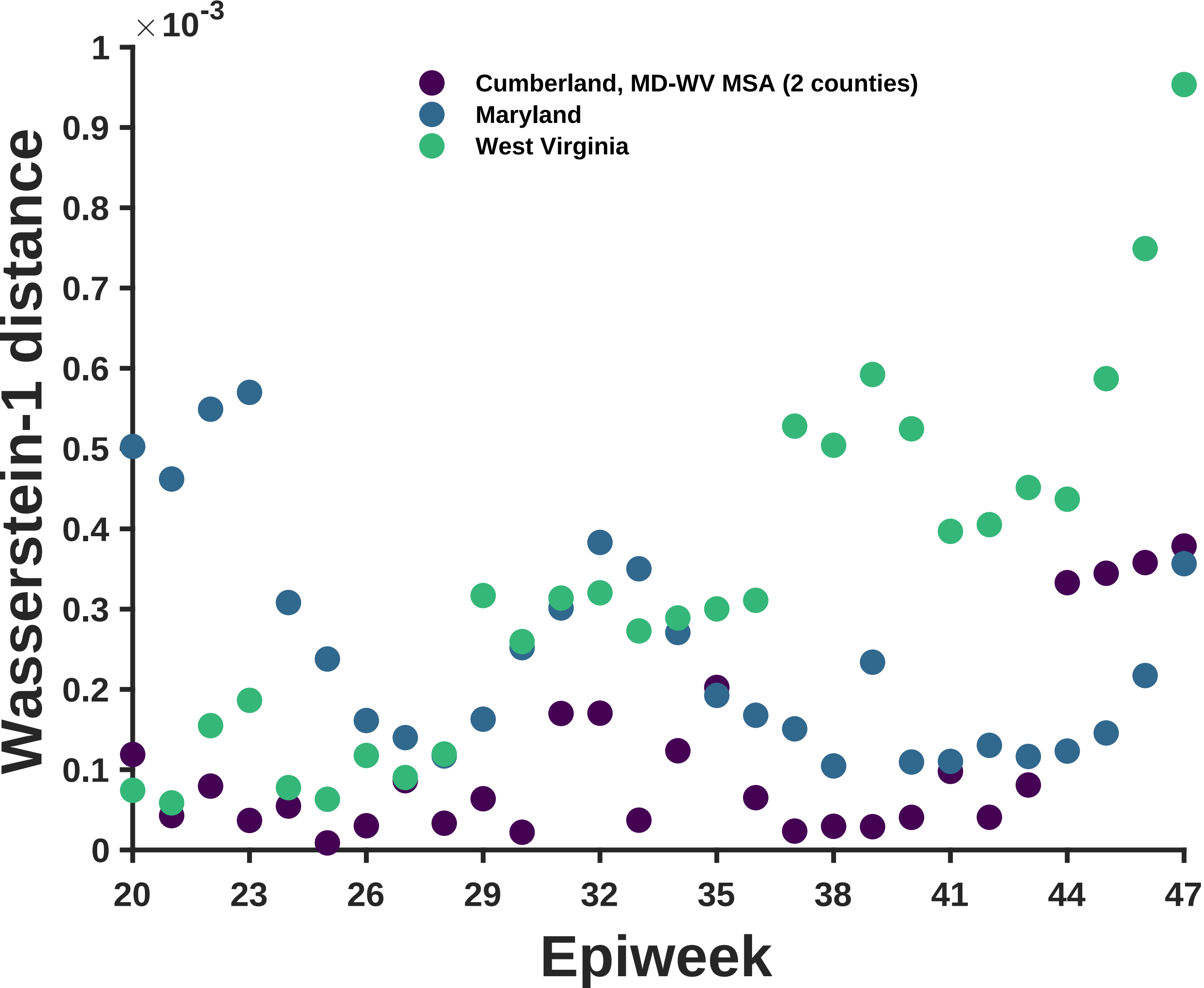
×10^-3
Cumberland, MD-WV MSA (2 counties)
Maryland
West Virginia
Wasserstein-1 distance
Epiweek
0
0.1
0.2
0.3
0.4
0.5
0.6
0.7
0.8
0.9
1
20
23
26
29
32
35
38
41
44
47

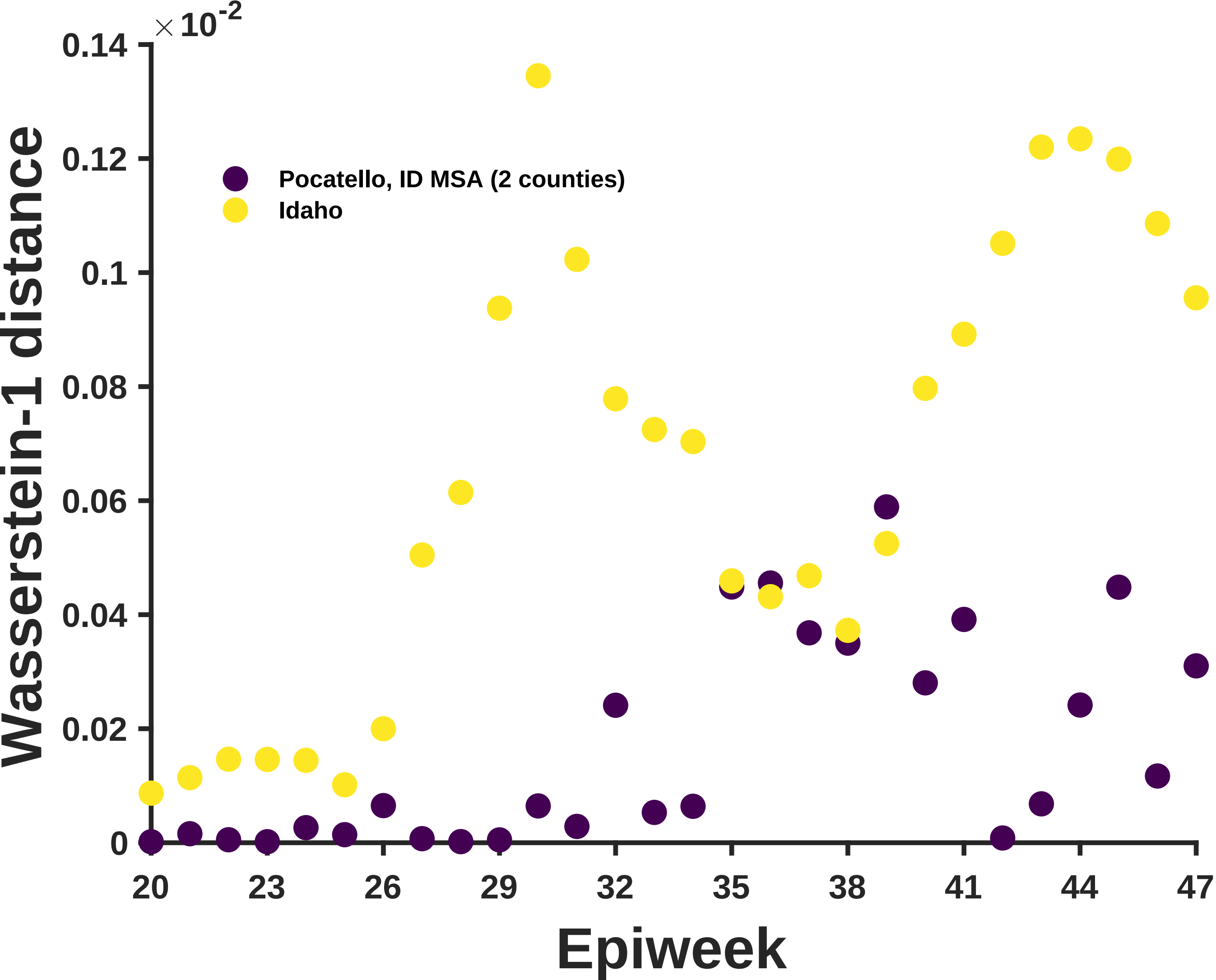

×10^-2
0.14
0.12
0.1
0.08
0.06
0.04
0.02
0
Wasserstein-1 distance
Pocatello, ID MSA (2 counties)
Idaho
20
23
26
29
32
35
38
41
44
47
Epiweek

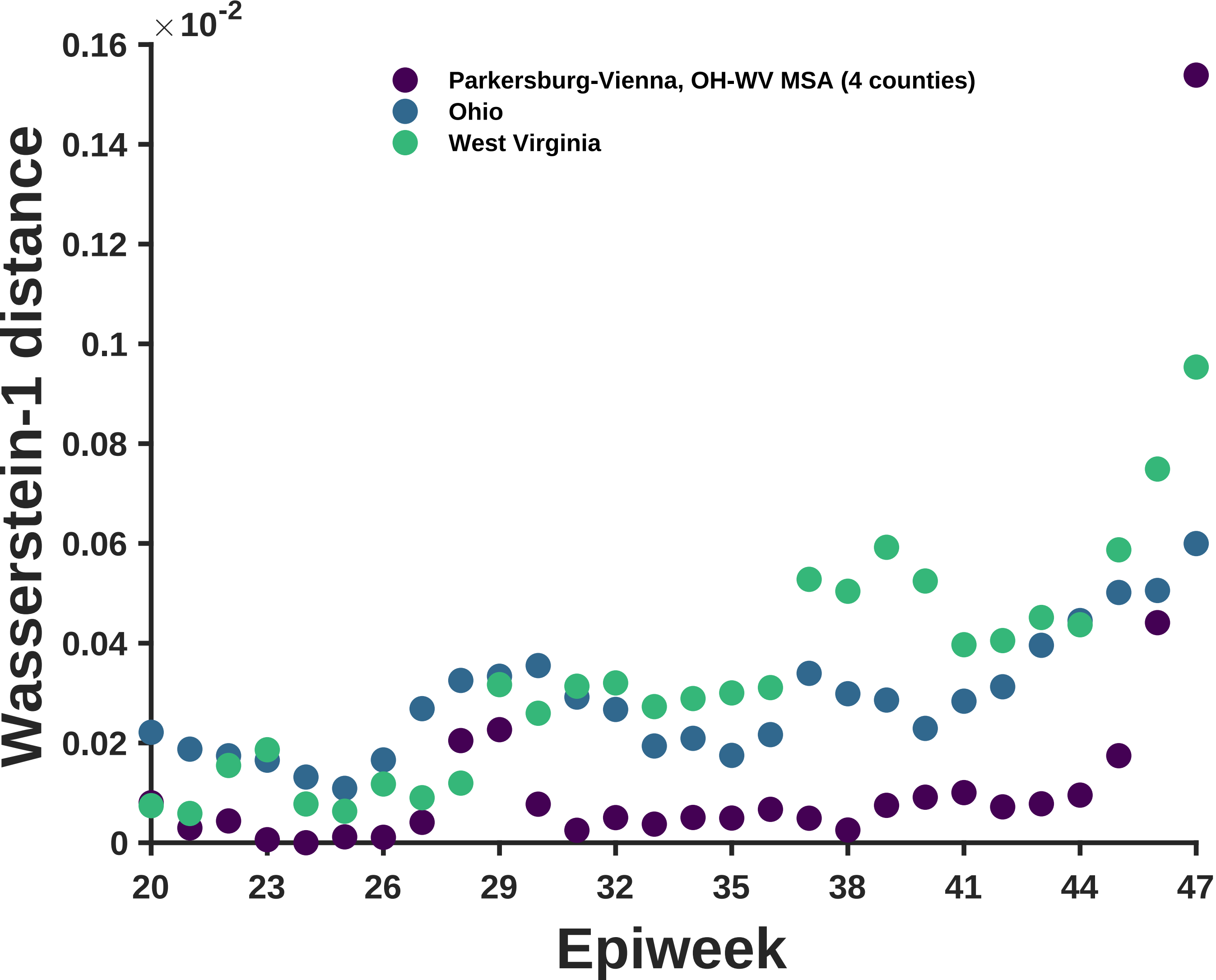
×10^-2
0.16
0.14
0.12
0.1
0.08
0.06
0.04
0.02
0
Wasserstein-1 distance
Parkersburg-Vienna, OH-WV MSA (4 counties)
Ohio
West Virginia
20
23
26
29
32
35
38
41
44
47
Epiweek

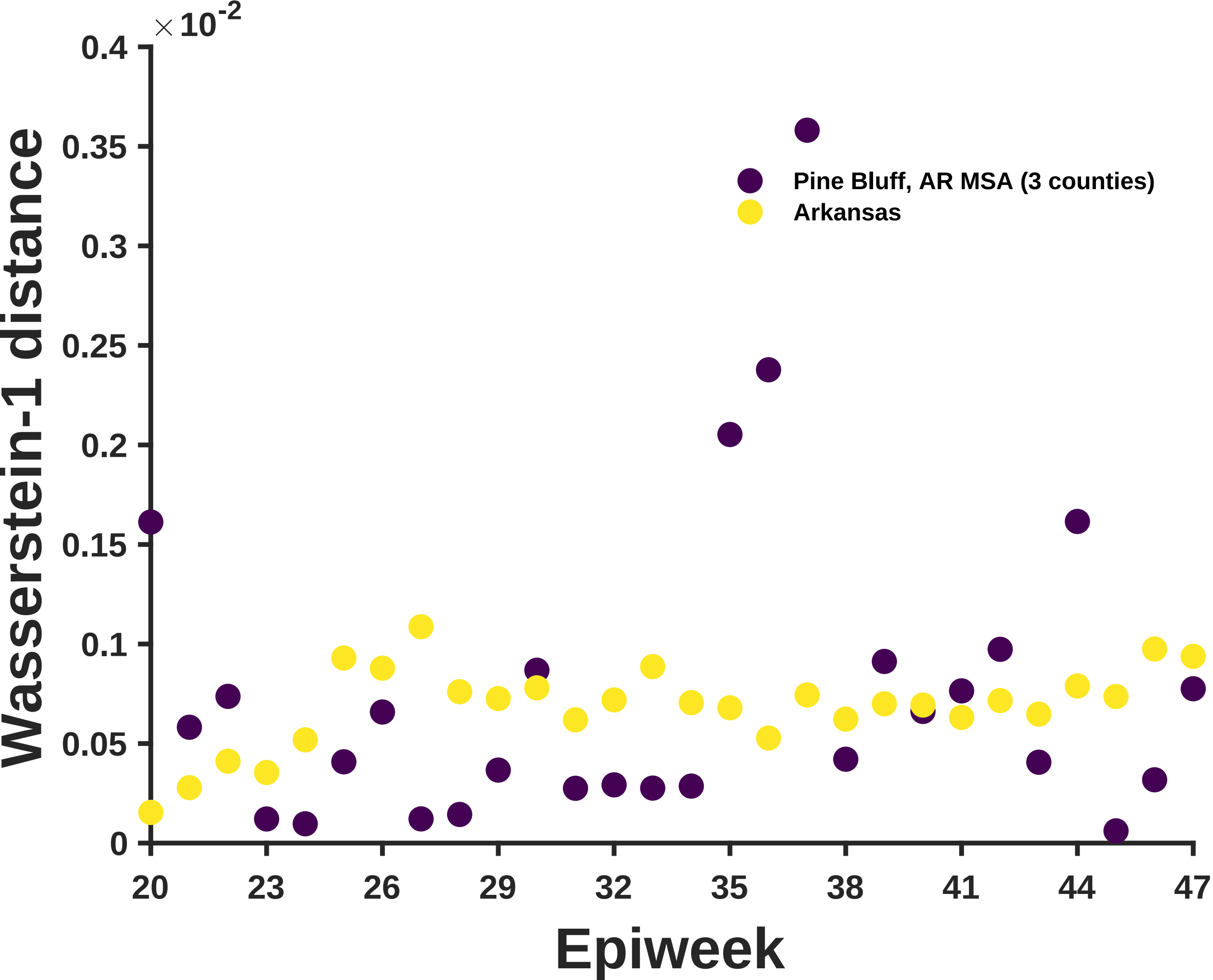
×10^-2
0.4
0.35
0.3
0.25
0.2
0.15
0.1
0.05
0
Wasserstein-1 distance
Pine Bluff, AR MSA (3 counties)
Arkansas
20
23
26
29
32
35
38
41
44
47
Epiweek

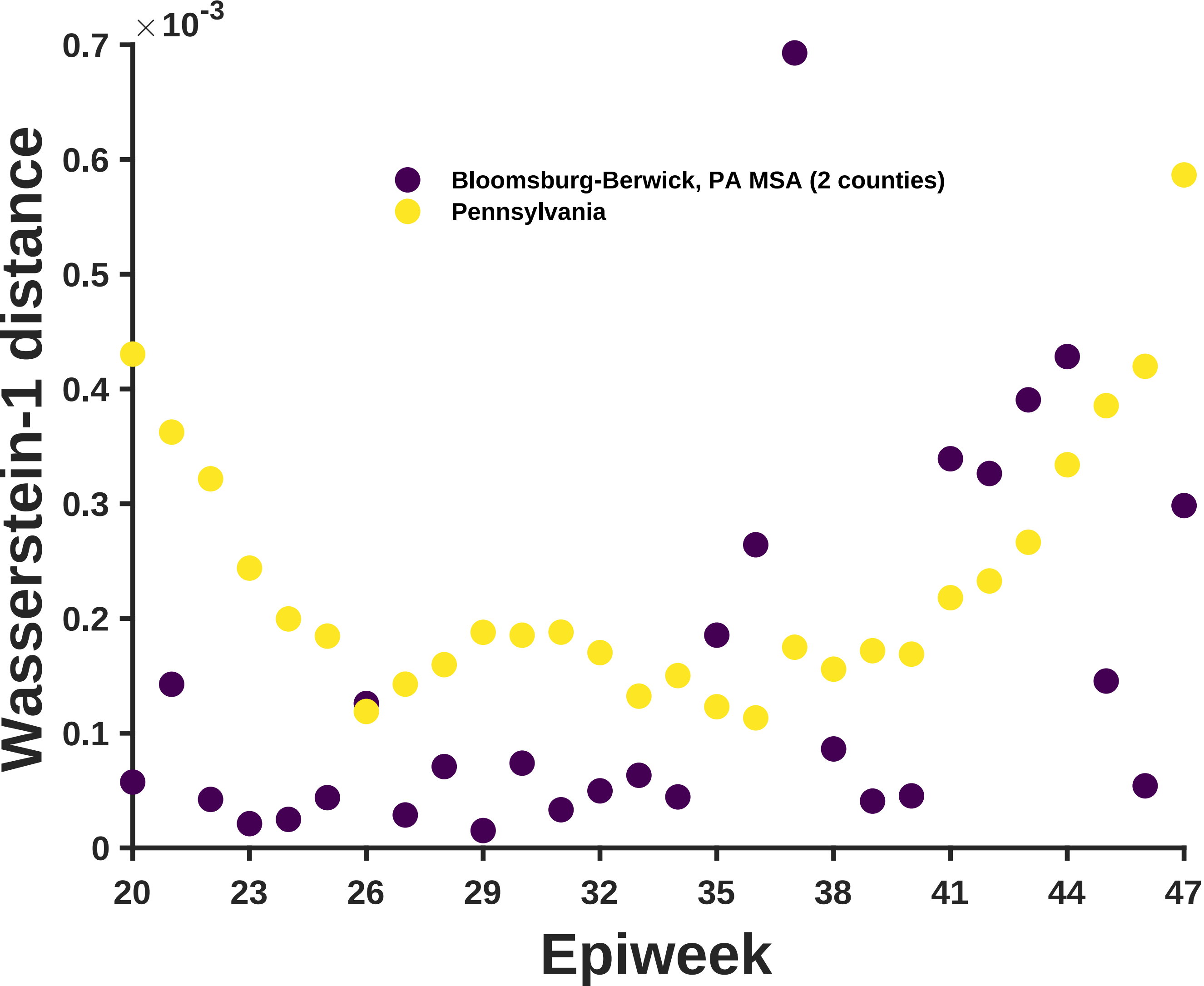

×10^-3
0.7
0.6
0.5
0.4
0.3
0.2
0.1
0
Wasserstein-1 distance
Bloomsburg-Berwick, PA MSA (2 counties)
Pennsylvania
20
23
26
29
32
35
38
41
44
47
Epiweek

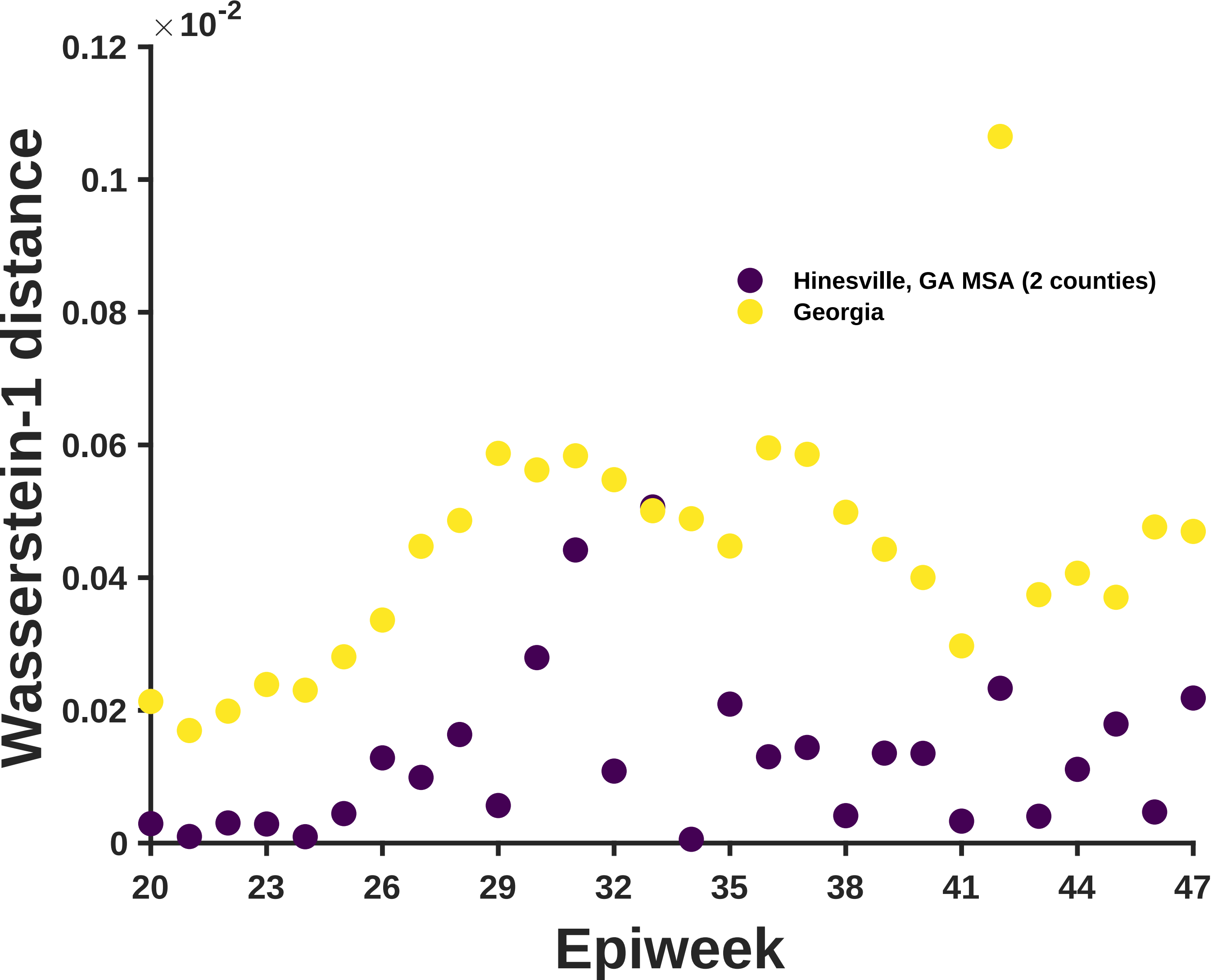

×10^-2
0.12
0.1
0.08
0.06
0.04
0.02
0
Wasserstein-1 distance
Hinesville, GA MSA (2 counties)
Georgia
20
23
26
29
32
35
38
41
44
47
Epiweek

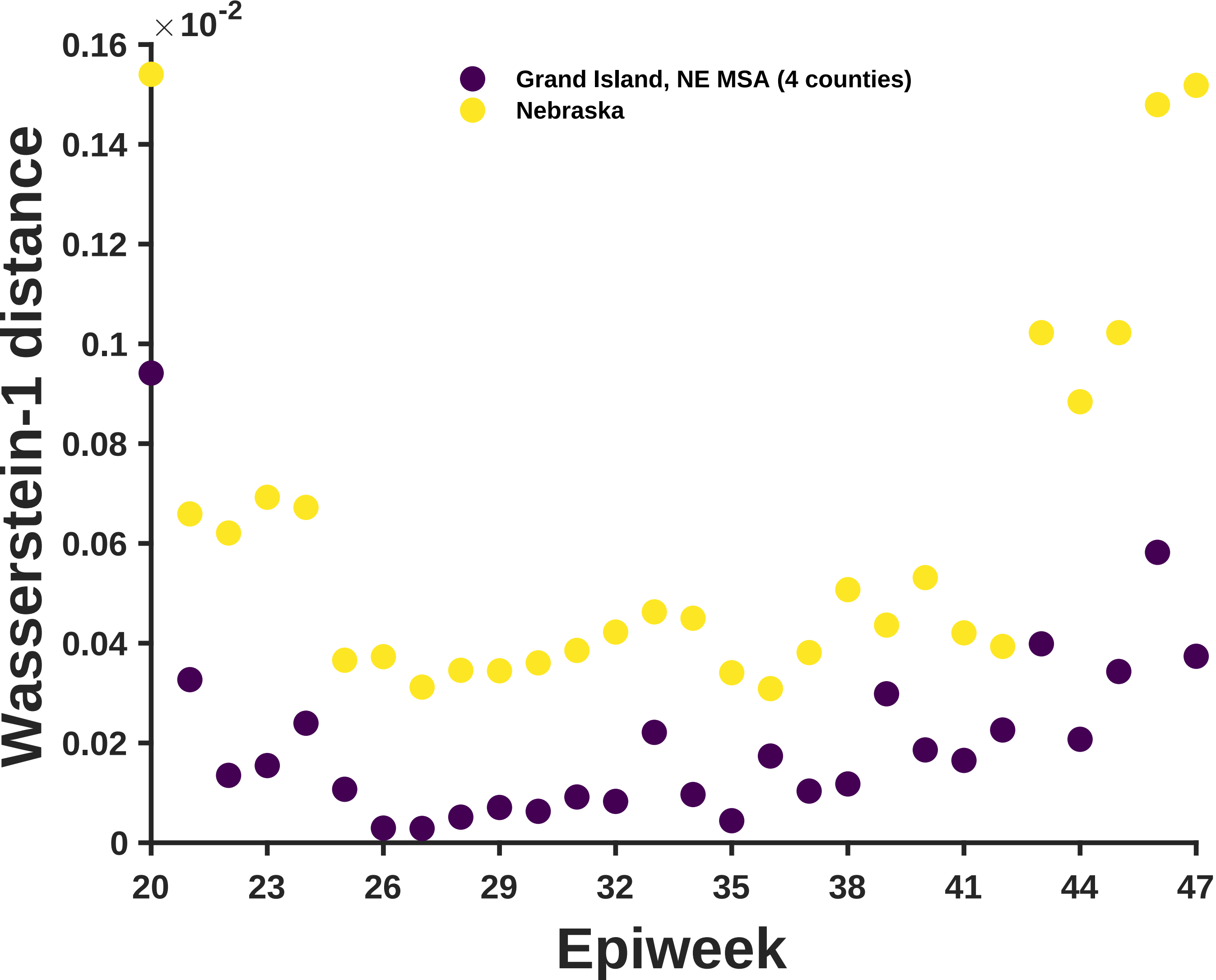

×10^-2
0.16
0.14
0.12
0.1
0.08
0.06
0.04
0.02
0
Wasserstein-1 distance
Grand Island, NE MSA (4 counties)
Nebraska
20
23
26
29
32
35
38
41
44
47
Epiweek

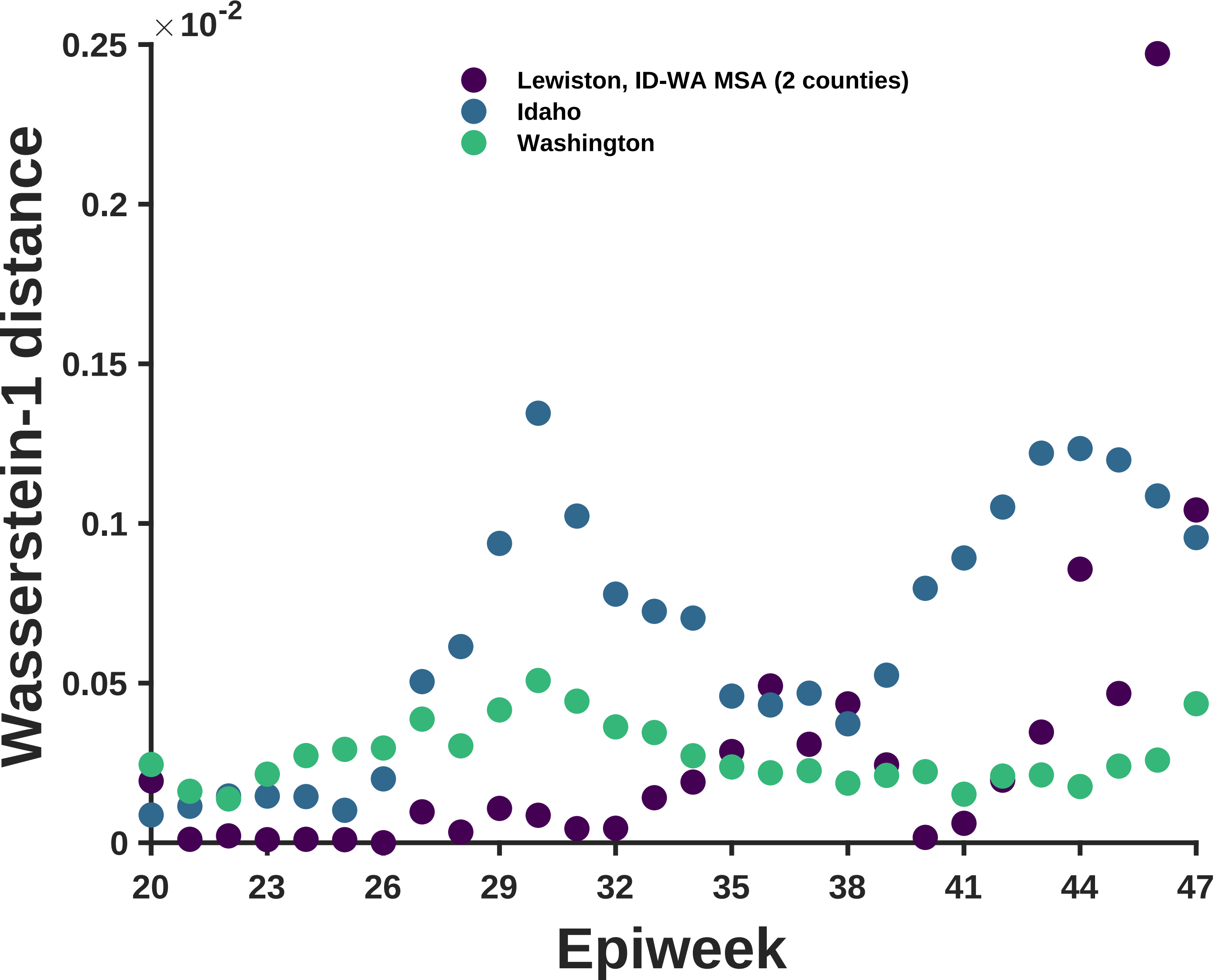

×10^-2
0.25
0.2
0.15
0.1
0.05
0
Wasserstein-1 distance
Lewiston, ID-WA MSA (2 counties)
Idaho
Washington
20
23
26
29
32
35
38
41
44
47
Epiweek
